# Revised estimates of the types and durations of long Covid symptoms based on claims records from 245 Million US patients

**DOI:** 10.64898/2026.02.17.26346448

**Authors:** Hamed Nilforoshan, Julia Reisler, Erfan Jahanparast, Michael Moor, Steven N. Goodman, Stefan Wager, Jure Leskovec

## Abstract

COVID-19 has been shown to cause a range of harmful long-term effects on nearly every organ system^1–3^. These findings are based on retrospective studies comparing COVID-19 patients to patients with similar medical histories and demographics but no COVID-19 diagnosis^4–16^. However, concerns have emerged that these comparisons may be biased if COVID-19 patients had unrelated health conditions or other factors not recorded in their medical records^17–21^. Here, using a massive dataset of 14.4 billion health insurance claims from 244.7 million U.S. patients, we find that the large majority of long-term effects attributed to COVID-19 by methods used in conventional studies are likely due to bias from selective testing. This bias arises because individuals with non-COVID health conditions producing long-term symptoms were more likely to seek care and be tested for COVID-19. As a result, their non-COVID symptoms are attributed to COVID-19. We develop a study design that reduces this bias by only considering individuals who have taken a COVID-19 PCR test, and then comparing similar patients whose first test came back positive vs. negative. This way the COVID-19 patients and control group are more similar and non-COVID factors play less of a role. We examine 614 clinical outcomes over a 2-year followup period and reveal an order of magnitude smaller—but still clinically significant—number of long-term effects attributed to COVID-19 that persist for up to one year after infection. We confirm that the long-term effects of COVID-19 span many organ systems, including respiratory, cardiovascular, musculoskeletal, and integumentary systems, but are significantly narrower in scope and duration than previously believed. Although some symptoms exist more than one year after COVID-19 infection, they occur at similar rates in individuals who tested negative and are therefore not attributable to COVID-19 infection. Our findings pinpoint the specific long-term effects of COVID-19 and show how large-scale data can be used to enable careful evaluation and design of population health studies.

## Introduction

Long COVID (post-acute sequelae of COVID-19, also known as PASC) describes a varied constellation of long-term adverse health effects after a severe acute respiratory syndrome coronavirus 2 (SARS-CoV-2) infection^1, 2, 22, 23^. Since the start of the global COVID-19 pandemic in early 2020, an extensive body of evidence has characterized long COVID as a multi-system disease with over 200 symptoms^4–6, 8, 9, 11, 12, 15, 24–30^. Long COVID manifests as both persistent symptoms after initial infection^6, 31^ and newonset symptoms^24, 32^, affecting almost all organ systems. New-onset long COVID symptoms include neurocognitive^12, 16, 33^, reproductive^5^, cardiovascular^9, 34–36^, and gastrointestinal effects^14^.

All studies of long COVID symptoms are observational and, therefore, susceptible to bias. These studies typically use electronic health records to compare individuals that were diagnosed with COVID-19 via testing to similar individuals without a confirmed COVID-19 diagnosis^6, 13, 24, 29, 32, 37–39^. This retrospective approach can be biased if there are differences between people with and without a COVID-19 diagnosis that are not recorded in the data^20^. For instance, individuals who receive a COVID-19 diagnosis may have actually been tested while seeking care for a non-COVID condition (e.g., diabetes). If non-COVID conditions are not well recorded in the patient’s medical history, symptoms resulting from these conditions could be falsely attributed to COVID-19. Another source of bias is careseeking behavior: individuals who frequently test for COVID-19 may also be more likely to seek medical care for non-COVID conditions. Overall, these non-COVID factors could cause COVID-19 patients to appear less healthy over time, independent of the effects of COVID-19 itself. Additionally, prior studies characterizing long COVID symptoms have reported mixed findings^4–6, 8, 9, 11–13, 15, 23–30, 32, 37–42^, partly due to methodological limitations such as small sample sizes, geographic variation, and differing study populations. Therefore, research with large, nationally representative samples is needed, as is thorough investigation into potential sources of bias inherent in conventional studies.

In this paper, we disentangle genuine long COVID symptoms from those mistakenly attributed to the disease. To achieve this, we use a prospective study design that is less subject to bias. Specifically, we enroll individuals at the time of their first recorded COVID-19 PCR test, comparing those who test positive (cases) with those who test negative (controls). We applied this design to evaluate 614 health outcomes over a two-year follow-up period. Compared to a conventional analysis using the same data, our test-based prospective design identifies substantially fewer and shorter-lasting long COVID effects. For instance, between days 120 and 360 postinfection, our method attributes 98.2% fewer symptoms to long COVID than a conventional analysis. Moreover, we show that conventional methods frequently overestimate the effects of long COVID, identifying implausible associations such as external injuries (e.g., firearm injuries) and congenital abnormalities in adults. These implausible associations become visible only due to the large scale and granularity of our dataset. Our proposed study design substantially reduces this bias: on a set of 49 outcomes not plausibly related to COVID-19, we cut the number of falsely identified associations from 53.1% down to 4.1%. This improvement allows us to confidently isolate a smaller but clinically meaningful set of long COVID effects persisting up to one year after infection. Finally, we demonstrate how selective COVID-19 testing, where individuals with non-COVID health conditions and related factors (e.g., a higher tendency to seek medical care) are more likely to be tested, explains why conventional methods are prone to detecting spurious associations.

## Results

To identify symptoms attributable to long COVID, we designed a study that corrects two key biases common in prior research (Figure 1a). First, conventional studies typically compare COVID-19 cases—who were all enrolled during a healthcare visit—with controls who neither tested nor visited a healthcare center at the time. This means that future health problems in the COVID-19 cases may be attributed to the COVID-19 infection, when they are actually due to non-COVID factors that prompted the healthcare visit and testing. Second, many conventional studies identify controls retrospectively, by selecting individuals who were never diagnosed with COVID-19. This allows future information about who remained uninfected to influence the selection of controls^43^.

**Figure 1:**
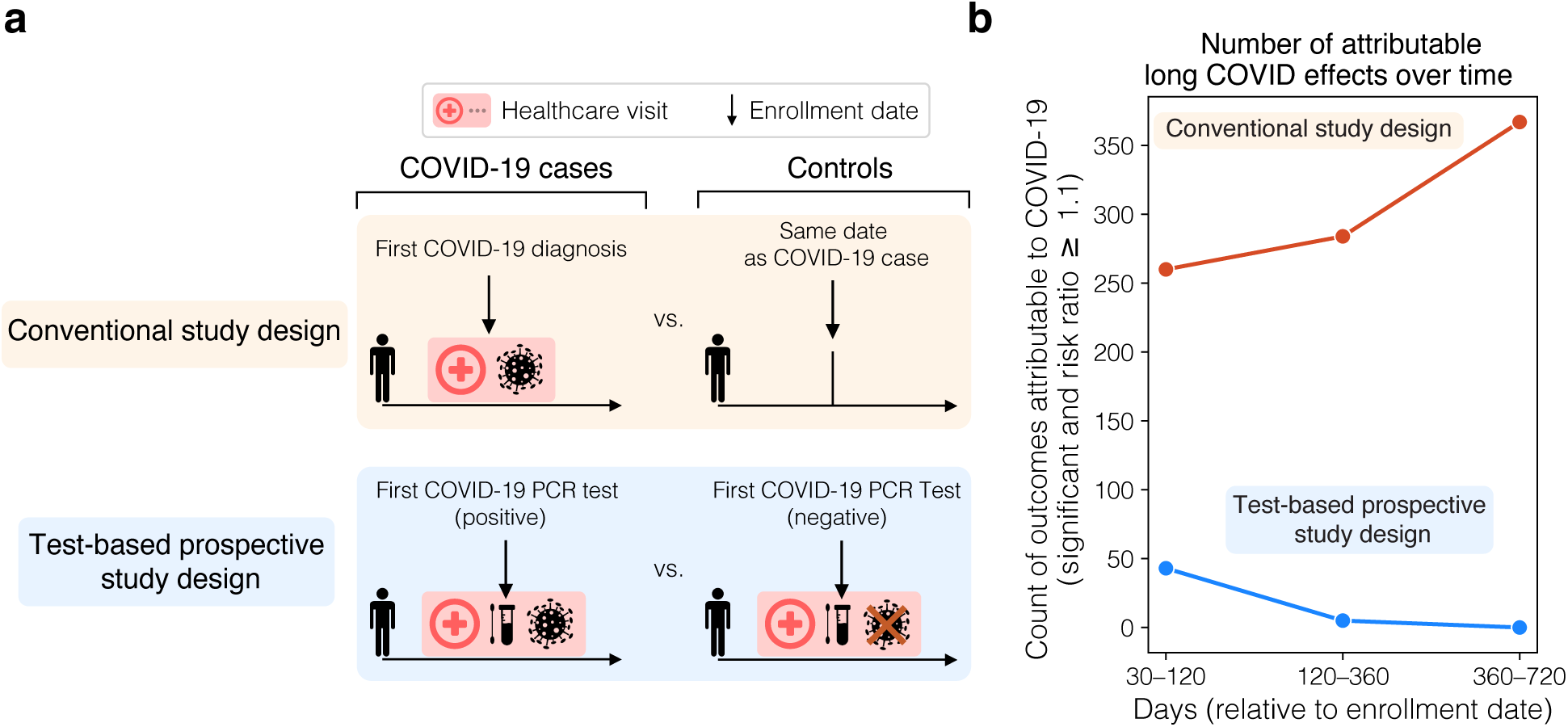
Test-based prospective design reduces the number of attributable long COVID effects by an order of magnitude. **(a)** *Top:* Conventional study designs (orange) compare individuals diagnosed with COVID-19 (cases) to matched individuals without a COVID-19 diagnosis but with similar medical histories and demographics (controls). This approach does not account for why individuals were having a healthcare visit. Since all COVID-19 cases are enrolled during a healthcare visit, any unrelated health conditions and other non-COVID factors that prompted the visit and COVID-19 testing^17–21^ may be attributed to the COVID-19 infection. In contrast, the control group is a set of random people, the large majority of whom were not visiting a healthcare center. As a result, COVID-19 cases are systematically more prone to exhibit future health problems due to non-COVID factors, despite having identical medical histories and demographics. Conventional designs may therefore attribute future health problems to COVID-19 infection, when these outcomes instead arise from non-COVID factors that led to the initial healthcare visit. *Bottom:* The test-based prospective design (blue) mitigates this bias by enrolling only individuals who had a healthcare visit and received their first COVID-19 PCR test. This design compares individuals with a positive COVID-19 PCR test result (cases) to matched individuals who also took a COVID-19 PCR test but received a negative test result (controls), and who again have similar medical histories and demographics. Now, the control group is much more similar to the COVID-19 cases, and thus the findings are less biased. **(b)** The test-based study design (blue) detects fewer long COVID symptoms (risk ratio ≥ 1.1 and p*<*0.05 with Bonferroni correction for multiple comparisons; Methods M3) with shorter duration compared to a conventional analysis of the same data (orange). Out of a set of 614 considered outcomes, the test-based study attributes 83.5% fewer outcomes to COVID-19 at 30–120 days than the conventional study design, and 98.2% fewer at 120–360 days post-infection. At 360–720 days, the test-based study finds no attributable outcomes, while the conventional study reported 369.

We address both sources of bias using a testing-based prospective design, where COVID-19 cases are compared to controls who also had a healthcare visit and received a COVID-19 test but tested negative. First, because our controls also visited a healthcare center and took a COVID-19 test, they are more comparable to the COVID-19 cases. Second, the fact that we enroll all participants at the time of their first PCR test—before knowing whether they would test positive or negative—ensures that our study design is not biased from future information about testing or patient outcomes.

To evaluate the impact of our proposed study design, we compare its results to a conventional analysis of the same dataset. For both analyses, we reweight the control group so that its medical history and demographics matches those of the COVID-19 cases (Methods M2), thereby reducing confounding^44, 45^ and collider bias^17^. Specifically, we use a synthetic control approach^46, 47^ that allows us to match medical histories over time (Methods M3; Methods Figure 1).

### Number and duration of long COVID symptoms

Our proposed study design detects significantly fewer long COVID symptoms—persisting for a shorter duration—than methods used in existing studies of long COVID (Figure 1b). Of the 614 health outcomes analyzed over two years, we find significant and harmful long-term effects of COVID-19 for only 43 outcomes on days 30-120 after infection, 5 outcomes on days 120-360 after infection, and no outcomes 360-720 days after infection (p*<*0.05 with Bonferroni correction for multiple comparisons, and risk ratio ≥ 1.1; Methods M3). By contrast, a conventional retrospective study of the same data, comparing COVID-19 patients to uninfected controls with similar medical histories, finds significant and harmful effects for 262, 286, and 369 outcomes on days 30-120, 120-360, and 360-720 respectively. These findings demonstrate that analyses of long COVID are highly sensitive to study design, and that there may be fewer symptoms of long COVID than previously thought^2, 4^. More-over, the conventional analysis, consistent with prior studies^48, 49^, suggests that there is a persistent burden of long COVID symptoms up to two years postinfection. However, our findings instead indicate a return to baseline health within approximately one year.

### Comparing the bias of our study versus conventional methods

We evaluate the susceptibility of methods used in existing studies of long COVID to detecting spurious symptoms—and whether our proposed study design addresses this issue (Figure 2). We do so by ruling out a causal effect of COVID on “negative control outcomes”. These are health outcomes that are highly unlikely to have been caused by COVID-19, such as congenital disorders (e.g., cerebral palsy in adults), or external events (e.g., firearm injuries). We regard any relationship between COVID-19 and these outcomes as due to design bias, and the magnitude of these estimated relationships allows us to compare the bias of different study designs. Our unprecedented data scale (Extended Data Figure 2) allowed us to analyze 49 negative control outcomes, many of which are too rare to be studied with smaller datasets.

**Figure 2:**
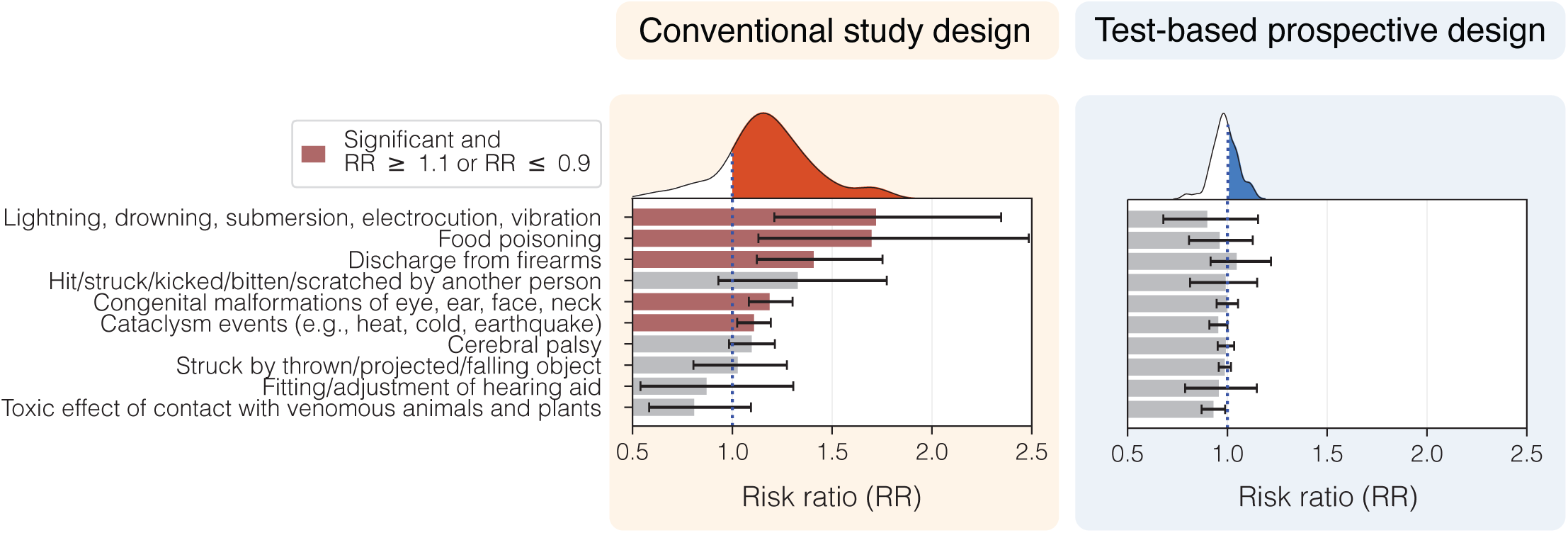
Test-based prospective design corrects for overestimation of effects found in conventional studies. We investigate outcomes not expected to be caused by COVID-19 infection (e.g., firearm injuries, congenital conditions in adults). A conventional analysis, where COVID-19 cases are compared to random individuals, suggests that COVID-19 causes these implausible effects (left panel). For instance, the conventional analysis shows that COVID-19 infection increases the likelihood of injury due to lightning, drowning, submersion, electrocution or vibration by 76% on days 30-120 post-infection (p*<*0.05). By contrast, our test-based study design corrects for this bias and does not detect such an effect (right panel). This example is one of 49 “negative control” outcomes that we examine—events that COVID-19 could not plausibly cause (10 shown here; full set in Supplementary Figures S45-47). These serve as a benchmark to measure bias. Overall, the conventional analysis shows strong positive bias: 53.1% of these outcomes are falsely detected as significant with risk ratios ≥1.1 (p*<*0.05). By contrast, the test-based study detects only 4.1% of these outcomes as significant with risk ratios ≤0.9 (p*<*0.05), suggesting only mild negative bias (Discussion). Conventional methods thus over-attribute effects to COVID-19, whereas the test-based design yields more reliable estimates. Red indicates statistically significant results (p*<*0.05; two-sided, unadjusted for multiple comparisons) with risk ratios that are ≥ 1.1 or ≤ 0.9. Error bars show 95% bootstrap confidence intervals (Methods M3).

We found that the conventional retrospective study, comparing COVID-19 cases to never-diagnosed controls, falsely detects associations for 53.1% of negative control outcomes, meaning the conventional study attributes positive and significant effects to COVID-19 for more than half of these negative control outcomes (RR≥1.1 and p*<*0.05; two-sided bootstrap, unadjusted for multiple comparisons). For instance, a conventional retrospective study of these data shows that COVID-19 infection increases the likelihood of future injury due to lightning, drowning, submersion, electrocution or vibration by 76%. Similarly, it shows that COVID-19 infection increases the likelihood of congenital malformations of the eye, ear, face, or neck in adults by 18.6%. We find that our test-based prospective design corrects for the detection of these spurious effects. This design results in a markedly lower false association rate of 4.1% across all negative control outcomes. For example, the test-based detects no effect of COVID-19 on the incidence of congenital malformations of the eye, ear, face, or neck—nor does it detect any increased incidence of injuries due to lightning, drowning, submersion, electrocution, or vibration. Thus, we find that the test-based design offers a more reliable way to detect long COVID symptoms.

### Characterizing long COVID symptoms and recovery trajectory

We investigate the long-term effect of COVID on all 614 health outcomes across three postacute time periods: early (30-120 days after infection, or 1-4 months), middle (120-360 days after infection, 4+ to 12 months) and late (360-720 days after infection, 12+ to 24 months). In the early postacute period (Figure 3a), we find that although the number of attributable effects is smaller than reported in previous studies, COVID-19 nevertheless impacts multiple organ systems: respiratory, cardiovascular, renal, immune, musculoskeletal, gastrointestinal, neurologic, and dermatological (hair loss). These findings confirm prior work showing that long COVID is a multisystem illness^1–3^.

**Figure 3:**
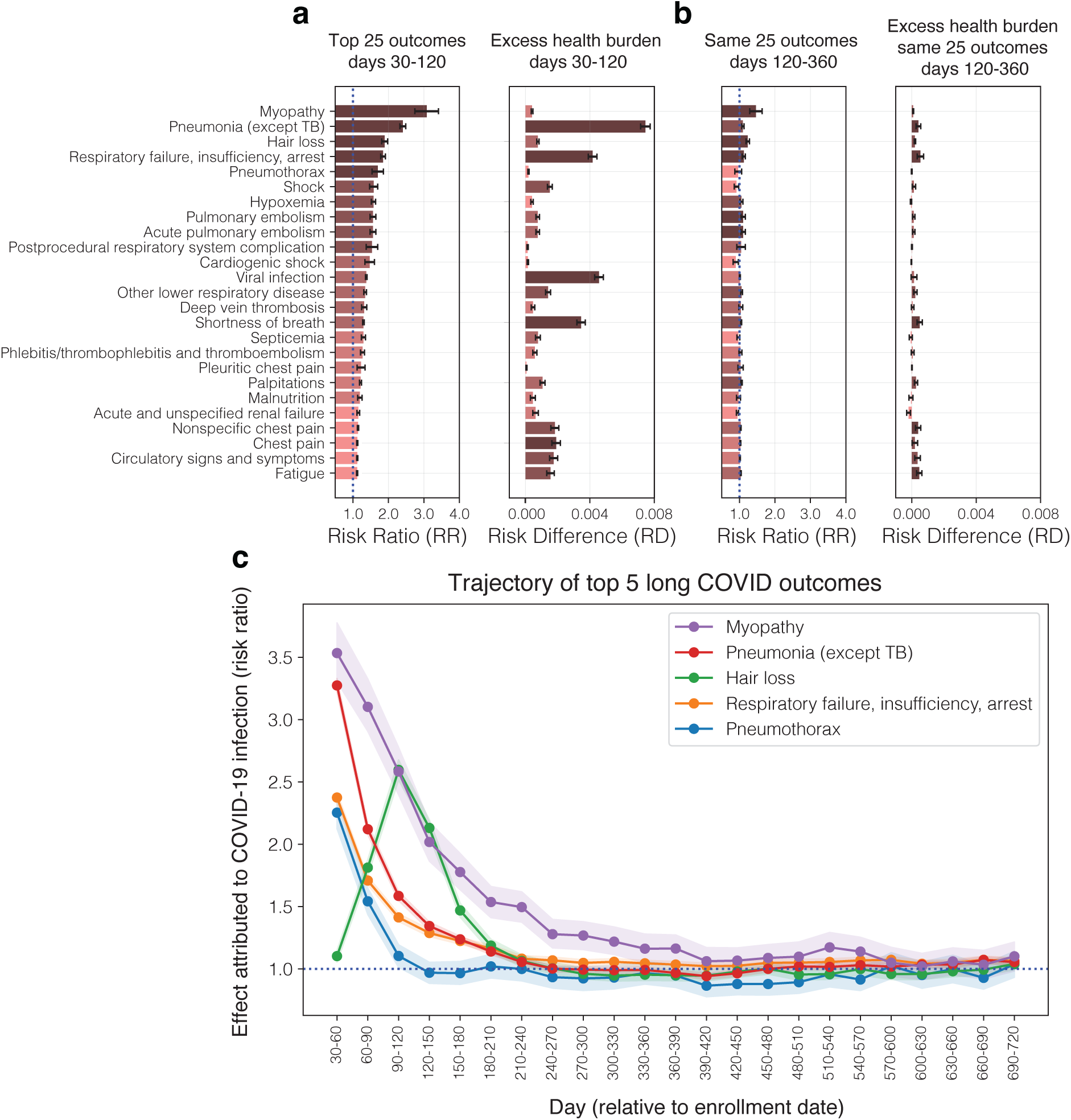
Long-term COVID-19 symptoms and sequelae detected by our test-based prospective study. **(a)** *Left:* Top 25 outcomes most attributable to long COVID, ranked by relative risk (RR) on days 30-120 after infection. We detect diverse, multi-system impacts of long COVID, which extend beyond the acute respiratory effects of the virus. For instance, conditions such as myopathy (RR 3.1), hair loss (RR 1.89), palpitations (RR 1.21), malnutrition (RR 1.20), and fatigue (RR 1.12) are substantially more likely after COVID-19 infection. All outcomes shown have RR ≥ 1.1, with 95% two-sided bootstrap confidence intervals (Bonferroni-corrected for multiple comparisons) also ≥ 1.1, indicating high-confidence attributable effects. *Right:* Corresponding risk differences (RDs in claims per month; Methods M3) show the absolute excess health risk attributable to long COVID symptoms. Respiratory (pneumonia, respiratory failure, shortness of breath), immune (viral infection, septicemia), and circulatory issues (chest pain, circulatory symptoms) pose the highest absolute burden, while conditions like myopathy, despite higher relative risks, have a smaller impact due to lower baseline prevalence. Overall, our findings validate prior research indicating that long COVID is a multi-systemic disease with persistent effects across organ systems. However, we find that its most significant population-level impacts are concentrated on a smaller subset of symptoms than previously reported. **(b)** The same outcomes as (a), but for days 120–360 post-infection, revealing a narrowed set of long COVID symptoms that persist up to one year after infection. *Left:* Risks of myopathy, hair loss, respiratory failure/insufficiency/arrest, and pulmonary embolism remain elevated (RR *>* 1.1), highlighting continued multi-system impacts of long COVID. *Right:* Across all outcomes, risk differences (RDs) fall to near zero, indicating low absolute excess risk. However, certain conditions such as pneumonia, shortness of breath, chest pain, and fatigue continue to contribute a small but statistically significant (p*<*0.05 with Bonferroni correction) risk due to high baseline prevalence. **(c)** Trajectories of RR for the top 5 long COVID symptoms shown in (a^3^). Unlike prior studies, we find that the average health of COVID-19 cases recovers to baseline within 1 year after infection. Bonferroni-corrected 95% confidence intervals are indicated by error bars (a-b) and shading (c) (Methods M3).

After the first 120 days, we find reduced, but still clinically meaningful impacts of long COVID (Figure 3b). In the middle post-acute period (days 120-360), the only symptoms with risk ratios greater than 1.1 are myopathy, hair loss, pulmonary embolism, acute pulmonary embolism, and respiratory failure, insufficiency, and respiratory arrest (two-sided bootstrap with Bonferroni correction). However, several outcomes—including pneumonia, shortness of breath, chest pain, and fatigue—continue to pose a significant public health burden (risk difference; Methods M3) despite exhibiting only modest risk ratios (between 1.0 and 1.1), owing to their high baseline prevalence in the general population. This suggests that while the intensity of risk fades, high-prevalence conditions remain burdensome at a population level up to a year after COVID-19 infection.

By one year post-infection, the measurable impact of long COVID appears to diminish significantly, with potential symptoms no longer clearly attributable to COVID-19. On days 360-720, there are no symptoms with significant risk ratios greater than 1.1 (two-sided bootstrap with Bonferroni correction). To further illustrate the trajectory of long COVID symptoms, we show the risk ratio of the top 5 early postacute period symptoms over the entire two-year followup period (Figure 3c). Downward trends indicate population-level recovery from COVID-19 by the end of the first year after infection. These findings differ from prior work showing significant harmful impacts of long COVID up to two^49^ or three years^48^ after infection. This is not to diminish the reality of such symptoms; rather, it suggests that they are unlikely to be specific to COVID-19. These symptoms may be due to a non-COVID condition that led to the initial hospital visit or COVID-19 testing, or some other factor. Ultimately, their cause is not discernible using these methods.

### Biases due to selective COVID-19 testing

We show that selection bias in who receives COVID-19 testing explains why our study design produces very different results compared to a conventional retrospective design using the same data. We assess these biases by rerunning a conventional retrospective study, but replacing the COVID-19 cases with “placebo cases”: individuals who were never diagnosed with COVID-19 and also received a negative (rather than positive) COVID-19 PCR test. We then compare these test-negative placebo cases to untested controls with the same medical history (Extended Data Figure 1a). Receiving a negative COVID-19 test cannot plausibly have harmful health effects. Thus, any difference in future outcomes between the placebo cases and controls must be because of non-COVID factors (Extended Data Figure 1b; x-axis). This allows us to measure the long-term effects of these non-COVID factors among tested individuals, an approach known as a negative control exposure in epidemiology^50, 51^.

We find that individuals who test negative for COVID-19 experienced more long-term health problems compared to untested individuals with similar medical histories. Notably, 82.5% of outcomes show elevated risk (RR ≥ 1.1) on days 360-720 after a negative COVID-19 test (Extended Data Figure 4). Furthermore, these elevated risks among test-negative individuals are positively correlated with those attributed to COVID-19 infection by a conventional analysis (Spearman Corr. 0.48; *p <* 10^−4^; Extended Data Figure 1b: left). This reveals that individuals that tested negative and positive share non-COVID risk factors that affect long-term outcomes. This selection bias—where some individuals are more likely to be tested due to non-COVID factors—undermines conventional retrospective designs and explains in part why they falsely attribute long-term effects to COVID-19 infection. On the other hand, our test-based design corrects for this selection bias. Our design significantly reduced the correlation between outcomes associated with negative testing and COVID-19 infection (*p <* 10^−4^; Steiger’s Z-test).

### Study design validation: detecting acute COVID-19 symptoms

We further validate the ability of our proposed study design to identify the known clinical effects of COVID-19 from insurance claims, by examining the well-established acute COVID-19 symptoms presenting within the first 30 days of infection (Extended Data Figure 5). Our analysis detected known acute symptoms of mild and severe COVID-19^52, 53^ across multiple systems. We observe only one symptom, anorexia, not known to be caused by COVID-19, which may be coded due to appetite decrease caused by COVID-19^54^. These findings demonstrate that, when properly analyzed using a test-based design, insurance claims data can identify clinical outcomes known to occur acutely. This supports the broader utility of insurance claims data for studying the effects of COVID-19.

## Discussion

As we continue to confront the impacts of the COVID-19 pandemic, it is important to pinpoint genuine long COVID effects from those identified by designs first used during the pandemic onset. We show that conventional studies are susceptible to selection bias, particularly due to differences in the health status of COVID-19 patients compared to controls who were not diagnosed. By using a prospective study design, where patients were enrolled at the time of their first PCR test, we mitigated many of these biases and more accurately attributed health outcomes to COVID-19. Our study reveals a smaller, yet still clinically significant, array of long COVID symptoms affecting multiple organ systems. This analysis suggests that while long COVID remains a serious concern, it may have fewer symptoms—and ones of shorter duration and lower overall severity—than previously thought.

Our approach, while an improvement over conventional retrospective studies, still has limitations. First, we rely on insurance claims data. While such insurance claims offer extensive coverage and longitudinal tracking, they are primarily designed for billing purposes rather than clinical research and may be lacking in clinical accuracy and granularity. It is thus reassuring that our study design is able to detect established acute COVID-19 symptoms (Extended Data Figure 5). It is similarly important to further study subpopulations we could not capture, such as healthy individuals taking at-home COVID-19 tests. Our analyses reveal that our sample is geographically representative (Supplementary Figure S2) and well-balanced along the dimensions of age, race, income, and education (Extended Data Tables 1-3), consistent with prior work^55^. Moreover, patients are linked longitudinally using a hash of their first name, last name, gender, and date of birth (Methods M1), which has the risk of misidentification and improper linkage. However, we show that our study results are robust to efforts to filter out these overlapping patients (Supplementary Figure S1).

While our prospective study mitigates the strong positive bias produced by previous designs, we in fact observe mild negative bias; our effects may be conservative by 10-20%. This is likely also due to selective testing, and is a manifestation of “collider bias”^17^. For instance, patients who tested negative in their first PCR test may be arriving at the hospital seeking treatment for other ailments (e.g., cancer) or for a non-COVID respiratory illness that could produce a non-specific prolonged viral syndrome, possible after many viruses^17^. We address this bias by reweighting test-negative controls to match COVID-19 cases on medical history. We note that relative risks approaching 1.0 indicate long-term symptoms at the same rate in COVID-19 cases and controls. Therefore, these data should not be interpreted as indicating that symptoms lasting more than a year in COVID-19 patients aren’t “real”, only that their etiology is uncertain. Furthermore, a small minority of individuals (*<* 1.1%) in the control group in each month later tested positive for COVID-19, again resulting in conservative effect estimates (Extended Data Figure 6). Finally, our requirement that relative risks exceed 1.1 raised the threshold for what is considered a meaningfully attributable effect. Thus, while the positive effects we detect are credible, there may be undetected small effects. Our study complements the RECOVER initiative^23^, which focused on defining a long COVID phenotype using a prospective clinical cohort. The RECOVER initiative identified 37 significant long COVID symptoms (RR *>* 1.5) from a narrow set of 44 self-reported symptoms. In contrast, we analyzed 614 outcomes in large-scale claims data and show that correcting for testing bias substantially reduces the number and duration of attributable effects, highlighting how study design influences long COVID estimates.

Overall, our findings suggest that the current understanding of long COVID may have to be reevaluated. They suggest that a more cautious and rigorous approach is needed to study the long-term effects of COVID-19. Future long COVID studies should continue to leverage large, diverse datasets and study designs that minimize the biases noted here. The prospective approach that we have taken could be applied to datasets used in previous conventional studies to develop a more complete understanding of the extent to which previous findings were due to bias or genuine long COVID effects. A deeper understanding of the biological mechanisms underlying long COVID, together with the development of new biomarkers, will be essential for tracking disease progression more precisely than is possible through population-based studies alone.

## Data Availability

The insurance claims data used in this study were obtained from the Komodo Health database under a data use agreement with Stanford University. Due to data use agreement restrictions and patient privacy requirements, the raw claims data cannot be shared publicly. Researchers interested in accessing similar data may contact Komodo Health to inquire about data access for academic research. Aggregate summary statistics, demographic tables, and risk ratio estimates supporting the findings are provided in the manuscript and Extended Data (Extended Data Tables E1-E3 and Extended Data Figures E1-E8).

## Methods

In Methods M1, we explain the datasets used in our analysis; in Methods M2, we explain data processing procedures to construct the cohorts and covariates in our subsequent observational studies, and in Methods M3, we explain the statistical analysis underlying our main results.

### M1 Datasets

#### Komodo

We use health insurance claims data from a sample of individuals in the United States, provided by the company Komodo Health (Komodo). The data are deidentified insurance claims which are collected and transmitted to Komodo by participating healthcare providers and payers^56^. Komodo data is deidentified per the standard specified in Section §164.514(a) of the Health Insurance Portability and Accountability Act of 1996 (HIPAA) Privacy Rule. Data deidentification is attested to through a formal determination by a qualified expert per Section §164.514(b)(1) of the HIPAA Privacy Rule. All data provided by Komodo is stored on a secure server behind a firewall. Data handling and analysis was conducted in accordance with Komodo policies and in accordance with the guidelines of the Stanford University Institutional Review Board.

Komodo traces the longitudinal patient journey of over 330 million unique U.S. patient lives, across multiple healthcare payers and providers. Komodo links patients across multiple data sources using a hash of the first name and last name coded by soundex^57^, date of birth, and gender of the patient. The Komodo dataset contains date-stamped hospital visits (inpatient, outpatient, emergency room, etc.), diagnosis codes (ICD-9, ICD-10), procedure codes (CPT, HCPCS, and ICD-10-PCS), drug prescription events (NDC codes). While the sample is not random sample, we find—consistent with prior work^55^—Komodo data is geographically representative (i.e. an approximately unbiased sample of different zip codes within each State), and representative along the dimensions of age, race, income, and education (Supplementary Figure S2 and Extended Data Tables 1-3). Furthermore, Komodo data is a widely used standard in largescale studies of health outcomes across many different areas including COVID-19 treatments ^59–61^, telehealth^55, 62^, drug safety^63^, mental health^64^, time-to-treatment^65^, cancer incidence^66^, and health resource utilization^67^.

The raw data provided by Komodo consist of two datasets. The first dataset, the Pandemic dataset, consists of 48,812,613,382 claims from 228,010,550 patients (mean 214.0 claims per patient). The Pandemic dataset consists of all patients with any COVID-related claim including COVID-19 testing, vaccination, diagnosis, etc. The second dataset, the Control dataset, consists of 5,759,362,767 claims from 33,749,570 patients (mean 170.6 claims per patient). The Control dataset is a random sample of all patients in the Komodo database that had no evidence of COVID-19 diagnosis, but includes individuals who received COVID-19 testing and vaccination (Methods M2). The observation period of both datasets ended on April 4th, 2023. Following prior work^4^, we filter these datasets to only include individuals who had at least one claim in the year 2019 (n = 18,084,992 individuals in the Pandemic dataset, n = 158,213,552 individuals in the Control dataset).

#### Area Deprivation Index

We use the Area Deprivation Index (ADI) as a measure of socioeconomic disadvantage based on patient residential location^69^. Patients in the cohort were linked via zip code to their residential ADI, a national percentile ranking from 1 to 100, where a higher ADI score indicates a higher level of socioeconomic disadvantage. Komodo data only includes the first three digits of the patient zip code to ensure anonymity. Thus, we aggregate neighborhood ADI measures for each five-digit zip codes to the first three digits using a population weighted average. Individuals who have multiple addresses with different zip codes, were assigned the average of all their assigned ADIs.

#### U.S. Census

We obtained data on demographic and socioeconomic features from the 2016-2020 American Community Survey (ACS) census estimates^70^ and 2020 Decennial Census^71^. Specifically, we retrieved mean family income, percentage of population with less than a Bachelor’s degree, and percentage of population that is non-white or Hispanic for each zip code. To ensure patient anonymity, Komodo data only includes the first three digits of the patient zip code. Thus, we aggregate five-digit zip codes to the first three digits using a population weighted average. While only zip code level information was available to describe patient socioeconomic and racial demographics, area-level measures have been demonstrated by prior work to meaningfully explain health outcomes^72, 73^.

#### High-dimensional outcomes

We consider 614 health outcomes which were aggregated from multiple sources, consisting of 569 outcomes which could plausibly be caused by COVID-19, and 49 outcomes which are negative control outcomes (outcomes which are not plausibly caused by COVID-19 and which are used to analyze the extent of bias in our estimates).

We obtained an initial set of 695 outcomes by combining (a) an outcomes list generated from the Clinical Classification Software Refined (CCSR)^74^, consistent with a previous high-dimensional study of long-term COVID-19 effects^4^ (b) additional outcomes plausibly caused by COVID-19 from previous studies^5, 6, 9^ and (c) manually generating a list of outcomes unlikely to be caused by COVID-19. The CCSR categorizes ICD-10-CM codes into clinically meaningful groups, and was developed as part of the Healthcare Cost and Utilization Project sponsored by the Agency for Healthcare Research and Quality. We categorized these 695 outcomes into 611 outcomes plausibly caused by COVID-19, and 84 outcomes which are negative control outcomes^78^. Our initial list of outcomes consisted of 695 outcomes, which we filtered down to 614 outcomes (including 49 negative controls) by excluding outcomes which were extremely sparse (i.e., those that occurred in less than 0.0001% of patients in the pre-enrollment period across all cohorts).

### M2 Data processing

#### COVID-related events

We used the following ICD-10, CPT, and HCPCS codes to identify COVID-19 related diagnoses and tests.

- **Confirmed COVID-19 positive diagnosis**: ICD-10 code B9729 before April 1, 2020, or ICD-10 Code U071 at any time.
- **Evidence of COVID-19**: confirmed COVID-19 diagnosis, or suspected COVID-19 infection, or history of COVID-19 from ICD-10 codes U072, U089, U099, or Z8616.
- **COVID-19 PCR Test**. CPT codes 87635, 87636, 87637, 0202U, 0223U, 0225U, 0240U, 0241U. HCPCS codes U0001, U0003, or U0005.

#### Observational study cohort construction

We construct three observational cohort studies.

#### Conventional retrospective study

We first use a conventional, retrospective cohort study design based on prior highdimensional studies of long COVID^4–6, 9^, in which COVID-positive patients (COVID-19 cases) are matched with patients for which there is no evidence of COVID-19 diagnosis (control). To construct the COVID-19 cases group, we sample all individuals who had a confirmed positive COVID-19 diagnosis between March 1st, 2020 and March 1st, 2021 (n = 10,445,894), filtering for only those who had no evidence of COVID-19 (e.g., suspected COVID-19) prior to their first confirmed COVID-19 positive diagnosis date (n = 10,431,860). To examine postacute outcomes, we then selected those that were alive 30 days after positive COVID-19 diagnosis (n = 10,118,877).

Following prior work^4, 9^, we generate a “control” group (no COVID-19 evidence) by selecting all individuals in our Control dataset who had no record of any COVID-19 diagnosis (n = 18,084,711). To eliminate any effect of seasonality in cohort enrollment, we sampled cohort enrollment dates for each individual in the control group from the distribution of cohort enrollment dates in the COVID-19 cases (i.e. the day in which the COVID-19 case tested positive). For the control group as well, we further filter for individuals that were alive on their enrollment date (n = 17,728,714), and finally for those who were alive 30 days after enrollment into the cohort (n = 17,717,308).

#### Prospective study with testing-based enrollment

In our proposed study design, patients are enrolled into the cohort the first time they take a PCR COVID-19 test between March 1st, 2020 and March 1st, 2021 (n = 39,967,054), if they had no prior evidence of COVID-19 diagnosis (n = 39,199,683). COVID-19 cases are defined as those who received a positive test result subsequent to testing, that is, they were diagnosed with COVID-19 within 7 days of testing (n = 2,866,888). We filter the COVID-19 cases for only those that remained alive 30 days after their enrollment date (n = 2,832,575). The control group is defined as those who received a negative test result, that is, they had no evidence of COVID-19 within 7 days of testing (n = 36,320,411). We further filter the control group for only individuals who were alive 30 days after the enrollment date (n = 36,095,303).

To mitigate seasonality, we down-sampled the control group to ensure alignment between its enrollment date distributions and the treatment group enrollment dates (n = 14,162,875). To do so, for each day, we calculate the minimum ratio between control and COVID-19 cases enrolled for each day, rounded down to an integer value (ratio = 5:1). We down-sample the control units for each enrollment day so that the ratio of control to COVID-19 cases is 5:1 for each of the enrollment dates.

#### Conventional retrospective study with placebo cases (negative COVID-19 PCR test recipients)

To validate the prior study design, we conduct an analogous cohort study in which we replace the COVID-19 cases with a group of patients that received a negative COVID-19 PCR test, which cannot plausibly have severe adverse health effects. We refer to these as “placebo” cases. This approach is also known as a negative control exposure in the epidemiology literature^50, 51^. Here, we use negative COVID-19 tests, by sampling all individuals who received their first PCR COVID-19 test between March 1st, 2020 and March 1st, 2021 (n = 39,967,054), and only including those for whom there was no evidence of a COVID-19 diagnosis before or after their test date (n = 27,439,975). As before, we further select for those that were alive 30 days after enrollment into the cohort, that is, those who were alive 30 days after receiving their first COVID-19 test (n = 27,263,919).

We generate a “control” group analogous to the prior study, which consists of all individuals in the Control dataset (n = 18,084,711). We use an enrollment date randomly sampled from the distribution of enrollment dates in the null treatment group, i.e. COVID-19 test dates. We further filter for those who were alive on their enrollment date (n = 17,737,596) and were alive for 30 days after enrollment into the study (n = 17,726,124). We use an enrollment date randomly sampled from the distribution of enrollment dates in the null treatment group, i.e. COVID-19 test dates.

#### Covariates

Across all experiments, we match on several static covariates used in prior work^4^: gender (one-hot encoded as male, female, and unknown/other), age at time of enrollment (years), and area deprivation index (ADI), which are taken or calculated from the patient data. We additionally match on U.S. Census demographic features described in Methods M1 (mean family income, non-white or Hispanic percentage, no-bachelor degree percentage) by first aggregating these features at the ZIP-3 level and then mapping them to respective patients residing in each ZIP-3 area (Extended Data Tables 1–3).

We also measure pre-enrollment health status from the four years prior to enrollment date. We follow prior work^4^ and use a high-dimensional set of health outcomes from before treatment. We also use several high-level proxies of healthcare usage: the number of inpatient visits, out-patient visits, home visits, emergency room visits, ambulance visits, pharmacy visits, and non-hospital institution visit. Lastly, we include a marker of data quality, the number of months prior to the testing date in which insurance data was payer-complete.

We aggregate each of the health status outcomes to the monthly granularity, creating 48 features for all of the 614 outcomes and proxies of healthcare usage for each of the four years (48 months) prior to study enrollment. To reduce noise from claim frequency variations, we binarize each outcome at the monthly level, indicating whether any claim occurred in that month. Before balancing covariates, we normalize the covariate matrix *X* to the unit norm. For missing age and ADI values, we impute the feature using the mean.

### M3 Analysis

#### Matching pre-enrollment medical histories and demographics

We reweight the control sample using a synthetic-control-type approach^46, 82–84^ to eliminate biases that can be explained away using observed pre-exposure covariates.^17, 44, 45^ Our reweighting is designed to achieve fine-grained balance of medical histories and patient demographics between COVID-19 (or placebo) cases and controls over the four-year period before enrollment, as illustrated in Methods Figure 1. We derive weights by solving the following optimization problem using the `cvxpy` package in `Python` with the `MOSEK` solver,

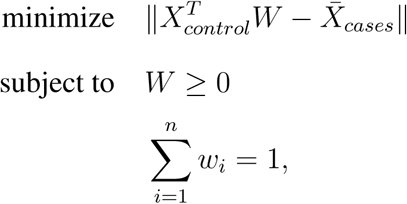

where:

- 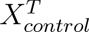 is the control matrix of covariates, transposed,
- *X̅_cases_* is the vector of treatment covariate means,
- *W_n_* is the weight vector consisting of *n* weights, one for each control unit,

We balance covariates (*X*) that capture individuals’ medical history prior to enrollment. These include health trends across all 614 outcomes and summary measures of healthcare utilization. Consistent with prior work^4^, we also include additional static demographic covariates in *X*, such as gender, age at enrollment, and the area deprivation index. A full description of the covariates used for balancing is provided in Methods M2: Covariates.

### Risk ratio and risk difference

We assess the effect of COVID-19 on each of the 614 outcomes by computing a relative risk ratio (RR) and risk difference (RD) for the occurrence of the outcome over several time windows in the subsequent two years. To calculate the RR and RD for each outcome, we compare the incidence of the outcome in the cases group to the incidence in the weighted control group. The risk ratio (RR) is defined as the ratio of the probability of the outcome occurring in the cases group to the probability of the outcome occurring in the weighted control group,

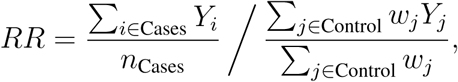

where *Y_i_* represents the number of claims for the outcome per time period (e.g., days 30-120) for individual *i*, *n*_cases_ is the number of individuals in the cases group, *w_j_* is the weight assigned to individual *j* in the control group, derived from the synthetic controls, and *Y_j_* is the number of claims per time period for individual *j* in the control group. The risk difference (RD, in claims per month) is defined as the difference in the average number of outcome occurrences per month, between the cases group and the weighted control group,

**Methods Figure 1:**
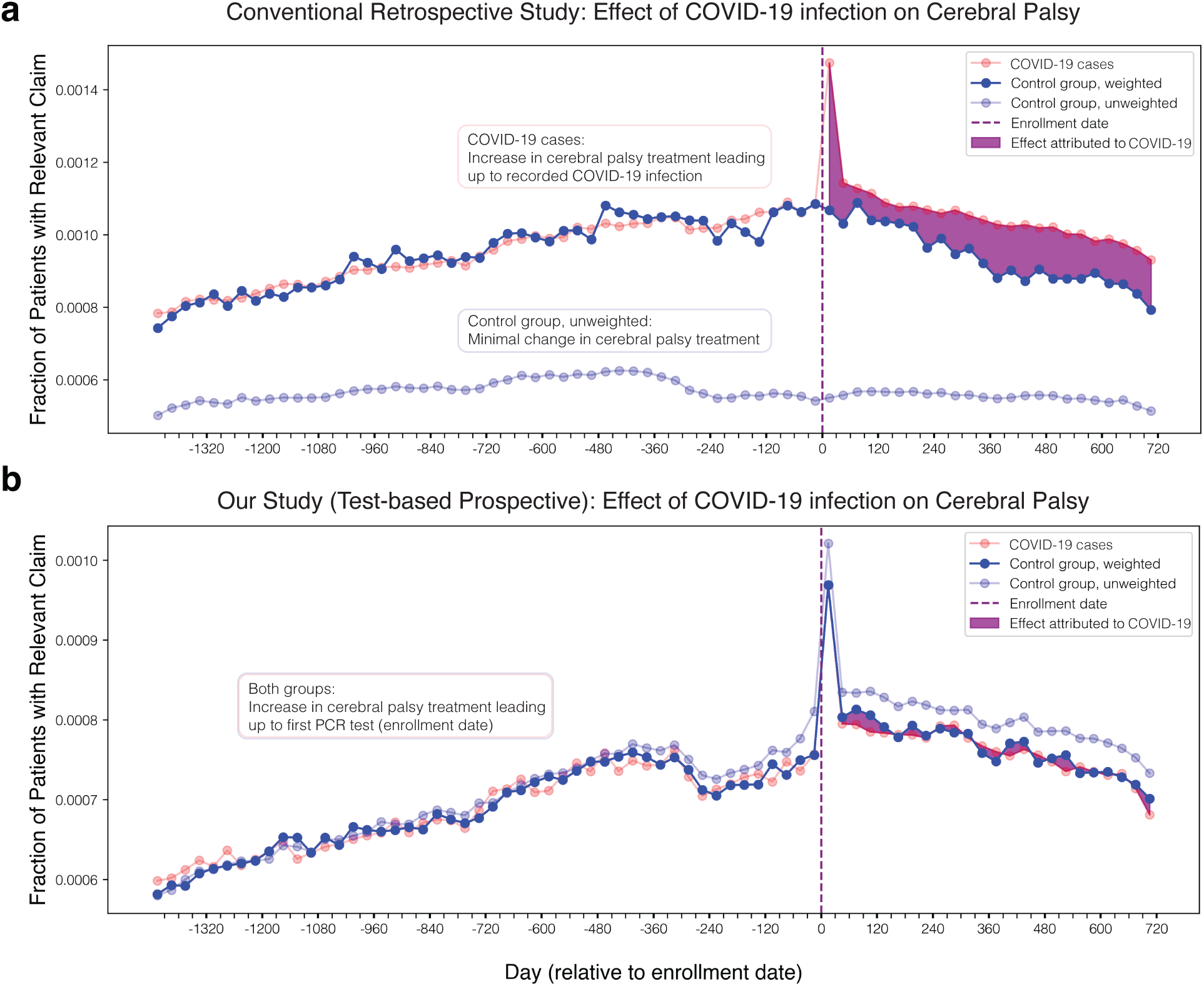
Comparison of a conventional retrospective study and our test-based prospective study, examining cerebral palsy in adults, an outcome that cannot plausibly be caused by COVID-19. **(a)** Conventional retrospective study design. Spurious effects (purple) persist for up to two years post-COVID. The control group differs substantially from COVID-19 cases. On one hand, COVID-19 cases show rising cerebral palsy diagnoses prior to infection, likely reflecting individuals already seeking care for cerebral palsy who incidentally tested positive for COVID-19. On the other hand, cerebral palsy diagnoses remain flat among unweighted controls—revealing a mismatch between groups. **(b)** Proposed design. No spurious effects observed. Both COVID-19 cases and test-negative controls (unweighted) exhibit similar pre-test increases in cerebral palsy diagnoses, suggesting comparable care-seeking behavior and populations with similar baseline health status.

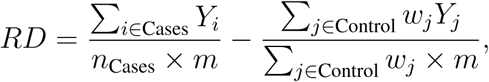

where *m* is the number of months in the followup period, that is, 3 months for days 30-120, 8 months for days 120-360, and 12 months for days 360-720.

Relative Risk (RR) and Risk Difference (RD) offer complementary insights into the impact of COVID-19 on health outcomes. RR indicates the proportional change in risk due to COVID-19 infection, making it valuable for comparing effects across varying baseline risks. However, its interpretation depends on baseline incidence: a large RR for rare outcomes might represent only a small absolute risk increase, while a moderate RR for common outcomes can imply substantial absolute differences. Conversely, RD measures the absolute change in risk directly, highlighting the actual disease burden. This makes RD especially relevant for public health decisions and resource allocation. However—unlike the RR which is unit free—the definition of the RD depends closely on the mechanics of how claims are recorded, e.g., if a healthcare system doubles the number of claims recorded per health event then the RD would double. RRs and RDs are computed for each outcome across the following four time periods for *Y* :

- **Acute period**: 1-30 days after COVID-19 test date.
- **Early post-acute period**: 30-120 days after COVID-19 test date.
- **Middle of post-acute period**. 120-360 days after COVID-19 test date
- **Late post-acute period**. 360-720 days after COVID-19 test date

### Statistical analysis

Confidence intervals and p-values were estimated using a bootstrap with 1,000 resamples. For each bootstrap replicate, patient records were sampled with replacement and the statistic of interest was computed; the standard error was then computed as the standard deviation of the bootstrap replicates.^88^ Statistical significance was assessed using Gaussian 95% confidence intervals obtained using the bootstrap standard error (i.e., estimate ± 1.96 standard errors). Multiple comparisons were controlled using a Bonferroni correction to maintain an overall significance level of *α* = 0.05. Steiger’s Z-test was used to compare pairs of Spearman correlations, and hypothesis tests for single Spearman correlation coefficients were computed using two-sided Student’s t-tests^89–91^.

## Acknowledgements

We thank Johann Gaebler, Serina Chang, Emma Pierson, and Emma Brunskill for helpful conversations; and David Rehkopf, Isabella Chu, Shafiq Poonja, Molly Wilson, Jane Huang, Tonya Brody and other staff at Komodo Health and the Stanford Center for Population Health Sciences for insurance claims data and feedback. H.N. was supported by a Stanford Knight-Hennessy Scholarship and the National Science Foundation Graduate Research Fellowship under grant no. DGE-1656518. We also gratefully acknowledge the support of NSF under Nos. OAC-1835598 (CINES), CCF-1918940 (Expeditions), DMS-2327709 (IHBEM), IIS-2403318 (III), SES-2242876; Stanford Data Applications Initiative, Wu Tsai Neurosciences Institute, Stanford Institute for Human-Centered AI, Chan Zuckerberg Initiative, Amazon, Genentech, GSK, Hitachi, SAP, and UCB.

## Extended Data

**Extended Data Figure 1:**
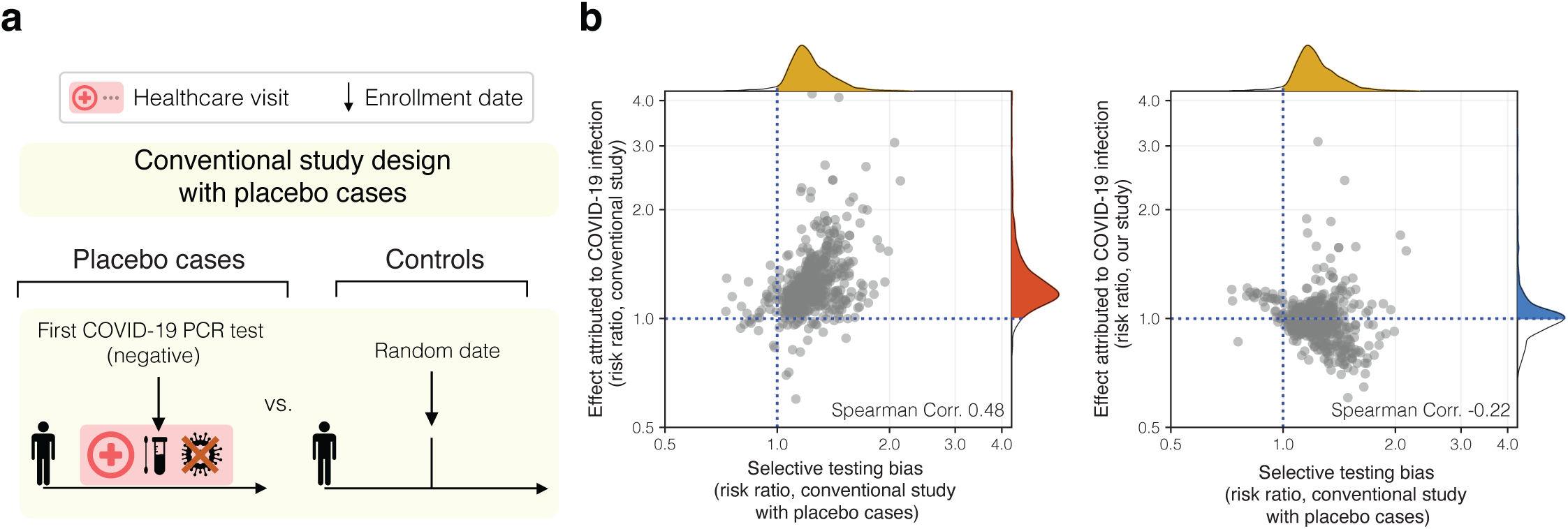
Selective testing bias explains results from conventional retrospective studies. **(a)** We measure bias introduced by selective testing of patients with non-COVID health problems. To do this, we replicate a conventional study design but replace COVID-19 cases with individuals who tested negative for COVID-19. These “placebo cases” had a healthcare visit and COVID-19 PCR test but were never diagnosed with COVID-19. We compare placebo cases to controls—individuals with similar histories, no COVID-19 diagnosis, and typically no visit or testing at enrollment. Because a negative test result cannot plausibly cause future health effects, any later differences between placebo cases and controls must be due to non-COVID factors among tested individuals. **(b)** Our study design appears less susceptible to selective testing bias compared to a conventional analysis. Each of the *N* = 614 points represents one outcome (e.g., headache, nausea, etc.) on days 30-120 post-infection. *Left:* Selective COVID-19 testing bias explains the effects attributed to COVID-19 by a conventional analysis (Spearman Corr 0.48). *Right:* In our study comparing COVID-19 cases to test-negative controls, this correlation is significantly weaker (Spearman Corr −0.22), indicating only mild bias^17^ (Discussion). Our findings show that accurate attribution of long-term health outcomes to COVID-19 requires accounting for testing behavior, not just medical history.

**Extended Data Figure 2:**
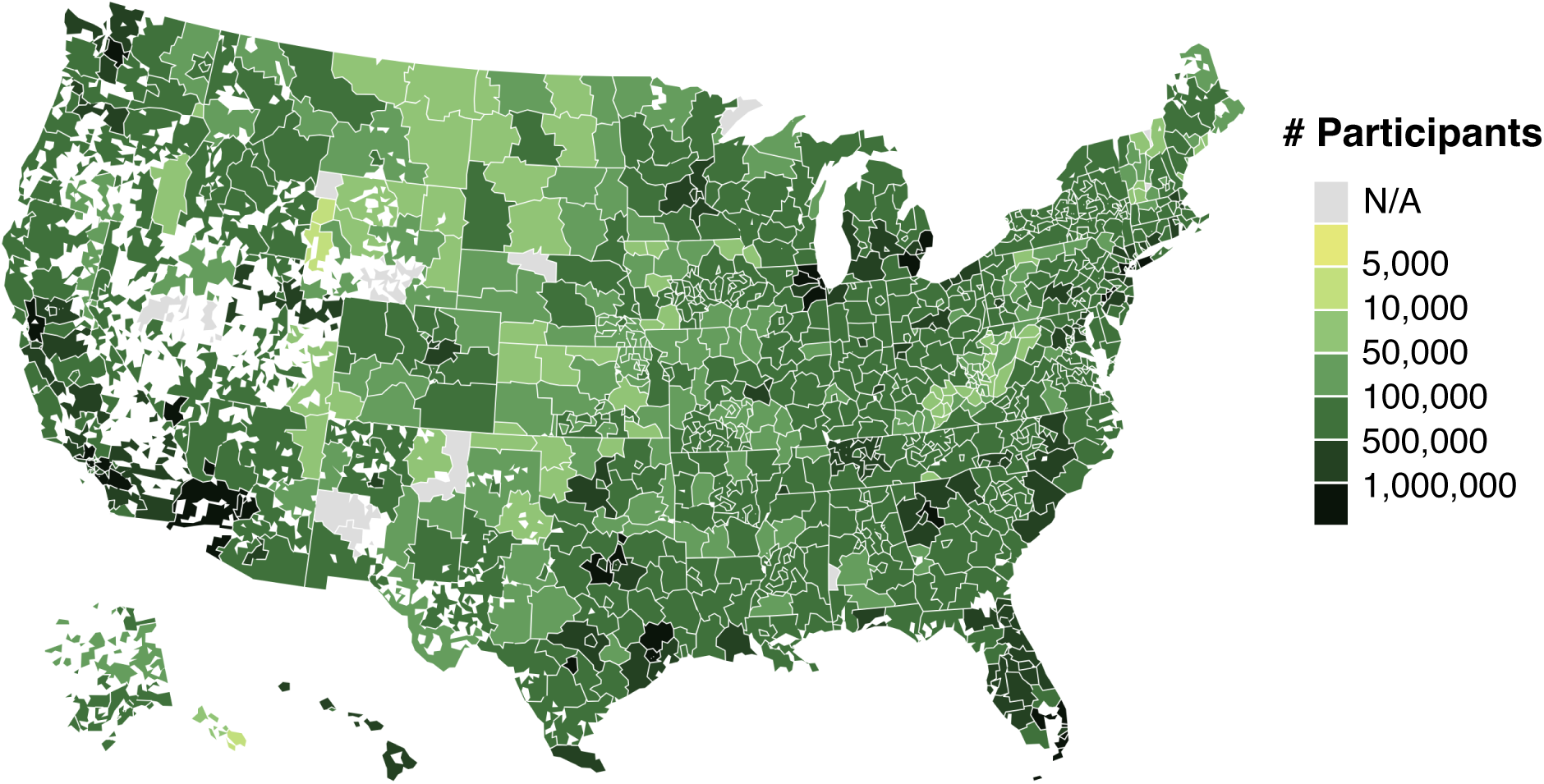
Number of participants across U.S. zipcodes. This nationwide study leverages a health-care map (Komodo) which aggregates 14.4 billion insurance claims across 244.7 million patients. Zipcodes are truncated to three digits to ensure patient anonymity.

**Extended Data Figure 3:**
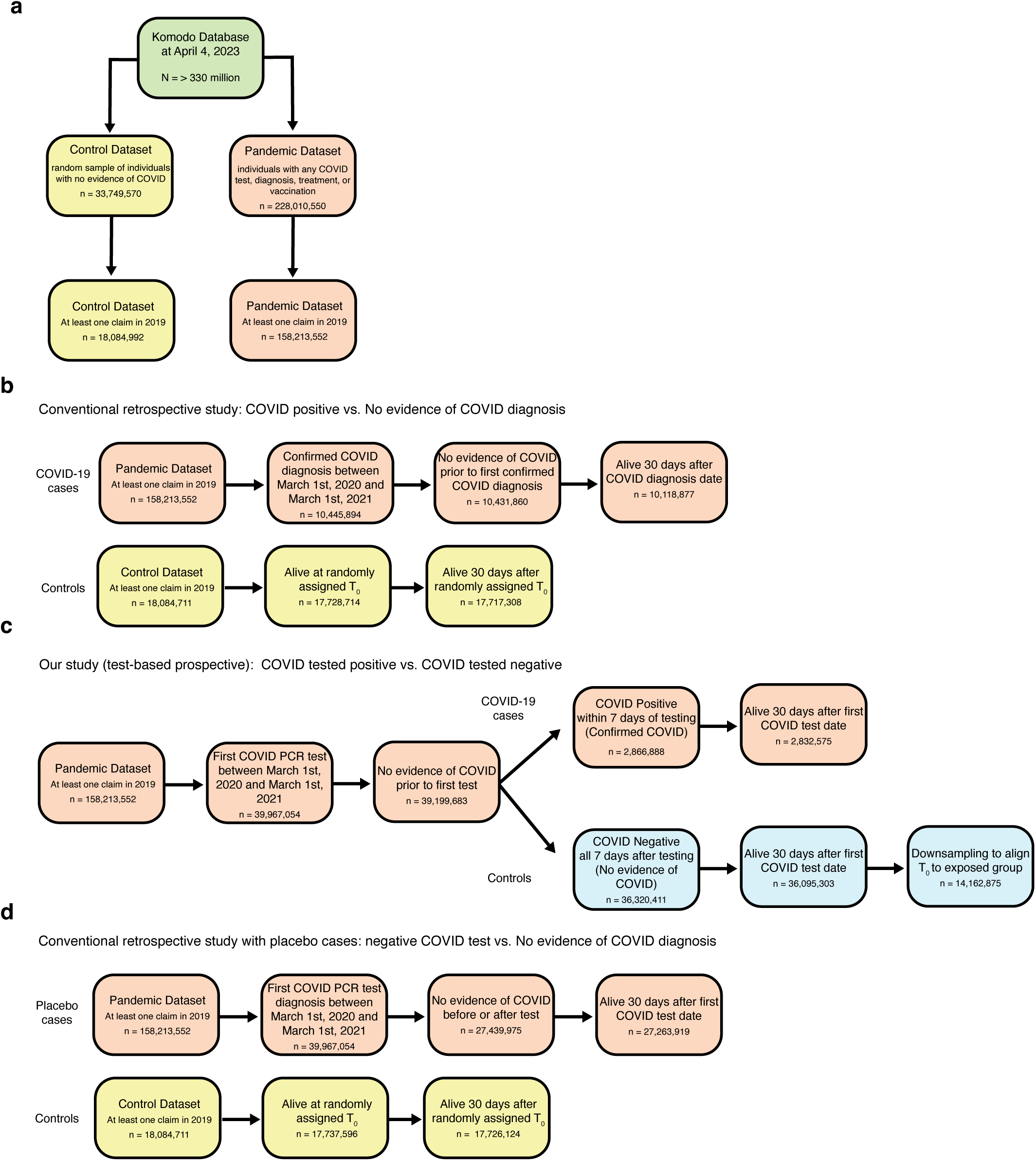
Large-scale health claims dataset enables varied observational studies designs to measure the long-term health effects of COVID. **(a)** Overall dataset. **(b)** The construction of the cohort for a conventional retrospective study **(c)**. The construction of the cohort for our test-based prospective study. **(d)** The construction of the cohort for a conventional retrospective study with placebo cases: an event that cannot plausibly have adverse health effects (negative COVID-19 PCR test).

**Extended Data Table 1:**
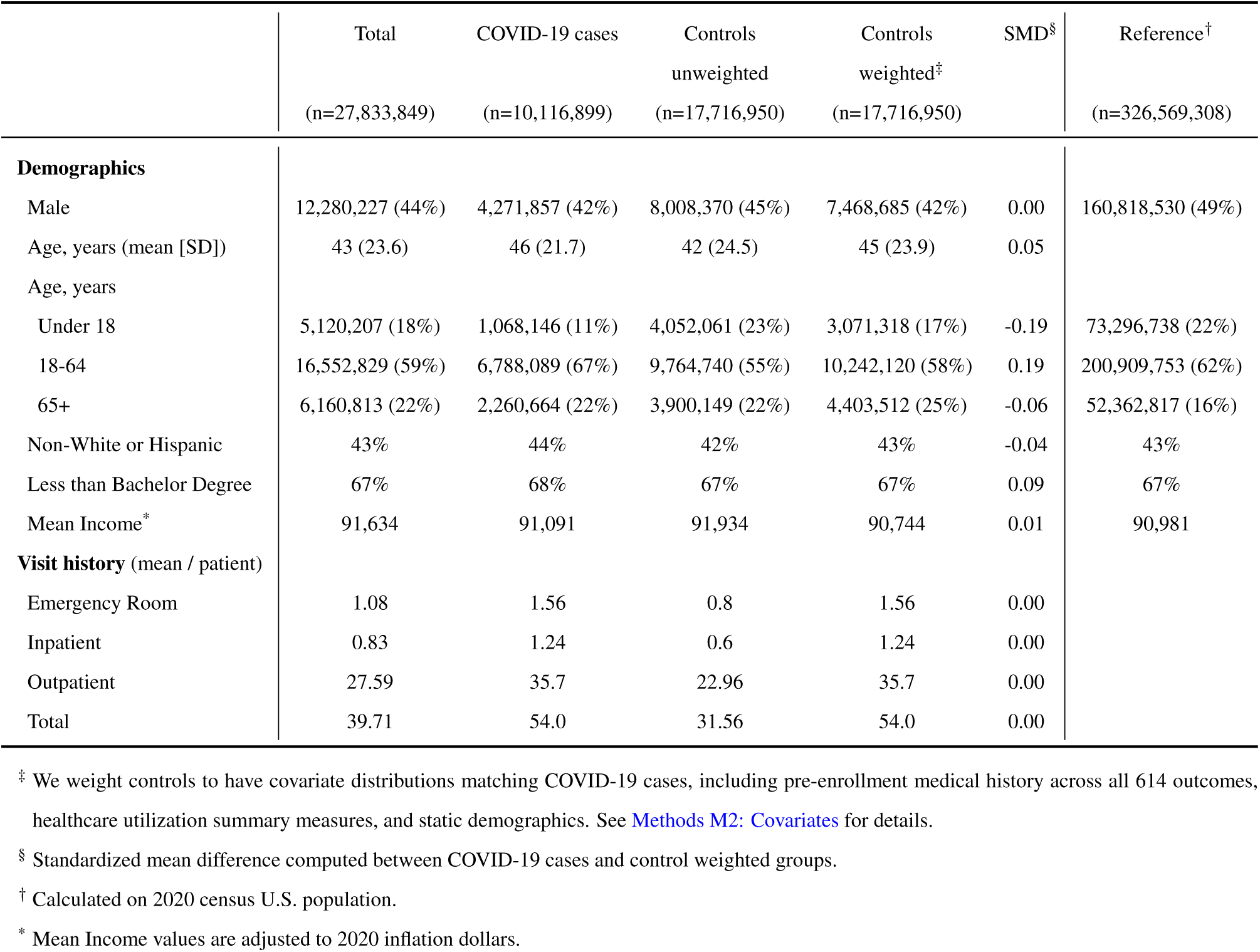
Conventional retrospective study; summary statistics of patient demographics and healthcare system interactions, stratified by total, COVID-19 cases, and control groups.

|  | Total<br>(n=27,833,849) | COVID-19 cases<br>(n=10,116,899) | Controls<br>unweighted<br>(n=17,716,950) | Controls<br>weighted<sup>‡</sup><br>(n=17,716,950) | SMD<sup>§</sup> | Reference<sup>†</sup><br>(n=326,569,308) |
| --- | --- | --- | --- | --- | --- | --- |
| **Demographics** |  |  |  |  |  |  |
| Male | 12,280,227 (44%) | 4,271,857 (42%) | 8,008,370 (45%) | 7,468,685 (42%) | 0.00 | 160,818,530 (49%) |
| Age, years (mean [SD]) | 43 (23.6) | 46 (21.7) | 42 (24.5) | 45 (23.9) | 0.05 |  |
| Age, years |  |  |  |  |  |  |
| Under 18 | 5,120,207 (18%) | 1,068,146 (11%) | 4,052,061 (23%) | 3,071,318 (17%) | -0.19 | 73,296,738 (22%) |
| 18-64 | 16,552,829 (59%) | 6,788,089 (67%) | 9,764,740 (55%) | 10,242,120 (58%) | 0.19 | 200,909,753 (62%) |
| 65+ | 6,160,813 (22%) | 2,260,664 (22%) | 3,900,149 (22%) | 4,403,512 (25%) | -0.06 | 52,362,817 (16%) |
| Non-White or Hispanic | 43% | 44% | 42% | 43% | -0.04 | 43% |
| Less than Bachelor Degree | 67% | 68% | 67% | 67% | 0.09 | 67% |
| Mean Income<sup>*</sup> | 91,634 | 91,091 | 91,934 | 90,744 | 0.01 | 90,981 |
| **Visit history** (mean / patient) |  |  |  |  |  |  |
| Emergency Room | 1.08 | 1.56 | 0.8 | 1.56 | 0.00 |  |
| Inpatient | 0.83 | 1.24 | 0.6 | 1.24 | 0.00 |  |
| Outpatient | 27.59 | 35.7 | 22.96 | 35.7 | 0.00 |  |
| Total | 39.71 | 54.0 | 31.56 | 54.0 | 0.00 |  |

<sup>‡</sup> We weight controls to have covariate distributions matching COVID-19 cases, including pre-enrollment medical history across all 614 outcomes, healthcare utilization summary measures, and static demographics. See Methods M2: Covariates for details.
<sup>§</sup> Standardized mean difference computed between COVID-19 cases and control weighted groups.
<sup>†</sup> Calculated on 2020 census U.S. population.
<sup>\*</sup> Mean Income values are adjusted to 2020 inflation dollars.

**Extended Data Table 2:** Our study (test-based prospective); summary statistics of patient demographics and health-care system interactions, stratified by total, COVID-19 cases, and control groups.

|  | Total<br>(n=16,994,746) | COVID-19 cases<br>(n=2,832,376) | Controls<br>unweighted<br>(n=14,162,370) | Controls<br>weighted<sup>‡</sup><br>(n=14,162,370) | SMD<sup>§</sup> | Reference<sup>†</sup><br>(n=326,569,308) |
| --- | --- | --- | --- | --- | --- | --- |
| **Demographics** |  |  |  |  |  |  |
| Male | 7,063,814 (42%) | 1,219,533 (43%) | 5,844,281 (41%) | 6,065,745 (43%) | 0.00 | 160,818,530 (49%) |
| Age, years (mean [SD]) | 41 (21.4) | 42 (21.0) | 41 (21.5) | 42 (21.5) | 0.02 |  |
| Age, years |  |  |  |  |  |  |
| Under 18 | 2,740,157 (16%) | 411,173 (15%) | 2,328,984 (16%) | 2,208,927 (16%) | -0.03 | 73,296,738 (22%) |
| 18-64 | 11,666,190 (69%) | 1,991,693 (70%) | 9,674,497 (68%) | 9,704,918 (69%) | 0.04 | 200,909,753 (62%) |
| 65+ | 2,588,399 (15%) | 429,510 (15%) | 2,158,889 (15%) | 2,248,525 (16%) | -0.02 | 52,362,817 (16%) |
| Non-White or Hispanic | 43% | 44% | 42% | 45% | 0.04 | 43% |
| Less than Bachelor Degree | 67% | 68% | 67% | 65% | 0.27 | 67% |
| Mean Income<sup>*</sup> | 93,225 | 90,539 | 93,761 | 88,819 | 0.07 | 90,981 |
| **Visit history** (mean / patient) |  |  |  |  |  |  |
| Emergency Room | 1.23 | 1.25 | 1.23 | 1.25 | 0.00 |  |
| Inpatient | 0.88 | 0.86 | 0.89 | 0.86 | 0.00 |  |
| Outpatient | 34.62 | 32.86 | 34.98 | 32.88 | 0.00 |  |
| Total | 47.69 | 46.53 | 47.92 | 46.55 | 0.00 |  |

<sup>‡</sup> We weight controls to have covariate distributions matching COVID-19 cases, including pre-enrollment medical history across all 614 outcomes, healthcare utilization summary measures, and static demographics. See Methods M2: Covariates for details.
<sup>§</sup> Standardized mean difference computed between COVID-19 cases and control weighted groups.
<sup>†</sup> Calculated on 2020 census U.S. population.
<sup>\*</sup> Mean Income values are adjusted to 2020 inflation dollars.

**Extended Data Table 3:** Conventional retrospective study with placebo cases (individuals that tested negative for COVID-19); summary statistics of patient demographics and healthcare system interactions, stratified by total, placebo cases, and control groups.

|  | Total<br>(n=44,965,453) | Placebo cases<br>(n=27,263,919) | Controls<br>unweighted<br>(n=17,701,534) | Controls<br>weighted<sup>‡</sup><br>(n=17,701,534) | SMD<sup>§</sup> | Reference<sup>†</sup><br>(n=326,569,308) |
| --- | --- | --- | --- | --- | --- | --- |
| **Demographics** |  |  |  |  |  |  |
| Male | 19,517,875 (43%) | 11,517,280 (42%) | 8,000,595 (45%) | 7,498,223 (42%) | -0.00 | 160,818,530 (49%) |
| Age, years (mean [SD]) | 41 (22.9) | 41 (21.8) | 41 (24.4) | 41 (23.7) | -0.01 |  |
| Age, years |  |  |  |  |  |  |
| Under 18 | 8,708,798 (19%) | 4,669,746 (17%) | 4,039,052 (23%) | 3,923,967 (22%) | -0.13 | 73,296,738 (22%) |
| 18-64 | 27,926,674 (62%) | 18,195,826 (67%) | 9,730,848 (55%) | 10,289,702 (58%) | 0.18 | 200,909,753 (62%) |
| 65+ | 8,186,881 (18%) | 4,311,375 (16%) | 3,875,506 (22%) | 3,430,882 (19%) | -0.09 | 52,362,817 (16%) |
| Non-White or Hispanic | 42% | 42% | 42% | 42% | 0.00 | 43% |
| Less than Bachelor Degree | 67% | 66% | 67% | 66% | 0.00 | 67% |
| Mean Income* | 93,162 | 93,976 | 91,939 | 92,333 | 0.00 | 90,981 |
| **Visit history** (mean / patient) |  |  |  |  |  |  |
| Emergency Room | 0.97 | 1.09 | 0.79 | 1.09 | 0.00 |  |
| Inpatient | 0.74 | 0.83 | 0.6 | 0.83 | 0.00 |  |
| Outpatient | 29.13 | 33.14 | 22.95 | 33.14 | 0.00 |  |
| Total | 39.74 | 45.07 | 31.53 | 45.08 | 0.00 |  |

<sup>‡</sup> We weight controls to have covariate distributions matching placebo cases, including pre-enrollment medical history across all 614 outcomes, health-care utilization summary measures, and static demographics. See Methods M2: Covariates for details.
<sup>§</sup> Standardized mean difference computed between placebo cases and weighted control groups.
<sup>†</sup> Calculated on 2020 census U.S. population.
\* Mean Income values are adjusted to 2020 inflation dollars.

**Extended Data Figure 4:**
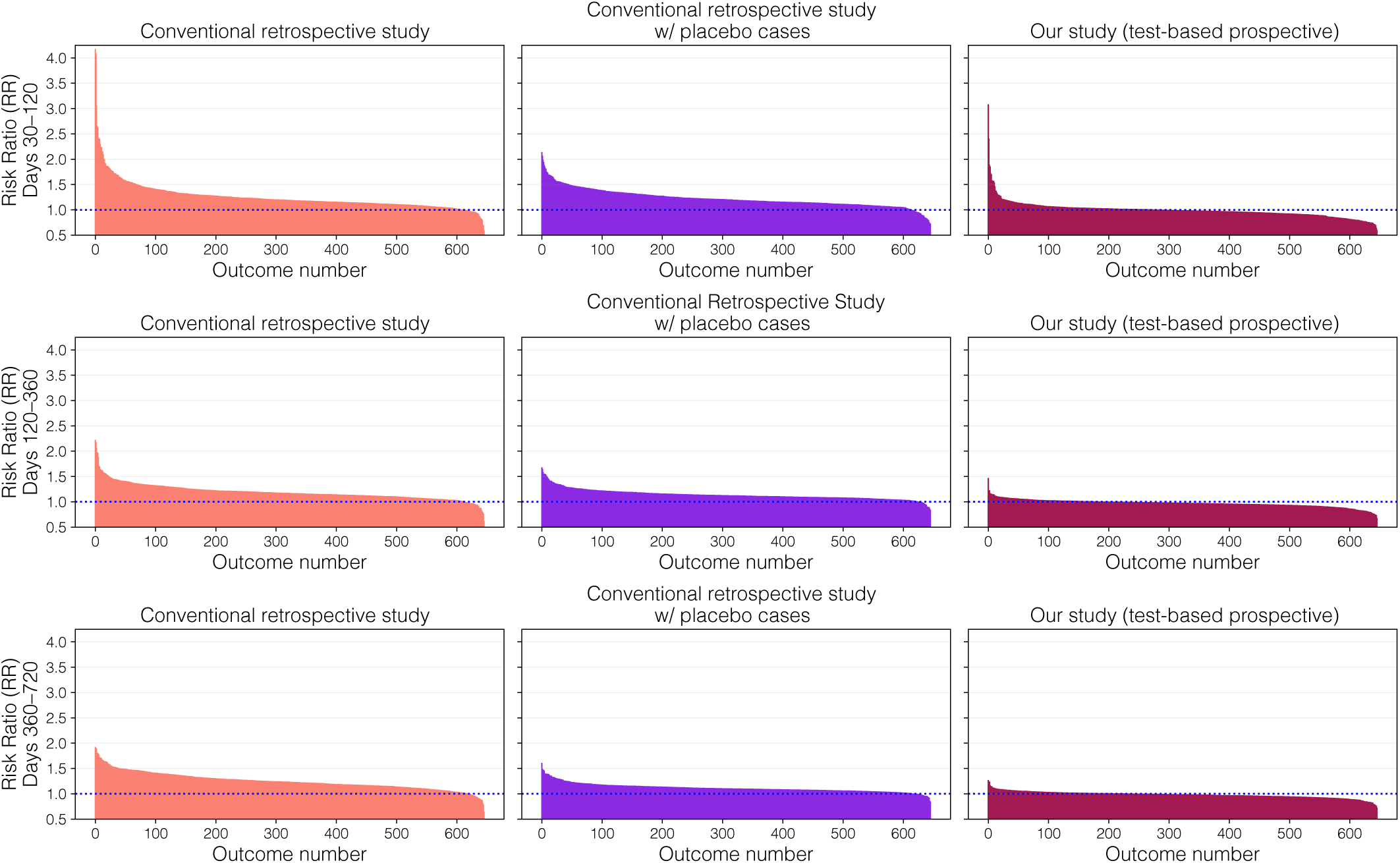
COVID symptoms and sequelae detected by a conventional retrospective study vs. our study: all 614 outcomes. Risk ratios (RRs; Methods M3) for the effect of COVID-19 on days 30-120 (top row), days 120-360 (middle row), and days 360-720 (bottom row) after infection. Outcomes are sorted in descending order by RR. Conventional study observes positive effects (RR ≥ 1.1) for the vast majority of outcomes: 82.7% on days 30-120, 77.5% on days 120-360, and 85.0% on days 360-720. However, a conventional retrospective study with placebo cases, a similar proportion of outcomes have positive effects: 82.5% on days 30-120, 66.6% on days 120-360, and 54.8% on days 360-720. This suggests that many effects detected by the conventional retrospective study may be spurious and an artifact of selective testing bias. By contrast, our study only detects positive effects for a minority of outcomes: 8.8% on days 30-120, 2.3% on days 120-360, and 1.6% on days 360-720, indicating that many previously reported effects may be overstated. These findings underscore the importance of adjusting for selective testing bias when assessing the long-term health consequences of COVID-19.

**Extended Data Figure 5:**
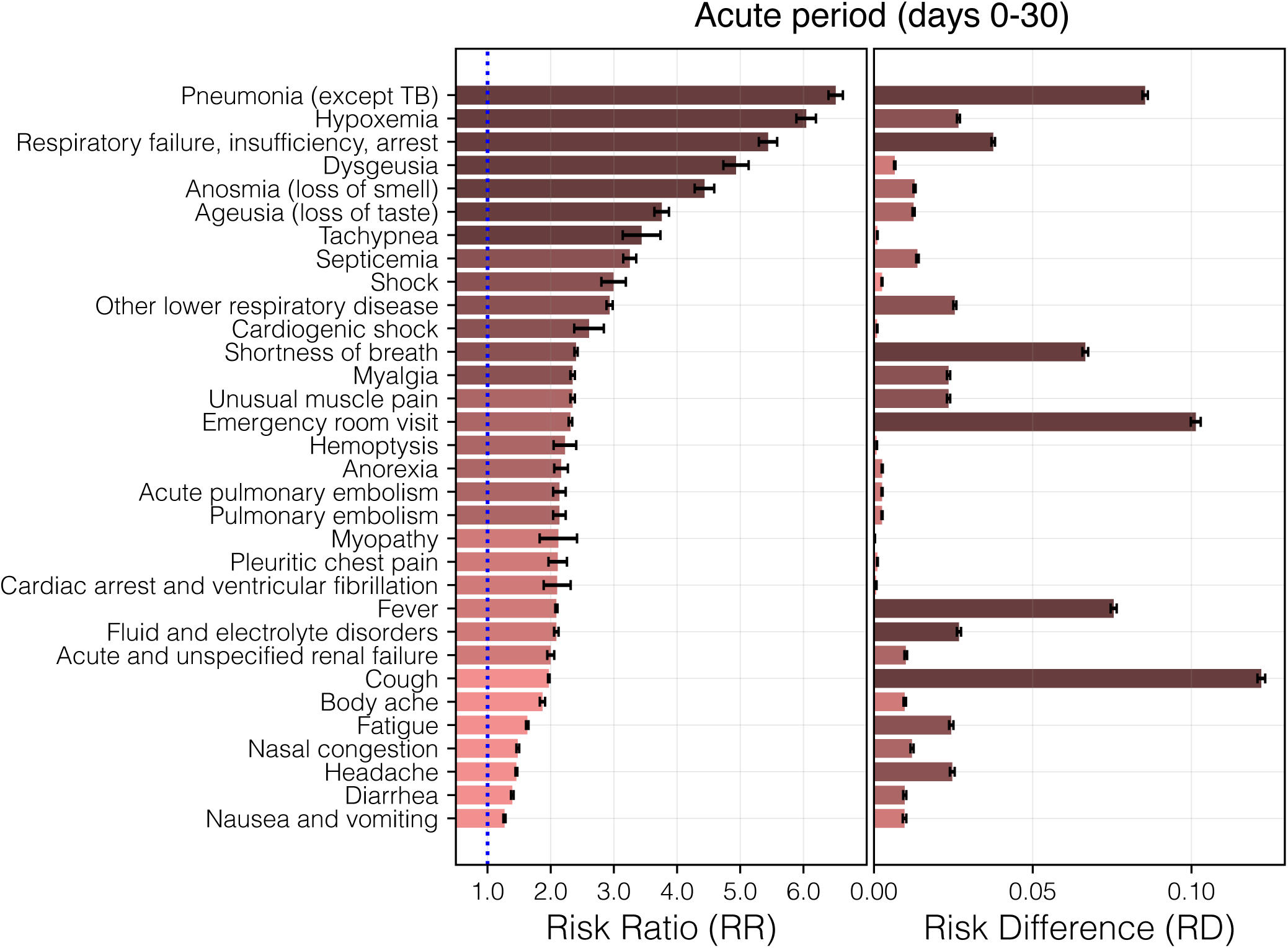
Acute COVID-19 symptoms and sequelae detected by our test-based prospective study. *Left:* relative risk ratios (RRs; Methods M3). *Right:* risk differences (RDs; Methods M3). Common COVID-19 symptoms^92^ and the top 25 most attributable (highest RR) adverse effects of COVID-19 are shown, during the acute period (days 0-30). Bars are colored by RR quintile; small effects (RR *<* 1.1) are not displayed. Our study recovers known acute COVID-19 symptoms from the clinical literature, validating that our study design can be used to an analyze insurance claims in order to infer population-level disease effects.

**Extended Data Figure 6:**
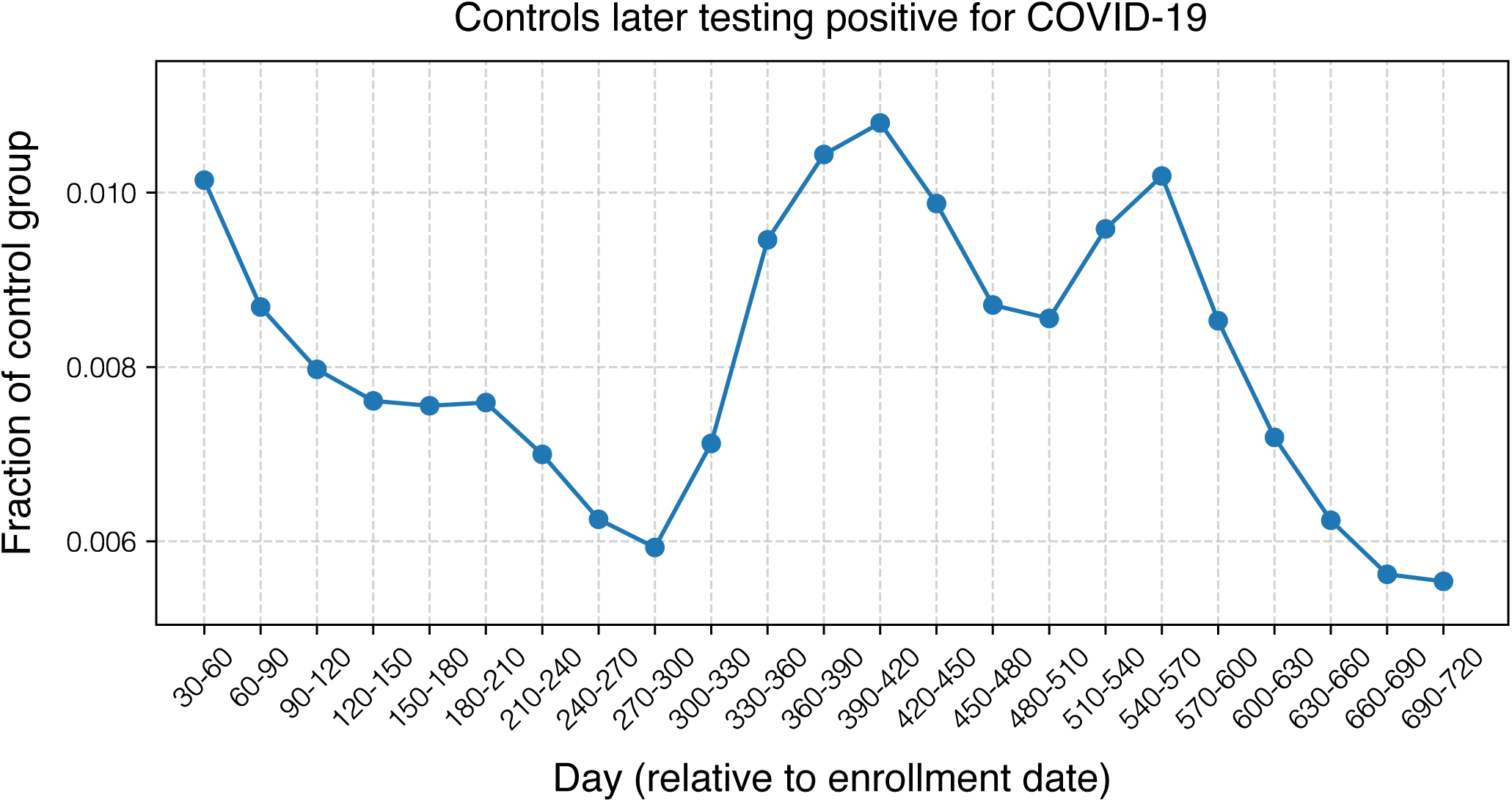
Proportion of controls in the test-based prospective study who later test positive for COVID-19. In our prospective study, individuals are enrolled into the control group at the time of their first negative PCR test. Some controls subsequently test positive for COVID-19. We retain these individuals to avoid selection bias that could arise from using future information to exclude participants, thereby preserving the integrity of a true prospective study design. While this approach prevents bias, it may dilute effect size estimates, making them more conservative. To quantify the potential impact, we plot the proportion of controls who later test positive over time. We observe that fewer than 1.1% of controls test positive in any given month, and 20.2% have tested positive after 720 days of enrollment. This suggests that our effect size estimates may be conservative by up to 20%, depending on the follow-up duration.

