## Supplemental Figures and Table for "Revised estimates of the types and durations of long Covid symptoms based on claims records from 245 Million US patients"

### Supplementary Information

**Robustness check for patient identification.** Supplementary Figure S1: filtering out individuals when there is a likely collision between two patients with the same name, gender, and date of birth. We do so by excluding patients with records occurring at two or more zipcodes (3-digit zip).

**Comparing Population Representations Across ZIP Codes.** We demonstrate minimal bias between the US Census population distribution by ZIP code and our dataset's ZIP-level distribution, ensuring representativeness and robustness for analyses in Supplementary Figure S2.

**Risk ratios for of all 614 outcomes and study designs.** We show the full list of 614 outcomes and the estimated effect across all 3 study designs (conventional retrospective, our test-based prospective, and conventional retrospective with placebo cases): Supplementary Figures S3-S68

**Medical histories across all outcomes.** We show the fraction of patients with relevant claims over time for all diagnostic categories within each experiment:

- Test-based prospective study: Supplementary Figures S69-S93
- Conventional retrospective study: Supplementary Figures S119 - S143
- Conventional retrospective study with placebo cases (negative COVID-19 test recipients): Supplementary Figures S94-S118

**Diagnosis Groups.** We include a comprehensive mapping of diagnosis groups to their corresponding ICD-10 and ICD-9 codes in Supplementary Table S1.

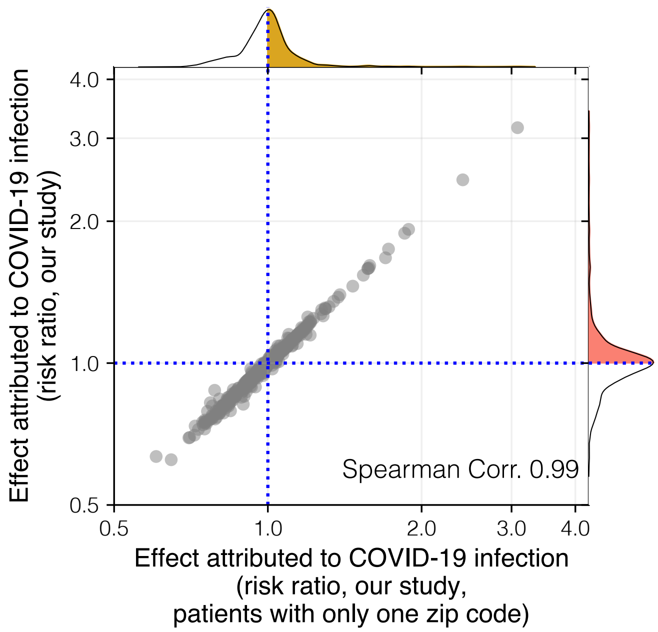


**Supplementary Figure S1: Robustness to patient identity collisions.** To assess whether our findings are affected by potential collisions between different patients sharing the same name, gender, and date of birth, we re-ran our analysis after excluding individuals with more than one zipcode (ZIP-3) in their medical history (a proxy for possible misidentification). The results remained nearly unchanged (Spearman correlation = 0.99), demonstrating that our findings are robust to patient identity resolution in the Komodo dataset.

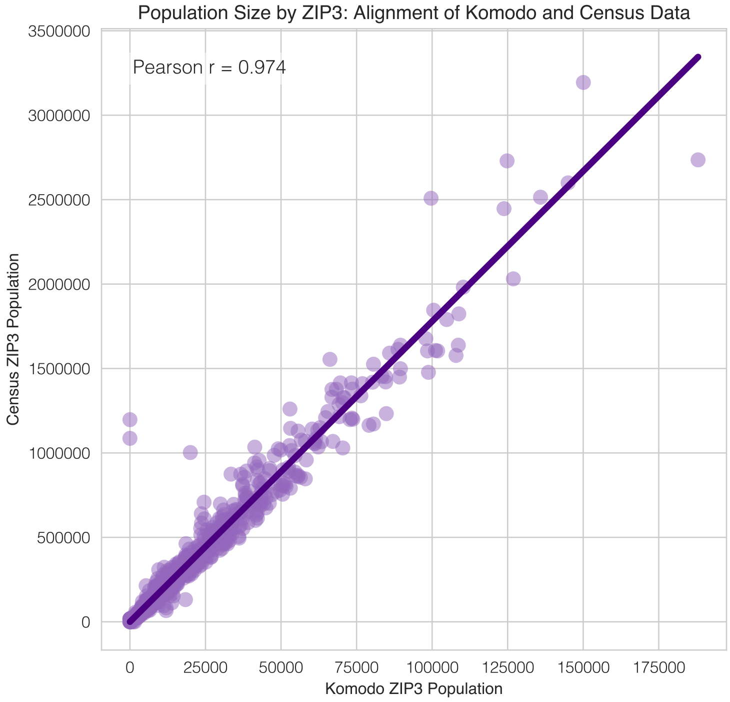


**Supplementary Figure S2: Geographic representativeness of the Komodo dataset**. We compared how people are distributed across ZIP-3 regions in our dataset to how they are distributed in the U.S. Census. The two distributions are highly aligned (Pearson R = 0.97), demonstrating that the Komodo dataset closely reflects the geographic makeup of the U.S. population and is not heavily biased toward certain regions.

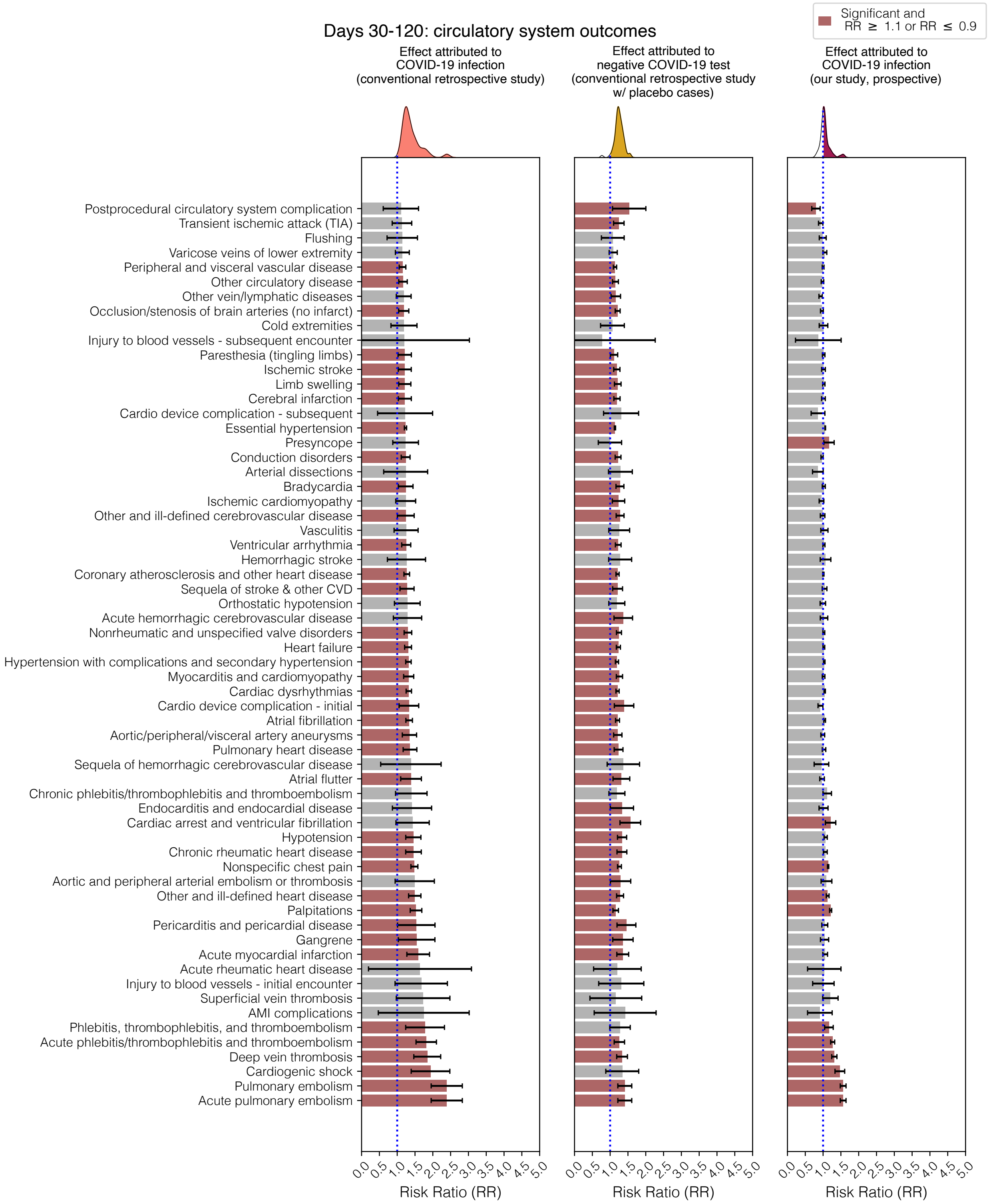


**Supplementary Figure S3:** Risk ratios for all outcomes related to the circulatory system on days 30–120 post-enrollment. Effects are shown for a conventional retrospective study of long COVID (left), our test-based prospective study (right), and a conventional retrospective study with placebo cases (middle). Red bars indicate significant effects ($p<0.05$ with Bonferroni correction for multiple comparisons; two-sided bootstrap) with RR≥1.1 or RR≤0.9.

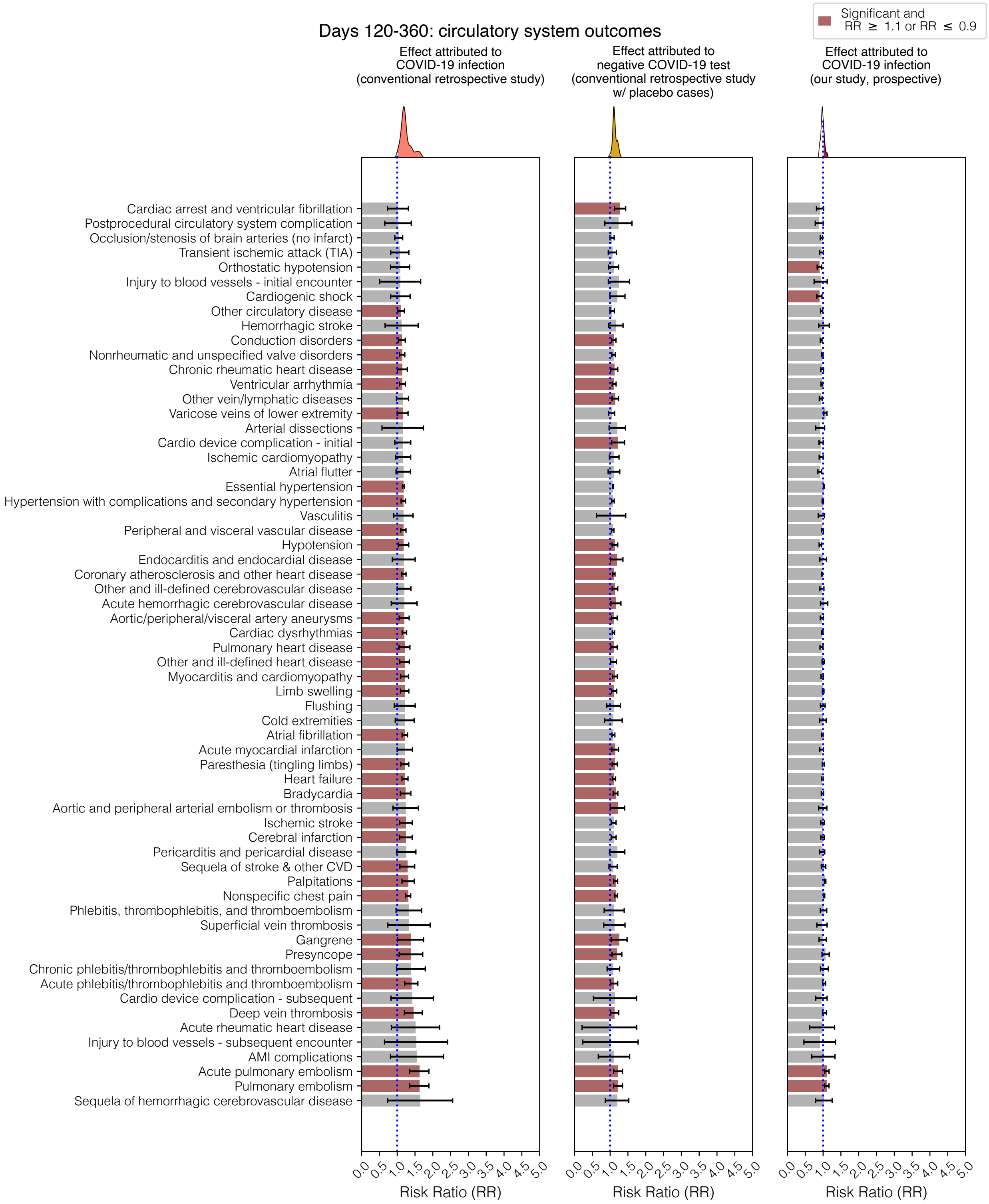


**Supplementary Figure S4:** Risk ratios for all outcomes related to the circulatory system on days 120–360 post-enrollment. Effects are shown for a conventional retrospective study of long COVID (left), our test-based prospective study (right), and a conventional retrospective study with placebo cases (middle). Red bars indicate significant effects ($p<0.05$ with Bonferroni correction for multiple comparisons; two-sided bootstrap) with RR$\geq$1.1 or RR$\leq$0.9.

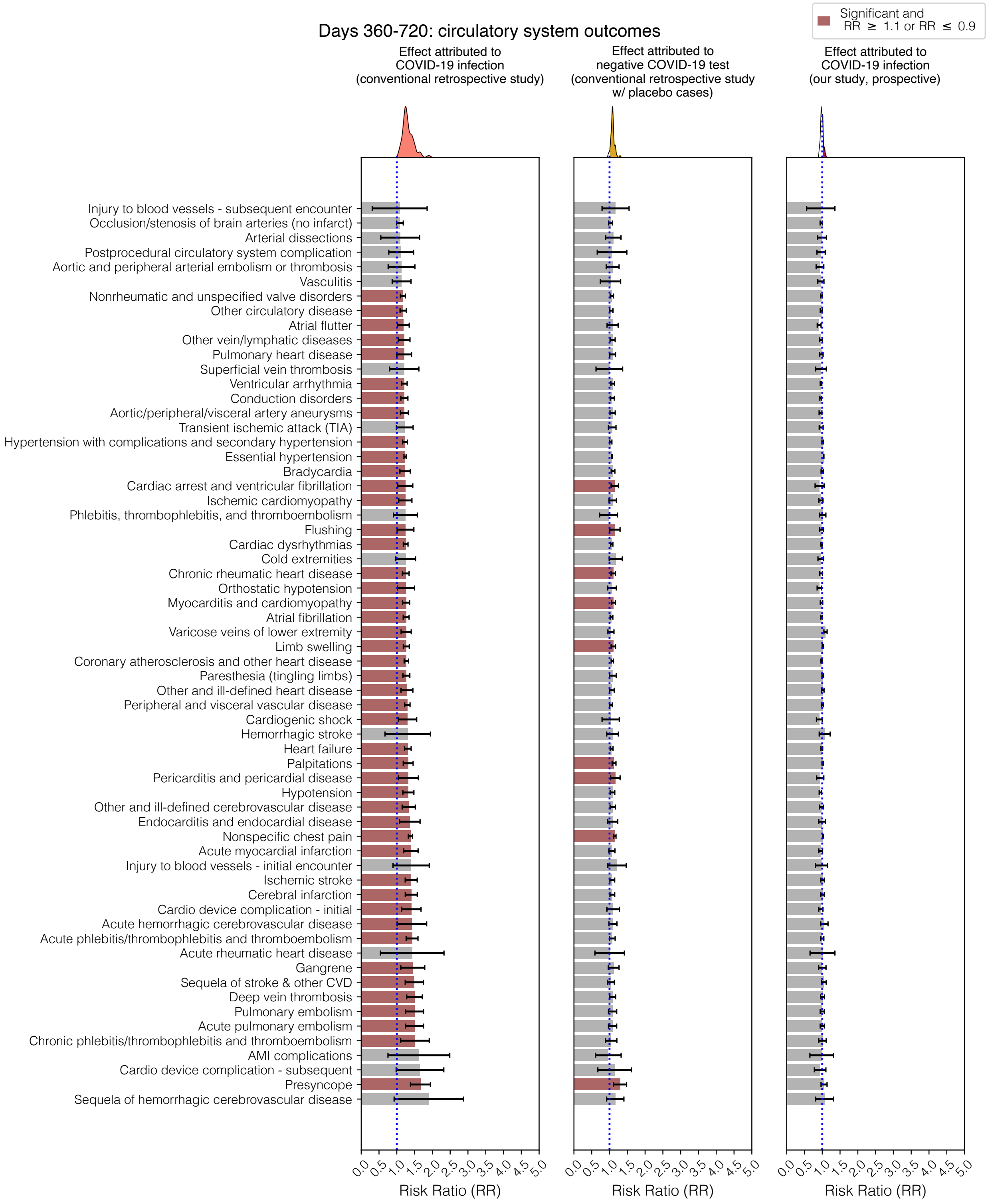


**Supplementary Figure S5:** Risk ratios for all outcomes related to the circulatory system on days 360–720 post-enrollment. Effects are shown for a conventional retrospective study of long COVID (left), our test-based prospective study (right), and a conventional retrospective study with placebo cases (middle). Red bars indicate significant effects ($p<0.05$ with Bonferroni correction for multiple comparisons; two-sided bootstrap) with $RR \geq 1.1$ or $RR \leq 0.9$.

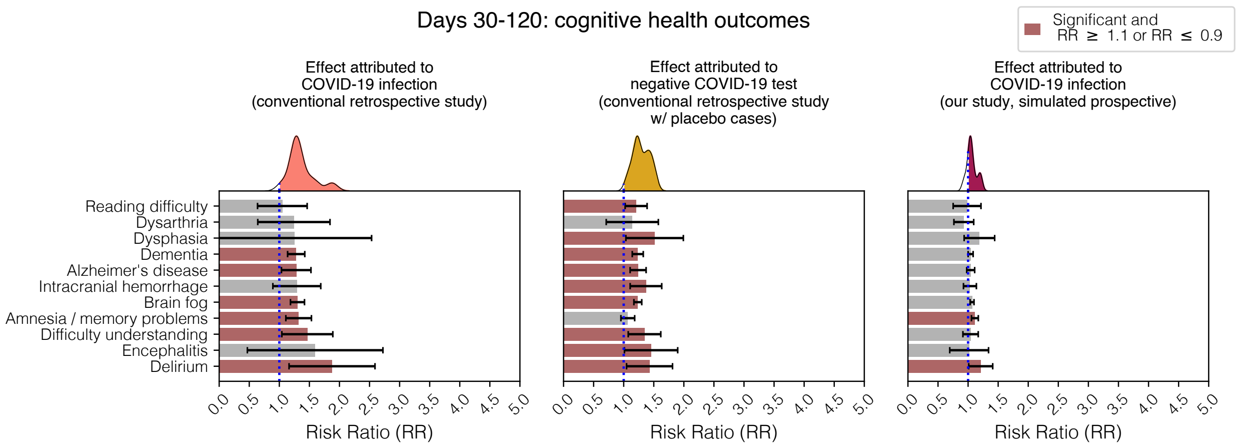


**Supplementary Figure S6:** Risk ratios for all outcomes related to cognitive health on days 30–120 post-enrollment. Effects are shown for a conventional retrospective study of long COVID (left), our test-based prospective study (right), and a conventional retrospective study with placebo cases (middle). Red bars indicate significant effects ($p<0.05$ with Bonferroni correction for multiple comparisons; two-sided bootstrap) with $RR \geq 1.1$ or $RR \leq 0.9$.

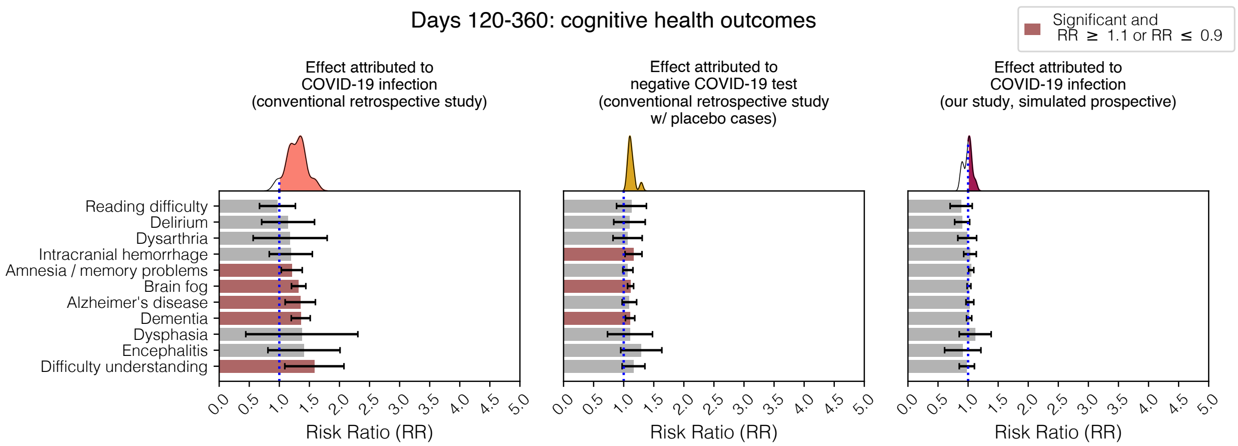


**Supplementary Figure S7:** Risk ratios for all outcomes related to cognitive health on days 120–360 post-enrollment. Effects are shown for a conventional retrospective study of long COVID (left), our test-based prospective study (right), and a conventional retrospective study with placebo cases (middle). Red bars indicate significant effects ($p<0.05$ with Bonferroni correction for multiple comparisons; two-sided bootstrap) with $RR \geq 1.1$ or $RR \leq 0.9$.

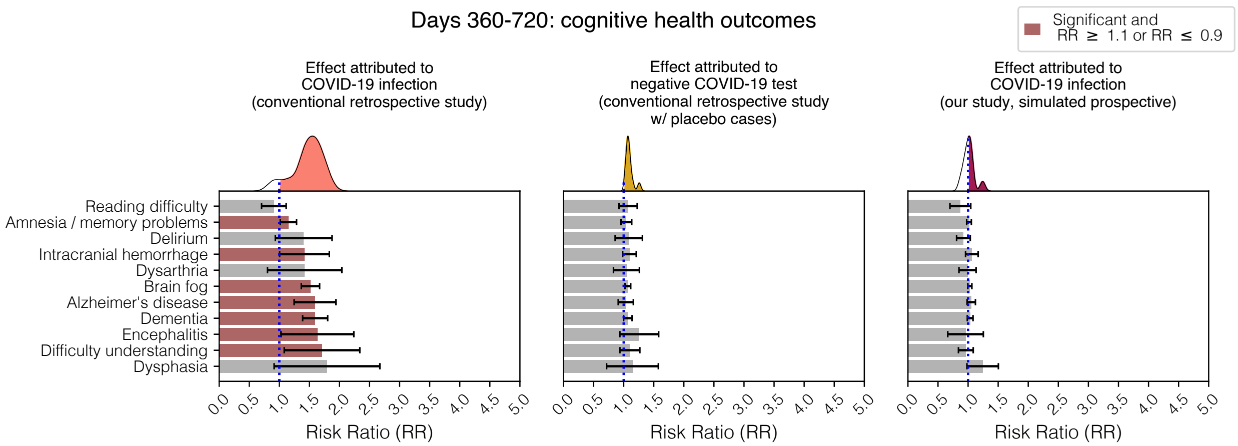


**Supplementary Figure S8:** Risk ratios for all outcomes related to cognitive health on days 360–720 post-enrollment. Effects are shown for a conventional retrospective study of long COVID (left), our test-based prospective study (right), and a conventional retrospective study with placebo cases (middle). Red bars indicate significant effects ($p<0.05$ with Bonferroni correction for multiple comparisons; two-sided bootstrap) with $RR \geq 1.1$ or $RR \leq 0.9$.

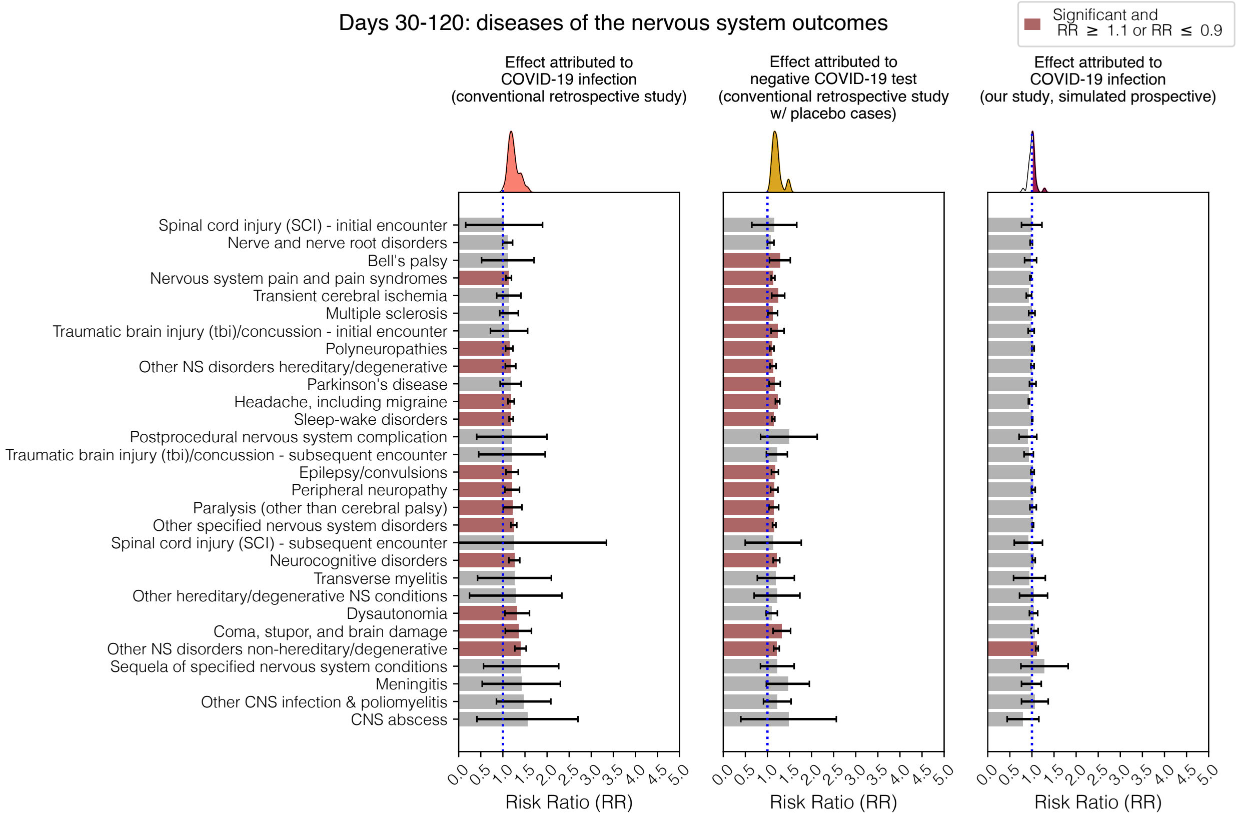


**Supplementary Figure S9:** Risk ratios for all outcomes related to diseases of the nervous system on days 30–120 post-enrollment. Effects are shown for a conventional retrospective study of long COVID (left), our test-based prospective study (right), and a conventional retrospective study with placebo cases (middle). Red bars indicate significant effects ($p<0.05$ with Bonferroni correction for multiple comparisons; two-sided bootstrap) with $RR \geq 1.1$ or $RR \leq 0.9$.

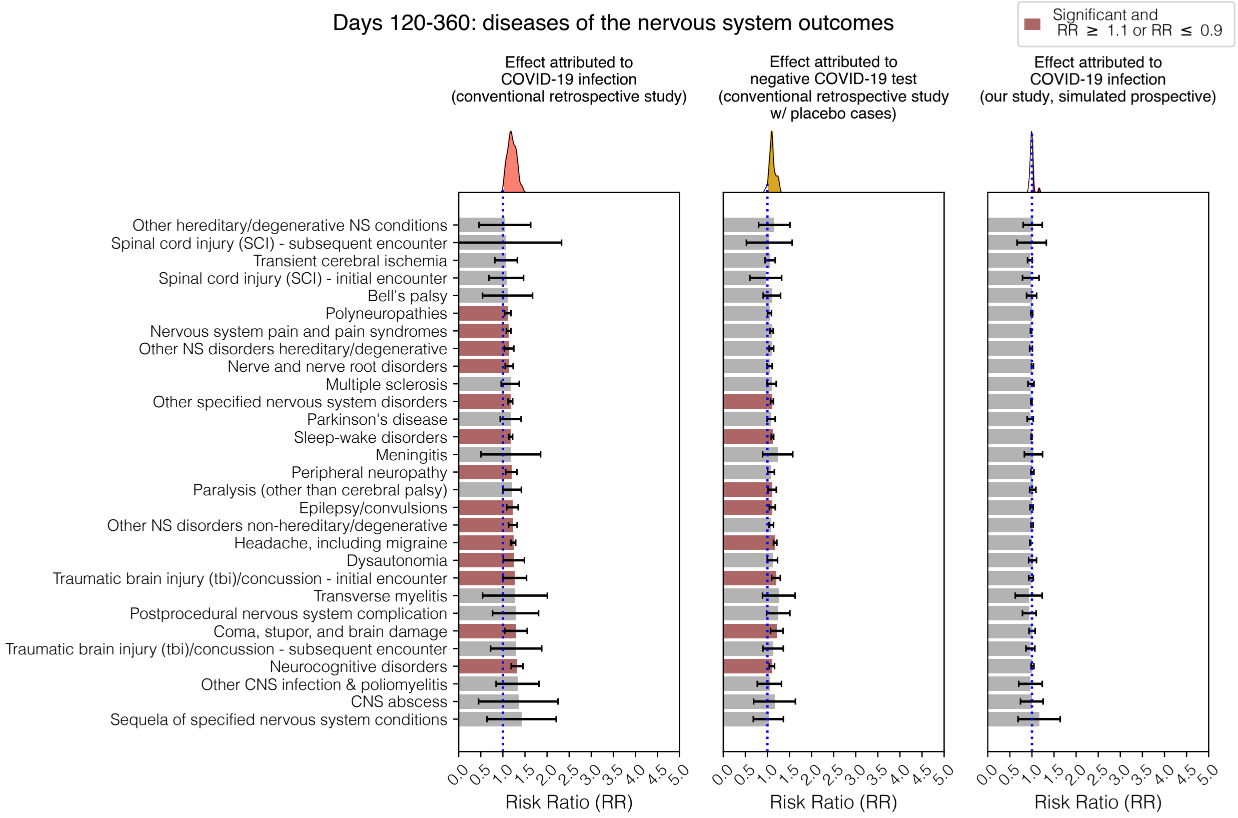


**Supplementary Figure S10:** Risk ratios for all outcomes related to diseases of the nervous system on days 120–360 post-enrollment. Effects are shown for a conventional retrospective study of long COVID (left), our test-based prospective study (right), and a conventional retrospective study with placebo cases (middle). Red bars indicate significant effects ($p<0.05$ with Bonferroni correction for multiple comparisons; two-sided bootstrap) with $RR \geq 1.1$ or $RR \leq 0.9$.

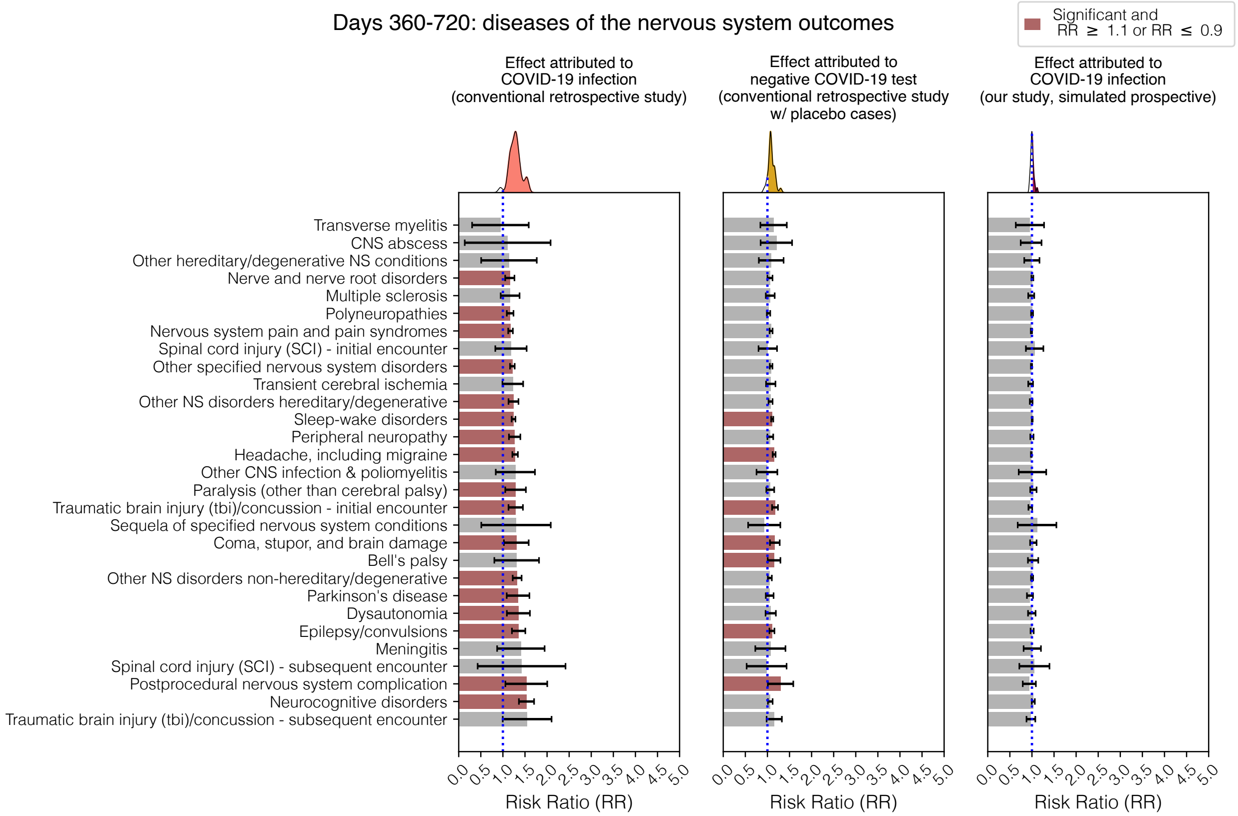


**Supplementary Figure S11:** Risk ratios for all outcomes related to diseases of the nervous system on days 360–720 post-enrollment. Effects are shown for a conventional retrospective study of long COVID (left), our test-based prospective study (right), and a conventional retrospective study with placebo cases (middle). Red bars indicate significant effects (p<0.05 with Bonferroni correction for multiple comparisons; two-sided bootstrap) with RR≥1.1 or RR≤0.9.

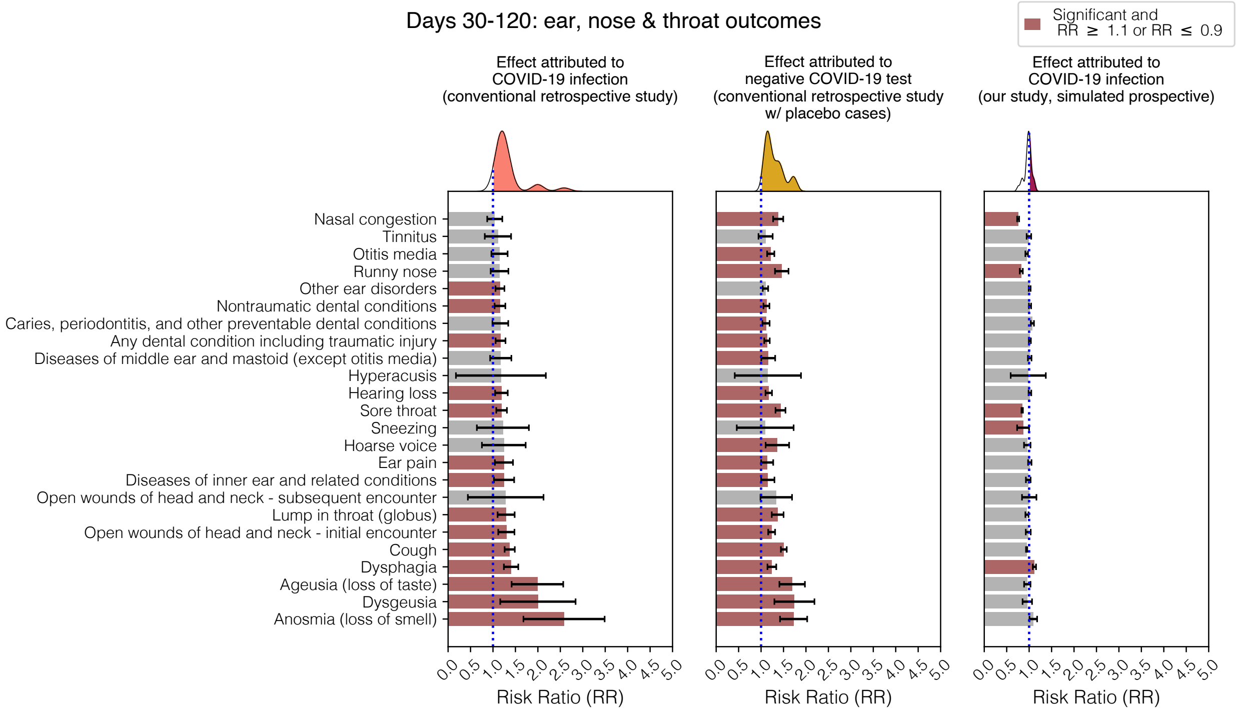


**Supplementary Figure S12:** Risk ratios for all outcomes related to ear, nose & throat on days 30–120 post-enrollment. Effects are shown for a conventional retrospective study of long COVID (left), our test-based prospective study (right), and a conventional retrospective study with placebo cases (middle). Red bars indicate significant effects ($p<0.05$ with Bonferroni correction for multiple comparisons; two-sided bootstrap) with $RR \geq 1.1$ or $RR \leq 0.9$.

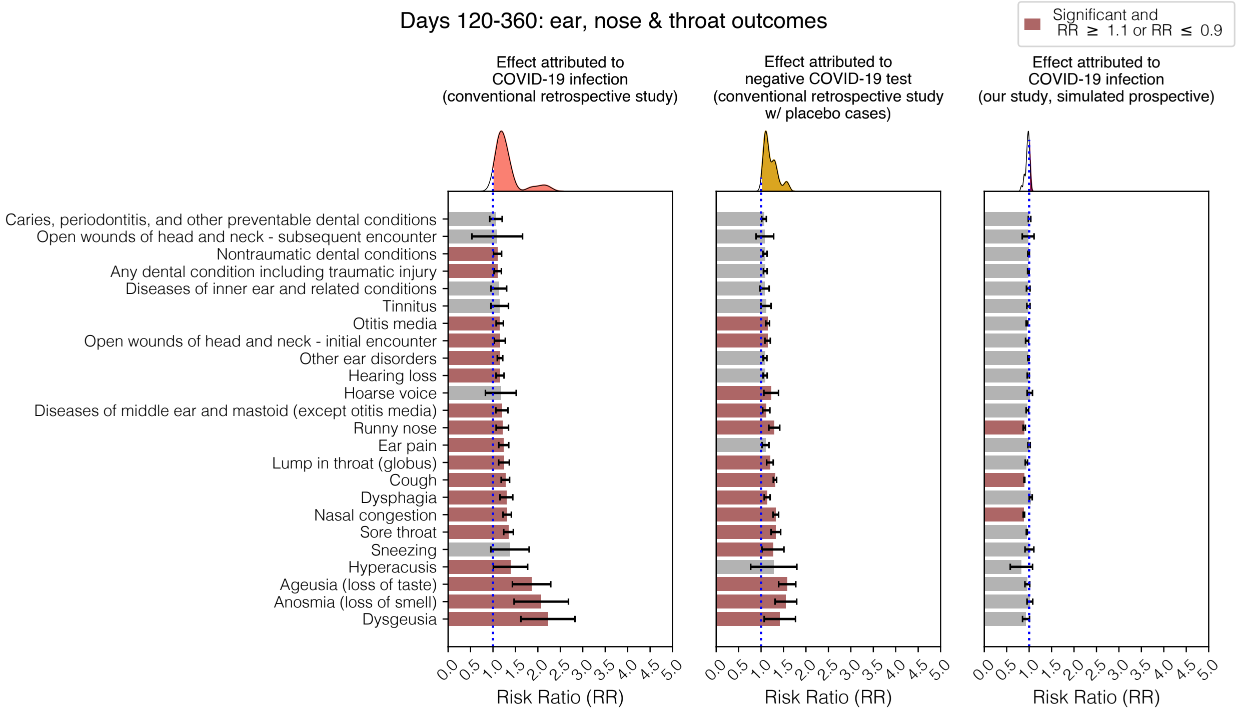


**Supplementary Figure S13:** Risk ratios for all outcomes related to ear, nose & throat on days 120–360 post-enrollment. Effects are shown for a conventional retrospective study of long COVID (left), our test-based prospective study (right), and a conventional retrospective study with placebo cases (middle). Red bars indicate significant effects ($p<0.05$ with Bonferroni correction for multiple comparisons; two-sided bootstrap) with $RR \geq 1.1$ or $RR \leq 0.9$.

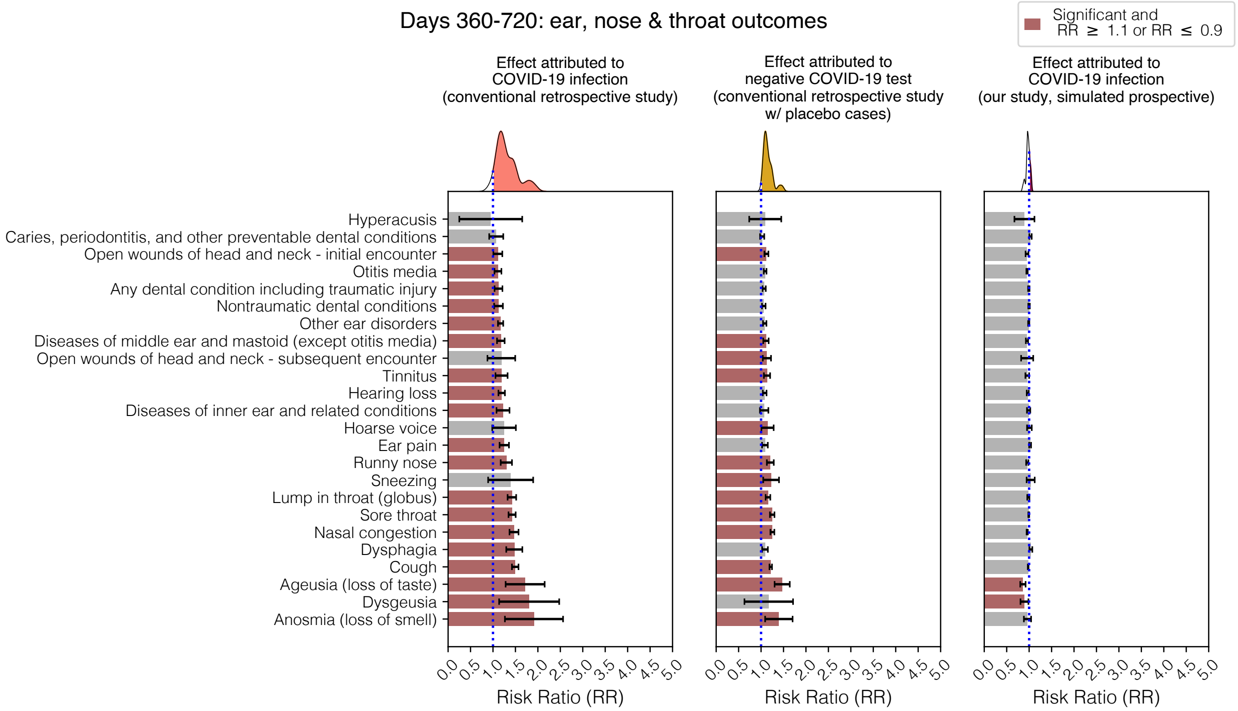


**Supplementary Figure S14:** Risk ratios for all outcomes related to ear, nose & throat on days 360–720 post-enrollment. Effects are shown for a conventional retrospective study of long COVID (left), our test-based prospective study (right), and a conventional retrospective study with placebo cases (middle). Red bars indicate significant effects ($p<0.05$ with Bonferroni correction for multiple comparisons; two-sided bootstrap) with $RR \geq 1.1$ or $RR \leq 0.9$.

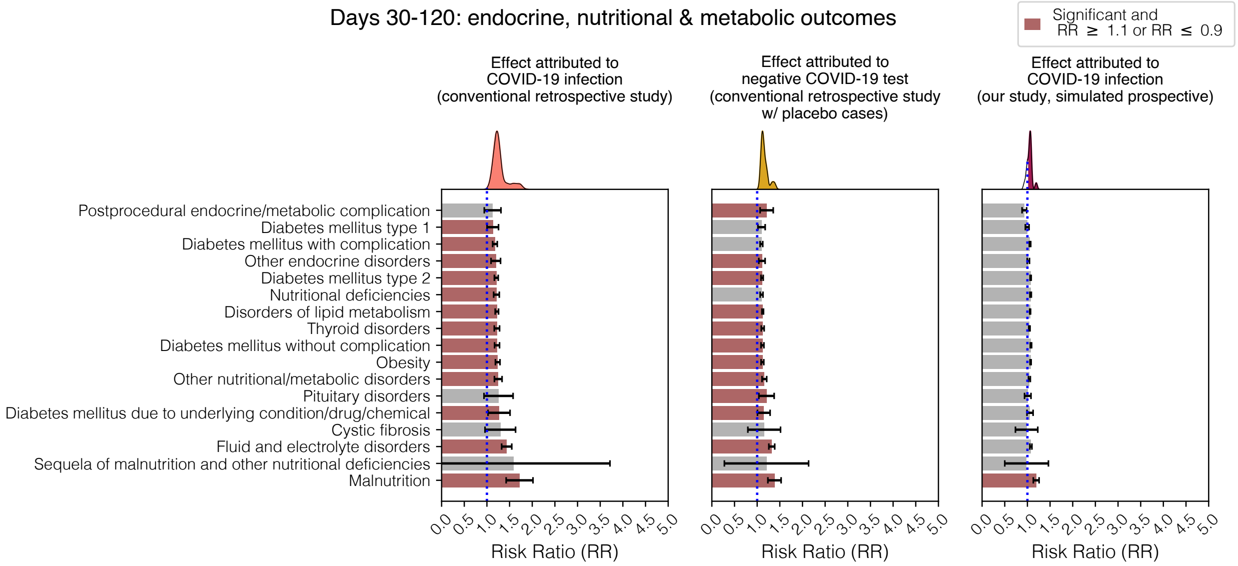


**Supplementary Figure S15:** Risk ratios for all outcomes related to the endocrine, nutritional & metabolic system on days 30–120 post-enrollment. Effects are shown for a conventional retrospective study of long COVID (left), our test-based prospective study (right), and a conventional retrospective study with placebo cases (middle). Red bars indicate significant effects ($p<0.05$ with Bonferroni correction for multiple comparisons; two-sided bootstrap) with $RR \geq 1.1$ or $RR \leq 0.9$.

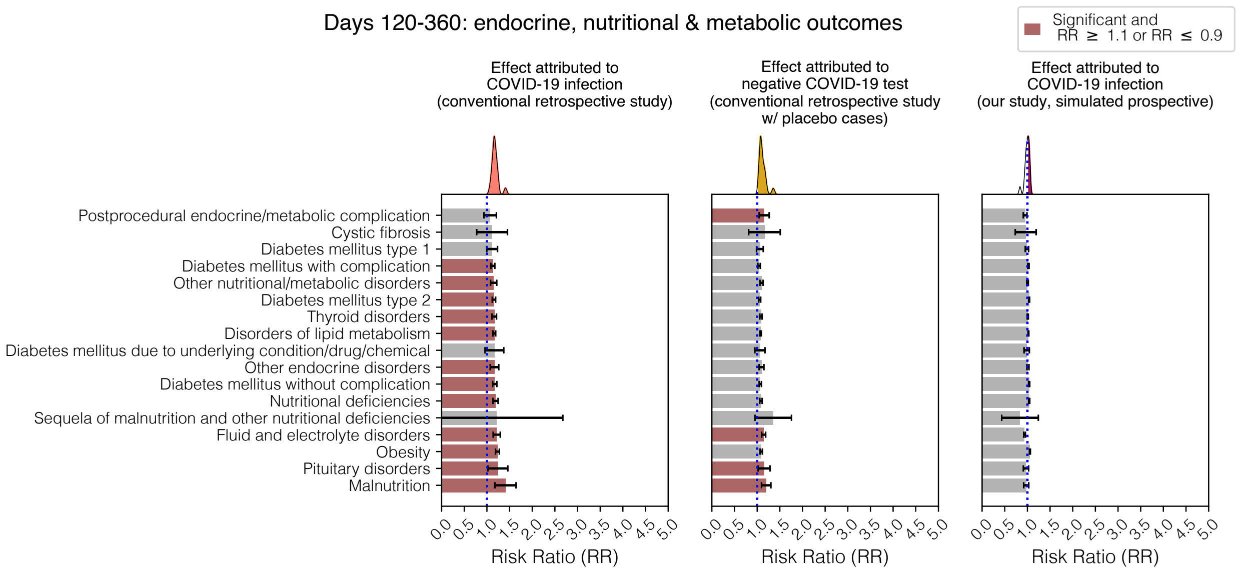


**Supplementary Figure S16:** Risk ratios for all outcomes related to the endocrine, nutritional & metabolic system on days 120–360 post-enrollment. Effects are shown for a conventional retrospective study of long COVID (left), our test-based prospective study (right), and a conventional retrospective study with placebo cases (middle). Red bars indicate significant effects (p<0.05 with Bonferroni correction for multiple comparisons; two-sided bootstrap) with RR≥1.1 or RR≤0.9.

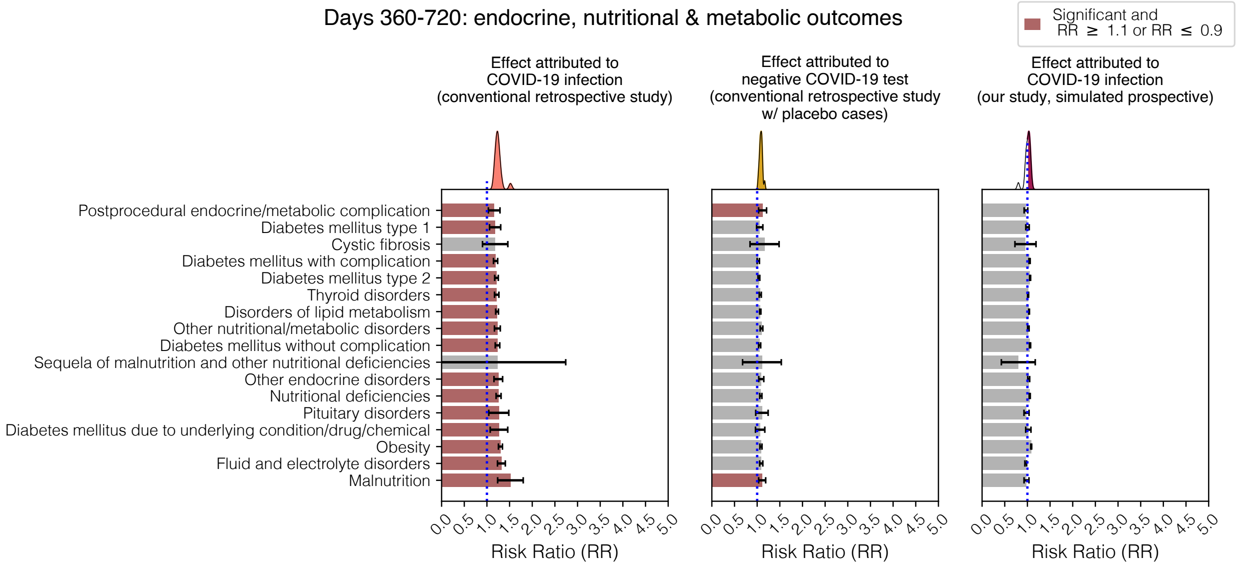


**Supplementary Figure S17:** Risk ratios for all outcomes related to the endocrine, nutritional & metabolic system on days 360–720 post-enrollment. Effects are shown for a conventional retrospective study of long COVID (left), our test-based prospective study (right), and a conventional retrospective study with placebo cases (middle). Red bars indicate significant effects ($p<0.05$ with Bonferroni correction for multiple comparisons; two-sided bootstrap) with $RR \geq 1.1$ or $RR \leq 0.9$.

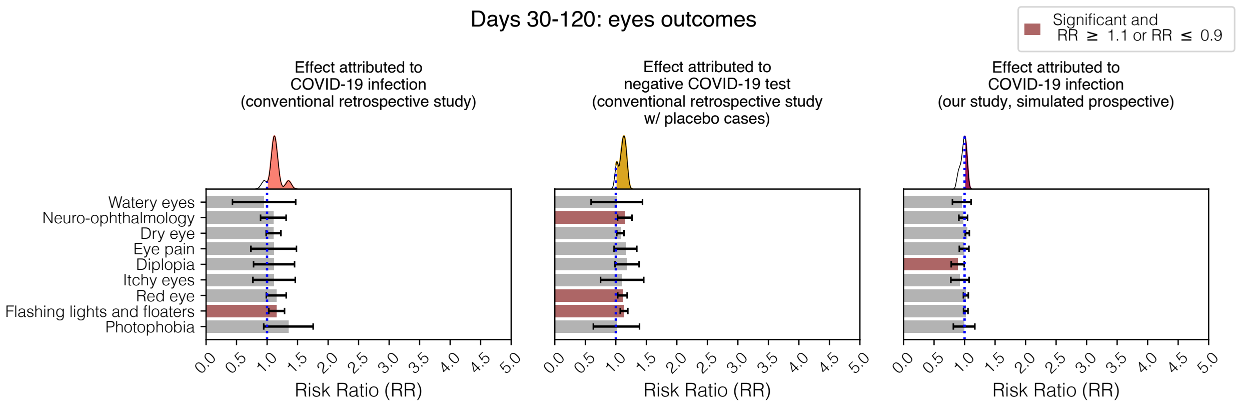


**Supplementary Figure S18:** Risk ratios for all outcomes related to eyes on days 30–120 post-enrollment. Effects are shown for a conventional retrospective study of long COVID (left), our test-based prospective study (right), and a conventional retrospective study with placebo cases (middle). Red bars indicate significant effects ($p<0.05$ with Bonferroni correction for multiple comparisons; two-sided bootstrap) with $RR \geq 1.1$ or $RR \leq 0.9$.

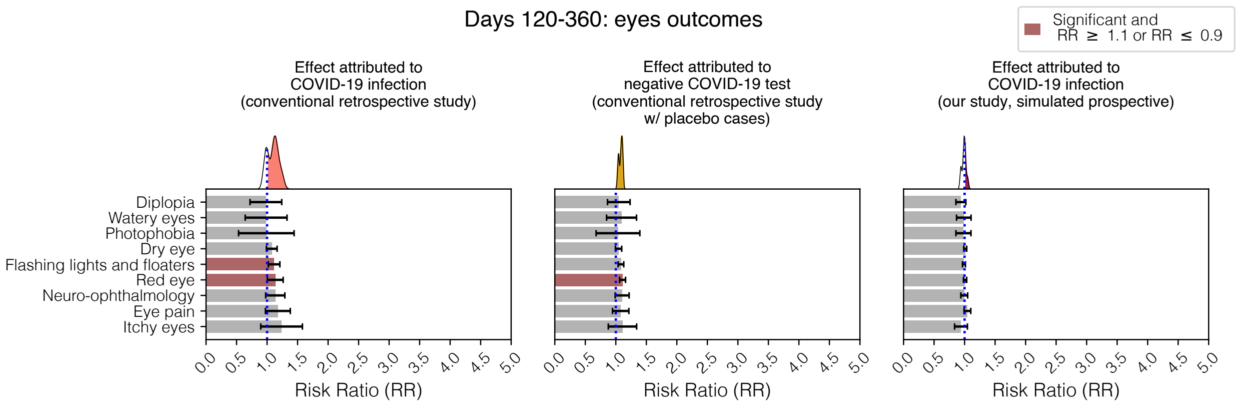


**Supplementary Figure S19:** Risk ratios for all outcomes related to eyes on days 120–360 post-enrollment. Effects are shown for a conventional retrospective study of long COVID (left), our test-based prospective study (right), and a conventional retrospective study with placebo cases (middle). Red bars indicate significant effects ($p<0.05$ with Bonferroni correction for multiple comparisons; two-sided bootstrap) with $RR \geq 1.1$ or $RR \leq 0.9$.

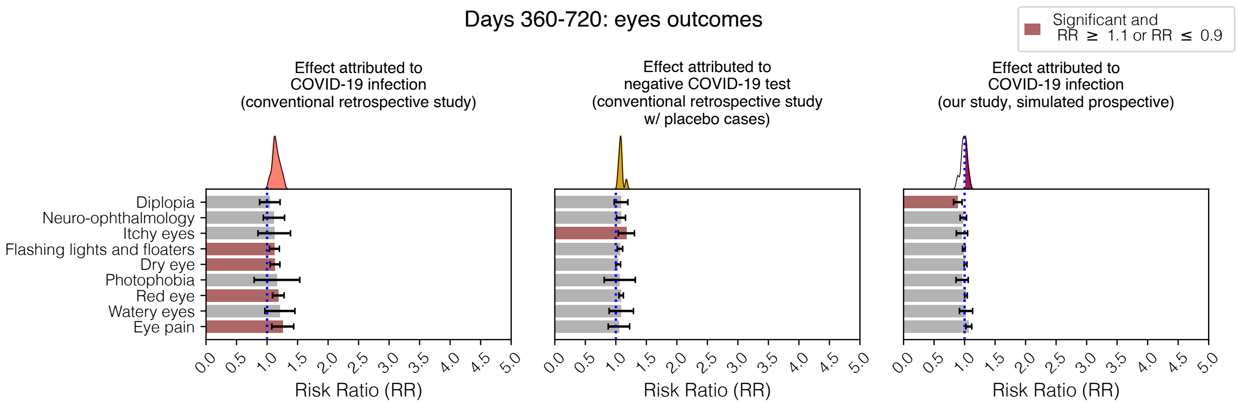


**Supplementary Figure S20:** Risk ratios for all outcomes related to eyes on days 360–720 post-enrollment. Effects are shown for a conventional retrospective study of long COVID (left), our test-based prospective study (right), and a conventional retrospective study with placebo cases (middle). Red bars indicate significant effects ($p<0.05$ with Bonferroni correction for multiple comparisons; two-sided bootstrap) with $RR \geq 1.1$ or $RR \leq 0.9$.

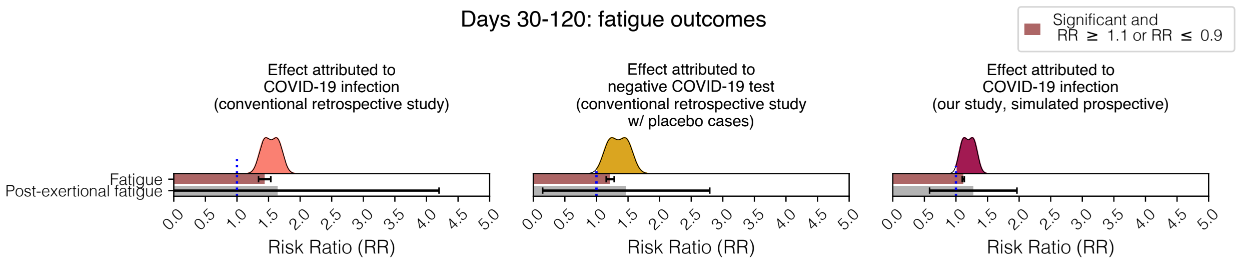


**Supplementary Figure S21:** Risk ratios for all outcomes related to fatigue on days 30–120 post-enrollment. Effects are shown for a conventional retrospective study of long COVID (left), our test-based prospective study (right), and a conventional retrospective study with placebo cases (middle). Red bars indicate significant effects ($p<0.05$ with Bonferroni correction for multiple comparisons; two-sided bootstrap) with $RR \geq 1.1$ or $RR \leq 0.9$.

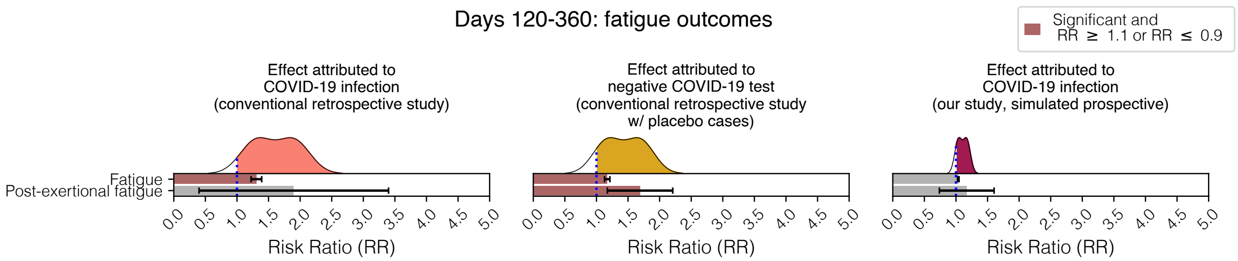


**Supplementary Figure S22:** Risk ratios for all outcomes related to fatigue on days 120–360 post-enrollment. Effects are shown for a conventional retrospective study of long COVID (left), our test-based prospective study (right), and a conventional retrospective study with placebo cases (middle). Red bars indicate significant effects ($p<0.05$ with Bonferroni correction for multiple comparisons; two-sided bootstrap) with $RR \geq 1.1$ or $RR \leq 0.9$.

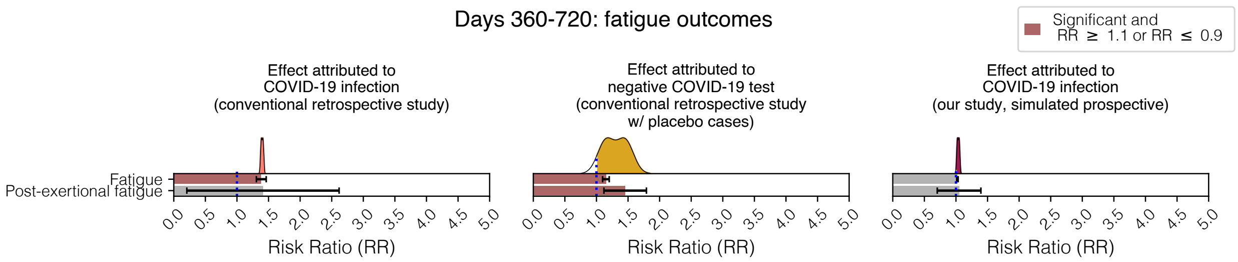


**Supplementary Figure S23:** Risk ratios for all outcomes related to fatigue on days 360–720 post-enrollment. Effects are shown for a conventional retrospective study of long COVID (left), our test-based prospective study (right), and a conventional retrospective study with placebo cases (middle). Red bars indicate significant effects ($p<0.05$ with Bonferroni correction for multiple comparisons; two-sided bootstrap) with $RR \geq 1.1$ or $RR \leq 0.9$.

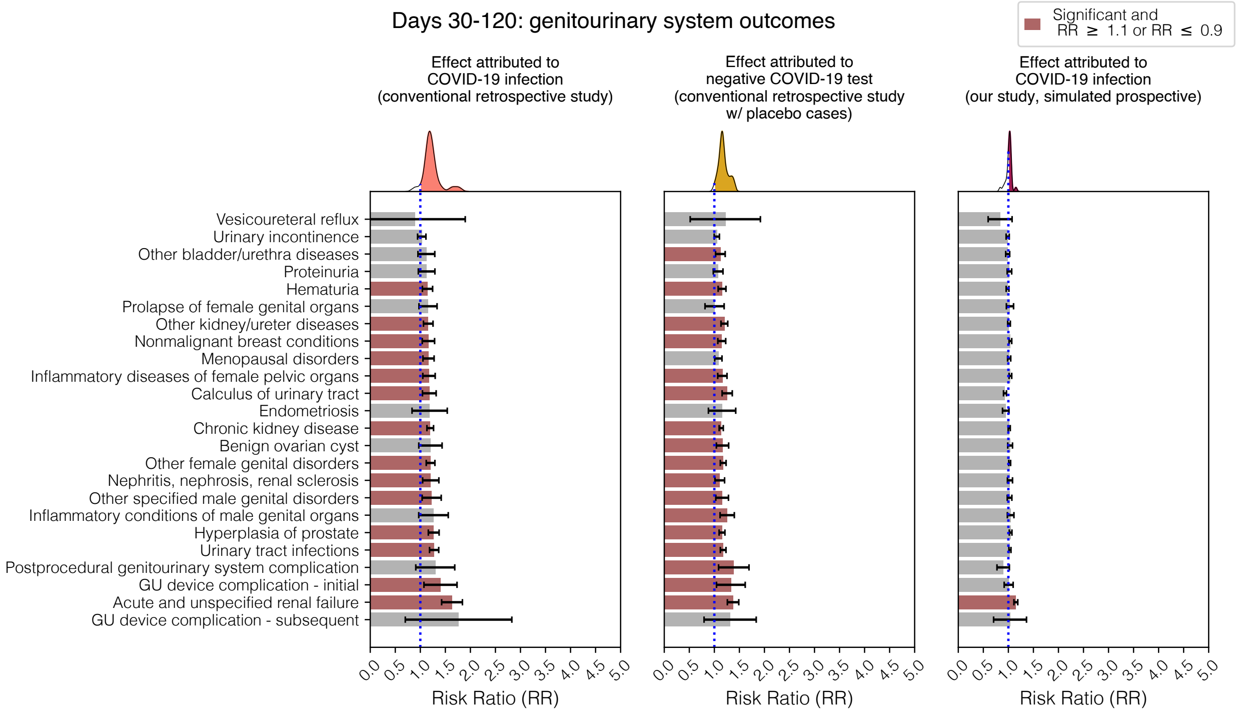


**Supplementary Figure S24:** Risk ratios for all outcomes related to the genitourinary system on days 30–120 post-enrollment. Effects are shown for a conventional retrospective study of long COVID (left), our test-based prospective study (right), and a conventional retrospective study with placebo cases (middle). Red bars indicate significant effects ($p<0.05$ with Bonferroni correction for multiple comparisons; two-sided bootstrap) with RR$\geq$1.1 or RR$\leq$0.9.

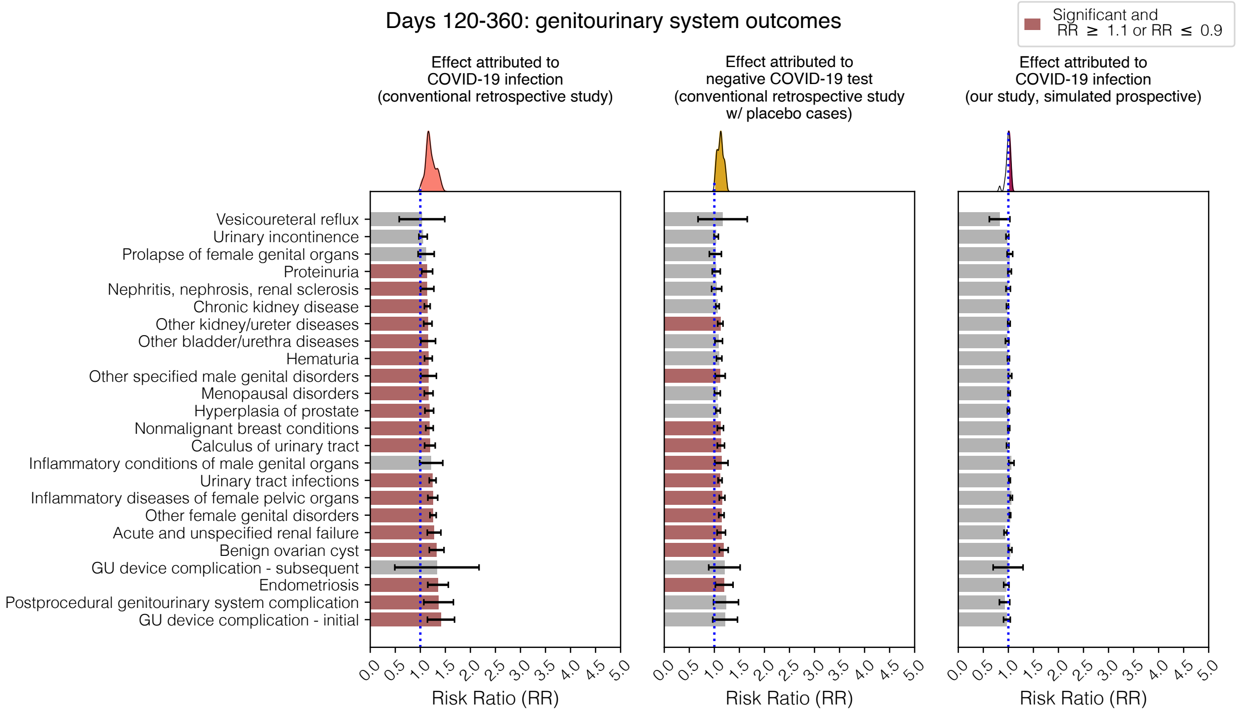


**Supplementary Figure S25:** Risk ratios for all outcomes related to the genitourinary system on days 120–360 post-enrollment. Effects are shown for a conventional retrospective study of long COVID (left), our test-based prospective study (right), and a conventional retrospective study with placebo cases (middle). Red bars indicate significant effects ($p<0.05$ with Bonferroni correction for multiple comparisons; two-sided bootstrap) with RR$\geq$1.1 or RR$\leq$0.9.

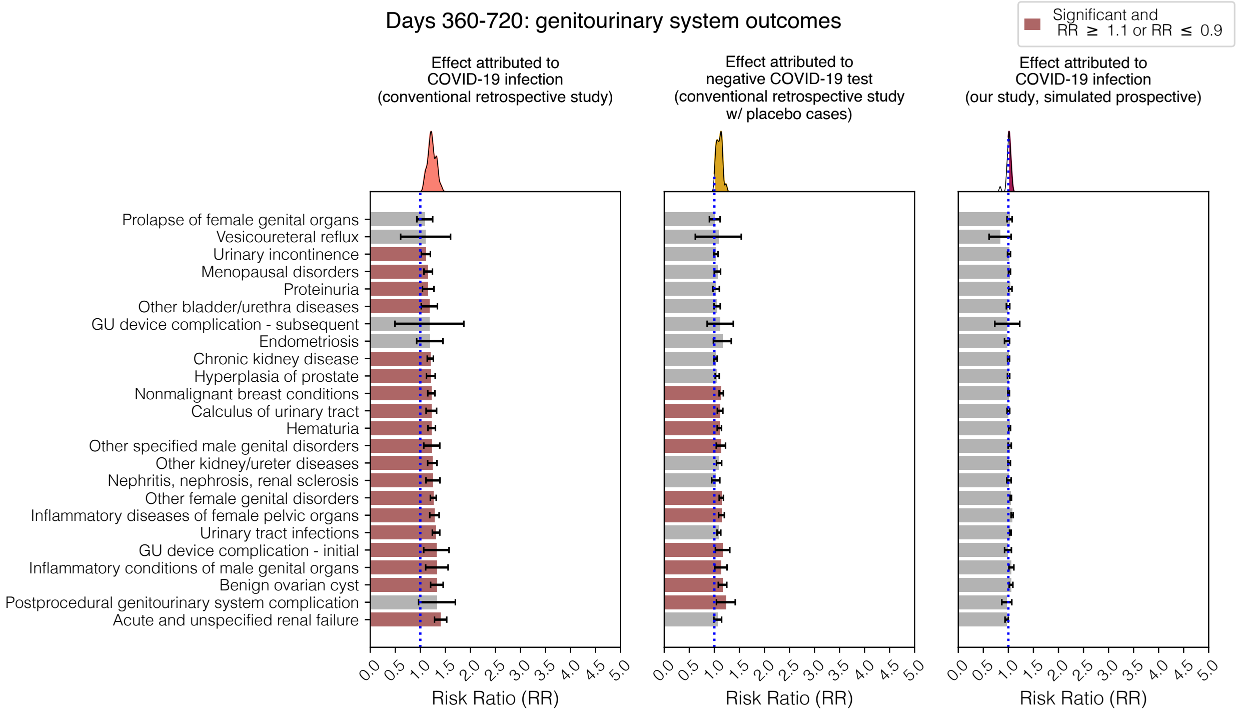


**Supplementary Figure S26:** Risk ratios for all outcomes related to the genitourinary system on days 360–720 post-enrollment. Effects are shown for a conventional retrospective study of long COVID (left), our test-based prospective study (right), and a conventional retrospective study with placebo cases (middle). Red bars indicate significant effects (p<0.05 with Bonferroni correction for multiple comparisons; two-sided bootstrap) with RR≥1.1 or RR≤0.9.

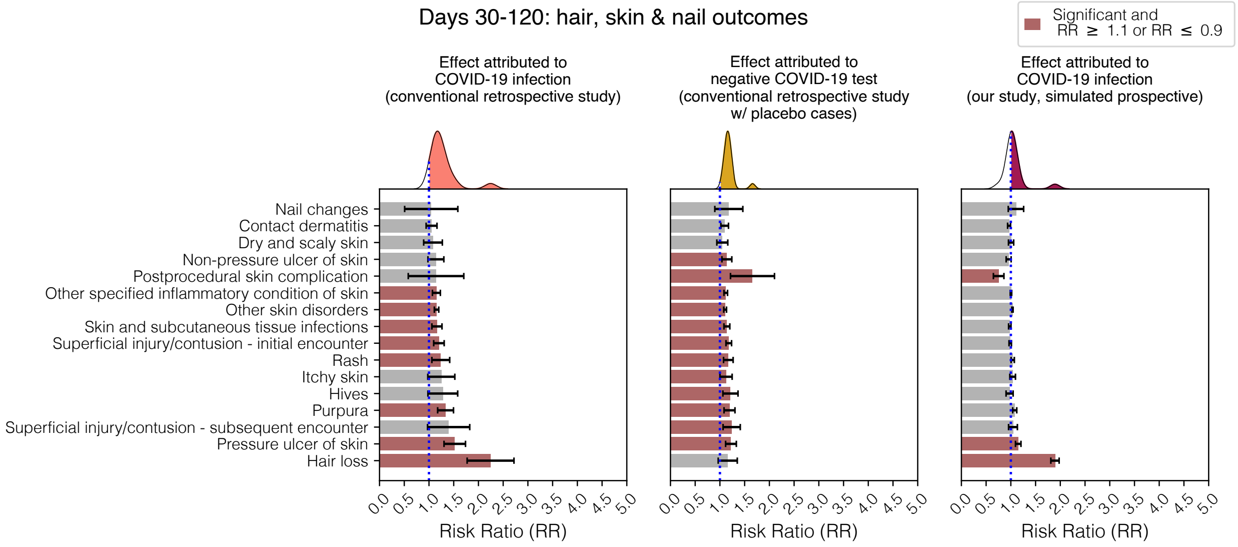


**Supplementary Figure S27:** Risk ratios for all outcomes related to hair, skin & nail on days 30–120 post-enrollment. Effects are shown for a conventional retrospective study of long COVID (left), our test-based prospective study (right), and a conventional retrospective study with placebo cases (middle). Red bars indicate significant effects ($p<0.05$ with Bonferroni correction for multiple comparisons; two-sided bootstrap) with $RR \geq 1.1$ or $RR \leq 0.9$.

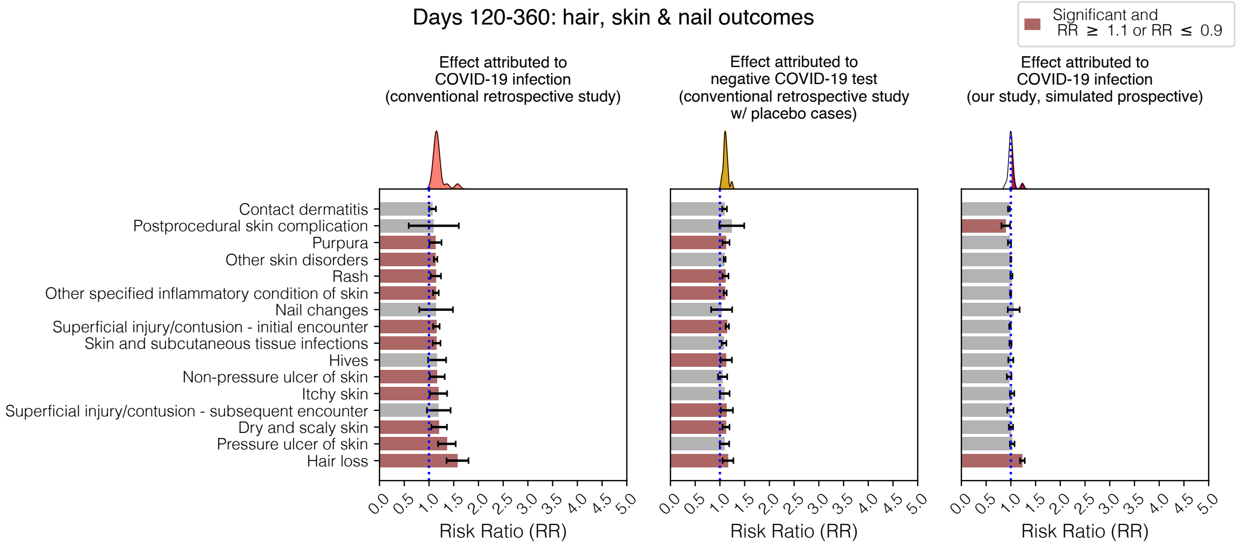


**Supplementary Figure S28:** Risk ratios for all outcomes related to hair, skin & nail on days 120–360 post-enrollment. Effects are shown for a conventional retrospective study of long COVID (left), our test-based prospective study (right), and a conventional retrospective study with placebo cases (middle). Red bars indicate significant effects ($p<0.05$ with Bonferroni correction for multiple comparisons; two-sided bootstrap) with RR$\geq$1.1 or RR$\leq$0.9.

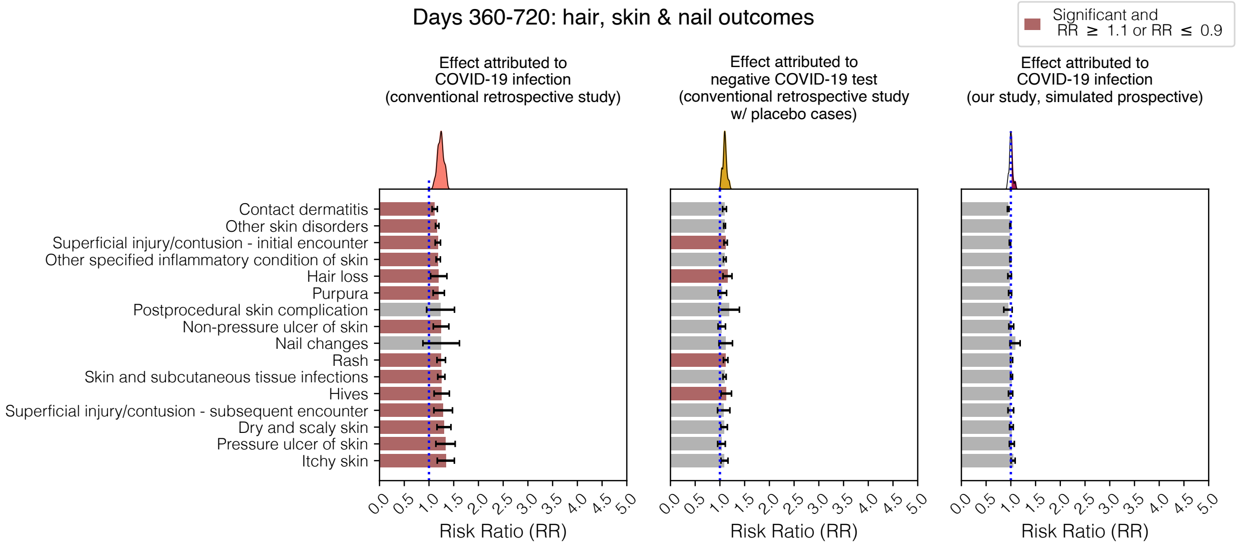


**Supplementary Figure S29:** Risk ratios for all outcomes related to hair, skin & nail on days 360–720 post-enrollment. Effects are shown for a conventional retrospective study of long COVID (left), our test-based prospective study (right), and a conventional retrospective study with placebo cases (middle). Red bars indicate significant effects ($p<0.05$ with Bonferroni correction for multiple comparisons; two-sided bootstrap) with RR$\geq$1.1 or RR$\leq$0.9.

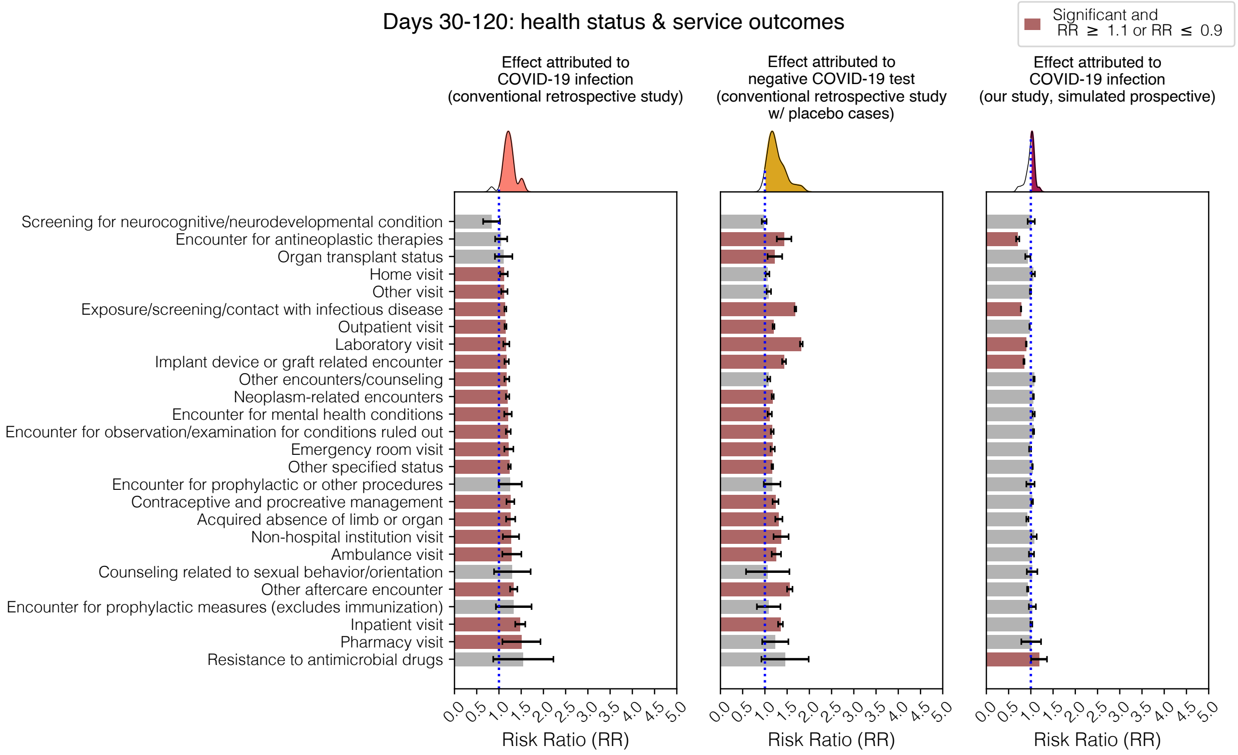


**Supplementary Figure S30:** Risk ratios for all outcomes related to health status & service on days 30–120 post-enrollment. Effects are shown for a conventional retrospective study of long COVID (left), our test-based prospective study (right), and a conventional retrospective study with placebo cases (middle). Red bars indicate significant effects ($p<0.05$ with Bonferroni correction for multiple comparisons; two-sided bootstrap) with $RR\geq1.1$ or $RR\leq0.9$.

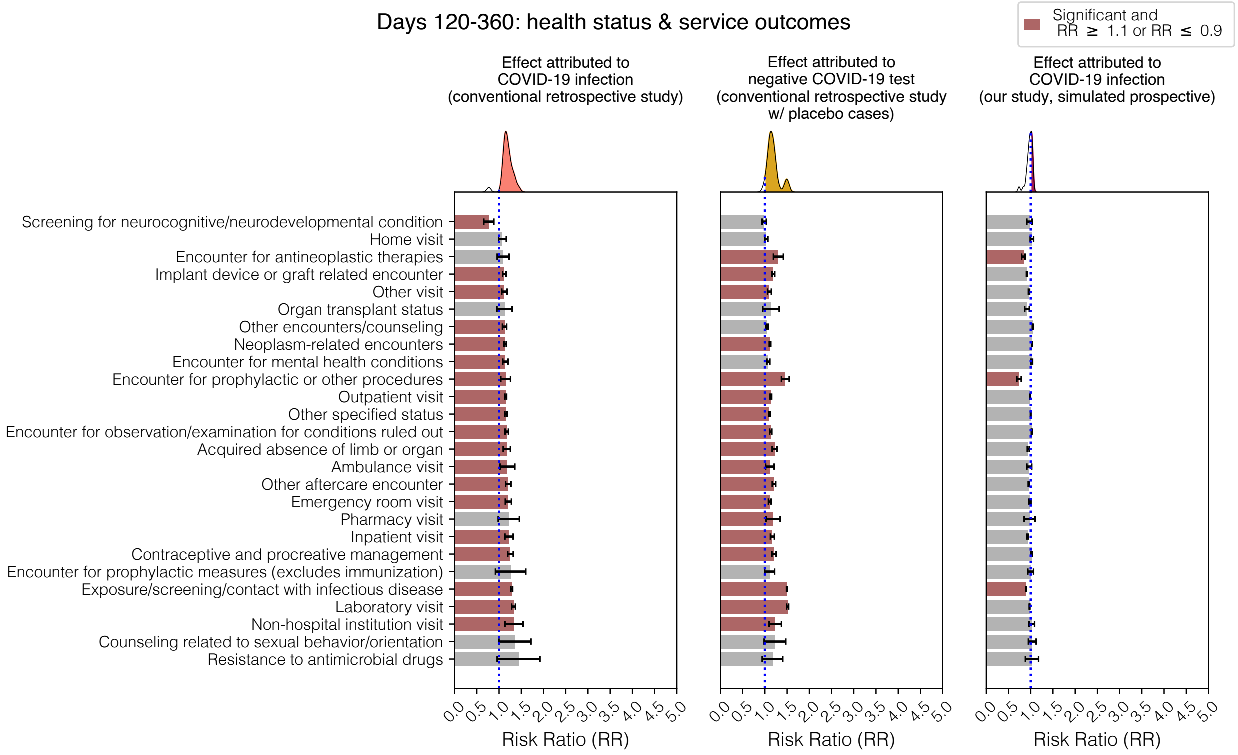


**Supplementary Figure S31:** Risk ratios for all outcomes related to health status & service on days 120–360 post-enrollment. Effects are shown for a conventional retrospective study of long COVID (left), our test-based prospective study (right), and a conventional retrospective study with placebo cases (middle). Red bars indicate significant effects ($p<0.05$ with Bonferroni correction for multiple comparisons; two-sided bootstrap) with $RR \geq 1.1$ or $RR \leq 0.9$.

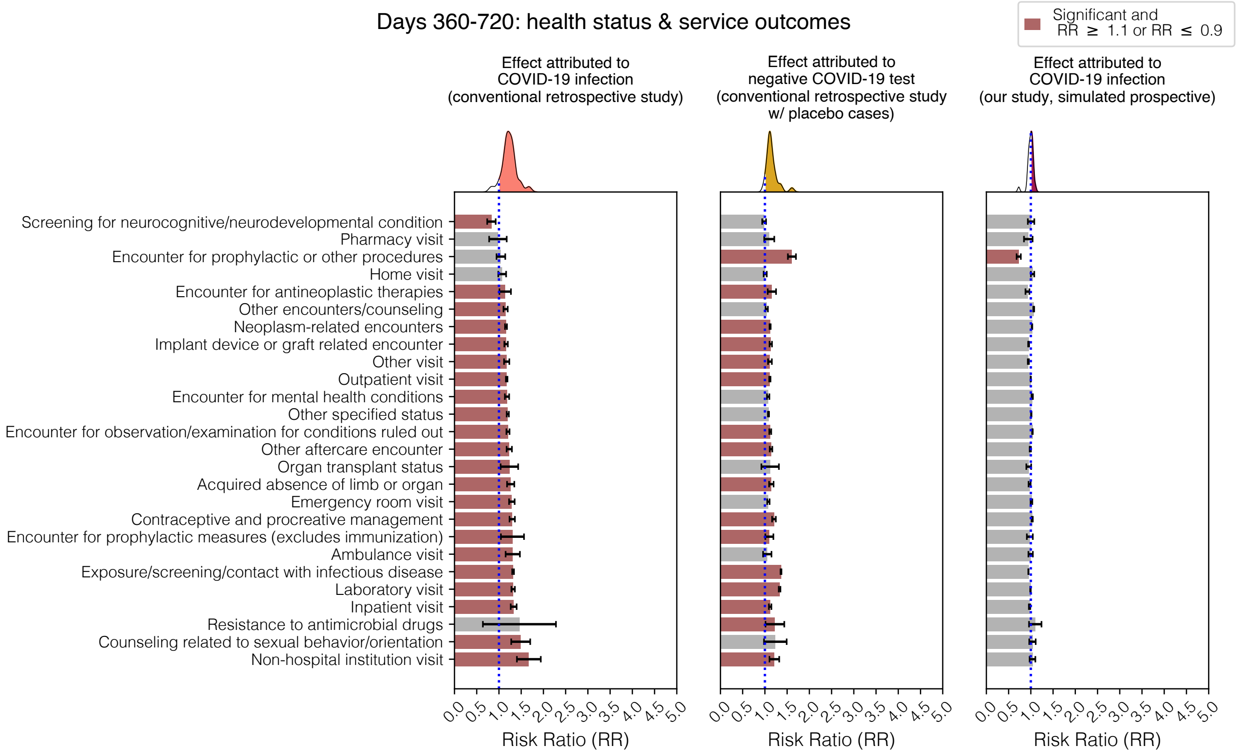


**Supplementary Figure S32:** Risk ratios for all outcomes related to health status & service on days 360–720 post-enrollment. Effects are shown for a conventional retrospective study of long COVID (left), our test-based prospective study (right), and a conventional retrospective study with placebo cases (middle). Red bars indicate significant effects ($p<0.05$ with Bonferroni correction for multiple comparisons; two-sided bootstrap) with $RR\geq1.1$ or $RR\leq0.9$.

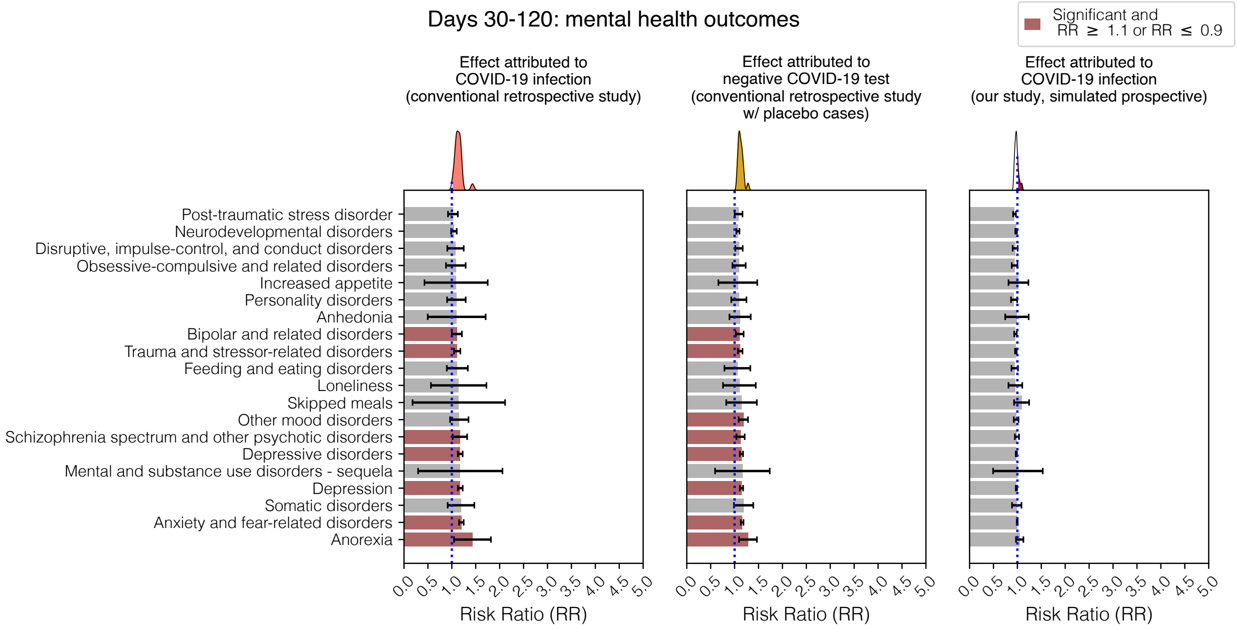


**Supplementary Figure S33:** Risk ratios for all outcomes related to mental health on days 30–120 post-enrollment. Effects are shown for a conventional retrospective study of long COVID (left), our test-based prospective study (right), and a conventional retrospective study with placebo cases (middle). Red bars indicate significant effects ($p<0.05$ with Bonferroni correction for multiple comparisons; two-sided bootstrap) with $RR \geq 1.1$ or $RR \leq 0.9$.

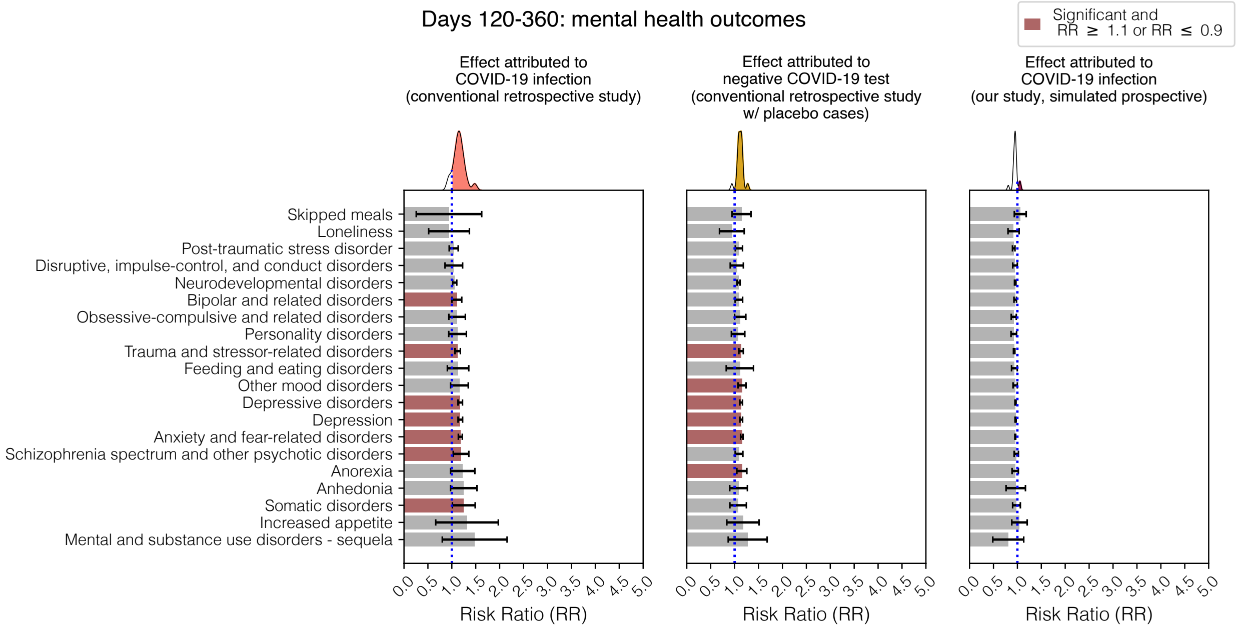


**Supplementary Figure S34:** Risk ratios for all outcomes related to mental health on days 120–360 post-enrollment. Effects are shown for a conventional retrospective study of long COVID (left), our test-based prospective study (right), and a conventional retrospective study with placebo cases (middle). Red bars indicate significant effects ($p<0.05$ with Bonferroni correction for multiple comparisons; two-sided bootstrap) with $RR \geq 1.1$ or $RR \leq 0.9$.

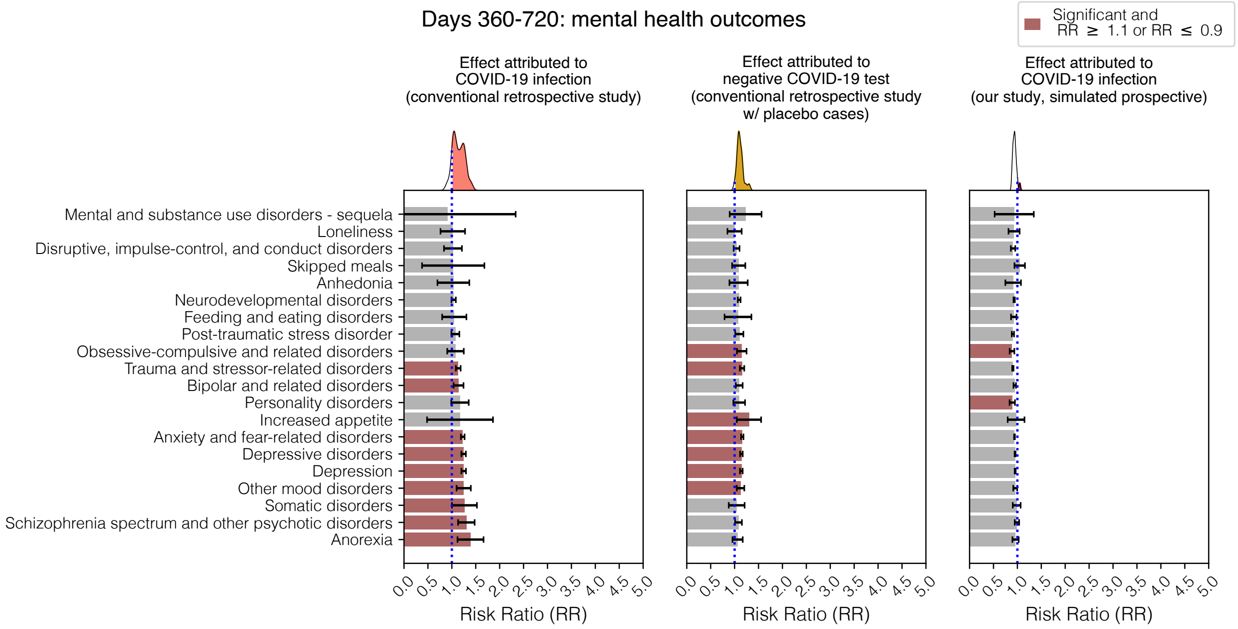


**Supplementary Figure S35:** Risk ratios for all outcomes related to mental health on days 360–720 post-enrollment. Effects are shown for a conventional retrospective study of long COVID (left), our test-based prospective study (right), and a conventional retrospective study with placebo cases (middle). Red bars indicate significant effects ($p<0.05$ with Bonferroni correction for multiple comparisons; two-sided bootstrap) with $RR \geq 1.1$ or $RR \leq 0.9$.

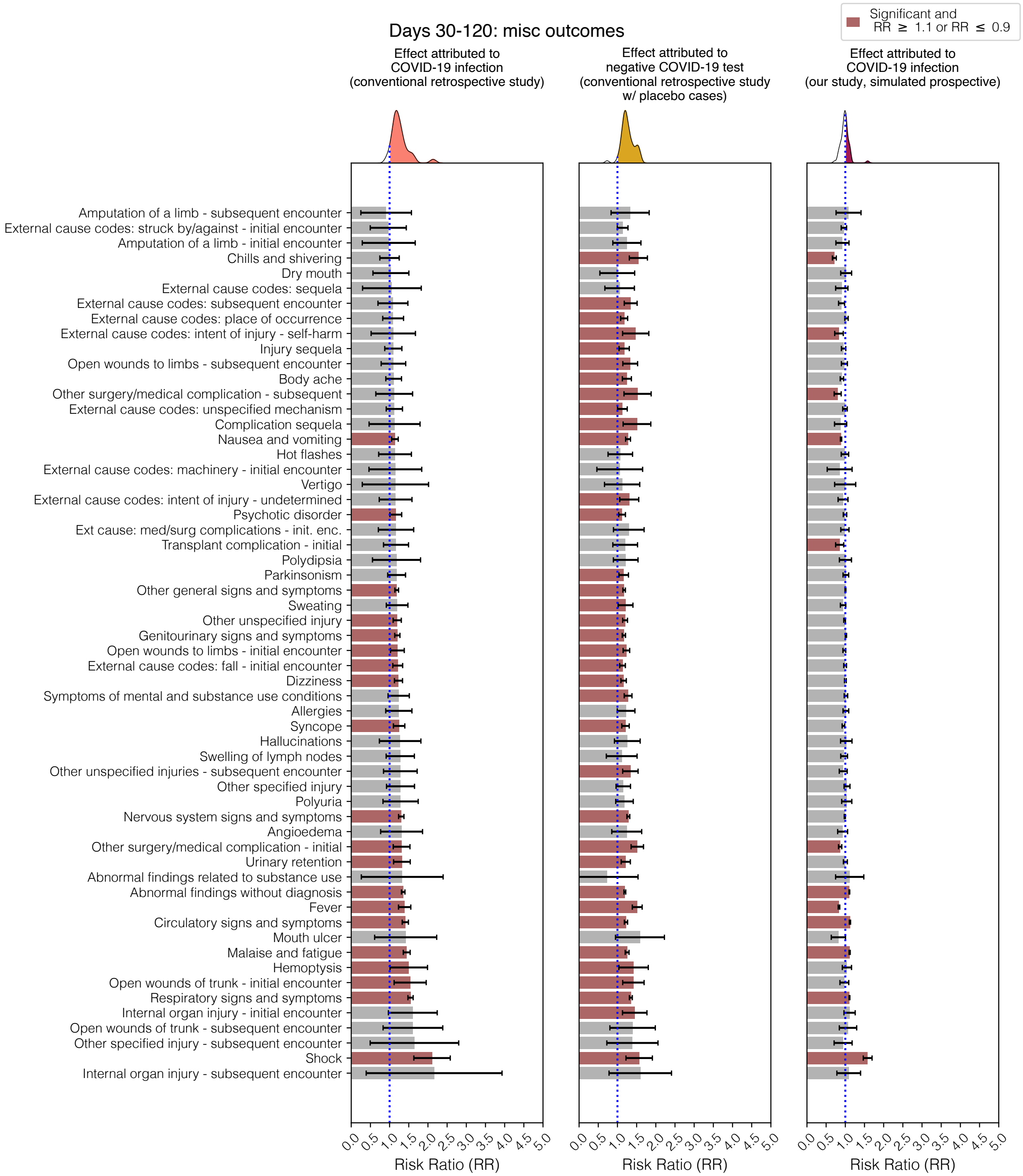


**Supplementary Figure S36:** Risk ratios for all miscellaneous outcomes on days 30–120 post-enrollment. Effects are shown for a conventional retrospective study of long COVID (left), our test-based prospective study (right), and a conventional retrospective study with placebo cases (middle). Red bars indicate significant effects ($p<0.05$ with Bonferroni correction for multiple comparisons; two-sided bootstrap) with $RR \geq 1.1$ or $RR \leq 0.9$.

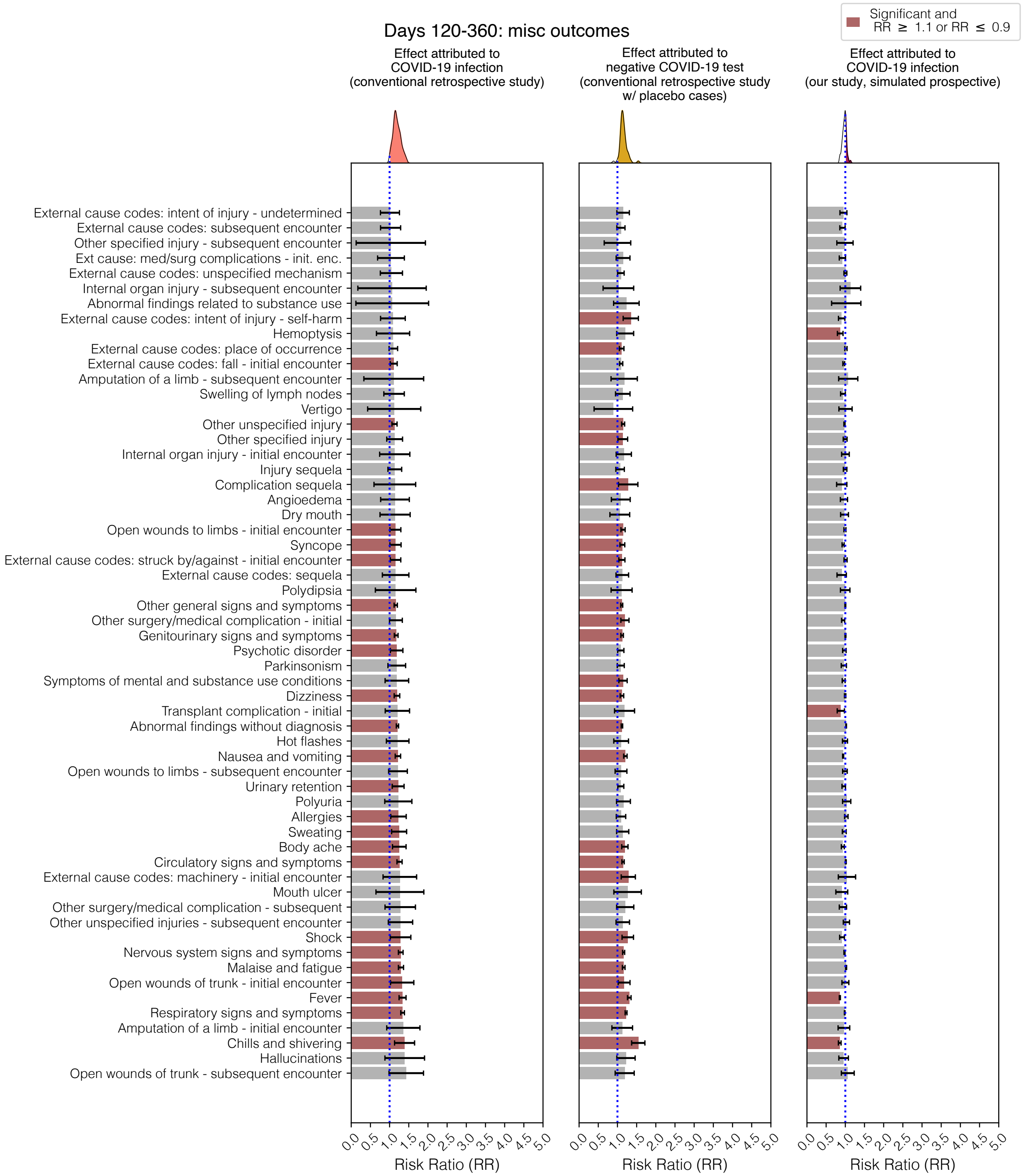


**Supplementary Figure S37:** Risk ratios for all miscellaneous outcomes on days 120–360 post-enrollment. Effects are shown for a conventional retrospective study of long COVID (left), our test-based prospective study (right), and a conventional retrospective study with placebo cases (middle). Red bars indicate significant effects ($p<0.05$ with Bonferroni correction for multiple comparisons; two-sided bootstrap) with $RR\geq1.1$ or $RR\leq0.9$.

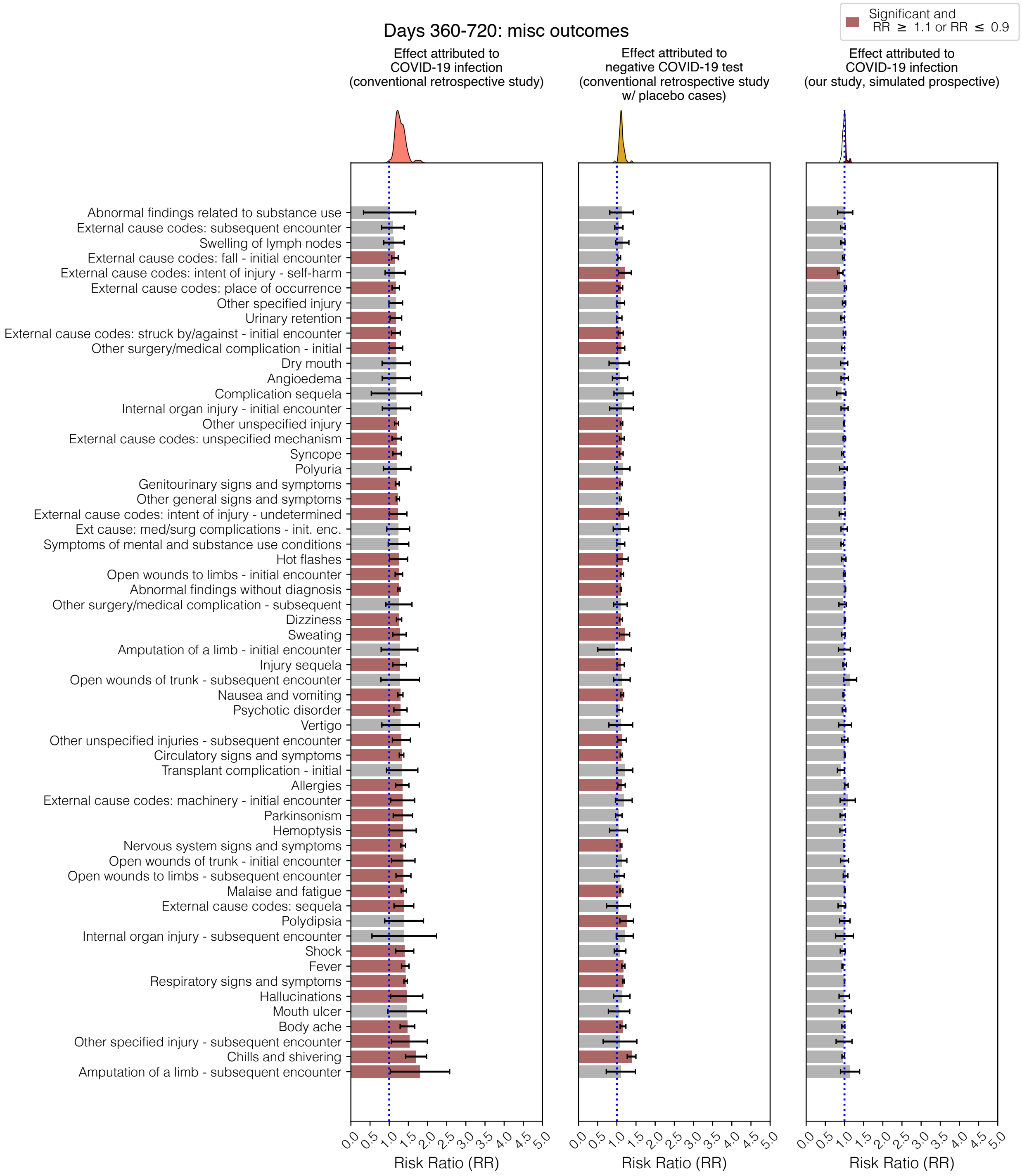


**Supplementary Figure S38:** Risk ratios for all miscellaneous outcomes on days 360–720 post-enrollment. Effects are shown for a conventional retrospective study of long COVID (left), our test-based prospective study (right), and a conventional retrospective study with placebo cases (middle). Red bars indicate significant effects ($p<0.05$ with Bonferroni correction for multiple comparisons; two-sided bootstrap) with RR$\geq$1.1 or RR$\leq$0.9.

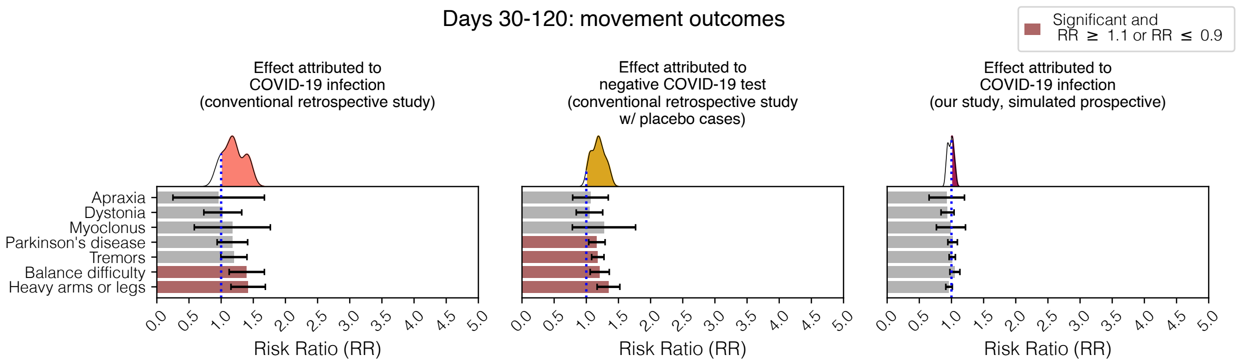


**Supplementary Figure S39:** Risk ratios for all outcomes related to movement on days 30–120 post-enrollment. Effects are shown for a conventional retrospective study of long COVID (left), our test-based prospective study (right), and a conventional retrospective study with placebo cases (middle). Red bars indicate significant effects ($p<0.05$ with Bonferroni correction for multiple comparisons; two-sided bootstrap) with $RR \geq 1.1$ or $RR \leq 0.9$.

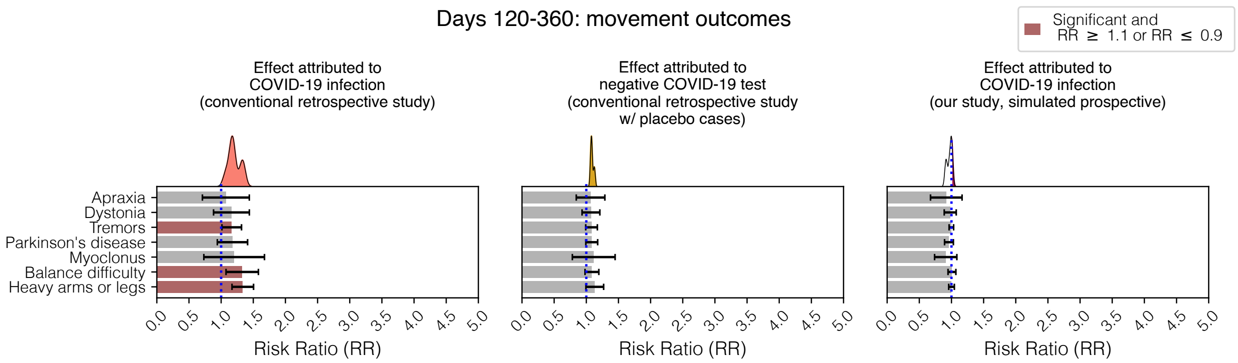


**Supplementary Figure S40:** Risk ratios for all outcomes related to movement on days 120–360 post-enrollment. Effects are shown for a conventional retrospective study of long COVID (left), our test-based prospective study (right), and a conventional retrospective study with placebo cases (middle). Red bars indicate significant effects ($p<0.05$ with Bonferroni correction for multiple comparisons; two-sided bootstrap) with $RR\geq1.1$ or $RR\leq0.9$.

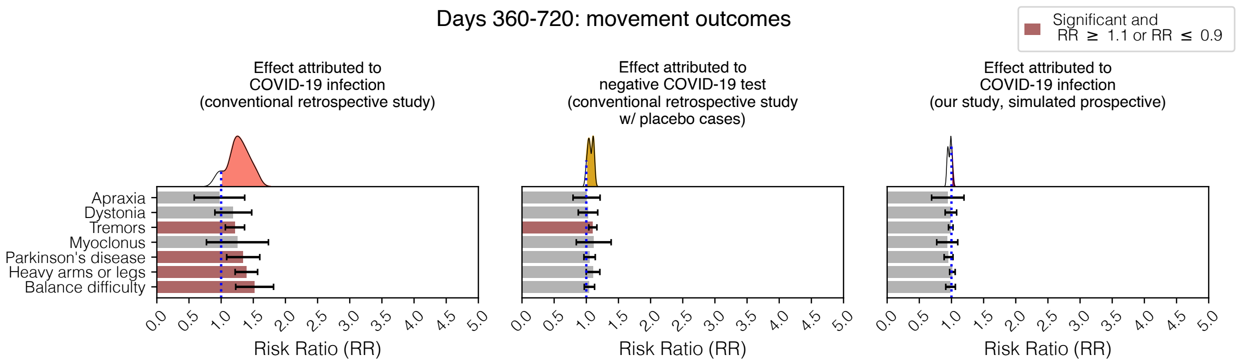


**Supplementary Figure S41:** Risk ratios for all outcomes related to movement on days 360–720 post-enrollment. Effects are shown for a conventional retrospective study of long COVID (left), our test-based prospective study (right), and a conventional retrospective study with placebo cases (middle). Red bars indicate significant effects ($p<0.05$ with Bonferroni correction for multiple comparisons; two-sided bootstrap) with RR$\geq$1.1 or RR$\leq$0.9.

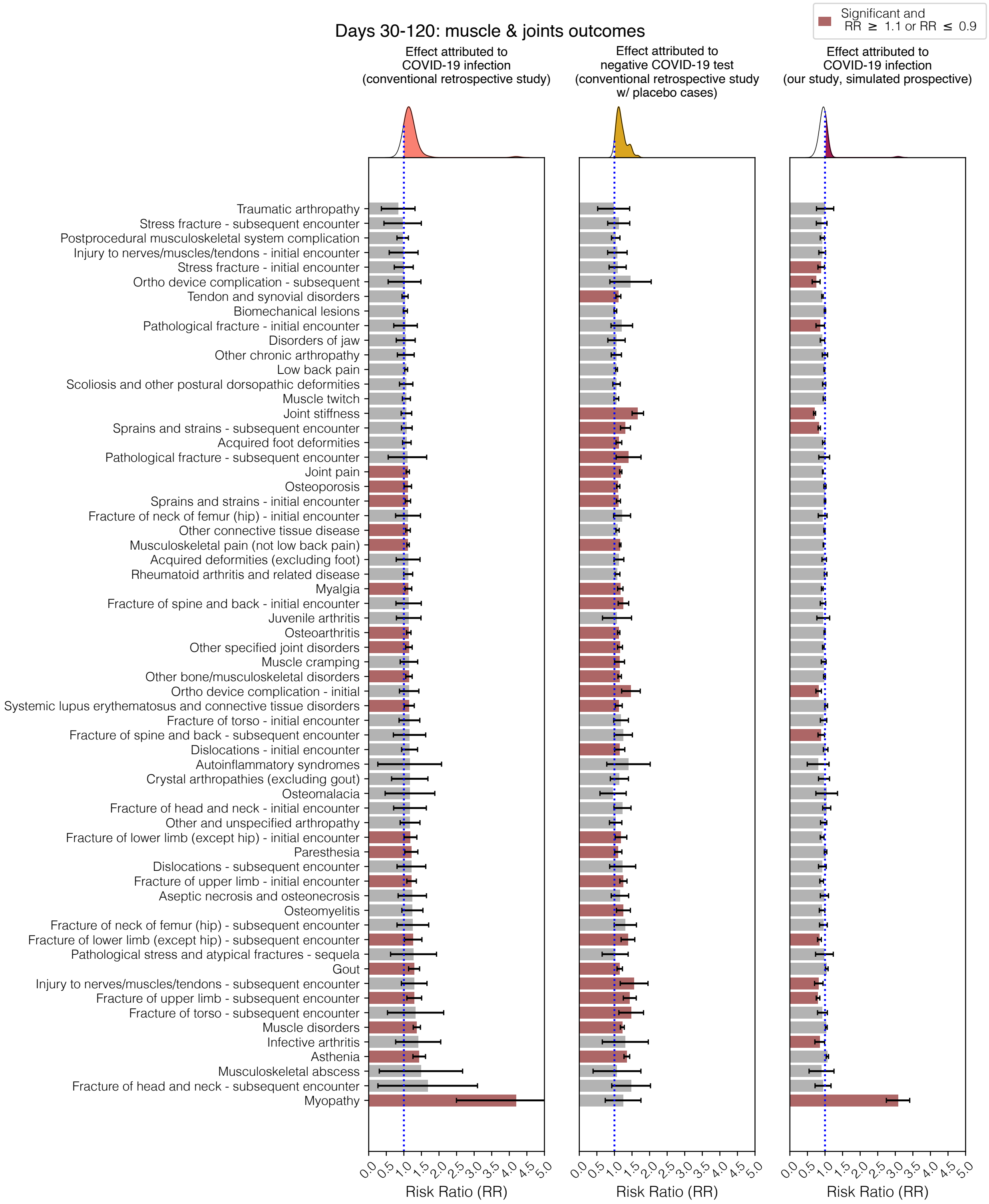


**Supplementary Figure S42:** Risk ratios for all outcomes related to muscle & joints on days 30–120 post-enrollment. Effects are shown for a conventional retrospective study of long COVID (left), our test-based prospective study (right), and a conventional retrospective study with placebo cases (middle). Red bars indicate significant effects ($p<0.05$ with Bonferroni correction for multiple comparisons; two-sided bootstrap) with $RR \geq 1.1$ or $RR \leq 0.9$.

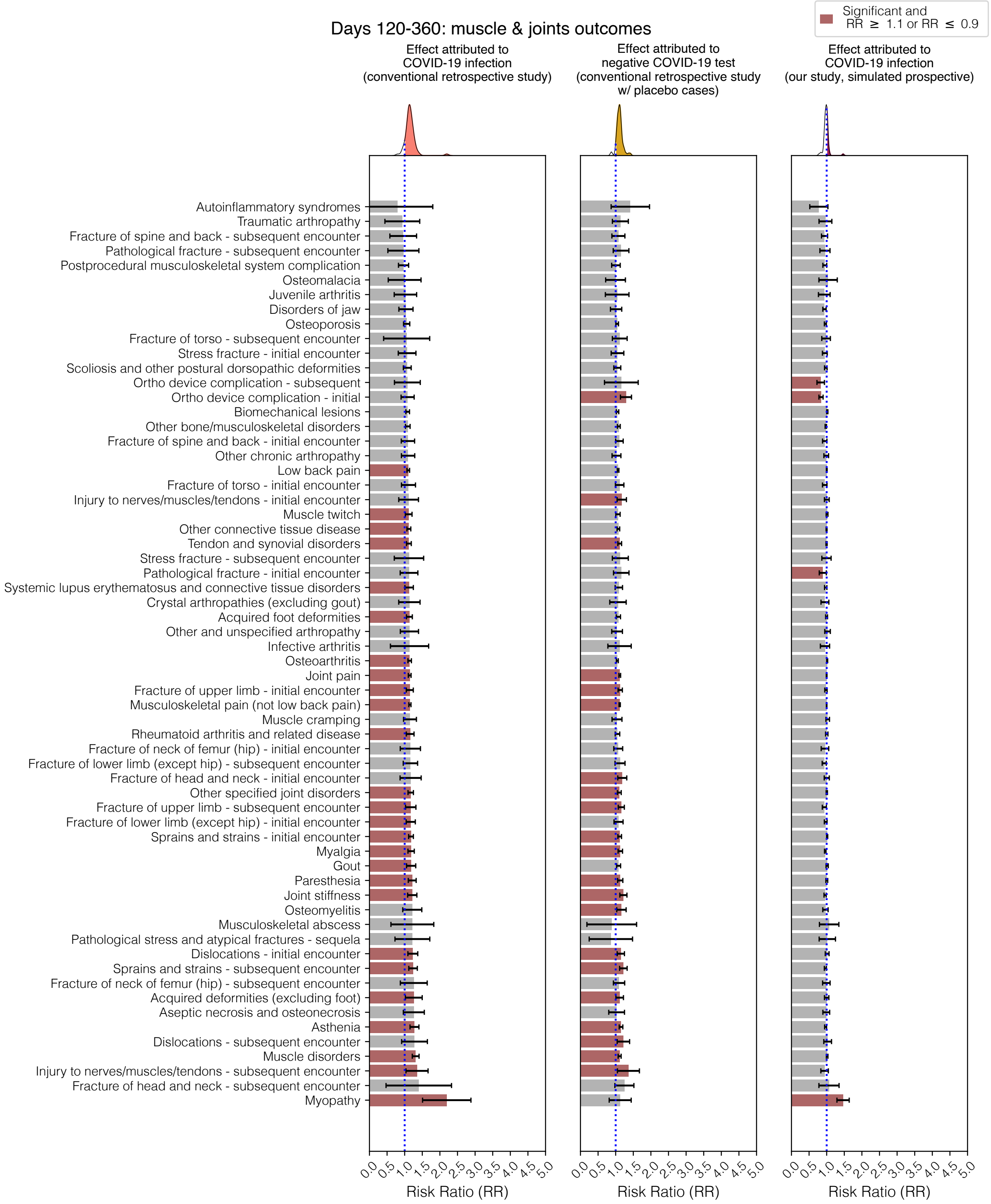


**Supplementary Figure S43:** Risk ratios for all outcomes related to muscle & joints on days 120–360 post-enrollment. Effects are shown for a conventional retrospective study of long COVID (left), our test-based prospective study (right), and a conventional retrospective study with placebo cases (middle). Red bars indicate significant effects ($p<0.05$ with Bonferroni correction for multiple comparisons; two-sided bootstrap) with $RR \geq 1.1$ or $RR \leq 0.9$.

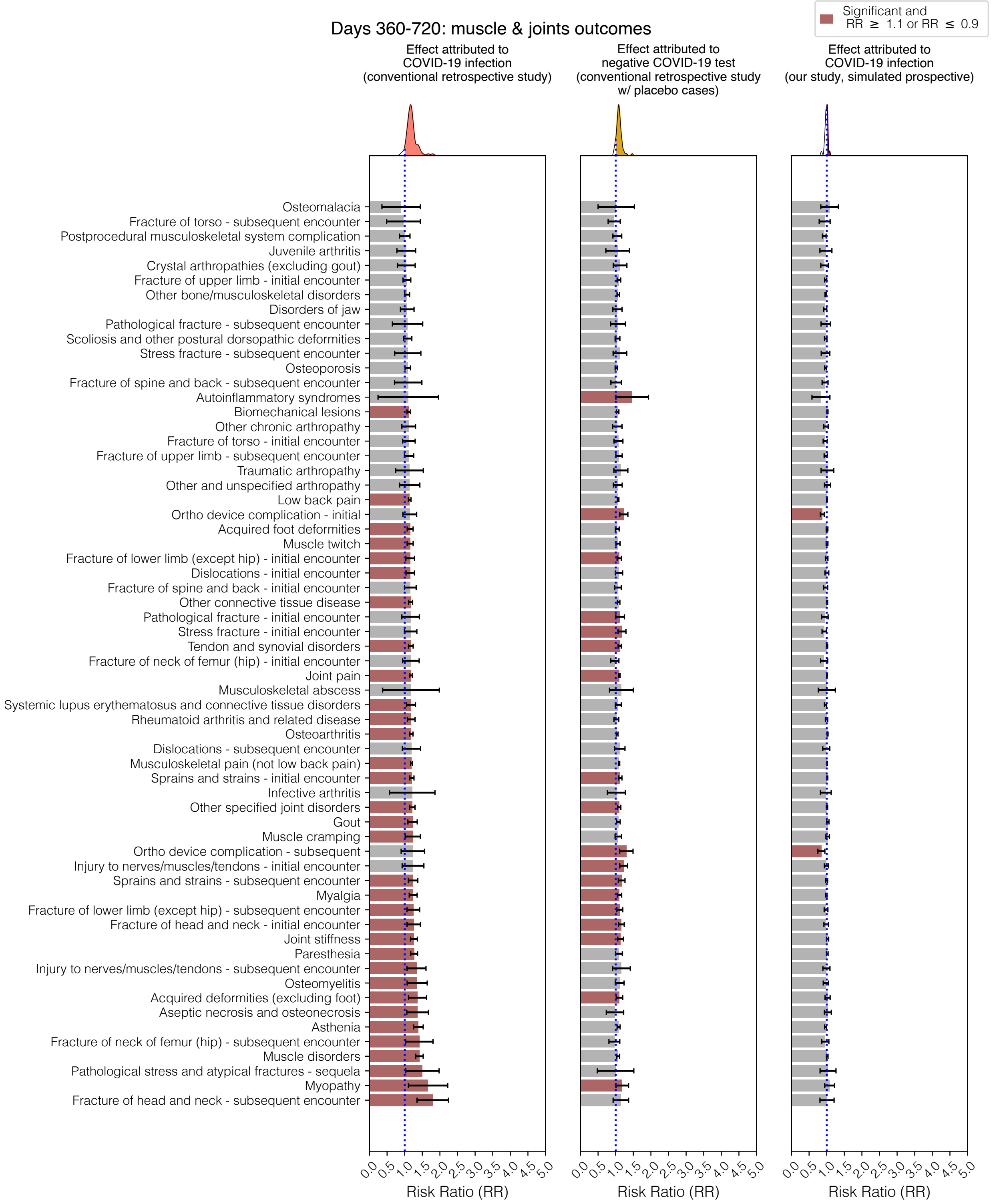


**Supplementary Figure S44:** Risk ratios for all outcomes related to muscle & joints on days 360–720 post-enrollment. Effects are shown for a conventional retrospective study of long COVID (left), our test-based prospective study (right), and a conventional retrospective study with placebo cases (middle). Red bars indicate significant effects (p<0.05 with Bonferroni correction for multiple comparisons; two-sided bootstrap) with RR≥1.1 or RR≤0.9.

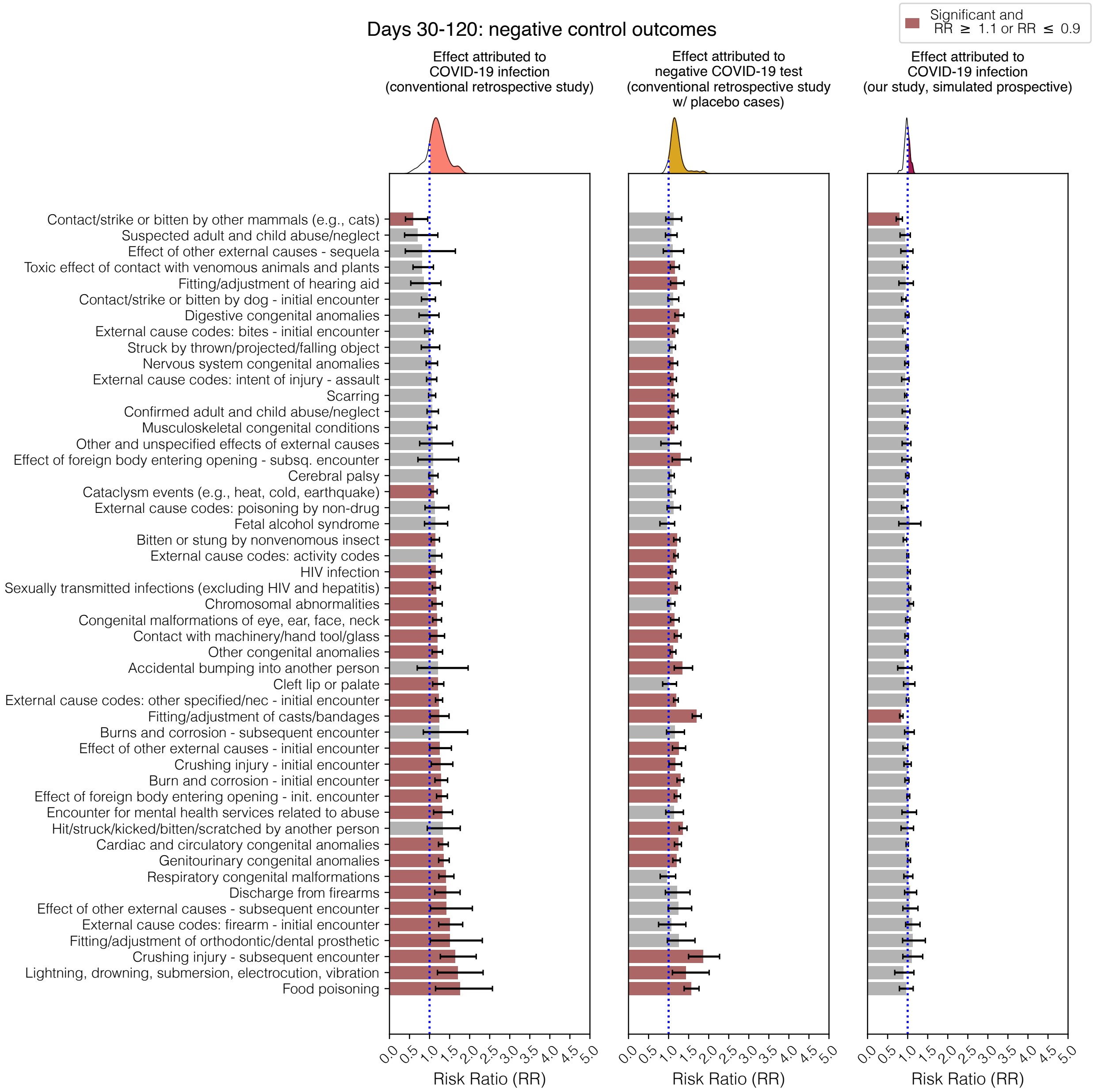


**Supplementary Figure S45:** Risk ratios for all negative control outcomes on days 30–120 post-enrollment. Effects are shown for a conventional retrospective study of long COVID (left), our test-based prospective study (right), and a conventional retrospective study with placebo cases (middle). Red bars indicate significant effects (p<0.05 unadjusted for multiple comparisons; two-sided bootstrap) with RR≥1.1 or RR≤0.9.

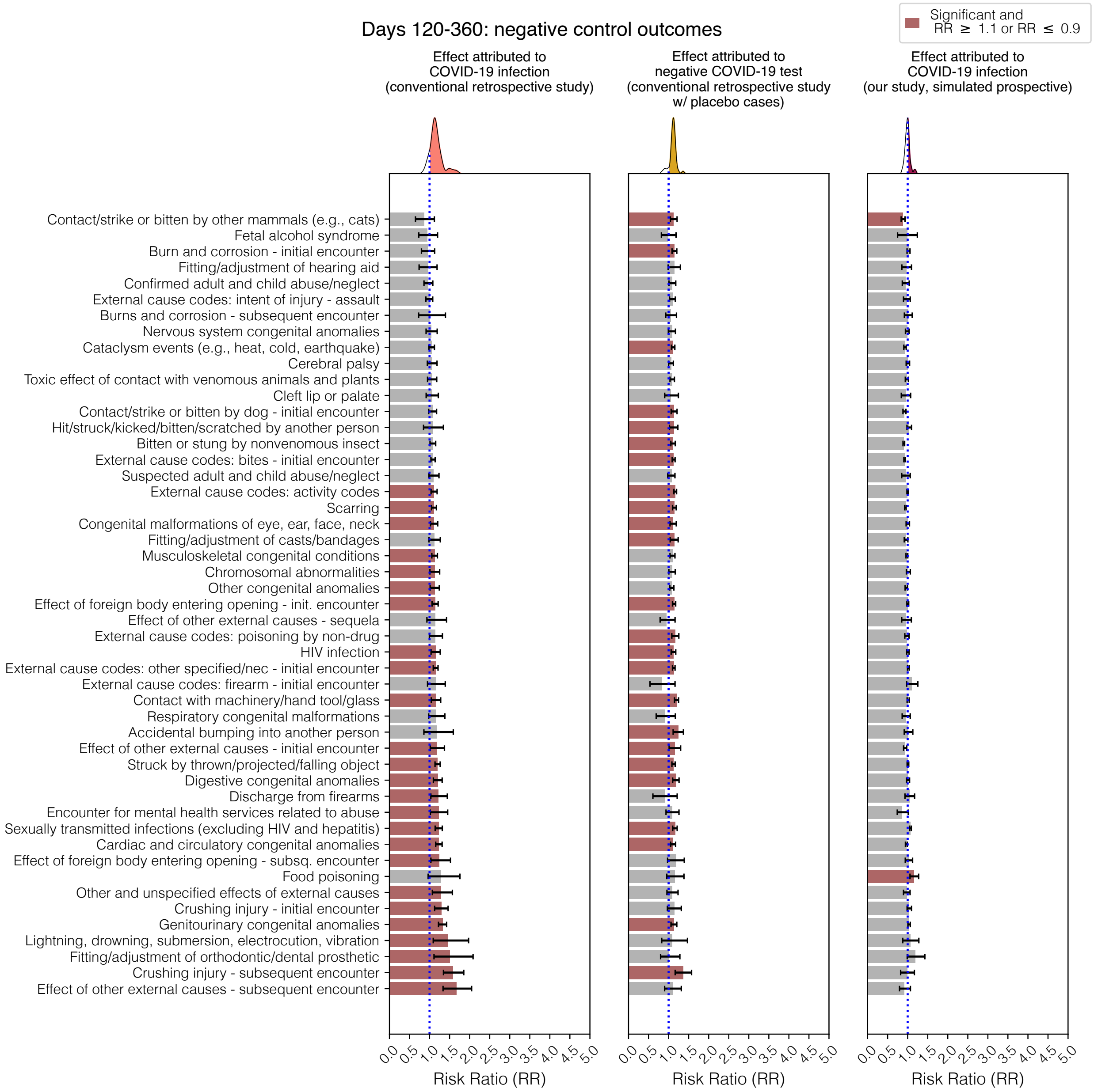


**Supplementary Figure S46:** Risk ratios for all negative control outcomes on days 120–360 post-enrollment. Effects are shown for a conventional retrospective study of long COVID (left), our test-based prospective study (right), and a conventional retrospective study with placebo cases (middle). Red bars indicate significant effects ($p<0.05$ unadjusted for multiple comparisons; two-sided bootstrap) with $RR \geq 1.1$ or $RR \leq 0.9$.

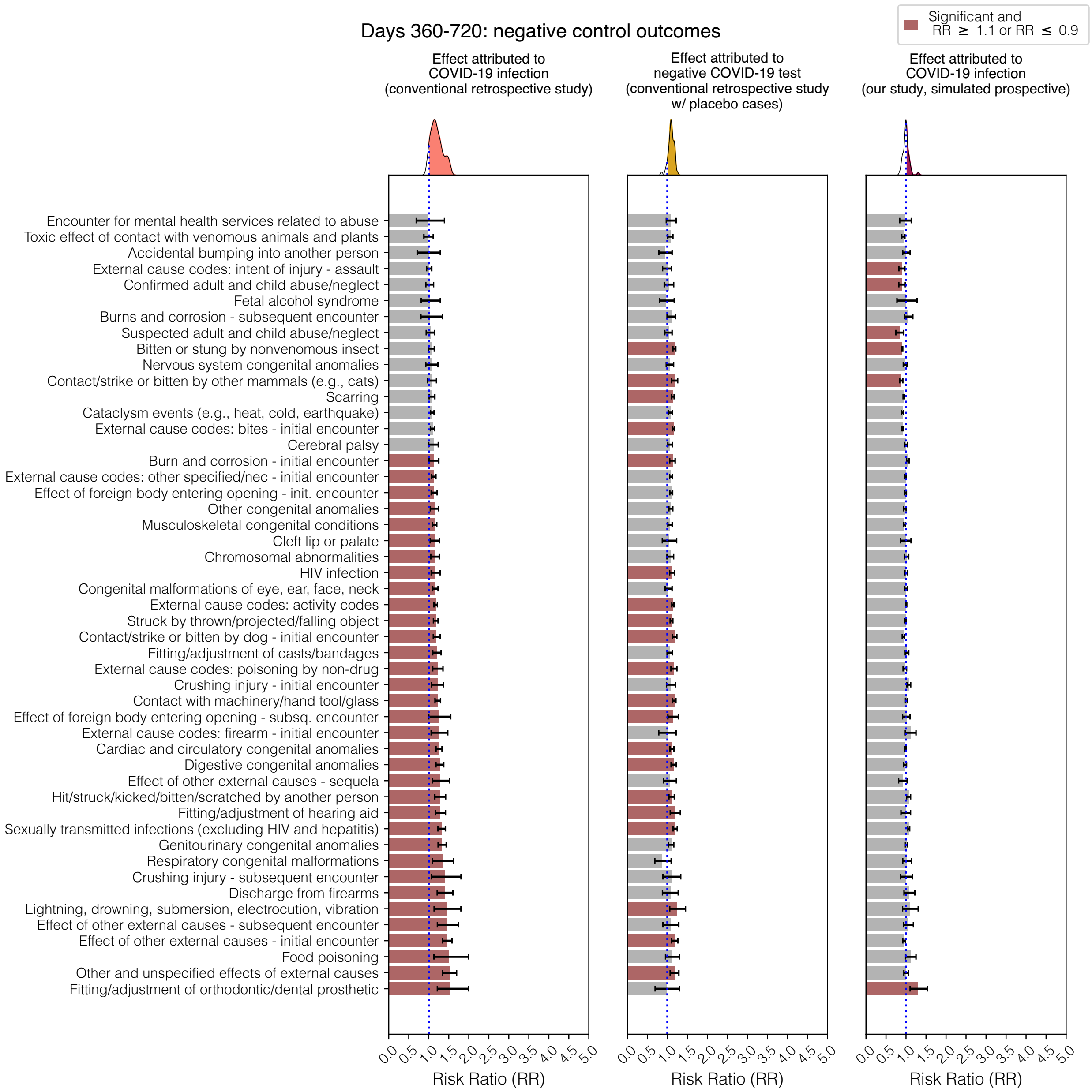


**Supplementary Figure S47:** Risk ratios for all negative control outcomes on days 360–720 post-enrollment. Effects are shown for a conventional retrospective study of long COVID (left), our test-based prospective study (right), and a conventional retrospective study with placebo cases (middle). Red bars indicate significant effects (p<0.05 unadjusted for multiple comparisons; two-sided bootstrap) with RR≥1.1 or RR≤0.9.

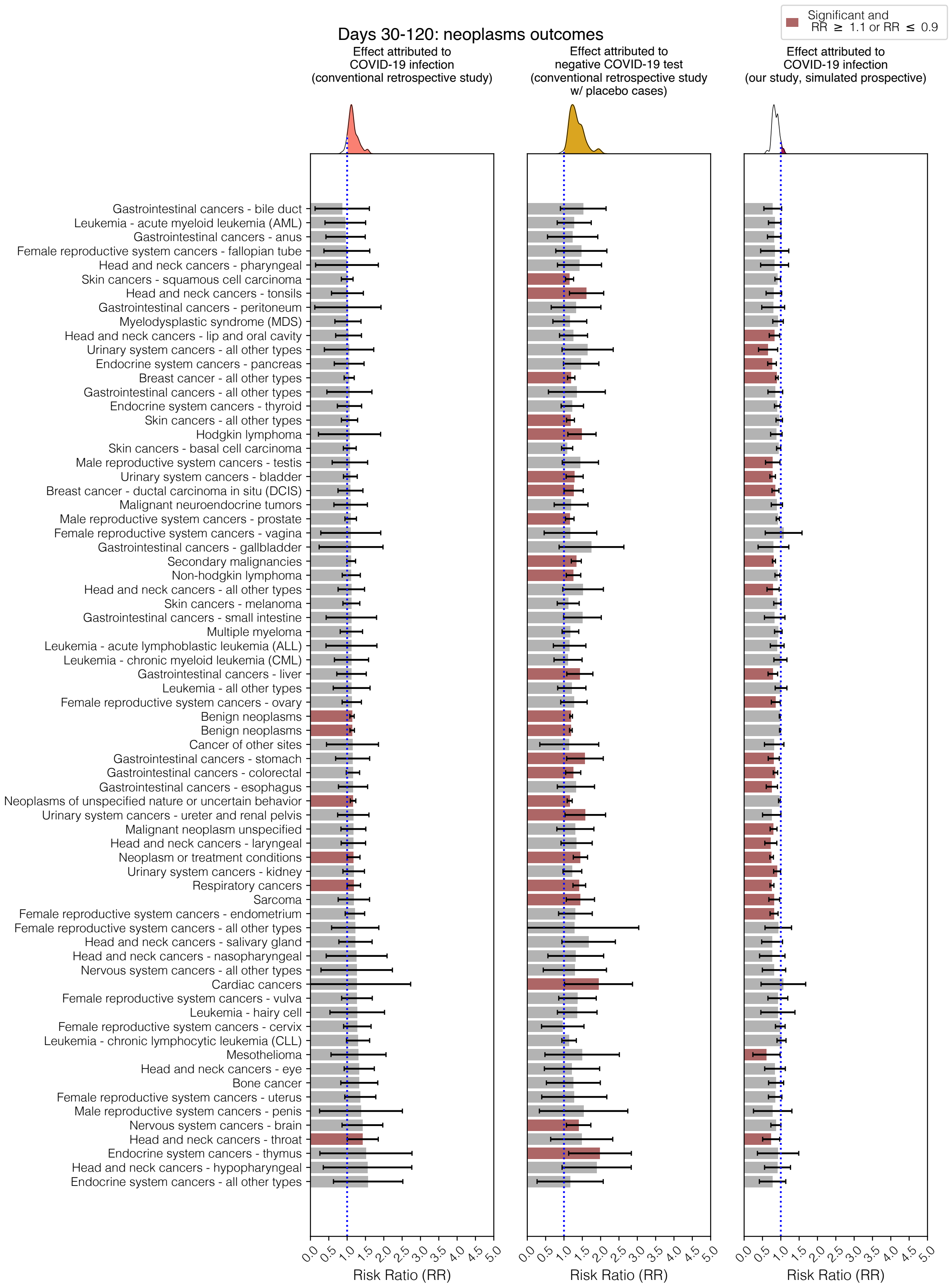


**Supplementary Figure S48:** Risk ratios for all outcomes related to neoplasms on days 30–120 post-enrollment. Effects are shown for a conventional retrospective study of long COVID (left), our test-based prospective study (right), and a conventional retrospective study with placebo cases (middle). Red bars indicate significant effects (p<0.05 with Bonferroni correction for multiple comparisons; two-sided bootstrap) with RR≥1.1 or RR≤0.9.

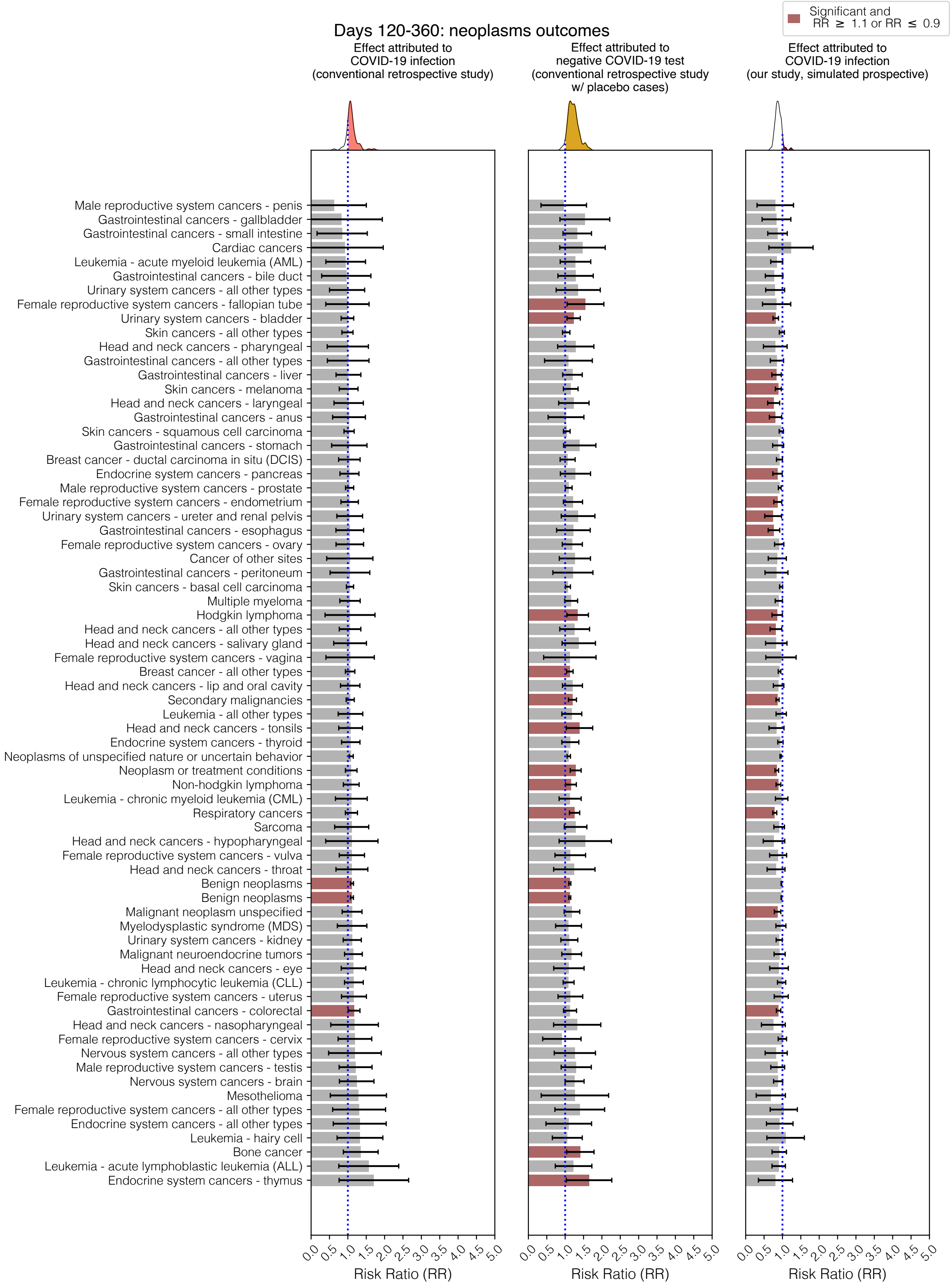


**Supplementary Figure S49:** Risk ratios for all outcomes related to neoplasms on days 120–360 post-enrollment. Effects are shown for a conventional retrospective study of long COVID (left), our test-based prospective study (right), and a conventional retrospective study with placebo cases (middle). Red bars indicate significant effects ($p<0.05$ with Bonferroni correction for multiple comparisons; two-sided bootstrap) with RR$\geq$1.1 or RR$\leq$0.9.

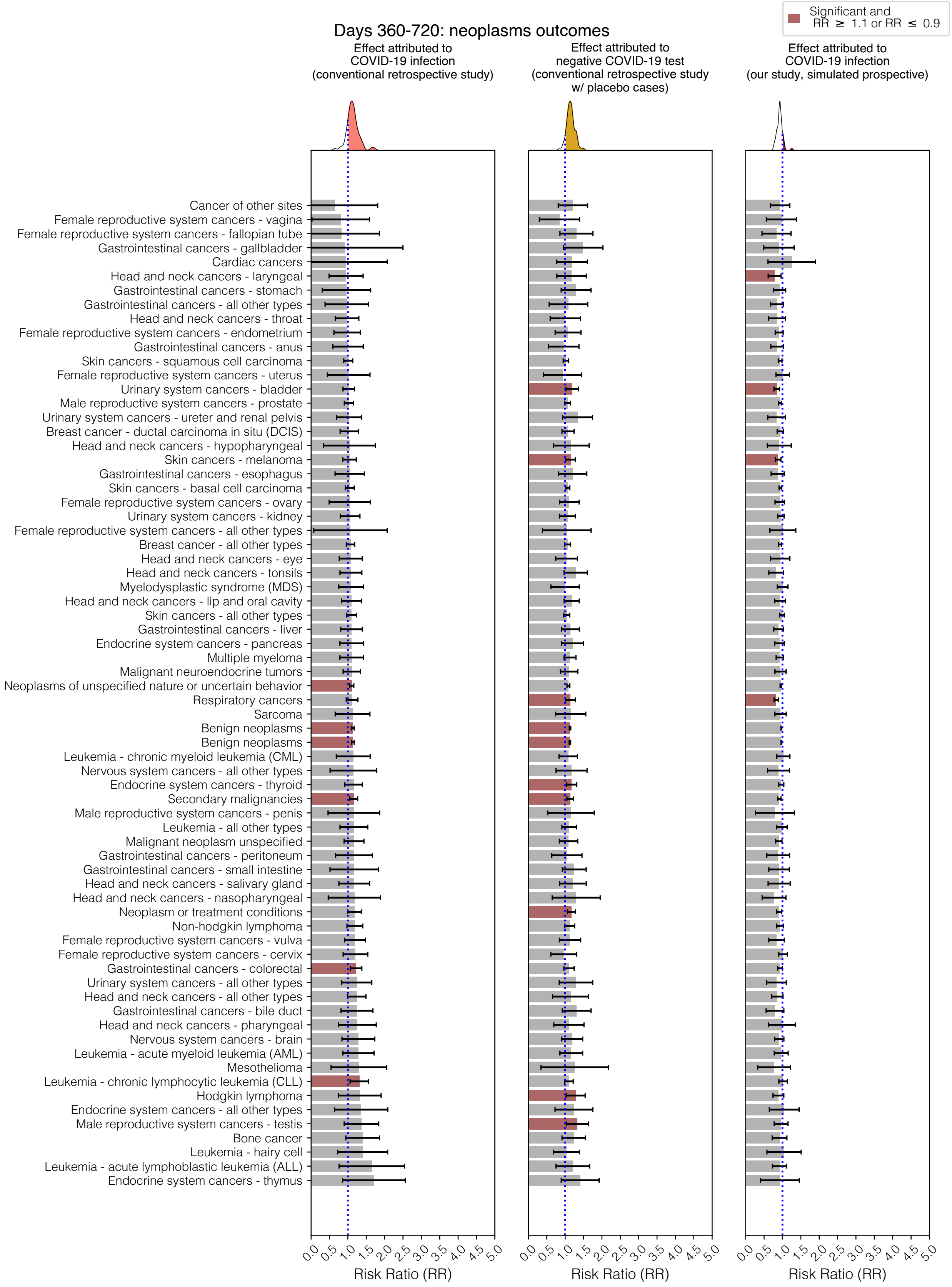


**Supplementary Figure S50:** Risk ratios for all outcomes related to neoplasms on days 360–720 post-enrollment. Effects are shown for a conventional retrospective study of long COVID (left), our test-based prospective study (right), and a conventional retrospective study with placebo cases (middle). Red bars indicate significant effects (p<0.05 with Bonferroni correction for multiple comparisons; two-sided bootstrap) with RR≥1.1 or RR≤0.9.

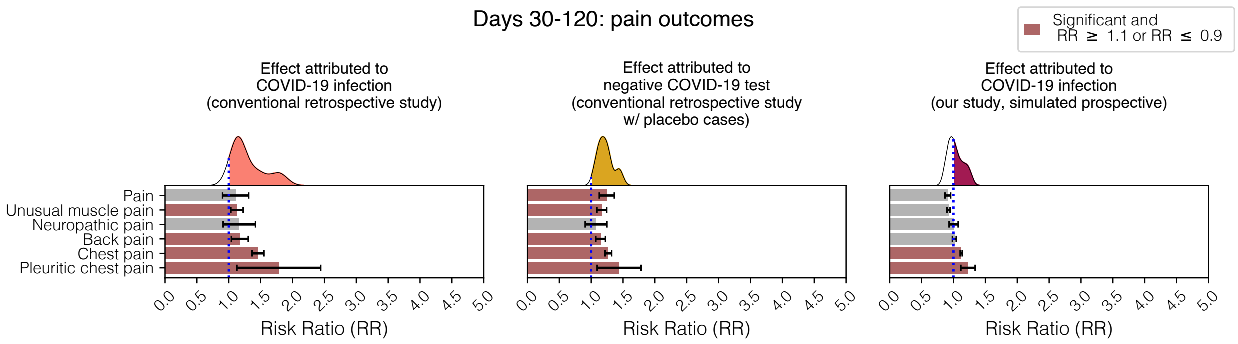


**Supplementary Figure S51:** Risk ratios for all outcomes related to pain on days 30–120 post-enrollment. Effects are shown for a conventional retrospective study of long COVID (left), our test-based prospective study (right), and a conventional retrospective study with placebo cases (middle). Red bars indicate significant effects (p<0.05 with Bonferroni correction for multiple comparisons; two-sided bootstrap) with RR≥1.1 or RR≤0.9.

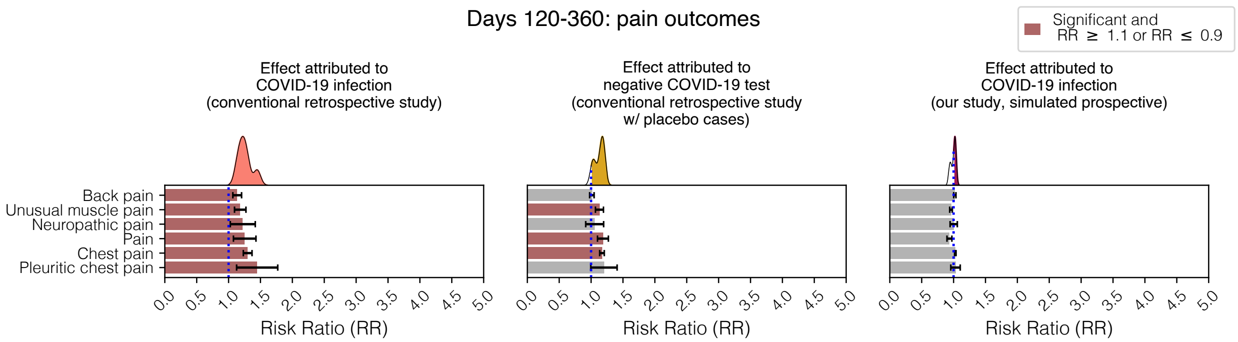


**Supplementary Figure S52:** Risk ratios for all outcomes related to pain on days 120–360 post-enrollment. Effects are shown for a conventional retrospective study of long COVID (left), our test-based prospective study (right), and a conventional retrospective study with placebo cases (middle). Red bars indicate significant effects ($p<0.05$ with Bonferroni correction for multiple comparisons; two-sided bootstrap) with RR$\geq$1.1 or RR$\leq$0.9.

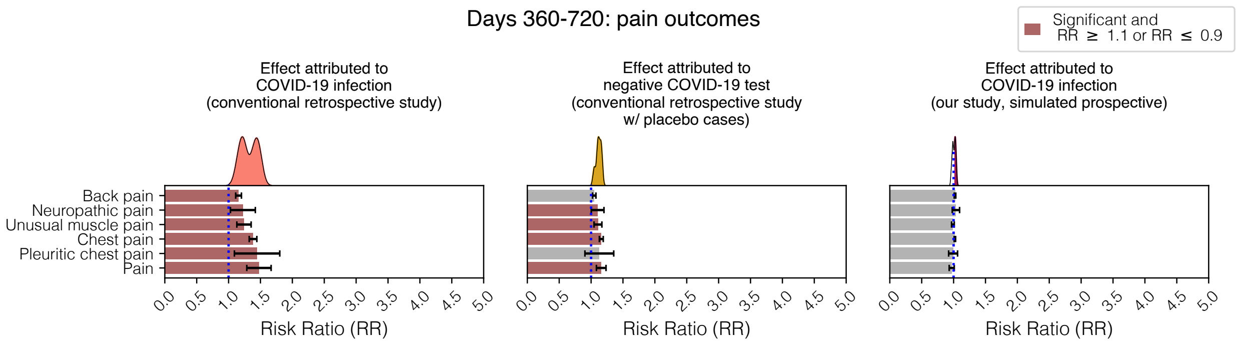


**Supplementary Figure S53:** Risk ratios for all outcomes related to pain on days 360–720 post-enrollment. Effects are shown for a conventional retrospective study of long COVID (left), our test-based prospective study (right), and a conventional retrospective study with placebo cases (middle). Red bars indicate significant effects ($p<0.05$ with Bonferroni correction for multiple comparisons; two-sided bootstrap) with $RR\geq1.1$ or $RR\leq0.9$.

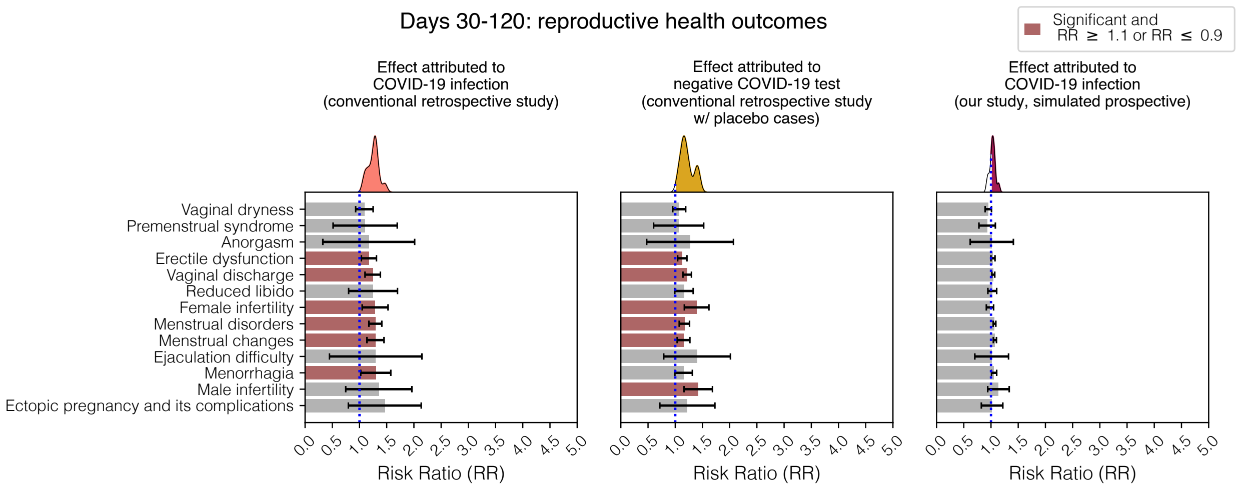


**Supplementary Figure S54:** Risk ratios for all outcomes related to reproductive health on days 30–120 post-enrollment. Effects are shown for a conventional retrospective study of long COVID (left), our test-based prospective study (right), and a conventional retrospective study with placebo cases (middle). Red bars indicate significant effects ($p<0.05$ with Bonferroni correction for multiple comparisons; two-sided bootstrap) with RR$\geq$1.1 or RR$\leq$0.9.

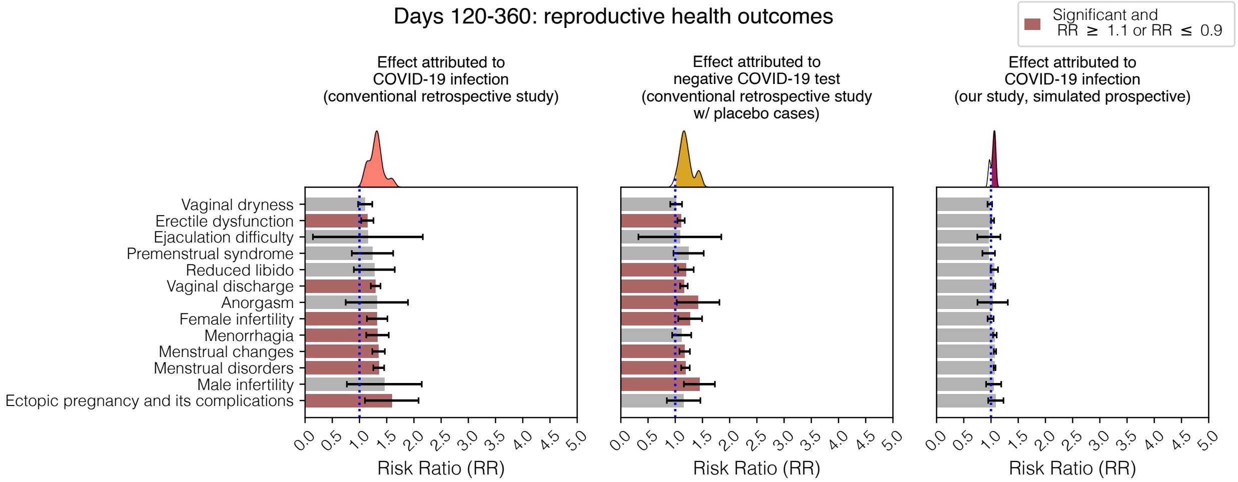


**Supplementary Figure S55:** Risk ratios for all outcomes related to reproductive health on days 120–360 post-enrollment. Effects are shown for a conventional retrospective study of long COVID (left), our test-based prospective study (right), and a conventional retrospective study with placebo cases (middle). Red bars indicate significant effects ($p<0.05$ with Bonferroni correction for multiple comparisons; two-sided bootstrap) with $RR \geq 1.1$ or $RR \leq 0.9$.

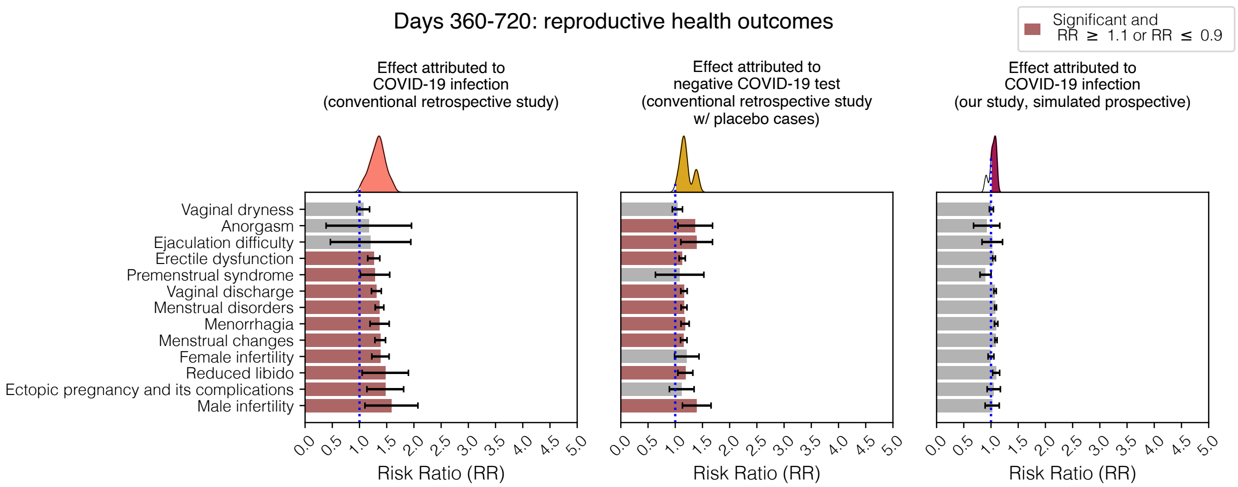


**Supplementary Figure S56:** Risk ratios for all outcomes related to reproductive health on days 360–720 post-enrollment. Effects are shown for a conventional retrospective study of long COVID (left), our test-based prospective study (right), and a conventional retrospective study with placebo cases (middle). Red bars indicate significant effects ($p<0.05$ with Bonferroni correction for multiple comparisons; two-sided bootstrap) with $RR \geq 1.1$ or $RR \leq 0.9$.

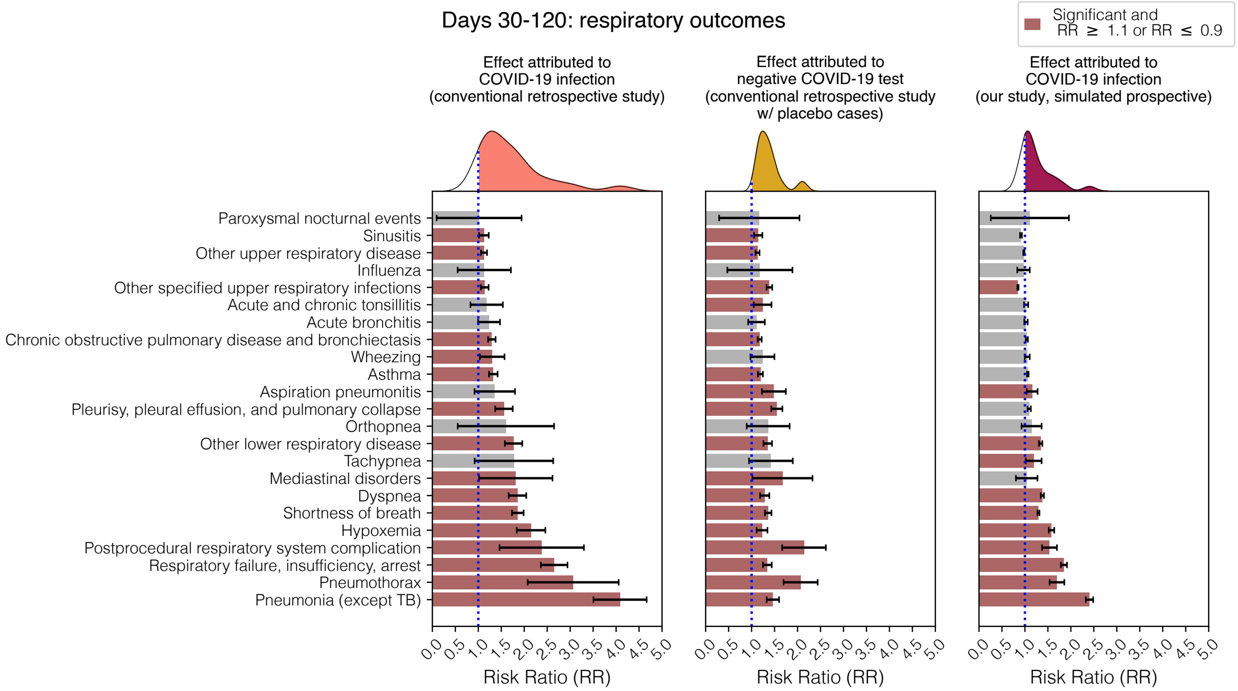


**Supplementary Figure S57:** Risk ratios for all outcomes related to the respiratory system on days 30–120 post-enrollment. Effects are shown for a conventional retrospective study of long COVID (left), our test-based prospective study (right), and a conventional retrospective study with placebo cases (middle). Red bars indicate significant effects ($p<0.05$ with Bonferroni correction for multiple comparisons; two-sided bootstrap) with RR≥1.1 or RR≤0.9.

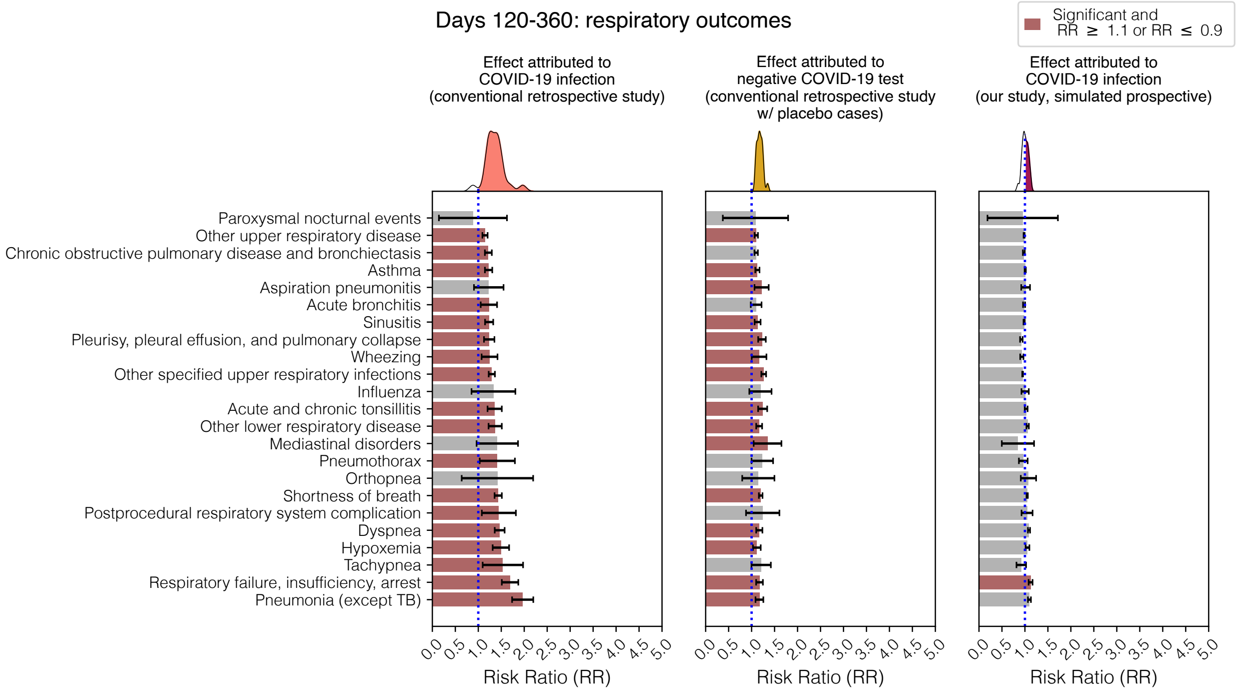


**Supplementary Figure S58:** Risk ratios for all outcomes related to the respiratory system on days 120–360 post-enrollment. Effects are shown for a conventional retrospective study of long COVID (left), our test-based prospective study (right), and a conventional retrospective study with placebo cases (middle). Red bars indicate significant effects (p<0.05 with Bonferroni correction for multiple comparisons; two-sided bootstrap) with RR≥1.1 or RR≤0.9.

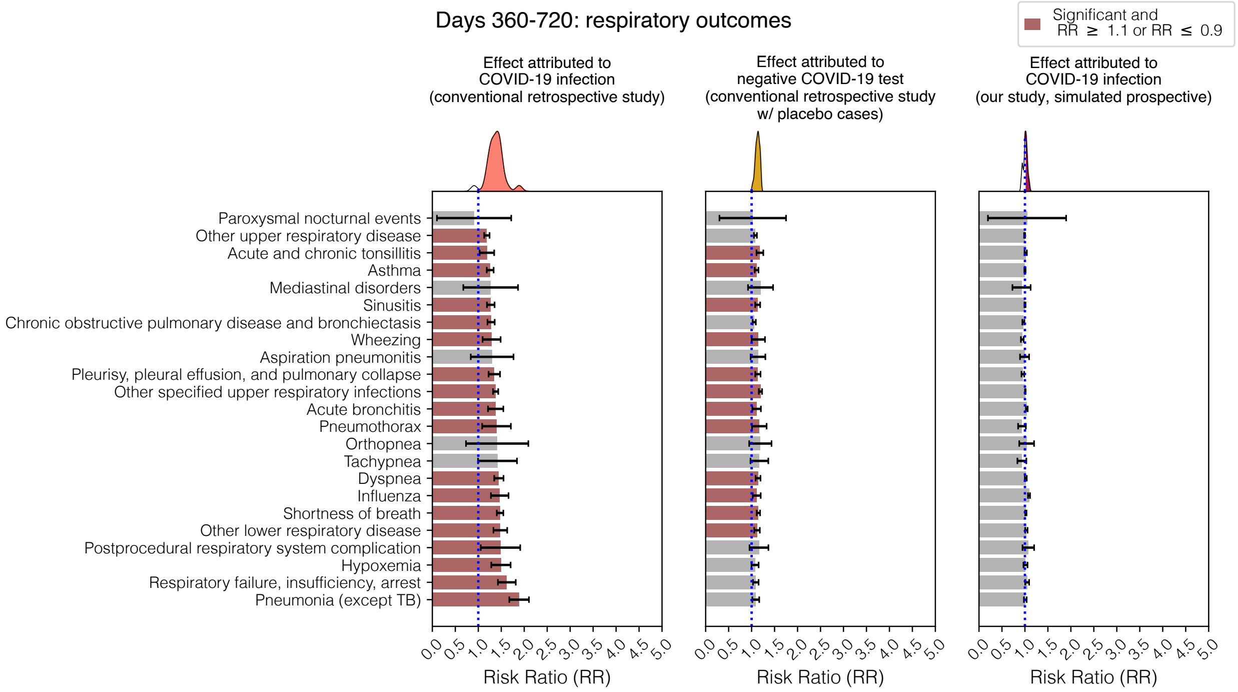


**Supplementary Figure S59:** Risk ratios for all outcomes related to the respiratory system on days 360–720 post-enrollment. Effects are shown for a conventional retrospective study of long COVID (left), our test-based prospective study (right), and a conventional retrospective study with placebo cases (middle). Red bars indicate significant effects ($p<0.05$ with Bonferroni correction for multiple comparisons; two-sided bootstrap) with $RR \geq 1.1$ or $RR \leq 0.9$.

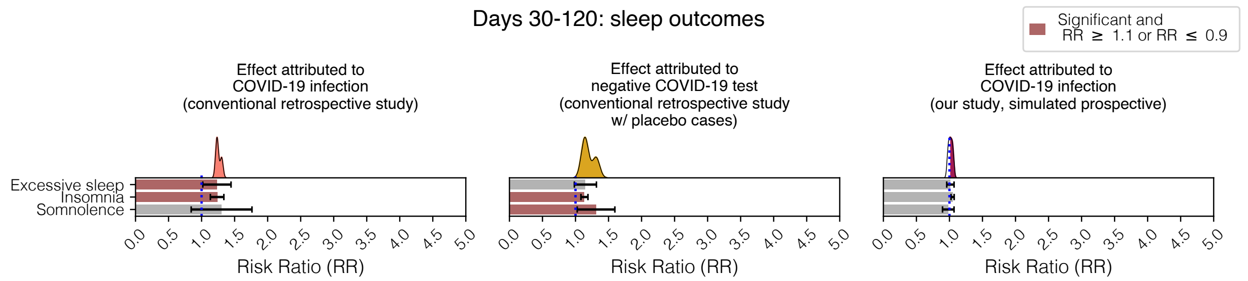


**Supplementary Figure S60:** Risk ratios for all outcomes related to sleep on days 30–120 post-enrollment. Effects are shown for a conventional retrospective study of long COVID (left), our test-based prospective study (right), and a conventional retrospective study with placebo cases (middle). Red bars indicate significant effects ($p<0.05$ with Bonferroni correction for multiple comparisons; two-sided bootstrap) with $RR \geq 1.1$ or $RR \leq 0.9$.

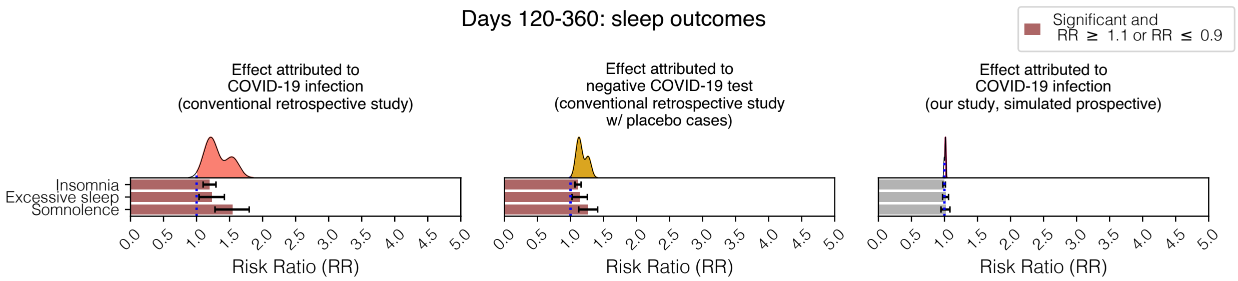


**Supplementary Figure S61:** Risk ratios for all outcomes related to sleep on days 120–360 post-enrollment. Effects are shown for a conventional retrospective study of long COVID (left), our test-based prospective study (right), and a conventional retrospective study with placebo cases (middle). Red bars indicate significant effects (p<0.05 with Bonferroni correction for multiple comparisons; two-sided bootstrap) with RR≥1.1 or RR≤0.9.

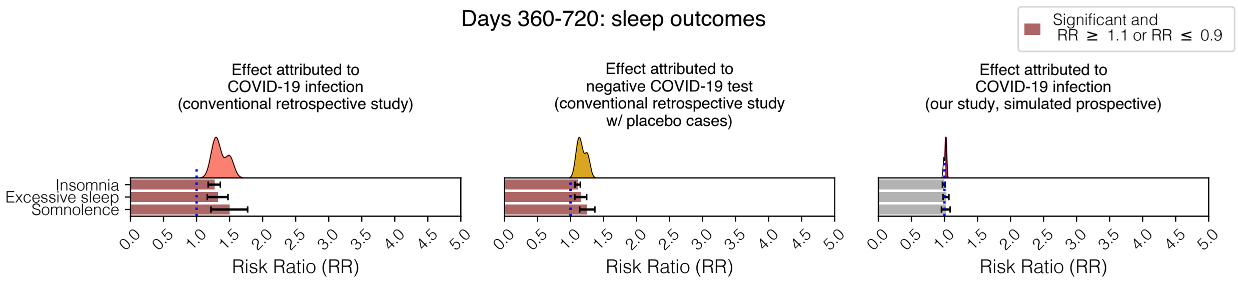


**Supplementary Figure S62:** Risk ratios for all outcomes related to sleep on days 360–720 post-enrollment. Effects are shown for a conventional retrospective study of long COVID (left), our test-based prospective study (right), and a conventional retrospective study with placebo cases (middle). Red bars indicate significant effects ($p<0.05$ with Bonferroni correction for multiple comparisons; two-sided bootstrap) with $RR \geq 1.1$ or $RR \leq 0.9$.

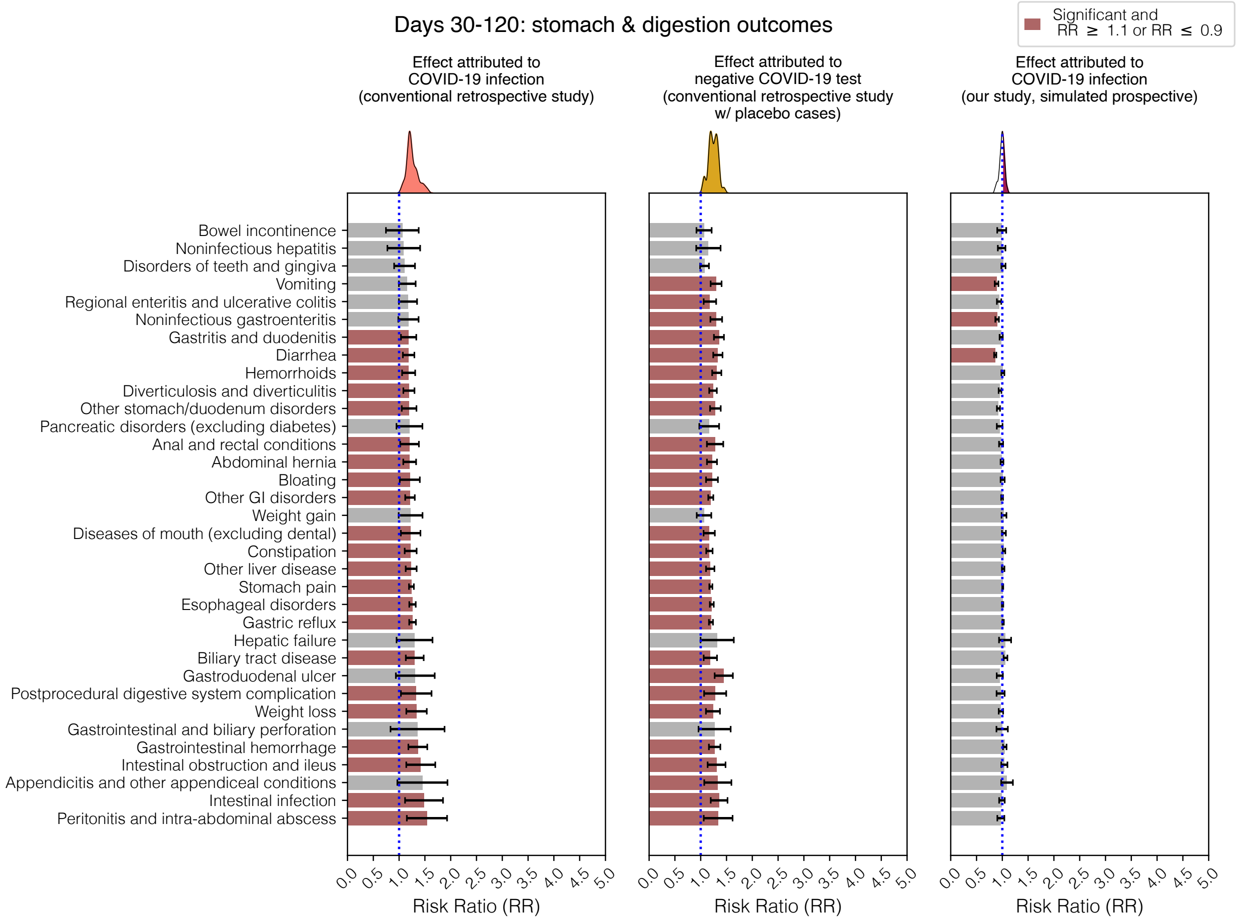


**Supplementary Figure S63:** Risk ratios for all outcomes related to stomach & digestion on days 30–120 post-enrollment. Effects are shown for a conventional retrospective study of long COVID (left), our test-based prospective study (right), and a conventional retrospective study with placebo cases (middle). Red bars indicate significant effects ($p<0.05$ with Bonferroni correction for multiple comparisons; two-sided bootstrap) with $RR \geq 1.1$ or $RR \leq 0.9$.

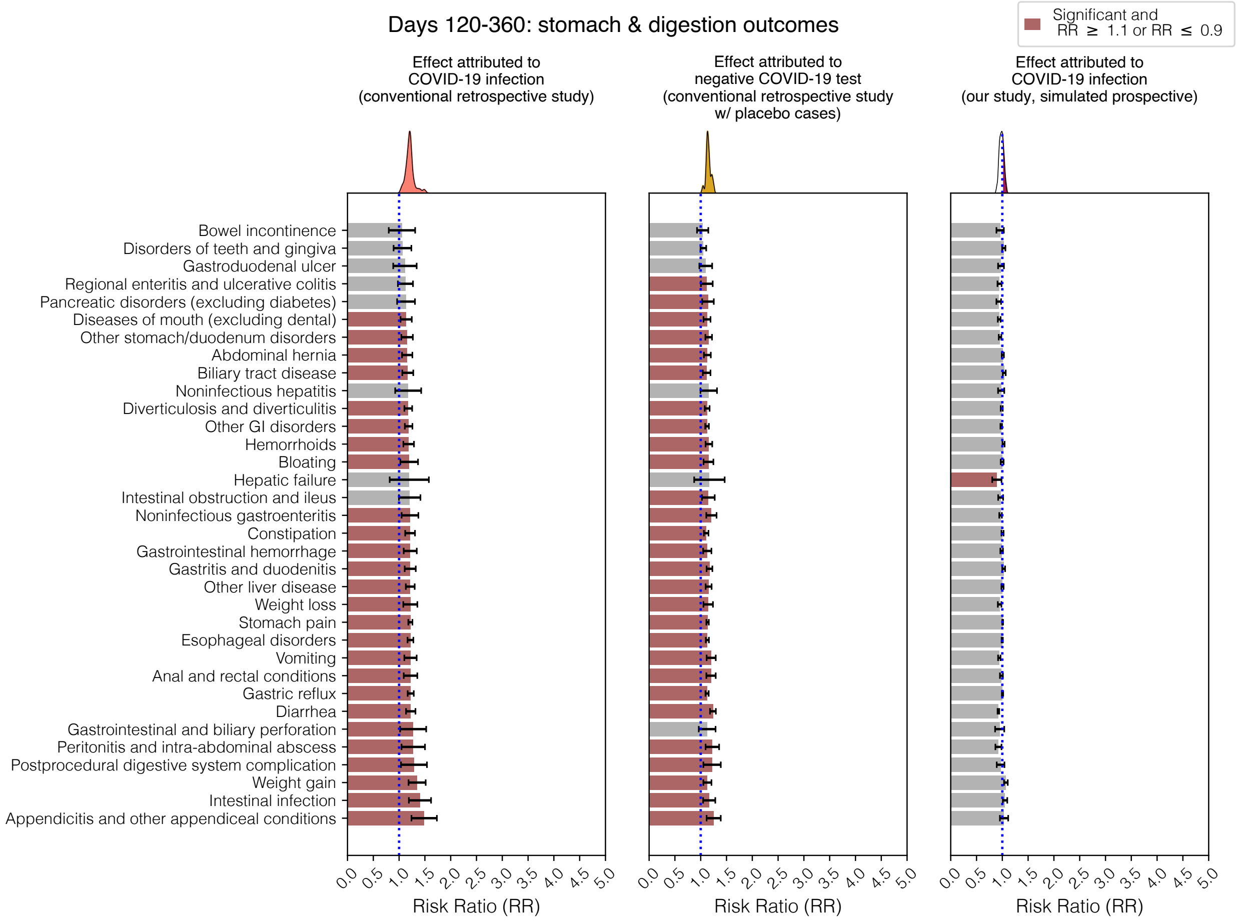


**Supplementary Figure S64:** Risk ratios for all outcomes related to stomach & digestion on days 120–360 post-enrollment. Effects are shown for a conventional retrospective study of long COVID (left), our test-based prospective study (right), and a conventional retrospective study with placebo cases (middle). Red bars indicate significant effects (p<0.05 with Bonferroni correction for multiple comparisons; two-sided bootstrap) with RR≥1.1 or RR≤0.9.

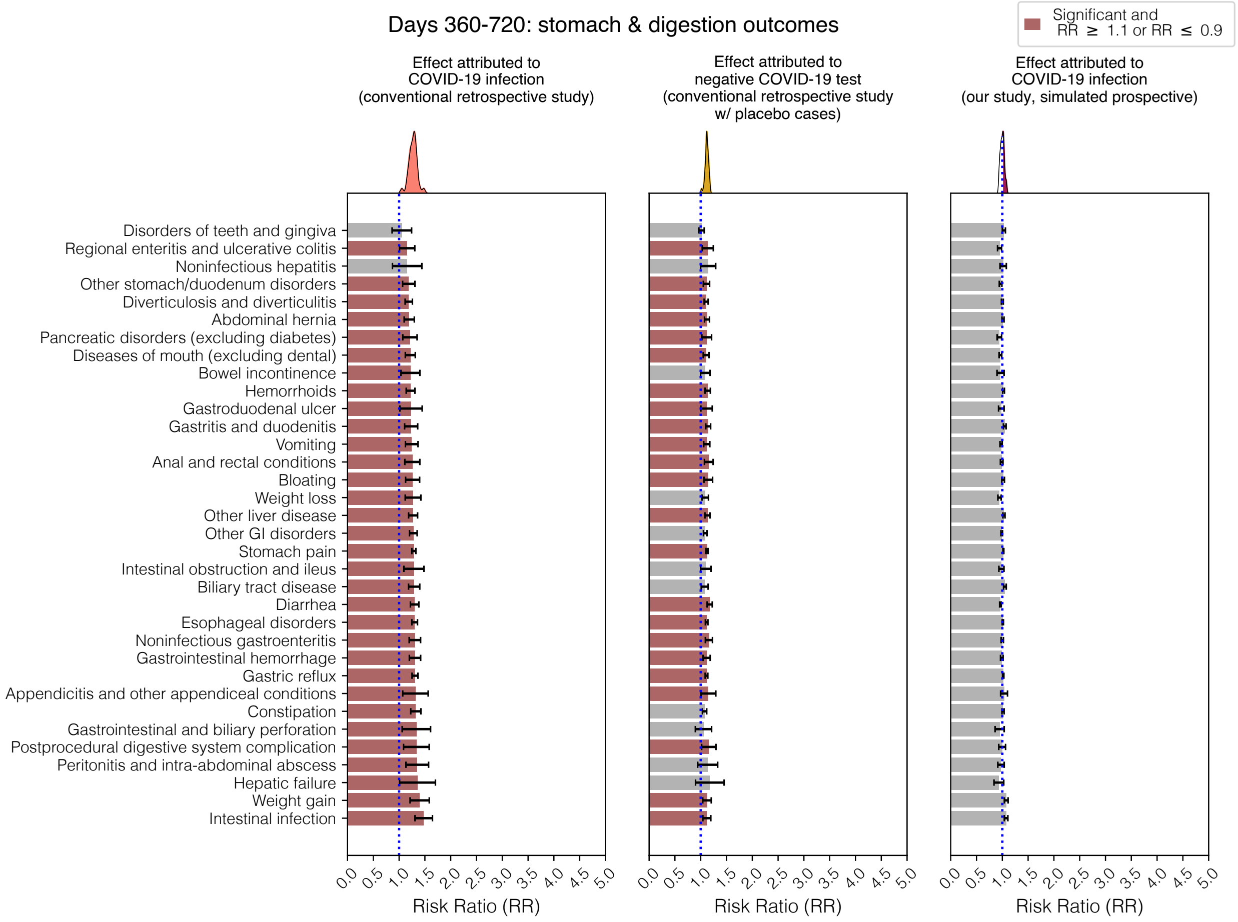


**Supplementary Figure S65:** Risk ratios for all outcomes related to stomach & digestion on days 360–720 post-enrollment. Effects are shown for a conventional retrospective study of long COVID (left), our test-based prospective study (right), and a conventional retrospective study with placebo cases (middle). Red bars indicate significant effects ($p<0.05$ with Bonferroni correction for multiple comparisons; two-sided bootstrap) with $RR \geq 1.1$ or $RR \leq 0.9$.

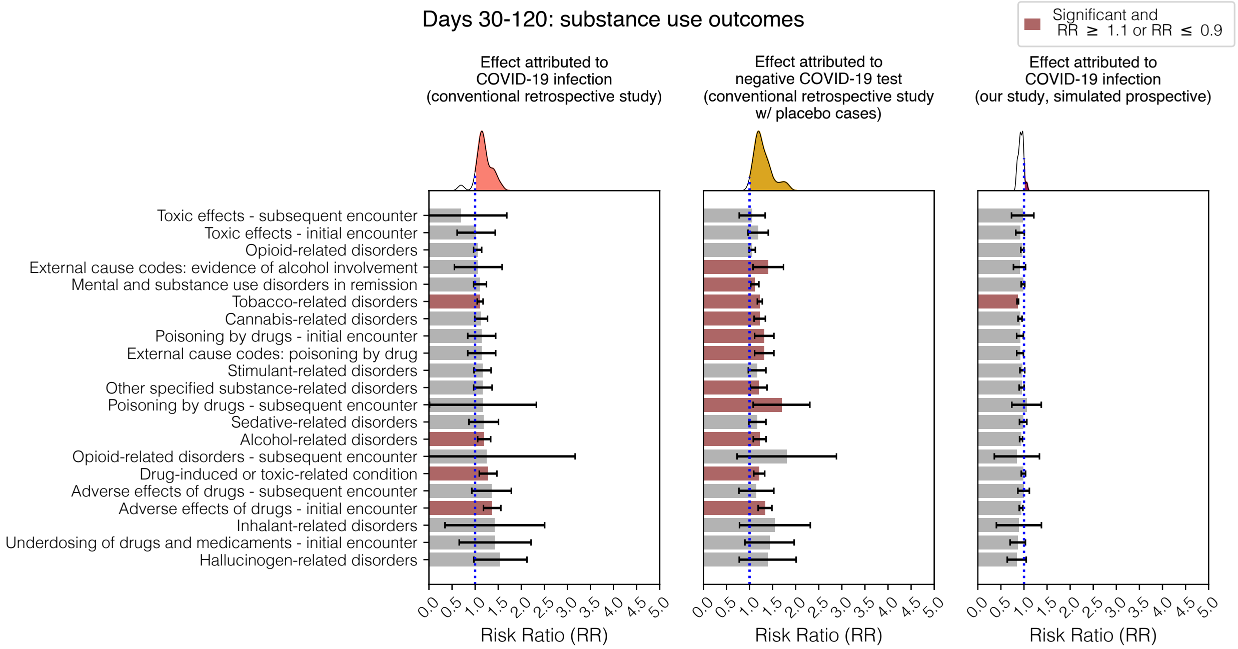


**Supplementary Figure S66:** Risk ratios for all outcomes related to substance use on days 30–120 post-enrollment. Effects are shown for a conventional retrospective study of long COVID (left), our test-based prospective study (right), and a conventional retrospective study with placebo cases (middle). Red bars indicate significant effects ($p<0.05$ with Bonferroni correction for multiple comparisons; two-sided bootstrap) with $RR \geq 1.1$ or $RR \leq 0.9$.

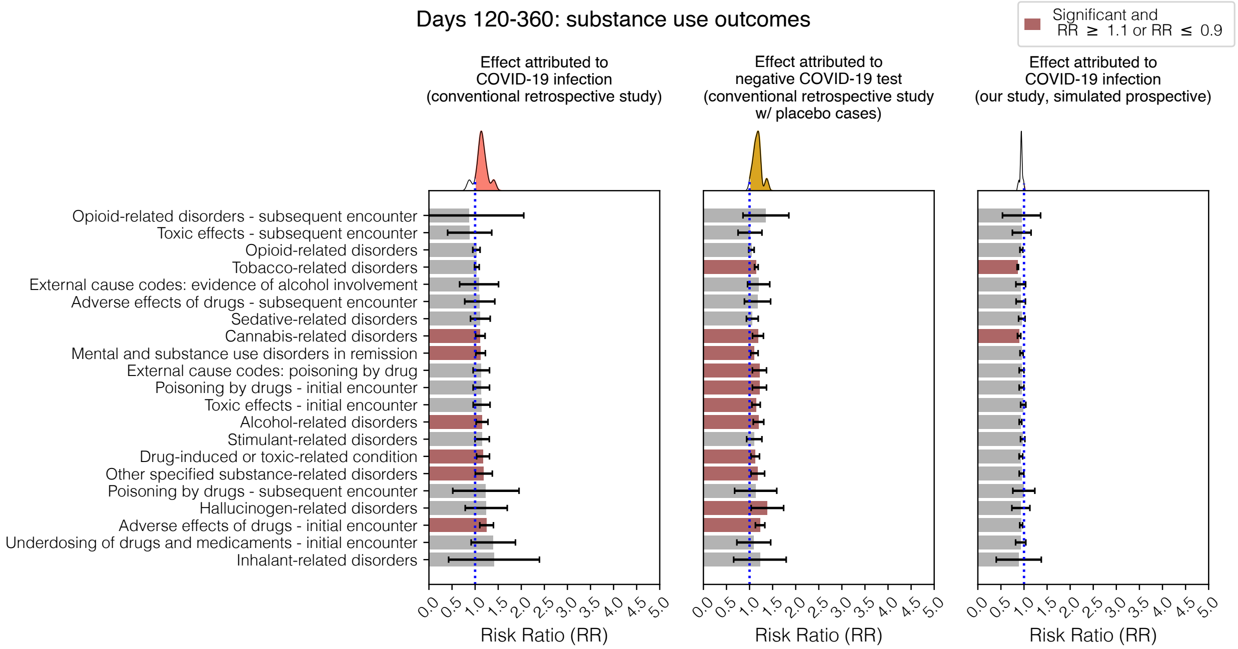


**Supplementary Figure S67:** Risk ratios for all outcomes related to substance use on days 120–360 post-enrollment. Effects are shown for a conventional retrospective study of long COVID (left), our test-based prospective study (right), and a conventional retrospective study with placebo cases (middle). Red bars indicate significant effects ($p<0.05$ with Bonferroni correction for multiple comparisons; two-sided bootstrap) with RR$\geq$1.1 or RR$\leq$0.9.

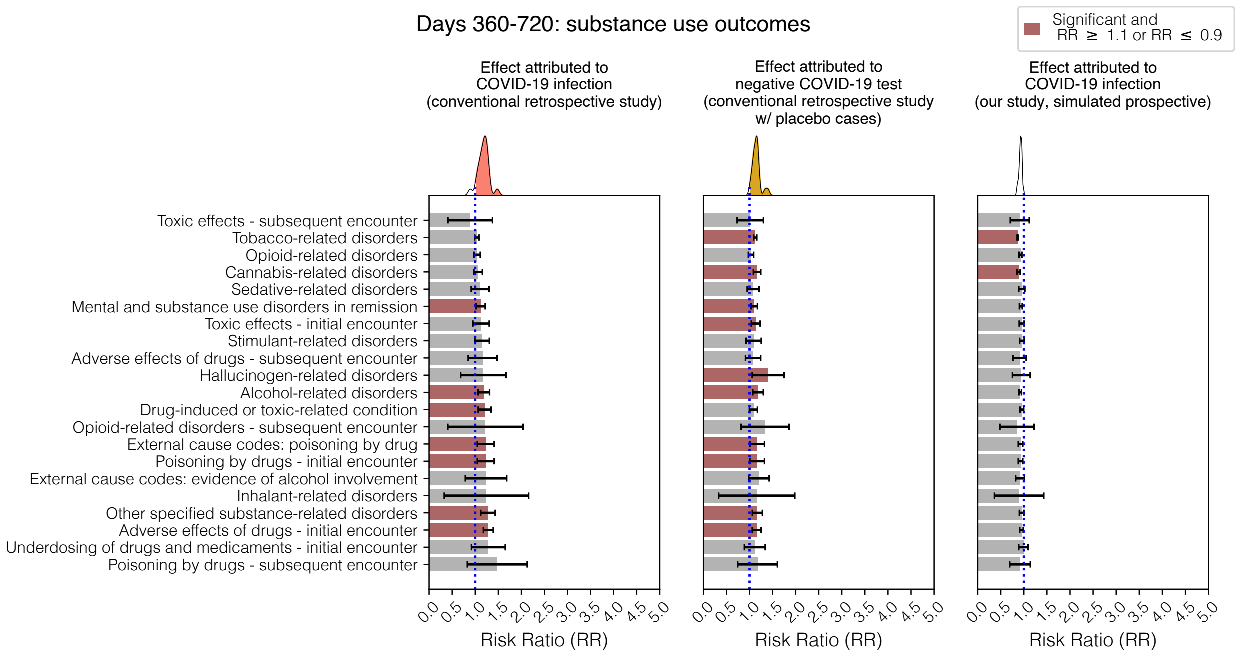


**Supplementary Figure S68:** Risk ratios for all outcomes related to substance use on days 360–720 post-enrollment. Effects are shown for a conventional retrospective study of long COVID (left), our test-based prospective study (right), and a conventional retrospective study with placebo cases (middle). Red bars indicate significant effects ($p<0.05$ with Bonferroni correction for multiple comparisons; two-sided bootstrap) with RR$\geq$1.1 or RR$\leq$0.9.

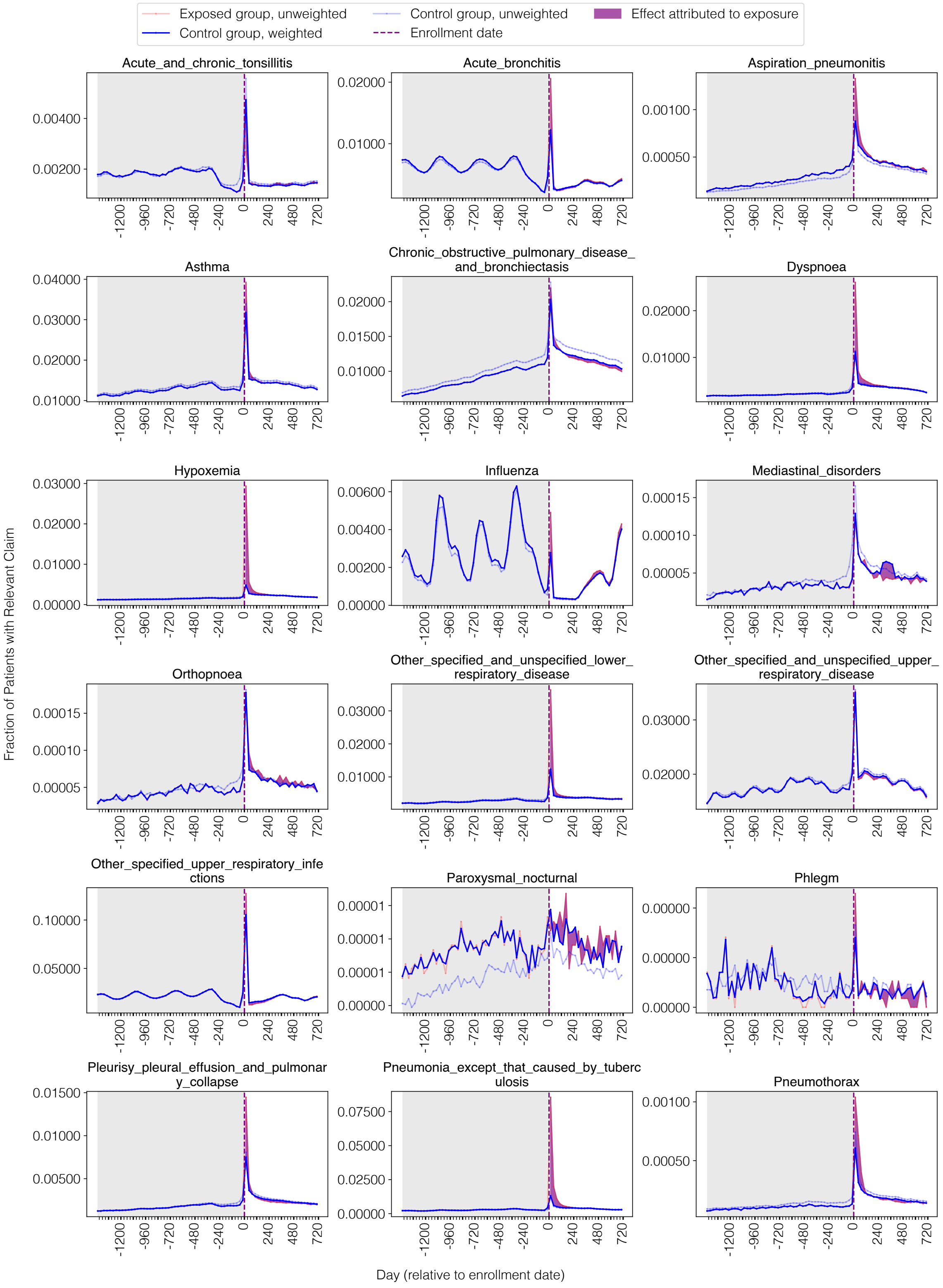


**Supplementary Figure S69:** Temporal trends in Breathing prevalence. The fraction of patients with relevant claims over time belonging for each outcome in the **prospective study** with testing-based enrollment experiment.

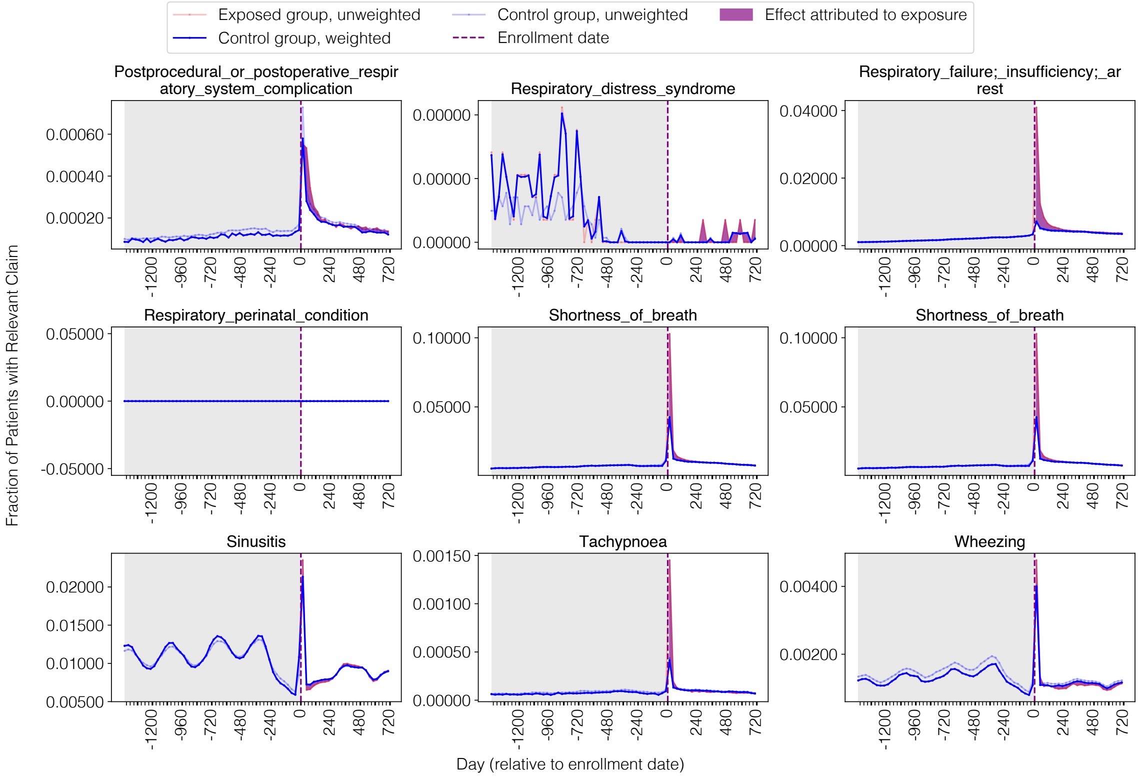


**Supplementary Figure S69:** (cont'd) Temporal trends in Breathing prevalence. The fraction of patients with relevant claims over time belonging for each outcome in the **prospective study** with testing-based enrollment experiment.

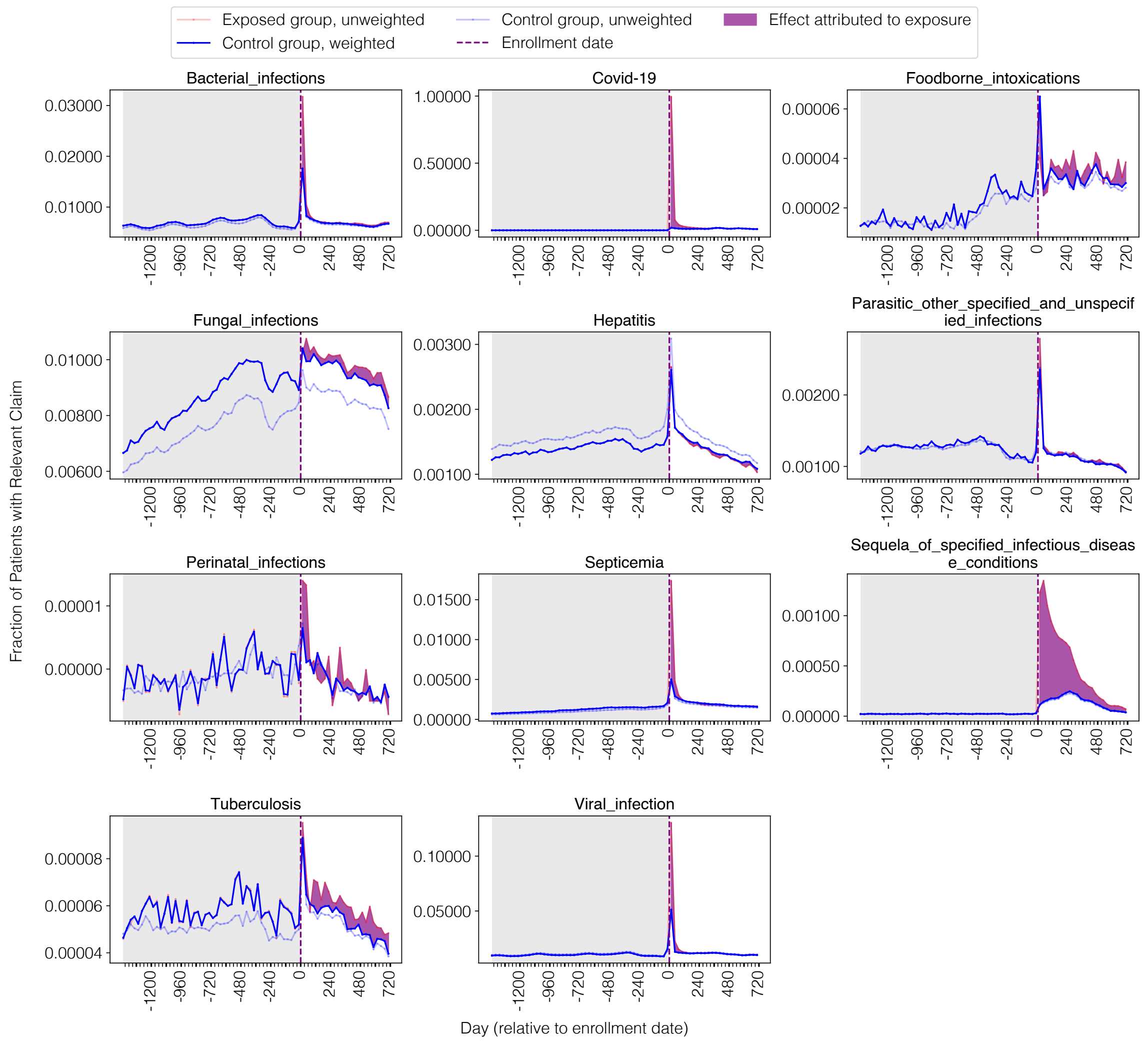


**Supplementary Figure S70:** Temporal trends in Certain Infectious and Parasitic Diseases prevalence. The fraction of patients with relevant claims over time belonging for each outcome in the **prospective study** with testing-based enrollment experiment.

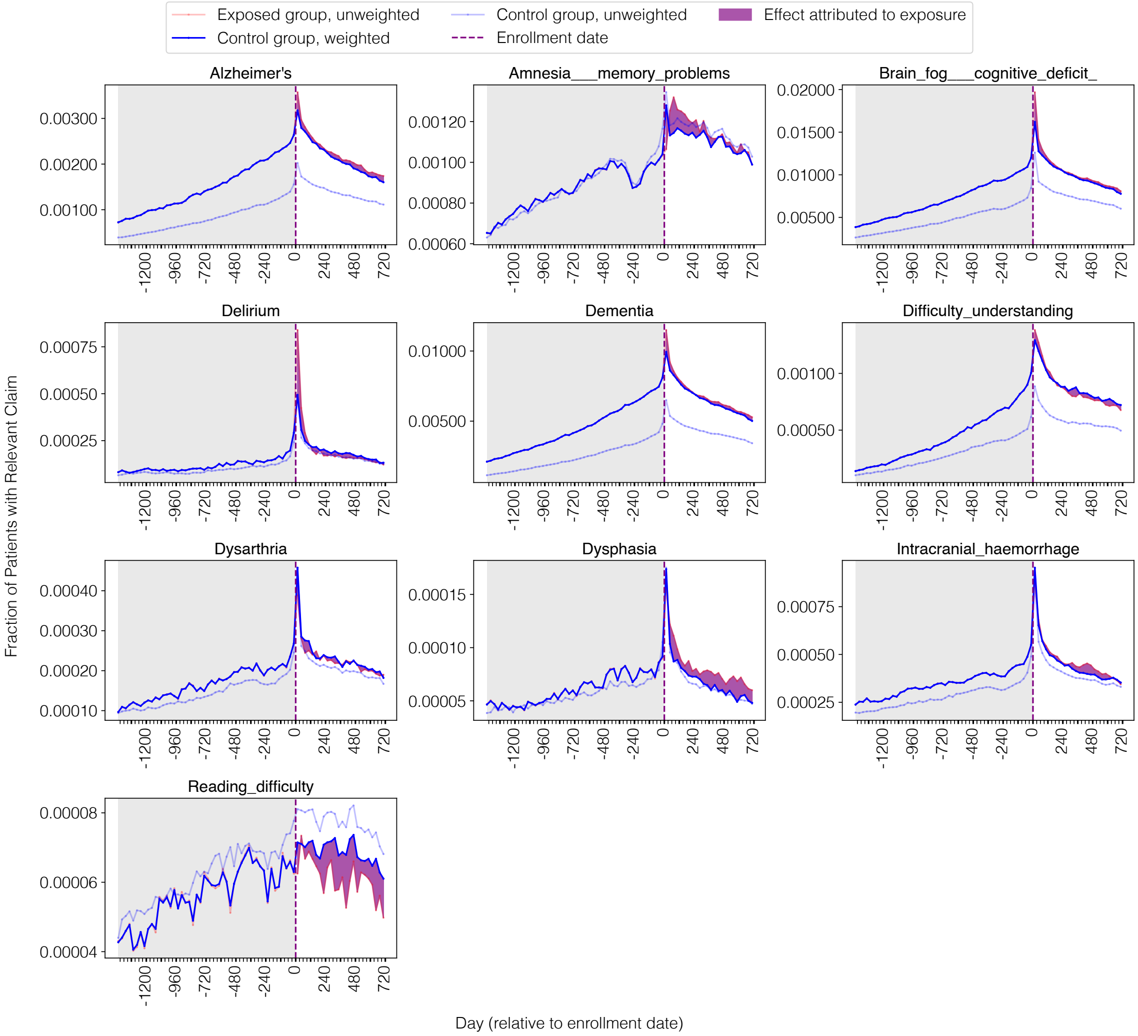


**Supplementary Figure S71:** Temporal trends in Cognitive health prevalence. The fraction of patients with relevant claims over time belonging for each outcome in the **prospective study** with testing-based enrollment experiment.

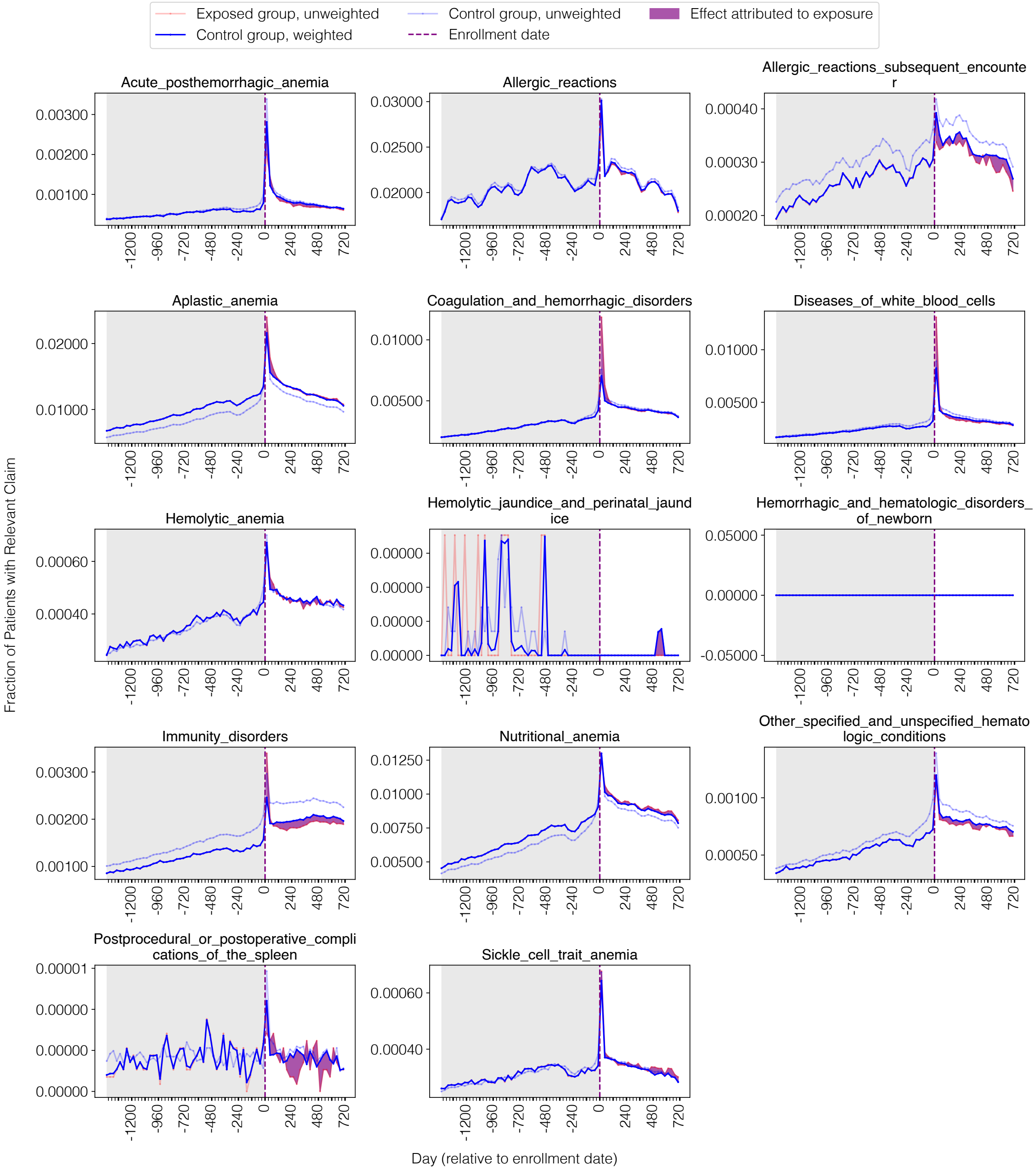


**Supplementary Figure S72:** Temporal trends in Diseases of the Blood and Blood Forming Organs and Certain Disorders Involving the Immune Mechanism prevalence. The fraction of patients with relevant claims over time belonging for each outcome in the **prospective study** with testing-based enrollment experiment.

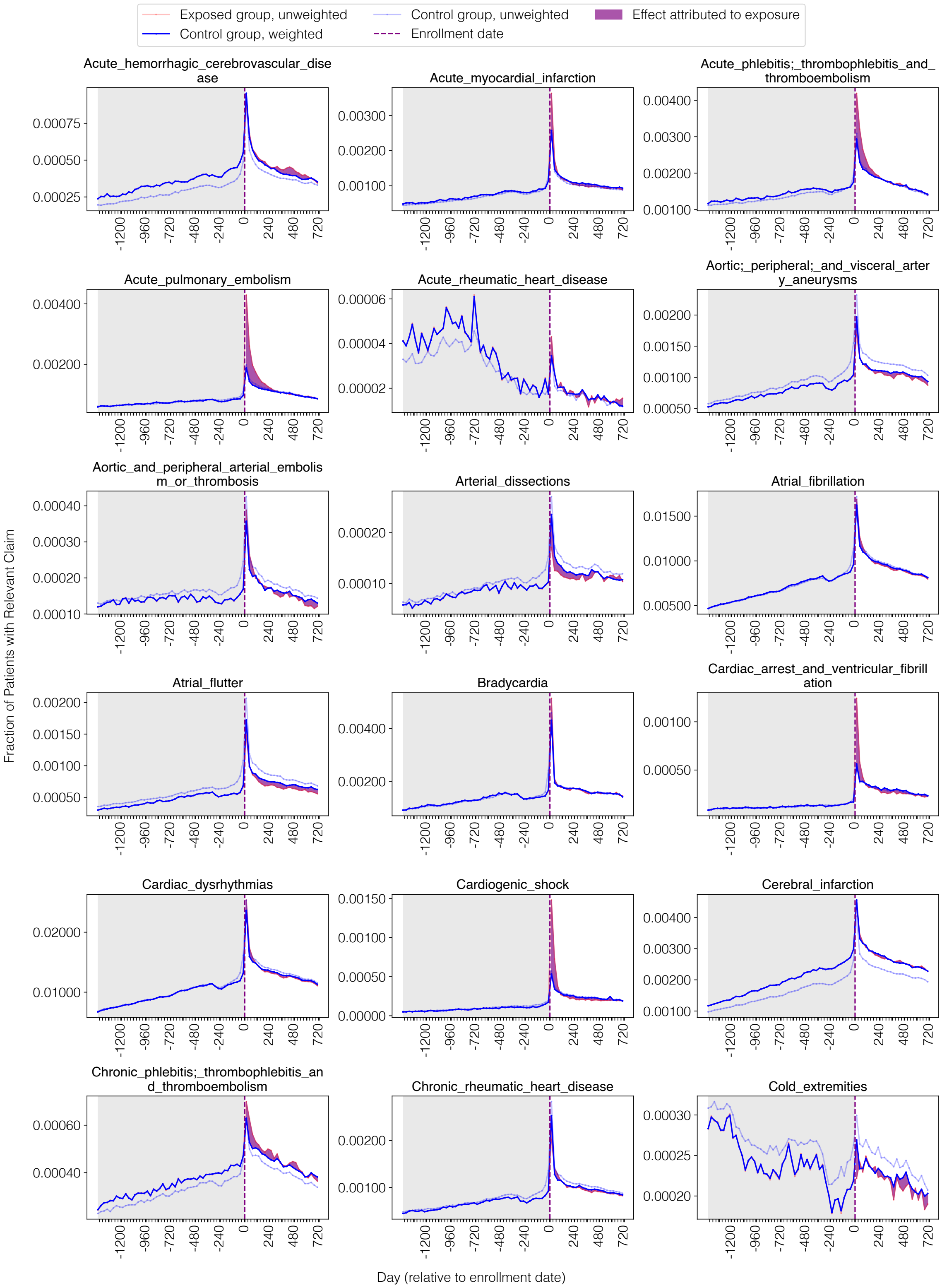


**Supplementary Figure S73:** Temporal trends in Diseases of the Circulatory System prevalence. The fraction of patients with relevant claims over time belonging for each outcome in the **prospective study** with testing-based enrollment experiment.

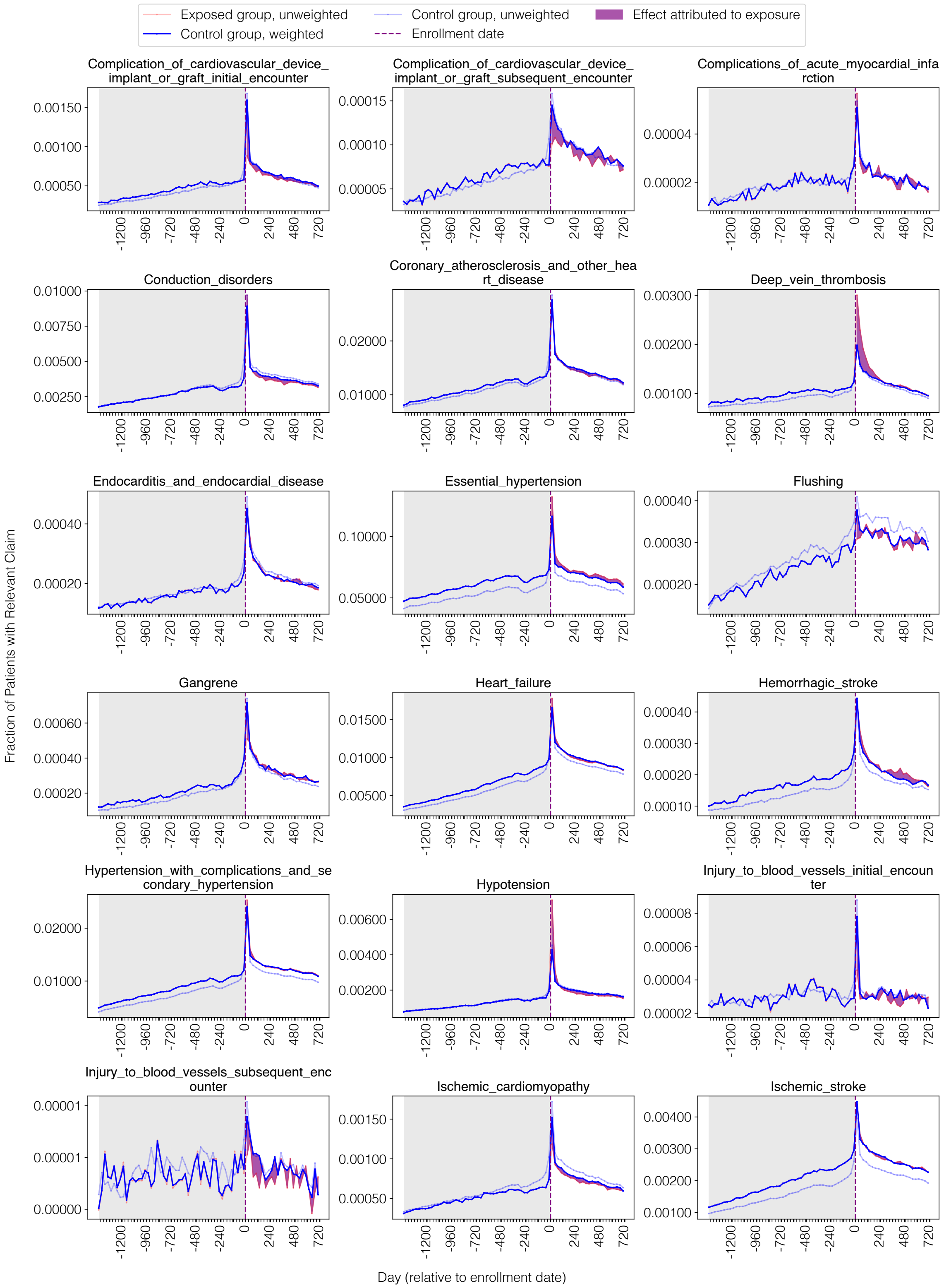


**Supplementary Figure S73:** (cont'd) Temporal trends in Diseases of the Circulatory System prevalence. The fraction of patients with relevant claims over time belonging for each outcome in the **prospective study** with testing-based enrollment experiment.

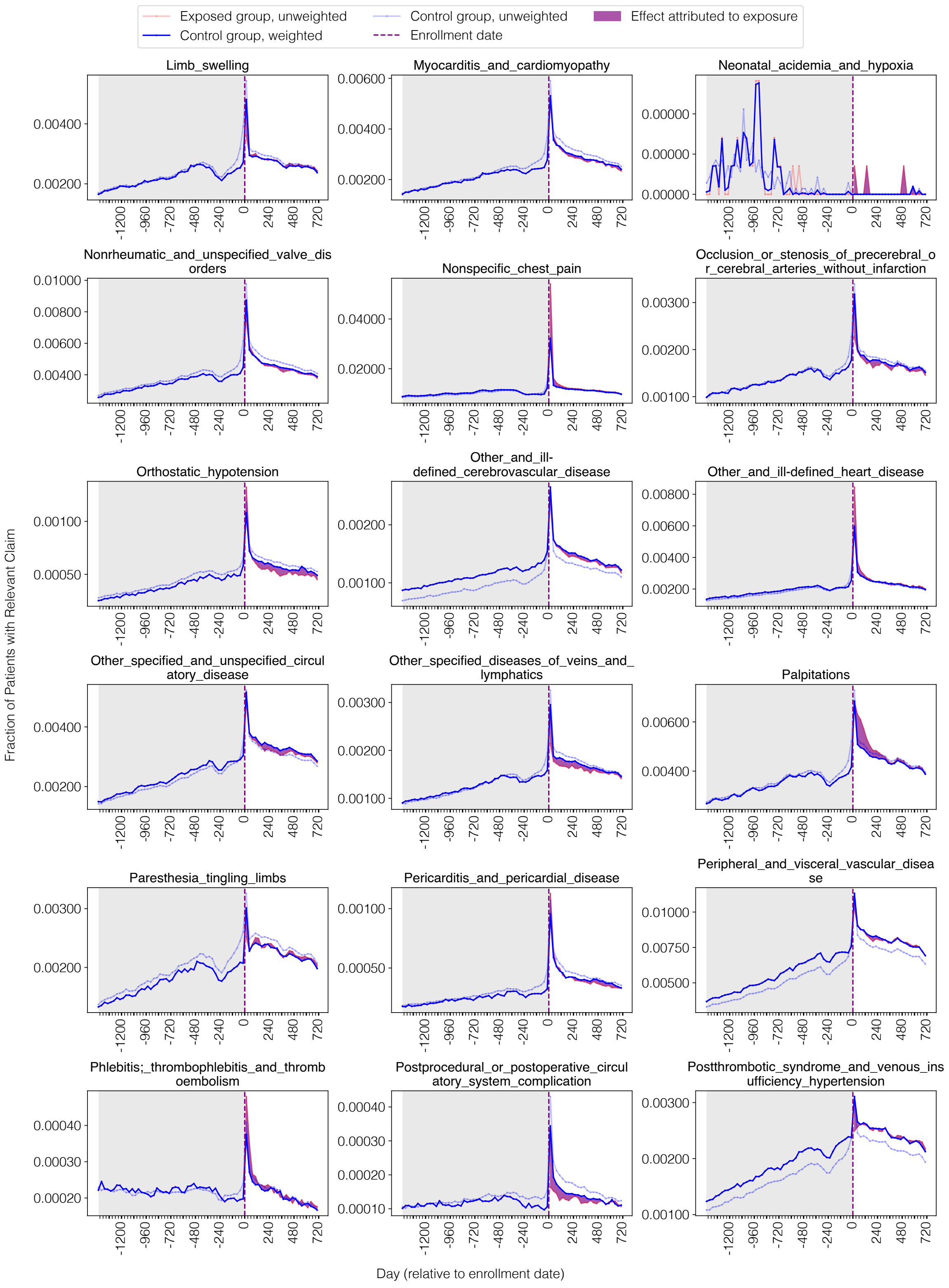


**Supplementary Figure S73:** (cont'd) Temporal trends in Diseases of the Circulatory System prevalence. The fraction of patients with relevant claims over time belonging for each outcome in the **prospective study** with testing-based enrollment experiment.

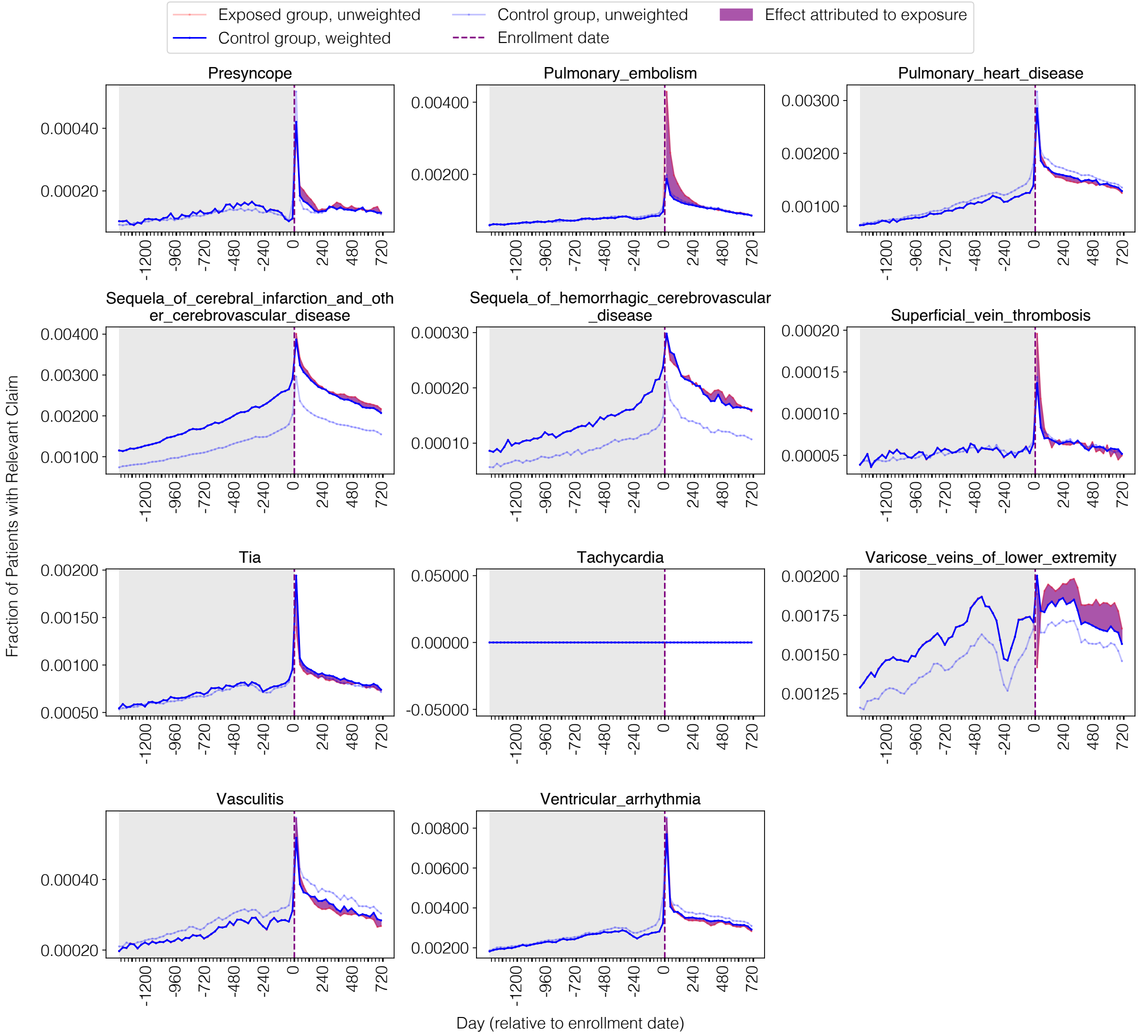


**Supplementary Figure S73:** (cont'd) Temporal trends in Diseases of the Circulatory System prevalence. The fraction of patients with relevant claims over time belonging for each outcome in the **prospective study** with testing-based enrollment experiment.

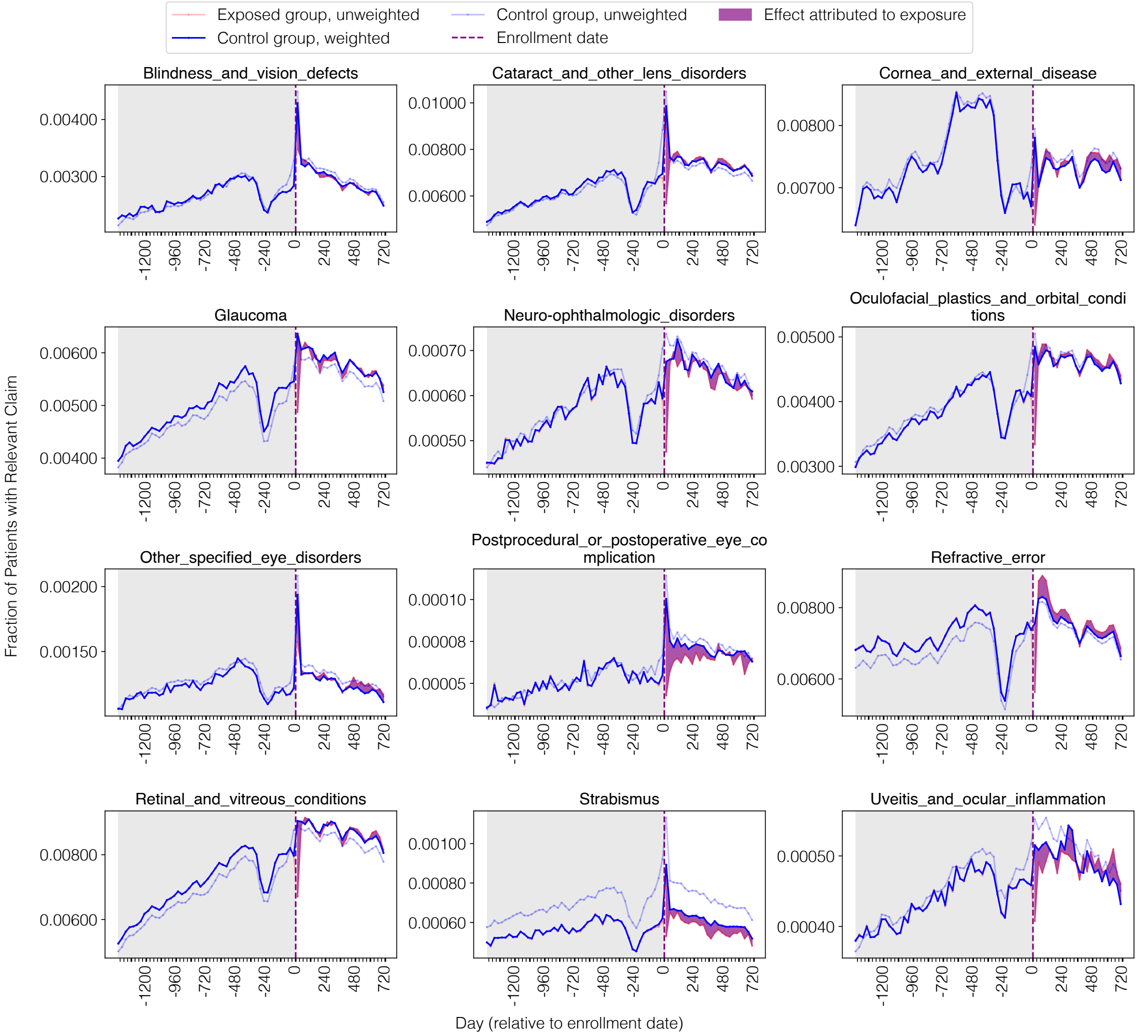


**Supplementary Figure S74:** Temporal trends in Diseases of the Eye and Adnexa prevalence. The fraction of patients with relevant claims over time belonging for each outcome in the **prospective study** with testing-based enrollment experiment.

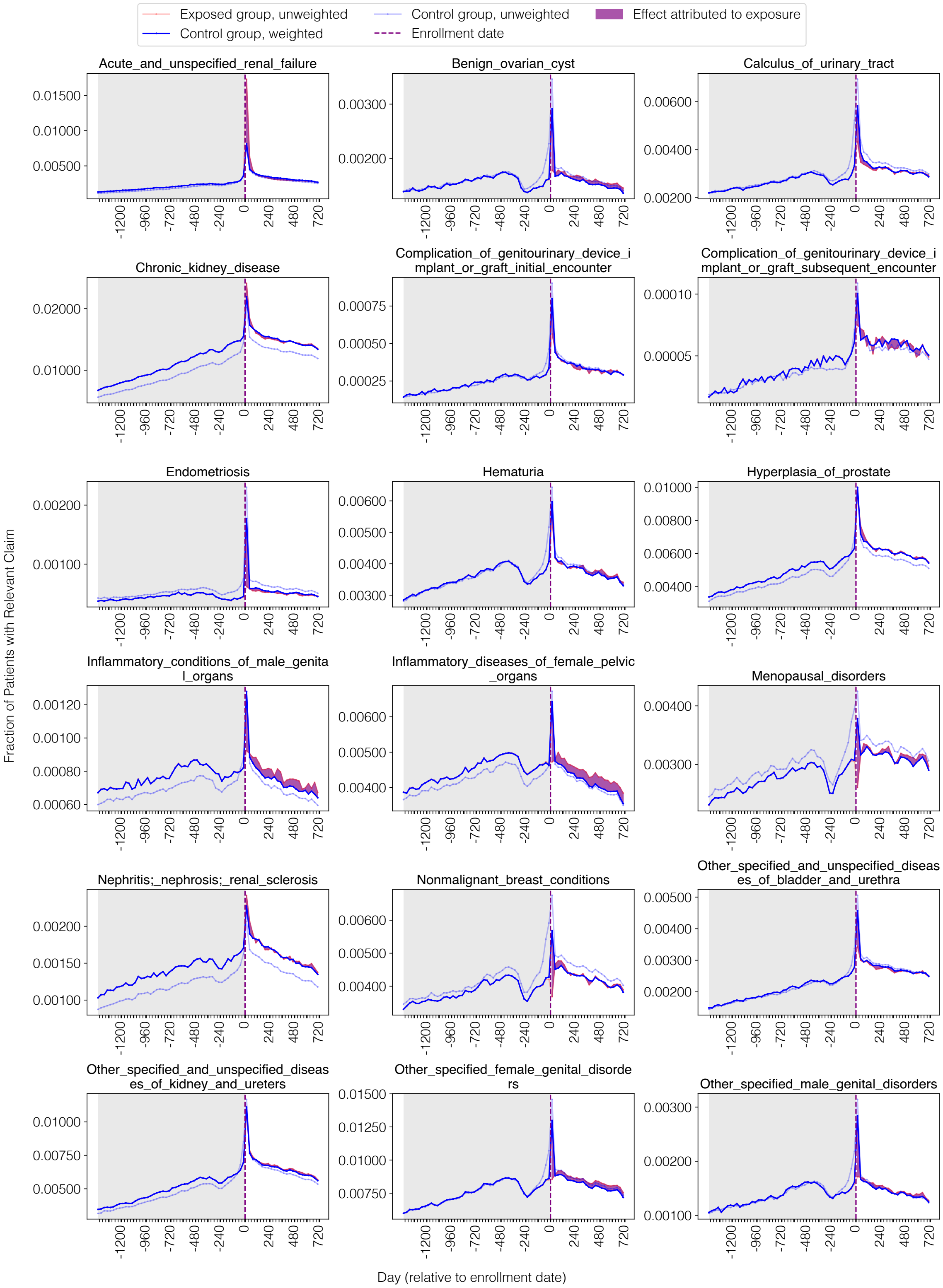


**Supplementary Figure S75:** Temporal trends in Diseases of the Genitourinary System prevalence. The fraction of patients with relevant claims over time belonging for each outcome in the **prospective study** with testing-based enrollment experiment.

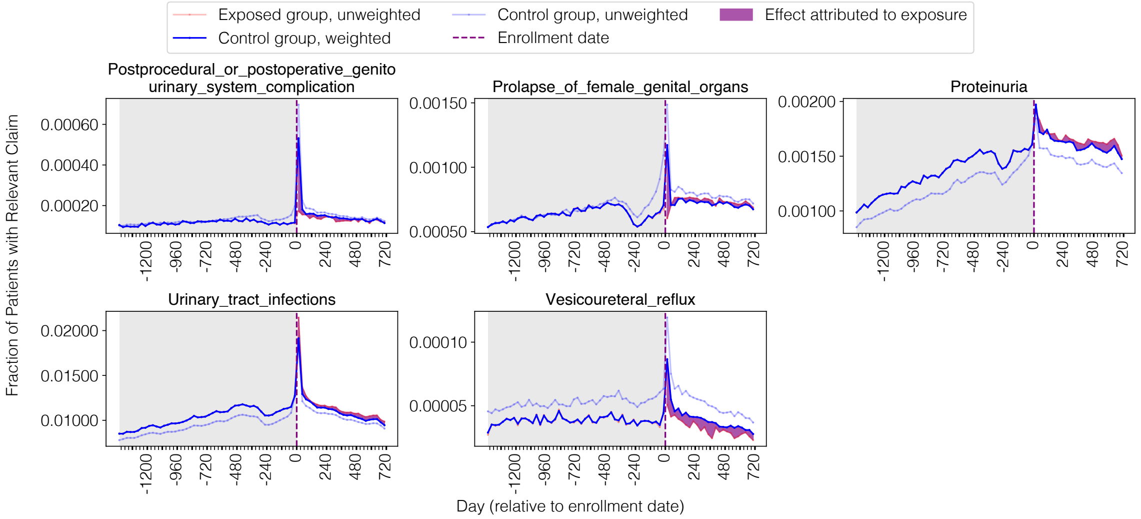


**Supplementary Figure S75:** (cont'd) Temporal trends in Diseases of the Genitourinary System prevalence. The fraction of patients with relevant claims over time belonging for each outcome in the **prospective study** with testing-based enrollment experiment.

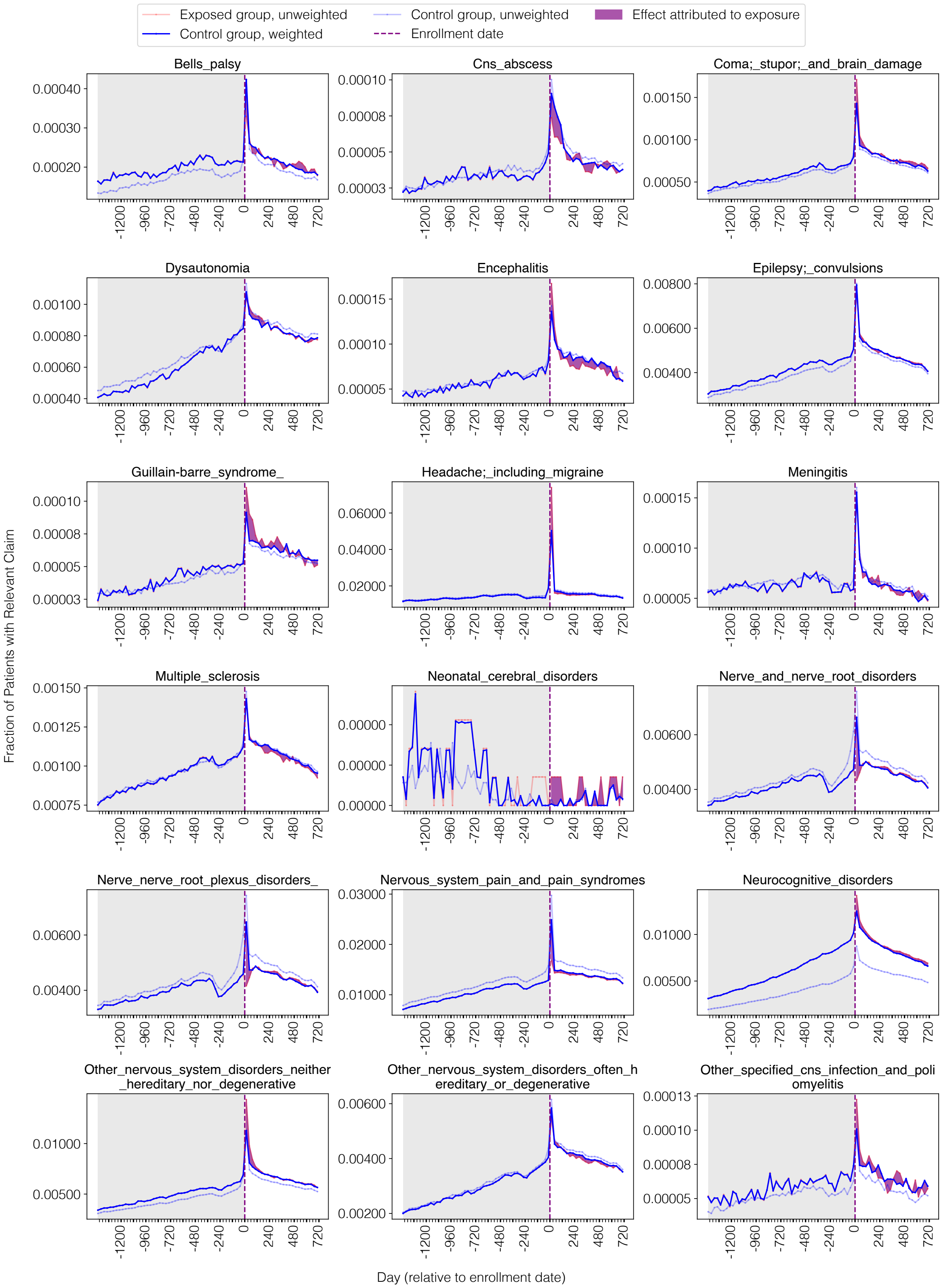


**Supplementary Figure S76:** Temporal trends in Diseases of the Nervous System prevalence. The fraction of patients with relevant claims over time belonging for each outcome in the **prospective study** with testing-based enrollment experiment.

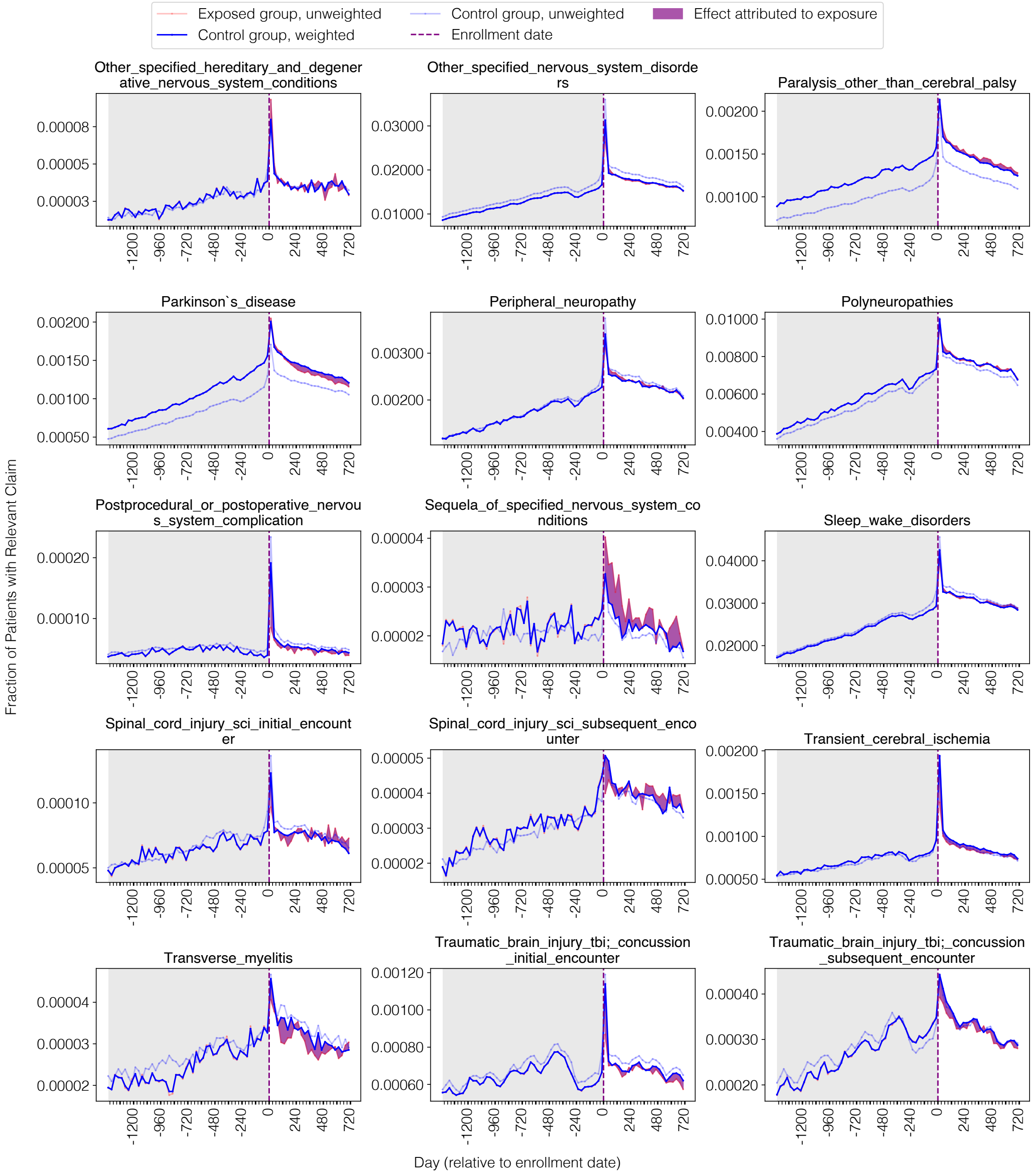


**Supplementary Figure S76:** (cont'd) Temporal trends in Diseases of the Nervous System prevalence. The fraction of patients with relevant claims over time belonging for each outcome in the **prospective study** with testing-based enrollment experiment.

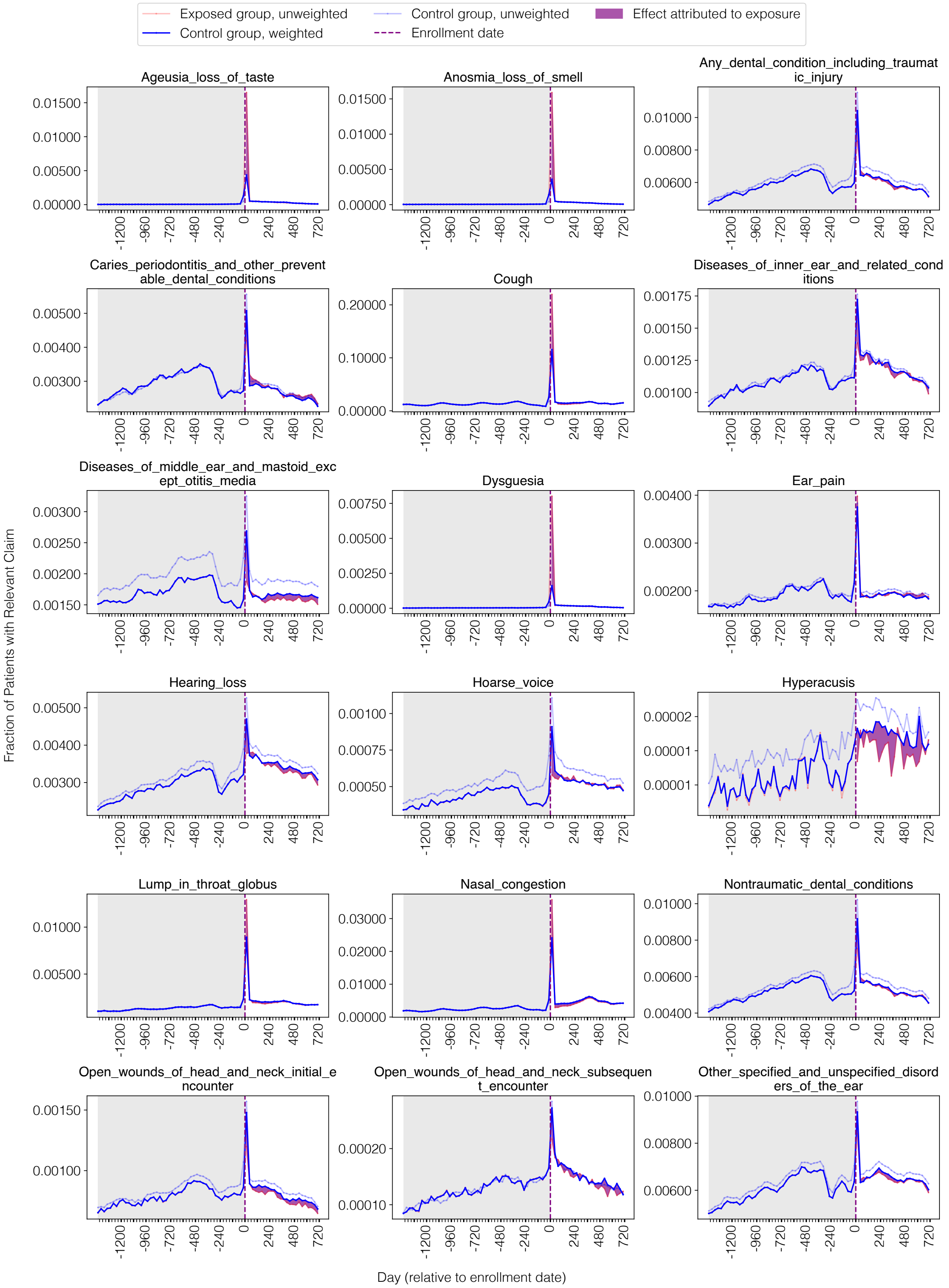


**Supplementary Figure S77:** Temporal trends in Ear, nose and throat prevalence. The fraction of patients with relevant claims over time belonging for each outcome in the **prospective study** with testing-based enrollment experiment.

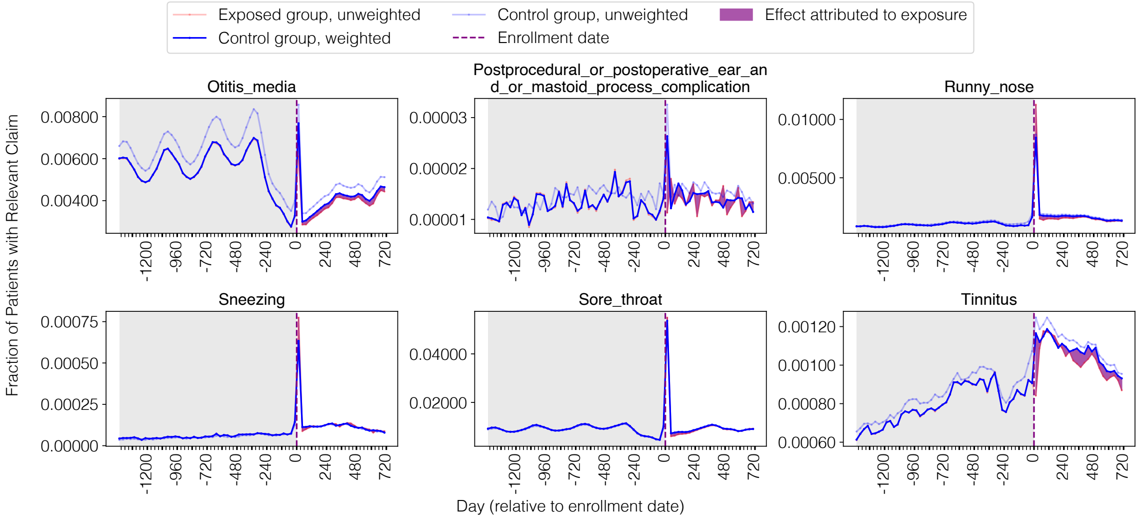


**Supplementary Figure S77:** (cont'd) Temporal trends in Ear, nose and throat prevalence. The fraction of patients with relevant claims over time belonging for each outcome in the **prospective study** with testing-based enrollment experiment.

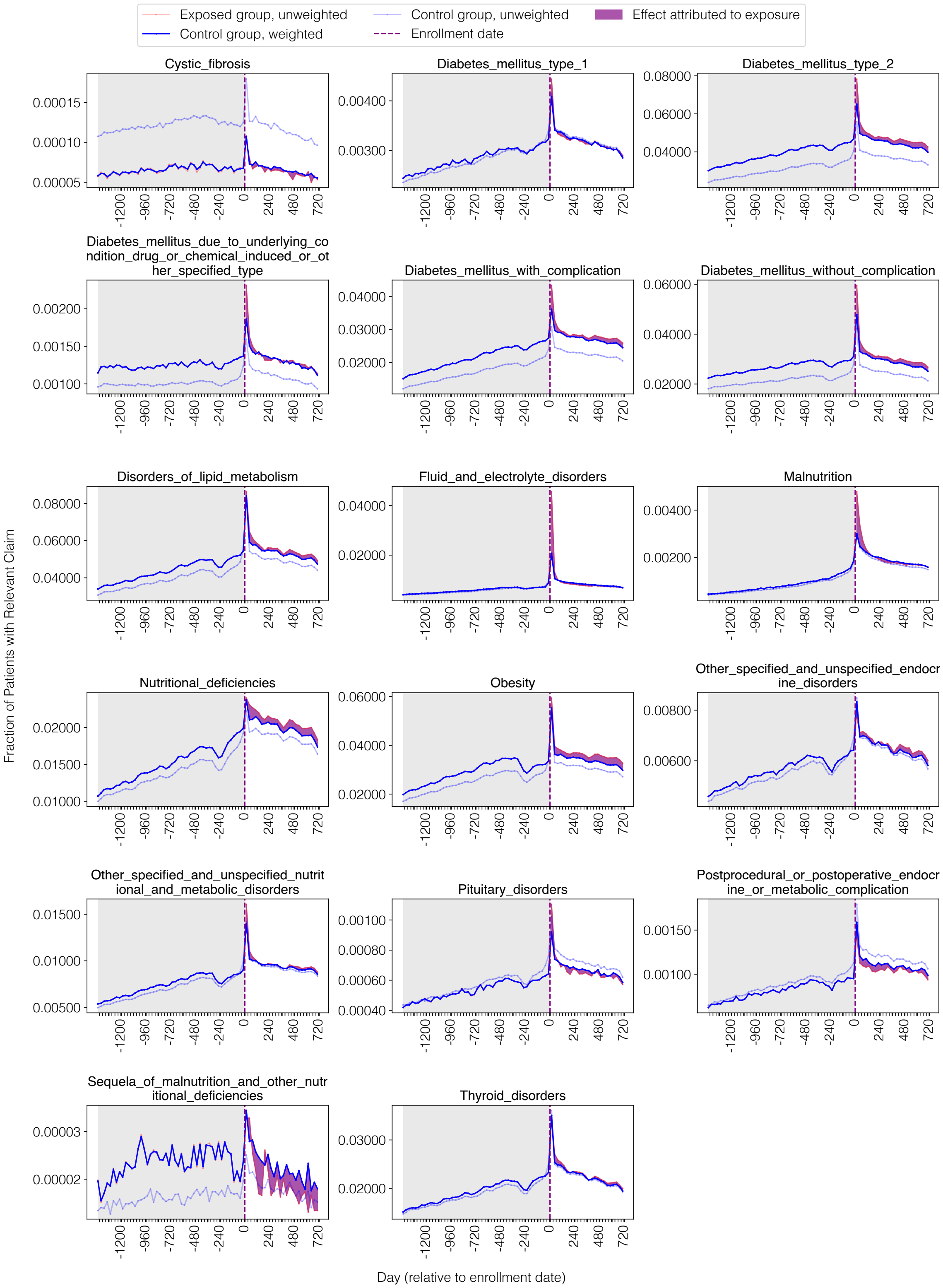


**Supplementary Figure S78:** Temporal trends in Endocrine, Nutritional and Metabolic Diseases prevalence. The fraction of patients with relevant claims over time belonging for each outcome in the **prospective study** with testing-based enrollment experiment.

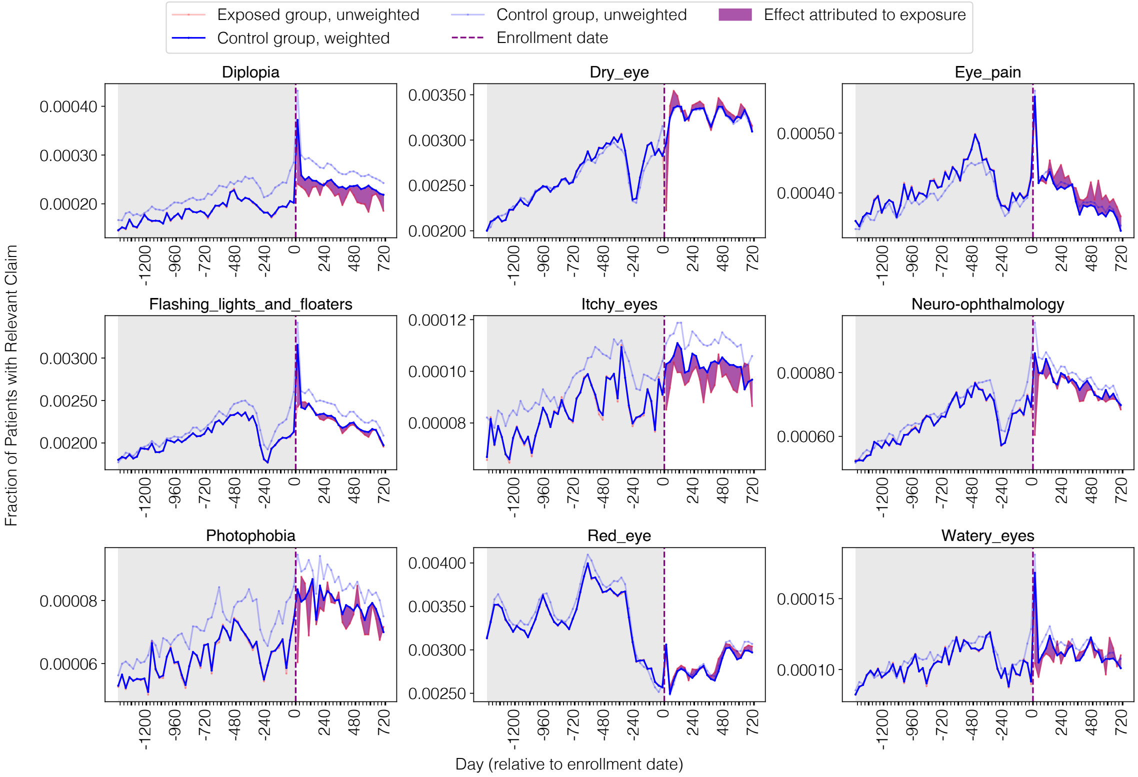


**Supplementary Figure S79:** Temporal trends in Eyes prevalence. The fraction of patients with relevant claims over time belonging for each outcome in the **prospective study** with testing-based enrollment experiment.

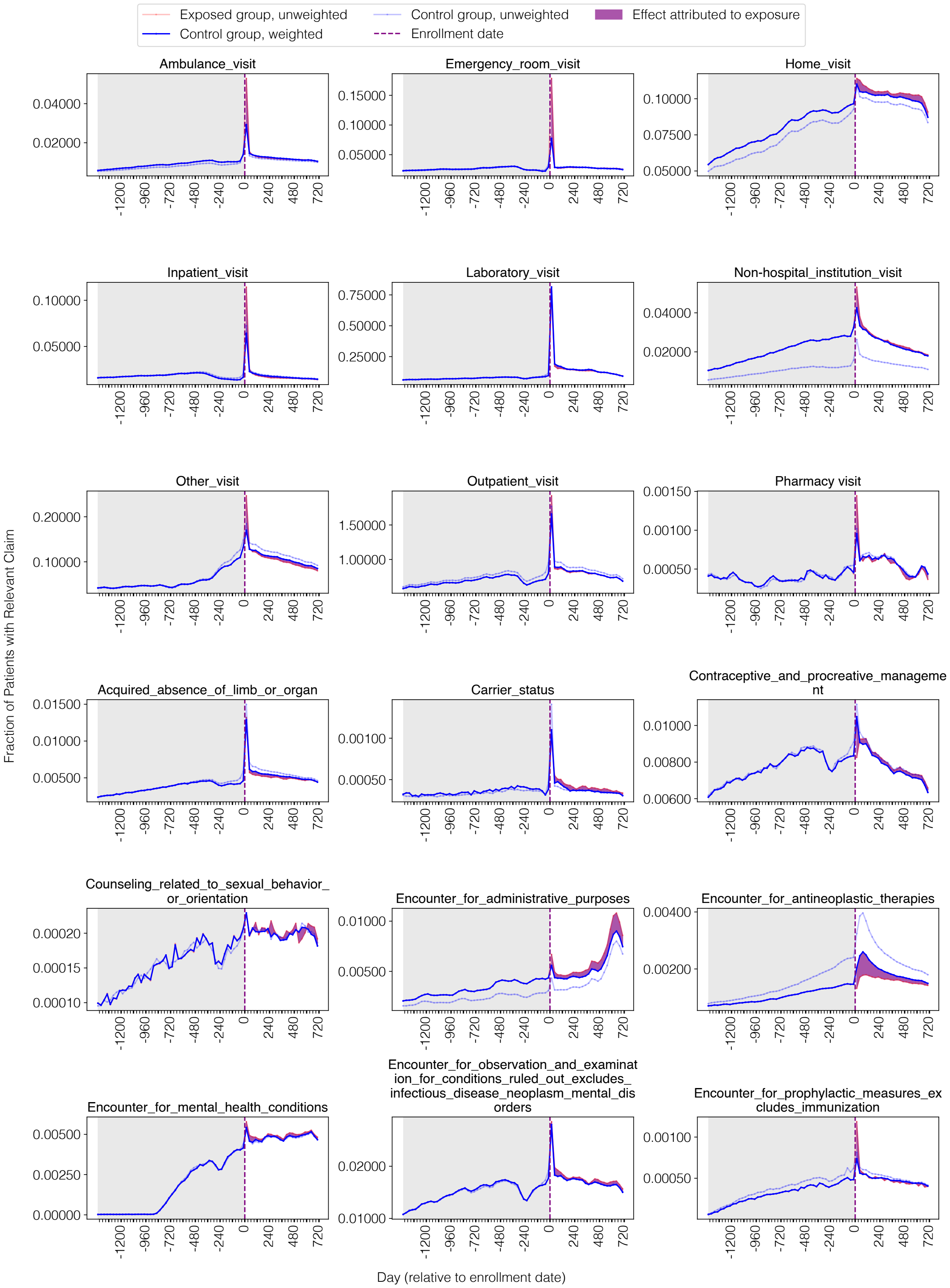


**Supplementary Figure S80:** Temporal trends in Factors Influencing Health Status and Contact with Health Services prevalence. The fraction of patients with relevant claims over time belonging for each outcome in the **prospective study** with testing-based enrollment experiment.

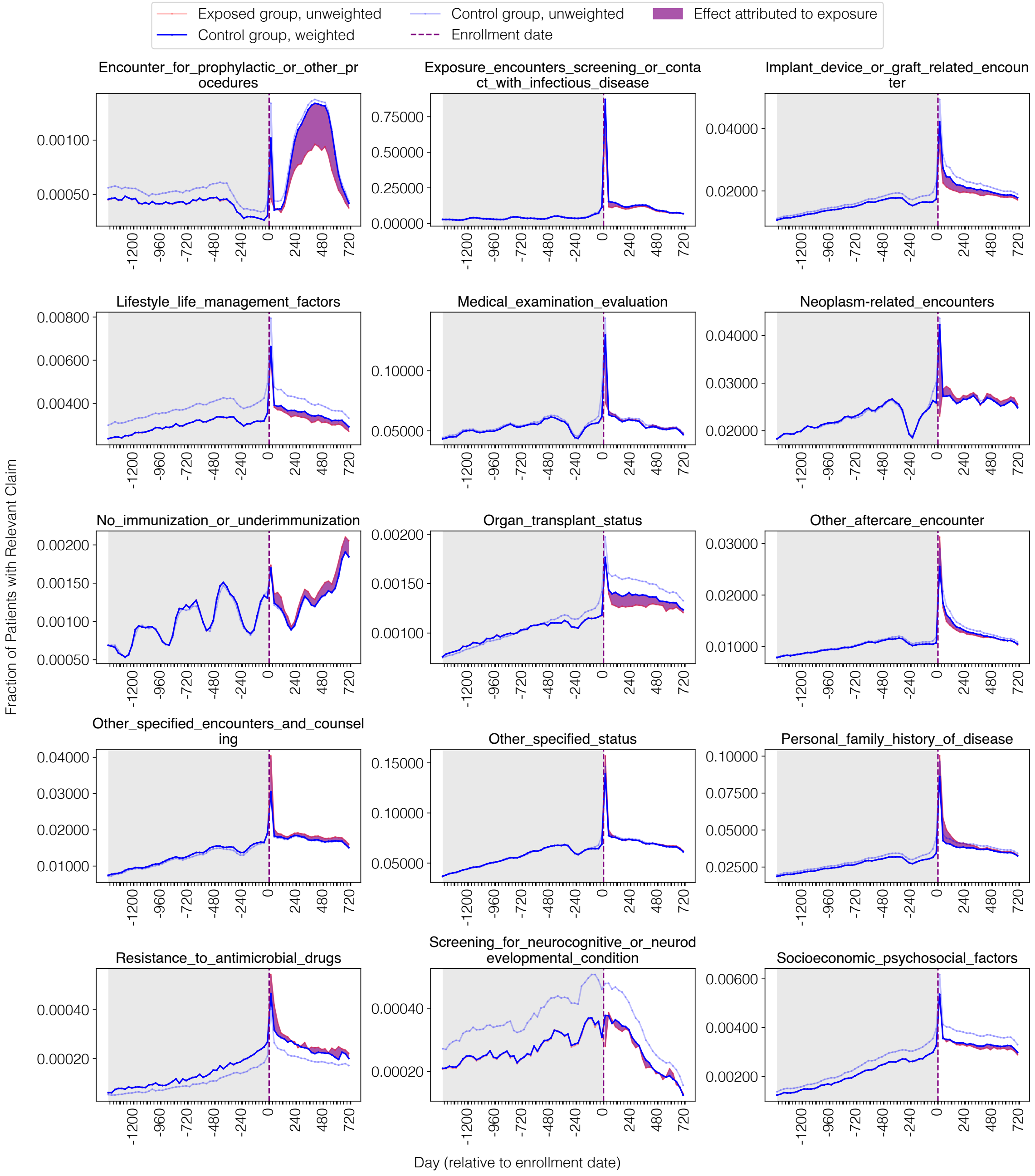


**Supplementary Figure S80:** (cont'd) Temporal trends in Factors Influencing Health Status and Contact with Health Services prevalence. The fraction of patients with relevant claims over time belonging for each outcome in the **prospective study** with testing-based enrollment experiment.

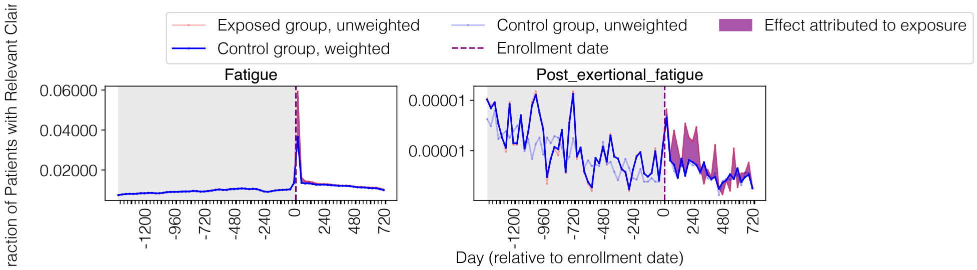


**Supplementary Figure S81:** Temporal trends in Fatigue prevalence. The fraction of patients with relevant claims over time belonging for each outcome in the **prospective study** with testing-based enrollment experiment.

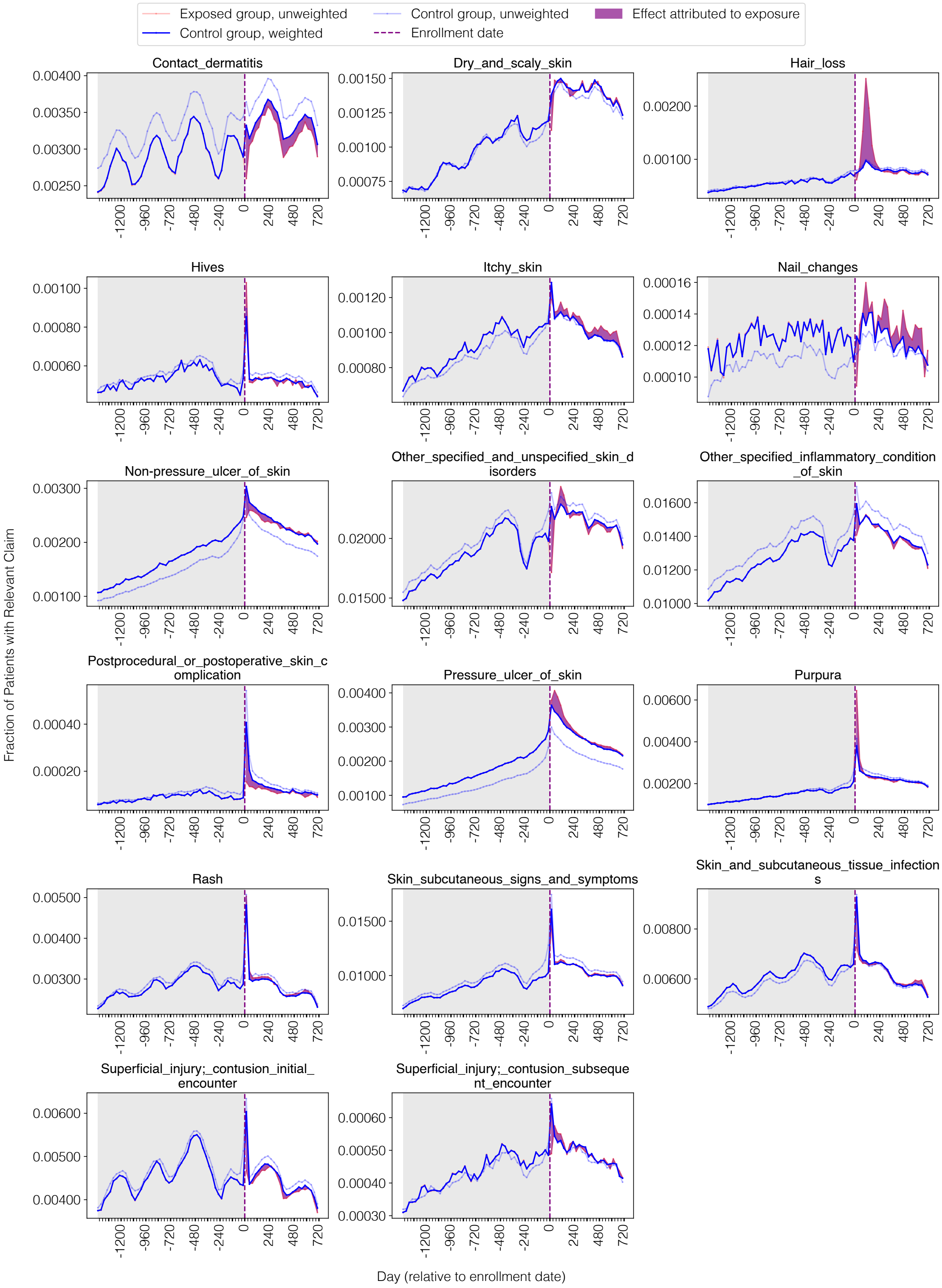


**Supplementary Figure S82:** Temporal trends in Hair, skin and nails prevalence. The fraction of patients with relevant claims over time belonging for each outcome in the **prospective study** with testing-based enrollment experiment.

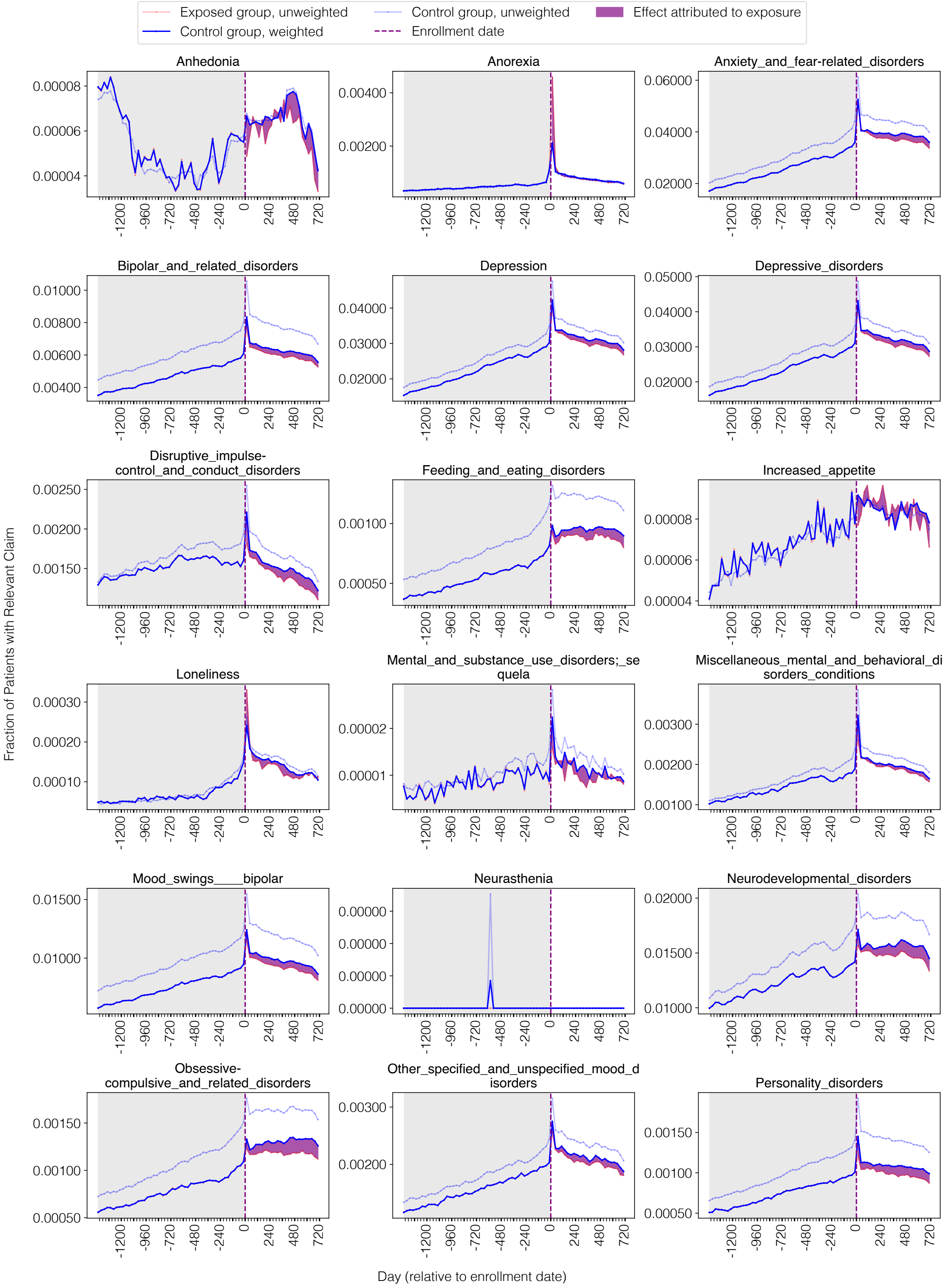


**Supplementary Figure S83:** Temporal trends in Mental health prevalence. The fraction of patients with relevant claims over time belonging for each outcome in the **prospective study** with testing-based enrollment experiment.

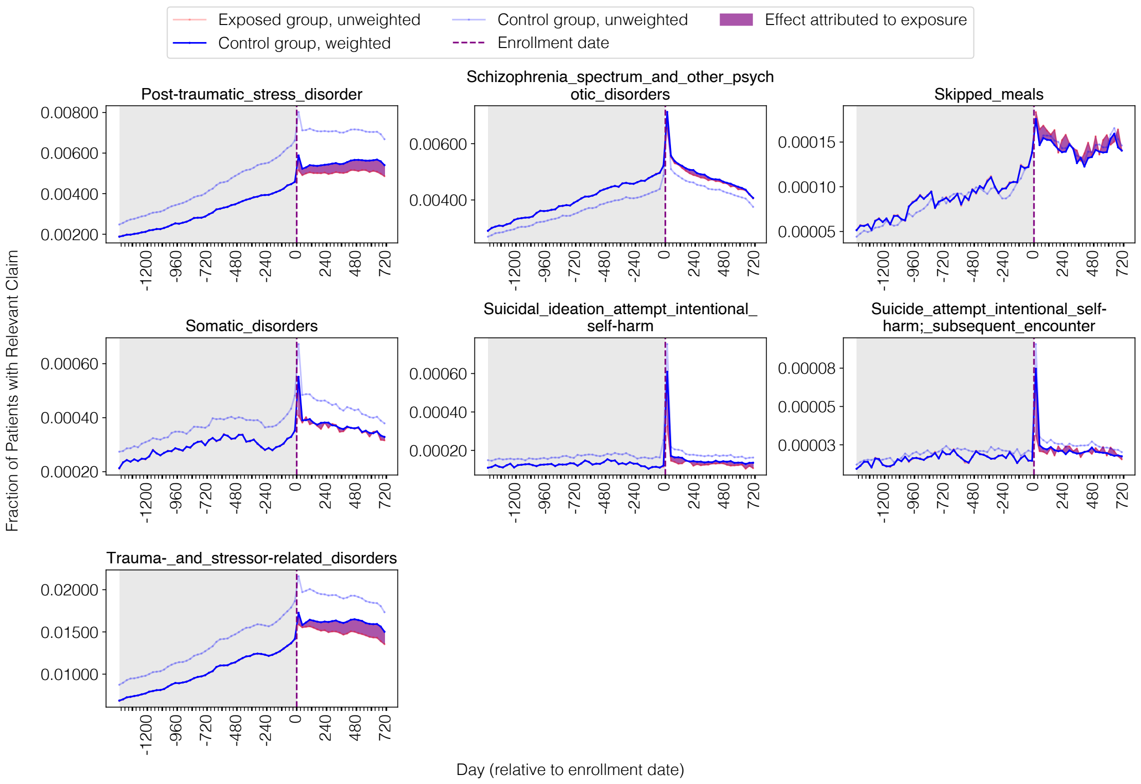


**Supplementary Figure S83:** (cont'd) Temporal trends in Mental health prevalence. The fraction of patients with relevant claims over time belonging for each outcome in the **prospective study** with testing-based enrollment experiment.

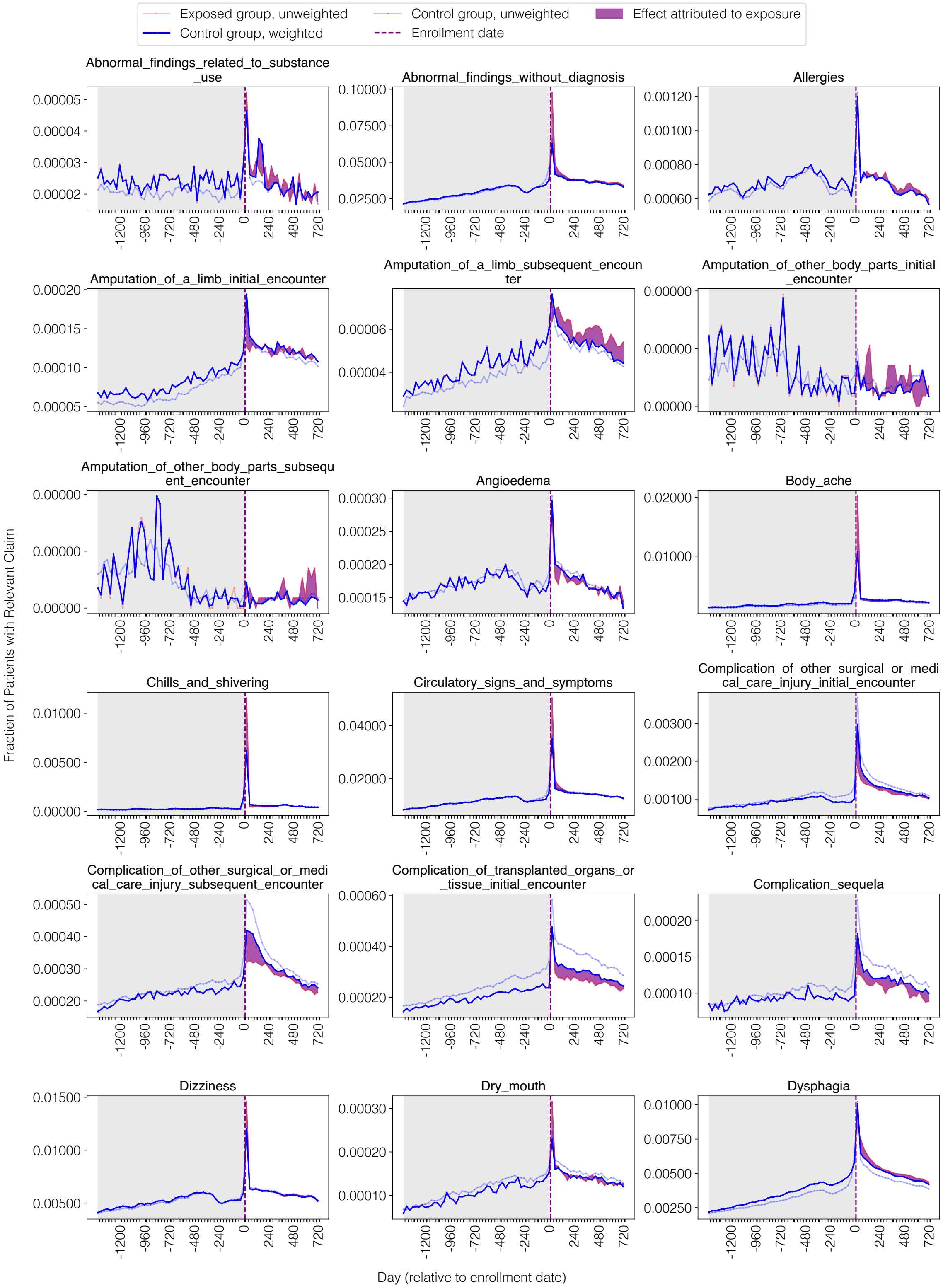


**Supplementary Figure S84:** Temporal trends in Misc prevalence. The fraction of patients with relevant claims over time belonging for each outcome in the **prospective study** with testing-based enrollment experiment.

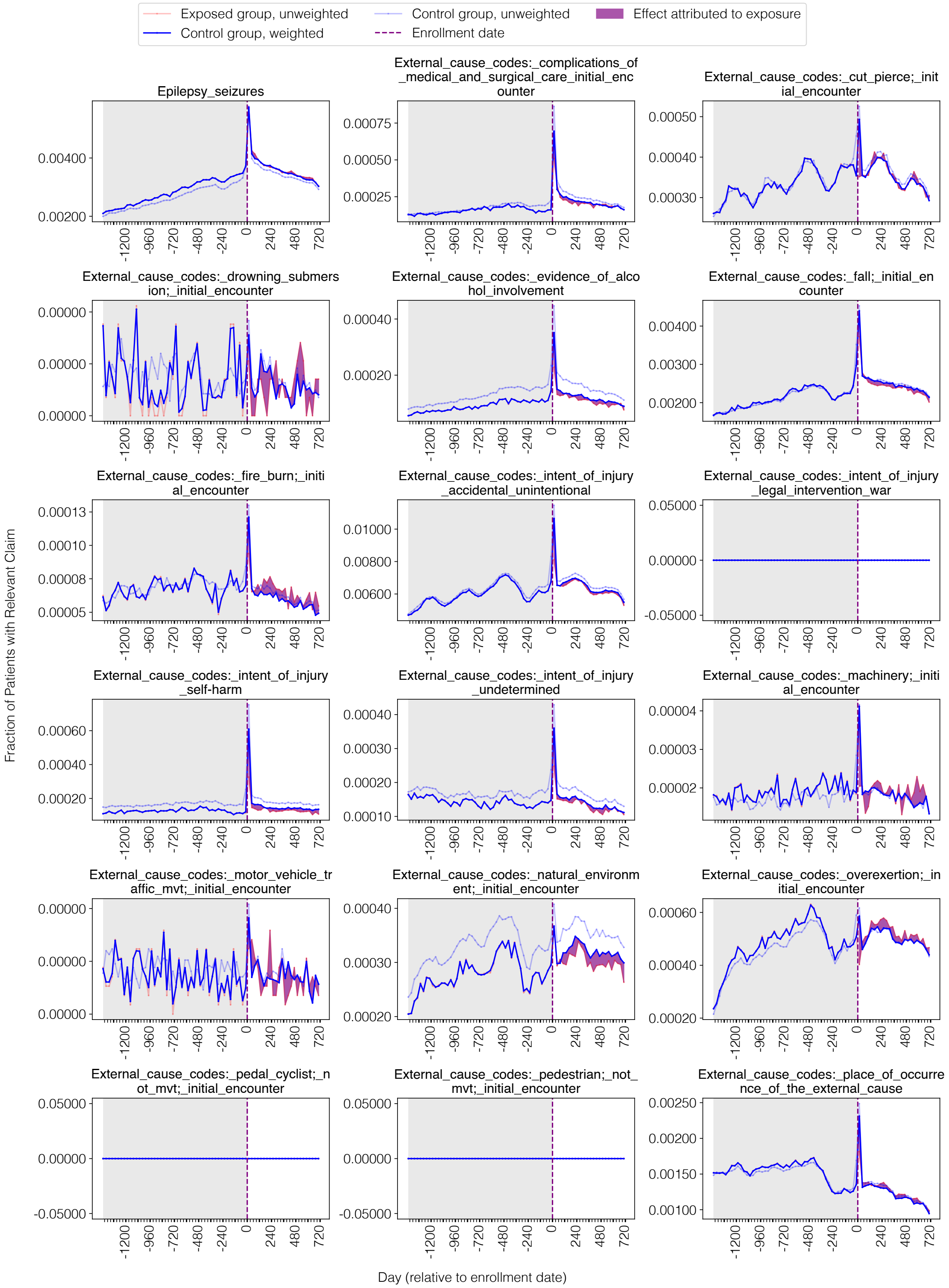


**Supplementary Figure S84:** (cont'd) Temporal trends in Misc prevalence. The fraction of patients with relevant claims over time belonging for each outcome in the **prospective study** with testing-based enrollment experiment.

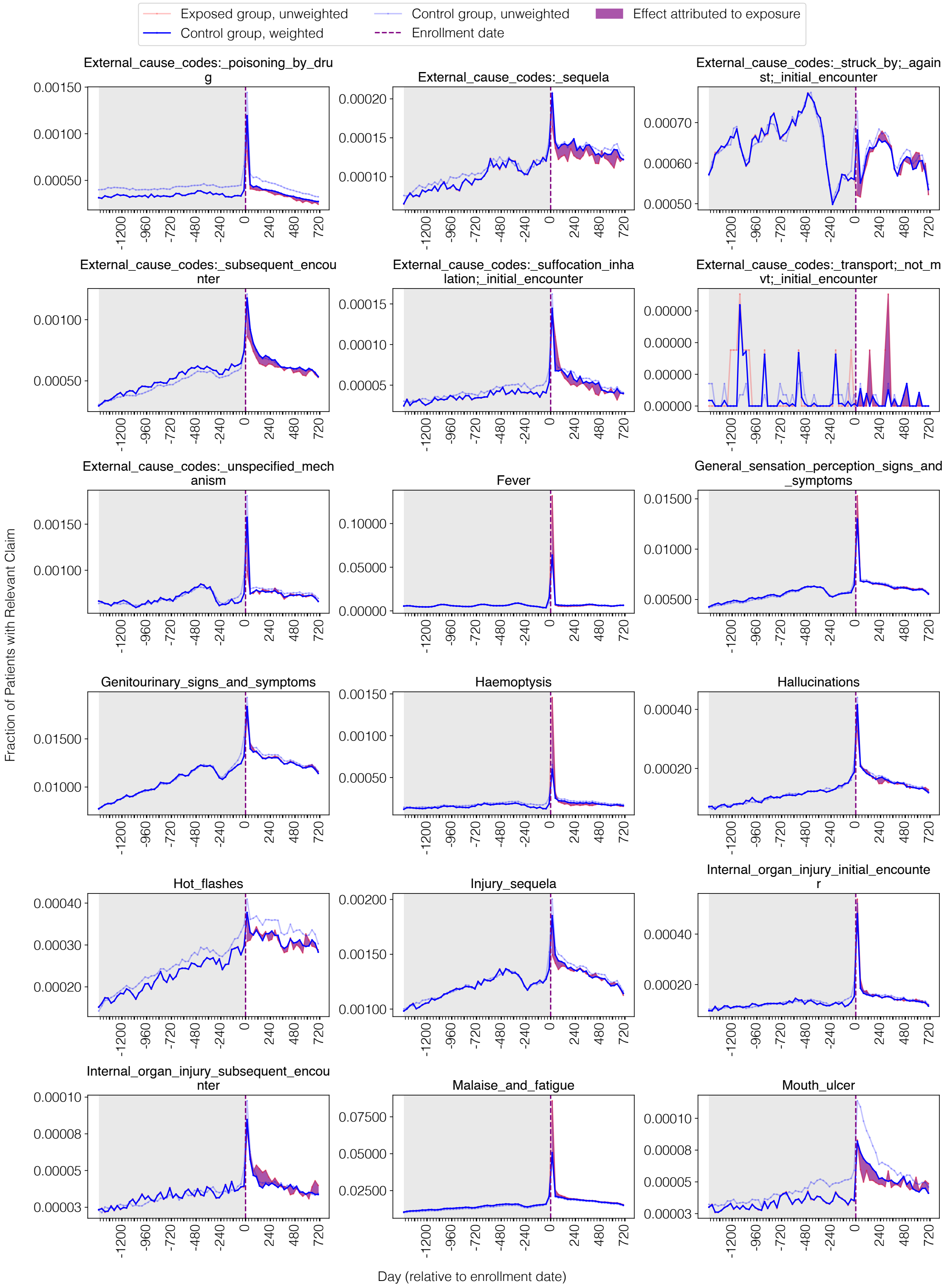


**Supplementary Figure S84:** (cont'd) Temporal trends in Misc prevalence. The fraction of patients with relevant claims over time belonging for each outcome in the **prospective study** with testing-based enrollment experiment.

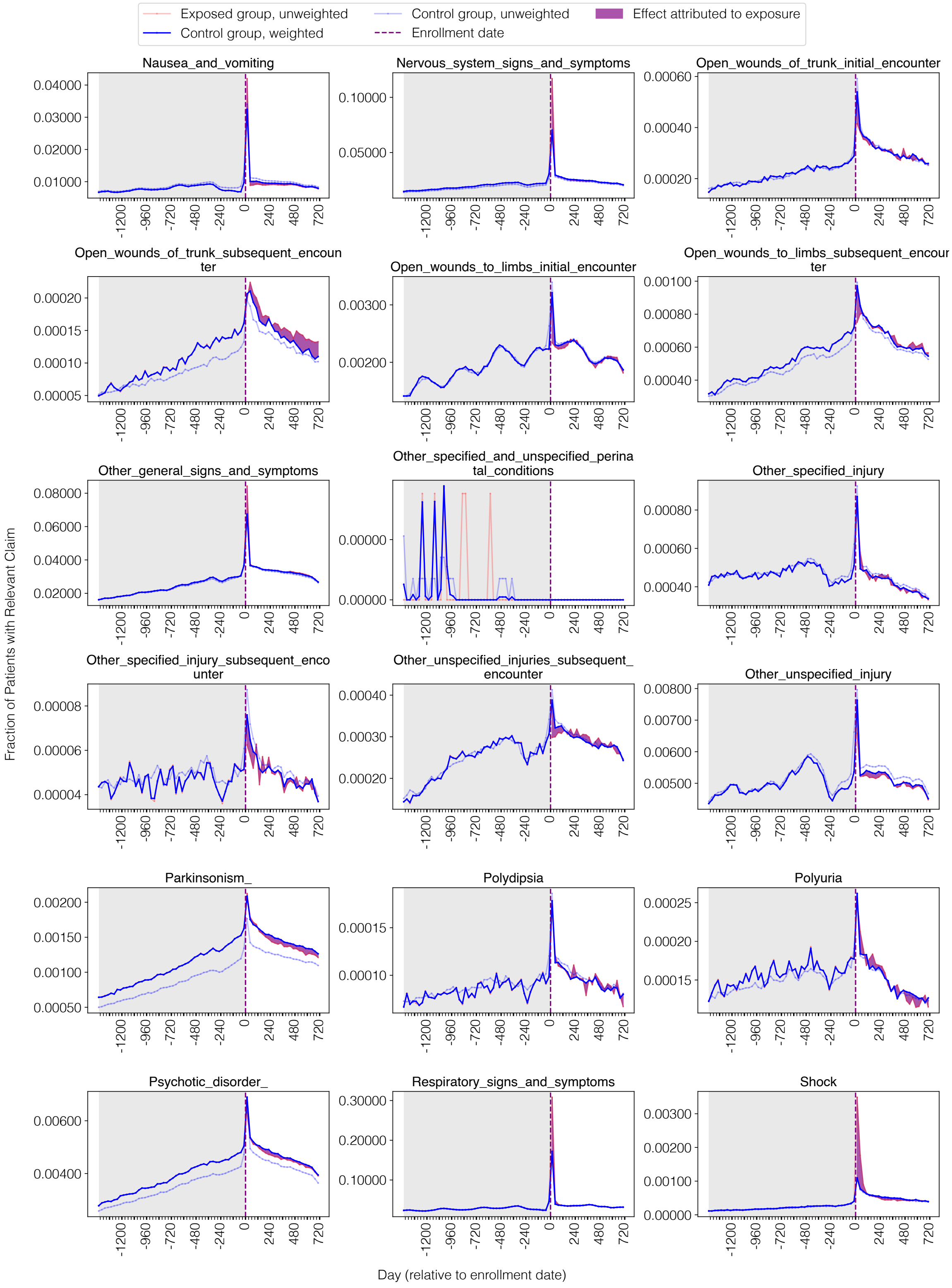


**Supplementary Figure S84:** (cont'd) Temporal trends in Misc prevalence. The fraction of patients with relevant claims over time belonging for each outcome in the **prospective study** with testing-based enrollment experiment.

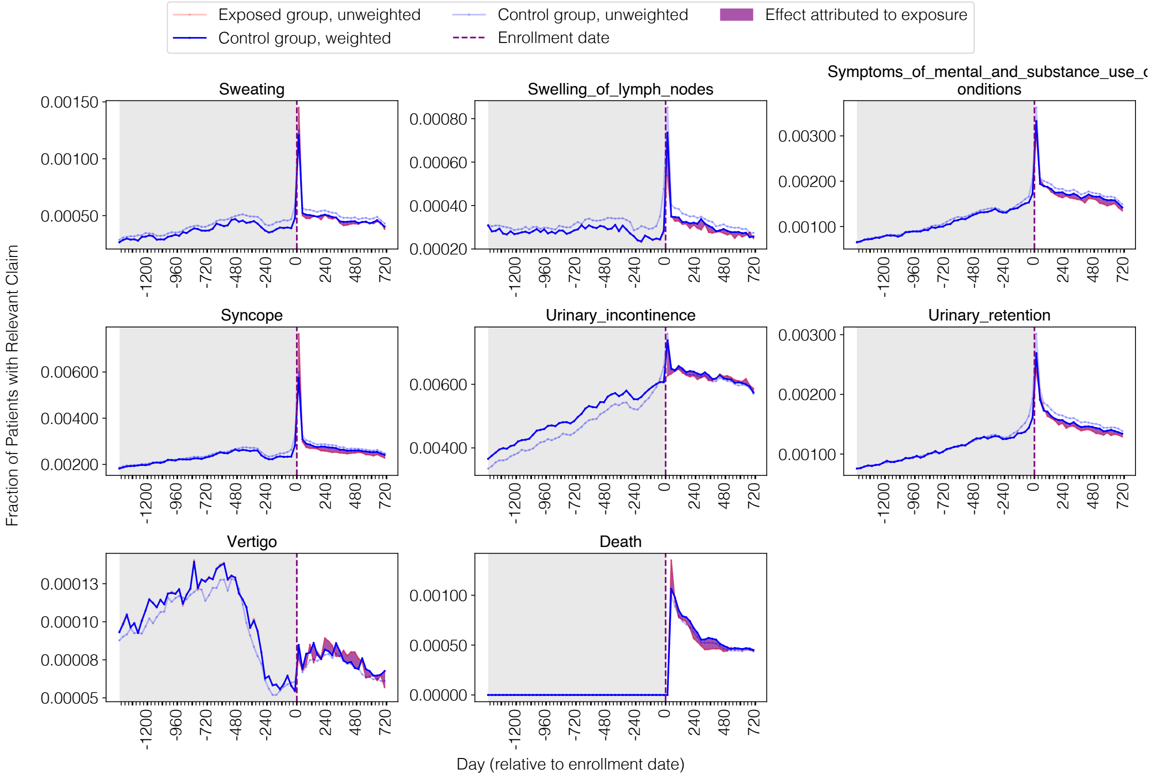


**Supplementary Figure S84:** (cont'd) Temporal trends in Misc prevalence. The fraction of patients with relevant claims over time belonging for each outcome in the **prospective study** with testing-based enrollment experiment.

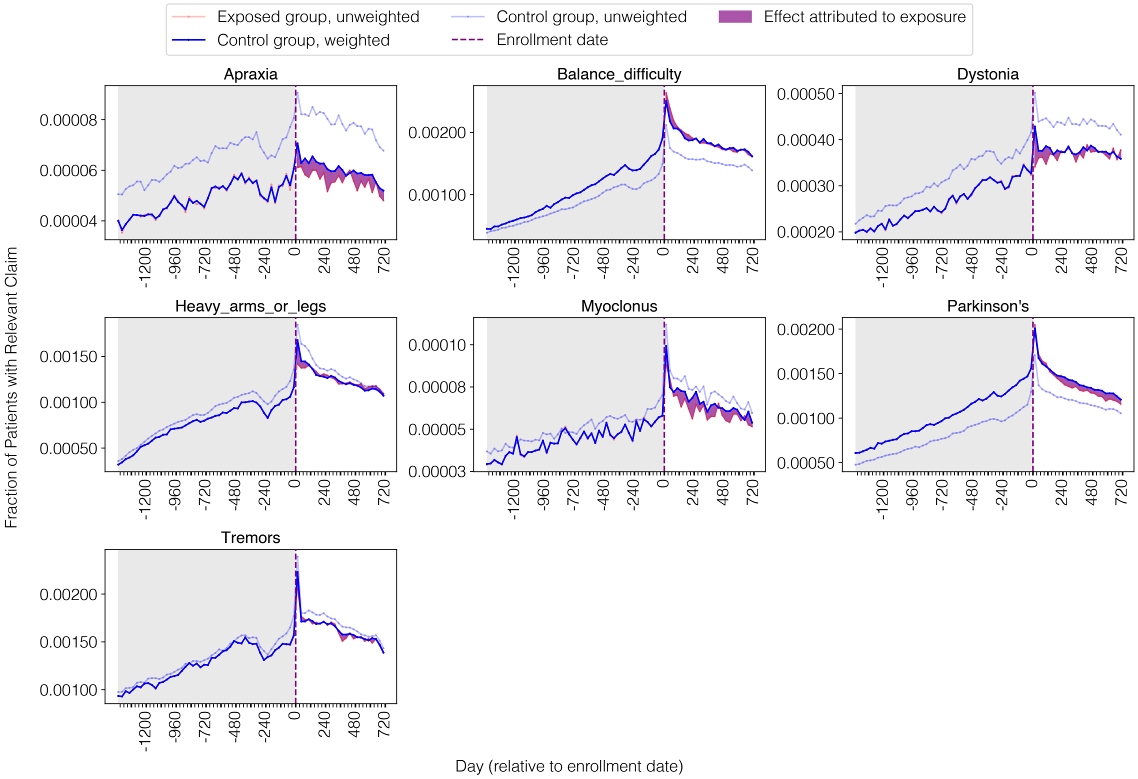


**Supplementary Figure S85:** Temporal trends in Movement prevalence. The fraction of patients with relevant claims over time belonging for each outcome in the **prospective study** with testing-based enrollment experiment.

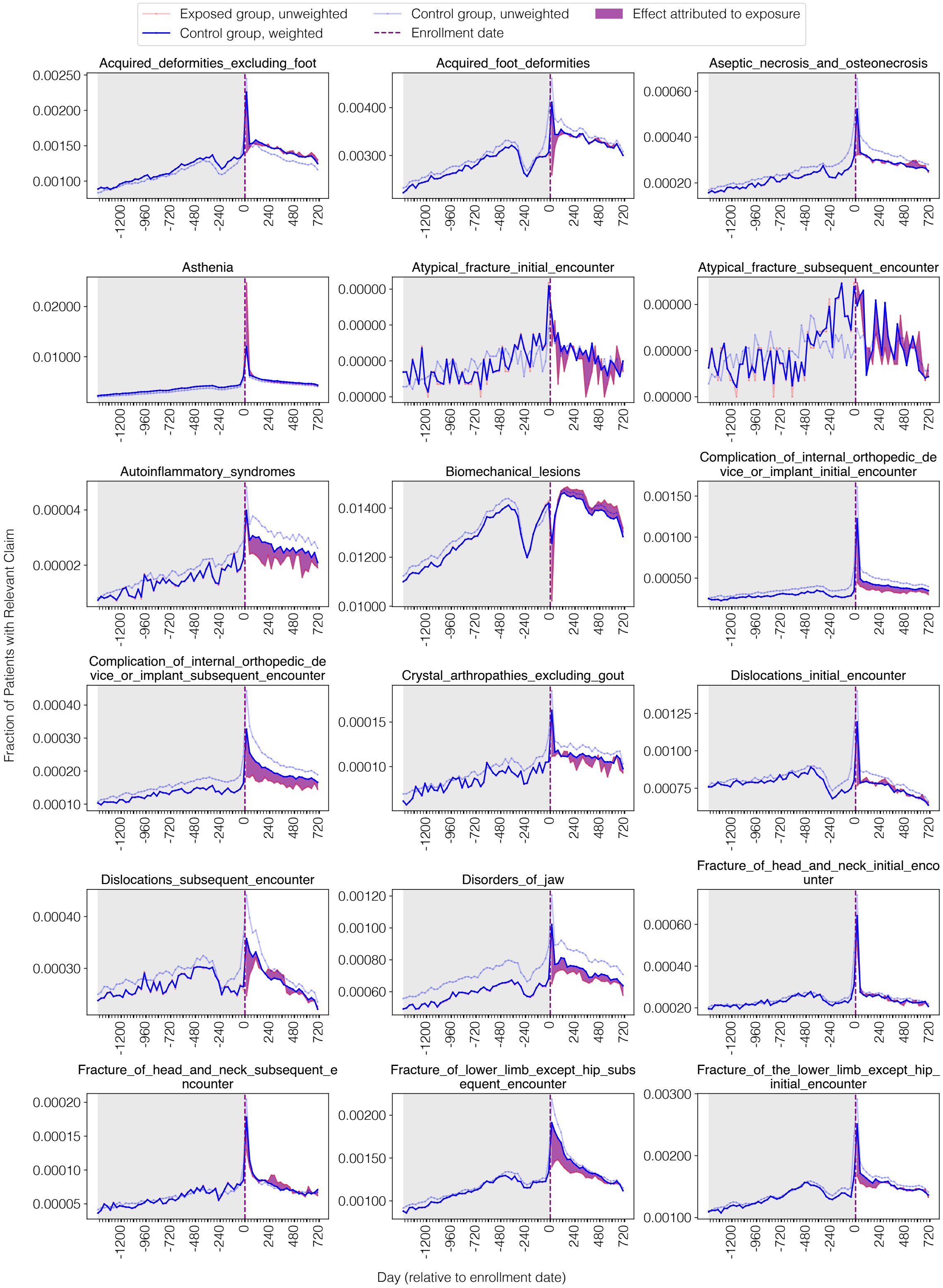


**Supplementary Figure S86:** Temporal trends in Muscles and joints prevalence. The fraction of patients with relevant claims over time belonging for each outcome in the **prospective study** with testing-based enrollment experiment.

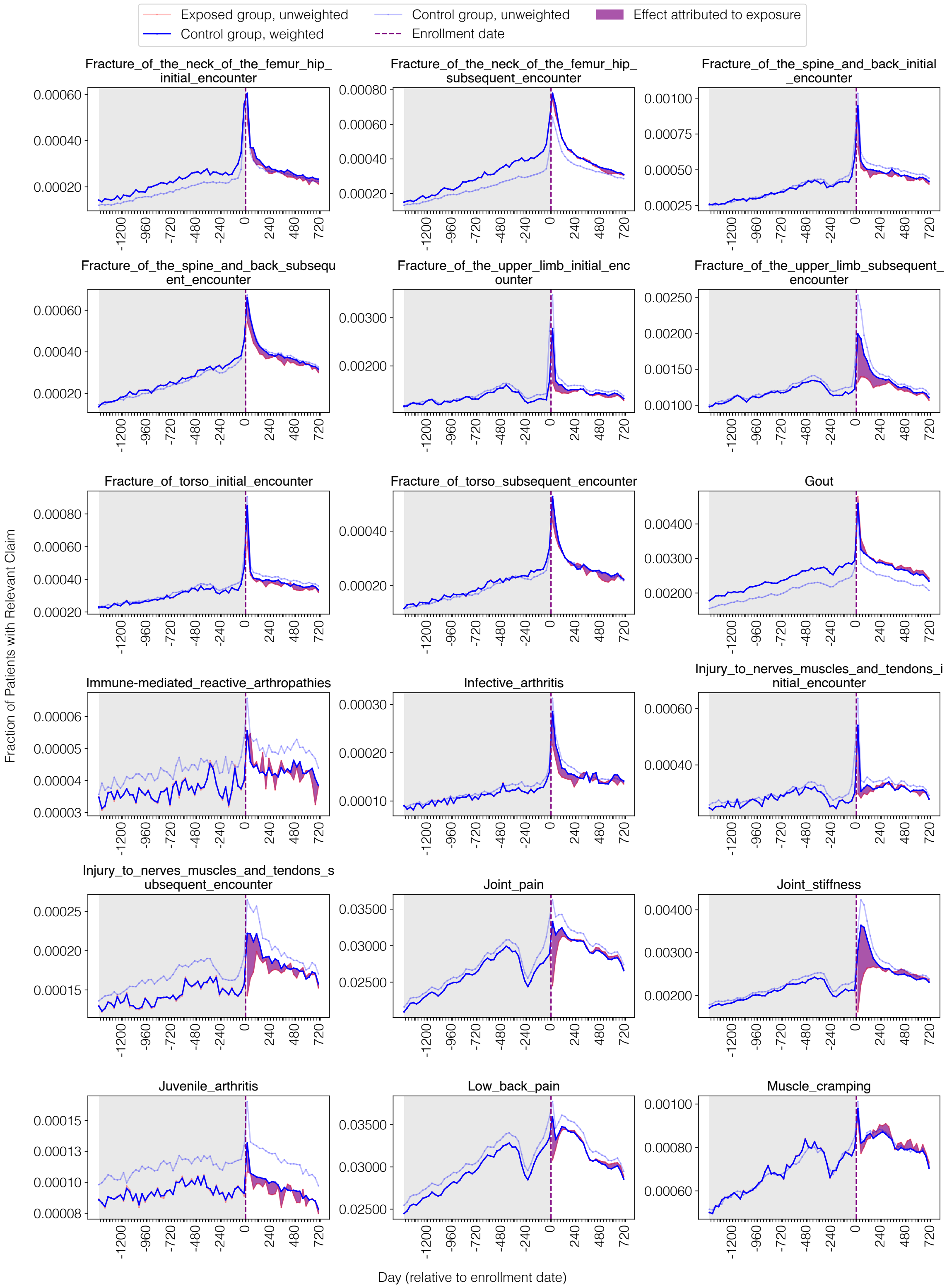


**Supplementary Figure S86:** (cont'd) Temporal trends in Muscles and joints prevalence. The fraction of patients with relevant claims over time belonging for each outcome in the **prospective study** with testing-based enrollment experiment.

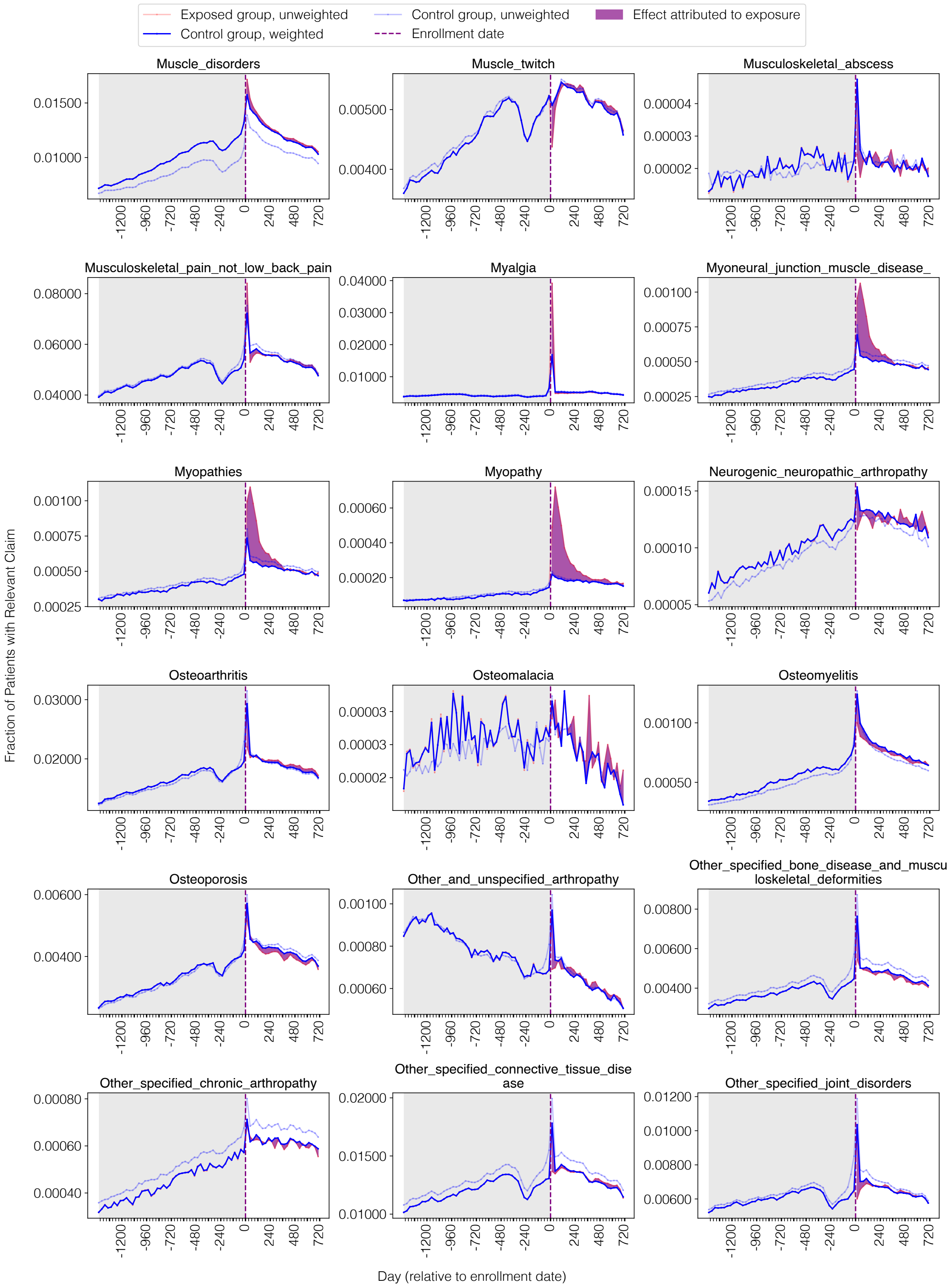


**Supplementary Figure S86:** (cont'd) Temporal trends in Muscles and joints prevalence. The fraction of patients with relevant claims over time belonging for each outcome in the **prospective study** with testing-based enrollment experiment.

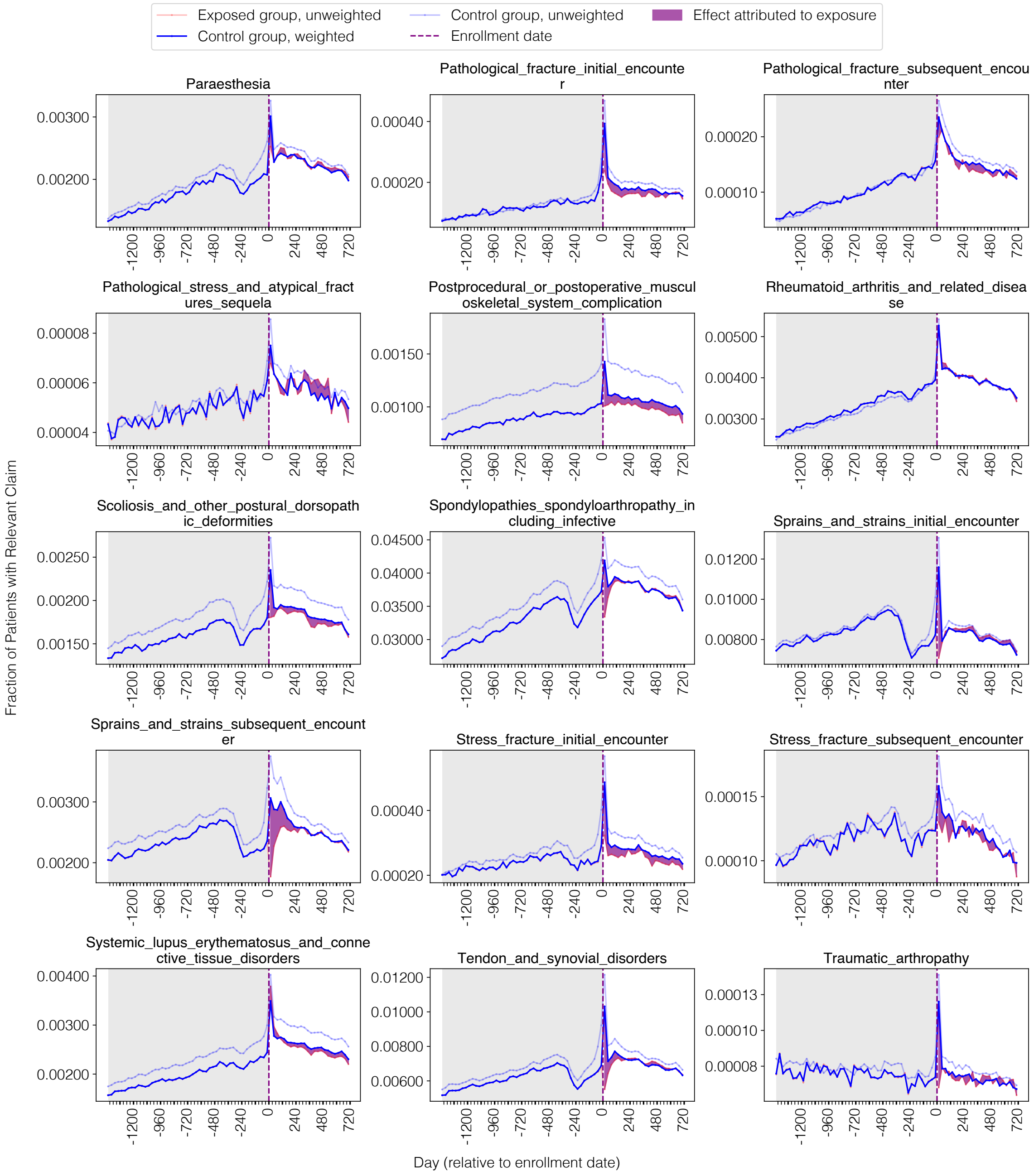


**Supplementary Figure S86:** (cont'd) Temporal trends in Muscles and joints prevalence. The fraction of patients with relevant claims over time belonging for each outcome in the **prospective study** with testing-based enrollment experiment.

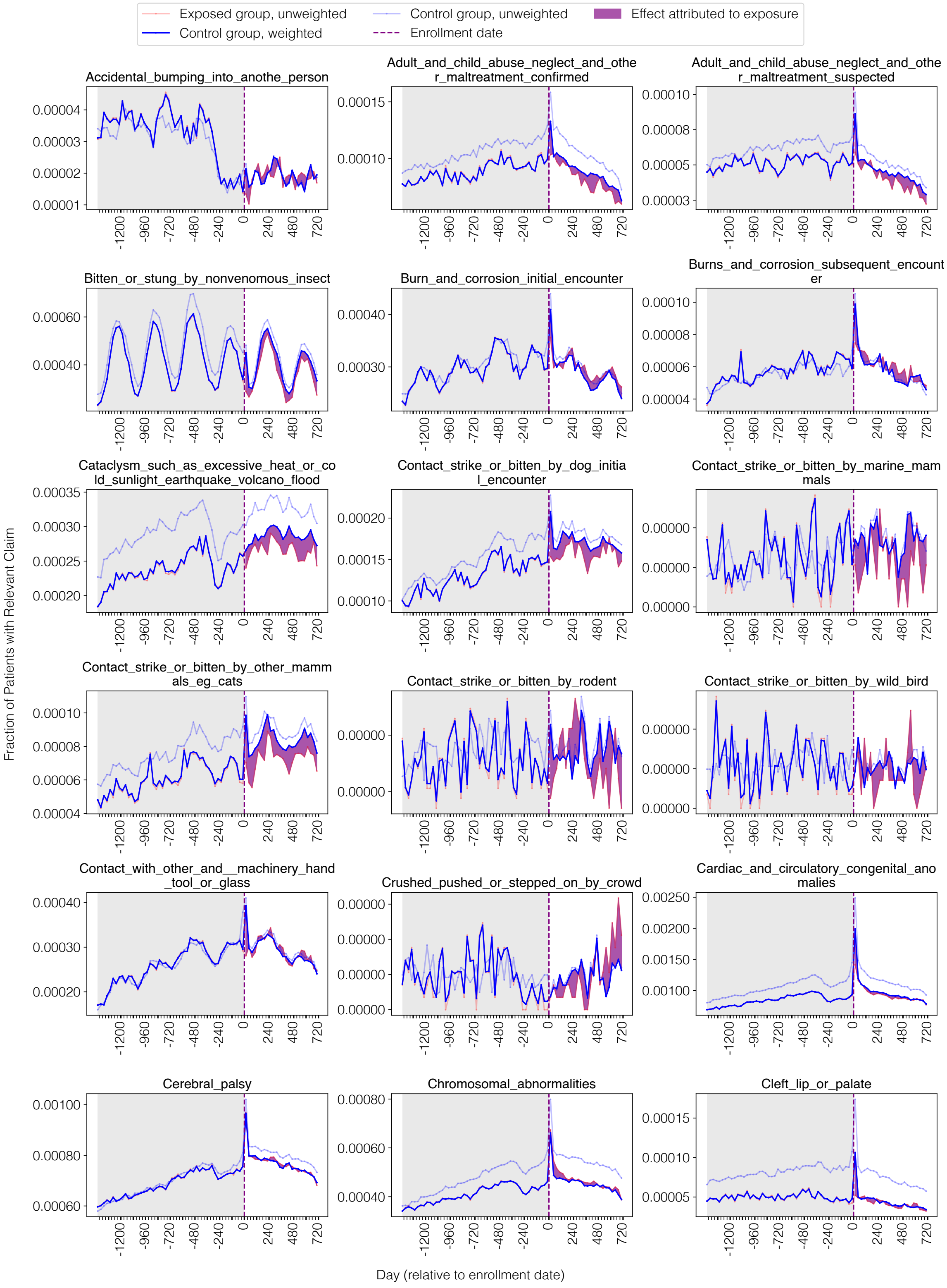


**Supplementary Figure S87:** Temporal trends in Negative prevalence. The fraction of patients with relevant claims over time belonging for each outcome in the **prospective study** with testing-based enrollment experiment.

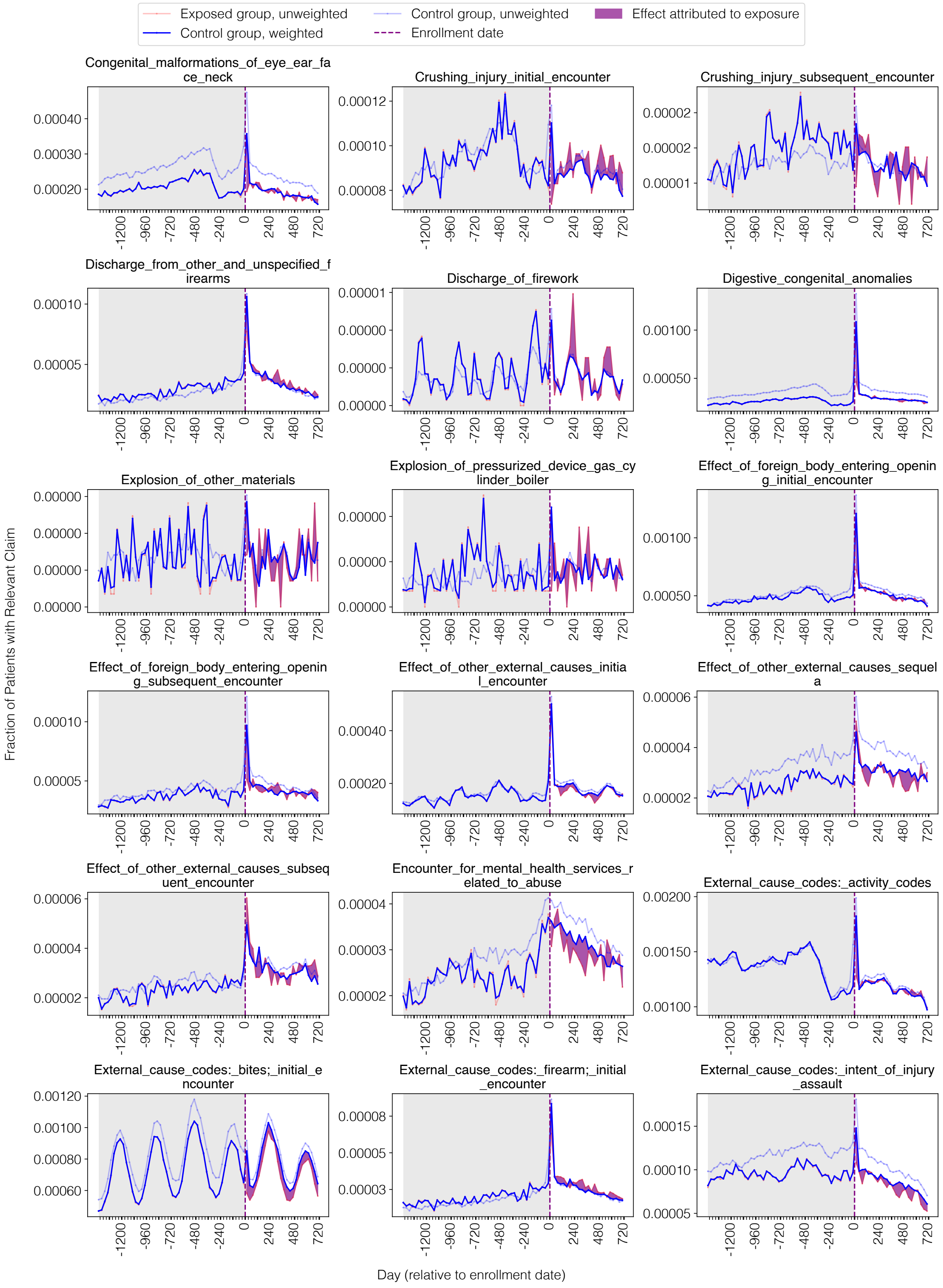


**Supplementary Figure S87:** (cont'd) Temporal trends in Negative prevalence. The fraction of patients with relevant claims over time belonging for each outcome in the **prospective study** with testing-based enrollment experiment.

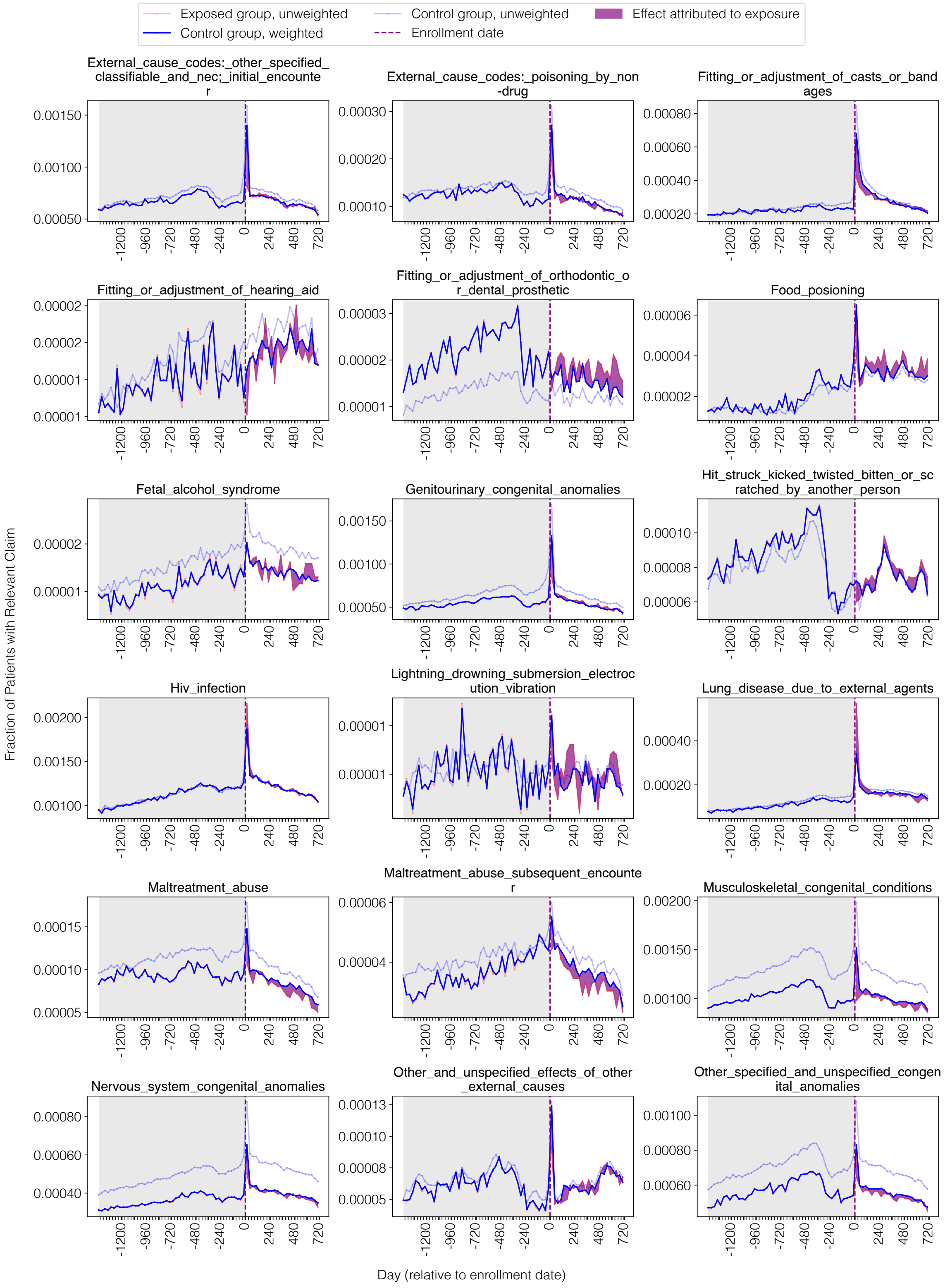


**Supplementary Figure S87:** (cont'd) Temporal trends in Negative prevalence. The fraction of patients with relevant claims over time belonging for each outcome in the **prospective study** with testing-based enrollment experiment.

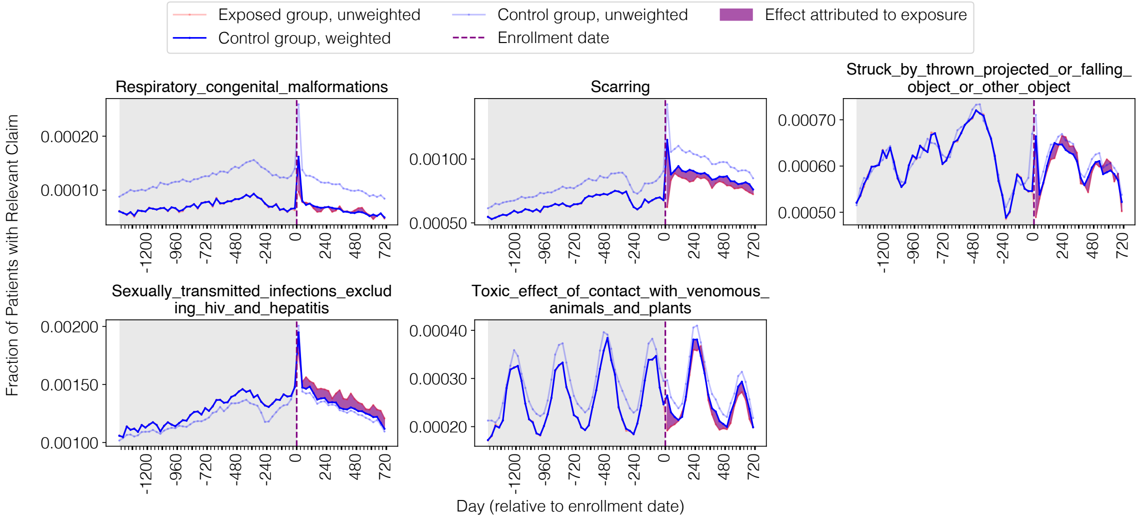


**Supplementary Figure S87:** (cont'd) Temporal trends in Negative prevalence. The fraction of patients with relevant claims over time belonging for each outcome in the **prospective study** with testing-based enrollment experiment.

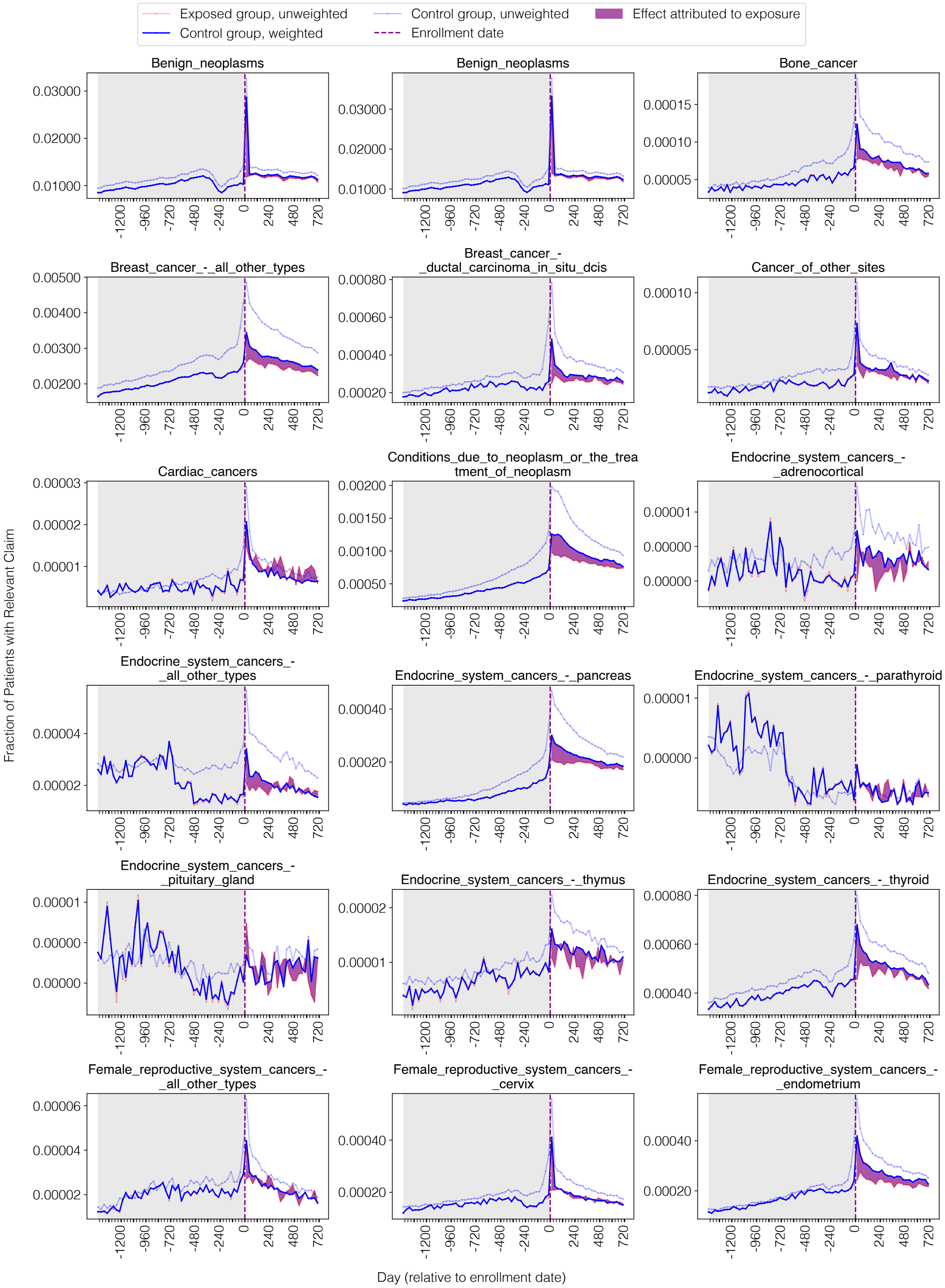


**Supplementary Figure S88:** Temporal trends in Neoplasms prevalence. The fraction of patients with relevant claims over time belonging for each outcome in the **prospective study** with testing-based enrollment experiment.

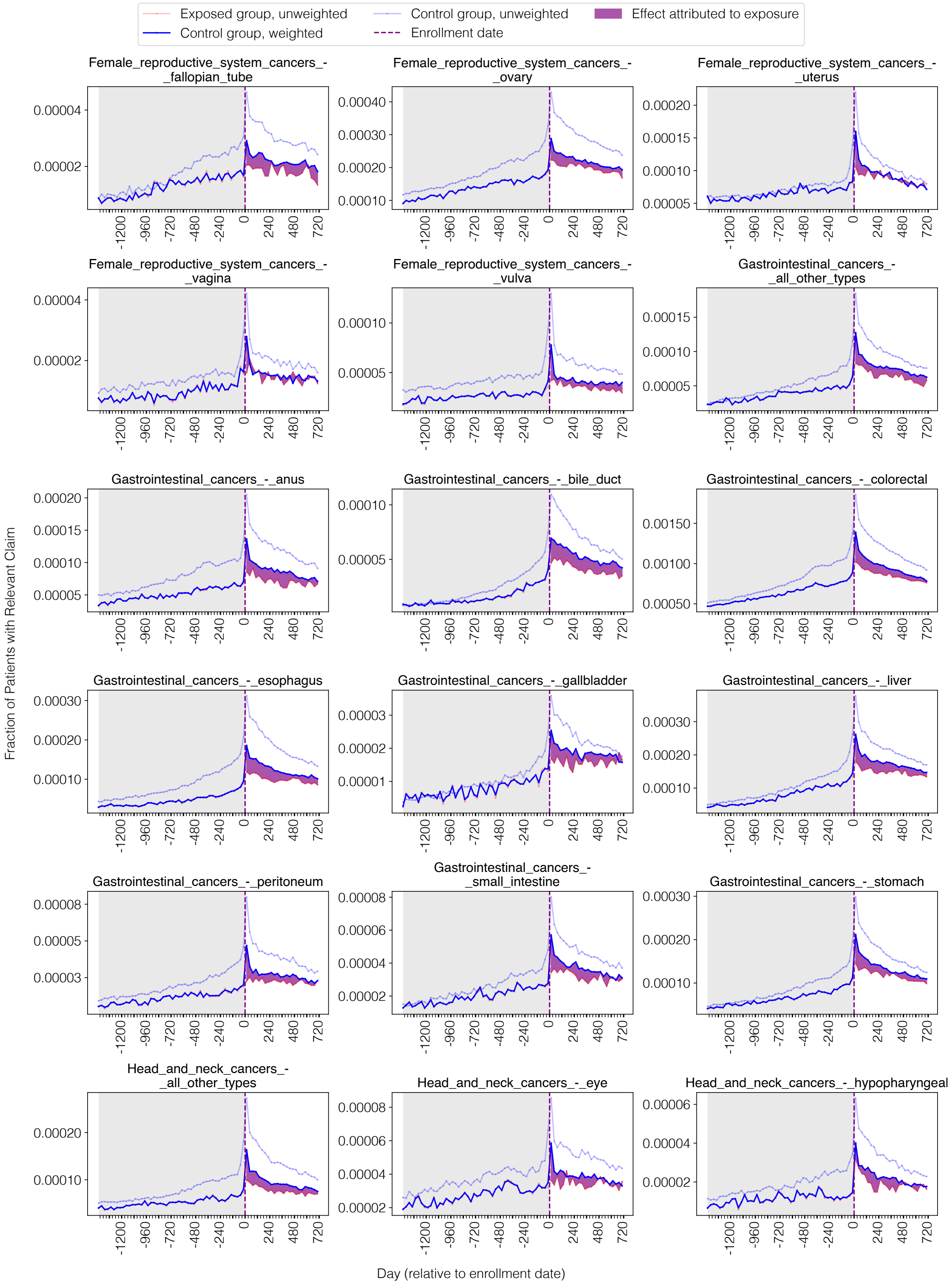


**Supplementary Figure S88:** (cont'd) Temporal trends in Neoplasms prevalence. The fraction of patients with relevant claims over time belonging for each outcome in the **prospective study** with testing-based enrollment experiment.

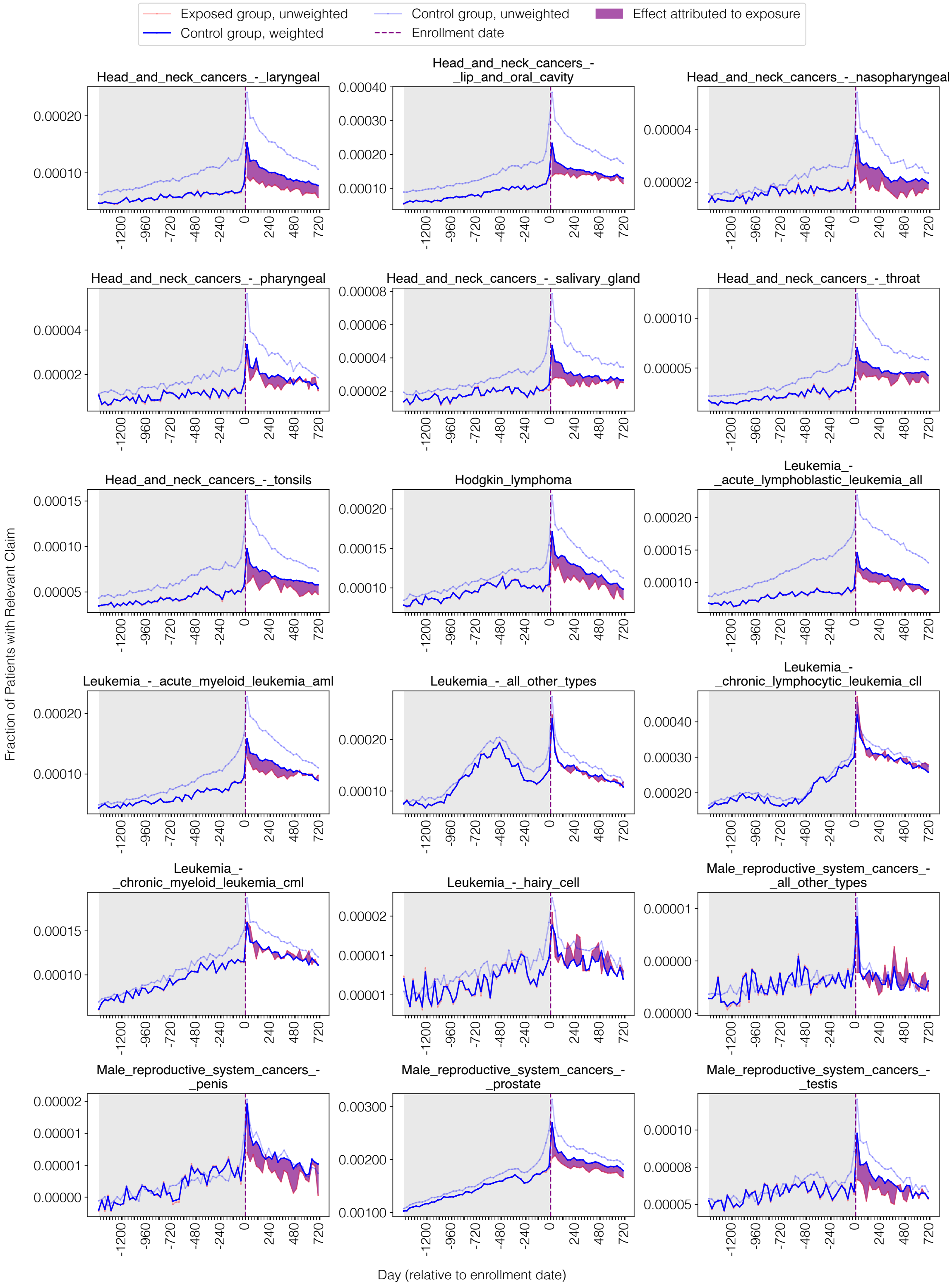


**Supplementary Figure S88:** (cont'd) Temporal trends in Neoplasms prevalence. The fraction of patients with relevant claims over time belonging for each outcome in the **prospective study** with testing-based enrollment experiment.

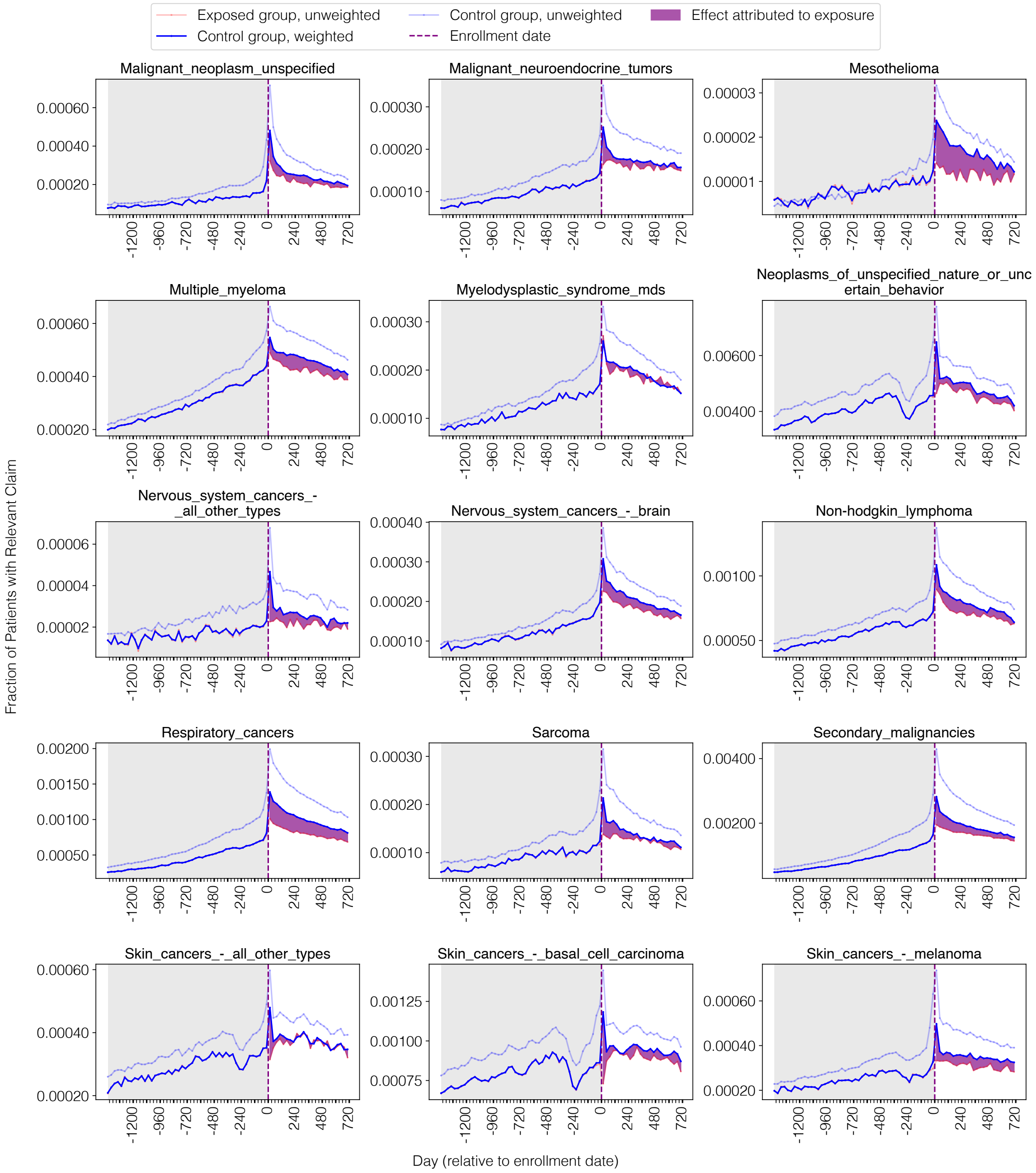


**Supplementary Figure S88:** (cont'd) Temporal trends in Neoplasms prevalence. The fraction of patients with relevant claims over time belonging for each outcome in the **prospective study** with testing-based enrollment experiment.

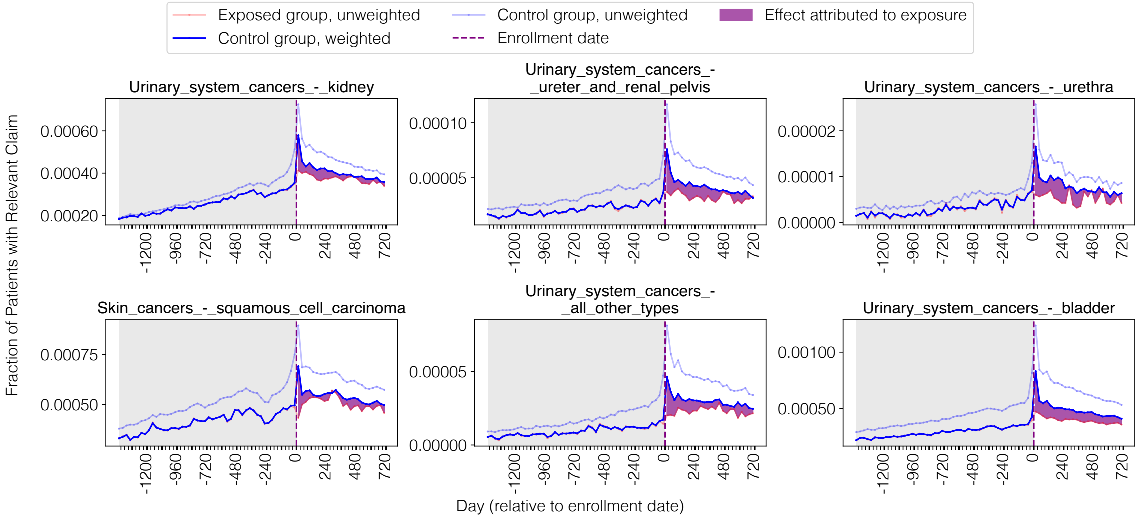


**Supplementary Figure S88:** (cont'd) Temporal trends in Neoplasms prevalence. The fraction of patients with relevant claims over time belonging for each outcome in the **prospective study** with testing-based enrollment experiment.

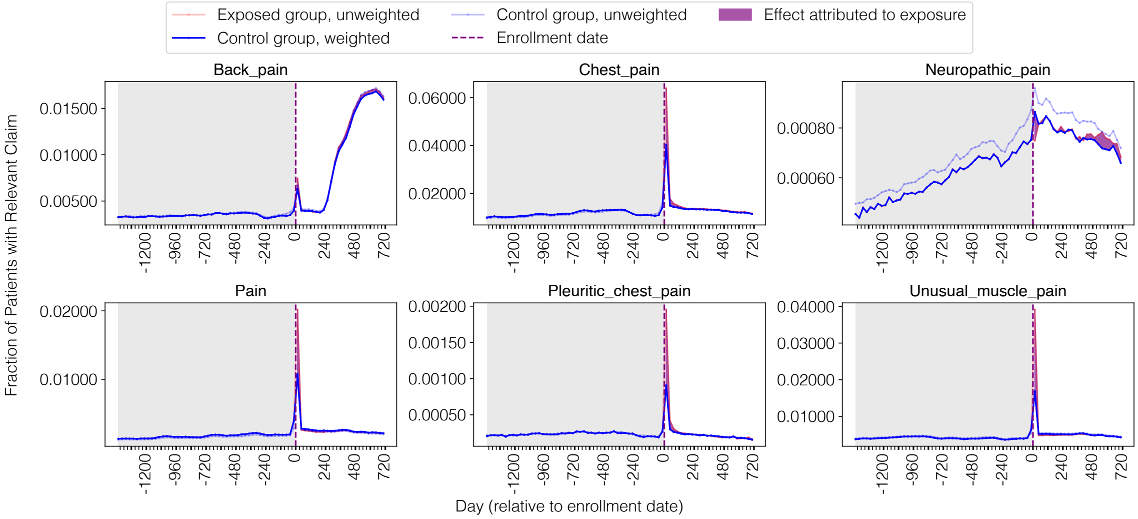


**Supplementary Figure S89:** Temporal trends in Pain prevalence. The fraction of patients with relevant claims over time belonging for each outcome in the **prospective study** with testing-based enrollment experiment.

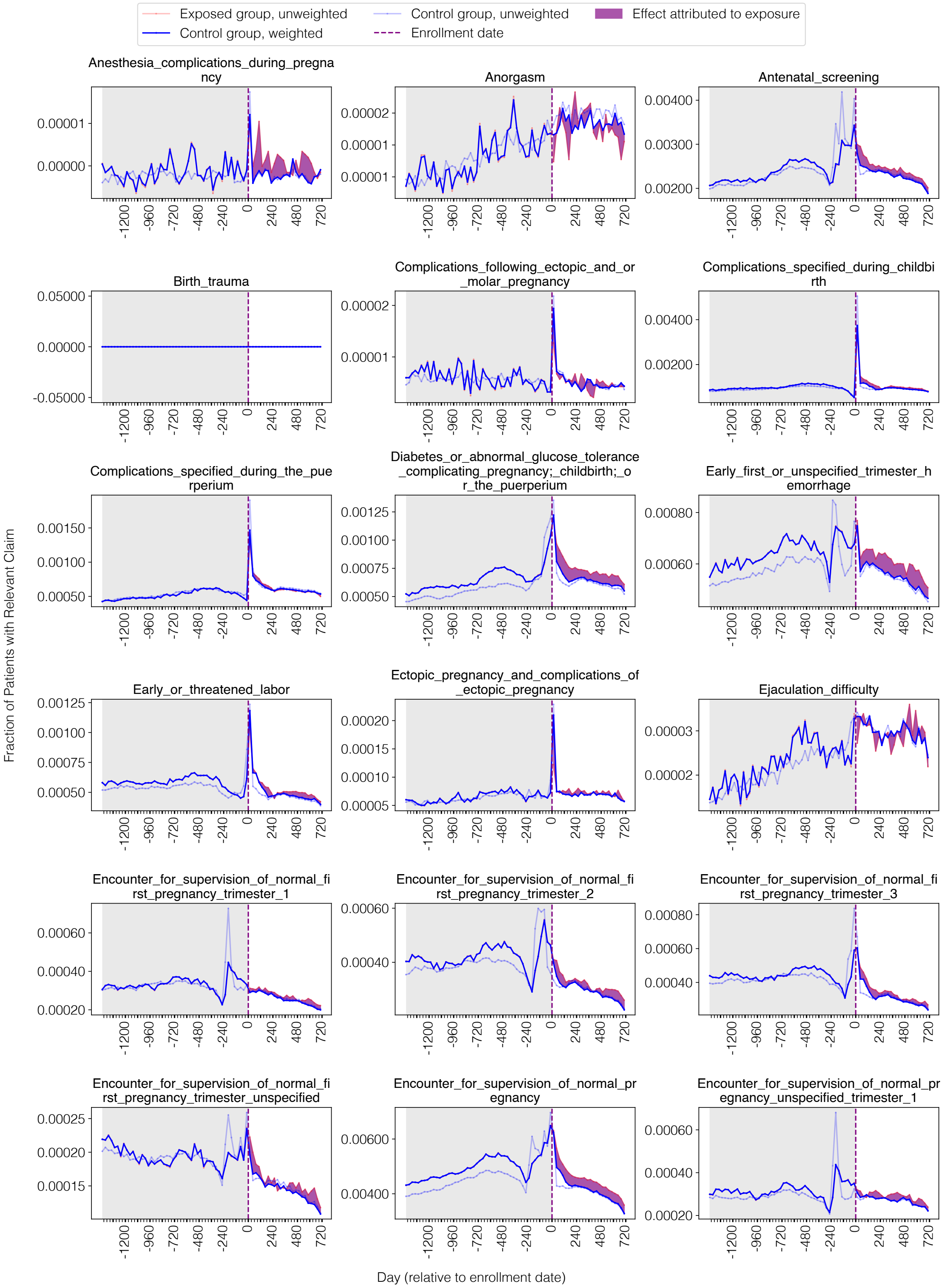


**Supplementary Figure S90:** Temporal trends in Reproductive health prevalence. The fraction of patients with relevant claims over time belonging for each outcome in the **prospective study** with testing-based enrollment experiment.

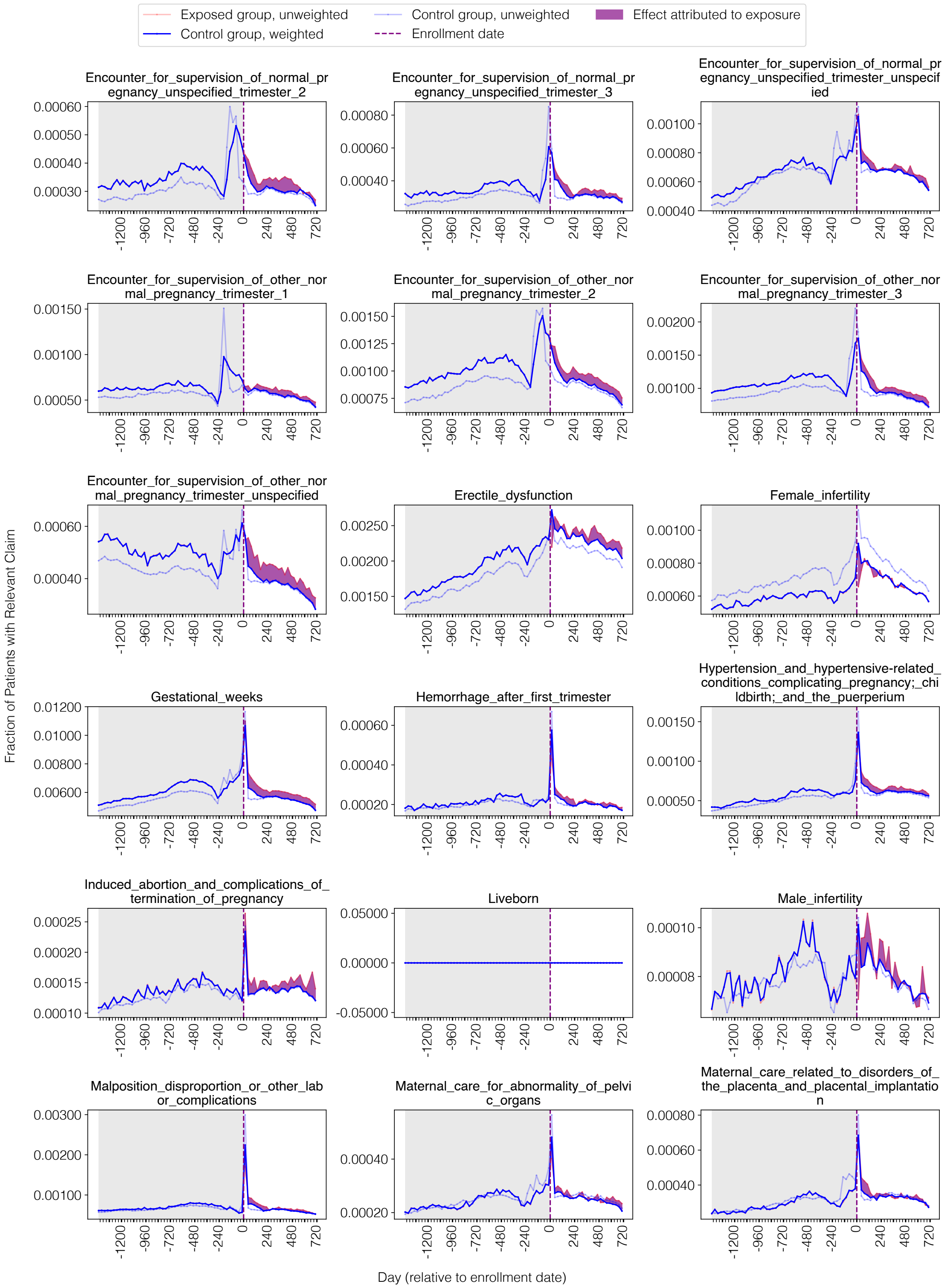


**Supplementary Figure S90:** (cont'd) Temporal trends in Reproductive health prevalence. The fraction of patients with relevant claims over time belonging for each outcome in the **prospective study** with testing-based enrollment experiment.

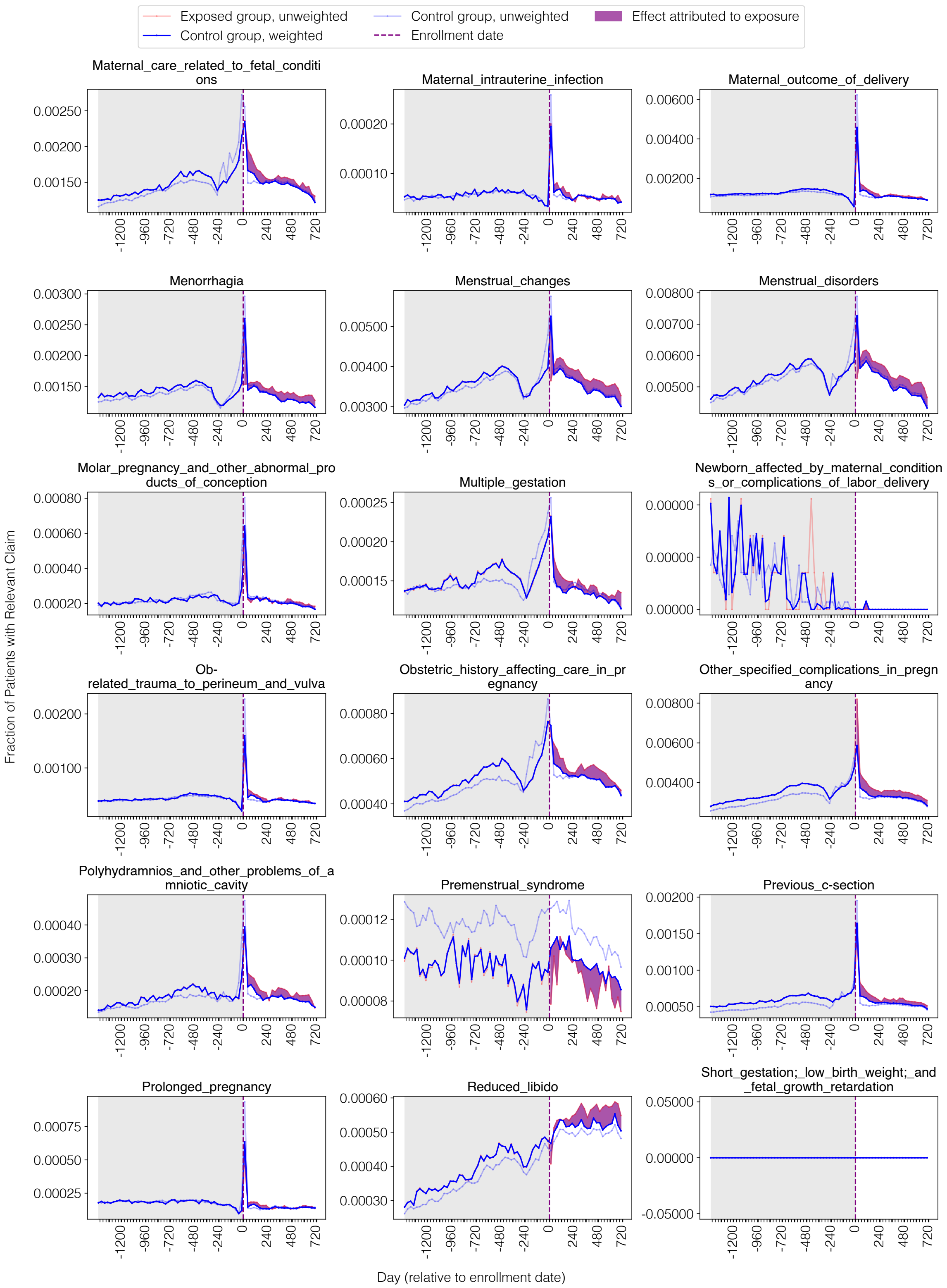


**Supplementary Figure S90:** (cont'd) Temporal trends in Reproductive health prevalence. The fraction of patients with relevant claims over time belonging for each outcome in the **prospective study** with testing-based enrollment experiment.

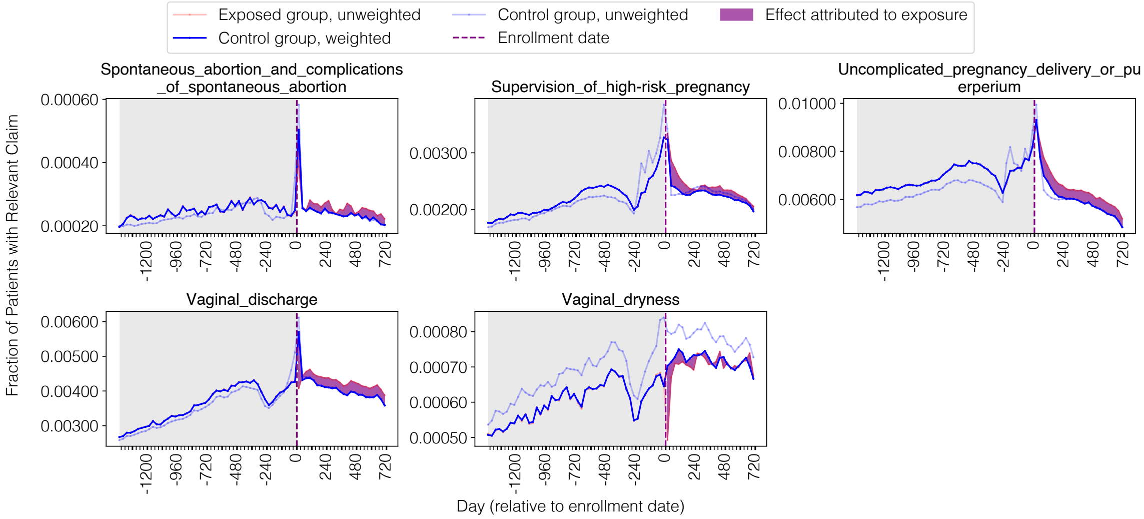


**Supplementary Figure S90:** (cont'd) Temporal trends in Reproductive health prevalence. The fraction of patients with relevant claims over time belonging for each outcome in the **prospective study** with testing-based enrollment experiment.

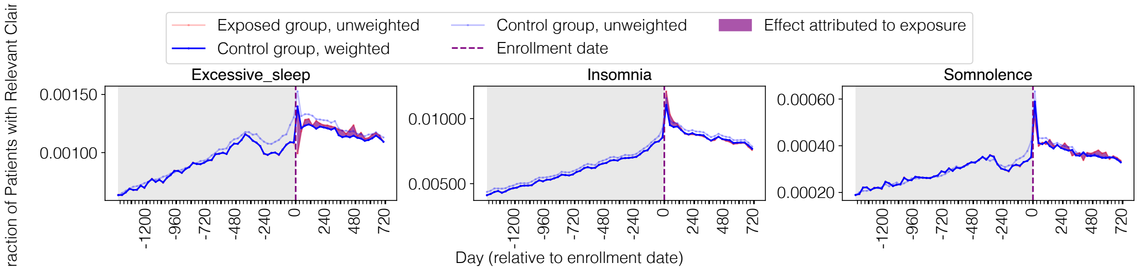


**Supplementary Figure S91:** Temporal trends in Sleep prevalence. The fraction of patients with relevant claims over time belonging for each outcome in the **prospective study** with testing-based enrollment experiment.

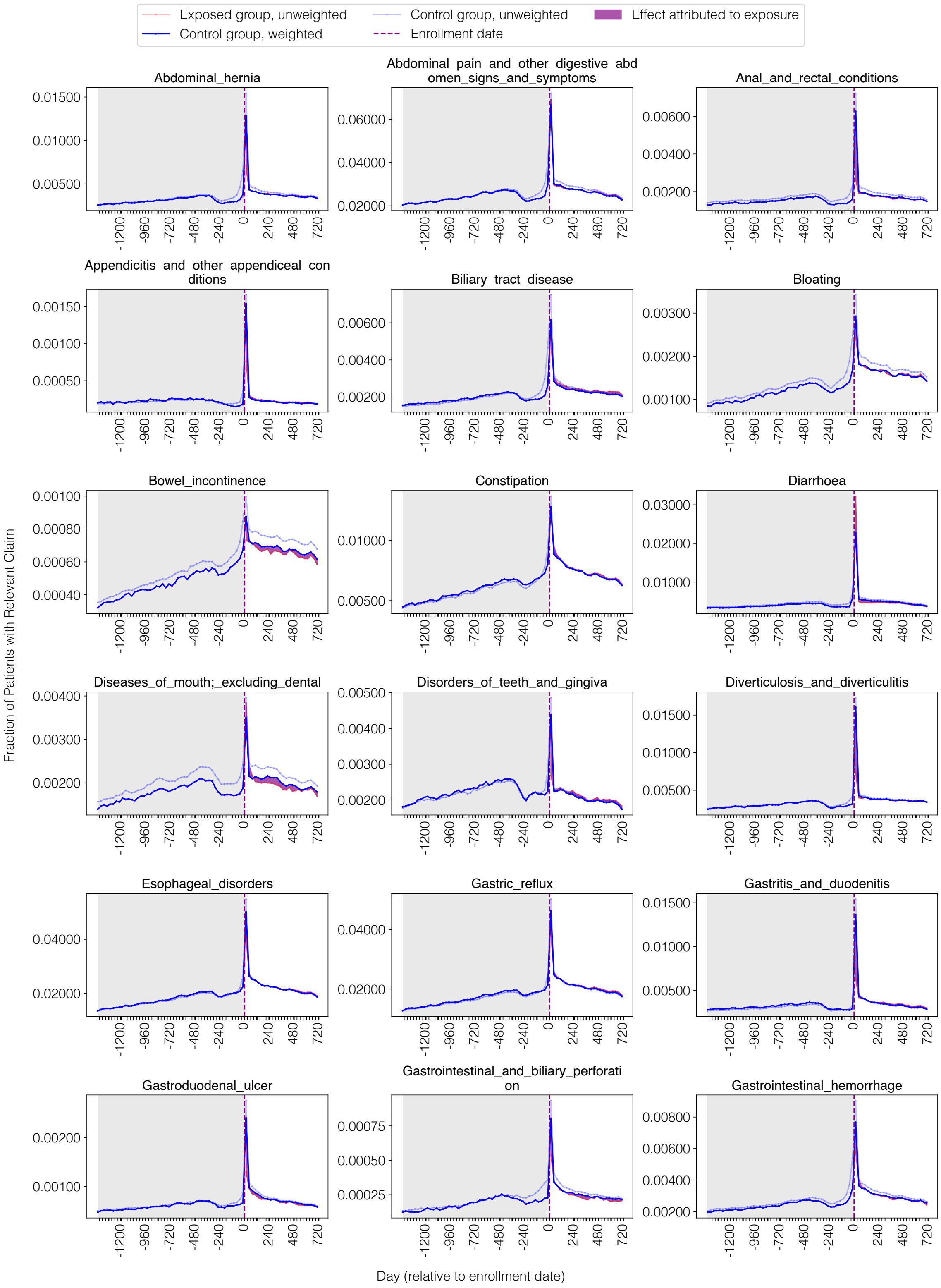


**Supplementary Figure S92:** Temporal trends in Stomach and digestion prevalence. The fraction of patients with relevant claims over time belonging for each outcome in the **prospective study** with testing-based enrollment experiment.

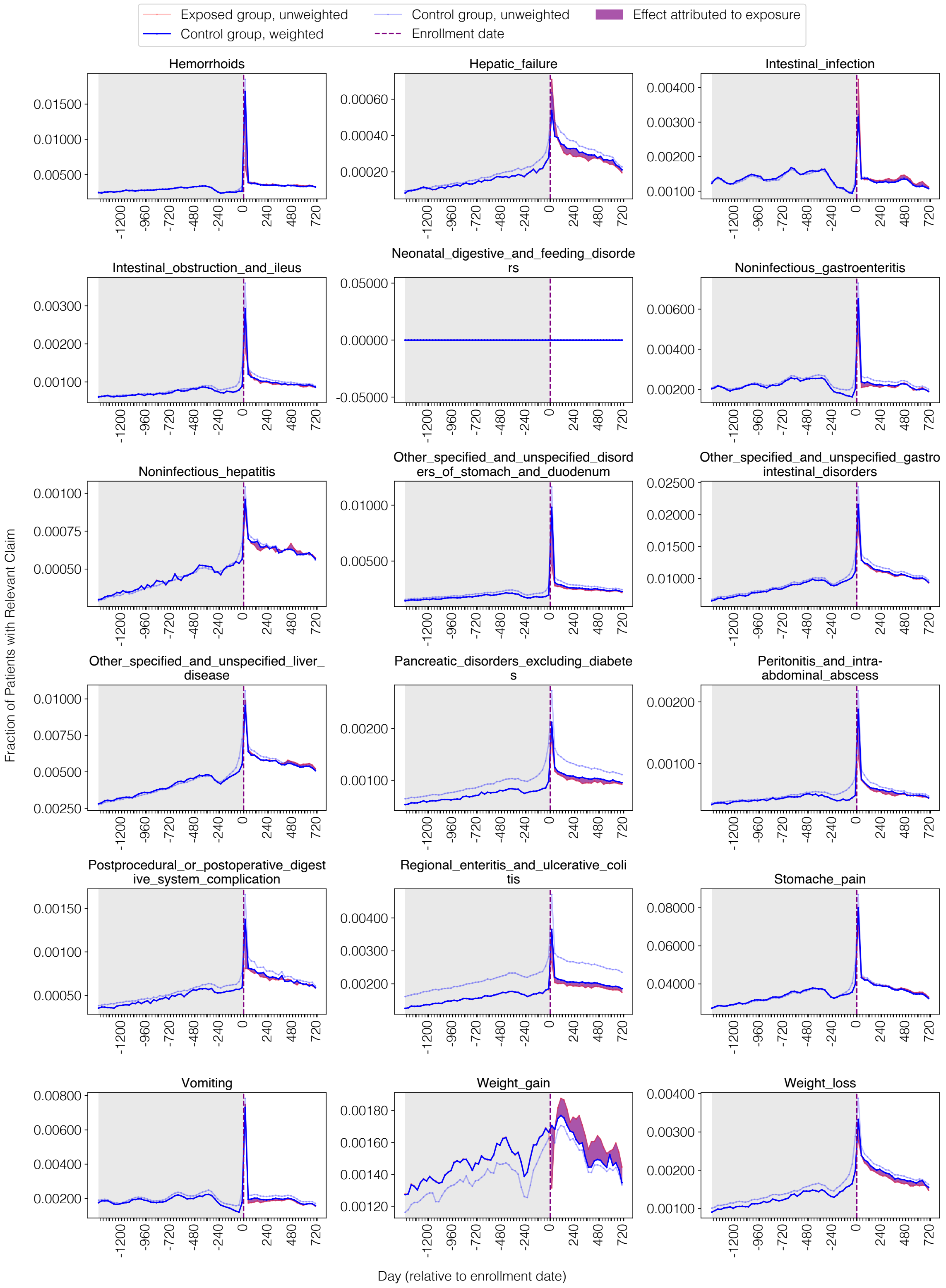


**Supplementary Figure S92:** (cont'd) Temporal trends in Stomach and digestion prevalence. The fraction of patients with relevant claims over time belonging for each outcome in the **prospective study** with testing-based enrollment experiment.

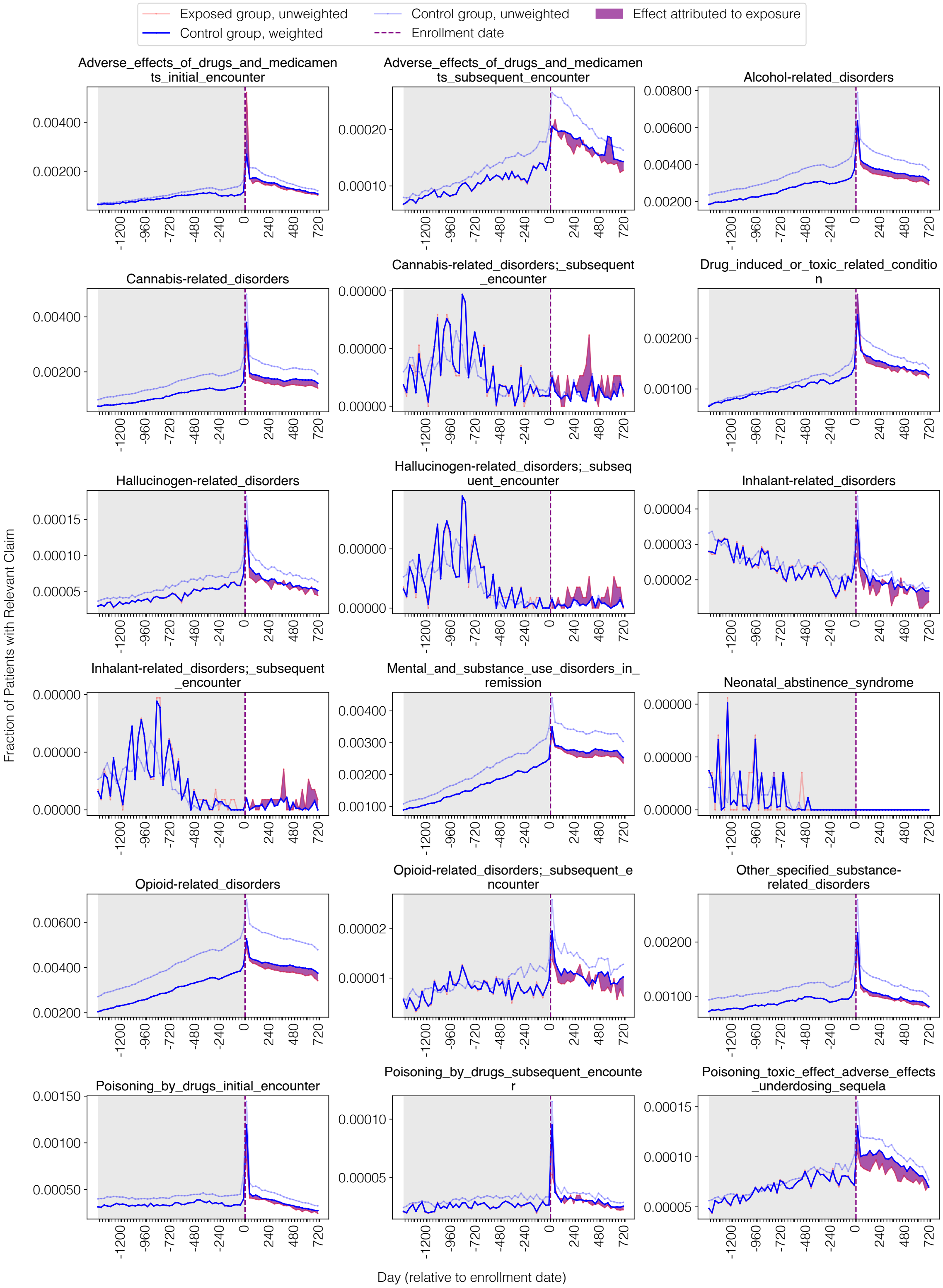


**Supplementary Figure S93:** Temporal trends in Substance use prevalence. The fraction of patients with relevant claims over time belonging for each outcome in the **prospective study** with testing-based enrollment experiment.

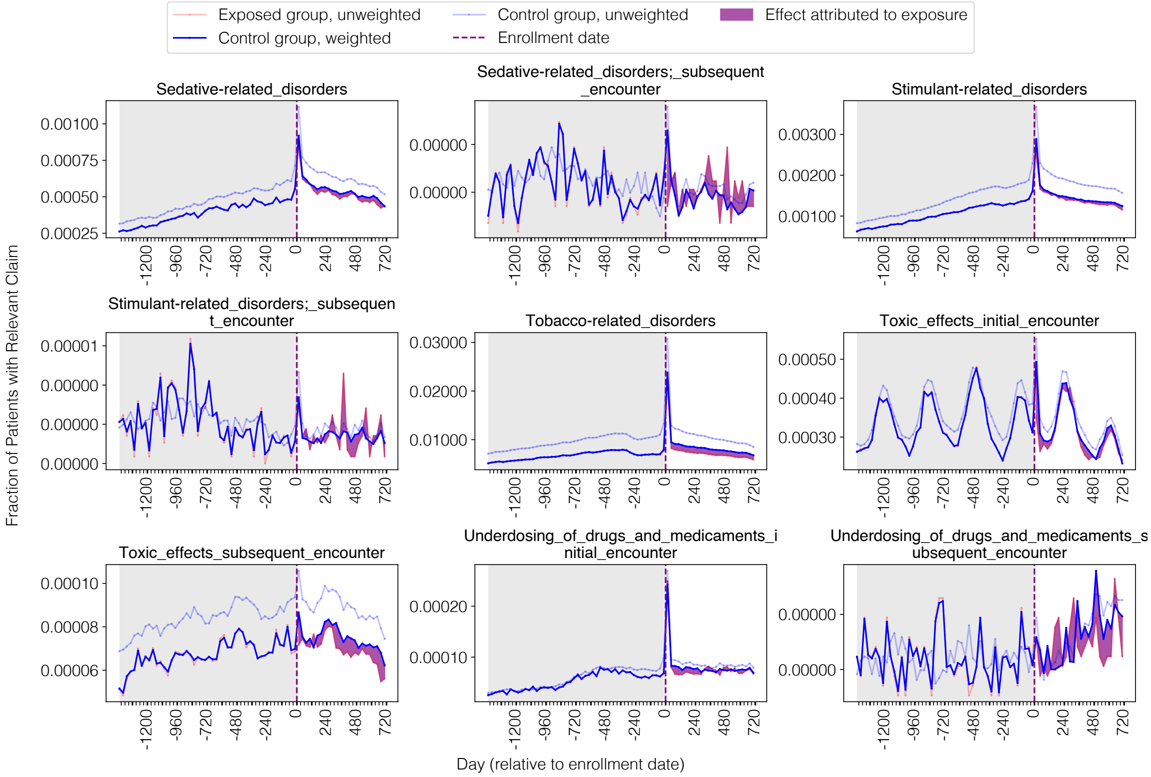


**Supplementary Figure S93:** (cont'd) Temporal trends in Substance use prevalence. The fraction of patients with relevant claims over time belonging for each outcome in the **prospective study** with testing-based enrollment experiment.

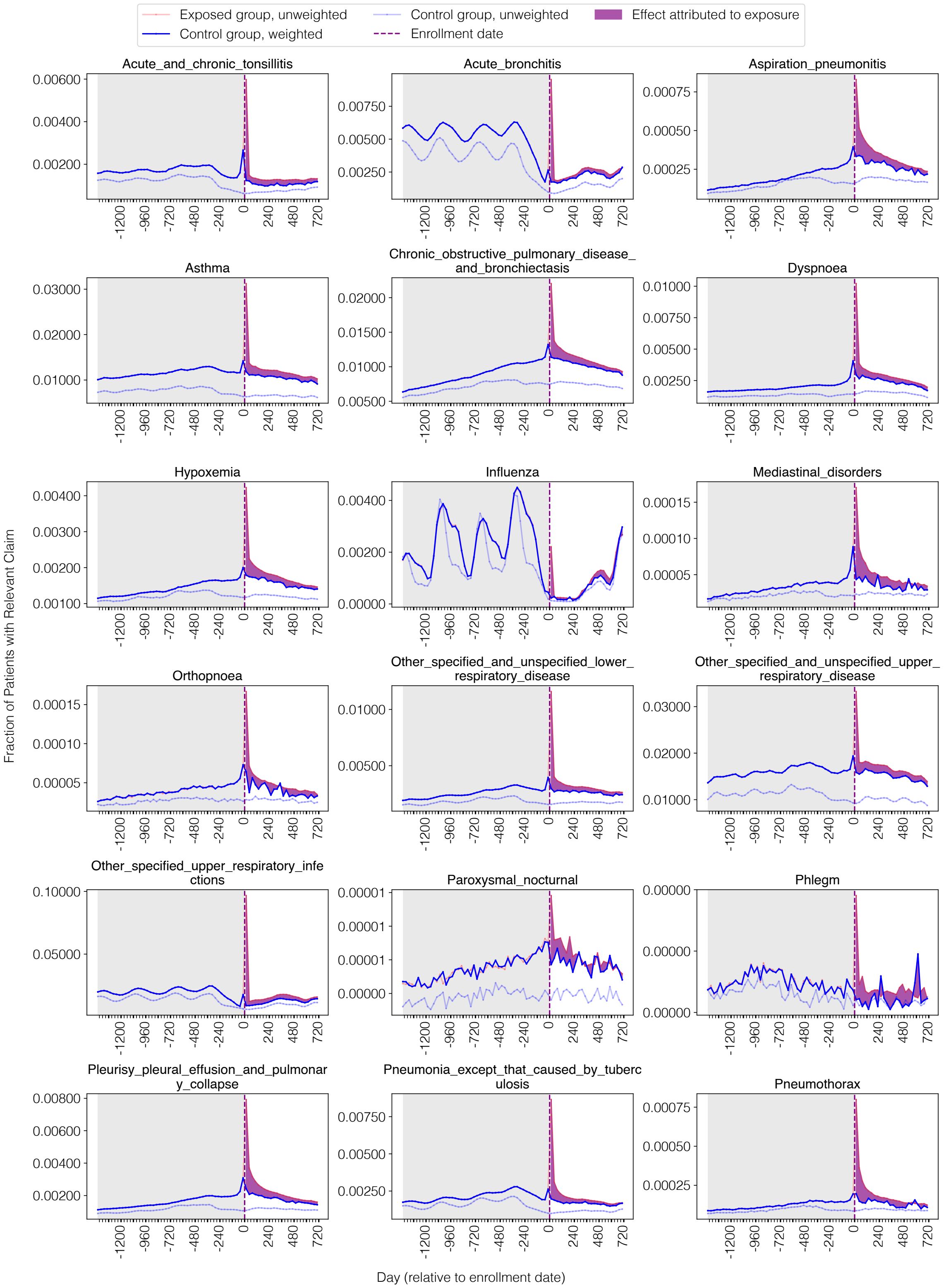


**Supplementary Figure S94:** Temporal trends in Breathing prevalence. The fraction of patients with relevant claims over time belonging for each outcome in the **conventional retrospective study with placebo cases** (negative COVID-19 PCR test) experiment.

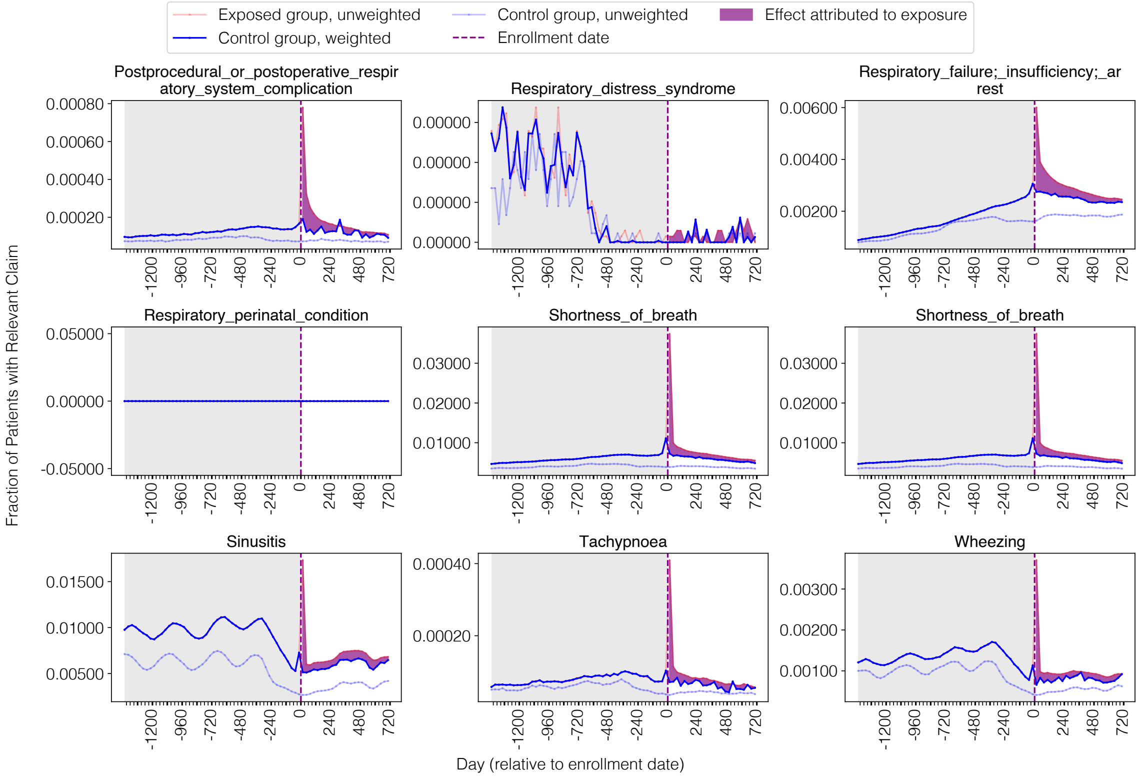


**Supplementary Figure S94:** (cont'd) Temporal trends in Breathing prevalence. The fraction of patients with relevant claims over time belonging for each outcome in the **conventional retrospective study with placebo cases** (negative COVID-19 PCR test) experiment.

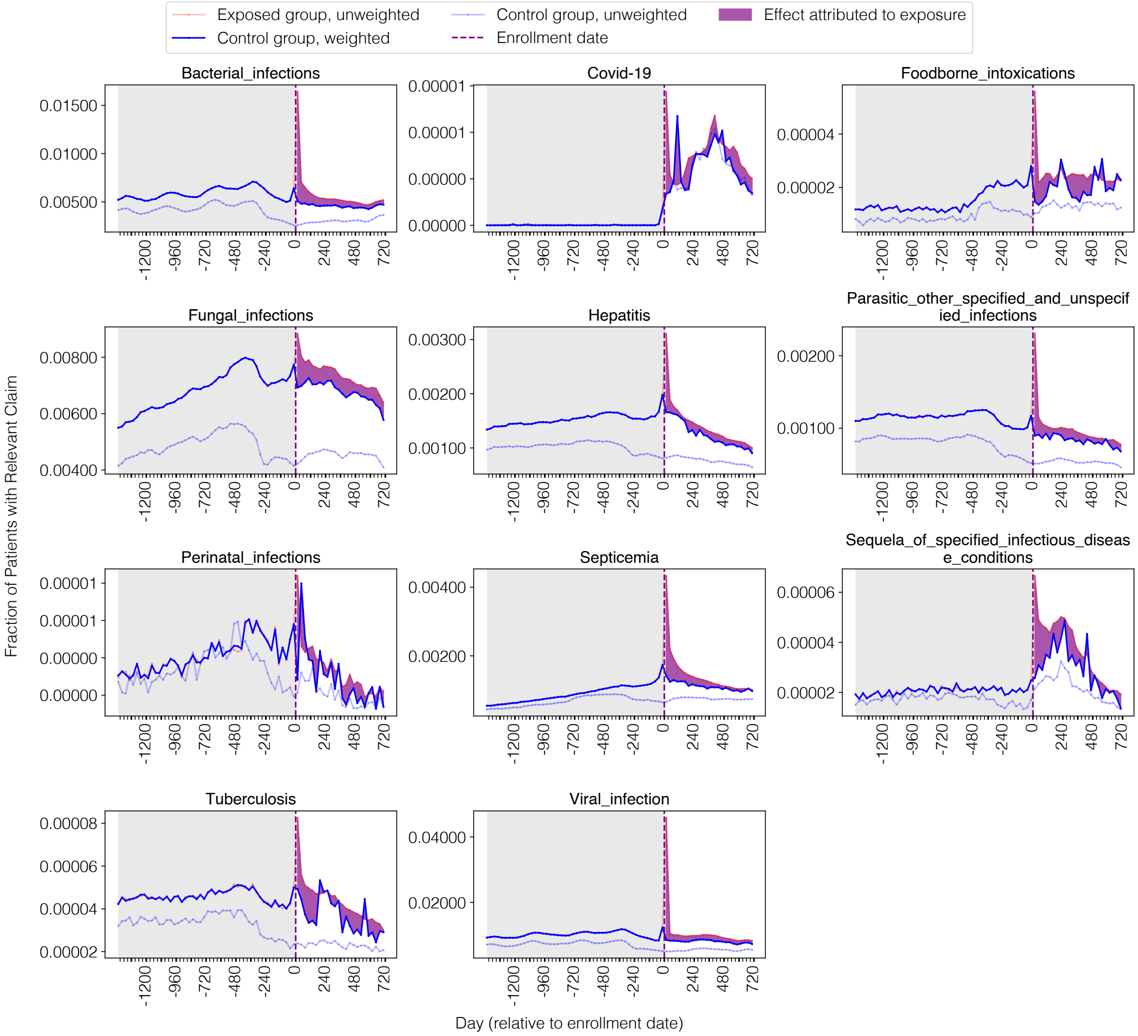


**Supplementary Figure S95:** Temporal trends in Certain Infectious and Parasitic Diseases prevalence. The fraction of patients with relevant claims over time belonging for each outcome in the **conventional retrospective study with placebo cases** (negative COVID-19 PCR test) experiment.

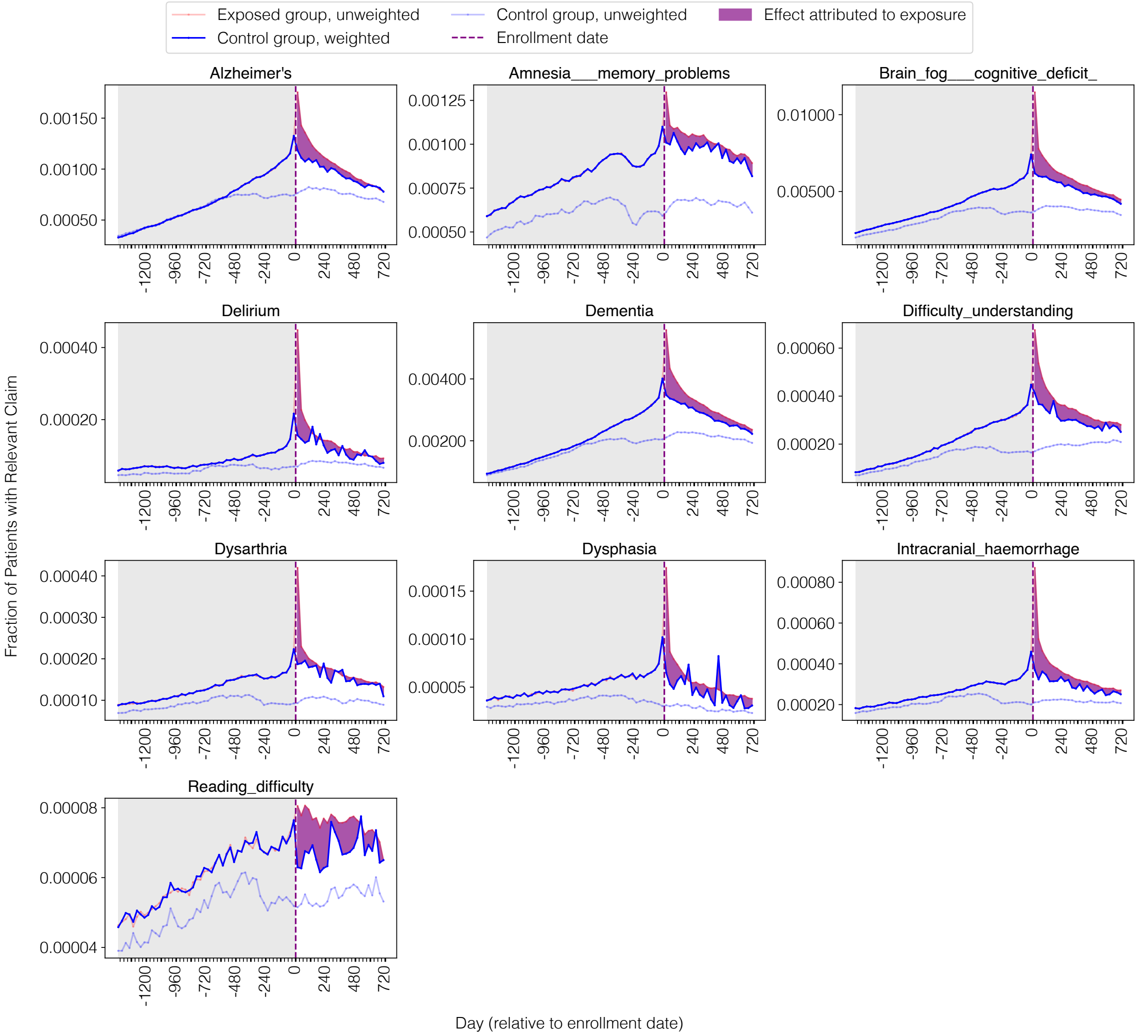


**Supplementary Figure S96:** Temporal trends in Cognitive health prevalence. The fraction of patients with relevant claims over time belonging for each outcome in the **conventional retrospective study with placebo cases** (negative COVID-19 PCR test) experiment.

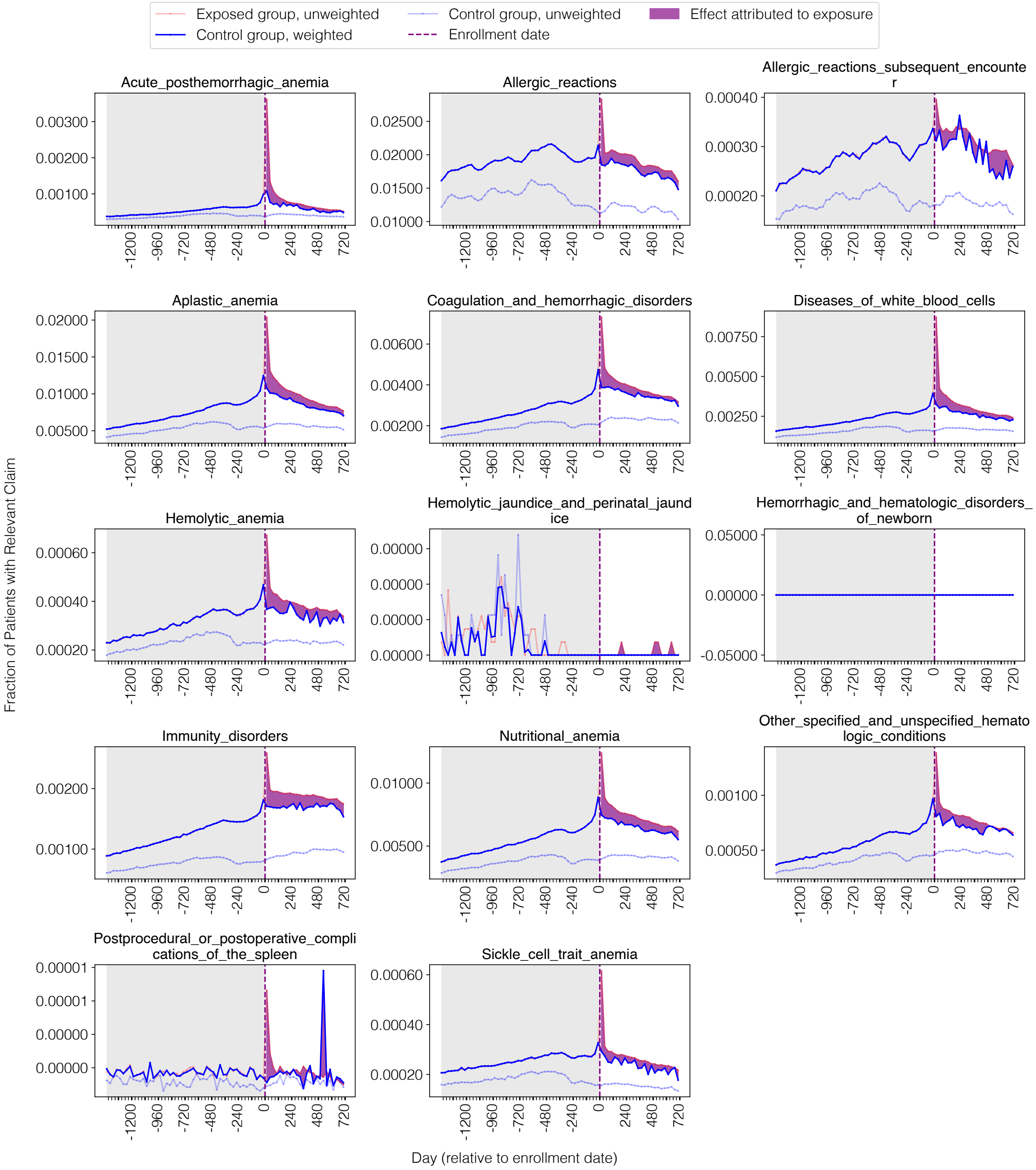


**Supplementary Figure S97:** Temporal trends in Diseases of the Blood and Blood Forming Organs and Certain Disorders Involving the Immune Mechanism prevalence. The fraction of patients with relevant claims over time belonging for each outcome in the **conventional retrospective study with placebo cases** (negative COVID-19 PCR test) experiment.

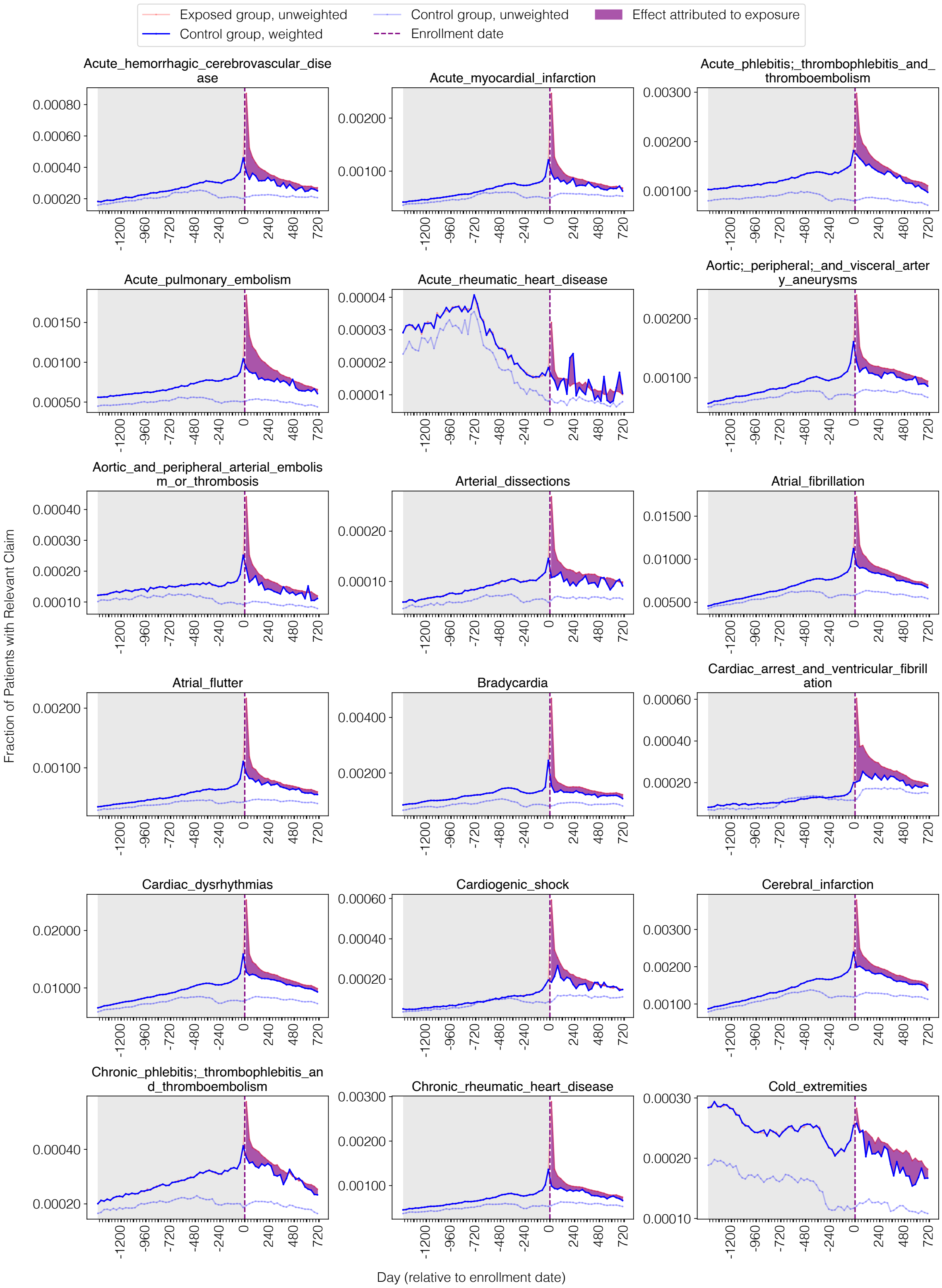


**Supplementary Figure S98:** Temporal trends in Diseases of the Circulatory System prevalence. The fraction of patients with relevant claims over time belonging for each outcome in the **conventional retrospective study with placebo cases** (negative COVID-19 PCR test) experiment.

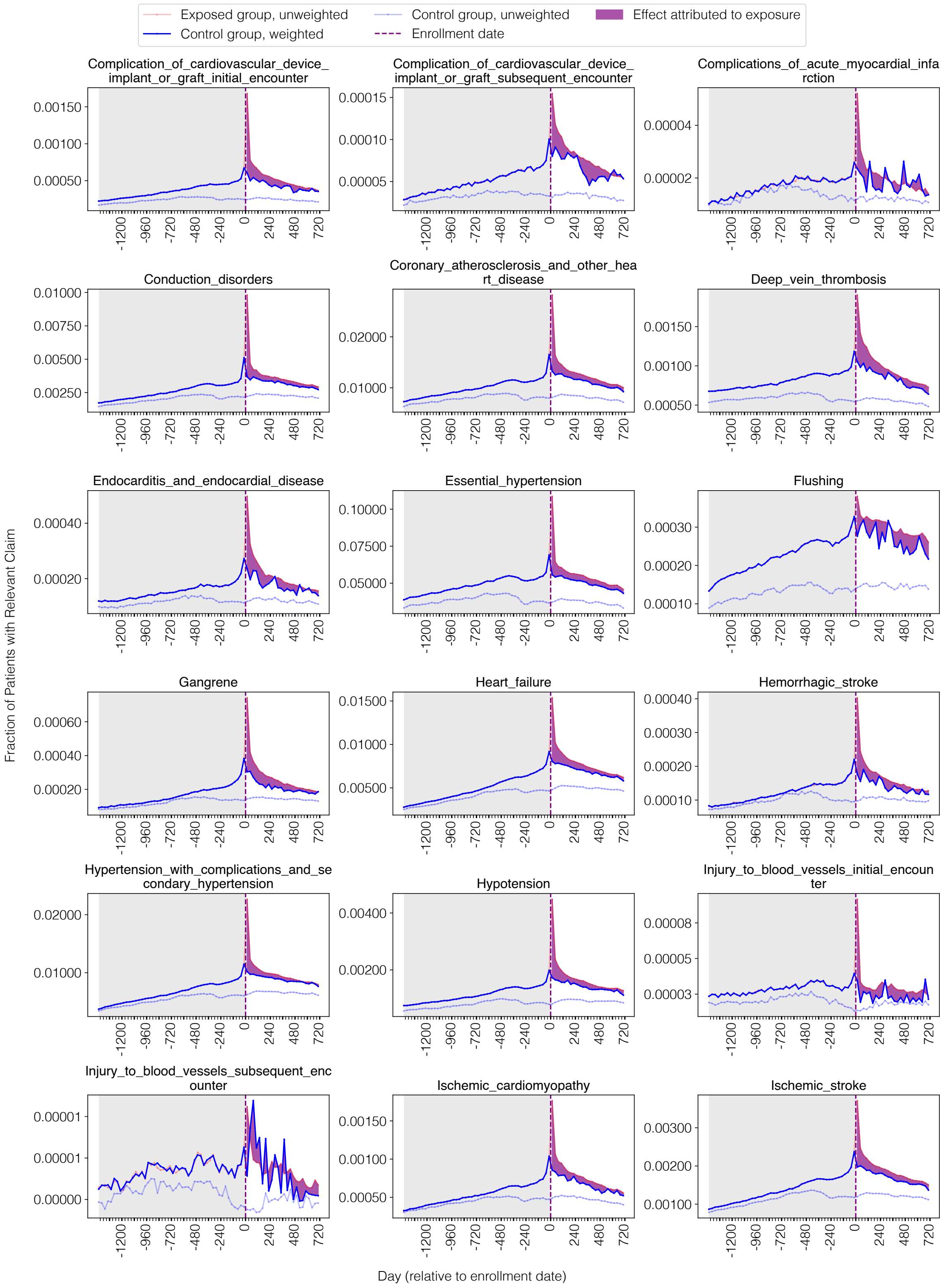


**Supplementary Figure S98:** (cont'd) Temporal trends in Diseases of the Circulatory System prevalence. The fraction of patients with relevant claims over time belonging for each outcome in the **conventional retrospective study with placebo cases** (negative COVID-19 PCR test) experiment.

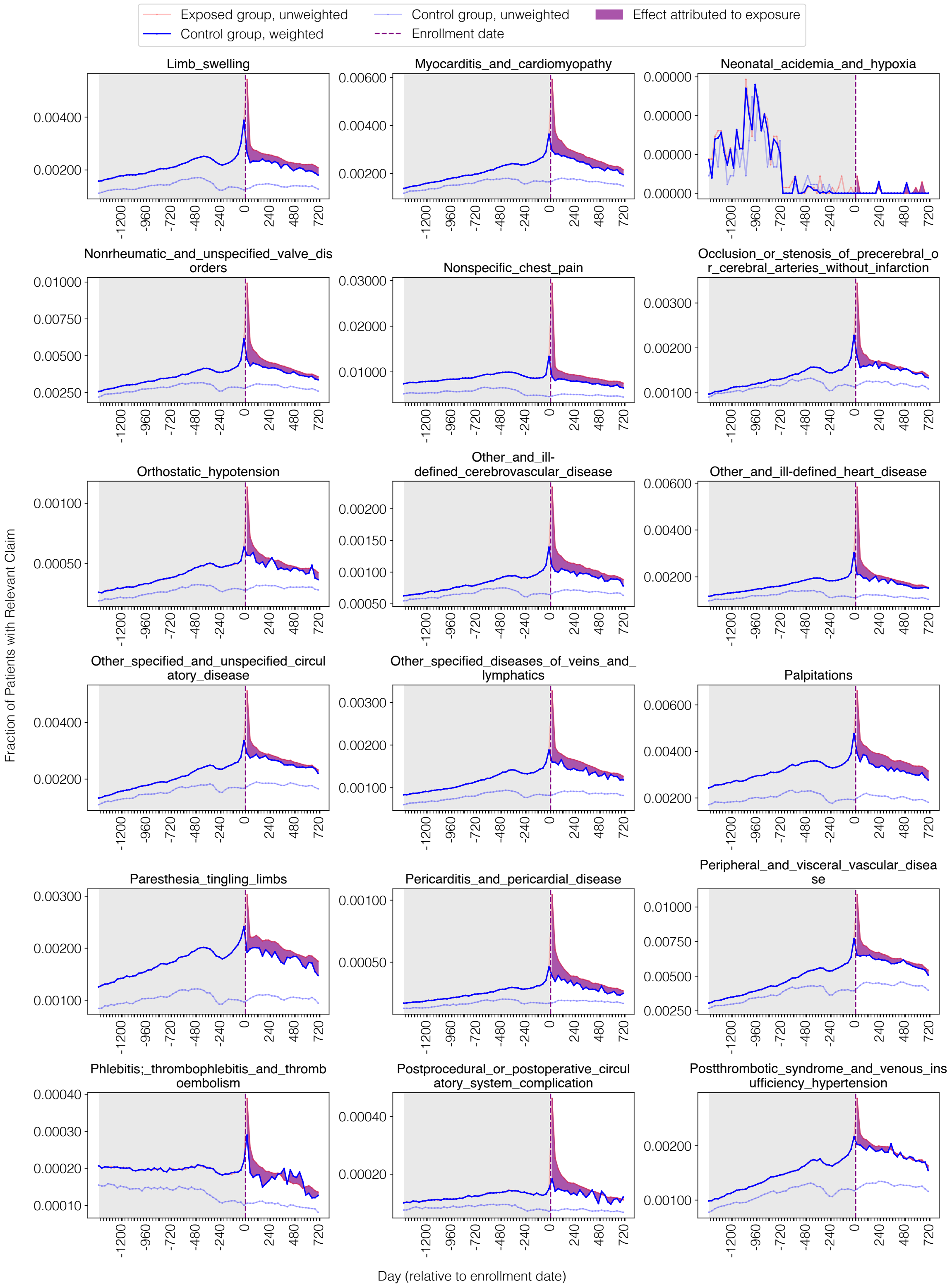


**Supplementary Figure S98:** (cont'd) Temporal trends in Diseases of the Circulatory System prevalence. The fraction of patients with relevant claims over time belonging for each outcome in the **conventional retrospective study with placebo cases** (negative COVID-19 PCR test) experiment.

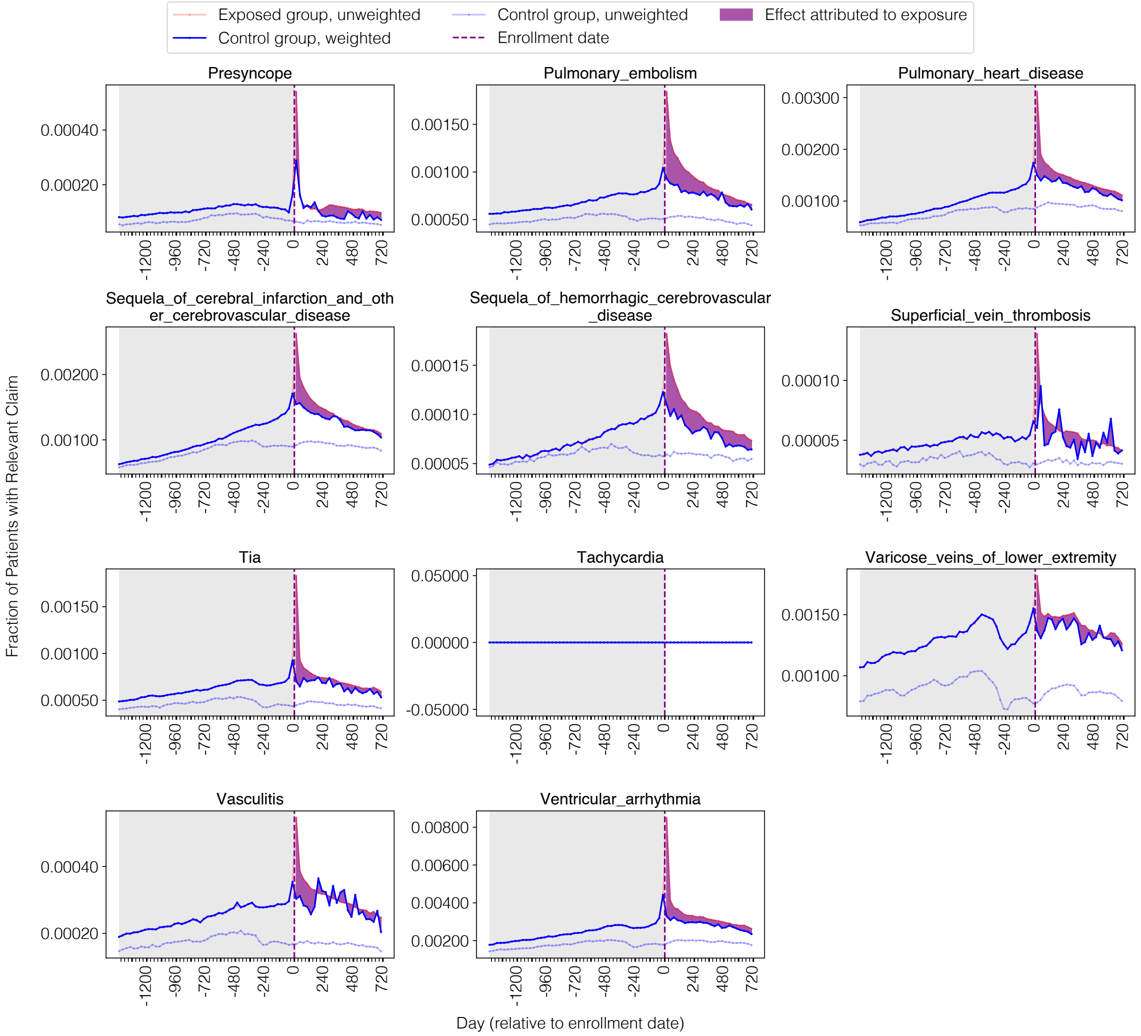


**Supplementary Figure S98:** (cont'd) Temporal trends in Diseases of the Circulatory System prevalence. The fraction of patients with relevant claims over time belonging for each outcome in the **conventional retrospective study with placebo cases** (negative COVID-19 PCR test) experiment.

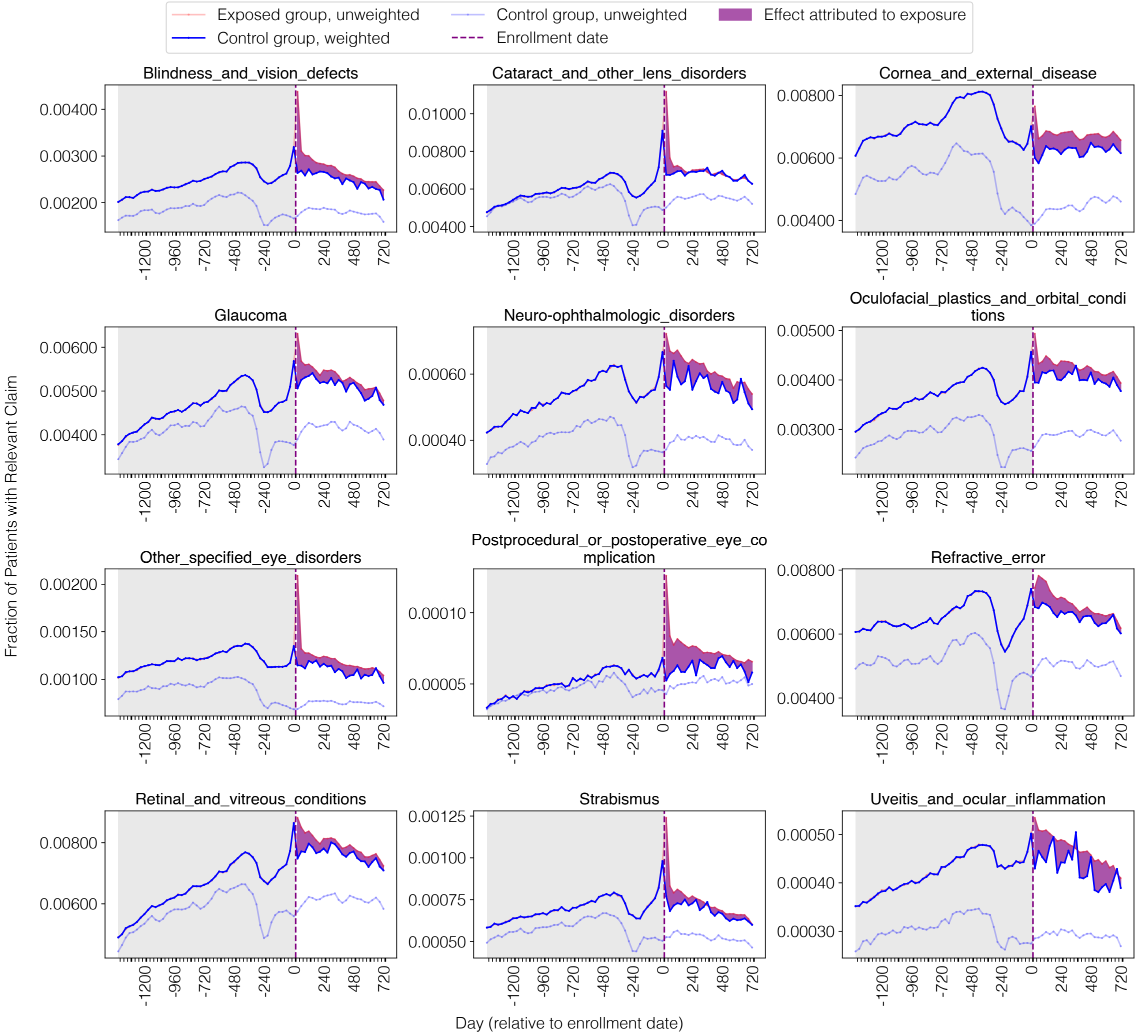


**Supplementary Figure S99:** Temporal trends in Diseases of the Eye and Adnexa prevalence. The fraction of patients with relevant claims over time belonging for each outcome in the **conventional retrospective study with placebo cases** (negative COVID-19 PCR test) experiment.

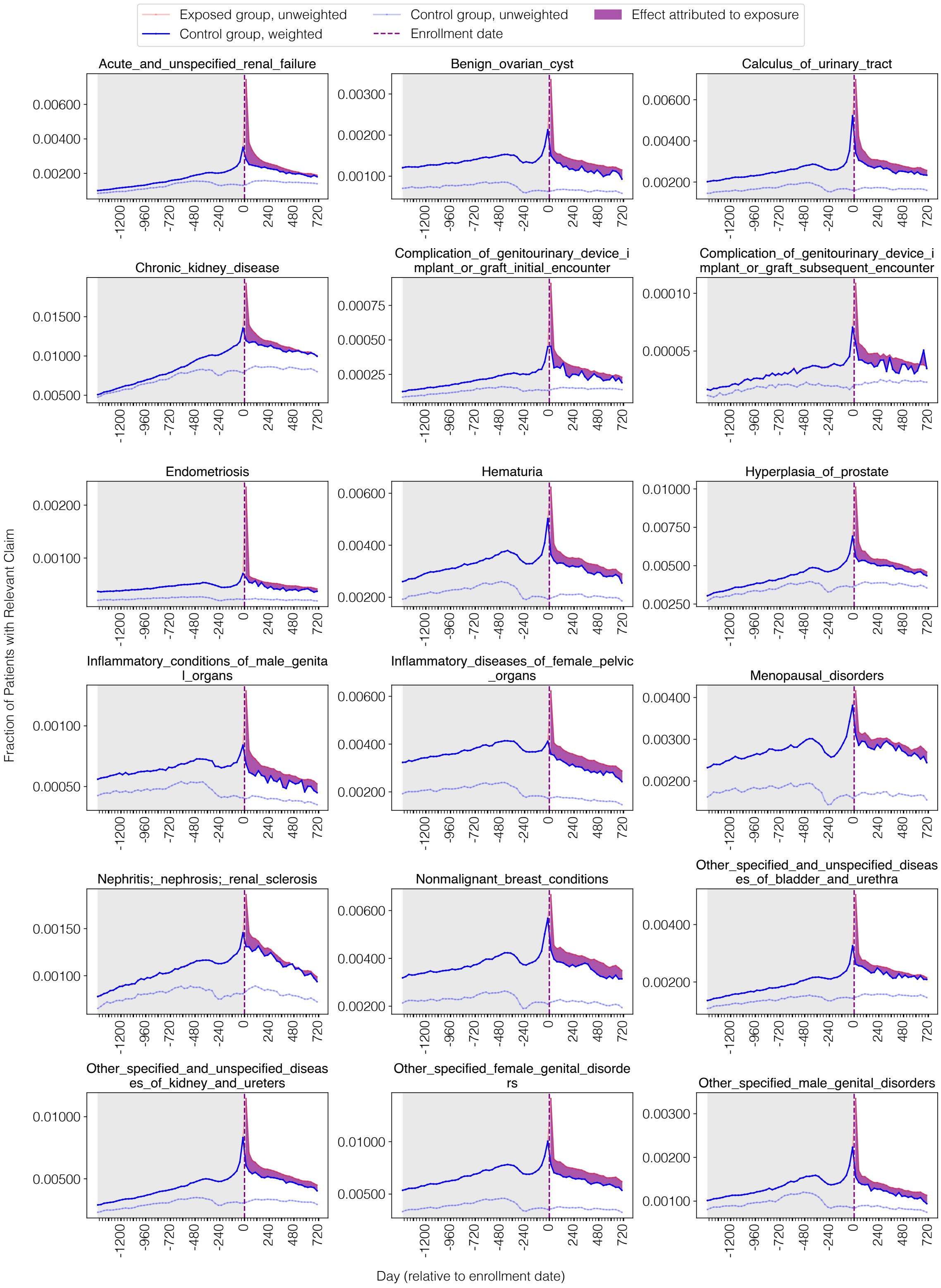


**Supplementary Figure S100:** Temporal trends in Diseases of the Genitourinary System prevalence. The fraction of patients with relevant claims over time belonging for each outcome in the **conventional retrospective study with placebo cases** (negative COVID-19 PCR test) experiment.

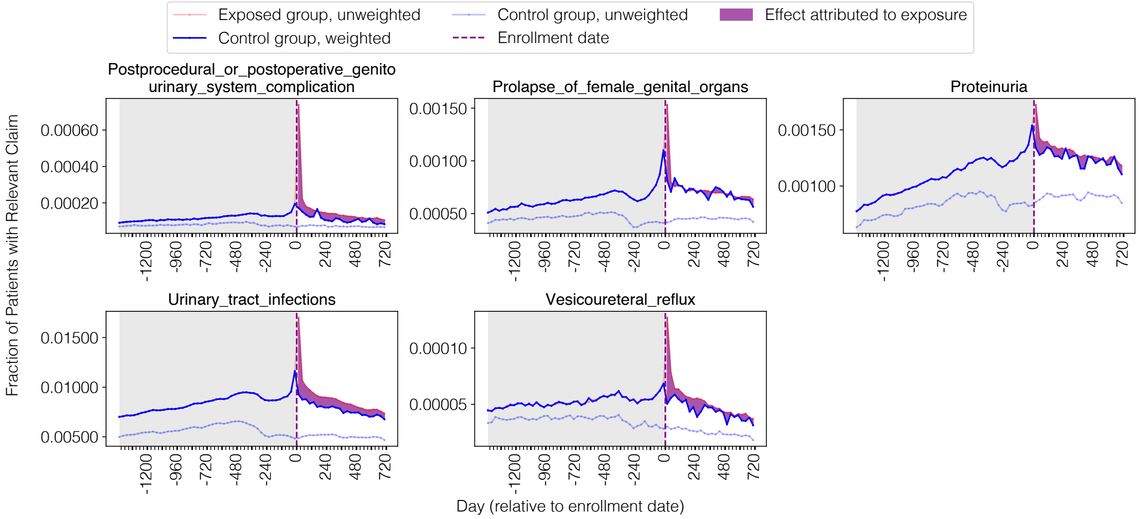


**Supplementary Figure S100:** (cont'd) Temporal trends in Diseases of the Genitourinary System prevalence. The fraction of patients with relevant claims over time belonging for each outcome in the **conventional retrospective study with placebo cases** (negative COVID-19 PCR test) experiment.

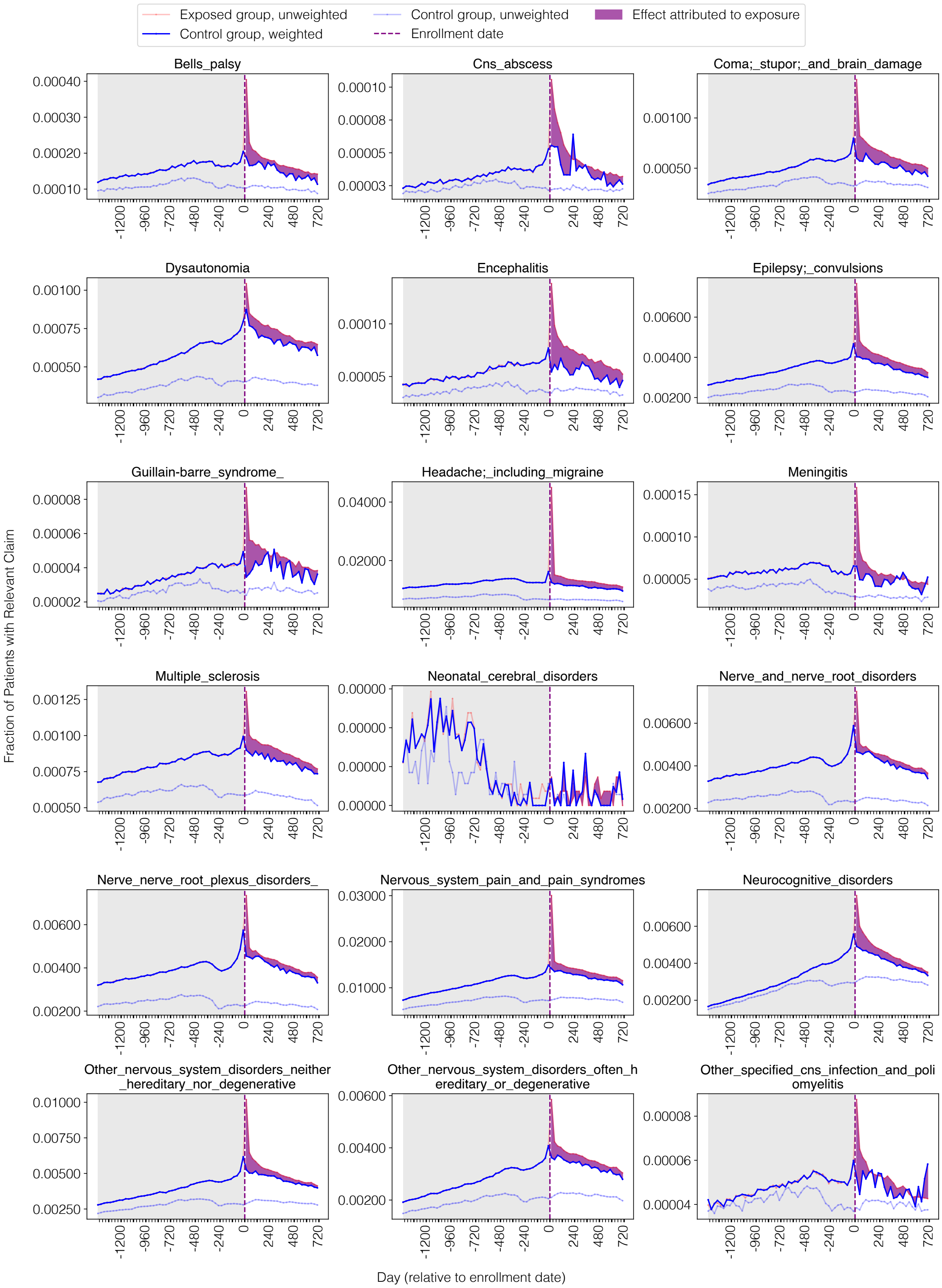


**Supplementary Figure S101:** Temporal trends in Diseases of the Nervous System prevalence. The fraction of patients with relevant claims over time belonging for each outcome in the **conventional retrospective study with placebo cases** (negative COVID-19 PCR test) experiment.

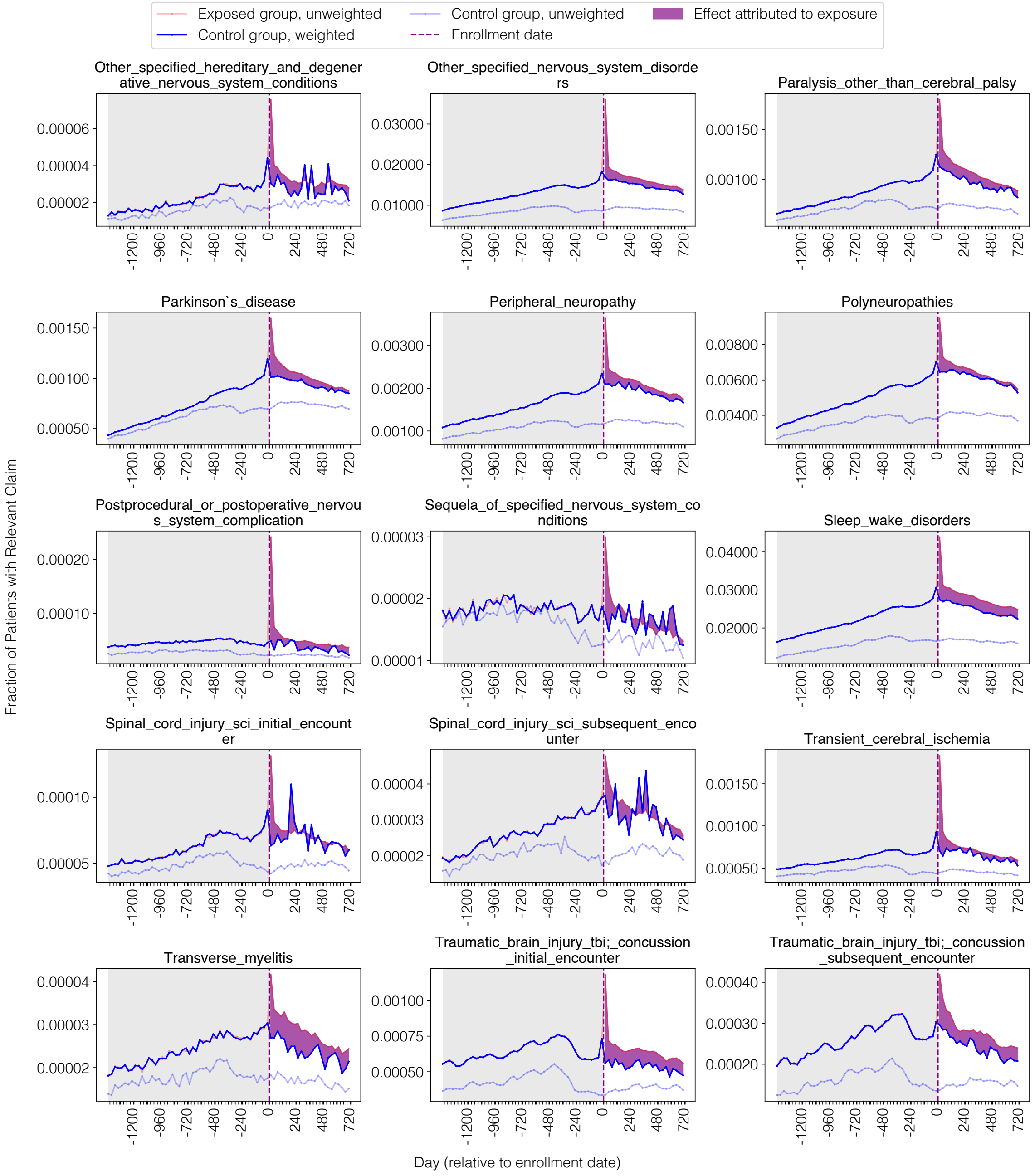


**Supplementary Figure S101:** (cont'd) Temporal trends in Diseases of the Nervous System prevalence. The fraction of patients with relevant claims over time belonging for each outcome in the **conventional retrospective study with placebo cases** (negative COVID-19 PCR test) experiment.

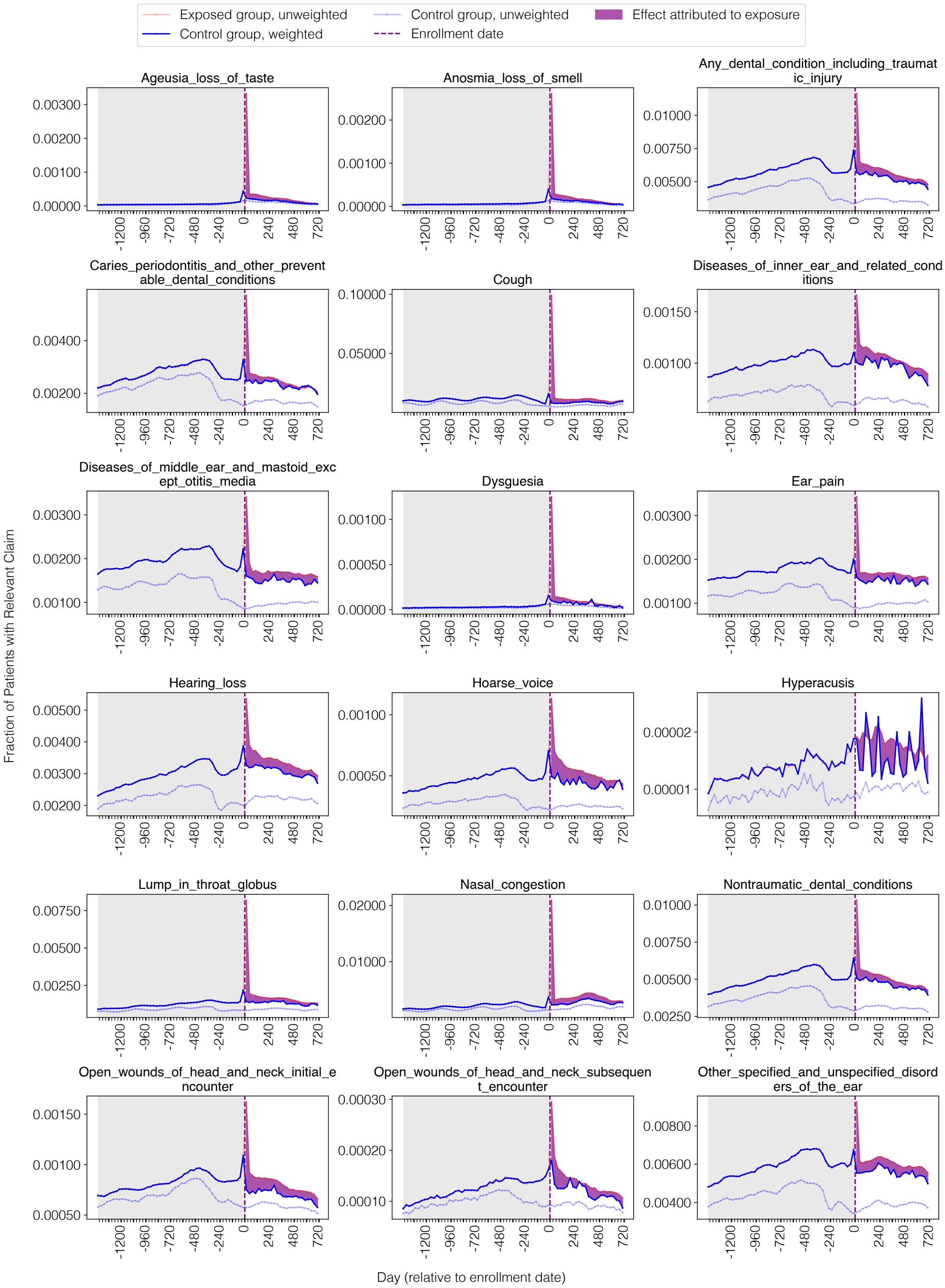


**Supplementary Figure S102:** Temporal trends in Ear, nose and throat prevalence. The fraction of patients with relevant claims over time belonging for each outcome in the **conventional retrospective study with placebo cases** (negative COVID-19 PCR test) experiment.

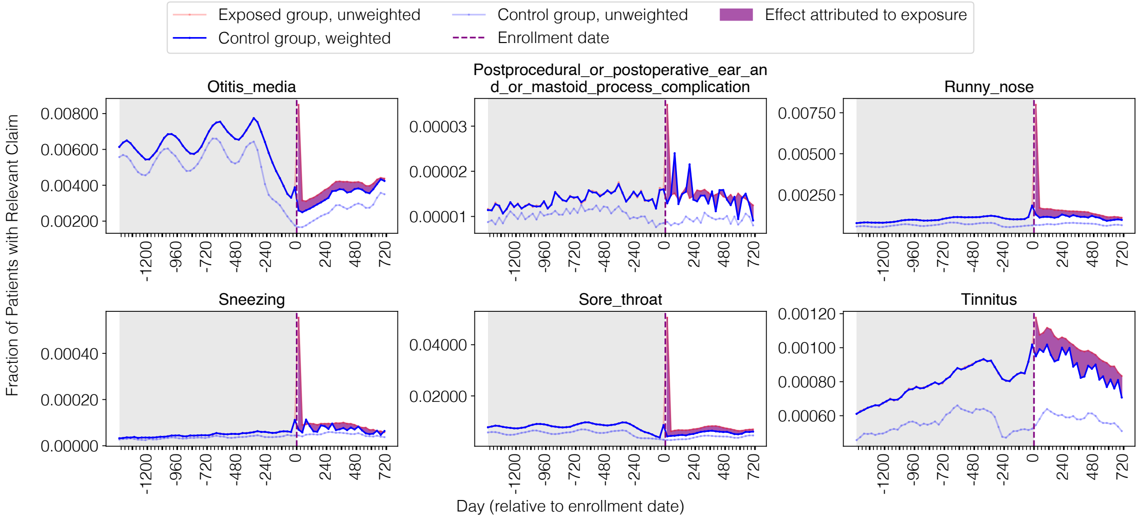


**Supplementary Figure S102:** (cont'd) Temporal trends in Ear, nose and throat prevalence. The fraction of patients with relevant claims over time belonging for each outcome in the **conventional retrospective study with placebo cases** (negative COVID-19 PCR test) experiment.

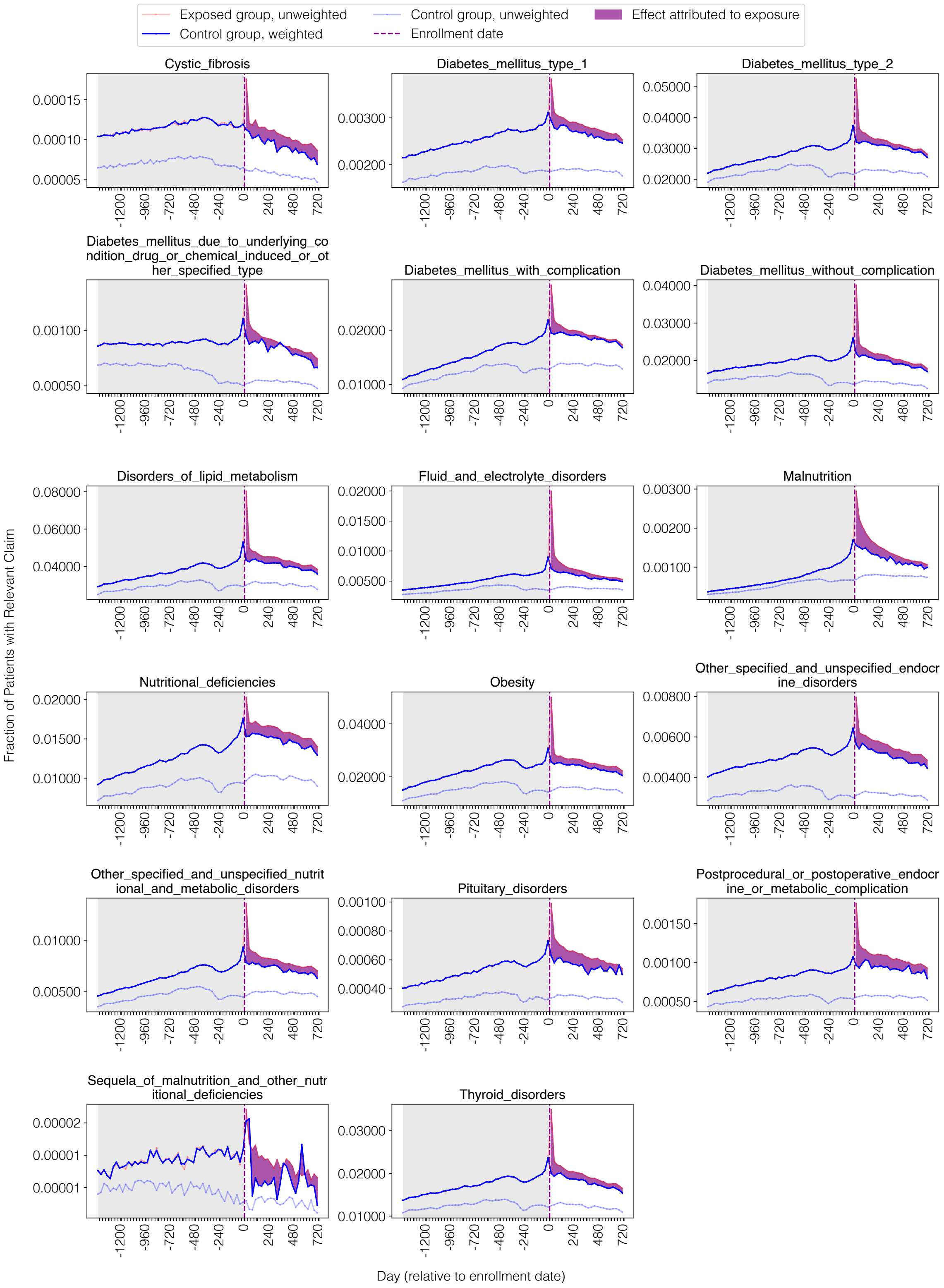


**Supplementary Figure S103:** Temporal trends in Endocrine, Nutritional and Metabolic Diseases prevalence. The fraction of patients with relevant claims over time belonging for each outcome in the **conventional retrospective study with placebo cases** (negative COVID-19 PCR test) experiment.

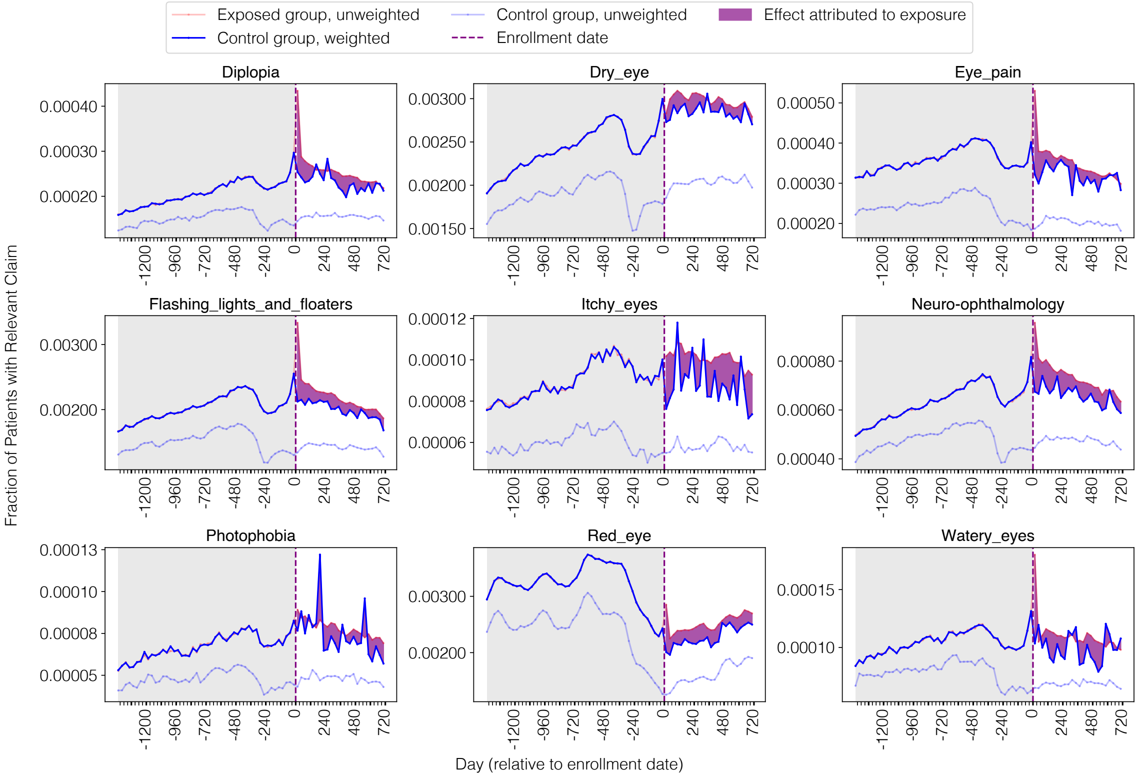


**Supplementary Figure S104:** Temporal trends in Eyes prevalence. The fraction of patients with relevant claims over time belonging for each outcome in the **conventional retrospective study with placebo cases** (negative COVID-19 PCR test) experiment.

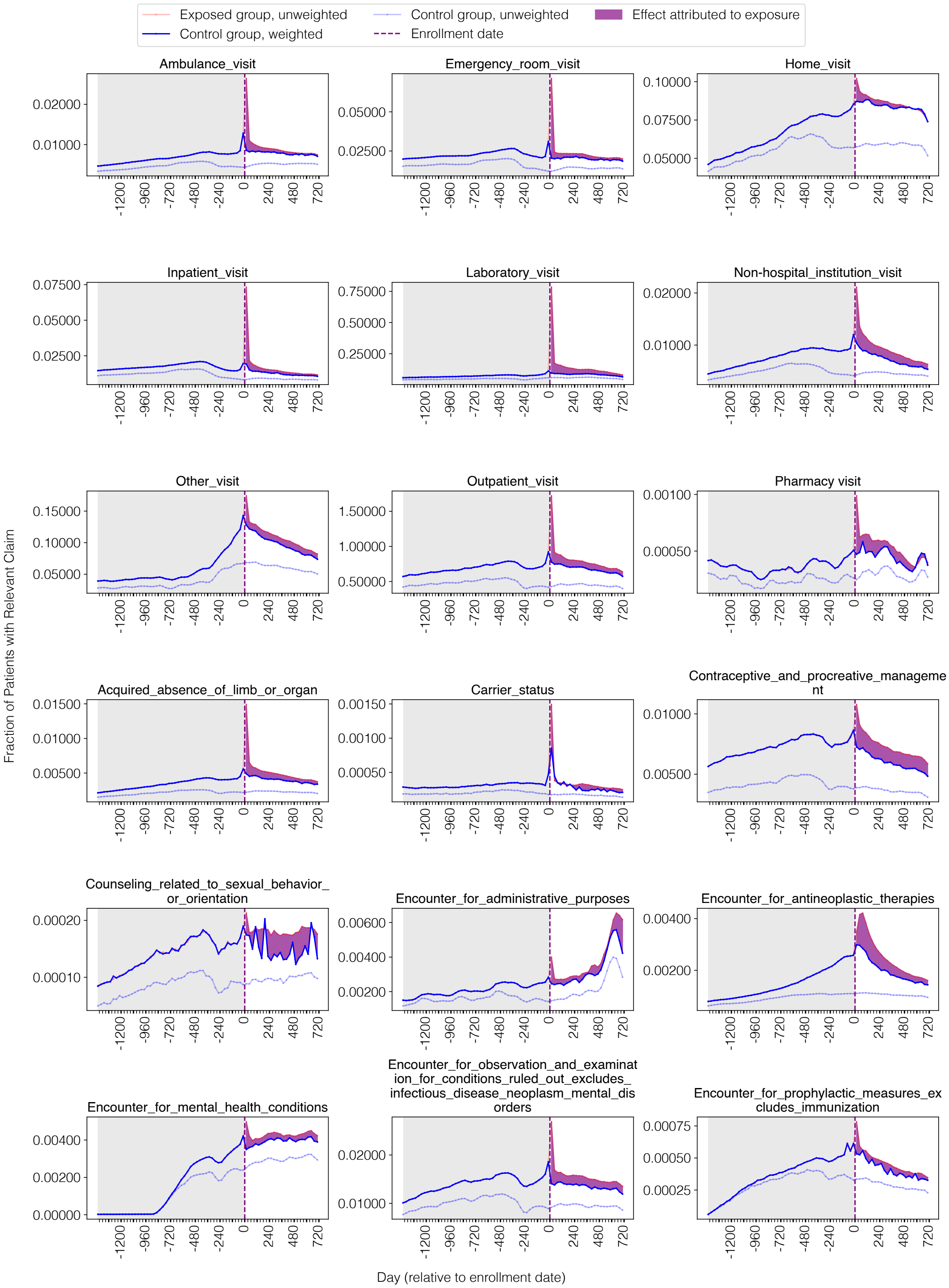


**Supplementary Figure S105:** Temporal trends in Factors Influencing Health Status and Contact with Health Services prevalence. The fraction of patients with relevant claims over time belonging for each outcome in the **conventional retrospective study with placebo cases** (negative COVID-19 PCR test) experiment.

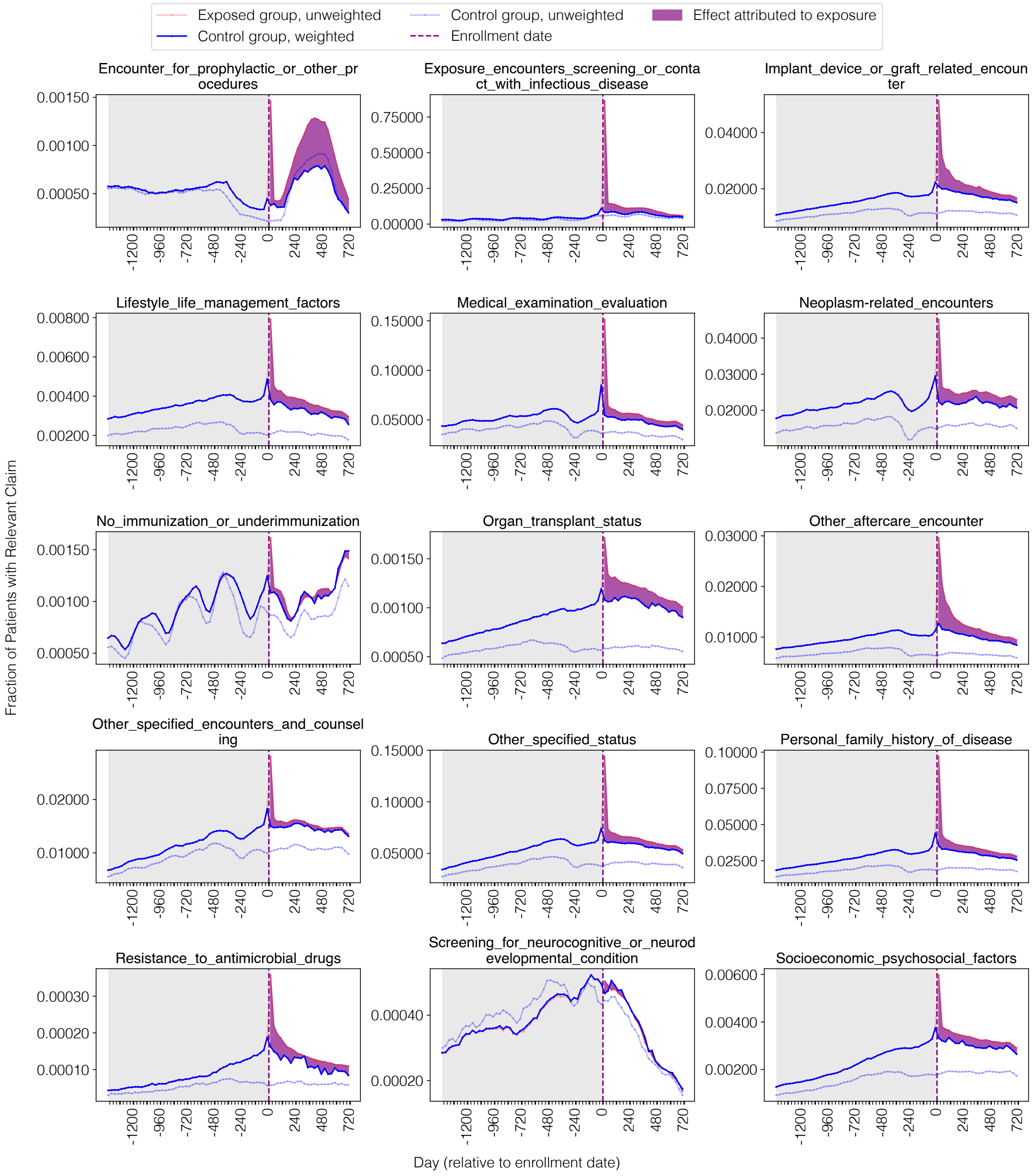


**Supplementary Figure S105:** (cont'd) Temporal trends in Factors Influencing Health Status and Contact with Health Services prevalence. The fraction of patients with relevant claims over time belonging for each outcome in the **conventional retrospective study with placebo cases** (negative COVID-19 PCR test) experiment.

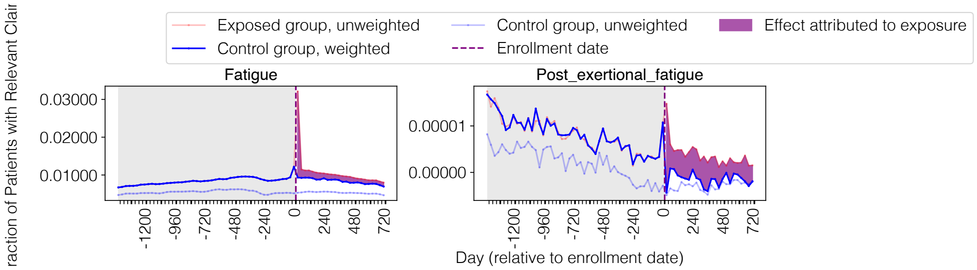


**Supplementary Figure S106:** Temporal trends in Fatigue prevalence. The fraction of patients with relevant claims over time belonging for each outcome in the **conventional retrospective study with placebo cases** (negative COVID-19 PCR test) experiment.

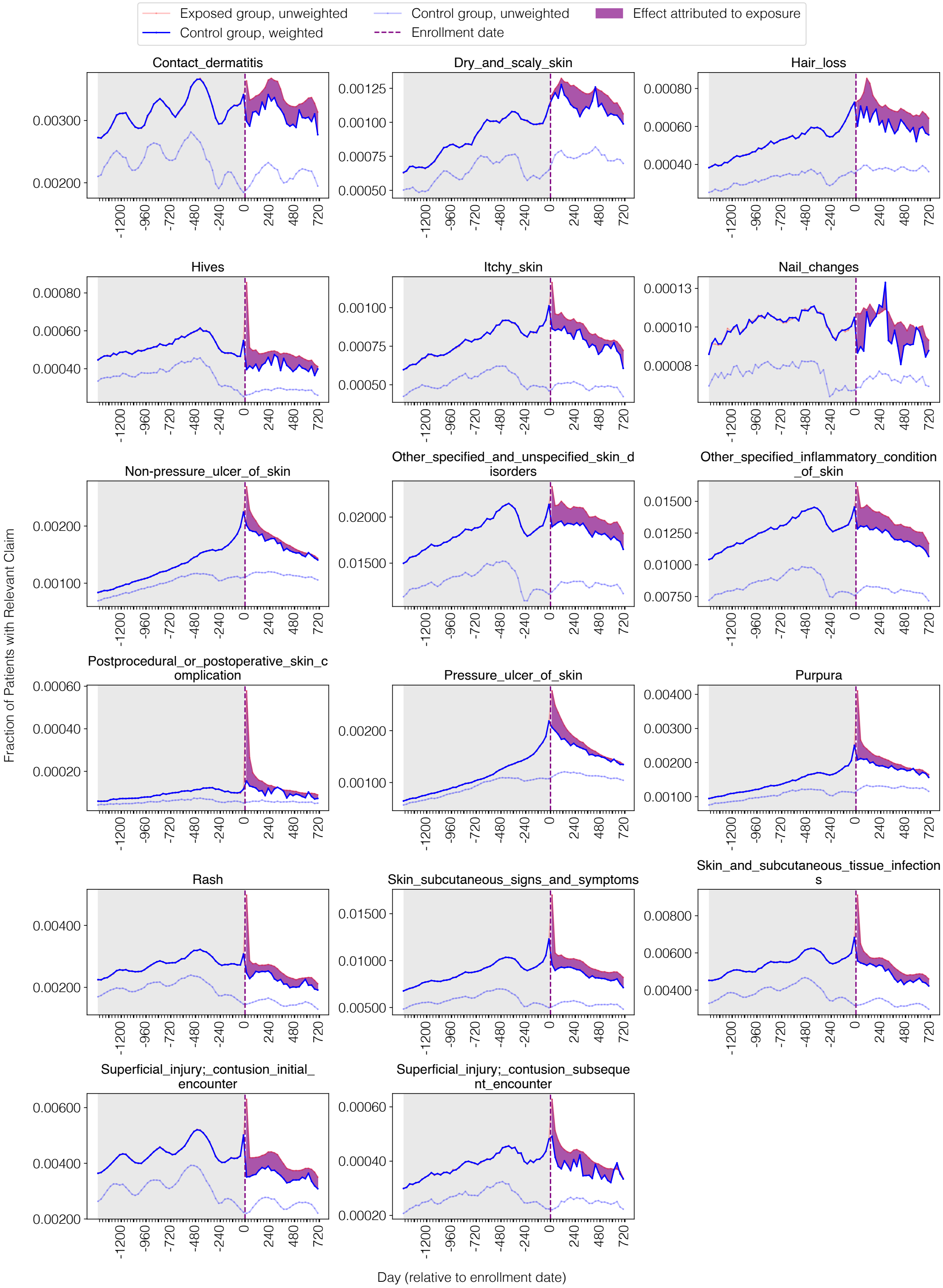


**Supplementary Figure S107:** Temporal trends in Hair, skin and nails prevalence. The fraction of patients with relevant claims over time belonging for each outcome in the **conventional retrospective study with placebo cases** (negative COVID-19 PCR test) experiment.

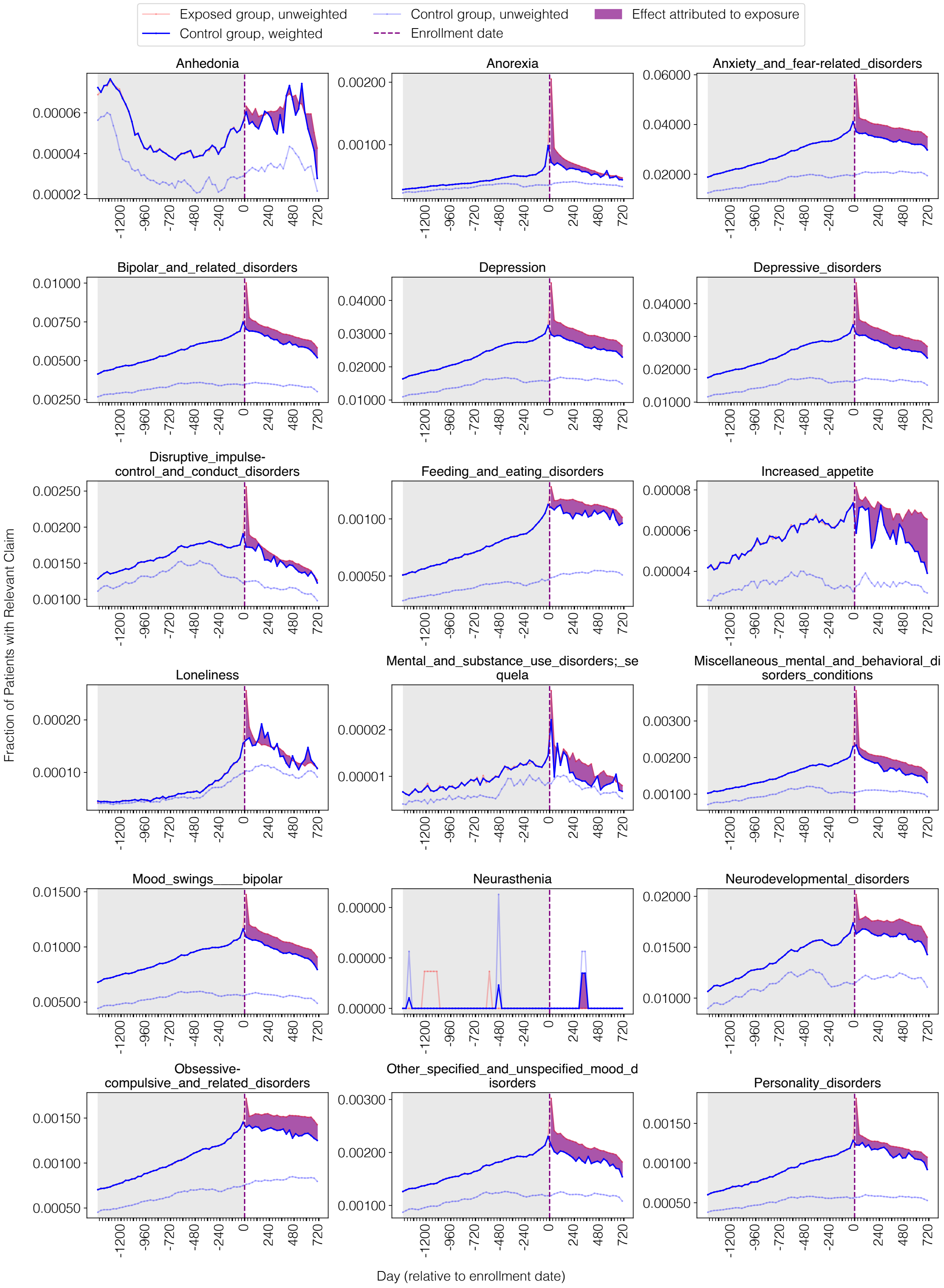


**Supplementary Figure S108:** Temporal trends in Mental health prevalence. The fraction of patients with relevant claims over time belonging for each outcome in the **conventional retrospective study with placebo cases** (negative COVID-19 PCR test) experiment.

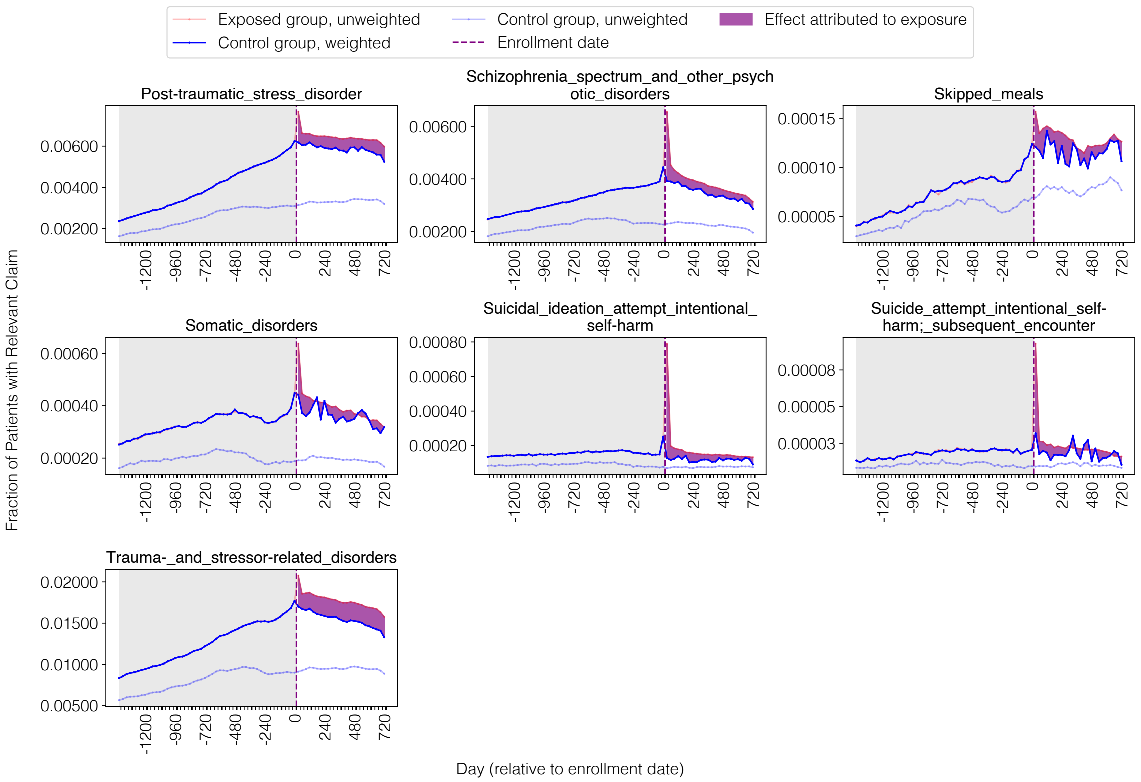


**Supplementary Figure S108:** (cont'd) Temporal trends in Mental health prevalence. The fraction of patients with relevant claims over time belonging for each outcome in the **conventional retrospective study with placebo cases** (negative COVID-19 PCR test) experiment.

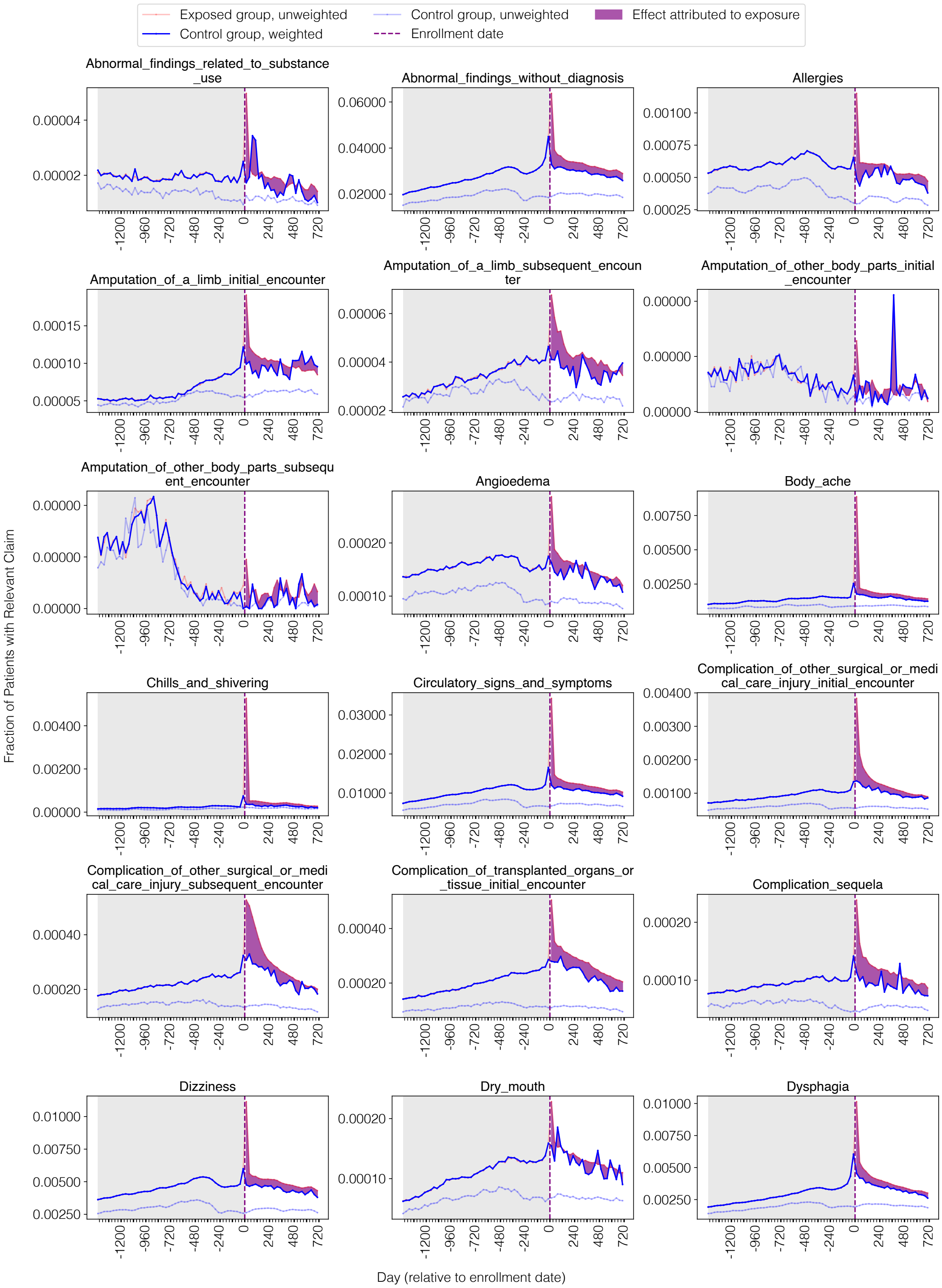


**Supplementary Figure S109:** Temporal trends in Misc prevalence. The fraction of patients with relevant claims over time belonging for each outcome in the **conventional retrospective study with placebo cases** (negative COVID-19 PCR test) experiment.

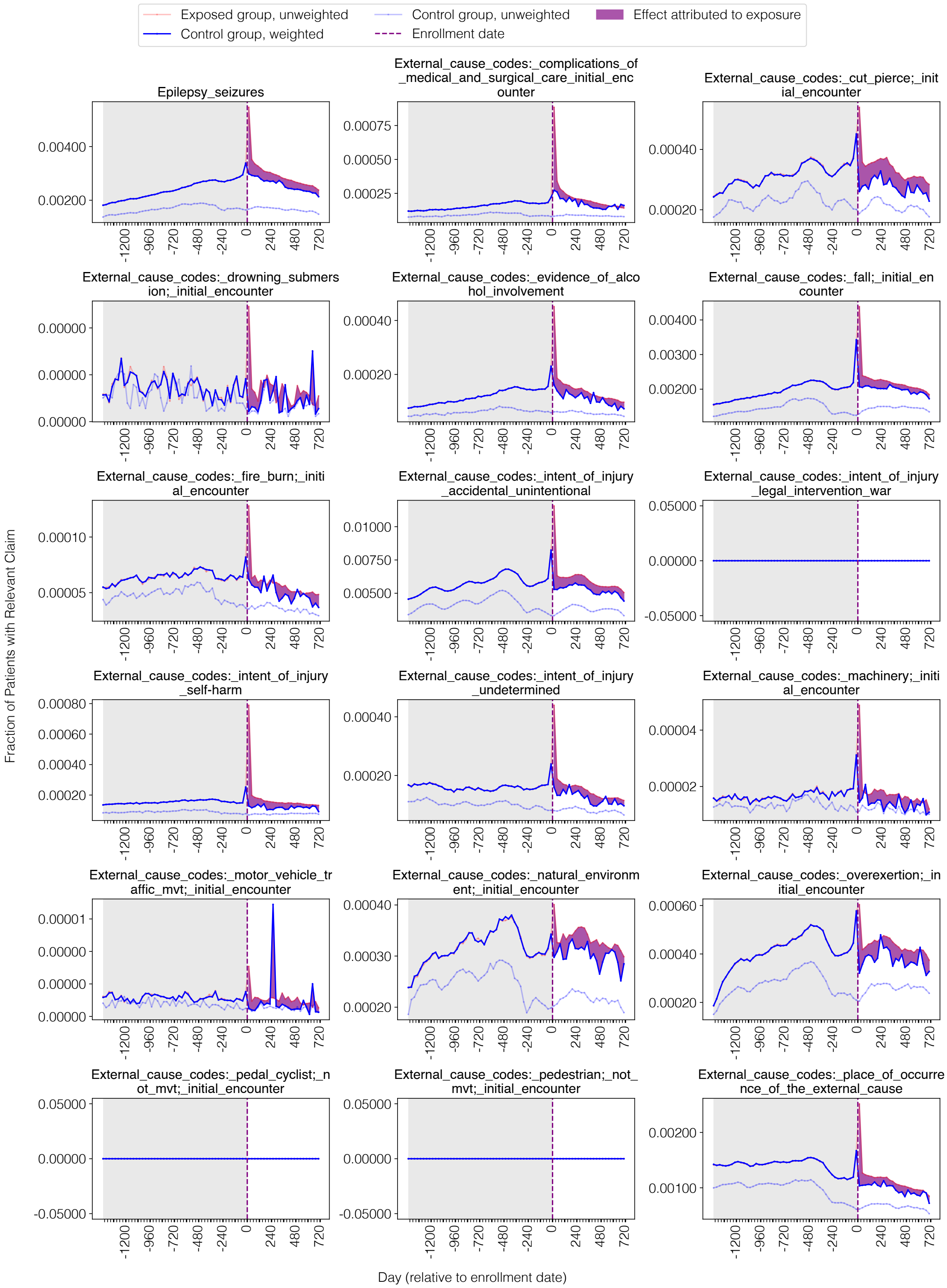


**Supplementary Figure S109:** (cont'd) Temporal trends in Misc prevalence. The fraction of patients with relevant claims over time belonging for each outcome in the **conventional retrospective study with placebo cases** (negative COVID-19 PCR test) experiment.

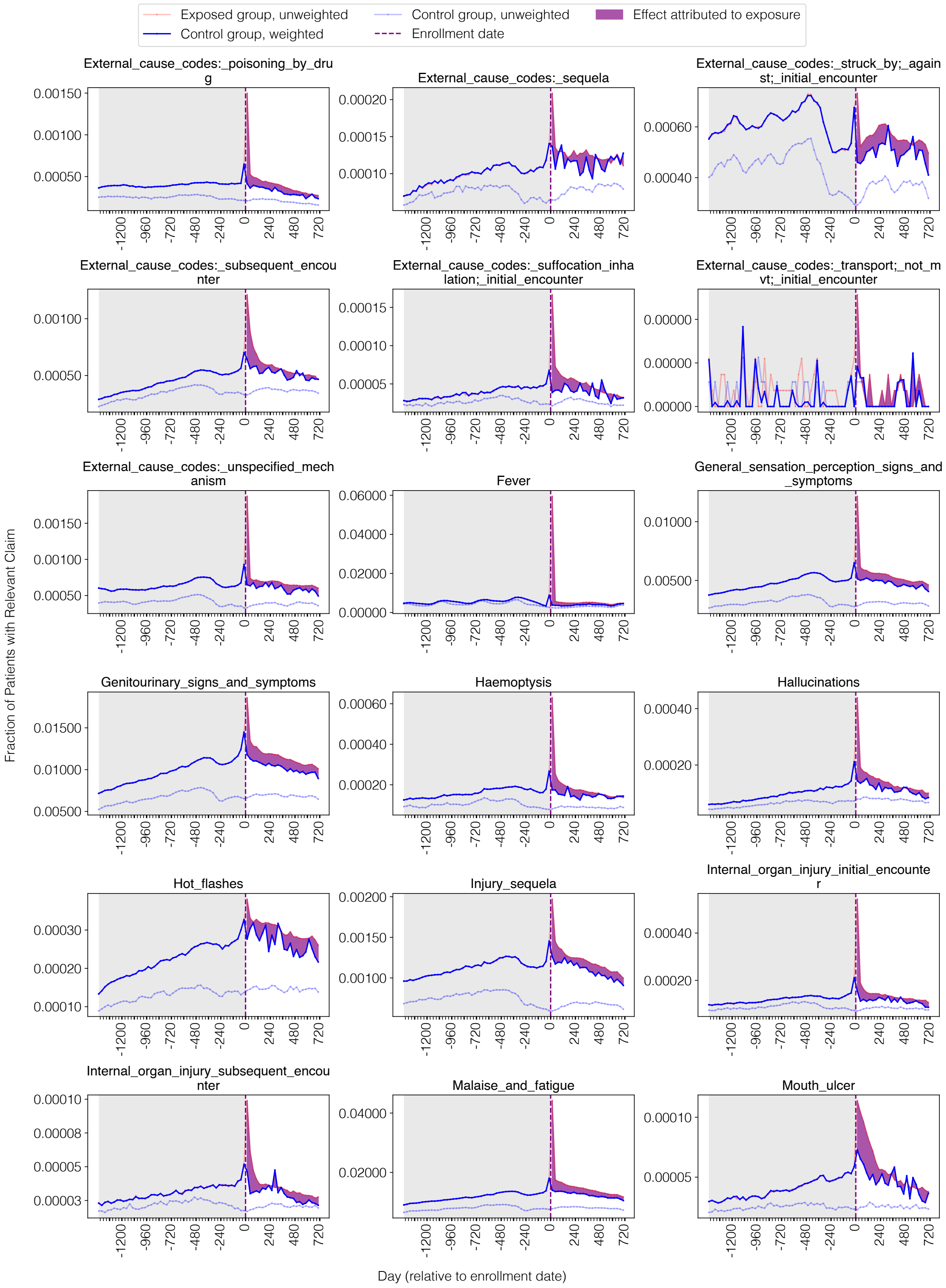


**Supplementary Figure S109:** (cont'd) Temporal trends in Misc prevalence. The fraction of patients with relevant claims over time belonging for each outcome in the **conventional retrospective study with placebo cases** (negative COVID-19 PCR test) experiment.

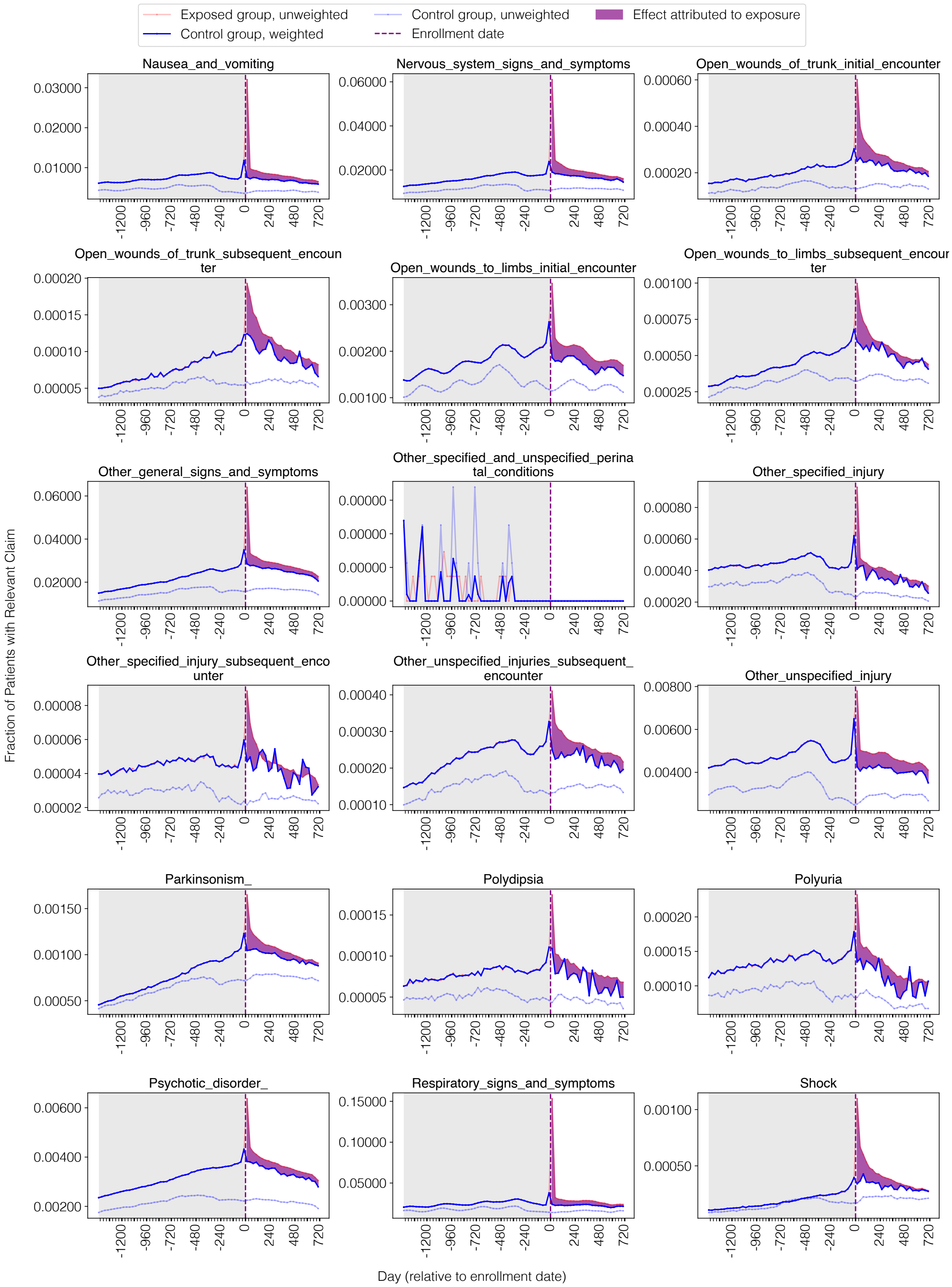


**Supplementary Figure S109:** (cont'd) Temporal trends in Misc prevalence. The fraction of patients with relevant claims over time belonging for each outcome in the **conventional retrospective study with placebo cases** (negative COVID-19 PCR test) experiment.

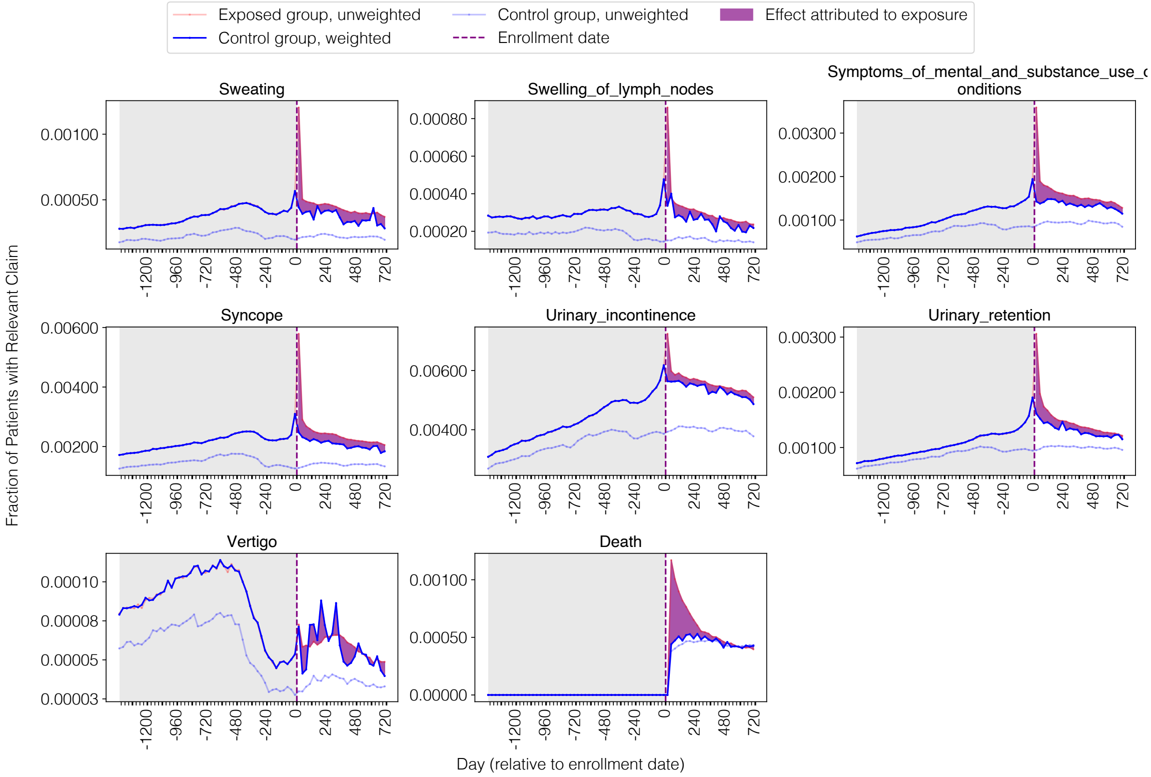


**Supplementary Figure S109:** (cont'd) Temporal trends in Misc prevalence. The fraction of patients with relevant claims over time belonging for each outcome in the **conventional retrospective study with placebo cases** (negative COVID-19 PCR test) experiment.

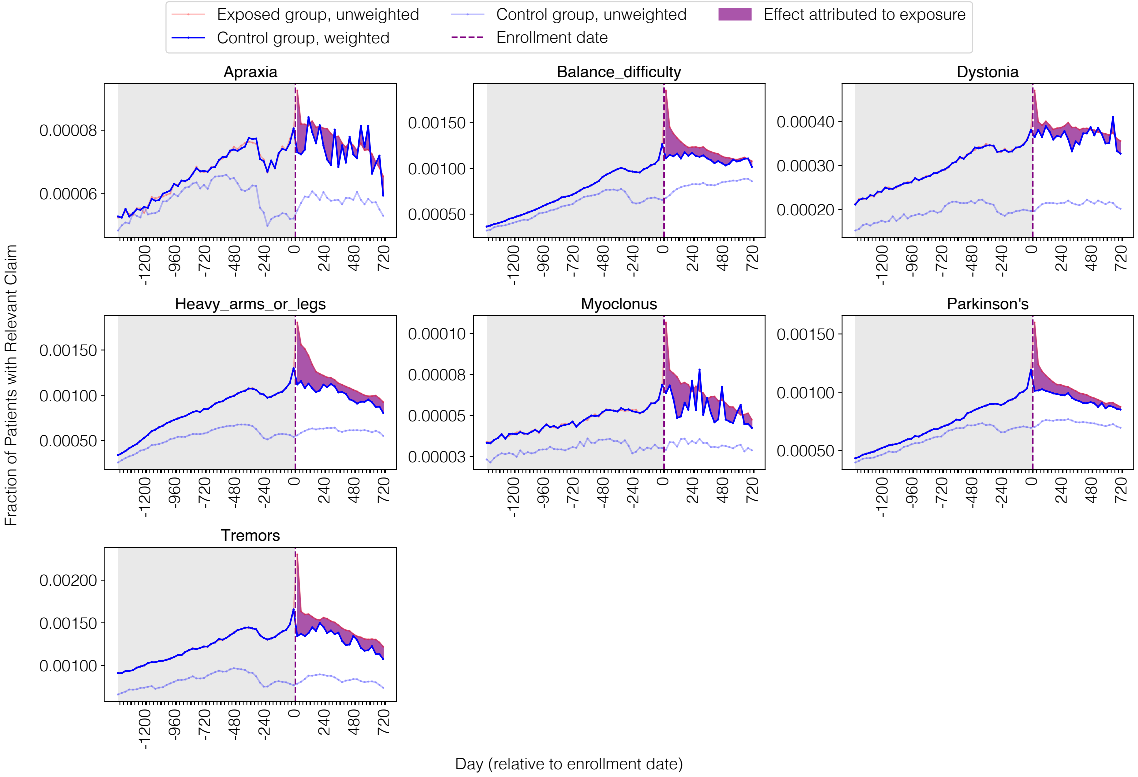


**Supplementary Figure S110:** Temporal trends in Movement prevalence. The fraction of patients with relevant claims over time belonging for each outcome in the **conventional retrospective study with placebo cases** (negative COVID-19 PCR test) experiment.

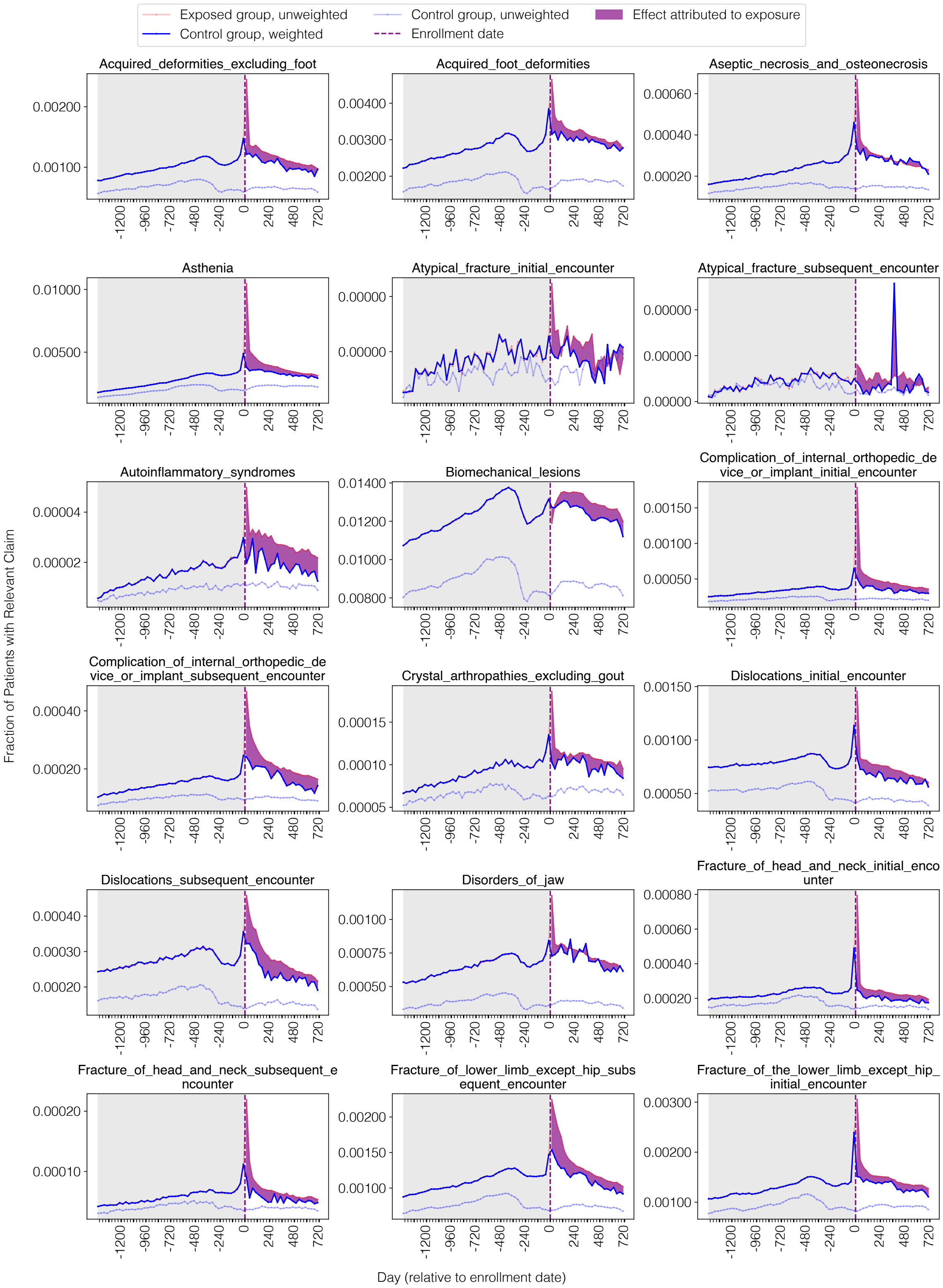


**Supplementary Figure S111:** Temporal trends in Muscles and joints prevalence. The fraction of patients with relevant claims over time belonging for each outcome in the **conventional retrospective study with placebo cases** (negative COVID-19 PCR test) experiment.

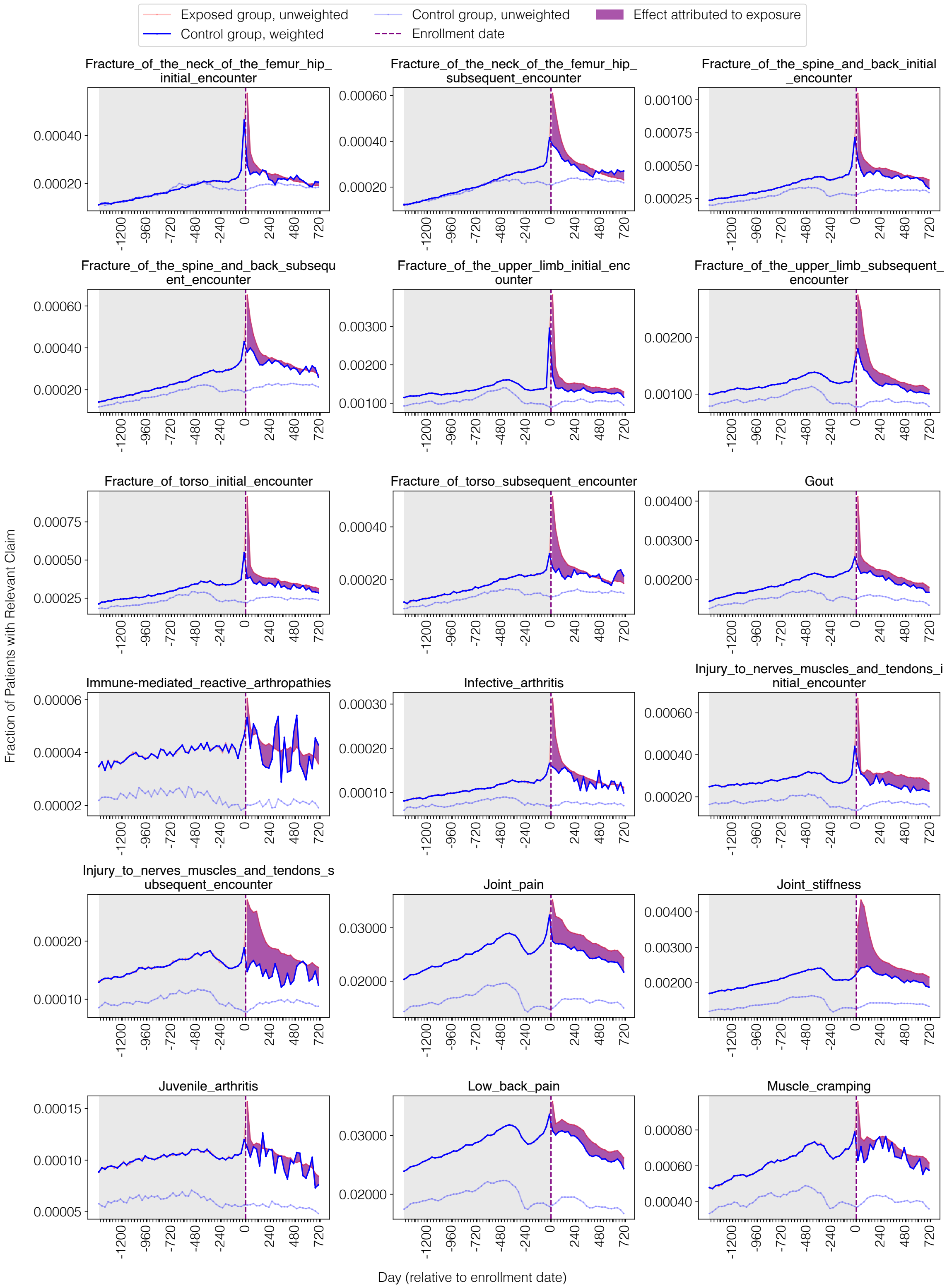


**Supplementary Figure S111:** (cont'd) Temporal trends in Muscles and joints prevalence. The fraction of patients with relevant claims over time belonging for each outcome in the **conventional retrospective study with placebo cases** (negative COVID-19 PCR test) experiment.

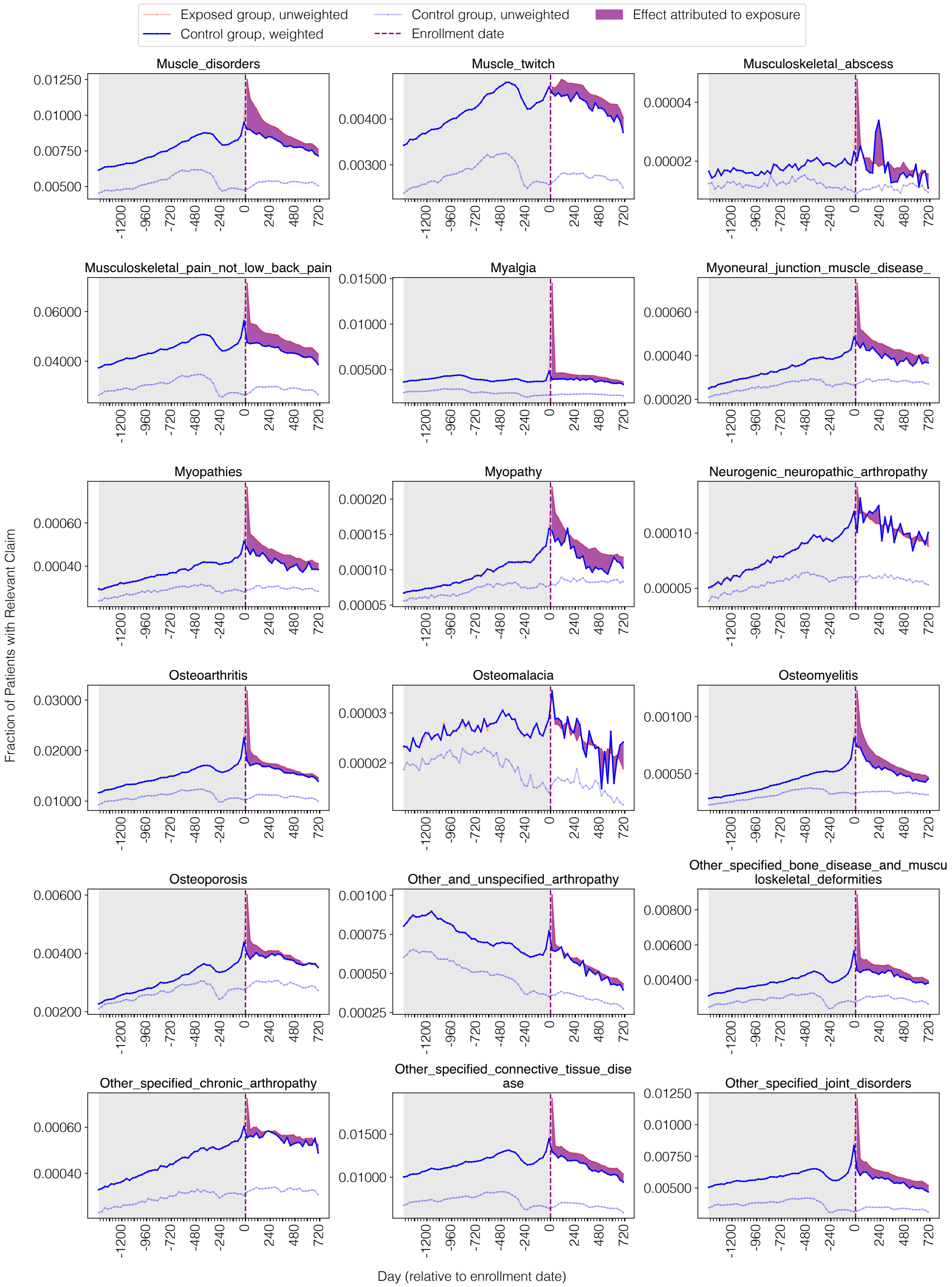


**Supplementary Figure S111:** (cont'd) Temporal trends in Muscles and joints prevalence. The fraction of patients with relevant claims over time belonging for each outcome in the **conventional retrospective study with placebo cases** (negative COVID-19 PCR test) experiment.

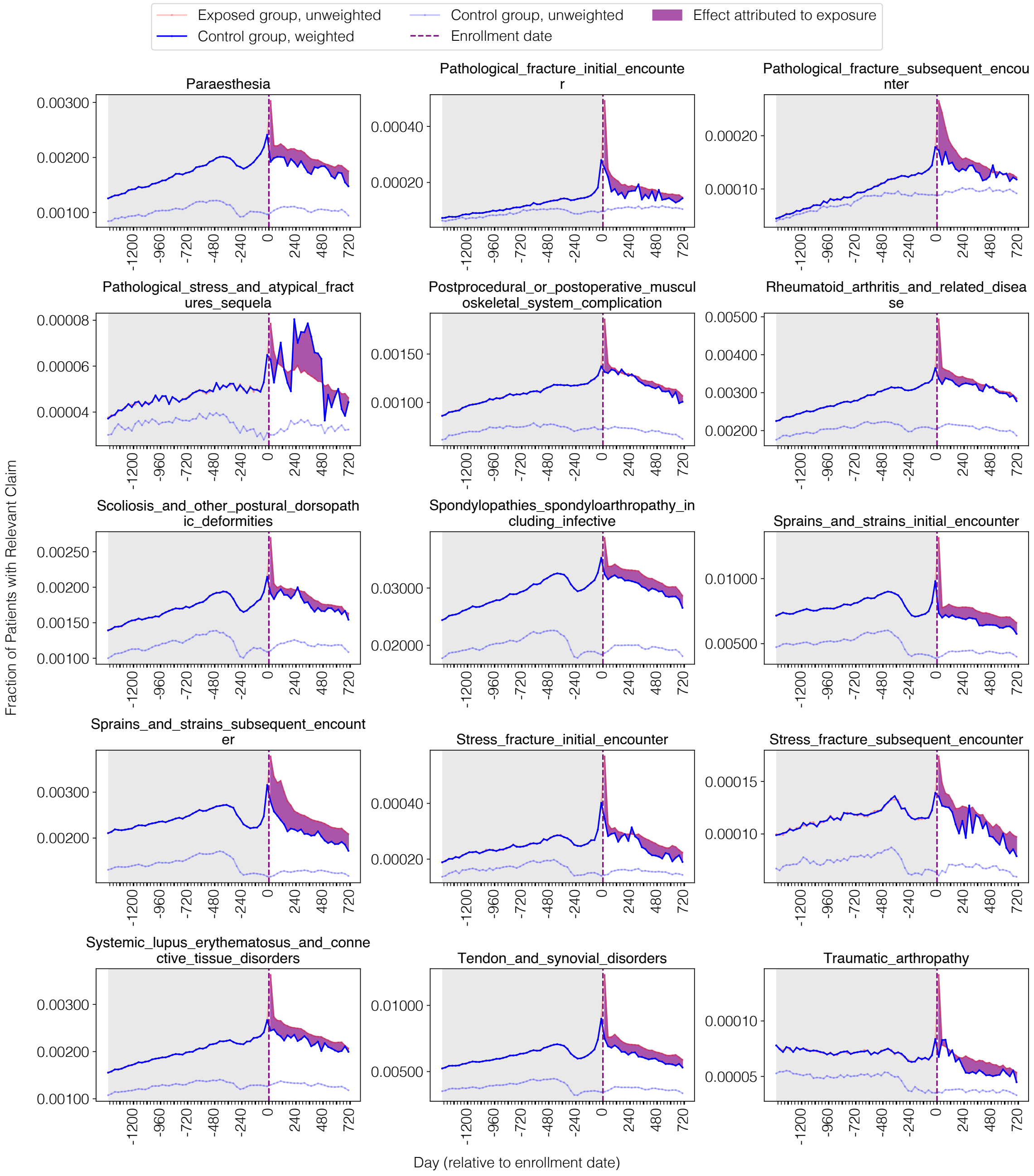


**Supplementary Figure S111:** (cont'd) Temporal trends in Muscles and joints prevalence. The fraction of patients with relevant claims over time belonging for each outcome in the **conventional retrospective study with placebo cases** (negative COVID-19 PCR test) experiment.

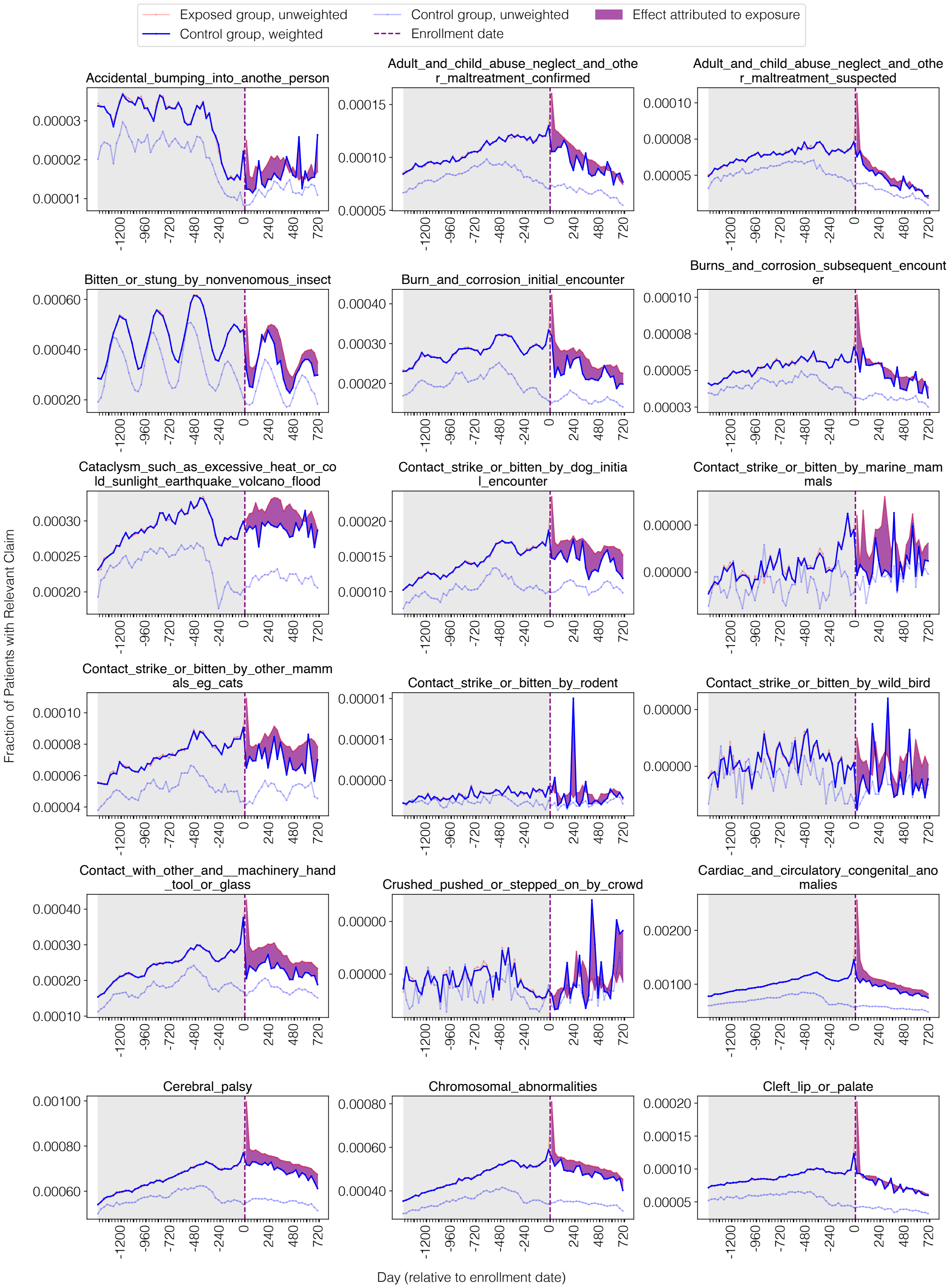


**Supplementary Figure S112:** Temporal trends in Negative prevalence. The fraction of patients with relevant claims over time belonging for each outcome in the **conventional retrospective study with placebo cases** (negative COVID-19 PCR test) experiment.

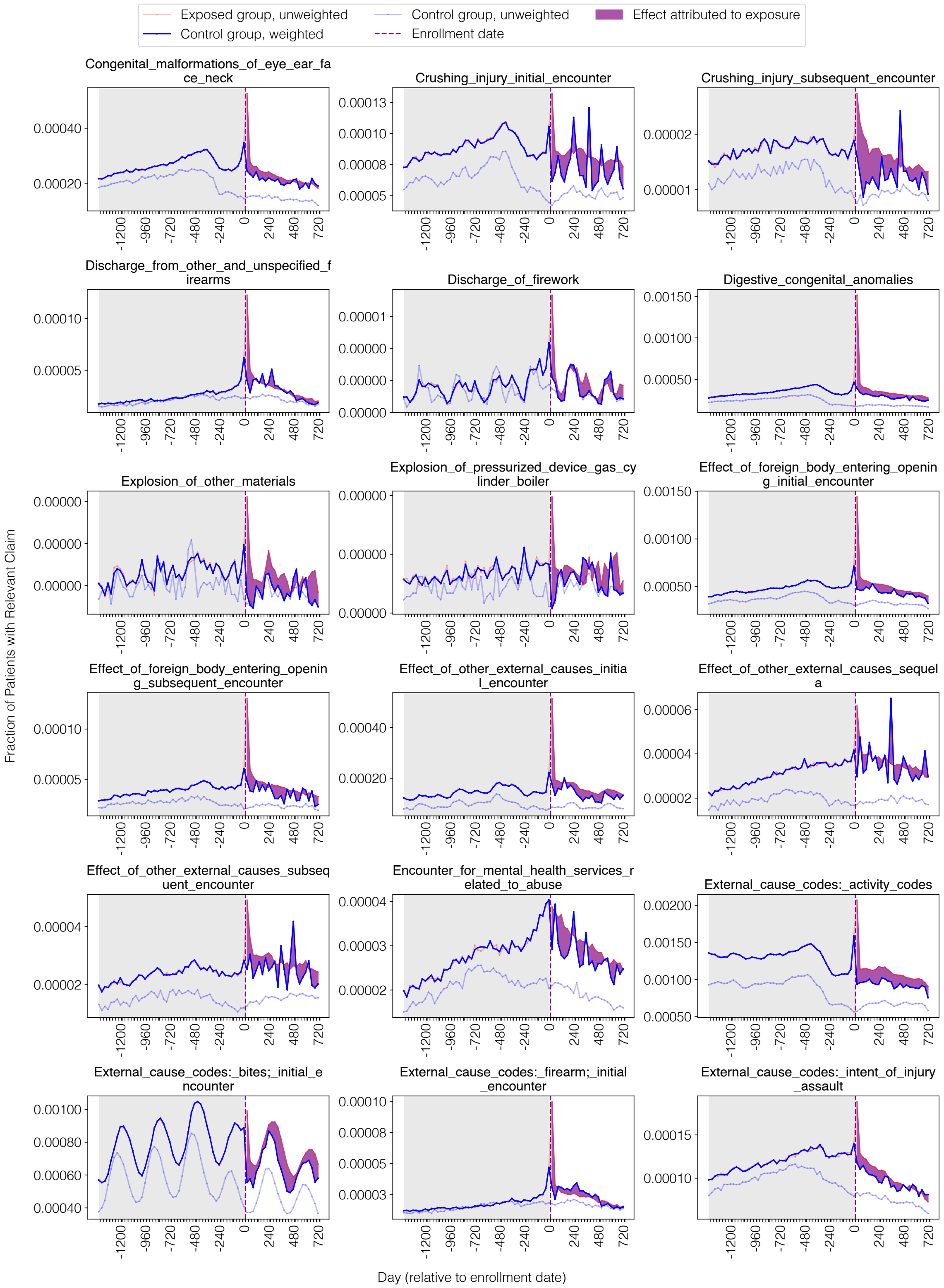


**Supplementary Figure S112:** (cont'd) Temporal trends in Negative prevalence. The fraction of patients with relevant claims over time belonging for each outcome in the **conventional retrospective study with placebo cases** (negative COVID-19 PCR test) experiment.

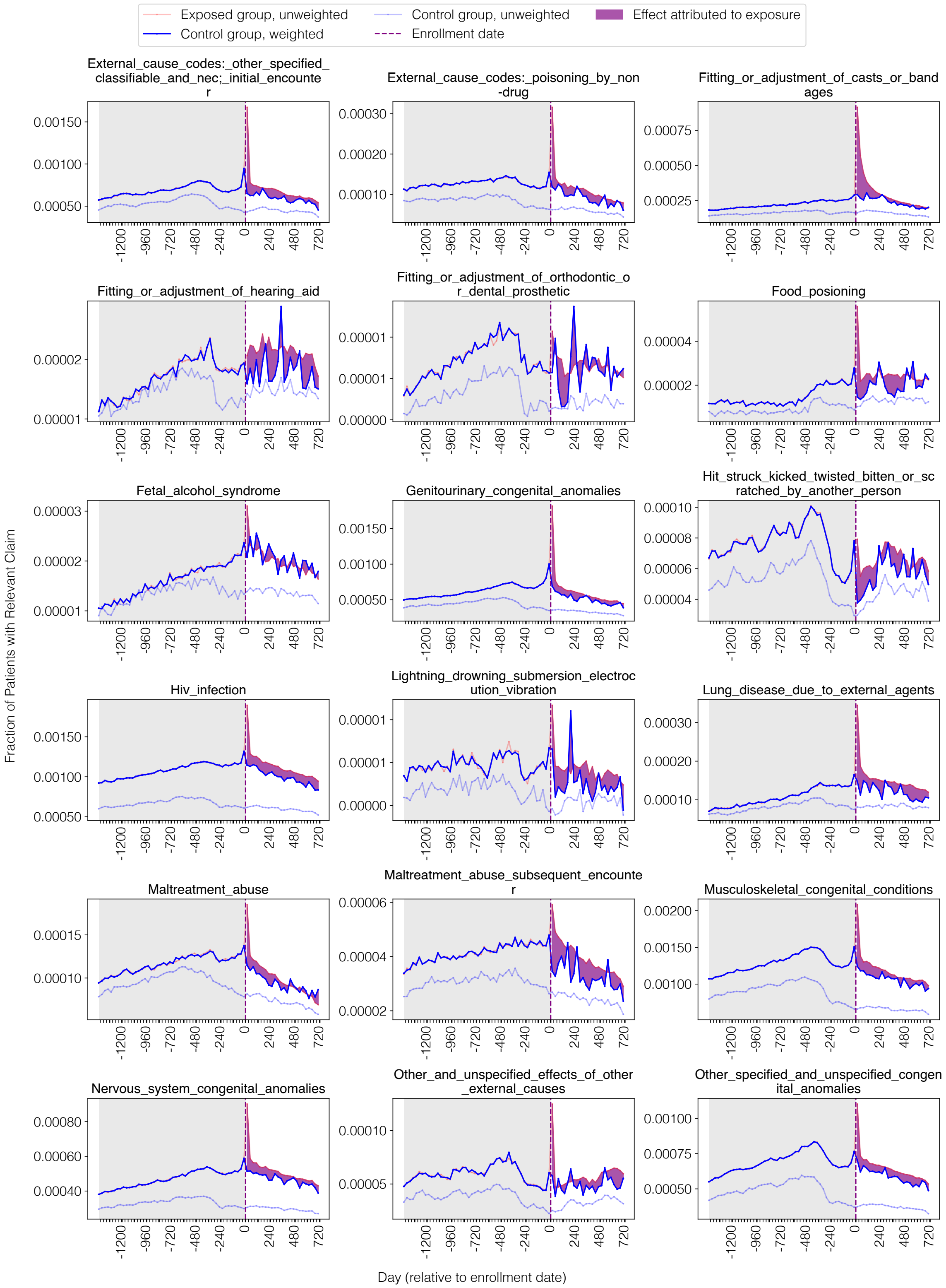


**Supplementary Figure S112:** (cont'd) Temporal trends in Negative prevalence. The fraction of patients with relevant claims over time belonging for each outcome in the **conventional retrospective study with placebo cases** (negative COVID-19 PCR test) experiment.

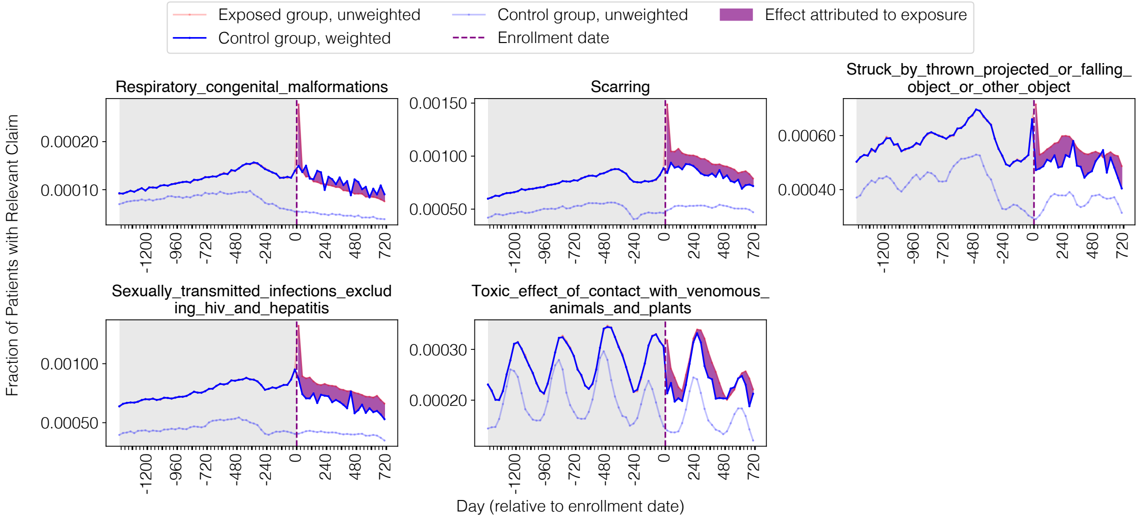


**Supplementary Figure S112:** (cont'd) Temporal trends in Negative prevalence. The fraction of patients with relevant claims over time belonging for each outcome in the **conventional retrospective study with placebo cases** (negative COVID-19 PCR test) experiment.

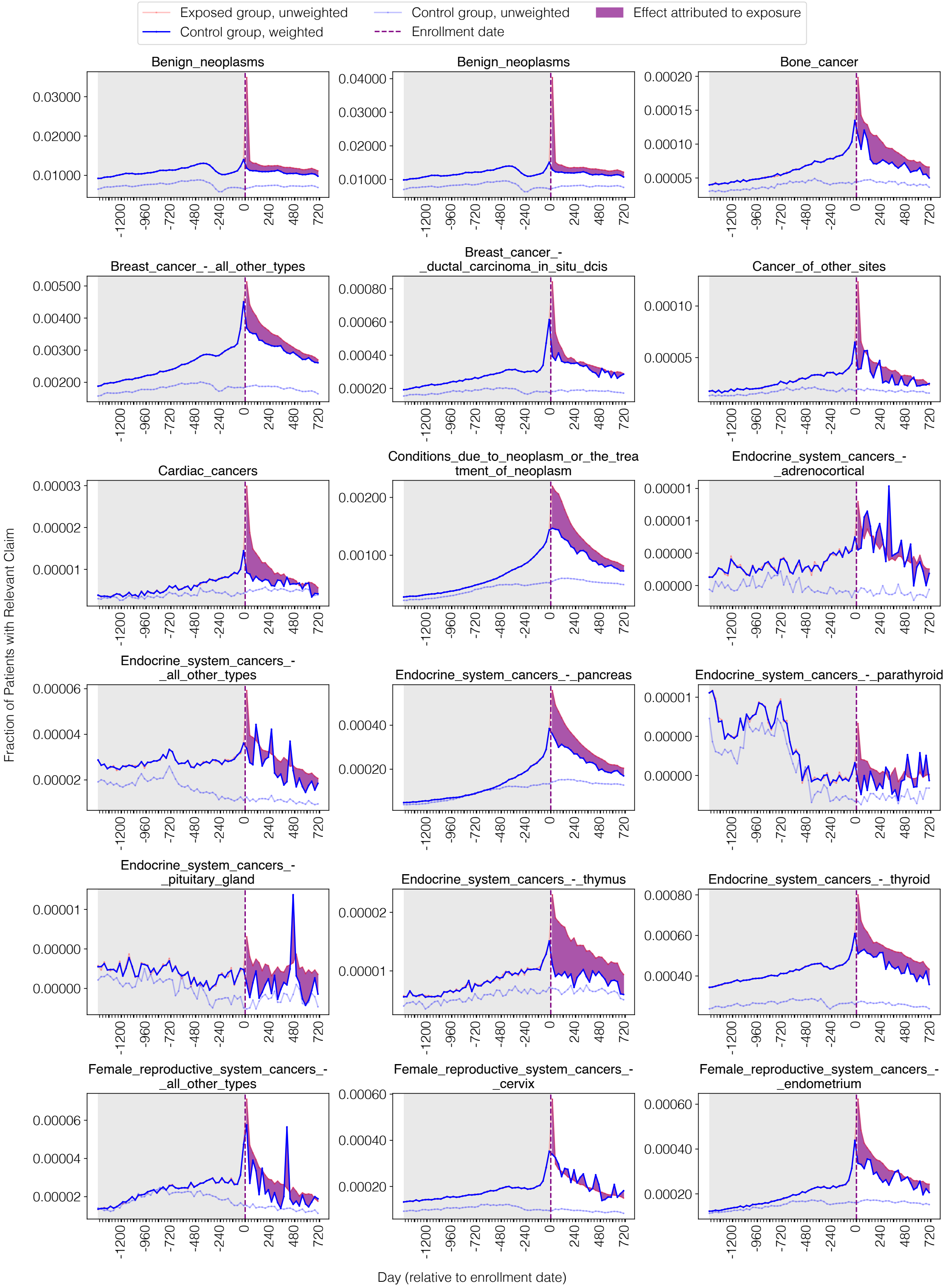


**Supplementary Figure S113:** Temporal trends in Neoplasms prevalence. The fraction of patients with relevant claims over time belonging for each outcome in the **conventional retrospective study with placebo cases** (negative COVID-19 PCR test) experiment.

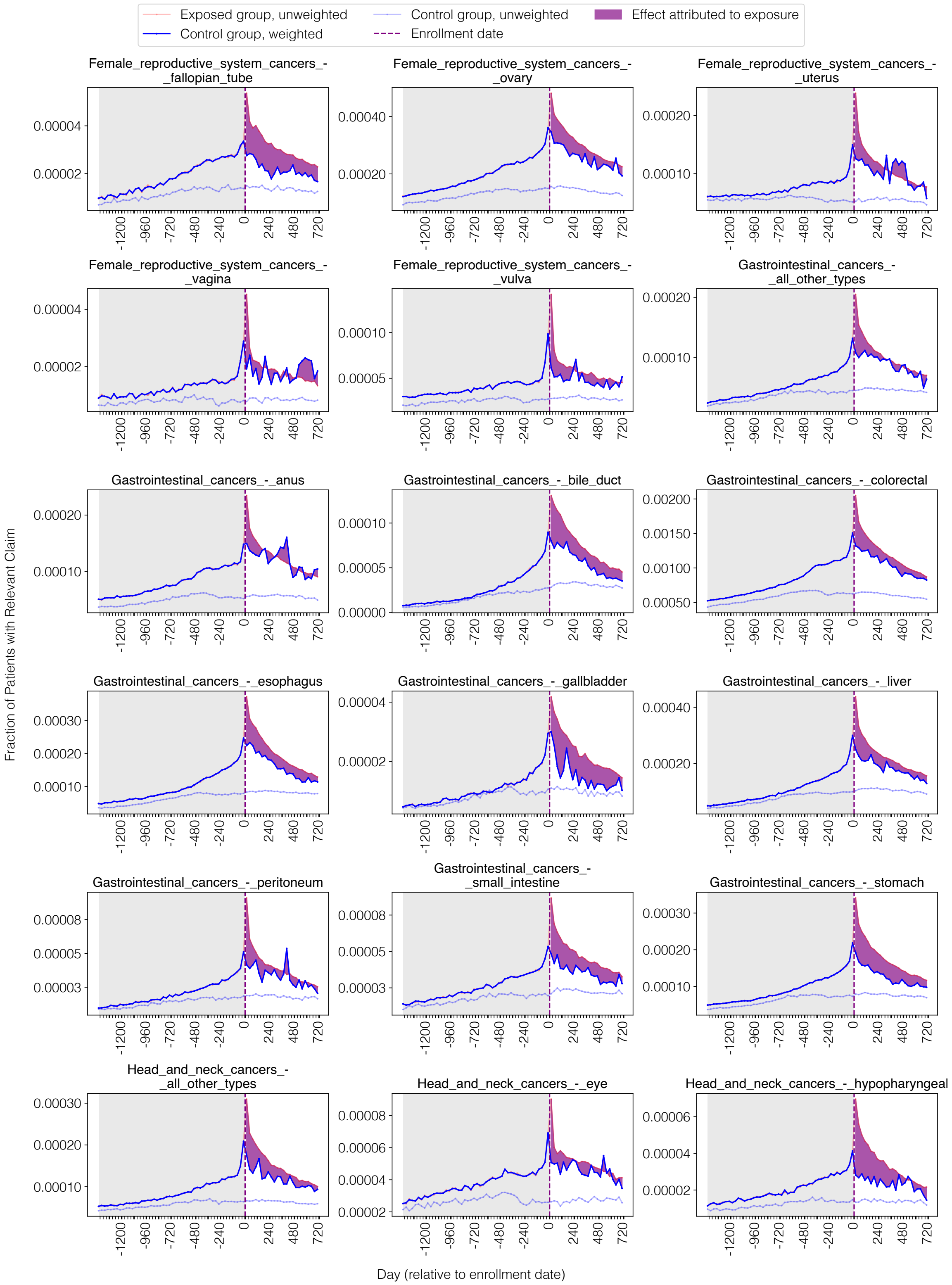


**Supplementary Figure S113:** (cont'd) Temporal trends in Neoplasms prevalence. The fraction of patients with relevant claims over time belonging for each outcome in the **conventional retrospective study with placebo cases** (negative COVID-19 PCR test) experiment.

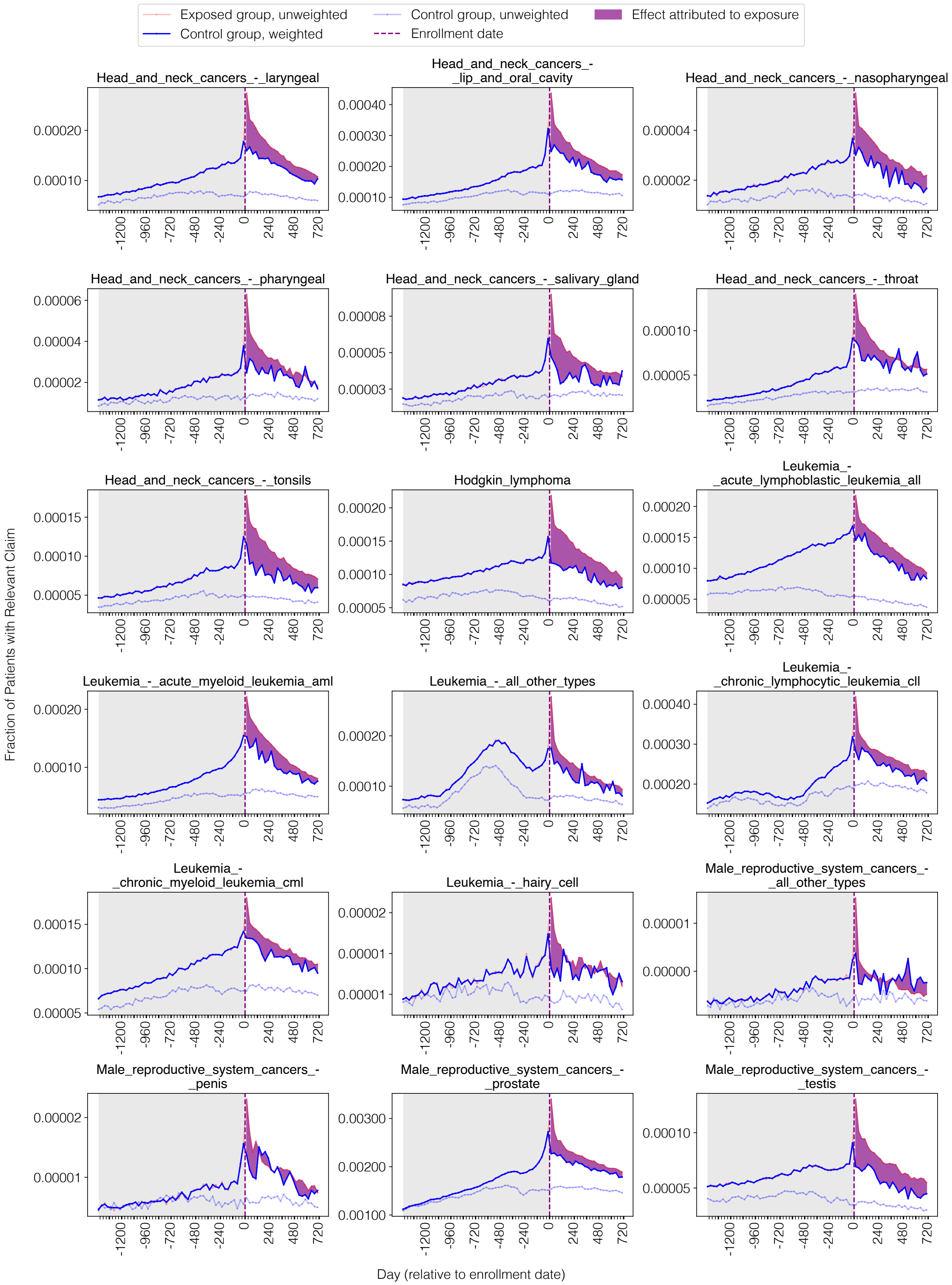


**Supplementary Figure S113:** (cont'd) Temporal trends in Neoplasms prevalence. The fraction of patients with relevant claims over time belonging for each outcome in the **conventional retrospective study with placebo cases** (negative COVID-19 PCR test) experiment.

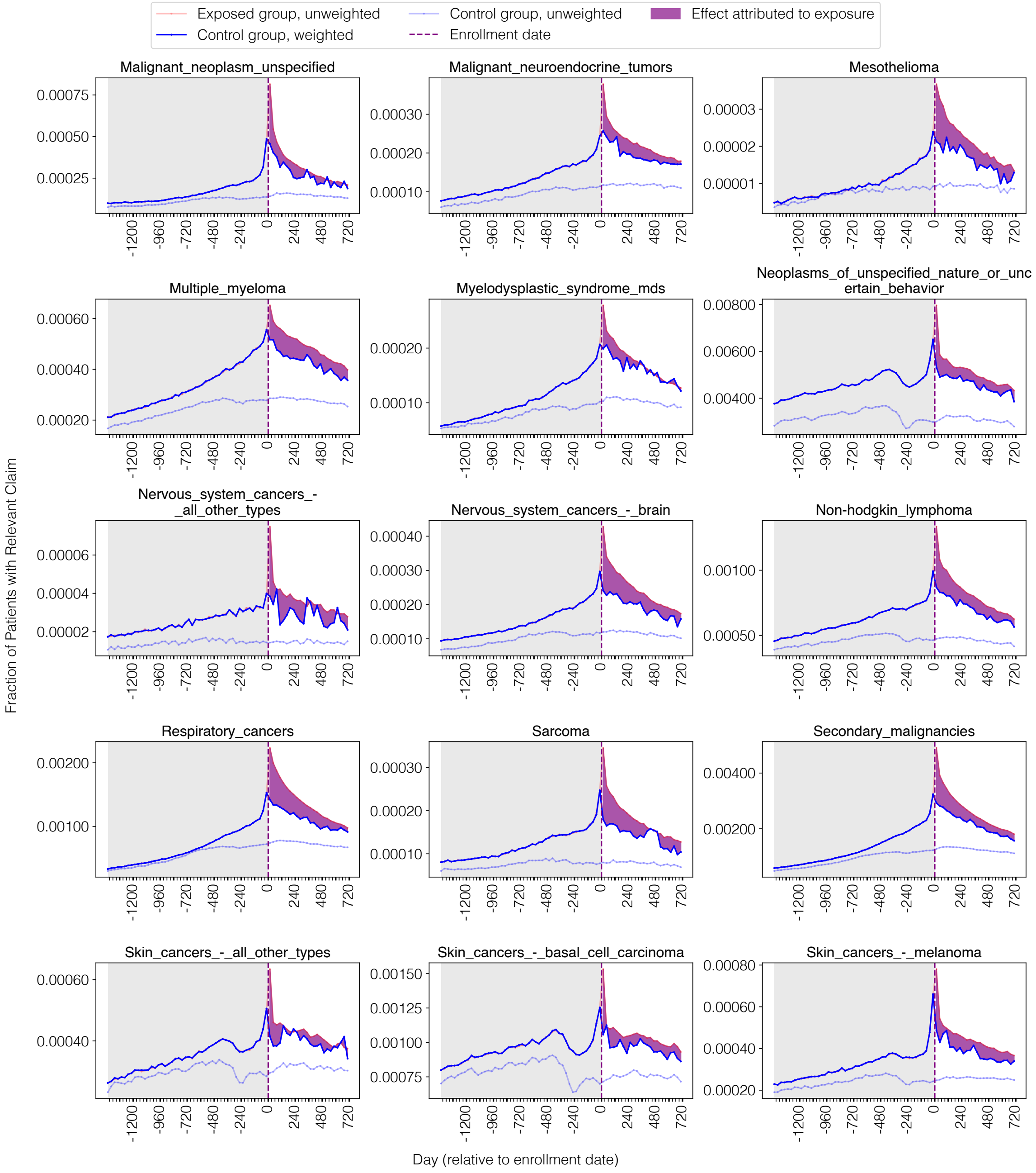


**Supplementary Figure S113:** (cont'd) Temporal trends in Neoplasms prevalence. The fraction of patients with relevant claims over time belonging for each outcome in the **conventional retrospective study with placebo cases** (negative COVID-19 PCR test) experiment.

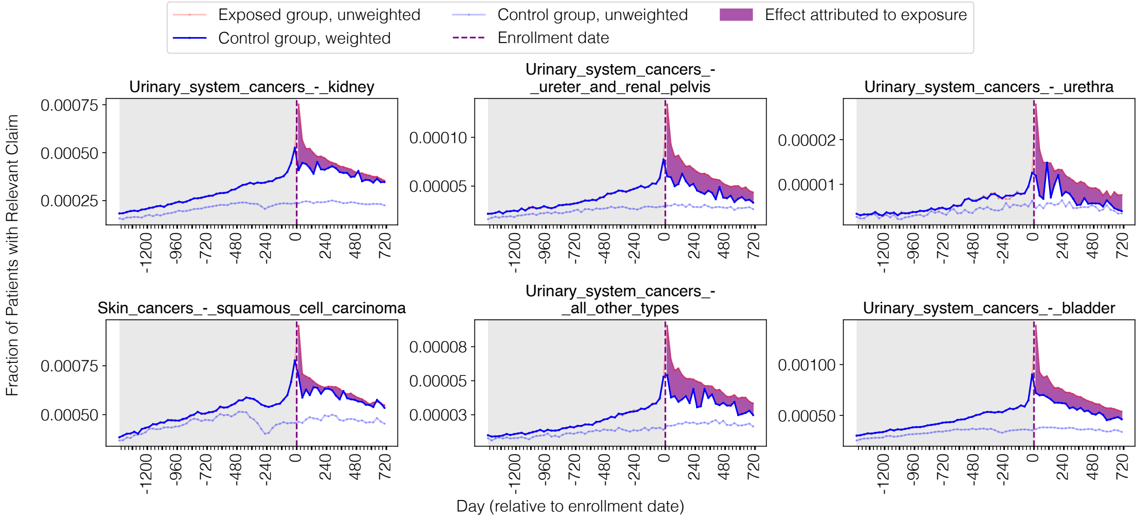


**Supplementary Figure S113:** (cont'd) Temporal trends in Neoplasms prevalence. The fraction of patients with relevant claims over time belonging for each outcome in the **conventional retrospective study with placebo cases** (negative COVID-19 PCR test) experiment.

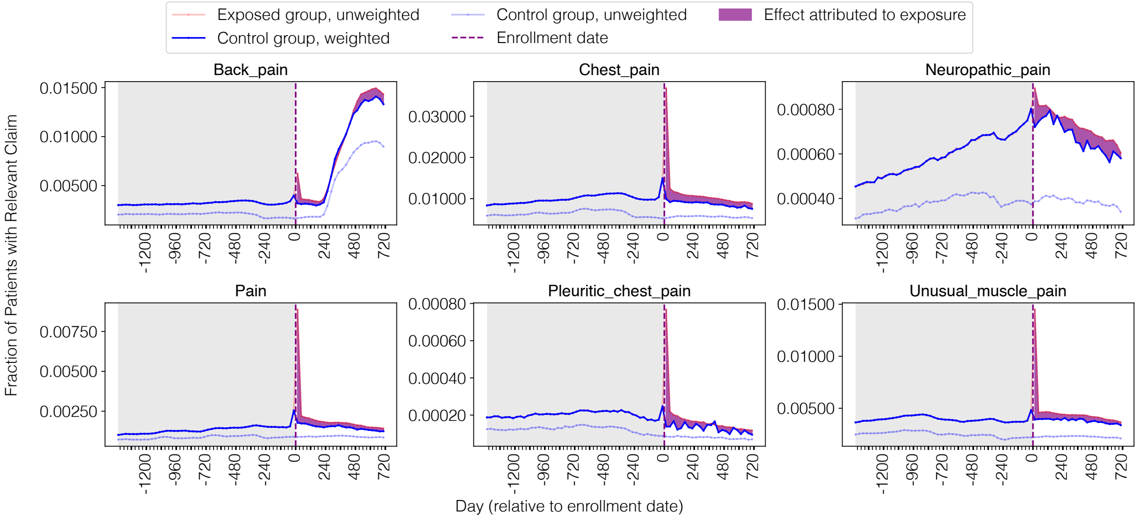


**Supplementary Figure S114:** Temporal trends in Pain prevalence. The fraction of patients with relevant claims over time belonging for each outcome in the **conventional retrospective study with placebo cases** (negative COVID-19 PCR test) experiment.

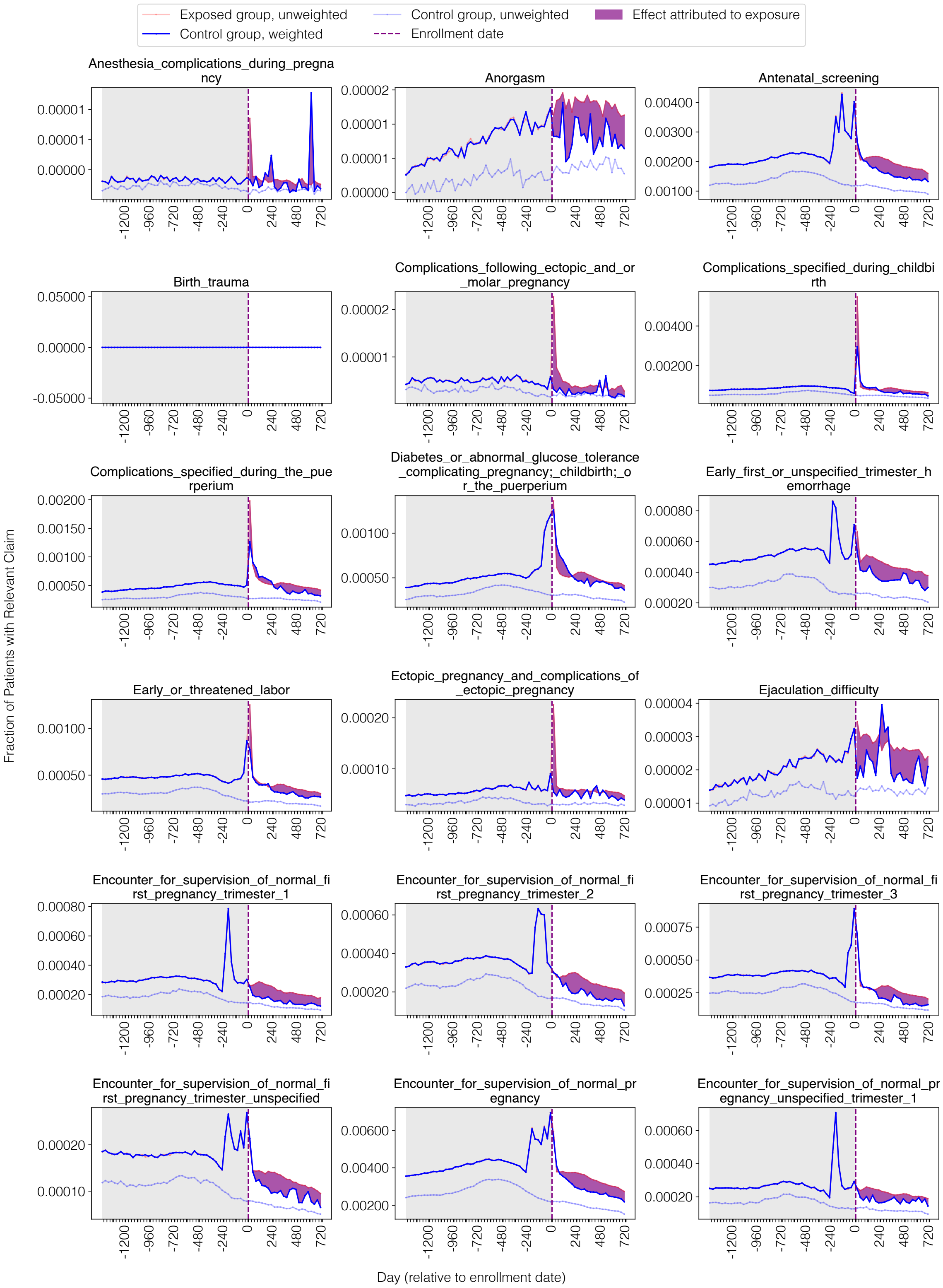


**Supplementary Figure S115:** Temporal trends in Reproductive health prevalence. The fraction of patients with relevant claims over time belonging for each outcome in the **conventional retrospective study with placebo cases** (negative COVID-19 PCR test) experiment.

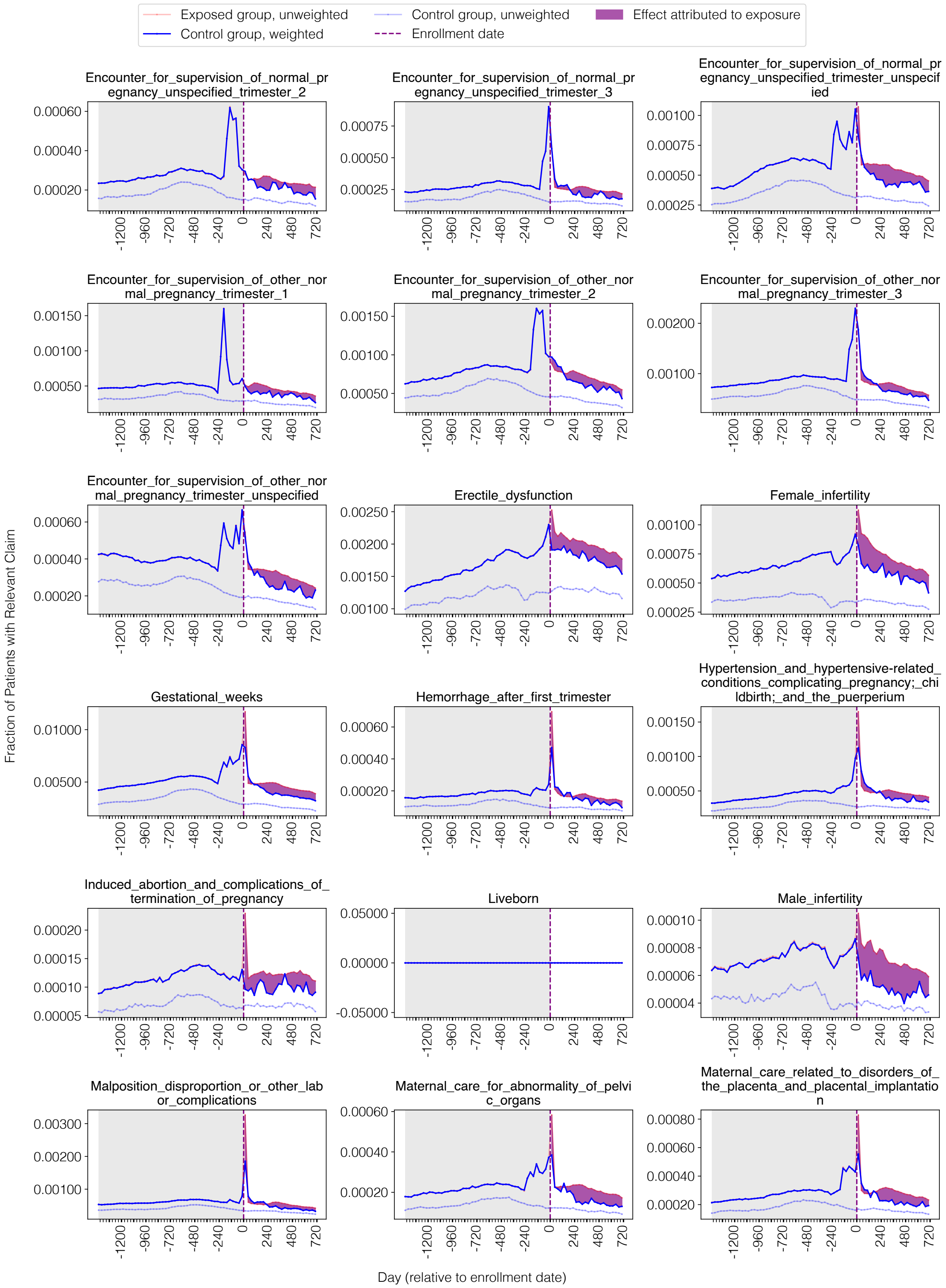


**Supplementary Figure S115:** (cont'd) Temporal trends in Reproductive health prevalence. The fraction of patients with relevant claims over time belonging for each outcome in the **conventional retrospective study with placebo cases** (negative COVID-19 PCR test) experiment.

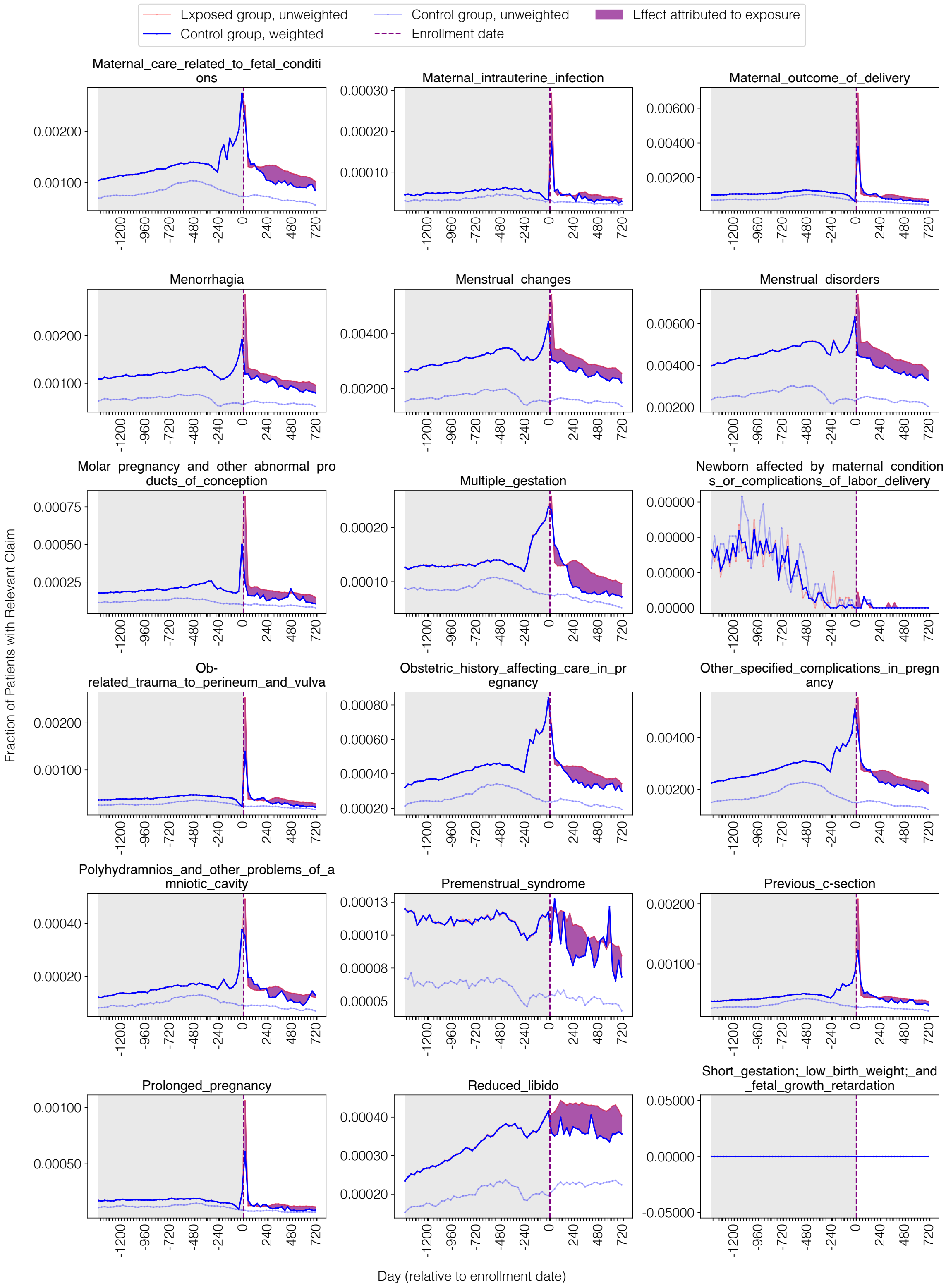


**Supplementary Figure S115:** (cont'd) Temporal trends in Reproductive health prevalence. The fraction of patients with relevant claims over time belonging for each outcome in the **conventional retrospective study with placebo cases** (negative COVID-19 PCR test) experiment.

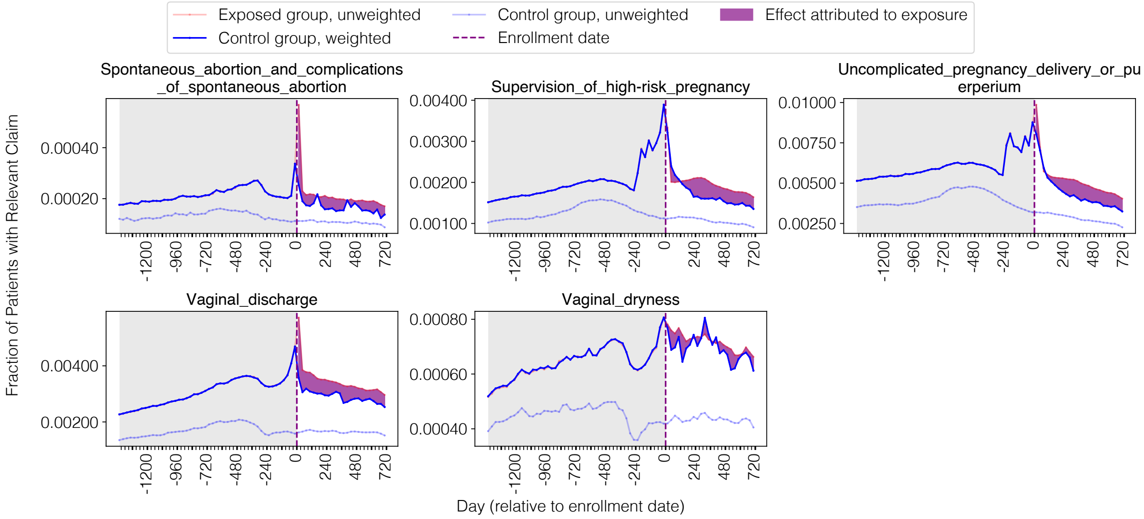


**Supplementary Figure S115:** (cont'd) Temporal trends in Reproductive health prevalence. The fraction of patients with relevant claims over time belonging for each outcome in the **conventional retrospective study with placebo cases** (negative COVID-19 PCR test) experiment.

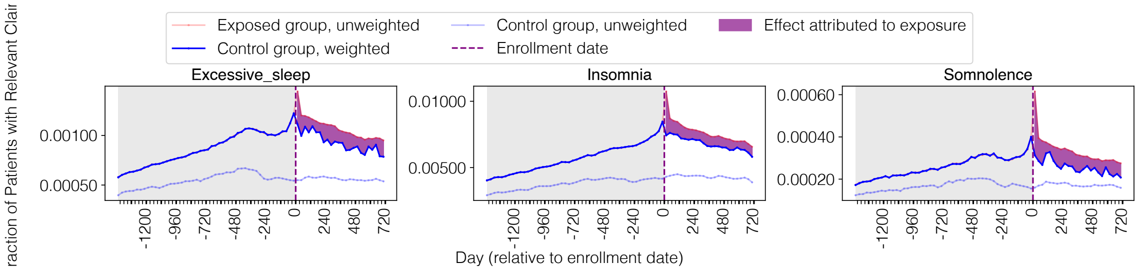


**Supplementary Figure S116:** Temporal trends in Sleep prevalence. The fraction of patients with relevant claims over time belonging for each outcome in the **conventional retrospective study with placebo cases** (negative COVID-19 PCR test) experiment.

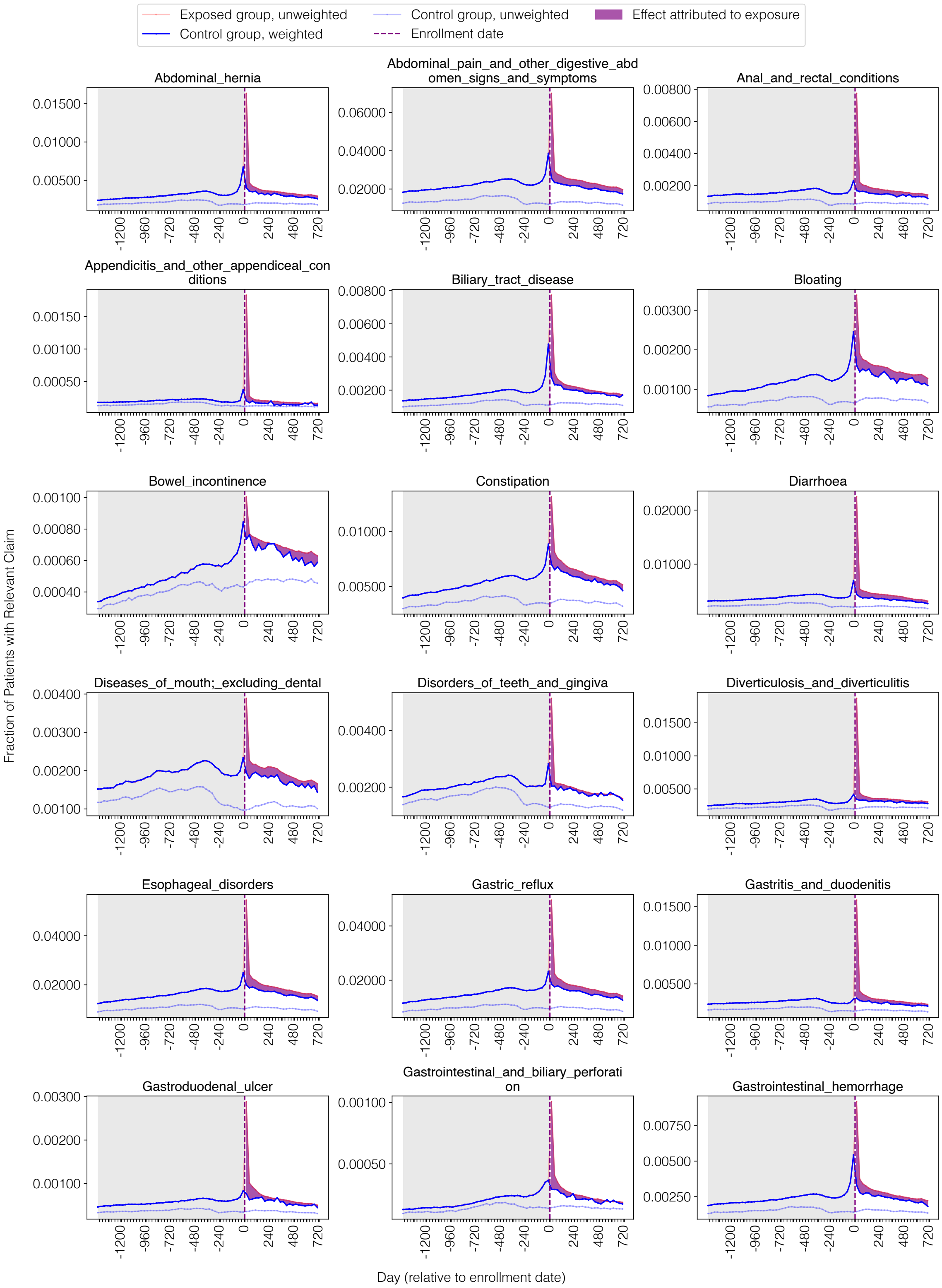


**Supplementary Figure S117:** Temporal trends in Stomach and digestion prevalence. The fraction of patients with relevant claims over time belonging for each outcome in the **conventional retrospective study with placebo cases** (negative COVID-19 PCR test) experiment.

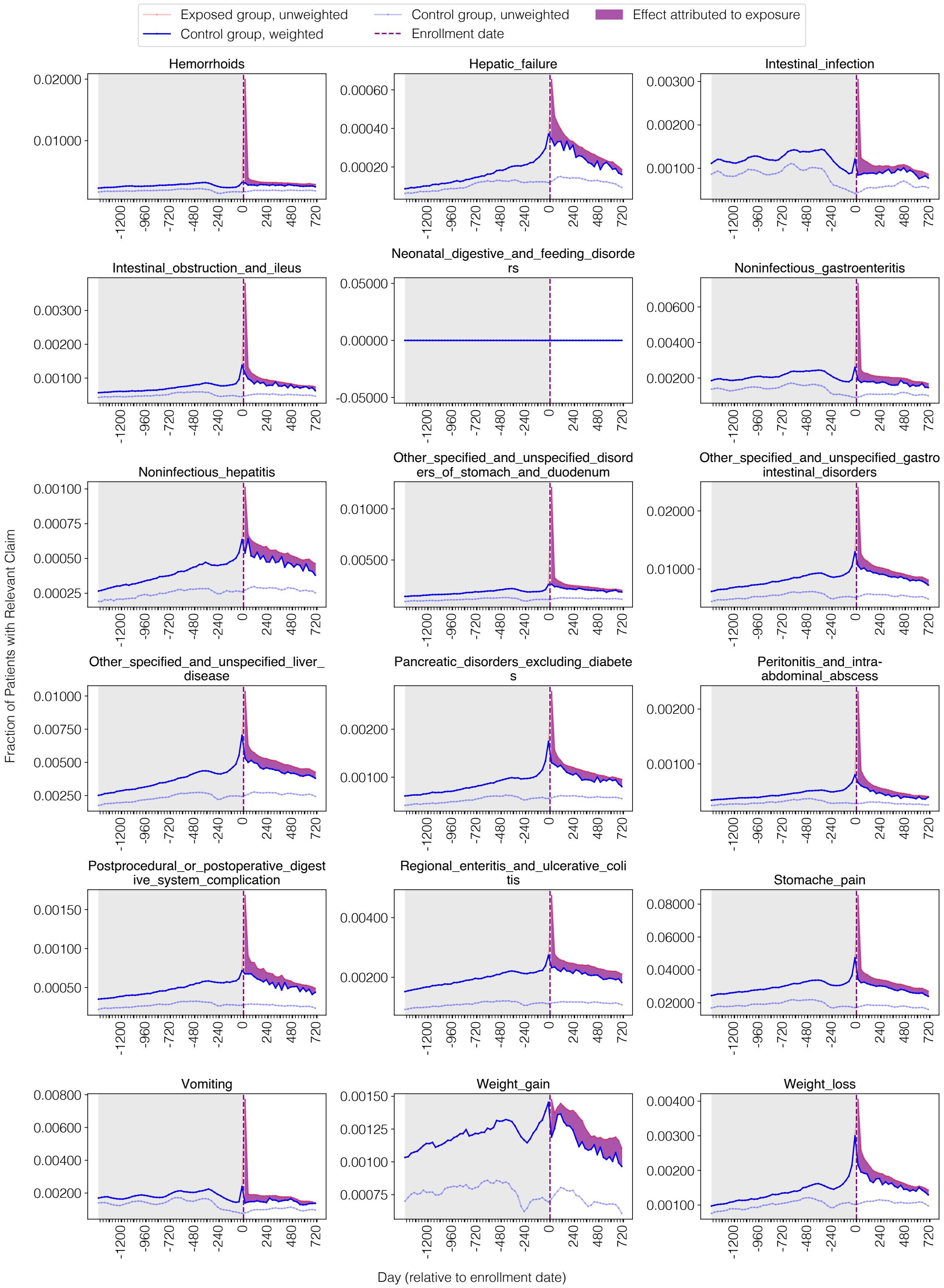


**Supplementary Figure S117:** (cont'd) Temporal trends in Stomach and digestion prevalence. The fraction of patients with relevant claims over time belonging for each outcome in the **conventional retrospective study with placebo cases** (negative COVID-19 PCR test) experiment.

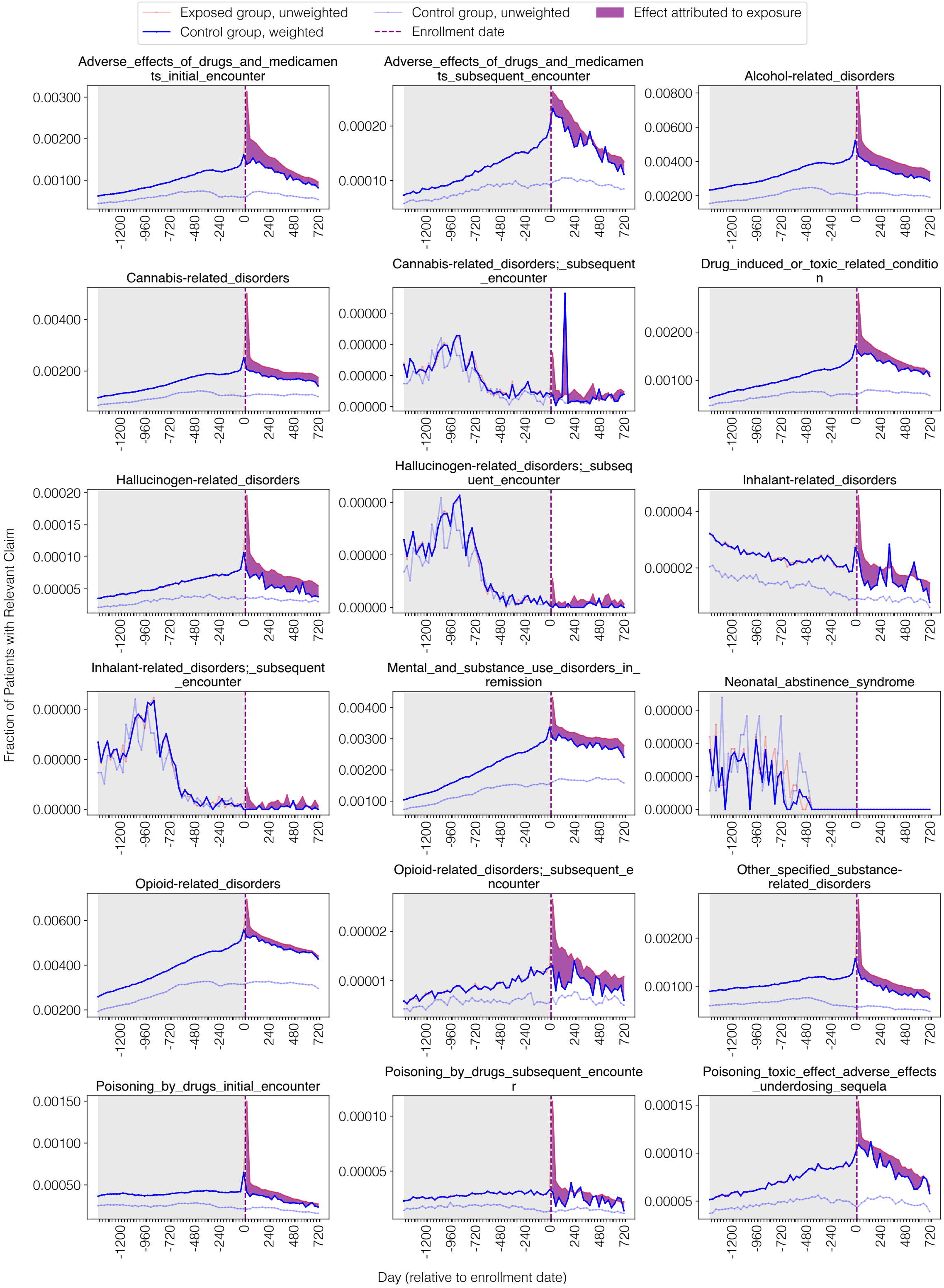


**Supplementary Figure S118:** Temporal trends in Substance use prevalence. The fraction of patients with relevant claims over time belonging for each outcome in the **conventional retrospective study with placebo cases** (negative COVID-19 PCR test) experiment.

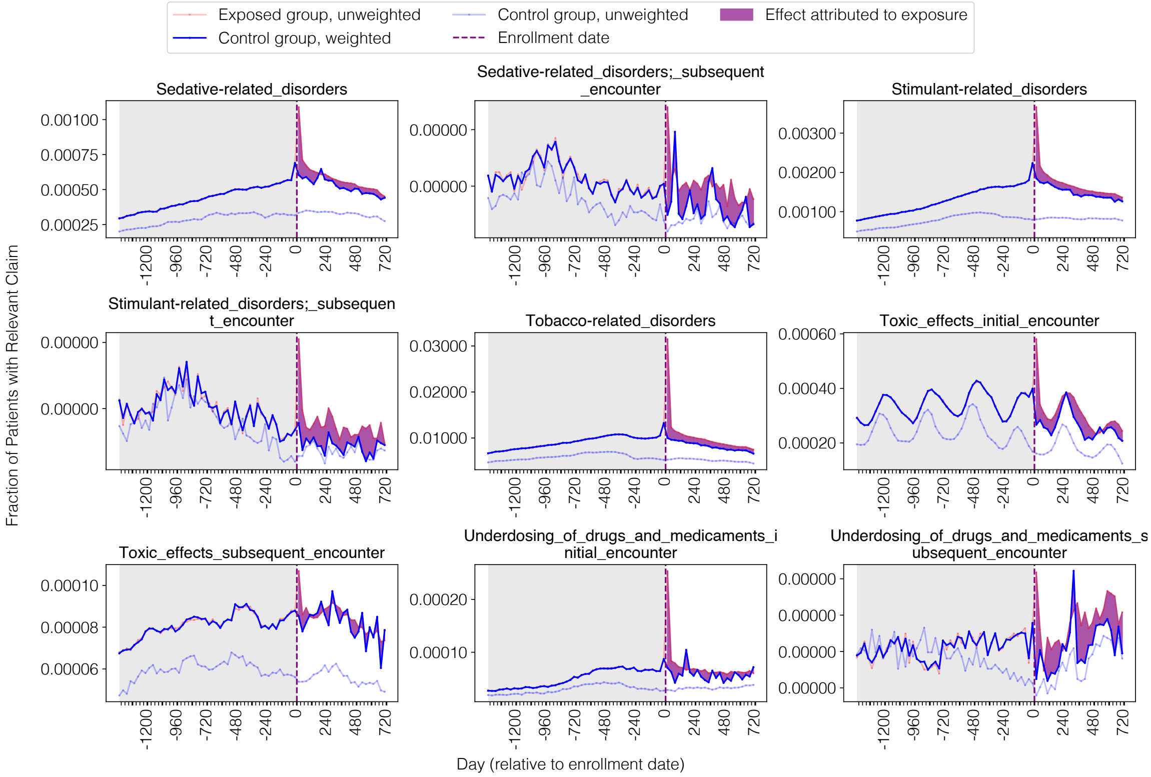


**Supplementary Figure S118:** (cont'd) Temporal trends in Substance use prevalence. The fraction of patients with relevant claims over time belonging for each outcome in the **conventional retrospective study with placebo cases** (negative COVID-19 PCR test) experiment.

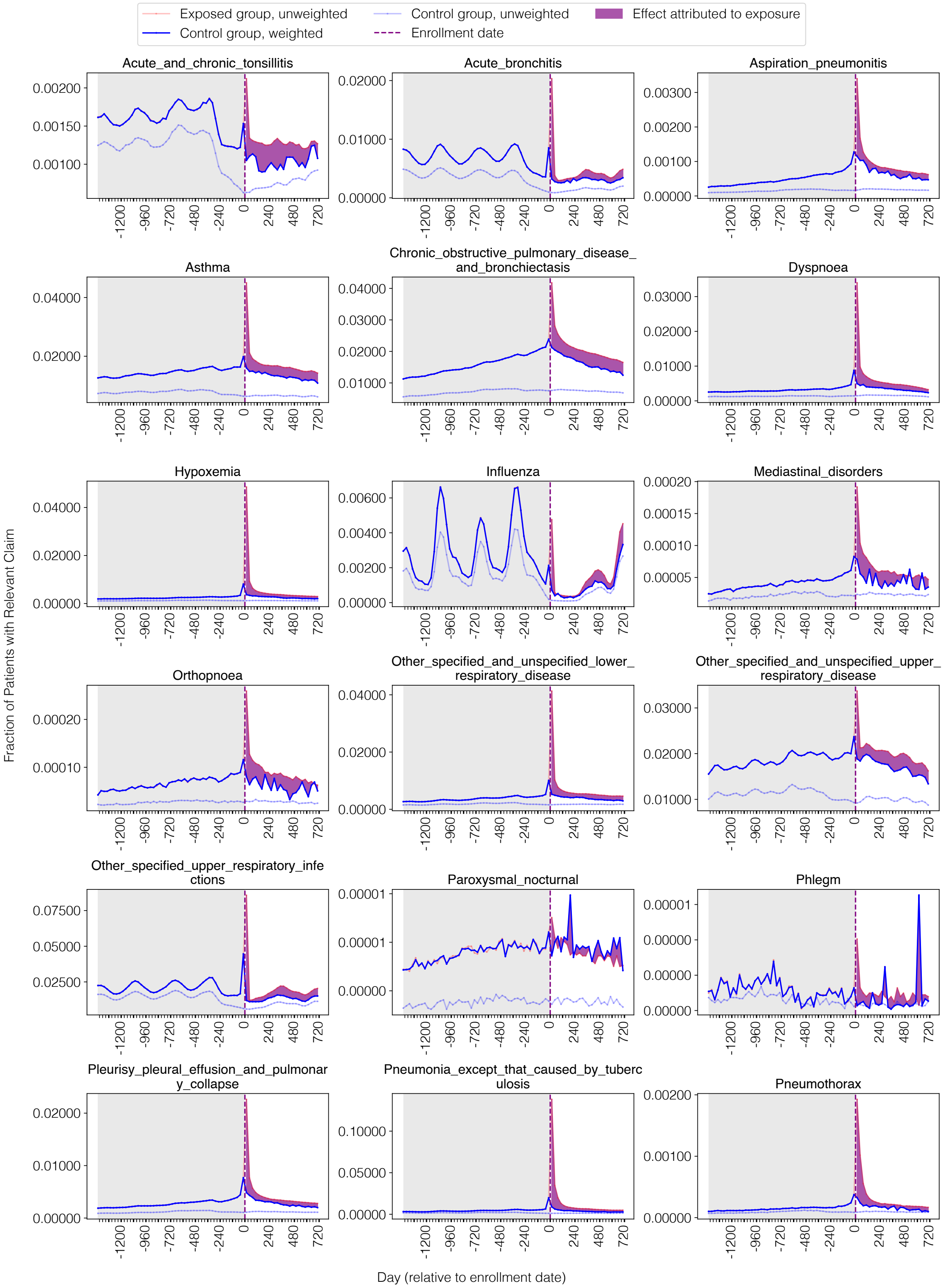


**Supplementary Figure S119:** Temporal trends in Breathing prevalence. The fraction of patients with relevant claims over time belonging for each outcome in the **conventional retrospective** study experiment.

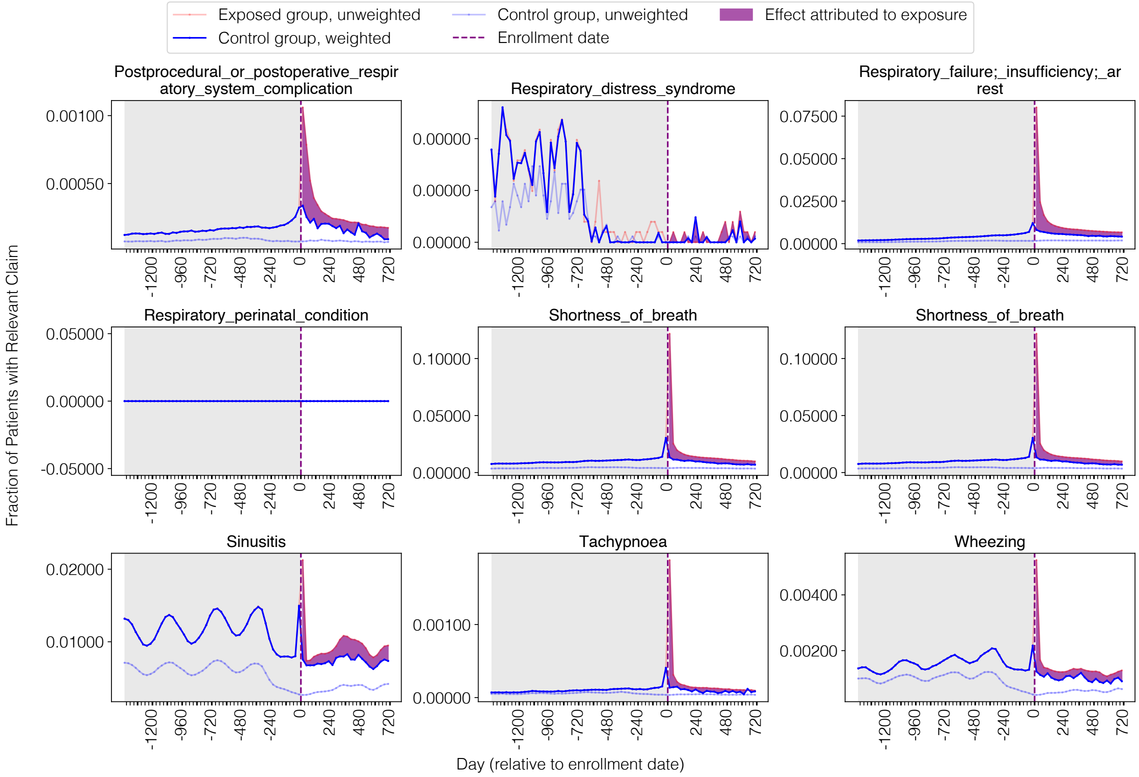


**Supplementary Figure S119:** (cont'd) Temporal trends in Breathing prevalence. The fraction of patients with relevant claims over time belonging for each outcome in the **conventional retrospective** study experiment.

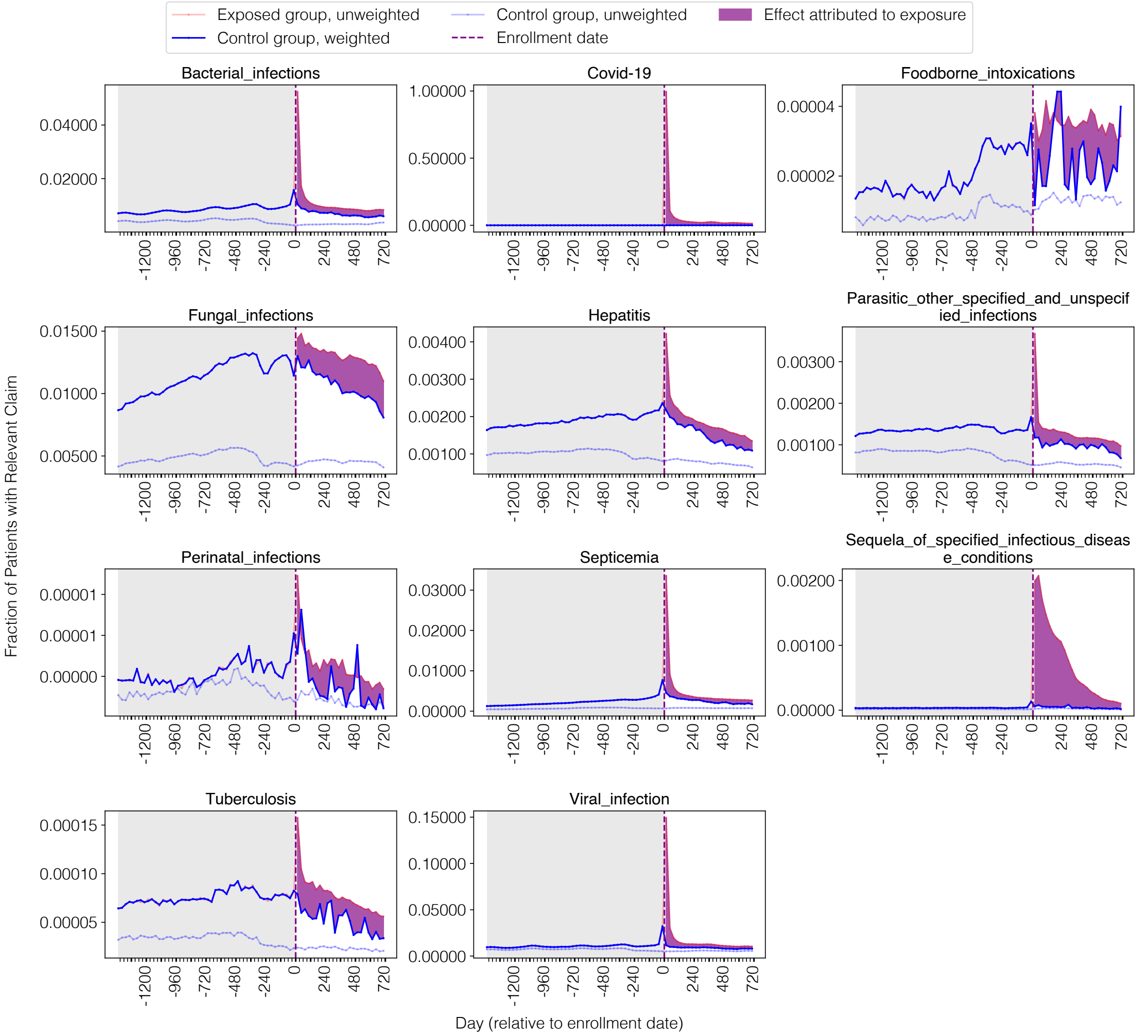


**Supplementary Figure S120:** Temporal trends in Certain Infectious and Parasitic Diseases prevalence. The fraction of patients with relevant claims over time belonging for each outcome in the **conventional retrospective** study experiment.

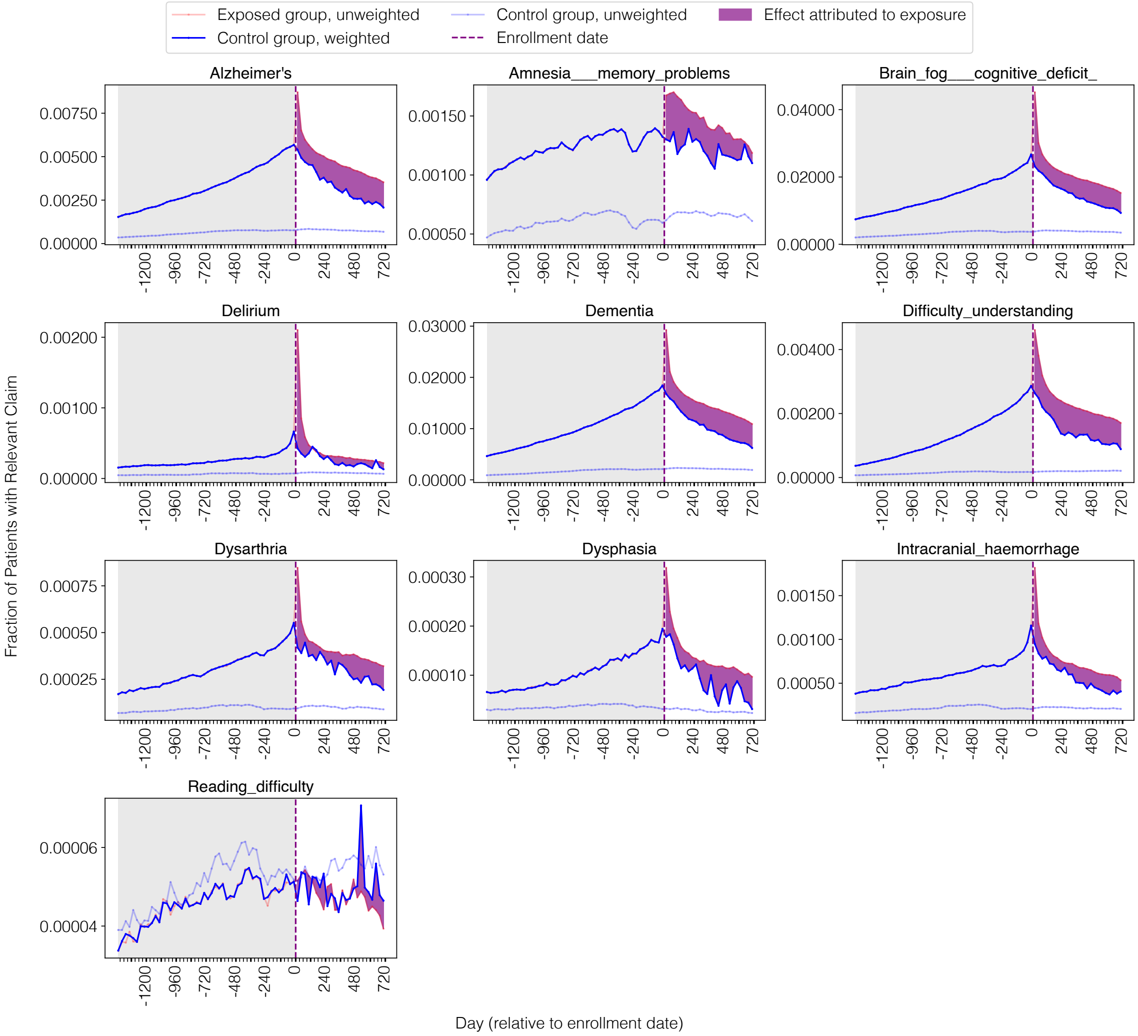


**Supplementary Figure S121:** Temporal trends in Cognitive health prevalence. The fraction of patients with relevant claims over time belonging for each outcome in the **conventional retrospective** study experiment.

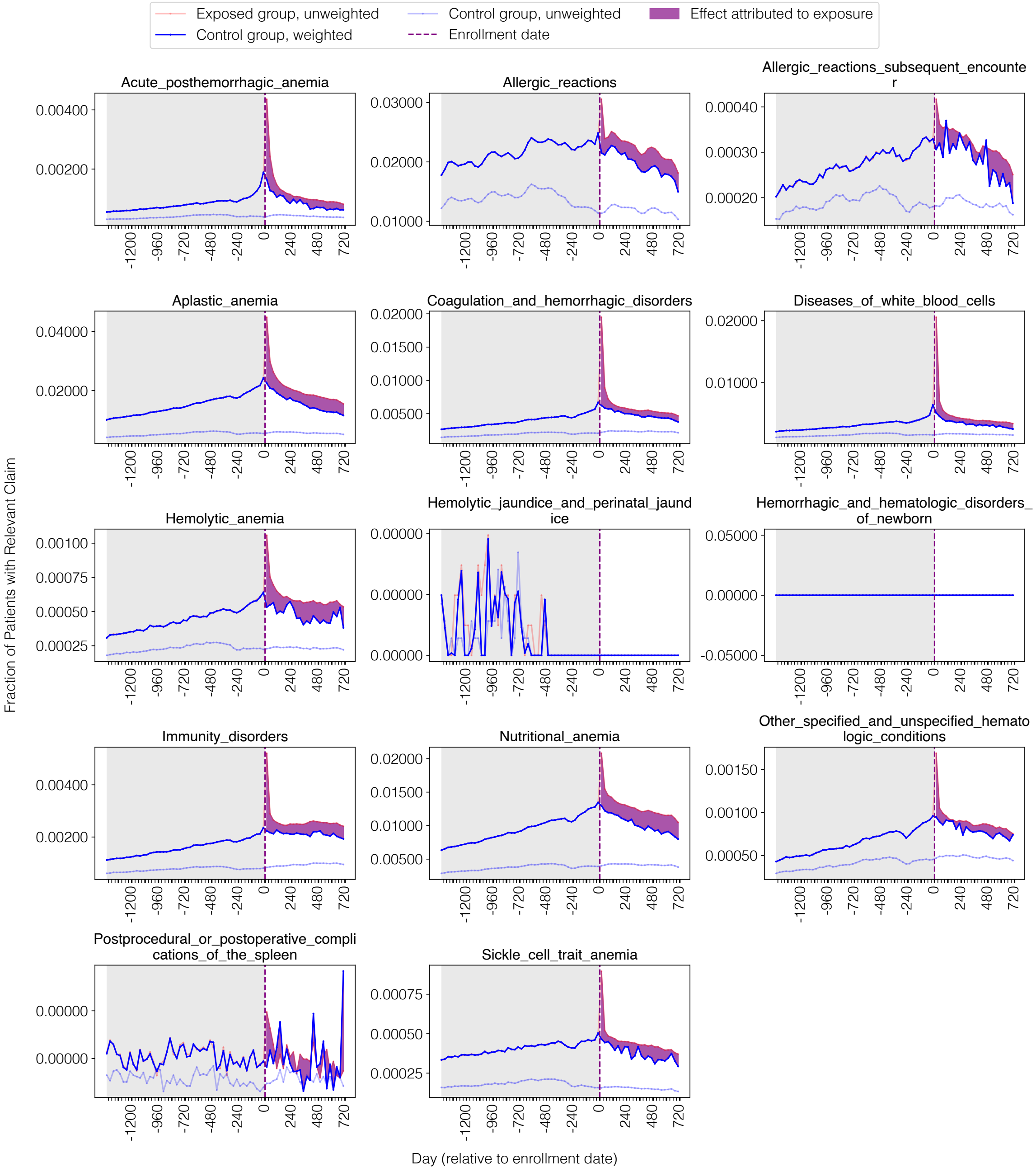


**Supplementary Figure S122:** Temporal trends in Diseases of the Blood and Blood Forming Organs and Certain Disorders Involving the Immune Mechanism prevalence. The fraction of patients with relevant claims over time belonging for each outcome in the **conventional retrospective** study experiment.

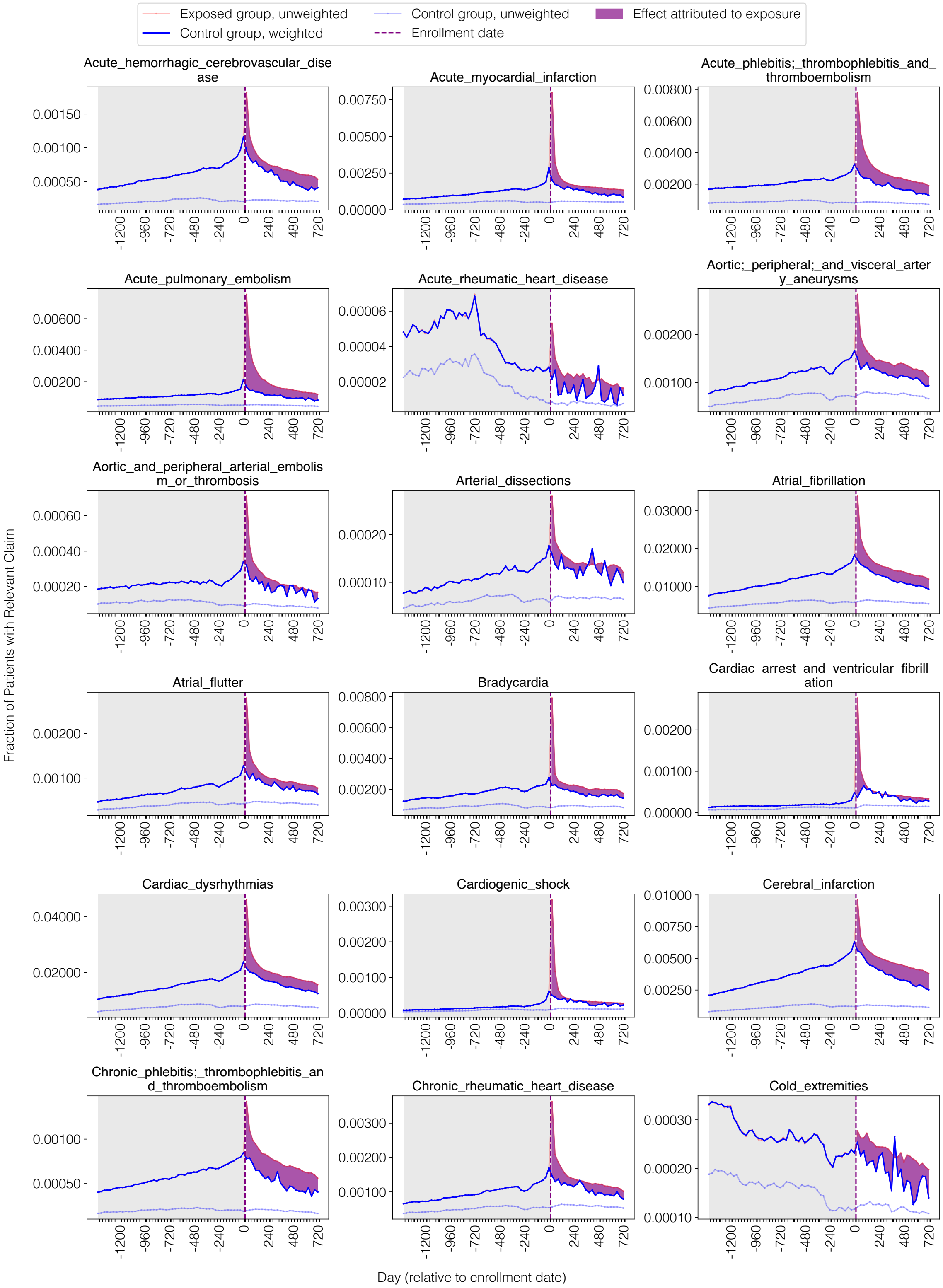


**Supplementary Figure S123:** Temporal trends in Diseases of the Circulatory System prevalence. The fraction of patients with relevant claims over time belonging for each outcome in the **conventional retrospective** study experiment.

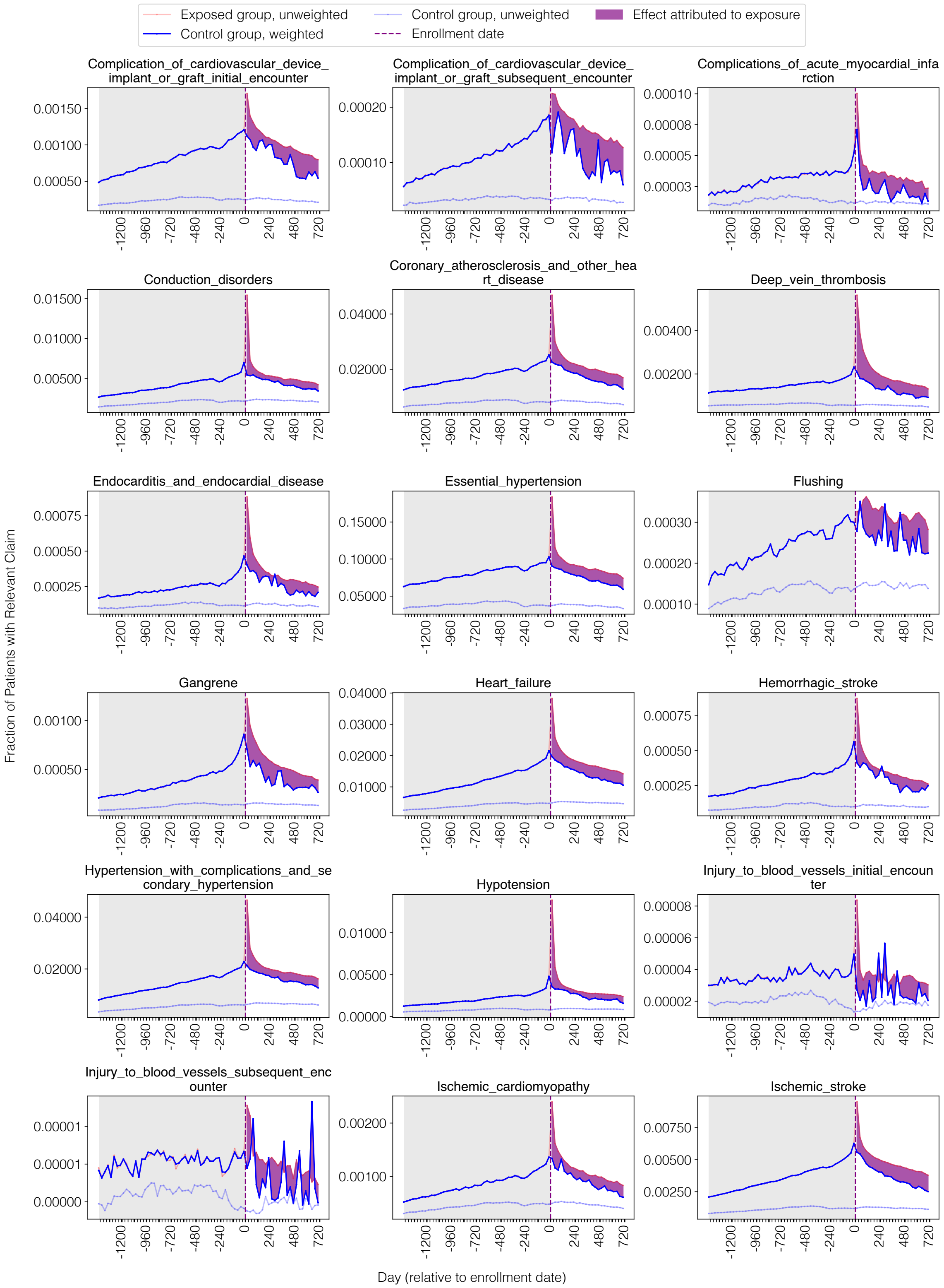


**Supplementary Figure S123:** (cont'd) Temporal trends in Diseases of the Circulatory System prevalence. The fraction of patients with relevant claims over time belonging for each outcome in the **conventional retrospective** study experiment.

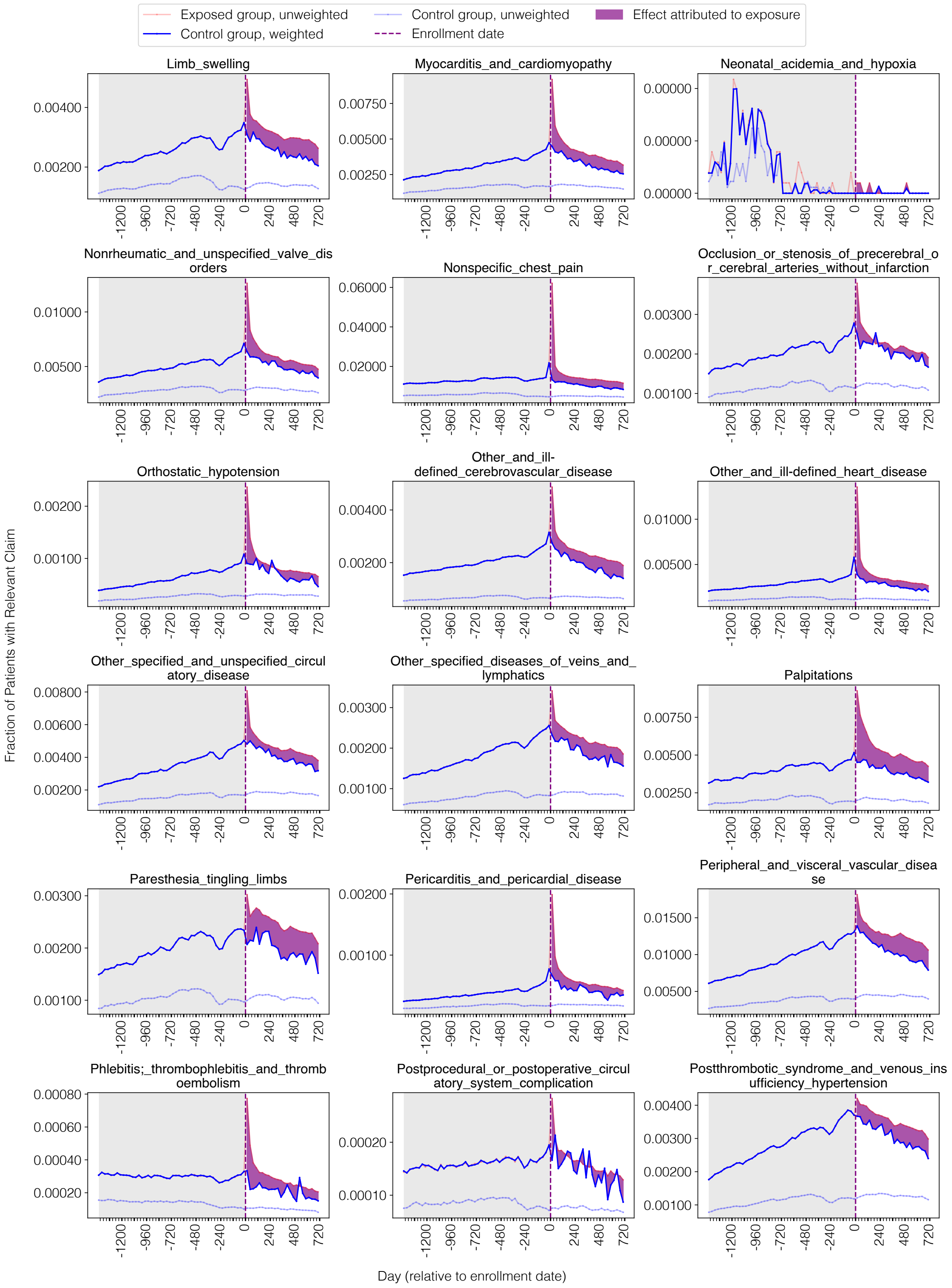


**Supplementary Figure S123:** (cont'd) Temporal trends in Diseases of the Circulatory System prevalence. The fraction of patients with relevant claims over time belonging for each outcome in the **conventional retrospective** study experiment.

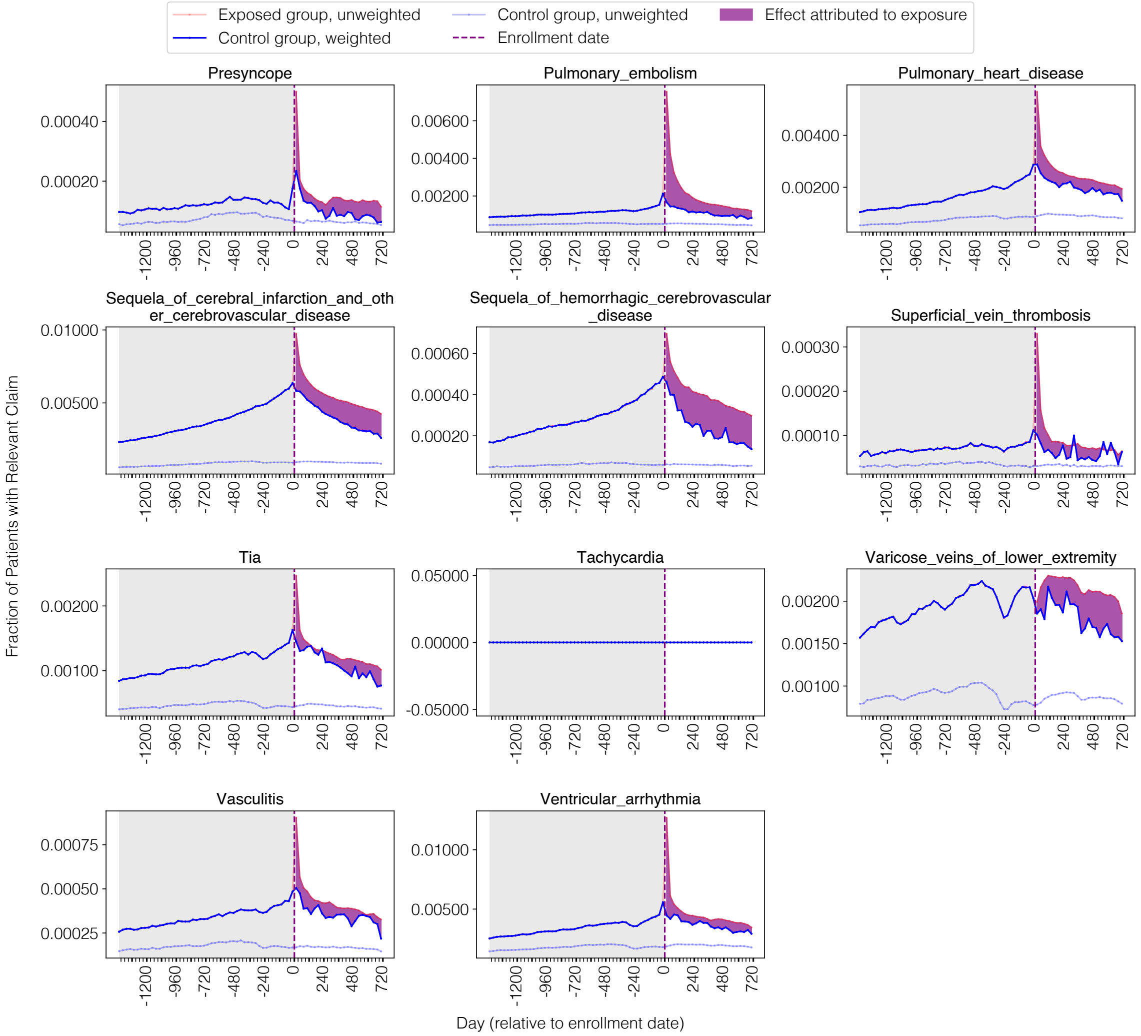


**Supplementary Figure S123:** (cont'd) Temporal trends in Diseases of the Circulatory System prevalence. The fraction of patients with relevant claims over time belonging for each outcome in the **conventional retrospective** study experiment.

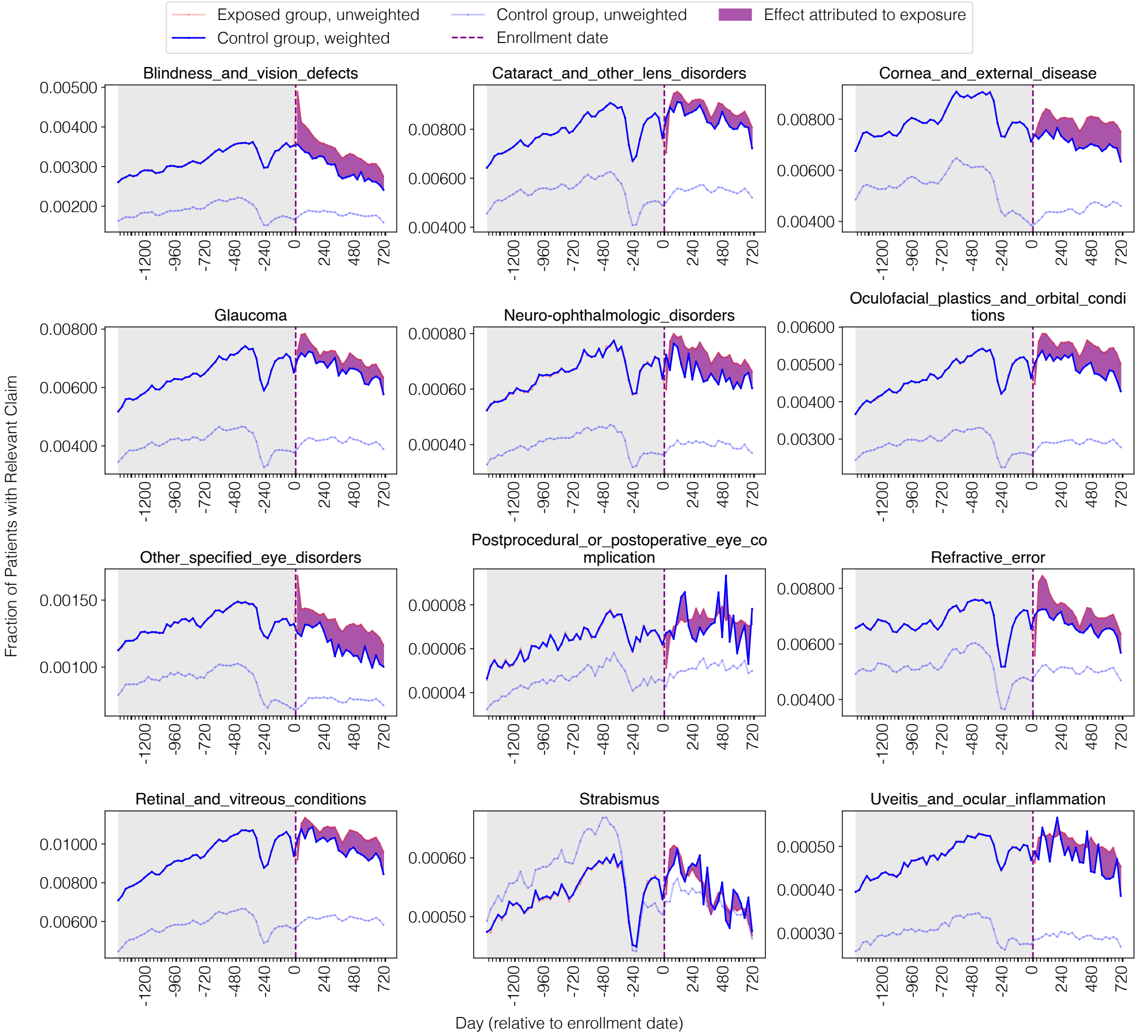


**Supplementary Figure S124:** Temporal trends in Diseases of the Eye and Adnexa prevalence. The fraction of patients with relevant claims over time belonging for each outcome in the **conventional retrospective** study experiment.

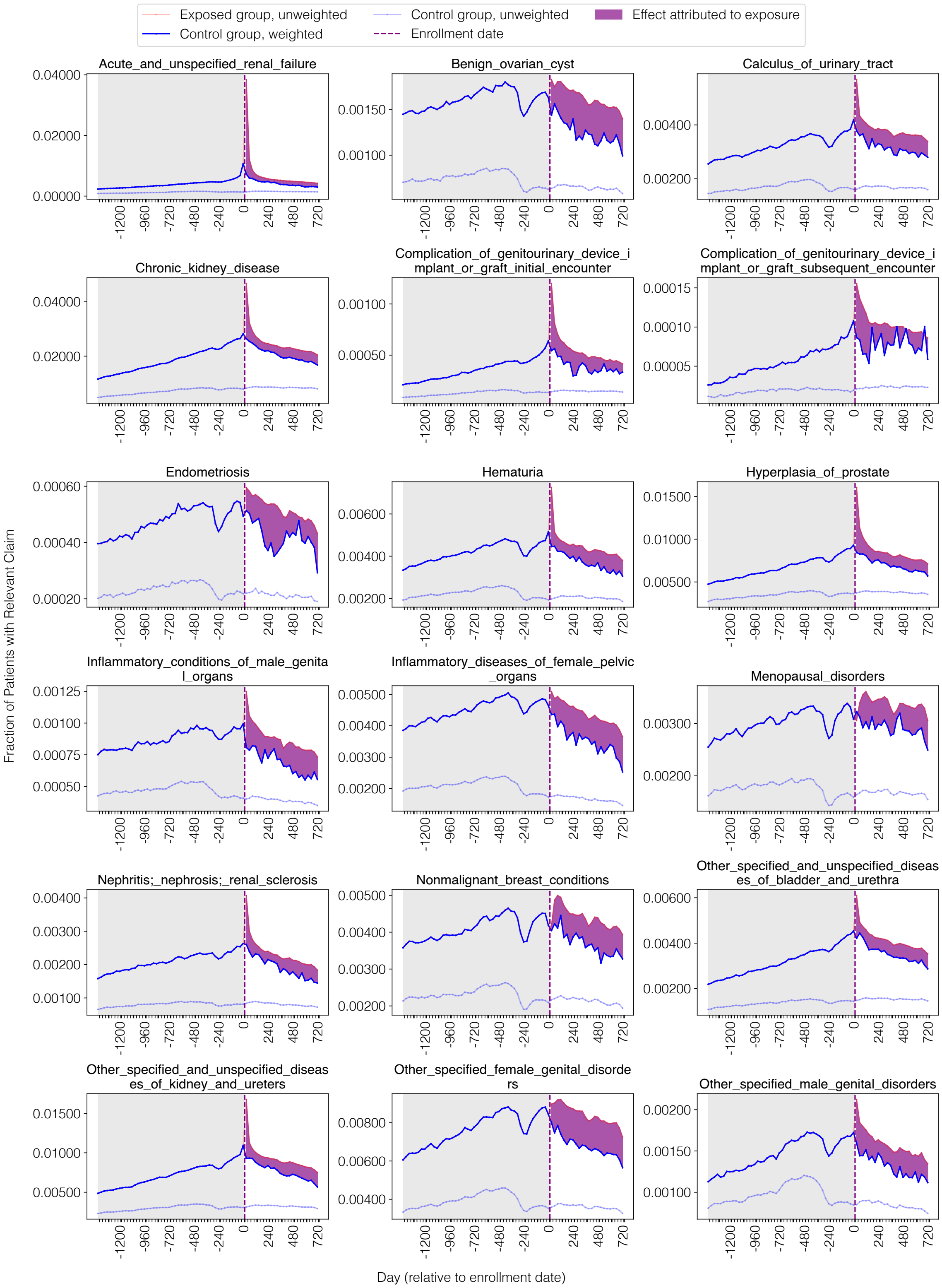


**Supplementary Figure S125:** Temporal trends in Diseases of the Genitourinary System prevalence. The fraction of patients with relevant claims over time belonging for each outcome in the **conventional retrospective** study experiment.

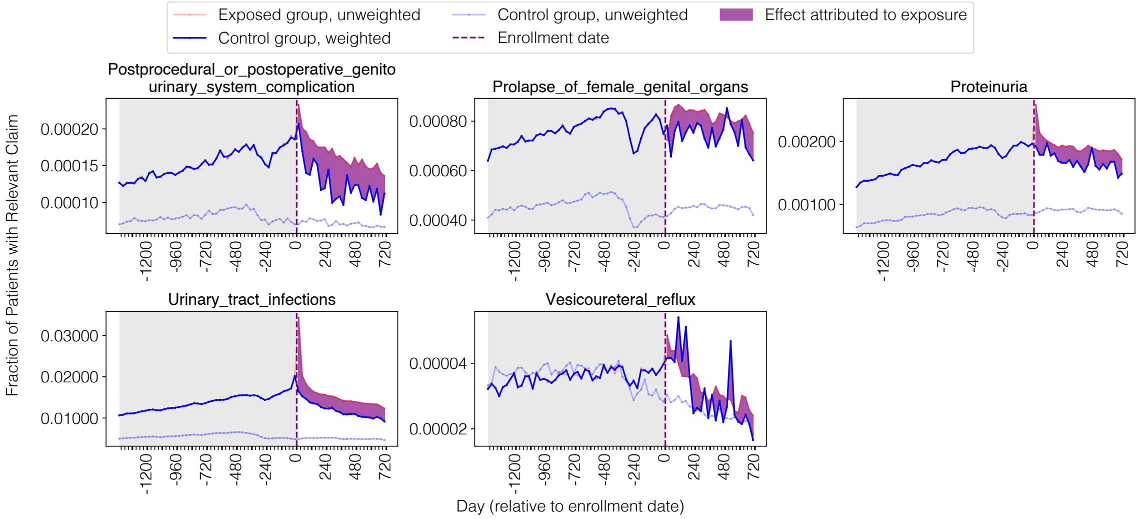


**Supplementary Figure S125:** (cont'd) Temporal trends in Diseases of the Genitourinary System prevalence. The fraction of patients with relevant claims over time belonging for each outcome in the **conventional retrospective** study experiment.

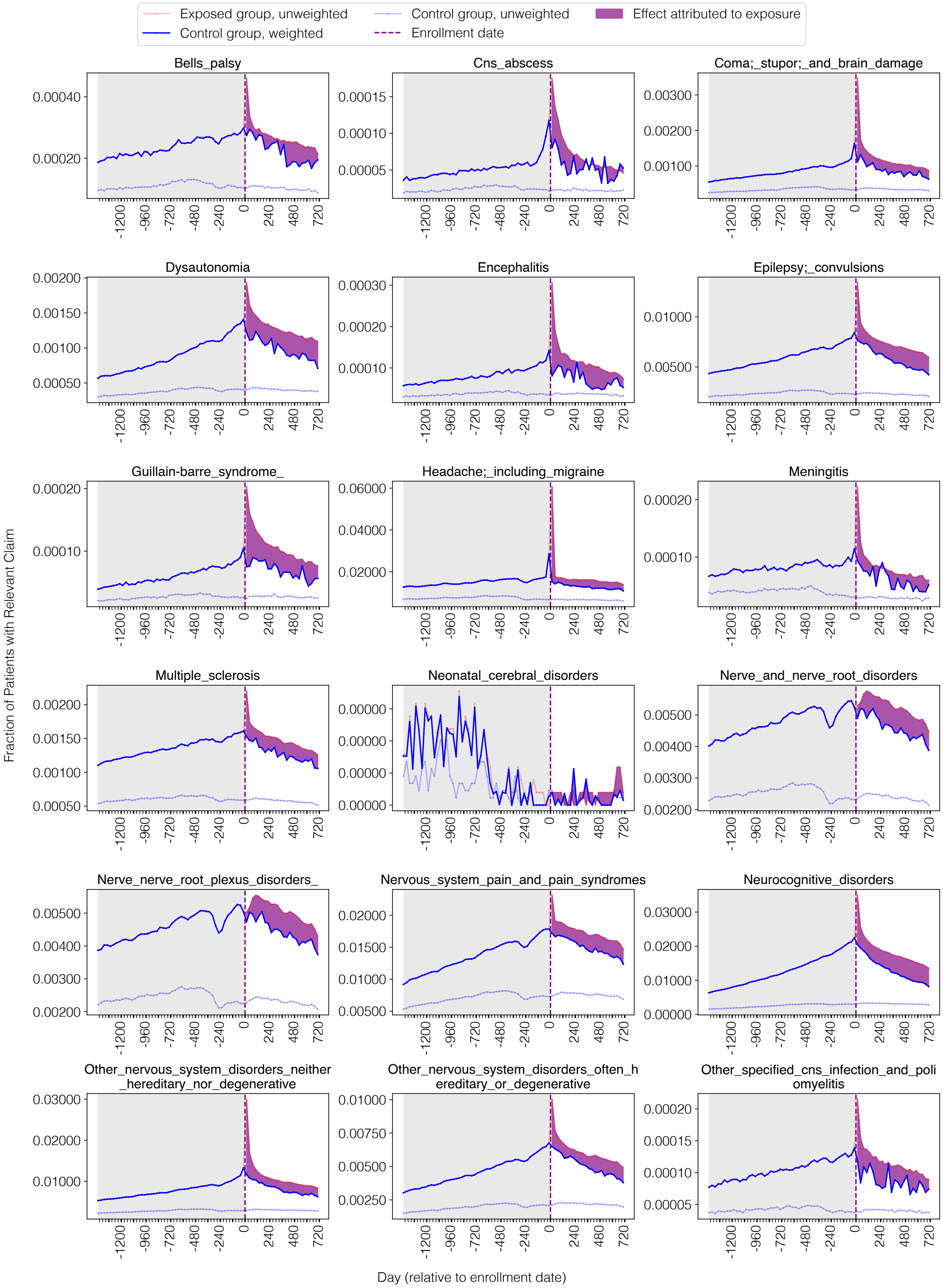


**Supplementary Figure S126:** Temporal trends in Diseases of the Nervous System prevalence. The fraction of patients with relevant claims over time belonging for each outcome in the **conventional retrospective** study experiment.

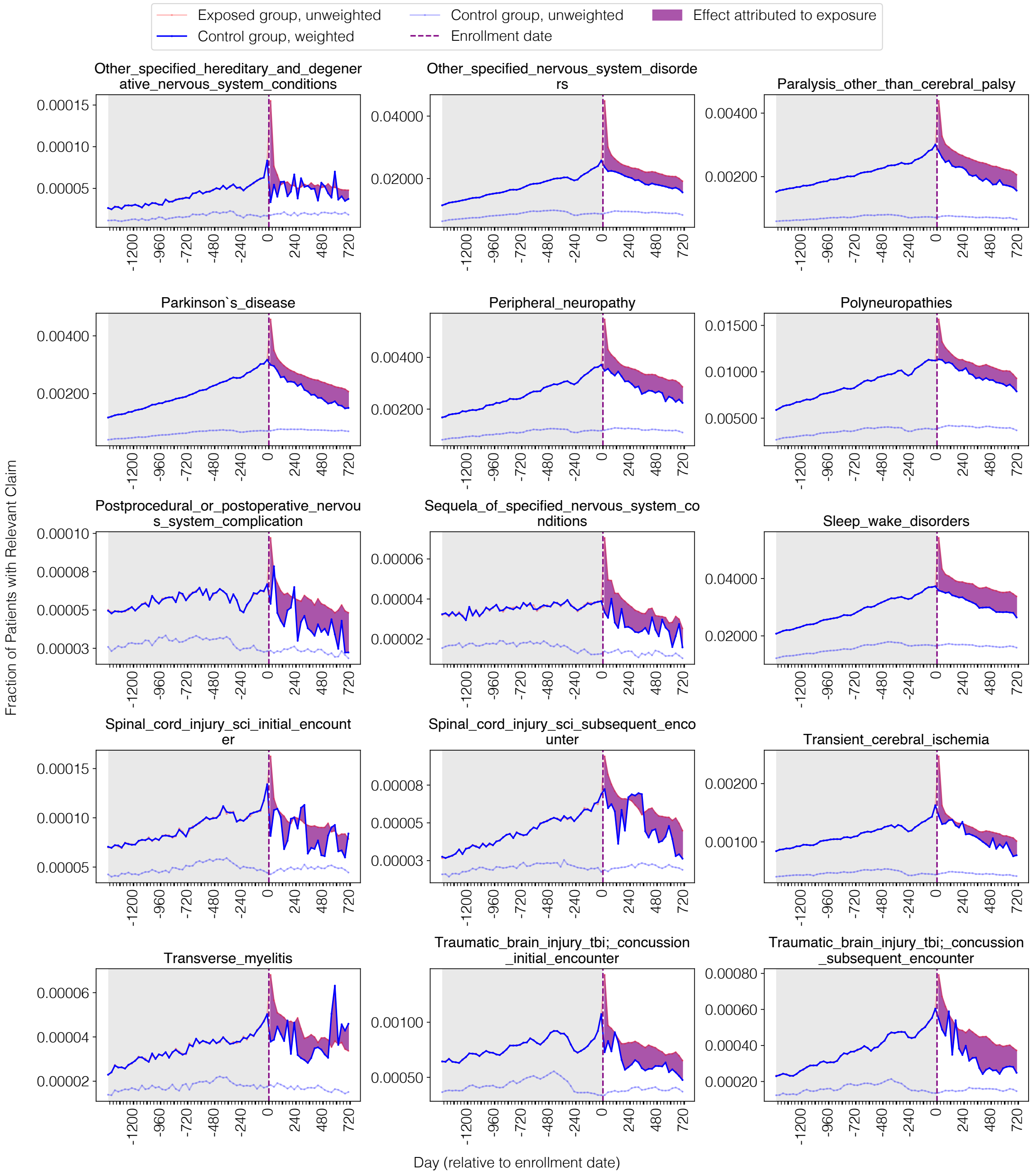


**Supplementary Figure S126:** (cont'd) Temporal trends in Diseases of the Nervous System prevalence. The fraction of patients with relevant claims over time belonging for each outcome in the **conventional retrospective** study experiment.

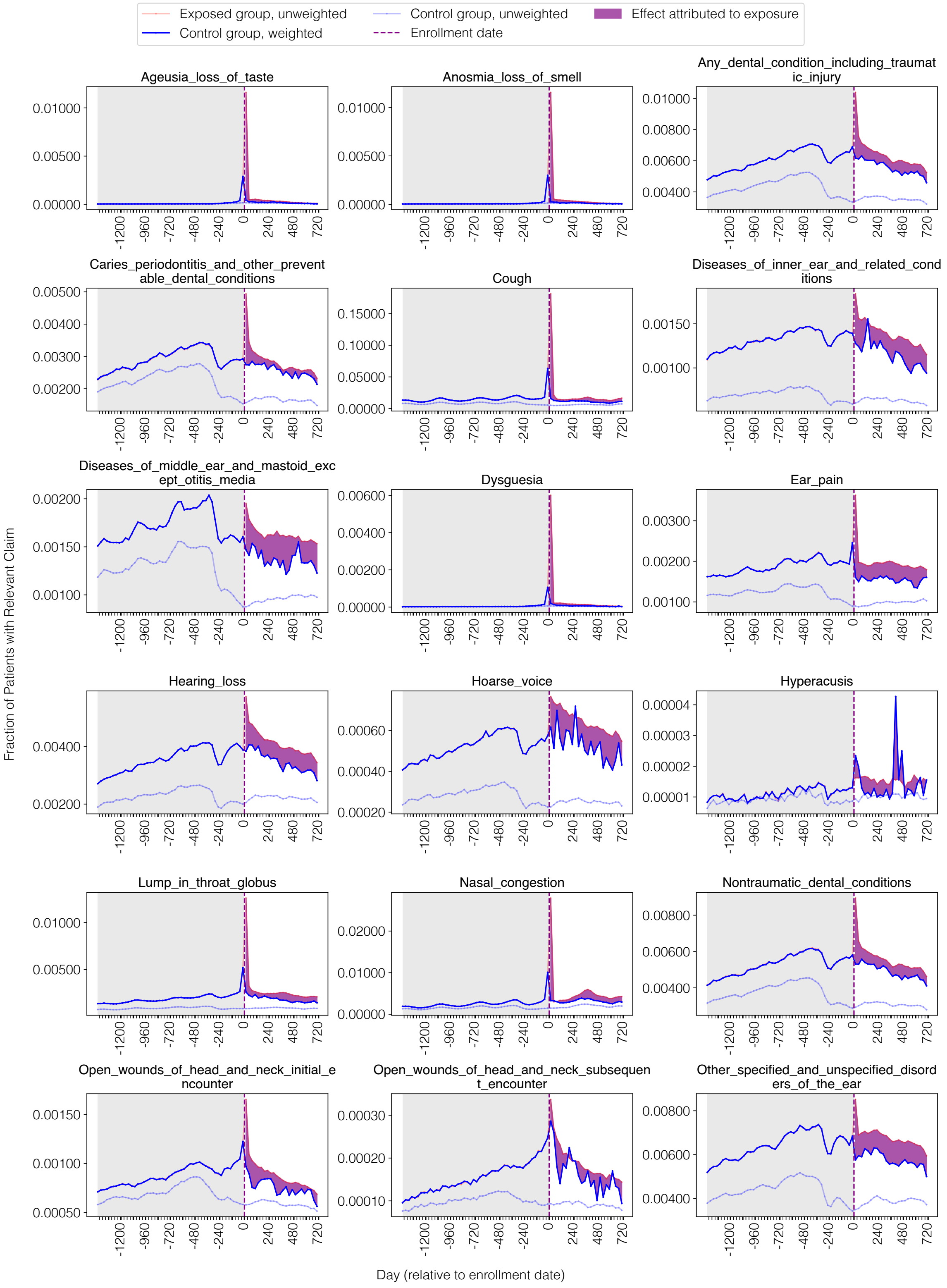


**Supplementary Figure S127:** Temporal trends in Ear, nose and throat prevalence. The fraction of patients with relevant claims over time belonging for each outcome in the **conventional retrospective** study experiment.

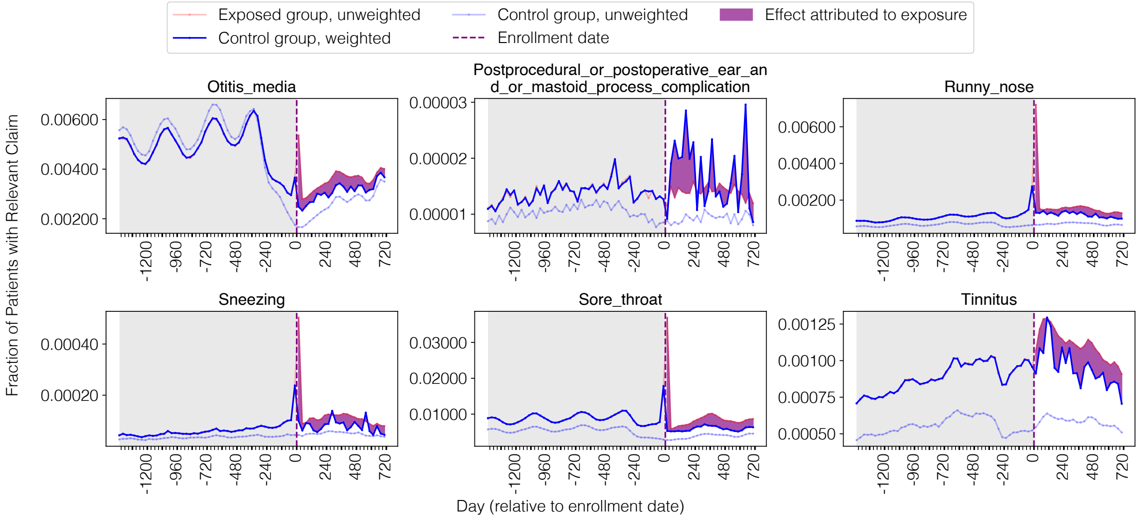


**Supplementary Figure S127:** (cont'd) Temporal trends in Ear, nose and throat prevalence. The fraction of patients with relevant claims over time belonging for each outcome in the **conventional retrospective** study experiment.

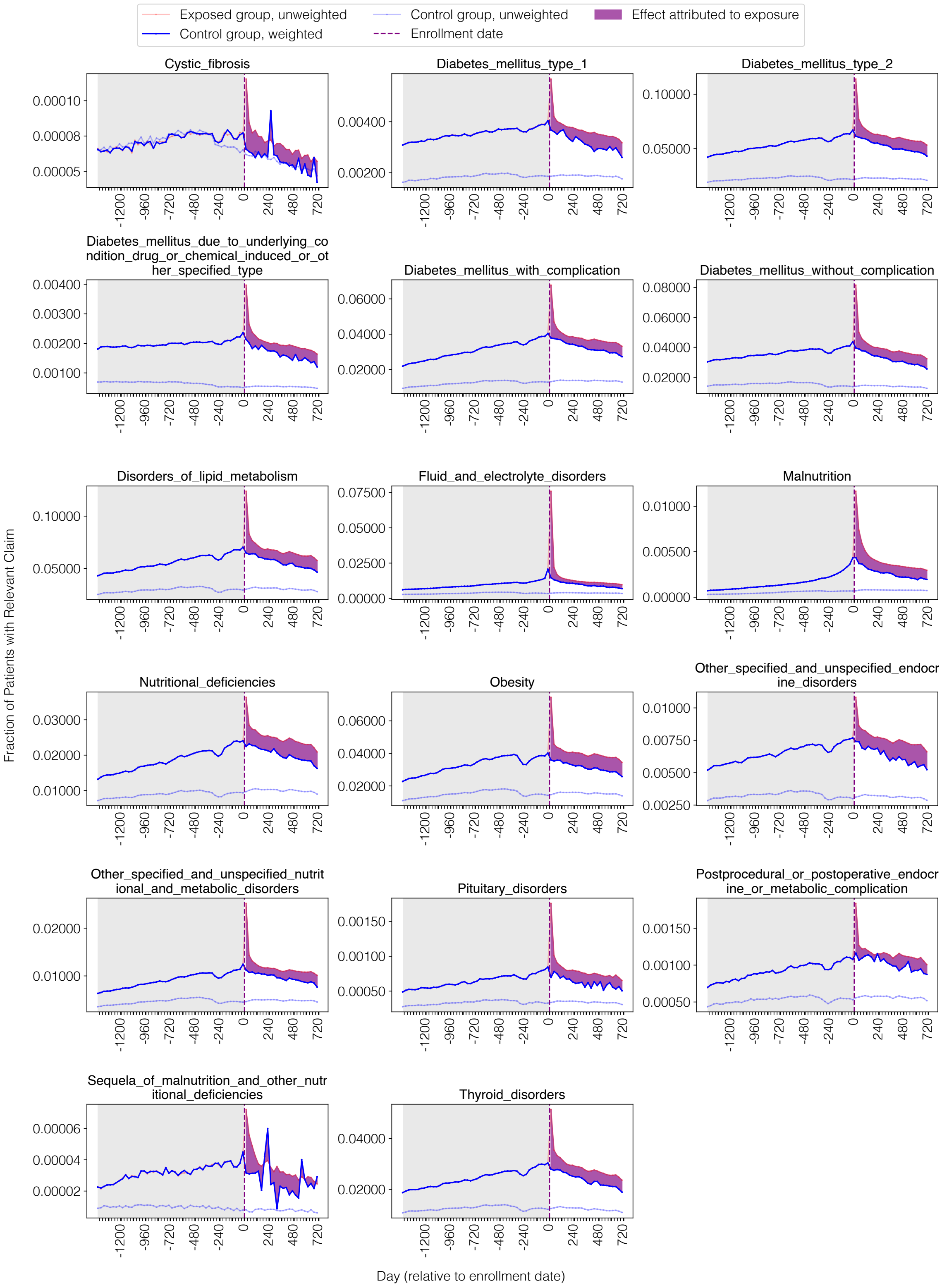


**Supplementary Figure S128:** Temporal trends in Endocrine, Nutritional and Metabolic Diseases prevalence. The fraction of patients with relevant claims over time belonging for each outcome in the **conventional retrospective** study experiment.

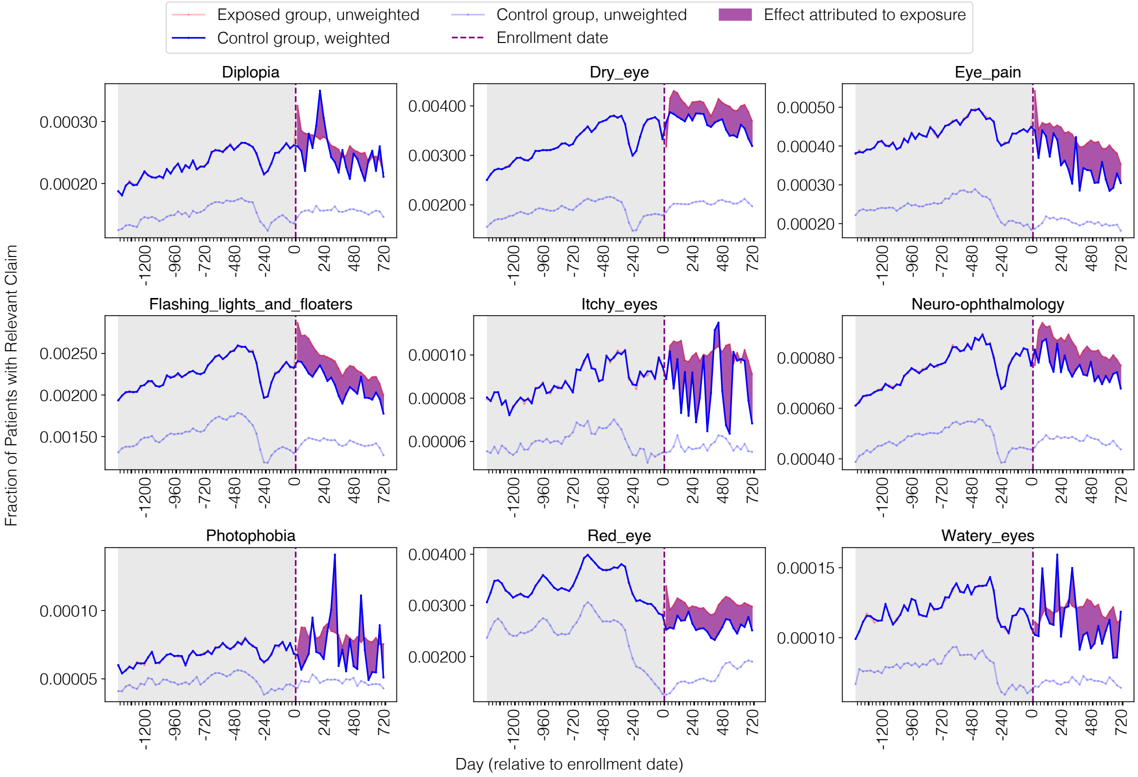


**Supplementary Figure S129:** Temporal trends in Eyes prevalence. The fraction of patients with relevant claims over time belonging for each outcome in the **conventional retrospective** study experiment.

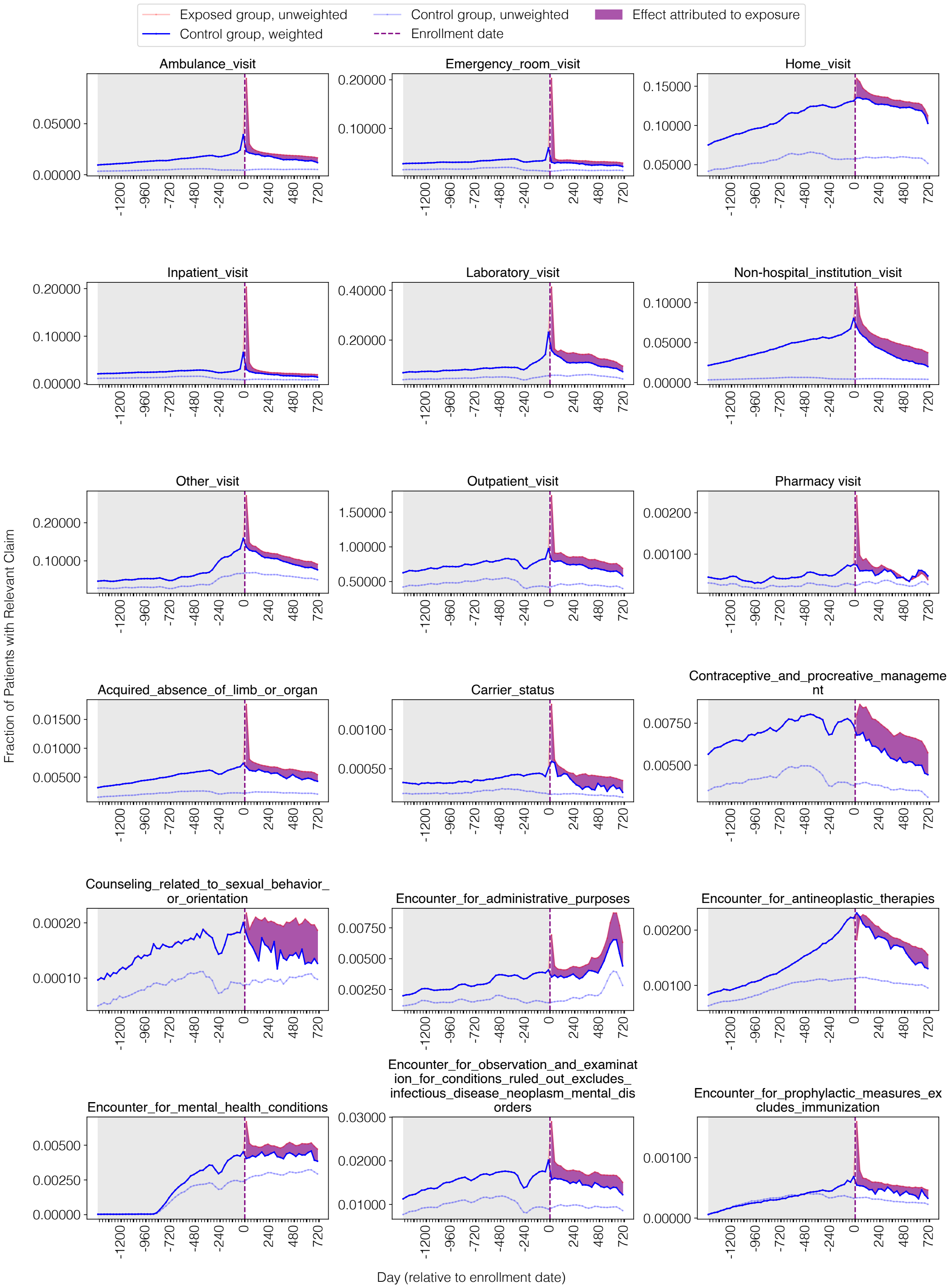


**Supplementary Figure S130:** Temporal trends in Factors Influencing Health Status and Contact with Health Services prevalence. The fraction of patients with relevant claims over time belonging for each outcome in the **conventional retrospective** study experiment.

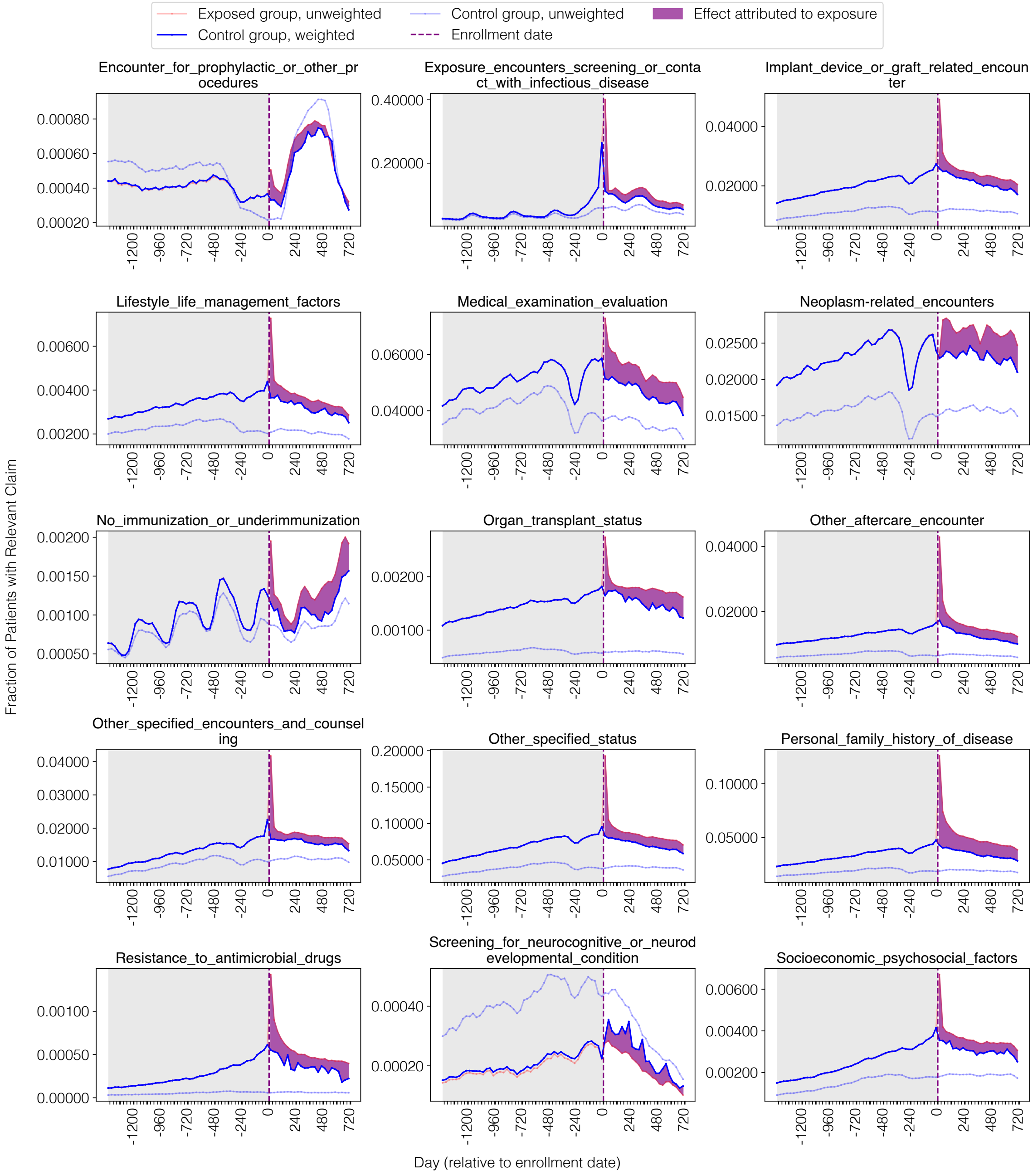


**Supplementary Figure S130:** (cont'd) Temporal trends in Factors Influencing Health Status and Contact with Health Services prevalence. The fraction of patients with relevant claims over time belonging for each outcome in the **conventional retrospective** study experiment.

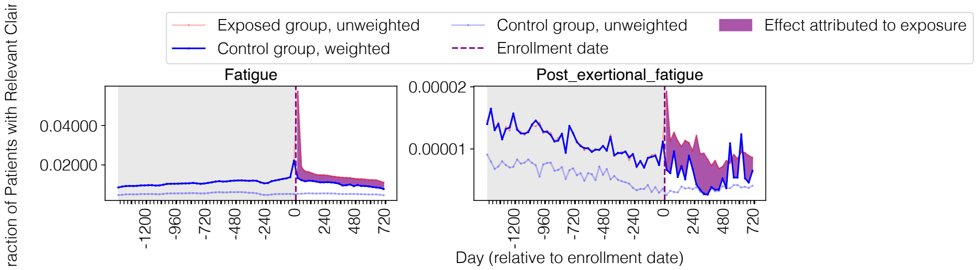


**Supplementary Figure S131:** Temporal trends in Fatigue prevalence. The fraction of patients with relevant claims over time belonging for each outcome in the **conventional retrospective** study experiment.

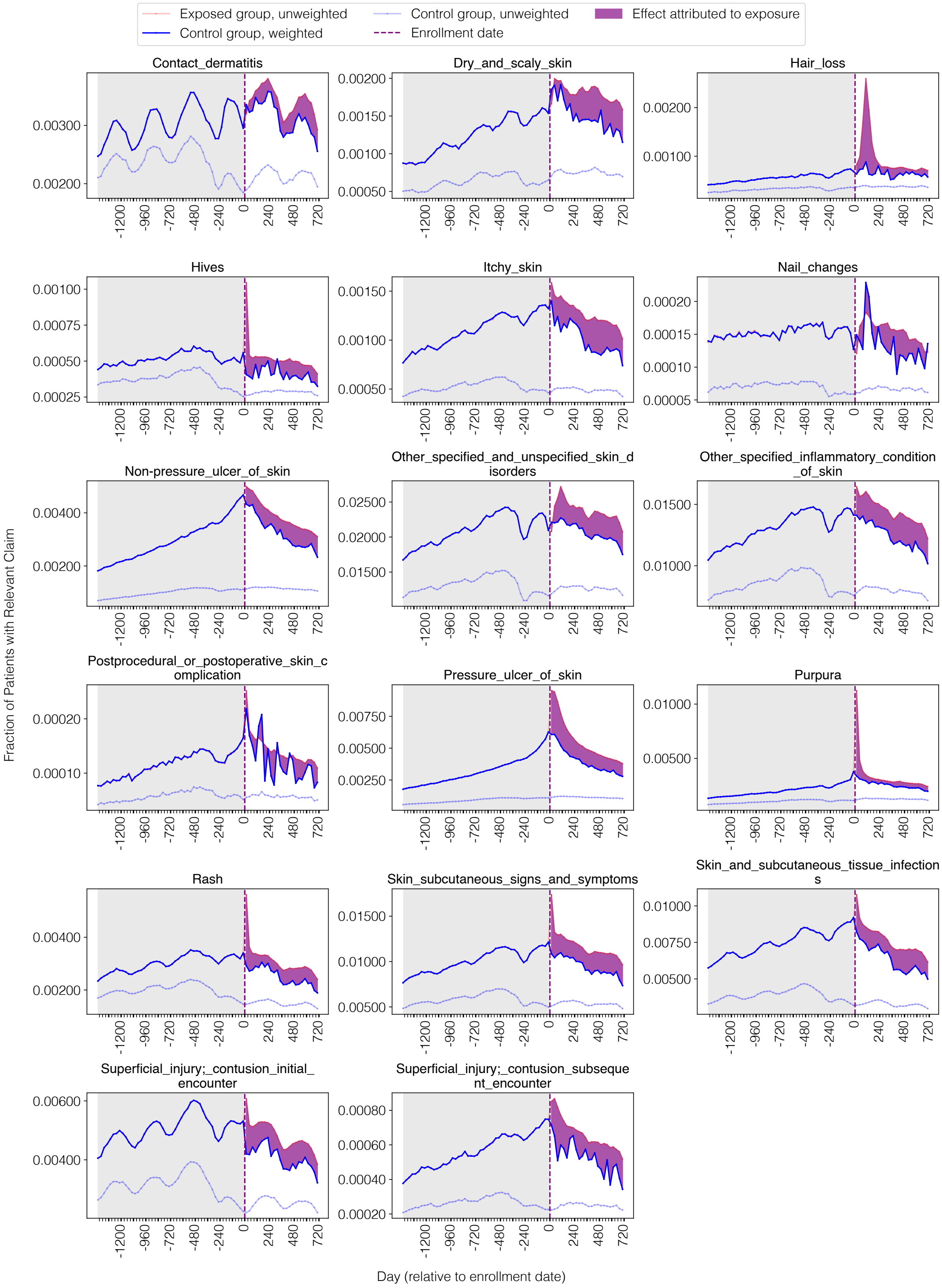


**Supplementary Figure S132:** Temporal trends in Hair, skin and nails prevalence. The fraction of patients with relevant claims over time belonging for each outcome in the **conventional retrospective** study experiment.

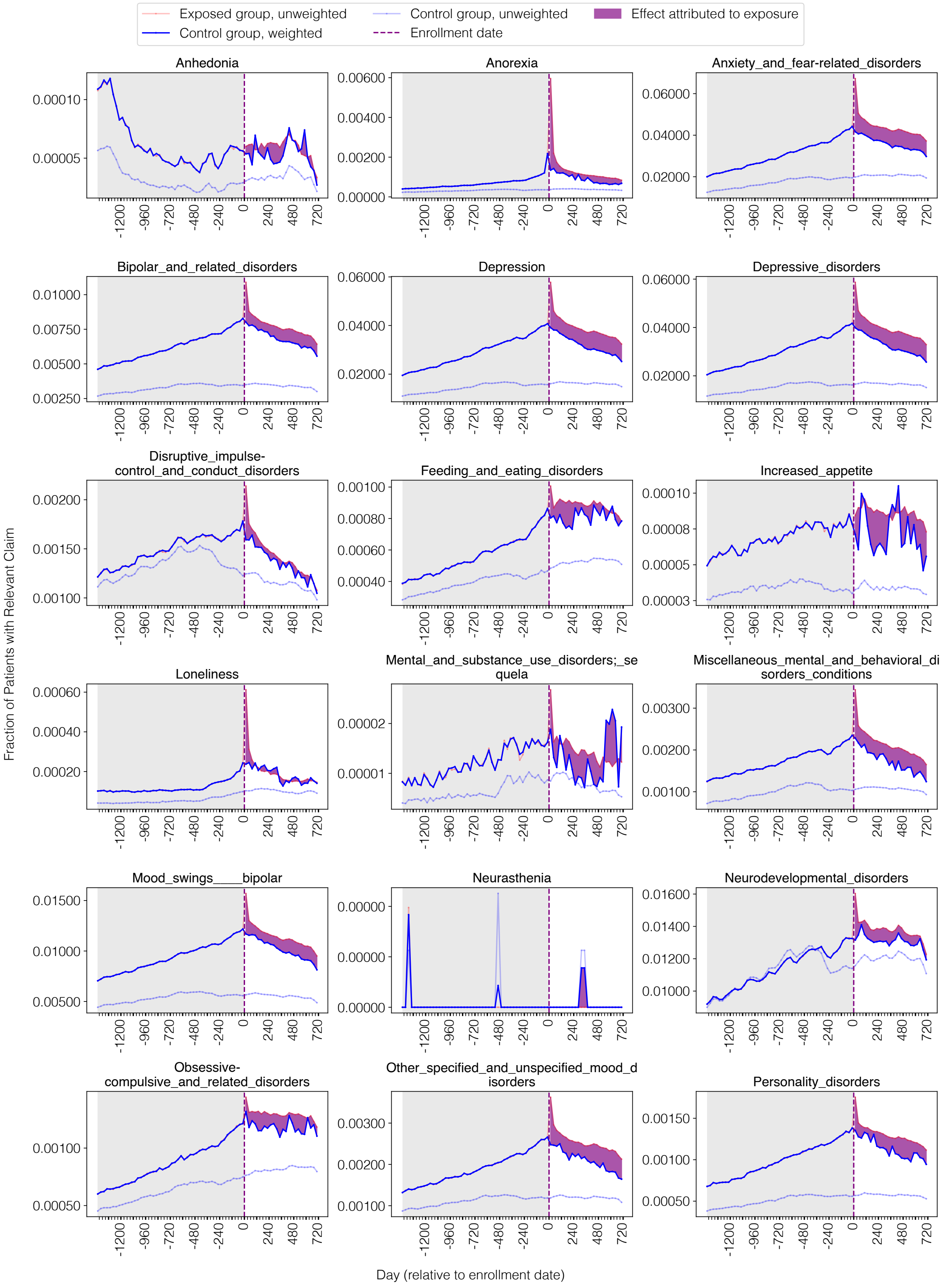


**Supplementary Figure S133:** Temporal trends in Mental health prevalence. The fraction of patients with relevant claims over time belonging for each outcome in the **conventional retrospective** study experiment.

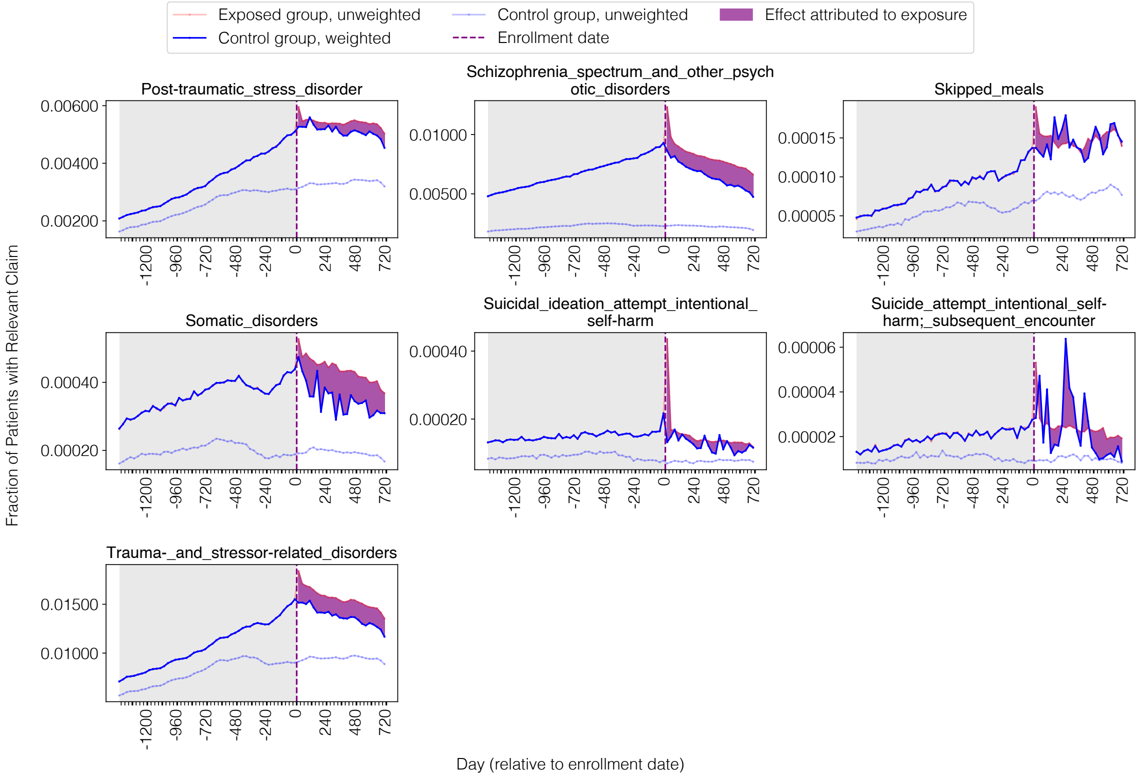


**Supplementary Figure S133:** (cont'd) Temporal trends in Mental health prevalence. The fraction of patients with relevant claims over time belonging for each outcome in the **conventional retrospective** study experiment.

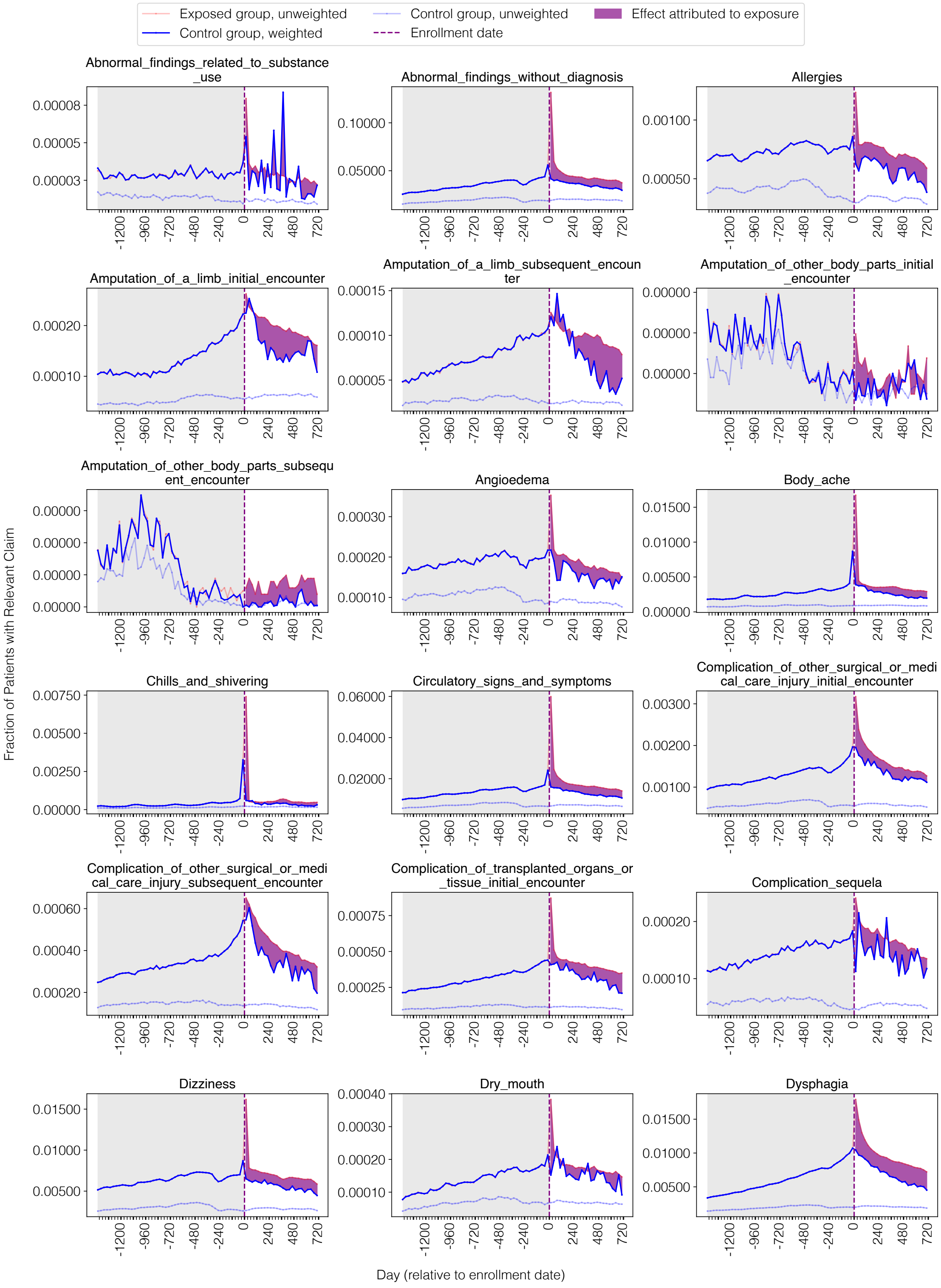


**Supplementary Figure S134:** Temporal trends in Misc prevalence. The fraction of patients with relevant claims over time belonging for each outcome in the **conventional retrospective** study experiment.

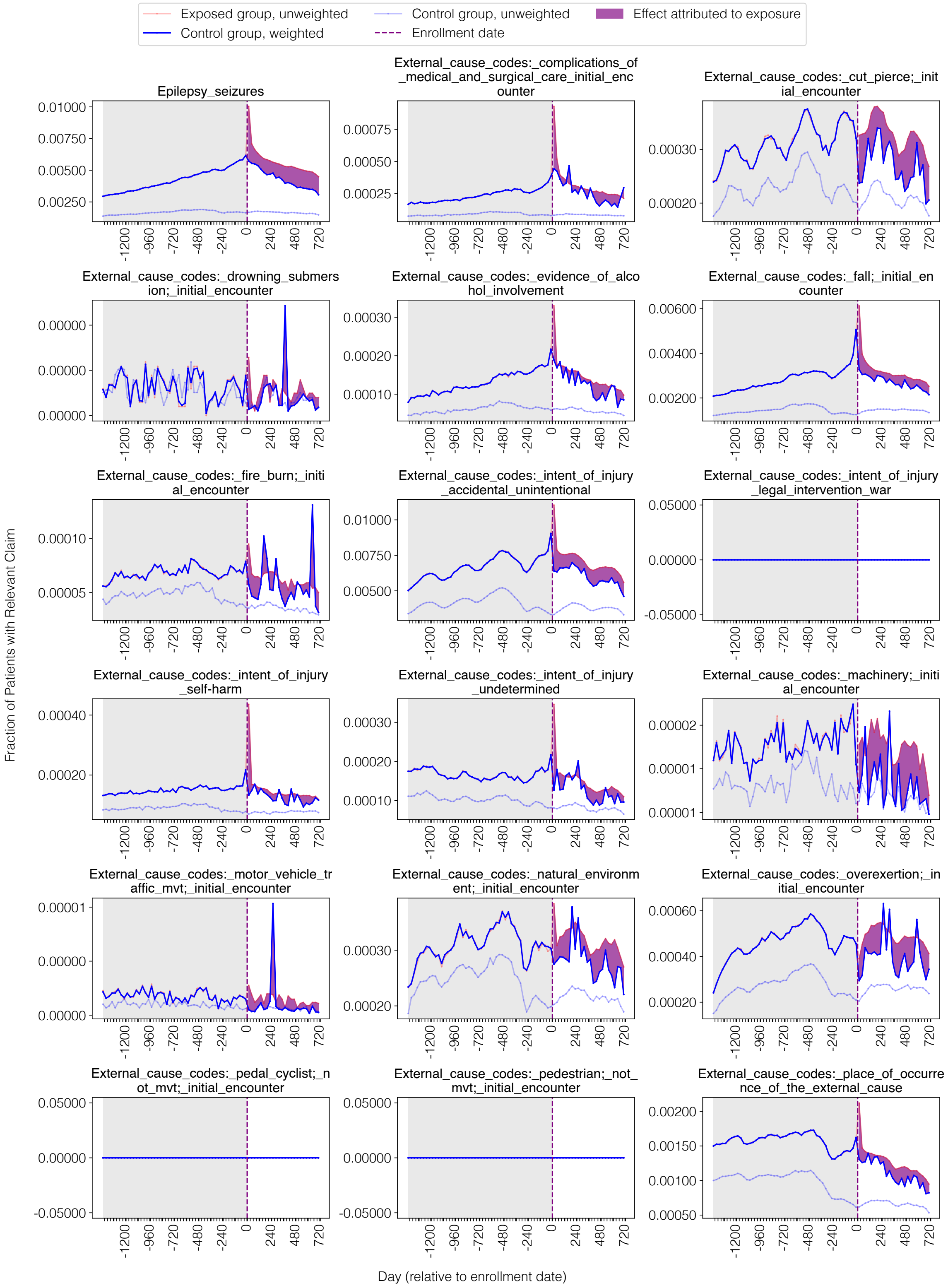


**Supplementary Figure S134:** (cont'd) Temporal trends in Misc prevalence. The fraction of patients with relevant claims over time belonging for each outcome in the **conventional retrospective** study experiment.

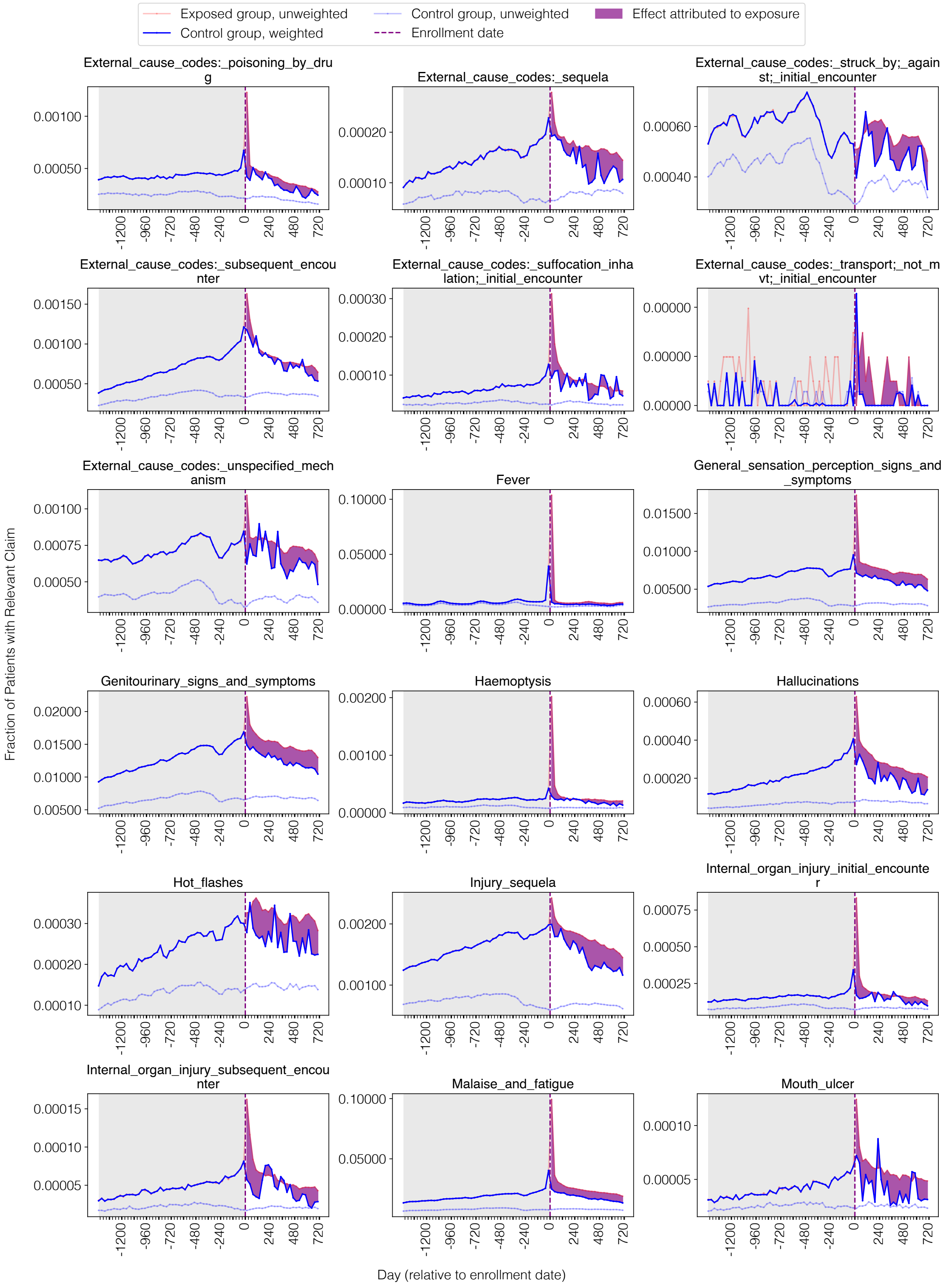


**Supplementary Figure S134:** (cont'd) Temporal trends in Misc prevalence. The fraction of patients with relevant claims over time belonging for each outcome in the **conventional retrospective** study experiment.

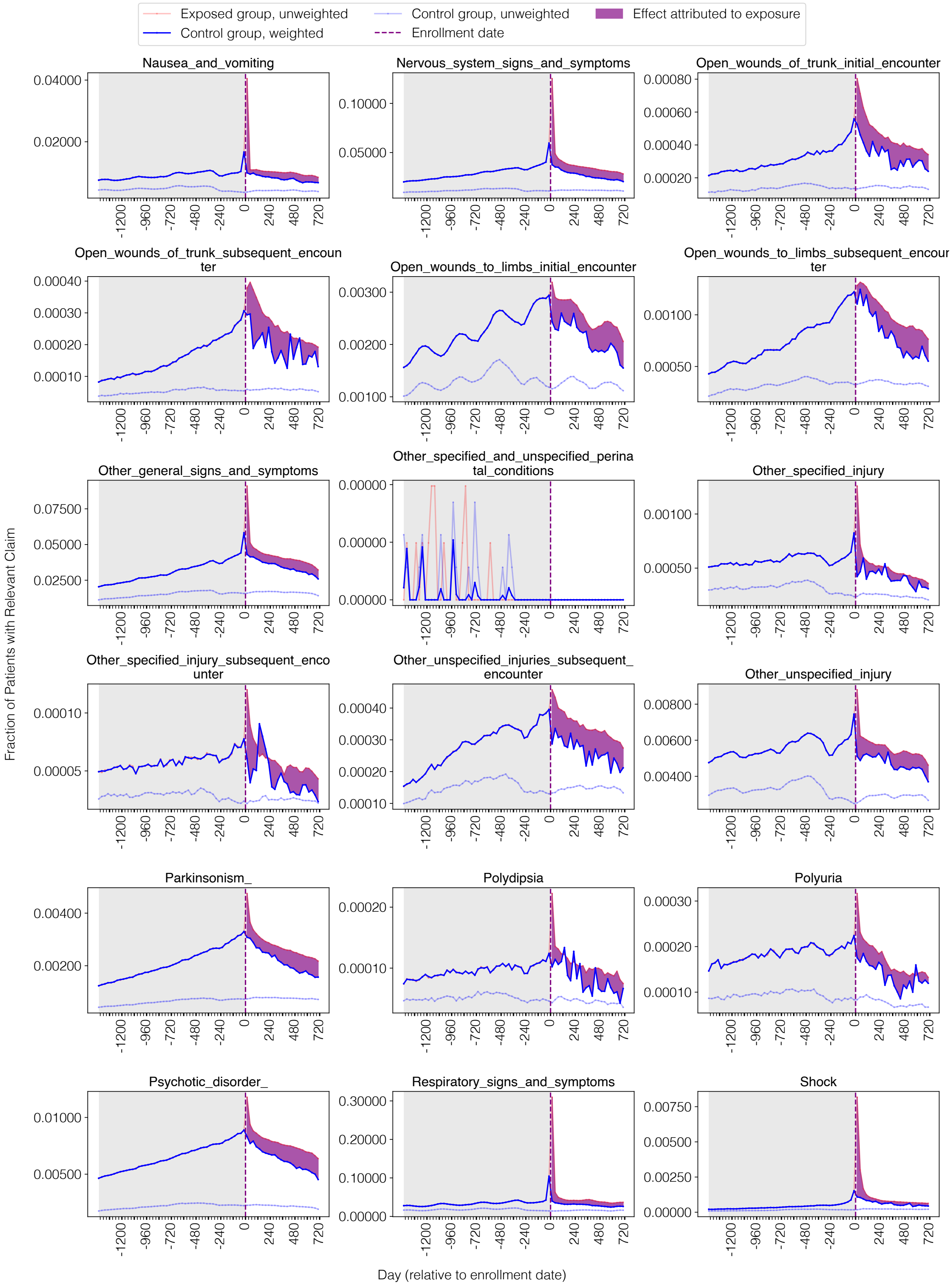


**Supplementary Figure S134:** (cont'd) Temporal trends in Misc prevalence. The fraction of patients with relevant claims over time belonging for each outcome in the **conventional retrospective** study experiment.

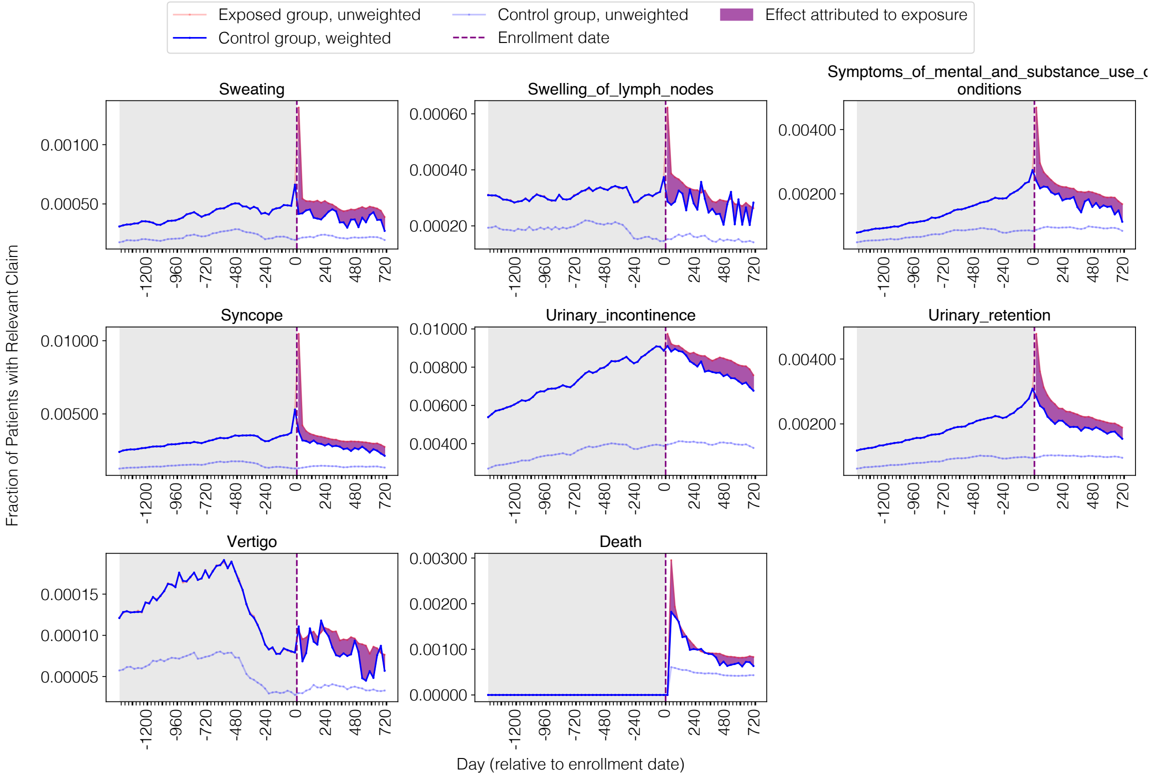


**Supplementary Figure S134:** (cont'd) Temporal trends in Misc prevalence. The fraction of patients with relevant claims over time belonging for each outcome in the **conventional retrospective** study experiment.

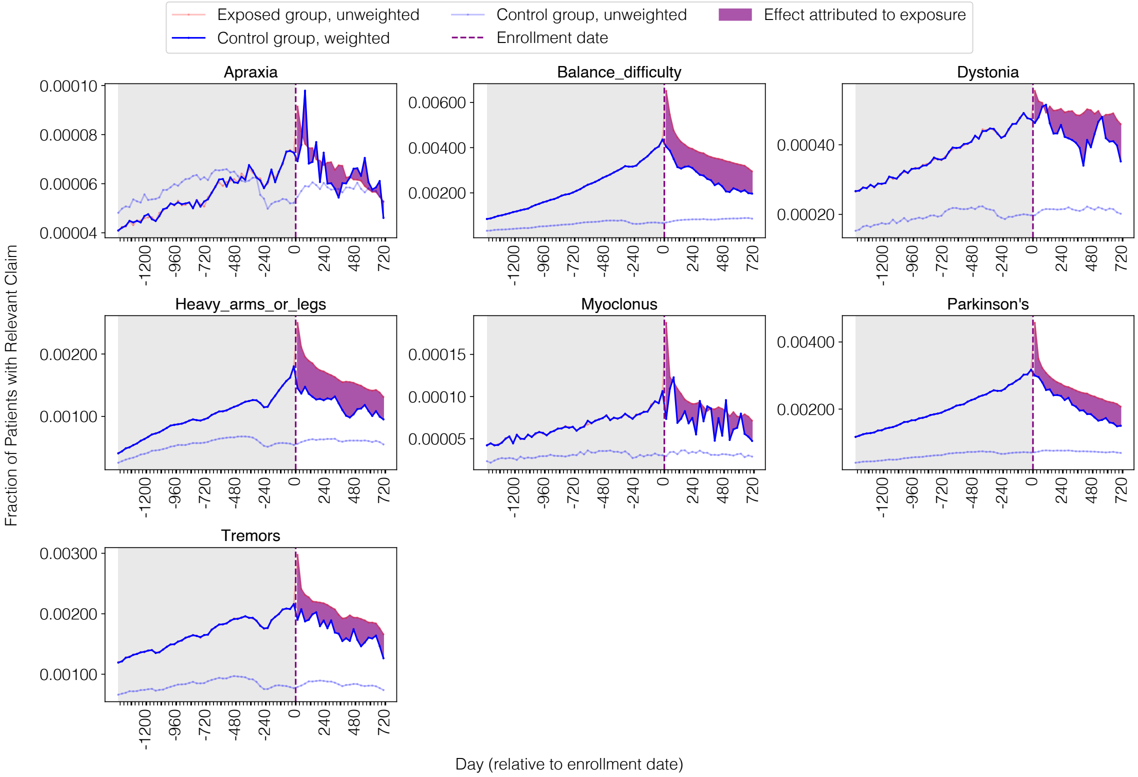


**Supplementary Figure S135:** Temporal trends in Movement prevalence. The fraction of patients with relevant claims over time belonging for each outcome in the **conventional retrospective** study experiment.

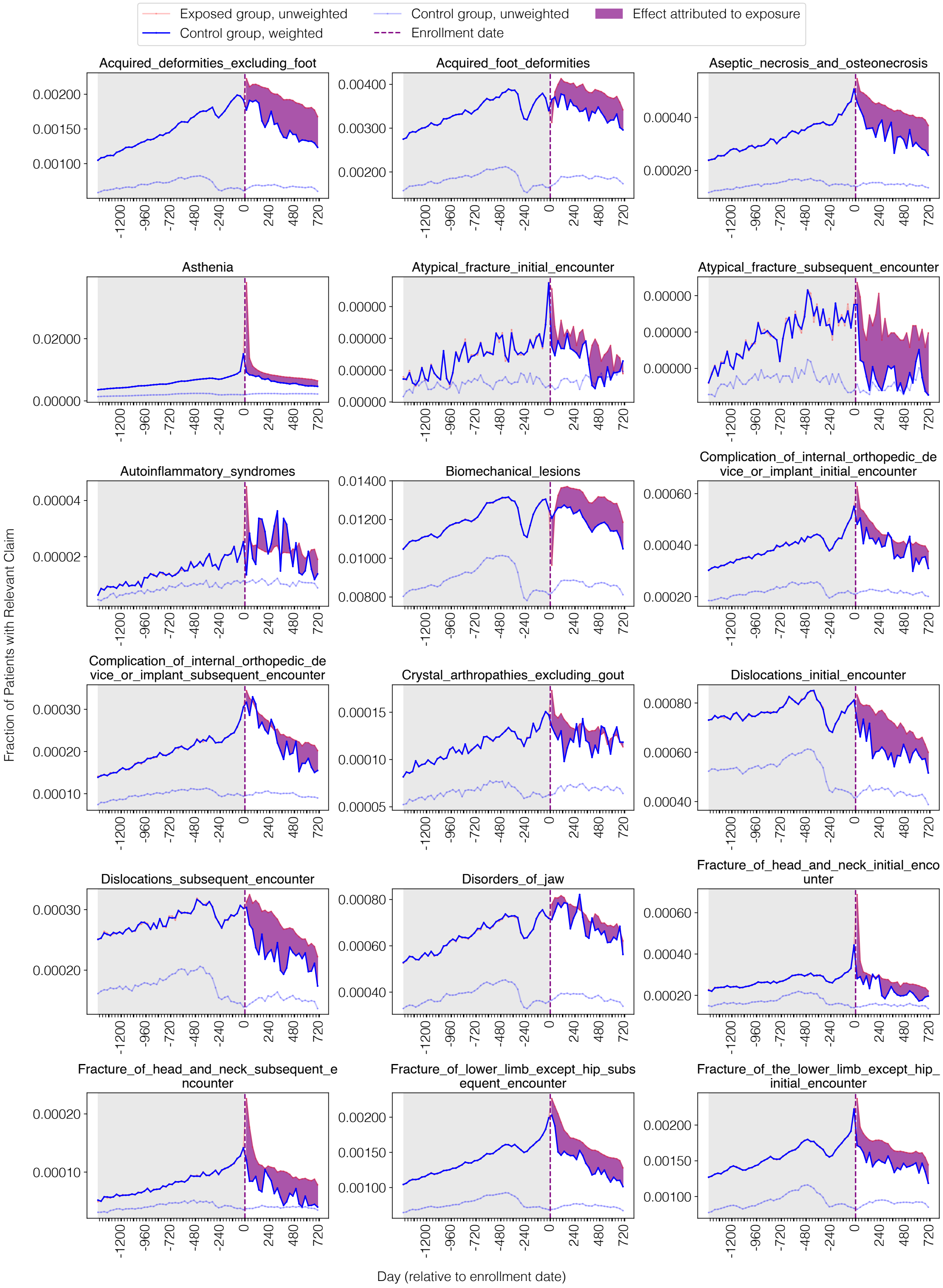


**Supplementary Figure S136:** Temporal trends in Muscles and joints prevalence. The fraction of patients with relevant claims over time belonging for each outcome in the **conventional retrospective** study experiment.

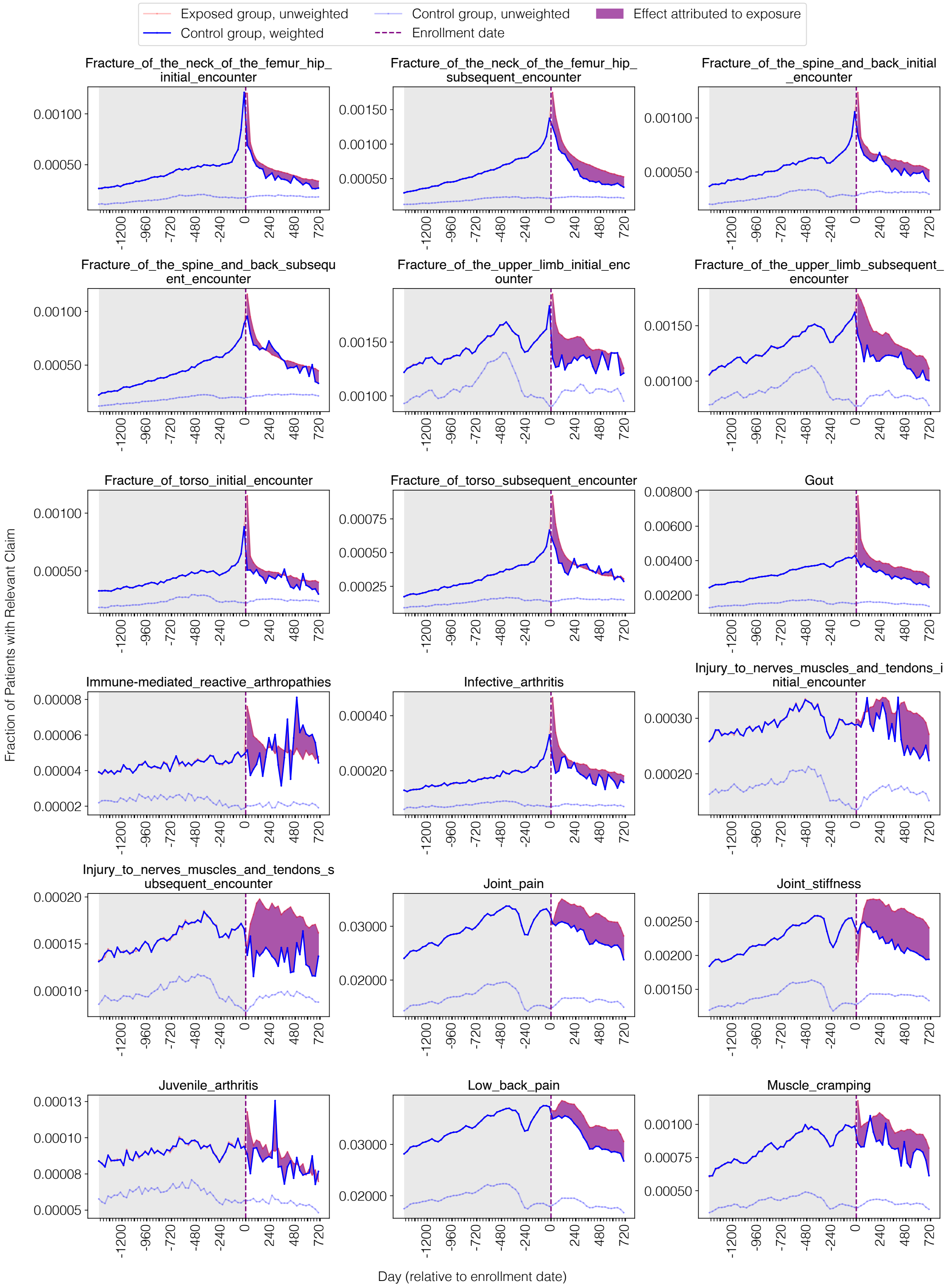


**Supplementary Figure S136:** (cont'd) Temporal trends in Muscles and joints prevalence. The fraction of patients with relevant claims over time belonging for each outcome in the **conventional retrospective** study experiment.

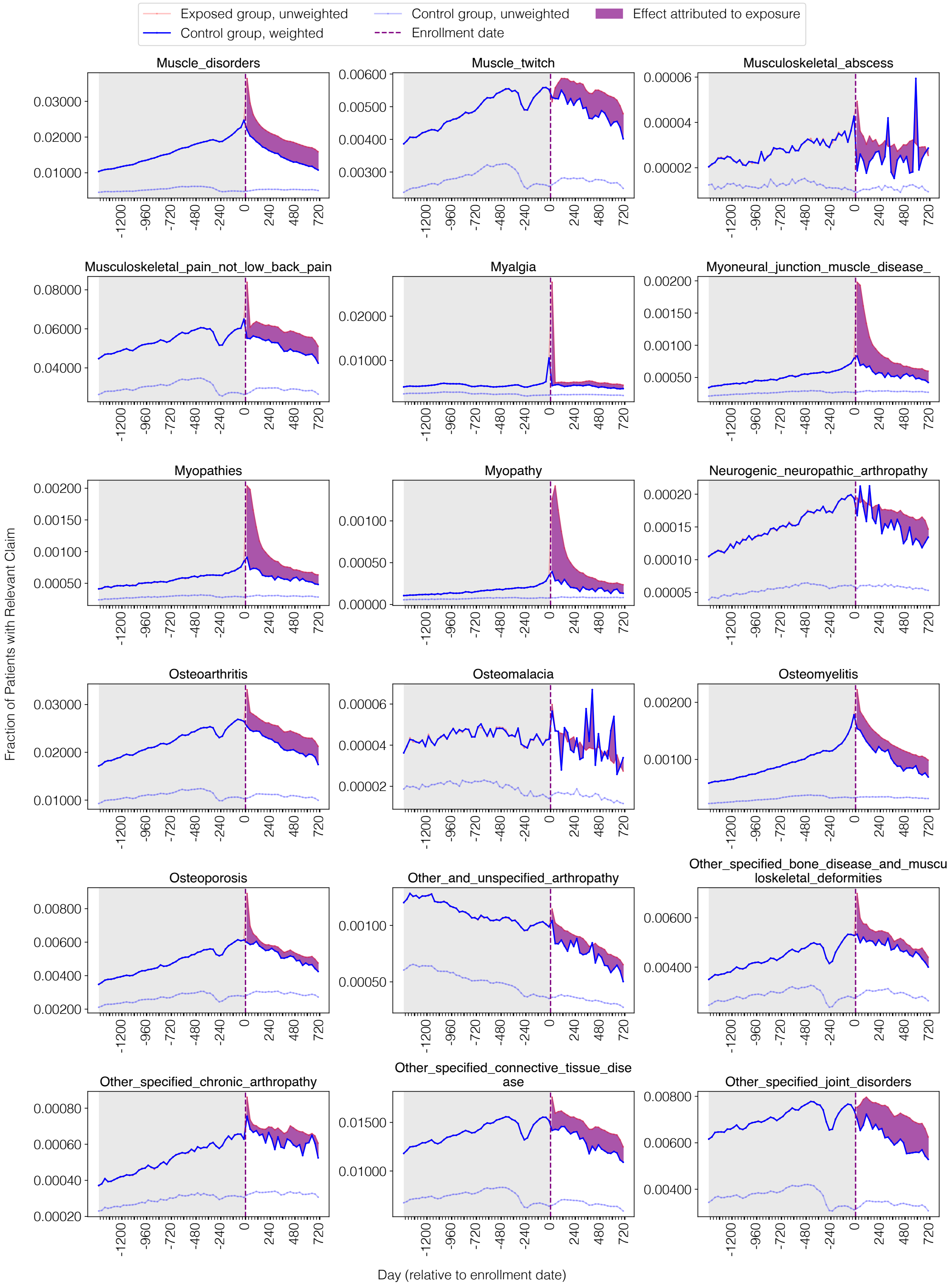


**Supplementary Figure S136:** (cont'd) Temporal trends in Muscles and joints prevalence. The fraction of patients with relevant claims over time belonging for each outcome in the **conventional retrospective** study experiment.

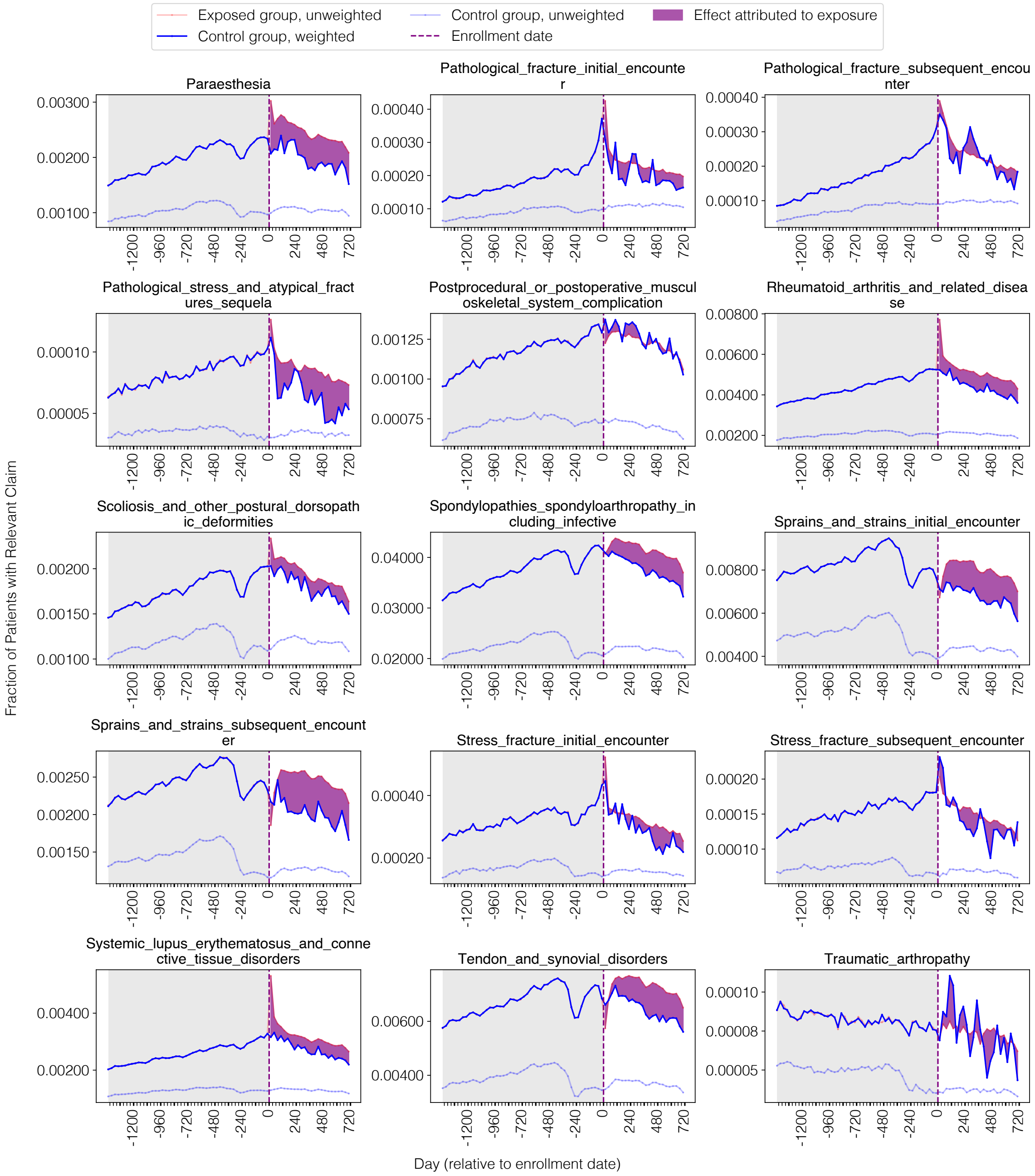


**Supplementary Figure S136:** (cont'd) Temporal trends in Muscles and joints prevalence. The fraction of patients with relevant claims over time belonging for each outcome in the **conventional retrospective** study experiment.

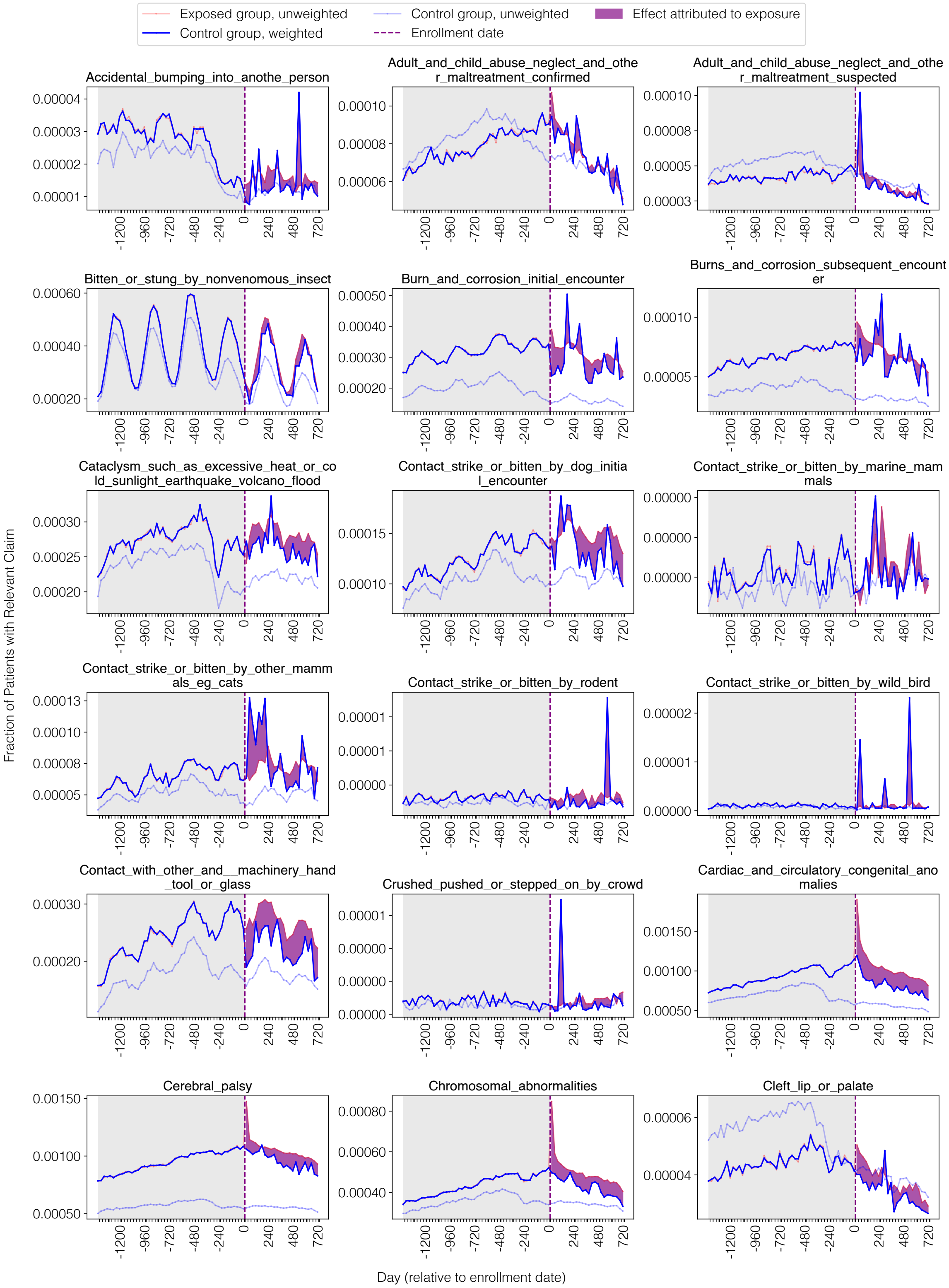


**Supplementary Figure S137:** Temporal trends in Negative prevalence. The fraction of patients with relevant claims over time belonging for each outcome in the **conventional retrospective** study experiment.

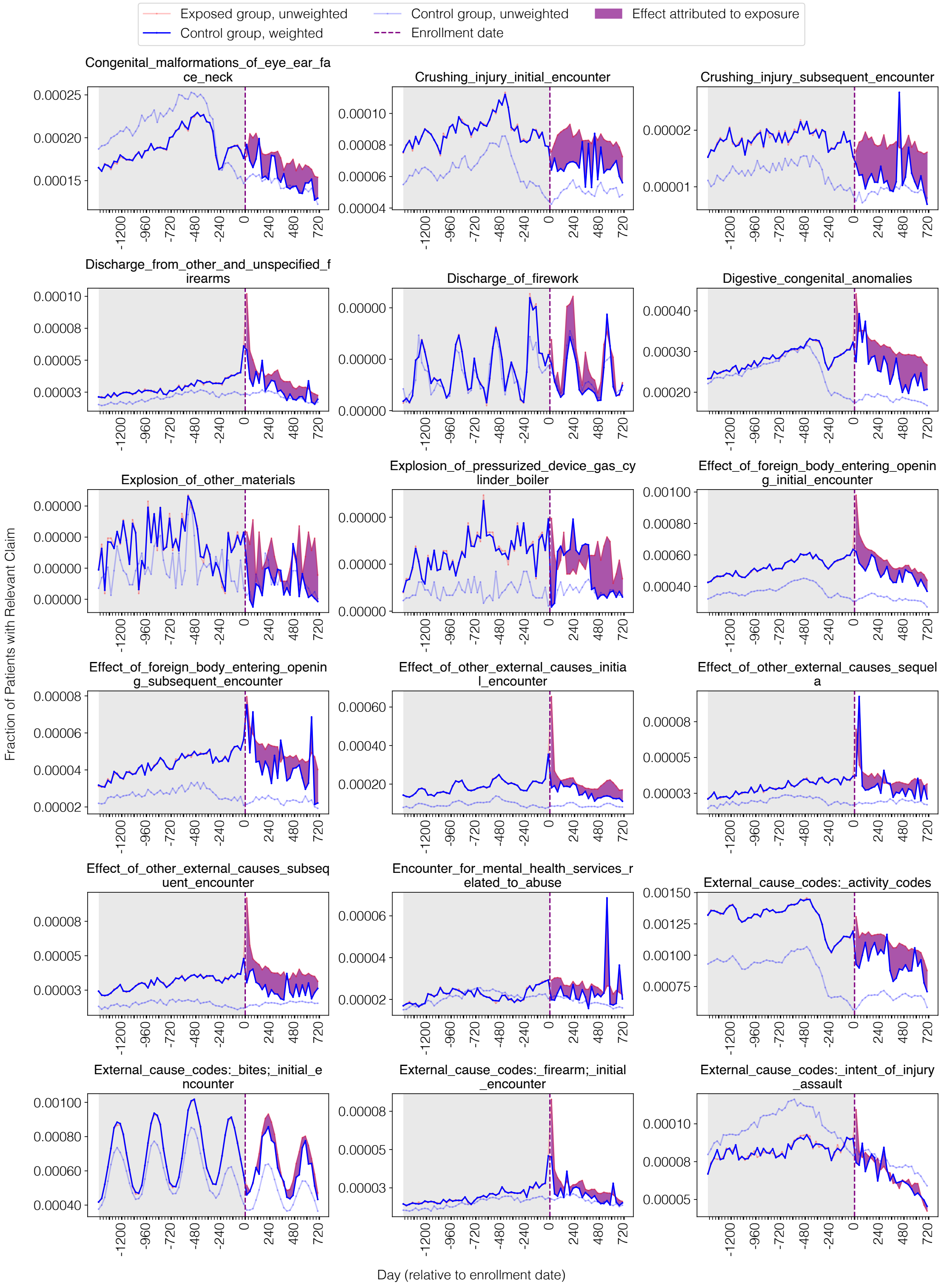


**Supplementary Figure S137:** (cont'd) Temporal trends in Negative prevalence. The fraction of patients with relevant claims over time belonging for each outcome in the **conventional retrospective** study experiment.

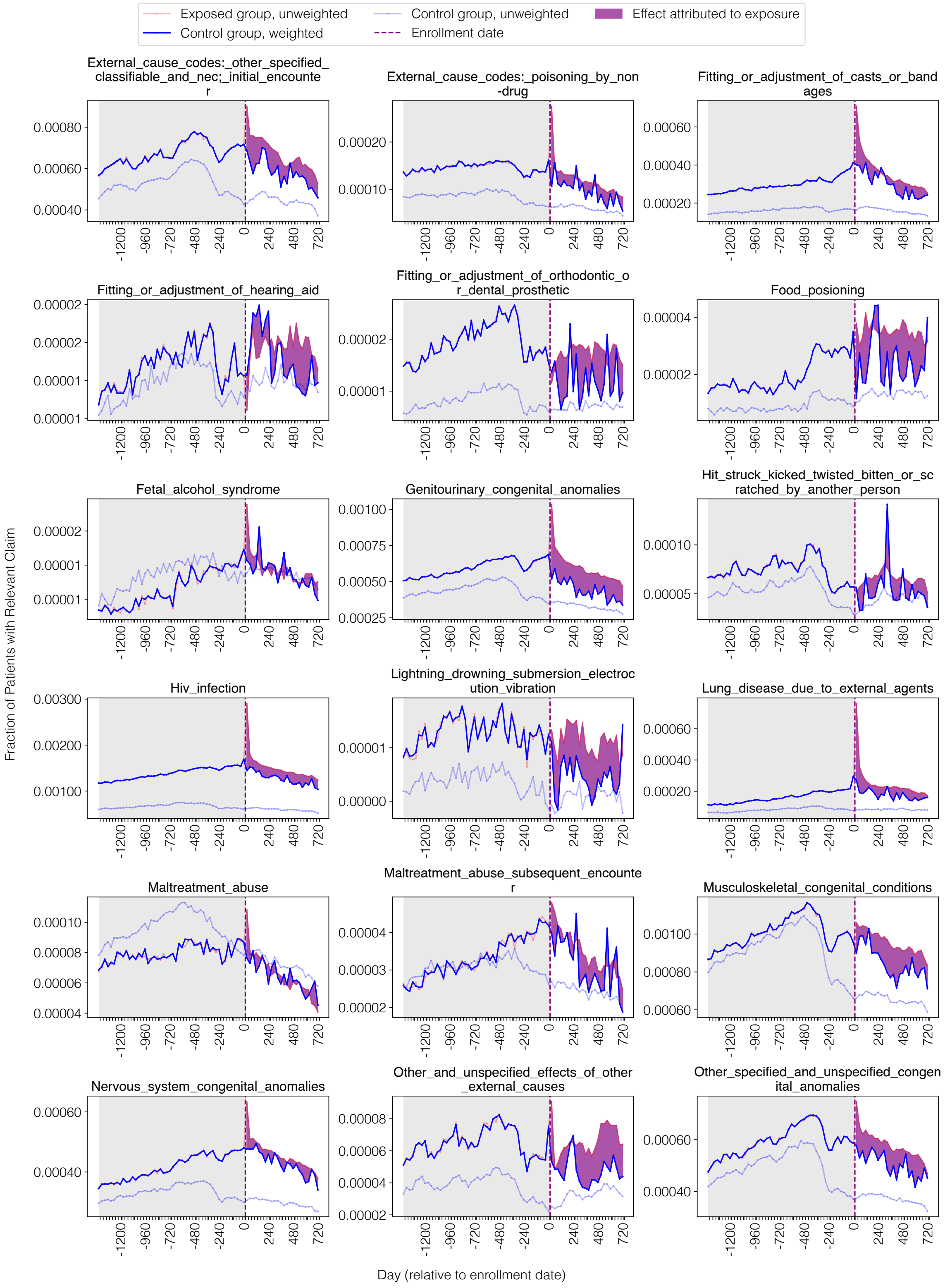


**Supplementary Figure S137:** (cont'd) Temporal trends in Negative prevalence. The fraction of patients with relevant claims over time belonging for each outcome in the **conventional retrospective** study experiment.

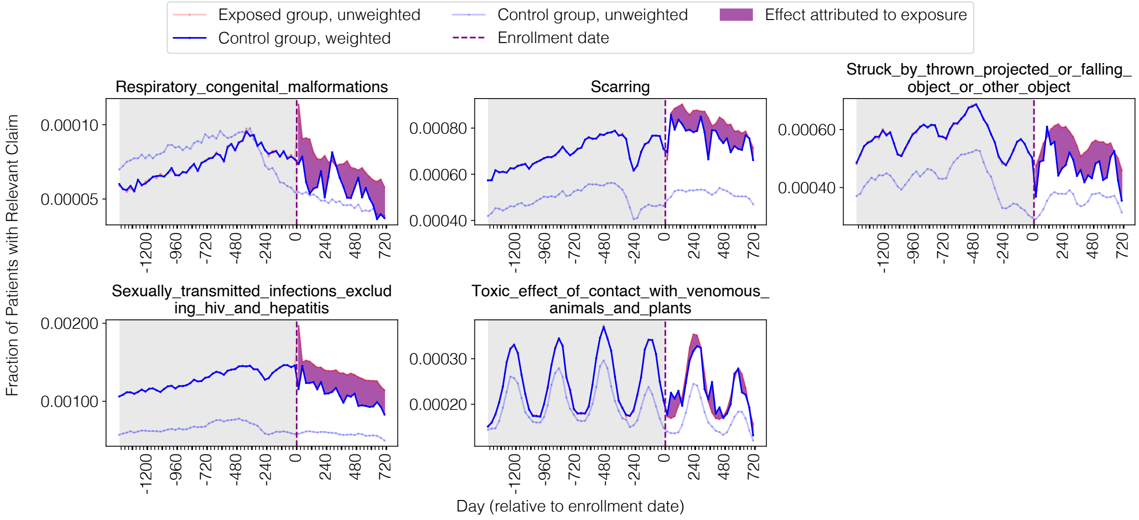


**Supplementary Figure S137:** (cont'd) Temporal trends in Negative prevalence. The fraction of patients with relevant claims over time belonging for each outcome in the **conventional retrospective** study experiment.

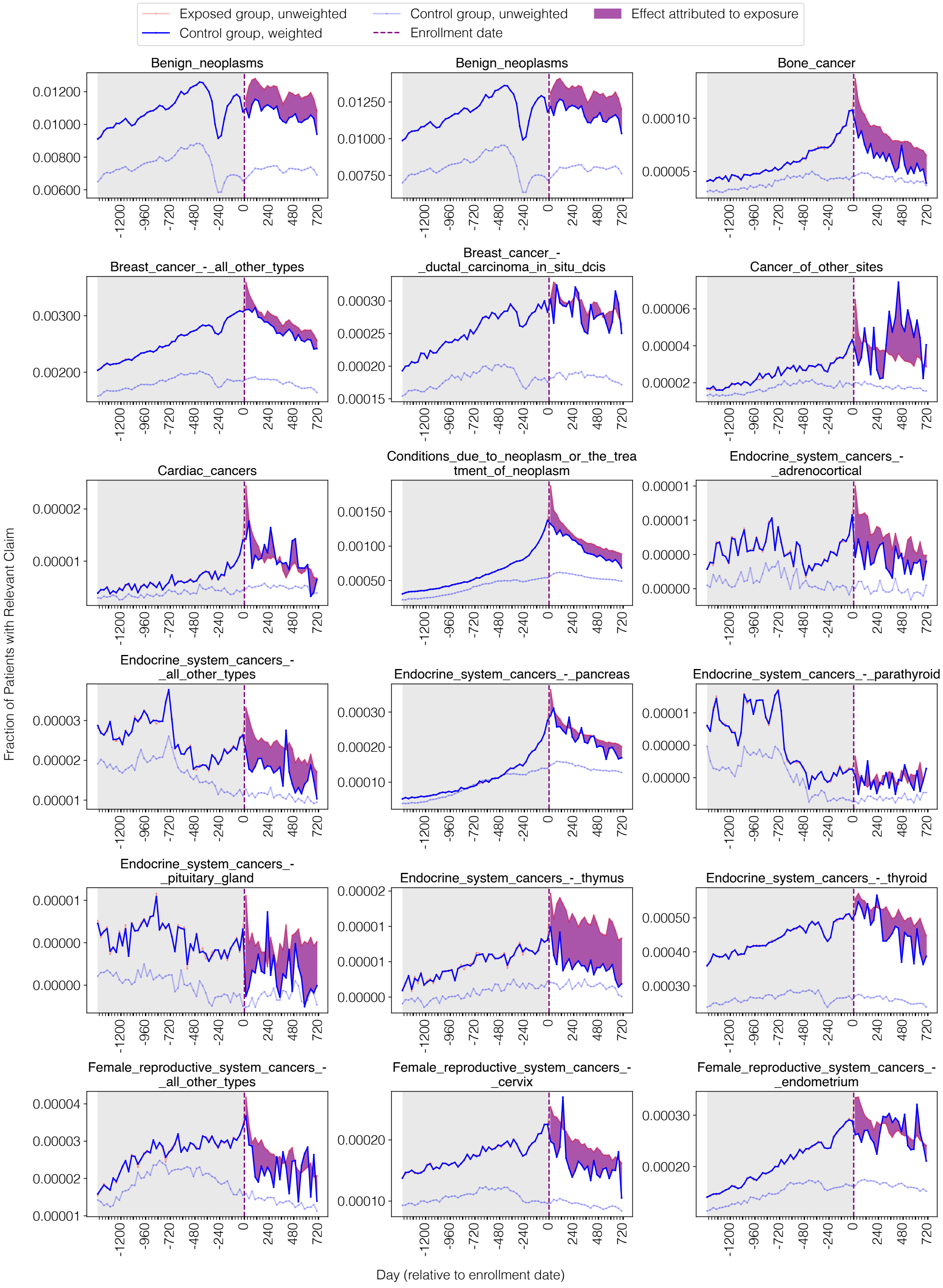


**Supplementary Figure S138:** Temporal trends in Neoplasms prevalence. The fraction of patients with relevant claims over time belonging for each outcome in the **conventional retrospective** study experiment.

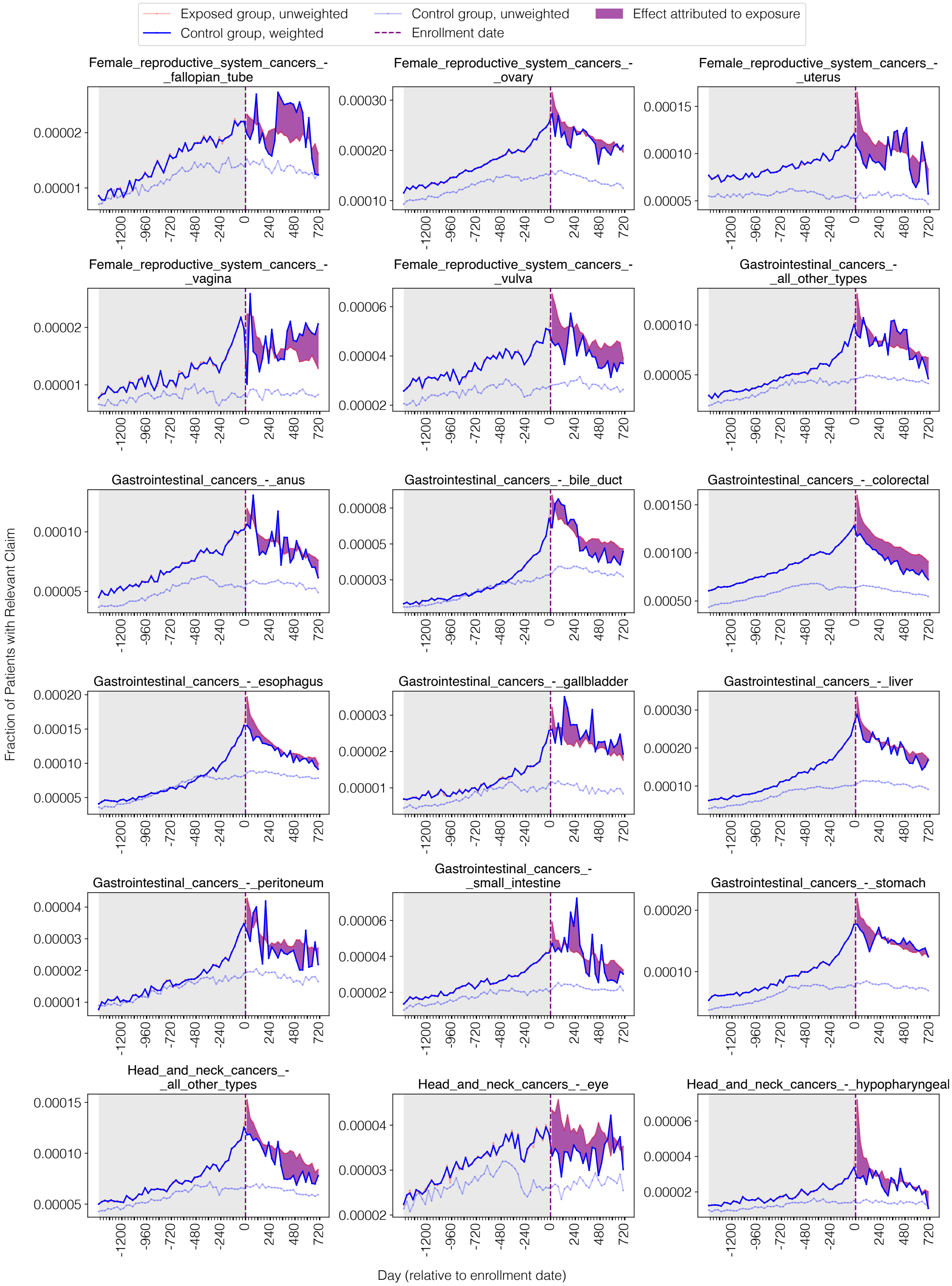


**Supplementary Figure S138:** (cont'd) Temporal trends in Neoplasms prevalence. The fraction of patients with relevant claims over time belonging for each outcome in the **conventional retrospective** study experiment.

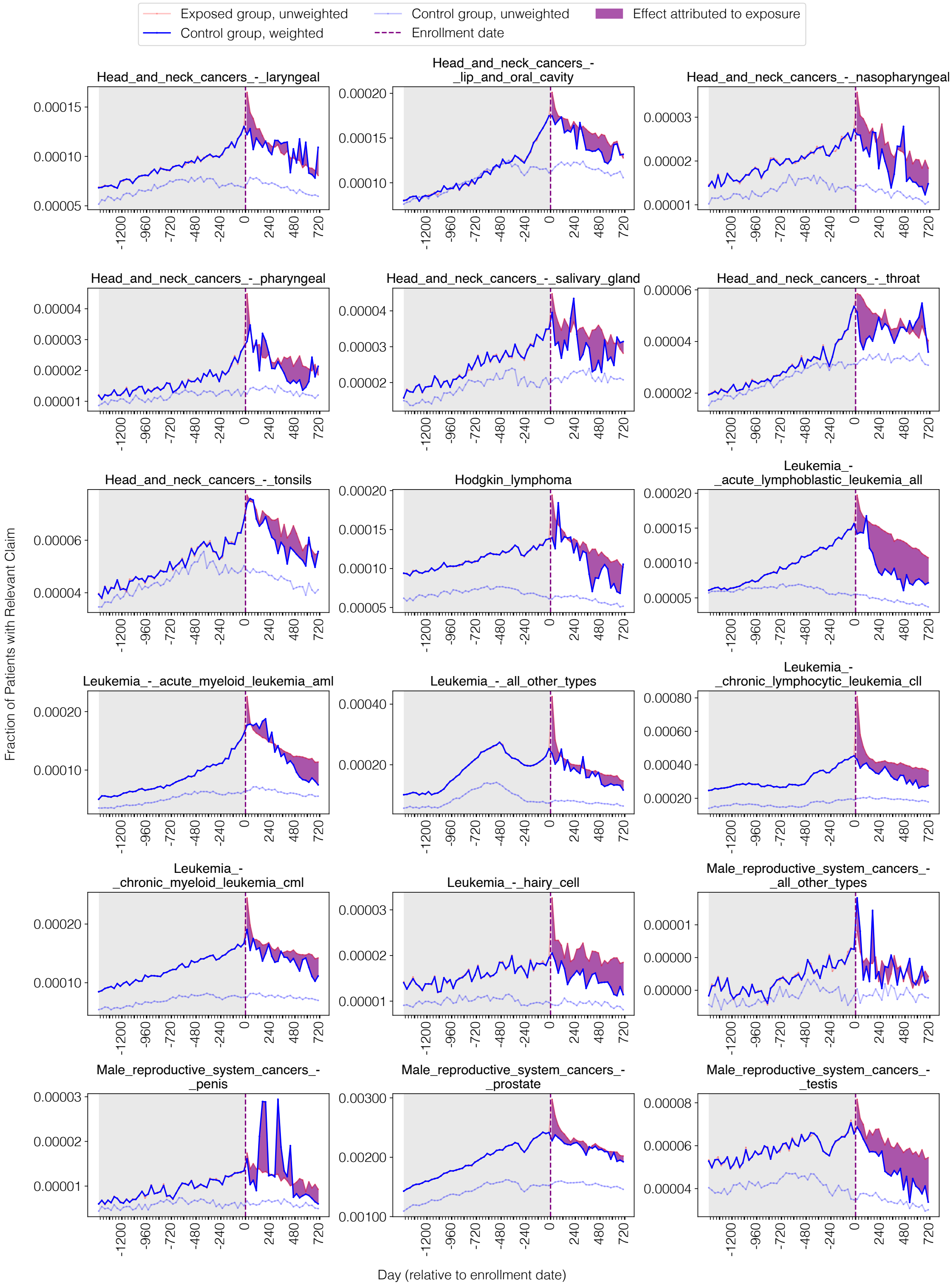


**Supplementary Figure S138:** (cont'd) Temporal trends in Neoplasms prevalence. The fraction of patients with relevant claims over time belonging for each outcome in the **conventional retrospective** study experiment.

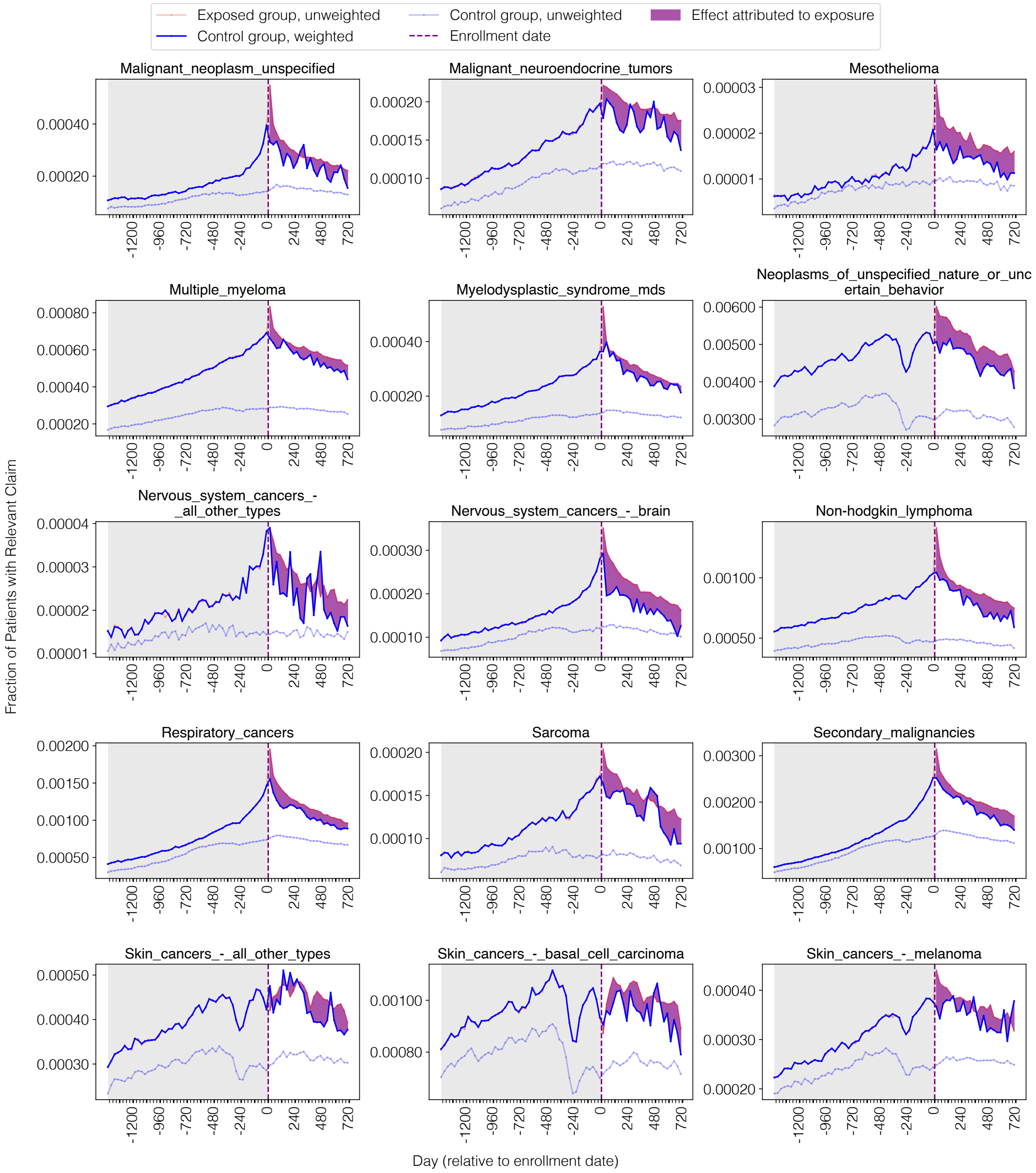


**Supplementary Figure S138:** (cont'd) Temporal trends in Neoplasms prevalence. The fraction of patients with relevant claims over time belonging for each outcome in the **conventional retrospective** study experiment.

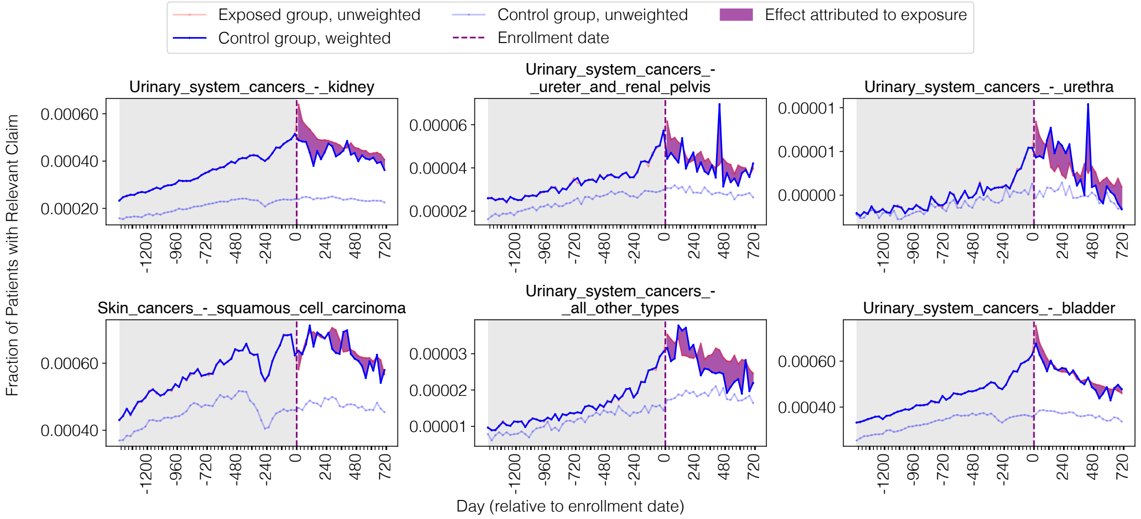


**Supplementary Figure S138:** (cont'd) Temporal trends in Neoplasms prevalence. The fraction of patients with relevant claims over time belonging for each outcome in the **conventional retrospective** study experiment.

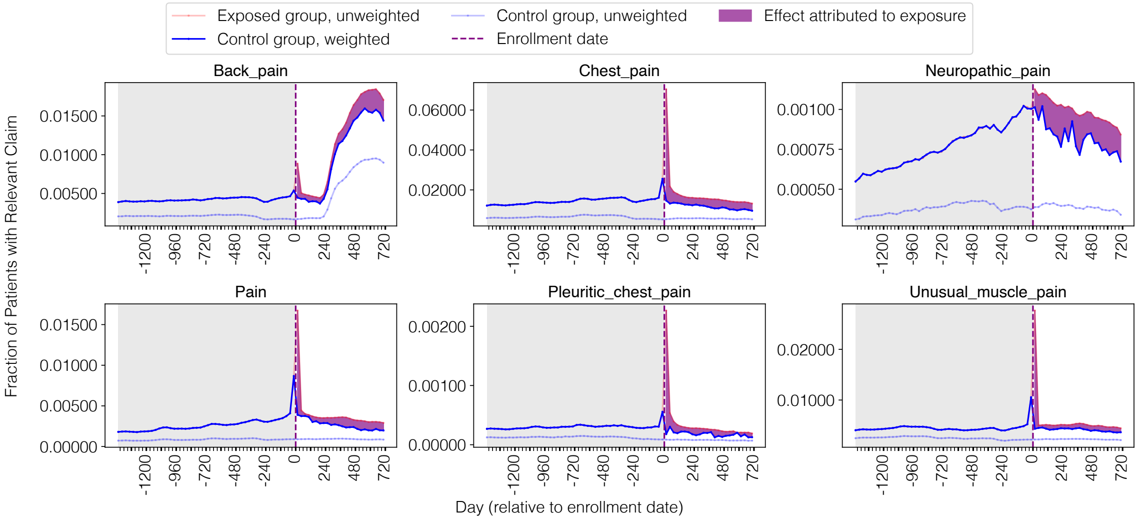


**Supplementary Figure S139:** Temporal trends in Pain prevalence. The fraction of patients with relevant claims over time belonging for each outcome in the **conventional retrospective** study experiment.

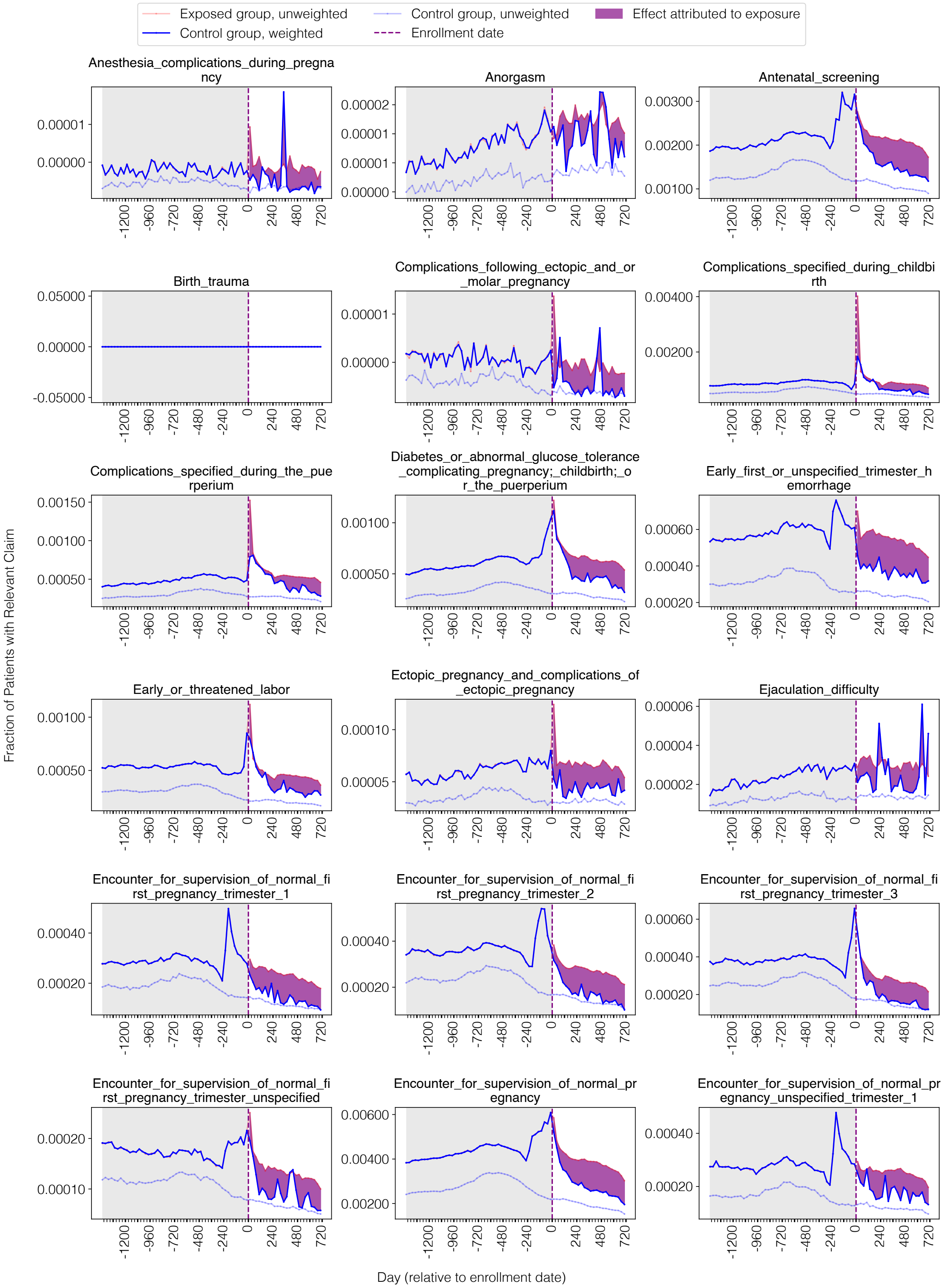


**Supplementary Figure S140:** Temporal trends in Reproductive health prevalence. The fraction of patients with relevant claims over time belonging for each outcome in the **conventional retrospective** study experiment.

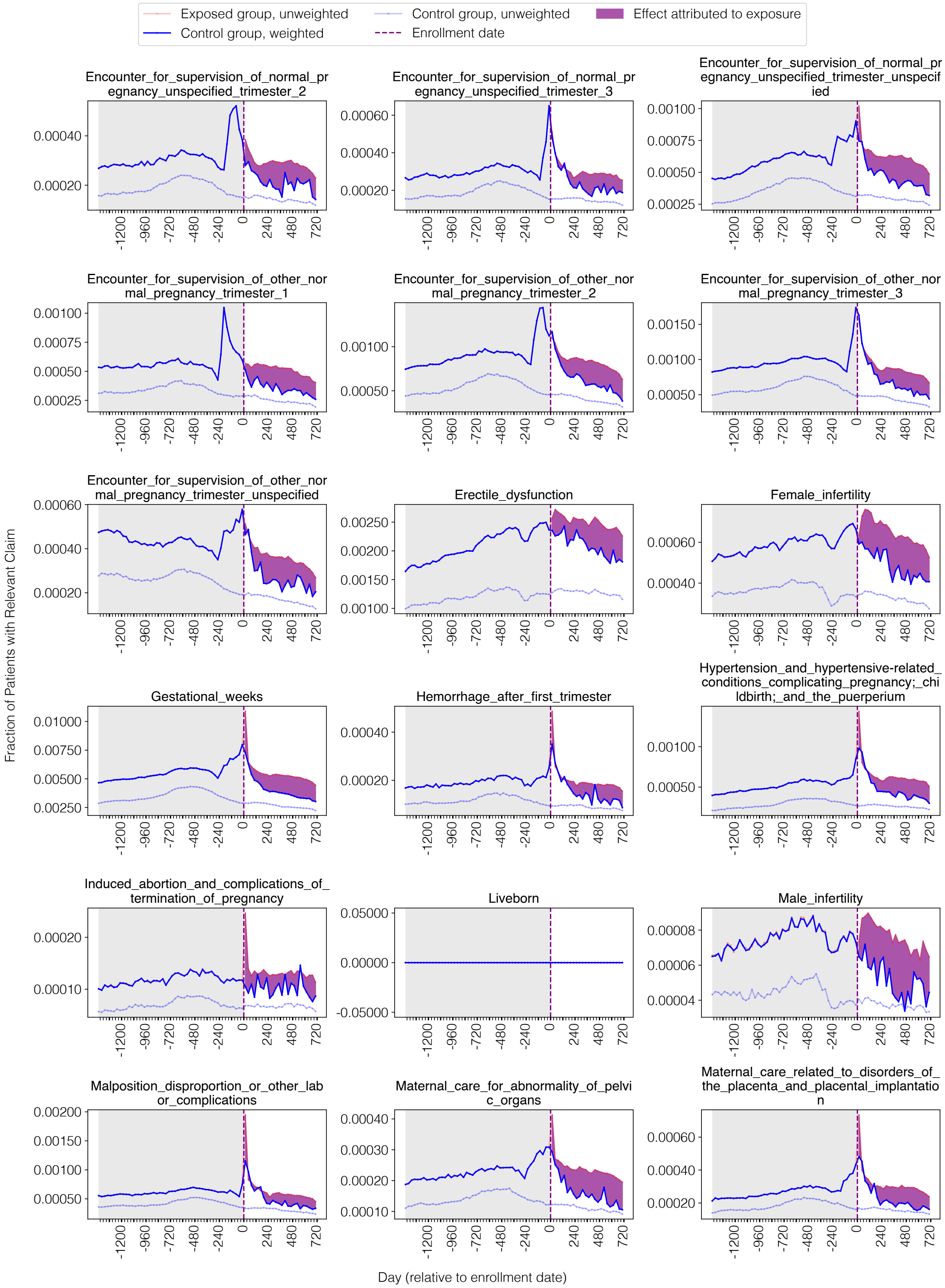


**Supplementary Figure S140:** (cont'd) Temporal trends in Reproductive health prevalence. The fraction of patients with relevant claims over time belonging for each outcome in the **conventional retrospective** study experiment.

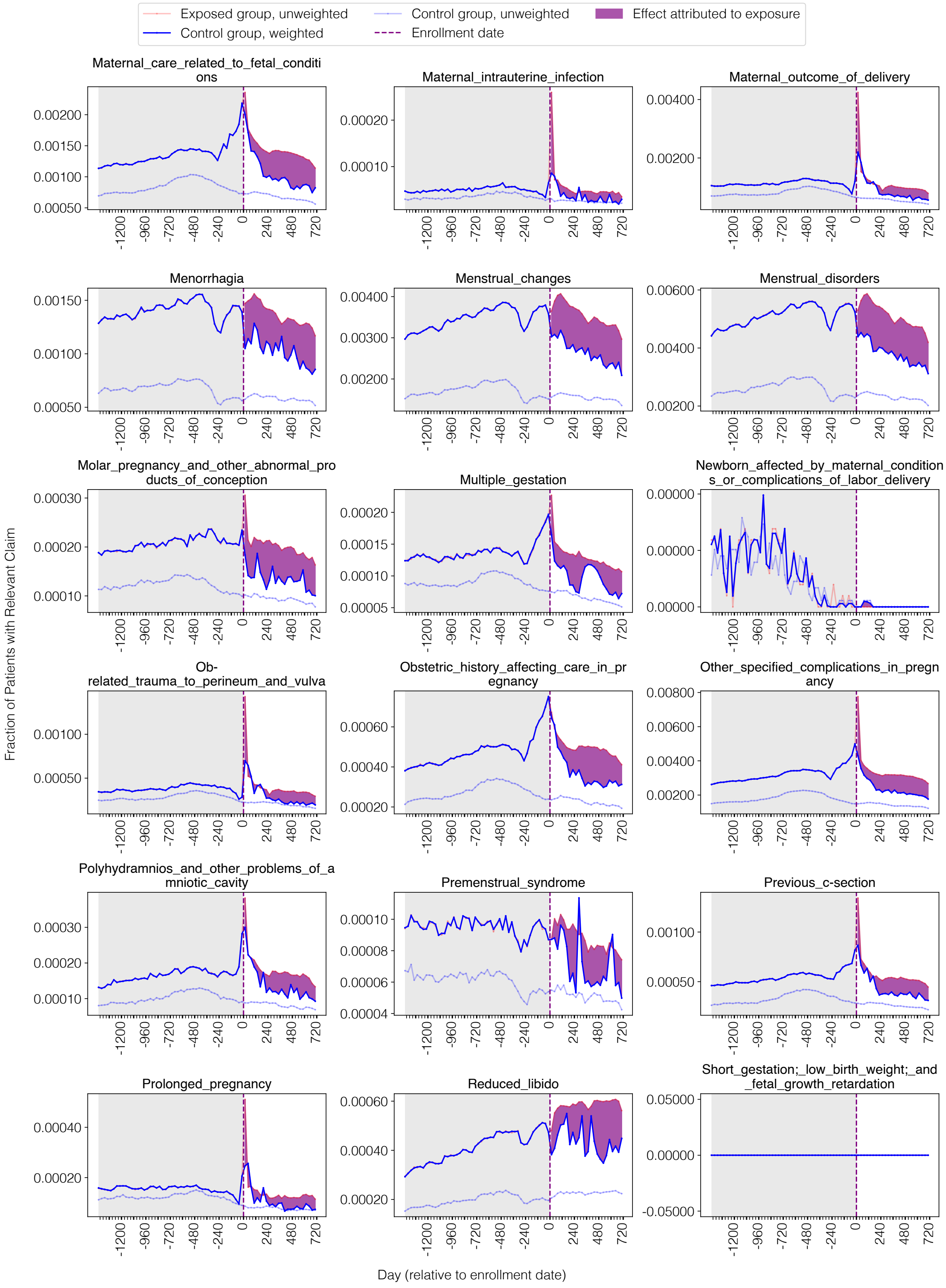


**Supplementary Figure S140:** (cont'd) Temporal trends in Reproductive health prevalence. The fraction of patients with relevant claims over time belonging for each outcome in the **conventional retrospective** study experiment.

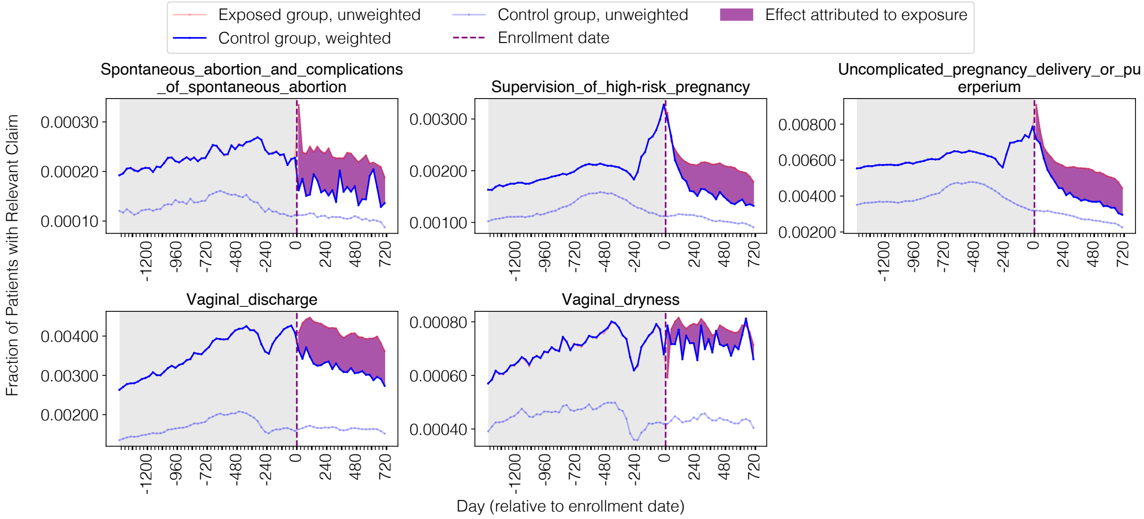


**Supplementary Figure S140:** (cont'd) Temporal trends in Reproductive health prevalence. The fraction of patients with relevant claims over time belonging for each outcome in the **conventional retrospective** study experiment.

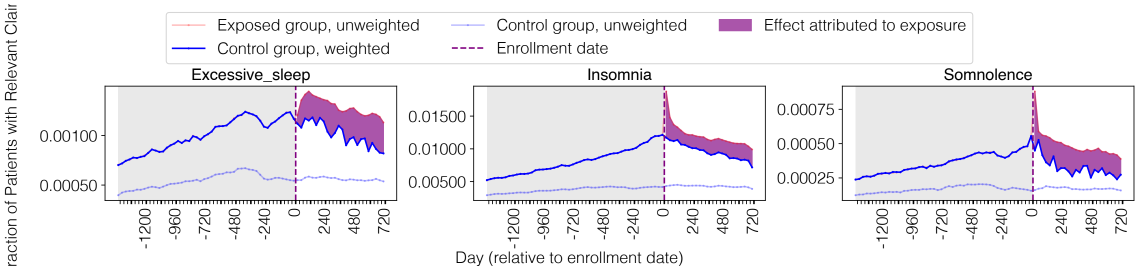


**Supplementary Figure S141:** Temporal trends in Sleep prevalence. The fraction of patients with relevant claims over time belonging for each outcome in the **conventional retrospective** study experiment.

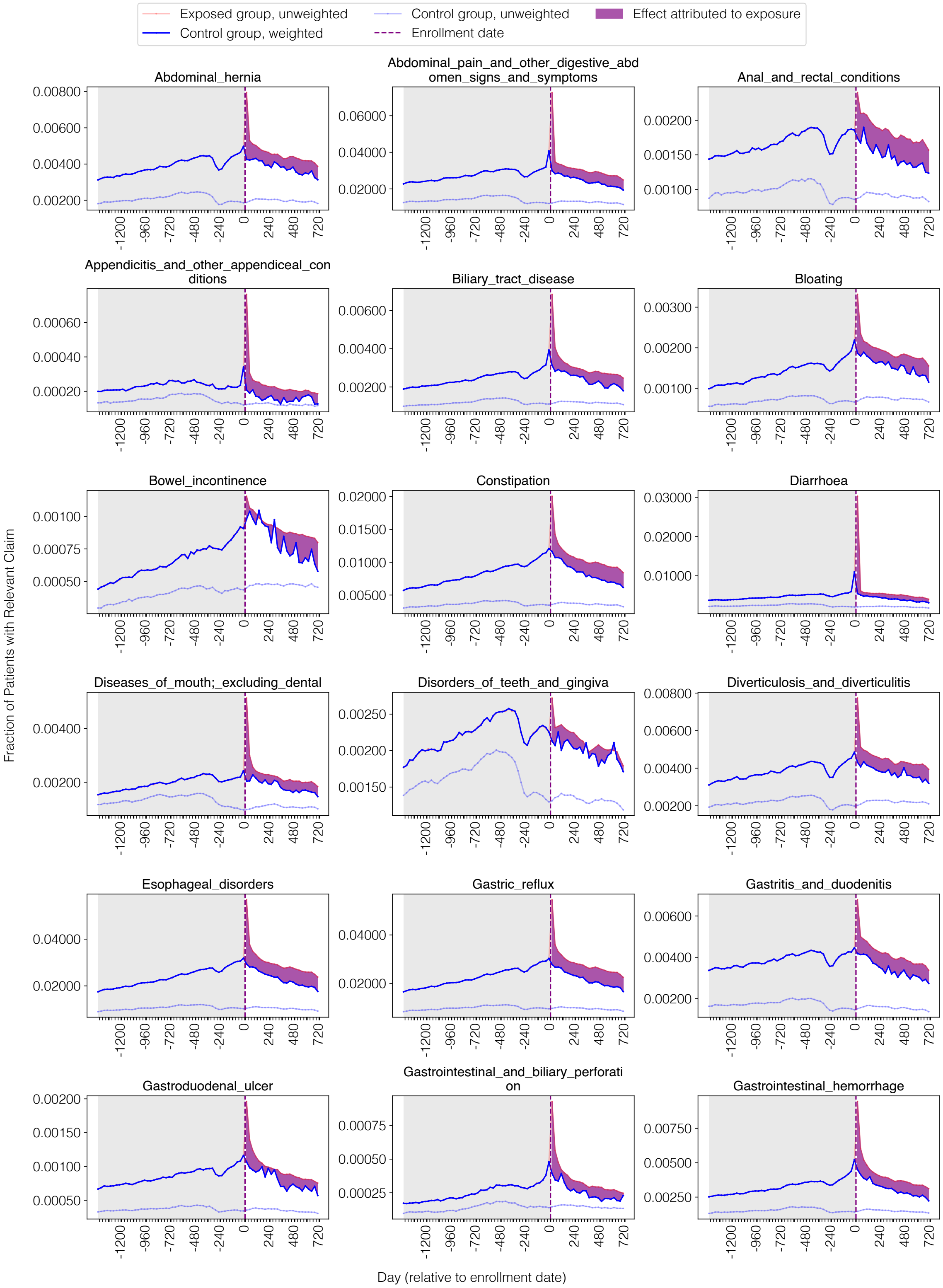


**Supplementary Figure S142:** Temporal trends in Stomach and digestion prevalence. The fraction of patients with relevant claims over time belonging for each outcome in the **conventional retrospective** study experiment.

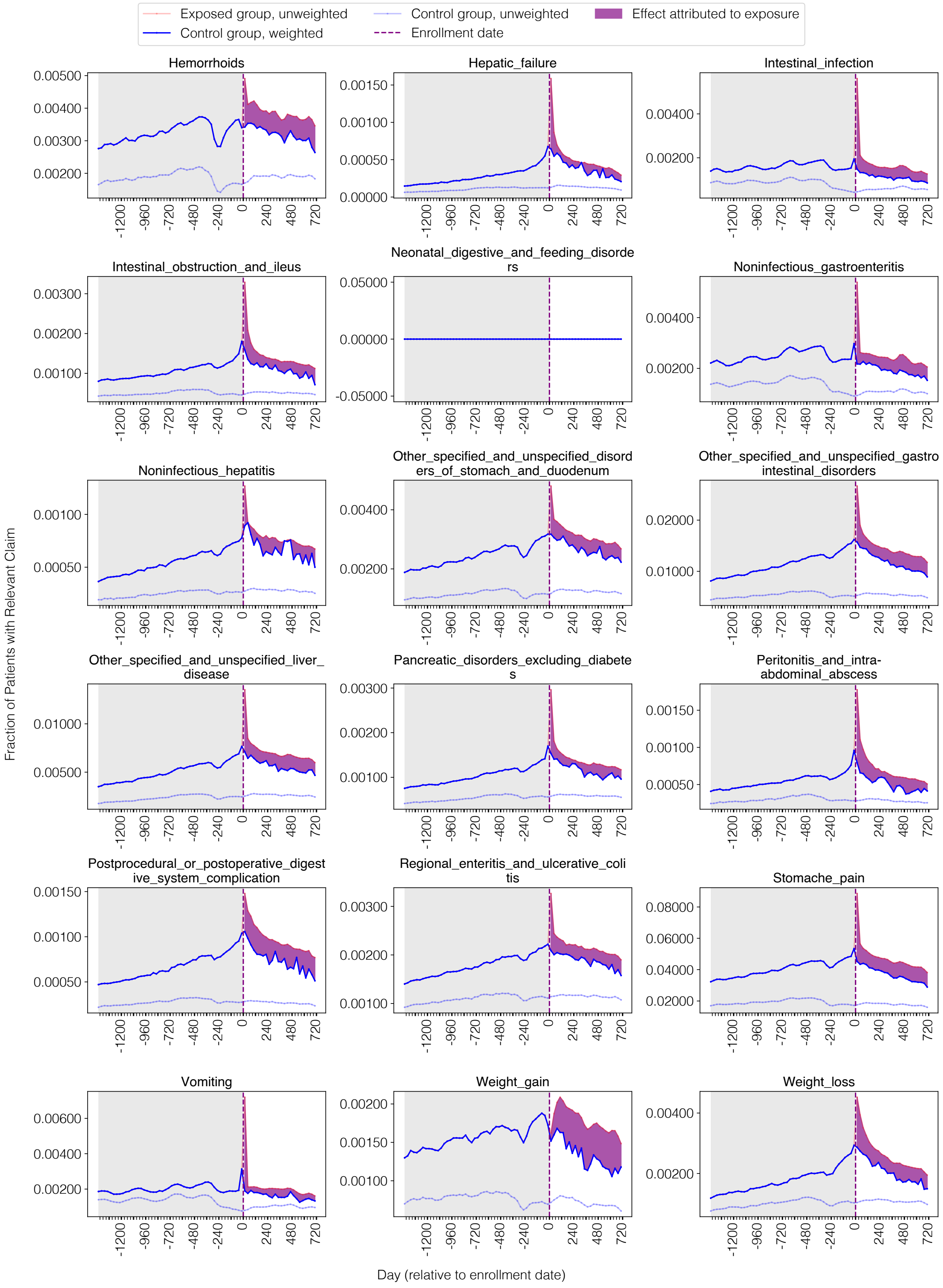


**Supplementary Figure S142:** (cont'd) Temporal trends in Stomach and digestion prevalence. The fraction of patients with relevant claims over time belonging for each outcome in the **conventional retrospective** study experiment.

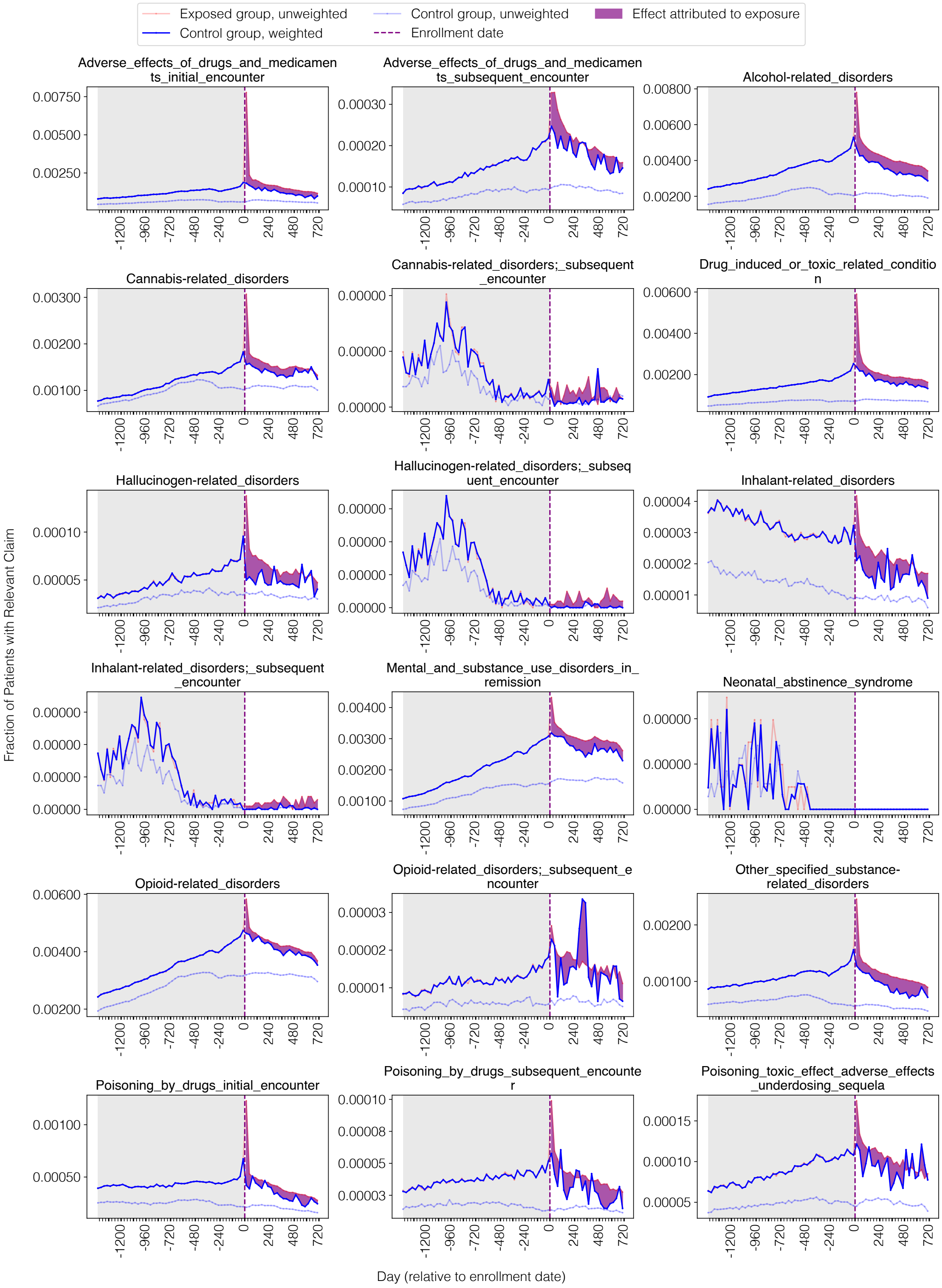


**Supplementary Figure S143:** Temporal trends in Substance use prevalence. The fraction of patients with relevant claims over time belonging for each outcome in the **conventional retrospective** study experiment.

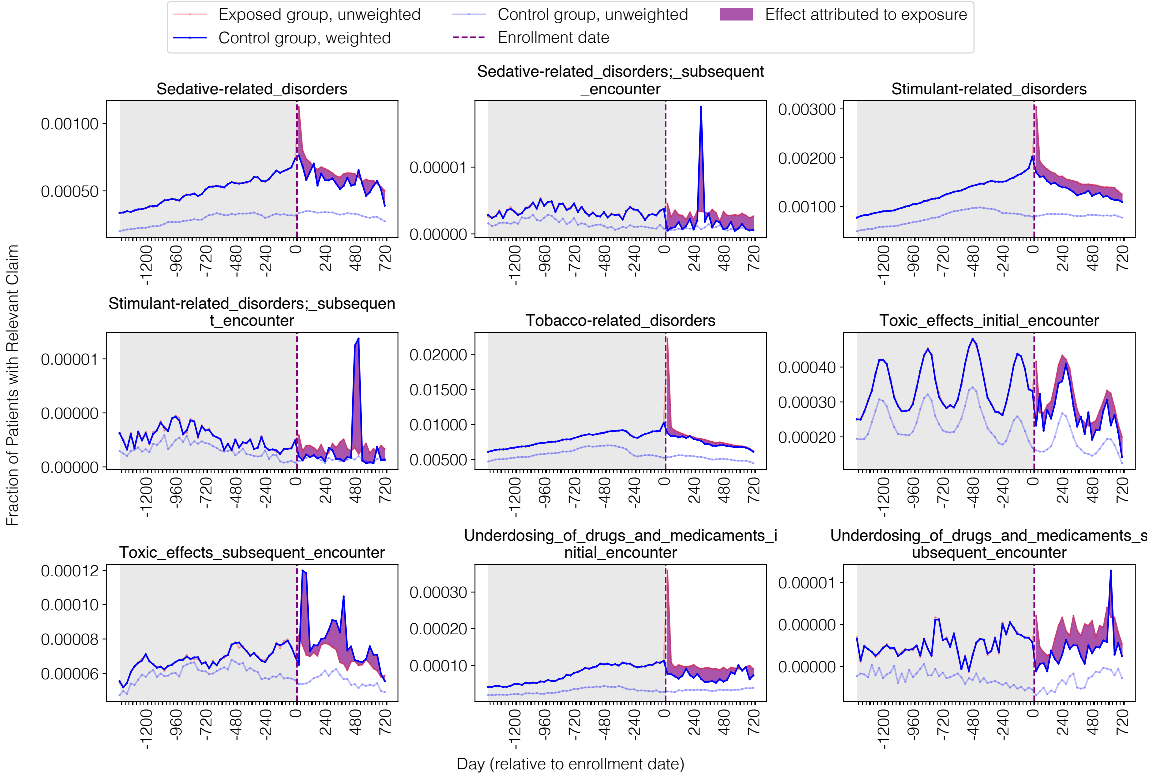


**Supplementary Figure S143:** (cont'd) Temporal trends in Substance use prevalence. The fraction of patients with relevant claims over time belonging for each outcome in the **conventional retrospective** study experiment.

**Supplementary Table S1:** A comprehensive mapping of diagnosis groups used in the experiments to their corresponding ICD-10 and ICD-9 codes.

| Diagnosis | ICD-10 | ICD-9 |
|---|---|---|
| Abdominal hernia | K4000,K4001,K4010,K4011,K4020,K4021,K4030,K4031,K4040,K4041 K4090,K4091,K4100,K4101,K4110,K4111,K4120,K4121,K4130,K4131 K4140,K4141,K4190,K4191,K420,K421,K429,K430,K431,K432,K433 K434,K435,K436,K437,K439,K440,K441,K449,K450,K451,K458,K460 K461,K469 | 55000,55001,55002,55003,55010,55011,55012,55013,55090,55091 55092,55093,55100,55101,55102,55103,5511,55120,55121,55129 5513,5518,5519,55200,55201,55202,55203,5521,55220,55221,55229 5523,5528,5529,55300,55301,55302,55303,5531,55320,55321,55329 5533,5538,5539 |
| Abdominal pain and other digestive abdomen signs and symptoms | G43D0,G43D1,R100,R1010,R1011,R1012,R1013,R102,R1030,R1031 R1032,R1033,R10811,R10812,R10813,R10814,R10815,R10816,R10817 R10819,R10821,R10822,R10823,R10824,R10825,R10826,R10827 R10829,R1083,R1084,R109,R12,R130,R140,R141,R142,R143,R150 R151,R152,R159,R160,R161,R162,R17,R180,R188,R1900,R1901,R1902 R1903,R1904,R1905,R1906,R1907,R1909,R1911,R1912,R1915,R192 R1930,R1931,R1932,R1933,R1934,R1935,R1936,R1937,R194,R195 R196,R197,R198 | 34620,34621,56882,6089,6259,7824,78499,7871,78720,7873,7874 7875,78760,78761,78762,78763,7877,78791,78799,78900,78901 78902,78903,78904,78905,78906,78907,78909,7891,7892,78930 78931,78932,78933,78934,78935,78936,78937,78939,78940,78941 78942,78943,78944,78945,78946,78947,78949,78951,78959,78960 78961,78962,78963,78964,78965,78966,78967,78969,7897,7899 7921 |
| Abnormal findings related to substance use | R780,R781,R782,R783,R784,R785,R786 | 7903,79099 |
| Abnormal findings without diagnosis | R700,R701,R710,R718,R7301,R7302,R7303,R7309,R739,R740,R7401 R7402,R748,R749,R75,R760,R7611,R7612,R768,R769,R770,R771,R772 R778,R779,R7871,R7879,R7881,R7889,R789,R790,R791,R7981,R7982 R7983,R7989,R799,R81,R830,R831,R832,R833,R834,R835,R836,R838 R839,R840,R841,R842,R843,R844,R845,R846,R847,R848,R849,R850 R851,R852,R853,R854,R855,R85610,R85611,R85612,R85613,R85614 R85615,R85616,R85618,R85619,R8569,R857,R8581,R8582,R8589,R859 R860,R861,R862,R863,R864,R865,R866,R867,R868,R869,R870,R871 R872,R873,R874,R875,R87610,R87611,R87612,R87613,R87614,R87615 R87616,R87618,R87619,R87620,R87621,R87622,R87623,R87624 R87625,R87628,R87629,R8769,R877,R87810,R87811,R87820,R87821 R8789,R879,R880,R888,R890,R891,R892,R893,R894,R895,R896,R897 R898,R899,R900,R9081,R9082,R9089,R911,R918,R920,R921,R922 R928,R930,R931,R932,R933,R934,R9341,R93421,R93422,R93429 R9349,R935,R936,R937,R938,R93811,R93812,R93813,R93819,R9389 R939,R9401,R9402,R9409,R94110,R94111,R94112,R94113,R94118 R94120,R94121,R94128,R94130,R94131,R94138,R942,R9430,R9431 R9439,R944,R945,R946,R947,R948,R970,R971,R972,R9720,R9721 R978 | 77183,7842,79001,79009,7901,79021,79022,79029,7904,7905,7906 7907,79091,79092,79093,79095,79099,7915,7920,7922,7924,7925 7929,7930,79311,79319,7932,7933,7934,7935,7936,7937,79380 79381,79382,79389,79391,79399,79400,79401,79402,79409,79410 79411,79412,79413,79414,79415,79416,79417,79419,7942,79430 79431,79439,7944,7945,7946,7947,7948,7949,79501,79502,79503 79504,79505,79506,79507,79508,79509,79510,79511,79512,79513 79514,79515,79516,79518,79519,7952,79539,7954,79551,79552 7956,79571,79579,79581,79582,79589,7960,79670,79671,79672 79673,79674,79675,79676,79677,79678,79679 |
| Acquired absence of | Z89011,Z89012,Z89019,Z89021,Z89022,Z89029,Z89111,Z89112 | V4571,V4572,V4573,V4574,V4575,V4576,V4577,V4578,V4579,V4960 |

Continued on next page

**Supplementary Table S1:** (cont'd) A comprehensive mapping of diagnosis groups used in the experiments to their corresponding ICD-10 and ICD-9 codes.

| Diagnosis | ICD-10 | ICD-9 |
|---|---|---|
| limb or organ | Z89119,Z89121,Z89122,Z89129,Z89201,Z89202,Z89209,Z89211 Z89212,Z89219,Z89221,Z89222,Z89229,Z89231,Z89232,Z89239 Z89411,Z89412,Z89419,Z89421,Z89422,Z89429,Z89431,Z89432 Z89439,Z89441,Z89442,Z89449,Z89511,Z89512,Z89519,Z89521 Z89522,Z89529,Z89611,Z89612,Z89619,Z89621,Z89622,Z89629,Z899 Z9001,Z9002,Z9009,Z9010,Z9011,Z9012,Z9013,Z902,Z903,Z90410 Z90411,Z9049,Z905,Z906,Z90710,Z90711,Z90712,Z90721,Z90722 Z9079,Z9081,Z9089 | V4961,V4962,V4963,V4964,V4965,V4966,V4967,V4970,V4971,V4972 V4973,V4974,V4975,V4976,V4977,V8801,V8802,V8803,V8811,V8812 V8821,V8822 |
| Acquired deformities (excluding foot) | M20001,M20002,M20009,M20011,M20012,M20019,M20021,M20022 M20029,M20031,M20032,M20039,M20091,M20092,M20099,M2100,M21021 M21022,M21029,M21051,M21052,M21059,M21061,M21062,M21069,M2110 M21121,M21122,M21129,M21151,M21152,M21159,M21161,M21162 M21169,M2120,M21211,M21212,M21219,M21221,M21222,M21229,M21231 M21232,M21239,M21241,M21242,M21249,M21251,M21252,M21259 M21261,M21262,M21269,M21271,M21272,M21279,M21331,M21332 M21339,M21511,M21512,M21519,M21521,M21522,M21529,M2170,M21721 M21722,M21729,M21731,M21732,M21733,M21734,M21739,M21751 M21752,M21759,M21761,M21762,M21763,M21764,M21769,M2180,M21821 M21822,M21829,M21831,M21832,M21839,M21851,M21852,M21859 M21861,M21862,M21869,M2190,M21921,M21922,M21929,M21931,M21932 M21939,M21941,M21942,M21949,M21951,M21952,M21959,M21961 M21962,M21969,M2450,M24511,M24512,M24519,M24521,M24522,M24529 M24531,M24532,M24539,M24541,M24542,M24549,M24551,M24552 M24559,M24561,M24562,M24569,M24571,M24572,M24573,M2459,M950 M9510,M9511,M9512,M952,M953,M954,M955,M958,M959 | 71840,71841,71842,71843,71844,71845,71846,71847,71848,71849 73600,73601,73602,73605,73606,73607,73609,7361,73620,73621 73622,73629,73630,73631,73632,73639,73641,73642,7366,73670 73681,73689,7369,7380,73819,7382,7383,7386,7387,7388,7389 |
| Acquired foot deformities | M2010,M2011,M2012,M2020,M2021,M2022,M2030,M2031,M2032,M2040 M2041,M2042,M205X1,M205X2,M205X9,M2060,M2061,M2062,M21071 M21072,M21079,M21171,M21172,M21179,M21371,M21372,M21379,M2140 M2141,M2142,M21531,M21532,M21539,M21541,M21542,M21549,M21611 M21612,M21619,M21621,M21622,M21629,M216X1,M216X2,M216X9 M24574,M24575,M24576 | 71847,7271,734,7350,7351,7352,7354,7358,7359,73671,73674 73679 |
| Acute and chronic tonsillitis | J0300,J0301,J0380,J0381,J0390,J0391,J3501,J3502,J3503,J351 J352,J353,J358,J359,J36 | 0340,463,47400,47401,47402,47410,47411,47412,4742,4748,4749 475 |
| Acute and unspecified renal failure | N170,N171,N172,N178,N179,N19,N990,O904 | 5836,5837,5845,5846,5847,5848,5849,586,66932,66934,9975 |

Continued on next page

**Supplementary Table S1:** (cont'd) A comprehensive mapping of diagnosis groups used in the experiments to their corresponding ICD-10 and ICD-9 codes.

| Diagnosis | ICD-10 | ICD-9 |
|---|---|---|
| Acute bronchitis | J200,J201,J202,J203,J204,J205,J206,J207,J208,J209,J210,J211<br>J218,J219,J40 | 04100,0415,04181,0791,0792,0793,0796,07989,4660,46611,46619<br>490 |
| Acute hemorrhagic cerebrovascular disease | I6000,I6001,I6002,I6010,I6011,I6012,I602,I6020,I6021,I6022<br>I6030,I6031,I6032,I604,I6050,I6051,I6052,I606,I607,I608,I609<br>I610,I611,I612,I613,I614,I615,I616,I618,I619,I6200,I6201<br>I6202,I6203,I621,I629 | 430,431,4320,4321,4329 |
| Acute myocardial infarction | I2101,I2102,I2109,I2111,I2119,I2121,I2129,I213,I214,I219<br>I21A1,I21A9,I220,I221,I222,I228,I229 | 41001,41011,41021,41031,41041,41051,41061,41071,41081,41091 |
| Acute phlebitis; thrombophlebitis and thromboembolism | I8000,I8001,I8002,I8003,I8010,I8011,I8012,I8013,I80201,I80202<br>I80203,I80209,I80211,I80212,I80213,I80219,I80221,I80222<br>I80223,I80229,I80231,I80232,I80233,I80239,I80241,I80242<br>I80243,I80249,I80251,I80252,I80253,I80259,I80291,I80292<br>I80293,I80299,I803,I808,I809,I81,I820,I821,I82210,I82220<br>I82290,I823,I82401,I82402,I82403,I82409,I82411,I82412,I82413<br>I82419,I82421,I82422,I82423,I82429,I82431,I82432,I82433<br>I82439,I82441,I82442,I82443,I82449,I82451,I82452,I82453<br>I82459,I82461,I82462,I82463,I82469,I82491,I82492,I82493<br>I82499,I824Y1,I824Y2,I824Y3,I824Y9,I824Z1,I824Z2,I824Z3<br>I824Z9,I82601,I82602,I82603,I82609,I82611,I82612,I82613<br>I82619,I82621,I82622,I82623,I82629,I82811,I82812,I82813<br>I82819,I82890,I8290,I82A11,I82A12,I82A13,I82A19,I82B11,I82B12<br>I82B13,I82B19,I82C11,I82C12,I82C13,I82C19,O2220,O2221,O2222<br>O2223,O2230,O2231,O2232,O2233,O8681,O870,O871 | 4510,45111,45119,4512,45181,45189,4519,452,4530,4531,4532<br>4533,45340,45341,45342,4536,45381,45382,45383,45384,45385<br>45386,45387,45389,67032,67034,67120,67121,67122,67123,67124<br>67130,67131,67133,67142,67144 |
| Acute posthemorrhagic anemia | D62 | 2851 |
| Acute pulmonary embolism | I2601,I2602,I2609,I2690,I2692,I2693,I2694,I2699 | 4150,41512,41513,41519 |
| Acute rheumatic heart disease | I00,I010,I011,I012,I018,I019,I020,I029 | 390,3910,3911,3912,3918,3919,3920,3929 |
| Adverse effects of drugs and medicaments, initial encounter | T360X5A,T361X5A,T362X5A,T363X5A,T364X5A,T365X5A,T366X5A<br>T367X5A,T368X5A,T3695XA,T370X5A,T371X5A,T372X5A,T373X5A<br>T374X5A,T375X5A,T378X5A,T3795XA,T380X5A,T381X5A,T382X5A<br>T383X5A,T384X5A,T385X5A,T386X5A,T387X5A,T38805A,T38815A | 99520,99522,99523,99529,E9300,E9301,E9302,E9303,E9304,E9305<br>E9306,E9308,E9310,E9314,E9315,E9316,E9317,E9319,E9320,E9321<br>E9322,E9323,E9324,E9327,E9328,E9329,E9330,E9331,E9334,E9335<br>E9336,E9337,E9338,E9339,E9340,E9342,E9344,E9345,E9346,E9348 |

Continued on next page

**Supplementary Table S1:** (cont'd) A comprehensive mapping of diagnosis groups used in the experiments to their corresponding ICD-10 and ICD-9 codes.

| Diagnosis | ICD-10 | ICD-9 |
|---|---|---|
| | T38895A,T38905A,T38995A,T39015A,T39095A,T391X5A,T392X5A<br>T39315A,T39395A,T394X5A,T398X5A,T3995XA,T400X5A,T401X5A<br>T402X5A,T403X5A,T40415A,T40425A,T40495A,T404X5A,T405X5A<br>T40605A,T40695A,T40715A,T40725A,T407X5A,T408X5A,T40905A<br>T40995A,T410X5A,T411X5A,T41205A,T41295A,T413X5A,T4145XA<br>T415X5A,T420X5A,T421X5A,T422X5A,T423X5A,T424X5A,T425X5A<br>T426X5A,T4275XA,T428X5A,T43015A,T43025A,T431X5A,T43205A<br>T43215A,T43225A,T43295A,T433X5A,T434X5A,T43505A,T43595A<br>T43605A,T43615A,T43625A,T43635A,T43655A,T43695A,T438X5A<br>T4395XA,T440X5A,T441X5A,T442X5A,T443X5A,T444X5A,T445X5A<br>T446X5A,T447X5A,T448X5A,T44905A,T44995A,T450X5A,T451X5A<br>T452X5A,T453X5A,T454X5A,T45515A,T45525A,T45605A,T45615A<br>T45625A,T45695A,T457X5A,T458X5A,T4595XA,T460X5A,T461X5A<br>T462X5A,T463X5A,T464X5A,T465X5A,T466X5A,T467X5A,T468X5A<br>T46905A,T46995A,T470X5A,T471X5A,T472X5A,T473X5A,T474X5A<br>T475X5A,T476X5A,T477X5A,T478X5A,T4795XA,T480X5A,T481X5A<br>T48205A,T48295A,T483X5A,T484X5A,T485X5A,T486X5A,T48905A<br>T48995A,T490X5A,T491X5A,T492X5A,T493X5A,T494X5A,T495X5A<br>T496X5A,T497X5A,T498X5A,T4995XA,T500X5A,T501X5A,T502X5A<br>T503X5A,T504X5A,T505X5A,T506X5A,T507X5A,T508X5A,T50905A<br>T50915A,T50995A,T50A15A,T50A25A,T50A95A,T50B15A,T50B95A<br>T50Z15A,T50Z95A | E9349,E9350,E9351,E9352,E9353,E9355,E9356,E9357,E9360,E9361<br>E9362,E9363,E9364,E9370,E9378,E9379,E9382,E9383,E9384,E9389<br>E9390,E9391,E9392,E9393,E9394,E9396,E9397,E9398,E9399,E9400<br>E9401,E9410,E9411,E9412,E9413,E9419,E9420,E9421,E9422,E9424<br>E9425,E9426,E9427,E9429,E9430,E9431,E9432,E9433,E9434,E9435<br>E9436,E9438,E9439,E9442,E9444,E9445,E9447,E9450,E9452,E9453<br>E9454,E9455,E9456,E9457,E9458,E9460,E9461,E9462,E9463,E9464<br>E9465,E9466,E9467,E9468,E9469,E9470,E9472,E9478,E9479,E9486<br>E9488,E9489,E9490,E9496,E9499 |
| Adverse effects of drugs and medicaments, subsequent encounter | T360X5D,T361X5D,T362X5D,T363X5D,T364X5D,T365X5D,T366X5D<br>T367X5D,T368X5D,T3695XD,T370X5D,T371X5D,T372X5D,T373X5D<br>T374X5D,T375X5D,T378X5D,T3795XD,T380X5D,T381X5D,T382X5D<br>T383X5D,T384X5D,T385X5D,T386X5D,T387X5D,T38805D,T38815D<br>T38895D,T38905D,T38995D,T39015D,T39095D,T391X5D,T392X5D<br>T39315D,T39395D,T394X5D,T398X5D,T3995XD,T400X5D,T401X5D<br>T402X5D,T403X5D,T40415D,T40425D,T40495D,T404X5D,T405X5D<br>T40605D,T40695D,T40715D,T40725D,T407X5D,T408X5D,T40905D<br>T40995D,T410X5D,T411X5D,T41205D,T41295D,T413X5D,T4145XD<br>T415X5D,T420X5D,T421X5D,T422X5D,T423X5D,T424X5D,T425X5D<br>T426X5D,T4275XD,T428X5D,T43015D,T43025D,T431X5D,T43205D<br>T43215D,T43225D,T43295D,T433X5D,T434X5D,T43505D,T43595D<br>T43605D,T43615D,T43625D,T43635D,T43655D,T43695D,T438X5D<br>T4395XD,T440X5D,T441X5D,T442X5D,T443X5D,T444X5D,T445X5D<br>T446X5D,T447X5D,T448X5D,T44905D,T44995D,T450X5D,T451X5D<br>T452X5D,T453X5D,T454X5D,T45515D,T45525D,T45605D,T45615D<br>T45625D,T45695D,T457X5D,T458X5D,T4595XD,T460X5D,T461X5D | V5889 |

Continued on next page

**Supplementary Table S1:** (cont'd) A comprehensive mapping of diagnosis groups used in the experiments to their corresponding ICD-10 and ICD-9 codes.

| Diagnosis | ICD-10 | ICD-9 |
|---|---|---|
| | T462X5D,T463X5D,T464X5D,T465X5D,T466X5D,T467X5D,T468X5D<br>T46905D,T46995D,T470X5D,T471X5D,T472X5D,T473X5D,T474X5D<br>T475X5D,T476X5D,T477X5D,T478X5D,T4795XD,T480X5D,T481X5D<br>T48205D,T48295D,T483X5D,T484X5D,T485X5D,T486X5D,T48905D<br>T48995D,T490X5D,T491X5D,T492X5D,T493X5D,T494X5D,T495X5D<br>T496X5D,T497X5D,T498X5D,T4995XD,T500X5D,T501X5D,T502X5D<br>T503X5D,T504X5D,T505X5D,T506X5D,T507X5D,T508X5D,T50905D<br>T50915D,T50995D,T50A15D,T50A25D,T50A95D,T50B15D,T50B95D<br>T50Z15D,T50Z95D | |
| Alcohol-related<br>disorders | F1010,F10120,F10121,F10129,F10130,F10131,F10132,F10139,F1014<br>F10150,F10151,F10159,F10180,F10181,F10182,F10188,F1019,F1020<br>F10220,F10221,F10229,F10230,F10231,F10232,F10239,F1024,F10250<br>F10251,F10259,F1026,F1027,F10280,F10281,F10282,F10288,F1029<br>F1090,F1091,F10920,F10921,F10929,F10930,F10931,F10932,F10939<br>F1094,F10950,F10951,F10959,F1096,F1097,F10980,F10981,F10982<br>F10988,F1099,G312,G621,I426,K2920,K2921,K700,K7010,K7011,K702<br>K7030,K7031,K7040,K7041,K709,O354XX0,O354XX1,O354XX2,O354XX3<br>O354XX4,O354XX5,O354XX9,O99310,O99311,O99312,O99313,O99314<br>O99315 | 2910,2911,2912,2913,2914,2915,29181,29182,29189,2919,30300<br>30301,30302,30303,30390,30391,30392,30500,30501,30502,30503<br>3317,3575,4255,53530,53531,5710,5711,5712,5713,5722,64840<br>64841,64843,64844,65541,65543 |
| Allergic reactions | B4481,H01111,H01112,H01113,H01114,H01115,H01116,H01119,H01131<br>H01132,H01133,H01134,H01135,H01136,H01139,H1045,H65111,H65112<br>H65113,H65114,H65115,H65116,H65117,H65119,H65411,H65412<br>H65413,H65419,J301,J302,J305,J3081,J3089,J309,K0855,K522<br>K5221,K5222,K5229,K9041,L200,L2081,L2082,L2083,L2084,L2089<br>L209,L230,L231,L232,L233,L234,L235,L236,L237,L2381,L2389,L239<br>L270,L271,L272,L278,L279,L300,L302,L308,L309,L500,L501,L502<br>L503,L504,L505,L506,L508,L509,L511,L512,L513,L518,L519,L560<br>L561,L562,L563,L564,M364,T7800XA,T7801XA,T7802XA,T7803XA<br>T7804XA,T7805XA,T7806XA,T7807XA,T7808XA,T7809XA,T781XXA<br>T782XXA,T783XXA,T7840XA,T7841XA,T7849XA,T788XXA | 37214,37331,37332,38102,38103,38104,38105,38106,3813,4770<br>4771,4772,4778,4779,5186,52566,5583,69012,6918,6923,6924,6925<br>6926,69272,69281,69282,69283,69284,69289,6929,6930,6931,6938<br>6939,69510,69511,69512,69513,69514,69515,69519,7080,7081,7082<br>7083,7084,7085,7088,7089,7136,78791,9889,9950,9951,99521,9953<br>99560,99561,99562,99563,99564,99565,99566,99567,99568,99569<br>9957,99589 |
| Allergic reactions,<br>subsequent encounter | T7800XD,T7801XD,T7802XD,T7803XD,T7804XD,T7805XD,T7806XD<br>T7807XD,T7808XD,T7809XD,T781XXD,T782XXD,T783XXD,T7840XD<br>T7841XD,T7849XD,T788XXD | V5889 |
| Amputation of a limb,<br>initial encounter | S48011A,S48012A,S48019A,S48021A,S48022A,S48029A,S48111A<br>S48112A,S48119A,S48121A,S48122A,S48129A,S48911A,S48912A<br>S48919A,S48921A,S48922A,S48929A,S58011A,S58012A,S58019A | 8850,8860,8870,8872,8874,8950,8960,8970,8972,8974 |

Continued on next page

**Supplementary Table S1:** (cont'd) A comprehensive mapping of diagnosis groups used in the experiments to their corresponding ICD-10 and ICD-9 codes.

| Diagnosis | ICD-10 | ICD-9 |
|---|---|---|
| | S58021A,S58022A,S58029A,S58111A,S58112A,S58119A,S58121A S58122A,S58129A,S58911A,S58912A,S58919A,S58921A,S58922A S58929A,S68011A,S68012A,S68019A,S68021A,S68022A,S68029A S68110A,S68111A,S68112A,S68113A,S68114A,S68115A,S68116A S68117A,S68118A,S68119A,S68120A,S68121A,S68122A,S68123A S68124A,S68125A,S68126A,S68127A,S68128A,S68129A,S68411A S68412A,S68419A,S68421A,S68422A,S68429A,S68511A,S68512A S68519A,S68521A,S68522A,S68529A,S68610A,S68611A,S68612A S68613A,S68614A,S68615A,S68616A,S68617A,S68618A,S68619A S68620A,S68621A,S68622A,S68623A,S68624A,S68625A,S68626A S68627A,S68628A,S68629A,S68711A,S68712A,S68719A,S68721A S68722A,S68729A,S78011A,S78012A,S78019A,S78021A,S78022A S78029A,S78111A,S78112A,S78119A,S78121A,S78122A,S78129A S78911A,S78912A,S78919A,S78921A,S78922A,S78929A,S88011A S88012A,S88019A,S88021A,S88022A,S88029A,S88111A,S88112A S88119A,S88121A,S88122A,S88129A,S88911A,S88912A,S88919A S88921A,S88922A,S88929A,S98011A,S98012A,S98019A,S98021A S98022A,S98029A,S98111A,S98112A,S98119A,S98121A,S98122A S98129A,S98131A,S98132A,S98139A,S98141A,S98142A,S98149A S98211A,S98212A,S98219A,S98221A,S98222A,S98229A,S98311A S98312A,S98319A,S98321A,S98322A,S98329A,S98911A,S98912A S98919A,S98921A,S98922A,S98929A | |
| Amputation of a limb, subsequent encounter | S48011D,S48012D,S48019D,S48021D,S48022D,S48029D,S48111D S48112D,S48119D,S48121D,S48122D,S48129D,S48911D,S48912D S48919D,S48921D,S48922D,S48929D,S58011D,S58012D,S58019D S58021D,S58022D,S58029D,S58111D,S58112D,S58119D,S58121D S58122D,S58129D,S58911D,S58912D,S58919D,S58921D,S58922D S58929D,S68011D,S68012D,S68019D,S68021D,S68022D,S68029D S68110D,S68111D,S68112D,S68113D,S68114D,S68115D,S68116D S68117D,S68118D,S68119D,S68120D,S68121D,S68122D,S68123D S68124D,S68125D,S68126D,S68127D,S68128D,S68129D,S68411D S68412D,S68419D,S68421D,S68422D,S68429D,S68511D,S68512D S68519D,S68521D,S68522D,S68529D,S68610D,S68611D,S68612D S68613D,S68614D,S68615D,S68616D,S68617D,S68618D,S68619D S68620D,S68621D,S68622D,S68623D,S68624D,S68625D,S68626D S68627D,S68628D,S68629D,S68711D,S68712D,S68719D,S68721D S68722D,S68729D,S78011D,S78012D,S78019D,S78021D,S78022D S78029D,S78111D,S78112D,S78119D,S78121D,S78122D,S78129D S78911D,S78912D,S78919D,S78921D,S78922D,S78929D,S88011D | V5889 |

Continued on next page

**Supplementary Table S1:** (cont'd) A comprehensive mapping of diagnosis groups used in the experiments to their corresponding ICD-10 and ICD-9 codes.

| Diagnosis | ICD-10 | ICD-9 |
|---|---|---|
| | S88012D,S88019D,S88021D,S88022D,S88029D,S88111D,S88112D<br>S88119D,S88121D,S88122D,S88129D,S88911D,S88912D,S88919D<br>S88921D,S88922D,S88929D,S98011D,S98012D,S98019D,S98021D<br>S98022D,S98029D,S98111D,S98112D,S98119D,S98121D,S98122D<br>S98129D,S98131D,S98132D,S98139D,S98141D,S98142D,S98149D<br>S98211D,S98212D,S98219D,S98221D,S98222D,S98229D,S98311D<br>S98312D,S98319D,S98321D,S98322D,S98329D,S98911D,S98912D<br>S98919D,S98921D,S98922D,S98929D | |
| Amputation of other body parts, initial encounter | S08111A,S08112A,S08119A,S08121A,S08122A,S08129A,S08811A<br>S08812A,S0889XA,S281XXA,S28211A,S28212A,S28219A,S28221A<br>S28222A,S28229A,S38211A,S38212A,S38221A,S38222A,S38231A<br>S38232A,S383XXA | 87201,87329,8738,8750,8780,8782,8788,8790,8798 |
| Amputation of other body parts, subsequent encounter | S08111D,S08112D,S08119D,S08121D,S08122D,S08129D,S08811D<br>S08812D,S0889XD,S281XXD,S28211D,S28212D,S28219D,S28221D<br>S28222D,S28229D,S38211D,S38212D,S38221D,S38222D,S38231D<br>S38232D,S383XXD | V5889 |
| Anal and rectal conditions | A511,A546,A563,A601,A609,A630,K594,K600,K601,K602,K603,K604<br>K605,K610,K611,K612,K613,K6131,K6139,K614,K615,K620,K621,K622<br>K623,K624,K626,K627,K6281,K6282,K6289,K629 | 05410,05419,07811,0911,0987,09952,5646,5650,5651,566,5690<br>5691,5692,56941,56942,56943,56944,56949 |
| Anesthesia complications during pregnancy | O29011,O29012,O29013,O29019,O29021,O29022,O29023,O29029<br>O29091,O29092,O29093,O29099,O29111,O29112,O29113,O29119<br>O29121,O29122,O29123,O29129,O29191,O29192,O29193,O29199<br>O29211,O29212,O29213,O29219,O29291,O29292,O29293,O29299<br>O293X1,O293X2,O293X3,O293X9,O2940,O2941,O2942,O2943,O295X1<br>O295X2,O295X3,O295X9,O2960,O2961,O2962,O2963,O298X1,O298X2<br>O298X3,O298X9,O2990,O2991,O2992,O2993,O740,O741,O742,O743<br>O744,O745,O746,O747,O748,O749 | 64680,64681,64683,66801,66802,66811,66812,66821,66822,66881<br>66882,66891,66892 |
| Antenatal screening | Z36,Z360,Z361,Z362,Z363,Z364,Z365,Z3681,Z3682,Z3683,Z3684<br>Z3685,Z3686,Z3687,Z3688,Z3689,Z368A,Z369 | V289 |
| Anxiety and fear-related disorders | F064,F10180,F10280,F10980,F12180,F12280,F12980,F13180,F13280<br>F13980,F14180,F14280,F14980,F15180,F15280,F15980,F16180<br>F16280,F16980,F18180,F18280,F18980,F19180,F19280,F19980,F4000<br>F4001,F4002,F4010,F4011,F40210,F40218,F40220,F40228,F40230<br>F40231,F40232,F40233,F40240,F40241,F40242,F40243,F40248 | 29189,29289,29384,30000,30001,30002,30009,30020,30021,30022<br>30023,30029,30410,30420,30430,30440,30450,30460,30921,31323 |

Continued on next page

**Supplementary Table S1:** (cont'd) A comprehensive mapping of diagnosis groups used in the experiments to their corresponding ICD-10 and ICD-9 codes.

| Diagnosis | ICD-10 | ICD-9 |
|---|---|---|
| | F40290,F40291,F40298,F408,F409,F410,F411,F413,F418,F419,F930 F940 | |
| Aortic and peripheral arterial embolism or thrombosis | I7401,I7409,I7410,I7411,I7419,I742,I743,I744,I745,I748,I749 I75011,I75012,I75013,I75019,I75021,I75022,I75023,I75029,I7581 I7589,I76 | 44401,44409,4441,44421,44422,44481,44489,4449,44501,44502 44581,44589,449 |
| Aortic; peripheral; and visceral artery aneurysms | A5201,I711,I7110,I7111,I7112,I7113,I712,I7120,I7121,I7122 I7123,I713,I7130,I7131,I7132,I7133,I714,I7140,I7141,I7142 I7143,I715,I7150,I7151,I7152,I716,I7160,I7161,I7162,I718,I719 I720,I721,I722,I723,I724,I725,I726,I728,I729,I790 | 0930,4411,4412,4413,4414,4415,4416,4417,4419,4420,4421,4422 4423,44281,44282,44283,44289,4429 |
| Aplastic anemia | D600,D601,D608,D609,D6101,D6109,D611,D612,D613,D61810,D61811 D61818,D6182,D6189,D619,D630,D631,D638,D640,D641,D642,D643 D644,D6481,D6489,D649 | 28401,28409,28411,28412,28419,2842,28481,28489,2849,2850 28521,28522,28529,2853,2858,2859 |
| Appendicitis and other appendiceal conditions | K352,K3520,K3521,K353,K3530,K3531,K3532,K3533,K3580,K3589 K35890,K35891,K36,K37,K380,K381,K382,K383,K388,K389 | 5400,5401,5409,541,542,5430,5439 |
| Arterial dissections | I2542,I670,I7100,I7101,I71010,I71011,I71012,I71019,I7102 I7103,I7770,I7771,I7772,I7773,I7774,I7775,I7776,I7777,I7779 | 41412,44100,44101,44102,44103,44321,44322,44323,44324,44329 |
| Aseptic necrosis and osteonecrosis | L97104,L97114,L97124,L97204,L97214,L97224,L97304,L97314 L97324,L97404,L97414,L97424,L97504,L97514,L97524,L97804 L97814,L97824,L97904,L97914,L97924,L98414,L98424,L98494,M8700 M87011,M87012,M87019,M87021,M87022,M87029,M87031,M87032 M87033,M87034,M87035,M87036,M87037,M87038,M87039,M87041 M87042,M87043,M87044,M87045,M87046,M87050,M87051,M87052 M87059,M87061,M87062,M87063,M87064,M87065,M87066,M87071 M87072,M87073,M87074,M87075,M87076,M87077,M87078,M87079,M8708 M8709,M8710,M87111,M87112,M87119,M87121,M87122,M87129,M87131 M87132,M87133,M87134,M87135,M87136,M87137,M87138,M87139 M87141,M87142,M87143,M87144,M87145,M87146,M87150,M87151 M87152,M87159,M87161,M87162,M87163,M87164,M87165,M87166 M87171,M87172,M87173,M87174,M87175,M87176,M87177,M87178 M87179,M87180,M87188,M8719,M8720,M87211,M87212,M87219,M87221 M87222,M87229,M87231,M87232,M87233,M87234,M87235,M87236 M87237,M87238,M87239,M87241,M87242,M87243,M87244,M87245 M87246,M87250,M87251,M87252,M87256,M87261,M87262,M87263 | 70710,70711,70712,70713,70714,70715,70719,7078,7079,7318 73340,73341,73342,73343,73344,73345,73349 |

Continued on next page

**Supplementary Table S1:** (cont'd) A comprehensive mapping of diagnosis groups used in the experiments to their corresponding ICD-10 and ICD-9 codes.

| Diagnosis | ICD-10 | ICD-9 |
|---|---|---|
| | M87264,M87265,M87266,M87271,M87272,M87273,M87274,M87275 M87276,M87277,M87278,M87279,M8728,M8729,M8730,M87311,M87312 M87319,M87321,M87322,M87329,M87331,M87332,M87333,M87334 M87335,M87336,M87337,M87338,M87339,M87341,M87342,M87343 M87344,M87345,M87346,M87350,M87351,M87352,M87353,M87361 M87362,M87363,M87364,M87365,M87366,M87371,M87372,M87373 M87374,M87375,M87376,M87377,M87378,M87379,M8738,M8739,M8780 M87811,M87812,M87819,M87821,M87822,M87829,M87831,M87832 M87833,M87834,M87835,M87836,M87837,M87838,M87839,M87841 M87842,M87843,M87844,M87845,M87849,M87850,M87851,M87852 M87859,M87861,M87862,M87863,M87864,M87865,M87869,M87871 M87872,M87873,M87874,M87875,M87876,M87877,M87878,M87879,M8788 M8789,M879,M9050,M90511,M90512,M90519,M90521,M90522,M90529 M90531,M90532,M90539,M90541,M90542,M90549,M90551,M90552 M90559,M90561,M90562,M90569,M90571,M90572,M90579,M9058,M9059 | |
| Aspiration pneumonitis | J690,J691,J698,J954,O29011,O29012,O29013,O29019,O740,O8901 | 5070,5071,5078,64680,64681,64683,66801,66802,66804,99739 |
| Asthma | J4520,J4521,J4522,J4530,J4531,J4532,J4540,J4541,J4542,J4550 J4551,J4552,J45901,J45902,J45909,J45990,J45991,J45998 | 49300,49301,49302,49310,49311,49312,49381,49382,49390,49391 49392 |
| Atypical fracture, initial encounter | M84750A,M84751A,M84752A,M84753A,M84754A,M84755A,M84756A M84757A,M84758A,M84759A | |
| Atypical fracture, subsequent encounter | M84750D,M84750G,M84750K,M84750P,M84751D,M84751G,M84751K M84751P,M84752D,M84752G,M84752K,M84752P,M84753D,M84753G M84753K,M84753P,M84754D,M84754G,M84754K,M84754P,M84755D M84755G,M84755K,M84755P,M84756D,M84756G,M84756K,M84756P M84757D,M84757G,M84757K,M84757P,M84758D,M84758G,M84758K M84758P,M84759D,M84759G,M84759K,M84759P | |
| Autoinflammatory syndromes | M041,M042,M048,M049 | |
| Bacterial infections | A000,A001,A009,A0100,A0101,A0102,A0103,A0104,A0105,A0109,A011 A012,A013,A014,A020,A021,A0220,A0221,A0222,A0223,A0224,A0225 A0229,A028,A029,A030,A031,A032,A033,A038,A039,A040,A041,A042 A043,A044,A045,A046,A047,A0471,A0472,A048,A049,A200,A201,A202 A203,A207,A208,A209,A210,A211,A212,A213,A217,A218,A219,A220 | 0010,0011,0019,0020,0021,0022,0023,0029,0030,0031,00320,00321 00322,00323,00324,00329,0038,0039,0040,0041,0042,0043,0048 0049,00800,00801,00802,00803,00804,00809,0082,00841,00843 00844,00845,00849,0085,0200,0201,0202,0203,0204,0205,0208 0209,0210,0211,0212,0213,0218,0219,0220,0221,0222,0223,0228 |

Continued on next page

**Supplementary Table S1:** (cont'd) A comprehensive mapping of diagnosis groups used in the experiments to their corresponding ICD-10 and ICD-9 codes.

| Diagnosis | ICD-10 | ICD-9 |
|---|---|---|
| | A221,A222,A227,A228,A229,A230,A231,A232,A233,A238,A239,A240 | 0229,0230,0231,0232,0233,0238,0239,024,025,0260,0261,0269 |
| | A241,A242,A243,A249,A250,A251,A259,A260,A267,A268,A269,A270 | 0270,0271,0272,0278,0279,0300,0301,0302,0303,0308,0309,0310 |
| | A2781,A2789,A279,A280,A281,A282,A288,A289,A300,A301,A302,A303 | 0311,0312,0318,0319,0320,0321,0322,0323,03281,03282,03283 |
| | A304,A305,A308,A309,A310,A311,A312,A318,A319,A320,A3211,A3212 | 03284,03285,03289,0329,0330,0331,0338,0339,0340,0341,035,0360 |
| | A327,A3281,A3282,A3289,A329,A33,A34,A35,A360,A361,A362,A363 | 0361,0362,0363,03640,03641,03642,03643,03681,03682,03689,0369 |
| | A3681,A3682,A3683,A3684,A3685,A3686,A3689,A369,A3700,A3701 | 037,0380,03810,03811,03812,03819,0382,0383,03840,03841,03842 |
| | A3710,A3711,A3780,A3781,A3790,A3791,A380,A381,A388,A389,A390 | 03843,03844,03849,0388,0389,0390,0391,0392,0393,0398,0399 |
| | A391,A392,A393,A394,A3950,A3951,A3952,A3953,A3981,A3982,A3983 | 0400,0401,04042,04082,04089,04100,04101,04102,04104,04109 |
| | A3984,A3989,A399,A400,A401,A403,A408,A409,A4101,A4102,A411 | 04110,04111,04112,04119,0412,0413,04141,04142,04143,04149 |
| | A412,A413,A414,A4150,A4151,A4152,A4153,A4159,A4181,A4189,A419 | 0415,0416,0417,04181,04182,04183,04184,04186,04189,0419,0739 |
| | A420,A421,A422,A427,A4281,A4282,A4289,A429,A430,A431,A438 | 0760,0761,0769,0770,07798,0783,07888,07988,07998,0830,0880 |
| | A439,A440,A441,A448,A449,A46,A480,A481,A482,A483,A484,A4852 | 1000,10081,10089,1009,3200,3201,3202,3203,3207,32082,32089 |
| | A488,A4901,A4902,A491,A492,A493,A498,A499,A70,A710,A711,A719 | 3209,390,3929,4660,481,4820,4821,4822,48231,48232,48239,48240 |
| | A740,A7481,A7489,A749,A78,B950,B951,B952,B953,B954,B955,B9561 | 48241,48242,48249,48281,48282,48283,48284,48289,4829,4830 |
| | B9562,B957,B958,B960,B961,B9620,B9621,B9622,B9623,B9629,B963 | 4831,4843,4845,56723,6390,71100,71101,71102,71103,71104,71105 |
| | B964,B965,B966,B967,B9681,B9682,B9689,D5931,G000,G001,G002 | 71106,71107,71108,71109,7700,7712,7713,77181,99591 |
| | G003,G008,G009,G01,G042,I00,I029,J020,J0300,J0301,J13,J14 | |
| | J150,J151,J1520,J15211,J15212,J1529,J153,J154,J155,J156,J157 | |
| | J158,J159,J160,J200,J201,J202,K652,M0000,M00011,M00012,M00019 | |
| | M00021,M00022,M00029,M00031,M00032,M00039,M00041,M00042 | |
| | M00049,M00051,M00052,M00059,M00061,M00062,M00069,M00071 | |
| | M00072,M00079,M0008,M0009,M0010,M00111,M00112,M00119,M00121 | |
| | M00122,M00129,M00131,M00132,M00139,M00141,M00142,M00149 | |
| | M00151,M00152,M00159,M00161,M00162,M00169,M00171,M00172 | |
| | M00179,M0018,M0019,M0020,M00211,M00212,M00219,M00221,M00222 | |
| | M00229,M00231,M00232,M00239,M00241,M00242,M00249,M00251 | |
| | M00252,M00259,M00261,M00262,M00269,M00271,M00272,M00279,M0028 | |
| | M0029,M0080,M00811,M00812,M00819,M00821,M00822,M00829,M00831 | |
| | M00832,M00839,M00841,M00842,M00849,M00851,M00852,M00859 | |
| | M00861,M00862,M00869,M00871,M00872,M00879,M0088,M0089,M009 | |
| | P0082,P231,P232,P233,P234,P235,P236,P360,P3610,P3619,P362 | |
| | P3630,P3639,P364,P365,P368,P369,P372 | |
| Benign neoplasms | D100,D101,D102,D1030,D1039,D104,D105,D106,D107,D109,D110,D117 | 20940,20941,20942,20943,20950,20951,20952,20953,20954,20955 |
| | D119,D120,D121,D122,D123,D124,D125,D126,D127,D128,D129,D130 | 20956,20957,20960,20961,20962,20963,20964,20965,20966,20967 |
| | D131,D132,D1330,D1339,D134,D135,D136,D137,D139,D140,D141,D142 | 20969,2100,2101,2102,2103,2104,2105,2106,2107,2108,2109,2110 |
| | D1430,D1431,D1432,D144,D150,D151,D152,D157,D159,D1600,D1601 | 2111,2112,2113,2114,2115,2116,2117,2118,2119,2120,2121,2122 |
| | D1602,D1610,D1611,D1612,D1620,D1621,D1622,D1630,D1631,D1632 | 2123,2124,2125,2126,2127,2128,2129,2130,2131,2132,2133,2134 |
| | D164,D165,D166,D167,D168,D169,D170,D171,D1720,D1721,D1722 | 2135,2136,2137,2138,2139,2140,2141,2142,2143,2144,2148,2149 |
| | D1723,D1724,D1730,D1739,D174,D175,D176,D1771,D1772,D1779,D179 | 2150,2152,2153,2154,2155,2156,2157,2159,2160,2161,2162,2163 |

Continued on next page

**Supplementary Table S1:** (cont'd) A comprehensive mapping of diagnosis groups used in the experiments to their corresponding ICD-10 and ICD-9 codes.

| Diagnosis | ICD-10 | ICD-9 |
|---|---|---|
| | D1800,D1801,D1802,D1803,D1809,D181,D190,D191,D197,D199,D200 | 2164,2165,2166,2167,2168,2169,217,2180,2181,2182,2189,2190 |
| | D201,D210,D2110,D2111,D2112,D2120,D2121,D2122,D213,D214,D215 | 2191,2198,2199,220,2210,2211,2212,2218,2219,2220,2221,2222 |
| | D216,D219,D220,D2210,D2211,D22111,D22112,D2212,D22121,D22122 | 2223,2224,2228,2229,2230,2231,2232,2233,22381,22389,2239,2240 |
| | D2220,D2221,D2222,D2230,D2239,D224,D225,D2260,D2261,D2262 | 2241,2242,2243,2244,2245,2246,2247,2248,2249,2250,2251,2252 |
| | D2270,D2271,D2272,D229,D230,D2310,D2311,D23111,D23112,D2312 | 2253,2254,2258,2259,226,2270,2271,2273,2274,2275,2276,2278 |
| | D23121,D23122,D2320,D2321,D2322,D2330,D2339,D234,D235,D2360 | 2279,22800,22801,22802,22804,22809,2281,2290,2298,2299 |
| | D2361,D2362,D2370,D2371,D2372,D239,D241,D242,D249,D250,D251 | |
| | D252,D259,D260,D261,D267,D269,D270,D271,D279,D280,D281,D282 | |
| | D287,D289,D290,D291,D2920,D2921,D2922,D2930,D2931,D2932,D294 | |
| | D298,D299,D3000,D3001,D3002,D3010,D3011,D3012,D3020,D3021 | |
| | D3022,D303,D304,D308,D309,D3100,D3101,D3102,D3110,D3111,D3112 | |
| | D3120,D3121,D3122,D3130,D3131,D3132,D3140,D3141,D3142,D3150 | |
| | D3151,D3152,D3160,D3161,D3162,D3190,D3191,D3192,D320,D321 | |
| | D329,D330,D331,D332,D333,D334,D337,D339,D34,D3500,D3501,D3502 | |
| | D351,D352,D353,D354,D355,D356,D357,D359,D360,D3610,D3611 | |
| | D3612,D3613,D3614,D3615,D3616,D3617,D367,D369,D3A00,D3A010 | |
| | D3A011,D3A012,D3A019,D3A020,D3A021,D3A022,D3A023,D3A024 | |
| | D3A025,D3A026,D3A029,D3A090,D3A091,D3A092,D3A093,D3A094 | |
| | D3A095,D3A096,D3A098,D3A8,K317,K635 | |
| Benign ovarian cyst | N801,N830,N8300,N8301,N8302,N831,N8310,N8311,N8312,N8320 | 6171,6200,6201,6202 |
| | N83201,N83202,N83209,N8329,N83291,N83292,N83299 | |
| Biliary tract disease | K563,K8000,K8001,K8010,K8011,K8012,K8013,K8018,K8019,K8020 | 56031,57400,57401,57410,57411,57420,57421,57430,57431,57440 |
| | K8021,K8030,K8031,K8032,K8033,K8034,K8035,K8036,K8037,K8040 | 57441,57450,57451,57460,57461,57470,57471,57480,57481,57490 |
| | K8041,K8042,K8043,K8044,K8045,K8046,K8047,K8050,K8051,K8060 | 57491,5750,57510,57511,57512,5752,5753,5754,5755,5756,5758 |
| | K8061,K8062,K8063,K8064,K8065,K8066,K8067,K8070,K8071,K8080 | 5759,5760,5761,5762,5763,5764,5765,5768,5769,5778 |
| | K8081,K810,K811,K812,K819,K820,K821,K822,K823,K824,K828,K829 | |
| | K82A1,K82A2,K830,K8301,K8309,K831,K832,K833,K834,K835,K838 | |
| | K839,K87,K915 | |
| Biomechanical lesions | M9900,M9901,M9902,M9903,M9904,M9905,M9906,M9907,M9908,M9909 | 7230,72401,72402,72409,73819,7382,7383,7385,7386,7388,7389 |
| | M9910,M9911,M9912,M9913,M9914,M9915,M9916,M9917,M9918,M9919 | 7390,7391,7392,7393,7394,7395,7396,7397,7398,7399,83900,83920 |
| | M9920,M9921,M9922,M9923,M9924,M9925,M9926,M9927,M9928,M9929 | 83921,83942,83949,83961,83969 |
| | M9930,M9931,M9932,M9933,M9934,M9935,M9936,M9937,M9938,M9939 | |
| | M9940,M9941,M9942,M9943,M9944,M9945,M9946,M9947,M9948,M9949 | |
| | M9950,M9951,M9952,M9953,M9954,M9955,M9956,M9957,M9958,M9959 | |
| | M9960,M9961,M9962,M9963,M9964,M9965,M9966,M9967,M9968,M9969 | |
| | M9970,M9971,M9972,M9973,M9974,M9975,M9976,M9977,M9978,M9979 | |
| | M9980,M9981,M9982,M9983,M9984,M9985,M9986,M9987,M9988,M9989 | |

Continued on next page

**Supplementary Table S1:** (cont'd) A comprehensive mapping of diagnosis groups used in the experiments to their corresponding ICD-10 and ICD-9 codes.

| Diagnosis | ICD-10 | ICD-9 |
|---|---|---|
| | M999 | |
| Bipolar and related disorders | F0633,F0634,F3010,F3011,F3012,F3013,F302,F303,F308,F309,F310<br>F3110,F3111,F3112,F3113,F312,F3130,F3131,F3132,F314,F315<br>F3160,F3161,F3162,F3163,F3164,F3171,F3173,F3175,F3177,F3181<br>F3189,F319,F340 | 29383,29600,29601,29602,29603,29604,29605,29640,29641,29642<br>29643,29644,29645,29650,29651,29652,29653,29654,29655,29660<br>29661,29662,29663,29664,29665,2967,29680,29681,29689,30110<br>30113 |
| Birth trauma | P100,P101,P102,P103,P104,P108,P109,P110,P111,P112,P113,P114<br>P115,P119,P120,P121,P122,P123,P124,P1281,P1289,P129,P130,P131<br>P132,P133,P134,P138,P139,P140,P141,P142,P143,P148,P149,P150<br>P151,P152,P153,P154,P155,P156,P158,P159 | 7670,76711,76719,7672,7673,7674,7675,7676,7677,7678,7679<br>77210,7722 |
| Blindness and vision defects | H53001,H53002,H53003,H53009,H53011,H53012,H53013,H53019<br>H53021,H53022,H53023,H53029,H53031,H53032,H53033,H53039<br>H53041,H53042,H53043,H53049,H5310,H5311,H53121,H53122,H53123<br>H53129,H53131,H53132,H53133,H53139,H53141,H53142,H53143<br>H53149,H5315,H5316,H5319,H532,H5330,H5331,H5332,H5333,H5334<br>H5340,H53411,H53412,H53413,H53419,H53421,H53422,H53423,H53429<br>H53431,H53432,H53433,H53439,H53451,H53452,H53453,H53459<br>H53461,H53462,H53469,H5347,H53481,H53482,H53483,H53489,H5350<br>H5351,H5352,H5353,H5354,H5355,H5359,H5360,H5361,H5362,H5363<br>H5369,H5371,H5372,H538,H539,H540,H540X33,H540X34,H540X35<br>H540X43,H540X44,H540X45,H540X53,H540X54,H540X55,H5410,H5411<br>H541131,H541132,H541141,H541142,H541151,H541152,H5412,H541213<br>H541214,H541215,H541223,H541224,H541225,H542,H542X11,H542X12<br>H542X21,H542X22,H543,H5440,H5441,H54413A,H54414A,H54415A<br>H5442,H5442A3,H5442A4,H5442A5,H5450,H5451,H54511A,H54512A<br>H5452,H5452A1,H5452A2,H5460,H5461,H5462,H547,H548,Z4531 | 36800,36801,36802,36803,36810,36811,36812,36813,36814,36815<br>36816,3682,36830,36831,36832,36833,36834,36840,36841,36842<br>36843,36844,36845,36846,36847,36851,36852,36853,36854,36855<br>36859,36860,36861,36862,36863,36869,3688,3689,36900,36901<br>36910,36920,36925,3693,3694,36960,36963,36966,36969,36970<br>36971,3698,3699,V410,V5309 |
| Bone cancer | C4000,C4001,C4002,C4010,C4011,C4012,C4020,C4021,C4022,C4030<br>C4031,C4032,C4080,C4081,C4082,C4090,C4091,C4092,C410,C411<br>C412,C413,C414,C419 | 1700,1701,1702,1703,1704,1705,1706,1707,1708,1709 |
| Breast cancer - all other types | C50011,C50012,C50019,C50021,C50022,C50029,C50111,C50112<br>C50119,C50121,C50122,C50129,C50211,C50212,C50219,C50221<br>C50222,C50229,C50311,C50312,C50319,C50321,C50322,C50329<br>C50411,C50412,C50419,C50421,C50422,C50429,C50511,C50512<br>C50519,C50521,C50522,C50529,C50611,C50612,C50619,C50621<br>C50622,C50629,C50811,C50812,C50819,C50821,C50822,C50829<br>C50911,C50912,C50919,C50921,C50922,C50929,D0500,D0501,D0502 | 1740,1741,1742,1743,1744,1745,1746,1748,1749,1750,1759,2330 |

Continued on next page

**Supplementary Table S1:** (cont'd) A comprehensive mapping of diagnosis groups used in the experiments to their corresponding ICD-10 and ICD-9 codes.

| Diagnosis | ICD-10 | ICD-9 |
|---|---|---|
| | D0580,D0581,D0582,D0590,D0591,D0592 | |
| Breast cancer - | D0510,D0511,D0512 | 2330 |
| ductal carcinoma in | | |
| situ (DCIS) | | |
| Burn and corrosion, | T2000XA,T20011A,T20012A,T20019A,T2002XA,T2003XA,T2004XA | 9400,9401,9402,9403,9404,9405,9409,94100,94101,94103,94104 |
| initial encounter | T2005XA,T2006XA,T2007XA,T2009XA,T2010XA,T20111A,T20112A | 94105,94106,94107,94108,94109,94110,94111,94113,94114,94115 |
| | T20119A,T2012XA,T2013XA,T2014XA,T2015XA,T2016XA,T2017XA | 94116,94117,94118,94119,94120,94121,94123,94124,94125,94126 |
| | T2019XA,T2020XA,T20211A,T20212A,T20219A,T2022XA,T2023XA | 94127,94128,94129,94130,94131,94133,94134,94135,94136,94137 |
| | T2024XA,T2025XA,T2026XA,T2027XA,T2029XA,T2030XA,T20311A | 94138,94139,94140,94141,94143,94144,94145,94146,94147,94148 |
| | T20312A,T20319A,T2032XA,T2033XA,T2034XA,T2035XA,T2036XA | 94149,94150,94151,94152,94153,94154,94155,94156,94157,94158 |
| | T2037XA,T2039XA,T2040XA,T20411A,T20412A,T20419A,T2042XA | 94159,94200,94201,94202,94203,94204,94205,94209,94210,94211 |
| | T2043XA,T2044XA,T2045XA,T2046XA,T2047XA,T2049XA,T2050XA | 94212,94213,94214,94215,94219,94220,94221,94222,94223,94224 |
| | T20511A,T20512A,T20519A,T2052XA,T2053XA,T2054XA,T2055XA | 94225,94229,94230,94231,94232,94233,94234,94235,94239,94240 |
| | T2056XA,T2057XA,T2059XA,T2060XA,T20611A,T20612A,T20619A | 94241,94242,94243,94244,94245,94249,94250,94251,94252,94253 |
| | T2062XA,T2063XA,T2064XA,T2065XA,T2066XA,T2067XA,T2069XA | 94254,94255,94259,94300,94301,94302,94303,94304,94305,94306 |
| | T2070XA,T20711A,T20712A,T20719A,T2072XA,T2073XA,T2074XA | 94309,94310,94311,94312,94313,94314,94315,94316,94319,94320 |
| | T2075XA,T2076XA,T2077XA,T2079XA,T2100XA,T2101XA,T2102XA | 94321,94322,94323,94324,94325,94326,94329,94330,94331,94332 |
| | T2103XA,T2104XA,T2105XA,T2106XA,T2107XA,T2109XA,T2110XA | 94333,94334,94335,94336,94339,94340,94341,94342,94343,94344 |
| | T2111XA,T2112XA,T2113XA,T2114XA,T2115XA,T2116XA,T2117XA | 94345,94346,94349,94351,94352,94353,94354,94355,94356,94359 |
| | T2119XA,T2120XA,T2121XA,T2122XA,T2123XA,T2124XA,T2125XA | 94400,94401,94402,94403,94404,94405,94406,94407,94408,94410 |
| | T2126XA,T2127XA,T2129XA,T2130XA,T2131XA,T2132XA,T2133XA | 94411,94412,94413,94414,94415,94416,94417,94418,94420,94421 |
| | T2134XA,T2135XA,T2136XA,T2137XA,T2139XA,T2140XA,T2141XA | 94422,94423,94424,94425,94426,94427,94428,94430,94431,94432 |
| | T2142XA,T2143XA,T2144XA,T2145XA,T2146XA,T2147XA,T2149XA | 94433,94434,94435,94436,94437,94438,94440,94441,94442,94443 |
| | T2150XA,T2151XA,T2152XA,T2153XA,T2154XA,T2155XA,T2156XA | 94444,94445,94446,94447,94448,94450,94451,94452,94453,94454 |
| | T2157XA,T2159XA,T2160XA,T2161XA,T2162XA,T2163XA,T2164XA | 94455,94456,94457,94458,94500,94501,94502,94503,94504,94505 |
| | T2165XA,T2166XA,T2167XA,T2169XA,T2170XA,T2171XA,T2172XA | 94506,94509,94510,94511,94512,94513,94514,94515,94516,94519 |
| | T2173XA,T2174XA,T2175XA,T2176XA,T2177XA,T2179XA,T2200XA | 94520,94521,94522,94523,94524,94525,94526,94529,94530,94531 |
| | T22011A,T22012A,T22019A,T22021A,T22022A,T22029A,T22031A | 94532,94533,94534,94535,94536,94539,94540,94541,94542,94543 |
| | T22032A,T22039A,T22041A,T22042A,T22049A,T22051A,T22052A | 94544,94545,94546,94549,94550,94551,94552,94553,94554,94555 |
| | T22059A,T22061A,T22062A,T22069A,T22091A,T22092A,T22099A | 94556,94559,9470,9471,9472,9473,9474,9478,9479,94800,94810 |
| | T2210XA,T22111A,T22112A,T22119A,T22121A,T22122A,T22129A | 94811,94820,94821,94822,94830,94831,94832,94833,94840,94841 |
| | T22131A,T22132A,T22139A,T22141A,T22142A,T22149A,T22151A | 94842,94843,94844,94850,94851,94852,94853,94854,94855,94860 |
| | T22152A,T22159A,T22161A,T22162A,T22169A,T22191A,T22192A | 94861,94862,94863,94864,94865,94866,94870,94871,94872,94873 |
| | T22199A,T2220XA,T22211A,T22212A,T22219A,T22221A,T22222A | 94874,94875,94876,94877,94880,94881,94882,94883,94884,94885 |
| | T22229A,T22231A,T22232A,T22239A,T22241A,T22242A,T22249A | 94886,94887,94888,94890,94891,94892,94893,94894,94895,94896 |
| | T22251A,T22252A,T22259A,T22261A,T22262A,T22269A,T22291A | 94897,94898,94899,9490 |
| | T22292A,T22299A,T2230XA,T22311A,T22312A,T22319A,T22321A | |
| | T22322A,T22329A,T22331A,T22332A,T22339A,T22341A,T22342A | |

Continued on next page

**Supplementary Table S1:** (cont'd) A comprehensive mapping of diagnosis groups used in the experiments to their corresponding ICD-10 and ICD-9 codes.

| Diagnosis | ICD-10 | ICD-9 |
|---|---|---|
| | T22349A,T22351A,T22352A,T22359A,T22361A,T22362A,T22369A | |
| | T22391A,T22392A,T22399A,T2240XA,T22411A,T22412A,T22419A | |
| | T22421A,T22422A,T22429A,T22431A,T22432A,T22439A,T22441A | |
| | T22442A,T22449A,T22451A,T22452A,T22459A,T22461A,T22462A | |
| | T22469A,T22491A,T22492A,T22499A,T2250XA,T22511A,T22512A | |
| | T22519A,T22521A,T22522A,T22529A,T22531A,T22532A,T22539A | |
| | T22541A,T22542A,T22549A,T22551A,T22552A,T22559A,T22561A | |
| | T22562A,T22569A,T22591A,T22592A,T22599A,T2260XA,T22611A | |
| | T22612A,T22619A,T22621A,T22622A,T22629A,T22631A,T22632A | |
| | T22639A,T22641A,T22642A,T22649A,T22651A,T22652A,T22659A | |
| | T22661A,T22662A,T22669A,T22691A,T22692A,T22699A,T2270XA | |
| | T22711A,T22712A,T22719A,T22721A,T22722A,T22729A,T22731A | |
| | T22732A,T22739A,T22741A,T22742A,T22749A,T22751A,T22752A | |
| | T22759A,T22761A,T22762A,T22769A,T22791A,T22792A,T22799A | |
| | T23001A,T23002A,T23009A,T23011A,T23012A,T23019A,T23021A | |
| | T23022A,T23029A,T23031A,T23032A,T23039A,T23041A,T23042A | |
| | T23049A,T23051A,T23052A,T23059A,T23061A,T23062A,T23069A | |
| | T23071A,T23072A,T23079A,T23091A,T23092A,T23099A,T23101A | |
| | T23102A,T23109A,T23111A,T23112A,T23119A,T23121A,T23122A | |
| | T23129A,T23131A,T23132A,T23139A,T23141A,T23142A,T23149A | |
| | T23151A,T23152A,T23159A,T23161A,T23162A,T23169A,T23171A | |
| | T23172A,T23179A,T23191A,T23192A,T23199A,T23201A,T23202A | |
| | T23209A,T23211A,T23212A,T23219A,T23221A,T23222A,T23229A | |
| | T23231A,T23232A,T23239A,T23241A,T23242A,T23249A,T23251A | |
| | T23252A,T23259A,T23261A,T23262A,T23269A,T23271A,T23272A | |
| | T23279A,T23291A,T23292A,T23299A,T23301A,T23302A,T23309A | |
| | T23311A,T23312A,T23319A,T23321A,T23322A,T23329A,T23331A | |
| | T23332A,T23339A,T23341A,T23342A,T23349A,T23351A,T23352A | |
| | T23359A,T23361A,T23362A,T23369A,T23371A,T23372A,T23379A | |
| | T23391A,T23392A,T23399A,T23401A,T23402A,T23409A,T23411A | |
| | T23412A,T23419A,T23421A,T23422A,T23429A,T23431A,T23432A | |
| | T23439A,T23441A,T23442A,T23449A,T23451A,T23452A,T23459A | |
| | T23461A,T23462A,T23469A,T23471A,T23472A,T23479A,T23491A | |
| | T23492A,T23499A,T23501A,T23502A,T23509A,T23511A,T23512A | |
| | T23519A,T23521A,T23522A,T23529A,T23531A,T23532A,T23539A | |
| | T23541A,T23542A,T23549A,T23551A,T23552A,T23559A,T23561A | |
| | T23562A,T23569A,T23571A,T23572A,T23579A,T23591A,T23592A | |
| | T23599A,T23601A,T23602A,T23609A,T23611A,T23612A,T23619A | |
| | T23621A,T23622A,T23629A,T23631A,T23632A,T23639A,T23641A | |
| | T23642A,T23649A,T23651A,T23652A,T23659A,T23661A,T23662A | |

Continued on next page

**Supplementary Table S1:** (cont'd) A comprehensive mapping of diagnosis groups used in the experiments to their corresponding ICD-10 and ICD-9 codes.

| Diagnosis | ICD-10 | ICD-9 |
|---|---|---|
| | T23669A,T23671A,T23672A,T23679A,T23691A,T23692A,T23699A | |
| | T23701A,T23702A,T23709A,T23711A,T23712A,T23719A,T23721A | |
| | T23722A,T23729A,T23731A,T23732A,T23739A,T23741A,T23742A | |
| | T23749A,T23751A,T23752A,T23759A,T23761A,T23762A,T23769A | |
| | T23771A,T23772A,T23779A,T23791A,T23792A,T23799A,T24001A | |
| | T24002A,T24009A,T24011A,T24012A,T24019A,T24021A,T24022A | |
| | T24029A,T24031A,T24032A,T24039A,T24091A,T24092A,T24099A | |
| | T24101A,T24102A,T24109A,T24111A,T24112A,T24119A,T24121A | |
| | T24122A,T24129A,T24131A,T24132A,T24139A,T24191A,T24192A | |
| | T24199A,T24201A,T24202A,T24209A,T24211A,T24212A,T24219A | |
| | T24221A,T24222A,T24229A,T24231A,T24232A,T24239A,T24291A | |
| | T24292A,T24299A,T24301A,T24302A,T24309A,T24311A,T24312A | |
| | T24319A,T24321A,T24322A,T24329A,T24331A,T24332A,T24339A | |
| | T24391A,T24392A,T24399A,T24401A,T24402A,T24409A,T24411A | |
| | T24412A,T24419A,T24421A,T24422A,T24429A,T24431A,T24432A | |
| | T24439A,T24491A,T24492A,T24499A,T24501A,T24502A,T24509A | |
| | T24511A,T24512A,T24519A,T24521A,T24522A,T24529A,T24531A | |
| | T24532A,T24539A,T24591A,T24592A,T24599A,T24601A,T24602A | |
| | T24609A,T24611A,T24612A,T24619A,T24621A,T24622A,T24629A | |
| | T24631A,T24632A,T24639A,T24691A,T24692A,T24699A,T24701A | |
| | T24702A,T24709A,T24711A,T24712A,T24719A,T24721A,T24722A | |
| | T24729A,T24731A,T24732A,T24739A,T24791A,T24792A,T24799A | |
| | T25011A,T25012A,T25019A,T25021A,T25022A,T25029A,T25031A | |
| | T25032A,T25039A,T25091A,T25092A,T25099A,T25111A,T25112A | |
| | T25119A,T25121A,T25122A,T25129A,T25131A,T25132A,T25139A | |
| | T25191A,T25192A,T25199A,T25211A,T25212A,T25219A,T25221A | |
| | T25222A,T25229A,T25231A,T25232A,T25239A,T25291A,T25292A | |
| | T25299A,T25311A,T25312A,T25319A,T25321A,T25322A,T25329A | |
| | T25331A,T25332A,T25339A,T25391A,T25392A,T25399A,T25411A | |
| | T25412A,T25419A,T25421A,T25422A,T25429A,T25431A,T25432A | |
| | T25439A,T25491A,T25492A,T25499A,T25511A,T25512A,T25519A | |
| | T25521A,T25522A,T25529A,T25531A,T25532A,T25539A,T25591A | |
| | T25592A,T25599A,T25611A,T25612A,T25619A,T25621A,T25622A | |
| | T25629A,T25631A,T25632A,T25639A,T25691A,T25692A,T25699A | |
| | T25711A,T25712A,T25719A,T25721A,T25722A,T25729A,T25731A | |
| | T25732A,T25739A,T25791A,T25792A,T25799A,T2600XA,T2601XA | |
| | T2602XA,T2610XA,T2611XA,T2612XA,T2620XA,T2621XA,T2622XA | |
| | T2630XA,T2631XA,T2632XA,T2640XA,T2641XA,T2642XA,T2650XA | |
| | T2651XA,T2652XA,T2660XA,T2661XA,T2662XA,T2670XA,T2671XA | |
| | T2672XA,T2680XA,T2681XA,T2682XA,T2690XA,T2691XA,T2692XA | |

Continued on next page

**Supplementary Table S1:** (cont'd) A comprehensive mapping of diagnosis groups used in the experiments to their corresponding ICD-10 and ICD-9 codes.

| Diagnosis | ICD-10 | ICD-9 |
|---|---|---|
| | T270XXA,T271XXA,T272XXA,T273XXA,T274XXA,T275XXA,T276XXA<br>T277XXA,T280XXA,T281XXA,T282XXA,T283XXA,T2840XA,T28411A<br>T28412A,T28419A,T2849XA,T285XXA,T286XXA,T287XXA,T288XXA<br>T2890XA,T28911A,T28912A,T28919A,T2899XA,T300,T304,T310,T3110<br>T3111,T3120,T3121,T3122,T3130,T3131,T3132,T3133,T3140,T3141<br>T3142,T3143,T3144,T3150,T3151,T3152,T3153,T3154,T3155,T3160<br>T3161,T3162,T3163,T3164,T3165,T3166,T3170,T3171,T3172,T3173<br>T3174,T3175,T3176,T3177,T3180,T3181,T3182,T3183,T3184,T3185<br>T3186,T3187,T3188,T3190,T3191,T3192,T3193,T3194,T3195,T3196<br>T3197,T3198,T3199,T320,T3210,T3211,T3220,T3221,T3222,T3230<br>T3231,T3232,T3233,T3240,T3241,T3242,T3243,T3244,T3250,T3251<br>T3252,T3253,T3254,T3255,T3260,T3261,T3262,T3263,T3264,T3265<br>T3266,T3270,T3271,T3272,T3273,T3274,T3275,T3276,T3277,T3280<br>T3281,T3282,T3283,T3284,T3285,T3286,T3287,T3288,T3290,T3291<br>T3292,T3293,T3294,T3295,T3296,T3297,T3298,T3299 | |
| Burns and corrosion,<br>subsequent encounter | T2000XD,T20011D,T20012D,T20019D,T2002XD,T2003XD,T2004XD<br>T2005XD,T2006XD,T2007XD,T2009XD,T2010XD,T20111D,T20112D<br>T20119D,T2012XD,T2013XD,T2014XD,T2015XD,T2016XD,T2017XD<br>T2019XD,T2020XD,T20211D,T20212D,T20219D,T2022XD,T2023XD<br>T2024XD,T2025XD,T2026XD,T2027XD,T2029XD,T2030XD,T20311D<br>T20312D,T20319D,T2032XD,T2033XD,T2034XD,T2035XD,T2036XD<br>T2037XD,T2039XD,T2040XD,T20411D,T20412D,T20419D,T2042XD<br>T2043XD,T2044XD,T2045XD,T2046XD,T2047XD,T2049XD,T2050XD<br>T20511D,T20512D,T20519D,T2052XD,T2053XD,T2054XD,T2055XD<br>T2056XD,T2057XD,T2059XD,T2060XD,T20611D,T20612D,T20619D<br>T2062XD,T2063XD,T2064XD,T2065XD,T2066XD,T2067XD,T2069XD<br>T2070XD,T20711D,T20712D,T20719D,T2072XD,T2073XD,T2074XD<br>T2075XD,T2076XD,T2077XD,T2079XD,T2100XD,T2101XD,T2102XD<br>T2103XD,T2104XD,T2105XD,T2106XD,T2107XD,T2109XD,T2110XD<br>T2111XD,T2112XD,T2113XD,T2114XD,T2115XD,T2116XD,T2117XD<br>T2119XD,T2120XD,T2121XD,T2122XD,T2123XD,T2124XD,T2125XD<br>T2126XD,T2127XD,T2129XD,T2130XD,T2131XD,T2132XD,T2133XD<br>T2134XD,T2135XD,T2136XD,T2137XD,T2139XD,T2140XD,T2141XD<br>T2142XD,T2143XD,T2144XD,T2145XD,T2146XD,T2147XD,T2149XD<br>T2150XD,T2151XD,T2152XD,T2153XD,T2154XD,T2155XD,T2156XD<br>T2157XD,T2159XD,T2160XD,T2161XD,T2162XD,T2163XD,T2164XD<br>T2165XD,T2166XD,T2167XD,T2169XD,T2170XD,T2171XD,T2172XD<br>T2173XD,T2174XD,T2175XD,T2176XD,T2177XD,T2179XD,T2200XD<br>T22011D,T22012D,T22019D,T22021D,T22022D,T22029D,T22031D | V5889 |

Continued on next page

**Supplementary Table S1:** (cont'd) A comprehensive mapping of diagnosis groups used in the experiments to their corresponding ICD-10 and ICD-9 codes.

| Diagnosis | ICD-10 | ICD-9 |
|---|---|---|
| | T22032D,T22039D,T22041D,T22042D,T22049D,T22051D,T22052D | |
| | T22059D,T22061D,T22062D,T22069D,T22091D,T22092D,T22099D | |
| | T2210XD,T22111D,T22112D,T22119D,T22121D,T22122D,T22129D | |
| | T22131D,T22132D,T22139D,T22141D,T22142D,T22149D,T22151D | |
| | T22152D,T22159D,T22161D,T22162D,T22169D,T22191D,T22192D | |
| | T22199D,T2220XD,T22211D,T22212D,T22219D,T22221D,T22222D | |
| | T22229D,T22231D,T22232D,T22239D,T22241D,T22242D,T22249D | |
| | T22251D,T22252D,T22259D,T22261D,T22262D,T22269D,T22291D | |
| | T22292D,T22299D,T2230XD,T22311D,T22312D,T22319D,T22321D | |
| | T22322D,T22329D,T22331D,T22332D,T22339D,T22341D,T22342D | |
| | T22349D,T22351D,T22352D,T22359D,T22361D,T22362D,T22369D | |
| | T22391D,T22392D,T22399D,T2240XD,T22411D,T22412D,T22419D | |
| | T22421D,T22422D,T22429D,T22431D,T22432D,T22439D,T22441D | |
| | T22442D,T22449D,T22451D,T22452D,T22459D,T22461D,T22462D | |
| | T22469D,T22491D,T22492D,T22499D,T2250XD,T22511D,T22512D | |
| | T22519D,T22521D,T22522D,T22529D,T22531D,T22532D,T22539D | |
| | T22541D,T22542D,T22549D,T22551D,T22552D,T22559D,T22561D | |
| | T22562D,T22569D,T22591D,T22592D,T22599D,T2260XD,T22611D | |
| | T22612D,T22619D,T22621D,T22622D,T22629D,T22631D,T22632D | |
| | T22639D,T22641D,T22642D,T22649D,T22651D,T22652D,T22659D | |
| | T22661D,T22662D,T22669D,T22691D,T22692D,T22699D,T2270XD | |
| | T22711D,T22712D,T22719D,T22721D,T22722D,T22729D,T22731D | |
| | T22732D,T22739D,T22741D,T22742D,T22749D,T22751D,T22752D | |
| | T22759D,T22761D,T22762D,T22769D,T22791D,T22792D,T22799D | |
| | T23001D,T23002D,T23009D,T23011D,T23012D,T23019D,T23021D | |
| | T23022D,T23029D,T23031D,T23032D,T23039D,T23041D,T23042D | |
| | T23049D,T23051D,T23052D,T23059D,T23061D,T23062D,T23069D | |
| | T23071D,T23072D,T23079D,T23091D,T23092D,T23099D,T23101D | |
| | T23102D,T23109D,T23111D,T23112D,T23119D,T23121D,T23122D | |
| | T23129D,T23131D,T23132D,T23139D,T23141D,T23142D,T23149D | |
| | T23151D,T23152D,T23159D,T23161D,T23162D,T23169D,T23171D | |
| | T23172D,T23179D,T23191D,T23192D,T23199D,T23201D,T23202D | |
| | T23209D,T23211D,T23212D,T23219D,T23221D,T23222D,T23229D | |
| | T23231D,T23232D,T23239D,T23241D,T23242D,T23249D,T23251D | |
| | T23252D,T23259D,T23261D,T23262D,T23269D,T23271D,T23272D | |
| | T23279D,T23291D,T23292D,T23299D,T23301D,T23302D,T23309D | |
| | T23311D,T23312D,T23319D,T23321D,T23322D,T23329D,T23331D | |
| | T23332D,T23339D,T23341D,T23342D,T23349D,T23351D,T23352D | |
| | T23359D,T23361D,T23362D,T23369D,T23371D,T23372D,T23379D | |
| | T23391D,T23392D,T23399D,T23401D,T23402D,T23409D,T23411D | |

Continued on next page

**Supplementary Table S1:** (cont'd) A comprehensive mapping of diagnosis groups used in the experiments to their corresponding ICD-10 and ICD-9 codes.

| Diagnosis | ICD-10 | ICD-9 |
|---|---|---|
| | T23412D,T23419D,T23421D,T23422D,T23429D,T23431D,T23432D | |
| | T23439D,T23441D,T23442D,T23449D,T23451D,T23452D,T23459D | |
| | T23461D,T23462D,T23469D,T23471D,T23472D,T23479D,T23491D | |
| | T23492D,T23499D,T23501D,T23502D,T23509D,T23511D,T23512D | |
| | T23519D,T23521D,T23522D,T23529D,T23531D,T23532D,T23539D | |
| | T23541D,T23542D,T23549D,T23551D,T23552D,T23559D,T23561D | |
| | T23562D,T23569D,T23571D,T23572D,T23579D,T23591D,T23592D | |
| | T23599D,T23601D,T23602D,T23609D,T23611D,T23612D,T23619D | |
| | T23621D,T23622D,T23629D,T23631D,T23632D,T23639D,T23641D | |
| | T23642D,T23649D,T23651D,T23652D,T23659D,T23661D,T23662D | |
| | T23669D,T23671D,T23672D,T23679D,T23691D,T23692D,T23699D | |
| | T23701D,T23702D,T23709D,T23711D,T23712D,T23719D,T23721D | |
| | T23722D,T23729D,T23731D,T23732D,T23739D,T23741D,T23742D | |
| | T23749D,T23751D,T23752D,T23759D,T23761D,T23762D,T23769D | |
| | T23771D,T23772D,T23779D,T23791D,T23792D,T23799D,T24001D | |
| | T24002D,T24009D,T24011D,T24012D,T24019D,T24021D,T24022D | |
| | T24029D,T24031D,T24032D,T24039D,T24091D,T24092D,T24099D | |
| | T24101D,T24102D,T24109D,T24111D,T24112D,T24119D,T24121D | |
| | T24122D,T24129D,T24131D,T24132D,T24139D,T24191D,T24192D | |
| | T24199D,T24201D,T24202D,T24209D,T24211D,T24212D,T24219D | |
| | T24221D,T24222D,T24229D,T24231D,T24232D,T24239D,T24291D | |
| | T24292D,T24299D,T24301D,T24302D,T24309D,T24311D,T24312D | |
| | T24319D,T24321D,T24322D,T24329D,T24331D,T24332D,T24339D | |
| | T24391D,T24392D,T24399D,T24401D,T24402D,T24409D,T24411D | |
| | T24412D,T24419D,T24421D,T24422D,T24429D,T24431D,T24432D | |
| | T24439D,T24491D,T24492D,T24499D,T24501D,T24502D,T24509D | |
| | T24511D,T24512D,T24519D,T24521D,T24522D,T24529D,T24531D | |
| | T24532D,T24539D,T24591D,T24592D,T24599D,T24601D,T24602D | |
| | T24609D,T24611D,T24612D,T24619D,T24621D,T24622D,T24629D | |
| | T24631D,T24632D,T24639D,T24691D,T24692D,T24699D,T24701D | |
| | T24702D,T24709D,T24711D,T24712D,T24719D,T24721D,T24722D | |
| | T24729D,T24731D,T24732D,T24739D,T24791D,T24792D,T24799D | |
| | T25011D,T25012D,T25019D,T25021D,T25022D,T25029D,T25031D | |
| | T25032D,T25039D,T25091D,T25092D,T25099D,T25111D,T25112D | |
| | T25119D,T25121D,T25122D,T25129D,T25131D,T25132D,T25139D | |
| | T25191D,T25192D,T25199D,T25211D,T25212D,T25219D,T25221D | |
| | T25222D,T25229D,T25231D,T25232D,T25239D,T25291D,T25292D | |
| | T25299D,T25311D,T25312D,T25319D,T25321D,T25322D,T25329D | |
| | T25331D,T25332D,T25339D,T25391D,T25392D,T25399D,T25411D | |
| | T25412D,T25419D,T25421D,T25422D,T25429D,T25431D,T25432D | |

Continued on next page

**Supplementary Table S1:** (cont'd) A comprehensive mapping of diagnosis groups used in the experiments to their corresponding ICD-10 and ICD-9 codes.

| Diagnosis | ICD-10 | ICD-9 |
|---|---|---|
| | T25439D,T25491D,T25492D,T25499D,T25511D,T25512D,T25519D<br>T25521D,T25522D,T25529D,T25531D,T25532D,T25539D,T25591D<br>T25592D,T25599D,T25611D,T25612D,T25619D,T25621D,T25622D<br>T25629D,T25631D,T25632D,T25639D,T25691D,T25692D,T25699D<br>T25711D,T25712D,T25719D,T25721D,T25722D,T25729D,T25731D<br>T25732D,T25739D,T25791D,T25792D,T25799D,T2600XD,T2601XD<br>T2602XD,T2610XD,T2611XD,T2612XD,T2620XD,T2621XD,T2622XD<br>T2630XD,T2631XD,T2632XD,T2640XD,T2641XD,T2642XD,T2650XD<br>T2651XD,T2652XD,T2660XD,T2661XD,T2662XD,T2670XD,T2671XD<br>T2672XD,T2680XD,T2681XD,T2682XD,T2690XD,T2691XD,T2692XD<br>T270XXD,T271XXD,T272XXD,T273XXD,T274XXD,T275XXD,T276XXD<br>T277XXD,T280XXD,T281XXD,T282XXD,T283XXD,T2840XD,T28411D<br>T28412D,T28419D,T2849XD,T285XXD,T286XXD,T287XXD,T288XXD<br>T2890XD,T28911D,T28912D,T28919D,T2899XD | |
| CNS abscess | A066,A1781,A5482,B431,G060,G061,G062,G07 | 0065,01320,01330,01340,01350,09889,1172,3240,3241,3249 |
| COVID-19 | J1282,U071,U099 | |
| COVID-19 acute | J1282,U071 | |
| COVID-19 long-term | U099 | |
| Calculus of urinary tract | N132,N200,N201,N202,N209,N210,N211,N218,N219,N22 | 591,5920,5921,5929,5940,5941,5942,5948,5949 |
| Cancer of other sites | C761,C762,C763,C7640,C7641,C7642,C7650,C7651,C7652,C768,D098 | 1951,1952,1953,1954,1955,1958,2348 |
| Cannabis-related disorders | F1210,F12120,F12121,F12122,F12129,F1213,F12150,F12151,F12159<br>F12180,F12188,F1219,F1220,F12220,F12221,F12222,F12229,F1223<br>F12250,F12251,F12259,F12280,F12288,F1229,F1290,F1291,F12920<br>F12921,F12922,F12929,F1293,F12950,F12951,F12959,F12980,F12988<br>F1299,T40711A,T40712A,T40713A,T40714A,T40715A,T40721A,T40722A<br>T40723A,T40724A,T40725A,T407X1A,T407X2A,T407X3A,T407X4A<br>T407X5A | 29211,29212,2922,29281,29289,2929,30430,30431,30432,30520<br>30521,30522,30523,9696,99529,E8541,E9396,E9502,E9620,E9802 |
| Cardiac and circulatory congenital anomalies | P095,Q200,Q201,Q202,Q203,Q204,Q205,Q206,Q208,Q209,Q210,Q211<br>Q2110,Q2111,Q2112,Q2113,Q2114,Q2115,Q2116,Q2119,Q212,Q2120<br>Q2121,Q2122,Q2123,Q213,Q214,Q218,Q219,Q220,Q221,Q222,Q223<br>Q224,Q225,Q226,Q228,Q229,Q230,Q231,Q232,Q233,Q234,Q238,Q239 | 7450,74510,74511,74512,74519,7452,7453,7454,7455,74560,74561<br>74569,7457,7458,7459,74600,74601,74602,74609,7461,7462,7463<br>7464,7465,7466,7467,74681,74682,74683,74684,74685,74686,74687<br>74689,7469,7470,74710,74720,74721,74722,74729,74731,74732 |

Continued on next page

**Supplementary Table S1:** (cont'd) A comprehensive mapping of diagnosis groups used in the experiments to their corresponding ICD-10 and ICD-9 codes.

| Diagnosis | ICD-10 | ICD-9 |
|---|---|---|
| | Q240,Q241,Q242,Q243,Q244,Q245,Q246,Q248,Q249,Q250,Q251,Q252<br>Q2521,Q2529,Q253,Q254,Q2540,Q2541,Q2542,Q2543,Q2544,Q2545<br>Q2546,Q2547,Q2548,Q2549,Q255,Q256,Q2571,Q2572,Q2579,Q258,Q259<br>Q260,Q261,Q262,Q263,Q264,Q265,Q266,Q268,Q269,Q270,Q271,Q272<br>Q2730,Q2731,Q2732,Q2733,Q2734,Q2739,Q274,Q278,Q279,Q280,Q281<br>Q282,Q283,Q288,Q289 | 74739,74740,74741,74742,74749,7475,74760,74761,74762,74763<br>74764,74769,74781,74789,7479 |
| Cardiac arrest and ventricular fibrillation | I462,I468,I469,I4901,I4902,I97120,I97121,I97710,I97711,O0336<br>O0386,O0486,O0736,O0881,O29111,O29112,O29113,O29119 | 42741,42742,4275,4294,63470,63471,63472,63572,6387,6398,64680<br>64681,64683,9971 |
| Cardiac cancers | C380,C381,C382,C383,C388 | 1641,1642,1643,1648,1649 |
| Cardiac dysrhythmias | I470,I471,I472,I4720,I4721,I4729,I479,I480,I481,I4811,I4819<br>I482,I4820,I4821,I483,I484,I4891,I4892,I491,I492,I493,I4940<br>I4949,I495,I498,I499 | 4270,4271,4272,42731,42732,42760,42761,42769,42781,42789,4279 |
| Carrier status | O99820,O99824,O99825,O99830,O99834,O99835,Z220,Z221,Z222<br>Z2231,Z22321,Z22322,Z22330,Z22338,Z2239,Z224,Z2250,Z2251<br>Z2252,Z2259,Z226,Z227,Z228,Z229 | 64781,64783,64784,64891,64893,64894,V020,V021,V022,V023,V024<br>V0251,V0252,V0253,V0254,V0259,V0260,V0261,V0262,V0269,V027<br>V028,V029 |
| Cataract and other lens disorders | E0836,E0936,E1036,E1136,E1336,H25011,H25012,H25013,H25019<br>H25031,H25032,H25033,H25039,H25041,H25042,H25043,H25049<br>H25091,H25092,H25093,H25099,H2510,H2511,H2512,H2513,H2520<br>H2521,H2522,H2523,H25811,H25812,H25813,H25819,H2589,H259<br>H26001,H26002,H26003,H26009,H26011,H26012,H26013,H26019<br>H26031,H26032,H26033,H26039,H26041,H26042,H26043,H26049<br>H26051,H26052,H26053,H26059,H26061,H26062,H26063,H26069,H2609<br>H26101,H26102,H26103,H26109,H26111,H26112,H26113,H26119<br>H26121,H26122,H26123,H26129,H26131,H26132,H26133,H26139,H2620<br>H26211,H26212,H26213,H26219,H26221,H26222,H26223,H26229<br>H26231,H26232,H26233,H26239,H2630,H2631,H2632,H2633,H2640<br>H26411,H26412,H26413,H26419,H26491,H26492,H26493,H26499,H268<br>H269,H2700,H2701,H2702,H2703,H2710,H27111,H27112,H27113<br>H27119,H27121,H27122,H27123,H27129,H27131,H27132,H27133<br>H27139,H278,H279,H28 | 24950,25050,25051,36600,36601,36602,36603,36604,36609,36610<br>36612,36613,36614,36615,36616,36618,36619,36620,36621,36622<br>36623,36630,36631,36632,36633,36634,36641,36644,36645,36646<br>36650,36651,36652,36653,3668,3669,37931,37932,37933,37934<br>37939 |
| Cerebral infarction | G43601,G43609,G43611,G43619,I6300,I63011,I63012,I63013,I63019<br>I6302,I63031,I63032,I63033,I63039,I6309,I6310,I63111,I63112<br>I63113,I63119,I6312,I63131,I63132,I63133,I63139,I6319,I6320 | 34660,34661,34662,34663,43301,43311,43321,43331,43381,43391<br>43401,43411,43491,99702 |

Continued on next page

**Supplementary Table S1:** (cont'd) A comprehensive mapping of diagnosis groups used in the experiments to their corresponding ICD-10 and ICD-9 codes.

| Diagnosis | ICD-10 | ICD-9 |
|---|---|---|
| | I63211,I63212,I63213,I63219,I6322,I63231,I63232,I63233,I63239<br>I6329,I6330,I63311,I63312,I63313,I63319,I63321,I63322,I63323<br>I63329,I63331,I63332,I63333,I63339,I63341,I63342,I63343<br>I63349,I6339,I6340,I63411,I63412,I63413,I63419,I63421,I63422<br>I63423,I63429,I63431,I63432,I63433,I63439,I63441,I63442<br>I63443,I63449,I6349,I6350,I63511,I63512,I63513,I63519,I63521<br>I63522,I63523,I63529,I63531,I63532,I63533,I63539,I63541<br>I63542,I63543,I63549,I6359,I636,I638,I6381,I6389,I639,I97810<br>I97811,I97820,I97821,R29700,R29701,R29702,R29703,R29704<br>R29705,R29706,R29707,R29708,R29709,R29710,R29711,R29712<br>R29713,R29714,R29715,R29716,R29717,R29718,R29719,R29720<br>R29721,R29722,R29723,R29724,R29725,R29726,R29727,R29728<br>R29729,R29730,R29731,R29732,R29733,R29734,R29735,R29736<br>R29737,R29738,R29739,R29740,R29741,R29742 | |
| Cerebral palsy | G800,G801,G802,G803,G804,G808,G809 | 33371,3430,3431,3432,3433,3434,3438,3439 |
| Chromosomal abnormalities | Q900,Q901,Q902,Q909,Q910,Q911,Q912,Q913,Q914,Q915,Q916,Q917<br>Q920,Q921,Q922,Q925,Q9261,Q9262,Q927,Q928,Q929,Q930,Q931,Q932<br>Q933,Q934,Q935,Q9351,Q9359,Q937,Q9381,Q9382,Q9388,Q9389,Q939<br>Q950,Q951,Q952,Q953,Q955,Q958,Q959,Q960,Q961,Q962,Q963,Q964<br>Q968,Q969,Q970,Q971,Q972,Q973,Q978,Q979,Q980,Q981,Q983,Q984<br>Q985,Q986,Q987,Q988,Q989,Q990,Q991,Q992,Q998,Q999 | 7580,7581,7582,75831,75832,75833,75839,7584,7585,7586,7587<br>75881,75889,7589,75983 |
| Chronic kidney disease | D631,E0822,E0922,E1022,E1122,E1322,I120,I129,I130,I1310,I1311<br>I132,N181,N182,N183,N1830,N1831,N1832,N184,N185,N186,N189<br>R880,Z4901,Z4902,Z4931,Z4932 | 24940,25040,25041,28521,40300,40301,40310,40311,40390,40391<br>40400,40401,40402,40403,40410,40411,40412,40413,40490,40491<br>40492,40493,58181,58381,5851,5852,5853,5854,5855,5856,5859<br>7925,V561,V562,V5631,V5632 |
| Chronic obstructive pulmonary disease and bronchiectasis | J410,J411,J418,J42,J430,J431,J432,J438,J439,J440,J441,J449<br>J470,J471,J479 | 4910,4911,49120,49121,49122,4918,4919,4920,4928,49320,49321<br>49322,4940,4941,496 |
| Chronic phlebitis; thrombophlebitis and thromboembolism | I82211,I82221,I82291,I82501,I82502,I82503,I82509,I82511<br>I82512,I82513,I82519,I82521,I82522,I82523,I82529,I82531<br>I82532,I82533,I82539,I82541,I82542,I82543,I82549,I82551<br>I82552,I82553,I82559,I82561,I82562,I82563,I82569,I82591<br>I82592,I82593,I82599,I825Y1,I825Y2,I825Y3,I825Y9,I825Z1<br>I825Z2,I825Z3,I825Z9,I82701,I82702,I82703,I82709,I82711<br>I82712,I82713,I82719,I82721,I82722,I82723,I82729,I82891,I8291 | 4532,45350,45351,45352,45371,45372,45373,45374,45375,45376<br>45377,45379 |

Continued on next page

**Supplementary Table S1:** (cont'd) A comprehensive mapping of diagnosis groups used in the experiments to their corresponding ICD-10 and ICD-9 codes.

| Diagnosis | ICD-10 | ICD-9 |
|---|---|---|
| | I82A21,I82A22,I82A23,I82A29,I82B21,I82B22,I82B23,I82B29 I82C21,I82C22,I82C23,I82C29 | |
| Chronic rheumatic heart disease | I050,I051,I052,I058,I059,I060,I061,I062,I068,I069,I070,I071 I072,I078,I079,I080,I081,I082,I083,I088,I089,I090,I091,I092 I0981,I0989,I099 | 393,3940,3941,3942,3949,3950,3951,3952,3959,3960,3961,3962 3963,3968,3969,3970,3971,3979,3980,39890,39891,39899 |
| Circulatory signs and symptoms | R000,R001,R002,R008,R009,R010,R011,R012,R030,R031,R0989,R58 | 42781,42789,4590,7850,7851,7852,7853,7859,7867,7962,7963 |
| Cleft lip or palate | Q351,Q353,Q355,Q357,Q359,Q360,Q361,Q369,Q370,Q371,Q372,Q373 Q374,Q375,Q378,Q379 | 74900,74902,74910,74911,74912,74913,74914,74920,74921,74922 74923,74924 |
| Coagulation and hemorrhagic disorders | D473,D65,D66,D67,D680,D6800,D6801,D68020,D68021,D68022,D68023 D68029,D6803,D6804,D6809,D681,D682,D68311,D68312,D68318,D6832 D684,D6851,D6852,D6859,D6861,D6862,D6869,D688,D689,D690,D691 D692,D693,D6941,D6942,D6949,D6951,D6959,D696,D698,D699,D7582 D75821,D75822,D75828,D75829,D75838,D75839,D7584,M362,O723 | 23871,2860,2861,2862,2863,2864,28652,28653,28659,2866,2867 2869,2870,2871,2872,28731,28732,28733,28739,28741,28749,2875 2878,2879,28981,28982,28984,66632,66634,7132 |
| Coma; stupor; and brain damage | B150,B160,B162,B1711,B190,B1911,B1921,E035,E0801,E0811,E08641 E0901,E0911,E09641,E1011,E10641,E1101,E1111,E11641,E1301 E1311,E13641,E15,G931,K7041,K7111,K7201,K7211,K7291,R400,R401 R4020,R403 | 0700,07020,07021,07041,0706,07071,24920,24930,25020,25030 25031,2510,3481,570,5713,5722,5728,5733,78001,78003,78009 |
| Complication of cardiovascular device, implant or graft, initial encounter | T8201XA,T8202XA,T8203XA,T8209XA,T82110A,T82111A,T82118A T82119A,T82120A,T82121A,T82128A,T82129A,T82190A,T82191A T82198A,T82199A,T82211A,T82212A,T82213A,T82218A,T82221A T82222A,T82223A,T82228A,T82310A,T82311A,T82312A,T82318A T82319A,T82320A,T82321A,T82322A,T82328A,T82329A,T82330A T82331A,T82332A,T82338A,T82339A,T82390A,T82391A,T82392A T82398A,T82399A,T8241XA,T8242XA,T8243XA,T8249XA,T82510A T82511A,T82512A,T82513A,T82514A,T82515A,T82518A,T82519A T82520A,T82521A,T82522A,T82523A,T82524A,T82525A,T82528A T82529A,T82530A,T82531A,T82532A,T82533A,T82534A,T82535A T82538A,T82539A,T82590A,T82591A,T82592A,T82593A,T82594A T82595A,T82598A,T82599A,T826XXA,T827XXA,T82817A,T82818A T82827A,T82828A,T82837A,T82838A,T82847A,T82848A,T82855A T82856A,T82857A,T82858A,T82867A,T82868A,T82897A,T82898A T829XXA | 99600,99601,99602,99603,99604,99609,9961,99661,99662,99671 99672,99673,99674 |

Continued on next page

**Supplementary Table S1:** (cont'd) A comprehensive mapping of diagnosis groups used in the experiments to their corresponding ICD-10 and ICD-9 codes.

| Diagnosis | ICD-10 | ICD-9 |
|---|---|---|
| Complication of cardiovascular device, implant or graft, subsequent encounter | T8201XD,T8202XD,T8203XD,T8209XD,T82110D,T82111D,T82118D T82119D,T82120D,T82121D,T82128D,T82129D,T82190D,T82191D T82198D,T82199D,T82211D,T82212D,T82213D,T82218D,T82221D T82222D,T82223D,T82228D,T82310D,T82311D,T82312D,T82318D T82319D,T82320D,T82321D,T82322D,T82328D,T82329D,T82330D T82331D,T82332D,T82338D,T82339D,T82390D,T82391D,T82392D T82398D,T82399D,T8241XD,T8242XD,T8243XD,T8249XD,T82510D T82511D,T82512D,T82513D,T82514D,T82515D,T82518D,T82519D T82520D,T82521D,T82522D,T82523D,T82524D,T82525D,T82528D T82529D,T82530D,T82531D,T82532D,T82533D,T82534D,T82535D T82538D,T82539D,T82590D,T82591D,T82592D,T82593D,T82594D T82595D,T82598D,T82599D,T826XXD,T827XXD,T82817D,T82818D T82827D,T82828D,T82837D,T82838D,T82847D,T82848D,T82855D T82856D,T82857D,T82858D,T82867D,T82868D,T82897D,T82898D T829XXD | V5889 |
| Complication of genitourinary device, implant or graft, initial encounter | T83010A,T83011A,T83012A,T83018A,T83020A,T83021A,T83022A T83028A,T83030A,T83031A,T83032A,T83038A,T83090A,T83091A T83092A,T83098A,T83110A,T83111A,T83112A,T83113A,T83118A T83120A,T83121A,T83122A,T83123A,T83128A,T83190A,T83191A T83192A,T83193A,T83198A,T8321XA,T8322XA,T8323XA,T8324XA T8325XA,T8329XA,T8331XA,T8332XA,T8339XA,T83410A,T83411A T83418A,T83420A,T83421A,T83428A,T83490A,T83491A,T83498A T83510A,T83511A,T83512A,T83518A,T8351XA,T83590A,T83591A T83592A,T83593A,T83598A,T8359XA,T8361XA,T8362XA,T8369XA T836XXA,T83711A,T83712A,T83713A,T83714A,T83718A,T83719A T83721A,T83722A,T83723A,T83724A,T83728A,T83729A,T8379XA T8381XA,T8382XA,T8383XA,T8384XA,T8385XA,T8386XA,T8389XA T839XXA | 62931,62932,99631,99632,99639,99664,99665,99676 |
| Complication of genitourinary device, implant or graft, subsequent encounter | T83010D,T83011D,T83012D,T83018D,T83020D,T83021D,T83022D T83028D,T83030D,T83031D,T83032D,T83038D,T83090D,T83091D T83092D,T83098D,T83110D,T83111D,T83112D,T83113D,T83118D T83120D,T83121D,T83122D,T83123D,T83128D,T83190D,T83191D T83192D,T83193D,T83198D,T8321XD,T8322XD,T8323XD,T8324XD T8325XD,T8329XD,T8331XD,T8332XD,T8339XD,T83410D,T83411D T83418D,T83420D,T83421D,T83428D,T83490D,T83491D,T83498D T83510D,T83511D,T83512D,T83518D,T8351XD,T83590D,T83591D T83592D,T83593D,T83598D,T8359XD,T8361XD,T8362XD,T8369XD T836XXD,T83711D,T83712D,T83713D,T83714D,T83718D,T83719D | V5889 |

Continued on next page

**Supplementary Table S1:** (cont'd) A comprehensive mapping of diagnosis groups used in the experiments to their corresponding ICD-10 and ICD-9 codes.

| Diagnosis | ICD-10 | ICD-9 |
|---|---|---|
| | T83721D,T83722D,T83723D,T83724D,T83728D,T83729D,T8379XD<br>T8381XD,T8382XD,T8383XD,T8384XD,T8385XD,T8386XD,T8389XD<br>T839XXD | |
| Complication of internal orthopedic device or implant, initial encounter | T84010A,T84011A,T84012A,T84013A,T84018A,T84019A,T84020A<br>T84021A,T84022A,T84023A,T84028A,T84029A,T84030A,T84031A<br>T84032A,T84033A,T84038A,T84039A,T84050A,T84051A,T84052A<br>T84053A,T84058A,T84059A,T84060A,T84061A,T84062A,T84063A<br>T84068A,T84069A,T84090A,T84091A,T84092A,T84093A,T84098A<br>T84099A,T84110A,T84111A,T84112A,T84113A,T84114A,T84115A<br>T84116A,T84117A,T84119A,T84120A,T84121A,T84122A,T84123A<br>T84124A,T84125A,T84126A,T84127A,T84129A,T84190A,T84191A<br>T84192A,T84193A,T84194A,T84195A,T84196A,T84197A,T84199A<br>T84210A,T84213A,T84216A,T84218A,T84220A,T84223A,T84226A<br>T84228A,T84290A,T84293A,T84296A,T84298A,T84310A,T84318A<br>T84320A,T84328A,T84390A,T84398A,T84428A,T84498A,T8450XA<br>T8451XA,T8452XA,T8453XA,T8454XA,T8459XA,T8460XA,T84610A<br>T84611A,T84612A,T84613A,T84614A,T84615A,T84619A,T84620A<br>T84621A,T84622A,T84623A,T84624A,T84625A,T84629A,T8463XA<br>T8469XA,T847XXA,T8481XA,T8482XA,T8483XA,T8484XA,T8485XA<br>T8486XA,T8489XA,T849XXA | 99641,99642,99643,99645,99646,99647,99649,99666,99667,99677<br>99678 |
| Complication of internal orthopedic device or implant, subsequent encounter | T84010D,T84011D,T84012D,T84013D,T84018D,T84019D,T84020D<br>T84021D,T84022D,T84023D,T84028D,T84029D,T84030D,T84031D<br>T84032D,T84033D,T84038D,T84039D,T84050D,T84051D,T84052D<br>T84053D,T84058D,T84059D,T84060D,T84061D,T84062D,T84063D<br>T84068D,T84069D,T84090D,T84091D,T84092D,T84093D,T84098D<br>T84099D,T84110D,T84111D,T84112D,T84113D,T84114D,T84115D<br>T84116D,T84117D,T84119D,T84120D,T84121D,T84122D,T84123D<br>T84124D,T84125D,T84126D,T84127D,T84129D,T84190D,T84191D<br>T84192D,T84193D,T84194D,T84195D,T84196D,T84197D,T84199D<br>T84210D,T84213D,T84216D,T84218D,T84220D,T84223D,T84226D<br>T84228D,T84290D,T84293D,T84296D,T84298D,T84310D,T84318D<br>T84320D,T84328D,T84390D,T84398D,T84418D,T84428D,T84498D<br>T8450XD,T8451XD,T8452XD,T8453XD,T8454XD,T8459XD,T8460XD<br>T84610D,T84611D,T84612D,T84613D,T84614D,T84615D,T84619D<br>T84620D,T84621D,T84622D,T84623D,T84624D,T84625D,T84629D<br>T8463XD,T8469XD,T847XXD,T8481XD,T8482XD,T8483XD,T8484XD<br>T8485XD,T8486XD,T8489XD,T849XXD | V5889 |

Continued on next page

**Supplementary Table S1:** (cont'd) A comprehensive mapping of diagnosis groups used in the experiments to their corresponding ICD-10 and ICD-9 codes.

| Diagnosis | ICD-10 | ICD-9 |
|---|---|---|
| Complication of other surgical or medical care, injury, initial encounter | T800XXA,T801XXA,T80211A,T80212A,T80218A,T80219A,T8022XA<br>T8029XA,T8030XA,T80310A,T80311A,T80319A,T8039XA,T8040XA<br>T80410A,T80411A,T80419A,T8049XA,T8051XA,T8052XA,T8059XA<br>T8061XA,T8062XA,T8069XA,T80810A,T80818A,T8082XA,T8089XA<br>T8090XA,T80910A,T80911A,T80919A,T8092XA,T80A0XA,T80A10A<br>T80A11A,T80A19A,T80A9XA,T8110XA,T8111XA,T8112XA,T8119XA<br>T8130XA,T8131XA,T8132XA,T8133XA,T8140XA,T8141XA,T8142XA<br>T8143XA,T8144XA,T8149XA,T814XXA,T81500A,T81501A,T81502A<br>T81503A,T81504A,T81505A,T81506A,T81507A,T81508A,T81509A<br>T81510A,T81511A,T81512A,T81513A,T81514A,T81515A,T81516A<br>T81517A,T81518A,T81519A,T81520A,T81521A,T81522A,T81523A<br>T81524A,T81525A,T81526A,T81527A,T81528A,T81529A,T81530A<br>T81531A,T81532A,T81533A,T81534A,T81535A,T81536A,T81537A<br>T81538A,T81539A,T81590A,T81591A,T81592A,T81593A,T81594A<br>T81595A,T81596A,T81597A,T81598A,T81599A,T8160XA,T8161XA<br>T8169XA,T81710A,T81711A,T81718A,T81719A,T8172XA,T8181XA<br>T8182XA,T8183XA,T8189XA,T819XXA,T84410A,T84418A,T84420A<br>T84490A,T8501XA,T8502XA,T8503XA,T8509XA,T85110A,T85111A<br>T85112A,T85113A,T85118A,T85120A,T85121A,T85122A,T85123A<br>T85128A,T85190A,T85191A,T85192A,T85193A,T85199A,T8521XA<br>T8522XA,T8529XA,T85310A,T85311A,T85318A,T85320A,T85321A<br>T85328A,T85390A,T85391A,T85398A,T8541XA,T8542XA,T8543XA<br>T8544XA,T8549XA,T85510A,T85511A,T85518A,T85520A,T85521A<br>T85528A,T85590A,T85591A,T85598A,T85610A,T85611A,T85612A<br>T85613A,T85614A,T85615A,T85618A,T85620A,T85621A,T85622A<br>T85623A,T85624A,T85625A,T85628A,T85630A,T85631A,T85633A<br>T85635A,T85638A,T85690A,T85691A,T85692A,T85693A,T85694A<br>T85695A,T85698A,T8571XA,T8572XA,T85730A,T85731A,T85732A<br>T85733A,T85734A,T85735A,T85738A,T8579XA,T85810A,T85818A<br>T8581XA,T85820A,T85828A,T8582XA,T85830A,T85838A,T8583XA<br>T85840A,T85848A,T8584XA,T85850A,T85858A,T8585XA,T85860A<br>T85868A,T8586XA,T85890A,T85898A,T8589XA,T859XXA,T8730,T8731<br>T8732,T8733,T8734,T8740,T8741,T8742,T8743,T8744,T8750,T8751<br>T8752,T8753,T8754,T8781,T8789,T879,T880XXA,T881XXA,T882XXA<br>T883XXA,T884XXA,T8851XA,T8852XA,T8853XA,T8859XA,T886XXA<br>T887XXA,T888XXA,T889XXA | 9950,99522,99524,9954,99586,99589,9962,99649,99651,99653<br>99654,99655,99656,99657,99659,99668,99669,99670,99675,99679<br>9972,99760,99761,99762,99769,99771,99772,99779,99800,99801<br>99802,99809,99830,99831,99832,99833,9984,99859,9986,9987<br>99881,99889,9989,9990,9991,9992,99931,99932,99933,99934,99939<br>99941,99942,99949,99951,99952,99959,99960,99961,99962,99963<br>99969,99970,99971,99972,99973,99974,99975,99976,99977,99978<br>99979,99980,99981,99982,99983,99984,99985,99988,99989,9999 |
| Complication of other surgical or medical care, injury, | T800XXD,T801XXD,T80211D,T80212D,T80218D,T80219D,T8022XD<br>T8029XD,T8030XD,T80310D,T80311D,T80319D,T8039XD,T8040XD<br>T80410D,T80411D,T80419D,T8049XD,T8051XD,T8052XD,T8059XD | V5889 |

Continued on next page

**Supplementary Table S1:** (cont'd) A comprehensive mapping of diagnosis groups used in the experiments to their corresponding ICD-10 and ICD-9 codes.

| Diagnosis | ICD-10 | ICD-9 |
|---|---|---|
| subsequent encounter | T8061XD,T8062XD,T8069XD,T80810D,T80818D,T8082XD,T8089XD<br>T8090XD,T80910D,T80911D,T80919D,T8092XD,T80A0XD,T80A10D<br>T80A11D,T80A19D,T80A9XD,T8110XD,T8111XD,T8112XD,T8119XD<br>T8130XD,T8131XD,T8132XD,T8133XD,T8140XD,T8141XD,T8142XD<br>T8143XD,T8144XD,T8149XD,T814XXD,T81500D,T81501D,T81502D<br>T81503D,T81504D,T81505D,T81506D,T81507D,T81508D,T81509D<br>T81510D,T81511D,T81512D,T81513D,T81514D,T81515D,T81516D<br>T81517D,T81518D,T81519D,T81520D,T81521D,T81522D,T81523D<br>T81524D,T81525D,T81526D,T81527D,T81528D,T81529D,T81530D<br>T81531D,T81532D,T81533D,T81534D,T81535D,T81536D,T81537D<br>T81538D,T81539D,T81590D,T81591D,T81592D,T81593D,T81594D<br>T81595D,T81596D,T81597D,T81598D,T81599D,T8160XD,T8161XD<br>T8169XD,T81710D,T81711D,T81718D,T81719D,T8172XD,T8181XD<br>T8182XD,T8183XD,T8189XD,T819XXD,T84410D,T84420D,T84490D<br>T8501XD,T8502XD,T8503XD,T8509XD,T85110D,T85111D,T85112D<br>T85113D,T85118D,T85120D,T85121D,T85122D,T85123D,T85128D<br>T85190D,T85191D,T85192D,T85193D,T85199D,T8521XD,T8522XD<br>T8529XD,T85310D,T85311D,T85318D,T85320D,T85321D,T85328D<br>T85390D,T85391D,T85398D,T8541XD,T8542XD,T8543XD,T8544XD<br>T8549XD,T85510D,T85511D,T85518D,T85520D,T85521D,T85528D<br>T85590D,T85591D,T85598D,T85610D,T85611D,T85612D,T85613D<br>T85614D,T85615D,T85618D,T85620D,T85621D,T85622D,T85623D<br>T85624D,T85625D,T85628D,T85630D,T85631D,T85633D,T85635D<br>T85638D,T85690D,T85691D,T85692D,T85693D,T85694D,T85695D<br>T85698D,T8571XD,T8572XD,T85730D,T85731D,T85732D,T85733D<br>T85734D,T85735D,T85738D,T8579XD,T85810D,T85818D,T8581XD<br>T85820D,T85828D,T8582XD,T85830D,T85838D,T8583XD,T85840D<br>T85848D,T8584XD,T85850D,T85858D,T8585XD,T85860D,T85868D<br>T8586XD,T85890D,T85898D,T8589XD,T859XXD,T880XXD,T881XXD<br>T882XXD,T883XXD,T884XXD,T8851XD,T8852XD,T8853XD,T8859XD<br>T886XXD,T887XXD,T888XXD,T889XXD | |
| Complication of transplanted organs or tissue, initial encounter | D89810,D89811,D89812,D89813,M3111,T8600,T8601,T8602,T8603<br>T8609,T8610,T8611,T8612,T8613,T8619,T8620,T8621,T8622,T8623<br>T86290,T86298,T8630,T8631,T8632,T8633,T8639,T8640,T8641,T8642<br>T8643,T8649,T865,T86810,T86811,T86812,T86818,T86819,T86820<br>T86821,T86822,T86828,T86829,T86830,T86831,T86832,T86838<br>T86839,T86840,T868401,T868402,T868403,T868409,T86841,T868411<br>T868412,T868413,T868419,T86842,T868421,T868422,T868423<br>T868429,T86848,T868481,T868482,T868483,T868489,T86849,T868491 | 27950,27951,27952,27953,99651,99652,99669,99679,99680,99681<br>99682,99683,99684,99685,99686,99687,99688,99689,99694,99696<br>99699 |

Continued on next page

**Supplementary Table S1:** (cont'd) A comprehensive mapping of diagnosis groups used in the experiments to their corresponding ICD-10 and ICD-9 codes.

| Diagnosis | ICD-10 | ICD-9 |
|---|---|---|
| | T868492,T868493,T868499,T86850,T86851,T86852,T86858,T86859 | |
| | T86890,T86891,T86892,T86898,T86899,T8690,T8691,T8692,T8693 | |
| | T8699,T870X1,T870X2,T870X9,T871X1,T871X2,T871X9,T872 | |
| Complication, sequela | T800XXS,T801XXS,T80211S,T80212S,T80218S,T80219S,T8022XS | 9093,9095 |
| | T8029XS,T8030XS,T80310S,T80311S,T80319S,T8039XS,T8040XS | |
| | T80410S,T80411S,T80419S,T8049XS,T8051XS,T8052XS,T8059XS | |
| | T8061XS,T8062XS,T8069XS,T80810S,T80818S,T8082XS,T8089XS | |
| | T8090XS,T80910S,T80911S,T80919S,T8092XS,T80A0XS,T80A10S | |
| | T80A11S,T80A19S,T80A9XS,T8110XS,T8111XS,T8112XS,T8119XS | |
| | T8130XS,T8131XS,T8132XS,T8133XS,T8140XS,T8141XS,T8142XS | |
| | T8143XS,T8144XS,T8149XS,T814XXS,T81500S,T81501S,T81502S | |
| | T81503S,T81504S,T81505S,T81506S,T81507S,T81508S,T81509S | |
| | T81510S,T81511S,T81512S,T81513S,T81514S,T81515S,T81516S | |
| | T81517S,T81518S,T81519S,T81520S,T81521S,T81522S,T81523S | |
| | T81524S,T81525S,T81526S,T81527S,T81528S,T81529S,T81530S | |
| | T81531S,T81532S,T81533S,T81534S,T81535S,T81536S,T81537S | |
| | T81538S,T81539S,T81590S,T81591S,T81592S,T81593S,T81594S | |
| | T81595S,T81596S,T81597S,T81598S,T81599S,T8160XS,T8161XS | |
| | T8169XS,T81710S,T81711S,T81718S,T81719S,T8172XS,T8181XS | |
| | T8182XS,T8183XS,T8189XS,T819XXS,T8201XS,T8202XS,T8203XS | |
| | T8209XS,T82110S,T82111S,T82118S,T82119S,T82120S,T82121S | |
| | T82128S,T82129S,T82190S,T82191S,T82198S,T82199S,T82211S | |
| | T82212S,T82213S,T82218S,T82221S,T82222S,T82223S,T82228S | |
| | T82310S,T82311S,T82312S,T82318S,T82319S,T82320S,T82321S | |
| | T82322S,T82328S,T82329S,T82330S,T82331S,T82332S,T82338S | |
| | T82339S,T82390S,T82391S,T82392S,T82398S,T82399S,T8241XS | |
| | T8242XS,T8243XS,T8249XS,T82510S,T82511S,T82512S,T82513S | |
| | T82514S,T82515S,T82518S,T82519S,T82520S,T82521S,T82522S | |
| | T82523S,T82524S,T82525S,T82528S,T82529S,T82530S,T82531S | |
| | T82532S,T82533S,T82534S,T82535S,T82538S,T82539S,T82590S | |
| | T82591S,T82592S,T82593S,T82594S,T82595S,T82598S,T82599S | |
| | T826XXS,T827XXS,T82817S,T82818S,T82827S,T82828S,T82837S | |
| | T82838S,T82847S,T82848S,T82855S,T82856S,T82857S,T82858S | |
| | T82867S,T82868S,T82897S,T82898S,T829XXS,T83010S,T83011S | |
| | T83012S,T83018S,T83020S,T83021S,T83022S,T83028S,T83030S | |
| | T83031S,T83032S,T83038S,T83090S,T83091S,T83092S,T83098S | |
| | T83110S,T83111S,T83112S,T83113S,T83118S,T83120S,T83121S | |
| | T83122S,T83123S,T83128S,T83190S,T83191S,T83192S,T83193S | |
| | T83198S,T8321XS,T8322XS,T8323XS,T8324XS,T8325XS,T8329XS | |

Continued on next page

**Supplementary Table S1:** (cont'd) A comprehensive mapping of diagnosis groups used in the experiments to their corresponding ICD-10 and ICD-9 codes.

| Diagnosis | ICD-10 | ICD-9 |
|---|---|---|
| | T8331XS,T8332XS,T8339XS,T83410S,T83411S,T83418S,T83420S | |
| | T83421S,T83428S,T83490S,T83491S,T83498S,T83510S,T83511S | |
| | T83512S,T83518S,T8351XS,T83590S,T83591S,T83592S,T83593S | |
| | T83598S,T8359XS,T8361XS,T8362XS,T8369XS,T836XXS,T83711S | |
| | T83712S,T83713S,T83714S,T83718S,T83719S,T83721S,T83722S | |
| | T83723S,T83724S,T83728S,T83729S,T8379XS,T8381XS,T8382XS | |
| | T8383XS,T8384XS,T8385XS,T8386XS,T8389XS,T839XXS,T84010S | |
| | T84011S,T84012S,T84013S,T84018S,T84019S,T84020S,T84021S | |
| | T84022S,T84023S,T84028S,T84029S,T84030S,T84031S,T84032S | |
| | T84033S,T84038S,T84039S,T84050S,T84051S,T84052S,T84053S | |
| | T84058S,T84059S,T84060S,T84061S,T84062S,T84063S,T84068S | |
| | T84069S,T84090S,T84091S,T84092S,T84093S,T84098S,T84099S | |
| | T84110S,T84111S,T84112S,T84113S,T84114S,T84115S,T84116S | |
| | T84117S,T84119S,T84120S,T84121S,T84122S,T84123S,T84124S | |
| | T84125S,T84126S,T84127S,T84129S,T84190S,T84191S,T84192S | |
| | T84193S,T84194S,T84195S,T84196S,T84197S,T84199S,T84210S | |
| | T84213S,T84216S,T84218S,T84220S,T84223S,T84226S,T84228S | |
| | T84290S,T84293S,T84296S,T84298S,T84310S,T84318S,T84320S | |
| | T84328S,T84390S,T84398S,T84410S,T84418S,T84420S,T84428S | |
| | T84490S,T84498S,T8450XS,T8451XS,T8452XS,T8453XS,T8454XS | |
| | T8459XS,T8460XS,T84610S,T84611S,T84612S,T84613S,T84614S | |
| | T84615S,T84619S,T84620S,T84621S,T84622S,T84623S,T84624S | |
| | T84625S,T84629S,T8463XS,T8469XS,T847XXS,T8481XS,T8482XS | |
| | T8483XS,T8484XS,T8485XS,T8486XS,T8489XS,T849XXS,T8501XS | |
| | T8502XS,T8503XS,T8509XS,T85110S,T85111S,T85112S,T85113S | |
| | T85118S,T85120S,T85121S,T85122S,T85123S,T85128S,T85190S | |
| | T85191S,T85192S,T85193S,T85199S,T8521XS,T8522XS,T8529XS | |
| | T85310S,T85311S,T85318S,T85320S,T85321S,T85328S,T85390S | |
| | T85391S,T85398S,T8541XS,T8542XS,T8543XS,T8544XS,T8549XS | |
| | T85510S,T85511S,T85518S,T85520S,T85521S,T85528S,T85590S | |
| | T85591S,T85598S,T85610S,T85611S,T85612S,T85613S,T85614S | |
| | T85615S,T85618S,T85620S,T85621S,T85622S,T85623S,T85624S | |
| | T85625S,T85628S,T85630S,T85631S,T85633S,T85635S,T85638S | |
| | T85690S,T85691S,T85692S,T85693S,T85694S,T85695S,T85698S | |
| | T8571XS,T8572XS,T85730S,T85731S,T85732S,T85733S,T85734S | |
| | T85735S,T85738S,T8579XS,T85810S,T85818S,T8581XS,T85820S | |
| | T85828S,T8582XS,T85830S,T85838S,T8583XS,T85840S,T85848S | |
| | T8584XS,T85850S,T85858S,T8585XS,T85860S,T85868S,T8586XS | |
| | T85890S,T85898S,T8589XS,T859XXS,T880XXS,T881XXS,T882XXS | |
| | T883XXS,T884XXS,T8851XS,T8852XS,T8853XS,T8859XS,T886XXS | |

Continued on next page

**Supplementary Table S1:** (cont'd) A comprehensive mapping of diagnosis groups used in the experiments to their corresponding ICD-10 and ICD-9 codes.

| Diagnosis | ICD-10 | ICD-9 |
|---|---|---|
| | T887XXS,T888XXS,T889XXS | |
| Complications following ectopic and or molar pregnancy | O080,O081,O082,O083,O084,O085,O086,O087,O0881,O0882,O0883<br>O0889,O089 | 6390,6391,6392,6393,6394,6395,6396,6398,6399 |
| Complications of acute myocardial infarction | I230,I231,I232,I233,I234,I235,I236,I237,I238,I241 | 4110,4295,4296,42971,42979 |
| Complications specified during childbirth | A34,O1002,O1012,O1022,O1032,O1042,O1092,O114,O1204,O1214<br>O1224,O134,O1404,O1414,O1424,O1494,O151,O164,O2402,O2412<br>O2432,O24420,O24424,O24425,O24429,O2482,O2492,O252,O2662<br>O2672,O670,O678,O679,O711,O713,O714,O715,O716,O717,O7181<br>O7189,O719,O730,O731,O740,O741,O742,O743,O744,O745,O746,O747<br>O748,O749,O750,O751,O752,O753,O754,O755,O7581,O7582,O7589<br>O759,O76,O770,O82,O860,O8600,O8601,O8602,O8603,O8604,O8609<br>O8802,O8812,O8822,O8832,O8882,O9802,O9812,O9822,O9832,O9842<br>O9852,O9862,O9872,O9882,O9892,O9902,O9912,O99214,O99284<br>O99314,O99324,O99334,O99344,O99354,O9942,O9952,O9962,O9972<br>O99814,O99824,O99834,O99844,O99892,O9A12,O9A22,O9A32,O9A42<br>O9A52 | 6390,64131,64181,64191,64201,64211,64221,64261,64671,64681<br>64701,64711,64721,64731,64741,64761,64781,64791,64801,64821<br>64831,64841,64881,64891,64901,64911,64921,64931,64981,64982<br>65681,65831,65921,65931,65971,65981,65991,66451,66454,66511<br>66531,66534,66541,66544,66551,66554,66561,66564,66571,66572<br>66574,66581,66582,66584,66591,66592,66594,66700,66702,66704<br>66710,66712,66714,66801,66802,66811,66812,66821,66822,66881<br>66882,66891,66892,66901,66902,66904,66911,66912,66914,66941<br>66942,66944,66971,66981,66982,66984,66991,66992,66994,67301<br>67302,67311,67312,67321,67322,67331,67332,67381,67382,67401<br>67402,67432,67434,V0251 |
| Complications specified during the puerperium | F53,F530,F531,M830,O1003,O1013,O1023,O1033,O1043,O1093,O115<br>O1205,O1215,O1225,O135,O1405,O1415,O1425,O1495,O152,O165<br>O2403,O2413,O2433,O24430,O24434,O24435,O24439,O2483,O2493<br>O253,O2663,O2673,O712,O720,O721,O722,O723,O85,O8611,O8612<br>O8613,O8619,O8620,O8621,O8622,O8629,O864,O8681,O8689,O870<br>O871,O872,O873,O874,O878,O879,O8803,O8813,O8823,O8833,O8883<br>O8901,O8909,O891,O892,O893,O894,O895,O896,O898,O899,O900,O901<br>O902,O904,O905,O906,O9081,O9089,O909,O9102,O9103,O9112,O9113<br>O9122,O9123,O9202,O9203,O9212,O9213,O9220,O9229,O923,O924<br>O925,O926,O9270,O9279,O9803,O9813,O9823,O9833,O9843,O9853<br>O9863,O9873,O9883,O9893,O9903,O9913,O99215,O99285,O99315<br>O99325,O99335,O99345,O99355,O9943,O9953,O9963,O9973,O99815<br>O99825,O99835,O99845,O99893,O9A13,O9A23,O9A33,O9A43,O9A53 | 2682,29389,64204,64214,64224,64244,64264,64272,64274,64644<br>64664,64682,64684,64702,64704,64712,64714,64722,64724,64732<br>64734,64742,64744,64762,64764,64782,64784,64792,64794,64802<br>64804,64812,64814,64822,64824,64832,64834,64842,64844,64882<br>64884,64894,64902,64904,64912,64914,64922,64924,64934,66522<br>66524,66602,66604,66612,66614,66622,66624,66632,66634,66802<br>66804,66814,66824,66882,66884,66892,66894,66932,66934,67012<br>67014,67022,67024,67032,67034,67082,67084,67102,67104,67122<br>67124,67142,67144,67152,67154,67182,67184,67192,67194,67202<br>67204,67302,67304,67312,67314,67322,67324,67332,67334,67382<br>67384,67404,67412,67414,67422,67424,67432,67444,67482,67484<br>67492,67494,67502,67504,67512,67514,67522,67524,67582,67584<br>67602,67604,67612,67614,67622,67632,67634,67642,67644,67652<br>67654,67662,67664,67682,67684,67692,67694,67904,V0251 |
| Conditions due to neoplasm or the | D61810,D630,D6481,D701,E883,G131,G731,G893,H4742,H47521<br>H47522,H47529,H47631,H47632,H47639,J910,K1231,M360,M361 | 27788,28411,28522,2853,28803,3383,3573,35831,37752,37761<br>37771,51181,52801,7103,7138,73310,73311,73312,73313,73314 |

Continued on next page

**Supplementary Table S1:** (cont'd) A comprehensive mapping of diagnosis groups used in the experiments to their corresponding ICD-10 and ICD-9 codes.

| Diagnosis | ICD-10 | ICD-9 |
|---|---|---|
| treatment of neoplasm | M8450XA,M84511A,M84512A,M84519A,M84521A,M84522A,M84529A<br>M84531A,M84532A,M84533A,M84534A,M84539A,M84541A,M84542A<br>M84549A,M84550A,M84551A,M84552A,M84553A,M84559A,M84561A<br>M84562A,M84563A,M84564A,M84569A,M84571A,M84572A,M84573A<br>M84574A,M84575A,M84576A,M8458XA,N5236,R180,R530,T80810A<br>T80810D,T80810S | 73315,73316,73319,78079,78951,9093,99981,V5889 |
| Conduction disorders | I440,I441,I442,I4430,I4439,I444,I445,I4460,I4469,I447,I450<br>I4510,I4519,I452,I453,I454,I455,I456,I4581,I4589,I459,Z45010<br>Z45018,Z950 | 4260,42610,42611,42612,42613,4262,4263,4264,42650,42651,42652<br>42653,42654,4266,4267,42681,42682,42689,4269,V4501,V5331 |
| Congenital malformations of eye, ear, face, neck | Q100,Q101,Q102,Q103,Q104,Q105,Q106,Q107,Q110,Q111,Q112,Q113<br>Q120,Q121,Q122,Q123,Q124,Q128,Q129,Q130,Q131,Q132,Q133,Q134<br>Q135,Q1381,Q1389,Q139,Q140,Q141,Q142,Q143,Q148,Q149,Q150,Q158<br>Q159,Q160,Q161,Q162,Q163,Q164,Q165,Q169,Q170,Q171,Q172,Q173<br>Q174,Q175,Q178,Q179,Q180,Q181,Q182,Q183,Q184,Q185,Q186,Q187<br>Q188,Q189 | 36514,74300,74303,74306,74310,74320,74321,74322,74330,74335<br>74336,74337,74339,74341,74343,74344,74345,74346,74347,74349<br>74351,74352,74353,74356,74357,74359,74361,74362,74363,74365<br>74366,74369,7438,7439,74400,74401,74402,74403,74404,74405<br>74409,7441,74421,74422,74423,74424,74429,7443,74441,74442<br>74443,74446,74447,74449,7445,74481,74482,74483,74484,74489<br>7449 |
| Contact dermatitis | L22,L230,L231,L232,L233,L234,L235,L236,L237,L2381,L2389,L239<br>L240,L241,L242,L243,L244,L245,L246,L247,L2481,L2489,L249<br>L24A0,L24A1,L24A2,L24A9,L24B0,L24B1,L24B2,L24B3,L250,L251<br>L252,L253,L254,L255,L258,L259,L568,L569,L578,L579,L580,L581<br>L589,L590,L598,L599 | 6910,6920,6921,6922,6923,6924,6925,6926,69270,69274,69279<br>69281,69282,69283,69284,69289,6929 |
| Contraceptive and procreative management | Z30011,Z30012,Z30013,Z30014,Z30015,Z30016,Z30017,Z30018<br>Z30019,Z3002,Z3009,Z302,Z3040,Z3041,Z3042,Z30430,Z30431<br>Z30432,Z30433,Z3044,Z3045,Z3046,Z3049,Z308,Z309,Z310,Z3141<br>Z3142,Z31430,Z31438,Z31440,Z31441,Z31448,Z3149,Z315,Z3161<br>Z3162,Z3169,Z317,Z3181,Z3182,Z3183,Z3184,Z3189,Z319,Z975<br>Z9851,Z9852 | V2501,V2502,V2503,V2504,V2509,V2511,V2512,V2513,V252,V253<br>V2540,V2541,V2542,V2543,V2549,V255,V258,V259,V260,V261,V2621<br>V2622,V2629,V2631,V2632,V2633,V2634,V2635,V2639,V2641,V2642<br>V2649,V2651,V2652,V2681,V2682,V2689,V269,V4551,V4559 |
| Cornea and external disease | A1851,A1852,A3686,A5031,A5431,A5433,A740,B0052,B0053,B0181<br>B0231,B0233,B0234,B0581,B300,B301,B302,B303,B308,B309,B6012<br>B6013,H00011,H00012,H00013,H00014,H00015,H00016,H00019,H00021<br>H00022,H00023,H00024,H00025,H00026,H00029,H00031,H00032<br>H00033,H00034,H00035,H00036,H00039,H0011,H0012,H0013,H0014<br>H0015,H0016,H0019,H01001,H01002,H01003,H01004,H01005,H01006<br>H01009,H0100A,H0100B,H01011,H01012,H01013,H01014,H01015<br>H01016,H01019,H0101A,H0101B,H01021,H01022,H01023,H01024 | 01730,03281,0527,05321,05329,05442,05443,05449,05571,0770<br>0771,0772,0773,0774,0778,07798,07799,0903,09840,09843,13621<br>36014,37000,37001,37002,37003,37004,37005,37006,37007,37020<br>37021,37022,37023,37024,37031,37032,37033,37034,37035,37040<br>37044,37049,37050,37052,37054,37055,37059,37060,37061,37062<br>37063,37064,3708,3709,37100,37101,37102,37103,37104,37110<br>37111,37112,37113,37114,37115,37116,37120,37121,37122,37123<br>37124,37130,37131,37132,37133,37140,37141,37142,37143,37144 |

Continued on next page

**Supplementary Table S1:** (cont'd) A comprehensive mapping of diagnosis groups used in the experiments to their corresponding ICD-10 and ICD-9 codes.

| Diagnosis | ICD-10 | ICD-9 |
|---|---|---|
| | H01025,H01026,H01029,H0102A,H0102B,H01111,H01112,H01113 | 37145,37146,37148,37149,37150,37151,37152,37153,37154,37155 |
| | H01114,H01115,H01116,H01119,H01121,H01122,H01123,H01124 | 37157,37160,37161,37162,37170,37171,37172,37173,37181,37182 |
| | H01125,H01126,H01129,H01131,H01132,H01133,H01134,H01135 | 37189,3719,37200,37201,37202,37203,37204,37205,37206,37210 |
| | H01136,H01139,H01141,H01142,H01143,H01144,H01145,H01146 | 37211,37212,37213,37214,37215,37220,37221,37222,37230,37234 |
| | H01149,H018,H019,H02001,H02002,H02003,H02004,H02005,H02006 | 37239,37240,37241,37242,37243,37244,37245,37250,37251,37252 |
| | H02009,H02011,H02012,H02013,H02014,H02015,H02016,H02019 | 37253,37254,37255,37256,37261,37262,37263,37264,37271,37272 |
| | H02021,H02022,H02023,H02024,H02025,H02026,H02029,H02031 | 37273,37274,37275,37281,37289,3729,37300,37301,37302,37311 |
| | H02032,H02033,H02034,H02035,H02036,H02039,H02041,H02042 | 37312,37313,3732,37331,37332,37333,37334,3738,3739,37400 |
| | H02043,H02044,H02045,H02046,H02049,H02051,H02052,H02053 | 37401,37402,37403,37404,37405,37410,37411,37412,37413,37414 |
| | H02054,H02055,H02056,H02059,H02101,H02102,H02103,H02104 | 37420,37421,37422,37423,37513,37514,37516,37520,37521,37522 |
| | H02105,H02106,H02109,H02111,H02112,H02113,H02114,H02115 | 37589,37900,37901,37902,37903,37904,37905,37906,37907,37909 |
| | H02116,H02119,H02121,H02122,H02123,H02124,H02125,H02126 | 37911,37912,37913,37914,37915,37916,37919,7102 |
| | H02129,H02131,H02132,H02133,H02134,H02135,H02136,H02139 | |
| | H02141,H02142,H02143,H02144,H02145,H02146,H02149,H02151 | |
| | H02152,H02153,H02154,H02155,H02156,H02159,H02201,H02202 | |
| | H02203,H02204,H02205,H02206,H02209,H0220A,H0220B,H0220C | |
| | H02211,H02212,H02213,H02214,H02215,H02216,H02219,H0221A | |
| | H0221B,H0221C,H02221,H02222,H02223,H02224,H02225,H02226 | |
| | H02229,H0222A,H0222B,H0222C,H02231,H02232,H02233,H02234 | |
| | H02235,H02236,H02239,H0223A,H0223B,H0223C,H02881,H02882 | |
| | H02883,H02884,H02885,H02886,H02889,H0288A,H0288B,H04141 | |
| | H04142,H04143,H04149,H04151,H04152,H04153,H04159,H04161 | |
| | H04162,H04163,H04169,H0419,H04201,H04202,H04203,H04209,H04211 | |
| | H04212,H04213,H04219,H04221,H04222,H04223,H04229,H10011 | |
| | H10012,H10013,H10019,H10021,H10022,H10023,H10029,H1010,H1011 | |
| | H1012,H1013,H10211,H10212,H10213,H10219,H10221,H10222,H10223 | |
| | H10229,H10231,H10232,H10233,H10239,H1030,H1031,H1032,H1033 | |
| | H10401,H10402,H10403,H10409,H10411,H10412,H10413,H10419 | |
| | H10421,H10422,H10423,H10429,H10431,H10432,H10433,H10439,H1044 | |
| | H1045,H10501,H10502,H10503,H10509,H10511,H10512,H10513,H10519 | |
| | H10521,H10522,H10523,H10529,H10531,H10532,H10533,H10539 | |
| | H10811,H10812,H10813,H10819,H10821,H10822,H10823,H10829,H1089 | |
| | H109,H11001,H11002,H11003,H11009,H11011,H11012,H11013,H11019 | |
| | H11021,H11022,H11023,H11029,H11031,H11032,H11033,H11039 | |
| | H11041,H11042,H11043,H11049,H11051,H11052,H11053,H11059 | |
| | H11061,H11062,H11063,H11069,H1110,H11111,H11112,H11113,H11119 | |
| | H11121,H11122,H11123,H11129,H11131,H11132,H11133,H11139 | |
| | H11141,H11142,H11143,H11149,H11151,H11152,H11153,H11159 | |
| | H11211,H11212,H11213,H11219,H11221,H11222,H11223,H11229 | |
| | H11231,H11232,H11233,H11239,H11241,H11242,H11243,H11249,H1130 | |

Continued on next page

**Supplementary Table S1:** (cont'd) A comprehensive mapping of diagnosis groups used in the experiments to their corresponding ICD-10 and ICD-9 codes.

| Diagnosis | ICD-10 | ICD-9 |
|---|---|---|
| | H1131,H1132,H1133,H11411,H11412,H11413,H11419,H11421,H11422 | |
| | H11423,H11429,H11431,H11432,H11433,H11439,H11441,H11442 | |
| | H11443,H11449,H11811,H11812,H11813,H11819,H11821,H11822 | |
| | H11823,H11829,H1189,H119,H15001,H15002,H15003,H15009,H15011 | |
| | H15012,H15013,H15019,H15021,H15022,H15023,H15029,H15031 | |
| | H15032,H15033,H15039,H15041,H15042,H15043,H15049,H15051 | |
| | H15052,H15053,H15059,H15091,H15092,H15093,H15099,H15101 | |
| | H15102,H15103,H15109,H15111,H15112,H15113,H15119,H15121 | |
| | H15122,H15123,H15129,H15811,H15812,H15813,H15819,H15821 | |
| | H15822,H15823,H15829,H15831,H15832,H15833,H15839,H15841 | |
| | H15842,H15843,H15849,H15851,H15852,H15853,H15859,H1589,H159 | |
| | H16001,H16002,H16003,H16009,H16011,H16012,H16013,H16019 | |
| | H16021,H16022,H16023,H16029,H16031,H16032,H16033,H16039 | |
| | H16041,H16042,H16043,H16049,H16051,H16052,H16053,H16059 | |
| | H16061,H16062,H16063,H16069,H16071,H16072,H16073,H16079 | |
| | H16101,H16102,H16103,H16109,H16111,H16112,H16113,H16119 | |
| | H16121,H16122,H16123,H16129,H16131,H16132,H16133,H16139 | |
| | H16141,H16142,H16143,H16149,H16201,H16202,H16203,H16209 | |
| | H16211,H16212,H16213,H16219,H16221,H16222,H16223,H16229 | |
| | H16231,H16232,H16233,H16239,H16241,H16242,H16243,H16249 | |
| | H16251,H16252,H16253,H16259,H16261,H16262,H16263,H16269 | |
| | H16291,H16292,H16293,H16299,H16301,H16302,H16303,H16309 | |
| | H16311,H16312,H16313,H16319,H16321,H16322,H16323,H16329 | |
| | H16331,H16332,H16333,H16339,H16391,H16392,H16393,H16399 | |
| | H16401,H16402,H16403,H16409,H16411,H16412,H16413,H16419 | |
| | H16421,H16422,H16423,H16429,H16431,H16432,H16433,H16439 | |
| | H16441,H16442,H16443,H16449,H168,H169,H1700,H1701,H1702,H1703 | |
| | H1710,H1711,H1712,H1713,H17811,H17812,H17813,H17819,H17821 | |
| | H17822,H17823,H17829,H1789,H179,H18001,H18002,H18003,H18009 | |
| | H18011,H18012,H18013,H18019,H18021,H18022,H18023,H18029 | |
| | H18031,H18032,H18033,H18039,H18041,H18042,H18043,H18049 | |
| | H18051,H18052,H18053,H18059,H18061,H18062,H18063,H18069,H1810 | |
| | H1811,H1812,H1813,H1820,H18211,H18212,H18213,H18219,H18221 | |
| | H18222,H18223,H18229,H18231,H18232,H18233,H18239,H1830,H18311 | |
| | H18312,H18313,H18319,H18321,H18322,H18323,H18329,H18331 | |
| | H18332,H18333,H18339,H1840,H18411,H18412,H18413,H18419,H18421 | |
| | H18422,H18423,H18429,H1843,H18441,H18442,H18443,H18449,H18451 | |
| | H18452,H18453,H18459,H18461,H18462,H18463,H18469,H1849,H1850 | |
| | H18501,H18502,H18503,H18509,H1851,H18511,H18512,H18513,H18519 | |
| | H1852,H18521,H18522,H18523,H18529,H1853,H18531,H18532,H18533 | |

Continued on next page

**Supplementary Table S1:** (cont'd) A comprehensive mapping of diagnosis groups used in the experiments to their corresponding ICD-10 and ICD-9 codes.

| Diagnosis | ICD-10 | ICD-9 |
|---|---|---|
| | H18539,H1854,H18541,H18542,H18543,H18549,H1855,H18551,H18552<br>H18553,H18559,H1859,H18591,H18592,H18593,H18599,H18601,H18602<br>H18603,H18609,H18611,H18612,H18613,H18619,H18621,H18622<br>H18623,H18629,H1870,H18711,H18712,H18713,H18719,H18721,H18722<br>H18723,H18729,H18731,H18732,H18733,H18739,H18791,H18792<br>H18793,H18799,H18811,H18812,H18813,H18819,H18821,H18822<br>H18823,H18829,H18831,H18832,H18833,H18839,H18891,H18892<br>H18893,H18899,H189,M3501 | |
| Coronary atherosclerosis and other heart disease | I200,I201,I202,I208,I209,I240,I248,I249,I2510,I25110,I25111<br>I25112,I25118,I25119,I252,I255,I256,I25700,I25701,I25702<br>I25708,I25709,I25710,I25711,I25712,I25718,I25719,I25720<br>I25721,I25722,I25728,I25729,I25730,I25731,I25732,I25738<br>I25739,I25750,I25751,I25752,I25758,I25759,I25760,I25761<br>I25762,I25768,I25769,I25790,I25791,I25792,I25798,I25799<br>I25810,I25811,I25812,I2582,I2583,I2584,I2589,I259,Z951,Z955<br>Z9861 | 4111,41181,41189,412,4131,4139,41401,41402,41403,41404,41405<br>41406,41407,4142,4143,4144,4148,4149,4292,V4581,V4582 |
| Crushing injury, initial encounter | S070XXA,S071XXA,S078XXA,S079XXA,S170XXA,S178XXA,S179XXA<br>S280XXA,S38001A,S38002A,S3801XA,S3802XA,S3803XA,S381XXA<br>S471XXA,S472XXA,S479XXA,S5700XA,S5701XA,S5702XA,S5780XA<br>S5781XA,S5782XA,S6700XA,S6701XA,S6702XA,S6710XA,S67190A<br>S67191A,S67192A,S67193A,S67194A,S67195A,S67196A,S67197A<br>S67198A,S6720XA,S6721XA,S6722XA,S6730XA,S6731XA,S6732XA<br>S6740XA,S6741XA,S6742XA,S6790XA,S6791XA,S6792XA,S7700XA<br>S7701XA,S7702XA,S7710XA,S7711XA,S7712XA,S7720XA,S7721XA<br>S7722XA,S8700XA,S8701XA,S8702XA,S8780XA,S8781XA,S8782XA<br>S9700XA,S9701XA,S9702XA,S97101A,S97102A,S97109A,S97111A<br>S97112A,S97119A,S97121A,S97122A,S97129A,S9780XA,S9781XA<br>S9782XA | 9251,9252,9260,92611,92619,92700,92701,92702,92703,92710<br>92711,92720,92721,9273,9278,9279,92800,92801,92810,92811<br>92820,92821,9283,9288,9289,9290,9299 |
| Crushing injury, subsequent encounter | S070XXD,S071XXD,S078XXD,S079XXD,S170XXD,S178XXD,S179XXD<br>S280XXD,S38001D,S38002D,S3801XD,S3802XD,S3803XD,S381XXD<br>S471XXD,S472XXD,S479XXD,S5700XD,S5701XD,S5702XD,S5780XD<br>S5781XD,S5782XD,S6700XD,S6701XD,S6702XD,S6710XD,S67190D<br>S67191D,S67192D,S67193D,S67194D,S67195D,S67196D,S67197D<br>S67198D,S6720XD,S6721XD,S6722XD,S6730XD,S6731XD,S6732XD<br>S6740XD,S6741XD,S6742XD,S6790XD,S6791XD,S6792XD,S7700XD<br>S7701XD,S7702XD,S7710XD,S7711XD,S7712XD,S7720XD,S7721XD<br>S7722XD,S8700XD,S8701XD,S8702XD,S8780XD,S8781XD,S8782XD | V5889 |

Continued on next page

**Supplementary Table S1:** (cont'd) A comprehensive mapping of diagnosis groups used in the experiments to their corresponding ICD-10 and ICD-9 codes.

| Diagnosis | ICD-10 | ICD-9 |
|---|---|---|
| | S9700XD,S9701XD,S9702XD,S97101D,S97102D,S97109D,S97111D S97112D,S97119D,S97121D,S97122D,S97129D,S9780XD,S9781XD S9782XD | |
| Crystal arthropathies (excluding gout) | M1100,M11011,M11012,M11019,M11021,M11022,M11029,M11031,M11032 M11039,M11041,M11042,M11049,M11051,M11052,M11059,M11061 M11062,M11069,M11071,M11072,M11079,M1108,M1109,M1110,M11111 M11112,M11119,M11121,M11122,M11129,M11131,M11132,M11139 M11141,M11142,M11149,M11151,M11152,M11159,M11161,M11162 M11169,M11171,M11172,M11179,M1118,M1119,M1120,M11211,M11212 M11219,M11221,M11222,M11229,M11231,M11232,M11239,M11241 M11242,M11249,M11251,M11252,M11259,M11261,M11262,M11269 M11271,M11272,M11279,M1128,M1129,M1180,M11811,M11812,M11819 M11821,M11822,M11829,M11831,M11832,M11839,M11841,M11842 M11849,M11851,M11852,M11859,M11861,M11862,M11869,M11871 M11872,M11879,M1188,M1189,M119 | 71230,71231,71232,71233,71234,71235,71236,71237,71238,71239 71280,71281,71282,71283,71284,71285,71286,71287,71288,71289 71290,71291,71292,71293,71294,71295,71296,71297,71298 |
| Cystic fibrosis | E840,E8411,E8419,E848,E849,P094 | 27700,27701,27702,27703,27709 |
| Depressive disorders | F0631,F0632,F0634,F320,F321,F322,F323,F324,F328,F3281,F3289 F329,F32A,F330,F331,F332,F333,F3341,F338,F339,F341,F530,O906 | 29383,29620,29621,29622,29623,29624,29625,29630,29631,29632 29633,29634,29635,29682,29699,2980,3004,30112,311,64842,64844 |
| Diabetes mellitus with complication | E0800,E0801,E0810,E0811,E0821,E0822,E0829,E08311,E08319 E08321,E083211,E083212,E083213,E083219,E08329,E083291,E083292 E083293,E083299,E08331,E083311,E083312,E083313,E083319,E08339 E083391,E083392,E083393,E083399,E08341,E083411,E083412 E083413,E083419,E08349,E083491,E083492,E083493,E083499,E08351 E083511,E083512,E083513,E083519,E083521,E083522,E083523 E083529,E083531,E083532,E083533,E083539,E083541,E083542 E083543,E083549,E083551,E083552,E083553,E083559,E08359 E083591,E083592,E083593,E083599,E0836,E0837X1,E0837X2,E0837X3 E0837X9,E0839,E0840,E0841,E0842,E0843,E0844,E0849,E0851,E0852 E0859,E08610,E08618,E08620,E08621,E08622,E08628,E08630,E08638 E08641,E08649,E0865,E0869,E088,E0900,E0901,E0910,E0911,E0921 E0922,E0929,E09311,E09319,E09321,E093211,E093212,E093213 E093219,E09329,E093291,E093292,E093293,E093299,E09331,E093311 E093312,E093313,E093319,E09339,E093391,E093392,E093393 E093399,E09341,E093411,E093412,E093413,E093419,E09349,E093491 E093492,E093493,E093499,E09351,E093511,E093512,E093513 E093519,E093521,E093522,E093523,E093529,E093531,E093532 | 24910,24920,24930,24940,24950,24960,24970,24980,24990,25010 25011,25020,25030,25031,25040,25041,25050,25051,25060,25061 25070,25071,25080,25081,25090,25091,2511,34989,3535,3559,3572 36201,36202,36204,36205,36206,36207,36641,44381,5238,5289 5363,58181,58381,7098,7135,71680,7854 |

Continued on next page

**Supplementary Table S1:** (cont'd) A comprehensive mapping of diagnosis groups used in the experiments to their corresponding ICD-10 and ICD-9 codes.

| Diagnosis | ICD-10 | ICD-9 |
|---|---|---|
| | E093533,E093539,E093541,E093542,E093543,E093549,E093551<br>E093552,E093553,E093559,E09359,E093591,E093592,E093593<br>E093599,E0936,E0937X1,E0937X2,E0937X3,E0937X9,E0939,E0940<br>E0941,E0942,E0943,E0944,E0949,E0951,E0952,E0959,E09610,E09618<br>E09620,E09621,E09622,E09628,E09630,E09638,E09641,E09649,E0965<br>E0969,E098,E1010,E1011,E1021,E1022,E1029,E10311,E10319,E10321<br>E103211,E103212,E103213,E103219,E10329,E103291,E103292<br>E103293,E103299,E10331,E103311,E103312,E103313,E103319,E10339<br>E103391,E103392,E103393,E103399,E10341,E103411,E103412<br>E103413,E103419,E10349,E103491,E103492,E103493,E103499,E10351<br>E103511,E103512,E103513,E103519,E103521,E103522,E103523<br>E103529,E103531,E103532,E103533,E103539,E103541,E103542<br>E103543,E103549,E103551,E103552,E103553,E103559,E10359<br>E103591,E103592,E103593,E103599,E1036,E1037X1,E1037X2,E1037X3<br>E1037X9,E1039,E1040,E1041,E1042,E1043,E1044,E1049,E1051,E1052<br>E1059,E10610,E10618,E10620,E10621,E10622,E10628,E10630,E10638<br>E10641,E10649,E1065,E1069,E108,E1100,E1101,E1110,E1111,E1121<br>E1122,E1129,E11311,E11319,E11321,E113211,E113212,E113213<br>E113219,E11329,E113291,E113292,E113293,E113299,E11331,E113311<br>E113312,E113313,E113319,E11339,E113391,E113392,E113393<br>E113399,E11341,E113411,E113412,E113413,E113419,E11349,E113491<br>E113492,E113493,E113499,E11351,E113511,E113512,E113513<br>E113519,E113521,E113522,E113523,E113529,E113531,E113532<br>E113533,E113539,E113541,E113542,E113543,E113549,E113551<br>E113552,E113553,E113559,E11359,E113591,E113592,E113593<br>E113599,E1136,E1137X1,E1137X2,E1137X3,E1137X9,E1139,E1140<br>E1141,E1142,E1143,E1144,E1149,E1151,E1152,E1159,E11610,E11618<br>E11620,E11621,E11622,E11628,E11630,E11638,E11641,E11649,E1165<br>E1169,E118,E1300,E1301,E1310,E1311,E1321,E1322,E1329,E13311<br>E13319,E13321,E133211,E133212,E133213,E133219,E13329,E133291<br>E133292,E133293,E133299,E13331,E133311,E133312,E133313<br>E133319,E13339,E133391,E133392,E133393,E133399,E13341,E133411<br>E133412,E133413,E133419,E13349,E133491,E133492,E133493<br>E133499,E13351,E133511,E133512,E133513,E133519,E133521<br>E133522,E133523,E133529,E133531,E133532,E133533,E133539<br>E133541,E133542,E133543,E133549,E133551,E133552,E133553<br>E133559,E13359,E133591,E133592,E133593,E133599,E1336,E1337X1<br>E1337X2,E1337X3,E1337X9,E1339,E1340,E1341,E1342,E1343,E1344<br>E1349,E1351,E1352,E1359,E13610,E13618,E13620,E13621,E13622<br>E13628,E13630,E13638,E13641,E13649,E1365,E1369,E138 | |

Continued on next page

**Supplementary Table S1:** (cont'd) A comprehensive mapping of diagnosis groups used in the experiments to their corresponding ICD-10 and ICD-9 codes.

| Diagnosis | ICD-10 | ICD-9 |
|---|---|---|
| Diabetes mellitus without complication | E089,E099,E109,E119,E139,E891,O24011,O24012,O24013,O24019 O2402,O2403,O24111,O24112,O24113,O24119,O2412,O2413,O24311 O24312,O24313,O24319,O2432,O2433,O24410,O24414,O24415,O24419 O24420,O24424,O24425,O24429,O24430,O24434,O24435,O24439 O24811,O24812,O24813,O24819,O2482,O2483,O24911,O24912,O24913 O24919,O2492,O2493,O99810,O99814,O99815 | 24900,25000,25001,2513,64800,64801,64802,64803,64804,64881 64882,64883,64884 |
| Diabetes or abnormal glucose tolerance complicating pregnancy; childbirth; or the puerperium | O24011,O24012,O24013,O24019,O2402,O2403,O24111,O24112,O24113 O24119,O2412,O2413,O24311,O24312,O24313,O24319,O2432,O2433 O24410,O24414,O24415,O24419,O24420,O24424,O24425,O24429 O24430,O24434,O24435,O24439,O24811,O24812,O24813,O24819,O2482 O2483,O24911,O24912,O24913,O24919,O2492,O2493,O99810,O99814 O99815 | 64800,64801,64802,64803,64804,64881,64882,64883,64884 |
| Digestive congenital anomalies | Q380,Q381,Q382,Q383,Q384,Q385,Q386,Q387,Q388,Q390,Q391,Q392 Q393,Q394,Q395,Q396,Q398,Q399,Q400,Q401,Q402,Q403,Q408,Q409 Q410,Q411,Q412,Q418,Q419,Q420,Q421,Q422,Q423,Q428,Q429,Q430 Q431,Q432,Q433,Q434,Q435,Q436,Q437,Q438,Q439,Q440,Q441,Q442 Q443,Q444,Q445,Q446,Q447,Q450,Q451,Q452,Q453,Q458,Q459 | 7500,75010,75011,75012,75013,75015,75016,75019,75021,75022 75023,75024,75025,75026,75027,75029,7503,7504,7505,7506,7507 7508,7509,7510,7511,7512,7513,7514,7515,75160,75161,75162 75169,7517,7518,7519 |
| Diseases of inner ear and related conditions | H8000,H8001,H8002,H8003,H8010,H8011,H8012,H8013,H8020,H8021 H8022,H8023,H8080,H8081,H8082,H8083,H8090,H8091,H8092,H8093 H8101,H8102,H8103,H8109,H8110,H8111,H8112,H8113,H8120,H8121 H8122,H8123,H81311,H81312,H81313,H81319,H81391,H81392,H81393 H81399,H814,H8141,H8142,H8143,H8149,H818X1,H818X2,H818X3 H818X9,H8190,H8191,H8192,H8193,H821,H822,H823,H829,H8301 H8302,H8303,H8309,H8311,H8312,H8313,H8319,H832X1,H832X2 H832X3,H832X9,H833X1,H833X2,H833X3,H833X9,H838X1,H838X2 H838X3,H838X9,H8390,H8391,H8392,H8393 | 38600,38601,38602,38603,38604,38610,38611,38612,38619,3862 38630,38631,38632,38633,38634,38635,38640,38641,38642,38643 38648,38650,38651,38652,38653,38654,38655,38656,38658,3868 3869,3870,3871,3872,3878,3879,38810,38812 |
| Diseases of middle ear and mastoid (except otitis media) | A186,H68001,H68002,H68003,H68009,H68011,H68012,H68013,H68019 H68021,H68022,H68023,H68029,H68101,H68102,H68103,H68109 H68111,H68112,H68113,H68119,H68121,H68122,H68123,H68129 H68131,H68132,H68133,H68139,H6900,H6901,H6902,H6903,H6980 H6981,H6982,H6983,H6990,H6991,H6992,H6993,H70001,H70002 H70003,H70009,H70011,H70012,H70013,H70019,H70091,H70092 H70093,H70099,H7010,H7011,H7012,H7013,H70201,H70202,H70203 H70209,H70211,H70212,H70213,H70219,H70221,H70222,H70223 H70229,H70811,H70812,H70813,H70819,H70891,H70892,H70893 | 01740,38150,38151,38152,38160,38161,38162,38163,3817,38181 38189,3819,38300,38301,38302,3831,38320,38321,38322,38330 38331,38332,38333,38381,38389,3839,38400,38401,38409,3841 38420,38421,38422,38423,38424,38425,38481,38482,3849,38500 38501,38502,38503,38509,38510,38511,38512,38513,38519,38521 38522,38523,38524,38530,38531,38532,38533,38535,38589,3859 |

Continued on next page

**Supplementary Table S1:** (cont'd) A comprehensive mapping of diagnosis groups used in the experiments to their corresponding ICD-10 and ICD-9 codes.

| Diagnosis | ICD-10 | ICD-9 |
|---|---|---|
| | H70899,H7090,H7091,H7092,H7093,H7100,H7101,H7102,H7103,H7110 | |
| | H7111,H7112,H7113,H7120,H7121,H7122,H7123,H7130,H7131,H7132 | |
| | H7133,H7190,H7191,H7192,H7193,H7200,H7201,H7202,H7203,H7210 | |
| | H7211,H7212,H7213,H722X1,H722X2,H722X3,H722X9,H72811,H72812 | |
| | H72813,H72819,H72821,H72822,H72823,H72829,H7290,H7291,H7292 | |
| | H7293,H73001,H73002,H73003,H73009,H73011,H73012,H73013,H73019 | |
| | H73091,H73092,H73093,H73099,H7310,H7311,H7312,H7313,H7320 | |
| | H7321,H7322,H7323,H73811,H73812,H73813,H73819,H73821,H73822 | |
| | H73823,H73829,H73891,H73892,H73893,H73899,H7390,H7391,H7392 | |
| | H7393,H7401,H7402,H7403,H7409,H7411,H7412,H7413,H7419,H7420 | |
| | H7421,H7422,H7423,H74311,H74312,H74313,H74319,H74321,H74322 | |
| | H74323,H74329,H74391,H74392,H74393,H74399,H7440,H7441,H7442 | |
| | H7443,H748X1,H748X2,H748X3,H748X9,H7490,H7491,H7492,H7493 | |
| | H7500,H7501,H7502,H7503,H7580,H7581,H7582,H7583,H9500,H9501 | |
| | H9502,H9503,H95111,H95112,H95113,H95119,H95121,H95122,H95123 | |
| | H95129,H95131,H95132,H95133,H95139,H95191,H95192,H95193 | |
| | H95199 | |
| Diseases of mouth; | A690,B002,B084,B370,B3783,K098,K099,K110,K111,K1120,K1121 | 0542,0743,101,1120,5270,5271,5272,5273,5274,5275,5276,5277 |
| excluding dental | K1122,K1123,K113,K114,K115,K116,K117,K118,K119,K120,K121,K122 | 5278,5279,52800,52801,52802,52809,5281,5282,5283,5284,5285 |
| | K1230,K1231,K1232,K1233,K1239,K130,K131,K1321,K1322,K1323 | 5286,52871,52872,52879,5288,5289,5290,5291,5292,5293,5294 |
| | K1324,K1329,K133,K134,K135,K136,K1370,K1379,K140,K141,K142 | 5295,5296,5298,5299,6820 |
| | K143,K144,K145,K146,K148,K149,R682 | |
| Diseases of white | D700,D701,D702,D703,D704,D708,D709,D71,D720,D721,D7210,D72110 | 28800,28801,28802,28803,28804,28809,2881,2882,2883,2884,28850 |
| blood cells | D72111,D72118,D72119,D7212,D7218,D7219,D72810,D72818,D72819 | 28851,28859,28860,28861,28862,28863,28864,28865,28866,28869 |
| | D72820,D72821,D72822,D72823,D72824,D72825,D72828,D72829,D7289 | 2888,2889,28953 |
| | D729,D7381,D761,D762,D763 | |
| Dislocations, initial | S0300XA,S0301XA,S0302XA,S0303XA,S030XXA,S031XXA,S032XXA | 8300,83100,83101,83102,83103,83104,83109,83200,83201,83202 |
| encounter | S130XXA,S13100A,S13101A,S13110A,S13111A,S13120A,S13121A | 83203,83204,83209,8322,83300,83301,83302,83303,83304,83305 |
| | S13130A,S13131A,S13140A,S13141A,S13150A,S13151A,S13160A | 83309,83400,83401,83402,83500,83501,83502,83503,8363,83650 |
| | S13161A,S13170A,S13171A,S13180A,S13181A,S1320XA,S1329XA | 83651,83652,83653,83654,83659,8370,83800,83802,83803,83804 |
| | S230XXA,S23100A,S23101A,S23110A,S23111A,S23120A,S23121A | 83805,83806,83809,83900,83901,83902,83903,83904,83905,83906 |
| | S23122A,S23123A,S23130A,S23131A,S23132A,S23133A,S23140A | 83907,83908,83920,83921,83940,83941,83942,83949,83961,83969 |
| | S23141A,S23142A,S23143A,S23150A,S23151A,S23152A,S23153A | 8480,8485,87363 |
| | S23160A,S23161A,S23162A,S23163A,S23170A,S23171A,S2320XA | |
| | S2329XA,S330XXA,S33100A,S33101A,S33110A,S33111A,S33120A | |
| | S33121A,S33130A,S33131A,S33140A,S33141A,S332XXA,S3330XA | |
| | S3339XA,S334XXA,S43001A,S43002A,S43003A,S43004A,S43005A | |

Continued on next page

**Supplementary Table S1:** (cont'd) A comprehensive mapping of diagnosis groups used in the experiments to their corresponding ICD-10 and ICD-9 codes.

| Diagnosis | ICD-10 | ICD-9 |
|---|---|---|
| | S43006A,S43011A,S43012A,S43013A,S43014A,S43015A,S43016A | |
| | S43021A,S43022A,S43023A,S43024A,S43025A,S43026A,S43031A | |
| | S43032A,S43033A,S43034A,S43035A,S43036A,S43081A,S43082A | |
| | S43083A,S43084A,S43085A,S43086A,S43101A,S43102A,S43109A | |
| | S43111A,S43112A,S43119A,S43121A,S43122A,S43129A,S43131A | |
| | S43132A,S43139A,S43141A,S43142A,S43149A,S43151A,S43152A | |
| | S43159A,S43201A,S43202A,S43203A,S43204A,S43205A,S43206A | |
| | S43211A,S43212A,S43213A,S43214A,S43215A,S43216A,S43221A | |
| | S43222A,S43223A,S43224A,S43225A,S43226A,S43301A,S43302A | |
| | S43303A,S43304A,S43305A,S43306A,S43311A,S43312A,S43313A | |
| | S43314A,S43315A,S43316A,S43391A,S43392A,S43393A,S43394A | |
| | S43395A,S43396A,S53001A,S53002A,S53003A,S53004A,S53005A | |
| | S53006A,S53011A,S53012A,S53013A,S53014A,S53015A,S53016A | |
| | S53021A,S53022A,S53023A,S53024A,S53025A,S53026A,S53031A | |
| | S53032A,S53033A,S53091A,S53092A,S53093A,S53094A,S53095A | |
| | S53096A,S53101A,S53102A,S53103A,S53104A,S53105A,S53106A | |
| | S53111A,S53112A,S53113A,S53114A,S53115A,S53116A,S53121A | |
| | S53122A,S53123A,S53124A,S53125A,S53126A,S53131A,S53132A | |
| | S53133A,S53134A,S53135A,S53136A,S53141A,S53142A,S53143A | |
| | S53144A,S53145A,S53146A,S53191A,S53192A,S53193A,S53194A | |
| | S53195A,S53196A,S63001A,S63002A,S63003A,S63004A,S63005A | |
| | S63006A,S63011A,S63012A,S63013A,S63014A,S63015A,S63016A | |
| | S63021A,S63022A,S63023A,S63024A,S63025A,S63026A,S63031A | |
| | S63032A,S63033A,S63034A,S63035A,S63036A,S63041A,S63042A | |
| | S63043A,S63044A,S63045A,S63046A,S63051A,S63052A,S63053A | |
| | S63054A,S63055A,S63056A,S63061A,S63062A,S63063A,S63064A | |
| | S63065A,S63066A,S63071A,S63072A,S63073A,S63074A,S63075A | |
| | S63076A,S63091A,S63092A,S63093A,S63094A,S63095A,S63096A | |
| | S63101A,S63102A,S63103A,S63104A,S63105A,S63106A,S63111A | |
| | S63112A,S63113A,S63114A,S63115A,S63116A,S63121A,S63122A | |
| | S63123A,S63124A,S63125A,S63126A,S63131A,S63132A,S63133A | |
| | S63134A,S63135A,S63136A,S63141A,S63142A,S63143A,S63144A | |
| | S63145A,S63146A,S63200A,S63201A,S63202A,S63203A,S63204A | |
| | S63205A,S63206A,S63207A,S63208A,S63209A,S63210A,S63211A | |
| | S63212A,S63213A,S63214A,S63215A,S63216A,S63217A,S63218A | |
| | S63219A,S63220A,S63221A,S63222A,S63223A,S63224A,S63225A | |
| | S63226A,S63227A,S63228A,S63229A,S63230A,S63231A,S63232A | |
| | S63233A,S63234A,S63235A,S63236A,S63237A,S63238A,S63239A | |
| | S63240A,S63241A,S63242A,S63243A,S63244A,S63245A,S63246A | |
| | S63247A,S63248A,S63249A,S63250A,S63251A,S63252A,S63253A | |

Continued on next page

**Supplementary Table S1:** (cont'd) A comprehensive mapping of diagnosis groups used in the experiments to their corresponding ICD-10 and ICD-9 codes.

| Diagnosis | ICD-10 | ICD-9 |
|---|---|---|
| | S63254A,S63255A,S63256A,S63257A,S63258A,S63259A,S63260A | |
| | S63261A,S63262A,S63263A,S63264A,S63265A,S63266A,S63267A | |
| | S63268A,S63269A,S63270A,S63271A,S63272A,S63273A,S63274A | |
| | S63275A,S63276A,S63277A,S63278A,S63279A,S63280A,S63281A | |
| | S63282A,S63283A,S63284A,S63285A,S63286A,S63287A,S63288A | |
| | S63289A,S63290A,S63291A,S63292A,S63293A,S63294A,S63295A | |
| | S63296A,S63297A,S63298A,S63299A,S73001A,S73002A,S73003A | |
| | S73004A,S73005A,S73006A,S73011A,S73012A,S73013A,S73014A | |
| | S73015A,S73016A,S73021A,S73022A,S73023A,S73024A,S73025A | |
| | S73026A,S73031A,S73032A,S73033A,S73034A,S73035A,S73036A | |
| | S73041A,S73042A,S73043A,S73044A,S73045A,S73046A,S83001A | |
| | S83002A,S83003A,S83004A,S83005A,S83006A,S83011A,S83012A | |
| | S83013A,S83014A,S83015A,S83016A,S83091A,S83092A,S83093A | |
| | S83094A,S83095A,S83096A,S83101A,S83102A,S83103A,S83104A | |
| | S83105A,S83106A,S83111A,S83112A,S83113A,S83114A,S83115A | |
| | S83116A,S83121A,S83122A,S83123A,S83124A,S83125A,S83126A | |
| | S83131A,S83132A,S83133A,S83134A,S83135A,S83136A,S83141A | |
| | S83142A,S83143A,S83144A,S83145A,S83146A,S83191A,S83192A | |
| | S83193A,S83194A,S83195A,S83196A,S9301XA,S9302XA,S9303XA | |
| | S9304XA,S9305XA,S9306XA,S93101A,S93102A,S93103A,S93104A | |
| | S93105A,S93106A,S93111A,S93112A,S93113A,S93114A,S93115A | |
| | S93116A,S93119A,S93121A,S93122A,S93123A,S93124A,S93125A | |
| | S93126A,S93129A,S93131A,S93132A,S93133A,S93134A,S93135A | |
| | S93136A,S93139A,S93141A,S93142A,S93143A,S93144A,S93145A | |
| | S93146A,S93149A,S93301A,S93302A,S93303A,S93304A,S93305A | |
| | S93306A,S93311A,S93312A,S93313A,S93314A,S93315A,S93316A | |
| | S93321A,S93322A,S93323A,S93324A,S93325A,S93326A,S93331A | |
| | S93332A,S93333A,S93334A,S93335A,S93336A | |
| Dislocations, | S0300XD,S0301XD,S0302XD,S0303XD,S030XXD,S031XXD,S032XXD | V5889 |
| subsequent encounter | S130XXD,S13100D,S13101D,S13110D,S13111D,S13120D,S13121D | |
| | S13130D,S13131D,S13140D,S13141D,S13150D,S13151D,S13160D | |
| | S13161D,S13170D,S13171D,S13180D,S13181D,S1320XD,S1329XD | |
| | S230XXD,S23100D,S23101D,S23110D,S23111D,S23120D,S23121D | |
| | S23122D,S23123D,S23130D,S23131D,S23132D,S23133D,S23140D | |
| | S23141D,S23142D,S23143D,S23150D,S23151D,S23152D,S23153D | |
| | S23160D,S23161D,S23162D,S23163D,S23170D,S23171D,S2320XD | |
| | S2329XD,S330XXD,S33100D,S33101D,S33110D,S33111D,S33120D | |
| | S33121D,S33130D,S33131D,S33140D,S33141D,S332XXD,S3330XD | |
| | S3339XD,S334XXD,S43001D,S43002D,S43003D,S43004D,S43005D | |

Continued on next page

Supplementary Table S1: (cont'd) A comprehensive mapping of diagnosis groups used in the experiments to their corresponding ICD-10 and ICD-9 codes.

| Diagnosis | ICD-10 | ICD-9 |
|---|---|---|
| | S43006D,S43011D,S43012D,S43013D,S43014D,S43015D,S43016D | |
| | S43021D,S43022D,S43023D,S43024D,S43025D,S43026D,S43031D | |
| | S43032D,S43033D,S43034D,S43035D,S43036D,S43081D,S43082D | |
| | S43083D,S43084D,S43085D,S43086D,S43101D,S43102D,S43109D | |
| | S43111D,S43112D,S43119D,S43121D,S43122D,S43129D,S43131D | |
| | S43132D,S43139D,S43141D,S43142D,S43149D,S43151D,S43152D | |
| | S43159D,S43201D,S43202D,S43203D,S43204D,S43205D,S43206D | |
| | S43211D,S43212D,S43213D,S43214D,S43215D,S43216D,S43221D | |
| | S43222D,S43223D,S43224D,S43225D,S43226D,S43301D,S43302D | |
| | S43303D,S43304D,S43305D,S43306D,S43311D,S43312D,S43313D | |
| | S43314D,S43315D,S43316D,S43391D,S43392D,S43393D,S43394D | |
| | S43395D,S43396D,S53001D,S53002D,S53003D,S53004D,S53005D | |
| | S53006D,S53011D,S53012D,S53013D,S53014D,S53015D,S53016D | |
| | S53021D,S53022D,S53023D,S53024D,S53025D,S53026D,S53031D | |
| | S53032D,S53033D,S53091D,S53092D,S53093D,S53094D,S53095D | |
| | S53096D,S53101D,S53102D,S53103D,S53104D,S53105D,S53106D | |
| | S53111D,S53112D,S53113D,S53114D,S53115D,S53116D,S53121D | |
| | S53122D,S53123D,S53124D,S53125D,S53126D,S53131D,S53132D | |
| | S53133D,S53134D,S53135D,S53136D,S53141D,S53142D,S53143D | |
| | S53144D,S53145D,S53146D,S53191D,S53192D,S53193D,S53194D | |
| | S53195D,S53196D,S63001D,S63002D,S63003D,S63004D,S63005D | |
| | S63006D,S63011D,S63012D,S63013D,S63014D,S63015D,S63016D | |
| | S63021D,S63022D,S63023D,S63024D,S63025D,S63026D,S63031D | |
| | S63032D,S63033D,S63034D,S63035D,S63036D,S63041D,S63042D | |
| | S63043D,S63044D,S63045D,S63046D,S63051D,S63052D,S63053D | |
| | S63054D,S63055D,S63056D,S63061D,S63062D,S63063D,S63064D | |
| | S63065D,S63066D,S63071D,S63072D,S63073D,S63074D,S63075D | |
| | S63076D,S63091D,S63092D,S63093D,S63094D,S63095D,S63096D | |
| | S63101D,S63102D,S63103D,S63104D,S63105D,S63106D,S63111D | |
| | S63112D,S63113D,S63114D,S63115D,S63116D,S63121D,S63122D | |
| | S63123D,S63124D,S63125D,S63126D,S63131D,S63132D,S63133D | |
| | S63134D,S63135D,S63136D,S63141D,S63142D,S63143D,S63144D | |
| | S63145D,S63146D,S63200D,S63201D,S63202D,S63203D,S63204D | |
| | S63205D,S63206D,S63207D,S63208D,S63209D,S63210D,S63211D | |
| | S63212D,S63213D,S63214D,S63215D,S63216D,S63217D,S63218D | |
| | S63219D,S63220D,S63221D,S63222D,S63223D,S63224D,S63225D | |
| | S63226D,S63227D,S63228D,S63229D,S63230D,S63231D,S63232D | |
| | S63233D,S63234D,S63235D,S63236D,S63237D,S63238D,S63239D | |
| | S63240D,S63241D,S63242D,S63243D,S63244D,S63245D,S63246D | |
| | S63247D,S63248D,S63249D,S63250D,S63251D,S63252D,S63253D | |

Continued on next page

**Supplementary Table S1:** (cont'd) A comprehensive mapping of diagnosis groups used in the experiments to their corresponding ICD-10 and ICD-9 codes.

| Diagnosis | ICD-10 | ICD-9 |
|---|---|---|
| | S63254D,S63255D,S63256D,S63257D,S63258D,S63259D,S63260D | |
| | S63261D,S63262D,S63263D,S63264D,S63265D,S63266D,S63267D | |
| | S63268D,S63269D,S63270D,S63271D,S63272D,S63273D,S63274D | |
| | S63275D,S63276D,S63277D,S63278D,S63279D,S63280D,S63281D | |
| | S63282D,S63283D,S63284D,S63285D,S63286D,S63287D,S63288D | |
| | S63289D,S63290D,S63291D,S63292D,S63293D,S63294D,S63295D | |
| | S63296D,S63297D,S63298D,S63299D,S73001D,S73002D,S73003D | |
| | S73004D,S73005D,S73006D,S73011D,S73012D,S73013D,S73014D | |
| | S73015D,S73016D,S73021D,S73022D,S73023D,S73024D,S73025D | |
| | S73026D,S73031D,S73032D,S73033D,S73034D,S73035D,S73036D | |
| | S73041D,S73042D,S73043D,S73044D,S73045D,S73046D,S83001D | |
| | S83002D,S83003D,S83004D,S83005D,S83006D,S83011D,S83012D | |
| | S83013D,S83014D,S83015D,S83016D,S83091D,S83092D,S83093D | |
| | S83094D,S83095D,S83096D,S83101D,S83102D,S83103D,S83104D | |
| | S83105D,S83106D,S83111D,S83112D,S83113D,S83114D,S83115D | |
| | S83116D,S83121D,S83122D,S83123D,S83124D,S83125D,S83126D | |
| | S83131D,S83132D,S83133D,S83134D,S83135D,S83136D,S83141D | |
| | S83142D,S83143D,S83144D,S83145D,S83146D,S83191D,S83192D | |
| | S83193D,S83194D,S83195D,S83196D,S9301XD,S9302XD,S9303XD | |
| | S9304XD,S9305XD,S9306XD,S93101D,S93102D,S93103D,S93104D | |
| | S93105D,S93106D,S93111D,S93112D,S93113D,S93114D,S93115D | |
| | S93116D,S93119D,S93121D,S93122D,S93123D,S93124D,S93125D | |
| | S93126D,S93129D,S93131D,S93132D,S93133D,S93134D,S93135D | |
| | S93136D,S93139D,S93141D,S93142D,S93143D,S93144D,S93145D | |
| | S93146D,S93149D,S93301D,S93302D,S93303D,S93304D,S93305D | |
| | S93306D,S93311D,S93312D,S93313D,S93314D,S93315D,S93316D | |
| | S93321D,S93322D,S93323D,S93324D,S93325D,S93326D,S93331D | |
| | S93332D,S93333D,S93334D,S93335D,S93336D | |
| Disorders of jaw | K0820,K0821,K0822,K0823,K0824,K0825,K0826,M2600,M2601,M2602 | 52400,52401,52402,52403,52404,52405,52406,52407,52409,52410 |
| | M2603,M2604,M2605,M2606,M2607,M2609,M2610,M2611,M2612,M2619 | 52411,52412,52419,52420,52421,52422,52423,52424,52425,52426 |
| | M2620,M26211,M26212,M26213,M26219,M26220,M26221,M2623,M2624 | 52427,52428,52429,52430,52431,52432,52433,52434,52435,52436 |
| | M2625,M2629,M2630,M2631,M2632,M2633,M2634,M2635,M2636,M2637 | 52437,52439,5244,52450,52451,52452,52453,52454,52455,52456 |
| | M2639,M264,M2650,M2651,M2652,M2653,M2654,M2655,M2656,M2657 | 52457,52459,52460,52461,52462,52463,52469,52470,52471,52472 |
| | M2659,M2660,M26601,M26602,M26603,M26609,M2661,M26611,M26612 | 52473,52474,52479,52481,52482,52489,5249,52520,52521,52522 |
| | M26613,M26619,M2662,M26621,M26622,M26623,M26629,M2663,M26631 | 52523,52524,52525,52526,52571,52572,52573,52579,5258,5262 |
| | M26632,M26633,M26639,M26641,M26642,M26643,M26649,M26651 | 5263,5264,5265,52661,52662,52663,52669,52681,52689,5269 |
| | M26652,M26653,M26659,M2669,M2670,M2671,M2672,M2673,M2674 | |
| | M2679,M2681,M2682,M2689,M269,M270,M271,M272,M273,M2740,M2749 | |
| | M2751,M2752,M2753,M2759,M2761,M2762,M2763,M2769,M278,M279 | |

Continued on next page

**Supplementary Table S1:** (cont'd) A comprehensive mapping of diagnosis groups used in the experiments to their corresponding ICD-10 and ICD-9 codes.

| Diagnosis | ICD-10 | ICD-9 |
|---|---|---|
| Disorders of lipid metabolism | E780,E7800,E7801,E781,E782,E783,E784,E7841,E7849,E785 | 2720,2721,2722,2723,2724 |
| Disorders of teeth and gingiva | K000,K001,K002,K003,K004,K005,K006,K007,K008,K009,K010,K011<br>K023,K0251,K0252,K0253,K0261,K0262,K0263,K027,K029,K030,K031<br>K032,K033,K034,K035,K036,K037,K0381,K0389,K039,K040,K0401<br>K0402,K041,K042,K043,K044,K045,K046,K047,K048,K0490,K0499<br>K0500,K0501,K0510,K0511,K0520,K0521,K05211,K05212,K05213<br>K05219,K0522,K05221,K05222,K05223,K05229,K0530,K0531,K05311<br>K05312,K05313,K05319,K0532,K05321,K05322,K05323,K05329,K054<br>K055,K056,K060,K06010,K06011,K06012,K06013,K06020,K06021<br>K06022,K06023,K061,K062,K063,K068,K069,K080,K08101,K08102<br>K08103,K08104,K08109,K08111,K08112,K08113,K08114,K08119<br>K08121,K08122,K08123,K08124,K08129,K08131,K08132,K08133<br>K08134,K08139,K08191,K08192,K08193,K08194,K08199,K0820,K0821<br>K0822,K0823,K0824,K0825,K0826,K083,K08401,K08402,K08403<br>K08404,K08409,K08411,K08412,K08413,K08414,K08419,K08421<br>K08422,K08423,K08424,K08429,K08431,K08432,K08433,K08434<br>K08439,K08491,K08492,K08493,K08494,K08499,K0850,K0851,K0852<br>K08530,K08531,K08539,K0854,K0855,K0856,K0859,K088,K0881,K0882<br>K0889,K089,K090,K091,M2600,M2601,M2602,M2603,M2604,M2605<br>M2606,M2607,M2609,M2610,M2611,M2612,M2619,M2620,M26211,M26212<br>M26213,M26219,M26220,M26221,M2623,M2624,M2625,M2629,M2630<br>M2631,M2632,M2633,M2634,M2635,M2636,M2637,M2639,M264,M2650<br>M2651,M2652,M2653,M2654,M2655,M2656,M2657,M2659,M2670,M2671<br>M2672,M2673,M2674,M2679,M2681,M2682,M2689,M269,M273,M2751<br>M2752,M2753,M2759,M2761,M2762,M2763,M2769,M350C,S032XXA<br>S032XXD,S032XXS | 5200,5201,5202,5203,5204,5205,5206,5207,5208,5209,52100,52101<br>52102,52103,52104,52105,52106,52107,52108,52110,52120,52130<br>52140,52141,52142,5215,5216,5217,52181,52189,5219,5220,5221<br>5222,5223,5224,5225,5226,5227,5228,5229,52300,52301,52310<br>52311,52320,52330,52331,52332,52333,52340,52341,52342,5235<br>5236,5238,5239,52400,52401,52402,52403,52404,52405,52406<br>52407,52409,52410,52411,52412,52419,52420,52421,52422,52423<br>52424,52425,52426,52427,52428,52429,52430,52431,52432,52433<br>52434,52435,52436,52437,52439,5244,52450,52451,52452,52453<br>52454,52455,52456,52457,52459,52470,52471,52472,52473,52474<br>52479,52481,52482,52489,5249,5250,52510,52511,52512,52513<br>52519,52520,52521,52522,52523,52524,52525,52526,5253,52540<br>52541,52542,52543,52544,52550,52551,52552,52553,52554,52560<br>52561,52562,52563,52564,52565,52566,52567,52569,52571,52572<br>52573,52579,5258,5259,5260,5261,5265,52661,52662,52663,52669<br>87363,9060,V5889 |
| Disruptive, impulse-control and conduct disorders | F602,F631,F632,F6381,F6389,F639,F910,F911,F912,F913,F918,F919 | 3017,31200,31210,31220,31230,31232,31233,31234,31235,31239<br>31281,31282,31289,3129,31381 |
| Diverticulosis and diverticulitis | K5700,K5701,K5710,K5711,K5712,K5713,K5720,K5721,K5730,K5731<br>K5732,K5733,K5740,K5741,K5750,K5751,K5752,K5753,K5780,K5781<br>K5790,K5791,K5792,K5793 | 56200,56201,56202,56203,56210,56211,56212,56213,5695 |
| Drug induced or toxic related condition | D521,D590,D592,D611,D612,D61811,D642,D702,E032,E064,E160,E231<br>E242,E273,E661,G210,G2111,G2119,G2401,G2402,G2409,G251,G254 | 2442,2443,2454,2511,2537,2550,25541,2682,27400,27401,27402<br>27403,27800,2812,2830,28319,28412,28489,2850,28803,32371 |

Continued on next page

**Supplementary Table S1:** (cont'd) A comprehensive mapping of diagnosis groups used in the experiments to their corresponding ICD-10 and ICD-9 codes.

| Diagnosis | ICD-10 | ICD-9 |
|---|---|---|
| | G2561,G2570,G2571,G2579,G4440,G4441,G620,G622,G701,G7114,G720<br>G722,G92,G9200,G9201,G9202,G9203,G9204,G9205,G928,G929,H10211<br>H10212,H10213,H10219,H2630,H2631,H2632,H2633,H35381,H35382<br>H35383,H35389,H4060X0,H4060X1,H4060X2,H4060X3,H4060X4,H4061X0<br>H4061X1,H4061X2,H4061X3,H4061X4,H4062X0,H4062X1,H4062X2<br>H4062X3,H4062X4,H4063X0,H4063X1,H4063X2,H4063X3,H4063X4,H463<br>H9101,H9102,H9103,H9109,I427,I952,J702,J703,J704,K1232,K521<br>K5903,K5931,K710,K7110,K7111,K712,K713,K714,K7150,K7151,K716<br>K717,K718,K719,K853,K8530,K8531,K8532,L105,L244,L251,L432<br>L530,L640,M1020,M10211,M10212,M10219,M10221,M10222,M10229<br>M10231,M10232,M10239,M10241,M10242,M10249,M10251,M10252<br>M10259,M10261,M10262,M10269,M10271,M10272,M10279,M1028,M1029<br>M1A20X0,M1A20X1,M1A2110,M1A2111,M1A2120,M1A2121,M1A2190<br>M1A2191,M1A2210,M1A2211,M1A2220,M1A2221,M1A2290,M1A2291<br>M1A2310,M1A2311,M1A2320,M1A2321,M1A2390,M1A2391,M1A2410<br>M1A2411,M1A2420,M1A2421,M1A2490,M1A2491,M1A2510,M1A2511<br>M1A2520,M1A2521,M1A2590,M1A2591,M1A2610,M1A2611,M1A2620<br>M1A2621,M1A2690,M1A2691,M1A2710,M1A2711,M1A2720,M1A2721<br>M1A2790,M1A2791,M1A28X0,M1A28X1,M1A29X0,M1A29X1,M320,M342<br>M835,M8710,M87111,M87112,M87119,M87121,M87122,M87129,M87131<br>M87132,M87133,M87134,M87135,M87136,M87137,M87138,M87139<br>M87141,M87142,M87143,M87144,M87145,M87146,M87150,M87151<br>M87152,M87159,M87161,M87162,M87163,M87164,M87165,M87166<br>M87171,M87172,M87173,M87174,M87175,M87176,M87177,M87178<br>M87179,M87180,M87188,M8719,N140,N141,N1411,N1419,N142,N143<br>N144,N46021,N46121,N4833,N522,O771,P5841,P5842,P938,R330,R502 | 32372,3321,3331,3333,3335,33372,33379,33385,33392,33399,3393<br>34982,3576,3577,3582,35924,3594,36255,36531,36532,36645,37206<br>37734,3898,4259,45829,5088,52802,5582,5722,5733,5770,58389<br>6060,6061,6073,60784,65681,6923,6944,6950,6970,70409,7100<br>7101,73340,73341,73342,73343,73344,73345,73349,7741,7794<br>78060,78829,E9479 |
| Dysphagia | D501,R1310,R1311,R1312,R1313,R1314,R1319 | 2808,78720,78721,78722,78723,78724,78729 |
| Early or threatened labor | O26872,O26873,O26879,O3430,O3431,O3432,O3433,O4200,O42011<br>O42012,O42013,O42019,O4210,O42111,O42112,O42113,O42119,O4290<br>O42911,O42912,O42913,O42919,O4700,O4702,O4703,O471,O479,O6000<br>O6002,O6003,O6010X0,O6010X1,O6010X2,O6010X3,O6010X4,O6010X5<br>O6010X9,O6012X0,O6012X1,O6012X2,O6012X3,O6012X4,O6012X5<br>O6012X9,O6013X0,O6013X1,O6013X2,O6013X3,O6013X4,O6013X5<br>O6013X9,O6014X0,O6014X1,O6014X2,O6014X3,O6014X4,O6014X5<br>O6014X9,O6020X0,O6020X1,O6020X2,O6020X3,O6020X4,O6020X5<br>O6020X9,O6022X0,O6022X1,O6022X2,O6022X3,O6022X4,O6022X5<br>O6022X9,O6023X0,O6023X1,O6023X2,O6023X3,O6023X4,O6023X5<br>O6023X9 | 64400,64403,64410,64413,64420,64421,64970,64971,64973,65450<br>65451,65452,65453,65454,65810,65811,65813,65820,65821,65823 |

Continued on next page

**Supplementary Table S1:** (cont'd) A comprehensive mapping of diagnosis groups used in the experiments to their corresponding ICD-10 and ICD-9 codes.

| Diagnosis | ICD-10 | ICD-9 |
|---|---|---|
| Early, first or unspecified trimester hemorrhage | O200,O208,O209,O26851,O26859,O4410,O4411,O4430,O4431,O4450 O4451,O45001,O45009,O45011,O45019,O45021,O45029,O45091,O45099 O458X1,O458X9,O4590,O4591,O46001,O46009,O46011,O46019,O46021 O46029,O46091,O46099,O468X1,O468X9,O4690,O4691 | 64003,64083,64093,64110,64111,64113,64120,64121,64123,64130 64131,64133,64180,64181,64183,64190,64191,64193,64950,64951 64953 |
| Ectopic pregnancy and complications of ectopic pregnancy | O000,O0000,O0001,O001,O0010,O00101,O00102,O00109,O0011,O00111 O00112,O00119,O002,O0020,O00201,O00202,O00209,O0021,O00211 O00212,O00219,O008,O0080,O0081,O009,O0090,O0091,O3670X0 O3670X1,O3670X2,O3670X3,O3670X4,O3670X5,O3670X9,O3671X0 O3671X1,O3671X2,O3671X3,O3671X4,O3671X5,O3671X9,O3672X0 O3672X1,O3672X2,O3672X3,O3672X4,O3672X5,O3672X9,O3673X0 O3673X1,O3673X2,O3673X3,O3673X4,O3673X5,O3673X9 | 63300,63310,63320,63380,63390,65680,65681,65683 |
| Effect of foreign body entering opening, initial encounter | T1500XA,T1501XA,T1502XA,T1510XA,T1511XA,T1512XA,T1580XA T1581XA,T1582XA,T1590XA,T1591XA,T1592XA,T161XXA,T162XXA T169XXA,T170XXA,T171XXA,T17200A,T17208A,T17210A,T17218A T17220A,T17228A,T17290A,T17298A,T17300A,T17308A,T17310A T17318A,T17320A,T17328A,T17390A,T17398A,T17400A,T17408A T17410A,T17418A,T17420A,T17428A,T17490A,T17498A,T17500A T17508A,T17510A,T17518A,T17520A,T17528A,T17590A,T17598A T17800A,T17808A,T17810A,T17818A,T17820A,T17828A,T17890A T17898A,T17900A,T17908A,T17910A,T17918A,T17920A,T17928A T17990A,T17998A,T180XXA,T18100A,T18108A,T18110A,T18118A T18120A,T18128A,T18190A,T18198A,T182XXA,T183XXA,T184XXA T185XXA,T188XXA,T189XXA,T190XXA,T191XXA,T192XXA,T193XXA T194XXA,T198XXA,T199XXA | 9300,9301,9302,9308,9309,931,932,9330,9331,9340,9341,9348 9349,9350,9351,9352,936,937,938,9390,9391,9392,9393,9399,E911 E912,E914,E915 |
| Effect of foreign body entering opening, subsequent encounter | T1500XD,T1501XD,T1502XD,T1510XD,T1511XD,T1512XD,T1580XD T1581XD,T1582XD,T1590XD,T1591XD,T1592XD,T161XXD,T162XXD T169XXD,T170XXD,T171XXD,T17200D,T17208D,T17210D,T17218D T17220D,T17228D,T17290D,T17298D,T17300D,T17308D,T17310D T17318D,T17320D,T17328D,T17390D,T17398D,T17400D,T17408D T17410D,T17418D,T17420D,T17428D,T17490D,T17498D,T17500D T17508D,T17510D,T17518D,T17520D,T17528D,T17590D,T17598D T17800D,T17808D,T17810D,T17818D,T17820D,T17828D,T17890D T17898D,T17900D,T17908D,T17910D,T17918D,T17920D,T17928D T17990D,T17998D,T180XXD,T18100D,T18108D,T18110D,T18118D T18120D,T18128D,T18190D,T18198D,T182XXD,T183XXD,T184XXD T185XXD,T188XXD,T189XXD,T190XXD,T191XXD,T192XXD,T193XXD | V5889 |

Continued on next page

**Supplementary Table S1:** (cont'd) A comprehensive mapping of diagnosis groups used in the experiments to their corresponding ICD-10 and ICD-9 codes.

| Diagnosis | ICD-10 | ICD-9 |
|---|---|---|
| | T194XXD,T198XXD,T199XXD | |
| Effect of other external causes, initial encounter | T33011A,T33012A,T33019A,T3302XA,T3309XA,T331XXA,T332XXA<br>T333XXA,T3340XA,T3341XA,T3342XA,T33511A,T33512A,T33519A<br>T33521A,T33522A,T33529A,T33531A,T33532A,T33539A,T3360XA<br>T3361XA,T3362XA,T3370XA,T3371XA,T3372XA,T33811A,T33812A<br>T33819A,T33821A,T33822A,T33829A,T33831A,T33832A,T33839A<br>T3390XA,T3399XA,T34011A,T34012A,T34019A,T3402XA,T3409XA<br>T341XXA,T342XXA,T343XXA,T3440XA,T3441XA,T3442XA,T34511A<br>T34512A,T34519A,T34521A,T34522A,T34529A,T34531A,T34532A<br>T34539A,T3460XA,T3461XA,T3462XA,T3470XA,T3471XA,T3472XA<br>T34811A,T34812A,T34819A,T34821A,T34822A,T34829A,T34831A<br>T34832A,T34839A,T3490XA,T3499XA,T66XXXA,T6701XA,T6702XA<br>T6709XA,T670XXA,T671XXA,T672XXA,T673XXA,T674XXA,T675XXA<br>T676XXA,T677XXA,T678XXA,T679XXA,T68XXXA,T69011A,T69012A<br>T69019A,T69021A,T69022A,T69029A,T691XXA,T698XXA,T699XXA<br>T700XXA,T701XXA,T7020XA,T7029XA,T703XXA,T704XXA,T708XXA<br>T709XXA,T71111A,T71112A,T71113A,T71114A,T71121A,T71122A<br>T71123A,T71124A,T71131A,T71132A,T71133A,T71134A,T71141A<br>T71143A,T71144A,T71151A,T71152A,T71153A,T71154A,T71161A<br>T71162A,T71163A,T71164A,T71191A,T71192A,T71193A,T71194A<br>T7120XA,T7121XA,T71221A,T71222A,T71223A,T71224A,T71231A<br>T71232A,T71233A,T71234A,T7129XA,T719XXA,T730XXA,T731XXA<br>T732XXA,T733XXA,T738XXA,T739XXA,T7500XA,T7501XA,T7509XA<br>T751XXA,T7520XA,T7521XA,T7522XA,T7523XA,T7529XA,T753XXA<br>T754XXA,T7581XA,T7582XA,T7589XA | 990,9910,9911,9912,9913,9914,9915,9916,9918,9919,9920,9921<br>9922,9923,9924,9925,9926,9927,9928,9929,9930,9931,9932,9933<br>9934,9938,9939,9940,9941,9942,9943,9944,9945,9946,9947,9948<br>9949,E903,E9041,E9042,E9043,E9049,E9130,E9131,E9132,E9133<br>E9138,E9139,E9272,E9530,E9531,E9538,E963,E9830,E9831,E9838 |
| Effect of other external causes, sequela | T33011S,T33012S,T33019S,T3302XS,T3309XS,T331XXS,T332XXS<br>T333XXS,T3340XS,T3341XS,T3342XS,T33511S,T33512S,T33519S<br>T33521S,T33522S,T33529S,T33531S,T33532S,T33539S,T3360XS<br>T3361XS,T3362XS,T3370XS,T3371XS,T3372XS,T33811S,T33812S<br>T33819S,T33821S,T33822S,T33829S,T33831S,T33832S,T33839S<br>T3390XS,T3399XS,T34011S,T34012S,T34019S,T3402XS,T3409XS<br>T341XXS,T342XXS,T343XXS,T3440XS,T3441XS,T3442XS,T34511S<br>T34512S,T34519S,T34521S,T34522S,T34529S,T34531S,T34532S<br>T34539S,T3460XS,T3461XS,T3462XS,T3470XS,T3471XS,T3472XS<br>T34811S,T34812S,T34819S,T34821S,T34822S,T34829S,T34831S<br>T34832S,T34839S,T3490XS,T3499XS,T66XXXS,T6701XS,T6702XS<br>T6709XS,T670XXS,T671XXS,T672XXS,T673XXS,T674XXS,T675XXS<br>T676XXS,T677XXS,T678XXS,T679XXS,T68XXXS,T69011S,T69012S | 9094,9099,E9298,E959,E969,E989 |

Continued on next page

**Supplementary Table S1:** (cont'd) A comprehensive mapping of diagnosis groups used in the experiments to their corresponding ICD-10 and ICD-9 codes.

| Diagnosis | ICD-10 | ICD-9 |
|---|---|---|
| | T69019S,T69021S,T69022S,T69029S,T691XXS,T698XXS,T699XXS | |
| | T700XXS,T701XXS,T7020XS,T7029XS,T703XXS,T704XXS,T708XXS | |
| | T709XXS,T71111S,T71112S,T71113S,T71114S,T71121S,T71122S | |
| | T71123S,T71124S,T71131S,T71132S,T71133S,T71134S,T71141S | |
| | T71143S,T71144S,T71151S,T71152S,T71153S,T71154S,T71161S | |
| | T71162S,T71163S,T71164S,T71191S,T71192S,T71193S,T71194S | |
| | T7120XS,T7121XS,T71221S,T71222S,T71223S,T71224S,T71231S | |
| | T71232S,T71233S,T71234S,T7129XS,T719XXS,T730XXS,T731XXS | |
| | T732XXS,T733XXS,T738XXS,T739XXS,T7401XS,T7402XS,T7411XS | |
| | T7412XS,T7421XS,T7422XS,T7431XS,T7432XS,T744XXS,T7451XS | |
| | T7452XS,T7461XS,T7462XS,T7491XS,T7492XS,T7500XS,T7501XS | |
| | T7509XS,T751XXS,T7520XS,T7521XS,T7522XS,T7523XS,T7529XS | |
| | T753XXS,T754XXS,T7581XS,T7582XS,T7589XS,T7601XS,T7602XS | |
| | T7611XS,T7612XS,T7621XS,T7622XS,T7631XS,T7632XS,T7651XS | |
| | T7652XS,T7661XS,T7662XS,T7691XS,T7692XS | |
| Effect of other | T33011D,T33012D,T33019D,T3302XD,T3309XD,T331XXD,T332XXD | V5889 |
| external causes, | T333XXD,T3340XD,T3341XD,T3342XD,T33511D,T33512D,T33519D | |
| subsequent encounter | T33521D,T33522D,T33529D,T33531D,T33532D,T33539D,T3360XD | |
| | T3361XD,T3362XD,T3370XD,T3371XD,T3372XD,T33811D,T33812D | |
| | T33819D,T33821D,T33822D,T33829D,T33831D,T33832D,T33839D | |
| | T3390XD,T3399XD,T34011D,T34012D,T34019D,T3402XD,T3409XD | |
| | T341XXD,T342XXD,T343XXD,T3440XD,T3441XD,T3442XD,T34511D | |
| | T34512D,T34519D,T34521D,T34522D,T34529D,T34531D,T34532D | |
| | T34539D,T3460XD,T3461XD,T3462XD,T3470XD,T3471XD,T3472XD | |
| | T34811D,T34812D,T34819D,T34821D,T34822D,T34829D,T34831D | |
| | T34832D,T34839D,T3490XD,T3499XD,T66XXXD,T6701XD,T6702XD | |
| | T6709XD,T670XXD,T671XXD,T672XXD,T673XXD,T674XXD,T675XXD | |
| | T676XXD,T677XXD,T678XXD,T679XXD,T68XXXD,T69011D,T69012D | |
| | T69019D,T69021D,T69022D,T69029D,T691XXD,T698XXD,T699XXD | |
| | T700XXD,T701XXD,T7020XD,T7029XD,T703XXD,T704XXD,T708XXD | |
| | T709XXD,T71111D,T71112D,T71113D,T71114D,T71121D,T71122D | |
| | T71123D,T71124D,T71131D,T71132D,T71133D,T71134D,T71141D | |
| | T71143D,T71144D,T71151D,T71152D,T71153D,T71154D,T71161D | |
| | T71162D,T71163D,T71164D,T71191D,T71192D,T71193D,T71194D | |
| | T7120XD,T7121XD,T71221D,T71222D,T71223D,T71224D,T71231D | |
| | T71232D,T71233D,T71234D,T7129XD,T719XXD,T730XXD,T731XXD | |
| | T732XXD,T733XXD,T738XXD,T739XXD,T7500XD,T7501XD,T7509XD | |
| | T751XXD,T7520XD,T7521XD,T7522XD,T7523XD,T7529XD,T753XXD | |
| | T754XXD,T7581XD,T7582XD,T7589XD | |

Continued on next page

**Supplementary Table S1:** (cont'd) A comprehensive mapping of diagnosis groups used in the experiments to their corresponding ICD-10 and ICD-9 codes.

| Diagnosis | ICD-10 | ICD-9 |
|---|---|---|
| Encephalitis | A1782,A3212,A3981,A4282,A5042,A5214,A811,A830,A831,A832,A833 A834,A835,A836,A838,A839,A840,A841,A848,A8481,A8489,A849,A850 A851,A852,A858,A86,A922,A9231,B004,B0111,B020,B050,B0601 B1001,B1009,B262,B4081,B5742,B582,B6011,G0400,G0401,G0402 G042,G0430,G0431,G0432,G0439,G0481,G0482,G0489,G0490,G0491 G053,G054 | 01360,0270,0361,0398,0462,0498,0499,0520,05319,0543,0550 05601,05821,05829,0620,0621,0622,0623,0624,0625,0628,0629 0630,0631,0632,0638,0639,064,0662,06641,0722,0861,09041,09481 1160,1300,13621,3209,32341,32342,32351,32352,32361,32362 32381,32382,3239 |
| Encounter for administrative purposes | Z020,Z021,Z022,Z023,Z024,Z025,Z026,Z0271,Z0279,Z0281,Z0282 Z0283,Z0289,Z029 | V6801,V6809,V6889,V689,V703,V704,V705 |
| Encounter for antineoplastic therapies | Z510,Z5111,Z5112 | V580,V5811,V5812 |
| Encounter for mental health conditions | Z046,Z1330,Z1331,Z1332,Z1339 | V701 |
| Encounter for mental health services related to abuse | Z69010,Z69011,Z69020,Z69021,Z6911,Z6912,Z6981,Z6982 | V6111,V6112,V6121,V6122,V6283,V6549 |
| Encounter for observation and examination for conditions ruled out (excludes infectious disease, neoplasm, mental disorders) | Z036,Z0371,Z0372,Z0373,Z0374,Z0375,Z0379,Z03821,Z03822,Z03823 Z0383,Z0389,Z041,Z042,Z043,Z0441,Z0442,Z0471,Z0472,Z048,Z0481 Z0482,Z0489,Z049,Z050,Z052,Z053,Z0541,Z0542,Z0543,Z055,Z056 Z0571,Z0572,Z0573,Z058,Z059,Z130,Z131,Z1321,Z13220,Z13228 Z1329,Z135,Z136,Z1371,Z1379,Z13810,Z13811,Z13818,Z13820 Z13828,Z1383,Z1384,Z13850,Z13858,Z1388,Z1389,Z139,Z3200,Z3202 | V682,V713,V714,V715,V716,V7181,V7189,V719,V7240,V7241,V770 V771,V773,V774,V776,V777,V7791,V7799,V782,V783,V789,V790,V791 V799,V8001,V8009,V801,V802,V803,V811,V812,V813,V814,V815,V816 V820,V821,V824,V825,V826,V8271,V8279,V8281,V8289,V829,V8901 V8902,V8903,V8904,V8905,V8909 |
| Encounter for prophylactic or other procedures | Z4000,Z4001,Z4002,Z4003,Z4009,Z408,Z409,Z411,Z412,Z413,Z418 Z419,Z421,Z428 | V501,V502,V503,V5041,V5042,V5049,V508,V509,V510,V518 |
| Endocarditis and endocardial disease | A1884,A3282,A3951,A5203,A5206,A5483,B3321,B376,I011,I018,I019 I020,I099,I330,I339,I38,I39,M3211 | 01790,0270,03642,07422,09320,09321,09322,09323,09324,09381 09382,09883,09884,09885,11281,3911,3918,3919,3920,39890,4200 4210,4211,4219,4220,42490,42491,42499,4258,7100 |
| Endocrine system | C7400,C7401,C7402 | 1940 |

Continued on next page

**Supplementary Table S1:** (cont'd) A comprehensive mapping of diagnosis groups used in the experiments to their corresponding ICD-10 and ICD-9 codes.

| Diagnosis | ICD-10 | ICD-9 |
|---|---|---|
| cancers - adrenocortical | | |
| Endocrine system cancers - all other types | C7410,C7411,C7412,C7490,C7491,C7492,C752,C753,C754,C755,C758 C759 | 1940,1943,1944,1945,1946,1948,1949 |
| Endocrine system cancers - pancreas | C250,C251,C252,C253,C254,C257,C258,C259 | 1570,1571,1572,1573,1574,1578,1579 |
| Endocrine system cancers - parathyroid | C750 | 1941 |
| Endocrine system cancers - pituitary gland | C751 | 1943 |
| Endocrine system cancers - thymus | C37 | 1640 |
| Endocrine system cancers - thyroid | C73,D093,E3122,E3123 | 193,2348,25802,25803 |
| Endometriosis | N800,N8000,N8001,N8002,N8003,N801,N80101,N80102,N80103,N80109 N80111,N80112,N80113,N80119,N80121,N80122,N80123,N80129,N802 N80201,N80202,N80203,N80209,N80211,N80212,N80213,N80219 N80221,N80222,N80223,N80229,N803,N8030,N80311,N80312,N80319 N80321,N80322,N80329,N80331,N80332,N80333,N80339,N80341 N80342,N80343,N80349,N80351,N80352,N80353,N80359,N80361 N80362,N80363,N80369,N80371,N80372,N80373,N80379,N80381 N80382,N80383,N80389,N80391,N80392,N80399,N803A1,N803A2 N803A3,N803A9,N803B1,N803B2,N803B3,N803B9,N803C1,N803C2 N803C3,N803C9,N804,N8040,N8041,N8042,N805,N8050,N80511,N80512 N80519,N80521,N80522,N80529,N80531,N80532,N80539,N80541 N80542,N80549,N80551,N80552,N80559,N80561,N80562,N80569,N806 N808,N809,N80A0,N80A1,N80A2,N80A41,N80A42,N80A43,N80A49 N80A51,N80A52,N80A53,N80A59,N80A61,N80A62,N80A63,N80A69,N80B1 N80B2,N80B31,N80B32,N80B39,N80B4,N80B5,N80B6,N80C0,N80C10 N80C11,N80C19,N80C2,N80C3,N80C4,N80C9,N80D0,N80D1,N80D2,N80D3 N80D4,N80D5,N80D6,N80D9 | 6170,6171,6172,6173,6174,6175,6176,6178,6179 |

Continued on next page

**Supplementary Table S1:** (cont'd) A comprehensive mapping of diagnosis groups used in the experiments to their corresponding ICD-10 and ICD-9 codes.

| Diagnosis | ICD-10 | ICD-9 |
|---|---|---|
| Epilepsy; convulsions | G40001,G40009,G40011,G40019,G40101,G40109,G40111,G40119 G40201,G40209,G40211,G40219,G40301,G40309,G40311,G40319 G40401,G40409,G40411,G40419,G4042,G40501,G40509,G40801,G40802 G40803,G40804,G40811,G40812,G40813,G40814,G40821,G40822 G40823,G40824,G40833,G40834,G4089,G40901,G40909,G40911,G40919 G40A01,G40A09,G40A11,G40A19,G40B01,G40B09,G40B11,G40B19,R5600 R5601,R561,R569 | 34500,34501,34510,34511,3452,3453,34540,34541,34550,34551 34560,34561,34570,34571,34580,34581,34590,34591,78031,78032 78033,78039 |
| Erectile dysfunction | N5201,N5202,N5203,N521,N522,N5231,N5232,N5233,N5234,N5235 N5236,N5237,N5239,N528,N529 | 60784 |
| Esophageal disorders | B3781,B5731,K200,K208,K2080,K2081,K209,K2090,K2091,K210,K2100 K2101,K219,K220,K2210,K2211,K222,K223,K224,K225,K226,K2270 K22710,K22711,K22719,K228,K2281,K2282,K2289,K229,K23 | 0861,11284,5300,53010,53011,53012,53013,53019,53020,53021 5303,5304,5305,5306,5307,53081,53082,53083,53085,53089,5309 |
| Essential hypertension | I10,O10011,O10012,O10013,O10019,O1002,O1003,O10911,O10912 O10913,O10919,O1092,O1093,O161,O162,O163,O164,O165,O169 | 4010,4011,4019,64200,64201,64203,64204,64290,64291,64293 64294 |
| Exposure, encounters, screening or contact with infectious disease | G0402,G0432,Z03810,Z03818,Z051,Z110,Z111,Z112,Z113,Z114,Z1151 Z1152,Z1159,Z116,Z117,Z118,Z119,Z2001,Z2009,Z201,Z202,Z203 Z204,Z205,Z206,Z207,Z20810,Z20811,Z20818,Z20820,Z20821,Z20822 Z20828,Z2089,Z209,Z23 | 32351,32352,V010,V011,V012,V013,V014,V015,V016,V0171,V0179 V0181,V0182,V0183,V0184,V0189,V019,V059,V7182,V7183,V730,V732 V733,V734,V735,V7381,V7388,V7389,V7398,V740,V741,V742,V743 V745,V748,V750,V751,V752,V753,V754,V755,V756,V757,V758,V759 |
| Feeding and eating disorders | F5000,F5001,F5002,F502,F508,F5081,F5082,F5089,F509,F9821 F9829,F983 | 3071,30750,30751,30752,30753,30754,30759 |
| Female infertility | E28310,E28319,E2839,N970,N971,N972,N978,N979 | 25631,25639,6280,6282,6283,6284,6288,6289 |
| Female reproductive system cancers - all other types | C510,C511,C512,C518,C5710,C5711,C5712,C5720,C5721,C5722,C573 C574,C577,C578,C579,C58,D0730,D0739 | 181,1833,1834,1835,1839,1841,1842,1843,1848,1849,23330,23339 |
| Female reproductive system cancers - cervix | C530,C531,C538,C539,D060,D061,D067,D069 | 1800,1801,1808,1809,2331 |
| Female reproductive system cancers - endometrium | C541,D070 | 1820,2332 |

Continued on next page

**Supplementary Table S1:** (cont'd) A comprehensive mapping of diagnosis groups used in the experiments to their corresponding ICD-10 and ICD-9 codes.

| Diagnosis | ICD-10 | ICD-9 |
|---|---|---|
| Female reproductive system cancers - fallopian tube | C5700,C5701,C5702 | 1832 |
| Female reproductive system cancers - ovary | C561,C562,C563,C569 | 1830 |
| Female reproductive system cancers - uterus | C540,C542,C543,C548,C549,C55 | 179,1820,1821,1828 |
| Female reproductive system cancers - vagina | C52,D072 | 1840,23331 |
| Female reproductive system cancers - vulva | C519,D071 | 1844,23332 |
| Fetal alcohol syndrome | P043,Q860 | 76071 |
| Fever | R502,R5081,R5082,R5083,R5084,R509 | 78060,78061,78062,78063,78066,E9479 |
| Fluid and electrolyte disorders | E860,E861,E869,E870,E871,E872,E8720,E8721,E8722,E8729,E873 E874,E875,E876,E8770,E8771,E8779,E878 | 2760,2761,2762,2763,2764,27650,27651,27652,27661,27669,2767 2768,2769 |
| Foodborne intoxications | A050,A051,A052,A053,A054,A055,A058,A059,A4851 | 0050,0051,0052,0054,00581,00589,0059,04041 |
| Fracture of head and neck, initial encounter | S020XXA,S020XXB,S02101A,S02101B,S02102A,S02102B,S02109A S02109B,S0210XA,S0210XB,S02110A,S02110B,S02111A,S02111B S02112A,S02112B,S02113A,S02113B,S02118A,S02118B,S02119A S02119B,S0211AA,S0211AB,S0211BA,S0211BB,S0211CA,S0211CB S0211DA,S0211DB,S0211EA,S0211EB,S0211FA,S0211FB,S0211GA S0211GB,S0211HA,S0211HB,S02121A,S02121B,S02122A,S02122B S02129A,S02129B,S0219XA,S0219XB,S022XXA,S022XXB,S0230XA S0230XB,S0231XA,S0231XB,S0232XA,S0232XB,S023XXA,S023XXB | 80000,80050,80100,80150,8020,8021,80220,80221,80222,80223 80224,80225,80226,80227,80228,80230,80231,80232,80233,80234 80235,80236,80237,80238,8024,8025,8026,8027,8028,8029,80300 80350,87363 |

Continued on next page

**Supplementary Table S1:** (cont'd) A comprehensive mapping of diagnosis groups used in the experiments to their corresponding ICD-10 and ICD-9 codes.

| Diagnosis | ICD-10 | ICD-9 |
|---|---|---|
| | S02400A,S02400B,S02401A,S02401B,S02402A,S02402B,S0240AA | |
| | S0240AB,S0240BA,S0240BB,S0240CA,S0240CB,S0240DA,S0240DB | |
| | S0240EA,S0240EB,S0240FA,S0240FB,S02411A,S02411B,S02412A | |
| | S02412B,S02413A,S02413B,S0242XA,S0242XB,S025XXA,S025XXB | |
| | S02600A,S02600B,S02601A,S02601B,S02602A,S02602B,S02609A | |
| | S02609B,S02610A,S02610B,S02611A,S02611B,S02612A,S02612B | |
| | S0261XA,S0261XB,S02620A,S02620B,S02621A,S02621B,S02622A | |
| | S02622B,S0262XA,S0262XB,S02630A,S02630B,S02631A,S02631B | |
| | S02632A,S02632B,S0263XA,S0263XB,S02640A,S02640B,S02641A | |
| | S02641B,S02642A,S02642B,S0264XA,S0264XB,S02650A,S02650B | |
| | S02651A,S02651B,S02652A,S02652B,S0265XA,S0265XB,S0266XA | |
| | S0266XB,S02670A,S02670B,S02671A,S02671B,S02672A,S02672B | |
| | S0267XA,S0267XB,S0269XA,S0269XB,S0280XA,S0280XB,S0281XA | |
| | S0281XB,S0282XA,S0282XB,S02831A,S02831B,S02832A,S02832B | |
| | S02839A,S02839B,S02841A,S02841B,S02842A,S02842B,S02849A | |
| | S02849B,S0285XA,S0285XB,S028XXA,S028XXB,S0291XA,S0291XB | |
| | S0292XA,S0292XB | |
| Fracture of head and | S020XXD,S020XXG,S020XXK,S02101D,S02101G,S02101K,S02102D | 73382,V5419 |
| neck, subsequent | S02102G,S02102K,S02109D,S02109G,S02109K,S0210XD,S0210XG | |
| encounter | S0210XK,S02110D,S02110G,S02110K,S02111D,S02111G,S02111K | |
| | S02112D,S02112G,S02112K,S02113D,S02113G,S02113K,S02118D | |
| | S02118G,S02118K,S02119D,S02119G,S02119K,S0211AD,S0211AG | |
| | S0211AK,S0211BD,S0211BG,S0211BK,S0211CD,S0211CG,S0211CK | |
| | S0211DD,S0211DG,S0211DK,S0211ED,S0211EG,S0211EK,S0211FD | |
| | S0211FG,S0211FK,S0211GD,S0211GG,S0211GK,S0211HD,S0211HG | |
| | S0211HK,S02121D,S02121G,S02121K,S02122D,S02122G,S02122K | |
| | S02129D,S02129G,S02129K,S0219XD,S0219XG,S0219XK,S022XXD | |
| | S022XXG,S022XXK,S0230XD,S0230XG,S0230XK,S0231XD,S0231XG | |
| | S0231XK,S0232XD,S0232XG,S0232XK,S023XXD,S023XXG,S023XXK | |
| | S02400D,S02400G,S02400K,S02401D,S02401G,S02401K,S02402D | |
| | S02402G,S02402K,S0240AD,S0240AG,S0240AK,S0240BD,S0240BG | |
| | S0240BK,S0240CD,S0240CG,S0240CK,S0240DD,S0240DG,S0240DK | |
| | S0240ED,S0240EG,S0240EK,S0240FD,S0240FG,S0240FK,S02411D | |
| | S02411G,S02411K,S02412D,S02412G,S02412K,S02413D,S02413G | |
| | S02413K,S0242XD,S0242XG,S0242XK,S025XXD,S025XXG,S025XXK | |
| | S02600D,S02600G,S02600K,S02601D,S02601G,S02601K,S02602D | |
| | S02602G,S02602K,S02609D,S02609G,S02609K,S02610D,S02610G | |
| | S02610K,S02611D,S02611G,S02611K,S02612D,S02612G,S02612K | |
| | S0261XD,S0261XG,S0261XK,S02620D,S02620G,S02620K,S02621D | |

Continued on next page

**Supplementary Table S1:** (cont'd) A comprehensive mapping of diagnosis groups used in the experiments to their corresponding ICD-10 and ICD-9 codes.

| Diagnosis | ICD-10 | ICD-9 |
|---|---|---|
| | S02621G,S02621K,S02622D,S02622G,S02622K,S0262XD,S0262XG<br>S0262XK,S02630D,S02630G,S02630K,S02631D,S02631G,S02631K<br>S02632D,S02632G,S02632K,S0263XD,S0263XG,S0263XK,S02640D<br>S02640G,S02640K,S02641D,S02641G,S02641K,S02642D,S02642G<br>S02642K,S0264XD,S0264XG,S0264XK,S02650D,S02650G,S02650K<br>S02651D,S02651G,S02651K,S02652D,S02652G,S02652K,S0265XD<br>S0265XG,S0265XK,S0266XD,S0266XG,S0266XK,S02670D,S02670G<br>S02670K,S02671D,S02671G,S02671K,S02672D,S02672G,S02672K<br>S0267XD,S0267XG,S0267XK,S0269XD,S0269XG,S0269XK,S0280XD<br>S0280XG,S0280XK,S0281XD,S0281XG,S0281XK,S0282XD,S0282XG<br>S0282XK,S02831D,S02831G,S02831K,S02832D,S02832G,S02832K<br>S02839D,S02839G,S02839K,S02841D,S02841G,S02841K,S02842D<br>S02842G,S02842K,S02849D,S02849G,S02849K,S0285XD,S0285XG<br>S0285XK,S028XXD,S028XXG,S028XXK,S0291XD,S0291XG,S0291XK<br>S0292XD,S0292XG,S0292XK | |
| Fracture of lower limb (except hip), subsequent encounter | M9711XD,M9712XD,M9721XD,M9722XD,M978XXD,M979XXD,S72301D<br>S72301E,S72301F,S72301G,S72301H,S72301J,S72301K,S72301M<br>S72301N,S72301P,S72301Q,S72301R,S72302D,S72302E,S72302F<br>S72302G,S72302H,S72302J,S72302K,S72302M,S72302N,S72302P<br>S72302Q,S72302R,S72309D,S72309E,S72309F,S72309G,S72309H<br>S72309J,S72309K,S72309M,S72309N,S72309P,S72309Q,S72309R<br>S72321D,S72321E,S72321F,S72321G,S72321H,S72321J,S72321K<br>S72321M,S72321N,S72321P,S72321Q,S72321R,S72322D,S72322E<br>S72322F,S72322G,S72322H,S72322J,S72322K,S72322M,S72322N<br>S72322P,S72322Q,S72322R,S72323D,S72323E,S72323F,S72323G<br>S72323H,S72323J,S72323K,S72323M,S72323N,S72323P,S72323Q<br>S72323R,S72324D,S72324E,S72324F,S72324G,S72324H,S72324J<br>S72324K,S72324M,S72324N,S72324P,S72324Q,S72324R,S72325D<br>S72325E,S72325F,S72325G,S72325H,S72325J,S72325K,S72325M<br>S72325N,S72325P,S72325Q,S72325R,S72326D,S72326E,S72326F<br>S72326G,S72326H,S72326J,S72326K,S72326M,S72326N,S72326P<br>S72326Q,S72326R,S72331D,S72331E,S72331F,S72331G,S72331H<br>S72331J,S72331K,S72331M,S72331N,S72331P,S72331Q,S72331R<br>S72332D,S72332E,S72332F,S72332G,S72332H,S72332J,S72332K<br>S72332M,S72332N,S72332P,S72332Q,S72332R,S72333D,S72333E<br>S72333F,S72333G,S72333H,S72333J,S72333K,S72333M,S72333N<br>S72333P,S72333Q,S72333R,S72334D,S72334E,S72334F,S72334G<br>S72334H,S72334J,S72334K,S72334M,S72334N,S72334P,S72334Q<br>S72334R,S72335D,S72335E,S72335F,S72335G,S72335H,S72335J | 73381,73382,V5415,V5416,V5426,V5429 |

Continued on next page

**Supplementary Table S1:** (cont'd) A comprehensive mapping of diagnosis groups used in the experiments to their corresponding ICD-10 and ICD-9 codes.

| Diagnosis | ICD-10 | ICD-9 |
|---|---|---|
| | S72335K,S72335M,S72335N,S72335P,S72335Q,S72335R,S72336D | |
| | S72336E,S72336F,S72336G,S72336H,S72336J,S72336K,S72336M | |
| | S72336N,S72336P,S72336Q,S72336R,S72341D,S72341E,S72341F | |
| | S72341G,S72341H,S72341J,S72341K,S72341M,S72341N,S72341P | |
| | S72341Q,S72341R,S72342D,S72342E,S72342F,S72342G,S72342H | |
| | S72342J,S72342K,S72342M,S72342N,S72342P,S72342Q,S72342R | |
| | S72343D,S72343E,S72343F,S72343G,S72343H,S72343J,S72343K | |
| | S72343M,S72343N,S72343P,S72343Q,S72343R,S72344D,S72344E | |
| | S72344F,S72344G,S72344H,S72344J,S72344K,S72344M,S72344N | |
| | S72344P,S72344Q,S72344R,S72345D,S72345E,S72345F,S72345G | |
| | S72345H,S72345J,S72345K,S72345M,S72345N,S72345P,S72345Q | |
| | S72345R,S72346D,S72346E,S72346F,S72346G,S72346H,S72346J | |
| | S72346K,S72346M,S72346N,S72346P,S72346Q,S72346R,S72351D | |
| | S72351E,S72351F,S72351G,S72351H,S72351J,S72351K,S72351M | |
| | S72351N,S72351P,S72351Q,S72351R,S72352D,S72352E,S72352F | |
| | S72352G,S72352H,S72352J,S72352K,S72352M,S72352N,S72352P | |
| | S72352Q,S72352R,S72353D,S72353E,S72353F,S72353G,S72353H | |
| | S72353J,S72353K,S72353M,S72353N,S72353P,S72353Q,S72353R | |
| | S72354D,S72354E,S72354F,S72354G,S72354H,S72354J,S72354K | |
| | S72354M,S72354N,S72354P,S72354Q,S72354R,S72355D,S72355E | |
| | S72355F,S72355G,S72355H,S72355J,S72355K,S72355M,S72355N | |
| | S72355P,S72355Q,S72355R,S72356D,S72356E,S72356F,S72356G | |
| | S72356H,S72356J,S72356K,S72356M,S72356N,S72356P,S72356Q | |
| | S72356R,S72361D,S72361E,S72361F,S72361G,S72361H,S72361J | |
| | S72361K,S72361M,S72361N,S72361P,S72361Q,S72361R,S72362D | |
| | S72362E,S72362F,S72362G,S72362H,S72362J,S72362K,S72362M | |
| | S72362N,S72362P,S72362Q,S72362R,S72363D,S72363E,S72363F | |
| | S72363G,S72363H,S72363J,S72363K,S72363M,S72363N,S72363P | |
| | S72363Q,S72363R,S72364D,S72364E,S72364F,S72364G,S72364H | |
| | S72364J,S72364K,S72364M,S72364N,S72364P,S72364Q,S72364R | |
| | S72365D,S72365E,S72365F,S72365G,S72365H,S72365J,S72365K | |
| | S72365M,S72365N,S72365P,S72365Q,S72365R,S72366D,S72366E | |
| | S72366F,S72366G,S72366H,S72366J,S72366K,S72366M,S72366N | |
| | S72366P,S72366Q,S72366R,S72391D,S72391E,S72391F,S72391G | |
| | S72391H,S72391J,S72391K,S72391M,S72391N,S72391P,S72391Q | |
| | S72391R,S72392D,S72392E,S72392F,S72392G,S72392H,S72392J | |
| | S72392K,S72392M,S72392N,S72392P,S72392Q,S72392R,S72399D | |
| | S72399E,S72399F,S72399G,S72399H,S72399J,S72399K,S72399M | |
| | S72399N,S72399P,S72399Q,S72399R,S72401D,S72401E,S72401F | |
| | S72401G,S72401H,S72401J,S72401K,S72401M,S72401N,S72401P | |

Continued on next page

**Supplementary Table S1:** (cont'd) A comprehensive mapping of diagnosis groups used in the experiments to their corresponding ICD-10 and ICD-9 codes.

| Diagnosis | ICD-10 | ICD-9 |
|---|---|---|
| | S72401Q,S72401R,S72402D,S72402E,S72402F,S72402G,S72402H | |
| | S72402J,S72402K,S72402M,S72402N,S72402P,S72402Q,S72402R | |
| | S72409D,S72409E,S72409F,S72409G,S72409H,S72409J,S72409K | |
| | S72409M,S72409N,S72409P,S72409Q,S72409R,S72411D,S72411E | |
| | S72411F,S72411G,S72411H,S72411J,S72411K,S72411M,S72411N | |
| | S72411P,S72411Q,S72411R,S72412D,S72412E,S72412F,S72412G | |
| | S72412H,S72412J,S72412K,S72412M,S72412N,S72412P,S72412Q | |
| | S72412R,S72413D,S72413E,S72413F,S72413G,S72413H,S72413J | |
| | S72413K,S72413M,S72413N,S72413P,S72413Q,S72413R,S72414D | |
| | S72414E,S72414F,S72414G,S72414H,S72414J,S72414K,S72414M | |
| | S72414N,S72414P,S72414Q,S72414R,S72415D,S72415E,S72415F | |
| | S72415G,S72415H,S72415J,S72415K,S72415M,S72415N,S72415P | |
| | S72415Q,S72415R,S72416D,S72416E,S72416F,S72416G,S72416H | |
| | S72416J,S72416K,S72416M,S72416N,S72416P,S72416Q,S72416R | |
| | S72421D,S72421E,S72421F,S72421G,S72421H,S72421J,S72421K | |
| | S72421M,S72421N,S72421P,S72421Q,S72421R,S72422D,S72422E | |
| | S72422F,S72422G,S72422H,S72422J,S72422K,S72422M,S72422N | |
| | S72422P,S72422Q,S72422R,S72423D,S72423E,S72423F,S72423G | |
| | S72423H,S72423J,S72423K,S72423M,S72423N,S72423P,S72423Q | |
| | S72423R,S72424D,S72424E,S72424F,S72424G,S72424H,S72424J | |
| | S72424K,S72424M,S72424N,S72424P,S72424Q,S72424R,S72425D | |
| | S72425E,S72425F,S72425G,S72425H,S72425J,S72425K,S72425M | |
| | S72425N,S72425P,S72425Q,S72425R,S72426D,S72426E,S72426F | |
| | S72426G,S72426H,S72426J,S72426K,S72426M,S72426N,S72426P | |
| | S72426Q,S72426R,S72431D,S72431E,S72431F,S72431G,S72431H | |
| | S72431J,S72431K,S72431M,S72431N,S72431P,S72431Q,S72431R | |
| | S72432D,S72432E,S72432F,S72432G,S72432H,S72432J,S72432K | |
| | S72432M,S72432N,S72432P,S72432Q,S72432R,S72433D,S72433E | |
| | S72433F,S72433G,S72433H,S72433J,S72433K,S72433M,S72433N | |
| | S72433P,S72433Q,S72433R,S72434D,S72434E,S72434F,S72434G | |
| | S72434H,S72434J,S72434K,S72434M,S72434N,S72434P,S72434Q | |
| | S72434R,S72435D,S72435E,S72435F,S72435G,S72435H,S72435J | |
| | S72435K,S72435M,S72435N,S72435P,S72435Q,S72435R,S72436D | |
| | S72436E,S72436F,S72436G,S72436H,S72436J,S72436K,S72436M | |
| | S72436N,S72436P,S72436Q,S72436R,S72441D,S72441E,S72441F | |
| | S72441G,S72441H,S72441J,S72441K,S72441M,S72441N,S72441P | |
| | S72441Q,S72441R,S72442D,S72442E,S72442F,S72442G,S72442H | |
| | S72442J,S72442K,S72442M,S72442N,S72442P,S72442Q,S72442R | |
| | S72443D,S72443E,S72443F,S72443G,S72443H,S72443J,S72443K | |
| | S72443M,S72443N,S72443P,S72443Q,S72443R,S72444D,S72444E | |

Continued on next page

**Supplementary Table S1:** (cont'd) A comprehensive mapping of diagnosis groups used in the experiments to their corresponding ICD-10 and ICD-9 codes.

| Diagnosis | ICD-10 | ICD-9 |
|---|---|---|
| | S72444F,S72444G,S72444H,S72444J,S72444K,S72444M,S72444N | |
| | S72444P,S72444Q,S72444R,S72445D,S72445E,S72445F,S72445G | |
| | S72445H,S72445J,S72445K,S72445M,S72445N,S72445P,S72445Q | |
| | S72445R,S72446D,S72446E,S72446F,S72446G,S72446H,S72446J | |
| | S72446K,S72446M,S72446N,S72446P,S72446Q,S72446R,S72451D | |
| | S72451E,S72451F,S72451G,S72451H,S72451J,S72451K,S72451M | |
| | S72451N,S72451P,S72451Q,S72451R,S72452D,S72452E,S72452F | |
| | S72452G,S72452H,S72452J,S72452K,S72452M,S72452N,S72452P | |
| | S72452Q,S72452R,S72453D,S72453E,S72453F,S72453G,S72453H | |
| | S72453J,S72453K,S72453M,S72453N,S72453P,S72453Q,S72453R | |
| | S72454D,S72454E,S72454F,S72454G,S72454H,S72454J,S72454K | |
| | S72454M,S72454N,S72454P,S72454Q,S72454R,S72455D,S72455E | |
| | S72455F,S72455G,S72455H,S72455J,S72455K,S72455M,S72455N | |
| | S72455P,S72455Q,S72455R,S72456D,S72456E,S72456F,S72456G | |
| | S72456H,S72456J,S72456K,S72456M,S72456N,S72456P,S72456Q | |
| | S72456R,S72461D,S72461E,S72461F,S72461G,S72461H,S72461J | |
| | S72461K,S72461M,S72461N,S72461P,S72461Q,S72461R,S72462D | |
| | S72462E,S72462F,S72462G,S72462H,S72462J,S72462K,S72462M | |
| | S72462N,S72462P,S72462Q,S72462R,S72463D,S72463E,S72463F | |
| | S72463G,S72463H,S72463J,S72463K,S72463M,S72463N,S72463P | |
| | S72463Q,S72463R,S72464D,S72464E,S72464F,S72464G,S72464H | |
| | S72464J,S72464K,S72464M,S72464N,S72464P,S72464Q,S72464R | |
| | S72465D,S72465E,S72465F,S72465G,S72465H,S72465J,S72465K | |
| | S72465M,S72465N,S72465P,S72465Q,S72465R,S72466D,S72466E | |
| | S72466F,S72466G,S72466H,S72466J,S72466K,S72466M,S72466N | |
| | S72466P,S72466Q,S72466R,S72471D,S72471G,S72471K,S72471P | |
| | S72472D,S72472G,S72472K,S72472P,S72479D,S72479G,S72479K | |
| | S72479P,S72491D,S72491E,S72491F,S72491G,S72491H,S72491J | |
| | S72491K,S72491M,S72491N,S72491P,S72491Q,S72491R,S72492D | |
| | S72492E,S72492F,S72492G,S72492H,S72492J,S72492K,S72492M | |
| | S72492N,S72492P,S72492Q,S72492R,S72499D,S72499E,S72499F | |
| | S72499G,S72499H,S72499J,S72499K,S72499M,S72499N,S72499P | |
| | S72499Q,S72499R,S728X1D,S728X1E,S728X1F,S728X1G,S728X1H | |
| | S728X1J,S728X1K,S728X1M,S728X1N,S728X1P,S728X1Q,S728X1R | |
| | S728X2D,S728X2E,S728X2F,S728X2G,S728X2H,S728X2J,S728X2K | |
| | S728X2M,S728X2N,S728X2P,S728X2Q,S728X2R,S728X9D,S728X9E | |
| | S728X9F,S728X9G,S728X9H,S728X9J,S728X9K,S728X9M,S728X9N | |
| | S728X9P,S728X9Q,S728X9R,S7290XD,S7290XE,S7290XF,S7290XG | |
| | S7290XH,S7290XJ,S7290XK,S7290XM,S7290XN,S7290XP,S7290XQ | |
| | S7290XR,S7291XD,S7291XE,S7291XF,S7291XG,S7291XH,S7291XJ | |

Continued on next page

**Supplementary Table S1:** (cont'd) A comprehensive mapping of diagnosis groups used in the experiments to their corresponding ICD-10 and ICD-9 codes.

| Diagnosis | ICD-10 | ICD-9 |
|---|---|---|
| | S7291XK,S7291XM,S7291XN,S7291XP,S7291XQ,S7291XR,S7292XD<br>S7292XE,S7292XF,S7292XG,S7292XH,S7292XJ,S7292XK,S7292XM<br>S7292XN,S7292XP,S7292XQ,S7292XR,S79001D,S79001G,S79001K<br>S79001P,S79002D,S79002G,S79002K,S79002P,S79009D,S79009G<br>S79009K,S79009P,S79011D,S79011G,S79011K,S79011P,S79012D<br>S79012G,S79012K,S79012P,S79019D,S79019G,S79019K,S79019P<br>S79091D,S79091G,S79091K,S79091P,S79092D,S79092G,S79092K<br>S79092P,S79099D,S79099G,S79099K,S79099P,S79101D,S79101G<br>S79101K,S79101P,S79102D,S79102G,S79102K,S79102P,S79109D<br>S79109G,S79109K,S79109P,S79111D,S79111G,S79111K,S79111P<br>S79112D,S79112G,S79112K,S79112P,S79119D,S79119G,S79119K<br>S79119P,S79121D,S79121G,S79121K,S79121P,S79122D,S79122G<br>S79122K,S79122P,S79129D,S79129G,S79129K,S79129P,S79131D<br>S79131G,S79131K,S79131P,S79132D,S79132G,S79132K,S79132P<br>S79139D,S79139G,S79139K,S79139P,S79141D,S79141G,S79141K<br>S79141P,S79142D,S79142G,S79142K,S79142P,S79149D,S79149G<br>S79149K,S79149P,S79191D,S79191G,S79191K,S79191P,S79192D<br>S79192G,S79192K,S79192P,S79199D,S79199G,S79199K,S79199P<br>S82001D,S82001E,S82001F,S82001G,S82001H,S82001J,S82001K<br>S82001M,S82001N,S82001P,S82001Q,S82001R,S82002D,S82002E<br>S82002F,S82002G,S82002H,S82002J,S82002K,S82002M,S82002N<br>S82002P,S82002Q,S82002R,S82009D,S82009E,S82009F,S82009G<br>S82009H,S82009J,S82009K,S82009M,S82009N,S82009P,S82009Q<br>S82009R,S82011D,S82011E,S82011F,S82011G,S82011H,S82011J<br>S82011K,S82011M,S82011N,S82011P,S82011Q,S82011R,S82012D<br>S82012E,S82012F,S82012G,S82012H,S82012J,S82012K,S82012M<br>S82012N,S82012P,S82012Q,S82012R,S82013D,S82013E,S82013F<br>S82013G,S82013H,S82013J,S82013K,S82013M,S82013N,S82013P<br>S82013Q,S82013R,S82014D,S82014E,S82014F,S82014G,S82014H<br>S82014J,S82014K,S82014M,S82014N,S82014P,S82014Q,S82014R<br>S82015D,S82015E,S82015F,S82015G,S82015H,S82015J,S82015K<br>S82015M,S82015N,S82015P,S82015Q,S82015R,S82016D,S82016E<br>S82016F,S82016G,S82016H,S82016J,S82016K,S82016M,S82016N<br>S82016P,S82016Q,S82016R,S82021D,S82021E,S82021F,S82021G<br>S82021H,S82021J,S82021K,S82021M,S82021N,S82021P,S82021Q<br>S82021R,S82022D,S82022E,S82022F,S82022G,S82022H,S82022J<br>S82022K,S82022M,S82022N,S82022P,S82022Q,S82022R,S82023D<br>S82023E,S82023F,S82023G,S82023H,S82023J,S82023K,S82023M<br>S82023N,S82023P,S82023Q,S82023R,S82024D,S82024E,S82024F<br>S82024G,S82024H,S82024J,S82024K,S82024M,S82024N,S82024P | |

Continued on next page

**Supplementary Table S1:** (cont'd) A comprehensive mapping of diagnosis groups used in the experiments to their corresponding ICD-10 and ICD-9 codes.

| Diagnosis | ICD-10 | ICD-9 |
|---|---|---|
| | S82024Q,S82024R,S82025D,S82025E,S82025F,S82025G,S82025H | |
| | S82025J,S82025K,S82025M,S82025N,S82025P,S82025Q,S82025R | |
| | S82026D,S82026E,S82026F,S82026G,S82026H,S82026J,S82026K | |
| | S82026M,S82026N,S82026P,S82026Q,S82026R,S82031D,S82031E | |
| | S82031F,S82031G,S82031H,S82031J,S82031K,S82031M,S82031N | |
| | S82031P,S82031Q,S82031R,S82032D,S82032E,S82032F,S82032G | |
| | S82032H,S82032J,S82032K,S82032M,S82032N,S82032P,S82032Q | |
| | S82032R,S82033D,S82033E,S82033F,S82033G,S82033H,S82033J | |
| | S82033K,S82033M,S82033N,S82033P,S82033Q,S82033R,S82034D | |
| | S82034E,S82034F,S82034G,S82034H,S82034J,S82034K,S82034M | |
| | S82034N,S82034P,S82034Q,S82034R,S82035D,S82035E,S82035F | |
| | S82035G,S82035H,S82035J,S82035K,S82035M,S82035N,S82035P | |
| | S82035Q,S82035R,S82036D,S82036E,S82036F,S82036G,S82036H | |
| | S82036J,S82036K,S82036M,S82036N,S82036P,S82036Q,S82036R | |
| | S82041D,S82041E,S82041F,S82041G,S82041H,S82041J,S82041K | |
| | S82041M,S82041N,S82041P,S82041Q,S82041R,S82042D,S82042E | |
| | S82042F,S82042G,S82042H,S82042J,S82042K,S82042M,S82042N | |
| | S82042P,S82042Q,S82042R,S82043D,S82043E,S82043F,S82043G | |
| | S82043H,S82043J,S82043K,S82043M,S82043N,S82043P,S82043Q | |
| | S82043R,S82044D,S82044E,S82044F,S82044G,S82044H,S82044J | |
| | S82044K,S82044M,S82044N,S82044P,S82044Q,S82044R,S82045D | |
| | S82045E,S82045F,S82045G,S82045H,S82045J,S82045K,S82045M | |
| | S82045N,S82045P,S82045Q,S82045R,S82046D,S82046E,S82046F | |
| | S82046G,S82046H,S82046J,S82046K,S82046M,S82046N,S82046P | |
| | S82046Q,S82046R,S82091D,S82091E,S82091F,S82091G,S82091H | |
| | S82091J,S82091K,S82091M,S82091N,S82091P,S82091Q,S82091R | |
| | S82092D,S82092E,S82092F,S82092G,S82092H,S82092J,S82092K | |
| | S82092M,S82092N,S82092P,S82092Q,S82092R,S82099D,S82099E | |
| | S82099F,S82099G,S82099H,S82099J,S82099K,S82099M,S82099N | |
| | S82099P,S82099Q,S82099R,S82101D,S82101E,S82101F,S82101G | |
| | S82101H,S82101J,S82101K,S82101M,S82101N,S82101P,S82101Q | |
| | S82101R,S82102D,S82102E,S82102F,S82102G,S82102H,S82102J | |
| | S82102K,S82102M,S82102N,S82102P,S82102Q,S82102R,S82109D | |
| | S82109E,S82109F,S82109G,S82109H,S82109J,S82109K,S82109M | |
| | S82109N,S82109P,S82109Q,S82109R,S82111D,S82111E,S82111F | |
| | S82111G,S82111H,S82111J,S82111K,S82111M,S82111N,S82111P | |
| | S82111Q,S82111R,S82112D,S82112E,S82112F,S82112G,S82112H | |
| | S82112J,S82112K,S82112M,S82112N,S82112P,S82112Q,S82112R | |
| | S82113D,S82113E,S82113F,S82113G,S82113H,S82113J,S82113K | |
| | S82113M,S82113N,S82113P,S82113Q,S82113R,S82114D,S82114E | |

Continued on next page

**Supplementary Table S1:** (cont'd) A comprehensive mapping of diagnosis groups used in the experiments to their corresponding ICD-10 and ICD-9 codes.

| Diagnosis | ICD-10 | ICD-9 |
|---|---|---|
| | S82114F,S82114G,S82114H,S82114J,S82114K,S82114M,S82114N | |
| | S82114P,S82114Q,S82114R,S82115D,S82115E,S82115F,S82115G | |
| | S82115H,S82115J,S82115K,S82115M,S82115N,S82115P,S82115Q | |
| | S82115R,S82116D,S82116E,S82116F,S82116G,S82116H,S82116J | |
| | S82116K,S82116M,S82116N,S82116P,S82116Q,S82116R,S82121D | |
| | S82121E,S82121F,S82121G,S82121H,S82121J,S82121K,S82121M | |
| | S82121N,S82121P,S82121Q,S82121R,S82122D,S82122E,S82122F | |
| | S82122G,S82122H,S82122J,S82122K,S82122M,S82122N,S82122P | |
| | S82122Q,S82122R,S82123D,S82123E,S82123F,S82123G,S82123H | |
| | S82123J,S82123K,S82123M,S82123N,S82123P,S82123Q,S82123R | |
| | S82124D,S82124E,S82124F,S82124G,S82124H,S82124J,S82124K | |
| | S82124M,S82124N,S82124P,S82124Q,S82124R,S82125D,S82125E | |
| | S82125F,S82125G,S82125H,S82125J,S82125K,S82125M,S82125N | |
| | S82125P,S82125Q,S82125R,S82126D,S82126E,S82126F,S82126G | |
| | S82126H,S82126J,S82126K,S82126M,S82126N,S82126P,S82126Q | |
| | S82126R,S82131D,S82131E,S82131F,S82131G,S82131H,S82131J | |
| | S82131K,S82131M,S82131N,S82131P,S82131Q,S82131R,S82132D | |
| | S82132E,S82132F,S82132G,S82132H,S82132J,S82132K,S82132M | |
| | S82132N,S82132P,S82132Q,S82132R,S82133D,S82133E,S82133F | |
| | S82133G,S82133H,S82133J,S82133K,S82133M,S82133N,S82133P | |
| | S82133Q,S82133R,S82134D,S82134E,S82134F,S82134G,S82134H | |
| | S82134J,S82134K,S82134M,S82134N,S82134P,S82134Q,S82134R | |
| | S82135D,S82135E,S82135F,S82135G,S82135H,S82135J,S82135K | |
| | S82135M,S82135N,S82135P,S82135Q,S82135R,S82136D,S82136E | |
| | S82136F,S82136G,S82136H,S82136J,S82136K,S82136M,S82136N | |
| | S82136P,S82136Q,S82136R,S82141D,S82141E,S82141F,S82141G | |
| | S82141H,S82141J,S82141K,S82141M,S82141N,S82141P,S82141Q | |
| | S82141R,S82142D,S82142E,S82142F,S82142G,S82142H,S82142J | |
| | S82142K,S82142M,S82142N,S82142P,S82142Q,S82142R,S82143D | |
| | S82143E,S82143F,S82143G,S82143H,S82143J,S82143K,S82143M | |
| | S82143N,S82143P,S82143Q,S82143R,S82144D,S82144E,S82144F | |
| | S82144G,S82144H,S82144J,S82144K,S82144M,S82144N,S82144P | |
| | S82144Q,S82144R,S82145D,S82145E,S82145F,S82145G,S82145H | |
| | S82145J,S82145K,S82145M,S82145N,S82145P,S82145Q,S82145R | |
| | S82146D,S82146E,S82146F,S82146G,S82146H,S82146J,S82146K | |
| | S82146M,S82146N,S82146P,S82146Q,S82146R,S82151D,S82151E | |
| | S82151F,S82151G,S82151H,S82151J,S82151K,S82151M,S82151N | |
| | S82151P,S82151Q,S82151R,S82152D,S82152E,S82152F,S82152G | |
| | S82152H,S82152J,S82152K,S82152M,S82152N,S82152P,S82152Q | |
| | S82152R,S82153D,S82153E,S82153F,S82153G,S82153H,S82153J | |

Continued on next page

**Supplementary Table S1:** (cont'd) A comprehensive mapping of diagnosis groups used in the experiments to their corresponding ICD-10 and ICD-9 codes.

| Diagnosis | ICD-10 | ICD-9 |
|---|---|---|
| | S82153K,S82153M,S82153N,S82153P,S82153Q,S82153R,S82154D | |
| | S82154E,S82154F,S82154G,S82154H,S82154J,S82154K,S82154M | |
| | S82154N,S82154P,S82154Q,S82154R,S82155D,S82155E,S82155F | |
| | S82155G,S82155H,S82155J,S82155K,S82155M,S82155N,S82155P | |
| | S82155Q,S82155R,S82156D,S82156E,S82156F,S82156G,S82156H | |
| | S82156J,S82156K,S82156M,S82156N,S82156P,S82156Q,S82156R | |
| | S82161D,S82161G,S82161K,S82161P,S82162D,S82162G,S82162K | |
| | S82162P,S82169D,S82169G,S82169K,S82169P,S82191D,S82191E | |
| | S82191F,S82191G,S82191H,S82191J,S82191K,S82191M,S82191N | |
| | S82191P,S82191Q,S82191R,S82192D,S82192E,S82192F,S82192G | |
| | S82192H,S82192J,S82192K,S82192M,S82192N,S82192P,S82192Q | |
| | S82192R,S82199D,S82199E,S82199F,S82199G,S82199H,S82199J | |
| | S82199K,S82199M,S82199N,S82199P,S82199Q,S82199R,S82201D | |
| | S82201E,S82201F,S82201G,S82201H,S82201J,S82201K,S82201M | |
| | S82201N,S82201P,S82201Q,S82201R,S82202D,S82202E,S82202F | |
| | S82202G,S82202H,S82202J,S82202K,S82202M,S82202N,S82202P | |
| | S82202Q,S82202R,S82209D,S82209E,S82209F,S82209G,S82209H | |
| | S82209J,S82209K,S82209M,S82209N,S82209P,S82209Q,S82209R | |
| | S82221D,S82221E,S82221F,S82221G,S82221H,S82221J,S82221K | |
| | S82221M,S82221N,S82221P,S82221Q,S82221R,S82222D,S82222E | |
| | S82222F,S82222G,S82222H,S82222J,S82222K,S82222M,S82222N | |
| | S82222P,S82222Q,S82222R,S82223D,S82223E,S82223F,S82223G | |
| | S82223H,S82223J,S82223K,S82223M,S82223N,S82223P,S82223Q | |
| | S82223R,S82224D,S82224E,S82224F,S82224G,S82224H,S82224J | |
| | S82224K,S82224M,S82224N,S82224P,S82224Q,S82224R,S82225D | |
| | S82225E,S82225F,S82225G,S82225H,S82225J,S82225K,S82225M | |
| | S82225N,S82225P,S82225Q,S82225R,S82226D,S82226E,S82226F | |
| | S82226G,S82226H,S82226J,S82226K,S82226M,S82226N,S82226P | |
| | S82226Q,S82226R,S82231D,S82231E,S82231F,S82231G,S82231H | |
| | S82231J,S82231K,S82231M,S82231N,S82231P,S82231Q,S82231R | |
| | S82232D,S82232E,S82232F,S82232G,S82232H,S82232J,S82232K | |
| | S82232M,S82232N,S82232P,S82232Q,S82232R,S82233D,S82233E | |
| | S82233F,S82233G,S82233H,S82233J,S82233K,S82233M,S82233N | |
| | S82233P,S82233Q,S82233R,S82234D,S82234E,S82234F,S82234G | |
| | S82234H,S82234J,S82234K,S82234M,S82234N,S82234P,S82234Q | |
| | S82234R,S82235D,S82235E,S82235F,S82235G,S82235H,S82235J | |
| | S82235K,S82235M,S82235N,S82235P,S82235Q,S82235R,S82236D | |
| | S82236E,S82236F,S82236G,S82236H,S82236J,S82236K,S82236M | |
| | S82236N,S82236P,S82236Q,S82236R,S82241D,S82241E,S82241F | |
| | S82241G,S82241H,S82241J,S82241K,S82241M,S82241N,S82241P | |

Continued on next page

**Supplementary Table S1:** (cont'd) A comprehensive mapping of diagnosis groups used in the experiments to their corresponding ICD-10 and ICD-9 codes.

| Diagnosis | ICD-10 | ICD-9 |
|---|---|---|
| | S82241Q,S82241R,S82242D,S82242E,S82242F,S82242G,S82242H | |
| | S82242J,S82242K,S82242M,S82242N,S82242P,S82242Q,S82242R | |
| | S82243D,S82243E,S82243F,S82243G,S82243H,S82243J,S82243K | |
| | S82243M,S82243N,S82243P,S82243Q,S82243R,S82244D,S82244E | |
| | S82244F,S82244G,S82244H,S82244J,S82244K,S82244M,S82244N | |
| | S82244P,S82244Q,S82244R,S82245D,S82245E,S82245F,S82245G | |
| | S82245H,S82245J,S82245K,S82245M,S82245N,S82245P,S82245Q | |
| | S82245R,S82246D,S82246E,S82246F,S82246G,S82246H,S82246J | |
| | S82246K,S82246M,S82246N,S82246P,S82246Q,S82246R,S82251D | |
| | S82251E,S82251F,S82251G,S82251H,S82251J,S82251K,S82251M | |
| | S82251N,S82251P,S82251Q,S82251R,S82252D,S82252E,S82252F | |
| | S82252G,S82252H,S82252J,S82252K,S82252M,S82252N,S82252P | |
| | S82252Q,S82252R,S82253D,S82253E,S82253F,S82253G,S82253H | |
| | S82253J,S82253K,S82253M,S82253N,S82253P,S82253Q,S82253R | |
| | S82254D,S82254E,S82254F,S82254G,S82254H,S82254J,S82254K | |
| | S82254M,S82254N,S82254P,S82254Q,S82254R,S82255D,S82255E | |
| | S82255F,S82255G,S82255H,S82255J,S82255K,S82255M,S82255N | |
| | S82255P,S82255Q,S82255R,S82256D,S82256E,S82256F,S82256G | |
| | S82256H,S82256J,S82256K,S82256M,S82256N,S82256P,S82256Q | |
| | S82256R,S82261D,S82261E,S82261F,S82261G,S82261H,S82261J | |
| | S82261K,S82261M,S82261N,S82261P,S82261Q,S82261R,S82262D | |
| | S82262E,S82262F,S82262G,S82262H,S82262J,S82262K,S82262M | |
| | S82262N,S82262P,S82262Q,S82262R,S82263D,S82263E,S82263F | |
| | S82263G,S82263H,S82263J,S82263K,S82263M,S82263N,S82263P | |
| | S82263Q,S82263R,S82264D,S82264E,S82264F,S82264G,S82264H | |
| | S82264J,S82264K,S82264M,S82264N,S82264P,S82264Q,S82264R | |
| | S82265D,S82265E,S82265F,S82265G,S82265H,S82265J,S82265K | |
| | S82265M,S82265N,S82265P,S82265Q,S82265R,S82266D,S82266E | |
| | S82266F,S82266G,S82266H,S82266J,S82266K,S82266M,S82266N | |
| | S82266P,S82266Q,S82266R,S82291D,S82291E,S82291F,S82291G | |
| | S82291H,S82291J,S82291K,S82291M,S82291N,S82291P,S82291Q | |
| | S82291R,S82292D,S82292E,S82292F,S82292G,S82292H,S82292J | |
| | S82292K,S82292M,S82292N,S82292P,S82292Q,S82292R,S82299D | |
| | S82299E,S82299F,S82299G,S82299H,S82299J,S82299K,S82299M | |
| | S82299N,S82299P,S82299Q,S82299R,S82301D,S82301E,S82301F | |
| | S82301G,S82301H,S82301J,S82301K,S82301M,S82301N,S82301P | |
| | S82301Q,S82301R,S82302D,S82302E,S82302F,S82302G,S82302H | |
| | S82302J,S82302K,S82302M,S82302N,S82302P,S82302Q,S82302R | |
| | S82309D,S82309E,S82309F,S82309G,S82309H,S82309J,S82309K | |
| | S82309M,S82309N,S82309P,S82309Q,S82309R,S82311D,S82311G | |

Continued on next page

**Supplementary Table S1:** (cont'd) A comprehensive mapping of diagnosis groups used in the experiments to their corresponding ICD-10 and ICD-9 codes.

| Diagnosis | ICD-10 | ICD-9 |
|---|---|---|
| | S82311K,S82311P,S82312D,S82312G,S82312K,S82312P,S82319D | |
| | S82319G,S82319K,S82319P,S82391D,S82391E,S82391F,S82391G | |
| | S82391H,S82391J,S82391K,S82391M,S82391N,S82391P,S82391Q | |
| | S82391R,S82392D,S82392E,S82392F,S82392G,S82392H,S82392J | |
| | S82392K,S82392M,S82392N,S82392P,S82392Q,S82392R,S82399D | |
| | S82399E,S82399F,S82399G,S82399H,S82399J,S82399K,S82399M | |
| | S82399N,S82399P,S82399Q,S82399R,S82401D,S82401E,S82401F | |
| | S82401G,S82401H,S82401J,S82401K,S82401M,S82401N,S82401P | |
| | S82401Q,S82401R,S82402D,S82402E,S82402F,S82402G,S82402H | |
| | S82402J,S82402K,S82402M,S82402N,S82402P,S82402Q,S82402R | |
| | S82409D,S82409E,S82409F,S82409G,S82409H,S82409J,S82409K | |
| | S82409M,S82409N,S82409P,S82409Q,S82409R,S82421D,S82421E | |
| | S82421F,S82421G,S82421H,S82421J,S82421K,S82421M,S82421N | |
| | S82421P,S82421Q,S82421R,S82422D,S82422E,S82422F,S82422G | |
| | S82422H,S82422J,S82422K,S82422M,S82422N,S82422P,S82422Q | |
| | S82422R,S82423D,S82423E,S82423F,S82423G,S82423H,S82423J | |
| | S82423K,S82423M,S82423N,S82423P,S82423Q,S82423R,S82424D | |
| | S82424E,S82424F,S82424G,S82424H,S82424J,S82424K,S82424M | |
| | S82424N,S82424P,S82424Q,S82424R,S82425D,S82425E,S82425F | |
| | S82425G,S82425H,S82425J,S82425K,S82425M,S82425N,S82425P | |
| | S82425Q,S82425R,S82426D,S82426E,S82426F,S82426G,S82426H | |
| | S82426J,S82426K,S82426M,S82426N,S82426P,S82426Q,S82426R | |
| | S82431D,S82431E,S82431F,S82431G,S82431H,S82431J,S82431K | |
| | S82431M,S82431N,S82431P,S82431Q,S82431R,S82432D,S82432E | |
| | S82432F,S82432G,S82432H,S82432J,S82432K,S82432M,S82432N | |
| | S82432P,S82432Q,S82432R,S82433D,S82433E,S82433F,S82433G | |
| | S82433H,S82433J,S82433K,S82433M,S82433N,S82433P,S82433Q | |
| | S82433R,S82434D,S82434E,S82434F,S82434G,S82434H,S82434J | |
| | S82434K,S82434M,S82434N,S82434P,S82434Q,S82434R,S82435D | |
| | S82435E,S82435F,S82435G,S82435H,S82435J,S82435K,S82435M | |
| | S82435N,S82435P,S82435Q,S82435R,S82436D,S82436E,S82436F | |
| | S82436G,S82436H,S82436J,S82436K,S82436M,S82436N,S82436P | |
| | S82436Q,S82436R,S82441D,S82441E,S82441F,S82441G,S82441H | |
| | S82441J,S82441K,S82441M,S82441N,S82441P,S82441Q,S82441R | |
| | S82442D,S82442E,S82442F,S82442G,S82442H,S82442J,S82442K | |
| | S82442M,S82442N,S82442P,S82442Q,S82442R,S82443D,S82443E | |
| | S82443F,S82443G,S82443H,S82443J,S82443K,S82443M,S82443N | |
| | S82443P,S82443Q,S82443R,S82444D,S82444E,S82444F,S82444G | |
| | S82444H,S82444J,S82444K,S82444M,S82444N,S82444P,S82444Q | |
| | S82444R,S82445D,S82445E,S82445F,S82445G,S82445H,S82445J | |

Continued on next page

**Supplementary Table S1:** (cont'd) A comprehensive mapping of diagnosis groups used in the experiments to their corresponding ICD-10 and ICD-9 codes.

| Diagnosis | ICD-10 | ICD-9 |
|---|---|---|
| | S82445K,S82445M,S82445N,S82445P,S82445Q,S82445R,S82446D | |
| | S82446E,S82446F,S82446G,S82446H,S82446J,S82446K,S82446M | |
| | S82446N,S82446P,S82446Q,S82446R,S82451D,S82451E,S82451F | |
| | S82451G,S82451H,S82451J,S82451K,S82451M,S82451N,S82451P | |
| | S82451Q,S82451R,S82452D,S82452E,S82452F,S82452G,S82452H | |
| | S82452J,S82452K,S82452M,S82452N,S82452P,S82452Q,S82452R | |
| | S82453D,S82453E,S82453F,S82453G,S82453H,S82453J,S82453K | |
| | S82453M,S82453N,S82453P,S82453Q,S82453R,S82454D,S82454E | |
| | S82454F,S82454G,S82454H,S82454J,S82454K,S82454M,S82454N | |
| | S82454P,S82454Q,S82454R,S82455D,S82455E,S82455F,S82455G | |
| | S82455H,S82455J,S82455K,S82455M,S82455N,S82455P,S82455Q | |
| | S82455R,S82456D,S82456E,S82456F,S82456G,S82456H,S82456J | |
| | S82456K,S82456M,S82456N,S82456P,S82456Q,S82456R,S82461D | |
| | S82461E,S82461F,S82461G,S82461H,S82461J,S82461K,S82461M | |
| | S82461N,S82461P,S82461Q,S82461R,S82462D,S82462E,S82462F | |
| | S82462G,S82462H,S82462J,S82462K,S82462M,S82462N,S82462P | |
| | S82462Q,S82462R,S82463D,S82463E,S82463F,S82463G,S82463H | |
| | S82463J,S82463K,S82463M,S82463N,S82463P,S82463Q,S82463R | |
| | S82464D,S82464E,S82464F,S82464G,S82464H,S82464J,S82464K | |
| | S82464M,S82464N,S82464P,S82464Q,S82464R,S82465D,S82465E | |
| | S82465F,S82465G,S82465H,S82465J,S82465K,S82465M,S82465N | |
| | S82465P,S82465Q,S82465R,S82466D,S82466E,S82466F,S82466G | |
| | S82466H,S82466J,S82466K,S82466M,S82466N,S82466P,S82466Q | |
| | S82466R,S82491D,S82491E,S82491F,S82491G,S82491H,S82491J | |
| | S82491K,S82491M,S82491N,S82491P,S82491Q,S82491R,S82492D | |
| | S82492E,S82492F,S82492G,S82492H,S82492J,S82492K,S82492M | |
| | S82492N,S82492P,S82492Q,S82492R,S82499D,S82499E,S82499F | |
| | S82499G,S82499H,S82499J,S82499K,S82499M,S82499N,S82499P | |
| | S82499Q,S82499R,S8251XD,S8251XE,S8251XF,S8251XG,S8251XH | |
| | S8251XJ,S8251XK,S8251XM,S8251XN,S8251XP,S8251XQ,S8251XR | |
| | S8252XD,S8252XE,S8252XF,S8252XG,S8252XH,S8252XJ,S8252XK | |
| | S8252XM,S8252XN,S8252XP,S8252XQ,S8252XR,S8253XD,S8253XE | |
| | S8253XF,S8253XG,S8253XH,S8253XJ,S8253XK,S8253XM,S8253XN | |
| | S8253XP,S8253XQ,S8253XR,S8254XD,S8254XE,S8254XF,S8254XG | |
| | S8254XH,S8254XJ,S8254XK,S8254XM,S8254XN,S8254XP,S8254XQ | |
| | S8254XR,S8255XD,S8255XE,S8255XF,S8255XG,S8255XH,S8255XJ | |
| | S8255XK,S8255XM,S8255XN,S8255XP,S8255XQ,S8255XR,S8256XD | |
| | S8256XE,S8256XF,S8256XG,S8256XH,S8256XJ,S8256XK,S8256XM | |
| | S8256XN,S8256XP,S8256XQ,S8256XR,S8261XD,S8261XE,S8261XF | |
| | S8261XG,S8261XH,S8261XJ,S8261XK,S8261XM,S8261XN,S8261XP | |

Continued on next page

**Supplementary Table S1:** (cont'd) A comprehensive mapping of diagnosis groups used in the experiments to their corresponding ICD-10 and ICD-9 codes.

| Diagnosis | ICD-10 | ICD-9 |
|---|---|---|
| | S8261XQ,S8261XR,S8262XD,S8262XE,S8262XF,S8262XG,S8262XH | |
| | S8262XJ,S8262XK,S8262XM,S8262XN,S8262XP,S8262XQ,S8262XR | |
| | S8263XD,S8263XE,S8263XF,S8263XG,S8263XH,S8263XJ,S8263XK | |
| | S8263XM,S8263XN,S8263XP,S8263XQ,S8263XR,S8264XD,S8264XE | |
| | S8264XF,S8264XG,S8264XH,S8264XJ,S8264XK,S8264XM,S8264XN | |
| | S8264XP,S8264XQ,S8264XR,S8265XD,S8265XE,S8265XF,S8265XG | |
| | S8265XH,S8265XJ,S8265XK,S8265XM,S8265XN,S8265XP,S8265XQ | |
| | S8265XR,S8266XD,S8266XE,S8266XF,S8266XG,S8266XH,S8266XJ | |
| | S8266XK,S8266XM,S8266XN,S8266XP,S8266XQ,S8266XR,S82811D | |
| | S82811G,S82811K,S82811P,S82812D,S82812G,S82812K,S82812P | |
| | S82819D,S82819G,S82819K,S82819P,S82821D,S82821G,S82821K | |
| | S82821P,S82822D,S82822G,S82822K,S82822P,S82829D,S82829G | |
| | S82829K,S82829P,S82831D,S82831E,S82831F,S82831G,S82831H | |
| | S82831J,S82831K,S82831M,S82831N,S82831P,S82831Q,S82831R | |
| | S82832D,S82832E,S82832F,S82832G,S82832H,S82832J,S82832K | |
| | S82832M,S82832N,S82832P,S82832Q,S82832R,S82839D,S82839E | |
| | S82839F,S82839G,S82839H,S82839J,S82839K,S82839M,S82839N | |
| | S82839P,S82839Q,S82839R,S82841D,S82841E,S82841F,S82841G | |
| | S82841H,S82841J,S82841K,S82841M,S82841N,S82841P,S82841Q | |
| | S82841R,S82842D,S82842E,S82842F,S82842G,S82842H,S82842J | |
| | S82842K,S82842M,S82842N,S82842P,S82842Q,S82842R,S82843D | |
| | S82843E,S82843F,S82843G,S82843H,S82843J,S82843K,S82843M | |
| | S82843N,S82843P,S82843Q,S82843R,S82844D,S82844E,S82844F | |
| | S82844G,S82844H,S82844J,S82844K,S82844M,S82844N,S82844P | |
| | S82844Q,S82844R,S82845D,S82845E,S82845F,S82845G,S82845H | |
| | S82845J,S82845K,S82845M,S82845N,S82845P,S82845Q,S82845R | |
| | S82846D,S82846E,S82846F,S82846G,S82846H,S82846J,S82846K | |
| | S82846M,S82846N,S82846P,S82846Q,S82846R,S82851D,S82851E | |
| | S82851F,S82851G,S82851H,S82851J,S82851K,S82851M,S82851N | |
| | S82851P,S82851Q,S82851R,S82852D,S82852E,S82852F,S82852G | |
| | S82852H,S82852J,S82852K,S82852M,S82852N,S82852P,S82852Q | |
| | S82852R,S82853D,S82853E,S82853F,S82853G,S82853H,S82853J | |
| | S82853K,S82853M,S82853N,S82853P,S82853Q,S82853R,S82854D | |
| | S82854E,S82854F,S82854G,S82854H,S82854J,S82854K,S82854M | |
| | S82854N,S82854P,S82854Q,S82854R,S82855D,S82855E,S82855F | |
| | S82855G,S82855H,S82855J,S82855K,S82855M,S82855N,S82855P | |
| | S82855Q,S82855R,S82856D,S82856E,S82856F,S82856G,S82856H | |
| | S82856J,S82856K,S82856M,S82856N,S82856P,S82856Q,S82856R | |
| | S82861D,S82861E,S82861F,S82861G,S82861H,S82861J,S82861K | |
| | S82861M,S82861N,S82861P,S82861Q,S82861R,S82862D,S82862E | |

Continued on next page

**Supplementary Table S1:** (cont'd) A comprehensive mapping of diagnosis groups used in the experiments to their corresponding ICD-10 and ICD-9 codes.

| Diagnosis | ICD-10 | ICD-9 |
|---|---|---|
| | S82862F,S82862G,S82862H,S82862J,S82862K,S82862M,S82862N | |
| | S82862P,S82862Q,S82862R,S82863D,S82863E,S82863F,S82863G | |
| | S82863H,S82863J,S82863K,S82863M,S82863N,S82863P,S82863Q | |
| | S82863R,S82864D,S82864E,S82864F,S82864G,S82864H,S82864J | |
| | S82864K,S82864M,S82864N,S82864P,S82864Q,S82864R,S82865D | |
| | S82865E,S82865F,S82865G,S82865H,S82865J,S82865K,S82865M | |
| | S82865N,S82865P,S82865Q,S82865R,S82866D,S82866E,S82866F | |
| | S82866G,S82866H,S82866J,S82866K,S82866M,S82866N,S82866P | |
| | S82866Q,S82866R,S82871D,S82871E,S82871F,S82871G,S82871H | |
| | S82871J,S82871K,S82871M,S82871N,S82871P,S82871Q,S82871R | |
| | S82872D,S82872E,S82872F,S82872G,S82872H,S82872J,S82872K | |
| | S82872M,S82872N,S82872P,S82872Q,S82872R,S82873D,S82873E | |
| | S82873F,S82873G,S82873H,S82873J,S82873K,S82873M,S82873N | |
| | S82873P,S82873Q,S82873R,S82874D,S82874E,S82874F,S82874G | |
| | S82874H,S82874J,S82874K,S82874M,S82874N,S82874P,S82874Q | |
| | S82874R,S82875D,S82875E,S82875F,S82875G,S82875H,S82875J | |
| | S82875K,S82875M,S82875N,S82875P,S82875Q,S82875R,S82876D | |
| | S82876E,S82876F,S82876G,S82876H,S82876J,S82876K,S82876M | |
| | S82876N,S82876P,S82876Q,S82876R,S82891D,S82891E,S82891F | |
| | S82891G,S82891H,S82891J,S82891K,S82891M,S82891N,S82891P | |
| | S82891Q,S82891R,S82892D,S82892E,S82892F,S82892G,S82892H | |
| | S82892J,S82892K,S82892M,S82892N,S82892P,S82892Q,S82892R | |
| | S82899D,S82899E,S82899F,S82899G,S82899H,S82899J,S82899K | |
| | S82899M,S82899N,S82899P,S82899Q,S82899R,S8290XD,S8290XE | |
| | S8290XF,S8290XG,S8290XH,S8290XJ,S8290XK,S8290XM,S8290XN | |
| | S8290XP,S8290XQ,S8290XR,S8291XD,S8291XE,S8291XF,S8291XG | |
| | S8291XH,S8291XJ,S8291XK,S8291XM,S8291XN,S8291XP,S8291XQ | |
| | S8291XR,S8292XD,S8292XE,S8292XF,S8292XG,S8292XH,S8292XJ | |
| | S8292XK,S8292XM,S8292XN,S8292XP,S8292XQ,S8292XR,S89001D | |
| | S89001G,S89001K,S89001P,S89002D,S89002G,S89002K,S89002P | |
| | S89009D,S89009G,S89009K,S89009P,S89011D,S89011G,S89011K | |
| | S89011P,S89012D,S89012G,S89012K,S89012P,S89019D,S89019G | |
| | S89019K,S89019P,S89021D,S89021G,S89021K,S89021P,S89022D | |
| | S89022G,S89022K,S89022P,S89029D,S89029G,S89029K,S89029P | |
| | S89031D,S89031G,S89031K,S89031P,S89032D,S89032G,S89032K | |
| | S89032P,S89039D,S89039G,S89039K,S89039P,S89041D,S89041G | |
| | S89041K,S89041P,S89042D,S89042G,S89042K,S89042P,S89049D | |
| | S89049G,S89049K,S89049P,S89091D,S89091G,S89091K,S89091P | |
| | S89092D,S89092G,S89092K,S89092P,S89099D,S89099G,S89099K | |
| | S89099P,S89101D,S89101G,S89101K,S89101P,S89102D,S89102G | |

Continued on next page

**Supplementary Table S1:** (cont'd) A comprehensive mapping of diagnosis groups used in the experiments to their corresponding ICD-10 and ICD-9 codes.

| Diagnosis | ICD-10 | ICD-9 |
|---|---|---|
| | S89102K,S89102P,S89109D,S89109G,S89109K,S89109P,S89111D | |
| | S89111G,S89111K,S89111P,S89112D,S89112G,S89112K,S89112P | |
| | S89119D,S89119G,S89119K,S89119P,S89121D,S89121G,S89121K | |
| | S89121P,S89122D,S89122G,S89122K,S89122P,S89129D,S89129G | |
| | S89129K,S89129P,S89131D,S89131G,S89131K,S89131P,S89132D | |
| | S89132G,S89132K,S89132P,S89139D,S89139G,S89139K,S89139P | |
| | S89141D,S89141G,S89141K,S89141P,S89142D,S89142G,S89142K | |
| | S89142P,S89149D,S89149G,S89149K,S89149P,S89191D,S89191G | |
| | S89191K,S89191P,S89192D,S89192G,S89192K,S89192P,S89199D | |
| | S89199G,S89199K,S89199P,S89201D,S89201G,S89201K,S89201P | |
| | S89202D,S89202G,S89202K,S89202P,S89209D,S89209G,S89209K | |
| | S89209P,S89211D,S89211G,S89211K,S89211P,S89212D,S89212G | |
| | S89212K,S89212P,S89219D,S89219G,S89219K,S89219P,S89221D | |
| | S89221G,S89221K,S89221P,S89222D,S89222G,S89222K,S89222P | |
| | S89229D,S89229G,S89229K,S89229P,S89291D,S89291G,S89291K | |
| | S89291P,S89292D,S89292G,S89292K,S89292P,S89299D,S89299G | |
| | S89299K,S89299P,S89301D,S89301G,S89301K,S89301P,S89302D | |
| | S89302G,S89302K,S89302P,S89309D,S89309G,S89309K,S89309P | |
| | S89311D,S89311G,S89311K,S89311P,S89312D,S89312G,S89312K | |
| | S89312P,S89319D,S89319G,S89319K,S89319P,S89321D,S89321G | |
| | S89321K,S89321P,S89322D,S89322G,S89322K,S89322P,S89329D | |
| | S89329G,S89329K,S89329P,S89391D,S89391G,S89391K,S89391P | |
| | S89392D,S89392G,S89392K,S89392P,S89399D,S89399G,S89399K | |
| | S89399P,S92001D,S92001G,S92001K,S92001P,S92002D,S92002G | |
| | S92002K,S92002P,S92009D,S92009G,S92009K,S92009P,S92011D | |
| | S92011G,S92011K,S92011P,S92012D,S92012G,S92012K,S92012P | |
| | S92013D,S92013G,S92013K,S92013P,S92014D,S92014G,S92014K | |
| | S92014P,S92015D,S92015G,S92015K,S92015P,S92016D,S92016G | |
| | S92016K,S92016P,S92021D,S92021G,S92021K,S92021P,S92022D | |
| | S92022G,S92022K,S92022P,S92023D,S92023G,S92023K,S92023P | |
| | S92024D,S92024G,S92024K,S92024P,S92025D,S92025G,S92025K | |
| | S92025P,S92026D,S92026G,S92026K,S92026P,S92031D,S92031G | |
| | S92031K,S92031P,S92032D,S92032G,S92032K,S92032P,S92033D | |
| | S92033G,S92033K,S92033P,S92034D,S92034G,S92034K,S92034P | |
| | S92035D,S92035G,S92035K,S92035P,S92036D,S92036G,S92036K | |
| | S92036P,S92041D,S92041G,S92041K,S92041P,S92042D,S92042G | |
| | S92042K,S92042P,S92043D,S92043G,S92043K,S92043P,S92044D | |
| | S92044G,S92044K,S92044P,S92045D,S92045G,S92045K,S92045P | |
| | S92046D,S92046G,S92046K,S92046P,S92051D,S92051G,S92051K | |
| | S92051P,S92052D,S92052G,S92052K,S92052P,S92053D,S92053G | |

Continued on next page

**Supplementary Table S1:** (cont'd) A comprehensive mapping of diagnosis groups used in the experiments to their corresponding ICD-10 and ICD-9 codes.

| Diagnosis | ICD-10 | ICD-9 |
|---|---|---|
| | S92053K,S92053P,S92054D,S92054G,S92054K,S92054P,S92055D | |
| | S92055G,S92055K,S92055P,S92056D,S92056G,S92056K,S92056P | |
| | S92061D,S92061G,S92061K,S92061P,S92062D,S92062G,S92062K | |
| | S92062P,S92063D,S92063G,S92063K,S92063P,S92064D,S92064G | |
| | S92064K,S92064P,S92065D,S92065G,S92065K,S92065P,S92066D | |
| | S92066G,S92066K,S92066P,S92101D,S92101G,S92101K,S92101P | |
| | S92102D,S92102G,S92102K,S92102P,S92109D,S92109G,S92109K | |
| | S92109P,S92111D,S92111G,S92111K,S92111P,S92112D,S92112G | |
| | S92112K,S92112P,S92113D,S92113G,S92113K,S92113P,S92114D | |
| | S92114G,S92114K,S92114P,S92115D,S92115G,S92115K,S92115P | |
| | S92116D,S92116G,S92116K,S92116P,S92121D,S92121G,S92121K | |
| | S92121P,S92122D,S92122G,S92122K,S92122P,S92123D,S92123G | |
| | S92123K,S92123P,S92124D,S92124G,S92124K,S92124P,S92125D | |
| | S92125G,S92125K,S92125P,S92126D,S92126G,S92126K,S92126P | |
| | S92131D,S92131G,S92131K,S92131P,S92132D,S92132G,S92132K | |
| | S92132P,S92133D,S92133G,S92133K,S92133P,S92134D,S92134G | |
| | S92134K,S92134P,S92135D,S92135G,S92135K,S92135P,S92136D | |
| | S92136G,S92136K,S92136P,S92141D,S92141G,S92141K,S92141P | |
| | S92142D,S92142G,S92142K,S92142P,S92143D,S92143G,S92143K | |
| | S92143P,S92144D,S92144G,S92144K,S92144P,S92145D,S92145G | |
| | S92145K,S92145P,S92146D,S92146G,S92146K,S92146P,S92151D | |
| | S92151G,S92151K,S92151P,S92152D,S92152G,S92152K,S92152P | |
| | S92153D,S92153G,S92153K,S92153P,S92154D,S92154G,S92154K | |
| | S92154P,S92155D,S92155G,S92155K,S92155P,S92156D,S92156G | |
| | S92156K,S92156P,S92191D,S92191G,S92191K,S92191P,S92192D | |
| | S92192G,S92192K,S92192P,S92199D,S92199G,S92199K,S92199P | |
| | S92201D,S92201G,S92201K,S92201P,S92202D,S92202G,S92202K | |
| | S92202P,S92209D,S92209G,S92209K,S92209P,S92211D,S92211G | |
| | S92211K,S92211P,S92212D,S92212G,S92212K,S92212P,S92213D | |
| | S92213G,S92213K,S92213P,S92214D,S92214G,S92214K,S92214P | |
| | S92215D,S92215G,S92215K,S92215P,S92216D,S92216G,S92216K | |
| | S92216P,S92221D,S92221G,S92221K,S92221P,S92222D,S92222G | |
| | S92222K,S92222P,S92223D,S92223G,S92223K,S92223P,S92224D | |
| | S92224G,S92224K,S92224P,S92225D,S92225G,S92225K,S92225P | |
| | S92226D,S92226G,S92226K,S92226P,S92231D,S92231G,S92231K | |
| | S92231P,S92232D,S92232G,S92232K,S92232P,S92233D,S92233G | |
| | S92233K,S92233P,S92234D,S92234G,S92234K,S92234P,S92235D | |
| | S92235G,S92235K,S92235P,S92236D,S92236G,S92236K,S92236P | |
| | S92241D,S92241G,S92241K,S92241P,S92242D,S92242G,S92242K | |
| | S92242P,S92243D,S92243G,S92243K,S92243P,S92244D,S92244G | |

Continued on next page

**Supplementary Table S1:** (cont'd) A comprehensive mapping of diagnosis groups used in the experiments to their corresponding ICD-10 and ICD-9 codes.

| Diagnosis | ICD-10 | ICD-9 |
|---|---|---|
| | S92244K,S92244P,S92245D,S92245G,S92245K,S92245P,S92246D | |
| | S92246G,S92246K,S92246P,S92251D,S92251G,S92251K,S92251P | |
| | S92252D,S92252G,S92252K,S92252P,S92253D,S92253G,S92253K | |
| | S92253P,S92254D,S92254G,S92254K,S92254P,S92255D,S92255G | |
| | S92255K,S92255P,S92256D,S92256G,S92256K,S92256P,S92301D | |
| | S92301G,S92301K,S92301P,S92302D,S92302G,S92302K,S92302P | |
| | S92309D,S92309G,S92309K,S92309P,S92311D,S92311G,S92311K | |
| | S92311P,S92312D,S92312G,S92312K,S92312P,S92313D,S92313G | |
| | S92313K,S92313P,S92314D,S92314G,S92314K,S92314P,S92315D | |
| | S92315G,S92315K,S92315P,S92316D,S92316G,S92316K,S92316P | |
| | S92321D,S92321G,S92321K,S92321P,S92322D,S92322G,S92322K | |
| | S92322P,S92323D,S92323G,S92323K,S92323P,S92324D,S92324G | |
| | S92324K,S92324P,S92325D,S92325G,S92325K,S92325P,S92326D | |
| | S92326G,S92326K,S92326P,S92331D,S92331G,S92331K,S92331P | |
| | S92332D,S92332G,S92332K,S92332P,S92333D,S92333G,S92333K | |
| | S92333P,S92334D,S92334G,S92334K,S92334P,S92335D,S92335G | |
| | S92335K,S92335P,S92336D,S92336G,S92336K,S92336P,S92341D | |
| | S92341G,S92341K,S92341P,S92342D,S92342G,S92342K,S92342P | |
| | S92343D,S92343G,S92343K,S92343P,S92344D,S92344G,S92344K | |
| | S92344P,S92345D,S92345G,S92345K,S92345P,S92346D,S92346G | |
| | S92346K,S92346P,S92351D,S92351G,S92351K,S92351P,S92352D | |
| | S92352G,S92352K,S92352P,S92353D,S92353G,S92353K,S92353P | |
| | S92354D,S92354G,S92354K,S92354P,S92355D,S92355G,S92355K | |
| | S92355P,S92356D,S92356G,S92356K,S92356P,S92401D,S92401G | |
| | S92401K,S92401P,S92402D,S92402G,S92402K,S92402P,S92403D | |
| | S92403G,S92403K,S92403P,S92404D,S92404G,S92404K,S92404P | |
| | S92405D,S92405G,S92405K,S92405P,S92406D,S92406G,S92406K | |
| | S92406P,S92411D,S92411G,S92411K,S92411P,S92412D,S92412G | |
| | S92412K,S92412P,S92413D,S92413G,S92413K,S92413P,S92414D | |
| | S92414G,S92414K,S92414P,S92415D,S92415G,S92415K,S92415P | |
| | S92416D,S92416G,S92416K,S92416P,S92421D,S92421G,S92421K | |
| | S92421P,S92422D,S92422G,S92422K,S92422P,S92423D,S92423G | |
| | S92423K,S92423P,S92424D,S92424G,S92424K,S92424P,S92425D | |
| | S92425G,S92425K,S92425P,S92426D,S92426G,S92426K,S92426P | |
| | S92491D,S92491G,S92491K,S92491P,S92492D,S92492G,S92492K | |
| | S92492P,S92499D,S92499G,S92499K,S92499P,S92501D,S92501G | |
| | S92501K,S92501P,S92502D,S92502G,S92502K,S92502P,S92503D | |
| | S92503G,S92503K,S92503P,S92504D,S92504G,S92504K,S92504P | |
| | S92505D,S92505G,S92505K,S92505P,S92506D,S92506G,S92506K | |
| | S92506P,S92511D,S92511G,S92511K,S92511P,S92512D,S92512G | |

Continued on next page

**Supplementary Table S1:** (cont'd) A comprehensive mapping of diagnosis groups used in the experiments to their corresponding ICD-10 and ICD-9 codes.

| Diagnosis | ICD-10 | ICD-9 |
|---|---|---|
| | S92512K,S92512P,S92513D,S92513G,S92513K,S92513P,S92514D | |
| | S92514G,S92514K,S92514P,S92515D,S92515G,S92515K,S92515P | |
| | S92516D,S92516G,S92516K,S92516P,S92521D,S92521G,S92521K | |
| | S92521P,S92522D,S92522G,S92522K,S92522P,S92523D,S92523G | |
| | S92523K,S92523P,S92524D,S92524G,S92524K,S92524P,S92525D | |
| | S92525G,S92525K,S92525P,S92526D,S92526G,S92526K,S92526P | |
| | S92531D,S92531G,S92531K,S92531P,S92532D,S92532G,S92532K | |
| | S92532P,S92533D,S92533G,S92533K,S92533P,S92534D,S92534G | |
| | S92534K,S92534P,S92535D,S92535G,S92535K,S92535P,S92536D | |
| | S92536G,S92536K,S92536P,S92591D,S92591G,S92591K,S92591P | |
| | S92592D,S92592G,S92592K,S92592P,S92599D,S92599G,S92599K | |
| | S92599P,S92811D,S92811G,S92811K,S92811P,S92812D,S92812G | |
| | S92812K,S92812P,S92819D,S92819G,S92819K,S92819P,S92901D | |
| | S92901G,S92901K,S92901P,S92902D,S92902G,S92902K,S92902P | |
| | S92909D,S92909G,S92909K,S92909P,S92911D,S92911G,S92911K | |
| | S92911P,S92912D,S92912G,S92912K,S92912P,S92919D,S92919G | |
| | S92919K,S92919P,S99001D,S99001G,S99001K,S99001P,S99002D | |
| | S99002G,S99002K,S99002P,S99009D,S99009G,S99009K,S99009P | |
| | S99011D,S99011G,S99011K,S99011P,S99012D,S99012G,S99012K | |
| | S99012P,S99019D,S99019G,S99019K,S99019P,S99021D,S99021G | |
| | S99021K,S99021P,S99022D,S99022G,S99022K,S99022P,S99029D | |
| | S99029G,S99029K,S99029P,S99031D,S99031G,S99031K,S99031P | |
| | S99032D,S99032G,S99032K,S99032P,S99039D,S99039G,S99039K | |
| | S99039P,S99041D,S99041G,S99041K,S99041P,S99042D,S99042G | |
| | S99042K,S99042P,S99049D,S99049G,S99049K,S99049P,S99091D | |
| | S99091G,S99091K,S99091P,S99092D,S99092G,S99092K,S99092P | |
| | S99099D,S99099G,S99099K,S99099P,S99101D,S99101G,S99101K | |
| | S99101P,S99102D,S99102G,S99102K,S99102P,S99109D,S99109G | |
| | S99109K,S99109P,S99111D,S99111G,S99111K,S99111P,S99112D | |
| | S99112G,S99112K,S99112P,S99119D,S99119G,S99119K,S99119P | |
| | S99121D,S99121G,S99121K,S99121P,S99122D,S99122G,S99122K | |
| | S99122P,S99129D,S99129G,S99129K,S99129P,S99131D,S99131G | |
| | S99131K,S99131P,S99132D,S99132G,S99132K,S99132P,S99139D | |
| | S99139G,S99139K,S99139P,S99141D,S99141G,S99141K,S99141P | |
| | S99142D,S99142G,S99142K,S99142P,S99149D,S99149G,S99149K | |
| | S99149P,S99191D,S99191G,S99191K,S99191P,S99192D,S99192G | |
| | S99192K,S99192P,S99199D,S99199G,S99199K,S99199P,S99201D | |
| | S99201G,S99201K,S99201P,S99202D,S99202G,S99202K,S99202P | |
| | S99209D,S99209G,S99209K,S99209P,S99211D,S99211G,S99211K | |
| | S99211P,S99212D,S99212G,S99212K,S99212P,S99219D,S99219G | |

Continued on next page

**Supplementary Table S1:** (cont'd) A comprehensive mapping of diagnosis groups used in the experiments to their corresponding ICD-10 and ICD-9 codes.

| Diagnosis | ICD-10 | ICD-9 |
|---|---|---|
| | S99219K,S99219P,S99221D,S99221G,S99221K,S99221P,S99222D | |
| | S99222G,S99222K,S99222P,S99229D,S99229G,S99229K,S99229P | |
| | S99231D,S99231G,S99231K,S99231P,S99232D,S99232G,S99232K | |
| | S99232P,S99239D,S99239G,S99239K,S99239P,S99241D,S99241G | |
| | S99241K,S99241P,S99242D,S99242G,S99242K,S99242P,S99249D | |
| | S99249G,S99249K,S99249P,S99291D,S99291G,S99291K,S99291P | |
| | S99292D,S99292G,S99292K,S99292P,S99299D,S99299G,S99299K | |
| | S99299P,T84042D,T84043D,T84048D,T84049D | |
| Fracture of the lower | M9711XA,M9712XA,M9721XA,M9722XA,M978XXA,M979XXA,S72301A | 82001,82100,82101,82110,82111,82120,82121,82122,82123,82129 |
| limb (except hip), | S72301B,S72301C,S72302A,S72302B,S72302C,S72309A,S72309B | 82130,82131,82132,82133,82139,8220,8221,82300,82301,82310 |
| initial encounter | S72309C,S72321A,S72321B,S72321C,S72322A,S72322B,S72322C | 82311,82320,82321,82330,82331,82340,82341,82380,82381,82390 |
| | S72323A,S72323B,S72323C,S72324A,S72324B,S72324C,S72325A | 82391,8240,8241,8242,8243,8244,8245,8246,8247,8248,8249,8250 |
| | S72325B,S72325C,S72326A,S72326B,S72326C,S72331A,S72331B | 8251,82520,82521,82522,82523,82524,82525,82529,82530,82531 |
| | S72331C,S72332A,S72332B,S72332C,S72333A,S72333B,S72333C | 82532,82533,82534,82535,82539,8260,8261,8270,8271,99644 |
| | S72334A,S72334B,S72334C,S72335A,S72335B,S72335C,S72336A | |
| | S72336B,S72336C,S72341A,S72341B,S72341C,S72342A,S72342B | |
| | S72342C,S72343A,S72343B,S72343C,S72344A,S72344B,S72344C | |
| | S72345A,S72345B,S72345C,S72346A,S72346B,S72346C,S72351A | |
| | S72351B,S72351C,S72352A,S72352B,S72352C,S72353A,S72353B | |
| | S72353C,S72354A,S72354B,S72354C,S72355A,S72355B,S72355C | |
| | S72356A,S72356B,S72356C,S72361A,S72361B,S72361C,S72362A | |
| | S72362B,S72362C,S72363A,S72363B,S72363C,S72364A,S72364B | |
| | S72364C,S72365A,S72365B,S72365C,S72366A,S72366B,S72366C | |
| | S72391A,S72391B,S72391C,S72392A,S72392B,S72392C,S72399A | |
| | S72399B,S72399C,S72401A,S72401B,S72401C,S72402A,S72402B | |
| | S72402C,S72409A,S72409B,S72409C,S72411A,S72411B,S72411C | |
| | S72412A,S72412B,S72412C,S72413A,S72413B,S72413C,S72414A | |
| | S72414B,S72414C,S72415A,S72415B,S72415C,S72416A,S72416B | |
| | S72416C,S72421A,S72421B,S72421C,S72422A,S72422B,S72422C | |
| | S72423A,S72423B,S72423C,S72424A,S72424B,S72424C,S72425A | |
| | S72425B,S72425C,S72426A,S72426B,S72426C,S72431A,S72431B | |
| | S72431C,S72432A,S72432B,S72432C,S72433A,S72433B,S72433C | |
| | S72434A,S72434B,S72434C,S72435A,S72435B,S72435C,S72436A | |
| | S72436B,S72436C,S72441A,S72441B,S72441C,S72442A,S72442B | |
| | S72442C,S72443A,S72443B,S72443C,S72444A,S72444B,S72444C | |
| | S72445A,S72445B,S72445C,S72446A,S72446B,S72446C,S72451A | |
| | S72451B,S72451C,S72452A,S72452B,S72452C,S72453A,S72453B | |
| | S72453C,S72454A,S72454B,S72454C,S72455A,S72455B,S72455C | |
| | S72456A,S72456B,S72456C,S72461A,S72461B,S72461C,S72462A | |

Continued on next page

**Supplementary Table S1:** (cont'd) A comprehensive mapping of diagnosis groups used in the experiments to their corresponding ICD-10 and ICD-9 codes.

| Diagnosis | ICD-10 | ICD-9 |
|---|---|---|
| | S72462B,S72462C,S72463A,S72463B,S72463C,S72464A,S72464B | |
| | S72464C,S72465A,S72465B,S72465C,S72466A,S72466B,S72466C | |
| | S72471A,S72472A,S72479A,S72491A,S72491B,S72491C,S72492A | |
| | S72492B,S72492C,S72499A,S72499B,S72499C,S728X1A,S728X1B | |
| | S728X1C,S728X2A,S728X2B,S728X2C,S728X9A,S728X9B,S728X9C | |
| | S7290XA,S7290XB,S7290XC,S7291XA,S7291XB,S7291XC,S7292XA | |
| | S7292XB,S7292XC,S79001A,S79002A,S79009A,S79011A,S79012A | |
| | S79019A,S79091A,S79092A,S79099A,S79101A,S79102A,S79109A | |
| | S79111A,S79112A,S79119A,S79121A,S79122A,S79129A,S79131A | |
| | S79132A,S79139A,S79141A,S79142A,S79149A,S79191A,S79192A | |
| | S79199A,S82001A,S82001B,S82001C,S82002A,S82002B,S82002C | |
| | S82009A,S82009B,S82009C,S82011A,S82011B,S82011C,S82012A | |
| | S82012B,S82012C,S82013A,S82013B,S82013C,S82014A,S82014B | |
| | S82014C,S82015A,S82015B,S82015C,S82016A,S82016B,S82016C | |
| | S82021A,S82021B,S82021C,S82022A,S82022B,S82022C,S82023A | |
| | S82023B,S82023C,S82024A,S82024B,S82024C,S82025A,S82025B | |
| | S82025C,S82026A,S82026B,S82026C,S82031A,S82031B,S82031C | |
| | S82032A,S82032B,S82032C,S82033A,S82033B,S82033C,S82034A | |
| | S82034B,S82034C,S82035A,S82035B,S82035C,S82036A,S82036B | |
| | S82036C,S82041A,S82041B,S82041C,S82042A,S82042B,S82042C | |
| | S82043A,S82043B,S82043C,S82044A,S82044B,S82044C,S82045A | |
| | S82045B,S82045C,S82046A,S82046B,S82046C,S82091A,S82091B | |
| | S82091C,S82092A,S82092B,S82092C,S82099A,S82099B,S82099C | |
| | S82101A,S82101B,S82101C,S82102A,S82102B,S82102C,S82109A | |
| | S82109B,S82109C,S82111A,S82111B,S82111C,S82112A,S82112B | |
| | S82112C,S82113A,S82113B,S82113C,S82114A,S82114B,S82114C | |
| | S82115A,S82115B,S82115C,S82116A,S82116B,S82116C,S82121A | |
| | S82121B,S82121C,S82122A,S82122B,S82122C,S82123A,S82123B | |
| | S82123C,S82124A,S82124B,S82124C,S82125A,S82125B,S82125C | |
| | S82126A,S82126B,S82126C,S82131A,S82131B,S82131C,S82132A | |
| | S82132B,S82132C,S82133A,S82133B,S82133C,S82134A,S82134B | |
| | S82134C,S82135A,S82135B,S82135C,S82136A,S82136B,S82136C | |
| | S82141A,S82141B,S82141C,S82142A,S82142B,S82142C,S82143A | |
| | S82143B,S82143C,S82144A,S82144B,S82144C,S82145A,S82145B | |
| | S82145C,S82146A,S82146B,S82146C,S82151A,S82151B,S82151C | |
| | S82152A,S82152B,S82152C,S82153A,S82153B,S82153C,S82154A | |
| | S82154B,S82154C,S82155A,S82155B,S82155C,S82156A,S82156B | |
| | S82156C,S82161A,S82162A,S82169A,S82191A,S82191B,S82191C | |
| | S82192A,S82192B,S82192C,S82199A,S82199B,S82199C,S82201A | |
| | S82201B,S82201C,S82202A,S82202B,S82202C,S82209A,S82209B | |

Continued on next page

**Supplementary Table S1:** (cont'd) A comprehensive mapping of diagnosis groups used in the experiments to their corresponding ICD-10 and ICD-9 codes.

| Diagnosis | ICD-10 | ICD-9 |
|---|---|---|
| | S82209C,S82221A,S82221B,S82221C,S82222A,S82222B,S82222C | |
| | S82223A,S82223B,S82223C,S82224A,S82224B,S82224C,S82225A | |
| | S82225B,S82225C,S82226A,S82226B,S82226C,S82231A,S82231B | |
| | S82231C,S82232A,S82232B,S82232C,S82233A,S82233B,S82233C | |
| | S82234A,S82234B,S82234C,S82235A,S82235B,S82235C,S82236A | |
| | S82236B,S82236C,S82241A,S82241B,S82241C,S82242A,S82242B | |
| | S82242C,S82243A,S82243B,S82243C,S82244A,S82244B,S82244C | |
| | S82245A,S82245B,S82245C,S82246A,S82246B,S82246C,S82251A | |
| | S82251B,S82251C,S82252A,S82252B,S82252C,S82253A,S82253B | |
| | S82253C,S82254A,S82254B,S82254C,S82255A,S82255B,S82255C | |
| | S82256A,S82256B,S82256C,S82261A,S82261B,S82261C,S82262A | |
| | S82262B,S82262C,S82263A,S82263B,S82263C,S82264A,S82264B | |
| | S82264C,S82265A,S82265B,S82265C,S82266A,S82266B,S82266C | |
| | S82291A,S82291B,S82291C,S82292A,S82292B,S82292C,S82299A | |
| | S82299B,S82299C,S82301A,S82301B,S82301C,S82302A,S82302B | |
| | S82302C,S82309A,S82309B,S82309C,S82311A,S82312A,S82319A | |
| | S82391A,S82391B,S82391C,S82392A,S82392B,S82392C,S82399A | |
| | S82399B,S82399C,S82401A,S82401B,S82401C,S82402A,S82402B | |
| | S82402C,S82409A,S82409B,S82409C,S82421A,S82421B,S82421C | |
| | S82422A,S82422B,S82422C,S82423A,S82423B,S82423C,S82424A | |
| | S82424B,S82424C,S82425A,S82425B,S82425C,S82426A,S82426B | |
| | S82426C,S82431A,S82431B,S82431C,S82432A,S82432B,S82432C | |
| | S82433A,S82433B,S82433C,S82434A,S82434B,S82434C,S82435A | |
| | S82435B,S82435C,S82436A,S82436B,S82436C,S82441A,S82441B | |
| | S82441C,S82442A,S82442B,S82442C,S82443A,S82443B,S82443C | |
| | S82444A,S82444B,S82444C,S82445A,S82445B,S82445C,S82446A | |
| | S82446B,S82446C,S82451A,S82451B,S82451C,S82452A,S82452B | |
| | S82452C,S82453A,S82453B,S82453C,S82454A,S82454B,S82454C | |
| | S82455A,S82455B,S82455C,S82456A,S82456B,S82456C,S82461A | |
| | S82461B,S82461C,S82462A,S82462B,S82462C,S82463A,S82463B | |
| | S82463C,S82464A,S82464B,S82464C,S82465A,S82465B,S82465C | |
| | S82466A,S82466B,S82466C,S82491A,S82491B,S82491C,S82492A | |
| | S82492B,S82492C,S82499A,S82499B,S82499C,S8251XA,S8251XB | |
| | S8251XC,S8252XA,S8252XB,S8252XC,S8253XA,S8253XB,S8253XC | |
| | S8254XA,S8254XB,S8254XC,S8255XA,S8255XB,S8255XC,S8256XA | |
| | S8256XB,S8256XC,S8261XA,S8261XB,S8261XC,S8262XA,S8262XB | |
| | S8262XC,S8263XA,S8263XB,S8263XC,S8264XA,S8264XB,S8264XC | |
| | S8265XA,S8265XB,S8265XC,S8266XA,S8266XB,S8266XC,S82811A | |
| | S82812A,S82819A,S82821A,S82822A,S82829A,S82831A,S82831B | |
| | S82831C,S82832A,S82832B,S82832C,S82839A,S82839B,S82839C | |

Continued on next page

**Supplementary Table S1:** (cont'd) A comprehensive mapping of diagnosis groups used in the experiments to their corresponding ICD-10 and ICD-9 codes.

| Diagnosis | ICD-10 | ICD-9 |
|---|---|---|
| | S82841A,S82841B,S82841C,S82842A,S82842B,S82842C,S82843A | |
| | S82843B,S82843C,S82844A,S82844B,S82844C,S82845A,S82845B | |
| | S82845C,S82846A,S82846B,S82846C,S82851A,S82851B,S82851C | |
| | S82852A,S82852B,S82852C,S82853A,S82853B,S82853C,S82854A | |
| | S82854B,S82854C,S82855A,S82855B,S82855C,S82856A,S82856B | |
| | S82856C,S82861A,S82861B,S82861C,S82862A,S82862B,S82862C | |
| | S82863A,S82863B,S82863C,S82864A,S82864B,S82864C,S82865A | |
| | S82865B,S82865C,S82866A,S82866B,S82866C,S82871A,S82871B | |
| | S82871C,S82872A,S82872B,S82872C,S82873A,S82873B,S82873C | |
| | S82874A,S82874B,S82874C,S82875A,S82875B,S82875C,S82876A | |
| | S82876B,S82876C,S82891A,S82891B,S82891C,S82892A,S82892B | |
| | S82892C,S82899A,S82899B,S82899C,S8290XA,S8290XB,S8290XC | |
| | S8291XA,S8291XB,S8291XC,S8292XA,S8292XB,S8292XC,S89001A | |
| | S89002A,S89009A,S89011A,S89012A,S89019A,S89021A,S89022A | |
| | S89029A,S89031A,S89032A,S89039A,S89041A,S89042A,S89049A | |
| | S89091A,S89092A,S89099A,S89101A,S89102A,S89109A,S89111A | |
| | S89112A,S89119A,S89121A,S89122A,S89129A,S89131A,S89132A | |
| | S89139A,S89141A,S89142A,S89149A,S89191A,S89192A,S89199A | |
| | S89201A,S89202A,S89209A,S89211A,S89212A,S89219A,S89221A | |
| | S89222A,S89229A,S89291A,S89292A,S89299A,S89301A,S89302A | |
| | S89309A,S89311A,S89312A,S89319A,S89321A,S89322A,S89329A | |
| | S89391A,S89392A,S89399A,S92001A,S92001B,S92002A,S92002B | |
| | S92009A,S92009B,S92011A,S92011B,S92012A,S92012B,S92013A | |
| | S92013B,S92014A,S92014B,S92015A,S92015B,S92016A,S92016B | |
| | S92021A,S92021B,S92022A,S92022B,S92023A,S92023B,S92024A | |
| | S92024B,S92025A,S92025B,S92026A,S92026B,S92031A,S92031B | |
| | S92032A,S92032B,S92033A,S92033B,S92034A,S92034B,S92035A | |
| | S92035B,S92036A,S92036B,S92041A,S92041B,S92042A,S92042B | |
| | S92043A,S92043B,S92044A,S92044B,S92045A,S92045B,S92046A | |
| | S92046B,S92051A,S92051B,S92052A,S92052B,S92053A,S92053B | |
| | S92054A,S92054B,S92055A,S92055B,S92056A,S92056B,S92061A | |
| | S92061B,S92062A,S92062B,S92063A,S92063B,S92064A,S92064B | |
| | S92065A,S92065B,S92066A,S92066B,S92101A,S92101B,S92102A | |
| | S92102B,S92109A,S92109B,S92111A,S92111B,S92112A,S92112B | |
| | S92113A,S92113B,S92114A,S92114B,S92115A,S92115B,S92116A | |
| | S92116B,S92121A,S92121B,S92122A,S92122B,S92123A,S92123B | |
| | S92124A,S92124B,S92125A,S92125B,S92126A,S92126B,S92131A | |
| | S92131B,S92132A,S92132B,S92133A,S92133B,S92134A,S92134B | |
| | S92135A,S92135B,S92136A,S92136B,S92141A,S92141B,S92142A | |
| | S92142B,S92143A,S92143B,S92144A,S92144B,S92145A,S92145B | |

Continued on next page

**Supplementary Table S1:** (cont'd) A comprehensive mapping of diagnosis groups used in the experiments to their corresponding ICD-10 and ICD-9 codes.

| Diagnosis | ICD-10 | ICD-9 |
|---|---|---|
| | S92146A,S92146B,S92151A,S92151B,S92152A,S92152B,S92153A | |
| | S92153B,S92154A,S92154B,S92155A,S92155B,S92156A,S92156B | |
| | S92191A,S92191B,S92192A,S92192B,S92199A,S92199B,S92201A | |
| | S92201B,S92202A,S92202B,S92209A,S92209B,S92211A,S92211B | |
| | S92212A,S92212B,S92213A,S92213B,S92214A,S92214B,S92215A | |
| | S92215B,S92216A,S92216B,S92221A,S92221B,S92222A,S92222B | |
| | S92223A,S92223B,S92224A,S92224B,S92225A,S92225B,S92226A | |
| | S92226B,S92231A,S92231B,S92232A,S92232B,S92233A,S92233B | |
| | S92234A,S92234B,S92235A,S92235B,S92236A,S92236B,S92241A | |
| | S92241B,S92242A,S92242B,S92243A,S92243B,S92244A,S92244B | |
| | S92245A,S92245B,S92246A,S92246B,S92251A,S92251B,S92252A | |
| | S92252B,S92253A,S92253B,S92254A,S92254B,S92255A,S92255B | |
| | S92256A,S92256B,S92301A,S92301B,S92302A,S92302B,S92309A | |
| | S92309B,S92311A,S92311B,S92312A,S92312B,S92313A,S92313B | |
| | S92314A,S92314B,S92315A,S92315B,S92316A,S92316B,S92321A | |
| | S92321B,S92322A,S92322B,S92323A,S92323B,S92324A,S92324B | |
| | S92325A,S92325B,S92326A,S92326B,S92331A,S92331B,S92332A | |
| | S92332B,S92333A,S92333B,S92334A,S92334B,S92335A,S92335B | |
| | S92336A,S92336B,S92341A,S92341B,S92342A,S92342B,S92343A | |
| | S92343B,S92344A,S92344B,S92345A,S92345B,S92346A,S92346B | |
| | S92351A,S92351B,S92352A,S92352B,S92353A,S92353B,S92354A | |
| | S92354B,S92355A,S92355B,S92356A,S92356B,S92401A,S92401B | |
| | S92402A,S92402B,S92403A,S92403B,S92404A,S92404B,S92405A | |
| | S92405B,S92406A,S92406B,S92411A,S92411B,S92412A,S92412B | |
| | S92413A,S92413B,S92414A,S92414B,S92415A,S92415B,S92416A | |
| | S92416B,S92421A,S92421B,S92422A,S92422B,S92423A,S92423B | |
| | S92424A,S92424B,S92425A,S92425B,S92426A,S92426B,S92491A | |
| | S92491B,S92492A,S92492B,S92499A,S92499B,S92501A,S92501B | |
| | S92502A,S92502B,S92503A,S92503B,S92504A,S92504B,S92505A | |
| | S92505B,S92506A,S92506B,S92511A,S92511B,S92512A,S92512B | |
| | S92513A,S92513B,S92514A,S92514B,S92515A,S92515B,S92516A | |
| | S92516B,S92521A,S92521B,S92522A,S92522B,S92523A,S92523B | |
| | S92524A,S92524B,S92525A,S92525B,S92526A,S92526B,S92531A | |
| | S92531B,S92532A,S92532B,S92533A,S92533B,S92534A,S92534B | |
| | S92535A,S92535B,S92536A,S92536B,S92591A,S92591B,S92592A | |
| | S92592B,S92599A,S92599B,S92811A,S92811B,S92812A,S92812B | |
| | S92819A,S92819B,S92901A,S92901B,S92902A,S92902B,S92909A | |
| | S92909B,S92911A,S92911B,S92912A,S92912B,S92919A,S92919B | |
| | S99001A,S99001B,S99002A,S99002B,S99009A,S99009B,S99011A | |
| | S99011B,S99012A,S99012B,S99019A,S99019B,S99021A,S99021B | |

Continued on next page

**Supplementary Table S1:** (cont'd) A comprehensive mapping of diagnosis groups used in the experiments to their corresponding ICD-10 and ICD-9 codes.

| Diagnosis | ICD-10 | ICD-9 |
|---|---|---|
| | S99022A,S99022B,S99029A,S99029B,S99031A,S99031B,S99032A | |
| | S99032B,S99039A,S99039B,S99041A,S99041B,S99042A,S99042B | |
| | S99049A,S99049B,S99091A,S99091B,S99092A,S99092B,S99099A | |
| | S99099B,S99101A,S99101B,S99102A,S99102B,S99109A,S99109B | |
| | S99111A,S99111B,S99112A,S99112B,S99119A,S99119B,S99121A | |
| | S99121B,S99122A,S99122B,S99129A,S99129B,S99131A,S99131B | |
| | S99132A,S99132B,S99139A,S99139B,S99141A,S99141B,S99142A | |
| | S99142B,S99149A,S99149B,S99191A,S99191B,S99192A,S99192B | |
| | S99199A,S99199B,S99201A,S99201B,S99202A,S99202B,S99209A | |
| | S99209B,S99211A,S99211B,S99212A,S99212B,S99219A,S99219B | |
| | S99221A,S99221B,S99222A,S99222B,S99229A,S99229B,S99231A | |
| | S99231B,S99232A,S99232B,S99239A,S99239B,S99241A,S99241B | |
| | S99242A,S99242B,S99249A,S99249B,S99291A,S99291B,S99292A | |
| | S99292B,S99299A,S99299B,T84042A,T84043A,T84048A,T84049A | |
| Fracture of the neck | M9701XA,M9702XA,S72001A,S72001B,S72001C,S72002A,S72002B | 82000,82001,82002,82003,82009,82010,82011,82012,82013,82019 |
| of the femur (hip), | S72002C,S72009A,S72009B,S72009C,S72011A,S72011B,S72011C | 82020,82021,82022,82030,82031,82032,8208,8209,99644 |
| initial encounter | S72012A,S72012B,S72012C,S72019A,S72019B,S72019C,S72021A | |
| | S72021B,S72021C,S72022A,S72022B,S72022C,S72023A,S72023B | |
| | S72023C,S72024A,S72024B,S72024C,S72025A,S72025B,S72025C | |
| | S72026A,S72026B,S72026C,S72031A,S72031B,S72031C,S72032A | |
| | S72032B,S72032C,S72033A,S72033B,S72033C,S72034A,S72034B | |
| | S72034C,S72035A,S72035B,S72035C,S72036A,S72036B,S72036C | |
| | S72041A,S72041B,S72041C,S72042A,S72042B,S72042C,S72043A | |
| | S72043B,S72043C,S72044A,S72044B,S72044C,S72045A,S72045B | |
| | S72045C,S72046A,S72046B,S72046C,S72051A,S72051B,S72051C | |
| | S72052A,S72052B,S72052C,S72059A,S72059B,S72059C,S72061A | |
| | S72061B,S72061C,S72062A,S72062B,S72062C,S72063A,S72063B | |
| | S72063C,S72064A,S72064B,S72064C,S72065A,S72065B,S72065C | |
| | S72066A,S72066B,S72066C,S72091A,S72091B,S72091C,S72092A | |
| | S72092B,S72092C,S72099A,S72099B,S72099C,S72101A,S72101B | |
| | S72101C,S72102A,S72102B,S72102C,S72109A,S72109B,S72109C | |
| | S72111A,S72111B,S72111C,S72112A,S72112B,S72112C,S72113A | |
| | S72113B,S72113C,S72114A,S72114B,S72114C,S72115A,S72115B | |
| | S72115C,S72116A,S72116B,S72116C,S72121A,S72121B,S72121C | |
| | S72122A,S72122B,S72122C,S72123A,S72123B,S72123C,S72124A | |
| | S72124B,S72124C,S72125A,S72125B,S72125C,S72126A,S72126B | |
| | S72126C,S72131A,S72131B,S72131C,S72132A,S72132B,S72132C | |
| | S72133A,S72133B,S72133C,S72134A,S72134B,S72134C,S72135A | |
| | S72135B,S72135C,S72136A,S72136B,S72136C,S72141A,S72141B | |

Continued on next page

**Supplementary Table S1:** (cont'd) A comprehensive mapping of diagnosis groups used in the experiments to their corresponding ICD-10 and ICD-9 codes.

| Diagnosis | ICD-10 | ICD-9 |
|---|---|---|
| | S72141C,S72142A,S72142B,S72142C,S72143A,S72143B,S72143C S72144A,S72144B,S72144C,S72145A,S72145B,S72145C,S72146A S72146B,S72146C,S7221XA,S7221XB,S7221XC,S7222XA,S7222XB S7222XC,S7223XA,S7223XB,S7223XC,S7224XA,S7224XB,S7224XC S7225XA,S7225XB,S7225XC,S7226XA,S7226XB,S7226XC,T84040A T84041A | |
| Fracture of the neck of the femur (hip), subsequent encounter | M9701XD,M9702XD,S72001D,S72001E,S72001F,S72001G,S72001H S72001J,S72001K,S72001M,S72001N,S72001P,S72001Q,S72001R S72002D,S72002E,S72002F,S72002G,S72002H,S72002J,S72002K S72002M,S72002N,S72002P,S72002Q,S72002R,S72009D,S72009E S72009F,S72009G,S72009H,S72009J,S72009K,S72009M,S72009N S72009P,S72009Q,S72009R,S72011D,S72011E,S72011F,S72011G S72011H,S72011J,S72011K,S72011M,S72011N,S72011P,S72011Q S72011R,S72012D,S72012E,S72012F,S72012G,S72012H,S72012J S72012K,S72012M,S72012N,S72012P,S72012Q,S72012R,S72019D S72019E,S72019F,S72019G,S72019H,S72019J,S72019K,S72019M S72019N,S72019P,S72019Q,S72019R,S72021D,S72021E,S72021F S72021G,S72021H,S72021J,S72021K,S72021M,S72021N,S72021P S72021Q,S72021R,S72022D,S72022E,S72022F,S72022G,S72022H S72022J,S72022K,S72022M,S72022N,S72022P,S72022Q,S72022R S72023D,S72023E,S72023F,S72023G,S72023H,S72023J,S72023K S72023M,S72023N,S72023P,S72023Q,S72023R,S72024D,S72024E S72024F,S72024G,S72024H,S72024J,S72024K,S72024M,S72024N S72024P,S72024Q,S72024R,S72025D,S72025E,S72025F,S72025G S72025H,S72025J,S72025K,S72025M,S72025N,S72025P,S72025Q S72025R,S72026D,S72026E,S72026F,S72026G,S72026H,S72026J S72026K,S72026M,S72026N,S72026P,S72026Q,S72026R,S72031D S72031E,S72031F,S72031G,S72031H,S72031J,S72031K,S72031M S72031N,S72031P,S72031Q,S72031R,S72032D,S72032E,S72032F S72032G,S72032H,S72032J,S72032K,S72032M,S72032N,S72032P S72032Q,S72032R,S72033D,S72033E,S72033F,S72033G,S72033H S72033J,S72033K,S72033M,S72033N,S72033P,S72033Q,S72033R S72034D,S72034E,S72034F,S72034G,S72034H,S72034J,S72034K S72034M,S72034N,S72034P,S72034Q,S72034R,S72035D,S72035E S72035F,S72035G,S72035H,S72035J,S72035K,S72035M,S72035N S72035P,S72035Q,S72035R,S72036D,S72036E,S72036F,S72036G S72036H,S72036J,S72036K,S72036M,S72036N,S72036P,S72036Q S72036R,S72041D,S72041E,S72041F,S72041G,S72041H,S72041J S72041K,S72041M,S72041N,S72041P,S72041Q,S72041R,S72042D | 73381,73382,V5413,V5415,V5423 |

Continued on next page

**Supplementary Table S1:** (cont'd) A comprehensive mapping of diagnosis groups used in the experiments to their corresponding ICD-10 and ICD-9 codes.

| Diagnosis | ICD-10 | ICD-9 |
|---|---|---|
| | S72042E,S72042F,S72042G,S72042H,S72042J,S72042K,S72042M | |
| | S72042N,S72042P,S72042Q,S72042R,S72043D,S72043E,S72043F | |
| | S72043G,S72043H,S72043J,S72043K,S72043M,S72043N,S72043P | |
| | S72043Q,S72043R,S72044D,S72044E,S72044F,S72044G,S72044H | |
| | S72044J,S72044K,S72044M,S72044N,S72044P,S72044Q,S72044R | |
| | S72045D,S72045E,S72045F,S72045G,S72045H,S72045J,S72045K | |
| | S72045M,S72045N,S72045P,S72045Q,S72045R,S72046D,S72046E | |
| | S72046F,S72046G,S72046H,S72046J,S72046K,S72046M,S72046N | |
| | S72046P,S72046Q,S72046R,S72051D,S72051E,S72051F,S72051G | |
| | S72051H,S72051J,S72051K,S72051M,S72051N,S72051P,S72051Q | |
| | S72051R,S72052D,S72052E,S72052F,S72052G,S72052H,S72052J | |
| | S72052K,S72052M,S72052N,S72052P,S72052Q,S72052R,S72059D | |
| | S72059E,S72059F,S72059G,S72059H,S72059J,S72059K,S72059M | |
| | S72059N,S72059P,S72059Q,S72059R,S72061D,S72061E,S72061F | |
| | S72061G,S72061H,S72061J,S72061K,S72061M,S72061N,S72061P | |
| | S72061Q,S72061R,S72062D,S72062E,S72062F,S72062G,S72062H | |
| | S72062J,S72062K,S72062M,S72062N,S72062P,S72062Q,S72062R | |
| | S72063D,S72063E,S72063F,S72063G,S72063H,S72063J,S72063K | |
| | S72063M,S72063N,S72063P,S72063Q,S72063R,S72064D,S72064E | |
| | S72064F,S72064G,S72064H,S72064J,S72064K,S72064M,S72064N | |
| | S72064P,S72064Q,S72064R,S72065D,S72065E,S72065F,S72065G | |
| | S72065H,S72065J,S72065K,S72065M,S72065N,S72065P,S72065Q | |
| | S72065R,S72066D,S72066E,S72066F,S72066G,S72066H,S72066J | |
| | S72066K,S72066M,S72066N,S72066P,S72066Q,S72066R,S72091D | |
| | S72091E,S72091F,S72091G,S72091H,S72091J,S72091K,S72091M | |
| | S72091N,S72091P,S72091Q,S72091R,S72092D,S72092E,S72092F | |
| | S72092G,S72092H,S72092J,S72092K,S72092M,S72092N,S72092P | |
| | S72092Q,S72092R,S72099D,S72099E,S72099F,S72099G,S72099H | |
| | S72099J,S72099K,S72099M,S72099N,S72099P,S72099Q,S72099R | |
| | S72101D,S72101E,S72101F,S72101G,S72101H,S72101J,S72101K | |
| | S72101M,S72101N,S72101P,S72101Q,S72101R,S72102D,S72102E | |
| | S72102F,S72102G,S72102H,S72102J,S72102K,S72102M,S72102N | |
| | S72102P,S72102Q,S72102R,S72109D,S72109E,S72109F,S72109G | |
| | S72109H,S72109J,S72109K,S72109M,S72109N,S72109P,S72109Q | |
| | S72109R,S72111D,S72111E,S72111F,S72111G,S72111H,S72111J | |
| | S72111K,S72111M,S72111N,S72111P,S72111Q,S72111R,S72112D | |
| | S72112E,S72112F,S72112G,S72112H,S72112J,S72112K,S72112M | |
| | S72112N,S72112P,S72112Q,S72112R,S72113D,S72113E,S72113F | |
| | S72113G,S72113H,S72113J,S72113K,S72113M,S72113N,S72113P | |
| | S72113Q,S72113R,S72114D,S72114E,S72114F,S72114G,S72114H | |

Continued on next page

**Supplementary Table S1:** (cont'd) A comprehensive mapping of diagnosis groups used in the experiments to their corresponding ICD-10 and ICD-9 codes.

| Diagnosis | ICD-10 | ICD-9 |
|---|---|---|
| | S72114J,S72114K,S72114M,S72114N,S72114P,S72114Q,S72114R | |
| | S72115D,S72115E,S72115F,S72115G,S72115H,S72115J,S72115K | |
| | S72115M,S72115N,S72115P,S72115Q,S72115R,S72116D,S72116E | |
| | S72116F,S72116G,S72116H,S72116J,S72116K,S72116M,S72116N | |
| | S72116P,S72116Q,S72116R,S72121D,S72121E,S72121F,S72121G | |
| | S72121H,S72121J,S72121K,S72121M,S72121N,S72121P,S72121Q | |
| | S72121R,S72122D,S72122E,S72122F,S72122G,S72122H,S72122J | |
| | S72122K,S72122M,S72122N,S72122P,S72122Q,S72122R,S72123D | |
| | S72123E,S72123F,S72123G,S72123H,S72123J,S72123K,S72123M | |
| | S72123N,S72123P,S72123Q,S72123R,S72124D,S72124E,S72124F | |
| | S72124G,S72124H,S72124J,S72124K,S72124M,S72124N,S72124P | |
| | S72124Q,S72124R,S72125D,S72125E,S72125F,S72125G,S72125H | |
| | S72125J,S72125K,S72125M,S72125N,S72125P,S72125Q,S72125R | |
| | S72126D,S72126E,S72126F,S72126G,S72126H,S72126J,S72126K | |
| | S72126M,S72126N,S72126P,S72126Q,S72126R,S72131D,S72131E | |
| | S72131F,S72131G,S72131H,S72131J,S72131K,S72131M,S72131N | |
| | S72131P,S72131Q,S72131R,S72132D,S72132E,S72132F,S72132G | |
| | S72132H,S72132J,S72132K,S72132M,S72132N,S72132P,S72132Q | |
| | S72132R,S72133D,S72133E,S72133F,S72133G,S72133H,S72133J | |
| | S72133K,S72133M,S72133N,S72133P,S72133Q,S72133R,S72134D | |
| | S72134E,S72134F,S72134G,S72134H,S72134J,S72134K,S72134M | |
| | S72134N,S72134P,S72134Q,S72134R,S72135D,S72135E,S72135F | |
| | S72135G,S72135H,S72135J,S72135K,S72135M,S72135N,S72135P | |
| | S72135Q,S72135R,S72136D,S72136E,S72136F,S72136G,S72136H | |
| | S72136J,S72136K,S72136M,S72136N,S72136P,S72136Q,S72136R | |
| | S72141D,S72141E,S72141F,S72141G,S72141H,S72141J,S72141K | |
| | S72141M,S72141N,S72141P,S72141Q,S72141R,S72142D,S72142E | |
| | S72142F,S72142G,S72142H,S72142J,S72142K,S72142M,S72142N | |
| | S72142P,S72142Q,S72142R,S72143D,S72143E,S72143F,S72143G | |
| | S72143H,S72143J,S72143K,S72143M,S72143N,S72143P,S72143Q | |
| | S72143R,S72144D,S72144E,S72144F,S72144G,S72144H,S72144J | |
| | S72144K,S72144M,S72144N,S72144P,S72144Q,S72144R,S72145D | |
| | S72145E,S72145F,S72145G,S72145H,S72145J,S72145K,S72145M | |
| | S72145N,S72145P,S72145Q,S72145R,S72146D,S72146E,S72146F | |
| | S72146G,S72146H,S72146J,S72146K,S72146M,S72146N,S72146P | |
| | S72146Q,S72146R,S7221XD,S7221XE,S7221XF,S7221XG,S7221XH | |
| | S7221XJ,S7221XK,S7221XM,S7221XN,S7221XP,S7221XQ,S7221XR | |
| | S7222XD,S7222XE,S7222XF,S7222XG,S7222XH,S7222XJ,S7222XK | |
| | S7222XM,S7222XN,S7222XP,S7222XQ,S7222XR,S7223XD,S7223XE | |
| | S7223XF,S7223XG,S7223XH,S7223XJ,S7223XK,S7223XM,S7223XN | |

Continued on next page

**Supplementary Table S1:** (cont'd) A comprehensive mapping of diagnosis groups used in the experiments to their corresponding ICD-10 and ICD-9 codes.

| Diagnosis | ICD-10 | ICD-9 |
|---|---|---|
| | S7223XP,S7223XQ,S7223XR,S7224XD,S7224XE,S7224XF,S7224XG | |
| | S7224XH,S7224XJ,S7224XK,S7224XM,S7224XN,S7224XP,S7224XQ | |
| | S7224XR,S7225XD,S7225XE,S7225XF,S7225XG,S7225XH,S7225XJ | |
| | S7225XK,S7225XM,S7225XN,S7225XP,S7225XQ,S7225XR,S7226XD | |
| | S7226XE,S7226XF,S7226XG,S7226XH,S7226XJ,S7226XK,S7226XM | |
| | S7226XN,S7226XP,S7226XQ,S7226XR,T84040D,T84041D | |
| Fracture of the spine | S12000A,S12000B,S12001A,S12001B,S1201XA,S1201XB,S1202XA | 80500,80501,80502,80503,80504,80505,80506,80507,80511,80512 |
| and back, initial | S1202XB,S12030A,S12030B,S12031A,S12031B,S12040A,S12040B | 80513,80514,80515,80516,80517,8052,8053,8054,8055,8056,8057 |
| encounter | S12041A,S12041B,S12090A,S12090B,S12091A,S12091B,S12100A | 8075,80849,80859,8088,8089 |
| | S12100B,S12101A,S12101B,S12110A,S12110B,S12111A,S12111B | |
| | S12112A,S12112B,S12120A,S12120B,S12121A,S12121B,S12130A | |
| | S12130B,S12131A,S12131B,S1214XA,S1214XB,S12150A,S12150B | |
| | S12151A,S12151B,S12190A,S12190B,S12191A,S12191B,S12200A | |
| | S12200B,S12201A,S12201B,S12230A,S12230B,S12231A,S12231B | |
| | S1224XA,S1224XB,S12250A,S12250B,S12251A,S12251B,S12290A | |
| | S12290B,S12291A,S12291B,S12300A,S12300B,S12301A,S12301B | |
| | S12330A,S12330B,S12331A,S12331B,S1234XA,S1234XB,S12350A | |
| | S12350B,S12351A,S12351B,S12390A,S12390B,S12391A,S12391B | |
| | S12400A,S12400B,S12401A,S12401B,S12430A,S12430B,S12431A | |
| | S12431B,S1244XA,S1244XB,S12450A,S12450B,S12451A,S12451B | |
| | S12490A,S12490B,S12491A,S12491B,S12500A,S12500B,S12501A | |
| | S12501B,S12530A,S12530B,S12531A,S12531B,S1254XA,S1254XB | |
| | S12550A,S12550B,S12551A,S12551B,S12590A,S12590B,S12591A | |
| | S12591B,S12600A,S12600B,S12601A,S12601B,S12630A,S12630B | |
| | S12631A,S12631B,S1264XA,S1264XB,S12650A,S12650B,S12651A | |
| | S12651B,S12690A,S12690B,S12691A,S12691B,S128XXA,S129XXA | |
| | S22000A,S22000B,S22001A,S22001B,S22002A,S22002B,S22008A | |
| | S22008B,S22009A,S22009B,S22010A,S22010B,S22011A,S22011B | |
| | S22012A,S22012B,S22018A,S22018B,S22019A,S22019B,S22020A | |
| | S22020B,S22021A,S22021B,S22022A,S22022B,S22028A,S22028B | |
| | S22029A,S22029B,S22030A,S22030B,S22031A,S22031B,S22032A | |
| | S22032B,S22038A,S22038B,S22039A,S22039B,S22040A,S22040B | |
| | S22041A,S22041B,S22042A,S22042B,S22048A,S22048B,S22049A | |
| | S22049B,S22050A,S22050B,S22051A,S22051B,S22052A,S22052B | |
| | S22058A,S22058B,S22059A,S22059B,S22060A,S22060B,S22061A | |
| | S22061B,S22062A,S22062B,S22068A,S22068B,S22069A,S22069B | |
| | S22070A,S22070B,S22071A,S22071B,S22072A,S22072B,S22078A | |
| | S22078B,S22079A,S22079B,S22080A,S22080B,S22081A,S22081B | |
| | S22082A,S22082B,S22088A,S22088B,S22089A,S22089B,S32000A | |

Continued on next page

**Supplementary Table S1:** (cont'd) A comprehensive mapping of diagnosis groups used in the experiments to their corresponding ICD-10 and ICD-9 codes.

| Diagnosis | ICD-10 | ICD-9 |
|---|---|---|
| | S32000B,S32001A,S32001B,S32002A,S32002B,S32008A,S32008B | |
| | S32009A,S32009B,S32010A,S32010B,S32011A,S32011B,S32012A | |
| | S32012B,S32018A,S32018B,S32019A,S32019B,S32020A,S32020B | |
| | S32021A,S32021B,S32022A,S32022B,S32028A,S32028B,S32029A | |
| | S32029B,S32030A,S32030B,S32031A,S32031B,S32032A,S32032B | |
| | S32038A,S32038B,S32039A,S32039B,S32040A,S32040B,S32041A | |
| | S32041B,S32042A,S32042B,S32048A,S32048B,S32049A,S32049B | |
| | S32050A,S32050B,S32051A,S32051B,S32052A,S32052B,S32058A | |
| | S32058B,S32059A,S32059B,S3210XA,S3210XB,S32110A,S32110B | |
| | S32111A,S32111B,S32112A,S32112B,S32119A,S32119B,S32120A | |
| | S32120B,S32121A,S32121B,S32122A,S32122B,S32129A,S32129B | |
| | S32130A,S32130B,S32131A,S32131B,S32132A,S32132B,S32139A | |
| | S32139B,S3214XA,S3214XB,S3215XA,S3215XB,S3216XA,S3216XB | |
| | S3217XA,S3217XB,S3219XA,S3219XB,S322XXA,S322XXB,S329XXA | |
| | S329XXB | |
| Fracture of the spine | S12000D,S12000G,S12000K,S12001D,S12001G,S12001K,S1201XD | 73382,V5417,V5419 |
| and back, subsequent | S1201XG,S1201XK,S1202XD,S1202XG,S1202XK,S12030D,S12030G | |
| encounter | S12030K,S12031D,S12031G,S12031K,S12040D,S12040G,S12040K | |
| | S12041D,S12041G,S12041K,S12090D,S12090G,S12090K,S12091D | |
| | S12091G,S12091K,S12100D,S12100G,S12100K,S12101D,S12101G | |
| | S12101K,S12110D,S12110G,S12110K,S12111D,S12111G,S12111K | |
| | S12112D,S12112G,S12112K,S12120D,S12120G,S12120K,S12121D | |
| | S12121G,S12121K,S12130D,S12130G,S12130K,S12131D,S12131G | |
| | S12131K,S1214XD,S1214XG,S1214XK,S12150D,S12150G,S12150K | |
| | S12151D,S12151G,S12151K,S12190D,S12190G,S12190K,S12191D | |
| | S12191G,S12191K,S12200D,S12200G,S12200K,S12201D,S12201G | |
| | S12201K,S12230D,S12230G,S12230K,S12231D,S12231G,S12231K | |
| | S1224XD,S1224XG,S1224XK,S12250D,S12250G,S12250K,S12251D | |
| | S12251G,S12251K,S12290D,S12290G,S12290K,S12291D,S12291G | |
| | S12291K,S12300D,S12300G,S12300K,S12301D,S12301G,S12301K | |
| | S12330D,S12330G,S12330K,S12331D,S12331G,S12331K,S1234XD | |
| | S1234XG,S1234XK,S12350D,S12350G,S12350K,S12351D,S12351G | |
| | S12351K,S12390D,S12390G,S12390K,S12391D,S12391G,S12391K | |
| | S12400D,S12400G,S12400K,S12401D,S12401G,S12401K,S12430D | |
| | S12430G,S12430K,S12431D,S12431G,S12431K,S1244XD,S1244XG | |
| | S1244XK,S12450D,S12450G,S12450K,S12451D,S12451G,S12451K | |
| | S12490D,S12490G,S12490K,S12491D,S12491G,S12491K,S12500D | |
| | S12500G,S12500K,S12501D,S12501G,S12501K,S12530D,S12530G | |
| | S12530K,S12531D,S12531G,S12531K,S1254XD,S1254XG,S1254XK | |

Continued on next page

**Supplementary Table S1:** (cont'd) A comprehensive mapping of diagnosis groups used in the experiments to their corresponding ICD-10 and ICD-9 codes.

| Diagnosis | ICD-10 | ICD-9 |
|---|---|---|
| | S12550D,S12550G,S12550K,S12551D,S12551G,S12551K,S12590D | |
| | S12590G,S12590K,S12591D,S12591G,S12591K,S12600D,S12600G | |
| | S12600K,S12601D,S12601G,S12601K,S12630D,S12630G,S12630K | |
| | S12631D,S12631G,S12631K,S1264XD,S1264XG,S1264XK,S12650D | |
| | S12650G,S12650K,S12651D,S12651G,S12651K,S12690D,S12690G | |
| | S12690K,S12691D,S12691G,S12691K,S128XXD,S129XXD,S22000D | |
| | S22000G,S22000K,S22001D,S22001G,S22001K,S22002D,S22002G | |
| | S22002K,S22008D,S22008G,S22008K,S22009D,S22009G,S22009K | |
| | S22010D,S22010G,S22010K,S22011D,S22011G,S22011K,S22012D | |
| | S22012G,S22012K,S22018D,S22018G,S22018K,S22019D,S22019G | |
| | S22019K,S22020D,S22020G,S22020K,S22021D,S22021G,S22021K | |
| | S22022D,S22022G,S22022K,S22028D,S22028G,S22028K,S22029D | |
| | S22029G,S22029K,S22030D,S22030G,S22030K,S22031D,S22031G | |
| | S22031K,S22032D,S22032G,S22032K,S22038D,S22038G,S22038K | |
| | S22039D,S22039G,S22039K,S22040D,S22040G,S22040K,S22041D | |
| | S22041G,S22041K,S22042D,S22042G,S22042K,S22048D,S22048G | |
| | S22048K,S22049D,S22049G,S22049K,S22050D,S22050G,S22050K | |
| | S22051D,S22051G,S22051K,S22052D,S22052G,S22052K,S22058D | |
| | S22058G,S22058K,S22059D,S22059G,S22059K,S22060D,S22060G | |
| | S22060K,S22061D,S22061G,S22061K,S22062D,S22062G,S22062K | |
| | S22068D,S22068G,S22068K,S22069D,S22069G,S22069K,S22070D | |
| | S22070G,S22070K,S22071D,S22071G,S22071K,S22072D,S22072G | |
| | S22072K,S22078D,S22078G,S22078K,S22079D,S22079G,S22079K | |
| | S22080D,S22080G,S22080K,S22081D,S22081G,S22081K,S22082D | |
| | S22082G,S22082K,S22088D,S22088G,S22088K,S22089D,S22089G | |
| | S22089K,S32000D,S32000G,S32000K,S32001D,S32001G,S32001K | |
| | S32002D,S32002G,S32002K,S32008D,S32008G,S32008K,S32009D | |
| | S32009G,S32009K,S32010D,S32010G,S32010K,S32011D,S32011G | |
| | S32011K,S32012D,S32012G,S32012K,S32018D,S32018G,S32018K | |
| | S32019D,S32019G,S32019K,S32020D,S32020G,S32020K,S32021D | |
| | S32021G,S32021K,S32022D,S32022G,S32022K,S32028D,S32028G | |
| | S32028K,S32029D,S32029G,S32029K,S32030D,S32030G,S32030K | |
| | S32031D,S32031G,S32031K,S32032D,S32032G,S32032K,S32038D | |
| | S32038G,S32038K,S32039D,S32039G,S32039K,S32040D,S32040G | |
| | S32040K,S32041D,S32041G,S32041K,S32042D,S32042G,S32042K | |
| | S32048D,S32048G,S32048K,S32049D,S32049G,S32049K,S32050D | |
| | S32050G,S32050K,S32051D,S32051G,S32051K,S32052D,S32052G | |
| | S32052K,S32058D,S32058G,S32058K,S32059D,S32059G,S32059K | |
| | S3210XD,S3210XG,S3210XK,S32110D,S32110G,S32110K,S32111D | |
| | S32111G,S32111K,S32112D,S32112G,S32112K,S32119D,S32119G | |

Continued on next page

**Supplementary Table S1:** (cont'd) A comprehensive mapping of diagnosis groups used in the experiments to their corresponding ICD-10 and ICD-9 codes.

| Diagnosis | ICD-10 | ICD-9 |
|---|---|---|
| | S32119K,S32120D,S32120G,S32120K,S32121D,S32121G,S32121K | |
| | S32122D,S32122G,S32122K,S32129D,S32129G,S32129K,S32130D | |
| | S32130G,S32130K,S32131D,S32131G,S32131K,S32132D,S32132G | |
| | S32132K,S32139D,S32139G,S32139K,S3214XD,S3214XG,S3214XK | |
| | S3215XD,S3215XG,S3215XK,S3216XD,S3216XG,S3216XK,S3217XD | |
| | S3217XG,S3217XK,S3219XD,S3219XG,S3219XK,S322XXD,S322XXG | |
| | S322XXK,S329XXD,S329XXG,S329XXK | |
| Fracture of the upper | M9731XA,M9732XA,M9741XA,M9742XA,S42001A,S42001B,S42002A | 81000,81001,81002,81003,81010,81011,81012,81013,81100,81101 |
| limb, initial | S42002B,S42009A,S42009B,S42011A,S42011B,S42012A,S42012B | 81102,81103,81109,81110,81111,81112,81113,81119,81200,81201 |
| encounter | S42013A,S42013B,S42014A,S42014B,S42015A,S42015B,S42016A | 81202,81203,81209,81210,81211,81212,81213,81219,81220,81221 |
| | S42016B,S42017A,S42017B,S42018A,S42018B,S42019A,S42019B | 81230,81231,81240,81241,81242,81243,81244,81249,81250,81251 |
| | S42021A,S42021B,S42022A,S42022B,S42023A,S42023B,S42024A | 81252,81253,81254,81259,81301,81302,81303,81304,81305,81306 |
| | S42024B,S42025A,S42025B,S42026A,S42026B,S42031A,S42031B | 81307,81311,81312,81313,81314,81315,81316,81317,81321,81322 |
| | S42032A,S42032B,S42033A,S42033B,S42034A,S42034B,S42035A | 81331,81332,81341,81342,81343,81345,81346,81351,81352,81353 |
| | S42035B,S42036A,S42036B,S42101A,S42101B,S42102A,S42102B | 81380,81390,81400,81401,81402,81403,81404,81405,81406,81407 |
| | S42109A,S42109B,S42111A,S42111B,S42112A,S42112B,S42113A | 81408,81409,81410,81411,81412,81413,81414,81415,81416,81417 |
| | S42113B,S42114A,S42114B,S42115A,S42115B,S42116A,S42116B | 81418,81419,81500,81501,81502,81503,81504,81509,81510,81511 |
| | S42121A,S42121B,S42122A,S42122B,S42123A,S42123B,S42124A | 81512,81513,81514,81519,81600,81601,81602,81603,81610,81611 |
| | S42124B,S42125A,S42125B,S42126A,S42126B,S42131A,S42131B | 81612,81613,8170,8181 |
| | S42132A,S42132B,S42133A,S42133B,S42134A,S42134B,S42135A | |
| | S42135B,S42136A,S42136B,S42141A,S42141B,S42142A,S42142B | |
| | S42143A,S42143B,S42144A,S42144B,S42145A,S42145B,S42146A | |
| | S42146B,S42151A,S42151B,S42152A,S42152B,S42153A,S42153B | |
| | S42154A,S42154B,S42155A,S42155B,S42156A,S42156B,S42191A | |
| | S42191B,S42192A,S42192B,S42199A,S42199B,S42201A,S42201B | |
| | S42202A,S42202B,S42209A,S42209B,S42211A,S42211B,S42212A | |
| | S42212B,S42213A,S42213B,S42214A,S42214B,S42215A,S42215B | |
| | S42216A,S42216B,S42221A,S42221B,S42222A,S42222B,S42223A | |
| | S42223B,S42224A,S42224B,S42225A,S42225B,S42226A,S42226B | |
| | S42231A,S42231B,S42232A,S42232B,S42239A,S42239B,S42241A | |
| | S42241B,S42242A,S42242B,S42249A,S42249B,S42251A,S42251B | |
| | S42252A,S42252B,S42253A,S42253B,S42254A,S42254B,S42255A | |
| | S42255B,S42256A,S42256B,S42261A,S42261B,S42262A,S42262B | |
| | S42263A,S42263B,S42264A,S42264B,S42265A,S42265B,S42266A | |
| | S42266B,S42271A,S42272A,S42279A,S42291A,S42291B,S42292A | |
| | S42292B,S42293A,S42293B,S42294A,S42294B,S42295A,S42295B | |
| | S42296A,S42296B,S42301A,S42301B,S42302A,S42302B,S42309A | |
| | S42309B,S42311A,S42312A,S42319A,S42321A,S42321B,S42322A | |
| | S42322B,S42323A,S42323B,S42324A,S42324B,S42325A,S42325B | |

Continued on next page

**Supplementary Table S1:** (cont'd) A comprehensive mapping of diagnosis groups used in the experiments to their corresponding ICD-10 and ICD-9 codes.

| Diagnosis | ICD-10 | ICD-9 |
|---|---|---|
| | S42326A,S42326B,S42331A,S42331B,S42332A,S42332B,S42333A | |
| | S42333B,S42334A,S42334B,S42335A,S42335B,S42336A,S42336B | |
| | S42341A,S42341B,S42342A,S42342B,S42343A,S42343B,S42344A | |
| | S42344B,S42345A,S42345B,S42346A,S42346B,S42351A,S42351B | |
| | S42352A,S42352B,S42353A,S42353B,S42354A,S42354B,S42355A | |
| | S42355B,S42356A,S42356B,S42361A,S42361B,S42362A,S42362B | |
| | S42363A,S42363B,S42364A,S42364B,S42365A,S42365B,S42366A | |
| | S42366B,S42391A,S42391B,S42392A,S42392B,S42399A,S42399B | |
| | S42401A,S42401B,S42402A,S42402B,S42409A,S42409B,S42411A | |
| | S42411B,S42412A,S42412B,S42413A,S42413B,S42414A,S42414B | |
| | S42415A,S42415B,S42416A,S42416B,S42421A,S42421B,S42422A | |
| | S42422B,S42423A,S42423B,S42424A,S42424B,S42425A,S42425B | |
| | S42426A,S42426B,S42431A,S42431B,S42432A,S42432B,S42433A | |
| | S42433B,S42434A,S42434B,S42435A,S42435B,S42436A,S42436B | |
| | S42441A,S42441B,S42442A,S42442B,S42443A,S42443B,S42444A | |
| | S42444B,S42445A,S42445B,S42446A,S42446B,S42447A,S42447B | |
| | S42448A,S42448B,S42449A,S42449B,S42451A,S42451B,S42452A | |
| | S42452B,S42453A,S42453B,S42454A,S42454B,S42455A,S42455B | |
| | S42456A,S42456B,S42461A,S42461B,S42462A,S42462B,S42463A | |
| | S42463B,S42464A,S42464B,S42465A,S42465B,S42466A,S42466B | |
| | S42471A,S42471B,S42472A,S42472B,S42473A,S42473B,S42474A | |
| | S42474B,S42475A,S42475B,S42476A,S42476B,S42481A,S42482A | |
| | S42489A,S42491A,S42491B,S42492A,S42492B,S42493A,S42493B | |
| | S42494A,S42494B,S42495A,S42495B,S42496A,S42496B,S4290XA | |
| | S4290XB,S4291XA,S4291XB,S4292XA,S4292XB,S49001A,S49002A | |
| | S49009A,S49011A,S49012A,S49019A,S49021A,S49022A,S49029A | |
| | S49031A,S49032A,S49039A,S49041A,S49042A,S49049A,S49091A | |
| | S49092A,S49099A,S49101A,S49102A,S49109A,S49111A,S49112A | |
| | S49119A,S49121A,S49122A,S49129A,S49131A,S49132A,S49139A | |
| | S49141A,S49142A,S49149A,S49191A,S49192A,S49199A,S52001A | |
| | S52001B,S52001C,S52002A,S52002B,S52002C,S52009A,S52009B | |
| | S52009C,S52011A,S52012A,S52019A,S52021A,S52021B,S52021C | |
| | S52022A,S52022B,S52022C,S52023A,S52023B,S52023C,S52024A | |
| | S52024B,S52024C,S52025A,S52025B,S52025C,S52026A,S52026B | |
| | S52026C,S52031A,S52031B,S52031C,S52032A,S52032B,S52032C | |
| | S52033A,S52033B,S52033C,S52034A,S52034B,S52034C,S52035A | |
| | S52035B,S52035C,S52036A,S52036B,S52036C,S52041A,S52041B | |
| | S52041C,S52042A,S52042B,S52042C,S52043A,S52043B,S52043C | |
| | S52044A,S52044B,S52044C,S52045A,S52045B,S52045C,S52046A | |
| | S52046B,S52046C,S52091A,S52091B,S52091C,S52092A,S52092B | |

Continued on next page

**Supplementary Table S1:** (cont'd) A comprehensive mapping of diagnosis groups used in the experiments to their corresponding ICD-10 and ICD-9 codes.

| Diagnosis | ICD-10 | ICD-9 |
|---|---|---|
| | S52092C,S52099A,S52099B,S52099C,S52101A,S52101B,S52101C<br>S52102A,S52102B,S52102C,S52109A,S52109B,S52109C,S52111A<br>S52112A,S52119A,S52121A,S52121B,S52121C,S52122A,S52122B<br>S52122C,S52123A,S52123B,S52123C,S52124A,S52124B,S52124C<br>S52125A,S52125B,S52125C,S52126A,S52126B,S52126C,S52131A<br>S52131B,S52131C,S52132A,S52132B,S52132C,S52133A,S52133B<br>S52133C,S52134A,S52134B,S52134C,S52135A,S52135B,S52135C<br>S52136A,S52136B,S52136C,S52181A,S52181B,S52181C,S52182A<br>S52182B,S52182C,S52189A,S52189B,S52189C,S52201A,S52201B<br>S52201C,S52202A,S52202B,S52202C,S52209A,S52209B,S52209C<br>S52211A,S52212A,S52219A,S52221A,S52221B,S52221C,S52222A<br>S52222B,S52222C,S52223A,S52223B,S52223C,S52224A,S52224B<br>S52224C,S52225A,S52225B,S52225C,S52226A,S52226B,S52226C<br>S52231A,S52231B,S52231C,S52232A,S52232B,S52232C,S52233A<br>S52233B,S52233C,S52234A,S52234B,S52234C,S52235A,S52235B<br>S52235C,S52236A,S52236B,S52236C,S52241A,S52241B,S52241C<br>S52242A,S52242B,S52242C,S52243A,S52243B,S52243C,S52244A<br>S52244B,S52244C,S52245A,S52245B,S52245C,S52246A,S52246B<br>S52246C,S52251A,S52251B,S52251C,S52252A,S52252B,S52252C<br>S52253A,S52253B,S52253C,S52254A,S52254B,S52254C,S52255A<br>S52255B,S52255C,S52256A,S52256B,S52256C,S52261A,S52261B<br>S52261C,S52262A,S52262B,S52262C,S52263A,S52263B,S52263C<br>S52264A,S52264B,S52264C,S52265A,S52265B,S52265C,S52266A<br>S52266B,S52266C,S52271A,S52271B,S52271C,S52272A,S52272B<br>S52272C,S52279A,S52279B,S52279C,S52281A,S52281B,S52281C<br>S52282A,S52282B,S52282C,S52283A,S52283B,S52283C,S52291A<br>S52291B,S52291C,S52292A,S52292B,S52292C,S52299A,S52299B<br>S52299C,S52301A,S52301B,S52301C,S52302A,S52302B,S52302C<br>S52309A,S52309B,S52309C,S52311A,S52312A,S52319A,S52321A<br>S52321B,S52321C,S52322A,S52322B,S52322C,S52323A,S52323B<br>S52323C,S52324A,S52324B,S52324C,S52325A,S52325B,S52325C<br>S52326A,S52326B,S52326C,S52331A,S52331B,S52331C,S52332A<br>S52332B,S52332C,S52333A,S52333B,S52333C,S52334A,S52334B<br>S52334C,S52335A,S52335B,S52335C,S52336A,S52336B,S52336C<br>S52341A,S52341B,S52341C,S52342A,S52342B,S52342C,S52343A<br>S52343B,S52343C,S52344A,S52344B,S52344C,S52345A,S52345B<br>S52345C,S52346A,S52346B,S52346C,S52351A,S52351B,S52351C<br>S52352A,S52352B,S52352C,S52353A,S52353B,S52353C,S52354A<br>S52354B,S52354C,S52355A,S52355B,S52355C,S52356A,S52356B<br>S52356C,S52361A,S52361B,S52361C,S52362A,S52362B,S52362C | |

Continued on next page

**Supplementary Table S1:** (cont'd) A comprehensive mapping of diagnosis groups used in the experiments to their corresponding ICD-10 and ICD-9 codes.

| Diagnosis | ICD-10 | ICD-9 |
|---|---|---|
| | S52363A,S52363B,S52363C,S52364A,S52364B,S52364C,S52365A | |
| | S52365B,S52365C,S52366A,S52366B,S52366C,S52371A,S52371B | |
| | S52371C,S52372A,S52372B,S52372C,S52379A,S52379B,S52379C | |
| | S52381A,S52381B,S52381C,S52382A,S52382B,S52382C,S52389A | |
| | S52389B,S52389C,S52391A,S52391B,S52391C,S52392A,S52392B | |
| | S52392C,S52399A,S52399B,S52399C,S52501A,S52501B,S52501C | |
| | S52502A,S52502B,S52502C,S52509A,S52509B,S52509C,S52511A | |
| | S52511B,S52511C,S52512A,S52512B,S52512C,S52513A,S52513B | |
| | S52513C,S52514A,S52514B,S52514C,S52515A,S52515B,S52515C | |
| | S52516A,S52516B,S52516C,S52521A,S52522A,S52529A,S52531A | |
| | S52531B,S52531C,S52532A,S52532B,S52532C,S52539A,S52539B | |
| | S52539C,S52541A,S52541B,S52541C,S52542A,S52542B,S52542C | |
| | S52549A,S52549B,S52549C,S52551A,S52551B,S52551C,S52552A | |
| | S52552B,S52552C,S52559A,S52559B,S52559C,S52561A,S52561B | |
| | S52561C,S52562A,S52562B,S52562C,S52569A,S52569B,S52569C | |
| | S52571A,S52571B,S52571C,S52572A,S52572B,S52572C,S52579A | |
| | S52579B,S52579C,S52591A,S52591B,S52591C,S52592A,S52592B | |
| | S52592C,S52599A,S52599B,S52599C,S52601A,S52601B,S52601C | |
| | S52602A,S52602B,S52602C,S52609A,S52609B,S52609C,S52611A | |
| | S52611B,S52611C,S52612A,S52612B,S52612C,S52613A,S52613B | |
| | S52613C,S52614A,S52614B,S52614C,S52615A,S52615B,S52615C | |
| | S52616A,S52616B,S52616C,S52621A,S52622A,S52629A,S52691A | |
| | S52691B,S52691C,S52692A,S52692B,S52692C,S52699A,S52699B | |
| | S52699C,S5290XA,S5290XB,S5290XC,S5291XA,S5291XB,S5291XC | |
| | S5292XA,S5292XB,S5292XC,S59001A,S59002A,S59009A,S59011A | |
| | S59012A,S59019A,S59021A,S59022A,S59029A,S59031A,S59032A | |
| | S59039A,S59041A,S59042A,S59049A,S59091A,S59092A,S59099A | |
| | S59101A,S59102A,S59109A,S59111A,S59112A,S59119A,S59121A | |
| | S59122A,S59129A,S59131A,S59132A,S59139A,S59141A,S59142A | |
| | S59149A,S59191A,S59192A,S59199A,S59201A,S59202A,S59209A | |
| | S59211A,S59212A,S59219A,S59221A,S59222A,S59229A,S59231A | |
| | S59232A,S59239A,S59241A,S59242A,S59249A,S59291A,S59292A | |
| | S59299A,S62001A,S62001B,S62002A,S62002B,S62009A,S62009B | |
| | S62011A,S62011B,S62012A,S62012B,S62013A,S62013B,S62014A | |
| | S62014B,S62015A,S62015B,S62016A,S62016B,S62021A,S62021B | |
| | S62022A,S62022B,S62023A,S62023B,S62024A,S62024B,S62025A | |
| | S62025B,S62026A,S62026B,S62031A,S62031B,S62032A,S62032B | |
| | S62033A,S62033B,S62034A,S62034B,S62035A,S62035B,S62036A | |
| | S62036B,S62101A,S62101B,S62102A,S62102B,S62109A,S62109B | |
| | S62111A,S62111B,S62112A,S62112B,S62113A,S62113B,S62114A | |

Continued on next page

**Supplementary Table S1:** (cont'd) A comprehensive mapping of diagnosis groups used in the experiments to their corresponding ICD-10 and ICD-9 codes.

| Diagnosis | ICD-10 | ICD-9 |
|---|---|---|
| | S62114B,S62115A,S62115B,S62116A,S62116B,S62121A,S62121B | |
| | S62122A,S62122B,S62123A,S62123B,S62124A,S62124B,S62125A | |
| | S62125B,S62126A,S62126B,S62131A,S62131B,S62132A,S62132B | |
| | S62133A,S62133B,S62134A,S62134B,S62135A,S62135B,S62136A | |
| | S62136B,S62141A,S62141B,S62142A,S62142B,S62143A,S62143B | |
| | S62144A,S62144B,S62145A,S62145B,S62146A,S62146B,S62151A | |
| | S62151B,S62152A,S62152B,S62153A,S62153B,S62154A,S62154B | |
| | S62155A,S62155B,S62156A,S62156B,S62161A,S62161B,S62162A | |
| | S62162B,S62163A,S62163B,S62164A,S62164B,S62165A,S62165B | |
| | S62166A,S62166B,S62171A,S62171B,S62172A,S62172B,S62173A | |
| | S62173B,S62174A,S62174B,S62175A,S62175B,S62176A,S62176B | |
| | S62181A,S62181B,S62182A,S62182B,S62183A,S62183B,S62184A | |
| | S62184B,S62185A,S62185B,S62186A,S62186B,S62201A,S62201B | |
| | S62202A,S62202B,S62209A,S62209B,S62211A,S62211B,S62212A | |
| | S62212B,S62213A,S62213B,S62221A,S62221B,S62222A,S62222B | |
| | S62223A,S62223B,S62224A,S62224B,S62225A,S62225B,S62226A | |
| | S62226B,S62231A,S62231B,S62232A,S62232B,S62233A,S62233B | |
| | S62234A,S62234B,S62235A,S62235B,S62236A,S62236B,S62241A | |
| | S62241B,S62242A,S62242B,S62243A,S62243B,S62244A,S62244B | |
| | S62245A,S62245B,S62246A,S62246B,S62251A,S62251B,S62252A | |
| | S62252B,S62253A,S62253B,S62254A,S62254B,S62255A,S62255B | |
| | S62256A,S62256B,S62291A,S62291B,S62292A,S62292B,S62299A | |
| | S62299B,S62300A,S62300B,S62301A,S62301B,S62302A,S62302B | |
| | S62303A,S62303B,S62304A,S62304B,S62305A,S62305B,S62306A | |
| | S62306B,S62307A,S62307B,S62308A,S62308B,S62309A,S62309B | |
| | S62310A,S62310B,S62311A,S62311B,S62312A,S62312B,S62313A | |
| | S62313B,S62314A,S62314B,S62315A,S62315B,S62316A,S62316B | |
| | S62317A,S62317B,S62318A,S62318B,S62319A,S62319B,S62320A | |
| | S62320B,S62321A,S62321B,S62322A,S62322B,S62323A,S62323B | |
| | S62324A,S62324B,S62325A,S62325B,S62326A,S62326B,S62327A | |
| | S62327B,S62328A,S62328B,S62329A,S62329B,S62330A,S62330B | |
| | S62331A,S62331B,S62332A,S62332B,S62333A,S62333B,S62334A | |
| | S62334B,S62335A,S62335B,S62336A,S62336B,S62337A,S62337B | |
| | S62338A,S62338B,S62339A,S62339B,S62340A,S62340B,S62341A | |
| | S62341B,S62342A,S62342B,S62343A,S62343B,S62344A,S62344B | |
| | S62345A,S62345B,S62346A,S62346B,S62347A,S62347B,S62348A | |
| | S62348B,S62349A,S62349B,S62350A,S62350B,S62351A,S62351B | |
| | S62352A,S62352B,S62353A,S62353B,S62354A,S62354B,S62355A | |
| | S62355B,S62356A,S62356B,S62357A,S62357B,S62358A,S62358B | |
| | S62359A,S62359B,S62360A,S62360B,S62361A,S62361B,S62362A | |

Continued on next page

**Supplementary Table S1:** (cont'd) A comprehensive mapping of diagnosis groups used in the experiments to their corresponding ICD-10 and ICD-9 codes.

| Diagnosis | ICD-10 | ICD-9 |
|---|---|---|
| | S62362B,S62363A,S62363B,S62364A,S62364B,S62365A,S62365B S62366A,S62366B,S62367A,S62367B,S62368A,S62368B,S62369A S62369B,S62390A,S62390B,S62391A,S62391B,S62392A,S62392B S62393A,S62393B,S62394A,S62394B,S62395A,S62395B,S62396A S62396B,S62397A,S62397B,S62398A,S62398B,S62399A,S62399B S62501A,S62501B,S62502A,S62502B,S62509A,S62509B,S62511A S62511B,S62512A,S62512B,S62513A,S62513B,S62514A,S62514B S62515A,S62515B,S62516A,S62516B,S62521A,S62521B,S62522A S62522B,S62523A,S62523B,S62524A,S62524B,S62525A,S62525B S62526A,S62526B,S62600A,S62600B,S62601A,S62601B,S62602A S62602B,S62603A,S62603B,S62604A,S62604B,S62605A,S62605B S62606A,S62606B,S62607A,S62607B,S62608A,S62608B,S62609A S62609B,S62610A,S62610B,S62611A,S62611B,S62612A,S62612B S62613A,S62613B,S62614A,S62614B,S62615A,S62615B,S62616A S62616B,S62617A,S62617B,S62618A,S62618B,S62619A,S62619B S62620A,S62620B,S62621A,S62621B,S62622A,S62622B,S62623A S62623B,S62624A,S62624B,S62625A,S62625B,S62626A,S62626B S62627A,S62627B,S62628A,S62628B,S62629A,S62629B,S62630A S62630B,S62631A,S62631B,S62632A,S62632B,S62633A,S62633B S62634A,S62634B,S62635A,S62635B,S62636A,S62636B,S62637A S62637B,S62638A,S62638B,S62639A,S62639B,S62640A,S62640B S62641A,S62641B,S62642A,S62642B,S62643A,S62643B,S62644A S62644B,S62645A,S62645B,S62646A,S62646B,S62647A,S62647B S62648A,S62648B,S62649A,S62649B,S62650A,S62650B,S62651A S62651B,S62652A,S62652B,S62653A,S62653B,S62654A,S62654B S62655A,S62655B,S62656A,S62656B,S62657A,S62657B,S62658A S62658B,S62659A,S62659B,S62660A,S62660B,S62661A,S62661B S62662A,S62662B,S62663A,S62663B,S62664A,S62664B,S62665A S62665B,S62666A,S62666B,S62667A,S62667B,S62668A,S62668B S62669A,S62669B,S6290XA,S6290XB,S6291XA,S6291XB,S6292XA S6292XB | |
| Fracture of the upper limb, subsequent encounter | M9731XD,M9732XD,M9741XD,M9742XD,S42001D,S42001G,S42001K S42001P,S42002D,S42002G,S42002K,S42002P,S42009D,S42009G S42009K,S42009P,S42011D,S42011G,S42011K,S42011P,S42012D S42012G,S42012K,S42012P,S42013D,S42013G,S42013K,S42013P S42014D,S42014G,S42014K,S42014P,S42015D,S42015G,S42015K S42015P,S42016D,S42016G,S42016K,S42016P,S42017D,S42017G S42017K,S42017P,S42018D,S42018G,S42018K,S42018P,S42019D S42019G,S42019K,S42019P,S42021D,S42021G,S42021K,S42021P | 73381,73382,V5410,V5411,V5412,V5419 |

Continued on next page

**Supplementary Table S1:** (cont'd) A comprehensive mapping of diagnosis groups used in the experiments to their corresponding ICD-10 and ICD-9 codes.

| Diagnosis | ICD-10 | ICD-9 |
|---|---|---|
| | S42022D,S42022G,S42022K,S42022P,S42023D,S42023G,S42023K | |
| | S42023P,S42024D,S42024G,S42024K,S42024P,S42025D,S42025G | |
| | S42025K,S42025P,S42026D,S42026G,S42026K,S42026P,S42031D | |
| | S42031G,S42031K,S42031P,S42032D,S42032G,S42032K,S42032P | |
| | S42033D,S42033G,S42033K,S42033P,S42034D,S42034G,S42034K | |
| | S42034P,S42035D,S42035G,S42035K,S42035P,S42036D,S42036G | |
| | S42036K,S42036P,S42101D,S42101G,S42101K,S42101P,S42102D | |
| | S42102G,S42102K,S42102P,S42109D,S42109G,S42109K,S42109P | |
| | S42111D,S42111G,S42111K,S42111P,S42112D,S42112G,S42112K | |
| | S42112P,S42113D,S42113G,S42113K,S42113P,S42114D,S42114G | |
| | S42114K,S42114P,S42115D,S42115G,S42115K,S42115P,S42116D | |
| | S42116G,S42116K,S42116P,S42121D,S42121G,S42121K,S42121P | |
| | S42122D,S42122G,S42122K,S42122P,S42123D,S42123G,S42123K | |
| | S42123P,S42124D,S42124G,S42124K,S42124P,S42125D,S42125G | |
| | S42125K,S42125P,S42126D,S42126G,S42126K,S42126P,S42131D | |
| | S42131G,S42131K,S42131P,S42132D,S42132G,S42132K,S42132P | |
| | S42133D,S42133G,S42133K,S42133P,S42134D,S42134G,S42134K | |
| | S42134P,S42135D,S42135G,S42135K,S42135P,S42136D,S42136G | |
| | S42136K,S42136P,S42141D,S42141G,S42141K,S42141P,S42142D | |
| | S42142G,S42142K,S42142P,S42143D,S42143G,S42143K,S42143P | |
| | S42144D,S42144G,S42144K,S42144P,S42145D,S42145G,S42145K | |
| | S42145P,S42146D,S42146G,S42146K,S42146P,S42151D,S42151G | |
| | S42151K,S42151P,S42152D,S42152G,S42152K,S42152P,S42153D | |
| | S42153G,S42153K,S42153P,S42154D,S42154G,S42154K,S42154P | |
| | S42155D,S42155G,S42155K,S42155P,S42156D,S42156G,S42156K | |
| | S42156P,S42191D,S42191G,S42191K,S42191P,S42192D,S42192G | |
| | S42192K,S42192P,S42199D,S42199G,S42199K,S42199P,S42201D | |
| | S42201G,S42201K,S42201P,S42202D,S42202G,S42202K,S42202P | |
| | S42209D,S42209G,S42209K,S42209P,S42211D,S42211G,S42211K | |
| | S42211P,S42212D,S42212G,S42212K,S42212P,S42213D,S42213G | |
| | S42213K,S42213P,S42214D,S42214G,S42214K,S42214P,S42215D | |
| | S42215G,S42215K,S42215P,S42216D,S42216G,S42216K,S42216P | |
| | S42221D,S42221G,S42221K,S42221P,S42222D,S42222G,S42222K | |
| | S42222P,S42223D,S42223G,S42223K,S42223P,S42224D,S42224G | |
| | S42224K,S42224P,S42225D,S42225G,S42225K,S42225P,S42226D | |
| | S42226G,S42226K,S42226P,S42231D,S42231G,S42231K,S42231P | |
| | S42232D,S42232G,S42232K,S42232P,S42239D,S42239G,S42239K | |
| | S42239P,S42241D,S42241G,S42241K,S42241P,S42242D,S42242G | |
| | S42242K,S42242P,S42249D,S42249G,S42249K,S42249P,S42251D | |
| | S42251G,S42251K,S42251P,S42252D,S42252G,S42252K,S42252P | |

Continued on next page

**Supplementary Table S1:** (cont'd) A comprehensive mapping of diagnosis groups used in the experiments to their corresponding ICD-10 and ICD-9 codes.

| Diagnosis | ICD-10 | ICD-9 |
|---|---|---|
| | S42253D,S42253G,S42253K,S42253P,S42254D,S42254G,S42254K | |
| | S42254P,S42255D,S42255G,S42255K,S42255P,S42256D,S42256G | |
| | S42256K,S42256P,S42261D,S42261G,S42261K,S42261P,S42262D | |
| | S42262G,S42262K,S42262P,S42263D,S42263G,S42263K,S42263P | |
| | S42264D,S42264G,S42264K,S42264P,S42265D,S42265G,S42265K | |
| | S42265P,S42266D,S42266G,S42266K,S42266P,S42271D,S42271G | |
| | S42271K,S42271P,S42272D,S42272G,S42272K,S42272P,S42279D | |
| | S42279G,S42279K,S42279P,S42291D,S42291G,S42291K,S42291P | |
| | S42292D,S42292G,S42292K,S42292P,S42293D,S42293G,S42293K | |
| | S42293P,S42294D,S42294G,S42294K,S42294P,S42295D,S42295G | |
| | S42295K,S42295P,S42296D,S42296G,S42296K,S42296P,S42301D | |
| | S42301G,S42301K,S42301P,S42302D,S42302G,S42302K,S42302P | |
| | S42309D,S42309G,S42309K,S42309P,S42311D,S42311G,S42311K | |
| | S42311P,S42312D,S42312G,S42312K,S42312P,S42319D,S42319G | |
| | S42319K,S42319P,S42321D,S42321G,S42321K,S42321P,S42322D | |
| | S42322G,S42322K,S42322P,S42323D,S42323G,S42323K,S42323P | |
| | S42324D,S42324G,S42324K,S42324P,S42325D,S42325G,S42325K | |
| | S42325P,S42326D,S42326G,S42326K,S42326P,S42331D,S42331G | |
| | S42331K,S42331P,S42332D,S42332G,S42332K,S42332P,S42333D | |
| | S42333G,S42333K,S42333P,S42334D,S42334G,S42334K,S42334P | |
| | S42335D,S42335G,S42335K,S42335P,S42336D,S42336G,S42336K | |
| | S42336P,S42341D,S42341G,S42341K,S42341P,S42342D,S42342G | |
| | S42342K,S42342P,S42343D,S42343G,S42343K,S42343P,S42344D | |
| | S42344G,S42344K,S42344P,S42345D,S42345G,S42345K,S42345P | |
| | S42346D,S42346G,S42346K,S42346P,S42351D,S42351G,S42351K | |
| | S42351P,S42352D,S42352G,S42352K,S42352P,S42353D,S42353G | |
| | S42353K,S42353P,S42354D,S42354G,S42354K,S42354P,S42355D | |
| | S42355G,S42355K,S42355P,S42356D,S42356G,S42356K,S42356P | |
| | S42361D,S42361G,S42361K,S42361P,S42362D,S42362G,S42362K | |
| | S42362P,S42363D,S42363G,S42363K,S42363P,S42364D,S42364G | |
| | S42364K,S42364P,S42365D,S42365G,S42365K,S42365P,S42366D | |
| | S42366G,S42366K,S42366P,S42391D,S42391G,S42391K,S42391P | |
| | S42392D,S42392G,S42392K,S42392P,S42399D,S42399G,S42399K | |
| | S42399P,S42401D,S42401G,S42401K,S42401P,S42402D,S42402G | |
| | S42402K,S42402P,S42409D,S42409G,S42409K,S42409P,S42411D | |
| | S42411G,S42411K,S42411P,S42412D,S42412G,S42412K,S42412P | |
| | S42413D,S42413G,S42413K,S42413P,S42414D,S42414G,S42414K | |
| | S42414P,S42415D,S42415G,S42415K,S42415P,S42416D,S42416G | |
| | S42416K,S42416P,S42421D,S42421G,S42421K,S42421P,S42422D | |
| | S42422G,S42422K,S42422P,S42423D,S42423G,S42423K,S42423P | |

Continued on next page

**Supplementary Table S1:** (cont'd) A comprehensive mapping of diagnosis groups used in the experiments to their corresponding ICD-10 and ICD-9 codes.

| Diagnosis | ICD-10 | ICD-9 |
|---|---|---|
| | S42424D,S42424G,S42424K,S42424P,S42425D,S42425G,S42425K | |
| | S42425P,S42426D,S42426G,S42426K,S42426P,S42431D,S42431G | |
| | S42431K,S42431P,S42432D,S42432G,S42432K,S42432P,S42433D | |
| | S42433G,S42433K,S42433P,S42434D,S42434G,S42434K,S42434P | |
| | S42435D,S42435G,S42435K,S42435P,S42436D,S42436G,S42436K | |
| | S42436P,S42441D,S42441G,S42441K,S42441P,S42442D,S42442G | |
| | S42442K,S42442P,S42443D,S42443G,S42443K,S42443P,S42444D | |
| | S42444G,S42444K,S42444P,S42445D,S42445G,S42445K,S42445P | |
| | S42446D,S42446G,S42446K,S42446P,S42447D,S42447G,S42447K | |
| | S42447P,S42448D,S42448G,S42448K,S42448P,S42449D,S42449G | |
| | S42449K,S42449P,S42451D,S42451G,S42451K,S42451P,S42452D | |
| | S42452G,S42452K,S42452P,S42453D,S42453G,S42453K,S42453P | |
| | S42454D,S42454G,S42454K,S42454P,S42455D,S42455G,S42455K | |
| | S42455P,S42456D,S42456G,S42456K,S42456P,S42461D,S42461G | |
| | S42461K,S42461P,S42462D,S42462G,S42462K,S42462P,S42463D | |
| | S42463G,S42463K,S42463P,S42464D,S42464G,S42464K,S42464P | |
| | S42465D,S42465G,S42465K,S42465P,S42466D,S42466G,S42466K | |
| | S42466P,S42471D,S42471G,S42471K,S42471P,S42472D,S42472G | |
| | S42472K,S42472P,S42473D,S42473G,S42473K,S42473P,S42474D | |
| | S42474G,S42474K,S42474P,S42475D,S42475G,S42475K,S42475P | |
| | S42476D,S42476G,S42476K,S42476P,S42481D,S42481G,S42481K | |
| | S42481P,S42482D,S42482G,S42482K,S42482P,S42489D,S42489G | |
| | S42489K,S42489P,S42491D,S42491G,S42491K,S42491P,S42492D | |
| | S42492G,S42492K,S42492P,S42493D,S42493G,S42493K,S42493P | |
| | S42494D,S42494G,S42494K,S42494P,S42495D,S42495G,S42495K | |
| | S42495P,S42496D,S42496G,S42496K,S42496P,S4290XD,S4290XG | |
| | S4290XK,S4290XP,S4291XD,S4291XG,S4291XK,S4291XP,S4292XD | |
| | S4292XG,S4292XK,S4292XP,S49001D,S49001G,S49001K,S49001P | |
| | S49002D,S49002G,S49002K,S49002P,S49009D,S49009G,S49009K | |
| | S49009P,S49011D,S49011G,S49011K,S49011P,S49012D,S49012G | |
| | S49012K,S49012P,S49019D,S49019G,S49019K,S49019P,S49021D | |
| | S49021G,S49021K,S49021P,S49022D,S49022G,S49022K,S49022P | |
| | S49029D,S49029G,S49029K,S49029P,S49031D,S49031G,S49031K | |
| | S49031P,S49032D,S49032G,S49032K,S49032P,S49039D,S49039G | |
| | S49039K,S49039P,S49041D,S49041G,S49041K,S49041P,S49042D | |
| | S49042G,S49042K,S49042P,S49049D,S49049G,S49049K,S49049P | |
| | S49091D,S49091G,S49091K,S49091P,S49092D,S49092G,S49092K | |
| | S49092P,S49099D,S49099G,S49099K,S49099P,S49101D,S49101G | |
| | S49101K,S49101P,S49102D,S49102G,S49102K,S49102P,S49109D | |
| | S49109G,S49109K,S49109P,S49111D,S49111G,S49111K,S49111P | |

Continued on next page

**Supplementary Table S1:** (cont'd) A comprehensive mapping of diagnosis groups used in the experiments to their corresponding ICD-10 and ICD-9 codes.

| Diagnosis | ICD-10 | ICD-9 |
|---|---|---|
| | S49112D,S49112G,S49112K,S49112P,S49119D,S49119G,S49119K | |
| | S49119P,S49121D,S49121G,S49121K,S49121P,S49122D,S49122G | |
| | S49122K,S49122P,S49129D,S49129G,S49129K,S49129P,S49131D | |
| | S49131G,S49131K,S49131P,S49132D,S49132G,S49132K,S49132P | |
| | S49139D,S49139G,S49139K,S49139P,S49141D,S49141G,S49141K | |
| | S49141P,S49142D,S49142G,S49142K,S49142P,S49149D,S49149G | |
| | S49149K,S49149P,S49191D,S49191G,S49191K,S49191P,S49192D | |
| | S49192G,S49192K,S49192P,S49199D,S49199G,S49199K,S49199P | |
| | S52001D,S52001E,S52001F,S52001G,S52001H,S52001J,S52001K | |
| | S52001M,S52001N,S52001P,S52001Q,S52001R,S52002D,S52002E | |
| | S52002F,S52002G,S52002H,S52002J,S52002K,S52002M,S52002N | |
| | S52002P,S52002Q,S52002R,S52009D,S52009E,S52009F,S52009G | |
| | S52009H,S52009J,S52009K,S52009M,S52009N,S52009P,S52009Q | |
| | S52009R,S52011D,S52011G,S52011K,S52011P,S52012D,S52012G | |
| | S52012K,S52012P,S52019D,S52019G,S52019K,S52019P,S52021D | |
| | S52021E,S52021F,S52021G,S52021H,S52021J,S52021K,S52021M | |
| | S52021N,S52021P,S52021Q,S52021R,S52022D,S52022E,S52022F | |
| | S52022G,S52022H,S52022J,S52022K,S52022M,S52022N,S52022P | |
| | S52022Q,S52022R,S52023D,S52023E,S52023F,S52023G,S52023H | |
| | S52023J,S52023K,S52023M,S52023N,S52023P,S52023Q,S52023R | |
| | S52024D,S52024E,S52024F,S52024G,S52024H,S52024J,S52024K | |
| | S52024M,S52024N,S52024P,S52024Q,S52024R,S52025D,S52025E | |
| | S52025F,S52025G,S52025H,S52025J,S52025K,S52025M,S52025N | |
| | S52025P,S52025Q,S52025R,S52026D,S52026E,S52026F,S52026G | |
| | S52026H,S52026J,S52026K,S52026M,S52026N,S52026P,S52026Q | |
| | S52026R,S52031D,S52031E,S52031F,S52031G,S52031H,S52031J | |
| | S52031K,S52031M,S52031N,S52031P,S52031Q,S52031R,S52032D | |
| | S52032E,S52032F,S52032G,S52032H,S52032J,S52032K,S52032M | |
| | S52032N,S52032P,S52032Q,S52032R,S52033D,S52033E,S52033F | |
| | S52033G,S52033H,S52033J,S52033K,S52033M,S52033N,S52033P | |
| | S52033Q,S52033R,S52034D,S52034E,S52034F,S52034G,S52034H | |
| | S52034J,S52034K,S52034M,S52034N,S52034P,S52034Q,S52034R | |
| | S52035D,S52035E,S52035F,S52035G,S52035H,S52035J,S52035K | |
| | S52035M,S52035N,S52035P,S52035Q,S52035R,S52036D,S52036E | |
| | S52036F,S52036G,S52036H,S52036J,S52036K,S52036M,S52036N | |
| | S52036P,S52036Q,S52036R,S52041D,S52041E,S52041F,S52041G | |
| | S52041H,S52041J,S52041K,S52041M,S52041N,S52041P,S52041Q | |
| | S52041R,S52042D,S52042E,S52042F,S52042G,S52042H,S52042J | |
| | S52042K,S52042M,S52042N,S52042P,S52042Q,S52042R,S52043D | |
| | S52043E,S52043F,S52043G,S52043H,S52043J,S52043K,S52043M | |

Continued on next page

**Supplementary Table S1:** (cont'd) A comprehensive mapping of diagnosis groups used in the experiments to their corresponding ICD-10 and ICD-9 codes.

| Diagnosis | ICD-10 | ICD-9 |
|---|---|---|
| | S52043N,S52043P,S52043Q,S52043R,S52044D,S52044E,S52044F | |
| | S52044G,S52044H,S52044J,S52044K,S52044M,S52044N,S52044P | |
| | S52044Q,S52044R,S52045D,S52045E,S52045F,S52045G,S52045H | |
| | S52045J,S52045K,S52045M,S52045N,S52045P,S52045Q,S52045R | |
| | S52046D,S52046E,S52046F,S52046G,S52046H,S52046J,S52046K | |
| | S52046M,S52046N,S52046P,S52046Q,S52046R,S52091D,S52091E | |
| | S52091F,S52091G,S52091H,S52091J,S52091K,S52091M,S52091N | |
| | S52091P,S52091Q,S52091R,S52092D,S52092E,S52092F,S52092G | |
| | S52092H,S52092J,S52092K,S52092M,S52092N,S52092P,S52092Q | |
| | S52092R,S52099D,S52099E,S52099F,S52099G,S52099H,S52099J | |
| | S52099K,S52099M,S52099N,S52099P,S52099Q,S52099R,S52101D | |
| | S52101E,S52101F,S52101G,S52101H,S52101J,S52101K,S52101M | |
| | S52101N,S52101P,S52101Q,S52101R,S52102D,S52102E,S52102F | |
| | S52102G,S52102H,S52102J,S52102K,S52102M,S52102N,S52102P | |
| | S52102Q,S52102R,S52109D,S52109E,S52109F,S52109G,S52109H | |
| | S52109J,S52109K,S52109M,S52109N,S52109P,S52109Q,S52109R | |
| | S52111D,S52111G,S52111K,S52111P,S52112D,S52112G,S52112K | |
| | S52112P,S52119D,S52119G,S52119K,S52119P,S52121D,S52121E | |
| | S52121F,S52121G,S52121H,S52121J,S52121K,S52121M,S52121N | |
| | S52121P,S52121Q,S52121R,S52122D,S52122E,S52122F,S52122G | |
| | S52122H,S52122J,S52122K,S52122M,S52122N,S52122P,S52122Q | |
| | S52122R,S52123D,S52123E,S52123F,S52123G,S52123H,S52123J | |
| | S52123K,S52123M,S52123N,S52123P,S52123Q,S52123R,S52124D | |
| | S52124E,S52124F,S52124G,S52124H,S52124J,S52124K,S52124M | |
| | S52124N,S52124P,S52124Q,S52124R,S52125D,S52125E,S52125F | |
| | S52125G,S52125H,S52125J,S52125K,S52125M,S52125N,S52125P | |
| | S52125Q,S52125R,S52126D,S52126E,S52126F,S52126G,S52126H | |
| | S52126J,S52126K,S52126M,S52126N,S52126P,S52126Q,S52126R | |
| | S52131D,S52131E,S52131F,S52131G,S52131H,S52131J,S52131K | |
| | S52131M,S52131N,S52131P,S52131Q,S52131R,S52132D,S52132E | |
| | S52132F,S52132G,S52132H,S52132J,S52132K,S52132M,S52132N | |
| | S52132P,S52132Q,S52132R,S52133D,S52133E,S52133F,S52133G | |
| | S52133H,S52133J,S52133K,S52133M,S52133N,S52133P,S52133Q | |
| | S52133R,S52134D,S52134E,S52134F,S52134G,S52134H,S52134J | |
| | S52134K,S52134M,S52134N,S52134P,S52134Q,S52134R,S52135D | |
| | S52135E,S52135F,S52135G,S52135H,S52135J,S52135K,S52135M | |
| | S52135N,S52135P,S52135Q,S52135R,S52136D,S52136E,S52136F | |
| | S52136G,S52136H,S52136J,S52136K,S52136M,S52136N,S52136P | |
| | S52136Q,S52136R,S52181D,S52181E,S52181F,S52181G,S52181H | |
| | S52181J,S52181K,S52181M,S52181N,S52181P,S52181Q,S52181R | |

Continued on next page

**Supplementary Table S1:** (cont'd) A comprehensive mapping of diagnosis groups used in the experiments to their corresponding ICD-10 and ICD-9 codes.

| Diagnosis | ICD-10 | ICD-9 |
|---|---|---|
| | S52182D,S52182E,S52182F,S52182G,S52182H,S52182J,S52182K | |
| | S52182M,S52182N,S52182P,S52182Q,S52182R,S52189D,S52189E | |
| | S52189F,S52189G,S52189H,S52189J,S52189K,S52189M,S52189N | |
| | S52189P,S52189Q,S52189R,S52201D,S52201E,S52201F,S52201G | |
| | S52201H,S52201J,S52201K,S52201M,S52201N,S52201P,S52201Q | |
| | S52201R,S52202D,S52202E,S52202F,S52202G,S52202H,S52202J | |
| | S52202K,S52202M,S52202N,S52202P,S52202Q,S52202R,S52209D | |
| | S52209E,S52209F,S52209G,S52209H,S52209J,S52209K,S52209M | |
| | S52209N,S52209P,S52209Q,S52209R,S52211D,S52211G,S52211K | |
| | S52211P,S52212D,S52212G,S52212K,S52212P,S52219D,S52219G | |
| | S52219K,S52219P,S52221D,S52221E,S52221F,S52221G,S52221H | |
| | S52221J,S52221K,S52221M,S52221N,S52221P,S52221Q,S52221R | |
| | S52222D,S52222E,S52222F,S52222G,S52222H,S52222J,S52222K | |
| | S52222M,S52222N,S52222P,S52222Q,S52222R,S52223D,S52223E | |
| | S52223F,S52223G,S52223H,S52223J,S52223K,S52223M,S52223N | |
| | S52223P,S52223Q,S52223R,S52224D,S52224E,S52224F,S52224G | |
| | S52224H,S52224J,S52224K,S52224M,S52224N,S52224P,S52224Q | |
| | S52224R,S52225D,S52225E,S52225F,S52225G,S52225H,S52225J | |
| | S52225K,S52225M,S52225N,S52225P,S52225Q,S52225R,S52226D | |
| | S52226E,S52226F,S52226G,S52226H,S52226J,S52226K,S52226M | |
| | S52226N,S52226P,S52226Q,S52226R,S52231D,S52231E,S52231F | |
| | S52231G,S52231H,S52231J,S52231K,S52231M,S52231N,S52231P | |
| | S52231Q,S52231R,S52232D,S52232E,S52232F,S52232G,S52232H | |
| | S52232J,S52232K,S52232M,S52232N,S52232P,S52232Q,S52232R | |
| | S52233D,S52233E,S52233F,S52233G,S52233H,S52233J,S52233K | |
| | S52233M,S52233N,S52233P,S52233Q,S52233R,S52234D,S52234E | |
| | S52234F,S52234G,S52234H,S52234J,S52234K,S52234M,S52234N | |
| | S52234P,S52234Q,S52234R,S52235D,S52235E,S52235F,S52235G | |
| | S52235H,S52235J,S52235K,S52235M,S52235N,S52235P,S52235Q | |
| | S52235R,S52236D,S52236E,S52236F,S52236G,S52236H,S52236J | |
| | S52236K,S52236M,S52236N,S52236P,S52236Q,S52236R,S52241D | |
| | S52241E,S52241F,S52241G,S52241H,S52241J,S52241K,S52241M | |
| | S52241N,S52241P,S52241Q,S52241R,S52242D,S52242E,S52242F | |
| | S52242G,S52242H,S52242J,S52242K,S52242M,S52242N,S52242P | |
| | S52242Q,S52242R,S52243D,S52243E,S52243F,S52243G,S52243H | |
| | S52243J,S52243K,S52243M,S52243N,S52243P,S52243Q,S52243R | |
| | S52244D,S52244E,S52244F,S52244G,S52244H,S52244J,S52244K | |
| | S52244M,S52244N,S52244P,S52244Q,S52244R,S52245D,S52245E | |
| | S52245F,S52245G,S52245H,S52245J,S52245K,S52245M,S52245N | |
| | S52245P,S52245Q,S52245R,S52246D,S52246E,S52246F,S52246G | |

Continued on next page

**Supplementary Table S1:** (cont'd) A comprehensive mapping of diagnosis groups used in the experiments to their corresponding ICD-10 and ICD-9 codes.

| Diagnosis | ICD-10 | ICD-9 |
|---|---|---|
| | S52246H,S52246J,S52246K,S52246M,S52246N,S52246P,S52246Q | |
| | S52246R,S52251D,S52251E,S52251F,S52251G,S52251H,S52251J | |
| | S52251K,S52251M,S52251N,S52251P,S52251Q,S52251R,S52252D | |
| | S52252E,S52252F,S52252G,S52252H,S52252J,S52252K,S52252M | |
| | S52252N,S52252P,S52252Q,S52252R,S52253D,S52253E,S52253F | |
| | S52253G,S52253H,S52253J,S52253K,S52253M,S52253N,S52253P | |
| | S52253Q,S52253R,S52254D,S52254E,S52254F,S52254G,S52254H | |
| | S52254J,S52254K,S52254M,S52254N,S52254P,S52254Q,S52254R | |
| | S52255D,S52255E,S52255F,S52255G,S52255H,S52255J,S52255K | |
| | S52255M,S52255N,S52255P,S52255Q,S52255R,S52256D,S52256E | |
| | S52256F,S52256G,S52256H,S52256J,S52256K,S52256M,S52256N | |
| | S52256P,S52256Q,S52256R,S52261D,S52261E,S52261F,S52261G | |
| | S52261H,S52261J,S52261K,S52261M,S52261N,S52261P,S52261Q | |
| | S52261R,S52262D,S52262E,S52262F,S52262G,S52262H,S52262J | |
| | S52262K,S52262M,S52262N,S52262P,S52262Q,S52262R,S52263D | |
| | S52263E,S52263F,S52263G,S52263H,S52263J,S52263K,S52263M | |
| | S52263N,S52263P,S52263Q,S52263R,S52264D,S52264E,S52264F | |
| | S52264G,S52264H,S52264J,S52264K,S52264M,S52264N,S52264P | |
| | S52264Q,S52264R,S52265D,S52265E,S52265F,S52265G,S52265H | |
| | S52265J,S52265K,S52265M,S52265N,S52265P,S52265Q,S52265R | |
| | S52266D,S52266E,S52266F,S52266G,S52266H,S52266J,S52266K | |
| | S52266M,S52266N,S52266P,S52266Q,S52266R,S52271D,S52271E | |
| | S52271F,S52271G,S52271H,S52271J,S52271K,S52271M,S52271N | |
| | S52271P,S52271Q,S52271R,S52272D,S52272E,S52272F,S52272G | |
| | S52272H,S52272J,S52272K,S52272M,S52272N,S52272P,S52272Q | |
| | S52272R,S52279D,S52279E,S52279F,S52279G,S52279H,S52279J | |
| | S52279K,S52279M,S52279N,S52279P,S52279Q,S52279R,S52281D | |
| | S52281E,S52281F,S52281G,S52281H,S52281J,S52281K,S52281M | |
| | S52281N,S52281P,S52281Q,S52281R,S52282D,S52282E,S52282F | |
| | S52282G,S52282H,S52282J,S52282K,S52282M,S52282N,S52282P | |
| | S52282Q,S52282R,S52283D,S52283E,S52283F,S52283G,S52283H | |
| | S52283J,S52283K,S52283M,S52283N,S52283P,S52283Q,S52283R | |
| | S52291D,S52291E,S52291F,S52291G,S52291H,S52291J,S52291K | |
| | S52291M,S52291N,S52291P,S52291Q,S52291R,S52292D,S52292E | |
| | S52292F,S52292G,S52292H,S52292J,S52292K,S52292M,S52292N | |
| | S52292P,S52292Q,S52292R,S52299D,S52299E,S52299F,S52299G | |
| | S52299H,S52299J,S52299K,S52299M,S52299N,S52299P,S52299Q | |
| | S52299R,S52301D,S52301E,S52301F,S52301G,S52301H,S52301J | |
| | S52301K,S52301M,S52301N,S52301P,S52301Q,S52301R,S52302D | |
| | S52302E,S52302F,S52302G,S52302H,S52302J,S52302K,S52302M | |

Continued on next page

**Supplementary Table S1:** (cont'd) A comprehensive mapping of diagnosis groups used in the experiments to their corresponding ICD-10 and ICD-9 codes.

| Diagnosis | ICD-10 | ICD-9 |
|---|---|---|
| | S52302N,S52302P,S52302Q,S52302R,S52309D,S52309E,S52309F | |
| | S52309G,S52309H,S52309J,S52309K,S52309M,S52309N,S52309P | |
| | S52309Q,S52309R,S52311D,S52311G,S52311K,S52311P,S52312D | |
| | S52312G,S52312K,S52312P,S52319D,S52319G,S52319K,S52319P | |
| | S52321D,S52321E,S52321F,S52321G,S52321H,S52321J,S52321K | |
| | S52321M,S52321N,S52321P,S52321Q,S52321R,S52322D,S52322E | |
| | S52322F,S52322G,S52322H,S52322J,S52322K,S52322M,S52322N | |
| | S52322P,S52322Q,S52322R,S52323D,S52323E,S52323F,S52323G | |
| | S52323H,S52323J,S52323K,S52323M,S52323N,S52323P,S52323Q | |
| | S52323R,S52324D,S52324E,S52324F,S52324G,S52324H,S52324J | |
| | S52324K,S52324M,S52324N,S52324P,S52324Q,S52324R,S52325D | |
| | S52325E,S52325F,S52325G,S52325H,S52325J,S52325K,S52325M | |
| | S52325N,S52325P,S52325Q,S52325R,S52326D,S52326E,S52326F | |
| | S52326G,S52326H,S52326J,S52326K,S52326M,S52326N,S52326P | |
| | S52326Q,S52326R,S52331D,S52331E,S52331F,S52331G,S52331H | |
| | S52331J,S52331K,S52331M,S52331N,S52331P,S52331Q,S52331R | |
| | S52332D,S52332E,S52332F,S52332G,S52332H,S52332J,S52332K | |
| | S52332M,S52332N,S52332P,S52332Q,S52332R,S52333D,S52333E | |
| | S52333F,S52333G,S52333H,S52333J,S52333K,S52333M,S52333N | |
| | S52333P,S52333Q,S52333R,S52334D,S52334E,S52334F,S52334G | |
| | S52334H,S52334J,S52334K,S52334M,S52334N,S52334P,S52334Q | |
| | S52334R,S52335D,S52335E,S52335F,S52335G,S52335H,S52335J | |
| | S52335K,S52335M,S52335N,S52335P,S52335Q,S52335R,S52336D | |
| | S52336E,S52336F,S52336G,S52336H,S52336J,S52336K,S52336M | |
| | S52336N,S52336P,S52336Q,S52336R,S52341D,S52341E,S52341F | |
| | S52341G,S52341H,S52341J,S52341K,S52341M,S52341N,S52341P | |
| | S52341Q,S52341R,S52342D,S52342E,S52342F,S52342G,S52342H | |
| | S52342J,S52342K,S52342M,S52342N,S52342P,S52342Q,S52342R | |
| | S52343D,S52343E,S52343F,S52343G,S52343H,S52343J,S52343K | |
| | S52343M,S52343N,S52343P,S52343Q,S52343R,S52344D,S52344E | |
| | S52344F,S52344G,S52344H,S52344J,S52344K,S52344M,S52344N | |
| | S52344P,S52344Q,S52344R,S52345D,S52345E,S52345F,S52345G | |
| | S52345H,S52345J,S52345K,S52345M,S52345N,S52345P,S52345Q | |
| | S52345R,S52346D,S52346E,S52346F,S52346G,S52346H,S52346J | |
| | S52346K,S52346M,S52346N,S52346P,S52346Q,S52346R,S52351D | |
| | S52351E,S52351F,S52351G,S52351H,S52351J,S52351K,S52351M | |
| | S52351N,S52351P,S52351Q,S52351R,S52352D,S52352E,S52352F | |
| | S52352G,S52352H,S52352J,S52352K,S52352M,S52352N,S52352P | |
| | S52352Q,S52352R,S52353D,S52353E,S52353F,S52353G,S52353H | |
| | S52353J,S52353K,S52353M,S52353N,S52353P,S52353Q,S52353R | |

Continued on next page

**Supplementary Table S1:** (cont'd) A comprehensive mapping of diagnosis groups used in the experiments to their corresponding ICD-10 and ICD-9 codes.

| Diagnosis | ICD-10 | ICD-9 |
|---|---|---|
| | S52354D,S52354E,S52354F,S52354G,S52354H,S52354J,S52354K | |
| | S52354M,S52354N,S52354P,S52354Q,S52354R,S52355D,S52355E | |
| | S52355F,S52355G,S52355H,S52355J,S52355K,S52355M,S52355N | |
| | S52355P,S52355Q,S52355R,S52356D,S52356E,S52356F,S52356G | |
| | S52356H,S52356J,S52356K,S52356M,S52356N,S52356P,S52356Q | |
| | S52356R,S52361D,S52361E,S52361F,S52361G,S52361H,S52361J | |
| | S52361K,S52361M,S52361N,S52361P,S52361Q,S52361R,S52362D | |
| | S52362E,S52362F,S52362G,S52362H,S52362J,S52362K,S52362M | |
| | S52362N,S52362P,S52362Q,S52362R,S52363D,S52363E,S52363F | |
| | S52363G,S52363H,S52363J,S52363K,S52363M,S52363N,S52363P | |
| | S52363Q,S52363R,S52364D,S52364E,S52364F,S52364G,S52364H | |
| | S52364J,S52364K,S52364M,S52364N,S52364P,S52364Q,S52364R | |
| | S52365D,S52365E,S52365F,S52365G,S52365H,S52365J,S52365K | |
| | S52365M,S52365N,S52365P,S52365Q,S52365R,S52366D,S52366E | |
| | S52366F,S52366G,S52366H,S52366J,S52366K,S52366M,S52366N | |
| | S52366P,S52366Q,S52366R,S52371D,S52371E,S52371F,S52371G | |
| | S52371H,S52371J,S52371K,S52371M,S52371N,S52371P,S52371Q | |
| | S52371R,S52372D,S52372E,S52372F,S52372G,S52372H,S52372J | |
| | S52372K,S52372M,S52372N,S52372P,S52372Q,S52372R,S52379D | |
| | S52379E,S52379F,S52379G,S52379H,S52379J,S52379K,S52379M | |
| | S52379N,S52379P,S52379Q,S52379R,S52381D,S52381E,S52381F | |
| | S52381G,S52381H,S52381J,S52381K,S52381M,S52381N,S52381P | |
| | S52381Q,S52381R,S52382D,S52382E,S52382F,S52382G,S52382H | |
| | S52382J,S52382K,S52382M,S52382N,S52382P,S52382Q,S52382R | |
| | S52389D,S52389E,S52389F,S52389G,S52389H,S52389J,S52389K | |
| | S52389M,S52389N,S52389P,S52389Q,S52389R,S52391D,S52391E | |
| | S52391F,S52391G,S52391H,S52391J,S52391K,S52391M,S52391N | |
| | S52391P,S52391Q,S52391R,S52392D,S52392E,S52392F,S52392G | |
| | S52392H,S52392J,S52392K,S52392M,S52392N,S52392P,S52392Q | |
| | S52392R,S52399D,S52399E,S52399F,S52399G,S52399H,S52399J | |
| | S52399K,S52399M,S52399N,S52399P,S52399Q,S52399R,S52501D | |
| | S52501E,S52501F,S52501G,S52501H,S52501J,S52501K,S52501M | |
| | S52501N,S52501P,S52501Q,S52501R,S52502D,S52502E,S52502F | |
| | S52502G,S52502H,S52502J,S52502K,S52502M,S52502N,S52502P | |
| | S52502Q,S52502R,S52509D,S52509E,S52509F,S52509G,S52509H | |
| | S52509J,S52509K,S52509M,S52509N,S52509P,S52509Q,S52509R | |
| | S52511D,S52511E,S52511F,S52511G,S52511H,S52511J,S52511K | |
| | S52511M,S52511N,S52511P,S52511Q,S52511R,S52512D,S52512E | |
| | S52512F,S52512G,S52512H,S52512J,S52512K,S52512M,S52512N | |
| | S52512P,S52512Q,S52512R,S52513D,S52513E,S52513F,S52513G | |

Continued on next page

**Supplementary Table S1:** (cont'd) A comprehensive mapping of diagnosis groups used in the experiments to their corresponding ICD-10 and ICD-9 codes.

| Diagnosis | ICD-10 | ICD-9 |
|---|---|---|
| | S52513H,S52513J,S52513K,S52513M,S52513N,S52513P,S52513Q | |
| | S52513R,S52514D,S52514E,S52514F,S52514G,S52514H,S52514J | |
| | S52514K,S52514M,S52514N,S52514P,S52514Q,S52514R,S52515D | |
| | S52515E,S52515F,S52515G,S52515H,S52515J,S52515K,S52515M | |
| | S52515N,S52515P,S52515Q,S52515R,S52516D,S52516E,S52516F | |
| | S52516G,S52516H,S52516J,S52516K,S52516M,S52516N,S52516P | |
| | S52516Q,S52516R,S52521D,S52521G,S52521K,S52521P,S52522D | |
| | S52522G,S52522K,S52522P,S52529D,S52529G,S52529K,S52529P | |
| | S52531D,S52531E,S52531F,S52531G,S52531H,S52531J,S52531K | |
| | S52531M,S52531N,S52531P,S52531Q,S52531R,S52532D,S52532E | |
| | S52532F,S52532G,S52532H,S52532J,S52532K,S52532M,S52532N | |
| | S52532P,S52532Q,S52532R,S52539D,S52539E,S52539F,S52539G | |
| | S52539H,S52539J,S52539K,S52539M,S52539N,S52539P,S52539Q | |
| | S52539R,S52541D,S52541E,S52541F,S52541G,S52541H,S52541J | |
| | S52541K,S52541M,S52541N,S52541P,S52541Q,S52541R,S52542D | |
| | S52542E,S52542F,S52542G,S52542H,S52542J,S52542K,S52542M | |
| | S52542N,S52542P,S52542Q,S52542R,S52549D,S52549E,S52549F | |
| | S52549G,S52549H,S52549J,S52549K,S52549M,S52549N,S52549P | |
| | S52549Q,S52549R,S52551D,S52551E,S52551F,S52551G,S52551H | |
| | S52551J,S52551K,S52551M,S52551N,S52551P,S52551Q,S52551R | |
| | S52552D,S52552E,S52552F,S52552G,S52552H,S52552J,S52552K | |
| | S52552M,S52552N,S52552P,S52552Q,S52552R,S52559D,S52559E | |
| | S52559F,S52559G,S52559H,S52559J,S52559K,S52559M,S52559N | |
| | S52559P,S52559Q,S52559R,S52561D,S52561E,S52561F,S52561G | |
| | S52561H,S52561J,S52561K,S52561M,S52561N,S52561P,S52561Q | |
| | S52561R,S52562D,S52562E,S52562F,S52562G,S52562H,S52562J | |
| | S52562K,S52562M,S52562N,S52562P,S52562Q,S52562R,S52569D | |
| | S52569E,S52569F,S52569G,S52569H,S52569J,S52569K,S52569M | |
| | S52569N,S52569P,S52569Q,S52569R,S52571D,S52571E,S52571F | |
| | S52571G,S52571H,S52571J,S52571K,S52571M,S52571N,S52571P | |
| | S52571Q,S52571R,S52572D,S52572E,S52572F,S52572G,S52572H | |
| | S52572J,S52572K,S52572M,S52572N,S52572P,S52572Q,S52572R | |
| | S52579D,S52579E,S52579F,S52579G,S52579H,S52579J,S52579K | |
| | S52579M,S52579N,S52579P,S52579Q,S52579R,S52591D,S52591E | |
| | S52591F,S52591G,S52591H,S52591J,S52591K,S52591M,S52591N | |
| | S52591P,S52591Q,S52591R,S52592D,S52592E,S52592F,S52592G | |
| | S52592H,S52592J,S52592K,S52592M,S52592N,S52592P,S52592Q | |
| | S52592R,S52599D,S52599E,S52599F,S52599G,S52599H,S52599J | |
| | S52599K,S52599M,S52599N,S52599P,S52599Q,S52599R,S52601D | |
| | S52601E,S52601F,S52601G,S52601H,S52601J,S52601K,S52601M | |

Continued on next page

**Supplementary Table S1:** (cont'd) A comprehensive mapping of diagnosis groups used in the experiments to their corresponding ICD-10 and ICD-9 codes.

| Diagnosis | ICD-10 | ICD-9 |
|---|---|---|
| | S52601N,S52601P,S52601Q,S52601R,S52602D,S52602E,S52602F | |
| | S52602G,S52602H,S52602J,S52602K,S52602M,S52602N,S52602P | |
| | S52602Q,S52602R,S52609D,S52609E,S52609F,S52609G,S52609H | |
| | S52609J,S52609K,S52609M,S52609N,S52609P,S52609Q,S52609R | |
| | S52611D,S52611E,S52611F,S52611G,S52611H,S52611J,S52611K | |
| | S52611M,S52611N,S52611P,S52611Q,S52611R,S52612D,S52612E | |
| | S52612F,S52612G,S52612H,S52612J,S52612K,S52612M,S52612N | |
| | S52612P,S52612Q,S52612R,S52613D,S52613E,S52613F,S52613G | |
| | S52613H,S52613J,S52613K,S52613M,S52613N,S52613P,S52613Q | |
| | S52613R,S52614D,S52614E,S52614F,S52614G,S52614H,S52614J | |
| | S52614K,S52614M,S52614N,S52614P,S52614Q,S52614R,S52615D | |
| | S52615E,S52615F,S52615G,S52615H,S52615J,S52615K,S52615M | |
| | S52615N,S52615P,S52615Q,S52615R,S52616D,S52616E,S52616F | |
| | S52616G,S52616H,S52616J,S52616K,S52616M,S52616N,S52616P | |
| | S52616Q,S52616R,S52621D,S52621G,S52621K,S52621P,S52622D | |
| | S52622G,S52622K,S52622P,S52629D,S52629G,S52629K,S52629P | |
| | S52691D,S52691E,S52691F,S52691G,S52691H,S52691J,S52691K | |
| | S52691M,S52691N,S52691P,S52691Q,S52691R,S52692D,S52692E | |
| | S52692F,S52692G,S52692H,S52692J,S52692K,S52692M,S52692N | |
| | S52692P,S52692Q,S52692R,S52699D,S52699E,S52699F,S52699G | |
| | S52699H,S52699J,S52699K,S52699M,S52699N,S52699P,S52699Q | |
| | S52699R,S5290XD,S5290XE,S5290XF,S5290XG,S5290XH,S5290XJ | |
| | S5290XK,S5290XM,S5290XN,S5290XP,S5290XQ,S5290XR,S5291XD | |
| | S5291XE,S5291XF,S5291XG,S5291XH,S5291XJ,S5291XK,S5291XM | |
| | S5291XN,S5291XP,S5291XQ,S5291XR,S5292XD,S5292XE,S5292XF | |
| | S5292XG,S5292XH,S5292XJ,S5292XK,S5292XM,S5292XN,S5292XP | |
| | S5292XQ,S5292XR,S59001D,S59001G,S59001K,S59001P,S59002D | |
| | S59002G,S59002K,S59002P,S59009D,S59009G,S59009K,S59009P | |
| | S59011D,S59011G,S59011K,S59011P,S59012D,S59012G,S59012K | |
| | S59012P,S59019D,S59019G,S59019K,S59019P,S59021D,S59021G | |
| | S59021K,S59021P,S59022D,S59022G,S59022K,S59022P,S59029D | |
| | S59029G,S59029K,S59029P,S59031D,S59031G,S59031K,S59031P | |
| | S59032D,S59032G,S59032K,S59032P,S59039D,S59039G,S59039K | |
| | S59039P,S59041D,S59041G,S59041K,S59041P,S59042D,S59042G | |
| | S59042K,S59042P,S59049D,S59049G,S59049K,S59049P,S59091D | |
| | S59091G,S59091K,S59091P,S59092D,S59092G,S59092K,S59092P | |
| | S59099D,S59099G,S59099K,S59099P,S59101D,S59101G,S59101K | |
| | S59101P,S59102D,S59102G,S59102K,S59102P,S59109D,S59109G | |
| | S59109K,S59109P,S59111D,S59111G,S59111K,S59111P,S59112D | |
| | S59112G,S59112K,S59112P,S59119D,S59119G,S59119K,S59119P | |

Continued on next page

**Supplementary Table S1:** (cont'd) A comprehensive mapping of diagnosis groups used in the experiments to their corresponding ICD-10 and ICD-9 codes.

| Diagnosis | ICD-10 | ICD-9 |
|---|---|---|
| | S59121D,S59121G,S59121K,S59121P,S59122D,S59122G,S59122K | |
| | S59122P,S59129D,S59129G,S59129K,S59129P,S59131D,S59131G | |
| | S59131K,S59131P,S59132D,S59132G,S59132K,S59132P,S59139D | |
| | S59139G,S59139K,S59139P,S59141D,S59141G,S59141K,S59141P | |
| | S59142D,S59142G,S59142K,S59142P,S59149D,S59149G,S59149K | |
| | S59149P,S59191D,S59191G,S59191K,S59191P,S59192D,S59192G | |
| | S59192K,S59192P,S59199D,S59199G,S59199K,S59199P,S59201D | |
| | S59201G,S59201K,S59201P,S59202D,S59202G,S59202K,S59202P | |
| | S59209D,S59209G,S59209K,S59209P,S59211D,S59211G,S59211K | |
| | S59211P,S59212D,S59212G,S59212K,S59212P,S59219D,S59219G | |
| | S59219K,S59219P,S59221D,S59221G,S59221K,S59221P,S59222D | |
| | S59222G,S59222K,S59222P,S59229D,S59229G,S59229K,S59229P | |
| | S59231D,S59231G,S59231K,S59231P,S59232D,S59232G,S59232K | |
| | S59232P,S59239D,S59239G,S59239K,S59239P,S59241D,S59241G | |
| | S59241K,S59241P,S59242D,S59242G,S59242K,S59242P,S59249D | |
| | S59249G,S59249K,S59249P,S59291D,S59291G,S59291K,S59291P | |
| | S59292D,S59292G,S59292K,S59292P,S59299D,S59299G,S59299K | |
| | S59299P,S62001D,S62001G,S62001K,S62001P,S62002D,S62002G | |
| | S62002K,S62002P,S62009D,S62009G,S62009K,S62009P,S62011D | |
| | S62011G,S62011K,S62011P,S62012D,S62012G,S62012K,S62012P | |
| | S62013D,S62013G,S62013K,S62013P,S62014D,S62014G,S62014K | |
| | S62014P,S62015D,S62015G,S62015K,S62015P,S62016D,S62016G | |
| | S62016K,S62016P,S62021D,S62021G,S62021K,S62021P,S62022D | |
| | S62022G,S62022K,S62022P,S62023D,S62023G,S62023K,S62023P | |
| | S62024D,S62024G,S62024K,S62024P,S62025D,S62025G,S62025K | |
| | S62025P,S62026D,S62026G,S62026K,S62026P,S62031D,S62031G | |
| | S62031K,S62031P,S62032D,S62032G,S62032K,S62032P,S62033D | |
| | S62033G,S62033K,S62033P,S62034D,S62034G,S62034K,S62034P | |
| | S62035D,S62035G,S62035K,S62035P,S62036D,S62036G,S62036K | |
| | S62036P,S62101D,S62101G,S62101K,S62101P,S62102D,S62102G | |
| | S62102K,S62102P,S62109D,S62109G,S62109K,S62109P,S62111D | |
| | S62111G,S62111K,S62111P,S62112D,S62112G,S62112K,S62112P | |
| | S62113D,S62113G,S62113K,S62113P,S62114D,S62114G,S62114K | |
| | S62114P,S62115D,S62115G,S62115K,S62115P,S62116D,S62116G | |
| | S62116K,S62116P,S62121D,S62121G,S62121K,S62121P,S62122D | |
| | S62122G,S62122K,S62122P,S62123D,S62123G,S62123K,S62123P | |
| | S62124D,S62124G,S62124K,S62124P,S62125D,S62125G,S62125K | |
| | S62125P,S62126D,S62126G,S62126K,S62126P,S62131D,S62131G | |
| | S62131K,S62131P,S62132D,S62132G,S62132K,S62132P,S62133D | |
| | S62133G,S62133K,S62133P,S62134D,S62134G,S62134K,S62134P | |

Continued on next page

**Supplementary Table S1:** (cont'd) A comprehensive mapping of diagnosis groups used in the experiments to their corresponding ICD-10 and ICD-9 codes.

| Diagnosis | ICD-10 | ICD-9 |
|---|---|---|
| | S62135D,S62135G,S62135K,S62135P,S62136D,S62136G,S62136K | |
| | S62136P,S62141D,S62141G,S62141K,S62141P,S62142D,S62142G | |
| | S62142K,S62142P,S62143D,S62143G,S62143K,S62143P,S62144D | |
| | S62144G,S62144K,S62144P,S62145D,S62145G,S62145K,S62145P | |
| | S62146D,S62146G,S62146K,S62146P,S62151D,S62151G,S62151K | |
| | S62151P,S62152D,S62152G,S62152K,S62152P,S62153D,S62153G | |
| | S62153K,S62153P,S62154D,S62154G,S62154K,S62154P,S62155D | |
| | S62155G,S62155K,S62155P,S62156D,S62156G,S62156K,S62156P | |
| | S62161D,S62161G,S62161K,S62161P,S62162D,S62162G,S62162K | |
| | S62162P,S62163D,S62163G,S62163K,S62163P,S62164D,S62164G | |
| | S62164K,S62164P,S62165D,S62165G,S62165K,S62165P,S62166D | |
| | S62166G,S62166K,S62166P,S62171D,S62171G,S62171K,S62171P | |
| | S62172D,S62172G,S62172K,S62172P,S62173D,S62173G,S62173K | |
| | S62173P,S62174D,S62174G,S62174K,S62174P,S62175D,S62175G | |
| | S62175K,S62175P,S62176D,S62176G,S62176K,S62176P,S62181D | |
| | S62181G,S62181K,S62181P,S62182D,S62182G,S62182K,S62182P | |
| | S62183D,S62183G,S62183K,S62183P,S62184D,S62184G,S62184K | |
| | S62184P,S62185D,S62185G,S62185K,S62185P,S62186D,S62186G | |
| | S62186K,S62186P,S62201D,S62201G,S62201K,S62201P,S62202D | |
| | S62202G,S62202K,S62202P,S62209D,S62209G,S62209K,S62209P | |
| | S62211D,S62211G,S62211K,S62211P,S62212D,S62212G,S62212K | |
| | S62212P,S62213D,S62213G,S62213K,S62213P,S62221D,S62221G | |
| | S62221K,S62221P,S62222D,S62222G,S62222K,S62222P,S62223D | |
| | S62223G,S62223K,S62223P,S62224D,S62224G,S62224K,S62224P | |
| | S62225D,S62225G,S62225K,S62225P,S62226D,S62226G,S62226K | |
| | S62226P,S62231D,S62231G,S62231K,S62231P,S62232D,S62232G | |
| | S62232K,S62232P,S62233D,S62233G,S62233K,S62233P,S62234D | |
| | S62234G,S62234K,S62234P,S62235D,S62235G,S62235K,S62235P | |
| | S62236D,S62236G,S62236K,S62236P,S62241D,S62241G,S62241K | |
| | S62241P,S62242D,S62242G,S62242K,S62242P,S62243D,S62243G | |
| | S62243K,S62243P,S62244D,S62244G,S62244K,S62244P,S62245D | |
| | S62245G,S62245K,S62245P,S62246D,S62246G,S62246K,S62246P | |
| | S62251D,S62251G,S62251K,S62251P,S62252D,S62252G,S62252K | |
| | S62252P,S62253D,S62253G,S62253K,S62253P,S62254D,S62254G | |
| | S62254K,S62254P,S62255D,S62255G,S62255K,S62255P,S62256D | |
| | S62256G,S62256K,S62256P,S62291D,S62291G,S62291K,S62291P | |
| | S62292D,S62292G,S62292K,S62292P,S62299D,S62299G,S62299K | |
| | S62299P,S62300D,S62300G,S62300K,S62300P,S62301D,S62301G | |
| | S62301K,S62301P,S62302D,S62302G,S62302K,S62302P,S62303D | |
| | S62303G,S62303K,S62303P,S62304D,S62304G,S62304K,S62304P | |

Continued on next page

**Supplementary Table S1:** (cont'd) A comprehensive mapping of diagnosis groups used in the experiments to their corresponding ICD-10 and ICD-9 codes.

| Diagnosis | ICD-10 | ICD-9 |
|---|---|---|
| | S62305D,S62305G,S62305K,S62305P,S62306D,S62306G,S62306K | |
| | S62306P,S62307D,S62307G,S62307K,S62307P,S62308D,S62308G | |
| | S62308K,S62308P,S62309D,S62309G,S62309K,S62309P,S62310D | |
| | S62310G,S62310K,S62310P,S62311D,S62311G,S62311K,S62311P | |
| | S62312D,S62312G,S62312K,S62312P,S62313D,S62313G,S62313K | |
| | S62313P,S62314D,S62314G,S62314K,S62314P,S62315D,S62315G | |
| | S62315K,S62315P,S62316D,S62316G,S62316K,S62316P,S62317D | |
| | S62317G,S62317K,S62317P,S62318D,S62318G,S62318K,S62318P | |
| | S62319D,S62319G,S62319K,S62319P,S62320D,S62320G,S62320K | |
| | S62320P,S62321D,S62321G,S62321K,S62321P,S62322D,S62322G | |
| | S62322K,S62322P,S62323D,S62323G,S62323K,S62323P,S62324D | |
| | S62324G,S62324K,S62324P,S62325D,S62325G,S62325K,S62325P | |
| | S62326D,S62326G,S62326K,S62326P,S62327D,S62327G,S62327K | |
| | S62327P,S62328D,S62328G,S62328K,S62328P,S62329D,S62329G | |
| | S62329K,S62329P,S62330D,S62330G,S62330K,S62330P,S62331D | |
| | S62331G,S62331K,S62331P,S62332D,S62332G,S62332K,S62332P | |
| | S62333D,S62333G,S62333K,S62333P,S62334D,S62334G,S62334K | |
| | S62334P,S62335D,S62335G,S62335K,S62335P,S62336D,S62336G | |
| | S62336K,S62336P,S62337D,S62337G,S62337K,S62337P,S62338D | |
| | S62338G,S62338K,S62338P,S62339D,S62339G,S62339K,S62339P | |
| | S62340D,S62340G,S62340K,S62340P,S62341D,S62341G,S62341K | |
| | S62341P,S62342D,S62342G,S62342K,S62342P,S62343D,S62343G | |
| | S62343K,S62343P,S62344D,S62344G,S62344K,S62344P,S62345D | |
| | S62345G,S62345K,S62345P,S62346D,S62346G,S62346K,S62346P | |
| | S62347D,S62347G,S62347K,S62347P,S62348D,S62348G,S62348K | |
| | S62348P,S62349D,S62349G,S62349K,S62349P,S62350D,S62350G | |
| | S62350K,S62350P,S62351D,S62351G,S62351K,S62351P,S62352D | |
| | S62352G,S62352K,S62352P,S62353D,S62353G,S62353K,S62353P | |
| | S62354D,S62354G,S62354K,S62354P,S62355D,S62355G,S62355K | |
| | S62355P,S62356D,S62356G,S62356K,S62356P,S62357D,S62357G | |
| | S62357K,S62357P,S62358D,S62358G,S62358K,S62358P,S62359D | |
| | S62359G,S62359K,S62359P,S62360D,S62360G,S62360K,S62360P | |
| | S62361D,S62361G,S62361K,S62361P,S62362D,S62362G,S62362K | |
| | S62362P,S62363D,S62363G,S62363K,S62363P,S62364D,S62364G | |
| | S62364K,S62364P,S62365D,S62365G,S62365K,S62365P,S62366D | |
| | S62366G,S62366K,S62366P,S62367D,S62367G,S62367K,S62367P | |
| | S62368D,S62368G,S62368K,S62368P,S62369D,S62369G,S62369K | |
| | S62369P,S62390D,S62390G,S62390K,S62390P,S62391D,S62391G | |
| | S62391K,S62391P,S62392D,S62392G,S62392K,S62392P,S62393D | |
| | S62393G,S62393K,S62393P,S62394D,S62394G,S62394K,S62394P | |

Continued on next page

**Supplementary Table S1:** (cont'd) A comprehensive mapping of diagnosis groups used in the experiments to their corresponding ICD-10 and ICD-9 codes.

| Diagnosis | ICD-10 | ICD-9 |
|---|---|---|
| | S62395D,S62395G,S62395K,S62395P,S62396D,S62396G,S62396K<br>S62396P,S62397D,S62397G,S62397K,S62397P,S62398D,S62398G<br>S62398K,S62398P,S62399D,S62399G,S62399K,S62399P,S62501D<br>S62501G,S62501K,S62501P,S62502D,S62502G,S62502K,S62502P<br>S62509D,S62509G,S62509K,S62509P,S62511D,S62511G,S62511K<br>S62511P,S62512D,S62512G,S62512K,S62512P,S62513D,S62513G<br>S62513K,S62513P,S62514D,S62514G,S62514K,S62514P,S62515D<br>S62515G,S62515K,S62515P,S62516D,S62516G,S62516K,S62516P<br>S62521D,S62521G,S62521K,S62521P,S62522D,S62522G,S62522K<br>S62522P,S62523D,S62523G,S62523K,S62523P,S62524D,S62524G<br>S62524K,S62524P,S62525D,S62525G,S62525K,S62525P,S62526D<br>S62526G,S62526K,S62526P,S62600D,S62600G,S62600K,S62600P<br>S62601D,S62601G,S62601K,S62601P,S62602D,S62602G,S62602K<br>S62602P,S62603D,S62603G,S62603K,S62603P,S62604D,S62604G<br>S62604K,S62604P,S62605D,S62605G,S62605K,S62605P,S62606D<br>S62606G,S62606K,S62606P,S62607D,S62607G,S62607K,S62607P<br>S62608D,S62608G,S62608K,S62608P,S62609D,S62609G,S62609K<br>S62609P,S62610D,S62610G,S62610K,S62610P,S62611D,S62611G<br>S62611K,S62611P,S62612D,S62612G,S62612K,S62612P,S62613D<br>S62613G,S62613K,S62613P,S62614D,S62614G,S62614K,S62614P<br>S62615D,S62615G,S62615K,S62615P,S62616D,S62616G,S62616K<br>S62616P,S62617D,S62617G,S62617K,S62617P,S62618D,S62618G<br>S62618K,S62618P,S62619D,S62619G,S62619K,S62619P,S62620D<br>S62620G,S62620K,S62620P,S62621D,S62621G,S62621K,S62621P<br>S62622D,S62622G,S62622K,S62622P,S62623D,S62623G,S62623K<br>S62623P,S62624D,S62624G,S62624K,S62624P,S62625D,S62625G<br>S62625K,S62625P,S62626D,S62626G,S62626K,S62626P,S62627D<br>S62627G,S62627K,S62627P,S62628D,S62628G,S62628K,S62628P<br>S62629D,S62629G,S62629K,S62629P,S62630D,S62630G,S62630K<br>S62630P,S62631D,S62631G,S62631K,S62631P,S62632D,S62632G<br>S62632K,S62632P,S62633D,S62633G,S62633K,S62633P,S62634D<br>S62634G,S62634K,S62634P,S62635D,S62635G,S62635K,S62635P<br>S62636D,S62636G,S62636K,S62636P,S62637D,S62637G,S62637K<br>S62637P,S62638D,S62638G,S62638K,S62638P,S62639D,S62639G<br>S62639K,S62639P,S62640D,S62640G,S62640K,S62640P,S62641D<br>S62641G,S62641K,S62641P,S62642D,S62642G,S62642K,S62642P<br>S62643D,S62643G,S62643K,S62643P,S62644D,S62644G,S62644K<br>S62644P,S62645D,S62645G,S62645K,S62645P,S62646D,S62646G<br>S62646K,S62646P,S62647D,S62647G,S62647K,S62647P,S62648D<br>S62648G,S62648K,S62648P,S62649D,S62649G,S62649K,S62649P | |

Continued on next page

**Supplementary Table S1:** (cont'd) A comprehensive mapping of diagnosis groups used in the experiments to their corresponding ICD-10 and ICD-9 codes.

| Diagnosis | ICD-10 | ICD-9 |
|---|---|---|
| | S62650D,S62650G,S62650K,S62650P,S62651D,S62651G,S62651K | |
| | S62651P,S62652D,S62652G,S62652K,S62652P,S62653D,S62653G | |
| | S62653K,S62653P,S62654D,S62654G,S62654K,S62654P,S62655D | |
| | S62655G,S62655K,S62655P,S62656D,S62656G,S62656K,S62656P | |
| | S62657D,S62657G,S62657K,S62657P,S62658D,S62658G,S62658K | |
| | S62658P,S62659D,S62659G,S62659K,S62659P,S62660D,S62660G | |
| | S62660K,S62660P,S62661D,S62661G,S62661K,S62661P,S62662D | |
| | S62662G,S62662K,S62662P,S62663D,S62663G,S62663K,S62663P | |
| | S62664D,S62664G,S62664K,S62664P,S62665D,S62665G,S62665K | |
| | S62665P,S62666D,S62666G,S62666K,S62666P,S62667D,S62667G | |
| | S62667K,S62667P,S62668D,S62668G,S62668K,S62668P,S62669D | |
| | S62669G,S62669K,S62669P,S6290XD,S6290XG,S6290XK,S6290XP | |
| | S6291XD,S6291XG,S6291XK,S6291XP,S6292XD,S6292XG,S6292XK | |
| | S6292XP | |
| Fracture of torso, | M96A1,M96A2,M96A3,M96A4,M96A9,S2220XA,S2220XB,S2221XA,S2221XB | 80700,80701,80702,80703,80704,80705,80706,80707,80708,80709 |
| initial encounter | S2222XA,S2222XB,S2223XA,S2223XB,S2224XA,S2224XB,S2231XA | 80711,80712,80713,80714,80715,80716,80717,80718,80719,8072 |
| | S2231XB,S2232XA,S2232XB,S2239XA,S2239XB,S2241XA,S2241XB | 8073,8074,8080,8081,8082,8083,80841,80842,80843,80844,80849 |
| | S2242XA,S2242XB,S2243XA,S2243XB,S2249XA,S2249XB,S225XXA | 80851,80852,80853,80854,80859,8090,8091 |
| | S225XXB,S229XXA,S229XXB,S32301A,S32301B,S32302A,S32302B | |
| | S32309A,S32309B,S32311A,S32311B,S32312A,S32312B,S32313A | |
| | S32313B,S32314A,S32314B,S32315A,S32315B,S32316A,S32316B | |
| | S32391A,S32391B,S32392A,S32392B,S32399A,S32399B,S32401A | |
| | S32401B,S32402A,S32402B,S32409A,S32409B,S32411A,S32411B | |
| | S32412A,S32412B,S32413A,S32413B,S32414A,S32414B,S32415A | |
| | S32415B,S32416A,S32416B,S32421A,S32421B,S32422A,S32422B | |
| | S32423A,S32423B,S32424A,S32424B,S32425A,S32425B,S32426A | |
| | S32426B,S32431A,S32431B,S32432A,S32432B,S32433A,S32433B | |
| | S32434A,S32434B,S32435A,S32435B,S32436A,S32436B,S32441A | |
| | S32441B,S32442A,S32442B,S32443A,S32443B,S32444A,S32444B | |
| | S32445A,S32445B,S32446A,S32446B,S32451A,S32451B,S32452A | |
| | S32452B,S32453A,S32453B,S32454A,S32454B,S32455A,S32455B | |
| | S32456A,S32456B,S32461A,S32461B,S32462A,S32462B,S32463A | |
| | S32463B,S32464A,S32464B,S32465A,S32465B,S32466A,S32466B | |
| | S32471A,S32471B,S32472A,S32472B,S32473A,S32473B,S32474A | |
| | S32474B,S32475A,S32475B,S32476A,S32476B,S32481A,S32481B | |
| | S32482A,S32482B,S32483A,S32483B,S32484A,S32484B,S32485A | |
| | S32485B,S32486A,S32486B,S32491A,S32491B,S32492A,S32492B | |
| | S32499A,S32499B,S32501A,S32501B,S32502A,S32502B,S32509A | |
| | S32509B,S32511A,S32511B,S32512A,S32512B,S32519A,S32519B | |

Continued on next page

**Supplementary Table S1:** (cont'd) A comprehensive mapping of diagnosis groups used in the experiments to their corresponding ICD-10 and ICD-9 codes.

| Diagnosis | ICD-10 | ICD-9 |
|---|---|---|
| | S32591A,S32591B,S32592A,S32592B,S32599A,S32599B,S32601A | |
| | S32601B,S32602A,S32602B,S32609A,S32609B,S32611A,S32611B | |
| | S32612A,S32612B,S32613A,S32613B,S32614A,S32614B,S32615A | |
| | S32615B,S32616A,S32616B,S32691A,S32691B,S32692A,S32692B | |
| | S32699A,S32699B,S32810A,S32810B,S32811A,S32811B,S3282XA | |
| | S3282XB,S3289XA,S3289XB | |
| Fracture of torso, | S2220XD,S2220XG,S2220XK,S2221XD,S2221XG,S2221XK,S2222XD | 73382,V5413,V5419 |
| subsequent encounter | S2222XG,S2222XK,S2223XD,S2223XG,S2223XK,S2224XD,S2224XG | |
| | S2224XK,S2231XD,S2231XG,S2231XK,S2232XD,S2232XG,S2232XK | |
| | S2239XD,S2239XG,S2239XK,S2241XD,S2241XG,S2241XK,S2242XD | |
| | S2242XG,S2242XK,S2243XD,S2243XG,S2243XK,S2249XD,S2249XG | |
| | S2249XK,S225XXD,S225XXG,S225XXK,S229XXD,S229XXG,S229XXK | |
| | S32301D,S32301G,S32301K,S32302D,S32302G,S32302K,S32309D | |
| | S32309G,S32309K,S32311D,S32311G,S32311K,S32312D,S32312G | |
| | S32312K,S32313D,S32313G,S32313K,S32314D,S32314G,S32314K | |
| | S32315D,S32315G,S32315K,S32316D,S32316G,S32316K,S32391D | |
| | S32391G,S32391K,S32392D,S32392G,S32392K,S32399D,S32399G | |
| | S32399K,S32401D,S32401G,S32401K,S32402D,S32402G,S32402K | |
| | S32409D,S32409G,S32409K,S32411D,S32411G,S32411K,S32412D | |
| | S32412G,S32412K,S32413D,S32413G,S32413K,S32414D,S32414G | |
| | S32414K,S32415D,S32415G,S32415K,S32416D,S32416G,S32416K | |
| | S32421D,S32421G,S32421K,S32422D,S32422G,S32422K,S32423D | |
| | S32423G,S32423K,S32424D,S32424G,S32424K,S32425D,S32425G | |
| | S32425K,S32426D,S32426G,S32426K,S32431D,S32431G,S32431K | |
| | S32432D,S32432G,S32432K,S32433D,S32433G,S32433K,S32434D | |
| | S32434G,S32434K,S32435D,S32435G,S32435K,S32436D,S32436G | |
| | S32436K,S32441D,S32441G,S32441K,S32442D,S32442G,S32442K | |
| | S32443D,S32443G,S32443K,S32444D,S32444G,S32444K,S32445D | |
| | S32445G,S32445K,S32446D,S32446G,S32446K,S32451D,S32451G | |
| | S32451K,S32452D,S32452G,S32452K,S32453D,S32453G,S32453K | |
| | S32454D,S32454G,S32454K,S32455D,S32455G,S32455K,S32456D | |
| | S32456G,S32456K,S32461D,S32461G,S32461K,S32462D,S32462G | |
| | S32462K,S32463D,S32463G,S32463K,S32464D,S32464G,S32464K | |
| | S32465D,S32465G,S32465K,S32466D,S32466G,S32466K,S32471D | |
| | S32471G,S32471K,S32472D,S32472G,S32472K,S32473D,S32473G | |
| | S32473K,S32474D,S32474G,S32474K,S32475D,S32475G,S32475K | |
| | S32476D,S32476G,S32476K,S32481D,S32481G,S32481K,S32482D | |
| | S32482G,S32482K,S32483D,S32483G,S32483K,S32484D,S32484G | |
| | S32484K,S32485D,S32485G,S32485K,S32486D,S32486G,S32486K | |

Continued on next page

**Supplementary Table S1:** (cont'd) A comprehensive mapping of diagnosis groups used in the experiments to their corresponding ICD-10 and ICD-9 codes.

| Diagnosis | ICD-10 | ICD-9 |
|---|---|---|
| | S32491D,S32491G,S32491K,S32492D,S32492G,S32492K,S32499D<br>S32499G,S32499K,S32501D,S32501G,S32501K,S32502D,S32502G<br>S32502K,S32509D,S32509G,S32509K,S32511D,S32511G,S32511K<br>S32512D,S32512G,S32512K,S32519D,S32519G,S32519K,S32591D<br>S32591G,S32591K,S32592D,S32592G,S32592K,S32599D,S32599G<br>S32599K,S32601D,S32601G,S32601K,S32602D,S32602G,S32602K<br>S32609D,S32609G,S32609K,S32611D,S32611G,S32611K,S32612D<br>S32612G,S32612K,S32613D,S32613G,S32613K,S32614D,S32614G<br>S32614K,S32615D,S32615G,S32615K,S32616D,S32616G,S32616K<br>S32691D,S32691G,S32691K,S32692D,S32692G,S32692K,S32699D<br>S32699G,S32699K,S32810D,S32810G,S32810K,S32811D,S32811G<br>S32811K,S3282XD,S3282XG,S3282XK,S3289XD,S3289XG,S3289XK | |
| Fungal infections | B350,B351,B352,B353,B354,B355,B356,B358,B359,B360,B361,B362<br>B363,B368,B369,B370,B371,B372,B373,B3731,B3732,B3741,B3742<br>B3749,B375,B376,B377,B3781,B3782,B3783,B3784,B3789,B379,B380<br>B381,B382,B383,B384,B387,B3881,B3889,B389,B390,B391,B392,B393<br>B394,B395,B399,B400,B401,B402,B403,B407,B4081,B4089,B409,B410<br>B417,B418,B419,B420,B421,B427,B4281,B4282,B4289,B429,B430<br>B431,B432,B438,B439,B440,B441,B442,B447,B4481,B4489,B449,B450<br>B451,B452,B453,B457,B458,B459,B460,B461,B462,B463,B464,B465<br>B468,B469,B470,B471,B479,B480,B481,B482,B483,B484,B488,B49<br>B59,P375 | 0394,0399,1100,1101,1102,1103,1104,1105,1106,1108,1109,1110<br>1111,1112,1113,1118,1119,1120,1121,1122,1123,1124,1125,11281<br>11282,11283,11284,11285,11289,1129,1140,1141,1142,1143,1144<br>1145,1149,11500,11505,11509,11510,11590,1160,1161,1162,1170<br>1171,1172,1173,1174,1175,1176,1177,1178,1179,118,1363,3210<br>4846,5186,7717,99591 |
| Gangrene | E0852,E0952,E1052,E1152,E1352,I70261,I70262,I70263,I70268<br>I70269,I70361,I70362,I70363,I70368,I70369,I70461,I70462<br>I70463,I70468,I70469,I70561,I70562,I70563,I70568,I70569<br>I70661,I70662,I70663,I70668,I70669,I70761,I70762,I70763<br>I70768,I70769,I7301,I96,J850,K4010,K4011,K4040,K4041,K4110<br>K4111,K4140,K4141,K421,K431,K434,K437,K441,K451,K461,K82A1<br>N493 | 24970,25070,25071,44024,44030,44031,44032,4430,44381,5130<br>55000,55001,55002,55003,55100,55101,55102,55103,5511,55120<br>55121,55129,5513,5518,5519,6084,7854 |
| Gastritis and duodenitis | K2900,K2901,K2920,K2921,K2930,K2931,K2940,K2941,K2950,K2951<br>K2960,K2961,K2970,K2971,K2980,K2981,K2990,K2991 | 53500,53501,53510,53511,53520,53521,53530,53531,53540,53541<br>53550,53551,53560,53561 |
| Gastroduodenal ulcer | K251,K253,K255,K257,K259,K261,K263,K265,K267,K269,K271,K273<br>K275,K277,K279,K281,K283,K285,K287,K289 | 53110,53130,53150,53170,53190,53210,53230,53250,53270,53290<br>53310,53330,53350,53370,53390,53410,53430,53450,53470,53490 |
| Gastrointestinal and biliary perforation | K223,K251,K255,K261,K265,K271,K275,K281,K285,K3532,K3533<br>K5700,K5701,K5720,K5721,K5740,K5741,K5780,K5781,K631,K822 | 5304,53110,53150,53210,53250,53310,53350,53410,53450,56201<br>56203,56211,56213,5695,56983,5754,5763 |

Continued on next page

**Supplementary Table S1:** (cont'd) A comprehensive mapping of diagnosis groups used in the experiments to their corresponding ICD-10 and ICD-9 codes.

| Diagnosis | ICD-10 | ICD-9 |
|---|---|---|
| | K82A2,K832 | |
| Gastrointestinal cancers - all other types | C240,C241,C248,C249,C261,C269,C480,C488,C49A0,C49A9,D017,D019 | 1561,1562,1568,1569,1580,1588,1591,1598,1599,2309 |
| Gastrointestinal cancers - anus | C210,C211,C212,C218,D013 | 1542,1543,1548,2305,2306 |
| Gastrointestinal cancers - bile duct | C221 | 1551 |
| Gastrointestinal cancers - colorectal | C180,C181,C182,C183,C184,C185,C186,C187,C188,C189,C19,C20<br>C260,C49A4,C49A5,D010,D011,D012,D0140 | 1530,1531,1532,1533,1534,1535,1536,1537,1538,1539,1540,1541<br>1590,2303,2304,2307 |
| Gastrointestinal cancers - esophagus | C153,C154,C155,C158,C159,C49A1,D001 | 1500,1501,1502,1503,1504,1505,1508,1509,2301 |
| Gastrointestinal cancers - gallbladder | C23 | 1560 |
| Gastrointestinal cancers - liver | C220,C222,C223,C224,C227,C228,C229,D015 | 1550,1552,2308 |
| Gastrointestinal cancers - peritoneum | C481,C482,C488 | 1588,1589 |
| Gastrointestinal cancers - small intestine | C170,C171,C172,C173,C178,C179,C49A3,D0149 | 1520,1521,1522,1523,1528,1529,2307 |
| Gastrointestinal cancers - stomach | C160,C161,C162,C163,C164,C165,C166,C168,C169,C49A2,D002 | 1510,1511,1512,1513,1514,1515,1516,1518,1519,2302 |
| Gastrointestinal hemorrhage | I8501,I8511,K2081,K2091,K2101,K2211,K250,K252,K254,K256,K260<br>K262,K264,K266,K270,K272,K274,K276,K280,K282,K284,K286,K2901<br>K2921,K2931,K2941,K2951,K2961,K2971,K2981,K2991,K31811,K50011<br>K50111,K50811,K50911,K51011,K51211,K51311,K51411,K51511<br>K51811,K51911,K5521,K5701,K5711,K5713,K5721,K5731,K5733,K5741<br>K5751,K5753,K5781,K5791,K5793,K625,K920,K921,K922 | 4560,45620,53021,53100,53120,53140,53160,53200,53220,53240<br>53260,53300,53320,53340,53360,53400,53420,53440,53460,53501<br>53511,53521,53531,53541,53551,53561,53783,5550,5551,5552,5559<br>5562,5563,5564,5565,5566,5568,5569,56202,56203,56212,56213<br>5693,5695,56985,5780,5781,5789 |

Continued on next page

**Supplementary Table S1:** (cont'd) A comprehensive mapping of diagnosis groups used in the experiments to their corresponding ICD-10 and ICD-9 codes.

| Diagnosis | ICD-10 | ICD-9 |
|---|---|---|
| General sensation perception signs and symptoms | R42,R440,R441,R442,R443,R448,R449 | 36816,7801,7804,79989 |
| Genitourinary congenital anomalies | Q5001,Q5002,Q501,Q502,Q5031,Q5032,Q5039,Q504,Q505,Q506,Q510 Q5110,Q5111,Q512,Q5120,Q5121,Q5122,Q5128,Q513,Q514,Q515,Q516 Q517,Q51810,Q51811,Q51818,Q51820,Q51821,Q51828,Q519,Q520 Q5210,Q5211,Q5212,Q52120,Q52121,Q52122,Q52123,Q52124,Q52129 Q522,Q523,Q524,Q525,Q526,Q5270,Q5271,Q5279,Q528,Q529,Q5300 Q5301,Q5302,Q5310,Q5311,Q53111,Q53112,Q5312,Q5313,Q5320,Q5321 Q53211,Q53212,Q5322,Q5323,Q539,Q540,Q541,Q542,Q543,Q544,Q548 Q549,Q550,Q551,Q5520,Q5521,Q5522,Q5523,Q5529,Q553,Q554,Q555 Q5561,Q5562,Q5563,Q5564,Q5569,Q557,Q558,Q559,Q560,Q561,Q562 Q563,Q564,Q600,Q601,Q602,Q603,Q604,Q605,Q606,Q6100,Q6101 Q6102,Q6111,Q6119,Q612,Q613,Q614,Q615,Q618,Q619,Q620,Q6210 Q6211,Q6212,Q622,Q6231,Q6232,Q6239,Q624,Q625,Q6260,Q6261 Q6262,Q6263,Q6269,Q627,Q628,Q630,Q631,Q632,Q633,Q638,Q639 Q640,Q6410,Q6411,Q6412,Q6419,Q642,Q6431,Q6432,Q6433,Q6439 Q644,Q645,Q646,Q6470,Q6471,Q6472,Q6473,Q6474,Q6475,Q6479,Q648 Q649 | 7520,75210,75211,75219,7522,75231,75232,75233,75234,75235 75236,75239,75240,75241,75242,75243,75244,75245,75246,75247 75249,75251,75252,75261,75262,75263,75264,75265,75269,7527 75281,75289,7529,7530,75310,75311,75312,75313,75314,75315 75316,75317,75319,75320,75321,75322,75323,75329,7533,7534 7535,7536,7537,7538,7539 |
| Genitourinary signs and symptoms | N398,N399,R300,R301,R309,R330,R338,R339,R34,R350,R351,R358 R3581,R3589,R360,R361,R369,R390,R3911,R3912,R3913,R3914,R3915 R3916,R3919,R39191,R39192,R39198,R392,R3982,R3983,R3984,R3989 R399,R820,R821,R822,R823,R824,R825,R826,R827,R8271,R8279,R828 R8281,R8289,R8290,R8291,R8299,R82991,R82992,R82993,R82994 R82998 | 59989,5999,60882,7881,78820,78821,78829,78841,78842,78843 7885,78861,78862,78863,78864,78865,78869,7887,7888,78899,7911 7912,7913,7914,7916,7917,7919,7960 |
| Glaucoma | H21521,H21522,H21523,H21529,H21531,H21532,H21533,H21539 H21541,H21542,H21543,H21549,H21551,H21552,H21553,H21559 H40001,H40002,H40003,H40009,H40011,H40012,H40013,H40019 H40021,H40022,H40023,H40029,H40031,H40032,H40033,H40039 H40041,H40042,H40043,H40049,H40051,H40052,H40053,H40059 H40061,H40062,H40063,H40069,H4010X0,H4010X1,H4010X2,H4010X3 H4010X4,H401110,H401111,H401112,H401113,H401114,H401120 H401121,H401122,H401123,H401124,H401130,H401131,H401132 H401133,H401134,H401190,H401191,H401192,H401193,H401194 H4011X0,H4011X1,H4011X2,H4011X3,H4011X4,H401210,H401211 H401212,H401213,H401214,H401220,H401221,H401222,H401223 | 36042,36471,36473,36476,36477,36500,36501,36502,36503,36504 36505,36506,36510,36511,36512,36513,36515,36520,36521,36522 36523,36524,36531,36532,36544,36552,36559,36560,36561,36562 36564,36565,36570,36571,36572,36573,36574,36581,36582,36583 36589,3659 |

Continued on next page

**Supplementary Table S1:** (cont'd) A comprehensive mapping of diagnosis groups used in the experiments to their corresponding ICD-10 and ICD-9 codes.

| Diagnosis | ICD-10 | ICD-9 |
|---|---|---|
| | H401224,H401230,H401231,H401232,H401233,H401234,H401290 | |
| | H401291,H401292,H401293,H401294,H401310,H401311,H401312 | |
| | H401313,H401314,H401320,H401321,H401322,H401323,H401324 | |
| | H401330,H401331,H401332,H401333,H401334,H401390,H401391 | |
| | H401392,H401393,H401394,H401410,H401411,H401412,H401413 | |
| | H401414,H401420,H401421,H401422,H401423,H401424,H401430 | |
| | H401431,H401432,H401433,H401434,H401490,H401491,H401492 | |
| | H401493,H401494,H40151,H40152,H40153,H40159,H4020X0,H4020X1 | |
| | H4020X2,H4020X3,H4020X4,H40211,H40212,H40213,H40219,H402210 | |
| | H402211,H402212,H402213,H402214,H402220,H402221,H402222 | |
| | H402223,H402224,H402230,H402231,H402232,H402233,H402234 | |
| | H402290,H402291,H402292,H402293,H402294,H40231,H40232,H40233 | |
| | H40239,H40241,H40242,H40243,H40249,H4030X0,H4030X1,H4030X2 | |
| | H4030X3,H4030X4,H4031X0,H4031X1,H4031X2,H4031X3,H4031X4 | |
| | H4032X0,H4032X1,H4032X2,H4032X3,H4032X4,H4033X0,H4033X1 | |
| | H4033X2,H4033X3,H4033X4,H4040X0,H4040X1,H4040X2,H4040X3 | |
| | H4040X4,H4041X0,H4041X1,H4041X2,H4041X3,H4041X4,H4042X0 | |
| | H4042X1,H4042X2,H4042X3,H4042X4,H4043X0,H4043X1,H4043X2 | |
| | H4043X3,H4043X4,H4050X0,H4050X1,H4050X2,H4050X3,H4050X4 | |
| | H4051X0,H4051X1,H4051X2,H4051X3,H4051X4,H4052X0,H4052X1 | |
| | H4052X2,H4052X3,H4052X4,H4053X0,H4053X1,H4053X2,H4053X3 | |
| | H4053X4,H4060X0,H4060X1,H4060X2,H4060X3,H4060X4,H4061X0 | |
| | H4061X1,H4061X2,H4061X3,H4061X4,H4062X0,H4062X1,H4062X2 | |
| | H4062X3,H4062X4,H4063X0,H4063X1,H4063X2,H4063X3,H4063X4 | |
| | H40811,H40812,H40813,H40819,H40821,H40822,H40823,H40829 | |
| | H40831,H40832,H40833,H40839,H4089,H409,H42,H44511,H44512 | |
| | H44513,H44519 | |
| Gout | M1000,M10011,M10012,M10019,M10021,M10022,M10029,M10031,M10032 | 27400,27401,27402,27403,27410,27489,2749,9849 |
| | M10039,M10041,M10042,M10049,M10051,M10052,M10059,M10061 | |
| | M10062,M10069,M10071,M10072,M10079,M1008,M1009,M1010,M10111 | |
| | M10112,M10119,M10121,M10122,M10129,M10131,M10132,M10139 | |
| | M10141,M10142,M10149,M10151,M10152,M10159,M10161,M10162 | |
| | M10169,M10171,M10172,M10179,M1018,M1019,M1020,M10211,M10212 | |
| | M10219,M10221,M10222,M10229,M10231,M10232,M10239,M10241 | |
| | M10242,M10249,M10251,M10252,M10259,M10261,M10262,M10269 | |
| | M10271,M10272,M10279,M1028,M1029,M1030,M10311,M10312,M10319 | |
| | M10321,M10322,M10329,M10331,M10332,M10339,M10341,M10342 | |
| | M10349,M10351,M10352,M10359,M10361,M10362,M10369,M10371 | |
| | M10372,M10379,M1038,M1039,M1040,M10411,M10412,M10419,M10421 | |

Continued on next page

**Supplementary Table S1:** (cont'd) A comprehensive mapping of diagnosis groups used in the experiments to their corresponding ICD-10 and ICD-9 codes.

| Diagnosis | ICD-10 | ICD-9 |
|---|---|---|
| | M10422,M10429,M10431,M10432,M10439,M10441,M10442,M10449 M10451,M10452,M10459,M10461,M10462,M10469,M10471,M10472 M10479,M1048,M1049,M109,M1A00X0,M1A00X1,M1A0110,M1A0111 M1A0120,M1A0121,M1A0190,M1A0191,M1A0210,M1A0211,M1A0220 M1A0221,M1A0290,M1A0291,M1A0310,M1A0311,M1A0320,M1A0321 M1A0390,M1A0391,M1A0410,M1A0411,M1A0420,M1A0421,M1A0490 M1A0491,M1A0510,M1A0511,M1A0520,M1A0521,M1A0590,M1A0591 M1A0610,M1A0611,M1A0620,M1A0621,M1A0690,M1A0691,M1A0710 M1A0711,M1A0720,M1A0721,M1A0790,M1A0791,M1A08X0,M1A08X1 M1A09X0,M1A09X1,M1A10X0,M1A10X1,M1A1110,M1A1111,M1A1120 M1A1121,M1A1190,M1A1191,M1A1210,M1A1211,M1A1220,M1A1221 M1A1290,M1A1291,M1A1310,M1A1311,M1A1320,M1A1321,M1A1390 M1A1391,M1A1410,M1A1411,M1A1420,M1A1421,M1A1490,M1A1491 M1A1510,M1A1511,M1A1520,M1A1521,M1A1590,M1A1591,M1A1610 M1A1611,M1A1620,M1A1621,M1A1690,M1A1691,M1A1710,M1A1711 M1A1720,M1A1721,M1A1790,M1A1791,M1A18X0,M1A18X1,M1A19X0 M1A19X1,M1A20X0,M1A20X1,M1A2110,M1A2111,M1A2120,M1A2121 M1A2190,M1A2191,M1A2210,M1A2211,M1A2220,M1A2221,M1A2290 M1A2291,M1A2310,M1A2311,M1A2320,M1A2321,M1A2390,M1A2391 M1A2410,M1A2411,M1A2420,M1A2421,M1A2490,M1A2491,M1A2510 M1A2511,M1A2520,M1A2521,M1A2590,M1A2591,M1A2610,M1A2611 M1A2620,M1A2621,M1A2690,M1A2691,M1A2710,M1A2711,M1A2720 M1A2721,M1A2790,M1A2791,M1A28X0,M1A28X1,M1A29X0,M1A29X1 M1A30X0,M1A30X1,M1A3110,M1A3111,M1A3120,M1A3121,M1A3190 M1A3191,M1A3210,M1A3211,M1A3220,M1A3221,M1A3290,M1A3291 M1A3310,M1A3311,M1A3320,M1A3321,M1A3390,M1A3391,M1A3410 M1A3411,M1A3420,M1A3421,M1A3490,M1A3491,M1A3510,M1A3511 M1A3520,M1A3521,M1A3590,M1A3591,M1A3610,M1A3611,M1A3620 M1A3621,M1A3690,M1A3691,M1A3710,M1A3711,M1A3720,M1A3721 M1A3790,M1A3791,M1A38X0,M1A38X1,M1A39X0,M1A39X1,M1A40X0 M1A40X1,M1A4110,M1A4111,M1A4120,M1A4121,M1A4190,M1A4191 M1A4210,M1A4211,M1A4220,M1A4221,M1A4290,M1A4291,M1A4310 M1A4311,M1A4320,M1A4321,M1A4390,M1A4391,M1A4410,M1A4411 M1A4420,M1A4421,M1A4490,M1A4491,M1A4510,M1A4511,M1A4520 M1A4521,M1A4590,M1A4591,M1A4610,M1A4611,M1A4620,M1A4621 M1A4690,M1A4691,M1A4710,M1A4711,M1A4720,M1A4721,M1A4790 M1A4791,M1A48X0,M1A48X1,M1A49X0,M1A49X1,M1A9XX0,M1A9XX1 | |
| HIV infection | B20,B9735,O98711,O98712,O98713,O98719,O9872,O9873,Z21 | 042,07953,64760,64761,64763,64764,V08 |

Continued on next page

**Supplementary Table S1:** (cont'd) A comprehensive mapping of diagnosis groups used in the experiments to their corresponding ICD-10 and ICD-9 codes.

| Diagnosis | ICD-10 | ICD-9 |
|---|---|---|
| Hallucinogen-related disorders | F1610,F16120,F16121,F16122,F16129,F1614,F16150,F16151,F16159 F16180,F16183,F16188,F1619,F1620,F16220,F16221,F16229,F1624 F16250,F16251,F16259,F16280,F16283,F16288,F1629,F1690,F1691 F16920,F16921,F16929,F1694,F16950,F16951,F16959,F16980,F16983 F16988,F1699,T408X1A,T408X2A,T408X3A,T408X4A,T40901A,T40902A T40903A,T40904A,T40905A,T40991A,T40992A,T40993A,T40994A T40995A,T43641A,T43642A,T43643A,T43644A | 29211,29212,2922,29281,29284,29289,2929,30450,30451,30452 30530,30531,30532,30533,9696,99529,E8541,E9396,E9503,E9620 E9802 |
| Head and neck cancers - all other types | C148,C300,C301,C310,C311,C312,C313,C318,C319,C760 | 1498,1499,1600,1601,1602,1603,1604,1605,1608,1609,1950 |
| Head and neck cancers - eye | C6900,C6901,C6902,C6910,C6911,C6912,C6920,C6921,C6922,C6930 C6931,C6932,C6940,C6941,C6942,C6950,C6951,C6952,C6960,C6961 C6962,C6980,C6981,C6982,C6990,C6991,C6992,D0920,D0921,D0922 | 1900,1901,1902,1903,1904,1905,1906,1907,1908,1909,2340 |
| Head and neck cancers - hypopharyngeal | C12,C130,C131,C132,C138,C139 | 1480,1481,1482,1483,1488,1489 |
| Head and neck cancers - laryngeal | C320,C321,C322,C323,C328,C329,D020 | 1610,1611,1612,1613,1618,1619,2310 |
| Head and neck cancers - lip and oral cavity | C000,C001,C002,C003,C004,C005,C006,C008,C009,C01,C020,C021 C022,C023,C024,C028,C029,C030,C031,C039,C040,C041,C048,C049 C050,C051,C052,C058,C059,C060,C061,C062,C0680,C0689,C069 D0000,D0001,D0002,D0003,D0004,D0005,D0006,D0007 | 1400,1401,1403,1404,1405,1406,1408,1409,1410,1411,1412,1413 1414,1416,1418,1419,1430,1431,1438,1439,1440,1441,1448,1449 1450,1451,1452,1453,1454,1455,1456,1458,1459,2300 |
| Head and neck cancers - nasopharyngeal | C110,C111,C112,C113,C118,C119 | 1470,1471,1472,1473,1478,1479 |
| Head and neck cancers - pharyngeal | C140,C142,D0008 | 1490,1491,2300 |
| Head and neck cancers - salivary gland | C07,C080,C081,C089 | 1420,1421,1422,1428,1429 |
| Head and neck cancers - throat | C100,C101,C102,C103,C104,C108,C109 | 1463,1464,1466,1467,1468,1469 |
| Head and neck cancers - tonsils | C090,C091,C098,C099 | 1460,1461,1462 |

Continued on next page

**Supplementary Table S1:** (cont'd) A comprehensive mapping of diagnosis groups used in the experiments to their corresponding ICD-10 and ICD-9 codes.

| Diagnosis | ICD-10 | ICD-9 |
|---|---|---|
| Headache; including migraine | G43001,G43009,G43011,G43019,G43101,G43109,G43111,G43119 G43401,G43409,G43411,G43419,G43501,G43509,G43511,G43519 G43601,G43609,G43611,G43619,G43701,G43709,G43711,G43719 G43801,G43809,G43811,G43819,G43821,G43829,G43831,G43839 G43901,G43909,G43911,G43919,G43A0,G43A1,G43B0,G43B1,G43C0 G43C1,G43D0,G43D1,G44001,G44009,G44011,G44019,G44021,G44029 G44031,G44039,G44041,G44049,G44051,G44059,G44091,G44099,G441 G44201,G44209,G44211,G44219,G44221,G44229,G44301,G44309 G44311,G44319,G44321,G44329,G4440,G4441,G4451,G4452,G4453 G4459,G4481,G4482,G4483,G4484,G4485,G4486,G4489,R51,R510,R519 | 30781,33900,33901,33902,33903,33904,33905,33909,33910,33911 33912,33920,33921,33922,3393,33941,33942,33943,33944,33981 33982,33983,33984,33985,33989,34600,34601,34602,34603,34610 34611,34612,34613,34620,34621,34622,34630,34631,34632,34633 34640,34641,34642,34643,34650,34651,34652,34653,34660,34661 34662,34663,34670,34671,34672,34673,34680,34681,34682,34683 34690,34691,34692,34693,7840 |
| Hearing loss | F804,H900,H9011,H9012,H902,H903,H9041,H9042,H905,H906,H9071 H9072,H908,H90A11,H90A12,H90A21,H90A22,H90A31,H90A32,H9101 H9102,H9103,H9109,H9110,H9111,H9112,H9113,H9120,H9121,H9122 H9123,H913,H918X1,H918X2,H918X3,H918X9,H9190,H9191,H9192 H9193,H93011,H93012,H93013,H93019,P096,Z45320,Z45321,Z45328 | 31534,38801,38802,38811,3882,38900,38901,38902,38903,38904 38905,38906,38908,38910,38911,38912,38913,38914,38915,38916 38917,38918,38920,38921,38922,3897,3898,3899,V5309 |
| Heart failure | I0981,I110,I130,I132,I501,I5020,I5021,I5022,I5023,I5030,I5031 I5032,I5033,I5040,I5041,I5042,I5043,I50810,I50811,I50812 I50813,I50814,I5082,I5083,I5084,I5089,I509,I97130,I97131 O29121,O29122,O29123,O29129,Z95811,Z95812 | 39891,40201,40211,40291,40401,40403,40411,40413,40491,40493 4280,4281,42820,42821,42822,42823,42830,42831,42832,42833 42840,42841,42842,42843,4289,4294,64680,64681,64683,9971 V4321,V4322 |
| Hematuria | N020,N021,N022,N023,N024,N025,N026,N027,N028,N029,N02A,N3001 N3011,N3021,N3031,N3041,N3081,N3091,R310,R311,R312,R3121 R3129,R319 | 5811,5812,5813,58189,5819,5950,5951,5952,5953,5954,59581 59582,59589,5959,59970,59971,59972 |
| Hemolytic anemia | D550,D551,D552,D5521,D5529,D553,D558,D559,D560,D561,D562,D563 D564,D565,D568,D569,D580,D581,D582,D588,D589,D590,D591,D5910 D5911,D5912,D5913,D5919,D592,D593,D5930,D5931,D5932,D5939 D594,D595,D596,D598,D599 | 2820,2821,2822,2823,28240,28243,28244,28245,28246,28247,28249 2827,2828,2829,2830,28310,28311,28319,2832,2839 |
| Hemolytic jaundice and perinatal jaundice | P550,P551,P558,P559,P560,P5690,P5699,P570,P578,P579,P580,P581 P582,P583,P5841,P5842,P585,P588,P589,P590,P591,P5920,P5929 P593,P598,P599 | 7730,7731,7732,7733,7734,7740,7741,7742,77430,77439,7744,7745 7746,7747 |
| Hemorrhage after first trimester | O26852,O26853,O4412,O4413,O4432,O4433,O4452,O4453,O45002 O45003,O45012,O45013,O45022,O45023,O45092,O45093,O458X2 O458X3,O4592,O4593,O46002,O46003,O46012,O46013,O46022,O46023 O46092,O46093,O468X2,O468X3,O4692,O4693,O670,O678,O679,O720 | 64111,64113,64121,64123,64131,64133,64181,64183,64191,64193 64951,64953,66602,66604,66612,66614,66622,66624 |

Continued on next page

**Supplementary Table S1:** (cont'd) A comprehensive mapping of diagnosis groups used in the experiments to their corresponding ICD-10 and ICD-9 codes.

| Diagnosis | ICD-10 | ICD-9 |
|---|---|---|
| | O721,O722 | |
| Hemorrhagic and hematologic disorders of newborn | P500,P501,P502,P503,P504,P505,P508,P509,P510,P518,P519,P520 P521,P5221,P5222,P523,P524,P525,P526,P528,P529,P53,P540,P541 P542,P543,P544,P545,P546,P548,P549,P60,P610,P611,P612,P613 P614,P615,P616,P618,P619 | 7670,7720,77210,77211,77212,77213,77214,7722,7723,7724,7725 7726,7728,7729,7760,7761,7762,7763,7764,7765,7766,7767,7768 7769 |
| Hemorrhoids | K640,K641,K642,K643,K644,K645,K648,K649,O2240,O2241,O2242 O2243,O872 | 4550,4551,4552,4553,4554,4555,4556,4557,4558,4559,67180,67181 67182,67183,67184 |
| Hepatic failure | K7040,K7041,K7200,K7201,K7210,K7211,K7290,K7291,K9182 | 570,5713,5722,5728,99749 |
| Hepatitis | A5145,B0081,B150,B159,B160,B161,B162,B169,B170,B1710,B1711 B172,B178,B179,B180,B181,B182,B188,B189,B190,B1910,B1911 B1920,B1921,B199,B251,B2681,B581,K7010,K7011,O98411,O98412 O98413,O98419,O9842,O9843,P353 | 05471,0700,0701,07020,07021,07030,07031,07032,07033,07041 07049,07051,07052,07053,07054,07059,0706,07070,07071,0709 07271,0785,09162,1305,5711,5731,64760,64761,64762,64763,64764 7712 |
| Hodgkin lymphoma | C8100,C8101,C8102,C8103,C8104,C8105,C8106,C8107,C8108,C8109 C8110,C8111,C8112,C8113,C8114,C8115,C8116,C8117,C8118,C8119 C8120,C8121,C8122,C8123,C8124,C8125,C8126,C8127,C8128,C8129 C8130,C8131,C8132,C8133,C8134,C8135,C8136,C8137,C8138,C8139 C8140,C8141,C8142,C8143,C8144,C8145,C8146,C8147,C8148,C8149 C8170,C8171,C8172,C8173,C8174,C8175,C8176,C8177,C8178,C8179 C8190,C8191,C8192,C8193,C8194,C8195,C8196,C8197,C8198,C8199 | 20100,20101,20102,20103,20104,20105,20106,20107,20108,20110 20111,20112,20113,20114,20115,20116,20117,20118,20120,20121 20122,20123,20124,20125,20126,20127,20128,20140,20141,20142 20143,20144,20145,20146,20147,20148,20150,20151,20152,20153 20154,20155,20156,20157,20158,20160,20161,20162,20163,20164 20165,20166,20167,20168,20170,20171,20172,20173,20174,20175 20176,20177,20178,20190,20191,20192,20193,20194,20195,20196 20197,20198 |
| Hyperplasia of prostate | N400,N401,N402,N403 | 60000,60001,60010,60011 |
| Hypertension and hypertensive-related conditions complicating pregnancy; childbirth; and the puerperium | O10011,O10012,O10013,O10019,O1002,O1003,O10111,O10112,O10113 O10119,O1012,O1013,O10211,O10212,O10213,O10219,O1022,O1023 O10311,O10312,O10313,O10319,O1032,O1033,O10411,O10412,O10413 O10419,O1042,O1043,O10911,O10912,O10913,O10919,O1092,O1093 O111,O112,O113,O114,O115,O119,O131,O132,O133,O134,O135,O139 O1400,O1402,O1403,O1404,O1405,O1410,O1412,O1413,O1414,O1415 O1420,O1422,O1423,O1424,O1425,O1490,O1492,O1493,O1494,O1495 O1500,O1502,O1503,O151,O152,O159,O161,O162,O163,O164,O165 O169 | 64200,64201,64203,64204,64210,64211,64213,64214,64220,64221 64223,64224,64230,64231,64232,64233,64234,64240,64241,64242 64243,64244,64250,64251,64252,64253,64254,64260,64261,64263 64264,64270,64271,64272,64273,64274,64290,64291,64293,64294 |
| Hypertension with | H35031,H35032,H35033,I110,I119,I120,I129,I130,I1310,I1311 | 36211,40200,40201,40210,40211,40290,40291,40300,40301,40310 |

Continued on next page

**Supplementary Table S1:** (cont'd) A comprehensive mapping of diagnosis groups used in the experiments to their corresponding ICD-10 and ICD-9 codes.

| Diagnosis | ICD-10 | ICD-9 |
|---|---|---|
| complications and | I132,I150,I151,I152,I158,I159,I160,I161,I169,I674,I973,O10111 | 40311,40390,40391,40400,40401,40402,40403,40410,40411,40412 |
| secondary | O10112,O10113,O10119,O1012,O1013,O10211,O10212,O10213,O10219 | 40413,40490,40491,40492,40493,40501,40509,40511,40519,40591 |
| hypertension | O1022,O1023,O10311,O10312,O10313,O10319,O1032,O1033,O10411 | 40599,4372,64210,64211,64213,64214,64220,64221,64223,64224 |
| | O10412,O10413,O10419,O1042,O1043,O111,O112,O113,O114,O115 | 64230,64231,64232,64233,64234,64270,64271,64273,99791 |
| | O119,O131,O132,O133,O134,O135,O139 | |
| Hypotension | I950,I951,I952,I953,I9581,I9589,I959,O2650,O2651,O2652,O2653 | 4580,4581,45821,45829,4588,4589,66920,66921,66922,66923,66924 |
| Immune-mediated | A3984,M0210,M02111,M02112,M02119,M02121,M02122,M02129,M02131 | 03682,0993,71110,71111,71112,71113,71114,71115,71116,71117 |
| reactive | M02132,M02139,M02141,M02142,M02149,M02151,M02152,M02159 | 71118,71119,71130,71131,71132,71133,71134,71135,71136,71137 |
| arthropathies | M02161,M02162,M02169,M02171,M02172,M02179,M0218,M0219,M0230 | 71138,71139,71140,71141,71142,71143,71144,71145,71146,71147 |
| | M02311,M02312,M02319,M02321,M02322,M02329,M02331,M02332 | 71148,71149,71150,71151,71152,71153,71154,71155,71156,71157 |
| | M02339,M02341,M02342,M02349,M02351,M02352,M02359,M02361 | 71158,71159,71180,71181,71182,71183,71184,71185,71186,71187 |
| | M02362,M02369,M02371,M02372,M02379,M0238,M0239,M0280,M02811 | 71188,71189,7131,7137,71680,71681,71682,71683,71684,71685 |
| | M02812,M02819,M02821,M02822,M02829,M02831,M02832,M02839 | 71686,71687,71688,71689 |
| | M02841,M02842,M02849,M02851,M02852,M02859,M02861,M02862 | |
| | M02869,M02871,M02872,M02879,M0288,M0289,M029,M0760,M07611 | |
| | M07612,M07619,M07621,M07622,M07629,M07631,M07632,M07639 | |
| | M07641,M07642,M07649,M07651,M07652,M07659,M07661,M07662 | |
| | M07669,M07671,M07672,M07679,M0768,M0769 | |
| Immunity disorders | D800,D801,D802,D803,D804,D805,D806,D807,D808,D809,D810,D811 | 135,2662,2730,2731,2732,2772,2776,27900,27901,27902,27903 |
| | D812,D813,D8130,D8131,D8132,D8139,D814,D815,D816,D817,D81810 | 27904,27905,27906,27909,27910,27911,27912,27913,27919,2792 |
| | D81818,D81819,D8182,D8189,D819,D820,D821,D822,D823,D824,D828 | 2793,27941,27950,27951,27952,27953,2798,2799,28989 |
| | D829,D830,D831,D832,D838,D839,D840,D841,D848,D8481,D84821 | |
| | D84822,D8489,D849,D860,D861,D862,D863,D8681,D8682,D8683,D8684 | |
| | D8685,D8686,D8687,D8689,D869,D890,D891,D892,D893,D8940,D8941 | |
| | D8942,D8943,D8944,D8949,D89810,D89811,D89812,D89813,D8982 | |
| | D89831,D89832,D89833,D89834,D89835,D89839,D8989,D899 | |
| Implant, device or | Z44001,Z44002,Z44009,Z44011,Z44012,Z44019,Z44021,Z44022 | V410,V412,V422,V430,V431,V4321,V4322,V433,V434,V435,V4360 |
| graft related | Z44029,Z44101,Z44102,Z44109,Z44111,Z44112,Z44119,Z44121 | V4361,V4362,V4363,V4364,V4365,V4366,V4369,V437,V4381,V4382 |
| encounter | Z44122,Z44129,Z4420,Z4421,Z4422,Z4430,Z4431,Z4432,Z448,Z449 | V4383,V4389,V4500,V4501,V4502,V4509,V452,V453,V454,V4561 |
| | Z45010,Z45018,Z4502,Z4509,Z451,Z452,Z4531,Z45320,Z45321 | V4569,V4581,V4582,V4584,V4585,V4586,V4589,V4982,V520,V521 |
| | Z45328,Z4541,Z4542,Z4549,Z45811,Z45812,Z45819,Z4582,Z4589 | V522,V523,V524,V528,V529,V5301,V5302,V5309,V531,V532,V5331 |
| | Z459,Z460,Z461,Z462,Z463,Z464,Z4651,Z4659,Z466,Z4681,Z4682 | V5332,V5339,V534,V5350,V5351,V5359,V536,V537,V538,V5390,V5391 |
| | Z4689,Z469,Z950,Z951,Z952,Z953,Z954,Z955,Z95810,Z95811,Z95812 | V5399,V585,V5881,V5882,V6546 |
| | Z95818,Z95820,Z95828,Z959,Z960,Z961,Z9620,Z9621,Z9622,Z9629 | |
| | Z963,Z9641,Z9649,Z965,Z9660,Z96611,Z96612,Z96619,Z96621 | |
| | Z96622,Z96629,Z96631,Z96632,Z96639,Z96641,Z96642,Z96643 | |

Continued on next page

**Supplementary Table S1:** (cont'd) A comprehensive mapping of diagnosis groups used in the experiments to their corresponding ICD-10 and ICD-9 codes.

| Diagnosis | ICD-10 | ICD-9 |
|---|---|---|
| | Z96649,Z96651,Z96652,Z96653,Z96659,Z96661,Z96662,Z96669<br>Z96691,Z96692,Z96693,Z96698,Z967,Z9681,Z9682,Z9689,Z969,Z970<br>Z9710,Z9711,Z9712,Z9713,Z9714,Z9715,Z9716,Z972,Z973,Z974,Z978<br>Z980,Z981,Z982,Z983,Z9841,Z9842,Z9849,Z9861,Z9862,Z98810<br>Z98811,Z98818,Z9882,Z9883,Z9884,Z9889,Z98890 | |
| Induced abortion and complications of termination of pregnancy | O045,O046,O047,O0480,O0481,O0482,O0483,O0484,O0485,O0486<br>O0487,O0488,O0489,O070,O071,O072,O0730,O0731,O0732,O0733<br>O0734,O0735,O0736,O0737,O0738,O0739,O074,Z332 | 63502,63512,63522,63532,63542,63552,63562,63572,63582,63590<br>6380,6381,6382,6383,6384,6385,6386,6387,6388,6389 |
| Infective arthritis | A0104,A0223,A1801,A1802,A1803,A3983,A5442,A6923,B0682,B2685<br>B4282,M0000,M00011,M00012,M00019,M00021,M00022,M00029,M00031<br>M00032,M00039,M00041,M00042,M00049,M00051,M00052,M00059<br>M00061,M00062,M00069,M00071,M00072,M00079,M0008,M0009,M0010<br>M00111,M00112,M00119,M00121,M00122,M00129,M00131,M00132<br>M00139,M00141,M00142,M00149,M00151,M00152,M00159,M00161<br>M00162,M00169,M00171,M00172,M00179,M0018,M0019,M0020,M00211<br>M00212,M00219,M00221,M00222,M00229,M00231,M00232,M00239<br>M00241,M00242,M00249,M00251,M00252,M00259,M00261,M00262<br>M00269,M00271,M00272,M00279,M0028,M0029,M0080,M00811,M00812<br>M00819,M00821,M00822,M00829,M00831,M00832,M00839,M00841<br>M00842,M00849,M00851,M00852,M00859,M00861,M00862,M00869<br>M00871,M00872,M00879,M0088,M0089,M009,M01X0,M01X11,M01X12<br>M01X19,M01X21,M01X22,M01X29,M01X31,M01X32,M01X39,M01X41<br>M01X42,M01X49,M01X51,M01X52,M01X59,M01X61,M01X62,M01X69<br>M01X71,M01X72,M01X79,M01X8,M01X9 | 0020,00323,01500,01510,01520,01560,01570,01580,01590,03682<br>0412,05671,07279,08881,09850,1171,71100,71101,71102,71103<br>71104,71105,71106,71107,71108,71109,71140,71141,71142,71143<br>71144,71145,71146,71147,71148,71149,71150,71151,71152,71153<br>71154,71155,71156,71157,71158,71159,71160,71161,71162,71163<br>71164,71165,71166,71167,71168,71169,71170,71171,71172,71173<br>71174,71175,71176,71177,71178,71179,71180,71181,71182,71183<br>71184,71185,71186,71187,71188,71189,72081,73088,73740 |
| Inflammatory conditions of male genital organs | A1814,A1815,A5422,A5423,A5902,A6001,A6002,B260,B3742,B3881<br>N410,N411,N412,N413,N414,N418,N419,N431,N451,N452,N453,N454<br>N476,N481,N4821,N4822,N4829,N490,N491,N492,N493,N498,N499 | 01650,05413,05419,0720,09812,09813,09814,09832,09833,09834<br>1122,1141,13103,6010,6011,6012,6013,6014,6018,6019,6031,6040<br>60490,60499,6071,6072,6080,6084 |
| Inflammatory diseases of female pelvic organs | A1816,A1817,A1818,A5142,A5402,A5403,A5424,A5602,A5611,A5901<br>A6003,A6004,B373,B3731,B3732,N7001,N7002,N7003,N7011,N7012<br>N7013,N7091,N7092,N7093,N710,N711,N719,N72,N730,N731,N732<br>N733,N734,N735,N736,N738,N739,N74,N750,N751,N758,N759,N760<br>N761,N762,N763,N764,N765,N766,N7681,N7682,N7689,N770,N771<br>O23511,O23512,O23513,O23519,O23591,O23592,O23593,O23599,O8611<br>O8619 | 01660,01670,05411,05412,05419,09189,0980,09815,09816,09835<br>09836,09953,09954,1121,13101,6140,6141,6142,6143,6144,6145<br>6146,6147,6148,6149,6150,6151,6159,6160,61610,61611,6162,6163<br>6164,61650,61651,61681,61689,6169,64660,64661,64663,64664 |

Continued on next page

**Supplementary Table S1:** (cont'd) A comprehensive mapping of diagnosis groups used in the experiments to their corresponding ICD-10 and ICD-9 codes.

| Diagnosis | ICD-10 | ICD-9 |
|---|---|---|
| Influenza | J09X1,J09X2,J09X3,J09X9,J1000,J1001,J1008,J101,J102,J1081<br>J1082,J1083,J1089,J1100,J1108,J111,J112,J1181,J1182,J1183<br>J1189 | 4870,4871,4878,48801,48802,48809,48811,48812,48819,48881<br>48882,48889 |
| Inhalant-related disorders | F1810,F18120,F18121,F18129,F1814,F18150,F18151,F18159,F1817<br>F18180,F18188,F1819,F1820,F18220,F18221,F18229,F1824,F18250<br>F18251,F18259,F1827,F18280,F18288,F1829,F1890,F1891,F18920<br>F18921,F18929,F1894,F18950,F18951,F18959,F1897,F18980,F18988<br>F1899,T410X1A,T410X2A,T410X3A,T410X4A,T410X5A | 29211,29212,2922,29281,29282,29284,29289,2929,30460,30590<br>30591,30592,30593,9682,99522,E8551,E9382,E9528,E9620,E9804 |
| Injury to blood vessels, initial encounter | S090XXA,S15001A,S15002A,S15009A,S15011A,S15012A,S15019A<br>S15021A,S15022A,S15029A,S15091A,S15092A,S15099A,S15101A<br>S15102A,S15109A,S15111A,S15112A,S15119A,S15121A,S15122A<br>S15129A,S15191A,S15192A,S15199A,S15201A,S15202A,S15209A<br>S15211A,S15212A,S15219A,S15221A,S15222A,S15229A,S15291A<br>S15292A,S15299A,S15301A,S15302A,S15309A,S15311A,S15312A<br>S15319A,S15321A,S15322A,S15329A,S15391A,S15392A,S15399A<br>S158XXA,S159XXA,S2500XA,S2501XA,S2502XA,S2509XA,S25101A<br>S25102A,S25109A,S25111A,S25112A,S25119A,S25121A,S25122A<br>S25129A,S25191A,S25192A,S25199A,S2520XA,S2521XA,S2522XA<br>S2529XA,S25301A,S25302A,S25309A,S25311A,S25312A,S25319A<br>S25321A,S25322A,S25329A,S25391A,S25392A,S25399A,S25401A<br>S25402A,S25409A,S25411A,S25412A,S25419A,S25421A,S25422A<br>S25429A,S25491A,S25492A,S25499A,S25501A,S25502A,S25509A<br>S25511A,S25512A,S25519A,S25591A,S25592A,S25599A,S25801A<br>S25802A,S25809A,S25811A,S25812A,S25819A,S25891A,S25892A<br>S25899A,S2590XA,S2591XA,S2599XA,S3500XA,S3501XA,S3502XA<br>S3509XA,S3510XA,S3511XA,S3512XA,S3519XA,S35211A,S35212A<br>S35218A,S35219A,S35221A,S35222A,S35228A,S35229A,S35231A<br>S35232A,S35238A,S35239A,S35291A,S35292A,S35298A,S35299A<br>S35311A,S35318A,S35319A,S35321A,S35328A,S35329A,S35331A<br>S35338A,S35339A,S35341A,S35348A,S35349A,S35401A,S35402A<br>S35403A,S35404A,S35405A,S35406A,S35411A,S35412A,S35413A<br>S35414A,S35415A,S35416A,S35491A,S35492A,S35493A,S35494A<br>S35495A,S35496A,S3550XA,S35511A,S35512A,S35513A,S35514A<br>S35515A,S35516A,S35531A,S35532A,S35533A,S35534A,S35535A<br>S35536A,S3559XA,S358X1A,S358X8A,S358X9A,S3590XA,S3591XA<br>S3599XA,S45001A,S45002A,S45009A,S45011A,S45012A,S45019A<br>S45091A,S45092A,S45099A,S45101A,S45102A,S45109A,S45111A<br>S45112A,S45119A,S45191A,S45192A,S45199A,S45201A,S45202A | 90000,90001,90002,90003,9001,90081,90082,90089,9009,9010,9011<br>9012,9013,90140,90141,90142,90181,90182,90189,9019,9020,90210<br>90211,90219,90220,90221,90222,90223,90224,90225,90226,90227<br>90229,90231,90232,90233,90234,90240,90241,90242,90249,90251<br>90252,90253,90254,90255,90256,90259,90281,90282,90289,9029<br>90300,90301,90302,9031,9032,9033,9034,9035,9038,9039,9040<br>9041,9042,9043,90441,90442,90450,90451,90452,90453,90454,9046<br>9047,9048 |

Continued on next page

**Supplementary Table S1:** (cont'd) A comprehensive mapping of diagnosis groups used in the experiments to their corresponding ICD-10 and ICD-9 codes.

| Diagnosis | ICD-10 | ICD-9 |
|---|---|---|
| | S45209A,S45211A,S45212A,S45219A,S45291A,S45292A,S45299A | |
| | S45301A,S45302A,S45309A,S45311A,S45312A,S45319A,S45391A | |
| | S45392A,S45399A,S45801A,S45802A,S45809A,S45811A,S45812A | |
| | S45819A,S45891A,S45892A,S45899A,S45901A,S45902A,S45909A | |
| | S45911A,S45912A,S45919A,S45991A,S45992A,S45999A,S55001A | |
| | S55002A,S55009A,S55011A,S55012A,S55019A,S55091A,S55092A | |
| | S55099A,S55101A,S55102A,S55109A,S55111A,S55112A,S55119A | |
| | S55191A,S55192A,S55199A,S55201A,S55202A,S55209A,S55211A | |
| | S55212A,S55219A,S55291A,S55292A,S55299A,S55801A,S55802A | |
| | S55809A,S55811A,S55812A,S55819A,S55891A,S55892A,S55899A | |
| | S55901A,S55902A,S55909A,S55911A,S55912A,S55919A,S55991A | |
| | S55992A,S55999A,S65001A,S65002A,S65009A,S65011A,S65012A | |
| | S65019A,S65091A,S65092A,S65099A,S65101A,S65102A,S65109A | |
| | S65111A,S65112A,S65119A,S65191A,S65192A,S65199A,S65201A | |
| | S65202A,S65209A,S65211A,S65212A,S65219A,S65291A,S65292A | |
| | S65299A,S65301A,S65302A,S65309A,S65311A,S65312A,S65319A | |
| | S65391A,S65392A,S65399A,S65401A,S65402A,S65409A,S65411A | |
| | S65412A,S65419A,S65491A,S65492A,S65499A,S65500A,S65501A | |
| | S65502A,S65503A,S65504A,S65505A,S65506A,S65507A,S65508A | |
| | S65509A,S65510A,S65511A,S65512A,S65513A,S65514A,S65515A | |
| | S65516A,S65517A,S65518A,S65519A,S65590A,S65591A,S65592A | |
| | S65593A,S65594A,S65595A,S65596A,S65597A,S65598A,S65599A | |
| | S65801A,S65802A,S65809A,S65811A,S65812A,S65819A,S65891A | |
| | S65892A,S65899A,S65901A,S65902A,S65909A,S65911A,S65912A | |
| | S65919A,S65991A,S65992A,S65999A,S75001A,S75002A,S75009A | |
| | S75011A,S75012A,S75019A,S75021A,S75022A,S75029A,S75091A | |
| | S75092A,S75099A,S75101A,S75102A,S75109A,S75111A,S75112A | |
| | S75119A,S75121A,S75122A,S75129A,S75191A,S75192A,S75199A | |
| | S75201A,S75202A,S75209A,S75211A,S75212A,S75219A,S75221A | |
| | S75222A,S75229A,S75291A,S75292A,S75299A,S75801A,S75802A | |
| | S75809A,S75811A,S75812A,S75819A,S75891A,S75892A,S75899A | |
| | S75901A,S75902A,S75909A,S75911A,S75912A,S75919A,S75991A | |
| | S75992A,S75999A,S85001A,S85002A,S85009A,S85011A,S85012A | |
| | S85019A,S85091A,S85092A,S85099A,S85101A,S85102A,S85109A | |
| | S85111A,S85112A,S85119A,S85121A,S85122A,S85129A,S85131A | |
| | S85132A,S85139A,S85141A,S85142A,S85149A,S85151A,S85152A | |
| | S85159A,S85161A,S85162A,S85169A,S85171A,S85172A,S85179A | |
| | S85181A,S85182A,S85189A,S85201A,S85202A,S85209A,S85211A | |
| | S85212A,S85219A,S85291A,S85292A,S85299A,S85301A,S85302A | |
| | S85309A,S85311A,S85312A,S85319A,S85391A,S85392A,S85399A | |

Continued on next page

**Supplementary Table S1:** (cont'd) A comprehensive mapping of diagnosis groups used in the experiments to their corresponding ICD-10 and ICD-9 codes.

| Diagnosis | ICD-10 | ICD-9 |
|---|---|---|
| | S85401A,S85402A,S85409A,S85411A,S85412A,S85419A,S85491A S85492A,S85499A,S85501A,S85502A,S85509A,S85511A,S85512A S85519A,S85591A,S85592A,S85599A,S85801A,S85802A,S85809A S85811A,S85812A,S85819A,S85891A,S85892A,S85899A,S85901A S85902A,S85909A,S85911A,S85912A,S85919A,S85991A,S85992A S85999A,S95001A,S95002A,S95009A,S95011A,S95012A,S95019A S95091A,S95092A,S95099A,S95101A,S95102A,S95109A,S95111A S95112A,S95119A,S95191A,S95192A,S95199A,S95201A,S95202A S95209A,S95211A,S95212A,S95219A,S95291A,S95292A,S95299A S95801A,S95802A,S95809A,S95811A,S95812A,S95819A,S95891A S95892A,S95899A,S95901A,S95902A,S95909A,S95911A,S95912A S95919A,S95991A,S95992A,S95999A | |
| Injury to blood vessels, subsequent encounter | S090XXD,S15001D,S15002D,S15009D,S15011D,S15012D,S15019D S15021D,S15022D,S15029D,S15091D,S15092D,S15099D,S15101D S15102D,S15109D,S15111D,S15112D,S15119D,S15121D,S15122D S15129D,S15191D,S15192D,S15199D,S15201D,S15202D,S15209D S15211D,S15212D,S15219D,S15221D,S15222D,S15229D,S15291D S15292D,S15299D,S15301D,S15302D,S15309D,S15311D,S15312D S15319D,S15321D,S15322D,S15329D,S15391D,S15392D,S15399D S158XXD,S159XXD,S2500XD,S2501XD,S2502XD,S2509XD,S25101D S25102D,S25109D,S25111D,S25112D,S25119D,S25121D,S25122D S25129D,S25191D,S25192D,S25199D,S2520XD,S2521XD,S2522XD S2529XD,S25301D,S25302D,S25309D,S25311D,S25312D,S25319D S25321D,S25322D,S25329D,S25391D,S25392D,S25399D,S25401D S25402D,S25409D,S25411D,S25412D,S25419D,S25421D,S25422D S25429D,S25491D,S25492D,S25499D,S25501D,S25502D,S25509D S25511D,S25512D,S25519D,S25591D,S25592D,S25599D,S25801D S25802D,S25809D,S25811D,S25812D,S25819D,S25891D,S25892D S25899D,S2590XD,S2591XD,S2599XD,S3500XD,S3501XD,S3502XD S3509XD,S3510XD,S3511XD,S3512XD,S3519XD,S35211D,S35212D S35218D,S35219D,S35221D,S35222D,S35228D,S35229D,S35231D S35232D,S35238D,S35239D,S35291D,S35292D,S35298D,S35299D S35311D,S35318D,S35319D,S35321D,S35328D,S35329D,S35331D S35338D,S35339D,S35341D,S35348D,S35349D,S35401D,S35402D S35403D,S35404D,S35405D,S35406D,S35411D,S35412D,S35413D S35414D,S35415D,S35416D,S35491D,S35492D,S35493D,S35494D S35495D,S35496D,S3550XD,S35511D,S35512D,S35513D,S35514D S35515D,S35516D,S35531D,S35532D,S35533D,S35534D,S35535D S35536D,S3559XD,S358X1D,S358X8D,S358X9D,S3590XD,S3591XD | V5889 |

Continued on next page

**Supplementary Table S1:** (cont'd) A comprehensive mapping of diagnosis groups used in the experiments to their corresponding ICD-10 and ICD-9 codes.

| Diagnosis | ICD-10 | ICD-9 |
|---|---|---|
| | S3599XD,S45001D,S45002D,S45009D,S45011D,S45012D,S45019D | |
| | S45091D,S45092D,S45099D,S45101D,S45102D,S45109D,S45111D | |
| | S45112D,S45119D,S45191D,S45192D,S45199D,S45201D,S45202D | |
| | S45209D,S45211D,S45212D,S45219D,S45291D,S45292D,S45299D | |
| | S45301D,S45302D,S45309D,S45311D,S45312D,S45319D,S45391D | |
| | S45392D,S45399D,S45801D,S45802D,S45809D,S45811D,S45812D | |
| | S45819D,S45891D,S45892D,S45899D,S45901D,S45902D,S45909D | |
| | S45911D,S45912D,S45919D,S45991D,S45992D,S45999D,S55001D | |
| | S55002D,S55009D,S55011D,S55012D,S55019D,S55091D,S55092D | |
| | S55099D,S55101D,S55102D,S55109D,S55111D,S55112D,S55119D | |
| | S55191D,S55192D,S55199D,S55201D,S55202D,S55209D,S55211D | |
| | S55212D,S55219D,S55291D,S55292D,S55299D,S55801D,S55802D | |
| | S55809D,S55811D,S55812D,S55819D,S55891D,S55892D,S55899D | |
| | S55901D,S55902D,S55909D,S55911D,S55912D,S55919D,S55991D | |
| | S55992D,S55999D,S65001D,S65002D,S65009D,S65011D,S65012D | |
| | S65019D,S65091D,S65092D,S65099D,S65101D,S65102D,S65109D | |
| | S65111D,S65112D,S65119D,S65191D,S65192D,S65199D,S65201D | |
| | S65202D,S65209D,S65211D,S65212D,S65219D,S65291D,S65292D | |
| | S65299D,S65301D,S65302D,S65309D,S65311D,S65312D,S65319D | |
| | S65391D,S65392D,S65399D,S65401D,S65402D,S65409D,S65411D | |
| | S65412D,S65419D,S65491D,S65492D,S65499D,S65500D,S65501D | |
| | S65502D,S65503D,S65504D,S65505D,S65506D,S65507D,S65508D | |
| | S65509D,S65510D,S65511D,S65512D,S65513D,S65514D,S65515D | |
| | S65516D,S65517D,S65518D,S65519D,S65590D,S65591D,S65592D | |
| | S65593D,S65594D,S65595D,S65596D,S65597D,S65598D,S65599D | |
| | S65801D,S65802D,S65809D,S65811D,S65812D,S65819D,S65891D | |
| | S65892D,S65899D,S65901D,S65902D,S65909D,S65911D,S65912D | |
| | S65919D,S65991D,S65992D,S65999D,S75001D,S75002D,S75009D | |
| | S75011D,S75012D,S75019D,S75021D,S75022D,S75029D,S75091D | |
| | S75092D,S75099D,S75101D,S75102D,S75109D,S75111D,S75112D | |
| | S75119D,S75121D,S75122D,S75129D,S75191D,S75192D,S75199D | |
| | S75201D,S75202D,S75209D,S75211D,S75212D,S75219D,S75221D | |
| | S75222D,S75229D,S75291D,S75292D,S75299D,S75801D,S75802D | |
| | S75809D,S75811D,S75812D,S75819D,S75891D,S75892D,S75899D | |
| | S75901D,S75902D,S75909D,S75911D,S75912D,S75919D,S75991D | |
| | S75992D,S75999D,S85001D,S85002D,S85009D,S85011D,S85012D | |
| | S85019D,S85091D,S85092D,S85099D,S85101D,S85102D,S85109D | |
| | S85111D,S85112D,S85119D,S85121D,S85122D,S85129D,S85131D | |
| | S85132D,S85139D,S85141D,S85142D,S85149D,S85151D,S85152D | |
| | S85159D,S85161D,S85162D,S85169D,S85171D,S85172D,S85179D | |

Continued on next page

**Supplementary Table S1:** (cont'd) A comprehensive mapping of diagnosis groups used in the experiments to their corresponding ICD-10 and ICD-9 codes.

| Diagnosis | ICD-10 | ICD-9 |
|---|---|---|
| | S85181D,S85182D,S85189D,S85201D,S85202D,S85209D,S85211D<br>S85212D,S85219D,S85291D,S85292D,S85299D,S85301D,S85302D<br>S85309D,S85311D,S85312D,S85319D,S85391D,S85392D,S85399D<br>S85401D,S85402D,S85409D,S85411D,S85412D,S85419D,S85491D<br>S85492D,S85499D,S85501D,S85502D,S85509D,S85511D,S85512D<br>S85519D,S85591D,S85592D,S85599D,S85801D,S85802D,S85809D<br>S85811D,S85812D,S85819D,S85891D,S85892D,S85899D,S85901D<br>S85902D,S85909D,S85911D,S85912D,S85919D,S85991D,S85992D<br>S85999D,S95001D,S95002D,S95009D,S95011D,S95012D,S95019D<br>S95091D,S95092D,S95099D,S95101D,S95102D,S95109D,S95111D<br>S95112D,S95119D,S95191D,S95192D,S95199D,S95201D,S95202D<br>S95209D,S95211D,S95212D,S95219D,S95291D,S95292D,S95299D<br>S95801D,S95802D,S95809D,S95811D,S95812D,S95819D,S95891D<br>S95892D,S95899D,S95901D,S95902D,S95909D,S95911D,S95912D<br>S95919D,S95991D,S95992D,S95999D | |
| Injury to nerves,<br>muscles and tendons,<br>initial encounter | S04011A,S04012A,S04019A,S0402XA,S04031A,S04032A,S04039A<br>S04041A,S04042A,S04049A,S0410XA,S0411XA,S0412XA,S0420XA<br>S0421XA,S0422XA,S0430XA,S0431XA,S0432XA,S0440XA,S0441XA<br>S0442XA,S0450XA,S0451XA,S0452XA,S0460XA,S0461XA,S0462XA<br>S0470XA,S0471XA,S0472XA,S04811A,S04812A,S04819A,S04891A<br>S04892A,S04899A,S049XXA,S0910XA,S0919XA,S142XXA,S143XXA<br>S144XXA,S145XXA,S148XXA,S149XXA,S168XXA,S169XXA,S242XXA<br>S243XXA,S244XXA,S248XXA,S249XXA,S29001A,S29002A,S29009A<br>S29091A,S29092A,S29099A,S3421XA,S3422XA,S344XXA,S345XXA<br>S346XXA,S348XXA,S349XXA,S39001A,S39002A,S39003A,S39091A<br>S39092A,S39093A,S4400XA,S4401XA,S4402XA,S4410XA,S4411XA<br>S4412XA,S4420XA,S4421XA,S4422XA,S4430XA,S4431XA,S4432XA<br>S4440XA,S4441XA,S4442XA,S4450XA,S4451XA,S4452XA,S448X1A<br>S448X2A,S448X9A,S4490XA,S4491XA,S4492XA,S46001A,S46002A<br>S46009A,S46091A,S46092A,S46099A,S46101A,S46102A,S46109A<br>S46191A,S46192A,S46199A,S46201A,S46202A,S46209A,S46291A<br>S46292A,S46299A,S46301A,S46302A,S46309A,S46391A,S46392A<br>S46399A,S46801A,S46802A,S46809A,S46891A,S46892A,S46899A<br>S46901A,S46902A,S46909A,S46991A,S46992A,S46999A,S5400XA<br>S5401XA,S5402XA,S5410XA,S5411XA,S5412XA,S5420XA,S5421XA<br>S5422XA,S5430XA,S5431XA,S5432XA,S548X1A,S548X2A,S548X9A<br>S5490XA,S5491XA,S5492XA,S56001A,S56002A,S56009A,S56091A<br>S56092A,S56099A,S56101A,S56102A,S56103A,S56104A,S56105A<br>S56106A,S56107A,S56108A,S56109A,S56191A,S56192A,S56193A | 9500,9501,9502,9503,9510,9511,9512,9513,9514,9515,9516,9517<br>9518,9519,9530,9531,9532,9533,9534,9535,9540,9541,9548,9549<br>9550,9551,9552,9553,9554,9555,9556,9557,9558,9559,9560,9561<br>9562,9563,9564,9565,9568,9569,9570,9571,9578,9579,95901,95909<br>95911,95912,95919,9592,9593,9594,9596,9597 |

Continued on next page

**Supplementary Table S1:** (cont'd) A comprehensive mapping of diagnosis groups used in the experiments to their corresponding ICD-10 and ICD-9 codes.

| Diagnosis | ICD-10 | ICD-9 |
|---|---|---|
| | S56194A,S56195A,S56196A,S56197A,S56198A,S56199A,S56201A | |
| | S56202A,S56209A,S56291A,S56292A,S56299A,S56301A,S56302A | |
| | S56309A,S56391A,S56392A,S56399A,S56401A,S56402A,S56403A | |
| | S56404A,S56405A,S56406A,S56407A,S56408A,S56409A,S56491A | |
| | S56492A,S56493A,S56494A,S56495A,S56496A,S56497A,S56498A | |
| | S56499A,S56501A,S56502A,S56509A,S56591A,S56592A,S56599A | |
| | S56801A,S56802A,S56809A,S56891A,S56892A,S56899A,S56901A | |
| | S56902A,S56909A,S56991A,S56992A,S56999A,S6400XA,S6401XA | |
| | S6402XA,S6410XA,S6411XA,S6412XA,S6420XA,S6421XA,S6422XA | |
| | S6430XA,S6431XA,S6432XA,S6440XA,S64490A,S64491A,S64492A | |
| | S64493A,S64494A,S64495A,S64496A,S64497A,S64498A,S648X1A | |
| | S648X2A,S648X9A,S6490XA,S6491XA,S6492XA,S66001A,S66002A | |
| | S66009A,S66091A,S66092A,S66099A,S66100A,S66101A,S66102A | |
| | S66103A,S66104A,S66105A,S66106A,S66107A,S66108A,S66109A | |
| | S66190A,S66191A,S66192A,S66193A,S66194A,S66195A,S66196A | |
| | S66197A,S66198A,S66199A,S66201A,S66202A,S66209A,S66291A | |
| | S66292A,S66299A,S66300A,S66301A,S66302A,S66303A,S66304A | |
| | S66305A,S66306A,S66307A,S66308A,S66309A,S66390A,S66391A | |
| | S66392A,S66393A,S66394A,S66395A,S66396A,S66397A,S66398A | |
| | S66399A,S66401A,S66402A,S66409A,S66491A,S66492A,S66499A | |
| | S66500A,S66501A,S66502A,S66503A,S66504A,S66505A,S66506A | |
| | S66507A,S66508A,S66509A,S66590A,S66591A,S66592A,S66593A | |
| | S66594A,S66595A,S66596A,S66597A,S66598A,S66599A,S66801A | |
| | S66802A,S66809A,S66891A,S66892A,S66899A,S66901A,S66902A | |
| | S66909A,S66991A,S66992A,S66999A,S7400XA,S7401XA,S7402XA | |
| | S7410XA,S7411XA,S7412XA,S7420XA,S7421XA,S7422XA,S748X1A | |
| | S748X2A,S748X9A,S7490XA,S7491XA,S7492XA,S76001A,S76002A | |
| | S76009A,S76091A,S76092A,S76099A,S76101A,S76102A,S76109A | |
| | S76191A,S76192A,S76199A,S76201A,S76202A,S76209A,S76291A | |
| | S76292A,S76299A,S76301A,S76302A,S76309A,S76391A,S76392A | |
| | S76399A,S76801A,S76802A,S76809A,S76891A,S76892A,S76899A | |
| | S76901A,S76902A,S76909A,S76991A,S76992A,S76999A,S8400XA | |
| | S8401XA,S8402XA,S8410XA,S8411XA,S8412XA,S8420XA,S8421XA | |
| | S8422XA,S84801A,S84802A,S84809A,S8490XA,S8491XA,S8492XA | |
| | S86001A,S86002A,S86009A,S86091A,S86092A,S86099A,S86101A | |
| | S86102A,S86109A,S86191A,S86192A,S86199A,S86201A,S86202A | |
| | S86209A,S86291A,S86292A,S86299A,S86301A,S86302A,S86309A | |
| | S86391A,S86392A,S86399A,S86801A,S86802A,S86809A,S86891A | |
| | S86892A,S86899A,S86901A,S86902A,S86909A,S86991A,S86992A | |
| | S86999A,S9400XA,S9401XA,S9402XA,S9410XA,S9411XA,S9412XA | |

Continued on next page

**Supplementary Table S1:** (cont'd) A comprehensive mapping of diagnosis groups used in the experiments to their corresponding ICD-10 and ICD-9 codes.

| Diagnosis | ICD-10 | ICD-9 |
|---|---|---|
| | S9420XA,S9421XA,S9422XA,S9430XA,S9431XA,S9432XA,S948X1A S948X2A,S948X9A,S9490XA,S9491XA,S9492XA,S96001A,S96002A S96009A,S96091A,S96092A,S96099A,S96101A,S96102A,S96109A S96191A,S96192A,S96199A,S96201A,S96202A,S96209A,S96291A S96292A,S96299A,S96801A,S96802A,S96809A,S96891A,S96892A S96899A,S96901A,S96902A,S96909A,S96991A,S96992A,S96999A | |
| Injury to nerves, muscles and tendons, subsequent encounter | S04011D,S04012D,S04019D,S0402XD,S04031D,S04032D,S04039D S04041D,S04042D,S04049D,S0410XD,S0411XD,S0412XD,S0420XD S0421XD,S0422XD,S0430XD,S0431XD,S0432XD,S0440XD,S0441XD S0442XD,S0450XD,S0451XD,S0452XD,S0460XD,S0461XD,S0462XD S0470XD,S0471XD,S0472XD,S04811D,S04812D,S04819D,S04891D S04892D,S04899D,S049XXD,S0910XD,S0919XD,S142XXD,S143XXD S144XXD,S145XXD,S148XXD,S149XXD,S168XXD,S169XXD,S242XXD S243XXD,S244XXD,S248XXD,S249XXD,S29001D,S29002D,S29009D S29091D,S29092D,S29099D,S3421XD,S3422XD,S344XXD,S345XXD S346XXD,S348XXD,S349XXD,S39001D,S39002D,S39003D,S39091D S39092D,S39093D,S4400XD,S4401XD,S4402XD,S4410XD,S4411XD S4412XD,S4420XD,S4421XD,S4422XD,S4430XD,S4431XD,S4432XD S4440XD,S4441XD,S4442XD,S4450XD,S4451XD,S4452XD,S448X1D S448X2D,S448X9D,S4490XD,S4491XD,S4492XD,S46001D,S46002D S46009D,S46091D,S46092D,S46099D,S46101D,S46102D,S46109D S46191D,S46192D,S46199D,S46201D,S46202D,S46209D,S46291D S46292D,S46299D,S46301D,S46302D,S46309D,S46391D,S46392D S46399D,S46801D,S46802D,S46809D,S46891D,S46892D,S46899D S46901D,S46902D,S46909D,S46991D,S46992D,S46999D,S5400XD S5401XD,S5402XD,S5410XD,S5411XD,S5412XD,S5420XD,S5421XD S5422XD,S5430XD,S5431XD,S5432XD,S548X1D,S548X2D,S548X9D S5490XD,S5491XD,S5492XD,S56001D,S56002D,S56009D,S56091D S56092D,S56099D,S56101D,S56102D,S56103D,S56104D,S56105D S56106D,S56107D,S56108D,S56109D,S56191D,S56192D,S56193D S56194D,S56195D,S56196D,S56197D,S56198D,S56199D,S56201D S56202D,S56209D,S56291D,S56292D,S56299D,S56301D,S56302D S56309D,S56391D,S56392D,S56399D,S56401D,S56402D,S56403D S56404D,S56405D,S56406D,S56407D,S56408D,S56409D,S56491D S56492D,S56493D,S56494D,S56495D,S56496D,S56497D,S56498D S56499D,S56501D,S56502D,S56509D,S56591D,S56592D,S56599D S56801D,S56802D,S56809D,S56891D,S56892D,S56899D,S56901D S56902D,S56909D,S56991D,S56992D,S56999D,S6400XD,S6401XD S6402XD,S6410XD,S6411XD,S6412XD,S6420XD,S6421XD,S6422XD | V5889 |

Continued on next page

**Supplementary Table S1:** (cont'd) A comprehensive mapping of diagnosis groups used in the experiments to their corresponding ICD-10 and ICD-9 codes.

| Diagnosis | ICD-10 | ICD-9 |
|---|---|---|
| | S6430XD,S6431XD,S6432XD,S6440XD,S64490D,S64491D,S64492D | |
| | S64493D,S64494D,S64495D,S64496D,S64497D,S64498D,S648X1D | |
| | S648X2D,S648X9D,S6490XD,S6491XD,S6492XD,S66001D,S66002D | |
| | S66009D,S66091D,S66092D,S66099D,S66100D,S66101D,S66102D | |
| | S66103D,S66104D,S66105D,S66106D,S66107D,S66108D,S66109D | |
| | S66190D,S66191D,S66192D,S66193D,S66194D,S66195D,S66196D | |
| | S66197D,S66198D,S66199D,S66201D,S66202D,S66209D,S66291D | |
| | S66292D,S66299D,S66300D,S66301D,S66302D,S66303D,S66304D | |
| | S66305D,S66306D,S66307D,S66308D,S66309D,S66390D,S66391D | |
| | S66392D,S66393D,S66394D,S66395D,S66396D,S66397D,S66398D | |
| | S66399D,S66401D,S66402D,S66409D,S66491D,S66492D,S66499D | |
| | S66500D,S66501D,S66502D,S66503D,S66504D,S66505D,S66506D | |
| | S66507D,S66508D,S66509D,S66590D,S66591D,S66592D,S66593D | |
| | S66594D,S66595D,S66596D,S66597D,S66598D,S66599D,S66801D | |
| | S66802D,S66809D,S66891D,S66892D,S66899D,S66901D,S66902D | |
| | S66909D,S66991D,S66992D,S66999D,S7400XD,S7401XD,S7402XD | |
| | S7410XD,S7411XD,S7412XD,S7420XD,S7421XD,S7422XD,S748X1D | |
| | S748X2D,S748X9D,S7490XD,S7491XD,S7492XD,S76001D,S76002D | |
| | S76009D,S76091D,S76092D,S76099D,S76101D,S76102D,S76109D | |
| | S76191D,S76192D,S76199D,S76201D,S76202D,S76209D,S76291D | |
| | S76292D,S76299D,S76301D,S76302D,S76309D,S76391D,S76392D | |
| | S76399D,S76801D,S76802D,S76809D,S76891D,S76892D,S76899D | |
| | S76901D,S76902D,S76909D,S76991D,S76992D,S76999D,S8400XD | |
| | S8401XD,S8402XD,S8410XD,S8411XD,S8412XD,S8420XD,S8421XD | |
| | S8422XD,S84801D,S84802D,S84809D,S8490XD,S8491XD,S8492XD | |
| | S86001D,S86002D,S86009D,S86091D,S86092D,S86099D,S86101D | |
| | S86102D,S86109D,S86191D,S86192D,S86199D,S86201D,S86202D | |
| | S86209D,S86291D,S86292D,S86299D,S86301D,S86302D,S86309D | |
| | S86391D,S86392D,S86399D,S86801D,S86802D,S86809D,S86891D | |
| | S86892D,S86899D,S86901D,S86902D,S86909D,S86991D,S86992D | |
| | S86999D,S9400XD,S9401XD,S9402XD,S9410XD,S9411XD,S9412XD | |
| | S9420XD,S9421XD,S9422XD,S9430XD,S9431XD,S9432XD,S948X1D | |
| | S948X2D,S948X9D,S9490XD,S9491XD,S9492XD,S96001D,S96002D | |
| | S96009D,S96091D,S96092D,S96099D,S96101D,S96102D,S96109D | |
| | S96191D,S96192D,S96199D,S96201D,S96202D,S96209D,S96291D | |
| | S96292D,S96299D,S96801D,S96802D,S96809D,S96891D,S96892D | |
| | S96899D,S96901D,S96902D,S96909D,S96991D,S96992D,S96999D | |
| Injury, sequela | M9701XS,M9702XS,M9711XS,M9712XS,M9721XS,M9722XS,M9731XS | 9050,9051,9052,9053,9054,9056,9057,9059,9060,9061,9062,9063 |
| | M9732XS,M9741XS,M9742XS,M978XXS,M979XXS,S0000XS,S0001XS | 9064,9065,9066,9067,9068,9070,9071,9072,9073,9074,9075,9080 |

Continued on next page

**Supplementary Table S1:** (cont'd) A comprehensive mapping of diagnosis groups used in the experiments to their corresponding ICD-10 and ICD-9 codes.

| Diagnosis | ICD-10 | ICD-9 |
|---|---|---|
| | S0002XS,S0003XS,S0004XS,S0005XS,S0006XS,S0007XS,S0010XS | 9081,9082,9083,9084,9085,9086,9089,9093,9099 |
| | S0011XS,S0012XS,S00201S,S00202S,S00209S,S00211S,S00212S | |
| | S00219S,S00221S,S00222S,S00229S,S00241S,S00242S,S00249S | |
| | S00251S,S00252S,S00259S,S00261S,S00262S,S00269S,S00271S | |
| | S00272S,S00279S,S0030XS,S0031XS,S0032XS,S0033XS,S0034XS | |
| | S0035XS,S0036XS,S0037XS,S00401S,S00402S,S00409S,S00411S | |
| | S00412S,S00419S,S00421S,S00422S,S00429S,S00431S,S00432S | |
| | S00439S,S00441S,S00442S,S00449S,S00451S,S00452S,S00459S | |
| | S00461S,S00462S,S00469S,S00471S,S00472S,S00479S,S00501S | |
| | S00502S,S00511S,S00512S,S00521S,S00522S,S00531S,S00532S | |
| | S00541S,S00542S,S00551S,S00552S,S00561S,S00562S,S00571S | |
| | S00572S,S0080XS,S0081XS,S0082XS,S0083XS,S0084XS,S0085XS | |
| | S0086XS,S0087XS,S0090XS,S0091XS,S0092XS,S0093XS,S0094XS | |
| | S0095XS,S0096XS,S0097XS,S0100XS,S0101XS,S0102XS,S0103XS | |
| | S0104XS,S0105XS,S01101S,S01102S,S01109S,S01111S,S01112S | |
| | S01119S,S01121S,S01122S,S01129S,S01131S,S01132S,S01139S | |
| | S01141S,S01142S,S01149S,S01151S,S01152S,S01159S,S0120XS | |
| | S0121XS,S0122XS,S0123XS,S0124XS,S0125XS,S01301S,S01302S | |
| | S01309S,S01311S,S01312S,S01319S,S01321S,S01322S,S01329S | |
| | S01331S,S01332S,S01339S,S01341S,S01342S,S01349S,S01351S | |
| | S01352S,S01359S,S01401S,S01402S,S01409S,S01411S,S01412S | |
| | S01419S,S01421S,S01422S,S01429S,S01431S,S01432S,S01439S | |
| | S01441S,S01442S,S01449S,S01451S,S01452S,S01459S,S01501S | |
| | S01502S,S01511S,S01512S,S01521S,S01522S,S01531S,S01532S | |
| | S01541S,S01542S,S01551S,S01552S,S0180XS,S0181XS,S0182XS | |
| | S0183XS,S0184XS,S0185XS,S0190XS,S0191XS,S0192XS,S0193XS | |
| | S0194XS,S0195XS,S020XXS,S02101S,S02102S,S02109S,S0210XS | |
| | S02110S,S02111S,S02112S,S02113S,S02118S,S02119S,S0211AS | |
| | S0211BS,S0211CS,S0211DS,S0211ES,S0211FS,S0211GS,S0211HS | |
| | S02121S,S02122S,S02129S,S0219XS,S022XXS,S0230XS,S0231XS | |
| | S0232XS,S023XXS,S02400S,S02401S,S02402S,S0240AS,S0240BS | |
| | S0240CS,S0240DS,S0240ES,S0240FS,S02411S,S02412S,S02413S | |
| | S0242XS,S025XXS,S02600S,S02601S,S02602S,S02609S,S02610S | |
| | S02611S,S02612S,S0261XS,S02620S,S02621S,S02622S,S0262XS | |
| | S02630S,S02631S,S02632S,S0263XS,S02640S,S02641S,S02642S | |
| | S0264XS,S02650S,S02651S,S02652S,S0265XS,S0266XS,S02670S | |
| | S02671S,S02672S,S0267XS,S0269XS,S0280XS,S0281XS,S0282XS | |
| | S02831S,S02832S,S02839S,S02841S,S02842S,S02849S,S0285XS | |
| | S028XXS,S0291XS,S0292XS,S0300XS,S0301XS,S0302XS,S0303XS | |
| | S030XXS,S031XXS,S032XXS,S0340XS,S0341XS,S0342XS,S0343XS | |

Continued on next page

**Supplementary Table S1:** (cont'd) A comprehensive mapping of diagnosis groups used in the experiments to their corresponding ICD-10 and ICD-9 codes.

| Diagnosis | ICD-10 | ICD-9 |
|---|---|---|
| | S034XXS,S038XXS,S039XXS,S04011S,S04012S,S04019S,S0402XS | |
| | S04031S,S04032S,S04039S,S04041S,S04042S,S04049S,S0410XS | |
| | S0411XS,S0412XS,S0420XS,S0421XS,S0422XS,S0430XS,S0431XS | |
| | S0432XS,S0440XS,S0441XS,S0442XS,S0450XS,S0451XS,S0452XS | |
| | S0460XS,S0461XS,S0462XS,S0470XS,S0471XS,S0472XS,S04811S | |
| | S04812S,S04819S,S04891S,S04892S,S04899S,S049XXS,S0500XS | |
| | S0501XS,S0502XS,S0510XS,S0511XS,S0512XS,S0520XS,S0521XS | |
| | S0522XS,S0530XS,S0531XS,S0532XS,S0540XS,S0541XS,S0542XS | |
| | S0550XS,S0551XS,S0552XS,S0560XS,S0561XS,S0562XS,S0570XS | |
| | S0571XS,S0572XS,S058X1S,S058X2S,S058X9S,S0590XS,S0591XS | |
| | S0592XS,S060X0S,S060X1S,S060X2S,S060X3S,S060X4S,S060X5S | |
| | S060X6S,S060X7S,S060X8S,S060X9S,S060XAS,S061X0S,S061X1S | |
| | S061X2S,S061X3S,S061X4S,S061X5S,S061X6S,S061X7S,S061X8S | |
| | S061X9S,S061XAS,S062X0S,S062X1S,S062X2S,S062X3S,S062X4S | |
| | S062X5S,S062X6S,S062X7S,S062X8S,S062X9S,S062XAS,S06300S | |
| | S06301S,S06302S,S06303S,S06304S,S06305S,S06306S,S06307S | |
| | S06308S,S06309S,S0630AS,S06310S,S06311S,S06312S,S06313S | |
| | S06314S,S06315S,S06316S,S06317S,S06318S,S06319S,S0631AS | |
| | S06320S,S06321S,S06322S,S06323S,S06324S,S06325S,S06326S | |
| | S06327S,S06328S,S06329S,S0632AS,S06330S,S06331S,S06332S | |
| | S06333S,S06334S,S06335S,S06336S,S06337S,S06338S,S06339S | |
| | S0633AS,S06340S,S06341S,S06342S,S06343S,S06344S,S06345S | |
| | S06346S,S06347S,S06348S,S06349S,S0634AS,S06350S,S06351S | |
| | S06352S,S06353S,S06354S,S06355S,S06356S,S06357S,S06358S | |
| | S06359S,S0635AS,S06360S,S06361S,S06362S,S06363S,S06364S | |
| | S06365S,S06366S,S06367S,S06368S,S06369S,S0636AS,S06370S | |
| | S06371S,S06372S,S06373S,S06374S,S06375S,S06376S,S06377S | |
| | S06378S,S06379S,S0637AS,S06380S,S06381S,S06382S,S06383S | |
| | S06384S,S06385S,S06386S,S06387S,S06388S,S06389S,S0638AS | |
| | S064X0S,S064X1S,S064X2S,S064X3S,S064X4S,S064X5S,S064X6S | |
| | S064X7S,S064X8S,S064X9S,S064XAS,S065X0S,S065X1S,S065X2S | |
| | S065X3S,S065X4S,S065X5S,S065X6S,S065X7S,S065X8S,S065X9S | |
| | S065XAS,S066X0S,S066X1S,S066X2S,S066X3S,S066X4S,S066X5S | |
| | S066X6S,S066X7S,S066X8S,S066X9S,S066XAS,S06810S,S06811S | |
| | S06812S,S06813S,S06814S,S06815S,S06816S,S06817S,S06818S | |
| | S06819S,S0681AS,S06820S,S06821S,S06822S,S06823S,S06824S | |
| | S06825S,S06826S,S06827S,S06828S,S06829S,S0682AS,S06890S | |
| | S06891S,S06892S,S06893S,S06894S,S06895S,S06896S,S06897S | |
| | S06898S,S06899S,S0689AS,S068A0S,S068A1S,S068A2S,S068A3S | |
| | S068A4S,S068A5S,S068A6S,S068A9S,S068AAS,S069X0S,S069X1S | |

Continued on next page

**Supplementary Table S1:** (cont'd) A comprehensive mapping of diagnosis groups used in the experiments to their corresponding ICD-10 and ICD-9 codes.

| Diagnosis | ICD-10 | ICD-9 |
|---|---|---|
| | S069X2S,S069X3S,S069X4S,S069X5S,S069X6S,S069X7S,S069X8S | |
| | S069X9S,S069XAS,S06A0XS,S06A1XS,S070XXS,S071XXS,S078XXS | |
| | S079XXS,S080XXS,S08111S,S08112S,S08119S,S08121S,S08122S | |
| | S08129S,S08811S,S08812S,S0889XS,S090XXS,S0910XS,S0911XS | |
| | S0912XS,S0919XS,S0920XS,S0921XS,S0922XS,S09301S,S09302S | |
| | S09309S,S09311S,S09312S,S09313S,S09319S,S09391S,S09392S | |
| | S09399S,S098XXS,S0990XS,S0991XS,S0992XS,S0993XS,S100XXS | |
| | S1010XS,S1011XS,S1012XS,S1014XS,S1015XS,S1016XS,S1017XS | |
| | S1080XS,S1081XS,S1082XS,S1083XS,S1084XS,S1085XS,S1086XS | |
| | S1087XS,S1090XS,S1091XS,S1092XS,S1093XS,S1094XS,S1095XS | |
| | S1096XS,S1097XS,S11011S,S11012S,S11013S,S11014S,S11015S | |
| | S11019S,S11021S,S11022S,S11023S,S11024S,S11025S,S11029S | |
| | S11031S,S11032S,S11033S,S11034S,S11035S,S11039S,S1110XS | |
| | S1111XS,S1112XS,S1113XS,S1114XS,S1115XS,S1120XS,S1121XS | |
| | S1122XS,S1123XS,S1124XS,S1125XS,S1180XS,S1181XS,S1182XS | |
| | S1183XS,S1184XS,S1185XS,S1189XS,S1190XS,S1191XS,S1192XS | |
| | S1193XS,S1194XS,S1195XS,S12000S,S12001S,S1201XS,S1202XS | |
| | S12030S,S12031S,S12040S,S12041S,S12090S,S12091S,S12100S | |
| | S12101S,S12110S,S12111S,S12112S,S12120S,S12121S,S12130S | |
| | S12131S,S1214XS,S12150S,S12151S,S12190S,S12191S,S12200S | |
| | S12201S,S12230S,S12231S,S1224XS,S12250S,S12251S,S12290S | |
| | S12291S,S12300S,S12301S,S12330S,S12331S,S1234XS,S12350S | |
| | S12351S,S12390S,S12391S,S12400S,S12401S,S12430S,S12431S | |
| | S1244XS,S12450S,S12451S,S12490S,S12491S,S12500S,S12501S | |
| | S12530S,S12531S,S1254XS,S12550S,S12551S,S12590S,S12591S | |
| | S12600S,S12601S,S12630S,S12631S,S1264XS,S12650S,S12651S | |
| | S12690S,S12691S,S128XXS,S129XXS,S130XXS,S13100S,S13101S | |
| | S13110S,S13111S,S13120S,S13121S,S13130S,S13131S,S13140S | |
| | S13141S,S13150S,S13151S,S13160S,S13161S,S13170S,S13171S | |
| | S13180S,S13181S,S1320XS,S1329XS,S134XXS,S135XXS,S138XXS | |
| | S139XXS,S140XXS,S14101S,S14102S,S14103S,S14104S,S14105S | |
| | S14106S,S14107S,S14108S,S14109S,S14111S,S14112S,S14113S | |
| | S14114S,S14115S,S14116S,S14117S,S14118S,S14119S,S14121S | |
| | S14122S,S14123S,S14124S,S14125S,S14126S,S14127S,S14128S | |
| | S14129S,S14131S,S14132S,S14133S,S14134S,S14135S,S14136S | |
| | S14137S,S14138S,S14139S,S14141S,S14142S,S14143S,S14144S | |
| | S14145S,S14146S,S14147S,S14148S,S14149S,S14151S,S14152S | |
| | S14153S,S14154S,S14155S,S14156S,S14157S,S14158S,S14159S | |
| | S142XXS,S143XXS,S144XXS,S145XXS,S148XXS,S149XXS,S15001S | |
| | S15002S,S15009S,S15011S,S15012S,S15019S,S15021S,S15022S | |

Continued on next page

**Supplementary Table S1:** (cont'd) A comprehensive mapping of diagnosis groups used in the experiments to their corresponding ICD-10 and ICD-9 codes.

| Diagnosis | ICD-10 | ICD-9 |
|---|---|---|
| | S15029S,S15091S,S15092S,S15099S,S15101S,S15102S,S15109S | |
| | S15111S,S15112S,S15119S,S15121S,S15122S,S15129S,S15191S | |
| | S15192S,S15199S,S15201S,S15202S,S15209S,S15211S,S15212S | |
| | S15219S,S15221S,S15222S,S15229S,S15291S,S15292S,S15299S | |
| | S15301S,S15302S,S15309S,S15311S,S15312S,S15319S,S15321S | |
| | S15322S,S15329S,S15391S,S15392S,S15399S,S158XXS,S159XXS | |
| | S161XXS,S162XXS,S168XXS,S169XXS,S170XXS,S178XXS,S179XXS | |
| | S1980XS,S1981XS,S1982XS,S1983XS,S1984XS,S1985XS,S1989XS | |
| | S199XXS,S2000XS,S2001XS,S2002XS,S20101S,S20102S,S20109S | |
| | S20111S,S20112S,S20119S,S20121S,S20122S,S20129S,S20141S | |
| | S20142S,S20149S,S20151S,S20152S,S20159S,S20161S,S20162S | |
| | S20169S,S20171S,S20172S,S20179S,S2020XS,S20211S,S20212S | |
| | S20213S,S20214S,S20219S,S20221S,S20222S,S20223S,S20224S | |
| | S20229S,S20301S,S20302S,S20303S,S20304S,S20309S,S20311S | |
| | S20312S,S20313S,S20314S,S20319S,S20321S,S20322S,S20323S | |
| | S20324S,S20329S,S20341S,S20342S,S20343S,S20344S,S20349S | |
| | S20351S,S20352S,S20353S,S20354S,S20359S,S20361S,S20362S | |
| | S20363S,S20364S,S20369S,S20371S,S20372S,S20373S,S20374S | |
| | S20379S,S20401S,S20402S,S20409S,S20411S,S20412S,S20419S | |
| | S20421S,S20422S,S20429S,S20441S,S20442S,S20449S,S20451S | |
| | S20452S,S20459S,S20461S,S20462S,S20469S,S20471S,S20472S | |
| | S20479S,S2090XS,S2091XS,S2092XS,S2094XS,S2095XS,S2096XS | |
| | S2097XS,S21001S,S21002S,S21009S,S21011S,S21012S,S21019S | |
| | S21021S,S21022S,S21029S,S21031S,S21032S,S21039S,S21041S | |
| | S21042S,S21049S,S21051S,S21052S,S21059S,S21101S,S21102S | |
| | S21109S,S21111S,S21112S,S21119S,S21121S,S21122S,S21129S | |
| | S21131S,S21132S,S21139S,S21141S,S21142S,S21149S,S21151S | |
| | S21152S,S21159S,S21201S,S21202S,S21209S,S21211S,S21212S | |
| | S21219S,S21221S,S21222S,S21229S,S21231S,S21232S,S21239S | |
| | S21241S,S21242S,S21249S,S21251S,S21252S,S21259S,S21301S | |
| | S21302S,S21309S,S21311S,S21312S,S21319S,S21321S,S21322S | |
| | S21329S,S21331S,S21332S,S21339S,S21341S,S21342S,S21349S | |
| | S21351S,S21352S,S21359S,S21401S,S21402S,S21409S,S21411S | |
| | S21412S,S21419S,S21421S,S21422S,S21429S,S21431S,S21432S | |
| | S21439S,S21441S,S21442S,S21449S,S21451S,S21452S,S21459S | |
| | S2190XS,S2191XS,S2192XS,S2193XS,S2194XS,S2195XS,S22000S | |
| | S22001S,S22002S,S22008S,S22009S,S22010S,S22011S,S22012S | |
| | S22018S,S22019S,S22020S,S22021S,S22022S,S22028S,S22029S | |
| | S22030S,S22031S,S22032S,S22038S,S22039S,S22040S,S22041S | |
| | S22042S,S22048S,S22049S,S22050S,S22051S,S22052S,S22058S | |

Continued on next page

**Supplementary Table S1:** (cont'd) A comprehensive mapping of diagnosis groups used in the experiments to their corresponding ICD-10 and ICD-9 codes.

| Diagnosis | ICD-10 | ICD-9 |
|---|---|---|
| | S22059S,S22060S,S22061S,S22062S,S22068S,S22069S,S22070S | |
| | S22071S,S22072S,S22078S,S22079S,S22080S,S22081S,S22082S | |
| | S22088S,S22089S,S2220XS,S2221XS,S2222XS,S2223XS,S2224XS | |
| | S2231XS,S2232XS,S2239XS,S2241XS,S2242XS,S2243XS,S2249XS | |
| | S225XXS,S229XXS,S230XXS,S23100S,S23101S,S23110S,S23111S | |
| | S23120S,S23121S,S23122S,S23123S,S23130S,S23131S,S23132S | |
| | S23133S,S23140S,S23141S,S23142S,S23143S,S23150S,S23151S | |
| | S23152S,S23153S,S23160S,S23161S,S23162S,S23163S,S23170S | |
| | S23171S,S2320XS,S2329XS,S233XXS,S2341XS,S23420S,S23421S | |
| | S23428S,S23429S,S238XXS,S239XXS,S240XXS,S24101S,S24102S | |
| | S24103S,S24104S,S24109S,S24111S,S24112S,S24113S,S24114S | |
| | S24119S,S24131S,S24132S,S24133S,S24134S,S24139S,S24141S | |
| | S24142S,S24143S,S24144S,S24149S,S24151S,S24152S,S24153S | |
| | S24154S,S24159S,S242XXS,S243XXS,S244XXS,S248XXS,S249XXS | |
| | S2500XS,S2501XS,S2502XS,S2509XS,S25101S,S25102S,S25109S | |
| | S25111S,S25112S,S25119S,S25121S,S25122S,S25129S,S25191S | |
| | S25192S,S25199S,S2520XS,S2521XS,S2522XS,S2529XS,S25301S | |
| | S25302S,S25309S,S25311S,S25312S,S25319S,S25321S,S25322S | |
| | S25329S,S25391S,S25392S,S25399S,S25401S,S25402S,S25409S | |
| | S25411S,S25412S,S25419S,S25421S,S25422S,S25429S,S25491S | |
| | S25492S,S25499S,S25501S,S25502S,S25509S,S25511S,S25512S | |
| | S25519S,S25591S,S25592S,S25599S,S25801S,S25802S,S25809S | |
| | S25811S,S25812S,S25819S,S25891S,S25892S,S25899S,S2590XS | |
| | S2591XS,S2599XS,S2600XS,S2601XS,S26020S,S26021S,S26022S | |
| | S2609XS,S2610XS,S2611XS,S2612XS,S2619XS,S2690XS,S2691XS | |
| | S2692XS,S2699XS,S270XXS,S271XXS,S272XXS,S27301S,S27302S | |
| | S27309S,S27311S,S27312S,S27319S,S27321S,S27322S,S27329S | |
| | S27331S,S27332S,S27339S,S27391S,S27392S,S27399S,S27401S | |
| | S27402S,S27409S,S27411S,S27412S,S27419S,S27421S,S27422S | |
| | S27429S,S27431S,S27432S,S27439S,S27491S,S27492S,S27499S | |
| | S2750XS,S2751XS,S2752XS,S2753XS,S2759XS,S2760XS,S2763XS | |
| | S2769XS,S27802S,S27803S,S27808S,S27809S,S27812S,S27813S | |
| | S27818S,S27819S,S27892S,S27893S,S27898S,S27899S,S279XXS | |
| | S280XXS,S281XXS,S28211S,S28212S,S28219S,S28221S,S28222S | |
| | S28229S,S29001S,S29002S,S29009S,S29011S,S29012S,S29019S | |
| | S29021S,S29022S,S29029S,S29091S,S29092S,S29099S,S298XXS | |
| | S299XXS,S300XXS,S301XXS,S30201S,S30202S,S3021XS,S3022XS | |
| | S3023XS,S303XXS,S30810S,S30811S,S30812S,S30813S,S30814S | |
| | S30815S,S30816S,S30817S,S30820S,S30821S,S30822S,S30823S | |
| | S30824S,S30825S,S30826S,S30827S,S30840S,S30841S,S30842S | |

Continued on next page

**Supplementary Table S1:** (cont'd) A comprehensive mapping of diagnosis groups used in the experiments to their corresponding ICD-10 and ICD-9 codes.

| Diagnosis | ICD-10 | ICD-9 |
|---|---|---|
| | S30843S,S30844S,S30845S,S30846S,S30850S,S30851S,S30852S | |
| | S30853S,S30854S,S30855S,S30856S,S30857S,S30860S,S30861S | |
| | S30862S,S30863S,S30864S,S30865S,S30866S,S30867S,S30870S | |
| | S30871S,S30872S,S30873S,S30874S,S30875S,S30876S,S30877S | |
| | S3091XS,S3092XS,S3093XS,S3094XS,S3095XS,S3096XS,S3097XS | |
| | S3098XS,S31000S,S31001S,S31010S,S31011S,S31020S,S31021S | |
| | S31030S,S31031S,S31040S,S31041S,S31050S,S31051S,S31100S | |
| | S31101S,S31102S,S31103S,S31104S,S31105S,S31109S,S31110S | |
| | S31111S,S31112S,S31113S,S31114S,S31115S,S31119S,S31120S | |
| | S31121S,S31122S,S31123S,S31124S,S31125S,S31129S,S31130S | |
| | S31131S,S31132S,S31133S,S31134S,S31135S,S31139S,S31140S | |
| | S31141S,S31142S,S31143S,S31144S,S31145S,S31149S,S31150S | |
| | S31151S,S31152S,S31153S,S31154S,S31155S,S31159S,S3120XS | |
| | S3121XS,S3122XS,S3123XS,S3124XS,S3125XS,S3130XS,S3131XS | |
| | S3132XS,S3133XS,S3134XS,S3135XS,S3140XS,S3141XS,S3142XS | |
| | S3143XS,S3144XS,S3145XS,S31501S,S31502S,S31511S,S31512S | |
| | S31521S,S31522S,S31531S,S31532S,S31541S,S31542S,S31551S | |
| | S31552S,S31600S,S31601S,S31602S,S31603S,S31604S,S31605S | |
| | S31609S,S31610S,S31611S,S31612S,S31613S,S31614S,S31615S | |
| | S31619S,S31620S,S31621S,S31622S,S31623S,S31624S,S31625S | |
| | S31629S,S31630S,S31631S,S31632S,S31633S,S31634S,S31635S | |
| | S31639S,S31640S,S31641S,S31642S,S31643S,S31644S,S31645S | |
| | S31649S,S31650S,S31651S,S31652S,S31653S,S31654S,S31655S | |
| | S31659S,S31801S,S31802S,S31803S,S31804S,S31805S,S31809S | |
| | S31811S,S31812S,S31813S,S31814S,S31815S,S31819S,S31821S | |
| | S31822S,S31823S,S31824S,S31825S,S31829S,S31831S,S31832S | |
| | S31833S,S31834S,S31835S,S31839S,S32000S,S32001S,S32002S | |
| | S32008S,S32009S,S32010S,S32011S,S32012S,S32018S,S32019S | |
| | S32020S,S32021S,S32022S,S32028S,S32029S,S32030S,S32031S | |
| | S32032S,S32038S,S32039S,S32040S,S32041S,S32042S,S32048S | |
| | S32049S,S32050S,S32051S,S32052S,S32058S,S32059S,S3210XS | |
| | S32110S,S32111S,S32112S,S32119S,S32120S,S32121S,S32122S | |
| | S32129S,S32130S,S32131S,S32132S,S32139S,S3214XS,S3215XS | |
| | S3216XS,S3217XS,S3219XS,S322XXS,S32301S,S32302S,S32309S | |
| | S32311S,S32312S,S32313S,S32314S,S32315S,S32316S,S32391S | |
| | S32392S,S32399S,S32401S,S32402S,S32409S,S32411S,S32412S | |
| | S32413S,S32414S,S32415S,S32416S,S32421S,S32422S,S32423S | |
| | S32424S,S32425S,S32426S,S32431S,S32432S,S32433S,S32434S | |
| | S32435S,S32436S,S32441S,S32442S,S32443S,S32444S,S32445S | |
| | S32446S,S32451S,S32452S,S32453S,S32454S,S32455S,S32456S | |

Continued on next page

**Supplementary Table S1:** (cont'd) A comprehensive mapping of diagnosis groups used in the experiments to their corresponding ICD-10 and ICD-9 codes.

| Diagnosis | ICD-10 | ICD-9 |
|---|---|---|
| | S32461S,S32462S,S32463S,S32464S,S32465S,S32466S,S32471S | |
| | S32472S,S32473S,S32474S,S32475S,S32476S,S32481S,S32482S | |
| | S32483S,S32484S,S32485S,S32486S,S32491S,S32492S,S32499S | |
| | S32501S,S32502S,S32509S,S32511S,S32512S,S32519S,S32591S | |
| | S32592S,S32599S,S32601S,S32602S,S32609S,S32611S,S32612S | |
| | S32613S,S32614S,S32615S,S32616S,S32691S,S32692S,S32699S | |
| | S32810S,S32811S,S3282XS,S3289XS,S329XXS,S330XXS,S33100S | |
| | S33101S,S33110S,S33111S,S33120S,S33121S,S33130S,S33131S | |
| | S33140S,S33141S,S332XXS,S3330XS,S3339XS,S334XXS,S335XXS | |
| | S336XXS,S338XXS,S339XXS,S3401XS,S3402XS,S34101S,S34102S | |
| | S34103S,S34104S,S34105S,S34109S,S34111S,S34112S,S34113S | |
| | S34114S,S34115S,S34119S,S34121S,S34122S,S34123S,S34124S | |
| | S34125S,S34129S,S34131S,S34132S,S34139S,S3421XS,S3422XS | |
| | S343XXS,S344XXS,S345XXS,S346XXS,S348XXS,S349XXS,S3500XS | |
| | S3501XS,S3502XS,S3509XS,S3510XS,S3511XS,S3512XS,S3519XS | |
| | S35211S,S35212S,S35218S,S35219S,S35221S,S35222S,S35228S | |
| | S35229S,S35231S,S35232S,S35238S,S35239S,S35291S,S35292S | |
| | S35298S,S35299S,S35311S,S35318S,S35319S,S35321S,S35328S | |
| | S35329S,S35331S,S35338S,S35339S,S35341S,S35348S,S35349S | |
| | S35401S,S35402S,S35403S,S35404S,S35405S,S35406S,S35411S | |
| | S35412S,S35413S,S35414S,S35415S,S35416S,S35491S,S35492S | |
| | S35493S,S35494S,S35495S,S35496S,S3550XS,S35511S,S35512S | |
| | S35513S,S35514S,S35515S,S35516S,S35531S,S35532S,S35533S | |
| | S35534S,S35535S,S35536S,S3559XS,S358X1S,S358X8S,S358X9S | |
| | S3590XS,S3591XS,S3599XS,S3600XS,S36020S,S36021S,S36029S | |
| | S36030S,S36031S,S36032S,S36039S,S3609XS,S36112S,S36113S | |
| | S36114S,S36115S,S36116S,S36118S,S36119S,S36122S,S36123S | |
| | S36128S,S36129S,S3613XS,S36200S,S36201S,S36202S,S36209S | |
| | S36220S,S36221S,S36222S,S36229S,S36230S,S36231S,S36232S | |
| | S36239S,S36240S,S36241S,S36242S,S36249S,S36250S,S36251S | |
| | S36252S,S36259S,S36260S,S36261S,S36262S,S36269S,S36290S | |
| | S36291S,S36292S,S36299S,S3630XS,S3632XS,S3633XS,S3639XS | |
| | S36400S,S36408S,S36409S,S36410S,S36418S,S36419S,S36420S | |
| | S36428S,S36429S,S36430S,S36438S,S36439S,S36490S,S36498S | |
| | S36499S,S36500S,S36501S,S36502S,S36503S,S36508S,S36509S | |
| | S36510S,S36511S,S36512S,S36513S,S36518S,S36519S,S36520S | |
| | S36521S,S36522S,S36523S,S36528S,S36529S,S36530S,S36531S | |
| | S36532S,S36533S,S36538S,S36539S,S36590S,S36591S,S36592S | |
| | S36593S,S36598S,S36599S,S3660XS,S3661XS,S3662XS,S3663XS | |
| | S3669XS,S3681XS,S36892S,S36893S,S36898S,S36899S,S3690XS | |

Continued on next page

**Supplementary Table S1:** (cont'd) A comprehensive mapping of diagnosis groups used in the experiments to their corresponding ICD-10 and ICD-9 codes.

| Diagnosis | ICD-10 | ICD-9 |
|---|---|---|
| | S3692XS,S3693XS,S3699XS,S37001S,S37002S,S37009S,S37011S | |
| | S37012S,S37019S,S37021S,S37022S,S37029S,S37031S,S37032S | |
| | S37039S,S37041S,S37042S,S37049S,S37051S,S37052S,S37059S | |
| | S37061S,S37062S,S37069S,S37091S,S37092S,S37099S,S3710XS | |
| | S3712XS,S3713XS,S3719XS,S3720XS,S3722XS,S3723XS,S3729XS | |
| | S3730XS,S3732XS,S3733XS,S3739XS,S37401S,S37402S,S37409S | |
| | S37421S,S37422S,S37429S,S37431S,S37432S,S37439S,S37491S | |
| | S37492S,S37499S,S37501S,S37502S,S37509S,S37511S,S37512S | |
| | S37519S,S37521S,S37522S,S37529S,S37531S,S37532S,S37539S | |
| | S37591S,S37592S,S37599S,S3760XS,S3762XS,S3763XS,S3769XS | |
| | S37812S,S37813S,S37818S,S37819S,S37822S,S37823S,S37828S | |
| | S37829S,S37892S,S37893S,S37898S,S37899S,S3790XS,S3792XS | |
| | S3793XS,S3799XS,S38001S,S38002S,S3801XS,S3802XS,S3803XS | |
| | S381XXS,S38211S,S38212S,S38221S,S38222S,S38231S,S38232S | |
| | S383XXS,S39001S,S39002S,S39003S,S39011S,S39012S,S39013S | |
| | S39021S,S39022S,S39023S,S39091S,S39092S,S39093S,S3981XS | |
| | S3982XS,S3983XS,S39840S,S39848S,S3991XS,S3992XS,S3993XS | |
| | S3994XS,S40011S,S40012S,S40019S,S40021S,S40022S,S40029S | |
| | S40211S,S40212S,S40219S,S40221S,S40222S,S40229S,S40241S | |
| | S40242S,S40249S,S40251S,S40252S,S40259S,S40261S,S40262S | |
| | S40269S,S40271S,S40272S,S40279S,S40811S,S40812S,S40819S | |
| | S40821S,S40822S,S40829S,S40841S,S40842S,S40849S,S40851S | |
| | S40852S,S40859S,S40861S,S40862S,S40869S,S40871S,S40872S | |
| | S40879S,S40911S,S40912S,S40919S,S40921S,S40922S,S40929S | |
| | S41001S,S41002S,S41009S,S41011S,S41012S,S41019S,S41021S | |
| | S41022S,S41029S,S41031S,S41032S,S41039S,S41041S,S41042S | |
| | S41049S,S41051S,S41052S,S41059S,S41101S,S41102S,S41109S | |
| | S41111S,S41112S,S41119S,S41121S,S41122S,S41129S,S41131S | |
| | S41132S,S41139S,S41141S,S41142S,S41149S,S41151S,S41152S | |
| | S41159S,S42001S,S42002S,S42009S,S42011S,S42012S,S42013S | |
| | S42014S,S42015S,S42016S,S42017S,S42018S,S42019S,S42021S | |
| | S42022S,S42023S,S42024S,S42025S,S42026S,S42031S,S42032S | |
| | S42033S,S42034S,S42035S,S42036S,S42101S,S42102S,S42109S | |
| | S42111S,S42112S,S42113S,S42114S,S42115S,S42116S,S42121S | |
| | S42122S,S42123S,S42124S,S42125S,S42126S,S42131S,S42132S | |
| | S42133S,S42134S,S42135S,S42136S,S42141S,S42142S,S42143S | |
| | S42144S,S42145S,S42146S,S42151S,S42152S,S42153S,S42154S | |
| | S42155S,S42156S,S42191S,S42192S,S42199S,S42201S,S42202S | |
| | S42209S,S42211S,S42212S,S42213S,S42214S,S42215S,S42216S | |
| | S42221S,S42222S,S42223S,S42224S,S42225S,S42226S,S42231S | |

Continued on next page

**Supplementary Table S1:** (cont'd) A comprehensive mapping of diagnosis groups used in the experiments to their corresponding ICD-10 and ICD-9 codes.

| Diagnosis | ICD-10 | ICD-9 |
|---|---|---|
| | S42232S,S42239S,S42241S,S42242S,S42249S,S42251S,S42252S | |
| | S42253S,S42254S,S42255S,S42256S,S42261S,S42262S,S42263S | |
| | S42264S,S42265S,S42266S,S42271S,S42272S,S42279S,S42291S | |
| | S42292S,S42293S,S42294S,S42295S,S42296S,S42301S,S42302S | |
| | S42309S,S42311S,S42312S,S42319S,S42321S,S42322S,S42323S | |
| | S42324S,S42325S,S42326S,S42331S,S42332S,S42333S,S42334S | |
| | S42335S,S42336S,S42341S,S42342S,S42343S,S42344S,S42345S | |
| | S42346S,S42351S,S42352S,S42353S,S42354S,S42355S,S42356S | |
| | S42361S,S42362S,S42363S,S42364S,S42365S,S42366S,S42391S | |
| | S42392S,S42399S,S42401S,S42402S,S42409S,S42411S,S42412S | |
| | S42413S,S42414S,S42415S,S42416S,S42421S,S42422S,S42423S | |
| | S42424S,S42425S,S42426S,S42431S,S42432S,S42433S,S42434S | |
| | S42435S,S42436S,S42441S,S42442S,S42443S,S42444S,S42445S | |
| | S42446S,S42447S,S42448S,S42449S,S42451S,S42452S,S42453S | |
| | S42454S,S42455S,S42456S,S42461S,S42462S,S42463S,S42464S | |
| | S42465S,S42466S,S42471S,S42472S,S42473S,S42474S,S42475S | |
| | S42476S,S42481S,S42482S,S42489S,S42491S,S42492S,S42493S | |
| | S42494S,S42495S,S42496S,S4290XS,S4291XS,S4292XS,S43001S | |
| | S43002S,S43003S,S43004S,S43005S,S43006S,S43011S,S43012S | |
| | S43013S,S43014S,S43015S,S43016S,S43021S,S43022S,S43023S | |
| | S43024S,S43025S,S43026S,S43031S,S43032S,S43033S,S43034S | |
| | S43035S,S43036S,S43081S,S43082S,S43083S,S43084S,S43085S | |
| | S43086S,S43101S,S43102S,S43109S,S43111S,S43112S,S43119S | |
| | S43121S,S43122S,S43129S,S43131S,S43132S,S43139S,S43141S | |
| | S43142S,S43149S,S43151S,S43152S,S43159S,S43201S,S43202S | |
| | S43203S,S43204S,S43205S,S43206S,S43211S,S43212S,S43213S | |
| | S43214S,S43215S,S43216S,S43221S,S43222S,S43223S,S43224S | |
| | S43225S,S43226S,S43301S,S43302S,S43303S,S43304S,S43305S | |
| | S43306S,S43311S,S43312S,S43313S,S43314S,S43315S,S43316S | |
| | S43391S,S43392S,S43393S,S43394S,S43395S,S43396S,S43401S | |
| | S43402S,S43409S,S43411S,S43412S,S43419S,S43421S,S43422S | |
| | S43429S,S43431S,S43432S,S43439S,S43491S,S43492S,S43499S | |
| | S4350XS,S4351XS,S4352XS,S4360XS,S4361XS,S4362XS,S4380XS | |
| | S4381XS,S4382XS,S4390XS,S4391XS,S4392XS,S4400XS,S4401XS | |
| | S4402XS,S4410XS,S4411XS,S4412XS,S4420XS,S4421XS,S4422XS | |
| | S4430XS,S4431XS,S4432XS,S4440XS,S4441XS,S4442XS,S4450XS | |
| | S4451XS,S4452XS,S448X1S,S448X2S,S448X9S,S4490XS,S4491XS | |
| | S4492XS,S45001S,S45002S,S45009S,S45011S,S45012S,S45019S | |
| | S45091S,S45092S,S45099S,S45101S,S45102S,S45109S,S45111S | |
| | S45112S,S45119S,S45191S,S45192S,S45199S,S45201S,S45202S | |

Continued on next page

**Supplementary Table S1:** (cont'd) A comprehensive mapping of diagnosis groups used in the experiments to their corresponding ICD-10 and ICD-9 codes.

| Diagnosis | ICD-10 | ICD-9 |
|---|---|---|
| | S45209S,S45211S,S45212S,S45219S,S45291S,S45292S,S45299S | |
| | S45301S,S45302S,S45309S,S45311S,S45312S,S45319S,S45391S | |
| | S45392S,S45399S,S45801S,S45802S,S45809S,S45811S,S45812S | |
| | S45819S,S45891S,S45892S,S45899S,S45901S,S45902S,S45909S | |
| | S45911S,S45912S,S45919S,S45991S,S45992S,S45999S,S46001S | |
| | S46002S,S46009S,S46011S,S46012S,S46019S,S46021S,S46022S | |
| | S46029S,S46091S,S46092S,S46099S,S46101S,S46102S,S46109S | |
| | S46111S,S46112S,S46119S,S46121S,S46122S,S46129S,S46191S | |
| | S46192S,S46199S,S46201S,S46202S,S46209S,S46211S,S46212S | |
| | S46219S,S46221S,S46222S,S46229S,S46291S,S46292S,S46299S | |
| | S46301S,S46302S,S46309S,S46311S,S46312S,S46319S,S46321S | |
| | S46322S,S46329S,S46391S,S46392S,S46399S,S46801S,S46802S | |
| | S46809S,S46811S,S46812S,S46819S,S46821S,S46822S,S46829S | |
| | S46891S,S46892S,S46899S,S46901S,S46902S,S46909S,S46911S | |
| | S46912S,S46919S,S46921S,S46922S,S46929S,S46991S,S46992S | |
| | S46999S,S471XXS,S472XXS,S479XXS,S48011S,S48012S,S48019S | |
| | S48021S,S48022S,S48029S,S48111S,S48112S,S48119S,S48121S | |
| | S48122S,S48129S,S48911S,S48912S,S48919S,S48921S,S48922S | |
| | S48929S,S49001S,S49002S,S49009S,S49011S,S49012S,S49019S | |
| | S49021S,S49022S,S49029S,S49031S,S49032S,S49039S,S49041S | |
| | S49042S,S49049S,S49091S,S49092S,S49099S,S49101S,S49102S | |
| | S49109S,S49111S,S49112S,S49119S,S49121S,S49122S,S49129S | |
| | S49131S,S49132S,S49139S,S49141S,S49142S,S49149S,S49191S | |
| | S49192S,S49199S,S4980XS,S4981XS,S4982XS,S4990XS,S4991XS | |
| | S4992XS,S5000XS,S5001XS,S5002XS,S5010XS,S5011XS,S5012XS | |
| | S50311S,S50312S,S50319S,S50321S,S50322S,S50329S,S50341S | |
| | S50342S,S50349S,S50351S,S50352S,S50359S,S50361S,S50362S | |
| | S50369S,S50371S,S50372S,S50379S,S50811S,S50812S,S50819S | |
| | S50821S,S50822S,S50829S,S50841S,S50842S,S50849S,S50851S | |
| | S50852S,S50859S,S50861S,S50862S,S50869S,S50871S,S50872S | |
| | S50879S,S50901S,S50902S,S50909S,S50911S,S50912S,S50919S | |
| | S51001S,S51002S,S51009S,S51011S,S51012S,S51019S,S51021S | |
| | S51022S,S51029S,S51031S,S51032S,S51039S,S51041S,S51042S | |
| | S51049S,S51051S,S51052S,S51059S,S51801S,S51802S,S51809S | |
| | S51811S,S51812S,S51819S,S51821S,S51822S,S51829S,S51831S | |
| | S51832S,S51839S,S51841S,S51842S,S51849S,S51851S,S51852S | |
| | S51859S,S52001S,S52002S,S52009S,S52011S,S52012S,S52019S | |
| | S52021S,S52022S,S52023S,S52024S,S52025S,S52026S,S52031S | |
| | S52032S,S52033S,S52034S,S52035S,S52036S,S52041S,S52042S | |
| | S52043S,S52044S,S52045S,S52046S,S52091S,S52092S,S52099S | |

Continued on next page

**Supplementary Table S1:** (cont'd) A comprehensive mapping of diagnosis groups used in the experiments to their corresponding ICD-10 and ICD-9 codes.

| Diagnosis | ICD-10 | ICD-9 |
|---|---|---|
| | S52101S,S52102S,S52109S,S52111S,S52112S,S52119S,S52121S | |
| | S52122S,S52123S,S52124S,S52125S,S52126S,S52131S,S52132S | |
| | S52133S,S52134S,S52135S,S52136S,S52181S,S52182S,S52189S | |
| | S52201S,S52202S,S52209S,S52211S,S52212S,S52219S,S52221S | |
| | S52222S,S52223S,S52224S,S52225S,S52226S,S52231S,S52232S | |
| | S52233S,S52234S,S52235S,S52236S,S52241S,S52242S,S52243S | |
| | S52244S,S52245S,S52246S,S52251S,S52252S,S52253S,S52254S | |
| | S52255S,S52256S,S52261S,S52262S,S52263S,S52264S,S52265S | |
| | S52266S,S52271S,S52272S,S52279S,S52281S,S52282S,S52283S | |
| | S52291S,S52292S,S52299S,S52301S,S52302S,S52309S,S52311S | |
| | S52312S,S52319S,S52321S,S52322S,S52323S,S52324S,S52325S | |
| | S52326S,S52331S,S52332S,S52333S,S52334S,S52335S,S52336S | |
| | S52341S,S52342S,S52343S,S52344S,S52345S,S52346S,S52351S | |
| | S52352S,S52353S,S52354S,S52355S,S52356S,S52361S,S52362S | |
| | S52363S,S52364S,S52365S,S52366S,S52371S,S52372S,S52379S | |
| | S52381S,S52382S,S52389S,S52391S,S52392S,S52399S,S52501S | |
| | S52502S,S52509S,S52511S,S52512S,S52513S,S52514S,S52515S | |
| | S52516S,S52521S,S52522S,S52529S,S52531S,S52532S,S52539S | |
| | S52541S,S52542S,S52549S,S52551S,S52552S,S52559S,S52561S | |
| | S52562S,S52569S,S52571S,S52572S,S52579S,S52591S,S52592S | |
| | S52599S,S52601S,S52602S,S52609S,S52611S,S52612S,S52613S | |
| | S52614S,S52615S,S52616S,S52621S,S52622S,S52629S,S52691S | |
| | S52692S,S52699S,S5290XS,S5291XS,S5292XS,S53001S,S53002S | |
| | S53003S,S53004S,S53005S,S53006S,S53011S,S53012S,S53013S | |
| | S53014S,S53015S,S53016S,S53021S,S53022S,S53023S,S53024S | |
| | S53025S,S53026S,S53031S,S53032S,S53033S,S53091S,S53092S | |
| | S53093S,S53094S,S53095S,S53096S,S53101S,S53102S,S53103S | |
| | S53104S,S53105S,S53106S,S53111S,S53112S,S53113S,S53114S | |
| | S53115S,S53116S,S53121S,S53122S,S53123S,S53124S,S53125S | |
| | S53126S,S53131S,S53132S,S53133S,S53134S,S53135S,S53136S | |
| | S53141S,S53142S,S53143S,S53144S,S53145S,S53146S,S53191S | |
| | S53192S,S53193S,S53194S,S53195S,S53196S,S5320XS,S5321XS | |
| | S5322XS,S5330XS,S5331XS,S5332XS,S53401S,S53402S,S53409S | |
| | S53411S,S53412S,S53419S,S53421S,S53422S,S53429S,S53431S | |
| | S53432S,S53439S,S53441S,S53442S,S53449S,S53491S,S53492S | |
| | S53499S,S5400XS,S5401XS,S5402XS,S5410XS,S5411XS,S5412XS | |
| | S5420XS,S5421XS,S5422XS,S5430XS,S5431XS,S5432XS,S548X1S | |
| | S548X2S,S548X9S,S5490XS,S5491XS,S5492XS,S55001S,S55002S | |
| | S55009S,S55011S,S55012S,S55019S,S55091S,S55092S,S55099S | |
| | S55101S,S55102S,S55109S,S55111S,S55112S,S55119S,S55191S | |

Continued on next page

**Supplementary Table S1:** (cont'd) A comprehensive mapping of diagnosis groups used in the experiments to their corresponding ICD-10 and ICD-9 codes.

| Diagnosis | ICD-10 | ICD-9 |
|---|---|---|
| | S55192S,S55199S,S55201S,S55202S,S55209S,S55211S,S55212S | |
| | S55219S,S55291S,S55292S,S55299S,S55801S,S55802S,S55809S | |
| | S55811S,S55812S,S55819S,S55891S,S55892S,S55899S,S55901S | |
| | S55902S,S55909S,S55911S,S55912S,S55919S,S55991S,S55992S | |
| | S55999S,S56001S,S56002S,S56009S,S56011S,S56012S,S56019S | |
| | S56021S,S56022S,S56029S,S56091S,S56092S,S56099S,S56101S | |
| | S56102S,S56103S,S56104S,S56105S,S56106S,S56107S,S56108S | |
| | S56109S,S56111S,S56112S,S56113S,S56114S,S56115S,S56116S | |
| | S56117S,S56118S,S56119S,S56121S,S56122S,S56123S,S56124S | |
| | S56125S,S56126S,S56127S,S56128S,S56129S,S56191S,S56192S | |
| | S56193S,S56194S,S56195S,S56196S,S56197S,S56198S,S56199S | |
| | S56201S,S56202S,S56209S,S56211S,S56212S,S56219S,S56221S | |
| | S56222S,S56229S,S56291S,S56292S,S56299S,S56301S,S56302S | |
| | S56309S,S56311S,S56312S,S56319S,S56321S,S56322S,S56329S | |
| | S56391S,S56392S,S56399S,S56401S,S56402S,S56403S,S56404S | |
| | S56405S,S56406S,S56407S,S56408S,S56409S,S56411S,S56412S | |
| | S56413S,S56414S,S56415S,S56416S,S56417S,S56418S,S56419S | |
| | S56421S,S56422S,S56423S,S56424S,S56425S,S56426S,S56427S | |
| | S56428S,S56429S,S56491S,S56492S,S56493S,S56494S,S56495S | |
| | S56496S,S56497S,S56498S,S56499S,S56501S,S56502S,S56509S | |
| | S56511S,S56512S,S56519S,S56521S,S56522S,S56529S,S56591S | |
| | S56592S,S56599S,S56801S,S56802S,S56809S,S56811S,S56812S | |
| | S56819S,S56821S,S56822S,S56829S,S56891S,S56892S,S56899S | |
| | S56901S,S56902S,S56909S,S56911S,S56912S,S56919S,S56921S | |
| | S56922S,S56929S,S56991S,S56992S,S56999S,S5700XS,S5701XS | |
| | S5702XS,S5780XS,S5781XS,S5782XS,S58011S,S58012S,S58019S | |
| | S58021S,S58022S,S58029S,S58111S,S58112S,S58119S,S58121S | |
| | S58122S,S58129S,S58911S,S58912S,S58919S,S58921S,S58922S | |
| | S58929S,S59001S,S59002S,S59009S,S59011S,S59012S,S59019S | |
| | S59021S,S59022S,S59029S,S59031S,S59032S,S59039S,S59041S | |
| | S59042S,S59049S,S59091S,S59092S,S59099S,S59101S,S59102S | |
| | S59109S,S59111S,S59112S,S59119S,S59121S,S59122S,S59129S | |
| | S59131S,S59132S,S59139S,S59141S,S59142S,S59149S,S59191S | |
| | S59192S,S59199S,S59201S,S59202S,S59209S,S59211S,S59212S | |
| | S59219S,S59221S,S59222S,S59229S,S59231S,S59232S,S59239S | |
| | S59241S,S59242S,S59249S,S59291S,S59292S,S59299S,S59801S | |
| | S59802S,S59809S,S59811S,S59812S,S59819S,S59901S,S59902S | |
| | S59909S,S59911S,S59912S,S59919S,S6000XS,S60011S,S60012S | |
| | S60019S,S60021S,S60022S,S60029S,S60031S,S60032S,S60039S | |
| | S60041S,S60042S,S60049S,S60051S,S60052S,S60059S,S6010XS | |

Continued on next page

**Supplementary Table S1:** (cont'd) A comprehensive mapping of diagnosis groups used in the experiments to their corresponding ICD-10 and ICD-9 codes.

| Diagnosis | ICD-10 | ICD-9 |
|---|---|---|
| | S60111S,S60112S,S60119S,S60121S,S60122S,S60129S,S60131S | |
| | S60132S,S60139S,S60141S,S60142S,S60149S,S60151S,S60152S | |
| | S60159S,S60211S,S60212S,S60219S,S60221S,S60222S,S60229S | |
| | S60311S,S60312S,S60319S,S60321S,S60322S,S60329S,S60341S | |
| | S60342S,S60349S,S60351S,S60352S,S60359S,S60361S,S60362S | |
| | S60369S,S60371S,S60372S,S60379S,S60391S,S60392S,S60399S | |
| | S60410S,S60411S,S60412S,S60413S,S60414S,S60415S,S60416S | |
| | S60417S,S60418S,S60419S,S60420S,S60421S,S60422S,S60423S | |
| | S60424S,S60425S,S60426S,S60427S,S60428S,S60429S,S60440S | |
| | S60441S,S60442S,S60443S,S60444S,S60445S,S60446S,S60447S | |
| | S60448S,S60449S,S60450S,S60451S,S60452S,S60453S,S60454S | |
| | S60455S,S60456S,S60457S,S60458S,S60459S,S60460S,S60461S | |
| | S60462S,S60463S,S60464S,S60465S,S60466S,S60467S,S60468S | |
| | S60469S,S60470S,S60471S,S60472S,S60473S,S60474S,S60475S | |
| | S60476S,S60477S,S60478S,S60479S,S60511S,S60512S,S60519S | |
| | S60521S,S60522S,S60529S,S60541S,S60542S,S60549S,S60551S | |
| | S60552S,S60559S,S60561S,S60562S,S60569S,S60571S,S60572S | |
| | S60579S,S60811S,S60812S,S60819S,S60821S,S60822S,S60829S | |
| | S60841S,S60842S,S60849S,S60851S,S60852S,S60859S,S60861S | |
| | S60862S,S60869S,S60871S,S60872S,S60879S,S60911S,S60912S | |
| | S60919S,S60921S,S60922S,S60929S,S60931S,S60932S,S60939S | |
| | S60940S,S60941S,S60942S,S60943S,S60944S,S60945S,S60946S | |
| | S60947S,S60948S,S60949S,S61001S,S61002S,S61009S,S61011S | |
| | S61012S,S61019S,S61021S,S61022S,S61029S,S61031S,S61032S | |
| | S61039S,S61041S,S61042S,S61049S,S61051S,S61052S,S61059S | |
| | S61101S,S61102S,S61109S,S61111S,S61112S,S61119S,S61121S | |
| | S61122S,S61129S,S61131S,S61132S,S61139S,S61141S,S61142S | |
| | S61149S,S61151S,S61152S,S61159S,S61200S,S61201S,S61202S | |
| | S61203S,S61204S,S61205S,S61206S,S61207S,S61208S,S61209S | |
| | S61210S,S61211S,S61212S,S61213S,S61214S,S61215S,S61216S | |
| | S61217S,S61218S,S61219S,S61220S,S61221S,S61222S,S61223S | |
| | S61224S,S61225S,S61226S,S61227S,S61228S,S61229S,S61230S | |
| | S61231S,S61232S,S61233S,S61234S,S61235S,S61236S,S61237S | |
| | S61238S,S61239S,S61240S,S61241S,S61242S,S61243S,S61244S | |
| | S61245S,S61246S,S61247S,S61248S,S61249S,S61250S,S61251S | |
| | S61252S,S61253S,S61254S,S61255S,S61256S,S61257S,S61258S | |
| | S61259S,S61300S,S61301S,S61302S,S61303S,S61304S,S61305S | |
| | S61306S,S61307S,S61308S,S61309S,S61310S,S61311S,S61312S | |
| | S61313S,S61314S,S61315S,S61316S,S61317S,S61318S,S61319S | |
| | S61320S,S61321S,S61322S,S61323S,S61324S,S61325S,S61326S | |

Continued on next page

**Supplementary Table S1:** (cont'd) A comprehensive mapping of diagnosis groups used in the experiments to their corresponding ICD-10 and ICD-9 codes.

| Diagnosis | ICD-10 | ICD-9 |
|---|---|---|
| | S61327S,S61328S,S61329S,S61330S,S61331S,S61332S,S61333S | |
| | S61334S,S61335S,S61336S,S61337S,S61338S,S61339S,S61340S | |
| | S61341S,S61342S,S61343S,S61344S,S61345S,S61346S,S61347S | |
| | S61348S,S61349S,S61350S,S61351S,S61352S,S61353S,S61354S | |
| | S61355S,S61356S,S61357S,S61358S,S61359S,S61401S,S61402S | |
| | S61409S,S61411S,S61412S,S61419S,S61421S,S61422S,S61429S | |
| | S61431S,S61432S,S61439S,S61441S,S61442S,S61449S,S61451S | |
| | S61452S,S61459S,S61501S,S61502S,S61509S,S61511S,S61512S | |
| | S61519S,S61521S,S61522S,S61529S,S61531S,S61532S,S61539S | |
| | S61541S,S61542S,S61549S,S61551S,S61552S,S61559S,S62001S | |
| | S62002S,S62009S,S62011S,S62012S,S62013S,S62014S,S62015S | |
| | S62016S,S62021S,S62022S,S62023S,S62024S,S62025S,S62026S | |
| | S62031S,S62032S,S62033S,S62034S,S62035S,S62036S,S62101S | |
| | S62102S,S62109S,S62111S,S62112S,S62113S,S62114S,S62115S | |
| | S62116S,S62121S,S62122S,S62123S,S62124S,S62125S,S62126S | |
| | S62131S,S62132S,S62133S,S62134S,S62135S,S62136S,S62141S | |
| | S62142S,S62143S,S62144S,S62145S,S62146S,S62151S,S62152S | |
| | S62153S,S62154S,S62155S,S62156S,S62161S,S62162S,S62163S | |
| | S62164S,S62165S,S62166S,S62171S,S62172S,S62173S,S62174S | |
| | S62175S,S62176S,S62181S,S62182S,S62183S,S62184S,S62185S | |
| | S62186S,S62201S,S62202S,S62209S,S62211S,S62212S,S62213S | |
| | S62221S,S62222S,S62223S,S62224S,S62225S,S62226S,S62231S | |
| | S62232S,S62233S,S62234S,S62235S,S62236S,S62241S,S62242S | |
| | S62243S,S62244S,S62245S,S62246S,S62251S,S62252S,S62253S | |
| | S62254S,S62255S,S62256S,S62291S,S62292S,S62299S,S62300S | |
| | S62301S,S62302S,S62303S,S62304S,S62305S,S62306S,S62307S | |
| | S62308S,S62309S,S62310S,S62311S,S62312S,S62313S,S62314S | |
| | S62315S,S62316S,S62317S,S62318S,S62319S,S62320S,S62321S | |
| | S62322S,S62323S,S62324S,S62325S,S62326S,S62327S,S62328S | |
| | S62329S,S62330S,S62331S,S62332S,S62333S,S62334S,S62335S | |
| | S62336S,S62337S,S62338S,S62339S,S62340S,S62341S,S62342S | |
| | S62343S,S62344S,S62345S,S62346S,S62347S,S62348S,S62349S | |
| | S62350S,S62351S,S62352S,S62353S,S62354S,S62355S,S62356S | |
| | S62357S,S62358S,S62359S,S62360S,S62361S,S62362S,S62363S | |
| | S62364S,S62365S,S62366S,S62367S,S62368S,S62369S,S62390S | |
| | S62391S,S62392S,S62393S,S62394S,S62395S,S62396S,S62397S | |
| | S62398S,S62399S,S62501S,S62502S,S62509S,S62511S,S62512S | |
| | S62513S,S62514S,S62515S,S62516S,S62521S,S62522S,S62523S | |
| | S62524S,S62525S,S62526S,S62600S,S62601S,S62602S,S62603S | |
| | S62604S,S62605S,S62606S,S62607S,S62608S,S62609S,S62610S | |

Continued on next page

**Supplementary Table S1:** (cont'd) A comprehensive mapping of diagnosis groups used in the experiments to their corresponding ICD-10 and ICD-9 codes.

| Diagnosis | ICD-10 | ICD-9 |
|---|---|---|
| | S62611S,S62612S,S62613S,S62614S,S62615S,S62616S,S62617S | |
| | S62618S,S62619S,S62620S,S62621S,S62622S,S62623S,S62624S | |
| | S62625S,S62626S,S62627S,S62628S,S62629S,S62630S,S62631S | |
| | S62632S,S62633S,S62634S,S62635S,S62636S,S62637S,S62638S | |
| | S62639S,S62640S,S62641S,S62642S,S62643S,S62644S,S62645S | |
| | S62646S,S62647S,S62648S,S62649S,S62650S,S62651S,S62652S | |
| | S62653S,S62654S,S62655S,S62656S,S62657S,S62658S,S62659S | |
| | S62660S,S62661S,S62662S,S62663S,S62664S,S62665S,S62666S | |
| | S62667S,S62668S,S62669S,S6290XS,S6291XS,S6292XS,S63001S | |
| | S63002S,S63003S,S63004S,S63005S,S63006S,S63011S,S63012S | |
| | S63013S,S63014S,S63015S,S63016S,S63021S,S63022S,S63023S | |
| | S63024S,S63025S,S63026S,S63031S,S63032S,S63033S,S63034S | |
| | S63035S,S63036S,S63041S,S63042S,S63043S,S63044S,S63045S | |
| | S63046S,S63051S,S63052S,S63053S,S63054S,S63055S,S63056S | |
| | S63061S,S63062S,S63063S,S63064S,S63065S,S63066S,S63071S | |
| | S63072S,S63073S,S63074S,S63075S,S63076S,S63091S,S63092S | |
| | S63093S,S63094S,S63095S,S63096S,S63101S,S63102S,S63103S | |
| | S63104S,S63105S,S63106S,S63111S,S63112S,S63113S,S63114S | |
| | S63115S,S63116S,S63121S,S63122S,S63123S,S63124S,S63125S | |
| | S63126S,S63131S,S63132S,S63133S,S63134S,S63135S,S63136S | |
| | S63141S,S63142S,S63143S,S63144S,S63145S,S63146S,S63200S | |
| | S63201S,S63202S,S63203S,S63204S,S63205S,S63206S,S63207S | |
| | S63208S,S63209S,S63210S,S63211S,S63212S,S63213S,S63214S | |
| | S63215S,S63216S,S63217S,S63218S,S63219S,S63220S,S63221S | |
| | S63222S,S63223S,S63224S,S63225S,S63226S,S63227S,S63228S | |
| | S63229S,S63230S,S63231S,S63232S,S63233S,S63234S,S63235S | |
| | S63236S,S63237S,S63238S,S63239S,S63240S,S63241S,S63242S | |
| | S63243S,S63244S,S63245S,S63246S,S63247S,S63248S,S63249S | |
| | S63250S,S63251S,S63252S,S63253S,S63254S,S63255S,S63256S | |
| | S63257S,S63258S,S63259S,S63260S,S63261S,S63262S,S63263S | |
| | S63264S,S63265S,S63266S,S63267S,S63268S,S63269S,S63270S | |
| | S63271S,S63272S,S63273S,S63274S,S63275S,S63276S,S63277S | |
| | S63278S,S63279S,S63280S,S63281S,S63282S,S63283S,S63284S | |
| | S63285S,S63286S,S63287S,S63288S,S63289S,S63290S,S63291S | |
| | S63292S,S63293S,S63294S,S63295S,S63296S,S63297S,S63298S | |
| | S63299S,S63301S,S63302S,S63309S,S63311S,S63312S,S63319S | |
| | S63321S,S63322S,S63329S,S63331S,S63332S,S63339S,S63391S | |
| | S63392S,S63399S,S63400S,S63401S,S63402S,S63403S,S63404S | |
| | S63405S,S63406S,S63407S,S63408S,S63409S,S63410S,S63411S | |
| | S63412S,S63413S,S63414S,S63415S,S63416S,S63417S,S63418S | |

Continued on next page

**Supplementary Table S1:** (cont'd) A comprehensive mapping of diagnosis groups used in the experiments to their corresponding ICD-10 and ICD-9 codes.

| Diagnosis | ICD-10 | ICD-9 |
|---|---|---|
| | S63419S,S63420S,S63421S,S63422S,S63423S,S63424S,S63425S | |
| | S63426S,S63427S,S63428S,S63429S,S63430S,S63431S,S63432S | |
| | S63433S,S63434S,S63435S,S63436S,S63437S,S63438S,S63439S | |
| | S63490S,S63491S,S63492S,S63493S,S63494S,S63495S,S63496S | |
| | S63497S,S63498S,S63499S,S63501S,S63502S,S63509S,S63511S | |
| | S63512S,S63519S,S63521S,S63522S,S63529S,S63591S,S63592S | |
| | S63599S,S63601S,S63602S,S63609S,S63610S,S63611S,S63612S | |
| | S63613S,S63614S,S63615S,S63616S,S63617S,S63618S,S63619S | |
| | S63621S,S63622S,S63629S,S63630S,S63631S,S63632S,S63633S | |
| | S63634S,S63635S,S63636S,S63637S,S63638S,S63639S,S63641S | |
| | S63642S,S63649S,S63650S,S63651S,S63652S,S63653S,S63654S | |
| | S63655S,S63656S,S63657S,S63658S,S63659S,S63681S,S63682S | |
| | S63689S,S63690S,S63691S,S63692S,S63693S,S63694S,S63695S | |
| | S63696S,S63697S,S63698S,S63699S,S638X1S,S638X2S,S638X9S | |
| | S6390XS,S6391XS,S6392XS,S6400XS,S6401XS,S6402XS,S6410XS | |
| | S6411XS,S6412XS,S6420XS,S6421XS,S6422XS,S6430XS,S6431XS | |
| | S6432XS,S6440XS,S64490S,S64491S,S64492S,S64493S,S64494S | |
| | S64495S,S64496S,S64497S,S64498S,S648X1S,S648X2S,S648X9S | |
| | S6490XS,S6491XS,S6492XS,S65001S,S65002S,S65009S,S65011S | |
| | S65012S,S65019S,S65091S,S65092S,S65099S,S65101S,S65102S | |
| | S65109S,S65111S,S65112S,S65119S,S65191S,S65192S,S65199S | |
| | S65201S,S65202S,S65209S,S65211S,S65212S,S65219S,S65291S | |
| | S65292S,S65299S,S65301S,S65302S,S65309S,S65311S,S65312S | |
| | S65319S,S65391S,S65392S,S65399S,S65401S,S65402S,S65409S | |
| | S65411S,S65412S,S65419S,S65491S,S65492S,S65499S,S65500S | |
| | S65501S,S65502S,S65503S,S65504S,S65505S,S65506S,S65507S | |
| | S65508S,S65509S,S65510S,S65511S,S65512S,S65513S,S65514S | |
| | S65515S,S65516S,S65517S,S65518S,S65519S,S65590S,S65591S | |
| | S65592S,S65593S,S65594S,S65595S,S65596S,S65597S,S65598S | |
| | S65599S,S65801S,S65802S,S65809S,S65811S,S65812S,S65819S | |
| | S65891S,S65892S,S65899S,S65901S,S65902S,S65909S,S65911S | |
| | S65912S,S65919S,S65991S,S65992S,S65999S,S66001S,S66002S | |
| | S66009S,S66011S,S66012S,S66019S,S66021S,S66022S,S66029S | |
| | S66091S,S66092S,S66099S,S66100S,S66101S,S66102S,S66103S | |
| | S66104S,S66105S,S66106S,S66107S,S66108S,S66109S,S66110S | |
| | S66111S,S66112S,S66113S,S66114S,S66115S,S66116S,S66117S | |
| | S66118S,S66119S,S66120S,S66121S,S66122S,S66123S,S66124S | |
| | S66125S,S66126S,S66127S,S66128S,S66129S,S66190S,S66191S | |
| | S66192S,S66193S,S66194S,S66195S,S66196S,S66197S,S66198S | |
| | S66199S,S66201S,S66202S,S66209S,S66211S,S66212S,S66219S | |

Continued on next page

**Supplementary Table S1:** (cont'd) A comprehensive mapping of diagnosis groups used in the experiments to their corresponding ICD-10 and ICD-9 codes.

| Diagnosis | ICD-10 | ICD-9 |
|---|---|---|
| | S66221S,S66222S,S66229S,S66291S,S66292S,S66299S,S66300S | |
| | S66301S,S66302S,S66303S,S66304S,S66305S,S66306S,S66307S | |
| | S66308S,S66309S,S66310S,S66311S,S66312S,S66313S,S66314S | |
| | S66315S,S66316S,S66317S,S66318S,S66319S,S66320S,S66321S | |
| | S66322S,S66323S,S66324S,S66325S,S66326S,S66327S,S66328S | |
| | S66329S,S66390S,S66391S,S66392S,S66393S,S66394S,S66395S | |
| | S66396S,S66397S,S66398S,S66399S,S66401S,S66402S,S66409S | |
| | S66411S,S66412S,S66419S,S66421S,S66422S,S66429S,S66491S | |
| | S66492S,S66499S,S66500S,S66501S,S66502S,S66503S,S66504S | |
| | S66505S,S66506S,S66507S,S66508S,S66509S,S66510S,S66511S | |
| | S66512S,S66513S,S66514S,S66515S,S66516S,S66517S,S66518S | |
| | S66519S,S66520S,S66521S,S66522S,S66523S,S66524S,S66525S | |
| | S66526S,S66527S,S66528S,S66529S,S66590S,S66591S,S66592S | |
| | S66593S,S66594S,S66595S,S66596S,S66597S,S66598S,S66599S | |
| | S66801S,S66802S,S66809S,S66811S,S66812S,S66819S,S66821S | |
| | S66822S,S66829S,S66891S,S66892S,S66899S,S66901S,S66902S | |
| | S66909S,S66911S,S66912S,S66919S,S66921S,S66922S,S66929S | |
| | S66991S,S66992S,S66999S,S6700XS,S6701XS,S6702XS,S6710XS | |
| | S67190S,S67191S,S67192S,S67193S,S67194S,S67195S,S67196S | |
| | S67197S,S67198S,S6720XS,S6721XS,S6722XS,S6730XS,S6731XS | |
| | S6732XS,S6740XS,S6741XS,S6742XS,S6790XS,S6791XS,S6792XS | |
| | S68011S,S68012S,S68019S,S68021S,S68022S,S68029S,S68110S | |
| | S68111S,S68112S,S68113S,S68114S,S68115S,S68116S,S68117S | |
| | S68118S,S68119S,S68120S,S68121S,S68122S,S68123S,S68124S | |
| | S68125S,S68126S,S68127S,S68128S,S68129S,S68411S,S68412S | |
| | S68419S,S68421S,S68422S,S68429S,S68511S,S68512S,S68519S | |
| | S68521S,S68522S,S68529S,S68610S,S68611S,S68612S,S68613S | |
| | S68614S,S68615S,S68616S,S68617S,S68618S,S68619S,S68620S | |
| | S68621S,S68622S,S68623S,S68624S,S68625S,S68626S,S68627S | |
| | S68628S,S68629S,S68711S,S68712S,S68719S,S68721S,S68722S | |
| | S68729S,S6980XS,S6981XS,S6982XS,S6990XS,S6991XS,S6992XS | |
| | S7000XS,S7001XS,S7002XS,S7010XS,S7011XS,S7012XS,S70211S | |
| | S70212S,S70219S,S70221S,S70222S,S70229S,S70241S,S70242S | |
| | S70249S,S70251S,S70252S,S70259S,S70261S,S70262S,S70269S | |
| | S70271S,S70272S,S70279S,S70311S,S70312S,S70319S,S70321S | |
| | S70322S,S70329S,S70341S,S70342S,S70349S,S70351S,S70352S | |
| | S70359S,S70361S,S70362S,S70369S,S70371S,S70372S,S70379S | |
| | S70911S,S70912S,S70919S,S70921S,S70922S,S70929S,S71001S | |
| | S71002S,S71009S,S71011S,S71012S,S71019S,S71021S,S71022S | |
| | S71029S,S71031S,S71032S,S71039S,S71041S,S71042S,S71049S | |

Continued on next page

**Supplementary Table S1:** (cont'd) A comprehensive mapping of diagnosis groups used in the experiments to their corresponding ICD-10 and ICD-9 codes.

| Diagnosis | ICD-10 | ICD-9 |
|---|---|---|
| | S71051S,S71052S,S71059S,S71101S,S71102S,S71109S,S71111S | |
| | S71112S,S71119S,S71121S,S71122S,S71129S,S71131S,S71132S | |
| | S71139S,S71141S,S71142S,S71149S,S71151S,S71152S,S71159S | |
| | S72001S,S72002S,S72009S,S72011S,S72012S,S72019S,S72021S | |
| | S72022S,S72023S,S72024S,S72025S,S72026S,S72031S,S72032S | |
| | S72033S,S72034S,S72035S,S72036S,S72041S,S72042S,S72043S | |
| | S72044S,S72045S,S72046S,S72051S,S72052S,S72059S,S72061S | |
| | S72062S,S72063S,S72064S,S72065S,S72066S,S72091S,S72092S | |
| | S72099S,S72101S,S72102S,S72109S,S72111S,S72112S,S72113S | |
| | S72114S,S72115S,S72116S,S72121S,S72122S,S72123S,S72124S | |
| | S72125S,S72126S,S72131S,S72132S,S72133S,S72134S,S72135S | |
| | S72136S,S72141S,S72142S,S72143S,S72144S,S72145S,S72146S | |
| | S7221XS,S7222XS,S7223XS,S7224XS,S7225XS,S7226XS,S72301S | |
| | S72302S,S72309S,S72321S,S72322S,S72323S,S72324S,S72325S | |
| | S72326S,S72331S,S72332S,S72333S,S72334S,S72335S,S72336S | |
| | S72341S,S72342S,S72343S,S72344S,S72345S,S72346S,S72351S | |
| | S72352S,S72353S,S72354S,S72355S,S72356S,S72361S,S72362S | |
| | S72363S,S72364S,S72365S,S72366S,S72391S,S72392S,S72399S | |
| | S72401S,S72402S,S72409S,S72411S,S72412S,S72413S,S72414S | |
| | S72415S,S72416S,S72421S,S72422S,S72423S,S72424S,S72425S | |
| | S72426S,S72431S,S72432S,S72433S,S72434S,S72435S,S72436S | |
| | S72441S,S72442S,S72443S,S72444S,S72445S,S72446S,S72451S | |
| | S72452S,S72453S,S72454S,S72455S,S72456S,S72461S,S72462S | |
| | S72463S,S72464S,S72465S,S72466S,S72471S,S72472S,S72479S | |
| | S72491S,S72492S,S72499S,S728X1S,S728X2S,S728X9S,S7290XS | |
| | S7291XS,S7292XS,S73001S,S73002S,S73003S,S73004S,S73005S | |
| | S73006S,S73011S,S73012S,S73013S,S73014S,S73015S,S73016S | |
| | S73021S,S73022S,S73023S,S73024S,S73025S,S73026S,S73031S | |
| | S73032S,S73033S,S73034S,S73035S,S73036S,S73041S,S73042S | |
| | S73043S,S73044S,S73045S,S73046S,S73101S,S73102S,S73109S | |
| | S73111S,S73112S,S73119S,S73121S,S73122S,S73129S,S73191S | |
| | S73192S,S73199S,S7400XS,S7401XS,S7402XS,S7410XS,S7411XS | |
| | S7412XS,S7420XS,S7421XS,S7422XS,S748X1S,S748X2S,S748X9S | |
| | S7490XS,S7491XS,S7492XS,S75001S,S75002S,S75009S,S75011S | |
| | S75012S,S75019S,S75021S,S75022S,S75029S,S75091S,S75092S | |
| | S75099S,S75101S,S75102S,S75109S,S75111S,S75112S,S75119S | |
| | S75121S,S75122S,S75129S,S75191S,S75192S,S75199S,S75201S | |
| | S75202S,S75209S,S75211S,S75212S,S75219S,S75221S,S75222S | |
| | S75229S,S75291S,S75292S,S75299S,S75801S,S75802S,S75809S | |
| | S75811S,S75812S,S75819S,S75891S,S75892S,S75899S,S75901S | |

Continued on next page

**Supplementary Table S1:** (cont'd) A comprehensive mapping of diagnosis groups used in the experiments to their corresponding ICD-10 and ICD-9 codes.

| Diagnosis | ICD-10 | ICD-9 |
|---|---|---|
| | S75902S,S75909S,S75911S,S75912S,S75919S,S75991S,S75992S | |
| | S75999S,S76001S,S76002S,S76009S,S76011S,S76012S,S76019S | |
| | S76021S,S76022S,S76029S,S76091S,S76092S,S76099S,S76101S | |
| | S76102S,S76109S,S76111S,S76112S,S76119S,S76121S,S76122S | |
| | S76129S,S76191S,S76192S,S76199S,S76201S,S76202S,S76209S | |
| | S76211S,S76212S,S76219S,S76221S,S76222S,S76229S,S76291S | |
| | S76292S,S76299S,S76301S,S76302S,S76309S,S76311S,S76312S | |
| | S76319S,S76321S,S76322S,S76329S,S76391S,S76392S,S76399S | |
| | S76801S,S76802S,S76809S,S76811S,S76812S,S76819S,S76821S | |
| | S76822S,S76829S,S76891S,S76892S,S76899S,S76901S,S76902S | |
| | S76909S,S76911S,S76912S,S76919S,S76921S,S76922S,S76929S | |
| | S76991S,S76992S,S76999S,S7700XS,S7701XS,S7702XS,S7710XS | |
| | S7711XS,S7712XS,S7720XS,S7721XS,S7722XS,S78011S,S78012S | |
| | S78019S,S78021S,S78022S,S78029S,S78111S,S78112S,S78119S | |
| | S78121S,S78122S,S78129S,S78911S,S78912S,S78919S,S78921S | |
| | S78922S,S78929S,S79001S,S79002S,S79009S,S79011S,S79012S | |
| | S79019S,S79091S,S79092S,S79099S,S79101S,S79102S,S79109S | |
| | S79111S,S79112S,S79119S,S79121S,S79122S,S79129S,S79131S | |
| | S79132S,S79139S,S79141S,S79142S,S79149S,S79191S,S79192S | |
| | S79199S,S79811S,S79812S,S79819S,S79821S,S79822S,S79829S | |
| | S79911S,S79912S,S79919S,S79921S,S79922S,S79929S,S8000XS | |
| | S8001XS,S8002XS,S8010XS,S8011XS,S8012XS,S80211S,S80212S | |
| | S80219S,S80221S,S80222S,S80229S,S80241S,S80242S,S80249S | |
| | S80251S,S80252S,S80259S,S80261S,S80262S,S80269S,S80271S | |
| | S80272S,S80279S,S80811S,S80812S,S80819S,S80821S,S80822S | |
| | S80829S,S80841S,S80842S,S80849S,S80851S,S80852S,S80859S | |
| | S80861S,S80862S,S80869S,S80871S,S80872S,S80879S,S80911S | |
| | S80912S,S80919S,S80921S,S80922S,S80929S,S81001S,S81002S | |
| | S81009S,S81011S,S81012S,S81019S,S81021S,S81022S,S81029S | |
| | S81031S,S81032S,S81039S,S81041S,S81042S,S81049S,S81051S | |
| | S81052S,S81059S,S81801S,S81802S,S81809S,S81811S,S81812S | |
| | S81819S,S81821S,S81822S,S81829S,S81831S,S81832S,S81839S | |
| | S81841S,S81842S,S81849S,S81851S,S81852S,S81859S,S82001S | |
| | S82002S,S82009S,S82011S,S82012S,S82013S,S82014S,S82015S | |
| | S82016S,S82021S,S82022S,S82023S,S82024S,S82025S,S82026S | |
| | S82031S,S82032S,S82033S,S82034S,S82035S,S82036S,S82041S | |
| | S82042S,S82043S,S82044S,S82045S,S82046S,S82091S,S82092S | |
| | S82099S,S82101S,S82102S,S82109S,S82111S,S82112S,S82113S | |
| | S82114S,S82115S,S82116S,S82121S,S82122S,S82123S,S82124S | |
| | S82125S,S82126S,S82131S,S82132S,S82133S,S82134S,S82135S | |

Continued on next page

**Supplementary Table S1:** (cont'd) A comprehensive mapping of diagnosis groups used in the experiments to their corresponding ICD-10 and ICD-9 codes.

| Diagnosis | ICD-10 | ICD-9 |
|---|---|---|
| | S82136S,S82141S,S82142S,S82143S,S82144S,S82145S,S82146S | |
| | S82151S,S82152S,S82153S,S82154S,S82155S,S82156S,S82161S | |
| | S82162S,S82169S,S82191S,S82192S,S82199S,S82201S,S82202S | |
| | S82209S,S82221S,S82222S,S82223S,S82224S,S82225S,S82226S | |
| | S82231S,S82232S,S82233S,S82234S,S82235S,S82236S,S82241S | |
| | S82242S,S82243S,S82244S,S82245S,S82246S,S82251S,S82252S | |
| | S82253S,S82254S,S82255S,S82256S,S82261S,S82262S,S82263S | |
| | S82264S,S82265S,S82266S,S82291S,S82292S,S82299S,S82301S | |
| | S82302S,S82309S,S82311S,S82312S,S82319S,S82391S,S82392S | |
| | S82399S,S82401S,S82402S,S82409S,S82421S,S82422S,S82423S | |
| | S82424S,S82425S,S82426S,S82431S,S82432S,S82433S,S82434S | |
| | S82435S,S82436S,S82441S,S82442S,S82443S,S82444S,S82445S | |
| | S82446S,S82451S,S82452S,S82453S,S82454S,S82455S,S82456S | |
| | S82461S,S82462S,S82463S,S82464S,S82465S,S82466S,S82491S | |
| | S82492S,S82499S,S8251XS,S8252XS,S8253XS,S8254XS,S8255XS | |
| | S8256XS,S8261XS,S8262XS,S8263XS,S8264XS,S8265XS,S8266XS | |
| | S82811S,S82812S,S82819S,S82821S,S82822S,S82829S,S82831S | |
| | S82832S,S82839S,S82841S,S82842S,S82843S,S82844S,S82845S | |
| | S82846S,S82851S,S82852S,S82853S,S82854S,S82855S,S82856S | |
| | S82861S,S82862S,S82863S,S82864S,S82865S,S82866S,S82871S | |
| | S82872S,S82873S,S82874S,S82875S,S82876S,S82891S,S82892S | |
| | S82899S,S8290XS,S8291XS,S8292XS,S83001S,S83002S,S83003S | |
| | S83004S,S83005S,S83006S,S83011S,S83012S,S83013S,S83014S | |
| | S83015S,S83016S,S83091S,S83092S,S83093S,S83094S,S83095S | |
| | S83096S,S83101S,S83102S,S83103S,S83104S,S83105S,S83106S | |
| | S83111S,S83112S,S83113S,S83114S,S83115S,S83116S,S83121S | |
| | S83122S,S83123S,S83124S,S83125S,S83126S,S83131S,S83132S | |
| | S83133S,S83134S,S83135S,S83136S,S83141S,S83142S,S83143S | |
| | S83144S,S83145S,S83146S,S83191S,S83192S,S83193S,S83194S | |
| | S83195S,S83196S,S83200S,S83201S,S83202S,S83203S,S83204S | |
| | S83205S,S83206S,S83207S,S83209S,S83211S,S83212S,S83219S | |
| | S83221S,S83222S,S83229S,S83231S,S83232S,S83239S,S83241S | |
| | S83242S,S83249S,S83251S,S83252S,S83259S,S83261S,S83262S | |
| | S83269S,S83271S,S83272S,S83279S,S83281S,S83282S,S83289S | |
| | S8330XS,S8331XS,S8332XS,S83401S,S83402S,S83409S,S83411S | |
| | S83412S,S83419S,S83421S,S83422S,S83429S,S83501S,S83502S | |
| | S83509S,S83511S,S83512S,S83519S,S83521S,S83522S,S83529S | |
| | S8360XS,S8361XS,S8362XS,S838X1S,S838X2S,S838X9S,S8390XS | |
| | S8391XS,S8392XS,S8400XS,S8401XS,S8402XS,S8410XS,S8411XS | |
| | S8412XS,S8420XS,S8421XS,S8422XS,S84801S,S84802S,S84809S | |

Continued on next page

**Supplementary Table S1:** (cont'd) A comprehensive mapping of diagnosis groups used in the experiments to their corresponding ICD-10 and ICD-9 codes.

| Diagnosis | ICD-10 | ICD-9 |
|---|---|---|
| | S8490XS,S8491XS,S8492XS,S85001S,S85002S,S85009S,S85011S | |
| | S85012S,S85019S,S85091S,S85092S,S85099S,S85101S,S85102S | |
| | S85109S,S85111S,S85112S,S85119S,S85121S,S85122S,S85129S | |
| | S85131S,S85132S,S85139S,S85141S,S85142S,S85149S,S85151S | |
| | S85152S,S85159S,S85161S,S85162S,S85169S,S85171S,S85172S | |
| | S85179S,S85181S,S85182S,S85189S,S85201S,S85202S,S85209S | |
| | S85211S,S85212S,S85219S,S85291S,S85292S,S85299S,S85301S | |
| | S85302S,S85309S,S85311S,S85312S,S85319S,S85391S,S85392S | |
| | S85399S,S85401S,S85402S,S85409S,S85411S,S85412S,S85419S | |
| | S85491S,S85492S,S85499S,S85501S,S85502S,S85509S,S85511S | |
| | S85512S,S85519S,S85591S,S85592S,S85599S,S85801S,S85802S | |
| | S85809S,S85811S,S85812S,S85819S,S85891S,S85892S,S85899S | |
| | S85901S,S85902S,S85909S,S85911S,S85912S,S85919S,S85991S | |
| | S85992S,S85999S,S86001S,S86002S,S86009S,S86011S,S86012S | |
| | S86019S,S86021S,S86022S,S86029S,S86091S,S86092S,S86099S | |
| | S86101S,S86102S,S86109S,S86111S,S86112S,S86119S,S86121S | |
| | S86122S,S86129S,S86191S,S86192S,S86199S,S86201S,S86202S | |
| | S86209S,S86211S,S86212S,S86219S,S86221S,S86222S,S86229S | |
| | S86291S,S86292S,S86299S,S86301S,S86302S,S86309S,S86311S | |
| | S86312S,S86319S,S86321S,S86322S,S86329S,S86391S,S86392S | |
| | S86399S,S86801S,S86802S,S86809S,S86811S,S86812S,S86819S | |
| | S86821S,S86822S,S86829S,S86891S,S86892S,S86899S,S86901S | |
| | S86902S,S86909S,S86911S,S86912S,S86919S,S86921S,S86922S | |
| | S86929S,S86991S,S86992S,S86999S,S8700XS,S8701XS,S8702XS | |
| | S8780XS,S8781XS,S8782XS,S88011S,S88012S,S88019S,S88021S | |
| | S88022S,S88029S,S88111S,S88112S,S88119S,S88121S,S88122S | |
| | S88129S,S88911S,S88912S,S88919S,S88921S,S88922S,S88929S | |
| | S89001S,S89002S,S89009S,S89011S,S89012S,S89019S,S89021S | |
| | S89022S,S89029S,S89031S,S89032S,S89039S,S89041S,S89042S | |
| | S89049S,S89091S,S89092S,S89099S,S89101S,S89102S,S89109S | |
| | S89111S,S89112S,S89119S,S89121S,S89122S,S89129S,S89131S | |
| | S89132S,S89139S,S89141S,S89142S,S89149S,S89191S,S89192S | |
| | S89199S,S89201S,S89202S,S89209S,S89211S,S89212S,S89219S | |
| | S89221S,S89222S,S89229S,S89291S,S89292S,S89299S,S89301S | |
| | S89302S,S89309S,S89311S,S89312S,S89319S,S89321S,S89322S | |
| | S89329S,S89391S,S89392S,S89399S,S8980XS,S8981XS,S8982XS | |
| | S8990XS,S8991XS,S8992XS,S9000XS,S9001XS,S9002XS,S90111S | |
| | S90112S,S90119S,S90121S,S90122S,S90129S,S90211S,S90212S | |
| | S90219S,S90221S,S90222S,S90229S,S9030XS,S9031XS,S9032XS | |
| | S90411S,S90412S,S90413S,S90414S,S90415S,S90416S,S90421S | |

Continued on next page

**Supplementary Table S1:** (cont'd) A comprehensive mapping of diagnosis groups used in the experiments to their corresponding ICD-10 and ICD-9 codes.

| Diagnosis | ICD-10 | ICD-9 |
|---|---|---|
| | S90422S,S90423S,S90424S,S90425S,S90426S,S90441S,S90442S | |
| | S90443S,S90444S,S90445S,S90446S,S90451S,S90452S,S90453S | |
| | S90454S,S90455S,S90456S,S90461S,S90462S,S90463S,S90464S | |
| | S90465S,S90466S,S90471S,S90472S,S90473S,S90474S,S90475S | |
| | S90476S,S90511S,S90512S,S90519S,S90521S,S90522S,S90529S | |
| | S90541S,S90542S,S90549S,S90551S,S90552S,S90559S,S90561S | |
| | S90562S,S90569S,S90571S,S90572S,S90579S,S90811S,S90812S | |
| | S90819S,S90821S,S90822S,S90829S,S90841S,S90842S,S90849S | |
| | S90851S,S90852S,S90859S,S90861S,S90862S,S90869S,S90871S | |
| | S90872S,S90879S,S90911S,S90912S,S90919S,S90921S,S90922S | |
| | S90929S,S90931S,S90932S,S90933S,S90934S,S90935S,S90936S | |
| | S91001S,S91002S,S91009S,S91011S,S91012S,S91019S,S91021S | |
| | S91022S,S91029S,S91031S,S91032S,S91039S,S91041S,S91042S | |
| | S91049S,S91051S,S91052S,S91059S,S91101S,S91102S,S91103S | |
| | S91104S,S91105S,S91106S,S91109S,S91111S,S91112S,S91113S | |
| | S91114S,S91115S,S91116S,S91119S,S91121S,S91122S,S91123S | |
| | S91124S,S91125S,S91126S,S91129S,S91131S,S91132S,S91133S | |
| | S91134S,S91135S,S91136S,S91139S,S91141S,S91142S,S91143S | |
| | S91144S,S91145S,S91146S,S91149S,S91151S,S91152S,S91153S | |
| | S91154S,S91155S,S91156S,S91159S,S91201S,S91202S,S91203S | |
| | S91204S,S91205S,S91206S,S91209S,S91211S,S91212S,S91213S | |
| | S91214S,S91215S,S91216S,S91219S,S91221S,S91222S,S91223S | |
| | S91224S,S91225S,S91226S,S91229S,S91231S,S91232S,S91233S | |
| | S91234S,S91235S,S91236S,S91239S,S91241S,S91242S,S91243S | |
| | S91244S,S91245S,S91246S,S91249S,S91251S,S91252S,S91253S | |
| | S91254S,S91255S,S91256S,S91259S,S91301S,S91302S,S91309S | |
| | S91311S,S91312S,S91319S,S91321S,S91322S,S91329S,S91331S | |
| | S91332S,S91339S,S91341S,S91342S,S91349S,S91351S,S91352S | |
| | S91359S,S92001S,S92002S,S92009S,S92011S,S92012S,S92013S | |
| | S92014S,S92015S,S92016S,S92021S,S92022S,S92023S,S92024S | |
| | S92025S,S92026S,S92031S,S92032S,S92033S,S92034S,S92035S | |
| | S92036S,S92041S,S92042S,S92043S,S92044S,S92045S,S92046S | |
| | S92051S,S92052S,S92053S,S92054S,S92055S,S92056S,S92061S | |
| | S92062S,S92063S,S92064S,S92065S,S92066S,S92101S,S92102S | |
| | S92109S,S92111S,S92112S,S92113S,S92114S,S92115S,S92116S | |
| | S92121S,S92122S,S92123S,S92124S,S92125S,S92126S,S92131S | |
| | S92132S,S92133S,S92134S,S92135S,S92136S,S92141S,S92142S | |
| | S92143S,S92144S,S92145S,S92146S,S92151S,S92152S,S92153S | |
| | S92154S,S92155S,S92156S,S92191S,S92192S,S92199S,S92201S | |
| | S92202S,S92209S,S92211S,S92212S,S92213S,S92214S,S92215S | |

Continued on next page

**Supplementary Table S1:** (cont'd) A comprehensive mapping of diagnosis groups used in the experiments to their corresponding ICD-10 and ICD-9 codes.

| Diagnosis | ICD-10 | ICD-9 |
|---|---|---|
| | S92216S,S92221S,S92222S,S92223S,S92224S,S92225S,S92226S | |
| | S92231S,S92232S,S92233S,S92234S,S92235S,S92236S,S92241S | |
| | S92242S,S92243S,S92244S,S92245S,S92246S,S92251S,S92252S | |
| | S92253S,S92254S,S92255S,S92256S,S92301S,S92302S,S92309S | |
| | S92311S,S92312S,S92313S,S92314S,S92315S,S92316S,S92321S | |
| | S92322S,S92323S,S92324S,S92325S,S92326S,S92331S,S92332S | |
| | S92333S,S92334S,S92335S,S92336S,S92341S,S92342S,S92343S | |
| | S92344S,S92345S,S92346S,S92351S,S92352S,S92353S,S92354S | |
| | S92355S,S92356S,S92401S,S92402S,S92403S,S92404S,S92405S | |
| | S92406S,S92411S,S92412S,S92413S,S92414S,S92415S,S92416S | |
| | S92421S,S92422S,S92423S,S92424S,S92425S,S92426S,S92491S | |
| | S92492S,S92499S,S92501S,S92502S,S92503S,S92504S,S92505S | |
| | S92506S,S92511S,S92512S,S92513S,S92514S,S92515S,S92516S | |
| | S92521S,S92522S,S92523S,S92524S,S92525S,S92526S,S92531S | |
| | S92532S,S92533S,S92534S,S92535S,S92536S,S92591S,S92592S | |
| | S92599S,S92811S,S92812S,S92819S,S92901S,S92902S,S92909S | |
| | S92911S,S92912S,S92919S,S9301XS,S9302XS,S9303XS,S9304XS | |
| | S9305XS,S9306XS,S93101S,S93102S,S93103S,S93104S,S93105S | |
| | S93106S,S93111S,S93112S,S93113S,S93114S,S93115S,S93116S | |
| | S93119S,S93121S,S93122S,S93123S,S93124S,S93125S,S93126S | |
| | S93129S,S93131S,S93132S,S93133S,S93134S,S93135S,S93136S | |
| | S93139S,S93141S,S93142S,S93143S,S93144S,S93145S,S93146S | |
| | S93149S,S93301S,S93302S,S93303S,S93304S,S93305S,S93306S | |
| | S93311S,S93312S,S93313S,S93314S,S93315S,S93316S,S93321S | |
| | S93322S,S93323S,S93324S,S93325S,S93326S,S93331S,S93332S | |
| | S93333S,S93334S,S93335S,S93336S,S93401S,S93402S,S93409S | |
| | S93411S,S93412S,S93419S,S93421S,S93422S,S93429S,S93431S | |
| | S93432S,S93439S,S93491S,S93492S,S93499S,S93501S,S93502S | |
| | S93503S,S93504S,S93505S,S93506S,S93509S,S93511S,S93512S | |
| | S93513S,S93514S,S93515S,S93516S,S93519S,S93521S,S93522S | |
| | S93523S,S93524S,S93525S,S93526S,S93529S,S93601S,S93602S | |
| | S93609S,S93611S,S93612S,S93619S,S93621S,S93622S,S93629S | |
| | S93691S,S93692S,S93699S,S9400XS,S9401XS,S9402XS,S9410XS | |
| | S9411XS,S9412XS,S9420XS,S9421XS,S9422XS,S9430XS,S9431XS | |
| | S9432XS,S948X1S,S948X2S,S948X9S,S9490XS,S9491XS,S9492XS | |
| | S95001S,S95002S,S95009S,S95011S,S95012S,S95019S,S95091S | |
| | S95092S,S95099S,S95101S,S95102S,S95109S,S95111S,S95112S | |
| | S95119S,S95191S,S95192S,S95199S,S95201S,S95202S,S95209S | |
| | S95211S,S95212S,S95219S,S95291S,S95292S,S95299S,S95801S | |
| | S95802S,S95809S,S95811S,S95812S,S95819S,S95891S,S95892S | |

Continued on next page

**Supplementary Table S1:** (cont'd) A comprehensive mapping of diagnosis groups used in the experiments to their corresponding ICD-10 and ICD-9 codes.

| Diagnosis | ICD-10 | ICD-9 |
|---|---|---|
| | S95899S,S95901S,S95902S,S95909S,S95911S,S95912S,S95919S | |
| | S95991S,S95992S,S95999S,S96001S,S96002S,S96009S,S96011S | |
| | S96012S,S96019S,S96021S,S96022S,S96029S,S96091S,S96092S | |
| | S96099S,S96101S,S96102S,S96109S,S96111S,S96112S,S96119S | |
| | S96121S,S96122S,S96129S,S96191S,S96192S,S96199S,S96201S | |
| | S96202S,S96209S,S96211S,S96212S,S96219S,S96221S,S96222S | |
| | S96229S,S96291S,S96292S,S96299S,S96801S,S96802S,S96809S | |
| | S96811S,S96812S,S96819S,S96821S,S96822S,S96829S,S96891S | |
| | S96892S,S96899S,S96901S,S96902S,S96909S,S96911S,S96912S | |
| | S96919S,S96921S,S96922S,S96929S,S96991S,S96992S,S96999S | |
| | S9700XS,S9701XS,S9702XS,S97101S,S97102S,S97109S,S97111S | |
| | S97112S,S97119S,S97121S,S97122S,S97129S,S9780XS,S9781XS | |
| | S9782XS,S98011S,S98012S,S98019S,S98021S,S98022S,S98029S | |
| | S98111S,S98112S,S98119S,S98121S,S98122S,S98129S,S98131S | |
| | S98132S,S98139S,S98141S,S98142S,S98149S,S98211S,S98212S | |
| | S98219S,S98221S,S98222S,S98229S,S98311S,S98312S,S98319S | |
| | S98321S,S98322S,S98329S,S98911S,S98912S,S98919S,S98921S | |
| | S98922S,S98929S,S99001S,S99002S,S99009S,S99011S,S99012S | |
| | S99019S,S99021S,S99022S,S99029S,S99031S,S99032S,S99039S | |
| | S99041S,S99042S,S99049S,S99091S,S99092S,S99099S,S99101S | |
| | S99102S,S99109S,S99111S,S99112S,S99119S,S99121S,S99122S | |
| | S99129S,S99131S,S99132S,S99139S,S99141S,S99142S,S99149S | |
| | S99191S,S99192S,S99199S,S99201S,S99202S,S99209S,S99211S | |
| | S99212S,S99219S,S99221S,S99222S,S99229S,S99231S,S99232S | |
| | S99239S,S99241S,S99242S,S99249S,S99291S,S99292S,S99299S | |
| | S99811S,S99812S,S99819S,S99821S,S99822S,S99829S,S99911S | |
| | S99912S,S99919S,S99921S,S99922S,S99929S,T07XXXS,T148XXS | |
| | T1490XS,T1491XS,T1500XS,T1501XS,T1502XS,T1510XS,T1511XS | |
| | T1512XS,T1580XS,T1581XS,T1582XS,T1590XS,T1591XS,T1592XS | |
| | T161XXS,T162XXS,T169XXS,T170XXS,T171XXS,T17200S,T17208S | |
| | T17210S,T17218S,T17220S,T17228S,T17290S,T17298S,T17300S | |
| | T17308S,T17310S,T17318S,T17320S,T17328S,T17390S,T17398S | |
| | T17400S,T17408S,T17410S,T17418S,T17420S,T17428S,T17490S | |
| | T17498S,T17500S,T17508S,T17510S,T17518S,T17520S,T17528S | |
| | T17590S,T17598S,T17800S,T17808S,T17810S,T17818S,T17820S | |
| | T17828S,T17890S,T17898S,T17900S,T17908S,T17910S,T17918S | |
| | T17920S,T17928S,T17990S,T17998S,T180XXS,T18100S,T18108S | |
| | T18110S,T18118S,T18120S,T18128S,T18190S,T18198S,T182XXS | |
| | T183XXS,T184XXS,T185XXS,T188XXS,T189XXS,T190XXS,T191XXS | |
| | T192XXS,T193XXS,T194XXS,T198XXS,T199XXS,T2000XS,T20011S | |

Continued on next page

**Supplementary Table S1:** (cont'd) A comprehensive mapping of diagnosis groups used in the experiments to their corresponding ICD-10 and ICD-9 codes.

| Diagnosis | ICD-10 | ICD-9 |
|---|---|---|
| | T20012S,T20019S,T2002XS,T2003XS,T2004XS,T2005XS,T2006XS | |
| | T2007XS,T2009XS,T2010XS,T20111S,T20112S,T20119S,T2012XS | |
| | T2013XS,T2014XS,T2015XS,T2016XS,T2017XS,T2019XS,T2020XS | |
| | T20211S,T20212S,T20219S,T2022XS,T2023XS,T2024XS,T2025XS | |
| | T2026XS,T2027XS,T2029XS,T2030XS,T20311S,T20312S,T20319S | |
| | T2032XS,T2033XS,T2034XS,T2035XS,T2036XS,T2037XS,T2039XS | |
| | T2040XS,T20411S,T20412S,T20419S,T2042XS,T2043XS,T2044XS | |
| | T2045XS,T2046XS,T2047XS,T2049XS,T2050XS,T20511S,T20512S | |
| | T20519S,T2052XS,T2053XS,T2054XS,T2055XS,T2056XS,T2057XS | |
| | T2059XS,T2060XS,T20611S,T20612S,T20619S,T2062XS,T2063XS | |
| | T2064XS,T2065XS,T2066XS,T2067XS,T2069XS,T2070XS,T20711S | |
| | T20712S,T20719S,T2072XS,T2073XS,T2074XS,T2075XS,T2076XS | |
| | T2077XS,T2079XS,T2100XS,T2101XS,T2102XS,T2103XS,T2104XS | |
| | T2105XS,T2106XS,T2107XS,T2109XS,T2110XS,T2111XS,T2112XS | |
| | T2113XS,T2114XS,T2115XS,T2116XS,T2117XS,T2119XS,T2120XS | |
| | T2121XS,T2122XS,T2123XS,T2124XS,T2125XS,T2126XS,T2127XS | |
| | T2129XS,T2130XS,T2131XS,T2132XS,T2133XS,T2134XS,T2135XS | |
| | T2136XS,T2137XS,T2139XS,T2140XS,T2141XS,T2142XS,T2143XS | |
| | T2144XS,T2145XS,T2146XS,T2147XS,T2149XS,T2150XS,T2151XS | |
| | T2152XS,T2153XS,T2154XS,T2155XS,T2156XS,T2157XS,T2159XS | |
| | T2160XS,T2161XS,T2162XS,T2163XS,T2164XS,T2165XS,T2166XS | |
| | T2167XS,T2169XS,T2170XS,T2171XS,T2172XS,T2173XS,T2174XS | |
| | T2175XS,T2176XS,T2177XS,T2179XS,T2200XS,T22011S,T22012S | |
| | T22019S,T22021S,T22022S,T22029S,T22031S,T22032S,T22039S | |
| | T22041S,T22042S,T22049S,T22051S,T22052S,T22059S,T22061S | |
| | T22062S,T22069S,T22091S,T22092S,T22099S,T2210XS,T22111S | |
| | T22112S,T22119S,T22121S,T22122S,T22129S,T22131S,T22132S | |
| | T22139S,T22141S,T22142S,T22149S,T22151S,T22152S,T22159S | |
| | T22161S,T22162S,T22169S,T22191S,T22192S,T22199S,T2220XS | |
| | T22211S,T22212S,T22219S,T22221S,T22222S,T22229S,T22231S | |
| | T22232S,T22239S,T22241S,T22242S,T22249S,T22251S,T22252S | |
| | T22259S,T22261S,T22262S,T22269S,T22291S,T22292S,T22299S | |
| | T2230XS,T22311S,T22312S,T22319S,T22321S,T22322S,T22329S | |
| | T22331S,T22332S,T22339S,T22341S,T22342S,T22349S,T22351S | |
| | T22352S,T22359S,T22361S,T22362S,T22369S,T22391S,T22392S | |
| | T22399S,T2240XS,T22411S,T22412S,T22419S,T22421S,T22422S | |
| | T22429S,T22431S,T22432S,T22439S,T22441S,T22442S,T22449S | |
| | T22451S,T22452S,T22459S,T22461S,T22462S,T22469S,T22491S | |
| | T22492S,T22499S,T2250XS,T22511S,T22512S,T22519S,T22521S | |
| | T22522S,T22529S,T22531S,T22532S,T22539S,T22541S,T22542S | |

Continued on next page

**Supplementary Table S1:** (cont'd) A comprehensive mapping of diagnosis groups used in the experiments to their corresponding ICD-10 and ICD-9 codes.

| Diagnosis | ICD-10 | ICD-9 |
|---|---|---|
| | T22549S,T22551S,T22552S,T22559S,T22561S,T22562S,T22569S | |
| | T22591S,T22592S,T22599S,T2260XS,T22611S,T22612S,T22619S | |
| | T22621S,T22622S,T22629S,T22631S,T22632S,T22639S,T22641S | |
| | T22642S,T22649S,T22651S,T22652S,T22659S,T22661S,T22662S | |
| | T22669S,T22691S,T22692S,T22699S,T2270XS,T22711S,T22712S | |
| | T22719S,T22721S,T22722S,T22729S,T22731S,T22732S,T22739S | |
| | T22741S,T22742S,T22749S,T22751S,T22752S,T22759S,T22761S | |
| | T22762S,T22769S,T22791S,T22792S,T22799S,T23001S,T23002S | |
| | T23009S,T23011S,T23012S,T23019S,T23021S,T23022S,T23029S | |
| | T23031S,T23032S,T23039S,T23041S,T23042S,T23049S,T23051S | |
| | T23052S,T23059S,T23061S,T23062S,T23069S,T23071S,T23072S | |
| | T23079S,T23091S,T23092S,T23099S,T23101S,T23102S,T23109S | |
| | T23111S,T23112S,T23119S,T23121S,T23122S,T23129S,T23131S | |
| | T23132S,T23139S,T23141S,T23142S,T23149S,T23151S,T23152S | |
| | T23159S,T23161S,T23162S,T23169S,T23171S,T23172S,T23179S | |
| | T23191S,T23192S,T23199S,T23201S,T23202S,T23209S,T23211S | |
| | T23212S,T23219S,T23221S,T23222S,T23229S,T23231S,T23232S | |
| | T23239S,T23241S,T23242S,T23249S,T23251S,T23252S,T23259S | |
| | T23261S,T23262S,T23269S,T23271S,T23272S,T23279S,T23291S | |
| | T23292S,T23299S,T23301S,T23302S,T23309S,T23311S,T23312S | |
| | T23319S,T23321S,T23322S,T23329S,T23331S,T23332S,T23339S | |
| | T23341S,T23342S,T23349S,T23351S,T23352S,T23359S,T23361S | |
| | T23362S,T23369S,T23371S,T23372S,T23379S,T23391S,T23392S | |
| | T23399S,T23401S,T23402S,T23409S,T23411S,T23412S,T23419S | |
| | T23421S,T23422S,T23429S,T23431S,T23432S,T23439S,T23441S | |
| | T23442S,T23449S,T23451S,T23452S,T23459S,T23461S,T23462S | |
| | T23469S,T23471S,T23472S,T23479S,T23491S,T23492S,T23499S | |
| | T23501S,T23502S,T23509S,T23511S,T23512S,T23519S,T23521S | |
| | T23522S,T23529S,T23531S,T23532S,T23539S,T23541S,T23542S | |
| | T23549S,T23551S,T23552S,T23559S,T23561S,T23562S,T23569S | |
| | T23571S,T23572S,T23579S,T23591S,T23592S,T23599S,T23601S | |
| | T23602S,T23609S,T23611S,T23612S,T23619S,T23621S,T23622S | |
| | T23629S,T23631S,T23632S,T23639S,T23641S,T23642S,T23649S | |
| | T23651S,T23652S,T23659S,T23661S,T23662S,T23669S,T23671S | |
| | T23672S,T23679S,T23691S,T23692S,T23699S,T23701S,T23702S | |
| | T23709S,T23711S,T23712S,T23719S,T23721S,T23722S,T23729S | |
| | T23731S,T23732S,T23739S,T23741S,T23742S,T23749S,T23751S | |
| | T23752S,T23759S,T23761S,T23762S,T23769S,T23771S,T23772S | |
| | T23779S,T23791S,T23792S,T23799S,T24001S,T24002S,T24009S | |
| | T24011S,T24012S,T24019S,T24021S,T24022S,T24029S,T24031S | |

Continued on next page

**Supplementary Table S1:** (cont'd) A comprehensive mapping of diagnosis groups used in the experiments to their corresponding ICD-10 and ICD-9 codes.

| Diagnosis | ICD-10 | ICD-9 |
|---|---|---|
| | T24032S,T24039S,T24091S,T24092S,T24099S,T24101S,T24102S | |
| | T24109S,T24111S,T24112S,T24119S,T24121S,T24122S,T24129S | |
| | T24131S,T24132S,T24139S,T24191S,T24192S,T24199S,T24201S | |
| | T24202S,T24209S,T24211S,T24212S,T24219S,T24221S,T24222S | |
| | T24229S,T24231S,T24232S,T24239S,T24291S,T24292S,T24299S | |
| | T24301S,T24302S,T24309S,T24311S,T24312S,T24319S,T24321S | |
| | T24322S,T24329S,T24331S,T24332S,T24339S,T24391S,T24392S | |
| | T24399S,T24401S,T24402S,T24409S,T24411S,T24412S,T24419S | |
| | T24421S,T24422S,T24429S,T24431S,T24432S,T24439S,T24491S | |
| | T24492S,T24499S,T24501S,T24502S,T24509S,T24511S,T24512S | |
| | T24519S,T24521S,T24522S,T24529S,T24531S,T24532S,T24539S | |
| | T24591S,T24592S,T24599S,T24601S,T24602S,T24609S,T24611S | |
| | T24612S,T24619S,T24621S,T24622S,T24629S,T24631S,T24632S | |
| | T24639S,T24691S,T24692S,T24699S,T24701S,T24702S,T24709S | |
| | T24711S,T24712S,T24719S,T24721S,T24722S,T24729S,T24731S | |
| | T24732S,T24739S,T24791S,T24792S,T24799S,T25011S,T25012S | |
| | T25019S,T25021S,T25022S,T25029S,T25031S,T25032S,T25039S | |
| | T25091S,T25092S,T25099S,T25111S,T25112S,T25119S,T25121S | |
| | T25122S,T25129S,T25131S,T25132S,T25139S,T25191S,T25192S | |
| | T25199S,T25211S,T25212S,T25219S,T25221S,T25222S,T25229S | |
| | T25231S,T25232S,T25239S,T25291S,T25292S,T25299S,T25311S | |
| | T25312S,T25319S,T25321S,T25322S,T25329S,T25331S,T25332S | |
| | T25339S,T25391S,T25392S,T25399S,T25411S,T25412S,T25419S | |
| | T25421S,T25422S,T25429S,T25431S,T25432S,T25439S,T25491S | |
| | T25492S,T25499S,T25511S,T25512S,T25519S,T25521S,T25522S | |
| | T25529S,T25531S,T25532S,T25539S,T25591S,T25592S,T25599S | |
| | T25611S,T25612S,T25619S,T25621S,T25622S,T25629S,T25631S | |
| | T25632S,T25639S,T25691S,T25692S,T25699S,T25711S,T25712S | |
| | T25719S,T25721S,T25722S,T25729S,T25731S,T25732S,T25739S | |
| | T25791S,T25792S,T25799S,T2600XS,T2601XS,T2602XS,T2610XS | |
| | T2611XS,T2612XS,T2620XS,T2621XS,T2622XS,T2630XS,T2631XS | |
| | T2632XS,T2640XS,T2641XS,T2642XS,T2650XS,T2651XS,T2652XS | |
| | T2660XS,T2661XS,T2662XS,T2670XS,T2671XS,T2672XS,T2680XS | |
| | T2681XS,T2682XS,T2690XS,T2691XS,T2692XS,T270XXS,T271XXS | |
| | T272XXS,T273XXS,T274XXS,T275XXS,T276XXS,T277XXS,T280XXS | |
| | T281XXS,T282XXS,T283XXS,T2840XS,T28411S,T28412S,T28419S | |
| | T2849XS,T285XXS,T286XXS,T287XXS,T288XXS,T2890XS,T28911S | |
| | T28912S,T28919S,T2899XS,T7800XS,T7801XS,T7802XS,T7803XS | |
| | T7804XS,T7805XS,T7806XS,T7807XS,T7808XS,T7809XS,T781XXS | |
| | T782XXS,T783XXS,T7840XS,T7841XS,T7849XS,T790XXS,T791XXS | |

Continued on next page

**Supplementary Table S1:** (cont'd) A comprehensive mapping of diagnosis groups used in the experiments to their corresponding ICD-10 and ICD-9 codes.

| Diagnosis | ICD-10 | ICD-9 |
|---|---|---|
| | T792XXS,T794XXS,T795XXS,T796XXS,T797XXS,T798XXS,T799XXS | |
| | T79A0XS,T79A11S,T79A12S,T79A19S,T79A21S,T79A22S,T79A29S | |
| | T79A3XS,T79A9XS,T84040S,T84041S,T84042S,T84043S,T84048S | |
| | T84049S | |
| Internal organ | S2600XA,S2601XA,S26020A,S26021A,S26022A,S2609XA,S2610XA | 8600,8602,8604,86100,86101,86102,86103,86120,86121,86122,8620 |
| injury, initial | S2611XA,S2612XA,S2619XA,S2690XA,S2691XA,S2692XA,S2699XA | 86221,86222,86229,8628,8630,86320,86321,86329,86340,86341 |
| encounter | S270XXA,S271XXA,S272XXA,S27301A,S27302A,S27309A,S27311A | 86342,86343,86344,86345,86346,86349,86381,86382,86383,86384 |
| | S27312A,S27319A,S27321A,S27322A,S27329A,S27331A,S27332A | 86385,86389,86400,86401,86402,86403,86404,86405,86409,86500 |
| | S27339A,S27391A,S27392A,S27399A,S27401A,S27402A,S27409A | 86501,86502,86503,86504,86509,86600,86601,86602,86603,8670 |
| | S27411A,S27412A,S27419A,S27421A,S27422A,S27429A,S27431A | 8672,8674,8676,8678,86800,86801,86802,86803,86804,86809,8690 |
| | S27432A,S27439A,S27491A,S27492A,S27499A,S2750XA,S2751XA | |
| | S2752XA,S2753XA,S2759XA,S2760XA,S2763XA,S2769XA,S27802A | |
| | S27803A,S27808A,S27809A,S27812A,S27813A,S27818A,S27819A | |
| | S27892A,S27893A,S27898A,S27899A,S279XXA,S3600XA,S36020A | |
| | S36021A,S36029A,S36030A,S36031A,S36032A,S36039A,S3609XA | |
| | S36112A,S36113A,S36114A,S36115A,S36116A,S36118A,S36119A | |
| | S36122A,S36123A,S36128A,S36129A,S3613XA,S36200A,S36201A | |
| | S36202A,S36209A,S36220A,S36221A,S36222A,S36229A,S36230A | |
| | S36231A,S36232A,S36239A,S36240A,S36241A,S36242A,S36249A | |
| | S36250A,S36251A,S36252A,S36259A,S36260A,S36261A,S36262A | |
| | S36269A,S36290A,S36291A,S36292A,S36299A,S3630XA,S3632XA | |
| | S3633XA,S3639XA,S36400A,S36408A,S36409A,S36410A,S36418A | |
| | S36419A,S36420A,S36428A,S36429A,S36430A,S36438A,S36439A | |
| | S36490A,S36498A,S36499A,S36500A,S36501A,S36502A,S36503A | |
| | S36508A,S36509A,S36510A,S36511A,S36512A,S36513A,S36518A | |
| | S36519A,S36520A,S36521A,S36522A,S36523A,S36528A,S36529A | |
| | S36530A,S36531A,S36532A,S36533A,S36538A,S36539A,S36590A | |
| | S36591A,S36592A,S36593A,S36598A,S36599A,S3660XA,S3661XA | |
| | S3662XA,S3663XA,S3669XA,S3681XA,S36892A,S36893A,S36898A | |
| | S36899A,S3690XA,S3692XA,S3693XA,S3699XA,S37001A,S37002A | |
| | S37009A,S37011A,S37012A,S37019A,S37021A,S37022A,S37029A | |
| | S37031A,S37032A,S37039A,S37041A,S37042A,S37049A,S37051A | |
| | S37052A,S37059A,S37061A,S37062A,S37069A,S37091A,S37092A | |
| | S37099A,S3710XA,S3712XA,S3713XA,S3719XA,S3720XA,S3722XA | |
| | S3723XA,S3729XA,S3730XA,S3732XA,S3733XA,S3739XA,S37401A | |
| | S37402A,S37409A,S37421A,S37422A,S37429A,S37431A,S37432A | |
| | S37439A,S37491A,S37492A,S37499A,S37501A,S37502A,S37509A | |
| | S37511A,S37512A,S37519A,S37521A,S37522A,S37529A,S37531A | |
| | S37532A,S37539A,S37591A,S37592A,S37599A,S3760XA,S3762XA | |

Continued on next page

**Supplementary Table S1:** (cont'd) A comprehensive mapping of diagnosis groups used in the experiments to their corresponding ICD-10 and ICD-9 codes.

| Diagnosis | ICD-10 | ICD-9 |
|---|---|---|
| | S3763XA,S3769XA,S37812A,S37813A,S37818A,S37819A,S37822A | |
| | S37823A,S37828A,S37829A,S37892A,S37893A,S37898A,S37899A | |
| | S3790XA,S3792XA,S3793XA,S3799XA,U070 | |
| Internal organ | S2600XD,S2601XD,S26020D,S26021D,S26022D,S2609XD,S2610XD | V5889 |
| injury, subsequent | S2611XD,S2612XD,S2619XD,S2690XD,S2691XD,S2692XD,S2699XD | |
| encounter | S270XXD,S271XXD,S272XXD,S27301D,S27302D,S27309D,S27311D | |
| | S27312D,S27319D,S27321D,S27322D,S27329D,S27331D,S27332D | |
| | S27339D,S27391D,S27392D,S27399D,S27401D,S27402D,S27409D | |
| | S27411D,S27412D,S27419D,S27421D,S27422D,S27429D,S27431D | |
| | S27432D,S27439D,S27491D,S27492D,S27499D,S2750XD,S2751XD | |
| | S2752XD,S2753XD,S2759XD,S2760XD,S2763XD,S2769XD,S27802D | |
| | S27803D,S27808D,S27809D,S27812D,S27813D,S27818D,S27819D | |
| | S27892D,S27893D,S27898D,S27899D,S279XXD,S3600XD,S36020D | |
| | S36021D,S36029D,S36030D,S36031D,S36032D,S36039D,S3609XD | |
| | S36112D,S36113D,S36114D,S36115D,S36116D,S36118D,S36119D | |
| | S36122D,S36123D,S36128D,S36129D,S3613XD,S36200D,S36201D | |
| | S36202D,S36209D,S36220D,S36221D,S36222D,S36229D,S36230D | |
| | S36231D,S36232D,S36239D,S36240D,S36241D,S36242D,S36249D | |
| | S36250D,S36251D,S36252D,S36259D,S36260D,S36261D,S36262D | |
| | S36269D,S36290D,S36291D,S36292D,S36299D,S3630XD,S3632XD | |
| | S3633XD,S3639XD,S36400D,S36408D,S36409D,S36410D,S36418D | |
| | S36419D,S36420D,S36428D,S36429D,S36430D,S36438D,S36439D | |
| | S36490D,S36498D,S36499D,S36500D,S36501D,S36502D,S36503D | |
| | S36508D,S36509D,S36510D,S36511D,S36512D,S36513D,S36518D | |
| | S36519D,S36520D,S36521D,S36522D,S36523D,S36528D,S36529D | |
| | S36530D,S36531D,S36532D,S36533D,S36538D,S36539D,S36590D | |
| | S36591D,S36592D,S36593D,S36598D,S36599D,S3660XD,S3661XD | |
| | S3662XD,S3663XD,S3669XD,S3681XD,S36892D,S36893D,S36898D | |
| | S36899D,S3690XD,S3692XD,S3693XD,S3699XD,S37001D,S37002D | |
| | S37009D,S37011D,S37012D,S37019D,S37021D,S37022D,S37029D | |
| | S37031D,S37032D,S37039D,S37041D,S37042D,S37049D,S37051D | |
| | S37052D,S37059D,S37061D,S37062D,S37069D,S37091D,S37092D | |
| | S37099D,S3710XD,S3712XD,S3713XD,S3719XD,S3720XD,S3722XD | |
| | S3723XD,S3729XD,S3730XD,S3732XD,S3733XD,S3739XD,S37401D | |
| | S37402D,S37409D,S37421D,S37422D,S37429D,S37431D,S37432D | |
| | S37439D,S37491D,S37492D,S37499D,S37501D,S37502D,S37509D | |
| | S37511D,S37512D,S37519D,S37521D,S37522D,S37529D,S37531D | |
| | S37532D,S37539D,S37591D,S37592D,S37599D,S3760XD,S3762XD | |
| | S3763XD,S3769XD,S37812D,S37813D,S37818D,S37819D,S37822D | |

Continued on next page

**Supplementary Table S1:** (cont'd) A comprehensive mapping of diagnosis groups used in the experiments to their corresponding ICD-10 and ICD-9 codes.

| Diagnosis | ICD-10 | ICD-9 |
| --- | --- | --- |
| | S37823D,S37828D,S37829D,S37892D,S37893D,S37898D,S37899D S3790XD,S3792XD,S3793XD,S3799XD | |
| Intestinal infection | A000,A001,A009,A0100,A0109,A011,A012,A013,A014,A020,A0229 A029,A030,A031,A032,A033,A038,A039,A040,A041,A042,A043,A044 A045,A046,A047,A0471,A0472,A048,A049,A060,A061,A062,A063,A064 A069,A070,A071,A072,A073,A074,A078,A079,A080,A0811,A0819,A082 A0831,A0832,A0839,A084,A088,A09,A1832,A213,A222,B3782 | 0010,0011,0019,0020,0021,0022,0023,0029,0030,00329,0039,0040 0041,0042,0043,0048,0049,0060,0061,0062,0063,0068,0069,0070 0071,0072,0073,0074,0075,0078,0079,00800,00801,00802,00803 00804,00809,0082,00841,00843,00844,00845,00849,0085,00861 00862,00863,00864,00865,00866,00867,00869,0088,0090,01480 0211,0222,11285,1365 |
| Intestinal obstruction and ileus | K315,K4000,K4001,K4030,K4031,K4100,K4101,K4130,K4131,K420 K430,K433,K436,K440,K450,K460,K50012,K50112,K50812,K50912 K51012,K51212,K51312,K51412,K51512,K51812,K51912,K560,K561 K562,K563,K5641,K5649,K565,K5650,K5651,K5652,K5660,K56600 K56601,K56609,K5669,K56690,K56691,K56699,K567,K913,K9130 K9131,K9132 | 5372,5373,55010,55011,55012,55013,55200,55201,55202,55203 5521,55220,55221,55229,5523,5528,5529,5550,5551,5552,5559 5562,5563,5564,5565,5566,5568,5569,5600,5601,5602,56030,56031 56032,56039,56081,56089,5609,99749 |
| Juvenile arthritis | M0800,M08011,M08012,M08019,M08021,M08022,M08029,M08031,M08032 M08039,M08041,M08042,M08049,M08051,M08052,M08059,M08061 M08062,M08069,M08071,M08072,M08079,M0808,M0809,M080A,M081 M0820,M08211,M08212,M08219,M08221,M08222,M08229,M08231,M08232 M08239,M08241,M08242,M08249,M08251,M08252,M08259,M08261 M08262,M08269,M08271,M08272,M08279,M0828,M0829,M082A,M083 M0840,M08411,M08412,M08419,M08421,M08422,M08429,M08431,M08432 M08439,M08441,M08442,M08449,M08451,M08452,M08459,M08461 M08462,M08469,M08471,M08472,M08479,M0848,M084A,M0880,M08811 M08812,M08819,M08821,M08822,M08829,M08831,M08832,M08839 M08841,M08842,M08849,M08851,M08852,M08859,M08861,M08862 M08869,M08871,M08872,M08879,M0888,M0889,M0890,M08911,M08912 M08919,M08921,M08922,M08929,M08931,M08932,M08939,M08941 M08942,M08949,M08951,M08952,M08959,M08961,M08962,M08969 M08971,M08972,M08979,M0898,M0899,M089A | 71430,71432,7200 |
| Leukemia - acute lymphoblastic leukemia (ALL) | C9100,C9101,C9102 | 20400,20401,20402 |
| Leukemia - acute myeloid leukemia (AML) | C9200,C9201,C9202,C9240,C9241,C9242,C9250,C9251,C9252,C9260 C9261,C9262,C92A0,C92A1,C92A2,C9300,C9301,C9302,C9400,C9401 C9402,C9420,C9421,C9422 | 20500,20501,20502,20600,20601,20602,20700,20701,20702,20720 20721,20722 |

Continued on next page

**Supplementary Table S1:** (cont'd) A comprehensive mapping of diagnosis groups used in the experiments to their corresponding ICD-10 and ICD-9 codes.

| Diagnosis | ICD-10 | ICD-9 |
|---|---|---|
| Leukemia - all other types | C9130,C9131,C9132,C9150,C9151,C9152,C9160,C9161,C9162,C9190 C9191,C9192,C91A0,C91A1,C91A2,C91Z0,C91Z1,C91Z2,C9290,C9291 C9292,C92Z0,C92Z1,C92Z2,C9330,C9331,C9332,C9390,C9391,C9392 C93Z0,C93Z1,C93Z2,C9430,C9431,C9432,C9440,C9441,C9442,C946 C9480,C9481,C9482,C9500,C9501,C9502,C9510,C9511,C9512,C9590 C9591,C9592 | 20480,20481,20482,20490,20491,20492,20580,20581,20582,20590 20591,20592,20620,20621,20622,20680,20681,20682,20690,20691 20692,20780,20781,20782,20800,20801,20802,20810,20811,20812 20820,20821,20822,20880,20881,20882,20890,20891,20892,23879 |
| Leukemia - chronic lymphocytic leukemia (CLL) | C9110,C9111,C9112 | 20410,20411,20412 |
| Leukemia - chronic myeloid leukemia (CML) | C9210,C9211,C9212,C9220,C9221,C9222,C9310,C9311,C9312 | 20510,20511,20512,20520,20521,20522,20610,20611,20612 |
| Leukemia - hairy cell | C9140,C9141,C9142 | 20240 |
| Lifestyle life management factors | Z720,Z723,Z724,Z7251,Z7252,Z7253,Z726,Z72810,Z72811,Z72820 Z72821,Z72823,Z7289,Z729,Z730,Z731,Z732,Z733,Z734,Z735,Z736 Z73810,Z73811,Z73812,Z73819,Z7382,Z7389,Z739 | V4985,V624,V6289,V690,V691,V692,V693,V694,V695,V698,V699 V7101,V7102 |
| Liveborn | Z3800,Z3801,Z381,Z382,Z3830,Z3831,Z384,Z385,Z3861,Z3862,Z3863 Z3864,Z3865,Z3866,Z3868,Z3869,Z387,Z388 | V3000,V3001,V301,V302,V3100,V3101,V311,V312,V3200,V3201,V321 V322,V3300,V3301,V331,V332,V3400,V3401,V341,V342,V3500,V3501 V351,V352,V3600,V3601,V361,V362,V3700,V3701,V371,V372,V3900 V391,V392 |
| Low back pain | M5106,M5116,M5117,M5126,M5127,M5136,M5137,M5146,M5147,M5186 M5187,M51A0,M51A1,M51A2,M51A3,M51A4,M51A5,M532X6,M532X7 M532X8,M533,M5386,M5387,M5388,M5416,M5417,M5418,M5430,M5431 M5432,M5440,M5441,M5442,M545,M5450,M5451,M5459,M62830 | 71888,72210,72232,72252,72273,72293,7242,7243,7244,7246,72470 72471,72479,7248,7292 |
| Lung disease due to external agents | J60,J61,J620,J628,J630,J631,J632,J633,J634,J635,J636,J64,J65 J660,J661,J662,J668,J670,J671,J672,J673,J674,J675,J676,J677 J678,J679,J680,J681,J682,J683,J684,J688,J689,J691,J698,J700 J701,J702,J703,J704,J705,J708,J709,U070 | 4950,4951,4952,4953,4954,4955,4956,4957,4958,4959,500,501,502 503,504,505,5060,5061,5062,5063,5064,5069,5071,5078,5080,5081 5082,5088,5089 |
| Malaise and fatigue | G933,R530,R531,R5381,R5382,R5383 | 78071,78079,7993 |
| Male infertility | N4601,N46021,N46022,N46023,N46024,N46025,N46029,N4611,N46121 | 6060,6061,6068,6069 |

Continued on next page

**Supplementary Table S1:** (cont'd) A comprehensive mapping of diagnosis groups used in the experiments to their corresponding ICD-10 and ICD-9 codes.

| Diagnosis | ICD-10 | ICD-9 |
|---|---|---|
| | N46122,N46123,N46124,N46125,N46129,N468,N469 | |
| Male reproductive system cancers - all other types | C6300,C6301,C6302,C6310,C6311,C6312,C632,C637,C638,C639,D0760 D0761,D0769 | 1875,1876,1877,1878,1879,2336 |
| Male reproductive system cancers - penis | C600,C601,C602,C608,C609,D074 | 1871,1872,1873,1874,1878,2335 |
| Male reproductive system cancers - prostate | C61,D075 | 185,2334 |
| Male reproductive system cancers - testis | C6200,C6201,C6202,C6210,C6211,C6212,C6290,C6291,C6292 | 1860,1869 |
| Malignant neoplasm, unspecified | C800,C801,C802,D099 | 1990,1991,1992,2349 |
| Malignant neuroendocrine tumors | C7A00,C7A010,C7A011,C7A012,C7A019,C7A020,C7A021,C7A022,C7A023 C7A024,C7A025,C7A026,C7A029,C7A090,C7A091,C7A092,C7A093 C7A094,C7A095,C7A096,C7A098,C7A1,C7A8,C7B00,C7B01,C7B02,C7B03 C7B04,C7B09,C7B1,C7B8 | 20900,20901,20902,20903,20910,20911,20912,20913,20914,20915 20916,20917,20920,20921,20922,20923,20924,20925,20926,20927 20929,20930,20970,20971,20972,20973,20974,20975,20979 |
| Malnutrition | E40,E41,E42,E43,E440,E441,E45,E46,O2510,O2511,O2512,O2513 O252,O253 | 260,261,262,2630,2631,2632,2638,2639,64890,64891,64893,64894 |
| Malposition, disproportion or other labor complications | O320XX0,O320XX1,O320XX2,O320XX3,O320XX4,O320XX5,O320XX9 O321XX0,O321XX1,O321XX2,O321XX3,O321XX4,O321XX5,O321XX9 O322XX0,O322XX1,O322XX2,O322XX3,O322XX4,O322XX5,O322XX9 O323XX0,O323XX1,O323XX2,O323XX3,O323XX4,O323XX5,O323XX9 O324XX0,O324XX1,O324XX2,O324XX3,O324XX4,O324XX5,O324XX9 O326XX0,O326XX1,O326XX2,O326XX3,O326XX4,O326XX5,O326XX9 O328XX0,O328XX1,O328XX2,O328XX3,O328XX4,O328XX5,O328XX9 O329XX0,O329XX1,O329XX2,O329XX3,O329XX4,O329XX5,O329XX9,O330 O331,O332,O333XX0,O333XX1,O333XX2,O333XX3,O333XX4,O333XX5 O333XX9,O334XX0,O334XX1,O334XX2,O334XX3,O334XX4,O334XX5 O334XX9,O335XX0,O335XX1,O335XX2,O335XX3,O335XX4,O335XX5 | 64871,64872,65201,65203,65211,65221,65223,65231,65233,65241 65243,65251,65253,65261,65281,65283,65291,65293,65301,65311 65313,65321,65323,65331,65333,65341,65343,65351,65353,65361 65363,65371,65373,65381,65383,65391,65393,65491,65631,65681 65901,65911,66001,66011,66021,66031,66041,66051,66061,66071 66081,66091,66101,66111,66121,66131,66141,66191,66201,66211 66221,66231,66301,66311,66321,66331,66341,66351,66361,66381 66391,66951 |

Continued on next page

**Supplementary Table S1:** (cont'd) A comprehensive mapping of diagnosis groups used in the experiments to their corresponding ICD-10 and ICD-9 codes.

| Diagnosis | ICD-10 | ICD-9 |
|---|---|---|
| | O335XX9,O336XX0,O336XX1,O336XX2,O336XX3,O336XX4,O336XX5 | |
| | O336XX9,O337,O337XX0,O337XX1,O337XX2,O337XX3,O337XX4,O337XX5 | |
| | O337XX9,O338,O339,O610,O611,O618,O619,O620,O621,O622,O623 | |
| | O624,O628,O629,O630,O631,O632,O639,O640XX0,O640XX1,O640XX2 | |
| | O640XX3,O640XX4,O640XX5,O640XX9,O641XX0,O641XX1,O641XX2 | |
| | O641XX3,O641XX4,O641XX5,O641XX9,O642XX0,O642XX1,O642XX2 | |
| | O642XX3,O642XX4,O642XX5,O642XX9,O643XX0,O643XX1,O643XX2 | |
| | O643XX3,O643XX4,O643XX5,O643XX9,O644XX0,O644XX1,O644XX2 | |
| | O644XX3,O644XX4,O644XX5,O644XX9,O645XX0,O645XX1,O645XX2 | |
| | O645XX3,O645XX4,O645XX5,O645XX9,O648XX0,O648XX1,O648XX2 | |
| | O648XX3,O648XX4,O648XX5,O648XX9,O649XX0,O649XX1,O649XX2 | |
| | O649XX3,O649XX4,O649XX5,O649XX9,O650,O651,O652,O653,O654,O655 | |
| | O658,O659,O660,O661,O662,O663,O6640,O6641,O665,O666,O668,O669 | |
| | O68,O690XX0,O690XX1,O690XX2,O690XX3,O690XX4,O690XX5,O690XX9 | |
| | O691XX0,O691XX1,O691XX2,O691XX3,O691XX4,O691XX5,O691XX9 | |
| | O692XX0,O692XX1,O692XX2,O692XX3,O692XX4,O692XX5,O692XX9 | |
| | O693XX0,O693XX1,O693XX2,O693XX3,O693XX4,O693XX5,O693XX9 | |
| | O694XX0,O694XX1,O694XX2,O694XX3,O694XX4,O694XX5,O694XX9 | |
| | O695XX0,O695XX1,O695XX2,O695XX3,O695XX4,O695XX5,O695XX9 | |
| | O6981X0,O6981X1,O6981X2,O6981X3,O6981X4,O6981X5,O6981X9 | |
| | O6982X0,O6982X1,O6982X2,O6982X3,O6982X4,O6982X5,O6982X9 | |
| | O6989X0,O6989X1,O6989X2,O6989X3,O6989X4,O6989X5,O6989X9 | |
| | O699XX0,O699XX1,O699XX2,O699XX3,O699XX4,O699XX5,O699XX9,O771 | |
| | O778,O779 | |
| Maltreatment abuse | O9A311,O9A312,O9A313,O9A319,O9A32,O9A33,O9A411,O9A412,O9A413 | 64890,64891,64893,64894,99550,99551,99552,99553,99554,99555 |
| | O9A419,O9A42,O9A43,O9A511,O9A512,O9A513,O9A519,O9A52,O9A53 | 99580,99581,99582,99583,99584,E9040 |
| | T7401XA,T7402XA,T7411XA,T7412XA,T7421XA,T7422XA,T7431XA | |
| | T7432XA,T744XXA,T7451XA,T7452XA,T7461XA,T7462XA,T7491XA | |
| | T7492XA,T7601XA,T7602XA,T7611XA,T7612XA,T7621XA,T7622XA | |
| | T7631XA,T7632XA,T7651XA,T7652XA,T7661XA,T7662XA,T7691XA | |
| | T7692XA | |
| Maltreatment abuse, | T7401XD,T7402XD,T7411XD,T7412XD,T7421XD,T7422XD,T7431XD | V5889 |
| subsequent encounter | T7432XD,T744XXD,T7451XD,T7452XD,T7461XD,T7462XD,T7491XD | |
| | T7492XD,T7601XD,T7602XD,T7611XD,T7612XD,T7621XD,T7622XD | |
| | T7631XD,T7632XD,T7651XD,T7652XD,T7661XD,T7662XD,T7691XD | |
| | T7692XD | |
| Maternal care for | O3400,O3401,O3402,O3403,O3410,O3411,O3412,O3413,O3429,O3440 | 65400,65401,65402,65403,65404,65410,65411,65412,65413,65414 |

Continued on next page

**Supplementary Table S1:** (cont'd) A comprehensive mapping of diagnosis groups used in the experiments to their corresponding ICD-10 and ICD-9 codes.

| Diagnosis | ICD-10 | ICD-9 |
|---|---|---|
| abnormality of pelvic<br>organs | O3441,O3442,O3443,O34511,O34512,O34513,O34519,O34521,O34522<br>O34523,O34529,O34531,O34532,O34533,O34539,O34591,O34592<br>O34593,O34599,O3460,O3461,O3462,O3463,O3470,O3471,O3472,O3473<br>O3480,O3481,O3482,O3483,O3490,O3491,O3492,O3493,O7100,O7102<br>O7103 | 65430,65431,65432,65433,65434,65440,65441,65442,65443,65444<br>65460,65461,65462,65463,65464,65470,65471,65472,65473,65474<br>65480,65481,65482,65483,65484,65490,65491,65492,65493,65494<br>66500,66501,66503 |
| Maternal care related<br>to disorders of the<br>placenta and<br>placental<br>implantation | O3670X0,O3670X1,O3670X2,O3670X3,O3670X4,O3670X5,O3670X9<br>O3671X0,O3671X1,O3671X2,O3671X3,O3671X4,O3671X5,O3671X9<br>O3672X0,O3672X1,O3672X2,O3672X3,O3672X4,O3672X5,O3672X9<br>O3673X0,O3673X1,O3673X2,O3673X3,O3673X4,O3673X5,O3673X9<br>O43011,O43012,O43013,O43019,O43021,O43022,O43023,O43029<br>O43101,O43102,O43103,O43109,O43111,O43112,O43113,O43119<br>O43121,O43122,O43123,O43129,O43191,O43192,O43193,O43199<br>O43211,O43212,O43213,O43219,O43221,O43222,O43223,O43229<br>O43231,O43232,O43233,O43239,O43811,O43812,O43813,O43819<br>O43891,O43892,O43893,O43899,O4390,O4391,O4392,O4393,O4400<br>O4401,O4402,O4403,O4410,O4411,O4412,O4413,O4420,O4421,O4422<br>O4423,O4430,O4431,O4432,O4433,O4440,O4441,O4442,O4443,O4450<br>O4451,O4452,O4453,O45001,O45002,O45003,O45009,O45011,O45012<br>O45013,O45019,O45021,O45022,O45023,O45029,O45091,O45092<br>O45093,O45099,O458X1,O458X2,O458X3,O458X9,O4590,O4591,O4592<br>O4593 | 64100,64101,64103,64110,64111,64113,64120,64121,64123,64130<br>64131,64133,65600,65601,65603,65670,65671,65673,65680,65681<br>65683,66380,66381,66700,66702,66704 |
| Maternal care related<br>to fetal conditions | O3500X0,O3500X1,O3500X2,O3500X3,O3500X4,O3500X5,O3500X9<br>O3501X0,O3501X1,O3501X2,O3501X3,O3501X4,O3501X5,O3501X9<br>O3502X0,O3502X1,O3502X2,O3502X3,O3502X4,O3502X5,O3502X9<br>O3503X0,O3503X1,O3503X2,O3503X3,O3503X4,O3503X5,O3503X9<br>O3504X0,O3504X1,O3504X2,O3504X3,O3504X4,O3504X5,O3504X9<br>O3505X0,O3505X1,O3505X2,O3505X3,O3505X4,O3505X5,O3505X9<br>O3506X0,O3506X1,O3506X2,O3506X3,O3506X4,O3506X5,O3506X9<br>O3507X0,O3507X1,O3507X2,O3507X3,O3507X4,O3507X5,O3507X9<br>O3508X0,O3508X1,O3508X2,O3508X3,O3508X4,O3508X5,O3508X9<br>O3509X0,O3509X1,O3509X2,O3509X3,O3509X4,O3509X5,O3509X9<br>O350XX0,O350XX1,O350XX2,O350XX3,O350XX4,O350XX5,O350XX9<br>O3510X0,O3510X1,O3510X2,O3510X3,O3510X4,O3510X5,O3510X9<br>O3511X0,O3511X1,O3511X2,O3511X3,O3511X4,O3511X5,O3511X9<br>O3512X0,O3512X1,O3512X2,O3512X3,O3512X4,O3512X5,O3512X9<br>O3513X0,O3513X1,O3513X2,O3513X3,O3513X4,O3513X5,O3513X9<br>O3514X0,O3514X1,O3514X2,O3514X3,O3514X4,O3514X5,O3514X9<br>O3515X0,O3515X1,O3515X2,O3515X3,O3515X4,O3515X5,O3515X9 | 65501,65503,65511,65513,65521,65523,65531,65533,65541,65543<br>65551,65553,65561,65563,65571,65573,65581,65583,65591,65593<br>65610,65611,65613,65620,65621,65623,65650,65651,65653,65660<br>65661,65663,65680,65681,65683,65690,65691,65693,67800,67801<br>67803,67910,67911,67912,67913,67914,V2387 |

Continued on next page

**Supplementary Table S1:** (cont'd) A comprehensive mapping of diagnosis groups used in the experiments to their corresponding ICD-10 and ICD-9 codes.

| Diagnosis | ICD-10 | ICD-9 |
|---|---|---|
| | O3519X0,O3519X1,O3519X2,O3519X3,O3519X4,O3519X5,O3519X9 | |
| | O351XX0,O351XX1,O351XX2,O351XX3,O351XX4,O351XX5,O351XX9 | |
| | O352XX0,O352XX1,O352XX2,O352XX3,O352XX4,O352XX5,O352XX9 | |
| | O353XX0,O353XX1,O353XX2,O353XX3,O353XX4,O353XX5,O353XX9 | |
| | O354XX0,O354XX1,O354XX2,O354XX3,O354XX4,O354XX5,O354XX9 | |
| | O355XX0,O355XX1,O355XX2,O355XX3,O355XX4,O355XX5,O355XX9 | |
| | O356XX0,O356XX1,O356XX2,O356XX3,O356XX4,O356XX5,O356XX9 | |
| | O357XX0,O357XX1,O357XX2,O357XX3,O357XX4,O357XX5,O357XX9 | |
| | O358XX0,O358XX1,O358XX2,O358XX3,O358XX4,O358XX5,O358XX9 | |
| | O359XX0,O359XX1,O359XX2,O359XX3,O359XX4,O359XX5,O359XX9 | |
| | O35AXX0,O35AXX1,O35AXX2,O35AXX3,O35AXX4,O35AXX5,O35AXX9 | |
| | O35BXX0,O35BXX1,O35BXX2,O35BXX3,O35BXX4,O35BXX5,O35BXX9 | |
| | O35CXX0,O35CXX1,O35CXX2,O35CXX3,O35CXX4,O35CXX5,O35CXX9 | |
| | O35DXX0,O35DXX1,O35DXX2,O35DXX3,O35DXX4,O35DXX5,O35DXX9 | |
| | O35EXX0,O35EXX1,O35EXX2,O35EXX3,O35EXX4,O35EXX5,O35EXX9 | |
| | O35FXX0,O35FXX1,O35FXX2,O35FXX3,O35FXX4,O35FXX5,O35FXX9 | |
| | O35GXX0,O35GXX1,O35GXX2,O35GXX3,O35GXX4,O35GXX5,O35GXX9 | |
| | O35HXX0,O35HXX1,O35HXX2,O35HXX3,O35HXX4,O35HXX5,O35HXX9 | |
| | O360110,O360111,O360112,O360113,O360114,O360115,O360119 | |
| | O360120,O360121,O360122,O360123,O360124,O360125,O360129 | |
| | O360130,O360131,O360132,O360133,O360134,O360135,O360139 | |
| | O360190,O360191,O360192,O360193,O360194,O360195,O360199 | |
| | O360910,O360911,O360912,O360913,O360914,O360915,O360919 | |
| | O360920,O360921,O360922,O360923,O360924,O360925,O360929 | |
| | O360930,O360931,O360932,O360933,O360934,O360935,O360939 | |
| | O360990,O360991,O360992,O360993,O360994,O360995,O360999 | |
| | O361110,O361111,O361112,O361113,O361114,O361115,O361119 | |
| | O361120,O361121,O361122,O361123,O361124,O361125,O361129 | |
| | O361130,O361131,O361132,O361133,O361134,O361135,O361139 | |
| | O361190,O361191,O361192,O361193,O361194,O361195,O361199 | |
| | O361910,O361911,O361912,O361913,O361914,O361915,O361919 | |
| | O361920,O361921,O361922,O361923,O361924,O361925,O361929 | |
| | O361930,O361931,O361932,O361933,O361934,O361935,O361939 | |
| | O361990,O361991,O361992,O361993,O361994,O361995,O361999 | |
| | O3620X0,O3620X1,O3620X2,O3620X3,O3620X4,O3620X5,O3620X9 | |
| | O3621X0,O3621X1,O3621X2,O3621X3,O3621X4,O3621X5,O3621X9 | |
| | O3622X0,O3622X1,O3622X2,O3622X3,O3622X4,O3622X5,O3622X9 | |
| | O3623X0,O3623X1,O3623X2,O3623X3,O3623X4,O3623X5,O3623X9 | |
| | O365110,O365111,O365112,O365113,O365114,O365115,O365119 | |
| | O365120,O365121,O365122,O365123,O365124,O365125,O365129 | |

Continued on next page

**Supplementary Table S1:** (cont'd) A comprehensive mapping of diagnosis groups used in the experiments to their corresponding ICD-10 and ICD-9 codes.

| Diagnosis | ICD-10 | ICD-9 |
|---|---|---|
| | O365130,O365131,O365132,O365133,O365134,O365135,O365139 | |
| | O365190,O365191,O365192,O365193,O365194,O365195,O365199 | |
| | O365910,O365911,O365912,O365913,O365914,O365915,O365919 | |
| | O365920,O365921,O365922,O365923,O365924,O365925,O365929 | |
| | O365930,O365931,O365932,O365933,O365934,O365935,O365939 | |
| | O365990,O365991,O365992,O365993,O365994,O365995,O365999 | |
| | O3660X0,O3660X1,O3660X2,O3660X3,O3660X4,O3660X5,O3660X9 | |
| | O3661X0,O3661X1,O3661X2,O3661X3,O3661X4,O3661X5,O3661X9 | |
| | O3662X0,O3662X1,O3662X2,O3662X3,O3662X4,O3662X5,O3662X9 | |
| | O3663X0,O3663X1,O3663X2,O3663X3,O3663X4,O3663X5,O3663X9 | |
| | O3680X0,O3680X1,O3680X2,O3680X3,O3680X4,O3680X5,O3680X9 | |
| | O368120,O368121,O368122,O368123,O368124,O368125,O368129 | |
| | O368130,O368131,O368132,O368133,O368134,O368135,O368139 | |
| | O368190,O368191,O368192,O368193,O368194,O368195,O368199 | |
| | O368210,O368211,O368212,O368213,O368214,O368215,O368219 | |
| | O368220,O368221,O368222,O368223,O368224,O368225,O368229 | |
| | O368230,O368231,O368232,O368233,O368234,O368235,O368239 | |
| | O368290,O368291,O368292,O368293,O368294,O368295,O368299 | |
| | O368310,O368311,O368312,O368313,O368314,O368315,O368319 | |
| | O368320,O368321,O368322,O368323,O368324,O368325,O368329 | |
| | O368330,O368331,O368332,O368333,O368334,O368335,O368339 | |
| | O368390,O368391,O368392,O368393,O368394,O368395,O368399 | |
| | O368910,O368911,O368912,O368913,O368914,O368915,O368919 | |
| | O368920,O368921,O368922,O368923,O368924,O368925,O368929 | |
| | O368930,O368931,O368932,O368933,O368934,O368935,O368939 | |
| | O368990,O368991,O368992,O368993,O368994,O368995,O368999 | |
| | O3690X0,O3690X1,O3690X2,O3690X3,O3690X4,O3690X5,O3690X9 | |
| | O3691X0,O3691X1,O3691X2,O3691X3,O3691X4,O3691X5,O3691X9 | |
| | O3692X0,O3692X1,O3692X2,O3692X3,O3692X4,O3692X5,O3692X9 | |
| | O3693X0,O3693X1,O3693X2,O3693X3,O3693X4,O3693X5,O3693X9 | |
| Maternal intrauterine infection | O411010,O411011,O411012,O411013,O411014,O411015,O411019 | 65840,65841,65843 |
| | O411020,O411021,O411022,O411023,O411024,O411025,O411029 | |
| | O411030,O411031,O411032,O411033,O411034,O411035,O411039 | |
| | O411090,O411091,O411092,O411093,O411094,O411095,O411099 | |
| | O411210,O411211,O411212,O411213,O411214,O411215,O411219 | |
| | O411220,O411221,O411222,O411223,O411224,O411225,O411229 | |
| | O411230,O411231,O411232,O411233,O411234,O411235,O411239 | |
| | O411290,O411291,O411292,O411293,O411294,O411295,O411299 | |
| | O411410,O411411,O411412,O411413,O411414,O411415,O411419 | |

Continued on next page

**Supplementary Table S1:** (cont'd) A comprehensive mapping of diagnosis groups used in the experiments to their corresponding ICD-10 and ICD-9 codes.

| Diagnosis | ICD-10 | ICD-9 |
|---|---|---|
| | O411420,O411421,O411422,O411423,O411424,O411425,O411429<br>O411430,O411431,O411432,O411433,O411434,O411435,O411439<br>O411490,O411491,O411492,O411493,O411494,O411495,O411499 | |
| Mediastinal disorders | J853,J985,J9851,J9859 | 5131,5192,5193 |
| Medical examination evaluation | Z0000,Z0001,Z00110,Z00111,Z00121,Z00129,Z002,Z003,Z005,Z006<br>Z0070,Z0071,Z008,Z0100,Z0101,Z01020,Z01021,Z0110,Z01110<br>Z01118,Z0112,Z0120,Z0121,Z0130,Z0131,Z01411,Z01419,Z0142<br>Z01810,Z01811,Z01812,Z01818,Z0182,Z0183,Z0184,Z0189 | V202,V2031,V2032,V210,V211,V212,V700,V706,V707,V708,V720<br>V7211,V7212,V7219,V722,V7231,V7232,V7261,V7263,V7281,V7282<br>V7283,V7284,V7285,V7286 |
| Meningitis | A0101,A0221,A170,A171,A203,A2781,A3211,A390,A4281,A5041,A5141<br>A5213,A5481,A6921,A870,A871,A872,A878,A879,B003,B010,B021<br>B051,B0602,B261,B2702,B2712,B2782,B2792,B375,B384,B4281,B451<br>B5741,D8681,G000,G001,G002,G003,G008,G009,G01,G02,G030,G031<br>G032,G038,G039 | 0020,00321,01300,01310,0208,0270,0360,0398,0470,0471,0478<br>0479,0490,0491,0527,0530,05472,05579,05609,0721,075,0861<br>08881,09042,09181,0942,09882,10081,11283,1142,1171,1175,135<br>3200,3201,3202,3203,3207,32082,32089,3209,3210,3211,3212,3218<br>3220,3221,3222,3229,99709 |
| Menopausal disorders | E28310,E28319,E2839,N924,N950,N951,N952,N958,N959 | 25631,25639,6270,6271,6272,6273,6278,6279 |
| Menstrual disorders | N897,N910,N911,N912,N913,N914,N915,N920,N921,N922,N923,N925<br>N926,N944,N945,N946 | 6253,6260,6261,6262,6263,6264,6265,6266,6268,6269 |
| Mental and substance use disorders in remission | F1011,F1021,F1111,F1121,F1211,F1221,F1311,F1321,F1411,F1421<br>F1511,F1521,F1611,F1621,F17201,F17211,F17221,F17291,F1811<br>F1821,F1911,F1921,F304,F3170,F3172,F3174,F3176,F3178,F325<br>F3340,F3342 | 29285,29606,29626,29630,29636,29646,29656,29666,2967,30393<br>30403,30413,30423,30433,30443,30453,30463,3051 |
| Mental and substance use disorders; sequela | T1491XS,T360X2S,T361X2S,T362X2S,T363X2S,T364X2S,T365X2S<br>T366X2S,T367X2S,T368X2S,T3692XS,T370X2S,T371X2S,T372X2S<br>T373X2S,T374X2S,T375X2S,T378X2S,T3792XS,T380X2S,T381X2S<br>T382X2S,T383X2S,T384X2S,T385X2S,T386X2S,T387X2S,T38802S<br>T38812S,T38892S,T38902S,T38992S,T39012S,T39092S,T391X2S<br>T392X2S,T39312S,T39392S,T394X2S,T398X2S,T3992XS,T400X1S<br>T400X2S,T400X3S,T400X4S,T400X5S,T401X1S,T401X2S,T401X3S<br>T401X4S,T402X1S,T402X2S,T402X3S,T402X4S,T402X5S,T403X1S<br>T403X2S,T403X3S,T403X4S,T403X5S,T40411S,T40412S,T40413S<br>T40414S,T40415S,T40421S,T40422S,T40423S,T40424S,T40425S<br>T40491S,T40492S,T40493S,T40494S,T40495S,T404X1S,T404X2S<br>T404X3S,T404X4S,T404X5S,T405X1S,T405X2S,T405X3S,T405X4S<br>T405X5S,T40601S,T40602S,T40603S,T40604S,T40605S,T40691S | 9090,9091,9094,9095,E9292,E9351,E9352,E9363,E9378,E9379,E9382<br>E9390,E9396,E9397,E959,E969,E989 |

Continued on next page

**Supplementary Table S1:** (cont'd) A comprehensive mapping of diagnosis groups used in the experiments to their corresponding ICD-10 and ICD-9 codes.

| Diagnosis | ICD-10 | ICD-9 |
|---|---|---|
| | T40692S,T40693S,T40694S,T40695S,T40711S,T40712S,T40713S | |
| | T40714S,T40715S,T40721S,T40722S,T40723S,T40724S,T40725S | |
| | T407X1S,T407X2S,T407X3S,T407X4S,T407X5S,T408X1S,T408X2S | |
| | T408X3S,T408X4S,T40901S,T40902S,T40903S,T40904S,T40905S | |
| | T40991S,T40992S,T40993S,T40994S,T40995S,T410X1S,T410X2S | |
| | T410X3S,T410X4S,T410X5S,T411X2S,T41202S,T41292S,T413X2S | |
| | T4142XS,T415X2S,T420X2S,T422X2S,T423X2S,T424X2S,T425X2S | |
| | T426X1S,T426X2S,T426X3S,T426X4S,T426X5S,T4271XS,T4272XS | |
| | T4273XS,T4274XS,T4275XS,T428X2S,T43012S,T43022S,T431X2S | |
| | T43202S,T43205S,T43212S,T43222S,T43292S,T433X2S,T434X2S | |
| | T43502S,T43592S,T43601S,T43602S,T43603S,T43604S,T43605S | |
| | T43611S,T43612S,T43613S,T43614S,T43615S,T43621S,T43622S | |
| | T43623S,T43624S,T43625S,T43631S,T43632S,T43633S,T43634S | |
| | T43635S,T43641S,T43642S,T43643S,T43644S,T43651S,T43652S | |
| | T43653S,T43654S,T43655S,T43691S,T43692S,T43693S,T43694S | |
| | T43695S,T438X2S,T4392XS,T440X2S,T441X2S,T442X2S,T443X2S | |
| | T444X2S,T445X2S,T446X2S,T447X2S,T448X2S,T44902S,T44992S | |
| | T450X2S,T451X2S,T452X2S,T453X2S,T454X2S,T45512S,T45522S | |
| | T45602S,T45612S,T45622S,T45692S,T457X2S,T458X2S,T4592XS | |
| | T460X2S,T461X2S,T462X2S,T463X2S,T464X2S,T465X2S,T466X2S | |
| | T467X2S,T468X2S,T46902S,T46992S,T470X2S,T471X2S,T472X2S | |
| | T473X2S,T474X2S,T475X2S,T476X2S,T477X2S,T478X2S,T4792XS | |
| | T480X2S,T481X2S,T48202S,T48292S,T483X2S,T484X2S,T485X2S | |
| | T486X2S,T48902S,T48992S,T490X2S,T491X2S,T492X2S,T493X2S | |
| | T494X2S,T495X2S,T496X2S,T497X2S,T498X2S,T4992XS,T500X2S | |
| | T501X2S,T502X2S,T503X2S,T504X2S,T505X2S,T506X2S,T507X2S | |
| | T508X2S,T50902S,T50912S,T50992S,T50A12S,T50A22S,T50A92S | |
| | T50B12S,T50B92S,T50Z12S,T50Z92S,T510X2S,T511X2S,T512X2S | |
| | T513X2S,T518X2S,T5192XS,T520X2S,T521X2S,T522X2S,T523X2S | |
| | T524X2S,T528X2S,T5292XS,T530X2S,T531X2S,T532X2S,T533X2S | |
| | T534X2S,T535X2S,T536X2S,T537X2S,T5392XS,T540X2S,T541X2S | |
| | T542X2S,T543X2S,T5492XS,T550X2S,T551X2S,T560X2S,T561X2S | |
| | T562X2S,T563X2S,T564X2S,T565X2S,T566X2S,T567X2S,T56812S | |
| | T56892S,T5692XS,T570X2S,T571X2S,T572X2S,T573X2S,T578X2S | |
| | T5792XS,T5802XS,T5812XS,T582X2S,T588X2S,T5892XS,T590X2S | |
| | T591X2S,T592X2S,T593X2S,T594X2S,T595X2S,T596X2S,T597X2S | |
| | T59812S,T59892S,T5992XS,T600X2S,T601X2S,T602X2S,T603X2S | |
| | T604X2S,T608X2S,T6092XS,T6102XS,T6112XS,T61772S,T61782S | |
| | T618X2S,T6192XS,T620X2S,T621X2S,T622X2S,T628X2S,T6292XS | |
| | T63002S,T63012S,T63022S,T63032S,T63042S,T63062S,T63072S | |

Continued on next page

**Supplementary Table S1:** (cont'd) A comprehensive mapping of diagnosis groups used in the experiments to their corresponding ICD-10 and ICD-9 codes.

| Diagnosis | ICD-10 | ICD-9 |
|---|---|---|
| | T63082S,T63092S,T63112S,T63122S,T63192S,T632X2S,T63302S T63312S,T63322S,T63332S,T63392S,T63412S,T63422S,T63432S T63442S,T63452S,T63462S,T63482S,T63512S,T63592S,T63612S T63622S,T63632S,T63692S,T63712S,T63792S,T63812S,T63822S T63832S,T63892S,T6392XS,T6402XS,T6482XS,T650X2S,T651X2S T65212S,T65222S,T65292S,T653X2S,T654X2S,T655X2S,T656X2S T65812S,T65822S,T65832S,T65892S,T6592XS,T71112S,T71122S T71132S,T71152S,T71162S,T71192S,T71222S,T71232S,X710XXS X711XXS,X712XXS,X713XXS,X718XXS,X719XXS,X72XXXS,X730XXS X731XXS,X732XXS,X738XXS,X739XXS,X7401XS,X7402XS,X7409XS X748XXS,X749XXS,X75XXXS,X76XXXS,X770XXS,X771XXS,X772XXS X773XXS,X778XXS,X779XXS,X780XXS,X781XXS,X782XXS,X788XXS X789XXS,X79XXXS,X80XXXS,X810XXS,X811XXS,X818XXS,X828XXS X830XXS,X831XXS,X832XXS,X838XXS | |
| Mesothelioma | C450,C451,C452,C457,C459 | 1588,1639,1641,1958,1991 |
| Miscellaneous mental and behavioral disorders conditions | F068,F079,F09,F488,F489,F520,F521,F5221,F5222,F5231,F5232 F524,F525,F526,F528,F529,F54,F550,F551,F552,F553,F554,F558 F59,F630,F640,F641,F642,F648,F649,F650,F651,F652,F653,F654 F6550,F6551,F6552,F6581,F6589,F659,F66,F938,F939,F989,F99 O99340,O99341,O99342,O99343,O99344,O99345,R37 | 2948,3005,30089,3009,3021,3022,3023,3024,30250,3026,30270 30271,30272,30273,30274,30275,30276,30279,30281,30282,30283 30284,30285,30289,3029,30590,30591,30592,30593,30651,3068 3109,31231,3130,3133,31382,31389,3139,316,64840,64841,64842 64843,64844,V402,V409,V417 |
| Molar pregnancy and other abnormal products of conception | O010,O011,O019,O020,O021,O0281,O0289,O029 | 630,6310,6318,632 |
| Multiple gestation | O30001,O30002,O30003,O30009,O30011,O30012,O30013,O30019 O30021,O30022,O30023,O30029,O30031,O30032,O30033,O30039 O30041,O30042,O30043,O30049,O30091,O30092,O30093,O30099 O30101,O30102,O30103,O30109,O30111,O30112,O30113,O30119 O30121,O30122,O30123,O30129,O30131,O30132,O30133,O30139 O30191,O30192,O30193,O30199,O30201,O30202,O30203,O30209 O30211,O30212,O30213,O30219,O30221,O30222,O30223,O30229 O30231,O30232,O30233,O30239,O30291,O30292,O30293,O30299 O30801,O30802,O30803,O30809,O30811,O30812,O30813,O30819 O30821,O30822,O30823,O30829,O30831,O30832,O30833,O30839 O30891,O30892,O30893,O30899,O3090,O3091,O3092,O3093,O3100X0 O3100X1,O3100X2,O3100X3,O3100X4,O3100X5,O3100X9,O3101X0 | 64600,64601,64603,65100,65101,65103,65110,65111,65113,65120 65121,65123,65130,65131,65133,65160,65161,65163,65170,65171 65173,65180,65181,65183,65190,65191,65193,67810,67811,67813 V9100,V9101,V9102,V9103,V9109,V9110,V9111,V9112,V9119,V9120 V9121,V9122,V9129,V9190,V9191,V9192,V9199 |

Continued on next page

**Supplementary Table S1:** (cont'd) A comprehensive mapping of diagnosis groups used in the experiments to their corresponding ICD-10 and ICD-9 codes.

| Diagnosis | ICD-10 | ICD-9 |
|---|---|---|
| | O3101X1,O3101X2,O3101X3,O3101X4,O3101X5,O3101X9,O3102X0 | |
| | O3102X1,O3102X2,O3102X3,O3102X4,O3102X5,O3102X9,O3103X0 | |
| | O3103X1,O3103X2,O3103X3,O3103X4,O3103X5,O3103X9,O3110X0 | |
| | O3110X1,O3110X2,O3110X3,O3110X4,O3110X5,O3110X9,O3111X0 | |
| | O3111X1,O3111X2,O3111X3,O3111X4,O3111X5,O3111X9,O3112X0 | |
| | O3112X1,O3112X2,O3112X3,O3112X4,O3112X5,O3112X9,O3113X0 | |
| | O3113X1,O3113X2,O3113X3,O3113X4,O3113X5,O3113X9,O3120X0 | |
| | O3120X1,O3120X2,O3120X3,O3120X4,O3120X5,O3120X9,O3121X0 | |
| | O3121X1,O3121X2,O3121X3,O3121X4,O3121X5,O3121X9,O3122X0 | |
| | O3122X1,O3122X2,O3122X3,O3122X4,O3122X5,O3122X9,O3123X0 | |
| | O3123X1,O3123X2,O3123X3,O3123X4,O3123X5,O3123X9,O3130X0 | |
| | O3130X1,O3130X2,O3130X3,O3130X4,O3130X5,O3130X9,O3131X0 | |
| | O3131X1,O3131X2,O3131X3,O3131X4,O3131X5,O3131X9,O3132X0 | |
| | O3132X1,O3132X2,O3132X3,O3132X4,O3132X5,O3132X9,O3133X0 | |
| | O3133X1,O3133X2,O3133X3,O3133X4,O3133X5,O3133X9,O318X10 | |
| | O318X11,O318X12,O318X13,O318X14,O318X15,O318X19,O318X20 | |
| | O318X21,O318X22,O318X23,O318X24,O318X25,O318X29,O318X30 | |
| | O318X31,O318X32,O318X33,O318X34,O318X35,O318X39,O318X90 | |
| | O318X91,O318X92,O318X93,O318X94,O318X95,O318X99 | |
| Multiple myeloma | C9000,C9001,C9002,C9010,C9011,C9012,C9020,C9021,C9022,C9030 | 20300,20301,20302,20310,20311,20312,20380,20381,20382 |
| | C9031,C9032 | |
| Multiple sclerosis | G35 | 340 |
| Muscle disorders | B5882,B6981,K6812,M60000,M60001,M60002,M60003,M60004,M60005 | 04081,1231,1307,56731,7280,72810,72811,72812,72819,7282,7283 |
| | M60009,M60011,M60012,M60019,M60021,M60022,M60029,M60031 | 72881,72883,72884,72885,72887,72888,72889,7289,7291 |
| | M60032,M60039,M60041,M60042,M60043,M60044,M60045,M60046 | |
| | M60051,M60052,M60059,M60061,M60062,M60069,M60070,M60071 | |
| | M60072,M60073,M60074,M60075,M60076,M60077,M60078,M6008,M6009 | |
| | M6010,M60111,M60112,M60119,M60121,M60122,M60129,M60131,M60132 | |
| | M60139,M60141,M60142,M60149,M60151,M60152,M60159,M60161 | |
| | M60162,M60169,M60171,M60172,M60179,M6018,M6019,M6080,M60811 | |
| | M60812,M60819,M60821,M60822,M60829,M60831,M60832,M60839 | |
| | M60841,M60842,M60849,M60851,M60852,M60859,M60861,M60862 | |
| | M60869,M60871,M60872,M60879,M6088,M6089,M609,M6100,M61011 | |
| | M61012,M61019,M61021,M61022,M61029,M61031,M61032,M61039 | |
| | M61041,M61042,M61049,M61051,M61052,M61059,M61061,M61062 | |
| | M61069,M61071,M61072,M61079,M6108,M6109,M6110,M61111,M61112 | |
| | M61119,M61121,M61122,M61129,M61131,M61132,M61139,M61141 | |

Continued on next page

**Supplementary Table S1:** (cont'd) A comprehensive mapping of diagnosis groups used in the experiments to their corresponding ICD-10 and ICD-9 codes.

| Diagnosis | ICD-10 | ICD-9 |
|---|---|---|
| | M61142,M61143,M61144,M61145,M61146,M61151,M61152,M61159 | |
| | M61161,M61162,M61169,M61171,M61172,M61173,M61174,M61175 | |
| | M61176,M61177,M61178,M61179,M6118,M6119,M6120,M61211,M61212 | |
| | M61219,M61221,M61222,M61229,M61231,M61232,M61239,M61241 | |
| | M61242,M61249,M61251,M61252,M61259,M61261,M61262,M61269 | |
| | M61271,M61272,M61279,M6128,M6129,M6130,M61311,M61312,M61319 | |
| | M61321,M61322,M61329,M61331,M61332,M61339,M61341,M61342 | |
| | M61349,M61351,M61352,M61359,M61361,M61362,M61369,M61371 | |
| | M61372,M61379,M6138,M6139,M6140,M61411,M61412,M61419,M61421 | |
| | M61422,M61429,M61431,M61432,M61439,M61441,M61442,M61449 | |
| | M61451,M61452,M61459,M61461,M61462,M61469,M61471,M61472 | |
| | M61479,M6148,M6149,M6150,M61511,M61512,M61519,M61521,M61522 | |
| | M61529,M61531,M61532,M61539,M61541,M61542,M61549,M61551 | |
| | M61552,M61559,M61561,M61562,M61569,M61571,M61572,M61579,M6158 | |
| | M6159,M619,M6200,M62011,M62012,M62019,M62021,M62022,M62029 | |
| | M62031,M62032,M62039,M62041,M62042,M62049,M62051,M62052 | |
| | M62059,M62061,M62062,M62069,M62071,M62072,M62079,M6208,M6210 | |
| | M62111,M62112,M62119,M62121,M62122,M62129,M62131,M62132 | |
| | M62139,M62141,M62142,M62149,M62151,M62152,M62159,M62161 | |
| | M62162,M62169,M62171,M62172,M62179,M6218,M6220,M62211,M62212 | |
| | M62219,M62221,M62222,M62229,M62231,M62232,M62239,M62241 | |
| | M62242,M62249,M62251,M62252,M62259,M62261,M62262,M62269 | |
| | M62271,M62272,M62279,M6228,M623,M6240,M62411,M62412,M62419 | |
| | M62421,M62422,M62429,M62431,M62432,M62439,M62441,M62442 | |
| | M62449,M62451,M62452,M62459,M62461,M62462,M62469,M62471 | |
| | M62472,M62479,M6248,M6249,M6250,M62511,M62512,M62519,M62521 | |
| | M62522,M62529,M62531,M62532,M62539,M62541,M62542,M62549 | |
| | M62551,M62552,M62559,M62561,M62562,M62569,M62571,M62572 | |
| | M62579,M6258,M6259,M625A0,M625A1,M625A2,M625A9,M6281,M6282 | |
| | M62831,M62838,M6284,M6289,M629,M6380,M63811,M63812,M63819 | |
| | M63821,M63822,M63829,M63831,M63832,M63839,M63841,M63842 | |
| | M63849,M63851,M63852,M63859,M63861,M63862,M63869,M63871 | |
| | M63872,M63879,M6388,M6389 | |
| Musculoskeletal | M6500,M65011,M65012,M65019,M65021,M65022,M65029,M65031,M65032 | 72789 |
| abscess | M65039,M65041,M65042,M65049,M65051,M65052,M65059,M65061 | |
| | M65062,M65069,M65071,M65072,M65079,M6508,M7100,M71011,M71012 | |
| | M71019,M71021,M71022,M71029,M71031,M71032,M71039,M71041 | |
| | M71042,M71049,M71051,M71052,M71059,M71061,M71062,M71069 | |
| | M71071,M71072,M71079,M7108,M7109 | |

Continued on next page

**Supplementary Table S1:** (cont'd) A comprehensive mapping of diagnosis groups used in the experiments to their corresponding ICD-10 and ICD-9 codes.

| Diagnosis | ICD-10 | ICD-9 |
|---|---|---|
| Musculoskeletal<br>congenital conditions | Q6500,Q6501,Q6502,Q651,Q652,Q6530,Q6531,Q6532,Q654,Q655,Q656<br>Q6581,Q6582,Q6589,Q659,Q660,Q6600,Q6601,Q6602,Q661,Q6610<br>Q6611,Q6612,Q662,Q6621,Q66211,Q66212,Q66219,Q6622,Q66221<br>Q66222,Q66229,Q663,Q6630,Q6631,Q6632,Q664,Q6640,Q6641,Q6642<br>Q6650,Q6651,Q6652,Q666,Q667,Q6670,Q6671,Q6672,Q6680,Q6681<br>Q6682,Q6689,Q669,Q6690,Q6691,Q6692,Q670,Q671,Q672,Q673,Q674<br>Q675,Q676,Q677,Q678,Q680,Q681,Q682,Q683,Q684,Q685,Q686,Q688<br>Q690,Q691,Q692,Q699,Q7000,Q7001,Q7002,Q7003,Q7010,Q7011,Q7012<br>Q7013,Q7020,Q7021,Q7022,Q7023,Q7030,Q7031,Q7032,Q7033,Q704<br>Q709,Q7100,Q7101,Q7102,Q7103,Q7110,Q7111,Q7112,Q7113,Q7120<br>Q7121,Q7122,Q7123,Q7130,Q7131,Q7132,Q7133,Q7140,Q7141,Q7142<br>Q7143,Q7150,Q7151,Q7152,Q7153,Q7160,Q7161,Q7162,Q7163,Q71811<br>Q71812,Q71813,Q71819,Q71891,Q71892,Q71893,Q71899,Q7190,Q7191<br>Q7192,Q7193,Q7200,Q7201,Q7202,Q7203,Q7210,Q7211,Q7212,Q7213<br>Q7220,Q7221,Q7222,Q7223,Q7230,Q7231,Q7232,Q7233,Q7240,Q7241<br>Q7242,Q7243,Q7250,Q7251,Q7252,Q7253,Q7260,Q7261,Q7262,Q7263<br>Q7270,Q7271,Q7272,Q7273,Q72811,Q72812,Q72813,Q72819,Q72891<br>Q72892,Q72893,Q72899,Q7290,Q7291,Q7292,Q7293,Q730,Q731,Q738<br>Q740,Q741,Q742,Q743,Q748,Q749,Q750,Q751,Q752,Q753,Q754,Q755<br>Q758,Q759,Q760,Q761,Q762,Q763,Q76411,Q76412,Q76413,Q76414<br>Q76415,Q76419,Q76425,Q76426,Q76427,Q76428,Q76429,Q7649,Q765<br>Q766,Q767,Q768,Q769,Q770,Q771,Q772,Q773,Q774,Q775,Q776,Q777<br>Q778,Q779,Q780,Q781,Q782,Q783,Q784,Q785,Q786,Q788,Q789,Q790<br>Q791,Q792,Q793,Q794,Q7951,Q7959,Q796,Q7960,Q7961,Q7962,Q7963<br>Q7969,Q798,Q799 | 7175,7540,7541,7542,75430,75431,75432,75433,75440,75441,75442<br>75443,75444,75451,75452,75453,75459,75461,75462,75469,75470<br>75471,75479,75481,75482,75489,75500,75501,75502,75510,75511<br>75512,75513,75514,75520,75521,75523,75525,75526,75527,75528<br>75529,75530,75531,75532,75533,75534,75535,75536,75537,75538<br>75539,7554,75550,75551,75552,75553,75554,75556,75557,75558<br>75559,75561,75562,75563,75564,75565,75566,75567,75569,7558<br>7559,7560,75612,75613,75614,75615,75616,75617,75619,7562,7563<br>7564,75650,75651,75652,75653,75654,75655,75656,75659,7566<br>75670,75671,75672,75673,75679,75681,75682,75683,75689,7569 |
| Musculoskeletal pain,<br>not low back pain | M2550,M25511,M25512,M25519,M25521,M25522,M25529,M25531,M25532<br>M25539,M25541,M25542,M25549,M25551,M25552,M25559,M25561<br>M25562,M25569,M25571,M25572,M25579,M2559,M2560,M25611,M25612<br>M25619,M25621,M25622,M25629,M25631,M25632,M25639,M25641<br>M25642,M25649,M25651,M25652,M25659,M25661,M25662,M25669<br>M25671,M25672,M25673,M25674,M25675,M25676,M2569,M546,M5481<br>M5489,M549,M791,M7910,M7911,M7912,M7918,M79601,M79602,M79603<br>M79604,M79605,M79606,M79609,M79621,M79622,M79629,M79631<br>M79632,M79639,M79641,M79642,M79643,M79644,M79645,M79646<br>M79651,M79652,M79659,M79661,M79662,M79669,M79671,M79672<br>M79673,M79674,M79675,M79676 | 71940,71941,71942,71943,71944,71945,71946,71947,71950,71951<br>71952,71953,71954,71955,71956,71957,7238,7241,7245,7291,7295 |
| Myelodysplastic | D460,D461,D4620,D4621,D4622,D464,D469,D46A,D46B,D46C,D46Z | 23872,23873,23874,23875 |

Continued on next page

**Supplementary Table S1:** (cont'd) A comprehensive mapping of diagnosis groups used in the experiments to their corresponding ICD-10 and ICD-9 codes.

| Diagnosis | ICD-10 | ICD-9 |
|---|---|---|
| syndrome (MDS) | | |
| Myocarditis and<br>cardiomyopathy | A3681,A381,A3950,A3952,B2682,B3320,B3322,B3324,B5881,D8685<br>I012,I090,I255,I400,I401,I408,I409,I41,I420,I421,I422,I423<br>I424,I425,I426,I427,I428,I429,I43,I514,J1082,J1182,O903 | 03282,0341,03640,03643,07279,07420,07423,07889,1303,135,3912<br>3980,4148,4220,42290,42291,42292,42293,42299,4250,42511,42518<br>4252,4253,4254,4255,4258,4259,4290,4878,67452,67454 |
| Myopathies | G7000,G7001,G701,G702,G7080,G7081,G7089,G709,G710,G7100,G7101<br>G7102,G71031,G71032,G71033,G710340,G710341,G710342,G710349<br>G71035,G71038,G71039,G7109,G7111,G7112,G7113,G7114,G7119,G712<br>G7120,G7121,G71220,G71228,G7129,G713,G718,G719,G720,G721,G722<br>G723,G7241,G7249,G7281,G7289,G729,G731,G733,G737,M0540,M05411<br>M05412,M05419,M05421,M05422,M05429,M05431,M05432,M05439<br>M05441,M05442,M05449,M05451,M05452,M05459,M05461,M05462<br>M05469,M05471,M05472,M05479,M0549,M3503 | 35800,35801,3581,3582,35830,35831,35839,3588,3589,3590,3591<br>35921,35922,35923,35924,35929,3593,3594,3595,3596,35971,35979<br>35981,35989,3599,7102,7140 |
| Nausea and vomiting | G43A0,G43A1,K910,O212,O218,O219,R110,R1110,R1111,R1112,R1113<br>R1114,R1115,R112 | 07882,34620,34621,5362,5643,56987,64323,64383,64393,78701<br>78702,78703,78704 |
| Neonatal abstinence<br>syndrome | P0414,P0415,P0416,P0417,P041A,P042,P0440,P0441,P0442,P0449<br>P0481,P961,P962 | 76072,76073,76075,76079,7795 |
| Neonatal acidemia and<br>hypoxia | P190,P191,P192,P199,P84 | 7682,7683,7684,7689,77088,77581 |
| Neonatal cerebral<br>disorders | P90,P910,P911,P912,P913,P914,P915,P9160,P9161,P9162,P9163<br>P918,P91811,P91819,P91821,P91822,P91823,P91829,P9188,P919 | 76870,76871,76872,76873,7790,7791,7792,7797 |
| Neonatal digestive<br>and feeding disorders | P760,P761,P762,P768,P769,P771,P772,P773,P779,P780,P781,P782<br>P783,P7881,P7882,P7883,P7884,P7889,P789,P9201,P9209,P921,P922<br>P923,P924,P925,P926,P928,P929 | 7771,7772,7773,7774,77750,77751,77752,77753,7776,7778,7779<br>77931,77932,77933,77934 |
| Neoplasm-related<br>encounters | Z08,Z120,Z1210,Z1211,Z1212,Z1213,Z122,Z1231,Z1239,Z124,Z125<br>Z126,Z1271,Z1272,Z1273,Z1279,Z1281,Z1282,Z1283,Z1289,Z129<br>Z483 | V5842,V6701,V671,V672,V6759,V676,V679,V760,V7610,V7612,V7619<br>V762,V763,V7641,V7642,V7643,V7644,V7645,V7646,V7647,V7649<br>V7650,V7651,V7652,V7681,V769 |
| Neoplasms of<br>unspecified nature or<br>uncertain behavior | D3701,D3702,D37030,D37031,D37032,D37039,D3704,D3705,D3709<br>D371,D372,D373,D374,D375,D376,D378,D379,D380,D381,D382,D383<br>D384,D385,D386,D390,D3910,D3911,D3912,D392,D398,D399,D400<br>D4010,D4011,D4012,D408,D409,D4100,D4101,D4102,D4110,D4111<br>D4112,D4120,D4121,D4122,D413,D414,D418,D419,D420,D421,D429 | 20710,20711,20712,2350,2351,2352,2353,2354,2355,2356,2357<br>2358,2359,2360,2361,2362,2363,2364,2365,2366,2367,23690,23691<br>23699,2370,2371,2372,2373,2374,2375,2376,2379,2380,2381,2382<br>2383,2384,2385,23871,23876,23877,23879,2388,2389,2390,2391<br>2392,2393,2394,2395,2396,2397,23981,23989,2399,2731,28989 |

Continued on next page

**Supplementary Table S1:** (cont'd) A comprehensive mapping of diagnosis groups used in the experiments to their corresponding ICD-10 and ICD-9 codes.

| Diagnosis | ICD-10 | ICD-9 |
|---|---|---|
| | D430,D431,D432,D433,D434,D438,D439,D440,D4410,D4411,D4412<br>D442,D443,D444,D445,D446,D447,D449,D45,D470,D4701,D4702,D4709<br>D471,D472,D473,D474,D479,D47Z1,D47Z2,D47Z9,D480,D481,D482<br>D483,D484,D485,D4860,D4861,D4862,D487,D489,D490,D491,D492<br>D493,D494,D495,D49511,D49512,D49519,D4959,D496,D497,D4981<br>D4989,D499 | |
| Nephritis; nephrosis; renal sclerosis | A3684,A5144,B2683,B520,B5883,E0821,E0921,E1021,E1121,E1321<br>M3504,M350A,N000,N001,N002,N003,N004,N005,N006,N007,N008,N009<br>N00A,N010,N011,N012,N013,N014,N015,N016,N017,N018,N019,N01A<br>N020,N021,N022,N023,N024,N025,N026,N027,N028,N029,N02A,N030<br>N031,N032,N033,N034,N035,N036,N037,N038,N039,N03A,N040,N041<br>N042,N043,N044,N045,N046,N047,N048,N049,N04A,N050,N051,N052<br>N053,N054,N055,N056,N057,N058,N059,N05A,N060,N061,N062,N063<br>N064,N065,N066,N067,N068,N069,N06A,N070,N071,N072,N073,N074<br>N075,N076,N077,N078,N079,N07A,N08,N140,N141,N1411,N1419,N142<br>N143,N144,N150,N158,N159,N16,N269 | 03289,07279,0849,09189,1307,24940,25040,25041,5800,5804,58089<br>5809,5810,5811,5812,5813,58181,58189,5819,5820,5821,5822,5824<br>58289,5829,5830,5831,5832,5834,58381,58389,5839,587,59081<br>5909,7102 |
| Nerve and nerve root disorders | A1783,B0221,B0222,E0841,E0941,E1041,E1141,E1341,G500,G501<br>G508,G509,G510,G511,G512,G513,G5131,G5132,G5133,G5139,G514<br>G518,G519,G520,G521,G522,G523,G527,G528,G529,G53,G540,G541<br>G542,G543,G544,G545,G546,G547,G548,G549,G55,G5600,G5601,G5602<br>G5603,G5610,G5611,G5612,G5613,G5620,G5621,G5622,G5623,G5630<br>G5631,G5632,G5633,G5640,G5641,G5642,G5643,G5680,G5681,G5682<br>G5683,G5690,G5691,G5692,G5693,G5700,G5701,G5702,G5703,G5710<br>G5711,G5712,G5713,G5720,G5721,G5722,G5723,G5730,G5731,G5732<br>G5733,G5740,G5741,G5742,G5743,G5750,G5751,G5752,G5753,G5760<br>G5761,G5762,G5763,G5770,G5771,G5772,G5773,G5780,G5781,G5782<br>G5783,G5790,G5791,G5792,G5793,G580,G587,G588,G589,G59 | 01362,01790,05311,05312,24960,25060,25061,3501,3502,3508,3509<br>3510,3511,3518,3519,3520,3521,3522,3523,3524,3525,3526,3529<br>3530,3531,3532,3533,3534,3535,3536,3538,3539,3540,3541,3542<br>3543,3544,3545,3548,3549,3550,3551,3552,3553,3554,3555,3556<br>35571,35579,3558,3559 |
| Nervous system cancers - all other types | C470,C4710,C4711,C4712,C4720,C4721,C4722,C473,C474,C475,C476<br>C478,C479,C720,C721,C7220,C7221,C7222,C7230,C7231,C7232,C7240<br>C7241,C7242,C7250,C7259,C729 | 1710,1712,1713,1714,1715,1716,1717,1718,1719,1920,1922,1928<br>1929 |
| Nervous system cancers - brain | C700,C701,C709,C710,C711,C712,C713,C714,C715,C716,C717,C718<br>C719 | 1910,1911,1912,1913,1914,1915,1916,1917,1918,1919,1921,1923 |
| Nervous system congenital anomalies | Q000,Q001,Q002,Q010,Q011,Q012,Q018,Q019,Q02,Q030,Q031,Q038<br>Q039,Q040,Q041,Q042,Q043,Q044,Q045,Q046,Q048,Q049,Q050,Q051<br>Q052,Q053,Q054,Q055,Q056,Q057,Q058,Q059,Q060,Q061,Q062,Q063 | 23770,23771,23772,23773,23779,7400,7401,7402,74100,74101<br>74102,74103,74190,74191,74192,74193,7420,7421,7422,7423,7424<br>74251,74253,74259,7428,7429 |

Continued on next page

**Supplementary Table S1:** (cont'd) A comprehensive mapping of diagnosis groups used in the experiments to their corresponding ICD-10 and ICD-9 codes.

| Diagnosis | ICD-10 | ICD-9 |
|---|---|---|
| | Q064,Q068,Q069,Q0700,Q0701,Q0702,Q0703,Q078,Q079,Q8500,Q8501<br>Q8502,Q8503,Q8509 | |
| Nervous system pain<br>and pain syndromes | G890,G8911,G8912,G8918,G8921,G8922,G8928,G8929,G893,G894,G902<br>G9050,G90511,G90512,G90513,G90519,G90521,G90522,G90523,G90529<br>G9059 | 33720,33721,33722,33729,3379,3380,33811,33812,33818,33821<br>33822,33828,33829,3383,3384 |
| Nervous system signs<br>and symptoms | R250,R251,R253,R258,R259,R260,R261,R262,R2681,R2689,R269,R270<br>R278,R279,R290,R291,R292,R295,R29810,R29818,R2990,R402110<br>R402111,R402112,R402113,R402114,R402120,R402121,R402122<br>R402123,R402124,R402130,R402131,R402132,R402133,R402134<br>R402140,R402141,R402142,R402143,R402144,R402210,R402211<br>R402212,R402213,R402214,R402220,R402221,R402222,R402223<br>R402224,R402230,R402231,R402232,R402233,R402234,R402240<br>R402241,R402242,R402243,R402244,R402250,R402251,R402252<br>R402253,R402254,R402310,R402311,R402312,R402313,R402314<br>R402320,R402321,R402322,R402323,R402324,R402330,R402331<br>R402332,R402333,R402334,R402340,R402341,R402342,R402343<br>R402344,R402350,R402351,R402352,R402353,R402354,R402360<br>R402361,R402362,R402363,R402364,R40241,R402410,R402411<br>R402412,R402413,R402414,R40242,R402420,R402421,R402422<br>R402423,R402424,R40243,R402430,R402431,R402432,R402433<br>R402434,R40244,R402440,R402441,R402442,R402443,R402444,R404<br>R410,R411,R412,R413,R414,R4181,R4182,R4183,R41840,R41841<br>R41842,R41843,R41844,R4189,R419,R430,R431,R432,R438,R439<br>R4701,R4702,R471,R4781,R4782,R4789,R479,R480,R481,R482,R483<br>R488,R489,R51,R510,R519,R532 | 31501,36816,7197,78001,78002,78072,78093,78097,78099,7810<br>7811,7812,7813,7814,7816,7817,7818,78194,78199,7840,7843<br>78451,78452,78459,78460,78461,78469,7961,797,79951,79952<br>79953,79954,79955,79959,NoDx,V414,V415,V6289 |
| Neuro-ophthalmology | A3982,A5044,H4600,H4601,H4602,H4603,H4610,H4611,H4612,H4613<br>H462,H463,H468,H469,H47011,H47012,H47013,H47019,H47021,H47022<br>H47023,H47029,H47031,H47032,H47033,H47039,H47091,H47092<br>H47093,H47099,H4710,H4711,H4712,H4713,H47141,H47142,H47143<br>H47149,H4720,H47211,H47212,H47213,H47219,H4722,H47231,H47232<br>H47233,H47239,H47291,H47292,H47293,H47299,H47311,H47312<br>H47313,H47319,H47321,H47322,H47323,H47329,H47331,H47332<br>H47333,H47339,H47391,H47392,H47393,H47399,H4741,H4742,H4743<br>H4749,H47511,H47512,H47519,H47521,H47522,H47529,H47531,H47532<br>H47539,H47611,H47612,H47619,H47621,H47622,H47629,H47631<br>H47632,H47639,H47641,H47642,H47649,H479,H4900,H4901,H4902<br>H4903,H4910,H4911,H4912,H4913,H4920,H4921,H4922,H4923,H4930 | 03681,0905,27787,37700,37701,37702,37703,37704,37710,37711<br>37712,37713,37714,37715,37716,37721,37722,37723,37724,37730<br>37731,37732,37733,37734,37739,37741,37742,37743,37749,37751<br>37752,37753,37754,37761,37762,37763,37771,37772,37773,37775<br>3779,37850,37851,37852,37853,37854,37855,37856,37885,37886<br>37887,3789 |

Continued on next page

**Supplementary Table S1:** (cont'd) A comprehensive mapping of diagnosis groups used in the experiments to their corresponding ICD-10 and ICD-9 codes.

| Diagnosis | ICD-10 | ICD-9 |
|---|---|---|
| | H4931,H4932,H4933,H4940,H4941,H4942,H4943,H49811,H49812 | |
| | H49813,H49819,H49881,H49882,H49883,H49889,H499,H5120,H5121 | |
| | H5122,H5123,H518,H519 | |
| Neurocognitive | F0150,F0151,F01511,F01518,F0152,F0153,F0154,F01A0,F01A11 | 2900,29010,29011,29013,29021,29040,29041,2908,2909,29282,2930 |
| disorders | F01A18,F01A2,F01A3,F01A4,F01B0,F01B11,F01B18,F01B2,F01B3 | 2931,2940,29410,29411,29420,29421,30460,3102,31081,31089,3308 |
| | F01B4,F01C0,F01C11,F01C18,F01C2,F01C3,F01C4,F0280,F0281 | 3310,33111,33119,3312,33182,33183,3319 |
| | F02811,F02818,F0282,F0283,F0284,F02A0,F02A11,F02A18,F02A2 | |
| | F02A3,F02A4,F02B0,F02B11,F02B18,F02B2,F02B3,F02B4,F02C0 | |
| | F02C11,F02C18,F02C2,F02C3,F02C4,F0390,F0391,F03911,F03918 | |
| | F0392,F0393,F0394,F03A0,F03A11,F03A18,F03A2,F03A3,F03A4,F03B0 | |
| | F03B11,F03B18,F03B2,F03B3,F03B4,F03C0,F03C11,F03C18,F03C2 | |
| | F03C3,F03C4,F04,F05,F0670,F0671,F0781,F0789,F1917,F1927,F1997 | |
| | F482,G300,G301,G308,G309,G3101,G3109,G311,G3181,G3183,G3184 | |
| | G319 | |
| Neurodevelopmental | F70,F71,F72,F73,F78,F78A1,F78A9,F79,F800,F801,F802,F804,F8081 | 29900,29901,29910,29980,29981,29990,3070,30720,30721,30722 |
| disorders | F8082,F8089,F809,F810,F812,F8181,F8189,F819,F82,F840,F842 | 30723,3073,30929,31389,3139,31400,31401,3142,3149,31500,31502 |
| | F843,F845,F848,F849,F88,F89,F900,F901,F902,F908,F909,F948 | 31509,3151,3152,31531,31532,31534,31535,31539,3154,3158,3159 |
| | F949,F950,F951,F952,F958,F959,F984,F985,F988 | 317,3180,3181,3182,319,3308,V400 |
| Neurogenic | G980,M1460,M14611,M14612,M14619,M14621,M14622,M14629,M14631 | 34989,7135 |
| neuropathic | M14632,M14639,M14641,M14642,M14649,M14651,M14652,M14659 | |
| arthropathy | M14661,M14662,M14669,M14671,M14672,M14679,M1468,M1469 | |
| Newborn affected by | P000,P001,P002,P003,P004,P005,P006,P007,P0081,P0082,P0089 | 7600,7601,7602,7603,7604,7605,76062,76063,76070,76071,76072 |
| maternal conditions | P009,P010,P011,P012,P013,P014,P015,P016,P017,P018,P019,P020 | 76073,76074,76075,76078,76079,7608,7609,7610,7611,7612,7613 |
| or complications of | P021,P0220,P0229,P023,P024,P025,P0260,P0269,P027,P0270,P0278 | 7614,7615,7616,7617,7618,7619,7620,7621,7622,7623,7624,7625 |
| labor delivery | P028,P029,P030,P031,P032,P033,P034,P035,P036,P03810,P03811 | 7626,7627,7628,7629,7630,7631,7632,7633,7634,7635,7636,7637 |
| | P03819,P0382,P0389,P039,P040,P041,P0411,P0412,P0413,P0414 | 76381,76382,76383,76384,76389,7639,7795 |
| | P0415,P0416,P0417,P0418,P0419,P041A,P042,P043,P0440,P0441 | |
| | P0442,P0449,P045,P046,P048,P0481,P0489,P049,P961 | |
| Non-Hodgkin lymphoma | C8200,C8201,C8202,C8203,C8204,C8205,C8206,C8207,C8208,C8209 | 20000,20001,20002,20003,20004,20005,20006,20007,20008,20010 |
| | C8210,C8211,C8212,C8213,C8214,C8215,C8216,C8217,C8218,C8219 | 20011,20012,20013,20014,20015,20016,20017,20018,20020,20021 |
| | C8220,C8221,C8222,C8223,C8224,C8225,C8226,C8227,C8228,C8229 | 20022,20023,20024,20025,20026,20027,20028,20030,20040,20041 |
| | C8230,C8231,C8232,C8233,C8234,C8235,C8236,C8237,C8238,C8239 | 20042,20043,20044,20045,20046,20047,20048,20050,20051,20052 |
| | C8240,C8241,C8242,C8243,C8244,C8245,C8246,C8247,C8248,C8249 | 20053,20054,20055,20056,20057,20058,20060,20061,20062,20063 |
| | C8250,C8251,C8252,C8253,C8254,C8255,C8256,C8257,C8258,C8259 | 20064,20065,20066,20067,20068,20070,20071,20072,20073,20074 |
| | C8260,C8261,C8262,C8263,C8264,C8265,C8266,C8267,C8268,C8269 | 20075,20076,20077,20078,20080,20081,20082,20083,20084,20085 |

Continued on next page

**Supplementary Table S1:** (cont'd) A comprehensive mapping of diagnosis groups used in the experiments to their corresponding ICD-10 and ICD-9 codes.

| Diagnosis | ICD-10 | ICD-9 |
|---|---|---|
| | C8280,C8281,C8282,C8283,C8284,C8285,C8286,C8287,C8288,C8289 | 20086,20087,20088,20200,20201,20202,20203,20204,20205,20206 |
| | C8290,C8291,C8292,C8293,C8294,C8295,C8296,C8297,C8298,C8299 | 20207,20208,20210,20211,20212,20213,20214,20215,20216,20217 |
| | C8300,C8301,C8302,C8303,C8304,C8305,C8306,C8307,C8308,C8309 | 20218,20220,20221,20222,20223,20224,20225,20226,20227,20228 |
| | C8310,C8311,C8312,C8313,C8314,C8315,C8316,C8317,C8318,C8319 | 20250,20260,20270,20271,20272,20273,20274,20275,20276,20277 |
| | C8330,C8331,C8332,C8333,C8334,C8335,C8336,C8337,C8338,C8339 | 20278,20280,20281,20282,20283,20284,20285,20286,20287,20288 |
| | C8350,C8351,C8352,C8353,C8354,C8355,C8356,C8357,C8358,C8359 | 20290,20291,20292,20293,20294,20295,20296,20297,20298,20380 |
| | C8370,C8371,C8372,C8373,C8374,C8375,C8376,C8377,C8378,C8379 | 20381,23879,2733,27789 |
| | C8380,C8381,C8382,C8383,C8384,C8385,C8386,C8387,C8388,C8389 | |
| | C8390,C8391,C8392,C8393,C8394,C8395,C8396,C8397,C8398,C8399 | |
| | C8400,C8401,C8402,C8403,C8404,C8405,C8406,C8407,C8408,C8409 | |
| | C8410,C8411,C8412,C8413,C8414,C8415,C8416,C8417,C8418,C8419 | |
| | C8440,C8441,C8442,C8443,C8444,C8445,C8446,C8447,C8448,C8449 | |
| | C8460,C8461,C8462,C8463,C8464,C8465,C8466,C8467,C8468,C8469 | |
| | C8470,C8471,C8472,C8473,C8474,C8475,C8476,C8477,C8478,C8479 | |
| | C847A,C8490,C8491,C8492,C8493,C8494,C8495,C8496,C8497,C8498 | |
| | C8499,C84A0,C84A1,C84A2,C84A3,C84A4,C84A5,C84A6,C84A7,C84A8 | |
| | C84A9,C84Z0,C84Z1,C84Z2,C84Z3,C84Z4,C84Z5,C84Z6,C84Z7,C84Z8 | |
| | C84Z9,C8510,C8511,C8512,C8513,C8514,C8515,C8516,C8517,C8518 | |
| | C8519,C8520,C8521,C8522,C8523,C8524,C8525,C8526,C8527,C8528 | |
| | C8529,C8580,C8581,C8582,C8583,C8584,C8585,C8586,C8587,C8588 | |
| | C8589,C8590,C8591,C8592,C8593,C8594,C8595,C8596,C8597,C8598 | |
| | C8599,C860,C861,C862,C863,C864,C865,C866,C880,C882,C883,C884 | |
| | C888,C889,C960,C962,C9620,C9621,C9629,C965,C966,C969,C96Z | |
| Non-pressure ulcer of | L97101,L97102,L97103,L97104,L97105,L97106,L97108,L97109 | 70710,70711,70712,70713,70714,70715,70719,7078,7079 |
| skin | L97111,L97112,L97113,L97114,L97115,L97116,L97118,L97119 | |
| | L97121,L97122,L97123,L97124,L97125,L97126,L97128,L97129 | |
| | L97201,L97202,L97203,L97204,L97205,L97206,L97208,L97209 | |
| | L97211,L97212,L97213,L97214,L97215,L97216,L97218,L97219 | |
| | L97221,L97222,L97223,L97224,L97225,L97226,L97228,L97229 | |
| | L97301,L97302,L97303,L97304,L97305,L97306,L97308,L97309 | |
| | L97311,L97312,L97313,L97314,L97315,L97316,L97318,L97319 | |
| | L97321,L97322,L97323,L97324,L97325,L97326,L97328,L97329 | |
| | L97401,L97402,L97403,L97404,L97405,L97406,L97408,L97409 | |
| | L97411,L97412,L97413,L97414,L97415,L97416,L97418,L97419 | |
| | L97421,L97422,L97423,L97424,L97425,L97426,L97428,L97429 | |
| | L97501,L97502,L97503,L97504,L97505,L97506,L97508,L97509 | |
| | L97511,L97512,L97513,L97514,L97515,L97516,L97518,L97519 | |
| | L97521,L97522,L97523,L97524,L97525,L97526,L97528,L97529 | |
| | L97801,L97802,L97803,L97804,L97805,L97806,L97808,L97809 | |

Continued on next page

**Supplementary Table S1:** (cont'd) A comprehensive mapping of diagnosis groups used in the experiments to their corresponding ICD-10 and ICD-9 codes.

| Diagnosis | ICD-10 | ICD-9 |
|---|---|---|
| | L97811,L97812,L97813,L97814,L97815,L97816,L97818,L97819<br>L97821,L97822,L97823,L97824,L97825,L97826,L97828,L97829<br>L97901,L97902,L97903,L97904,L97905,L97906,L97908,L97909<br>L97911,L97912,L97913,L97914,L97915,L97916,L97918,L97919<br>L97921,L97922,L97923,L97924,L97925,L97926,L97928,L97929<br>L98411,L98412,L98413,L98414,L98415,L98416,L98418,L98419<br>L98421,L98422,L98423,L98424,L98425,L98426,L98428,L98429<br>L98491,L98492,L98493,L98494,L98495,L98496,L98498,L98499 | |
| Noninfectious gastroenteritis | K520,K521,K522,K5221,K5222,K5229,K523,K5281,K5282,K52831<br>K52832,K52838,K52839,K5289,K529 | 53570,53571,5581,5582,5583,55841,55842,5589,78791 |
| Noninfectious hepatitis | K712,K713,K714,K7150,K7151,K716,K730,K731,K732,K738,K739,K752<br>K753,K754,K7581 | 57140,57141,57142,57149,5718,5733 |
| Nonmalignant breast conditions | N6001,N6002,N6009,N6011,N6012,N6019,N6021,N6022,N6029,N6031<br>N6032,N6039,N6041,N6042,N6049,N6081,N6082,N6089,N6091,N6092<br>N6099,N61,N610,N611,N6120,N6121,N6122,N6123,N62,N63,N630<br>N6310,N6311,N6312,N6313,N6314,N6315,N6320,N6321,N6322,N6323<br>N6324,N6325,N6331,N6332,N6341,N6342,N640,N641,N642,N643,N644<br>N6451,N6452,N6453,N6459,N6481,N6482,N6489,N649,N650,N651<br>O91011,O91012,O91013,O91019,O9102,O9103,O91111,O91112,O91113<br>O91119,O9112,O9113,O91211,O91212,O91213,O91219,O9122,O9123<br>O92011,O92012,O92013,O92019,O9202,O9203 | 6100,6101,6102,6103,6104,6108,6109,6110,6111,6112,6113,6114<br>6115,6116,61171,61172,61179,61181,61182,61189,6119,6120,6121<br>67500,67501,67502,67503,67504,67510,67511,67512,67513,67514<br>67520,67521,67522,67523,67524,67582,67584,67600,67601,67602<br>67603,67604 |
| Nonrheumatic and unspecified valve disorders | I340,I341,I342,I348,I3481,I3489,I349,I350,I351,I352,I358,I359<br>I360,I361,I362,I368,I369,I370,I371,I372,I378,I379,Z952,Z953<br>Z954 | 4240,4241,4242,4243,V422,V433 |
| Nonspecific chest pain | R072,R0789,R079 | 78650,78651,78659 |
| Nutritional anemia | D500,D501,D508,D509,D510,D511,D512,D513,D518,D519,D520,D521<br>D528,D529,D530,D531,D532,D538,D539 | 2800,2801,2808,2809,2810,2811,2812,2813,2814,2818,2819 |
| Nutritional deficiencies | E500,E501,E502,E503,E504,E505,E506,E507,E508,E509,E5111,E5112<br>E512,E518,E519,E52,E530,E531,E538,E539,E54,E550,E559,E560<br>E561,E568,E569,E58,E59,E60,E610,E611,E612,E613,E614,E615,E616<br>E617,E618,E619,E630,E631,E638,E639 | 2640,2641,2642,2643,2644,2645,2646,2647,2648,2649,2650,2651<br>2652,2660,2661,2662,2669,267,2680,2689,2690,2691,2692,2693<br>2698,2699 |

Continued on next page

**Supplementary Table S1:** (cont'd) A comprehensive mapping of diagnosis groups used in the experiments to their corresponding ICD-10 and ICD-9 codes.

| Diagnosis | ICD-10 | ICD-9 |
|---|---|---|
| OB-related trauma to perineum and vulva | O700,O701,O702,O7020,O7021,O7022,O7023,O703,O704,O709,O7182 | 66401,66404,66411,66414,66421,66424,66431,66434,66441,66444<br>66461,66464,66481,66484 |
| Obesity | E6601,E6609,E661,E662,E668,E669,O99210,O99211,O99212,O99213<br>O99214,O99215,Z6830,Z6831,Z6832,Z6833,Z6834,Z6835,Z6836,Z6837<br>Z6838,Z6839,Z6841,Z6842,Z6843,Z6844,Z6845,Z6854 | 27800,27801,27803,64910,64911,64912,64913,64914,V8530,V8531<br>V8532,V8533,V8534,V8535,V8536,V8537,V8538,V8539,V8541,V8542<br>V8543,V8544,V8545,V8554 |
| Obsessive-compulsive and related disorders | F42,F422,F423,F424,F428,F429,F4522,F633 | 3003,3007,3079,31239 |
| Obstetric history affecting care in pregnancy | O09211,O09212,O09213,O09219,O09291,O09292,O09293,O09299,O2620<br>O2621,O2622,O2623,O2630,O2631,O2632,O2633 | 64630,64631,64633,64680,64681,64683,V2341,V2349,V235 |
| Occlusion or stenosis of precerebral or cerebral arteries without infarction | I6501,I6502,I6503,I6509,I651,I6521,I6522,I6523,I6529,I658<br>I659,I6601,I6602,I6603,I6609,I6611,I6612,I6613,I6619,I6621<br>I6622,I6623,I6629,I663,I668,I669 | 43300,43310,43320,43380,43390,43400,43410,43490 |
| Oculofacial plastics and orbital conditions | H00031,H00032,H00033,H00034,H00035,H00036,H00039,H01111<br>H01112,H01113,H01114,H01115,H01116,H01119,H01121,H01122<br>H01123,H01124,H01125,H01126,H01129,H01131,H01132,H01133<br>H01134,H01135,H01136,H01139,H01141,H01142,H01143,H01144<br>H01145,H01146,H01149,H018,H019,H02001,H02002,H02003,H02004<br>H02005,H02006,H02009,H02011,H02012,H02013,H02014,H02015<br>H02016,H02019,H02021,H02022,H02023,H02024,H02025,H02026<br>H02029,H02031,H02032,H02033,H02034,H02035,H02036,H02039<br>H02041,H02042,H02043,H02044,H02045,H02046,H02049,H02051<br>H02052,H02053,H02054,H02055,H02056,H02059,H02101,H02102<br>H02103,H02104,H02105,H02106,H02109,H02111,H02112,H02113<br>H02114,H02115,H02116,H02119,H02121,H02122,H02123,H02124<br>H02125,H02126,H02129,H02131,H02132,H02133,H02134,H02135<br>H02136,H02139,H02141,H02142,H02143,H02144,H02145,H02146<br>H02149,H02151,H02152,H02153,H02154,H02155,H02156,H02159<br>H02201,H02202,H02203,H02204,H02205,H02206,H02209,H0220A<br>H0220B,H0220C,H02211,H02212,H02213,H02214,H02215,H02216<br>H02219,H0221A,H0221B,H0221C,H02221,H02222,H02223,H02224<br>H02225,H02226,H02229,H0222A,H0222B,H0222C,H02231,H02232<br>H02233,H02234,H02235,H02236,H02239,H0223A,H0223B,H0223C,H0230<br>H0231,H0232,H0233,H0234,H0235,H0236,H02401,H02402,H02403 | 36081,36089,3609,37313,37331,37332,37333,37334,3738,3739<br>37400,37401,37402,37403,37404,37405,37410,37411,37412,37413<br>37414,37420,37421,37422,37423,37430,37431,37432,37433,37434<br>37441,37443,37444,37445,37446,37450,37451,37452,37453,37454<br>37455,37456,37481,37482,37483,37484,37485,37486,37487,37489<br>3749,37500,37501,37502,37503,37511,37512,37513,37514,37515<br>37516,37520,37521,37522,37530,37531,37532,37533,37541,37542<br>37543,37551,37552,37553,37554,37555,37556,37557,37561,37569<br>37581,37589,3759,37600,37601,37602,37603,37604,37610,37611<br>37612,37630,37631,37632,37633,37634,37635,37636,37640,37642<br>37643,37645,37646,37647,37650,37651,37652,3766,37681,37682<br>37689,3769 |

Continued on next page

**Supplementary Table S1:** (cont'd) A comprehensive mapping of diagnosis groups used in the experiments to their corresponding ICD-10 and ICD-9 codes.

| Diagnosis | ICD-10 | ICD-9 |
|---|---|---|
| | H02409,H02411,H02412,H02413,H02419,H02421,H02422,H02423 | |
| | H02429,H02431,H02432,H02433,H02439,H02511,H02512,H02513 | |
| | H02514,H02515,H02516,H02519,H02521,H02522,H02523,H02524 | |
| | H02525,H02526,H02529,H02531,H02532,H02533,H02534,H02535 | |
| | H02536,H02539,H0259,H0260,H0261,H0262,H0263,H0264,H0265,H0266 | |
| | H0270,H02711,H02712,H02713,H02714,H02715,H02716,H02719,H02721 | |
| | H02722,H02723,H02724,H02725,H02726,H02729,H02731,H02732 | |
| | H02733,H02734,H02735,H02736,H02739,H0279,H02811,H02812,H02813 | |
| | H02814,H02815,H02816,H02819,H02821,H02822,H02823,H02824 | |
| | H02825,H02826,H02829,H02831,H02832,H02833,H02834,H02835 | |
| | H02836,H02839,H02841,H02842,H02843,H02844,H02845,H02846 | |
| | H02849,H02851,H02852,H02853,H02854,H02855,H02856,H02859 | |
| | H02861,H02862,H02863,H02864,H02865,H02866,H02869,H02871 | |
| | H02872,H02873,H02874,H02875,H02876,H02879,H0289,H029,H04001 | |
| | H04002,H04003,H04009,H04011,H04012,H04013,H04019,H04021 | |
| | H04022,H04023,H04029,H04031,H04032,H04033,H04039,H04111 | |
| | H04112,H04113,H04119,H04121,H04122,H04123,H04129,H04131 | |
| | H04132,H04133,H04139,H04141,H04142,H04143,H04149,H04151 | |
| | H04152,H04153,H04159,H04161,H04162,H04163,H04169,H0419,H04201 | |
| | H04202,H04203,H04209,H04211,H04212,H04213,H04219,H04221 | |
| | H04222,H04223,H04229,H04301,H04302,H04303,H04309,H04311 | |
| | H04312,H04313,H04319,H04321,H04322,H04323,H04329,H04331 | |
| | H04332,H04333,H04339,H04411,H04412,H04413,H04419,H04421 | |
| | H04422,H04423,H04429,H04431,H04432,H04433,H04439,H04511 | |
| | H04512,H04513,H04519,H04521,H04522,H04523,H04529,H04531 | |
| | H04532,H04533,H04539,H04541,H04542,H04543,H04549,H04551 | |
| | H04552,H04553,H04559,H04561,H04562,H04563,H04569,H04571 | |
| | H04572,H04573,H04579,H04611,H04612,H04613,H04619,H0469,H04811 | |
| | H04812,H04813,H04819,H0489,H049,H0500,H05011,H05012,H05013 | |
| | H05019,H05021,H05022,H05023,H05029,H05031,H05032,H05033 | |
| | H05039,H05041,H05042,H05043,H05049,H0510,H05111,H05112,H05113 | |
| | H05119,H05121,H05122,H05123,H05129,H0520,H05211,H05212,H05213 | |
| | H05219,H05221,H05222,H05223,H05229,H05231,H05232,H05233 | |
| | H05239,H05241,H05242,H05243,H05249,H05251,H05252,H05253 | |
| | H05259,H05261,H05262,H05263,H05269,H0530,H05311,H05312,H05313 | |
| | H05319,H05321,H05322,H05323,H05329,H05331,H05332,H05333 | |
| | H05339,H05341,H05342,H05343,H05349,H05351,H05352,H05353 | |
| | H05359,H05401,H05402,H05403,H05409,H05411,H05412,H05413 | |
| | H05419,H05421,H05422,H05423,H05429,H0550,H0551,H0552,H0553 | |
| | H05811,H05812,H05813,H05819,H05821,H05822,H05823,H05829,H0589 | |

Continued on next page

**Supplementary Table S1:** (cont'd) A comprehensive mapping of diagnosis groups used in the experiments to their corresponding ICD-10 and ICD-9 codes.

| Diagnosis | ICD-10 | ICD-9 |
|---|---|---|
| | H059,H44821,H44822,H44823,H44829,H4489,H449,H57811,H57812 H57813,H57819 | |
| Open wounds of head and neck, initial encounter | S0100XA,S0101XA,S0102XA,S0103XA,S0104XA,S0105XA,S01101A S01102A,S01109A,S01111A,S01112A,S01119A,S01121A,S01122A S01129A,S01131A,S01132A,S01139A,S01141A,S01142A,S01149A S01151A,S01152A,S01159A,S0120XA,S0121XA,S0122XA,S0123XA S0124XA,S0125XA,S01301A,S01302A,S01309A,S01311A,S01312A S01319A,S01321A,S01322A,S01329A,S01331A,S01332A,S01339A S01341A,S01342A,S01349A,S01351A,S01352A,S01359A,S01401A S01402A,S01409A,S01411A,S01412A,S01419A,S01421A,S01422A S01429A,S01431A,S01432A,S01439A,S01441A,S01442A,S01449A S01451A,S01452A,S01459A,S01501A,S01502A,S01511A,S01512A S01521A,S01522A,S01531A,S01532A,S01541A,S01542A,S01551A S01552A,S0180XA,S0181XA,S0182XA,S0183XA,S0184XA,S0185XA S0190XA,S0191XA,S0192XA,S0193XA,S0194XA,S0195XA,S0520XA S0521XA,S0522XA,S0530XA,S0531XA,S0532XA,S0540XA,S0541XA S0542XA,S0550XA,S0551XA,S0552XA,S0560XA,S0561XA,S0562XA S0570XA,S0571XA,S0572XA,S080XXA,S0912XA,S0920XA,S0921XA S0922XA,S11011A,S11012A,S11013A,S11014A,S11015A,S11019A S11021A,S11022A,S11023A,S11024A,S11025A,S11029A,S11031A S11032A,S11033A,S11034A,S11035A,S11039A,S1110XA,S1111XA S1112XA,S1113XA,S1114XA,S1115XA,S1120XA,S1121XA,S1122XA S1123XA,S1124XA,S1125XA,S1180XA,S1181XA,S1182XA,S1183XA S1184XA,S1185XA,S1189XA,S1190XA,S1191XA,S1192XA,S1193XA S1194XA,S1195XA,S162XXA | 8700,8702,8703,8704,8708,8709,8710,8711,8712,8713,8715,8716 8717,87200,87201,87202,87210,87261,87271,8730,8731,87320 87330,87341,87342,87343,87344,87349,87351,87352,87353,87354 87359,87360,87361,87362,87364,87365,87370,87371,87372,87374 87375,8738,8739,87401,87402,87411,87412,8742,8743,8744,8745 8748,8749 |
| Open wounds of head and neck, subsequent encounter | S0100XD,S0101XD,S0102XD,S0103XD,S0104XD,S0105XD,S01101D S01102D,S01109D,S01111D,S01112D,S01119D,S01121D,S01122D S01129D,S01131D,S01132D,S01139D,S01141D,S01142D,S01149D S01151D,S01152D,S01159D,S0120XD,S0121XD,S0122XD,S0123XD S0124XD,S0125XD,S01301D,S01302D,S01309D,S01311D,S01312D S01319D,S01321D,S01322D,S01329D,S01331D,S01332D,S01339D S01341D,S01342D,S01349D,S01351D,S01352D,S01359D,S01401D S01402D,S01409D,S01411D,S01412D,S01419D,S01421D,S01422D S01429D,S01431D,S01432D,S01439D,S01441D,S01442D,S01449D S01451D,S01452D,S01459D,S01501D,S01502D,S01511D,S01512D S01521D,S01522D,S01531D,S01532D,S01541D,S01542D,S01551D S01552D,S0180XD,S0181XD,S0182XD,S0183XD,S0184XD,S0185XD S0190XD,S0191XD,S0192XD,S0193XD,S0194XD,S0195XD,S0520XD | V5889 |

Continued on next page

**Supplementary Table S1:** (cont'd) A comprehensive mapping of diagnosis groups used in the experiments to their corresponding ICD-10 and ICD-9 codes.

| Diagnosis | ICD-10 | ICD-9 |
|---|---|---|
| | S0521XD,S0522XD,S0530XD,S0531XD,S0532XD,S0540XD,S0541XD | |
| | S0542XD,S0550XD,S0551XD,S0552XD,S0560XD,S0561XD,S0562XD | |
| | S0570XD,S0571XD,S0572XD,S080XXD,S0912XD,S0920XD,S0921XD | |
| | S0922XD,S11011D,S11012D,S11013D,S11014D,S11015D,S11019D | |
| | S11021D,S11022D,S11023D,S11024D,S11025D,S11029D,S11031D | |
| | S11032D,S11033D,S11034D,S11035D,S11039D,S1110XD,S1111XD | |
| | S1112XD,S1113XD,S1114XD,S1115XD,S1120XD,S1121XD,S1122XD | |
| | S1123XD,S1124XD,S1125XD,S1180XD,S1181XD,S1182XD,S1183XD | |
| | S1184XD,S1185XD,S1189XD,S1190XD,S1191XD,S1192XD,S1193XD | |
| | S1194XD,S1195XD,S162XXD | |
| Open wounds of trunk, | S21001A,S21002A,S21009A,S21011A,S21012A,S21019A,S21021A | 8629,86813,86814,8750,8751,8760,8761,8770,8771,8780,8781,8782 |
| initial encounter | S21022A,S21029A,S21031A,S21032A,S21039A,S21041A,S21042A | 8783,8784,8785,8786,8787,8788,8789,8790,8791,8792,8793,8794 |
| | S21049A,S21051A,S21052A,S21059A,S21101A,S21102A,S21109A | 8795,8796,8797,8798,8799 |
| | S21111A,S21112A,S21119A,S21121A,S21122A,S21129A,S21131A | |
| | S21132A,S21139A,S21141A,S21142A,S21149A,S21151A,S21152A | |
| | S21159A,S21201A,S21202A,S21209A,S21211A,S21212A,S21219A | |
| | S21221A,S21222A,S21229A,S21231A,S21232A,S21239A,S21241A | |
| | S21242A,S21249A,S21251A,S21252A,S21259A,S21301A,S21302A | |
| | S21309A,S21311A,S21312A,S21319A,S21321A,S21322A,S21329A | |
| | S21331A,S21332A,S21339A,S21341A,S21342A,S21349A,S21351A | |
| | S21352A,S21359A,S21401A,S21402A,S21409A,S21411A,S21412A | |
| | S21419A,S21421A,S21422A,S21429A,S21431A,S21432A,S21439A | |
| | S21441A,S21442A,S21449A,S21451A,S21452A,S21459A,S2190XA | |
| | S2191XA,S2192XA,S2193XA,S2194XA,S2195XA,S29021A,S29022A | |
| | S29029A,S31000A,S31001A,S31010A,S31011A,S31020A,S31021A | |
| | S31030A,S31031A,S31040A,S31041A,S31050A,S31051A,S31100A | |
| | S31101A,S31102A,S31103A,S31104A,S31105A,S31109A,S31110A | |
| | S31111A,S31112A,S31113A,S31114A,S31115A,S31119A,S31120A | |
| | S31121A,S31122A,S31123A,S31124A,S31125A,S31129A,S31130A | |
| | S31131A,S31132A,S31133A,S31134A,S31135A,S31139A,S31140A | |
| | S31141A,S31142A,S31143A,S31144A,S31145A,S31149A,S31150A | |
| | S31151A,S31152A,S31153A,S31154A,S31155A,S31159A,S3120XA | |
| | S3121XA,S3122XA,S3123XA,S3124XA,S3125XA,S3130XA,S3131XA | |
| | S3132XA,S3133XA,S3134XA,S3135XA,S3140XA,S3141XA,S3142XA | |
| | S3143XA,S3144XA,S3145XA,S31501A,S31502A,S31511A,S31512A | |
| | S31521A,S31522A,S31531A,S31532A,S31541A,S31542A,S31551A | |
| | S31552A,S31600A,S31601A,S31602A,S31603A,S31604A,S31605A | |
| | S31609A,S31610A,S31611A,S31612A,S31613A,S31614A,S31615A | |
| | S31619A,S31620A,S31621A,S31622A,S31623A,S31624A,S31625A | |

Continued on next page

**Supplementary Table S1:** (cont'd) A comprehensive mapping of diagnosis groups used in the experiments to their corresponding ICD-10 and ICD-9 codes.

| Diagnosis | ICD-10 | ICD-9 |
|---|---|---|
| | S31629A,S31630A,S31631A,S31632A,S31633A,S31634A,S31635A | |
| | S31639A,S31640A,S31641A,S31642A,S31643A,S31644A,S31645A | |
| | S31649A,S31650A,S31651A,S31652A,S31653A,S31654A,S31655A | |
| | S31659A,S31801A,S31802A,S31803A,S31804A,S31805A,S31809A | |
| | S31811A,S31812A,S31813A,S31814A,S31815A,S31819A,S31821A | |
| | S31822A,S31823A,S31824A,S31825A,S31829A,S31831A,S31832A | |
| | S31833A,S31834A,S31835A,S31839A,S39021A,S39022A,S39023A | |
| Open wounds of trunk, | S21001D,S21002D,S21009D,S21011D,S21012D,S21019D,S21021D | V5889 |
| subsequent encounter | S21022D,S21029D,S21031D,S21032D,S21039D,S21041D,S21042D | |
| | S21049D,S21051D,S21052D,S21059D,S21101D,S21102D,S21109D | |
| | S21111D,S21112D,S21119D,S21121D,S21122D,S21129D,S21131D | |
| | S21132D,S21139D,S21141D,S21142D,S21149D,S21151D,S21152D | |
| | S21159D,S21201D,S21202D,S21209D,S21211D,S21212D,S21219D | |
| | S21221D,S21222D,S21229D,S21231D,S21232D,S21239D,S21241D | |
| | S21242D,S21249D,S21251D,S21252D,S21259D,S21301D,S21302D | |
| | S21309D,S21311D,S21312D,S21319D,S21321D,S21322D,S21329D | |
| | S21331D,S21332D,S21339D,S21341D,S21342D,S21349D,S21351D | |
| | S21352D,S21359D,S21401D,S21402D,S21409D,S21411D,S21412D | |
| | S21419D,S21421D,S21422D,S21429D,S21431D,S21432D,S21439D | |
| | S21441D,S21442D,S21449D,S21451D,S21452D,S21459D,S2190XD | |
| | S2191XD,S2192XD,S2193XD,S2194XD,S2195XD,S29021D,S29022D | |
| | S29029D,S31000D,S31001D,S31010D,S31011D,S31020D,S31021D | |
| | S31030D,S31031D,S31040D,S31041D,S31050D,S31051D,S31100D | |
| | S31101D,S31102D,S31103D,S31104D,S31105D,S31109D,S31110D | |
| | S31111D,S31112D,S31113D,S31114D,S31115D,S31119D,S31120D | |
| | S31121D,S31122D,S31123D,S31124D,S31125D,S31129D,S31130D | |
| | S31131D,S31132D,S31133D,S31134D,S31135D,S31139D,S31140D | |
| | S31141D,S31142D,S31143D,S31144D,S31145D,S31149D,S31150D | |
| | S31151D,S31152D,S31153D,S31154D,S31155D,S31159D,S3120XD | |
| | S3121XD,S3122XD,S3123XD,S3124XD,S3125XD,S3130XD,S3131XD | |
| | S3132XD,S3133XD,S3134XD,S3135XD,S3140XD,S3141XD,S3142XD | |
| | S3143XD,S3144XD,S3145XD,S31501D,S31502D,S31511D,S31512D | |
| | S31521D,S31522D,S31531D,S31532D,S31541D,S31542D,S31551D | |
| | S31552D,S31600D,S31601D,S31602D,S31603D,S31604D,S31605D | |
| | S31609D,S31610D,S31611D,S31612D,S31613D,S31614D,S31615D | |
| | S31619D,S31620D,S31621D,S31622D,S31623D,S31624D,S31625D | |
| | S31629D,S31630D,S31631D,S31632D,S31633D,S31634D,S31635D | |
| | S31639D,S31640D,S31641D,S31642D,S31643D,S31644D,S31645D | |
| | S31649D,S31650D,S31651D,S31652D,S31653D,S31654D,S31655D | |

Continued on next page

**Supplementary Table S1:** (cont'd) A comprehensive mapping of diagnosis groups used in the experiments to their corresponding ICD-10 and ICD-9 codes.

| Diagnosis | ICD-10 | ICD-9 |
|---|---|---|
| | S31659D,S31801D,S31802D,S31803D,S31804D,S31805D,S31809D | |
| | S31811D,S31812D,S31813D,S31814D,S31815D,S31819D,S31821D | |
| | S31822D,S31823D,S31824D,S31825D,S31829D,S31831D,S31832D | |
| | S31833D,S31834D,S31835D,S31839D,S39021D,S39022D,S39023D | |
| Open wounds to limbs, | S41001A,S41002A,S41009A,S41011A,S41012A,S41019A,S41021A | 88000,88001,88002,88003,88009,88010,88011,88012,88013,88019 |
| initial encounter | S41022A,S41029A,S41031A,S41032A,S41039A,S41041A,S41042A | 88020,88100,88101,88102,88110,88111,88112,88120,88122,8820 |
| | S41049A,S41051A,S41052A,S41059A,S41101A,S41102A,S41109A | 8821,8822,8830,8831,8840,8841,8900,8901,8902,8910,8911,8912 |
| | S41111A,S41112A,S41119A,S41121A,S41122A,S41129A,S41131A | 8920,8921,8930,8931,8941 |
| | S41132A,S41139A,S41141A,S41142A,S41149A,S41151A,S41152A | |
| | S41159A,S46021A,S46022A,S46029A,S46121A,S46122A,S46129A | |
| | S46221A,S46222A,S46229A,S46321A,S46322A,S46329A,S46821A | |
| | S46822A,S46829A,S46921A,S46922A,S46929A,S51001A,S51002A | |
| | S51009A,S51011A,S51012A,S51019A,S51021A,S51022A,S51029A | |
| | S51031A,S51032A,S51039A,S51041A,S51042A,S51049A,S51051A | |
| | S51052A,S51059A,S51801A,S51802A,S51809A,S51811A,S51812A | |
| | S51819A,S51821A,S51822A,S51829A,S51831A,S51832A,S51839A | |
| | S51841A,S51842A,S51849A,S51851A,S51852A,S51859A,S56021A | |
| | S56022A,S56029A,S56121A,S56122A,S56123A,S56124A,S56125A | |
| | S56126A,S56127A,S56128A,S56129A,S56221A,S56222A,S56229A | |
| | S56321A,S56322A,S56329A,S56421A,S56422A,S56423A,S56424A | |
| | S56425A,S56426A,S56427A,S56428A,S56429A,S56521A,S56522A | |
| | S56529A,S56821A,S56822A,S56829A,S56921A,S56922A,S56929A | |
| | S61001A,S61002A,S61009A,S61011A,S61012A,S61019A,S61021A | |
| | S61022A,S61029A,S61031A,S61032A,S61039A,S61041A,S61042A | |
| | S61049A,S61051A,S61052A,S61059A,S61101A,S61102A,S61109A | |
| | S61111A,S61112A,S61119A,S61121A,S61122A,S61129A,S61131A | |
| | S61132A,S61139A,S61141A,S61142A,S61149A,S61151A,S61152A | |
| | S61159A,S61200A,S61201A,S61202A,S61203A,S61204A,S61205A | |
| | S61206A,S61207A,S61208A,S61209A,S61210A,S61211A,S61212A | |
| | S61213A,S61214A,S61215A,S61216A,S61217A,S61218A,S61219A | |
| | S61220A,S61221A,S61222A,S61223A,S61224A,S61225A,S61226A | |
| | S61227A,S61228A,S61229A,S61230A,S61231A,S61232A,S61233A | |
| | S61234A,S61235A,S61236A,S61237A,S61238A,S61239A,S61240A | |
| | S61241A,S61242A,S61243A,S61244A,S61245A,S61246A,S61247A | |
| | S61248A,S61249A,S61250A,S61251A,S61252A,S61253A,S61254A | |
| | S61255A,S61256A,S61257A,S61258A,S61259A,S61300A,S61301A | |
| | S61302A,S61303A,S61304A,S61305A,S61306A,S61307A,S61308A | |
| | S61309A,S61310A,S61311A,S61312A,S61313A,S61314A,S61315A | |
| | S61316A,S61317A,S61318A,S61319A,S61320A,S61321A,S61322A | |

Continued on next page

**Supplementary Table S1:** (cont'd) A comprehensive mapping of diagnosis groups used in the experiments to their corresponding ICD-10 and ICD-9 codes.

| Diagnosis | ICD-10 | ICD-9 |
|---|---|---|
| | S61323A,S61324A,S61325A,S61326A,S61327A,S61328A,S61329A | |
| | S61330A,S61331A,S61332A,S61333A,S61334A,S61335A,S61336A | |
| | S61337A,S61338A,S61339A,S61340A,S61341A,S61342A,S61343A | |
| | S61344A,S61345A,S61346A,S61347A,S61348A,S61349A,S61350A | |
| | S61351A,S61352A,S61353A,S61354A,S61355A,S61356A,S61357A | |
| | S61358A,S61359A,S61401A,S61402A,S61409A,S61411A,S61412A | |
| | S61419A,S61421A,S61422A,S61429A,S61431A,S61432A,S61439A | |
| | S61441A,S61442A,S61449A,S61451A,S61452A,S61459A,S61501A | |
| | S61502A,S61509A,S61511A,S61512A,S61519A,S61521A,S61522A | |
| | S61529A,S61531A,S61532A,S61539A,S61541A,S61542A,S61549A | |
| | S61551A,S61552A,S61559A,S66021A,S66022A,S66029A,S66120A | |
| | S66121A,S66122A,S66123A,S66124A,S66125A,S66126A,S66127A | |
| | S66128A,S66129A,S66221A,S66222A,S66229A,S66320A,S66321A | |
| | S66322A,S66323A,S66324A,S66325A,S66326A,S66327A,S66328A | |
| | S66329A,S66421A,S66422A,S66429A,S66520A,S66521A,S66522A | |
| | S66523A,S66524A,S66525A,S66526A,S66527A,S66528A,S66529A | |
| | S66821A,S66822A,S66829A,S66921A,S66922A,S66929A,S71001A | |
| | S71002A,S71009A,S71011A,S71012A,S71019A,S71021A,S71022A | |
| | S71029A,S71031A,S71032A,S71039A,S71041A,S71042A,S71049A | |
| | S71051A,S71052A,S71059A,S71101A,S71102A,S71109A,S71111A | |
| | S71112A,S71119A,S71121A,S71122A,S71129A,S71131A,S71132A | |
| | S71139A,S71141A,S71142A,S71149A,S71151A,S71152A,S71159A | |
| | S76021A,S76022A,S76029A,S76121A,S76122A,S76129A,S76221A | |
| | S76222A,S76229A,S76321A,S76322A,S76329A,S76821A,S76822A | |
| | S76829A,S76921A,S76922A,S76929A,S81001A,S81002A,S81009A | |
| | S81011A,S81012A,S81019A,S81021A,S81022A,S81029A,S81031A | |
| | S81032A,S81039A,S81041A,S81042A,S81049A,S81051A,S81052A | |
| | S81059A,S81801A,S81802A,S81809A,S81811A,S81812A,S81819A | |
| | S81821A,S81822A,S81829A,S81831A,S81832A,S81839A,S81841A | |
| | S81842A,S81849A,S81851A,S81852A,S81859A,S86021A,S86022A | |
| | S86029A,S86121A,S86122A,S86129A,S86221A,S86222A,S86229A | |
| | S86321A,S86322A,S86329A,S86821A,S86822A,S86829A,S86921A | |
| | S86922A,S86929A,S91001A,S91002A,S91009A,S91011A,S91012A | |
| | S91019A,S91021A,S91022A,S91029A,S91031A,S91032A,S91039A | |
| | S91041A,S91042A,S91049A,S91051A,S91052A,S91059A,S91101A | |
| | S91102A,S91103A,S91104A,S91105A,S91106A,S91109A,S91111A | |
| | S91112A,S91113A,S91114A,S91115A,S91116A,S91119A,S91121A | |
| | S91122A,S91123A,S91124A,S91125A,S91126A,S91129A,S91131A | |
| | S91132A,S91133A,S91134A,S91135A,S91136A,S91139A,S91141A | |
| | S91142A,S91143A,S91144A,S91145A,S91146A,S91149A,S91151A | |

Continued on next page

**Supplementary Table S1:** (cont'd) A comprehensive mapping of diagnosis groups used in the experiments to their corresponding ICD-10 and ICD-9 codes.

| Diagnosis | ICD-10 | ICD-9 |
|---|---|---|
| | S91152A,S91153A,S91154A,S91155A,S91156A,S91159A,S91201A | |
| | S91202A,S91203A,S91204A,S91205A,S91206A,S91209A,S91211A | |
| | S91212A,S91213A,S91214A,S91215A,S91216A,S91219A,S91221A | |
| | S91222A,S91223A,S91224A,S91225A,S91226A,S91229A,S91231A | |
| | S91232A,S91233A,S91234A,S91235A,S91236A,S91239A,S91241A | |
| | S91242A,S91243A,S91244A,S91245A,S91246A,S91249A,S91251A | |
| | S91252A,S91253A,S91254A,S91255A,S91256A,S91259A,S91301A | |
| | S91302A,S91309A,S91311A,S91312A,S91319A,S91321A,S91322A | |
| | S91329A,S91331A,S91332A,S91339A,S91341A,S91342A,S91349A | |
| | S91351A,S91352A,S91359A,S96021A,S96022A,S96029A,S96121A | |
| | S96122A,S96129A,S96221A,S96222A,S96229A,S96821A,S96822A | |
| | S96829A,S96921A,S96922A,S96929A | |
| Open wounds to limbs, | S41001D,S41002D,S41009D,S41011D,S41012D,S41019D,S41021D | V5889 |
| subsequent encounter | S41022D,S41029D,S41031D,S41032D,S41039D,S41041D,S41042D | |
| | S41049D,S41051D,S41052D,S41059D,S41101D,S41102D,S41109D | |
| | S41111D,S41112D,S41119D,S41121D,S41122D,S41129D,S41131D | |
| | S41132D,S41139D,S41141D,S41142D,S41149D,S41151D,S41152D | |
| | S41159D,S46021D,S46022D,S46029D,S46121D,S46122D,S46129D | |
| | S46221D,S46222D,S46229D,S46321D,S46322D,S46329D,S46821D | |
| | S46822D,S46829D,S46921D,S46922D,S46929D,S51001D,S51002D | |
| | S51009D,S51011D,S51012D,S51019D,S51021D,S51022D,S51029D | |
| | S51031D,S51032D,S51039D,S51041D,S51042D,S51049D,S51051D | |
| | S51052D,S51059D,S51801D,S51802D,S51809D,S51811D,S51812D | |
| | S51819D,S51821D,S51822D,S51829D,S51831D,S51832D,S51839D | |
| | S51841D,S51842D,S51849D,S51851D,S51852D,S51859D,S56021D | |
| | S56022D,S56029D,S56121D,S56122D,S56123D,S56124D,S56125D | |
| | S56126D,S56127D,S56128D,S56129D,S56221D,S56222D,S56229D | |
| | S56321D,S56322D,S56329D,S56421D,S56422D,S56423D,S56424D | |
| | S56425D,S56426D,S56427D,S56428D,S56429D,S56521D,S56522D | |
| | S56529D,S56821D,S56822D,S56829D,S56921D,S56922D,S56929D | |
| | S61001D,S61002D,S61009D,S61011D,S61012D,S61019D,S61021D | |
| | S61022D,S61029D,S61031D,S61032D,S61039D,S61041D,S61042D | |
| | S61049D,S61051D,S61052D,S61059D,S61101D,S61102D,S61109D | |
| | S61111D,S61112D,S61119D,S61121D,S61122D,S61129D,S61131D | |
| | S61132D,S61139D,S61141D,S61142D,S61149D,S61151D,S61152D | |
| | S61159D,S61200D,S61201D,S61202D,S61203D,S61204D,S61205D | |
| | S61206D,S61207D,S61208D,S61209D,S61210D,S61211D,S61212D | |
| | S61213D,S61214D,S61215D,S61216D,S61217D,S61218D,S61219D | |
| | S61220D,S61221D,S61222D,S61223D,S61224D,S61225D,S61226D | |

Continued on next page

**Supplementary Table S1:** (cont'd) A comprehensive mapping of diagnosis groups used in the experiments to their corresponding ICD-10 and ICD-9 codes.

| Diagnosis | ICD-10 | ICD-9 |
|---|---|---|
| | S61227D,S61228D,S61229D,S61230D,S61231D,S61232D,S61233D | |
| | S61234D,S61235D,S61236D,S61237D,S61238D,S61239D,S61240D | |
| | S61241D,S61242D,S61243D,S61244D,S61245D,S61246D,S61247D | |
| | S61248D,S61249D,S61250D,S61251D,S61252D,S61253D,S61254D | |
| | S61255D,S61256D,S61257D,S61258D,S61259D,S61300D,S61301D | |
| | S61302D,S61303D,S61304D,S61305D,S61306D,S61307D,S61308D | |
| | S61309D,S61310D,S61311D,S61312D,S61313D,S61314D,S61315D | |
| | S61316D,S61317D,S61318D,S61319D,S61320D,S61321D,S61322D | |
| | S61323D,S61324D,S61325D,S61326D,S61327D,S61328D,S61329D | |
| | S61330D,S61331D,S61332D,S61333D,S61334D,S61335D,S61336D | |
| | S61337D,S61338D,S61339D,S61340D,S61341D,S61342D,S61343D | |
| | S61344D,S61345D,S61346D,S61347D,S61348D,S61349D,S61350D | |
| | S61351D,S61352D,S61353D,S61354D,S61355D,S61356D,S61357D | |
| | S61358D,S61359D,S61401D,S61402D,S61409D,S61411D,S61412D | |
| | S61419D,S61421D,S61422D,S61429D,S61431D,S61432D,S61439D | |
| | S61441D,S61442D,S61449D,S61451D,S61452D,S61459D,S61501D | |
| | S61502D,S61509D,S61511D,S61512D,S61519D,S61521D,S61522D | |
| | S61529D,S61531D,S61532D,S61539D,S61541D,S61542D,S61549D | |
| | S61551D,S61552D,S61559D,S66021D,S66022D,S66029D,S66120D | |
| | S66121D,S66122D,S66123D,S66124D,S66125D,S66126D,S66127D | |
| | S66128D,S66129D,S66221D,S66222D,S66229D,S66320D,S66321D | |
| | S66322D,S66323D,S66324D,S66325D,S66326D,S66327D,S66328D | |
| | S66329D,S66421D,S66422D,S66429D,S66520D,S66521D,S66522D | |
| | S66523D,S66524D,S66525D,S66526D,S66527D,S66528D,S66529D | |
| | S66821D,S66822D,S66829D,S66921D,S66922D,S66929D,S71001D | |
| | S71002D,S71009D,S71011D,S71012D,S71019D,S71021D,S71022D | |
| | S71029D,S71031D,S71032D,S71039D,S71041D,S71042D,S71049D | |
| | S71051D,S71052D,S71059D,S71101D,S71102D,S71109D,S71111D | |
| | S71112D,S71119D,S71121D,S71122D,S71129D,S71131D,S71132D | |
| | S71139D,S71141D,S71142D,S71149D,S71151D,S71152D,S71159D | |
| | S76021D,S76022D,S76029D,S76121D,S76122D,S76129D,S76221D | |
| | S76222D,S76229D,S76321D,S76322D,S76329D,S76821D,S76822D | |
| | S76829D,S76921D,S76922D,S76929D,S81001D,S81002D,S81009D | |
| | S81011D,S81012D,S81019D,S81021D,S81022D,S81029D,S81031D | |
| | S81032D,S81039D,S81041D,S81042D,S81049D,S81051D,S81052D | |
| | S81059D,S81801D,S81802D,S81809D,S81811D,S81812D,S81819D | |
| | S81821D,S81822D,S81829D,S81831D,S81832D,S81839D,S81841D | |
| | S81842D,S81849D,S81851D,S81852D,S81859D,S86021D,S86022D | |
| | S86029D,S86121D,S86122D,S86129D,S86221D,S86222D,S86229D | |
| | S86321D,S86322D,S86329D,S86821D,S86822D,S86829D,S86921D | |

Continued on next page

**Supplementary Table S1:** (cont'd) A comprehensive mapping of diagnosis groups used in the experiments to their corresponding ICD-10 and ICD-9 codes.

| Diagnosis | ICD-10 | ICD-9 |
|---|---|---|
| | S86922D,S86929D,S91001D,S91002D,S91009D,S91011D,S91012D | |
| | S91019D,S91021D,S91022D,S91029D,S91031D,S91032D,S91039D | |
| | S91041D,S91042D,S91049D,S91051D,S91052D,S91059D,S91101D | |
| | S91102D,S91103D,S91104D,S91105D,S91106D,S91109D,S91111D | |
| | S91112D,S91113D,S91114D,S91115D,S91116D,S91119D,S91121D | |
| | S91122D,S91123D,S91124D,S91125D,S91126D,S91129D,S91131D | |
| | S91132D,S91133D,S91134D,S91135D,S91136D,S91139D,S91141D | |
| | S91142D,S91143D,S91144D,S91145D,S91146D,S91149D,S91151D | |
| | S91152D,S91153D,S91154D,S91155D,S91156D,S91159D,S91201D | |
| | S91202D,S91203D,S91204D,S91205D,S91206D,S91209D,S91211D | |
| | S91212D,S91213D,S91214D,S91215D,S91216D,S91219D,S91221D | |
| | S91222D,S91223D,S91224D,S91225D,S91226D,S91229D,S91231D | |
| | S91232D,S91233D,S91234D,S91235D,S91236D,S91239D,S91241D | |
| | S91242D,S91243D,S91244D,S91245D,S91246D,S91249D,S91251D | |
| | S91252D,S91253D,S91254D,S91255D,S91256D,S91259D,S91301D | |
| | S91302D,S91309D,S91311D,S91312D,S91319D,S91321D,S91322D | |
| | S91329D,S91331D,S91332D,S91339D,S91341D,S91342D,S91349D | |
| | S91351D,S91352D,S91359D,S96021D,S96022D,S96029D,S96121D | |
| | S96122D,S96129D,S96221D,S96222D,S96229D,S96821D,S96822D | |
| | S96829D,S96921D,S96922D,S96929D | |
| Opioid-related | F1110,F11120,F11121,F11122,F11129,F1113,F1114,F11150,F11151 | 2920,29211,29212,2922,29281,29282,29284,29285,29289,2929 |
| disorders | F11159,F11181,F11182,F11188,F1119,F1120,F11220,F11221,F11222 | 30400,30401,30402,30550,30551,30552,30553,96500,96501,96502 |
| | F11229,F1123,F1124,F11250,F11251,F11259,F11281,F11282,F11288 | 96509,99529,E8500,E8501,E8502,E9351,E9352,E9500,E9504,E9620 |
| | F1129,F1190,F1191,F11920,F11921,F11922,F11929,F1193,F1194 | E9800,E9804 |
| | F11950,F11951,F11959,F11981,F11982,F11988,F1199,T400X1A | |
| | T400X2A,T400X3A,T400X4A,T400X5A,T401X1A,T401X2A,T401X3A | |
| | T401X4A,T402X1A,T402X2A,T402X3A,T402X4A,T402X5A,T403X1A | |
| | T403X2A,T403X3A,T403X4A,T403X5A,T40411A,T40412A,T40413A | |
| | T40414A,T40415A,T40422A,T40423A,T40424A,T40425A,T40491A | |
| | T40492A,T40493A,T40494A,T40495A,T404X1A,T404X2A,T404X3A | |
| | T404X4A,T404X5A,T40601A,T40602A,T40603A,T40604A,T40605A | |
| | T40691A,T40692A,T40693A,T40694A,T40695A | |
| Osteoarthritis | M150,M151,M152,M153,M154,M158,M159,M160,M1610,M1611,M1612 | 71500,71504,71509,71510,71511,71512,71513,71514,71515,71516 |
| | M162,M1630,M1631,M1632,M164,M1650,M1651,M1652,M166,M167,M169 | 71517,71520,71521,71522,71523,71524,71525,71526,71527,71528 |
| | M170,M1710,M1711,M1712,M172,M1730,M1731,M1732,M174,M175,M179 | 71530,71534,71535,71536,71580,71589,71590,71594,71595,71596 |
| | M180,M1810,M1811,M1812,M182,M1830,M1831,M1832,M184,M1850 | |
| | M1851,M1852,M189,M19011,M19012,M19019,M19021,M19022,M19029 | |
| | M19031,M19032,M19039,M19041,M19042,M19049,M19071,M19072 | |

Continued on next page

**Supplementary Table S1:** (cont'd) A comprehensive mapping of diagnosis groups used in the experiments to their corresponding ICD-10 and ICD-9 codes.

| Diagnosis | ICD-10 | ICD-9 |
|---|---|---|
| | M19079,M1909,M19111,M19112,M19119,M19121,M19122,M19129,M19131<br>M19132,M19139,M19141,M19142,M19149,M19171,M19172,M19179,M1919<br>M19211,M19212,M19219,M19221,M19222,M19229,M19231,M19232<br>M19239,M19241,M19242,M19249,M19271,M19272,M19279,M1929,M1990<br>M1991,M1992,M1993 | |
| Osteomalacia | M830,M831,M832,M833,M834,M835,M838,M839 | 2682 |
| Osteomyelitis | A0105,A0224,A5443,H05021,H05022,H05023,H05029,M4620,M4621<br>M4622,M4623,M4624,M4625,M4626,M4627,M4628,M8600,M86011,M86012<br>M86019,M86021,M86022,M86029,M86031,M86032,M86039,M86041<br>M86042,M86049,M86051,M86052,M86059,M86061,M86062,M86069<br>M86071,M86072,M86079,M8608,M8609,M8610,M86111,M86112,M86119<br>M86121,M86122,M86129,M86131,M86132,M86139,M86141,M86142<br>M86149,M86151,M86152,M86159,M86161,M86162,M86169,M86171<br>M86172,M86179,M8618,M8619,M8620,M86211,M86212,M86219,M86221<br>M86222,M86229,M86231,M86232,M86239,M86241,M86242,M86249<br>M86251,M86252,M86259,M86261,M86262,M86269,M86271,M86272<br>M86279,M8628,M8629,M8630,M86311,M86312,M86319,M86321,M86322<br>M86329,M86331,M86332,M86339,M86341,M86342,M86349,M86351<br>M86352,M86359,M86361,M86362,M86369,M86371,M86372,M86379,M8638<br>M8639,M8640,M86411,M86412,M86419,M86421,M86422,M86429,M86431<br>M86432,M86439,M86441,M86442,M86449,M86451,M86452,M86459<br>M86461,M86462,M86469,M86471,M86472,M86479,M8648,M8649,M8650<br>M86511,M86512,M86519,M86521,M86522,M86529,M86531,M86532<br>M86539,M86541,M86542,M86549,M86551,M86552,M86559,M86561<br>M86562,M86569,M86571,M86572,M86579,M8658,M8659,M8660,M86611<br>M86612,M86619,M86621,M86622,M86629,M86631,M86632,M86639<br>M86641,M86642,M86649,M86651,M86652,M86659,M86661,M86662<br>M86669,M86671,M86672,M86679,M8668,M8669,M868X0,M868X1,M868X2<br>M868X3,M868X4,M868X5,M868X6,M868X7,M868X8,M868X9,M869 | 0020,00324,09859,37603,73000,73001,73002,73003,73004,73005<br>73006,73007,73008,73009,73010,73011,73012,73013,73014,73015<br>73016,73017,73018,73019,73020,73028,73030,73090,73098 |
| Osteoporosis | M8000XA,M80011A,M80012A,M80019A,M80021A,M80022A,M80029A<br>M80031A,M80032A,M80039A,M80041A,M80042A,M80049A,M80051A<br>M80052A,M80059A,M80061A,M80062A,M80069A,M80071A,M80072A<br>M80079A,M8008XA,M800AXA,M8080XA,M80811A,M80812A,M80819A<br>M80821A,M80822A,M80829A,M80831A,M80832A,M80839A,M80841A<br>M80842A,M80849A,M80851A,M80852A,M80859A,M80861A,M80862A<br>M80869A,M80871A,M80872A,M80879A,M8088XA,M808AXA,M810,M816<br>M818 | 73300,73301,73302,73303,73309,73310,73311,73312,73313,73314<br>73315,73316,73319 |

Continued on next page

**Supplementary Table S1:** (cont'd) A comprehensive mapping of diagnosis groups used in the experiments to their corresponding ICD-10 and ICD-9 codes.

| Diagnosis | ICD-10 | ICD-9 |
|---|---|---|
| Other aftercare encounter | Z09,Z430,Z431,Z432,Z433,Z434,Z435,Z436,Z437,Z438,Z439,Z471<br>Z472,Z4731,Z4732,Z4733,Z4781,Z4782,Z4789,Z4800,Z4801,Z4802<br>Z4803,Z481,Z4821,Z4822,Z4823,Z4824,Z48280,Z48288,Z48290<br>Z48298,Z48810,Z48811,Z48812,Z48813,Z48814,Z48815,Z48816<br>Z48817,Z4889,Z4901,Z4902,Z4931,Z4932,Z515,Z516,Z5181,Z5189 | V420,V421,V426,V427,V4281,V4289,V429,V5401,V5481,V5482,V5489<br>V550,V551,V552,V553,V554,V555,V556,V557,V558,V559,V561,V562<br>V5631,V5632,V5830,V5831,V5832,V5841,V5844,V5849,V5871,V5872<br>V5873,V5874,V5875,V5876,V5877,V5883,V5889,V589,V667,V6700<br>V6709,V671,V672,V673,V674,V6751,V6759,V676,V679 |
| Other and ill-defined cerebrovascular disease | A5204,A5205,G460,G461,G462,G463,G464,G465,G466,G467,G468,I671<br>I672,I673,I675,I676,I677,I6781,I6782,I6783,I67841,I67848<br>I67850,I67858,I6789,I679,I680,I682,I688 | 0463,09389,34839,4358,4359,436,4370,4371,4373,4374,4375,4376<br>4378,4379 |
| Other and ill-defined heart disease | I253,I2541,I510,I511,I512,I513,I515,I517,I5181,I5189,I519,I52<br>I5A,M0530,M05311,M05312,M05319,M05321,M05322,M05329,M05331<br>M05332,M05339,M05341,M05342,M05349,M05351,M05352,M05359<br>M05361,M05362,M05369,M05371,M05372,M05379,M0539 | 41410,41411,41419,4291,4293,4295,4296,42971,42979,42981,42982<br>42983,42989,4299,7142 |
| Other general signs and symptoms | R252,R293,R294,R296,R29890,R29891,R29898,R2991,R490,R491<br>R4921,R4922,R498,R499,R52,R54,R590,R591,R599,R600,R601,R609<br>R61,R620,R6250,R6251,R6252,R6259,R627,R630,R631,R632,R633<br>R6330,R6331,R6332,R6339,R634,R635,R636,R638,R64,R6510,R6511<br>R680,R6811,R6812,R6813,R6819,R682,R683,R6881,R6882,R6883<br>R6884,R6889,R69,R99 | 33819,5277,71965,72982,72989,78064,78065,7808,78091,78092<br>78094,78096,78099,7810,7815,78191,78192,78193,78199,7823,7830<br>7831,78321,78322,7833,78340,78341,78342,78343,7835,7836,7837<br>7839,78440,78441,78442,78443,78444,78449,78492,7856,7964,7969<br>797,7980,7994,79981,79982,79989,7999,99593,99594,V419 |
| Other nervous system disorders (neither hereditary nor degenerative) | A1789,A179,B2701,B2711,B2781,B2791,E0840,E0844,E0940,E0944<br>E1040,E1044,E1140,E1144,E1340,E1344,G08,G130,G131,G132,G138<br>G360,G361,G368,G369,G370,G371,G372,G373,G374,G375,G378,G379<br>G460,G461,G462,G463,G464,G465,G466,G467,G468,G64,G908,G909<br>G90A,G910,G911,G912,G913,G914,G918,G919,G92,G9200,G9201,G9202<br>G9203,G9204,G9205,G928,G929,G930,G932,G933,G9331,G9332,G9339<br>G9340,G9341,G9349,G935,G936,G937,G9381,G9382,G9389,G939,G94<br>G950,G9511,G9519,G9520,G9529,G9581,G9589,G959,G960,G9600<br>G9601,G9602,G9608,G9609,G9611,G9612,G9619,G96191,G96198,G968<br>G96810,G96811,G96819,G9689,G969,G980,G988,G998,I673,I674<br>J1081,J1181,K7682,M3506,M3507 | 01380,01790,0463,075,24960,25060,25061,32371,32372,3239,325<br>3313,3314,3315,3317,33181,3360,3361,3368,3369,3379,3410,3411<br>34120,34121,3418,3419,3480,3482,34830,34831,34839,3484,3485<br>34881,34882,34889,3489,3492,34939,34981,34982,34989,3499,3535<br>3572,3573,35789,38861,4358,4372,4378,4878,78079 |
| Other nervous system disorders (often hereditary or degenerative) | E7500,E7501,E7502,E7509,E7510,E7511,E7519,E7523,E7525,E7526<br>E7529,E754,G10,G110,G111,G1110,G1111,G1119,G112,G113,G114<br>G118,G119,G120,G121,G1220,G1221,G1222,G1223,G1224,G1225,G1229<br>G128,G129,G210,G2111,G2119,G212,G213,G214,G218,G219,G230,G231<br>G232,G238,G239,G2401,G2402,G2409,G241,G242,G243,G244,G245 | 3300,3301,3308,3316,3317,33183,33189,3319,3320,3321,3330,3331<br>3332,3333,3334,3335,3336,33372,33379,33381,33382,33383,33384<br>33385,33389,33390,33391,33392,33393,33394,33399,3340,3341<br>3342,3343,3344,3348,3349,3350,33510,33511,33519,33520,33521<br>33522,33524,33529,3359,3362,3363,33700,33701,33709,3371,3373 |

Continued on next page

**Supplementary Table S1:** (cont'd) A comprehensive mapping of diagnosis groups used in the experiments to their corresponding ICD-10 and ICD-9 codes.

| Diagnosis | ICD-10 | ICD-9 |
|---|---|---|
| | G248,G249,G250,G251,G252,G253,G254,G255,G2561,G2569,G2570 G2571,G2579,G2581,G2582,G2583,G2589,G259,G26,G312,G3181,G3182 G3184,G3185,G3189,G319,G320,G3281,G3289,G9001,G9009,G901,G903 G904,G990,G992 | 34989,7428 |
| Other specified CNS infection and poliomyelitis | A5210,A5211,A5212,A5217,A5219,A522,A523,A800,A801,A802,A8030 A8039,A804,A809,A8100,A8101,A8109,A812,A8181,A8182,A8183 A8189,A819,A820,A821,A829,A880,A881,A888,A89,G14 | 04510,04520,04590,0460,04611,04619,0463,04671,04672,04679 0468,0469,048,0498,0499,071,07881,0940,0941,0943,09482,09483 09489,0949,138 |
| Other specified and unspecified circulatory disease | A5200,A5202,A5209,E0851,E0951,E1051,E1151,E1351,I770,I771 I772,I773,I774,I775,I776,I77810,I77811,I77812,I77819,I7782 I7789,I779,I780,I781,I788,I789,I791,I798,I998,I999 | 0931,09389,0939,24970,25070,25071,44381,4470,4471,4472,4473 4474,4475,4476,44770,44771,44772,44773,4478,4479,4480,4481 4489,45989,4599 |
| Other specified and unspecified congenital anomalies | P092,P093,Q800,Q801,Q802,Q803,Q804,Q808,Q809,Q810,Q811,Q812 Q818,Q819,Q820,Q821,Q822,Q823,Q824,Q825,Q826,Q828,Q829,Q830 Q831,Q832,Q833,Q838,Q839,Q840,Q841,Q842,Q843,Q844,Q845,Q846 Q848,Q849,Q851,Q858,Q8581,Q8582,Q8583,Q8589,Q859,Q860,Q861 Q862,Q868,Q870,Q871,Q8711,Q8719,Q872,Q873,Q8740,Q87410,Q87418 Q8742,Q8743,Q875,Q8781,Q8782,Q8789,Q8901,Q8909,Q891,Q892,Q893 Q894,Q897,Q898,Q899 | 75555,7560,7570,7571,7572,75731,75732,75733,75739,7574,7575 7576,7578,7579,7590,7591,7592,7593,7594,7595,7596,7597,75981 75982,75989,7599,76071,76077,76079 |
| Other specified and unspecified diseases of bladder and urethra | A1812,N310,N311,N312,N318,N319,N320,N321,N322,N323,N3281 N3289,N329,N33,N35010,N35011,N35012,N35013,N35014,N35016 N35021,N35028,N35111,N35112,N35113,N35114,N35116,N35119,N3512 N358,N35811,N35812,N35813,N35814,N35816,N35819,N3582,N359 N35911,N35912,N35913,N35914,N35916,N35919,N3592,N360,N361 N362,N3641,N3642,N3643,N3644,N365,N368,N369,N37,N99110,N99111 N99112,N99113,N99114,N99115,N99116,N9912 | 01610,5960,5961,5962,5963,5964,59651,59652,59653,59654,59655 59659,5966,5967,59689,5969,59800,59801,5981,5982,5988,5989 5991,5992,5993,5994,5995,59981,59982,59984,5999 |
| Other specified and unspecified diseases of kidney and ureters | A1811,A5421,N130,N131,N132,N1330,N1339,N134,N135,N138,N139 N23,N250,N251,N2581,N2589,N259,N261,N262,N270,N271,N279,N280 N281,N2881,N2882,N2883,N2884,N2885,N2886,N2889,N289,N29 O26831,O26832,O26833,O26839 | 01600,01620,09819,40591,587,5880,5881,58881,58889,5889,5890 5891,5899,5903,591,5930,5931,5932,5933,5934,5935,59373,59381 59382,59389,5939,59960,59969,64620,64621,64622,64623,7880 |
| Other specified and unspecified disorders of stomach and duodenum | K30,K310,K311,K312,K313,K314,K316,K317,K31811,K31819,K3182 K3183,K3184,K3189,K319,K31A0,K31A11,K31A12,K31A13,K31A14 K31A15,K31A19,K31A21,K31A22,K31A29 | 2111,5360,5361,5363,5368,5369,5370,5371,5374,5375,5376,53781 53782,53783,53784,53789,5379 |
| Other specified and | A186,B3784,H6000,H6001,H6002,H6003,H6010,H6011,H6012,H6013 | 01740,11282,31532,38000,38001,38002,38003,38010,38011,38012 |

Continued on next page

**Supplementary Table S1:** (cont'd) A comprehensive mapping of diagnosis groups used in the experiments to their corresponding ICD-10 and ICD-9 codes.

| Diagnosis | ICD-10 | ICD-9 |
|---|---|---|
| unspecified disorders of the ear | H6020,H6021,H6022,H6023,H60311,H60312,H60313,H60319,H60321 H60322,H60323,H60329,H60331,H60332,H60333,H60339,H60391 H60392,H60393,H60399,H6040,H6041,H6042,H6043,H60501,H60502 H60503,H60509,H60511,H60512,H60513,H60519,H60521,H60522 H60523,H60529,H60531,H60532,H60533,H60539,H60541,H60542 H60543,H60549,H60551,H60552,H60553,H60559,H60591,H60592 H60593,H60599,H6060,H6061,H6062,H6063,H608X1,H608X2,H608X3 H608X9,H6090,H6091,H6092,H6093,H61001,H61002,H61003,H61009 H61011,H61012,H61013,H61019,H61021,H61022,H61023,H61029 H61031,H61032,H61033,H61039,H61101,H61102,H61103,H61109 H61111,H61112,H61113,H61119,H61121,H61122,H61123,H61129 H61191,H61192,H61193,H61199,H6120,H6121,H6122,H6123,H61301 H61302,H61303,H61309,H61311,H61312,H61313,H61319,H61321 H61322,H61323,H61329,H61391,H61392,H61393,H61399,H61811 H61812,H61813,H61819,H61891,H61892,H61893,H61899,H6190,H6191 H6192,H6193,H6240,H6241,H6242,H6243,H628X1,H628X2,H628X3 H628X9,H9201,H9202,H9203,H9209,H9210,H9211,H9212,H9213,H9220 H9221,H9222,H9223,H93091,H93092,H93093,H93099,H9311,H9312 H9313,H9319,H93211,H93212,H93213,H93219,H93221,H93222,H93223 H93229,H93231,H93232,H93233,H93239,H93241,H93242,H93243 H93249,H9325,H93291,H93292,H93293,H93299,H933X1,H933X2,H933X3 H933X9,H938X1,H938X2,H938X3,H938X9,H9390,H9391,H9392,H9393 H93A1,H93A2,H93A3,H93A9,H9400,H9401,H9402,H9403,H9480,H9481 H9482,H9483 | 38013,38014,38015,38021,38022,38023,38030,38031,38032,38039 3804,38050,38051,38052,38053,38081,38089,3809,38800,38830 38831,38832,38840,38841,38842,38844,3885,38860,38869,38870 3888,3889,V413 |
| Other specified and unspecified endocrine disorders | A187,A1882,A391,E15,E160,E161,E162,E163,E164,E168,E169,E200 E201,E208,E209,E210,E211,E212,E213,E214,E215,E241,E242,E243 E244,E248,E249,E250,E258,E259,E2601,E2602,E2609,E261,E2681 E2689,E269,E270,E271,E272,E273,E2740,E2749,E275,E278,E279 E280,E281,E282,E28310,E28319,E2839,E288,E289,E290,E291,E298 E299,E300,E301,E308,E309,E310,E311,E3120,E3121,E3122,E3123 E318,E319,E320,E321,E328,E329,E340,E341,E342,E343,E3430,E3431 E34321,E34322,E34328,E34329,E3439,E344,E3450,E3451,E3452,E348 E349,E35 | 01760,01790,0363,2468,2510,2511,2512,2514,2515,2518,2519 25200,25201,25202,25208,2521,2528,2529,2530,2538,2540,2541 2548,2549,2550,25510,25511,25512,25513,25514,2552,2553,25541 25542,2556,2558,2559,2560,2561,25631,25639,2564,2568,2569 2570,2572,2578,2579,25801,25802,25803,2581,2588,2589,2590 2591,2592,2593,2594,25950,25951,25952,2598,2599,27549 |
| Other specified and unspecified gastrointestinal disorders | A1883,B5732,K550,K55011,K55012,K55019,K55021,K55022,K55029 K55031,K55032,K55039,K55041,K55042,K55049,K55051,K55052 K55059,K55061,K55062,K55069,K551,K5520,K5521,K5530,K5531 K5532,K5533,K558,K559,K580,K581,K582,K588,K589,K5900,K5901 K5902,K5903,K5904,K5909,K591,K592,K593,K5931,K5939,K598,K5981 | 01480,0402,0861,538,5570,5571,5579,56400,56401,56402,56409 5641,5645,5647,56481,56489,5649,56739,5680,56881,56889,5689 56981,56982,56984,56985,56986,56989,5699,5790,5791,5792,5798 5799 |

Continued on next page

**Supplementary Table S1:** (cont'd) A comprehensive mapping of diagnosis groups used in the experiments to their corresponding ICD-10 and ICD-9 codes.

| Diagnosis | ICD-10 | ICD-9 |
|---|---|---|
| | K5989,K599,K632,K633,K634,K6381,K6389,K639,K660,K661,K668<br>K669,K689,K900,K901,K902,K904,K9041,K9049,K9081,K9089,K909<br>K9281,K9289,K929,M3508 | |
| Other specified and unspecified hematologic conditions | A1885,B510,D730,D731,D732,D733,D734,D735,D7389,D739,D740,D748<br>D749,D750,D751,D7581,D7589,D759,D75A,D77,M363 | 01770,0849,2890,2894,28950,28951,28959,2896,2897,28983,28989<br>2899,7132 |
| Other specified and unspecified liver disease | A5274,B670,B675,K700,K702,K7030,K7031,K709,K710,K7110,K7111<br>K717,K718,K719,K740,K7400,K7401,K7402,K741,K742,K743,K744<br>K745,K7460,K7469,K751,K7589,K759,K760,K761,K762,K763,K764<br>K765,K766,K767,K7681,K7682,K7689,K769,K77,O26611,O26612<br>O26613,O26619,O2662,O2663 | 0952,0953,1220,1225,570,5710,5712,5713,5715,5716,5718,5719<br>5721,5722,5723,5724,5730,5733,5734,5735,5738,5739,64670,64671<br>64673,64684 |
| Other specified and unspecified lower respiratory disease | A065,A157,A158,A159,B400,B401,B402,B410,B420,B440,B441,B450<br>B460,B671,J182,J22,J810,J811,J82,J8281,J8282,J8283,J8289<br>J8401,J8402,J8403,J8409,J8410,J84111,J84112,J84113,J84114<br>J84115,J84116,J84117,J8417,J84170,J84178,J842,J8481,J8482<br>J8483,J84841,J84842,J84843,J84848,J8489,J849,J852,J9801,J9809<br>J984,J986,J988,J989,J99,M0510,M05111,M05112,M05119,M05121<br>M05122,M05129,M05131,M05132,M05139,M05141,M05142,M05149<br>M05151,M05152,M05159,M05161,M05162,M05169,M05171,M05172<br>M05179,M0519 | 0064,01090,01280,1160,1161,1171,1173,1175,1177,1221,4846,5130<br>514,515,5160,5161,5162,51630,51631,51632,51633,51634,51635<br>51636,51637,5164,5165,51661,51662,51663,51664,51669,5168,5169<br>5178,5183,5184,51889,51911,51919,5194,5198,5199,71481 |
| Other specified and unspecified mood disorders | F0630,F348,F3481,F3489,F349,F39 | 29383,29690,29699 |
| Other specified and unspecified nutritional and metabolic disorders | D813,D8130,D8131,D8132,D8139,D815,D81810,D81818,D81819,E65<br>E663,E670,E671,E672,E673,E678,E700,E701,E7020,E7021,E7029<br>E7030,E70310,E70311,E70318,E70319,E70320,E70321,E70328,E70329<br>E70330,E70331,E70338,E70339,E7039,E7040,E7041,E7049,E705,E708<br>E7081,E7089,E709,E710,E71110,E71111,E71118,E71120,E71121<br>E71128,E7119,E712,E7130,E71310,E71311,E71312,E71313,E71314<br>E71318,E7132,E7139,E7140,E7141,E7142,E7143,E71440,E71448<br>E7150,E71510,E71511,E71518,E71520,E71521,E71522,E71528,E71529<br>E7153,E71540,E71541,E71542,E71548,E7200,E7201,E7202,E7203<br>E7204,E7209,E7210,E7211,E7212,E7219,E7220,E7221,E7222,E7223<br>E7229,E723,E724,E7250,E7251,E7252,E7253,E7259,E728,E7281 | 2662,2700,2701,2702,2703,2704,2705,2706,2707,2708,2709,2710<br>2711,2712,2713,2714,2718,2719,2725,2726,2727,2728,2729,2734<br>2738,27501,27502,27503,27509,2751,2752,2753,27540,27541,27542<br>27549,2755,2758,2759,2771,2772,27730,27731,27739,2774,2775<br>2776,2777,27781,27782,27783,27784,27785,27786,27787,27788<br>27789,2779,27802,2781,2782,2783,2784,2788,3300,3301,6868<br>75989,7906 |

Continued on next page

**Supplementary Table S1:** (cont'd) A comprehensive mapping of diagnosis groups used in the experiments to their corresponding ICD-10 and ICD-9 codes.

| Diagnosis | ICD-10 | ICD-9 |
|---|---|---|
| | E7289,E729,E730,E731,E738,E739,E7400,E7401,E7402,E7403,E7404 | |
| | E7409,E7410,E7411,E7412,E7419,E7420,E7421,E7429,E7431,E7439 | |
| | E744,E748,E74810,E74818,E74819,E7489,E749,E7500,E7501,E7502 | |
| | E7509,E7510,E7511,E7519,E7521,E7522,E7523,E75240,E75241 | |
| | E75242,E75243,E75244,E75248,E75249,E7525,E7526,E7529,E753 | |
| | E754,E755,E756,E7601,E7602,E7603,E761,E76210,E76211,E76219 | |
| | E7622,E7629,E763,E768,E769,E770,E771,E778,E779,E786,E7870 | |
| | E7871,E7872,E7879,E7881,E7889,E789,E790,E791,E792,E798,E799 | |
| | E800,E801,E8020,E8021,E8029,E803,E804,E805,E806,E807,E8300 | |
| | E8301,E8309,E8310,E83110,E83111,E83118,E83119,E8319,E832 | |
| | E8330,E8331,E8332,E8339,E8340,E8341,E8342,E8349,E8350,E8351 | |
| | E8352,E8359,E8381,E8389,E839,E850,E851,E852,E853,E854,E858 | |
| | E8581,E8582,E8589,E859,E8801,E8802,E8809,E881,E882,E883,E8840 | |
| | E8841,E8842,E8849,E8881,E8889,E889,P091 | |
| Other specified and | P080,P081,P0821,P0822,P09,P091,P092,P093,P094,P095,P096,P098 | 74783,7608,7660,7661,76621,76622,7706,7750,7751,7752,7753 |
| unspecified perinatal | P099,P221,P290,P2911,P2912,P292,P293,P2930,P2938,P294,P2981 | 7754,7755,7756,7757,77589,7759,7780,7781,7782,7783,7784,7785 |
| conditions | P2989,P299,P700,P701,P702,P703,P704,P708,P709,P710,P711,P712 | 7786,7787,7788,7789,7794,77981,77982,77983,77984,77985,77989 |
| | P713,P714,P718,P719,P720,P721,P722,P728,P729,P740,P741,P742 | 7799,7966 |
| | P7421,P7422,P743,P7431,P7432,P744,P7441,P74421,P74422,P7449 | |
| | P745,P746,P748,P749,P800,P808,P809,P810,P818,P819,P830,P831 | |
| | P832,P8330,P8339,P834,P835,P836,P838,P8381,P8388,P839,P930 | |
| | P938,P940,P941,P942,P948,P949,P95,P960,P963,P965,P9681,P9682 | |
| | P9683,P9689,P969 | |
| Other specified and | A5132,B000,B001,B372,B383,B403,B421,B430,B432,B452,B463,B781 | 0540,05473,09182,1123,1141,1160,1171,1172,1175,1177,1360,6861 |
| unspecified skin | L300,L301,L302,L308,L309,L560,L561,L562,L563,L564,L565,L570 | 6868,69272,69273,69275,69282,6929,700,7010,7011,7012,7013 |
| disorders | L571,L572,L573,L574,L575,L600,L601,L602,L603,L604,L605,L608 | 7014,7015,7018,7019,7020,70211,70219,7030,7038,7039,70400 |
| | L609,L62,L630,L631,L632,L638,L639,L640,L648,L649,L650,L651 | 70401,70402,70409,7041,7042,7043,70441,70442,7048,7049,7050 |
| | L652,L658,L659,L660,L662,L663,L664,L668,L669,L670,L671,L678 | 7051,70521,70522,70581,70582,70589,7059,7061,7062,7068,7069 |
| | L679,L680,L681,L682,L683,L688,L689,L720,L7211,L7212,L722,L723 | 70900,70901,70909,7092,7093,7094,7098,7099 |
| | L728,L729,L730,L731,L739,L740,L741,L742,L743,L744,L74510 | |
| | L74511,L74512,L74513,L74519,L7452,L748,L749,L750,L751,L752 | |
| | L758,L759,L80,L810,L811,L812,L813,L814,L815,L816,L817,L818 | |
| | L819,L820,L821,L83,L84,L850,L851,L852,L853,L858,L859,L86,L870 | |
| | L871,L872,L878,L879,L900,L901,L902,L903,L904,L905,L906,L908 | |
| | L909,L910,L918,L919,L921,L922,L923,L928,L929,L940,L941,L942 | |
| | L943,L944,L946,L948,L949,L980,L985,L986,L987,L988,L989,L99 | |
| Other specified and | A5005,J300,J301,J302,J305,J3081,J3089,J309,J310,J311,J312 | 0900,470,4710,4711,4718,4719,4720,4721,4722,4760,4761,4770 |

Continued on next page

**Supplementary Table S1:** (cont'd) A comprehensive mapping of diagnosis groups used in the experiments to their corresponding ICD-10 and ICD-9 codes.

| Diagnosis | ICD-10 | ICD-9 |
|---|---|---|
| unspecified upper | J330,J331,J338,J339,J340,J341,J342,J343,J3481,J3489,J349,J370 | 4771,4772,4778,4779,4780,47811,47819,47825,47826,47829,47830 |
| respiratory disease | J371,J3800,J3801,J3802,J381,J382,J383,J384,J385,J386,J387 | 47831,47832,47833,47834,4784,4785,4786,47871,47874,47875 |
| | J392,J393,J398,J399 | 47879,4788,4789,51919 |
| Other specified bone | A5002,A5051,A5055,A5056,A5146,A5216,A5277,B672,L4052,M830 | 0900,0905,09161,0940,0955,1223,2682,6960,7135,73081,73082 |
| disease and | M831,M832,M833,M834,M838,M839,M8480,M84811,M84812,M84819 | 73083,73084,73085,73086,73087,73088,73089,7310,7311,7312,7313 |
| musculoskeletal | M84821,M84822,M84829,M84831,M84832,M84833,M84834,M84839 | 7318,7321,7322,7323,7324,7325,7326,7327,7328,7329,73321,73322 |
| deformities | M84841,M84842,M84849,M84851,M84852,M84859,M84861,M84862 | 73329,7333,7335,7336,7337,73390,73391,73392,73399,73811,73812 |
| | M84863,M84864,M84869,M84871,M84872,M84879,M8488,M849,M8500 | V454 |
| | M85011,M85012,M85019,M85021,M85022,M85029,M85031,M85032 | |
| | M85039,M85041,M85042,M85049,M85051,M85052,M85059,M85061 | |
| | M85062,M85069,M85071,M85072,M85079,M8508,M8509,M8510,M85111 | |
| | M85112,M85119,M85121,M85122,M85129,M85131,M85132,M85139 | |
| | M85141,M85142,M85149,M85151,M85152,M85159,M85161,M85162 | |
| | M85169,M85171,M85172,M85179,M8518,M8519,M852,M8530,M85311 | |
| | M85312,M85319,M85321,M85322,M85329,M85331,M85332,M85339 | |
| | M85341,M85342,M85349,M85351,M85352,M85359,M85361,M85362 | |
| | M85369,M85371,M85372,M85379,M8538,M8539,M8540,M85411,M85412 | |
| | M85419,M85421,M85422,M85429,M85431,M85432,M85439,M85441 | |
| | M85442,M85449,M85451,M85452,M85459,M85461,M85462,M85469 | |
| | M85471,M85472,M85479,M8548,M8550,M85511,M85512,M85519,M85521 | |
| | M85522,M85529,M85531,M85532,M85539,M85541,M85542,M85549 | |
| | M85551,M85552,M85559,M85561,M85562,M85569,M85571,M85572 | |
| | M85579,M8558,M8559,M8560,M85611,M85612,M85619,M85621,M85622 | |
| | M85629,M85631,M85632,M85639,M85641,M85642,M85649,M85651 | |
| | M85652,M85659,M85661,M85662,M85669,M85671,M85672,M85679,M8568 | |
| | M8569,M8580,M85811,M85812,M85819,M85821,M85822,M85829,M85831 | |
| | M85832,M85839,M85841,M85842,M85849,M85851,M85852,M85859 | |
| | M85861,M85862,M85869,M85871,M85872,M85879,M8588,M8589,M859 | |
| | M880,M881,M88811,M88812,M88819,M88821,M88822,M88829,M88831 | |
| | M88832,M88839,M88841,M88842,M88849,M88851,M88852,M88859 | |
| | M88861,M88862,M88869,M88871,M88872,M88879,M8888,M8889,M889 | |
| | M8900,M89011,M89012,M89019,M89021,M89022,M89029,M89031,M89032 | |
| | M89039,M89041,M89042,M89049,M89051,M89052,M89059,M89061 | |
| | M89062,M89069,M89071,M89072,M89079,M8908,M8909,M89121,M89122 | |
| | M89123,M89124,M89125,M89126,M89127,M89128,M89129,M89131 | |
| | M89132,M89133,M89134,M89138,M89139,M89151,M89152,M89153 | |
| | M89154,M89155,M89156,M89157,M89158,M89159,M89160,M89161 | |
| | M89162,M89163,M89164,M89165,M89166,M89167,M89168,M89169,M8918 | |
| | M8920,M89211,M89212,M89219,M89221,M89222,M89229,M89231,M89232 | |

Continued on next page

**Supplementary Table S1:** (cont'd) A comprehensive mapping of diagnosis groups used in the experiments to their corresponding ICD-10 and ICD-9 codes.

| Diagnosis | ICD-10 | ICD-9 |
|---|---|---|
| | M89233,M89234,M89239,M89241,M89242,M89249,M89251,M89252 | |
| | M89259,M89261,M89262,M89263,M89264,M89269,M89271,M89272 | |
| | M89279,M8928,M8929,M8930,M89311,M89312,M89319,M89321,M89322 | |
| | M89329,M89331,M89332,M89333,M89334,M89339,M89341,M89342 | |
| | M89349,M89351,M89352,M89359,M89361,M89362,M89363,M89364 | |
| | M89369,M89371,M89372,M89379,M8938,M8939,M8940,M89411,M89412 | |
| | M89419,M89421,M89422,M89429,M89431,M89432,M89439,M89441 | |
| | M89442,M89449,M89451,M89452,M89459,M89461,M89462,M89469 | |
| | M89471,M89472,M89479,M8948,M8949,M8950,M89511,M89512,M89519 | |
| | M89521,M89522,M89529,M89531,M89532,M89539,M89541,M89542 | |
| | M89549,M89551,M89552,M89559,M89561,M89562,M89569,M89571 | |
| | M89572,M89579,M8958,M8959,M8970,M89711,M89712,M89719,M89721 | |
| | M89722,M89729,M89731,M89732,M89739,M89741,M89742,M89749 | |
| | M89751,M89752,M89759,M89761,M89762,M89769,M89771,M89772 | |
| | M89779,M8978,M8979,M898X0,M898X1,M898X2,M898X3,M898X4,M898X5 | |
| | M898X6,M898X7,M898X8,M898X9,M899,M9060,M90611,M90612,M90619 | |
| | M90621,M90622,M90629,M90631,M90632,M90639,M90641,M90642 | |
| | M90649,M90651,M90652,M90659,M90661,M90662,M90669,M90671 | |
| | M90672,M90679,M9068,M9069,M9080,M90811,M90812,M90819,M90821 | |
| | M90822,M90829,M90831,M90832,M90839,M90841,M90842,M90849 | |
| | M90851,M90852,M90859,M90861,M90862,M90869,M90871,M90872 | |
| | M90879,M9088,M9089,M910,M9110,M9111,M9112,M9120,M9121,M9122 | |
| | M9130,M9131,M9132,M9140,M9141,M9142,M9180,M9181,M9182,M9190 | |
| | M9191,M9192,M9200,M9201,M9202,M9210,M9211,M9212,M92201,M92202 | |
| | M92209,M92211,M92212,M92219,M92221,M92222,M92229,M92291 | |
| | M92292,M92299,M9230,M9231,M9232,M9240,M9241,M9242,M9250 | |
| | M92501,M92502,M92503,M92509,M9251,M92511,M92512,M92513,M92519 | |
| | M9252,M92521,M92522,M92523,M92529,M92591,M92592,M92593,M92599 | |
| | M9260,M9261,M9262,M9270,M9271,M9272,M928,M929,M93001,M93002 | |
| | M93003,M93004,M93011,M93012,M93013,M93014,M93021,M93022 | |
| | M93023,M93024,M93031,M93032,M93033,M93034,M93041,M93042 | |
| | M93043,M93044,M93051,M93052,M93053,M93054,M93061,M93062 | |
| | M93063,M93064,M93071,M93072,M93073,M93074,M931,M9320,M93211 | |
| | M93212,M93219,M93221,M93222,M93229,M93231,M93232,M93239 | |
| | M93241,M93242,M93249,M93251,M93252,M93259,M93261,M93262 | |
| | M93269,M93271,M93272,M93279,M9328,M9329,M9380,M93811,M93812 | |
| | M93819,M93821,M93822,M93829,M93831,M93832,M93839,M93841 | |
| | M93842,M93849,M93851,M93852,M93859,M93861,M93862,M93869 | |
| | M93871,M93872,M93879,M9388,M9389,M9390,M93911,M93912,M93919 | |
| | M93921,M93922,M93929,M93931,M93932,M93939,M93941,M93942 | |

Continued on next page

**Supplementary Table S1:** (cont'd) A comprehensive mapping of diagnosis groups used in the experiments to their corresponding ICD-10 and ICD-9 codes.

| Diagnosis | ICD-10 | ICD-9 |
|---|---|---|
| | M93949,M93951,M93952,M93959,M93961,M93962,M93969,M93971 M93972,M93979,M9398,M9399,M940,M941,M9420,M94211,M94212 M94219,M94221,M94222,M94229,M94231,M94232,M94239,M94241 M94242,M94249,M94251,M94252,M94259,M94261,M94262,M94269 M94271,M94272,M94279,M9428,M9429,M94351,M94352,M94359,M948X0 M948X1,M948X2,M948X3,M948X4,M948X5,M948X6,M948X7,M948X8 M948X9,M949,M960 | |
| Other specified chronic arthropathy | L4050,L4051,L4054,L4059,M1200,M12011,M12012,M12019,M12021 M12022,M12029,M12031,M12032,M12039,M12041,M12042,M12049 M12051,M12052,M12059,M12061,M12062,M12069,M12071,M12072 M12079,M1208,M1209 | 6960,7144 |
| Other specified complications in pregnancy | O1200,O1201,O1202,O1203,O1210,O1211,O1212,O1213,O1220,O1221 O1222,O1223,O210,O211,O212,O218,O219,O2200,O2201,O2202,O2203 O2210,O2211,O2212,O2213,O2220,O2221,O2222,O2223,O2230,O2231 O2232,O2233,O2240,O2241,O2242,O2243,O2250,O2251,O2252,O2253 O228X1,O228X2,O228X3,O228X9,O2290,O2291,O2292,O2293,O2300 O2301,O2302,O2303,O2310,O2311,O2312,O2313,O2320,O2321,O2322 O2323,O2330,O2331,O2332,O2333,O2340,O2341,O2342,O2343,O23511 O23512,O23513,O23519,O23521,O23522,O23523,O23529,O23591 O23592,O23593,O23599,O2390,O2391,O2392,O2393,O2510,O2511 O2512,O2513,O2600,O2601,O2602,O2603,O2610,O2611,O2612,O2613 O2640,O2641,O2642,O2643,O2650,O2651,O2652,O2653,O26611,O26612 O26613,O26619,O26711,O26712,O26713,O26719,O26811,O26812 O26813,O26819,O26821,O26822,O26823,O26829,O26831,O26832 O26833,O26839,O26841,O26842,O26843,O26849,O2686,O26891,O26892 O26893,O26899,O2690,O2691,O2692,O2693,O280,O281,O282,O283 O284,O285,O288,O289,O364XX0,O364XX1,O364XX2,O364XX3,O364XX4 O364XX5,O364XX9,O88011,O88012,O88013,O88019,O88111,O88112 O88113,O88119,O88211,O88212,O88213,O88219,O88311,O88312 O88313,O88319,O88811,O88812,O88813,O88819,O903,O91011,O91012 O91013,O91019,O91111,O91112,O91113,O91119,O91211,O91212 O91213,O91219,O92011,O92012,O92013,O92019,O92111,O92112 O92113,O92119,O94,O98011,O98012,O98013,O98019,O98111,O98112 O98113,O98119,O98211,O98212,O98213,O98219,O98311,O98312 O98313,O98319,O98411,O98412,O98413,O98419,O98511,O98512 O98513,O98519,O98611,O98612,O98613,O98619,O98711,O98712 O98713,O98719,O98811,O98812,O98813,O98819,O98911,O98912 O98913,O98919,O99011,O99012,O99013,O99019,O99111,O99112 | 64303,64313,64323,64383,64393,64610,64611,64612,64613,64614 64620,64621,64622,64623,64640,64641,64642,64643,64660,64661 64662,64663,64670,64671,64673,64680,64681,64682,64683,64690 64693,64700,64701,64703,64710,64711,64713,64720,64721,64723 64730,64731,64733,64740,64760,64761,64763,64780,64781,64783 64790,64791,64793,64820,64821,64823,64830,64831,64833,64840 64841,64843,64890,64891,64893,64900,64901,64903,64910,64911 64913,64920,64921,64923,64930,64931,64933,64960,64961,64962 64963,64964,65641,65643,66920,66921,66922,66923,66924,67100 67101,67103,67110,67111,67112,67113,67114,67120,67121,67123 67130,67131,67133,67150,67151,67153,67180,67181,67183,67190 67191,67193,67300,67301,67303,67310,67311,67313,67320,67321 67323,67330,67331,67333,67380,67381,67382,67383,67400,67401 67403,67452,67454,67500,67501,67503,67510,67511,67513,67520 67521,67523,67600,67601,67603,67610,67611,67613,677,67903 7965,V0251 |

Continued on next page

**Supplementary Table S1:** (cont'd) A comprehensive mapping of diagnosis groups used in the experiments to their corresponding ICD-10 and ICD-9 codes.

| Diagnosis | ICD-10 | ICD-9 |
|---|---|---|
| | O99113,O99119,O99210,O99211,O99212,O99213,O99280,O99281 O99282,O99283,O99310,O99311,O99312,O99313,O99320,O99321 O99322,O99323,O99330,O99331,O99332,O99333,O99340,O99341 O99342,O99343,O99350,O99351,O99352,O99353,O99411,O99412 O99413,O99419,O99511,O99512,O99513,O99519,O99611,O99612 O99613,O99619,O99711,O99712,O99713,O99719,O99820,O99830 O99840,O99841,O99842,O99843,O9989,O99891,O9A111,O9A112,O9A113 O9A119,O9A211,O9A212,O9A213,O9A219,O9A311,O9A312,O9A313 O9A319,O9A411,O9A412,O9A413,O9A419,O9A511,O9A512,O9A513 O9A519 | |
| Other specified connective tissue disease | A1809,L945,M5400,M5401,M5402,M5403,M5404,M5405,M5406,M5407 M5408,M5409,M6020,M60211,M60212,M60219,M60221,M60222,M60229 M60231,M60232,M60239,M60241,M60242,M60249,M60251,M60252 M60259,M60261,M60262,M60269,M60271,M60272,M60279,M6028,M7010 M7011,M7012,M7020,M7021,M7022,M7030,M7031,M7032,M7040,M7041 M7042,M7050,M7051,M7052,M7060,M7061,M7062,M7070,M7071,M7072 M7080,M70811,M70812,M70819,M70821,M70822,M70829,M70831,M70832 M70839,M70841,M70842,M70849,M70851,M70852,M70859,M70861 M70862,M70869,M70871,M70872,M70879,M7088,M7089,M7090,M70911 M70912,M70919,M70921,M70922,M70929,M70931,M70932,M70939 M70941,M70942,M70949,M70951,M70952,M70959,M70961,M70962 M70969,M70971,M70972,M70979,M7098,M7099,M7100,M71011,M71012 M71019,M71021,M71022,M71029,M71031,M71032,M71039,M71041 M71042,M71049,M71051,M71052,M71059,M71061,M71062,M71069 M71071,M71072,M71079,M7108,M7109,M7110,M71111,M71112,M71119 M71121,M71122,M71129,M71131,M71132,M71139,M71141,M71142 M71149,M71151,M71152,M71159,M71161,M71162,M71169,M71171 M71172,M71179,M7118,M7119,M7130,M71311,M71312,M71319,M71321 M71322,M71329,M71331,M71332,M71339,M71341,M71342,M71349 M71351,M71352,M71359,M71371,M71372,M71379,M7138,M7139,M7140 M71421,M71422,M71429,M71431,M71432,M71439,M71441,M71442 M71449,M71451,M71452,M71459,M71461,M71462,M71469,M71471 M71472,M71479,M7148,M7149,M7150,M71521,M71522,M71529,M71531 M71532,M71539,M71541,M71542,M71549,M71551,M71552,M71559 M71561,M71562,M71569,M71571,M71572,M71579,M7158,M7180,M71811 M71812,M71819,M71821,M71822,M71829,M71831,M71832,M71839 M71841,M71842,M71849,M71851,M71852,M71859,M71861,M71862 M71869,M71871,M71872,M71879,M7188,M7189,M719,M720,M721,M722 M724,M726,M728,M729,M7500,M7501,M7502,M7540,M7541,M7542,M7550 | 01590,6962,7236,7248,7260,72610,72619,7262,72631,72632,72633 72639,7264,7265,72660,72662,72665,72670,72672,72673,7272,7273 72740,72749,72782,72789,7279,7286,72871,72879,72882,72886 72889,7290,7291,7292,72930,72931,7294,7296,72971,72972,72973 72979,72981,72990,72992,72999 |

Continued on next page

**Supplementary Table S1:** (cont'd) A comprehensive mapping of diagnosis groups used in the experiments to their corresponding ICD-10 and ICD-9 codes.

| Diagnosis | ICD-10 | ICD-9 |
|---|---|---|
| | M7551,M7552,M7580,M7581,M7582,M7590,M7591,M7592,M7620,M7621<br>M7622,M7630,M7631,M7632,M7640,M7641,M7642,M76811,M76812<br>M76819,M7700,M7701,M7702,M7710,M7711,M7712,M7720,M7721,M7722<br>M7730,M7731,M7732,M7740,M7741,M7742,M790,M792,M793,M794,M795<br>M797,M7981,M7989,M799,M79A11,M79A12,M79A19,M79A21,M79A22<br>M79A29,M79A3,M79A9 | |
| Other specified diseases of veins and lymphatics | I8500,I8501,I8510,I8511,I860,I861,I862,I863,I864,I868,I878<br>I879,I880,I881,I888,I889,I890,I891,I898,I899,O2210,O2211<br>O2212,O2213 | 2891,2892,2893,4560,4561,45620,45621,4563,4564,4565,4566,4568<br>4571,4572,4578,4579,45981,45989,67110,67111,67112,67113,67114 |
| Other specified encounters and counseling | Z322,Z323,Z710,Z713,Z7141,Z7142,Z7151,Z7152,Z716,Z717,Z7181<br>Z7182,Z7183,Z7184,Z7185,Z7187,Z7188,Z7189,Z719,Z760,Z761,Z762<br>Z7689,Z9912 | V200,V201,V4612,V6519,V653,V6540,V6542,V6544,V6549,V658,V659<br>V681 |
| Other specified eye disorders | A1850,A1854,A1859,A211,A5001,A5030,A5039,A5143,A5271,A5430<br>A5432,A5439,A710,A711,A719,B0050,B0051,B0059,B0230,B0232<br>B0239,B5800,B5809,B691,B7300,B7301,B7309,H2100,H2101,H2102<br>H2103,H211X1,H211X2,H211X3,H211X9,H21211,H21212,H21213,H21219<br>H21221,H21222,H21223,H21229,H21231,H21232,H21233,H21239<br>H21241,H21242,H21243,H21249,H21251,H21252,H21253,H21259<br>H21261,H21262,H21263,H21269,H21271,H21272,H21273,H21279,H2129<br>H21301,H21302,H21303,H21309,H21311,H21312,H21313,H21319<br>H21321,H21322,H21323,H21329,H21331,H21332,H21333,H21339<br>H21341,H21342,H21343,H21349,H21351,H21352,H21353,H21359,H2140<br>H2141,H2142,H2143,H21501,H21502,H21503,H21509,H21511,H21512<br>H21513,H21519,H21521,H21522,H21523,H21529,H21531,H21532<br>H21533,H21539,H21541,H21542,H21543,H21549,H21551,H21552<br>H21553,H21559,H21561,H21562,H21563,H21569,H2181,H2182,H2189<br>H219,H22,H4420,H4421,H4422,H4423,H442A1,H442A2,H442A3,H442A9<br>H442B1,H442B2,H442B3,H442B9,H442C1,H442C2,H442C3,H442C9<br>H442D1,H442D2,H442D3,H442D9,H442E1,H442E2,H442E3,H442E9,H4430<br>H44311,H44312,H44313,H44319,H44321,H44322,H44323,H44329<br>H44391,H44392,H44393,H44399,H4440,H44411,H44412,H44413,H44419<br>H44421,H44422,H44423,H44429,H44431,H44432,H44433,H44439<br>H44441,H44442,H44443,H44449,H4450,H44521,H44522,H44523,H44529<br>H44531,H44532,H44533,H44539,H44601,H44602,H44603,H44609<br>H44611,H44612,H44613,H44619,H44621,H44622,H44623,H44629<br>H44631,H44632,H44633,H44639,H44641,H44642,H44643,H44649<br>H44651,H44652,H44653,H44659,H44691,H44692,H44693,H44699 | 01730,0213,05320,05322,05329,05440,05441,05444,05449,0760<br>0761,0769,0900,0905,09150,09151,09152,0950,09841,09842,09849<br>1231,1253,1301,1307,36013,36020,36021,36023,36024,36029,36030<br>36031,36032,36033,36034,36040,36041,36043,36044,36050,36051<br>36052,36053,36054,36055,36059,36060,36061,36062,36063,36064<br>36065,36069,36411,36441,36442,36451,36452,36453,36454,36455<br>36456,36457,36459,36460,36461,36462,36463,36464,36470,36471<br>36472,36473,36474,36475,36476,36477,36481,36482,36489,3649<br>37881,37882,37883,37884,37940,37941,37942,37943,37945,37946<br>37949,37950,37951,37952,37953,37955,37956,37957,37959,3798<br>37990,37991,V411 |

Continued on next page

**Supplementary Table S1:** (cont'd) A comprehensive mapping of diagnosis groups used in the experiments to their corresponding ICD-10 and ICD-9 codes.

| Diagnosis | ICD-10 | ICD-9 |
|---|---|---|
| | H44701,H44702,H44703,H44709,H44711,H44712,H44713,H44719 | |
| | H44721,H44722,H44723,H44729,H44731,H44732,H44733,H44739 | |
| | H44741,H44742,H44743,H44749,H44751,H44752,H44753,H44759 | |
| | H44791,H44792,H44793,H44799,H44811,H44812,H44813,H44819,H510 | |
| | H5111,H5112,H5500,H5501,H5502,H5503,H5504,H5509,H5581,H5582 | |
| | H5589,H5700,H5701,H5702,H5703,H5704,H57051,H57052,H57053 | |
| | H57059,H5709,H5710,H5711,H5712,H5713,H578,H5789,H579 | |
| Other specified | B373,B3731,B3732,N820,N821,N822,N823,N824,N825,N828,N829 | 1121,6185,6190,6191,6192,6198,6199,6203,6204,6205,6206,6207 |
| female genital | N8331,N83311,N83312,N83319,N8332,N83321,N83322,N83329,N8333 | 6208,6209,6210,6211,6212,62130,62131,62132,62133,62134,62135 |
| disorders | N83331,N83332,N83339,N834,N8340,N8341,N8342,N8351,N83511 | 6214,6215,6216,6217,6218,6219,6220,62210,62211,62212,6222 |
| | N83512,N83519,N8352,N83521,N83522,N83529,N8353,N836,N837,N838 | 6223,6224,6225,6226,6227,6228,6229,6230,6231,6232,6233,6234 |
| | N839,N840,N841,N842,N843,N848,N849,N8500,N8501,N8502,N852 | 6235,6236,6237,6238,6239,62401,62402,62409,6241,6242,6243 |
| | N853,N854,N855,N856,N857,N858,N859,N85A,N86,N870,N871,N879 | 6246,6248,6249,6250,6251,6252,6254,6255,62570,62571,62579 |
| | N880,N881,N882,N883,N884,N888,N889,N890,N891,N893,N894,N895 | 6258,6267,6268,6269,6273,6290,6291,62920,62921,62922,62923 |
| | N896,N898,N899,N900,N901,N903,N904,N905,N906,N9060,N9061 | 62929,62981,62989,6299 |
| | N9069,N907,N90810,N90811,N90812,N90813,N90818,N9089,N909,N930 | |
| | N931,N938,N939,N940,N941,N9410,N9411,N9412,N9419,N942,N943 | |
| | N94810,N94818,N94819,N9489,N949,N952,N96,N992,N993 | |
| Other specified | B653,E08620,E09620,E10620,E11620,E13620,L100,L101,L102,L103 | 0578,1203,24980,25080,25081,6829,68601,69010,69011,69012 |
| inflammatory | L104,L105,L1081,L1089,L109,L110,L111,L118,L119,L120,L121,L122 | 69018,6918,69271,69276,69277,6930,6931,6938,6939,6940,6941 |
| condition of skin | L1230,L1231,L1235,L128,L129,L130,L131,L138,L139,L14,L200 | 6942,6943,6944,6945,69460,69461,6948,6949,6950,69510,69511 |
| | L2081,L2082,L2083,L2084,L2089,L209,L210,L211,L218,L219,L26 | 69512,69513,69514,69515,69519,6952,6953,6954,69550,69551 |
| | L270,L271,L272,L278,L279,L280,L281,L282,L290,L291,L292,L293 | 69552,69553,69554,69555,69556,69557,69558,69559,69589,6959 |
| | L298,L299,L304,L305,L400,L401,L402,L403,L404,L4050,L4051 | 6960,6961,6962,6963,6964,6965,6968,6970,6971,6978,6980,6981 |
| | L4052,L4053,L4054,L4059,L408,L409,L410,L411,L413,L414,L415 | 6982,6983,6984,6988,6989,7011,7018,7028,7048,70583,7060,7061 |
| | L418,L419,L42,L430,L431,L432,L433,L438,L439,L440,L441,L442 | 7063,7080,7081,7082,7083,7084,7085,7088,7089,7091,7098 |
| | L443,L444,L448,L449,L45,L490,L491,L492,L493,L494,L495,L496 | |
| | L497,L498,L499,L500,L501,L502,L503,L504,L505,L506,L508,L509 | |
| | L510,L511,L512,L513,L518,L519,L52,L530,L531,L532,L533,L538 | |
| | L539,L54,L550,L551,L552,L559,L661,L700,L701,L702,L703,L704 | |
| | L705,L708,L709,L710,L711,L718,L719,L732,L738,L88,L920,L930 | |
| | L931,L932,L945,L950,L951,L958,L959,L981,L982,L983 | |
| Other specified | S058X1A,S058X2A,S058X9A,S09311A,S09312A,S09313A,S09319A | 8719,87262,87263,87264,87269,87272,87273,87274,8738,8739,9580 |
| injury | S09391A,S09392A,S09399A,S098XXA,S1980XA,S1981XA,S1982XA | 9581,9582,9584,9585,9586,9587,9588,95890,95891,95892,95893 |
| | S1983XA,S1984XA,S1985XA,S1989XA,S298XXA,S3981XA,S3982XA | 95899,95901,95909,95911,95912,95913,95914,95919,9592,9593 |
| | S3983XA,S39840A,S39848A,S4980XA,S4981XA,S4982XA,S59801A | 9594,9595,9596,9597 |
| | S59802A,S59809A,S59811A,S59812A,S59819A,S6980XA,S6981XA | |

Continued on next page

**Supplementary Table S1:** (cont'd) A comprehensive mapping of diagnosis groups used in the experiments to their corresponding ICD-10 and ICD-9 codes.

| Diagnosis | ICD-10 | ICD-9 |
|---|---|---|
| | S6982XA,S79811A,S79812A,S79819A,S79821A,S79822A,S79829A | |
| | S8980XA,S8981XA,S8982XA,S99811A,S99812A,S99819A,S99821A | |
| | S99822A,S99829A,T790XXA,T791XXA,T792XXA,T794XXA,T795XXA | |
| | T796XXA,T797XXA,T798XXA,T799XXA,T79A0XA,T79A11A,T79A12A | |
| | T79A19A,T79A21A,T79A22A,T79A29A,T79A3XA,T79A9XA | |
| Other specified | S058X1D,S058X2D,S058X9D,S09311D,S09312D,S09313D,S09319D | V5889 |
| injury, subsequent | S09391D,S09392D,S09399D,S098XXD,S1980XD,S1981XD,S1982XD | |
| encounter | S1983XD,S1984XD,S1985XD,S1989XD,S298XXD,S3981XD,S3982XD | |
| | S3983XD,S39840D,S39848D,S4980XD,S4981XD,S4982XD,S59801D | |
| | S59802D,S59809D,S59811D,S59812D,S59819D,S6980XD,S6981XD | |
| | S6982XD,S79811D,S79812D,S79819D,S79821D,S79822D,S79829D | |
| | S8980XD,S8981XD,S8982XD,S99811D,S99812D,S99819D,S99821D | |
| | S99822D,S99829D,T790XXD,T791XXD,T792XXD,T794XXD,T795XXD | |
| | T796XXD,T797XXD,T798XXD,T799XXD,T79A0XD,T79A11D,T79A12D | |
| | T79A19D,T79A21D,T79A22D,T79A29D,T79A3XD,T79A9XD | |
| Other specified joint | E08610,E08618,E09610,E09618,E10610,E10618,E11610,E11618 | 24960,24980,25060,25061,25080,25081,7131,7132,7135,7136,7138 |
| disorders | E13610,E13618,M0200,M02011,M02012,M02019,M02021,M02022,M02029 | 71600,71601,71602,71603,71604,71605,71606,71607,71608,71609 |
| | M02031,M02032,M02039,M02041,M02042,M02049,M02051,M02052 | 71620,71621,71622,71623,71624,71625,71626,71627,71628,71629 |
| | M02059,M02061,M02062,M02069,M02071,M02072,M02079,M0208,M0209 | 71640,71641,71642,71643,71644,71645,71646,71647,71648,71649 |
| | M0220,M02211,M02212,M02219,M02221,M02222,M02229,M02231,M02232 | 71650,71660,71661,71662,71663,71664,71665,71666,71667,71668 |
| | M02239,M02241,M02242,M02249,M02251,M02252,M02259,M02261 | 71680,71681,71682,71683,71684,71685,71686,71687,71688,71689 |
| | M02262,M02269,M02271,M02272,M02279,M0228,M0229,M1210,M12111 | 71690,7171,7172,7173,71740,71742,71743,71749,7175,7176,7177 |
| | M12112,M12119,M12121,M12122,M12129,M12131,M12132,M12139 | 71785,71789,7179,71800,71801,71802,71803,71804,71805,71807 |
| | M12141,M12142,M12149,M12151,M12152,M12159,M12161,M12162 | 71810,71811,71812,71813,71814,71815,71817,71818,71820,71821 |
| | M12169,M12171,M12172,M12179,M1218,M1219,M1230,M12311,M12312 | 71822,71823,71824,71825,71826,71827,71828,71829,71830,71831 |
| | M12319,M12321,M12322,M12329,M12331,M12332,M12339,M12341 | 71832,71833,71834,71835,71836,71837,71850,71851,71852,71853 |
| | M12342,M12349,M12351,M12352,M12359,M12361,M12362,M12369 | 71854,71855,71856,71857,71865,71871,71872,71873,71874,71875 |
| | M12371,M12372,M12379,M1238,M1239,M1240,M12411,M12412,M12419 | 71877,71880,71881,71882,71883,71884,71885,71886,71887,71888 |
| | M12421,M12422,M12429,M12431,M12432,M12439,M12441,M12442 | 71890,71900,71901,71902,71903,71904,71905,71906,71907,71908 |
| | M12449,M12451,M12452,M12459,M12461,M12462,M12469,M12471 | 71910,71911,71912,71913,71914,71915,71916,71917,71918,71930 |
| | M12472,M12479,M1248,M1249,M1280,M12811,M12812,M12819,M12821 | 71931,71932,71933,71934,71935,71936,71937,71938,71939,71980 |
| | M12822,M12829,M12831,M12832,M12839,M12841,M12842,M12849 | 71981,71982,71983,71984,71985,71986,71987,71988,71990,7218 |
| | M12851,M12852,M12859,M12861,M12862,M12869,M12871,M12872 | 7262,72630,7264,7265,72660,72670,72691,7284,73070,73071,73072 |
| | M12879,M1288,M1289,M129,M130,M1310,M13111,M13112,M13119 | 73073,73074,73075,73076,73077,73078,73079,99952 |
| | M13121,M13122,M13129,M13131,M13132,M13139,M13141,M13142 | |
| | M13149,M13151,M13152,M13159,M13161,M13162,M13169,M13171 | |
| | M13172,M13179,M1380,M13811,M13812,M13819,M13821,M13822,M13829 | |
| | M13831,M13832,M13839,M13841,M13842,M13849,M13851,M13852 | |

Continued on next page

**Supplementary Table S1:** (cont'd) A comprehensive mapping of diagnosis groups used in the experiments to their corresponding ICD-10 and ICD-9 codes.

| Diagnosis | ICD-10 | ICD-9 |
|---|---|---|
| | M13859,M13861,M13862,M13869,M13871,M13872,M13879,M1388,M1389<br>M1480,M14811,M14812,M14819,M14821,M14822,M14829,M14831,M14832<br>M14839,M14841,M14842,M14849,M14851,M14852,M14859,M14861<br>M14862,M14869,M14871,M14872,M14879,M1488,M1489,M2200,M2201<br>M2202,M2210,M2211,M2212,M222X1,M222X2,M222X9,M223X1,M223X2<br>M223X9,M2240,M2241,M2242,M228X1,M228X2,M228X9,M2290,M2291<br>M2292,M23000,M23001,M23002,M23003,M23004,M23005,M23006,M23007<br>M23009,M23011,M23012,M23019,M23021,M23022,M23029,M23031<br>M23032,M23039,M23041,M23042,M23049,M23051,M23052,M23059<br>M23061,M23062,M23069,M23200,M23201,M23202,M23203,M23204<br>M23205,M23206,M23207,M23209,M23211,M23212,M23219,M23221<br>M23222,M23229,M23231,M23232,M23239,M23241,M23242,M23249<br>M23251,M23252,M23259,M23261,M23262,M23269,M23300,M23301<br>M23302,M23303,M23304,M23305,M23306,M23307,M23309,M23311<br>M23312,M23319,M23321,M23322,M23329,M23331,M23332,M23339<br>M23341,M23342,M23349,M23351,M23352,M23359,M23361,M23362<br>M23369,M2340,M2341,M2342,M2350,M2351,M2352,M23601,M23602<br>M23609,M23611,M23612,M23619,M23621,M23622,M23629,M23631<br>M23632,M23639,M23641,M23642,M23649,M23671,M23672,M23679<br>M238X1,M238X2,M238X9,M2390,M2391,M2392,M2400,M24011,M24012<br>M24019,M24021,M24022,M24029,M24031,M24032,M24039,M24041<br>M24042,M24049,M24051,M24052,M24059,M24071,M24072,M24073<br>M24074,M24075,M24076,M2408,M2410,M24111,M24112,M24119,M24121<br>M24122,M24129,M24131,M24132,M24139,M24141,M24142,M24149<br>M24151,M24152,M24159,M24171,M24172,M24173,M24174,M24175<br>M24176,M2419,M2420,M24211,M24212,M24219,M24221,M24222,M24229<br>M24231,M24232,M24239,M24241,M24242,M24249,M24251,M24252<br>M24259,M24271,M24272,M24273,M24274,M24275,M24276,M2428,M2429<br>M2430,M24311,M24312,M24319,M24321,M24322,M24329,M24331,M24332<br>M24339,M24341,M24342,M24349,M24351,M24352,M24359,M24361<br>M24362,M24369,M24371,M24372,M24373,M24374,M24375,M24376,M2439<br>M2440,M24411,M24412,M24419,M24421,M24422,M24429,M24431,M24432<br>M24439,M24441,M24442,M24443,M24444,M24445,M24446,M24451<br>M24452,M24459,M24461,M24462,M24469,M24471,M24472,M24473<br>M24474,M24475,M24476,M24477,M24478,M24479,M2449,M2460,M24611<br>M24612,M24619,M24621,M24622,M24629,M24631,M24632,M24639<br>M24641,M24642,M24649,M24651,M24652,M24659,M24661,M24662<br>M24669,M24671,M24672,M24673,M24674,M24675,M24676,M2469,M247<br>M2480,M24811,M24812,M24819,M24821,M24822,M24829,M24831,M24832<br>M24839,M24841,M24842,M24849,M24851,M24852,M24859,M24871 | |

Continued on next page

**Supplementary Table S1:** (cont'd) A comprehensive mapping of diagnosis groups used in the experiments to their corresponding ICD-10 and ICD-9 codes.

| Diagnosis | ICD-10 | ICD-9 |
|---|---|---|
| | M24872,M24873,M24874,M24875,M24876,M2489,M249,M2500,M25011 | |
| | M25012,M25019,M25021,M25022,M25029,M25031,M25032,M25039 | |
| | M25041,M25042,M25049,M25051,M25052,M25059,M25061,M25062 | |
| | M25069,M25071,M25072,M25073,M25074,M25075,M25076,M2508,M2510 | |
| | M25111,M25112,M25119,M25121,M25122,M25129,M25131,M25132 | |
| | M25139,M25141,M25142,M25149,M25151,M25152,M25159,M25161 | |
| | M25162,M25169,M25171,M25172,M25173,M25174,M25175,M25176,M2518 | |
| | M2520,M25211,M25212,M25219,M25221,M25222,M25229,M25231,M25232 | |
| | M25239,M25241,M25242,M25249,M25251,M25252,M25259,M25261 | |
| | M25262,M25269,M25271,M25272,M25279,M2528,M2530,M25311,M25312 | |
| | M25319,M25321,M25322,M25329,M25331,M25332,M25339,M25341 | |
| | M25342,M25349,M25351,M25352,M25359,M25361,M25362,M25369 | |
| | M25371,M25372,M25373,M25374,M25375,M25376,M2539,M2540,M25411 | |
| | M25412,M25419,M25421,M25422,M25429,M25431,M25432,M25439 | |
| | M25441,M25442,M25449,M25451,M25452,M25459,M25461,M25462 | |
| | M25469,M25471,M25472,M25473,M25474,M25475,M25476,M2548,M2570 | |
| | M25711,M25712,M25719,M25721,M25722,M25729,M25731,M25732 | |
| | M25739,M25741,M25742,M25749,M25751,M25752,M25759,M25761 | |
| | M25762,M25769,M25771,M25772,M25773,M25774,M25775,M25776,M2578 | |
| | M2580,M25811,M25812,M25819,M25821,M25822,M25829,M25831,M25832 | |
| | M25839,M25841,M25842,M25849,M25851,M25852,M25859,M25861 | |
| | M25862,M25869,M25871,M25872,M25879,M259,M3505,M361,M362,M363 | |
| | M364,M8960,M89611,M89612,M89619,M89621,M89622,M89629,M89631 | |
| | M89632,M89639,M89641,M89642,M89649,M89651,M89652,M89659 | |
| | M89661,M89662,M89669,M89671,M89672,M89679,M8968,M8969 | |
| Other specified male | N420,N421,N423,N4230,N4231,N4232,N4239,N4281,N4282,N4283 | 6003,6020,6021,6022,6023,6028,6029,6030,6031,6038,6039,605 |
| genital disorders | N4289,N429,N430,N431,N432,N433,N4340,N4341,N4342,N4400,N4401 | 6070,6073,60781,60782,60783,60785,60789,6079,6081,60820,60821 |
| | N4402,N4403,N4404,N441,N442,N448,N470,N471,N472,N473,N474 | 60822,60823,60824,6083,60881,60883,60884,60885,60886,60887 |
| | N475,N477,N478,N480,N4830,N4831,N4832,N4833,N4839,N485,N486 | 60889,6089 |
| | N4881,N4882,N4883,N4889,N489,N500,N501,N503,N508,N50811 | |
| | N50812,N50819,N5082,N5089,N509,N51,N5311,N5312,N5313,N5314 | |
| | N5319,N538,N539 | |
| Other specified | Z1401,Z1402,Z141,Z148,Z1801,Z1809,Z1810,Z1811,Z1812,Z182 | NoDx,V074,V0751,V0752,V0759,V140,V141,V142,V143,V144,V145 |
| status | Z1831,Z1832,Z1833,Z1839,Z1881,Z1883,Z1889,Z189,Z52000,Z52001 | V146,V147,V148,V149,V1501,V1502,V1503,V1504,V1505,V1506,V1507 |
| | Z52008,Z52010,Z52011,Z52018,Z52090,Z52091,Z52098,Z5210,Z5211 | V1508,V1509,V1581,V1584,V1586,V1589,V4031,V440,V441,V442,V443 |
| | Z5219,Z5220,Z5221,Z5229,Z523,Z524,Z525,Z526,Z52810,Z52811 | V444,V4450,V4451,V4452,V4459,V446,V447,V448,V449,V4511,V4512 |
| | Z52812,Z52813,Z52819,Z5289,Z529,Z5301,Z5309,Z531,Z5320,Z5321 | V460,V4611,V462,V463,V468,V469,V4981,V4983,V4984,V4986,V4987 |
| | Z5329,Z5331,Z5332,Z5333,Z5339,Z538,Z539,Z66,Z6710,Z6711,Z6720 | V4989,V5861,V5862,V5863,V5864,V5865,V5866,V5867,V5868,V5869 |

Continued on next page

**Supplementary Table S1:** (cont'd) A comprehensive mapping of diagnosis groups used in the experiments to their corresponding ICD-10 and ICD-9 codes.

| Diagnosis | ICD-10 | ICD-9 |
|---|---|---|
| | Z6721,Z6730,Z6731,Z6740,Z6741,Z6790,Z6791,Z681,Z6820,Z6821<br>Z6822,Z6823,Z6824,Z6825,Z6826,Z6827,Z6828,Z6829,Z6851,Z6852<br>Z6853,Z711,Z712,Z7401,Z7409,Z741,Z742,Z743,Z748,Z749,Z750<br>Z751,Z752,Z753,Z754,Z755,Z758,Z759,Z763,Z764,Z765,Z7681,Z7682<br>Z77010,Z77011,Z77012,Z77018,Z77020,Z77021,Z77028,Z77090<br>Z77098,Z77110,Z77111,Z77112,Z77118,Z77120,Z77121,Z77122<br>Z77123,Z77128,Z7721,Z7722,Z7729,Z779,Z780,Z781,Z789,Z7901<br>Z7902,Z791,Z792,Z793,Z794,Z7951,Z7952,Z7960,Z7961,Z79620<br>Z79621,Z79622,Z79623,Z79624,Z79630,Z79631,Z79632,Z79633<br>Z79634,Z7964,Z7969,Z79810,Z79811,Z79818,Z7982,Z7983,Z7984<br>Z7985,Z79890,Z79891,Z79899,Z880,Z881,Z882,Z883,Z884,Z885,Z886<br>Z887,Z888,Z889,Z91010,Z91011,Z91012,Z91013,Z91014,Z91018<br>Z9102,Z91030,Z91038,Z91040,Z91041,Z91048,Z9109,Z9111,Z91110<br>Z91118,Z91119,Z91120,Z91128,Z91130,Z91138,Z9114,Z9115,Z9119<br>Z91190,Z91198,Z91199,Z9183,Z91841,Z91842,Z91843,Z91849,Z9189<br>Z91A10,Z91A18,Z91A20,Z91A28,Z91A3,Z91A4,Z91A5,Z91A9,Z930,Z931<br>Z932,Z933,Z934,Z9350,Z9351,Z9352,Z9359,Z936,Z938,Z939,Z990<br>Z9911,Z992,Z993,Z9981,Z9989 | V5901,V5902,V5909,V591,V592,V593,V594,V595,V596,V5970,V5971<br>V5972,V5973,V5974,V598,V599,V604,V605,V6089,V609,V626,V631<br>V632,V638,V639,V6406,V641,V642,V643,V650,V6511,V652,V655,V658<br>V8301,V8302,V8381,V8389,V850,V851,V8521,V8522,V8523,V8524<br>V8525,V8551,V8552,V8553,V8701,V8702,V8709,V8711,V8712,V8719<br>V872,V8731,V8732,V8739,V9001,V9009,V9010,V9011,V9012,V902<br>V9031,V9032,V9033,V9039,V9081,V9083,V9089,V909 |
| Other specified substance-related disorders | F1910,F19120,F19121,F19122,F19129,F19130,F19131,F19132,F19139<br>F1914,F19150,F19151,F19159,F1916,F1917,F19180,F19181,F19182<br>F19188,F1919,F1920,F19220,F19221,F19222,F19229,F19230,F19231<br>F19232,F19239,F1924,F19250,F19251,F19259,F1926,F1927,F19280<br>F19281,F19282,F19288,F1929,F1990,F1991,F19920,F19921,F19922<br>F19929,F19930,F19931,F19932,F19939,F1994,F19950,F19951,F19959<br>F1996,F1997,F19980,F19981,F19982,F19988,F1999,O355XX0,O355XX1<br>O355XX2,O355XX3,O355XX4,O355XX5,O355XX9,O99320,O99321,O99322<br>O99323,O99324,O99325 | 2920,29211,29212,2922,29281,29282,29283,29284,29285,29289<br>2929,30460,30461,30462,30580,30581,30582,30583,64830,64831<br>64832,64833,64834,65551,65553 |
| Other specified upper respiratory infections | A155,A360,A361,A362,A5003,A5273,A545,A564,B002,B085,J00,J020<br>J028,J029,J040,J0410,J0411,J042,J0430,J0431,J050,J0510,J0511<br>J060,J069,J390,J391 | 01130,01220,01230,0320,0321,0323,0340,0542,0740,0900,0958<br>0986,09951,460,462,46400,46401,46410,46411,46420,46421,46430<br>46431,4644,46450,46451,4650,4658,4659,47821,47822,47824 |
| Other unspecified injuries, subsequent encounter | S0590XD,S0591XD,S0592XD,S09301D,S09302D,S09309D,S0990XD<br>S0991XD,S0992XD,S0993XD,S199XXD,S299XXD,S3991XD,S3992XD<br>S3993XD,S3994XD,S4990XD,S4991XD,S4992XD,S59901D,S59902D<br>S59909D,S59911D,S59912D,S59919D,S6990XD,S6991XD,S6992XD<br>S79911D,S79912D,S79919D,S79921D,S79922D,S79929D,S8990XD<br>S8991XD,S8992XD,S99911D,S99912D,S99919D,S99921D,S99922D<br>S99929D,T07XXXD,T148XXD,T1490XD,T1491XD | V5889 |

Continued on next page

**Supplementary Table S1:** (cont'd) A comprehensive mapping of diagnosis groups used in the experiments to their corresponding ICD-10 and ICD-9 codes.

| Diagnosis | ICD-10 | ICD-9 |
|---|---|---|
| Other unspecified | O9A211,O9A212,O9A213,O9A219,O9A22,O9A23,R4588,S0590XA,S0591XA | 64890,64891,64893,64894,8719,87262,87263,87264,87272,87273 |
| injury | S0592XA,S09301A,S09302A,S09309A,S0990XA,S0991XA,S0992XA | 87274,8728,8738,8739,9189,9219,95901,95909,95911,95912,95914 |
| | S0993XA,S199XXA,S299XXA,S3991XA,S3992XA,S3993XA,S3994XA | 95919,9592,9593,9594,9595,9596,9597,9598,9599,E9589 |
| | S4990XA,S4991XA,S4992XA,S59901A,S59902A,S59909A,S59911A | |
| | S59912A,S59919A,S6990XA,S6991XA,S6992XA,S79911A,S79912A | |
| | S79919A,S79921A,S79922A,S79929A,S8990XA,S8991XA,S8992XA | |
| | S99911A,S99912A,S99919A,S99921A,S99922A,S99929A,T07,T07XXXA | |
| | T148,T148XXA,T1490,T1490XA,T1491,T1491XA | |
| Otitis media | A380,B053,H6500,H6501,H6502,H6503,H6504,H6505,H6506,H6507 | 0341,0552,38100,38101,38102,38103,38104,38105,38106,38110 |
| | H65111,H65112,H65113,H65114,H65115,H65116,H65117,H65119 | 38119,38120,38129,3813,3814,38200,38201,38202,3821,3822,3823 |
| | H65191,H65192,H65193,H65194,H65195,H65196,H65197,H65199,H6520 | 3824,3829,4878 |
| | H6521,H6522,H6523,H6530,H6531,H6532,H6533,H65411,H65412 | |
| | H65413,H65419,H65491,H65492,H65493,H65499,H6590,H6591,H6592 | |
| | H6593,H66001,H66002,H66003,H66004,H66005,H66006,H66007,H66009 | |
| | H66011,H66012,H66013,H66014,H66015,H66016,H66017,H66019,H6610 | |
| | H6611,H6612,H6613,H6620,H6621,H6622,H6623,H663X1,H663X2 | |
| | H663X3,H663X9,H6640,H6641,H6642,H6643,H6690,H6691,H6692,H6693 | |
| | H671,H672,H673,H679,J1083,J1183 | |
| Pancreatic disorders | B252,B263,K850,K8500,K8501,K8502,K851,K8510,K8511,K8512,K852 | 0723,0785,5770,5771,5772,5778,5779,5794 |
| (excluding diabetes) | K8520,K8521,K8522,K853,K8530,K8531,K8532,K858,K8580,K8581 | |
| | K8582,K859,K8590,K8591,K8592,K860,K861,K862,K863,K868,K8681 | |
| | K8689,K869,K903 | |
| Paralysis (other than | G041,G8100,G8101,G8102,G8103,G8104,G8110,G8111,G8112,G8113 | 34200,34201,34202,34210,34211,34212,34280,34281,34282,34290 |
| cerebral palsy) | G8114,G8190,G8191,G8192,G8193,G8194,G8220,G8221,G8222,G8250 | 34291,34292,34400,34401,34402,34403,34404,34409,3441,3442 |
| | G8251,G8252,G8253,G8254,G830,G8310,G8311,G8312,G8313,G8314 | 34430,34431,34432,34440,34441,34442,3445,34460,34461,34481 |
| | G8320,G8321,G8322,G8323,G8324,G8330,G8331,G8332,G8333,G8334 | 34489,3449 |
| | G834,G835,G8381,G8382,G8383,G8384,G8389,G839 | |
| Parasitic, other | A060,A061,A062,A063,A064,A065,A066,A067,A0681,A0682,A0689 | 0060,0061,0062,0063,0064,0065,0066,0068,0069,0070,0071,0072 |
| specified and | A069,A070,A071,A072,A073,A074,A078,A079,A09,A5900,A5901,A5902 | 0073,0074,0075,0078,0079,0090,080,0810,0811,0812,0819,0820 |
| unspecified | A5903,A5909,A598,A599,A65,A660,A661,A662,A663,A664,A665,A666 | 0821,0822,0823,08240,08241,08249,0828,0831,0832,0838,0839 |
| infections | A667,A668,A669,A670,A671,A672,A673,A679,A680,A681,A689,A690 | 0840,0841,0842,0843,0844,0845,0846,0848,0849,0850,0851,0852 |
| | A691,A6920,A6921,A6922,A6923,A6929,A698,A699,A750,A751,A752 | 0853,0854,0855,0859,0860,0861,0862,0863,0864,0865,0869,0870 |
| | A753,A759,A770,A771,A772,A773,A7740,A7741,A7749,A778,A779 | 0871,0879,08881,08882,0889,101,1020,1021,1022,1023,1024,1025 |
| | A790,A791,A7981,A7982,A7989,A799,B500,B508,B509,B510,B518 | 1026,1027,1028,1029,1030,1031,1032,1033,1039,1040,1048,1049 |
| | B519,B520,B528,B529,B530,B531,B538,B54,B550,B551,B552,B559 | 1200,1201,1202,1203,1208,1209,1210,1211,1212,1213,1214,1215 |

Continued on next page

**Supplementary Table S1:** (cont'd) A comprehensive mapping of diagnosis groups used in the experiments to their corresponding ICD-10 and ICD-9 codes.

| Diagnosis | ICD-10 | ICD-9 |
|---|---|---|
| | B560,B561,B569,B570,B571,B572,B5730,B5731,B5732,B5739,B5740<br>B5741,B5742,B5749,B575,B5800,B5801,B5809,B581,B582,B583,B5881<br>B5882,B5883,B5889,B589,B600,B6000,B6001,B6002,B6003,B6009<br>B6010,B6011,B6012,B6013,B6019,B602,B608,B64,B650,B651,B652<br>B653,B658,B659,B660,B661,B662,B663,B664,B665,B668,B669,B670<br>B671,B672,B6731,B6732,B6739,B674,B675,B6761,B6769,B677,B678<br>B6790,B6799,B680,B681,B689,B690,B691,B6981,B6989,B699,B700<br>B701,B710,B711,B718,B719,B72,B7300,B7301,B7302,B7309,B731<br>B740,B741,B742,B743,B744,B748,B749,B75,B760,B761,B768,B769<br>B770,B7781,B7789,B779,B780,B781,B787,B789,B79,B80,B810,B811<br>B812,B813,B814,B818,B820,B829,B830,B831,B832,B833,B834,B838<br>B839,B850,B851,B852,B853,B854,B86,B870,B871,B872,B873,B874<br>B8781,B8782,B8789,B879,B880,B881,B882,B883,B888,B889,B89,B998<br>B999,G02,O98611,O98612,O98613,O98619,O9862,O9863,P238,P371<br>P373,P374,P378,P379 | 1216,1218,1219,1220,1221,1222,1223,1224,1225,1226,1227,1228<br>1229,1230,1231,1232,1233,1234,1235,1236,1238,1239,124,1250<br>1251,1252,1253,1255,1256,1257,1259,1260,1261,1262,1263,1268<br>1269,1270,1271,1272,1273,1274,1275,1276,1277,1278,1279,1280<br>1281,1288,1289,129,1300,1301,1302,1303,1304,1305,1307,1309<br>13100,13101,13102,13103,13109,1318,1319,1320,1321,1322,1323<br>1329,1330,1338,1340,1341,1342,1348,1349,13621,13629,1365,1368<br>1369,3211,3212,3218,3708,37215,4848,5281,58181,64740,64741<br>64742,64744,64781,64783,6868,7700,7712 |
| Parkinson's disease | G20 | 3320 |
| Pathological fracture, initial encounter | M8000XA,M80011A,M80012A,M80019A,M80021A,M80022A,M80029A<br>M80031A,M80032A,M80039A,M80041A,M80042A,M80049A,M80051A<br>M80052A,M80059A,M80061A,M80062A,M80069A,M80071A,M80072A<br>M80079A,M8008XA,M800AXA,M8080XA,M80811A,M80812A,M80819A<br>M80821A,M80822A,M80829A,M80831A,M80832A,M80839A,M80841A<br>M80842A,M80849A,M80851A,M80852A,M80859A,M80861A,M80862A<br>M80869A,M80871A,M80872A,M80879A,M8088XA,M808AXA,M8440XA<br>M84411A,M84412A,M84419A,M84421A,M84422A,M84429A,M84431A<br>M84432A,M84433A,M84434A,M84439A,M84441A,M84442A,M84443A<br>M84444A,M84445A,M84446A,M84451A,M84452A,M84453A,M84454A<br>M84459A,M84461A,M84462A,M84463A,M84464A,M84469A,M84471A<br>M84472A,M84473A,M84474A,M84475A,M84476A,M84477A,M84478A<br>M84479A,M8448XA,M8450XA,M84511A,M84512A,M84519A,M84521A<br>M84522A,M84529A,M84531A,M84532A,M84533A,M84534A,M84539A<br>M84541A,M84542A,M84549A,M84550A,M84551A,M84552A,M84553A<br>M84559A,M84561A,M84562A,M84563A,M84564A,M84569A,M84571A<br>M84572A,M84573A,M84574A,M84575A,M84576A,M8458XA,M8460XA<br>M84611A,M84612A,M84619A,M84621A,M84622A,M84629A,M84631A<br>M84632A,M84633A,M84634A,M84639A,M84641A,M84642A,M84649A<br>M84650A,M84651A,M84652A,M84653A,M84659A,M84661A,M84662A<br>M84663A,M84664A,M84669A,M84671A,M84672A,M84673A,M84674A<br>M84675A,M84676A,M8468XA | 73310,73311,73312,73313,73314,73315,73316,73319 |

Continued on next page

**Supplementary Table S1:** (cont'd) A comprehensive mapping of diagnosis groups used in the experiments to their corresponding ICD-10 and ICD-9 codes.

| Diagnosis | ICD-10 | ICD-9 |
|---|---|---|
| Pathological | M8000XD,M8000XG,M8000XK,M8000XP,M80011D,M80011G,M80011K | 73381,73382,V5420,V5421,V5422,V5423,V5424,V5425,V5426,V5427 |
| fracture, subsequent | M80011P,M80012D,M80012G,M80012K,M80012P,M80019D,M80019G | V5429 |
| encounter | M80019K,M80019P,M80021D,M80021G,M80021K,M80021P,M80022D | |
| | M80022G,M80022K,M80022P,M80029D,M80029G,M80029K,M80029P | |
| | M80031D,M80031G,M80031K,M80031P,M80032D,M80032G,M80032K | |
| | M80032P,M80039D,M80039G,M80039K,M80039P,M80041D,M80041G | |
| | M80041K,M80041P,M80042D,M80042G,M80042K,M80042P,M80049D | |
| | M80049G,M80049K,M80049P,M80051D,M80051G,M80051K,M80051P | |
| | M80052D,M80052G,M80052K,M80052P,M80059D,M80059G,M80059K | |
| | M80059P,M80061D,M80061G,M80061K,M80061P,M80062D,M80062G | |
| | M80062K,M80062P,M80069D,M80069G,M80069K,M80069P,M80071D | |
| | M80071G,M80071K,M80071P,M80072D,M80072G,M80072K,M80072P | |
| | M80079D,M80079G,M80079K,M80079P,M8008XD,M8008XG,M8008XK | |
| | M8008XP,M800AXD,M800AXG,M800AXK,M800AXP,M8080XD,M8080XG | |
| | M8080XK,M8080XP,M80811D,M80811G,M80811K,M80811P,M80812D | |
| | M80812G,M80812K,M80812P,M80819D,M80819G,M80819K,M80819P | |
| | M80821D,M80821G,M80821K,M80821P,M80822D,M80822G,M80822K | |
| | M80822P,M80829D,M80829G,M80829K,M80829P,M80831D,M80831G | |
| | M80831K,M80831P,M80832D,M80832G,M80832K,M80832P,M80839D | |
| | M80839G,M80839K,M80839P,M80841D,M80841G,M80841K,M80841P | |
| | M80842D,M80842G,M80842K,M80842P,M80849D,M80849G,M80849K | |
| | M80849P,M80851D,M80851G,M80851K,M80851P,M80852D,M80852G | |
| | M80852K,M80852P,M80859D,M80859G,M80859K,M80859P,M80861D | |
| | M80861G,M80861K,M80861P,M80862D,M80862G,M80862K,M80862P | |
| | M80869D,M80869G,M80869K,M80869P,M80871D,M80871G,M80871K | |
| | M80871P,M80872D,M80872G,M80872K,M80872P,M80879D,M80879G | |
| | M80879K,M80879P,M8088XD,M8088XG,M8088XK,M8088XP,M808AXD | |
| | M808AXG,M808AXK,M808AXP,M8440XD,M8440XG,M8440XK,M8440XP | |
| | M84411D,M84411G,M84411K,M84411P,M84412D,M84412G,M84412K | |
| | M84412P,M84419D,M84419G,M84419K,M84419P,M84421D,M84421G | |
| | M84421K,M84421P,M84422D,M84422G,M84422K,M84422P,M84429D | |
| | M84429G,M84429K,M84429P,M84431D,M84431G,M84431K,M84431P | |
| | M84432D,M84432G,M84432K,M84432P,M84433D,M84433G,M84433K | |
| | M84433P,M84434D,M84434G,M84434K,M84434P,M84439D,M84439G | |
| | M84439K,M84439P,M84441D,M84441G,M84441K,M84441P,M84442D | |
| | M84442G,M84442K,M84442P,M84443D,M84443G,M84443K,M84443P | |
| | M84444D,M84444G,M84444K,M84444P,M84445D,M84445G,M84445K | |
| | M84445P,M84446D,M84446G,M84446K,M84446P,M84451D,M84451G | |
| | M84451K,M84451P,M84452D,M84452G,M84452K,M84452P,M84453D | |

Continued on next page

**Supplementary Table S1:** (cont'd) A comprehensive mapping of diagnosis groups used in the experiments to their corresponding ICD-10 and ICD-9 codes.

| Diagnosis | ICD-10 | ICD-9 |
|---|---|---|
| | M84453G,M84453K,M84453P,M84454D,M84454G,M84454K,M84454P | |
| | M84459D,M84459G,M84459K,M84459P,M84461D,M84461G,M84461K | |
| | M84461P,M84462D,M84462G,M84462K,M84462P,M84463D,M84463G | |
| | M84463K,M84463P,M84464D,M84464G,M84464K,M84464P,M84469D | |
| | M84469G,M84469K,M84469P,M84471D,M84471G,M84471K,M84471P | |
| | M84472D,M84472G,M84472K,M84472P,M84473D,M84473G,M84473K | |
| | M84473P,M84474D,M84474G,M84474K,M84474P,M84475D,M84475G | |
| | M84475K,M84475P,M84476D,M84476G,M84476K,M84476P,M84477D | |
| | M84477G,M84477K,M84477P,M84478D,M84478G,M84478K,M84478P | |
| | M84479D,M84479G,M84479K,M84479P,M8448XD,M8448XG,M8448XK | |
| | M8448XP,M8450XD,M8450XG,M8450XK,M8450XP,M84511D,M84511G | |
| | M84511K,M84511P,M84512D,M84512G,M84512K,M84512P,M84519D | |
| | M84519G,M84519K,M84519P,M84521D,M84521G,M84521K,M84521P | |
| | M84522D,M84522G,M84522K,M84522P,M84529D,M84529G,M84529K | |
| | M84529P,M84531D,M84531G,M84531K,M84531P,M84532D,M84532G | |
| | M84532K,M84532P,M84533D,M84533G,M84533K,M84533P,M84534D | |
| | M84534G,M84534K,M84534P,M84539D,M84539G,M84539K,M84539P | |
| | M84541D,M84541G,M84541K,M84541P,M84542D,M84542G,M84542K | |
| | M84542P,M84549D,M84549G,M84549K,M84549P,M84550D,M84550G | |
| | M84550K,M84550P,M84551D,M84551G,M84551K,M84551P,M84552D | |
| | M84552G,M84552K,M84552P,M84553D,M84553G,M84553K,M84553P | |
| | M84559D,M84559G,M84559K,M84559P,M84561D,M84561G,M84561K | |
| | M84561P,M84562D,M84562G,M84562K,M84562P,M84563D,M84563G | |
| | M84563K,M84563P,M84564D,M84564G,M84564K,M84564P,M84569D | |
| | M84569G,M84569K,M84569P,M84571D,M84571G,M84571K,M84571P | |
| | M84572D,M84572G,M84572K,M84572P,M84573D,M84573G,M84573K | |
| | M84573P,M84574D,M84574G,M84574K,M84574P,M84575D,M84575G | |
| | M84575K,M84575P,M84576D,M84576G,M84576K,M84576P,M8458XD | |
| | M8458XG,M8458XK,M8458XP,M8460XD,M8460XG,M8460XK,M8460XP | |
| | M84611D,M84611G,M84611K,M84611P,M84612D,M84612G,M84612K | |
| | M84612P,M84619D,M84619G,M84619K,M84619P,M84621D,M84621G | |
| | M84621K,M84621P,M84622D,M84622G,M84622K,M84622P,M84629D | |
| | M84629G,M84629K,M84629P,M84631D,M84631G,M84631K,M84631P | |
| | M84632D,M84632G,M84632K,M84632P,M84633D,M84633G,M84633K | |
| | M84633P,M84634D,M84634G,M84634K,M84634P,M84639D,M84639G | |
| | M84639K,M84639P,M84641D,M84641G,M84641K,M84641P,M84642D | |
| | M84642G,M84642K,M84642P,M84649D,M84649G,M84649K,M84649P | |
| | M84650D,M84650G,M84650K,M84650P,M84651D,M84651G,M84651K | |
| | M84651P,M84652D,M84652G,M84652K,M84652P,M84653D,M84653G | |
| | M84653K,M84653P,M84659D,M84659G,M84659K,M84659P,M84661D | |

Continued on next page

**Supplementary Table S1:** (cont'd) A comprehensive mapping of diagnosis groups used in the experiments to their corresponding ICD-10 and ICD-9 codes.

| Diagnosis | ICD-10 | ICD-9 |
|---|---|---|
| | M84661G,M84661K,M84661P,M84662D,M84662G,M84662K,M84662P M84663D,M84663G,M84663K,M84663P,M84664D,M84664G,M84664K M84664P,M84669D,M84669G,M84669K,M84669P,M84671D,M84671G M84671K,M84671P,M84672D,M84672G,M84672K,M84672P,M84673D M84673G,M84673K,M84673P,M84674D,M84674G,M84674K,M84674P M84675D,M84675G,M84675K,M84675P,M84676D,M84676G,M84676K M84676P,M8468XD,M8468XG,M8468XK,M8468XP | |
| Pathological, stress and atypical fractures, sequela | M4840XS,M4841XS,M4842XS,M4843XS,M4844XS,M4845XS,M4846XS M4847XS,M4848XS,M4850XS,M4851XS,M4852XS,M4853XS,M4854XS M4855XS,M4856XS,M4857XS,M4858XS,M8000XS,M80011S,M80012S M80019S,M80021S,M80022S,M80029S,M80031S,M80032S,M80039S M80041S,M80042S,M80049S,M80051S,M80052S,M80059S,M80061S M80062S,M80069S,M80071S,M80072S,M80079S,M8008XS,M800AXS M8080XS,M80811S,M80812S,M80819S,M80821S,M80822S,M80829S M80831S,M80832S,M80839S,M80841S,M80842S,M80849S,M80851S M80852S,M80859S,M80861S,M80862S,M80869S,M80871S,M80872S M80879S,M8088XS,M808AXS,M8430XS,M84311S,M84312S,M84319S M84321S,M84322S,M84329S,M84331S,M84332S,M84333S,M84334S M84339S,M84341S,M84342S,M84343S,M84344S,M84345S,M84346S M84350S,M84351S,M84352S,M84353S,M84359S,M84361S,M84362S M84363S,M84364S,M84369S,M84371S,M84372S,M84373S,M84374S M84375S,M84376S,M84377S,M84378S,M84379S,M8438XS,M8440XS M84411S,M84412S,M84419S,M84421S,M84422S,M84429S,M84431S M84432S,M84433S,M84434S,M84439S,M84441S,M84442S,M84443S M84444S,M84445S,M84446S,M84451S,M84452S,M84453S,M84454S M84459S,M84461S,M84462S,M84463S,M84464S,M84469S,M84471S M84472S,M84473S,M84474S,M84475S,M84476S,M84477S,M84478S M84479S,M8448XS,M8450XS,M84511S,M84512S,M84519S,M84521S M84522S,M84529S,M84531S,M84532S,M84533S,M84534S,M84539S M84541S,M84542S,M84549S,M84550S,M84551S,M84552S,M84553S M84559S,M84561S,M84562S,M84563S,M84564S,M84569S,M84571S M84572S,M84573S,M84574S,M84575S,M84576S,M8458XS,M8460XS M84611S,M84612S,M84619S,M84621S,M84622S,M84629S,M84631S M84632S,M84633S,M84634S,M84639S,M84641S,M84642S,M84649S M84650S,M84651S,M84652S,M84653S,M84659S,M84661S,M84662S M84663S,M84664S,M84669S,M84671S,M84672S,M84673S,M84674S M84675S,M84676S,M8468XS,M84750S,M84751S,M84752S,M84753S M84754S,M84755S,M84756S,M84757S,M84758S,M84759S | 9051,9052,9053,9054,9055 |

Continued on next page

**Supplementary Table S1:** (cont'd) A comprehensive mapping of diagnosis groups used in the experiments to their corresponding ICD-10 and ICD-9 codes.

| Diagnosis | ICD-10 | ICD-9 |
|---|---|---|
| Pericarditis and pericardial disease | A3953,B3323,I010,I092,I300,I301,I308,I309,I310,I311,I312,I313<br>I3131,I3139,I314,I318,I319,I32,M3212 | 03641,07421,3910,393,4200,42090,42091,42099,4230,4231,4232<br>4233,4238,4239,7100 |
| Perinatal infections | A33,A4851,A5001,A5002,A5003,A5004,A5005,A5006,A5007,A5008<br>A5009,A501,A502,A5030,A5031,A5032,A5039,A5040,A5041,A5042<br>A5043,A5044,A5045,A5049,A5051,A5052,A5053,A5054,A5055,A5056<br>A5057,A5059,A506,A507,A509,P230,P231,P232,P233,P234,P235,P236<br>P238,P239,P350,P351,P352,P353,P354,P358,P359,P360,P3610,P3619<br>P362,P3630,P3639,P364,P365,P368,P369,P370,P371,P372,P373,P374<br>P375,P378,P379,P381,P389,P390,P391,P392,P393,P394,P398,P399 | 04041,0900,0901,0902,0903,09040,09041,09042,09049,0905,0906<br>0907,0909,7700,7710,7711,7712,7713,7714,7715,7716,7717,77181<br>77182,77189 |
| Peripheral and visceral vascular disease | I700,I701,I70201,I70202,I70203,I70208,I70209,I70211,I70212<br>I70213,I70218,I70219,I70221,I70222,I70223,I70228,I70229<br>I70231,I70232,I70233,I70234,I70235,I70238,I70239,I70241<br>I70242,I70243,I70244,I70245,I70248,I70249,I7025,I70261,I70262<br>I70263,I70268,I70269,I70291,I70292,I70293,I70298,I70299<br>I70301,I70302,I70303,I70308,I70309,I70311,I70312,I70313<br>I70318,I70319,I70321,I70322,I70323,I70328,I70329,I70331<br>I70332,I70333,I70334,I70335,I70338,I70339,I70341,I70342<br>I70343,I70344,I70345,I70348,I70349,I7035,I70361,I70362,I70363<br>I70368,I70369,I70391,I70392,I70393,I70398,I70399,I70401<br>I70402,I70403,I70408,I70409,I70411,I70412,I70413,I70418<br>I70419,I70421,I70422,I70423,I70428,I70429,I70431,I70432<br>I70433,I70434,I70435,I70438,I70439,I70441,I70442,I70443<br>I70444,I70445,I70448,I70449,I7045,I70461,I70462,I70463,I70468<br>I70469,I70491,I70492,I70493,I70498,I70499,I70501,I70502<br>I70503,I70508,I70509,I70511,I70512,I70513,I70518,I70519<br>I70521,I70522,I70523,I70528,I70529,I70531,I70532,I70533<br>I70534,I70535,I70538,I70539,I70541,I70542,I70543,I70544<br>I70545,I70548,I70549,I7055,I70561,I70562,I70563,I70568,I70569<br>I70591,I70592,I70593,I70598,I70599,I70601,I70602,I70603<br>I70608,I70609,I70611,I70612,I70613,I70618,I70619,I70621<br>I70622,I70623,I70628,I70629,I70631,I70632,I70633,I70634<br>I70635,I70638,I70639,I70641,I70642,I70643,I70644,I70645<br>I70648,I70649,I7065,I70661,I70662,I70663,I70668,I70669,I70691<br>I70692,I70693,I70698,I70699,I70701,I70702,I70703,I70708<br>I70709,I70711,I70712,I70713,I70718,I70719,I70721,I70722<br>I70723,I70728,I70729,I70731,I70732,I70733,I70734,I70735<br>I70738,I70739,I70741,I70742,I70743,I70744,I70745,I70748<br>I70749,I7075,I70761,I70762,I70763,I70768,I70769,I70791,I70792 | 4400,4401,44020,44021,44022,44023,44024,44029,44030,44031<br>44032,4404,4408,4409,4430,4431,44382,44389,4439,5570,5571<br>56984,56985,70711,70712,70713,70714,70715,70719,7079,7854<br>V434,V4589 |

Continued on next page

**Supplementary Table S1:** (cont'd) A comprehensive mapping of diagnosis groups used in the experiments to their corresponding ICD-10 and ICD-9 codes.

| Diagnosis | ICD-10 | ICD-9 |
|---|---|---|
| | I70793,I70798,I70799,I708,I7090,I7091,I7092,I7300,I7301,I731<br>I7381,I7389,I739,K550,K55011,K55012,K55019,K55021,K55022<br>K55029,K55031,K55032,K55039,K55041,K55042,K55049,K55051<br>K55052,K55059,K55061,K55062,K55069,K551,K5520,K5521,Z95820<br>Z9862 | |
| Peritonitis and intra-abdominal abscess | A1831,A5485,A7481,K352,K3520,K3521,K353,K3530,K3531,K3532<br>K3533,K50014,K50114,K50814,K50914,K51014,K51214,K51314,K51414<br>K51514,K51814,K51914,K5700,K5720,K5740,K5780,K610,K611,K612<br>K613,K6131,K6139,K614,K615,K630,K650,K651,K652,K653,K654,K658<br>K659,K67,K6811,K6812,K6819,K750 | 01400,07988,09886,5400,5401,5550,5551,5552,5559,5562,5563<br>5564,5565,5566,5568,5569,56201,56211,566,5670,56721,56722<br>56723,56729,56731,56738,56781,56782,56789,5679,5695,5720<br>99859 |
| Personal family history of disease | Z1501,Z1502,Z1503,Z1504,Z1509,Z1581,Z1589,Z170,Z171,Z191,Z192<br>Z800,Z801,Z802,Z803,Z8041,Z8042,Z8043,Z8049,Z8051,Z8052,Z8059<br>Z806,Z807,Z808,Z809,Z810,Z811,Z812,Z813,Z814,Z818,Z820,Z821<br>Z822,Z823,Z8241,Z8249,Z825,Z8261,Z8262,Z8269,Z8271,Z8279,Z828<br>Z830,Z831,Z832,Z833,Z8341,Z8342,Z83430,Z83438,Z8349,Z83511<br>Z83518,Z8352,Z836,Z8371,Z8379,Z840,Z841,Z842,Z843,Z8481,Z8482<br>Z8489,Z8500,Z8501,Z85020,Z85028,Z85030,Z85038,Z85040,Z85048<br>Z8505,Z85060,Z85068,Z8507,Z8509,Z85110,Z85118,Z8512,Z8520<br>Z8521,Z8522,Z85230,Z85238,Z8529,Z853,Z8540,Z8541,Z8542,Z8543<br>Z8544,Z8545,Z8546,Z8547,Z8548,Z8549,Z8550,Z8551,Z85520,Z85528<br>Z8553,Z8554,Z8559,Z856,Z8571,Z8572,Z8579,Z85810,Z85818,Z85819<br>Z85820,Z85821,Z85828,Z85830,Z85831,Z85840,Z85841,Z85848<br>Z85850,Z85858,Z8589,Z859,Z86000,Z86001,Z86002,Z86003,Z86004<br>Z86005,Z86006,Z86007,Z86008,Z86010,Z86011,Z86012,Z86018,Z8603<br>Z8611,Z8612,Z8613,Z8614,Z8615,Z8616,Z8619,Z862,Z8631,Z8632<br>Z8639,Z8651,Z8659,Z8661,Z8669,Z86711,Z86718,Z8672,Z8673,Z8674<br>Z8679,Z8701,Z8709,Z8711,Z8719,Z872,Z87310,Z87311,Z87312,Z8739<br>Z87410,Z87411,Z87412,Z8742,Z87430,Z87438,Z87440,Z87441,Z87442<br>Z87448,Z8751,Z8759,Z8761,Z8768,Z87710,Z87718,Z87720,Z87721<br>Z87728,Z87730,Z87731,Z87732,Z87738,Z8774,Z8775,Z8776,Z87760<br>Z87761,Z87762,Z87763,Z87768,Z87790,Z87798,Z8781,Z87820,Z87821<br>Z87828,Z87890,Z87891,Z87892,Z87898,Z91410,Z91411,Z91412<br>Z91419,Z9142,Z9149,Z915,Z9151,Z9152,Z9181,Z9182,Z920,Z9221<br>Z9222,Z9223,Z92240,Z92241,Z9225,Z9229,Z923,Z9281,Z9282,Z9283<br>Z9284,Z92850,Z92858,Z92859,Z9286,Z9289,Z9886,Z98870,Z98871<br>Z98891 | 30250,V1000,V1001,V1002,V1003,V1004,V1005,V1006,V1007,V1009<br>V1011,V1012,V1020,V1021,V1022,V1029,V103,V1040,V1041,V1042<br>V1043,V1044,V1045,V1046,V1047,V1048,V1049,V1050,V1051,V1052<br>V1053,V1059,V1060,V1072,V1079,V1081,V1082,V1083,V1084,V1085<br>V1086,V1087,V1088,V1089,V1090,V1091,V114,V118,V1200,V1201<br>V1202,V1203,V1204,V1209,V121,V1221,V1229,V123,V1240,V1241<br>V1242,V1249,V1250,V1251,V1252,V1253,V1254,V1255,V1259,V1260<br>V1261,V1269,V1270,V1271,V1272,V1279,V1300,V1301,V1302,V1303<br>V1309,V131,V1321,V1322,V1323,V1324,V1329,V133,V1351,V1352<br>V1359,V1361,V1362,V1363,V1364,V1365,V1366,V1367,V1368,V1369<br>V1381,V1389,V1521,V1522,V153,V1541,V1542,V1549,V1551,V1552<br>V1553,V1559,V157,V1580,V1582,V1587,V1588,V1589,V160,V161,V162<br>V163,V1640,V1641,V1642,V1643,V1649,V1651,V1652,V1659,V166<br>V167,V168,V169,V170,V171,V172,V173,V1741,V1749,V175,V176,V177<br>V1781,V1789,V180,V1811,V1819,V182,V183,V184,V1851,V1859,V1861<br>V1869,V187,V188,V189,V190,V1911,V1919,V192,V193,V194,V195<br>V196,V197,V198,V4583,V4588,V8401,V8402,V8403,V8404,V8409<br>V8481,V8489,V860,V861,V8741,V8742,V8743,V8744,V8745,V8746<br>V8749 |
| Personality disorders | F070,F21,F600,F601,F602,F603,F604,F605,F606,F607,F6081,F6089 | 30019,3010,30120,30122,3013,3014,30150,30159,3016,3017,30181 |

Continued on next page

**Supplementary Table S1:** (cont'd) A comprehensive mapping of diagnosis groups used in the experiments to their corresponding ICD-10 and ICD-9 codes.

| Diagnosis | ICD-10 | ICD-9 |
|---|---|---|
| | F609,F688,F69 | 30182,30183,30184,30189,3019,3100,3101 |
| Pituitary disorders | E220,E221,E222,E228,E229,E230,E231,E232,E233,E236,E237,E240 | 2530,2531,2532,2533,2535,2536,2537,2538,2539,2550,6281 |
| Pleurisy, pleural effusion and pulmonary collapse | A156,J860,J869,J90,J910,J918,J920,J929,J940,J941,J942,J948<br>J949,J9811,J9819,J982,J983,R091 | 01200,5100,5109,5110,5111,51181,51189,5119,5180,5181,5182<br>53084 |
| Pneumonia (except that caused by tuberculosis) | A0103,A0222,A202,A212,A221,A310,A3701,A3711,A3781,A3791,A430<br>A481,A5004,A5272,A5484,B012,B052,B0681,B250,B371,B380,B381<br>B382,B390,B391,B392,B583,B59,B7781,J09X1,J1000,J1001,J1008<br>J1100,J1108,J120,J121,J122,J123,J1281,J1282,J1289,J129,J13<br>J14,J150,J151,J1520,J15211,J15212,J1529,J153,J154,J155,J156<br>J157,J158,J159,J160,J168,J17,J180,J181,J188,J189,J851,J95851 | 0020,00322,0203,0204,0205,0212,0221,0310,0330,0331,0338,0339<br>0391,0521,0551,05679,0785,0900,0951,09889,1124,1140,1144,1145<br>11505,1270,1304,1363,4800,4801,4802,4803,4808,4809,481,4820<br>4821,4822,48231,48232,48239,48240,48241,48242,48249,48281<br>48282,48283,48284,48289,4829,4830,4831,4838,4841,4843,4845<br>4847,4848,485,486,4870,4871,48801,48811,48881,5130,5171,99731 |
| Pneumothorax | J930,J9311,J9312,J9381,J9382,J9383,J939,J95811 | 5120,5121,51281,51282,51283,51284,51289 |
| Poisoning by drugs, initial encounter | T360X1A,T360X2A,T360X3A,T360X4A,T361X1A,T361X2A,T361X3A<br>T361X4A,T362X1A,T362X2A,T362X3A,T362X4A,T363X1A,T363X2A<br>T363X3A,T363X4A,T364X1A,T364X2A,T364X3A,T364X4A,T365X1A<br>T365X2A,T365X3A,T365X4A,T366X1A,T366X2A,T366X3A,T366X4A<br>T367X1A,T367X2A,T367X3A,T367X4A,T368X1A,T368X2A,T368X3A<br>T368X4A,T3691XA,T3692XA,T3693XA,T3694XA,T370X1A,T370X2A<br>T370X3A,T370X4A,T371X1A,T371X2A,T371X3A,T371X4A,T372X1A<br>T372X2A,T372X3A,T372X4A,T373X1A,T373X2A,T373X3A,T373X4A<br>T374X1A,T374X2A,T374X3A,T374X4A,T375X1A,T375X2A,T375X3A<br>T375X4A,T378X1A,T378X2A,T378X3A,T378X4A,T3791XA,T3792XA<br>T3793XA,T3794XA,T380X1A,T380X2A,T380X3A,T380X4A,T381X1A<br>T381X2A,T381X3A,T381X4A,T382X1A,T382X2A,T382X3A,T382X4A<br>T383X1A,T383X2A,T383X3A,T383X4A,T384X1A,T384X2A,T384X3A<br>T384X4A,T385X1A,T385X2A,T385X3A,T385X4A,T386X1A,T386X2A<br>T386X3A,T386X4A,T387X1A,T387X2A,T387X3A,T387X4A,T38801A<br>T38802A,T38803A,T38804A,T38811A,T38812A,T38813A,T38814A<br>T38891A,T38892A,T38893A,T38894A,T38901A,T38902A,T38903A<br>T38904A,T38991A,T38992A,T38993A,T38994A,T39011A,T39012A<br>T39013A,T39014A,T39091A,T39092A,T39093A,T39094A,T391X1A<br>T391X2A,T391X3A,T391X4A,T392X1A,T392X2A,T392X3A,T392X4A<br>T39311A,T39312A,T39313A,T39314A,T39391A,T39392A,T39393A<br>T39394A,T394X1A,T394X2A,T394X3A,T394X4A,T398X1A,T398X2A<br>T398X3A,T398X4A,T3991XA,T3992XA,T3993XA,T3994XA,T400X1A | 9600,9601,9602,9603,9604,9605,9606,9608,9609,9610,9614,9615<br>9616,9617,9618,9619,9620,9621,9622,9623,9624,9627,9628,9629<br>9630,9631,9634,9635,9638,9639,9640,9642,9643,9644,9645,9646<br>9648,9649,96500,96501,96502,96509,9651,9654,9655,96561,96569<br>9657,9659,9660,9661,9662,9663,9664,9670,9678,9679,9680,9682<br>9683,9684,9685,9689,96900,96901,96902,96903,96904,96905,96909<br>9691,9692,9693,9694,9696,96970,96971,96972,96973,96979,9698<br>9699,9700,9701,97081,9710,9711,9712,9713,9719,9720,9721,9722<br>9723,9724,9725,9726,9727,9729,9730,9731,9732,9733,9734,9735<br>9736,9738,9739,9742,9744,9745,9747,9750,9751,9752,9753,9754<br>9755,9756,9757,9758,9760,9761,9762,9763,9764,9765,9766,9767<br>9768,9769,9770,9772,9778,9779,9786,9788,9789,9790,9796,9799<br>E8500,E8501,E8502,E8503,E8504,E8505,E8506,E8507,E851,E8528<br>E8530,E8531,E8532,E8540,E8541,E8542,E8543,E8548,E8550,E8551<br>E8552,E8553,E8554,E8555,E8556,E8558,E856,E857,E8580,E8581<br>E8582,E8583,E8584,E8585,E8586,E8587,E8588,E9500,E9501,E9502<br>E9503,E9504,E9528,E9620,E9800,E9801,E9802,E9803,E9804,E9805 |

Continued on next page

**Supplementary Table S1:** (cont'd) A comprehensive mapping of diagnosis groups used in the experiments to their corresponding ICD-10 and ICD-9 codes.

| Diagnosis | ICD-10 | ICD-9 |
|---|---|---|
| | T400X2A,T400X3A,T400X4A,T401X1A,T401X2A,T401X3A,T401X4A | |
| | T402X1A,T402X2A,T402X3A,T402X4A,T403X1A,T403X2A,T403X3A | |
| | T403X4A,T40411A,T40412A,T40413A,T40414A,T40421A,T40422A | |
| | T40423A,T40424A,T40491A,T40492A,T40493A,T40494A,T404X1A | |
| | T404X2A,T404X3A,T404X4A,T405X1A,T405X2A,T405X3A,T405X4A | |
| | T40601A,T40602A,T40603A,T40604A,T40691A,T40692A,T40693A | |
| | T40694A,T40711A,T40712A,T40713A,T40714A,T40721A,T40722A | |
| | T40723A,T40724A,T407X1A,T407X2A,T407X3A,T407X4A,T408X1A | |
| | T408X2A,T408X3A,T408X4A,T40901A,T40902A,T40903A,T40904A | |
| | T40991A,T40992A,T40993A,T40994A,T410X1A,T410X2A,T410X3A | |
| | T410X4A,T411X1A,T411X2A,T411X3A,T411X4A,T41201A,T41202A | |
| | T41203A,T41204A,T41291A,T41292A,T41293A,T41294A,T413X1A | |
| | T413X2A,T413X3A,T413X4A,T4141XA,T4142XA,T4143XA,T4144XA | |
| | T415X1A,T415X2A,T415X3A,T415X4A,T420X1A,T420X2A,T420X3A | |
| | T420X4A,T421X1A,T421X2A,T421X3A,T421X4A,T422X1A,T422X2A | |
| | T422X3A,T422X4A,T423X1A,T423X2A,T423X3A,T423X4A,T424X1A | |
| | T424X2A,T424X3A,T424X4A,T425X1A,T425X2A,T425X3A,T425X4A | |
| | T426X1A,T426X2A,T426X3A,T426X4A,T4271XA,T4272XA,T4273XA | |
| | T4274XA,T428X1A,T428X2A,T428X3A,T428X4A,T43011A,T43012A | |
| | T43013A,T43014A,T43021A,T43022A,T43023A,T43024A,T431X1A | |
| | T431X2A,T431X3A,T431X4A,T43201A,T43202A,T43203A,T43204A | |
| | T43211A,T43212A,T43213A,T43214A,T43221A,T43222A,T43223A | |
| | T43224A,T43291A,T43292A,T43293A,T43294A,T433X1A,T433X2A | |
| | T433X3A,T433X4A,T434X1A,T434X2A,T434X3A,T434X4A,T43501A | |
| | T43502A,T43503A,T43504A,T43591A,T43592A,T43593A,T43594A | |
| | T43601A,T43602A,T43603A,T43604A,T43611A,T43612A,T43613A | |
| | T43614A,T43621A,T43622A,T43623A,T43624A,T43631A,T43632A | |
| | T43633A,T43634A,T43641A,T43642A,T43643A,T43644A,T43651A | |
| | T43652A,T43653A,T43654A,T43691A,T43692A,T43693A,T43694A | |
| | T438X1A,T438X2A,T438X3A,T438X4A,T4391XA,T4392XA,T4393XA | |
| | T4394XA,T440X1A,T440X2A,T440X3A,T440X4A,T441X1A,T441X2A | |
| | T441X3A,T441X4A,T442X1A,T442X2A,T442X3A,T442X4A,T443X1A | |
| | T443X2A,T443X3A,T443X4A,T444X1A,T444X2A,T444X3A,T444X4A | |
| | T445X1A,T445X2A,T445X3A,T445X4A,T446X1A,T446X2A,T446X3A | |
| | T446X4A,T447X1A,T447X2A,T447X3A,T447X4A,T448X1A,T448X2A | |
| | T448X3A,T448X4A,T44901A,T44902A,T44903A,T44904A,T44991A | |
| | T44992A,T44993A,T44994A,T450X1A,T450X2A,T450X3A,T450X4A | |
| | T451X1A,T451X2A,T451X3A,T451X4A,T452X1A,T452X2A,T452X3A | |
| | T452X4A,T453X1A,T453X2A,T453X3A,T453X4A,T454X1A,T454X2A | |
| | T454X3A,T454X4A,T45511A,T45512A,T45513A,T45514A,T45521A | |

Continued on next page

**Supplementary Table S1:** (cont'd) A comprehensive mapping of diagnosis groups used in the experiments to their corresponding ICD-10 and ICD-9 codes.

| Diagnosis | ICD-10 | ICD-9 |
|---|---|---|
| | T45522A,T45523A,T45524A,T45601A,T45602A,T45603A,T45604A | |
| | T45611A,T45612A,T45613A,T45614A,T45621A,T45622A,T45623A | |
| | T45624A,T45691A,T45692A,T45693A,T45694A,T457X1A,T457X2A | |
| | T457X3A,T457X4A,T458X1A,T458X2A,T458X3A,T458X4A,T4591XA | |
| | T4592XA,T4593XA,T4594XA,T460X1A,T460X2A,T460X3A,T460X4A | |
| | T461X1A,T461X2A,T461X3A,T461X4A,T462X1A,T462X2A,T462X3A | |
| | T462X4A,T463X1A,T463X2A,T463X3A,T463X4A,T464X1A,T464X2A | |
| | T464X3A,T464X4A,T465X1A,T465X2A,T465X3A,T465X4A,T466X1A | |
| | T466X2A,T466X3A,T466X4A,T467X1A,T467X2A,T467X3A,T467X4A | |
| | T468X1A,T468X2A,T468X3A,T468X4A,T46901A,T46902A,T46903A | |
| | T46904A,T46991A,T46992A,T46993A,T46994A,T470X1A,T470X2A | |
| | T470X3A,T470X4A,T471X1A,T471X2A,T471X3A,T471X4A,T472X1A | |
| | T472X2A,T472X3A,T472X4A,T473X1A,T473X2A,T473X3A,T473X4A | |
| | T474X1A,T474X2A,T474X3A,T474X4A,T475X1A,T475X2A,T475X3A | |
| | T475X4A,T476X1A,T476X2A,T476X3A,T476X4A,T477X1A,T477X2A | |
| | T477X3A,T477X4A,T478X1A,T478X2A,T478X3A,T478X4A,T4791XA | |
| | T4792XA,T4793XA,T4794XA,T480X1A,T480X2A,T480X3A,T480X4A | |
| | T481X1A,T481X2A,T481X3A,T481X4A,T48201A,T48202A,T48203A | |
| | T48204A,T48291A,T48292A,T48293A,T48294A,T483X1A,T483X2A | |
| | T483X3A,T483X4A,T484X1A,T484X2A,T484X3A,T484X4A,T485X1A | |
| | T485X2A,T485X3A,T485X4A,T486X1A,T486X2A,T486X3A,T486X4A | |
| | T48901A,T48902A,T48903A,T48904A,T48991A,T48992A,T48993A | |
| | T48994A,T490X1A,T490X2A,T490X3A,T490X4A,T491X1A,T491X2A | |
| | T491X3A,T491X4A,T492X1A,T492X2A,T492X3A,T492X4A,T493X1A | |
| | T493X2A,T493X3A,T493X4A,T494X1A,T494X2A,T494X3A,T494X4A | |
| | T495X1A,T495X2A,T495X3A,T495X4A,T496X1A,T496X2A,T496X3A | |
| | T496X4A,T497X1A,T497X2A,T497X3A,T497X4A,T498X1A,T498X2A | |
| | T498X3A,T498X4A,T4991XA,T4992XA,T4993XA,T4994XA,T500X1A | |
| | T500X2A,T500X3A,T500X4A,T501X1A,T501X2A,T501X3A,T501X4A | |
| | T502X1A,T502X2A,T502X3A,T502X4A,T503X1A,T503X2A,T503X3A | |
| | T503X4A,T504X1A,T504X2A,T504X3A,T504X4A,T505X1A,T505X2A | |
| | T505X3A,T505X4A,T506X1A,T506X2A,T506X3A,T506X4A,T507X1A | |
| | T507X2A,T507X3A,T507X4A,T508X1A,T508X2A,T508X3A,T508X4A | |
| | T50901A,T50902A,T50903A,T50904A,T50911A,T50912A,T50913A | |
| | T50914A,T50991A,T50992A,T50993A,T50994A,T50A11A,T50A12A | |
| | T50A13A,T50A14A,T50A21A,T50A22A,T50A23A,T50A24A,T50A91A | |
| | T50A92A,T50A93A,T50A94A,T50B11A,T50B12A,T50B13A,T50B14A | |
| | T50B91A,T50B92A,T50B93A,T50B94A,T50Z11A,T50Z12A,T50Z13A | |
| | T50Z14A,T50Z91A,T50Z92A,T50Z93A,T50Z94A | |

Continued on next page

**Supplementary Table S1:** (cont'd) A comprehensive mapping of diagnosis groups used in the experiments to their corresponding ICD-10 and ICD-9 codes.

| Diagnosis | ICD-10 | ICD-9 |
|---|---|---|
| Poisoning by drugs,<br>subsequent encounter | T360X1D,T360X2D,T360X3D,T360X4D,T361X1D,T361X2D,T361X3D<br>T361X4D,T362X1D,T362X2D,T362X3D,T362X4D,T363X1D,T363X2D<br>T363X3D,T363X4D,T364X1D,T364X2D,T364X3D,T364X4D,T365X1D<br>T365X2D,T365X3D,T365X4D,T366X1D,T366X2D,T366X3D,T366X4D<br>T367X1D,T367X2D,T367X3D,T367X4D,T368X1D,T368X2D,T368X3D<br>T368X4D,T3691XD,T3692XD,T3693XD,T3694XD,T370X1D,T370X2D<br>T370X3D,T370X4D,T371X1D,T371X2D,T371X3D,T371X4D,T372X1D<br>T372X2D,T372X3D,T372X4D,T373X1D,T373X2D,T373X3D,T373X4D<br>T374X1D,T374X2D,T374X3D,T374X4D,T375X1D,T375X2D,T375X3D<br>T375X4D,T378X1D,T378X2D,T378X3D,T378X4D,T3791XD,T3792XD<br>T3793XD,T3794XD,T380X1D,T380X2D,T380X3D,T380X4D,T381X1D<br>T381X2D,T381X3D,T381X4D,T382X1D,T382X2D,T382X3D,T382X4D<br>T383X1D,T383X2D,T383X3D,T383X4D,T384X1D,T384X2D,T384X3D<br>T384X4D,T385X1D,T385X2D,T385X3D,T385X4D,T386X1D,T386X2D<br>T386X3D,T386X4D,T387X1D,T387X2D,T387X3D,T387X4D,T38801D<br>T38802D,T38803D,T38804D,T38811D,T38812D,T38813D,T38814D<br>T38891D,T38892D,T38893D,T38894D,T38901D,T38902D,T38903D<br>T38904D,T38991D,T38992D,T38993D,T38994D,T39011D,T39012D<br>T39013D,T39014D,T39091D,T39092D,T39093D,T39094D,T391X1D<br>T391X2D,T391X3D,T391X4D,T392X1D,T392X2D,T392X3D,T392X4D<br>T39311D,T39312D,T39313D,T39314D,T39391D,T39392D,T39393D<br>T39394D,T394X1D,T394X2D,T394X3D,T394X4D,T398X1D,T398X2D<br>T398X3D,T398X4D,T3991XD,T3992XD,T3993XD,T3994XD,T400X1D<br>T400X2D,T400X3D,T400X4D,T401X1D,T401X2D,T401X3D,T401X4D<br>T402X1D,T402X2D,T402X3D,T402X4D,T403X1D,T403X2D,T403X3D<br>T403X4D,T40411D,T40412D,T40413D,T40414D,T40421D,T40422D<br>T40423D,T40424D,T40491D,T40492D,T40493D,T40494D,T404X1D<br>T404X2D,T404X3D,T404X4D,T405X1D,T405X2D,T405X3D,T405X4D<br>T40601D,T40602D,T40603D,T40604D,T40691D,T40692D,T40693D<br>T40694D,T40711D,T40712D,T40713D,T40714D,T40721D,T40722D<br>T40723D,T40724D,T407X1D,T407X2D,T407X3D,T407X4D,T408X1D<br>T408X2D,T408X3D,T408X4D,T40901D,T40902D,T40903D,T40904D<br>T40991D,T40992D,T40993D,T40994D,T410X1D,T410X2D,T410X3D<br>T410X4D,T411X1D,T411X2D,T411X3D,T411X4D,T41201D,T41202D<br>T41203D,T41204D,T41291D,T41292D,T41293D,T41294D,T413X1D<br>T413X2D,T413X3D,T413X4D,T4141XD,T4142XD,T4143XD,T4144XD<br>T415X1D,T415X2D,T415X3D,T415X4D,T420X1D,T420X2D,T420X3D<br>T420X4D,T421X1D,T421X2D,T421X3D,T421X4D,T422X1D,T422X2D<br>T422X3D,T422X4D,T423X1D,T423X2D,T423X3D,T423X4D,T424X1D<br>T424X2D,T424X3D,T424X4D,T425X1D,T425X2D,T425X3D,T425X4D | V5889 |

Continued on next page

**Supplementary Table S1:** (cont'd) A comprehensive mapping of diagnosis groups used in the experiments to their corresponding ICD-10 and ICD-9 codes.

| Diagnosis | ICD-10 | ICD-9 |
|---|---|---|
| | T426X1D,T426X2D,T426X3D,T426X4D,T4271XD,T4272XD,T4273XD | |
| | T4274XD,T428X1D,T428X2D,T428X3D,T428X4D,T43011D,T43012D | |
| | T43013D,T43014D,T43021D,T43022D,T43023D,T43024D,T431X1D | |
| | T431X2D,T431X3D,T431X4D,T43201D,T43202D,T43203D,T43204D | |
| | T43211D,T43212D,T43213D,T43214D,T43221D,T43222D,T43223D | |
| | T43224D,T43291D,T43292D,T43293D,T43294D,T433X1D,T433X2D | |
| | T433X3D,T433X4D,T434X1D,T434X2D,T434X3D,T434X4D,T43501D | |
| | T43502D,T43503D,T43504D,T43591D,T43592D,T43593D,T43594D | |
| | T43601D,T43602D,T43603D,T43604D,T43611D,T43612D,T43613D | |
| | T43614D,T43621D,T43622D,T43623D,T43624D,T43631D,T43632D | |
| | T43633D,T43634D,T43641D,T43642D,T43643D,T43644D,T43651D | |
| | T43652D,T43653D,T43654D,T43691D,T43692D,T43693D,T43694D | |
| | T438X1D,T438X2D,T438X3D,T438X4D,T4391XD,T4392XD,T4393XD | |
| | T4394XD,T440X1D,T440X2D,T440X3D,T440X4D,T441X1D,T441X2D | |
| | T441X3D,T441X4D,T442X1D,T442X2D,T442X3D,T442X4D,T443X1D | |
| | T443X2D,T443X3D,T443X4D,T444X1D,T444X2D,T444X3D,T444X4D | |
| | T445X1D,T445X2D,T445X3D,T445X4D,T446X1D,T446X2D,T446X3D | |
| | T446X4D,T447X1D,T447X2D,T447X3D,T447X4D,T448X1D,T448X2D | |
| | T448X3D,T448X4D,T44901D,T44902D,T44903D,T44904D,T44991D | |
| | T44992D,T44993D,T44994D,T450X1D,T450X2D,T450X3D,T450X4D | |
| | T451X1D,T451X2D,T451X3D,T451X4D,T452X1D,T452X2D,T452X3D | |
| | T452X4D,T453X1D,T453X2D,T453X3D,T453X4D,T454X1D,T454X2D | |
| | T454X3D,T454X4D,T45511D,T45512D,T45513D,T45514D,T45521D | |
| | T45522D,T45523D,T45524D,T45601D,T45602D,T45603D,T45604D | |
| | T45611D,T45612D,T45613D,T45614D,T45621D,T45622D,T45623D | |
| | T45624D,T45691D,T45692D,T45693D,T45694D,T457X1D,T457X2D | |
| | T457X3D,T457X4D,T458X1D,T458X2D,T458X3D,T458X4D,T4591XD | |
| | T4592XD,T4593XD,T4594XD,T460X1D,T460X2D,T460X3D,T460X4D | |
| | T461X1D,T461X2D,T461X3D,T461X4D,T462X1D,T462X2D,T462X3D | |
| | T462X4D,T463X1D,T463X2D,T463X3D,T463X4D,T464X1D,T464X2D | |
| | T464X3D,T464X4D,T465X1D,T465X2D,T465X3D,T465X4D,T466X1D | |
| | T466X2D,T466X3D,T466X4D,T467X1D,T467X2D,T467X3D,T467X4D | |
| | T468X1D,T468X2D,T468X3D,T468X4D,T46901D,T46902D,T46903D | |
| | T46904D,T46991D,T46992D,T46993D,T46994D,T470X1D,T470X2D | |
| | T470X3D,T470X4D,T471X1D,T471X2D,T471X3D,T471X4D,T472X1D | |
| | T472X2D,T472X3D,T472X4D,T473X1D,T473X2D,T473X3D,T473X4D | |
| | T474X1D,T474X2D,T474X3D,T474X4D,T475X1D,T475X2D,T475X3D | |
| | T475X4D,T476X1D,T476X2D,T476X3D,T476X4D,T477X1D,T477X2D | |
| | T477X3D,T477X4D,T478X1D,T478X2D,T478X3D,T478X4D,T4791XD | |
| | T4792XD,T4793XD,T4794XD,T480X1D,T480X2D,T480X3D,T480X4D | |

Continued on next page

**Supplementary Table S1:** (cont'd) A comprehensive mapping of diagnosis groups used in the experiments to their corresponding ICD-10 and ICD-9 codes.

| Diagnosis | ICD-10 | ICD-9 |
|---|---|---|
| | T481X1D,T481X2D,T481X3D,T481X4D,T48201D,T48202D,T48203D<br>T48204D,T48291D,T48292D,T48293D,T48294D,T483X1D,T483X2D<br>T483X3D,T483X4D,T484X1D,T484X2D,T484X3D,T484X4D,T485X1D<br>T485X2D,T485X3D,T485X4D,T486X1D,T486X2D,T486X3D,T486X4D<br>T48901D,T48902D,T48903D,T48904D,T48991D,T48992D,T48993D<br>T48994D,T490X1D,T490X2D,T490X3D,T490X4D,T491X1D,T491X2D<br>T491X3D,T491X4D,T492X1D,T492X2D,T492X3D,T492X4D,T493X1D<br>T493X2D,T493X3D,T493X4D,T494X1D,T494X2D,T494X3D,T494X4D<br>T495X1D,T495X2D,T495X3D,T495X4D,T496X1D,T496X2D,T496X3D<br>T496X4D,T497X1D,T497X2D,T497X3D,T497X4D,T498X1D,T498X2D<br>T498X3D,T498X4D,T4991XD,T4992XD,T4993XD,T4994XD,T500X1D<br>T500X2D,T500X3D,T500X4D,T501X1D,T501X2D,T501X3D,T501X4D<br>T502X1D,T502X2D,T502X3D,T502X4D,T503X1D,T503X2D,T503X3D<br>T503X4D,T504X1D,T504X2D,T504X3D,T504X4D,T505X1D,T505X2D<br>T505X3D,T505X4D,T506X1D,T506X2D,T506X3D,T506X4D,T507X1D<br>T507X2D,T507X3D,T507X4D,T508X1D,T508X2D,T508X3D,T508X4D<br>T50901D,T50902D,T50903D,T50904D,T50911D,T50912D,T50913D<br>T50914D,T50991D,T50992D,T50993D,T50994D,T50A11D,T50A12D<br>T50A13D,T50A14D,T50A21D,T50A22D,T50A23D,T50A24D,T50A91D<br>T50A92D,T50A93D,T50A94D,T50B11D,T50B12D,T50B13D,T50B14D<br>T50B91D,T50B92D,T50B93D,T50B94D,T50Z11D,T50Z12D,T50Z13D<br>T50Z14D,T50Z91D,T50Z92D,T50Z93D,T50Z94D | |
| Poisoning toxic<br>effect adverse<br>effects underdosing,<br>sequela | T360X1S,T360X2S,T360X3S,T360X4S,T360X5S,T360X6S,T361X1S<br>T361X2S,T361X3S,T361X4S,T361X5S,T361X6S,T362X1S,T362X2S<br>T362X3S,T362X4S,T362X5S,T362X6S,T363X1S,T363X2S,T363X3S<br>T363X4S,T363X5S,T363X6S,T364X1S,T364X2S,T364X3S,T364X4S<br>T364X5S,T364X6S,T365X1S,T365X2S,T365X3S,T365X4S,T365X5S<br>T365X6S,T366X1S,T366X2S,T366X3S,T366X4S,T366X5S,T366X6S<br>T367X1S,T367X2S,T367X3S,T367X4S,T367X5S,T367X6S,T368X1S<br>T368X2S,T368X3S,T368X4S,T368X5S,T368X6S,T3691XS,T3692XS<br>T3693XS,T3694XS,T3695XS,T3696XS,T370X1S,T370X2S,T370X3S<br>T370X4S,T370X5S,T370X6S,T371X1S,T371X2S,T371X3S,T371X4S<br>T371X5S,T371X6S,T372X1S,T372X2S,T372X3S,T372X4S,T372X5S<br>T372X6S,T373X1S,T373X2S,T373X3S,T373X4S,T373X5S,T373X6S<br>T374X1S,T374X2S,T374X3S,T374X4S,T374X5S,T374X6S,T375X1S<br>T375X2S,T375X3S,T375X4S,T375X5S,T375X6S,T378X1S,T378X2S<br>T378X3S,T378X4S,T378X5S,T378X6S,T3791XS,T3792XS,T3793XS<br>T3794XS,T3795XS,T3796XS,T380X1S,T380X2S,T380X3S,T380X4S<br>T380X5S,T380X6S,T381X1S,T381X2S,T381X3S,T381X4S,T381X5S | 9090,9091,9095,9099,E9292,E9300,E9301,E9302,E9303,E9304,E9305<br>E9306,E9308,E9310,E9314,E9315,E9316,E9317,E9319,E9320,E9321<br>E9322,E9323,E9324,E9327,E9328,E9329,E9330,E9331,E9334,E9335<br>E9336,E9337,E9338,E9340,E9342,E9344,E9345,E9346,E9348,E9349<br>E9350,E9351,E9352,E9353,E9355,E9356,E9357,E9360,E9361,E9362<br>E9363,E9364,E9370,E9378,E9379,E9382,E9383,E9384,E9389,E9390<br>E9391,E9392,E9393,E9394,E9396,E9397,E9398,E9399,E9400,E9401<br>E9410,E9411,E9412,E9413,E9419,E9420,E9421,E9422,E9424,E9425<br>E9426,E9427,E9429,E9430,E9431,E9432,E9433,E9434,E9435,E9436<br>E9438,E9439,E9442,E9444,E9445,E9447,E9450,E9452,E9453,E9454<br>E9455,E9456,E9457,E9458,E9460,E9461,E9462,E9463,E9464,E9465<br>E9466,E9467,E9468,E9469,E9470,E9472,E9478,E9479,E9486,E9488<br>E9489,E9490,E9496,E9499,E959,E969,E989,NoDx |

Continued on next page

**Supplementary Table S1:** (cont'd) A comprehensive mapping of diagnosis groups used in the experiments to their corresponding ICD-10 and ICD-9 codes.

| Diagnosis | ICD-10 | ICD-9 |
|---|---|---|
| | T381X6S,T382X1S,T382X2S,T382X3S,T382X4S,T382X5S,T382X6S | |
| | T383X1S,T383X2S,T383X3S,T383X4S,T383X5S,T383X6S,T384X1S | |
| | T384X2S,T384X3S,T384X4S,T384X5S,T384X6S,T385X1S,T385X2S | |
| | T385X3S,T385X4S,T385X5S,T385X6S,T386X1S,T386X2S,T386X3S | |
| | T386X4S,T386X5S,T386X6S,T387X1S,T387X2S,T387X3S,T387X4S | |
| | T387X5S,T387X6S,T38801S,T38802S,T38803S,T38804S,T38805S | |
| | T38806S,T38811S,T38812S,T38813S,T38814S,T38815S,T38816S | |
| | T38891S,T38892S,T38893S,T38894S,T38895S,T38896S,T38901S | |
| | T38902S,T38903S,T38904S,T38905S,T38906S,T38991S,T38992S | |
| | T38993S,T38994S,T38995S,T38996S,T39011S,T39012S,T39013S | |
| | T39014S,T39015S,T39016S,T39091S,T39092S,T39093S,T39094S | |
| | T39095S,T39096S,T391X1S,T391X2S,T391X3S,T391X4S,T391X5S | |
| | T391X6S,T392X1S,T392X2S,T392X3S,T392X4S,T392X5S,T392X6S | |
| | T39311S,T39312S,T39313S,T39314S,T39315S,T39316S,T39391S | |
| | T39392S,T39393S,T39394S,T39395S,T39396S,T394X1S,T394X2S | |
| | T394X3S,T394X4S,T394X5S,T394X6S,T398X1S,T398X2S,T398X3S | |
| | T398X4S,T398X5S,T398X6S,T3991XS,T3992XS,T3993XS,T3994XS | |
| | T3995XS,T3996XS,T400X1S,T400X2S,T400X3S,T400X4S,T400X5S | |
| | T400X6S,T401X1S,T401X2S,T401X3S,T401X4S,T401X5S,T402X1S | |
| | T402X2S,T402X3S,T402X4S,T402X5S,T402X6S,T403X1S,T403X2S | |
| | T403X3S,T403X4S,T403X5S,T403X6S,T40411S,T40412S,T40413S | |
| | T40414S,T40415S,T40416S,T40421S,T40422S,T40423S,T40424S | |
| | T40425S,T40426S,T40491S,T40492S,T40493S,T40494S,T40495S | |
| | T40496S,T404X1S,T404X2S,T404X3S,T404X4S,T404X5S,T404X6S | |
| | T405X1S,T405X2S,T405X3S,T405X4S,T405X5S,T405X6S,T40601S | |
| | T40602S,T40603S,T40604S,T40605S,T40606S,T40691S,T40692S | |
| | T40693S,T40694S,T40695S,T40696S,T40711S,T40712S,T40713S | |
| | T40714S,T40715S,T40716S,T40721S,T40722S,T40723S,T40724S | |
| | T40725S,T40726S,T407X1S,T407X2S,T407X3S,T407X4S,T407X5S | |
| | T407X6S,T408X1S,T408X2S,T408X3S,T408X4S,T408X5S,T40901S | |
| | T40902S,T40903S,T40904S,T40905S,T40906S,T40991S,T40992S | |
| | T40993S,T40994S,T40995S,T40996S,T410X1S,T410X2S,T410X3S | |
| | T410X4S,T410X5S,T410X6S,T411X1S,T411X2S,T411X3S,T411X4S | |
| | T411X5S,T411X6S,T41201S,T41202S,T41203S,T41204S,T41205S | |
| | T41206S,T41291S,T41292S,T41293S,T41294S,T41295S,T41296S | |
| | T413X1S,T413X2S,T413X3S,T413X4S,T413X5S,T413X6S,T4141XS | |
| | T4142XS,T4143XS,T4144XS,T4145XS,T4146XS,T415X1S,T415X2S | |
| | T415X3S,T415X4S,T415X5S,T415X6S,T420X1S,T420X2S,T420X3S | |
| | T420X4S,T420X5S,T420X6S,T421X1S,T421X2S,T421X3S,T421X4S | |
| | T421X5S,T421X6S,T422X1S,T422X2S,T422X3S,T422X4S,T422X5S | |

Continued on next page

**Supplementary Table S1:** (cont'd) A comprehensive mapping of diagnosis groups used in the experiments to their corresponding ICD-10 and ICD-9 codes.

| Diagnosis | ICD-10 | ICD-9 |
|---|---|---|
| | T422X6S,T423X1S,T423X2S,T423X3S,T423X4S,T423X5S,T423X6S | |
| | T424X1S,T424X2S,T424X3S,T424X4S,T424X5S,T424X6S,T425X1S | |
| | T425X2S,T425X3S,T425X4S,T425X5S,T425X6S,T426X1S,T426X2S | |
| | T426X3S,T426X4S,T426X5S,T426X6S,T4271XS,T4272XS,T4273XS | |
| | T4274XS,T4275XS,T4276XS,T428X1S,T428X2S,T428X3S,T428X4S | |
| | T428X5S,T428X6S,T43011S,T43012S,T43013S,T43014S,T43015S | |
| | T43016S,T43021S,T43022S,T43023S,T43024S,T43025S,T43026S | |
| | T431X1S,T431X2S,T431X3S,T431X4S,T431X5S,T431X6S,T43201S | |
| | T43202S,T43203S,T43204S,T43205S,T43206S,T43211S,T43212S | |
| | T43213S,T43214S,T43215S,T43216S,T43221S,T43222S,T43223S | |
| | T43224S,T43225S,T43226S,T43291S,T43292S,T43293S,T43294S | |
| | T43295S,T43296S,T433X1S,T433X2S,T433X3S,T433X4S,T433X5S | |
| | T433X6S,T434X1S,T434X2S,T434X3S,T434X4S,T434X5S,T434X6S | |
| | T43501S,T43502S,T43503S,T43504S,T43505S,T43506S,T43591S | |
| | T43592S,T43593S,T43594S,T43595S,T43596S,T43601S,T43602S | |
| | T43603S,T43604S,T43605S,T43606S,T43611S,T43612S,T43613S | |
| | T43614S,T43615S,T43616S,T43621S,T43622S,T43623S,T43624S | |
| | T43625S,T43626S,T43631S,T43632S,T43633S,T43634S,T43635S | |
| | T43636S,T43641S,T43642S,T43643S,T43644S,T43651S,T43652S | |
| | T43653S,T43654S,T43655S,T43656S,T43691S,T43692S,T43693S | |
| | T43694S,T43695S,T43696S,T438X1S,T438X2S,T438X3S,T438X4S | |
| | T438X5S,T438X6S,T4391XS,T4392XS,T4393XS,T4394XS,T4395XS | |
| | T4396XS,T440X1S,T440X2S,T440X3S,T440X4S,T440X5S,T440X6S | |
| | T441X1S,T441X2S,T441X3S,T441X4S,T441X5S,T441X6S,T442X1S | |
| | T442X2S,T442X3S,T442X4S,T442X5S,T442X6S,T443X1S,T443X2S | |
| | T443X3S,T443X4S,T443X5S,T443X6S,T444X1S,T444X2S,T444X3S | |
| | T444X4S,T444X5S,T444X6S,T445X1S,T445X2S,T445X3S,T445X4S | |
| | T445X5S,T445X6S,T446X1S,T446X2S,T446X3S,T446X4S,T446X5S | |
| | T446X6S,T447X1S,T447X2S,T447X3S,T447X4S,T447X5S,T447X6S | |
| | T448X1S,T448X2S,T448X3S,T448X4S,T448X5S,T448X6S,T44901S | |
| | T44902S,T44903S,T44904S,T44905S,T44906S,T44991S,T44992S | |
| | T44993S,T44994S,T44995S,T44996S,T450X1S,T450X2S,T450X3S | |
| | T450X4S,T450X5S,T450X6S,T451X1S,T451X2S,T451X3S,T451X4S | |
| | T451X5S,T451X6S,T452X1S,T452X2S,T452X3S,T452X4S,T452X5S | |
| | T452X6S,T453X1S,T453X2S,T453X3S,T453X4S,T453X5S,T453X6S | |
| | T454X1S,T454X2S,T454X3S,T454X4S,T454X5S,T454X6S,T45511S | |
| | T45512S,T45513S,T45514S,T45515S,T45516S,T45521S,T45522S | |
| | T45523S,T45524S,T45525S,T45526S,T45601S,T45602S,T45603S | |
| | T45604S,T45605S,T45606S,T45611S,T45612S,T45613S,T45614S | |
| | T45615S,T45616S,T45621S,T45622S,T45623S,T45624S,T45625S | |

Continued on next page

**Supplementary Table S1:** (cont'd) A comprehensive mapping of diagnosis groups used in the experiments to their corresponding ICD-10 and ICD-9 codes.

| Diagnosis | ICD-10 | ICD-9 |
|---|---|---|
| | T45626S,T45691S,T45692S,T45693S,T45694S,T45695S,T45696S | |
| | T457X1S,T457X2S,T457X3S,T457X4S,T457X5S,T457X6S,T458X1S | |
| | T458X2S,T458X3S,T458X4S,T458X5S,T458X6S,T4591XS,T4592XS | |
| | T4593XS,T4594XS,T4595XS,T4596XS,T460X1S,T460X2S,T460X3S | |
| | T460X4S,T460X5S,T460X6S,T461X1S,T461X2S,T461X3S,T461X4S | |
| | T461X5S,T461X6S,T462X1S,T462X2S,T462X3S,T462X4S,T462X5S | |
| | T462X6S,T463X1S,T463X2S,T463X3S,T463X4S,T463X5S,T463X6S | |
| | T464X1S,T464X2S,T464X3S,T464X4S,T464X5S,T464X6S,T465X1S | |
| | T465X2S,T465X3S,T465X4S,T465X5S,T465X6S,T466X1S,T466X2S | |
| | T466X3S,T466X4S,T466X5S,T466X6S,T467X1S,T467X2S,T467X3S | |
| | T467X4S,T467X5S,T467X6S,T468X1S,T468X2S,T468X3S,T468X4S | |
| | T468X5S,T468X6S,T46901S,T46902S,T46903S,T46904S,T46905S | |
| | T46906S,T46991S,T46992S,T46993S,T46994S,T46995S,T46996S | |
| | T470X1S,T470X2S,T470X3S,T470X4S,T470X5S,T470X6S,T471X1S | |
| | T471X2S,T471X3S,T471X4S,T471X5S,T471X6S,T472X1S,T472X2S | |
| | T472X3S,T472X4S,T472X5S,T472X6S,T473X1S,T473X2S,T473X3S | |
| | T473X4S,T473X5S,T473X6S,T474X1S,T474X2S,T474X3S,T474X4S | |
| | T474X5S,T474X6S,T475X1S,T475X2S,T475X3S,T475X4S,T475X5S | |
| | T475X6S,T476X1S,T476X2S,T476X3S,T476X4S,T476X5S,T476X6S | |
| | T477X1S,T477X2S,T477X3S,T477X4S,T477X5S,T477X6S,T478X1S | |
| | T478X2S,T478X3S,T478X4S,T478X5S,T478X6S,T4791XS,T4792XS | |
| | T4793XS,T4794XS,T4795XS,T4796XS,T480X1S,T480X2S,T480X3S | |
| | T480X4S,T480X5S,T480X6S,T481X1S,T481X2S,T481X3S,T481X4S | |
| | T481X5S,T481X6S,T48201S,T48202S,T48203S,T48204S,T48205S | |
| | T48206S,T48291S,T48292S,T48293S,T48294S,T48295S,T48296S | |
| | T483X1S,T483X2S,T483X3S,T483X4S,T483X5S,T483X6S,T484X1S | |
| | T484X2S,T484X3S,T484X4S,T484X5S,T484X6S,T485X1S,T485X2S | |
| | T485X3S,T485X4S,T485X5S,T485X6S,T486X1S,T486X2S,T486X3S | |
| | T486X4S,T486X5S,T486X6S,T48901S,T48902S,T48903S,T48904S | |
| | T48905S,T48906S,T48991S,T48992S,T48993S,T48994S,T48995S | |
| | T48996S,T490X1S,T490X2S,T490X3S,T490X4S,T490X5S,T490X6S | |
| | T491X1S,T491X2S,T491X3S,T491X4S,T491X5S,T491X6S,T492X1S | |
| | T492X2S,T492X3S,T492X4S,T492X5S,T492X6S,T493X1S,T493X2S | |
| | T493X3S,T493X4S,T493X5S,T493X6S,T494X1S,T494X2S,T494X3S | |
| | T494X4S,T494X5S,T494X6S,T495X1S,T495X2S,T495X3S,T495X4S | |
| | T495X5S,T495X6S,T496X1S,T496X2S,T496X3S,T496X4S,T496X5S | |
| | T496X6S,T497X1S,T497X2S,T497X3S,T497X4S,T497X5S,T497X6S | |
| | T498X1S,T498X2S,T498X3S,T498X4S,T498X5S,T498X6S,T4991XS | |
| | T4992XS,T4993XS,T4994XS,T4995XS,T4996XS,T500X1S,T500X2S | |
| | T500X3S,T500X4S,T500X5S,T500X6S,T501X1S,T501X2S,T501X3S | |

Continued on next page

**Supplementary Table S1:** (cont'd) A comprehensive mapping of diagnosis groups used in the experiments to their corresponding ICD-10 and ICD-9 codes.

| Diagnosis | ICD-10 | ICD-9 |
|---|---|---|
| | T501X4S,T501X5S,T501X6S,T502X1S,T502X2S,T502X3S,T502X4S | |
| | T502X5S,T502X6S,T503X1S,T503X2S,T503X3S,T503X4S,T503X5S | |
| | T503X6S,T504X1S,T504X2S,T504X3S,T504X4S,T504X5S,T504X6S | |
| | T505X1S,T505X2S,T505X3S,T505X4S,T505X5S,T505X6S,T506X1S | |
| | T506X2S,T506X3S,T506X4S,T506X5S,T506X6S,T507X1S,T507X2S | |
| | T507X3S,T507X4S,T507X5S,T507X6S,T508X1S,T508X2S,T508X3S | |
| | T508X4S,T508X5S,T508X6S,T50901S,T50902S,T50903S,T50904S | |
| | T50905S,T50906S,T50911S,T50912S,T50913S,T50914S,T50915S | |
| | T50916S,T50991S,T50992S,T50993S,T50994S,T50995S,T50996S | |
| | T50A11S,T50A12S,T50A13S,T50A14S,T50A15S,T50A16S,T50A21S | |
| | T50A22S,T50A23S,T50A24S,T50A25S,T50A26S,T50A91S,T50A92S | |
| | T50A93S,T50A94S,T50A95S,T50A96S,T50B11S,T50B12S,T50B13S | |
| | T50B14S,T50B15S,T50B16S,T50B91S,T50B92S,T50B93S,T50B94S | |
| | T50B95S,T50B96S,T50Z11S,T50Z12S,T50Z13S,T50Z14S,T50Z15S | |
| | T50Z16S,T50Z91S,T50Z92S,T50Z93S,T50Z94S,T50Z95S,T50Z96S | |
| | T510X1S,T510X2S,T510X3S,T510X4S,T511X1S,T511X2S,T511X3S | |
| | T511X4S,T512X1S,T512X2S,T512X3S,T512X4S,T513X1S,T513X2S | |
| | T513X3S,T513X4S,T518X1S,T518X2S,T518X3S,T518X4S,T5191XS | |
| | T5192XS,T5193XS,T5194XS,T520X1S,T520X2S,T520X3S,T520X4S | |
| | T521X1S,T521X2S,T521X3S,T521X4S,T522X1S,T522X2S,T522X3S | |
| | T522X4S,T523X1S,T523X2S,T523X3S,T523X4S,T524X1S,T524X2S | |
| | T524X3S,T524X4S,T528X1S,T528X2S,T528X3S,T528X4S,T5291XS | |
| | T5292XS,T5293XS,T5294XS,T530X1S,T530X2S,T530X3S,T530X4S | |
| | T531X1S,T531X2S,T531X3S,T531X4S,T532X1S,T532X2S,T532X3S | |
| | T532X4S,T533X1S,T533X2S,T533X3S,T533X4S,T534X1S,T534X2S | |
| | T534X3S,T534X4S,T535X1S,T535X2S,T535X3S,T535X4S,T536X1S | |
| | T536X2S,T536X3S,T536X4S,T537X1S,T537X2S,T537X3S,T537X4S | |
| | T5391XS,T5392XS,T5393XS,T5394XS,T540X1S,T540X2S,T540X3S | |
| | T540X4S,T541X1S,T541X2S,T541X3S,T541X4S,T542X1S,T542X2S | |
| | T542X3S,T542X4S,T543X1S,T543X2S,T543X3S,T543X4S,T5491XS | |
| | T5492XS,T5493XS,T5494XS,T550X1S,T550X2S,T550X3S,T550X4S | |
| | T551X1S,T551X2S,T551X3S,T551X4S,T560X1S,T560X2S,T560X3S | |
| | T560X4S,T561X1S,T561X2S,T561X3S,T561X4S,T562X1S,T562X2S | |
| | T562X3S,T562X4S,T563X1S,T563X2S,T563X3S,T563X4S,T564X1S | |
| | T564X2S,T564X3S,T564X4S,T565X1S,T565X2S,T565X3S,T565X4S | |
| | T566X1S,T566X2S,T566X3S,T566X4S,T567X1S,T567X2S,T567X3S | |
| | T567X4S,T56811S,T56812S,T56813S,T56814S,T56891S,T56892S | |
| | T56893S,T56894S,T5691XS,T5692XS,T5693XS,T5694XS,T570X1S | |
| | T570X2S,T570X3S,T570X4S,T571X1S,T571X2S,T571X3S,T571X4S | |
| | T572X1S,T572X2S,T572X3S,T572X4S,T573X1S,T573X2S,T573X3S | |

Continued on next page

**Supplementary Table S1:** (cont'd) A comprehensive mapping of diagnosis groups used in the experiments to their corresponding ICD-10 and ICD-9 codes.

| Diagnosis | ICD-10 | ICD-9 |
|---|---|---|
| | T573X4S,T578X1S,T578X2S,T578X3S,T578X4S,T5791XS,T5792XS<br>T5793XS,T5794XS,T5801XS,T5802XS,T5803XS,T5804XS,T5811XS<br>T5812XS,T5813XS,T5814XS,T582X1S,T582X2S,T582X3S,T582X4S<br>T588X1S,T588X2S,T588X3S,T588X4S,T5891XS,T5892XS,T5893XS<br>T5894XS,T590X1S,T590X2S,T590X3S,T590X4S,T591X1S,T591X2S<br>T591X3S,T591X4S,T592X1S,T592X2S,T592X3S,T592X4S,T593X1S<br>T593X2S,T593X3S,T593X4S,T594X1S,T594X2S,T594X3S,T594X4S<br>T595X1S,T595X2S,T595X3S,T595X4S,T596X1S,T596X2S,T596X3S<br>T596X4S,T597X1S,T597X2S,T597X3S,T597X4S,T59811S,T59812S<br>T59813S,T59814S,T59891S,T59892S,T59893S,T59894S,T5991XS<br>T5992XS,T5993XS,T5994XS,T600X1S,T600X2S,T600X3S,T600X4S<br>T601X1S,T601X2S,T601X3S,T601X4S,T602X1S,T602X2S,T602X3S<br>T602X4S,T603X1S,T603X2S,T603X3S,T603X4S,T604X1S,T604X2S<br>T604X3S,T604X4S,T608X1S,T608X2S,T608X3S,T608X4S,T6091XS<br>T6092XS,T6093XS,T6094XS,T6101XS,T6102XS,T6103XS,T6104XS<br>T6111XS,T6112XS,T6113XS,T6114XS,T61771S,T61772S,T61773S<br>T61774S,T61781S,T61782S,T61783S,T61784S,T618X1S,T618X2S<br>T618X3S,T618X4S,T6191XS,T6192XS,T6193XS,T6194XS,T620X1S<br>T620X2S,T620X3S,T620X4S,T621X1S,T621X2S,T621X3S,T621X4S<br>T622X1S,T622X2S,T622X3S,T622X4S,T628X1S,T628X2S,T628X3S<br>T628X4S,T6291XS,T6292XS,T6293XS,T6294XS,T63001S,T63002S<br>T63003S,T63004S,T63011S,T63012S,T63013S,T63014S,T63021S<br>T63022S,T63023S,T63024S,T63031S,T63032S,T63033S,T63034S<br>T63041S,T63042S,T63043S,T63044S,T63061S,T63062S,T63063S<br>T63064S,T63071S,T63072S,T63073S,T63074S,T63081S,T63082S<br>T63083S,T63084S,T63091S,T63092S,T63093S,T63094S,T63111S<br>T63112S,T63113S,T63114S,T63121S,T63122S,T63123S,T63124S<br>T63191S,T63192S,T63193S,T63194S,T632X1S,T632X2S,T632X3S<br>T632X4S,T63301S,T63302S,T63303S,T63304S,T63311S,T63312S<br>T63313S,T63314S,T63321S,T63322S,T63323S,T63324S,T63331S<br>T63332S,T63333S,T63334S,T63391S,T63392S,T63393S,T63394S<br>T63411S,T63412S,T63413S,T63414S,T63421S,T63422S,T63423S<br>T63424S,T63431S,T63432S,T63433S,T63434S,T63441S,T63442S<br>T63443S,T63444S,T63451S,T63452S,T63453S,T63454S,T63461S<br>T63462S,T63463S,T63464S,T63481S,T63482S,T63483S,T63484S<br>T63511S,T63512S,T63513S,T63514S,T63591S,T63592S,T63593S<br>T63594S,T63611S,T63612S,T63613S,T63614S,T63621S,T63622S<br>T63623S,T63624S,T63631S,T63632S,T63633S,T63634S,T63691S<br>T63692S,T63693S,T63694S,T63711S,T63712S,T63713S,T63714S<br>T63791S,T63792S,T63793S,T63794S,T63811S,T63812S,T63813S | |

Continued on next page

**Supplementary Table S1:** (cont'd) A comprehensive mapping of diagnosis groups used in the experiments to their corresponding ICD-10 and ICD-9 codes.

| Diagnosis | ICD-10 | ICD-9 |
|---|---|---|
| | T63814S,T63821S,T63822S,T63823S,T63824S,T63831S,T63832S | |
| | T63833S,T63834S,T63891S,T63892S,T63893S,T63894S,T6391XS | |
| | T6392XS,T6393XS,T6394XS,T6401XS,T6402XS,T6403XS,T6404XS | |
| | T6481XS,T6482XS,T6483XS,T6484XS,T650X1S,T650X2S,T650X3S | |
| | T650X4S,T651X1S,T651X2S,T651X3S,T651X4S,T65211S,T65212S | |
| | T65213S,T65214S,T65221S,T65222S,T65223S,T65224S,T65291S | |
| | T65292S,T65293S,T65294S,T653X1S,T653X2S,T653X3S,T653X4S | |
| | T654X1S,T654X2S,T654X3S,T654X4S,T655X1S,T655X2S,T655X3S | |
| | T655X4S,T656X1S,T656X2S,T656X3S,T656X4S,T65811S,T65812S | |
| | T65813S,T65814S,T65821S,T65822S,T65823S,T65824S,T65831S | |
| | T65832S,T65833S,T65834S,T65891S,T65892S,T65893S,T65894S | |
| | T6591XS,T6592XS,T6593XS,T6594XS,T788XXS | |
| Polyhydramnios and | O401XX0,O401XX1,O401XX2,O401XX3,O401XX4,O401XX5,O401XX9 | 65700,65701,65703,65800,65801,65803,65880,65881,65883,65890 |
| other problems of | O402XX0,O402XX1,O402XX2,O402XX3,O402XX4,O402XX5,O402XX9 | 65891,65893 |
| amniotic cavity | O403XX0,O403XX1,O403XX2,O403XX3,O403XX4,O403XX5,O403XX9 | |
| | O409XX0,O409XX1,O409XX2,O409XX3,O409XX4,O409XX5,O409XX9 | |
| | O4100X0,O4100X1,O4100X2,O4100X3,O4100X4,O4100X5,O4100X9 | |
| | O4101X0,O4101X1,O4101X2,O4101X3,O4101X4,O4101X5,O4101X9 | |
| | O4102X0,O4102X1,O4102X2,O4102X3,O4102X4,O4102X5,O4102X9 | |
| | O4103X0,O4103X1,O4103X2,O4103X3,O4103X4,O4103X5,O4103X9 | |
| | O418X10,O418X11,O418X12,O418X13,O418X14,O418X15,O418X19 | |
| | O418X20,O418X21,O418X22,O418X23,O418X24,O418X25,O418X29 | |
| | O418X30,O418X31,O418X32,O418X33,O418X34,O418X35,O418X39 | |
| | O418X90,O418X91,O418X92,O418X93,O418X94,O418X95,O418X99 | |
| | O4190X0,O4190X1,O4190X2,O4190X3,O4190X4,O4190X5,O4190X9 | |
| | O4191X0,O4191X1,O4191X2,O4191X3,O4191X4,O4191X5,O4191X9 | |
| | O4192X0,O4192X1,O4192X2,O4192X3,O4192X4,O4192X5,O4192X9 | |
| | O4193X0,O4193X1,O4193X2,O4193X3,O4193X4,O4193X5,O4193X9 | |
| Polyneuropathies | A3683,A5043,A5215,B0223,B2684,B2701,B2711,B2781,B2791,E0842 | 03289,05313,07272,075,09040,09484,09485,09486,09489,24960 |
| | E0843,E0942,E0943,E1042,E1043,E1142,E1143,E1342,E1343,G600 | 25060,25061,3560,3561,3562,3563,3564,3568,3569,3570,3571,3572 |
| | G601,G602,G603,G608,G609,G610,G611,G6181,G6182,G6189,G619 | 3574,3575,3576,3577,35781,35782,35789,3579,5363,7101,7140 |
| | G620,G621,G622,G6281,G6282,G6289,G629,G63,M0550,M05511,M05512 | |
| | M05519,M05521,M05522,M05529,M05531,M05532,M05539,M05541 | |
| | M05542,M05549,M05551,M05552,M05559,M05561,M05562,M05569 | |
| | M05571,M05572,M05579,M0559,M3483 | |
| Postprocedural or | I970,I97110,I97111,I97120,I97121,I97130,I97131,I97190,I97191 | 4294,4570,99702,9971,99791,99811,99812,9982 |
| postoperative | I972,I973,I97410,I97411,I97418,I9742,I9751,I9752,I97610 | |

Continued on next page

**Supplementary Table S1:** (cont'd) A comprehensive mapping of diagnosis groups used in the experiments to their corresponding ICD-10 and ICD-9 codes.

| Diagnosis | ICD-10 | ICD-9 |
|---|---|---|
| circulatory system complication | I97611,I97618,I9762,I97620,I97621,I97622,I97630,I97631,I97638<br>I97640,I97641,I97648,I97710,I97711,I97790,I97791,I97810<br>I97811,I97820,I97821,I9788,I9789 | |
| Postprocedural or postoperative complications of the spleen | D7801,D7802,D7811,D7812,D7821,D7822,D7831,D7832,D7833,D7834<br>D7881,D7889 | 99799,99811,99812,9982 |
| Postprocedural or postoperative digestive system complication | K910,K911,K912,K913,K9130,K9131,K9132,K915,K9161,K9162,K9171<br>K9172,K9181,K9182,K9183,K91840,K91841,K91850,K91858,K9186<br>K91870,K91871,K91872,K91873,K9189,K9400,K9401,K9402,K9403<br>K9409,K9410,K9411,K9412,K9413,K9419,K9420,K9421,K9422,K9423<br>K9429,K9430,K9431,K9432,K9433,K9439,K9501,K9509,K9581,K9589 | 53086,53087,53640,53641,53642,53649,53901,53909,53981,53989<br>5642,5643,5644,56960,56961,56962,56969,56971,56979,5760,5793<br>99741,99749,99811,99812,9982 |
| Postprocedural or postoperative ear and or mastoid process complication | H9500,H9501,H9502,H9503,H95111,H95112,H95113,H95119,H95121<br>H95122,H95123,H95129,H95131,H95132,H95133,H95139,H95191<br>H95192,H95193,H95199,H9521,H9522,H9531,H9532,H9541,H9542<br>H9551,H9552,H9553,H9554,H95811,H95812,H95813,H95819,H9588<br>H9589 | 38330,38331,38332,38333,99799,99811,99812,9982 |
| Postprocedural or postoperative endocrine or metabolic complication | E3601,E3602,E3611,E3612,E368,E890,E891,E892,E893,E8940,E8941<br>E895,E896,E89810,E89811,E89820,E89821,E89822,E89823,E8989 | 2440,2441,2513,2521,2537,25541,2562,2571,99799,99811,99812<br>9982,99889 |
| Postprocedural or postoperative eye complication | H59011,H59012,H59013,H59019,H59021,H59022,H59023,H59029<br>H59031,H59032,H59033,H59039,H59091,H59092,H59093,H59099<br>H59111,H59112,H59113,H59119,H59121,H59122,H59123,H59129<br>H59211,H59212,H59213,H59219,H59221,H59222,H59223,H59229<br>H59311,H59312,H59313,H59319,H59321,H59322,H59323,H59329<br>H59331,H59332,H59333,H59339,H59341,H59342,H59343,H59349<br>H59351,H59352,H59353,H59359,H59361,H59362,H59363,H59369,H5940<br>H5941,H5942,H5943,H59811,H59812,H59813,H59819,H5988,H5989 | 37960,37961,37962,37963,99799,99811,99812,9982,99882 |
| Postprocedural or postoperative genitourinary system complication | N980,N981,N982,N983,N988,N989,N990,N99110,N99111,N99112<br>N99113,N99114,N99115,N99116,N9912,N992,N993,N994,N99510<br>N99511,N99512,N99518,N99520,N99521,N99522,N99523,N99524<br>N99528,N99530,N99531,N99532,N99533,N99534,N99538,N9961,N9962 | 2561,5680,59681,59682,59683,5982,6185,6208,6232,9975,99799<br>99811,99812,9982,99939 |

Continued on next page

**Supplementary Table S1:** (cont'd) A comprehensive mapping of diagnosis groups used in the experiments to their corresponding ICD-10 and ICD-9 codes.

| Diagnosis | ICD-10 | ICD-9 |
|---|---|---|
| | N9971,N9972,N9981,N99820,N99821,N9983,N99840,N99841,N99842 N99843,N9985,N9989 | |
| Postprocedural or postoperative musculoskeletal system complication | M960,M961,M962,M963,M964,M965,M96621,M96622,M96629,M96631 M96632,M96639,M9665,M96661,M96662,M96669,M96671,M96672,M96679 M9669,M96810,M96811,M96820,M96821,M96830,M96831,M96840,M96841 M96842,M96843,M9689 | 72280,72281,72282,72283,73711,73712,73721,73722,73733,99649 99799,99811,99812,9982,V454 |
| Postprocedural or postoperative nervous system complication | G970,G971,G972,G9731,G9732,G9741,G9748,G9749,G9751,G9752 G9761,G9762,G9763,G9764,G9781,G9782,G9783,G9784 | 3490,3491,34931,99701,99702,99709,99811,99812,9982 |
| Postprocedural or postoperative respiratory system complication | J9500,J9501,J9502,J9503,J9504,J9509,J951,J952,J953,J954,J955 J9561,J9562,J9571,J9572,J95811,J95812,J95821,J95822,J95830 J95831,J9584,J95850,J95851,J95859,J95860,J95861,J95862,J95863 J9587,J9588,J9589 | 5121,5122,51851,51852,51853,5187,51900,51901,51902,51909 99731,99732,99739,99811,99812,9982,V4614 |
| Postprocedural or postoperative skin complication | L7601,L7602,L7611,L7612,L7621,L7622,L7631,L7632,L7633,L7634 L7681,L7682 | 99811,99812,9982,99889 |
| Postthrombotic syndrome and venous insufficiency hypertension | I868,I87001,I87002,I87003,I87009,I87011,I87012,I87013,I87019 I87021,I87022,I87023,I87029,I87031,I87032,I87033,I87039 I87091,I87092,I87093,I87099,I871,I872,I87301,I87302,I87303 I87309,I87311,I87312,I87313,I87319,I87321,I87322,I87323 I87329,I87331,I87332,I87333,I87339,I87391,I87392,I87393 I87399 | 4568,45910,45911,45912,45913,45919,4592,45930,45931,45932 45933,45939,45981 |
| Pressure ulcer of skin | E08621,E08622,E09621,E09622,E10621,E10622,E11621,E11622 E13621,E13622,L89000,L89001,L89002,L89003,L89004,L89006 L89009,L89010,L89011,L89012,L89013,L89014,L89016,L89019 L89020,L89021,L89022,L89023,L89024,L89026,L89029,L89100 L89101,L89102,L89103,L89104,L89106,L89109,L89110,L89111 L89112,L89113,L89114,L89116,L89119,L89120,L89121,L89122 L89123,L89124,L89126,L89129,L89130,L89131,L89132,L89133 L89134,L89136,L89139,L89140,L89141,L89142,L89143,L89144 L89146,L89149,L89150,L89151,L89152,L89153,L89154,L89156 L89159,L89200,L89201,L89202,L89203,L89204,L89206,L89209 L89210,L89211,L89212,L89213,L89214,L89216,L89219,L89220 L89221,L89222,L89223,L89224,L89226,L89229,L89300,L89301 | 24980,25080,25081,70700,70701,70702,70703,70704,70705,70706 70707,70709,70720,70721,70722,70723,70724,70725 |

Continued on next page

**Supplementary Table S1:** (cont'd) A comprehensive mapping of diagnosis groups used in the experiments to their corresponding ICD-10 and ICD-9 codes.

| Diagnosis | ICD-10 | ICD-9 |
|---|---|---|
| | L89302,L89303,L89304,L89306,L89309,L89310,L89311,L89312<br>L89313,L89314,L89316,L89319,L89320,L89321,L89322,L89323<br>L89324,L89326,L89329,L8940,L8941,L8942,L8943,L8944,L8945<br>L8946,L89500,L89501,L89502,L89503,L89504,L89506,L89509,L89510<br>L89511,L89512,L89513,L89514,L89516,L89519,L89520,L89521<br>L89522,L89523,L89524,L89526,L89529,L89600,L89601,L89602<br>L89603,L89604,L89606,L89609,L89610,L89611,L89612,L89613<br>L89614,L89616,L89619,L89620,L89621,L89622,L89623,L89624<br>L89626,L89629,L89810,L89811,L89812,L89813,L89814,L89816<br>L89819,L89890,L89891,L89892,L89893,L89894,L89896,L89899,L8990<br>L8991,L8992,L8993,L8994,L8995,L8996 | |
| Previous C-section | O3421,O34211,O34212,O34218,O34219,O3422 | 65421,65423 |
| Prolapse of female genital organs | N810,N8110,N8111,N8112,N812,N813,N814,N815,N816,N8181,N8182<br>N8183,N8184,N8185,N8189,N819 | 61800,61801,61802,61803,61804,61805,6182,6183,6184,6186,6187<br>61881,61882,61883,61884,61889,6189 |
| Prolonged pregnancy | O480,O481 | 64511,64513,64523 |
| Proteinuria | N060,N061,N062,N063,N064,N065,N066,N067,N068,N069,N06A,O1210<br>O1211,O1212,O1213,O1214,O1215,O1220,O1221,O1222,O1223,O1224<br>O1225,R800,R801,R802,R803,R808,R809 | 5831,5832,58389,5839,5936,64610,64611,64613,64620,64621,64623<br>7910 |
| Pulmonary heart disease | I270,I271,I272,I2720,I2721,I2722,I2723,I2724,I2729,I2781<br>I2782,I2783,I2789,I279,I280,I281,I288,I289 | 4160,4161,4162,4168,4169,4170,4171,4178,4179 |
| Refractive error | H5200,H5201,H5202,H5203,H5210,H5211,H5212,H5213,H52201,H52202<br>H52203,H52209,H52211,H52212,H52213,H52219,H52221,H52222<br>H52223,H52229,H5231,H5232,H524,H52511,H52512,H52513,H52519<br>H52521,H52522,H52523,H52529,H52531,H52532,H52533,H52539,H526<br>H527 | 3670,3671,36720,36721,36722,36731,36732,3674,36751,36752<br>36753,36789,3679 |
| Regional enteritis and ulcerative colitis | K5000,K50011,K50012,K50013,K50014,K50018,K50019,K5010,K50111<br>K50112,K50113,K50114,K50118,K50119,K5080,K50811,K50812,K50813<br>K50814,K50818,K50819,K5090,K50911,K50912,K50913,K50914,K50918<br>K50919,K5100,K51011,K51012,K51013,K51014,K51018,K51019,K5120<br>K51211,K51212,K51213,K51214,K51218,K51219,K5130,K51311,K51312<br>K51313,K51314,K51318,K51319,K5140,K51411,K51412,K51413,K51414<br>K51418,K51419,K5150,K51511,K51512,K51513,K51514,K51518,K51519<br>K5180,K51811,K51812,K51813,K51814,K51818,K51819,K5190,K51911 | 5550,5551,5552,5559,5562,5563,5564,5565,5566,5568,5569,56089<br>5609,566,5695,56981 |

Continued on next page

**Supplementary Table S1:** (cont'd) A comprehensive mapping of diagnosis groups used in the experiments to their corresponding ICD-10 and ICD-9 codes.

| Diagnosis | ICD-10 | ICD-9 |
|---|---|---|
| | K51912,K51913,K51914,K51918,K51919 | |
| Respiratory cancers | C33,C3400,C3401,C3402,C3410,C3411,C3412,C342,C3430,C3431<br>C3432,C3480,C3481,C3482,C3490,C3491,C3492,C384,C390,C399,D021<br>D0220,D0221,D0222,D023,D024 | 1620,1622,1623,1624,1625,1628,1629,1639,1650,1658,1659,2311<br>2312,2318,2319 |
| Respiratory congenital malformations | Q300,Q301,Q302,Q303,Q308,Q309,Q310,Q311,Q312,Q313,Q315,Q318<br>Q319,Q320,Q321,Q322,Q323,Q324,Q330,Q331,Q332,Q333,Q334,Q335<br>Q336,Q338,Q339,Q340,Q341,Q348,Q349 | 7480,7481,7482,7483,7484,7485,74860,74861,74869,7488,7489 |
| Respiratory distress syndrome | P220,P228,P229 | 769,77089 |
| Respiratory failure; insufficiency; arrest | J80,J95821,J95822,J9600,J9601,J9602,J9610,J9611,J9612,J9620<br>J9621,J9622,J9690,J9691,J9692,R092 | 51851,51853,51881,51882,51883,51884,7991 |
| Respiratory perinatal condition | P230,P231,P232,P233,P234,P235,P236,P238,P239,P2400,P2401<br>P2410,P2411,P2420,P2421,P2430,P2431,P2480,P2481,P249,P250<br>P251,P252,P253,P258,P260,P261,P268,P269,P270,P271,P278,P279<br>P280,P2810,P2811,P2819,P282,P283,P2830,P2831,P2832,P2833<br>P2839,P284,P2840,P2841,P2842,P2843,P2849,P285,P2881,P2889<br>P289 | 7700,77010,77011,77012,77013,77014,77015,77016,77017,77018<br>7702,7703,7704,7705,7707,77081,77082,77083,77084,77085,77086<br>77087,77089,7709 |
| Respiratory signs and symptoms | R040,R041,R042,R0481,R0489,R049,R05,R051,R052,R053,R054,R058<br>R059,R0600,R0601,R0602,R0603,R0609,R061,R062,R063,R064,R065<br>R066,R067,R0681,R0682,R0683,R0689,R069,R070,R071,R0781,R0782<br>R0901,R0902,R093,R0981,R0982 | 47819,7841,7847,7848,78491,78499,78600,78601,78602,78603<br>78604,78605,78606,78607,78609,7861,7862,78630,78631,78639<br>7864,78652,78659,7868,7869,79901,79902 |
| Retinal and vitreous conditions | A1853,A5032,B5801,E08311,E08319,E08321,E083211,E083212<br>E083213,E083219,E08329,E083291,E083292,E083293,E083299,E08331<br>E083311,E083312,E083313,E083319,E08339,E083391,E083392<br>E083393,E083399,E08341,E083411,E083412,E083413,E083419,E08349<br>E083491,E083492,E083493,E083499,E08351,E083511,E083512<br>E083513,E083519,E083521,E083522,E083523,E083529,E083531<br>E083532,E083533,E083539,E083541,E083542,E083543,E083549<br>E083551,E083552,E083553,E083559,E08359,E083591,E083592<br>E083593,E083599,E09311,E09319,E09321,E093211,E093212,E093213<br>E093219,E09329,E093291,E093292,E093293,E093299,E09331,E093311<br>E093312,E093313,E093319,E09339,E093391,E093392,E093393<br>E093399,E09341,E093411,E093412,E093413,E093419,E09349,E093491 | 01730,0905,1302,24950,25050,25051,36013,36100,36101,36102<br>36103,36104,36105,36107,36110,36111,36112,36113,36114,36119<br>3612,36130,36131,36132,36133,36181,36189,36201,36202,36204<br>36205,36206,36207,36210,36211,36212,36213,36214,36215,36216<br>36217,36218,36220,36221,36222,36223,36224,36225,36226,36227<br>36229,36230,36231,36232,36233,36234,36235,36236,36237,36240<br>36241,36242,36243,36250,36251,36252,36253,36254,36255,36256<br>36257,36260,36261,36262,36263,36264,36265,36266,36270,36271<br>36272,36273,36274,36275,36276,36277,36281,36282,36283,36284<br>36285,36289,3629,36300,36301,36303,36304,36305,36306,36307<br>36308,36310,36311,36312,36313,36314,36315,36320,36321,36322<br>36330,36331,36332,36333,36334,36335,36340,36341,36342,36343 |

Continued on next page

**Supplementary Table S1:** (cont'd) A comprehensive mapping of diagnosis groups used in the experiments to their corresponding ICD-10 and ICD-9 codes.

| Diagnosis | ICD-10 | ICD-9 |
|---|---|---|
| | E093492,E093493,E093499,E09351,E093511,E093512,E093513 | 36350,36353,36354,36355,36356,36357,36361,36362,36363,36370 |
| | E093519,E093521,E093522,E093523,E093529,E093531,E093532 | 36371,36372,3638,3639,37913,37921,37922,37923,37924,37925 |
| | E093533,E093539,E093541,E093542,E093543,E093549,E093551 | 37926,37927,37929 |
| | E093552,E093553,E093559,E09359,E093591,E093592,E093593 | |
| | E093599,E10311,E10319,E10321,E103211,E103212,E103213,E103219 | |
| | E10329,E103291,E103292,E103293,E103299,E10331,E103311,E103312 | |
| | E103313,E103319,E10339,E103391,E103392,E103393,E103399,E10341 | |
| | E103411,E103412,E103413,E103419,E10349,E103491,E103492 | |
| | E103493,E103499,E10351,E103511,E103512,E103513,E103519 | |
| | E103521,E103522,E103523,E103529,E103531,E103532,E103533 | |
| | E103539,E103541,E103542,E103543,E103549,E103551,E103552 | |
| | E103553,E103559,E10359,E103591,E103592,E103593,E103599,E11311 | |
| | E11319,E11321,E113211,E113212,E113213,E113219,E11329,E113291 | |
| | E113292,E113293,E113299,E11331,E113311,E113312,E113313 | |
| | E113319,E11339,E113391,E113392,E113393,E113399,E11341,E113411 | |
| | E113412,E113413,E113419,E11349,E113491,E113492,E113493 | |
| | E113499,E11351,E113511,E113512,E113513,E113519,E113521 | |
| | E113522,E113523,E113529,E113531,E113532,E113533,E113539 | |
| | E113541,E113542,E113543,E113549,E113551,E113552,E113553 | |
| | E113559,E11359,E113591,E113592,E113593,E113599,E13311,E13319 | |
| | E13321,E133211,E133212,E133213,E133219,E13329,E133291,E133292 | |
| | E133293,E133299,E13331,E133311,E133312,E133313,E133319,E13339 | |
| | E133391,E133392,E133393,E133399,E13341,E133411,E133412 | |
| | E133413,E133419,E13349,E133491,E133492,E133493,E133499,E13351 | |
| | E133511,E133512,E133513,E133519,E133521,E133522,E133523 | |
| | E133529,E133531,E133532,E133533,E133539,E133541,E133542 | |
| | E133543,E133549,E133551,E133552,E133553,E133559,E13359 | |
| | E133591,E133592,E133593,E133599,H15811,H15812,H15813,H15819 | |
| | H30001,H30002,H30003,H30009,H30011,H30012,H30013,H30019 | |
| | H30021,H30022,H30023,H30029,H30031,H30032,H30033,H30039 | |
| | H30041,H30042,H30043,H30049,H30101,H30102,H30103,H30109 | |
| | H30111,H30112,H30113,H30119,H30121,H30122,H30123,H30129 | |
| | H30131,H30132,H30133,H30139,H30141,H30142,H30143,H30149,H3020 | |
| | H3021,H3022,H3023,H30811,H30812,H30813,H30819,H30891,H30892 | |
| | H30893,H30899,H3090,H3091,H3092,H3093,H31001,H31002,H31003 | |
| | H31009,H31011,H31012,H31013,H31019,H31021,H31022,H31023 | |
| | H31029,H31091,H31092,H31093,H31099,H31101,H31102,H31103 | |
| | H31109,H31111,H31112,H31113,H31119,H31121,H31122,H31123 | |
| | H31129,H3120,H3121,H3122,H3123,H3129,H31301,H31302,H31303 | |
| | H31309,H31311,H31312,H31313,H31319,H31321,H31322,H31323 | |

Continued on next page

**Supplementary Table S1:** (cont'd) A comprehensive mapping of diagnosis groups used in the experiments to their corresponding ICD-10 and ICD-9 codes.

| Diagnosis | ICD-10 | ICD-9 |
|---|---|---|
| | H31329,H31401,H31402,H31403,H31409,H31411,H31412,H31413 | |
| | H31419,H31421,H31422,H31423,H31429,H318,H319,H32,H33001 | |
| | H33002,H33003,H33009,H33011,H33012,H33013,H33019,H33021 | |
| | H33022,H33023,H33029,H33031,H33032,H33033,H33039,H33041 | |
| | H33042,H33043,H33049,H33051,H33052,H33053,H33059,H33101 | |
| | H33102,H33103,H33109,H33111,H33112,H33113,H33119,H33121 | |
| | H33122,H33123,H33129,H33191,H33192,H33193,H33199,H3320,H3321 | |
| | H3322,H3323,H33301,H33302,H33303,H33309,H33311,H33312,H33313 | |
| | H33319,H33321,H33322,H33323,H33329,H33331,H33332,H33333 | |
| | H33339,H3340,H3341,H3342,H3343,H338,H3400,H3401,H3402,H3403 | |
| | H3410,H3411,H3412,H3413,H34211,H34212,H34213,H34219,H34231 | |
| | H34232,H34233,H34239,H34811,H348110,H348111,H348112,H34812 | |
| | H348120,H348121,H348122,H34813,H348130,H348131,H348132,H34819 | |
| | H348190,H348191,H348192,H34821,H34822,H34823,H34829,H34831 | |
| | H348310,H348311,H348312,H34832,H348320,H348321,H348322,H34833 | |
| | H348330,H348331,H348332,H34839,H348390,H348391,H348392,H349 | |
| | H3500,H35011,H35012,H35013,H35019,H35021,H35022,H35023,H35029 | |
| | H35031,H35032,H35033,H35039,H35041,H35042,H35043,H35049 | |
| | H35051,H35052,H35053,H35059,H35061,H35062,H35063,H35069 | |
| | H35071,H35072,H35073,H35079,H3509,H35101,H35102,H35103,H35109 | |
| | H35111,H35112,H35113,H35119,H35121,H35122,H35123,H35129 | |
| | H35131,H35132,H35133,H35139,H35141,H35142,H35143,H35149 | |
| | H35151,H35152,H35153,H35159,H35161,H35162,H35163,H35169 | |
| | H35171,H35172,H35173,H35179,H3520,H3521,H3522,H3523,H3530 | |
| | H3531,H353110,H353111,H353112,H353113,H353114,H353120,H353121 | |
| | H353122,H353123,H353124,H353130,H353131,H353132,H353133 | |
| | H353134,H353190,H353191,H353192,H353193,H353194,H3532,H353210 | |
| | H353211,H353212,H353213,H353220,H353221,H353222,H353223 | |
| | H353230,H353231,H353232,H353233,H353290,H353291,H353292 | |
| | H353293,H3533,H35341,H35342,H35343,H35349,H35351,H35352 | |
| | H35353,H35359,H35361,H35362,H35363,H35369,H35371,H35372 | |
| | H35373,H35379,H35381,H35382,H35383,H35389,H3540,H35411,H35412 | |
| | H35413,H35419,H35421,H35422,H35423,H35429,H35431,H35432 | |
| | H35433,H35439,H35441,H35442,H35443,H35449,H35451,H35452 | |
| | H35453,H35459,H35461,H35462,H35463,H35469,H3550,H3551,H3552 | |
| | H3553,H3554,H3560,H3561,H3562,H3563,H3570,H35711,H35712 | |
| | H35713,H35719,H35721,H35722,H35723,H35729,H35731,H35732 | |
| | H35733,H35739,H3581,H3582,H3589,H359,H36,H4300,H4301,H4302 | |
| | H4303,H4310,H4311,H4312,H4313,H4320,H4321,H4322,H4323,H43311 | |
| | H43312,H43313,H43319,H43391,H43392,H43393,H43399,H43811 | |

Continued on next page

**Supplementary Table S1:** (cont'd) A comprehensive mapping of diagnosis groups used in the experiments to their corresponding ICD-10 and ICD-9 codes.

| Diagnosis | ICD-10 | ICD-9 |
|---|---|---|
| | H43812,H43813,H43819,H43821,H43822,H43823,H43829,H4389,H439 | |
| Rheumatoid arthritis | M0500,M05011,M05012,M05019,M05021,M05022,M05029,M05031,M05032 | 3571,3596,7140,7141,7142,71481,71489,7149,7200 |
| and related disease | M05039,M05041,M05042,M05049,M05051,M05052,M05059,M05061 | |
| | M05062,M05069,M05071,M05072,M05079,M0509,M0510,M05111,M05112 | |
| | M05119,M05121,M05122,M05129,M05131,M05132,M05139,M05141 | |
| | M05142,M05149,M05151,M05152,M05159,M05161,M05162,M05169 | |
| | M05171,M05172,M05179,M0519,M0520,M05211,M05212,M05219,M05221 | |
| | M05222,M05229,M05231,M05232,M05239,M05241,M05242,M05249 | |
| | M05251,M05252,M05259,M05261,M05262,M05269,M05271,M05272 | |
| | M05279,M0529,M0530,M05311,M05312,M05319,M05321,M05322,M05329 | |
| | M05331,M05332,M05339,M05341,M05342,M05349,M05351,M05352 | |
| | M05359,M05361,M05362,M05369,M05371,M05372,M05379,M0539,M0540 | |
| | M05411,M05412,M05419,M05421,M05422,M05429,M05431,M05432 | |
| | M05439,M05441,M05442,M05449,M05451,M05452,M05459,M05461 | |
| | M05462,M05469,M05471,M05472,M05479,M0549,M0550,M05511,M05512 | |
| | M05519,M05521,M05522,M05529,M05531,M05532,M05539,M05541 | |
| | M05542,M05549,M05551,M05552,M05559,M05561,M05562,M05569 | |
| | M05571,M05572,M05579,M0559,M0560,M05611,M05612,M05619,M05621 | |
| | M05622,M05629,M05631,M05632,M05639,M05641,M05642,M05649 | |
| | M05651,M05652,M05659,M05661,M05662,M05669,M05671,M05672 | |
| | M05679,M0569,M0570,M05711,M05712,M05719,M05721,M05722,M05729 | |
| | M05731,M05732,M05739,M05741,M05742,M05749,M05751,M05752 | |
| | M05759,M05761,M05762,M05769,M05771,M05772,M05779,M0579,M057A | |
| | M0580,M05811,M05812,M05819,M05821,M05822,M05829,M05831,M05832 | |
| | M05839,M05841,M05842,M05849,M05851,M05852,M05859,M05861 | |
| | M05862,M05869,M05871,M05872,M05879,M0589,M058A,M059,M0600 | |
| | M06011,M06012,M06019,M06021,M06022,M06029,M06031,M06032 | |
| | M06039,M06041,M06042,M06049,M06051,M06052,M06059,M06061 | |
| | M06062,M06069,M06071,M06072,M06079,M0608,M0609,M060A,M061 | |
| | M0620,M06211,M06212,M06219,M06221,M06222,M06229,M06231,M06232 | |
| | M06239,M06241,M06242,M06249,M06251,M06252,M06259,M06261 | |
| | M06262,M06269,M06271,M06272,M06279,M0628,M0629,M0630,M06311 | |
| | M06312,M06319,M06321,M06322,M06329,M06331,M06332,M06339 | |
| | M06341,M06342,M06349,M06351,M06352,M06359,M06361,M06362 | |
| | M06369,M06371,M06372,M06379,M0638,M0639,M064,M0680,M06811 | |
| | M06812,M06819,M06821,M06822,M06829,M06831,M06832,M06839 | |
| | M06841,M06842,M06849,M06851,M06852,M06859,M06861,M06862 | |
| | M06869,M06871,M06872,M06879,M0688,M0689,M068A,M069,M450,M451 | |
| | M452,M453,M454,M455,M456,M457,M458,M459,M45A0,M45A1,M45A2 | |

Continued on next page

**Supplementary Table S1:** (cont'd) A comprehensive mapping of diagnosis groups used in the experiments to their corresponding ICD-10 and ICD-9 codes.

| Diagnosis | ICD-10 | ICD-9 |
|---|---|---|
| | M45A3,M45A4,M45A5,M45A6,M45A7,M45A8,M45AB,M488X1,M488X2<br>M488X3,M488X4,M488X5,M488X6,M488X7,M488X8,M488X9 | |
| Sarcoma | C460,C461,C462,C463,C464,C4650,C4651,C4652,C467,C469,C490<br>C4910,C4911,C4912,C4920,C4921,C4922,C493,C494,C495,C496,C498<br>C499,C9230,C9231,C9232,C9622,C964,C96A | 1710,1712,1713,1714,1715,1716,1717,1718,1719,1760,1761,1762<br>1763,1764,1765,1768,1769,20230,20290,20530,20531,20532 |
| Schizophrenia spectrum and other psychotic disorders | F060,F061,F062,F10150,F10151,F10159,F10250,F10251,F10259<br>F10950,F10951,F10959,F11150,F11151,F11159,F11250,F11251<br>F11259,F11950,F11951,F11959,F12150,F12151,F12159,F12250<br>F12251,F12259,F12950,F12951,F12959,F13150,F13151,F13159<br>F13250,F13251,F13259,F13950,F13951,F13959,F14150,F14151<br>F14159,F14250,F14251,F14259,F14950,F14951,F14959,F15150<br>F15151,F15159,F15250,F15251,F15259,F15950,F15951,F15959<br>F16150,F16151,F16159,F16250,F16251,F16259,F16950,F16951<br>F16959,F18150,F18151,F18159,F18250,F18251,F18259,F18950<br>F18951,F18959,F19150,F19151,F19159,F19250,F19251,F19259<br>F19950,F19951,F19959,F200,F201,F202,F203,F205,F2081,F2089<br>F209,F21,F22,F23,F24,F250,F251,F258,F259,F28,F29,F53,F531 | 2913,2915,29189,29211,29212,29289,29381,29382,29389,2948<br>29510,29520,29530,29540,29560,29570,29580,29590,2970,2971<br>2972,2973,2983,2984,2988,2989,30122,30400,30410,30420,30430<br>30440,30450,30460 |
| Scoliosis and other postural dorsopathic deformities | M4000,M4003,M4004,M4005,M4010,M4012,M4013,M4014,M4015,M40202<br>M40203,M40204,M40205,M40209,M40292,M40293,M40294,M40295<br>M40299,M4030,M4035,M4036,M4037,M4040,M4045,M4046,M4047,M4050<br>M4055,M4056,M4057,M4100,M4102,M4103,M4104,M4105,M4106,M4107<br>M4108,M41112,M41113,M41114,M41115,M41116,M41117,M41119,M41122<br>M41123,M41124,M41125,M41126,M41127,M41129,M4120,M4122,M4123<br>M4124,M4125,M4126,M4127,M4130,M4134,M4135,M4140,M4141,M4142<br>M4143,M4144,M4145,M4146,M4147,M4150,M4152,M4153,M4154,M4155<br>M4156,M4157,M4180,M4182,M4183,M4184,M4185,M4186,M4187,M419<br>M4200,M4201,M4202,M4203,M4204,M4205,M4206,M4207,M4208,M4209<br>M4210,M4211,M4212,M4213,M4214,M4215,M4216,M4217,M4218,M4219<br>M429,M962,M963,M964,M965 | 7320,7328,7329,7370,73710,73711,73712,73719,73720,73721,73722<br>73729,73730,73731,73732,73733,73734,73739,73741,73742,73743 |
| Screening for neurocognitive or neurodevelopmental condition | Z134,Z1340,Z1341,Z1342,Z1349 | V792,V793,V798 |
| Secondary malignancies | C770,C771,C772,C773,C774,C775,C778,C779,C7800,C7801,C7802<br>C781,C782,C7830,C7839,C784,C785,C786,C787,C7880,C7889,C7900 | 1960,1961,1962,1963,1965,1966,1968,1969,1970,1971,1972,1973<br>1974,1975,1976,1977,1978,1980,1981,1982,1983,1984,1985,1986 |

Continued on next page

**Supplementary Table S1:** (cont'd) A comprehensive mapping of diagnosis groups used in the experiments to their corresponding ICD-10 and ICD-9 codes.

| Diagnosis | ICD-10 | ICD-9 |
|---|---|---|
| | C7901,C7902,C7910,C7911,C7919,C792,C7931,C7932,C7940,C7949<br>C7951,C7952,C7960,C7961,C7962,C7963,C7970,C7971,C7972,C7981<br>C7982,C7989,C799 | 1987,19881,19882,19889 |
| Sedative-related disorders | F1310,F13120,F13121,F13129,F13130,F13131,F13132,F13139,F1314<br>F13150,F13151,F13159,F13180,F13181,F13182,F13188,F1319,F1320<br>F13220,F13221,F13229,F13230,F13231,F13232,F13239,F1324,F13250<br>F13251,F13259,F1326,F1327,F13280,F13281,F13282,F13288,F1329<br>F1390,F1391,F13920,F13921,F13929,F13930,F13931,F13932,F13939<br>F1394,F13950,F13951,F13959,F1396,F1397,F13980,F13981,F13982<br>F13988,F1399,T426X1A,T426X2A,T426X3A,T426X4A,T426X5A,T4271XA<br>T4272XA,T4273XA,T4274XA,T4275XA | 2920,29211,29212,2922,29281,29282,29283,29284,29285,29289<br>2929,30410,30411,30412,30540,30541,30542,30543,9663,9678,9679<br>99529,E8528,E8588,E9363,E9378,E9379,E9502,E9503,E9504,E9620<br>E9804 |
| Septicemia | A021,A207,A227,A267,A327,A392,A393,A394,A400,A401,A403,A408<br>A409,A4101,A4102,A411,A412,A413,A414,A4150,A4151,A4152,A4153<br>A4159,A4181,A4189,A419,A427,A5486,B007,B377,I76,O0337,O0387<br>O0487,O0737,O0882,O85,O8604,P360,P3610,P3619,P362,P3630,P3639<br>P364,P365,P368,P369,R6520,R6521,T8112XA,T8144XA | 0031,0202,0223,0270,0271,0362,0380,03810,03811,03812,03819<br>0382,0383,03840,03841,03842,03843,03844,03849,0388,0389,0545<br>09889,1125,449,63400,63401,63402,63502,6380,6390,67022,67024<br>77181,78552,99591,99592,99802 |
| Sequela of cerebral infarction and other cerebrovascular disease | I6930,I6931,I69310,I69311,I69312,I69313,I69314,I69315,I69318<br>I69319,I69320,I69321,I69322,I69323,I69328,I69331,I69332<br>I69333,I69334,I69339,I69341,I69342,I69343,I69344,I69349<br>I69351,I69352,I69353,I69354,I69359,I69361,I69362,I69363<br>I69364,I69365,I69369,I69390,I69391,I69392,I69393,I69398,I6980<br>I6981,I69810,I69811,I69812,I69813,I69814,I69815,I69818,I69819<br>I69820,I69821,I69822,I69823,I69828,I69831,I69832,I69833<br>I69834,I69839,I69841,I69842,I69843,I69844,I69849,I69851<br>I69852,I69853,I69854,I69859,I69861,I69862,I69863,I69864<br>I69865,I69869,I69890,I69891,I69892,I69893,I69898,I6990,I6991<br>I69910,I69911,I69912,I69913,I69914,I69915,I69918,I69919<br>I69920,I69921,I69922,I69923,I69928,I69931,I69932,I69933<br>I69934,I69939,I69941,I69942,I69943,I69944,I69949,I69951<br>I69952,I69953,I69954,I69959,I69961,I69962,I69963,I69964<br>I69965,I69969,I69990,I69991,I69992,I69993,I69998 | 4380,43810,43811,43812,43813,43814,43819,43820,43821,43822<br>43830,43831,43832,43840,43841,43842,43850,43851,43852,43853<br>4386,4387,43881,43882,43883,43884,43889,4389 |
| Sequela of hemorrhagic cerebrovascular disease | I6900,I6901,I69010,I69011,I69012,I69013,I69014,I69015,I69018<br>I69019,I69020,I69021,I69022,I69023,I69028,I69031,I69032<br>I69033,I69034,I69039,I69041,I69042,I69043,I69044,I69049<br>I69051,I69052,I69053,I69054,I69059,I69061,I69062,I69063<br>I69064,I69065,I69069,I69090,I69091,I69092,I69093,I69098,I6910 | 4380,43811,43812,43813,43814,43819,43820,43821,43822,43830<br>43831,43832,43840,43841,43842,43850,43851,43852,43853,43881<br>43882,43883,43884,43889,4389 |

Continued on next page

**Supplementary Table S1:** (cont'd) A comprehensive mapping of diagnosis groups used in the experiments to their corresponding ICD-10 and ICD-9 codes.

| Diagnosis | ICD-10 | ICD-9 |
|---|---|---|
| | I6911,I69110,I69111,I69112,I69113,I69114,I69115,I69118,I69119<br>I69120,I69121,I69122,I69123,I69128,I69131,I69132,I69133<br>I69134,I69139,I69141,I69142,I69143,I69144,I69149,I69151<br>I69152,I69153,I69154,I69159,I69161,I69162,I69163,I69164<br>I69165,I69169,I69190,I69191,I69192,I69193,I69198,I6920,I6921<br>I69210,I69211,I69212,I69213,I69214,I69215,I69218,I69219<br>I69220,I69221,I69222,I69223,I69228,I69231,I69232,I69233<br>I69234,I69239,I69241,I69242,I69243,I69244,I69249,I69251<br>I69252,I69253,I69254,I69259,I69261,I69262,I69263,I69264<br>I69265,I69269,I69290,I69291,I69292,I69293,I69298 | |
| Sequela of malnutrition and other nutritional deficiencies | E640,E641,E642,E643,E648,E649,E68 | 2639,2649,267,2681,2698,2699,2788 |
| Sequela of specified infectious disease conditions | B900,B901,B902,B908,B909,B91,B92,B940,B941,B942,B948,B949 | 1370,1371,1372,1373,1374,138,1390,1391,1398 |
| Sequela of specified nervous system conditions | B91,B941,G09,G650,G651,G652 | 138,1390,326,3574 |
| Sexually transmitted infections (excluding HIV and hepatitis) | A5001,A5002,A5003,A5004,A5005,A5006,A5007,A5008,A5009,A501<br>A502,A5030,A5031,A5032,A5039,A5040,A5041,A5042,A5043,A5044<br>A5045,A5049,A5051,A5052,A5053,A5054,A5055,A5056,A5057,A5059<br>A506,A507,A509,A510,A511,A512,A5131,A5132,A5139,A5141,A5142<br>A5143,A5144,A5145,A5146,A5149,A515,A519,A5200,A5201,A5202<br>A5203,A5204,A5205,A5206,A5209,A5210,A5211,A5212,A5213,A5214<br>A5215,A5216,A5217,A5219,A522,A523,A5271,A5272,A5273,A5274<br>A5275,A5276,A5277,A5278,A5279,A528,A529,A530,A539,A5400,A5401<br>A5402,A5403,A5409,A541,A5421,A5422,A5423,A5424,A5429,A5430<br>A5431,A5432,A5433,A5439,A5440,A5441,A5442,A5443,A5449,A545<br>A546,A5481,A5482,A5483,A5484,A5485,A5486,A5489,A549,A55,A5600<br>A5601,A5602,A5609,A5611,A5619,A562,A563,A564,A568,A57,A58<br>A5900,A5901,A5902,A5903,A5909,A598,A599,A6000,A6001,A6002<br>A6003,A6004,A6009,A601,A609,A630,A638,A64,O98111,O98112<br>O98113,O98119,O9812,O9813,O98211,O98212,O98213,O98219,O9822<br>O9823,O98311,O98312,O98313,O98319,O9832,O9833 | 05410,05411,05412,05413,05419,07811,0900,0901,0902,0903,09040<br>09041,09042,09049,0905,0906,0907,0909,0910,0911,0912,0913<br>0914,09150,09151,09152,09161,09162,09169,0917,09181,09182<br>09189,0920,0929,0930,0931,09320,09321,09322,09323,09324,09381<br>09382,09389,0939,0940,0941,0942,0943,09481,09482,09483,09484<br>09485,09486,09489,0949,0950,0951,0952,0953,0954,0955,0956<br>0957,0958,0959,096,0970,0971,0979,0980,09811,09812,09813<br>09814,09815,09816,09819,0982,09831,09832,09833,09834,09835<br>09836,09840,09841,09842,09843,09849,09850,09851,09852,09853<br>09859,0986,0987,09881,09882,09883,09884,09885,09886,09889<br>0990,0991,0992,09951,09952,09953,09954,09955,09959,0998,0999<br>13100,13101,13102,13103,13109,1318,1319,3574,5954,6149,61611<br>64700,64701,64702,64703,64704,64710,64711,64712,64713,64714<br>64720,64721,64722,64723,64724,7135,99591 |

Continued on next page

**Supplementary Table S1:** (cont'd) A comprehensive mapping of diagnosis groups used in the experiments to their corresponding ICD-10 and ICD-9 codes.

| Diagnosis | ICD-10 | ICD-9 |
|---|---|---|
| Shock | O0331,O0381,O0481,O0731,O083,O751,R570,R571,R578,R579,R6521<br>T8110XA,T8111XA,T8112XA,T8119XA,T882XXA | 63450,63451,63452,63552,6385,6395,66911,66912,66914,78550<br>78551,78552,78559,9954,99592,99800,99801,99802,99809 |
| Short gestation; low birth weight; and fetal growth retardation | P0500,P0501,P0502,P0503,P0504,P0505,P0506,P0507,P0508,P0509<br>P0510,P0511,P0512,P0513,P0514,P0515,P0516,P0517,P0518,P0519<br>P052,P059,P0700,P0701,P0702,P0703,P0710,P0714,P0715,P0716<br>P0717,P0718,P0720,P0721,P0722,P0723,P0724,P0725,P0726,P0730<br>P0731,P0732,P0733,P0734,P0735,P0736,P0737,P0738,P0739 | 76400,76401,76402,76403,76404,76405,76406,76407,76408,76410<br>76411,76412,76413,76414,76415,76416,76417,76418,76420,76490<br>76500,76501,76502,76503,76504,76505,76506,76507,76508,76509<br>76510,76511,76512,76513,76514,76515,76516,76517,76518,76519<br>76520,76521,76522,76523,76524,76525,76526,76527,76528 |
| Sickle cell trait anemia | D5700,D5701,D5702,D5703,D5709,D571,D5720,D57211,D57212,D57213<br>D57218,D57219,D573,D5740,D57411,D57412,D57413,D57418,D57419<br>D5742,D57431,D57432,D57433,D57438,D57439,D5744,D57451,D57452<br>D57453,D57458,D57459,D5780,D57811,D57812,D57813,D57818,D57819 | 28241,28242,2825,28260,28261,28262,28263,28264,28268,28269<br>28952,5173 |
| Sinusitis | J0100,J0101,J0110,J0111,J0120,J0121,J0130,J0131,J0140,J0141<br>J0180,J0181,J0190,J0191,J320,J321,J322,J323,J324,J328,J329 | 4610,4611,4612,4613,4618,4619,4730,4731,4732,4733,4738,4739 |
| Skin and subcutaneous tissue infections | A201,A210,A220,A311,A363,A431,A46,A5139,H05011,H05012,H05013<br>H05019,L00,L0100,L0101,L0102,L0103,L0109,L011,L0201,L0202<br>L0203,L0211,L0212,L0213,L02211,L02212,L02213,L02214,L02215<br>L02216,L02219,L02221,L02222,L02223,L02224,L02225,L02226<br>L02229,L02231,L02232,L02233,L02234,L02235,L02236,L02239,L0231<br>L0232,L0233,L02411,L02412,L02413,L02414,L02415,L02416,L02419<br>L02421,L02422,L02423,L02424,L02425,L02426,L02429,L02431<br>L02432,L02433,L02434,L02435,L02436,L02439,L02511,L02512<br>L02519,L02521,L02522,L02529,L02531,L02532,L02539,L02611<br>L02612,L02619,L02621,L02622,L02629,L02631,L02632,L02639<br>L02811,L02818,L02821,L02828,L02831,L02838,L0291,L0292,L0293<br>L03011,L03012,L03019,L03021,L03022,L03029,L03031,L03032<br>L03039,L03041,L03042,L03049,L03111,L03112,L03113,L03114<br>L03115,L03116,L03119,L03121,L03122,L03123,L03124,L03125<br>L03126,L03129,L03211,L03212,L03213,L03221,L03222,L03311<br>L03312,L03313,L03314,L03315,L03316,L03317,L03319,L03321<br>L03322,L03323,L03324,L03325,L03326,L03327,L03329,L03811<br>L03818,L03891,L03898,L0390,L0391,L040,L041,L042,L043,L048<br>L049,L0501,L0502,L0591,L0592,L080,L081,L0881,L0882,L0889,L089<br>L303 | 0201,0210,0220,0311,03285,035,0390,0913,37601,6800,6801,6802<br>6803,6804,6805,6806,6807,6808,6809,68100,68101,68102,68110<br>68111,6819,6820,6821,6822,6823,6824,6825,6826,6827,6828,6829<br>683,684,6850,6851,68600,68609,6868,6869,6908,69581 |
| Skin cancers - all | C4400,C4409,C44101,C44102,C441021,C441022,C44109,C441091 | 17300,17309,17310,17319,17320,17329,17330,17339,17340,17349 |

Continued on next page

**Supplementary Table S1:** (cont'd) A comprehensive mapping of diagnosis groups used in the experiments to their corresponding ICD-10 and ICD-9 codes.

| Diagnosis | ICD-10 | ICD-9 |
|---|---|---|
| other types | C441092,C44131,C441321,C441322,C441391,C441392,C44191,C44192<br>C441921,C441922,C44199,C441991,C441992,C44201,C44202,C44209<br>C44291,C44292,C44299,C44300,C44301,C44309,C44390,C44391<br>C44399,C4440,C4449,C44500,C44501,C44509,C44590,C44591,C44599<br>C44601,C44602,C44609,C44691,C44692,C44699,C44701,C44702<br>C44709,C44791,C44792,C44799,C4480,C4489,C4490,C4499,C4A0<br>C4A10,C4A11,C4A111,C4A112,C4A12,C4A121,C4A122,C4A20,C4A21<br>C4A22,C4A30,C4A31,C4A39,C4A4,C4A51,C4A52,C4A59,C4A60,C4A61<br>C4A62,C4A70,C4A71,C4A72,C4A8,C4A9,D040,D0410,D0411,D04111<br>D04112,D0412,D04121,D04122,D0420,D0421,D0422,D0430,D0439,D044<br>D045,D0460,D0461,D0462,D0470,D0471,D0472,D048,D049 | 17350,17359,17360,17369,17370,17379,17380,17389,17390,17399<br>20931,20932,20933,20934,20935,20936,2320,2321,2322,2323,2324<br>2325,2326,2327,2328,2329 |
| Skin cancers - basal cell carcinoma | C4401,C44111,C44112,C441121,C441122,C44119,C441191,C441192<br>C44211,C44212,C44219,C44310,C44311,C44319,C4441,C44510,C44511<br>C44519,C44611,C44612,C44619,C44711,C44712,C44719,C4481,C4491 | 17301,17311,17321,17331,17341,17351,17361,17371,17381,17391 |
| Skin cancers - melanoma | C430,C4310,C4311,C43111,C43112,C4312,C43121,C43122,C4320<br>C4321,C4322,C4330,C4331,C4339,C434,C4351,C4352,C4359,C4360<br>C4361,C4362,C4370,C4371,C4372,C438,C439,D030,D0310,D0311<br>D03111,D03112,D0312,D03121,D03122,D0320,D0321,D0322,D0330<br>D0339,D034,D0351,D0352,D0359,D0360,D0361,D0362,D0370,D0371<br>D0372,D038,D039 | 1720,1721,1722,1723,1724,1725,1726,1727,1728,1729 |
| Skin cancers - squamous cell carcinoma | C4402,C44121,C44122,C441221,C441222,C44129,C441291,C441292<br>C44221,C44222,C44229,C44320,C44321,C44329,C4442,C44520,C44521<br>C44529,C44621,C44622,C44629,C44721,C44722,C44729,C4482,C4492 | 17302,17312,17322,17332,17342,17352,17362,17372,17382,17392 |
| Skin Subcutaneous signs and symptoms | R200,R201,R202,R203,R208,R209,R21,R220,R221,R222,R2230,R2231<br>R2232,R2233,R2240,R2241,R2242,R2243,R229,R230,R231,R232,R233<br>R234,R238,R239 | 7820,7821,7822,7825,78261,78262,7827,7828,7829,7842,7866 |
| Sleep wake disorders | F19182,F19282,F19982,F5101,F5102,F5103,F5104,F5105,F5109<br>F5111,F5112,F5113,F5119,F513,F514,F515,F518,F519,G2581,G4700<br>G4701,G4709,G4710,G4711,G4712,G4713,G4714,G4719,G4720,G4721<br>G4722,G4723,G4724,G4725,G4726,G4727,G4729,G4730,G4731,G4732<br>G4733,G4734,G4735,G4736,G4737,G4739,G47411,G47419,G47421<br>G47429,G4750,G4751,G4752,G4753,G4754,G4759,G4761,G4762,G4763<br>G4769,G478,G479,R063 | 29285,30460,30740,30741,30742,30744,30746,30747,30749,32700<br>32701,32702,32709,32710,32711,32712,32713,32714,32715,32719<br>32720,32721,32722,32723,32724,32725,32726,32727,32729,32730<br>32731,32732,32733,32734,32735,32736,32737,32739,32740,32741<br>32742,32743,32744,32749,32751,32752,32753,32759,3278,33394<br>34700,34701,34710,34711,78050,78051,78052,78053,78054,78057<br>78604 |
| Somatic disorders | F450,F451,F4520,F4521,F4529,F4541,F4542,F458,F459,F6810,F6811 | 30016,3007,30081,30082,30089,30151,3060,3061,3062,3063,3064 |

Continued on next page

**Supplementary Table S1:** (cont'd) A comprehensive mapping of diagnosis groups used in the experiments to their corresponding ICD-10 and ICD-9 codes.

| Diagnosis | ICD-10 | ICD-9 |
|---|---|---|
| | F6812,F6813,F68A | 30650,30652,30653,30659,3067,3068,3069,30780,30789 |
| Spinal cord injury (SCI), initial encounter | S140XXA,S14101A,S14102A,S14103A,S14104A,S14105A,S14106A S14107A,S14108A,S14109A,S14111A,S14112A,S14113A,S14114A S14115A,S14116A,S14117A,S14118A,S14119A,S14121A,S14122A S14123A,S14124A,S14125A,S14126A,S14127A,S14128A,S14129A S14131A,S14132A,S14133A,S14134A,S14135A,S14136A,S14137A S14138A,S14139A,S14141A,S14142A,S14143A,S14144A,S14145A S14146A,S14147A,S14148A,S14149A,S14151A,S14152A,S14153A S14154A,S14155A,S14156A,S14157A,S14158A,S14159A,S240XXA S24101A,S24102A,S24103A,S24104A,S24109A,S24111A,S24112A S24113A,S24114A,S24119A,S24131A,S24132A,S24133A,S24134A S24139A,S24141A,S24142A,S24143A,S24144A,S24149A,S24151A S24152A,S24153A,S24154A,S24159A,S3401XA,S3402XA,S34101A S34102A,S34103A,S34104A,S34105A,S34109A,S34111A,S34112A S34113A,S34114A,S34115A,S34119A,S34121A,S34122A,S34123A S34124A,S34125A,S34129A,S34131A,S34132A,S34139A,S343XXA | 95200,95201,95202,95203,95204,95205,95206,95207,95208,95209 95210,95211,95212,95213,95214,95215,95216,95217,95218,95219 9522,9523,9524,9529 |
| Spinal cord injury (SCI), subsequent encounter | S140XXD,S14101D,S14102D,S14103D,S14104D,S14105D,S14106D S14107D,S14108D,S14109D,S14111D,S14112D,S14113D,S14114D S14115D,S14116D,S14117D,S14118D,S14119D,S14121D,S14122D S14123D,S14124D,S14125D,S14126D,S14127D,S14128D,S14129D S14131D,S14132D,S14133D,S14134D,S14135D,S14136D,S14137D S14138D,S14139D,S14141D,S14142D,S14143D,S14144D,S14145D S14146D,S14147D,S14148D,S14149D,S14151D,S14152D,S14153D S14154D,S14155D,S14156D,S14157D,S14158D,S14159D,S240XXD S24101D,S24102D,S24103D,S24104D,S24109D,S24111D,S24112D S24113D,S24114D,S24119D,S24131D,S24132D,S24133D,S24134D S24139D,S24141D,S24142D,S24143D,S24144D,S24149D,S24151D S24152D,S24153D,S24154D,S24159D,S3401XD,S3402XD,S34101D S34102D,S34103D,S34104D,S34105D,S34109D,S34111D,S34112D S34113D,S34114D,S34115D,S34119D,S34121D,S34122D,S34123D S34124D,S34125D,S34129D,S34131D,S34132D,S34139D,S343XXD | V5889 |
| Spondylopathies spondyloarthropathy (including infective) | A5441,L4053,M081,M4300,M4301,M4302,M4303,M4304,M4305,M4306 M4307,M4308,M4309,M4310,M4311,M4312,M4313,M4314,M4315,M4316 M4317,M4318,M4319,M4320,M4321,M4322,M4323,M4324,M4325,M4326 M4327,M4328,M433,M434,M435X2,M435X3,M435X4,M435X5,M435X6 M435X7,M435X8,M435X9,M436,M438X1,M438X2,M438X3,M438X4,M438X5 M438X6,M438X7,M438X8,M438X9,M439,M450,M451,M452,M453,M454 | 09853,6960,71838,71880,71888,7200,7201,7202,72081,72089,7209 7210,7211,7212,7213,72141,72142,7215,7216,7217,7218,72190 72191,7220,72210,72211,72231,72232,7224,72251,72252,72271 72272,72273,72280,72281,72282,72283,72290,72291,72292,72293 7230,7231,7232,7233,7234,7235,7238,72400,72401,72402,72409 7243,7244,7246,72470,72471,72479,7249,7292,73098,73740,7378 |

Continued on next page

**Supplementary Table S1:** (cont'd) A comprehensive mapping of diagnosis groups used in the experiments to their corresponding ICD-10 and ICD-9 codes.

| Diagnosis | ICD-10 | ICD-9 |
|---|---|---|
| | M455,M456,M457,M458,M459,M45A0,M45A1,M45A2,M45A3,M45A4,M45A5 | 7379,7384 |
| | M45A6,M45A7,M45A8,M45AB,M4600,M4601,M4602,M4603,M4604,M4605 | |
| | M4606,M4607,M4608,M4609,M461,M4630,M4631,M4632,M4633,M4634 | |
| | M4635,M4636,M4637,M4638,M4639,M4640,M4641,M4642,M4643,M4644 | |
| | M4645,M4646,M4647,M4648,M4649,M4650,M4651,M4652,M4653,M4654 | |
| | M4655,M4656,M4657,M4658,M4659,M4680,M4681,M4682,M4683,M4684 | |
| | M4685,M4686,M4687,M4688,M4689,M4690,M4691,M4692,M4693,M4694 | |
| | M4695,M4696,M4697,M4698,M4699,M47011,M47012,M47013,M47014 | |
| | M47015,M47016,M47019,M47021,M47022,M47029,M4710,M4711,M4712 | |
| | M4713,M4714,M4715,M4716,M4720,M4721,M4722,M4723,M4724,M4725 | |
| | M4726,M4727,M4728,M47811,M47812,M47813,M47814,M47815,M47816 | |
| | M47817,M47818,M47819,M47891,M47892,M47893,M47894,M47895 | |
| | M47896,M47897,M47898,M47899,M479,M4800,M4801,M4802,M4803 | |
| | M4804,M4805,M4806,M48061,M48062,M4807,M4808,M4810,M4811,M4812 | |
| | M4813,M4814,M4815,M4816,M4817,M4818,M4819,M4820,M4821,M4822 | |
| | M4823,M4824,M4825,M4826,M4827,M4830,M4831,M4832,M4833,M4834 | |
| | M4835,M4836,M4837,M4838,M488X1,M488X2,M488X3,M488X4,M488X5 | |
| | M488X6,M488X7,M488X8,M488X9,M489,M4980,M4981,M4982,M4983 | |
| | M4984,M4985,M4986,M4987,M4988,M4989,M5000,M5001,M5002,M50020 | |
| | M50021,M50022,M50023,M5003,M5010,M5011,M5012,M50120,M50121 | |
| | M50122,M50123,M5013,M5020,M5021,M5022,M50220,M50221,M50222 | |
| | M50223,M5023,M5030,M5031,M5032,M50320,M50321,M50322,M50323 | |
| | M5033,M5080,M5081,M5082,M50820,M50821,M50822,M50823,M5083 | |
| | M5090,M5091,M5092,M50920,M50921,M50922,M50923,M5093,M5104 | |
| | M5105,M5106,M5114,M5115,M5116,M5117,M5124,M5125,M5126,M5127 | |
| | M5134,M5135,M5136,M5137,M5144,M5145,M5146,M5147,M5184,M5185 | |
| | M5186,M5187,M519,M51A0,M51A1,M51A2,M51A3,M51A4,M51A5,M530 | |
| | M531,M532X1,M532X2,M532X3,M532X4,M532X5,M532X6,M532X7,M532X8 | |
| | M532X9,M533,M5380,M5381,M5382,M5383,M5384,M5385,M5386,M5387 | |
| | M5388,M539,M5410,M5411,M5412,M5413,M5414,M5415,M5416,M5417 | |
| | M5418,M542,M5430,M5431,M5432,M5440,M5441,M5442,M961 | |
| Spontaneous abortion | O030,O031,O032,O0330,O0331,O0332,O0333,O0334,O0335,O0336 | 63400,63401,63402,63410,63411,63412,63420,63421,63422,63430 |
| and complications of | O0337,O0338,O0339,O034,O035,O036,O037,O0380,O0381,O0382,O0383 | 63431,63432,63440,63441,63442,63450,63451,63452,63460,63461 |
| spontaneous abortion | O0384,O0385,O0386,O0387,O0388,O0389,O039 | 63462,63470,63471,63472,63480,63481,63482,63490,63491,63492 |
| Sprains and strains, | S0340XA,S0341XA,S0342XA,S0343XA,S034XXA,S038XXA,S039XXA | 8360,8361,8362,8400,8401,8402,8403,8404,8405,8406,8407,8408 |
| initial encounter | S0911XA,S134XXA,S135XXA,S138XXA,S139XXA,S161XXA,S233XXA | 8409,8410,8411,8412,8413,8418,8419,84200,84201,84202,84209 |
| | S2341XA,S23420A,S23421A,S23428A,S23429A,S238XXA,S239XXA | 84210,84212,84213,84219,8430,8431,8438,8439,8440,8441,8442 |
| | S29011A,S29012A,S29019A,S335XXA,S336XXA,S338XXA,S339XXA | 8443,8448,8449,84500,84501,84502,84503,84509,84510,84511 |

Continued on next page

**Supplementary Table S1:** (cont'd) A comprehensive mapping of diagnosis groups used in the experiments to their corresponding ICD-10 and ICD-9 codes.

| Diagnosis | ICD-10 | ICD-9 |
|---|---|---|
| | S39011A,S39012A,S39013A,S43401A,S43402A,S43409A,S43411A | 84512,84513,84519,8460,8461,8469,8470,8471,8472,8479,8481 |
| | S43412A,S43419A,S43421A,S43422A,S43429A,S43431A,S43432A | 8482,8483,84840,84841,84842,84849,8485,8488,95901 |
| | S43439A,S43491A,S43492A,S43499A,S4350XA,S4351XA,S4352XA | |
| | S4360XA,S4361XA,S4362XA,S4380XA,S4381XA,S4382XA,S4390XA | |
| | S4391XA,S4392XA,S46011A,S46012A,S46019A,S46111A,S46112A | |
| | S46119A,S46211A,S46212A,S46219A,S46311A,S46312A,S46319A | |
| | S46811A,S46812A,S46819A,S46911A,S46912A,S46919A,S5320XA | |
| | S5321XA,S5322XA,S5330XA,S5331XA,S5332XA,S53401A,S53402A | |
| | S53409A,S53411A,S53412A,S53419A,S53421A,S53422A,S53429A | |
| | S53431A,S53432A,S53439A,S53441A,S53442A,S53449A,S53491A | |
| | S53492A,S53499A,S56011A,S56012A,S56019A,S56111A,S56112A | |
| | S56113A,S56114A,S56115A,S56116A,S56117A,S56118A,S56119A | |
| | S56211A,S56212A,S56219A,S56311A,S56312A,S56319A,S56411A | |
| | S56412A,S56413A,S56414A,S56415A,S56416A,S56417A,S56418A | |
| | S56419A,S56511A,S56512A,S56519A,S56811A,S56812A,S56819A | |
| | S56911A,S56912A,S56919A,S63301A,S63302A,S63309A,S63311A | |
| | S63312A,S63319A,S63321A,S63322A,S63329A,S63331A,S63332A | |
| | S63339A,S63391A,S63392A,S63399A,S63400A,S63401A,S63402A | |
| | S63403A,S63404A,S63405A,S63406A,S63407A,S63408A,S63409A | |
| | S63410A,S63411A,S63412A,S63413A,S63414A,S63415A,S63416A | |
| | S63417A,S63418A,S63419A,S63420A,S63421A,S63422A,S63423A | |
| | S63424A,S63425A,S63426A,S63427A,S63428A,S63429A,S63430A | |
| | S63431A,S63432A,S63433A,S63434A,S63435A,S63436A,S63437A | |
| | S63438A,S63439A,S63490A,S63491A,S63492A,S63493A,S63494A | |
| | S63495A,S63496A,S63497A,S63498A,S63499A,S63501A,S63502A | |
| | S63509A,S63511A,S63512A,S63519A,S63521A,S63522A,S63529A | |
| | S63591A,S63592A,S63599A,S63601A,S63602A,S63609A,S63610A | |
| | S63611A,S63612A,S63613A,S63614A,S63615A,S63616A,S63617A | |
| | S63618A,S63619A,S63621A,S63622A,S63629A,S63630A,S63631A | |
| | S63632A,S63633A,S63634A,S63635A,S63636A,S63637A,S63638A | |
| | S63639A,S63641A,S63642A,S63649A,S63650A,S63651A,S63652A | |
| | S63653A,S63654A,S63655A,S63656A,S63657A,S63658A,S63659A | |
| | S63681A,S63682A,S63689A,S63690A,S63691A,S63692A,S63693A | |
| | S63694A,S63695A,S63696A,S63697A,S63698A,S63699A,S638X1A | |
| | S638X2A,S638X9A,S6390XA,S6391XA,S6392XA,S66011A,S66012A | |
| | S66019A,S66110A,S66111A,S66112A,S66113A,S66114A,S66115A | |
| | S66116A,S66117A,S66118A,S66119A,S66211A,S66212A,S66219A | |
| | S66310A,S66311A,S66312A,S66313A,S66314A,S66315A,S66316A | |
| | S66317A,S66318A,S66319A,S66411A,S66412A,S66419A,S66510A | |
| | S66511A,S66512A,S66513A,S66514A,S66515A,S66516A,S66517A | |

Continued on next page

**Supplementary Table S1:** (cont'd) A comprehensive mapping of diagnosis groups used in the experiments to their corresponding ICD-10 and ICD-9 codes.

| Diagnosis | ICD-10 | ICD-9 |
|---|---|---|
| | S66518A,S66519A,S66811A,S66812A,S66819A,S66911A,S66912A | |
| | S66919A,S73101A,S73102A,S73109A,S73111A,S73112A,S73119A | |
| | S73121A,S73122A,S73129A,S73191A,S73192A,S73199A,S76011A | |
| | S76012A,S76019A,S76111A,S76112A,S76119A,S76211A,S76212A | |
| | S76219A,S76311A,S76312A,S76319A,S76811A,S76812A,S76819A | |
| | S76911A,S76912A,S76919A,S83200A,S83201A,S83202A,S83203A | |
| | S83204A,S83205A,S83206A,S83207A,S83209A,S83211A,S83212A | |
| | S83219A,S83221A,S83222A,S83229A,S83231A,S83232A,S83239A | |
| | S83241A,S83242A,S83249A,S83251A,S83252A,S83259A,S83261A | |
| | S83262A,S83269A,S83271A,S83272A,S83279A,S83281A,S83282A | |
| | S83289A,S8330XA,S8331XA,S8332XA,S83401A,S83402A,S83409A | |
| | S83411A,S83412A,S83419A,S83421A,S83422A,S83429A,S83501A | |
| | S83502A,S83509A,S83511A,S83512A,S83519A,S83521A,S83522A | |
| | S83529A,S8360XA,S8361XA,S8362XA,S838X1A,S838X2A,S838X9A | |
| | S8390XA,S8391XA,S8392XA,S86011A,S86012A,S86019A,S86111A | |
| | S86112A,S86119A,S86211A,S86212A,S86219A,S86311A,S86312A | |
| | S86319A,S86811A,S86812A,S86819A,S86911A,S86912A,S86919A | |
| | S93401A,S93402A,S93409A,S93411A,S93412A,S93419A,S93421A | |
| | S93422A,S93429A,S93431A,S93432A,S93439A,S93491A,S93492A | |
| | S93499A,S93501A,S93502A,S93503A,S93504A,S93505A,S93506A | |
| | S93509A,S93511A,S93512A,S93513A,S93514A,S93515A,S93516A | |
| | S93519A,S93521A,S93522A,S93523A,S93524A,S93525A,S93526A | |
| | S93529A,S93601A,S93602A,S93609A,S93611A,S93612A,S93619A | |
| | S93621A,S93622A,S93629A,S93691A,S93692A,S93699A,S96011A | |
| | S96012A,S96019A,S96111A,S96112A,S96119A,S96211A,S96212A | |
| | S96219A,S96811A,S96812A,S96819A,S96911A,S96912A,S96919A | |
| Sprains and strains, | S0340XD,S0341XD,S0342XD,S0343XD,S034XXD,S038XXD,S039XXD | V5889 |
| subsequent encounter | S0911XD,S134XXD,S135XXD,S138XXD,S139XXD,S161XXD,S233XXD | |
| | S2341XD,S23420D,S23421D,S23428D,S23429D,S238XXD,S239XXD | |
| | S29011D,S29012D,S29019D,S335XXD,S336XXD,S338XXD,S339XXD | |
| | S39011D,S39012D,S39013D,S43401D,S43402D,S43409D,S43411D | |
| | S43412D,S43419D,S43421D,S43422D,S43429D,S43431D,S43432D | |
| | S43439D,S43491D,S43492D,S43499D,S4350XD,S4351XD,S4352XD | |
| | S4360XD,S4361XD,S4362XD,S4380XD,S4381XD,S4382XD,S4390XD | |
| | S4391XD,S4392XD,S46011D,S46012D,S46019D,S46111D,S46112D | |
| | S46119D,S46211D,S46212D,S46219D,S46311D,S46312D,S46319D | |
| | S46811D,S46812D,S46819D,S46911D,S46912D,S46919D,S5320XD | |
| | S5321XD,S5322XD,S5330XD,S5331XD,S5332XD,S53401D,S53402D | |
| | S53409D,S53411D,S53412D,S53419D,S53421D,S53422D,S53429D | |

Continued on next page

**Supplementary Table S1:** (cont'd) A comprehensive mapping of diagnosis groups used in the experiments to their corresponding ICD-10 and ICD-9 codes.

| Diagnosis | ICD-10 | ICD-9 |
|---|---|---|
| | S53431D,S53432D,S53439D,S53441D,S53442D,S53449D,S53491D | |
| | S53492D,S53499D,S56011D,S56012D,S56019D,S56111D,S56112D | |
| | S56113D,S56114D,S56115D,S56116D,S56117D,S56118D,S56119D | |
| | S56211D,S56212D,S56219D,S56311D,S56312D,S56319D,S56411D | |
| | S56412D,S56413D,S56414D,S56415D,S56416D,S56417D,S56418D | |
| | S56419D,S56511D,S56512D,S56519D,S56811D,S56812D,S56819D | |
| | S56911D,S56912D,S56919D,S63301D,S63302D,S63309D,S63311D | |
| | S63312D,S63319D,S63321D,S63322D,S63329D,S63331D,S63332D | |
| | S63339D,S63391D,S63392D,S63399D,S63400D,S63401D,S63402D | |
| | S63403D,S63404D,S63405D,S63406D,S63407D,S63408D,S63409D | |
| | S63410D,S63411D,S63412D,S63413D,S63414D,S63415D,S63416D | |
| | S63417D,S63418D,S63419D,S63420D,S63421D,S63422D,S63423D | |
| | S63424D,S63425D,S63426D,S63427D,S63428D,S63429D,S63430D | |
| | S63431D,S63432D,S63433D,S63434D,S63435D,S63436D,S63437D | |
| | S63438D,S63439D,S63490D,S63491D,S63492D,S63493D,S63494D | |
| | S63495D,S63496D,S63497D,S63498D,S63499D,S63501D,S63502D | |
| | S63509D,S63511D,S63512D,S63519D,S63521D,S63522D,S63529D | |
| | S63591D,S63592D,S63599D,S63601D,S63602D,S63609D,S63610D | |
| | S63611D,S63612D,S63613D,S63614D,S63615D,S63616D,S63617D | |
| | S63618D,S63619D,S63621D,S63622D,S63629D,S63630D,S63631D | |
| | S63632D,S63633D,S63634D,S63635D,S63636D,S63637D,S63638D | |
| | S63639D,S63641D,S63642D,S63649D,S63650D,S63651D,S63652D | |
| | S63653D,S63654D,S63655D,S63656D,S63657D,S63658D,S63659D | |
| | S63681D,S63682D,S63689D,S63690D,S63691D,S63692D,S63693D | |
| | S63694D,S63695D,S63696D,S63697D,S63698D,S63699D,S638X1D | |
| | S638X2D,S638X9D,S6390XD,S6391XD,S6392XD,S66011D,S66012D | |
| | S66019D,S66110D,S66111D,S66112D,S66113D,S66114D,S66115D | |
| | S66116D,S66117D,S66118D,S66119D,S66211D,S66212D,S66219D | |
| | S66310D,S66311D,S66312D,S66313D,S66314D,S66315D,S66316D | |
| | S66317D,S66318D,S66319D,S66411D,S66412D,S66419D,S66510D | |
| | S66511D,S66512D,S66513D,S66514D,S66515D,S66516D,S66517D | |
| | S66518D,S66519D,S66811D,S66812D,S66819D,S66911D,S66912D | |
| | S66919D,S73101D,S73102D,S73109D,S73111D,S73112D,S73119D | |
| | S73121D,S73122D,S73129D,S73191D,S73192D,S73199D,S76011D | |
| | S76012D,S76019D,S76111D,S76112D,S76119D,S76211D,S76212D | |
| | S76219D,S76311D,S76312D,S76319D,S76811D,S76812D,S76819D | |
| | S76911D,S76912D,S76919D,S83200D,S83201D,S83202D,S83203D | |
| | S83204D,S83205D,S83206D,S83207D,S83209D,S83211D,S83212D | |
| | S83219D,S83221D,S83222D,S83229D,S83231D,S83232D,S83239D | |
| | S83241D,S83242D,S83249D,S83251D,S83252D,S83259D,S83261D | |

Continued on next page

**Supplementary Table S1:** (cont'd) A comprehensive mapping of diagnosis groups used in the experiments to their corresponding ICD-10 and ICD-9 codes.

| Diagnosis | ICD-10 | ICD-9 |
|---|---|---|
| | S83262D,S83269D,S83271D,S83272D,S83279D,S83281D,S83282D | |
| | S83289D,S8330XD,S8331XD,S8332XD,S83401D,S83402D,S83409D | |
| | S83411D,S83412D,S83419D,S83421D,S83422D,S83429D,S83501D | |
| | S83502D,S83509D,S83511D,S83512D,S83519D,S83521D,S83522D | |
| | S83529D,S8360XD,S8361XD,S8362XD,S838X1D,S838X2D,S838X9D | |
| | S8390XD,S8391XD,S8392XD,S86011D,S86012D,S86019D,S86111D | |
| | S86112D,S86119D,S86211D,S86212D,S86219D,S86311D,S86312D | |
| | S86319D,S86811D,S86812D,S86819D,S86911D,S86912D,S86919D | |
| | S93401D,S93402D,S93409D,S93411D,S93412D,S93419D,S93421D | |
| | S93422D,S93429D,S93431D,S93432D,S93439D,S93491D,S93492D | |
| | S93499D,S93501D,S93502D,S93503D,S93504D,S93505D,S93506D | |
| | S93509D,S93511D,S93512D,S93513D,S93514D,S93515D,S93516D | |
| | S93519D,S93521D,S93522D,S93523D,S93524D,S93525D,S93526D | |
| | S93529D,S93601D,S93602D,S93609D,S93611D,S93612D,S93619D | |
| | S93621D,S93622D,S93629D,S93691D,S93692D,S93699D,S96011D | |
| | S96012D,S96019D,S96111D,S96112D,S96119D,S96211D,S96212D | |
| | S96219D,S96811D,S96812D,S96819D,S96911D,S96912D,S96919D | |
| Stimulant-related | F1410,F14120,F14121,F14122,F14129,F1413,F1414,F14150,F14151 | 2920,29211,29212,2922,29281,29284,29285,29289,2929,30420 |
| disorders | F14159,F14180,F14181,F14182,F14188,F1419,F1420,F14220,F14221 | 30421,30422,30440,30441,30442,30560,30561,30562,30563,30570 |
| | F14222,F14229,F1423,F1424,F14250,F14251,F14259,F14280,F14281 | 30571,30572,30573,96970,96971,96972,96973,96979,97081,99529 |
| | F14282,F14288,F1429,F1490,F1491,F14920,F14921,F14922,F14929 | E8542,E9397,E9503,E9504,E9620,E9803,E9804 |
| | F1493,F1494,F14950,F14951,F14959,F14980,F14981,F14982,F14988 | |
| | F1499,F1510,F15120,F15121,F15122,F15129,F1513,F1514,F15150 | |
| | F15151,F15159,F15180,F15181,F15182,F15188,F1519,F1520,F15220 | |
| | F15221,F15222,F15229,F1523,F1524,F15250,F15251,F15259,F15280 | |
| | F15281,F15282,F15288,F1529,F1590,F1591,F15920,F15921,F15922 | |
| | F15929,F1593,F1594,F15950,F15951,F15959,F15980,F15981,F15982 | |
| | F15988,F1599,T405X1A,T405X2A,T405X3A,T405X4A,T405X5A,T43601A | |
| | T43602A,T43603A,T43604A,T43605A,T43611A,T43612A,T43613A | |
| | T43614A,T43615A,T43621A,T43622A,T43623A,T43624A,T43625A | |
| | T43631A,T43632A,T43633A,T43634A,T43635A,T43641A,T43642A | |
| | T43643A,T43644A,T43651A,T43652A,T43653A,T43654A,T43655A | |
| | T43691A,T43692A,T43693A,T43694A,T43695A | |
| Strabismus | H4900,H4901,H4902,H4903,H4910,H4911,H4912,H4913,H4920,H4921 | 27787,37800,37801,37802,37803,37804,37805,37806,37807,37808 |
| | H4922,H4923,H4930,H4931,H4932,H4933,H4940,H4941,H4942,H4943 | 37810,37811,37812,37813,37814,37815,37816,37817,37818,37820 |
| | H49811,H49812,H49813,H49819,H49881,H49882,H49883,H49889,H499 | 37821,37822,37823,37824,37830,37831,37833,37834,37835,37840 |
| | H5000,H50011,H50012,H50021,H50022,H50031,H50032,H50041,H50042 | 37841,37842,37843,37844,37845,37850,37851,37852,37853,37854 |
| | H5005,H5006,H5007,H5008,H5010,H50111,H50112,H50121,H50122 | 37855,37856,37860,37861,37862,37863,37871,37873,3789 |

Continued on next page

**Supplementary Table S1:** (cont'd) A comprehensive mapping of diagnosis groups used in the experiments to their corresponding ICD-10 and ICD-9 codes.

| Diagnosis | ICD-10 | ICD-9 |
|---|---|---|
| | H50131,H50132,H50141,H50142,H5015,H5016,H5017,H5018,H5021 H5022,H5030,H50311,H50312,H5032,H50331,H50332,H5034,H5040 H50411,H50412,H5042,H5043,H5050,H5051,H5052,H5053,H5054,H5055 H5060,H50611,H50612,H5069,H50811,H50812,H5089,H509 | |
| Stress fracture, initial encounter | M4840XA,M4841XA,M4842XA,M4843XA,M4844XA,M4845XA,M4846XA M4847XA,M4848XA,M4850XA,M4851XA,M4852XA,M4853XA,M4854XA M4855XA,M4856XA,M4857XA,M4858XA,M8430XA,M84311A,M84312A M84319A,M84321A,M84322A,M84329A,M84331A,M84332A,M84333A M84334A,M84339A,M84341A,M84342A,M84343A,M84344A,M84345A M84346A,M84350A,M84351A,M84352A,M84353A,M84359A,M84361A M84362A,M84363A,M84364A,M84369A,M84371A,M84372A,M84373A M84374A,M84375A,M84376A,M84377A,M84378A,M84379A,M8438XA | 73313,73393,73394,73395,73396,73397,73398 |
| Stress fracture, subsequent encounter | M4840XD,M4840XG,M4841XD,M4841XG,M4842XD,M4842XG,M4843XD M4843XG,M4844XD,M4844XG,M4845XD,M4845XG,M4846XD,M4846XG M4847XD,M4847XG,M4848XD,M4848XG,M4850XD,M4850XG,M4851XD M4851XG,M4852XD,M4852XG,M4853XD,M4853XG,M4854XD,M4854XG M4855XD,M4855XG,M4856XD,M4856XG,M4857XD,M4857XG,M4858XD M4858XG,M8430XD,M8430XG,M8430XK,M8430XP,M84311D,M84311G M84311K,M84311P,M84312D,M84312G,M84312K,M84312P,M84319D M84319G,M84319K,M84319P,M84321D,M84321G,M84321K,M84321P M84322D,M84322G,M84322K,M84322P,M84329D,M84329G,M84329K M84329P,M84331D,M84331G,M84331K,M84331P,M84332D,M84332G M84332K,M84332P,M84333D,M84333G,M84333K,M84333P,M84334D M84334G,M84334K,M84334P,M84339D,M84339G,M84339K,M84339P M84341D,M84341G,M84341K,M84341P,M84342D,M84342G,M84342K M84342P,M84343D,M84343G,M84343K,M84343P,M84344D,M84344G M84344K,M84344P,M84345D,M84345G,M84345K,M84345P,M84346D M84346G,M84346K,M84346P,M84350D,M84350G,M84350K,M84350P M84351D,M84351G,M84351K,M84351P,M84352D,M84352G,M84352K M84352P,M84353D,M84353G,M84353K,M84353P,M84359D,M84359G M84359K,M84359P,M84361D,M84361G,M84361K,M84361P,M84362D M84362G,M84362K,M84362P,M84363D,M84363G,M84363K,M84363P M84364D,M84364G,M84364K,M84364P,M84369D,M84369G,M84369K M84369P,M84371D,M84371G,M84371K,M84371P,M84372D,M84372G M84372K,M84372P,M84373D,M84373G,M84373K,M84373P,M84374D M84374G,M84374K,M84374P,M84375D,M84375G,M84375K,M84375P M84376D,M84376G,M84376K,M84376P,M84377D,M84377G,M84377K M84377P,M84378D,M84378G,M84378K,M84378P,M84379D,M84379G | 73381,73382,V5421,V5422,V5423,V5425,V5426,V5427,V5429 |

Continued on next page

**Supplementary Table S1:** (cont'd) A comprehensive mapping of diagnosis groups used in the experiments to their corresponding ICD-10 and ICD-9 codes.

| Diagnosis | ICD-10 | ICD-9 |
|---|---|---|
| | M84379K,M84379P,M8438XD,M8438XG,M8438XK,M8438XP | |
| Suicidal ideation | R45851,R4588,T1491,T1491XA,T360X2A,T361X2A,T362X2A,T363X2A | 9599,9600,9601,9602,9603,9604,9605,9606,9608,9609,9610,9614 |
| attempt intentional | T364X2A,T365X2A,T366X2A,T367X2A,T368X2A,T3692XA,T370X2A | 9615,9616,9617,9618,9619,9620,9621,9622,9623,9624,9627,9628 |
| self-harm | T371X2A,T372X2A,T373X2A,T374X2A,T375X2A,T378X2A,T3792XA | 9629,9630,9631,9634,9635,9638,9639,9640,9642,9643,9644,9645 |
| | T380X2A,T381X2A,T382X2A,T383X2A,T384X2A,T385X2A,T386X2A | 9646,9648,9649,96500,96501,96502,96509,9651,9654,9655,96561 |
| | T387X2A,T38802A,T38812A,T38892A,T38902A,T38992A,T39012A | 96569,9657,9659,9660,9661,9662,9663,9664,9670,9678,9679,9680 |
| | T39092A,T391X2A,T392X2A,T39312A,T39392A,T394X2A,T398X2A | 9682,9683,9684,9685,9689,96900,96901,96902,96903,96904,96905 |
| | T3992XA,T400X2A,T401X2A,T402X2A,T403X2A,T40412A,T40422A | 96909,9691,9692,9693,9694,9696,96970,96971,96972,96973,96979 |
| | T40492A,T404X2A,T405X2A,T40602A,T40692A,T40712A,T40722A | 9698,9699,9700,9701,97081,9710,9711,9712,9713,9719,9720,9721 |
| | T407X2A,T408X2A,T40902A,T40992A,T410X2A,T411X2A,T41202A | 9722,9723,9724,9725,9726,9727,9729,9730,9731,9732,9733,9734 |
| | T41292A,T413X2A,T4142XA,T415X2A,T420X2A,T421X2A,T422X2A | 9735,9736,9738,9739,9742,9744,9745,9747,9750,9752,9753,9754 |
| | T423X2A,T424X2A,T425X2A,T426X2A,T4272XA,T428X2A,T43012A | 9755,9756,9757,9758,9760,9761,9762,9763,9764,9765,9766,9767 |
| | T43022A,T431X2A,T43202A,T43212A,T43222A,T43292A,T433X2A | 9768,9769,9770,9772,9778,9779,9786,9788,9789,9790,9796,9799 |
| | T434X2A,T43502A,T43592A,T43602A,T43612A,T43622A,T43632A | 9800,9801,9802,9803,9808,9809,981,9820,9821,9822,9823,9824 |
| | T43642A,T43652A,T43692A,T438X2A,T4392XA,T440X2A,T441X2A | 9828,9830,9831,9832,9839,9849,9850,9851,9852,9853,9855,9856 |
| | T442X2A,T443X2A,T444X2A,T445X2A,T446X2A,T447X2A,T448X2A | 9858,9859,986,9871,9872,9873,9874,9875,9876,9877,9878,9879 |
| | T44902A,T44992A,T450X2A,T451X2A,T452X2A,T453X2A,T454X2A | 9880,9881,9882,9888,9889,9890,9891,9893,9894,9895,9896,9897 |
| | T45512A,T45522A,T45602A,T45612A,T45622A,T45692A,T457X2A | 98982,98984,98989,9899,9947,E9500,E9501,E9502,E9503,E9504 |
| | T458X2A,T4592XA,T460X2A,T461X2A,T462X2A,T463X2A,T464X2A | E9506,E9507,E9508,E9509,E9520,E9521,E9528,E9529,E9530,E9531 |
| | T465X2A,T466X2A,T467X2A,T468X2A,T46902A,T46992A,T470X2A | E9538,E954,E9550,E9551,E9552,E9553,E9554,E9555,E9556,E9557 |
| | T471X2A,T472X2A,T473X2A,T474X2A,T475X2A,T476X2A,T477X2A | E9559,E956,E9579,E9580,E9581,E9582,E9583,E9584,E9585,E9586 |
| | T478X2A,T4792XA,T480X2A,T481X2A,T48202A,T48292A,T483X2A | E9587,E9588,E9589,V6284 |
| | T484X2A,T485X2A,T486X2A,T48902A,T48992A,T490X2A,T491X2A | |
| | T492X2A,T493X2A,T494X2A,T495X2A,T496X2A,T497X2A,T498X2A | |
| | T4992XA,T500X2A,T501X2A,T502X2A,T503X2A,T504X2A,T505X2A | |
| | T506X2A,T507X2A,T508X2A,T50902A,T50912A,T50992A,T50A12A | |
| | T50A22A,T50A92A,T50B12A,T50B92A,T50Z12A,T50Z92A,T510X2A | |
| | T511X2A,T512X2A,T513X2A,T518X2A,T5192XA,T520X2A,T521X2A | |
| | T522X2A,T523X2A,T524X2A,T528X2A,T5292XA,T530X2A,T531X2A | |
| | T532X2A,T533X2A,T534X2A,T535X2A,T536X2A,T537X2A,T5392XA | |
| | T540X2A,T541X2A,T542X2A,T543X2A,T5492XA,T550X2A,T551X2A | |
| | T560X2A,T561X2A,T562X2A,T563X2A,T564X2A,T565X2A,T566X2A | |
| | T567X2A,T56812A,T56892A,T5692XA,T570X2A,T571X2A,T572X2A | |
| | T573X2A,T578X2A,T5792XA,T5802XA,T5812XA,T582X2A,T588X2A | |
| | T5892XA,T590X2A,T591X2A,T592X2A,T593X2A,T594X2A,T595X2A | |
| | T596X2A,T597X2A,T59812A,T59892A,T5992XA,T600X2A,T601X2A | |
| | T602X2A,T603X2A,T604X2A,T608X2A,T6092XA,T6102XA,T6112XA | |
| | T61772A,T61782A,T618X2A,T6192XA,T620X2A,T621X2A,T622X2A | |
| | T628X2A,T6292XA,T63002A,T63012A,T63022A,T63032A,T63042A | |

Continued on next page

**Supplementary Table S1:** (cont'd) A comprehensive mapping of diagnosis groups used in the experiments to their corresponding ICD-10 and ICD-9 codes.

| Diagnosis | ICD-10 | ICD-9 |
|---|---|---|
| | T63062A,T63072A,T63082A,T63092A,T63112A,T63122A,T63192A<br>T632X2A,T63302A,T63312A,T63322A,T63332A,T63392A,T63412A<br>T63422A,T63432A,T63442A,T63452A,T63462A,T63482A,T63512A<br>T63592A,T63612A,T63622A,T63632A,T63692A,T63712A,T63792A<br>T63812A,T63822A,T63832A,T63892A,T6392XA,T6402XA,T6482XA<br>T650X2A,T651X2A,T65212A,T65222A,T65292A,T653X2A,T654X2A<br>T655X2A,T656X2A,T65812A,T65822A,T65832A,T65892A,T6592XA<br>T71112A,T71122A,T71132A,T71152A,T71162A,T71192A,T71222A<br>T71232A,X710XXA,X711XXA,X712XXA,X713XXA,X718XXA,X719XXA<br>X72XXXA,X730XXA,X731XXA,X732XXA,X738XXA,X739XXA,X7401XA<br>X7402XA,X7409XA,X748XXA,X749XXA,X75XXXA,X76XXXA,X770XXA<br>X771XXA,X772XXA,X773XXA,X778XXA,X779XXA,X780XXA,X781XXA<br>X782XXA,X788XXA,X789XXA,X79XXXA,X80XXXA,X810XXA,X811XXA<br>X818XXA,X820XXA,X821XXA,X822XXA,X828XXA,X830XXA,X831XXA<br>X832XXA,X838XXA | |
| Suicide attempt intentional self-harm; subsequent encounter | T1491XD,T360X2D,T361X2D,T362X2D,T363X2D,T364X2D,T365X2D<br>T366X2D,T367X2D,T368X2D,T3692XD,T370X2D,T371X2D,T372X2D<br>T373X2D,T374X2D,T375X2D,T378X2D,T3792XD,T380X2D,T381X2D<br>T382X2D,T383X2D,T384X2D,T385X2D,T386X2D,T387X2D,T38802D<br>T38812D,T38892D,T38902D,T38992D,T39012D,T39092D,T391X2D<br>T392X2D,T39312D,T39392D,T394X2D,T398X2D,T3992XD,T400X2D<br>T401X2D,T402X2D,T403X2D,T40412D,T40422D,T40492D,T404X2D<br>T405X2D,T40602D,T40692D,T40712D,T40722D,T407X2D,T408X2D<br>T40902D,T40992D,T410X2D,T411X2D,T41202D,T41292D,T413X2D<br>T4142XD,T415X2D,T420X2D,T421X2D,T422X2D,T423X2D,T424X2D<br>T425X2D,T426X2D,T4272XD,T428X2D,T43012D,T43022D,T431X2D<br>T43202D,T43212D,T43222D,T43292D,T433X2D,T434X2D,T43502D<br>T43592D,T43602D,T43612D,T43622D,T43632D,T43642D,T43652D<br>T43692D,T438X2D,T4392XD,T440X2D,T441X2D,T442X2D,T443X2D<br>T444X2D,T445X2D,T446X2D,T447X2D,T448X2D,T44902D,T44992D<br>T450X2D,T451X2D,T452X2D,T453X2D,T454X2D,T45512D,T45522D<br>T45602D,T45612D,T45622D,T45692D,T457X2D,T458X2D,T4592XD<br>T460X2D,T461X2D,T462X2D,T463X2D,T464X2D,T465X2D,T466X2D<br>T467X2D,T468X2D,T46902D,T46992D,T470X2D,T471X2D,T472X2D<br>T473X2D,T474X2D,T475X2D,T476X2D,T477X2D,T478X2D,T4792XD<br>T480X2D,T481X2D,T48202D,T48292D,T483X2D,T484X2D,T485X2D<br>T486X2D,T48902D,T48992D,T490X2D,T491X2D,T492X2D,T493X2D<br>T494X2D,T495X2D,T496X2D,T497X2D,T498X2D,T4992XD,T500X2D<br>T501X2D,T502X2D,T503X2D,T504X2D,T505X2D,T506X2D,T507X2D | E954,E9550,E9551,E9552,E9553,E9554,E9555,E9556,E9557,E9559<br>E956,E9579,E9580,E9581,E9582,E9583,E9584,E9585,E9586,E9587<br>E9588,V5889 |

Continued on next page

**Supplementary Table S1:** (cont'd) A comprehensive mapping of diagnosis groups used in the experiments to their corresponding ICD-10 and ICD-9 codes.

| Diagnosis | ICD-10 | ICD-9 |
|---|---|---|
| | T508X2D,T50902D,T50912D,T50992D,T50A12D,T50A22D,T50A92D | |
| | T50B12D,T50B92D,T50Z12D,T50Z92D,T510X2D,T511X2D,T512X2D | |
| | T513X2D,T518X2D,T5192XD,T520X2D,T521X2D,T522X2D,T523X2D | |
| | T524X2D,T528X2D,T5292XD,T530X2D,T531X2D,T532X2D,T533X2D | |
| | T534X2D,T535X2D,T536X2D,T537X2D,T5392XD,T540X2D,T541X2D | |
| | T542X2D,T543X2D,T5492XD,T550X2D,T551X2D,T560X2D,T561X2D | |
| | T562X2D,T563X2D,T564X2D,T565X2D,T566X2D,T567X2D,T56812D | |
| | T56892D,T5692XD,T570X2D,T571X2D,T572X2D,T573X2D,T578X2D | |
| | T5792XD,T5802XD,T5812XD,T582X2D,T588X2D,T5892XD,T590X2D | |
| | T591X2D,T592X2D,T593X2D,T594X2D,T595X2D,T596X2D,T597X2D | |
| | T59812D,T59892D,T5992XD,T600X2D,T601X2D,T602X2D,T603X2D | |
| | T604X2D,T608X2D,T6092XD,T6102XD,T6112XD,T61772D,T61782D | |
| | T618X2D,T6192XD,T620X2D,T621X2D,T622X2D,T628X2D,T6292XD | |
| | T63002D,T63012D,T63022D,T63032D,T63042D,T63062D,T63072D | |
| | T63082D,T63092D,T63112D,T63122D,T63192D,T632X2D,T63302D | |
| | T63312D,T63322D,T63332D,T63392D,T63412D,T63422D,T63432D | |
| | T63442D,T63452D,T63462D,T63482D,T63512D,T63592D,T63612D | |
| | T63622D,T63632D,T63692D,T63712D,T63792D,T63812D,T63822D | |
| | T63832D,T63892D,T6392XD,T6402XD,T6482XD,T650X2D,T651X2D | |
| | T65212D,T65222D,T65292D,T653X2D,T654X2D,T655X2D,T656X2D | |
| | T65812D,T65822D,T65832D,T65892D,T6592XD,T71112D,T71122D | |
| | T71132D,T71152D,T71162D,T71192D,T71222D,T71232D,X710XXD | |
| | X711XXD,X712XXD,X713XXD,X718XXD,X719XXD,X72XXXD,X730XXD | |
| | X731XXD,X732XXD,X738XXD,X739XXD,X7401XD,X7402XD,X7409XD | |
| | X748XXD,X749XXD,X75XXXD,X76XXXD,X770XXD,X771XXD,X772XXD | |
| | X773XXD,X778XXD,X779XXD,X780XXD,X781XXD,X782XXD,X788XXD | |
| | X789XXD,X79XXXD,X80XXXD,X810XXD,X811XXD,X818XXD,X828XXD | |
| | X830XXD,X831XXD,X832XXD,X838XXD | |
| Superficial injury; | S0000XA,S0001XA,S0002XA,S0003XA,S0004XA,S0005XA,S0006XA | 9100,9102,9104,9106,9108,9110,9112,9114,9116,9118,9120,9122 |
| contusion, initial | S0007XA,S0010XA,S0011XA,S0012XA,S00201A,S00202A,S00209A | 9124,9126,9128,9130,9132,9134,9136,9138,9140,9142,9144,9146 |
| encounter | S00211A,S00212A,S00219A,S00221A,S00222A,S00229A,S00241A | 9148,9150,9152,9154,9156,9158,9160,9162,9164,9166,9168,9170 |
| | S00242A,S00249A,S00251A,S00252A,S00259A,S00261A,S00262A | 9172,9174,9176,9178,9180,9181,9182,920,9210,9211,9212,9213 |
| | S00269A,S00271A,S00272A,S00279A,S0030XA,S0031XA,S0032XA | 9220,9221,9222,92231,92232,92233,9224,9228,9229,92300,92301 |
| | S0033XA,S0034XA,S0035XA,S0036XA,S0037XA,S00401A,S00402A | 92302,92303,92310,92311,92320,92321,9233,92400,92401,92410 |
| | S00409A,S00411A,S00412A,S00419A,S00421A,S00422A,S00429A | 92411,92420,92421,9243,9244,9245,9248 |
| | S00431A,S00432A,S00439A,S00441A,S00442A,S00449A,S00451A | |
| | S00452A,S00459A,S00461A,S00462A,S00469A,S00471A,S00472A | |
| | S00479A,S00501A,S00502A,S00511A,S00512A,S00521A,S00522A | |
| | S00531A,S00532A,S00541A,S00542A,S00551A,S00552A,S00561A | |

Continued on next page

**Supplementary Table S1:** (cont'd) A comprehensive mapping of diagnosis groups used in the experiments to their corresponding ICD-10 and ICD-9 codes.

| Diagnosis | ICD-10 | ICD-9 |
|---|---|---|
| | S00562A,S00571A,S00572A,S0080XA,S0081XA,S0082XA,S0083XA | |
| | S0084XA,S0085XA,S0086XA,S0087XA,S0090XA,S0091XA,S0092XA | |
| | S0093XA,S0094XA,S0095XA,S0096XA,S0097XA,S0500XA,S0501XA | |
| | S0502XA,S0510XA,S0511XA,S0512XA,S100XXA,S1010XA,S1011XA | |
| | S1012XA,S1014XA,S1015XA,S1016XA,S1017XA,S1080XA,S1081XA | |
| | S1082XA,S1083XA,S1084XA,S1085XA,S1086XA,S1087XA,S1090XA | |
| | S1091XA,S1092XA,S1093XA,S1094XA,S1095XA,S1096XA,S1097XA | |
| | S2000XA,S2001XA,S2002XA,S20101A,S20102A,S20109A,S20111A | |
| | S20112A,S20119A,S20121A,S20122A,S20129A,S20141A,S20142A | |
| | S20149A,S20151A,S20152A,S20159A,S20161A,S20162A,S20169A | |
| | S20171A,S20172A,S20179A,S2020XA,S20211A,S20212A,S20213A | |
| | S20214A,S20219A,S20221A,S20222A,S20223A,S20224A,S20229A | |
| | S20301A,S20302A,S20303A,S20304A,S20309A,S20311A,S20312A | |
| | S20313A,S20314A,S20319A,S20321A,S20322A,S20323A,S20324A | |
| | S20329A,S20341A,S20342A,S20343A,S20344A,S20349A,S20351A | |
| | S20352A,S20353A,S20354A,S20359A,S20361A,S20362A,S20363A | |
| | S20364A,S20369A,S20371A,S20372A,S20373A,S20374A,S20379A | |
| | S20401A,S20402A,S20409A,S20411A,S20412A,S20419A,S20421A | |
| | S20422A,S20429A,S20441A,S20442A,S20449A,S20451A,S20452A | |
| | S20459A,S20461A,S20462A,S20469A,S20471A,S20472A,S20479A | |
| | S2090XA,S2091XA,S2092XA,S2094XA,S2095XA,S2096XA,S2097XA | |
| | S300XXA,S301XXA,S30201A,S30202A,S3021XA,S3022XA,S3023XA | |
| | S303XXA,S30810A,S30811A,S30812A,S30813A,S30814A,S30815A | |
| | S30816A,S30817A,S30820A,S30821A,S30822A,S30823A,S30824A | |
| | S30825A,S30826A,S30827A,S30840A,S30841A,S30842A,S30843A | |
| | S30844A,S30845A,S30846A,S30850A,S30851A,S30852A,S30853A | |
| | S30854A,S30855A,S30856A,S30857A,S30860A,S30861A,S30862A | |
| | S30863A,S30864A,S30865A,S30866A,S30867A,S30870A,S30871A | |
| | S30872A,S30873A,S30874A,S30875A,S30876A,S30877A,S3091XA | |
| | S3092XA,S3093XA,S3094XA,S3095XA,S3096XA,S3097XA,S3098XA | |
| | S40011A,S40012A,S40019A,S40021A,S40022A,S40029A,S40211A | |
| | S40212A,S40219A,S40221A,S40222A,S40229A,S40241A,S40242A | |
| | S40249A,S40251A,S40252A,S40259A,S40261A,S40262A,S40269A | |
| | S40271A,S40272A,S40279A,S40811A,S40812A,S40819A,S40821A | |
| | S40822A,S40829A,S40841A,S40842A,S40849A,S40851A,S40852A | |
| | S40859A,S40861A,S40862A,S40869A,S40871A,S40872A,S40879A | |
| | S40911A,S40912A,S40919A,S40921A,S40922A,S40929A,S5000XA | |
| | S5001XA,S5002XA,S5010XA,S5011XA,S5012XA,S50311A,S50312A | |
| | S50319A,S50321A,S50322A,S50329A,S50341A,S50342A,S50349A | |
| | S50351A,S50352A,S50359A,S50361A,S50362A,S50369A,S50371A | |

Continued on next page

**Supplementary Table S1:** (cont'd) A comprehensive mapping of diagnosis groups used in the experiments to their corresponding ICD-10 and ICD-9 codes.

| Diagnosis | ICD-10 | ICD-9 |
|---|---|---|
| | S50372A,S50379A,S50811A,S50812A,S50819A,S50821A,S50822A | |
| | S50829A,S50841A,S50842A,S50849A,S50851A,S50852A,S50859A | |
| | S50861A,S50862A,S50869A,S50871A,S50872A,S50879A,S50901A | |
| | S50902A,S50909A,S50911A,S50912A,S50919A,S6000XA,S60011A | |
| | S60012A,S60019A,S60021A,S60022A,S60029A,S60031A,S60032A | |
| | S60039A,S60041A,S60042A,S60049A,S60051A,S60052A,S60059A | |
| | S6010XA,S60111A,S60112A,S60119A,S60121A,S60122A,S60129A | |
| | S60131A,S60132A,S60139A,S60141A,S60142A,S60149A,S60151A | |
| | S60152A,S60159A,S60211A,S60212A,S60219A,S60221A,S60222A | |
| | S60229A,S60311A,S60312A,S60319A,S60321A,S60322A,S60329A | |
| | S60341A,S60342A,S60349A,S60351A,S60352A,S60359A,S60361A | |
| | S60362A,S60369A,S60371A,S60372A,S60379A,S60391A,S60392A | |
| | S60399A,S60410A,S60411A,S60412A,S60413A,S60414A,S60415A | |
| | S60416A,S60417A,S60418A,S60419A,S60420A,S60421A,S60422A | |
| | S60423A,S60424A,S60425A,S60426A,S60427A,S60428A,S60429A | |
| | S60440A,S60441A,S60442A,S60443A,S60444A,S60445A,S60446A | |
| | S60447A,S60448A,S60449A,S60450A,S60451A,S60452A,S60453A | |
| | S60454A,S60455A,S60456A,S60457A,S60458A,S60459A,S60460A | |
| | S60461A,S60462A,S60463A,S60464A,S60465A,S60466A,S60467A | |
| | S60468A,S60469A,S60470A,S60471A,S60472A,S60473A,S60474A | |
| | S60475A,S60476A,S60477A,S60478A,S60479A,S60511A,S60512A | |
| | S60519A,S60521A,S60522A,S60529A,S60541A,S60542A,S60549A | |
| | S60551A,S60552A,S60559A,S60561A,S60562A,S60569A,S60571A | |
| | S60572A,S60579A,S60811A,S60812A,S60819A,S60821A,S60822A | |
| | S60829A,S60841A,S60842A,S60849A,S60851A,S60852A,S60859A | |
| | S60861A,S60862A,S60869A,S60871A,S60872A,S60879A,S60911A | |
| | S60912A,S60919A,S60921A,S60922A,S60929A,S60931A,S60932A | |
| | S60939A,S60940A,S60941A,S60942A,S60943A,S60944A,S60945A | |
| | S60946A,S60947A,S60948A,S60949A,S7000XA,S7001XA,S7002XA | |
| | S7010XA,S7011XA,S7012XA,S70211A,S70212A,S70219A,S70221A | |
| | S70222A,S70229A,S70241A,S70242A,S70249A,S70251A,S70252A | |
| | S70259A,S70261A,S70262A,S70269A,S70271A,S70272A,S70279A | |
| | S70311A,S70312A,S70319A,S70321A,S70322A,S70329A,S70341A | |
| | S70342A,S70349A,S70351A,S70352A,S70359A,S70361A,S70362A | |
| | S70369A,S70371A,S70372A,S70379A,S70911A,S70912A,S70919A | |
| | S70921A,S70922A,S70929A,S8000XA,S8001XA,S8002XA,S8010XA | |
| | S8011XA,S8012XA,S80211A,S80212A,S80219A,S80221A,S80222A | |
| | S80229A,S80241A,S80242A,S80249A,S80251A,S80252A,S80259A | |
| | S80261A,S80262A,S80269A,S80271A,S80272A,S80279A,S80811A | |
| | S80812A,S80819A,S80821A,S80822A,S80829A,S80841A,S80842A | |

Continued on next page

**Supplementary Table S1:** (cont'd) A comprehensive mapping of diagnosis groups used in the experiments to their corresponding ICD-10 and ICD-9 codes.

| Diagnosis | ICD-10 | ICD-9 |
|---|---|---|
| | S80849A,S80851A,S80852A,S80859A,S80861A,S80862A,S80869A | |
| | S80871A,S80872A,S80879A,S80911A,S80912A,S80919A,S80921A | |
| | S80922A,S80929A,S9000XA,S9001XA,S9002XA,S90111A,S90112A | |
| | S90119A,S90121A,S90122A,S90129A,S90211A,S90212A,S90219A | |
| | S90221A,S90222A,S90229A,S9030XA,S9031XA,S9032XA,S90411A | |
| | S90412A,S90413A,S90414A,S90415A,S90416A,S90421A,S90422A | |
| | S90423A,S90424A,S90425A,S90426A,S90441A,S90442A,S90443A | |
| | S90444A,S90445A,S90446A,S90451A,S90452A,S90453A,S90454A | |
| | S90455A,S90456A,S90461A,S90462A,S90463A,S90464A,S90465A | |
| | S90466A,S90471A,S90472A,S90473A,S90474A,S90475A,S90476A | |
| | S90511A,S90512A,S90519A,S90521A,S90522A,S90529A,S90541A | |
| | S90542A,S90549A,S90551A,S90552A,S90559A,S90561A,S90562A | |
| | S90569A,S90571A,S90572A,S90579A,S90811A,S90812A,S90819A | |
| | S90821A,S90822A,S90829A,S90841A,S90842A,S90849A,S90851A | |
| | S90852A,S90859A,S90861A,S90862A,S90869A,S90871A,S90872A | |
| | S90879A,S90911A,S90912A,S90919A,S90921A,S90922A,S90929A | |
| | S90931A,S90932A,S90933A,S90934A,S90935A,S90936A | |
| Superficial injury; | S0000XD,S0001XD,S0002XD,S0003XD,S0004XD,S0005XD,S0006XD | V5889 |
| contusion, subsequent | S0007XD,S0010XD,S0011XD,S0012XD,S00201D,S00202D,S00209D | |
| encounter | S00211D,S00212D,S00219D,S00221D,S00222D,S00229D,S00241D | |
| | S00242D,S00249D,S00251D,S00252D,S00259D,S00261D,S00262D | |
| | S00269D,S00271D,S00272D,S00279D,S0030XD,S0031XD,S0032XD | |
| | S0033XD,S0034XD,S0035XD,S0036XD,S0037XD,S00401D,S00402D | |
| | S00409D,S00411D,S00412D,S00419D,S00421D,S00422D,S00429D | |
| | S00431D,S00432D,S00439D,S00441D,S00442D,S00449D,S00451D | |
| | S00452D,S00459D,S00461D,S00462D,S00469D,S00471D,S00472D | |
| | S00479D,S00501D,S00502D,S00511D,S00512D,S00521D,S00522D | |
| | S00531D,S00532D,S00541D,S00542D,S00551D,S00552D,S00561D | |
| | S00562D,S00571D,S00572D,S0080XD,S0081XD,S0082XD,S0083XD | |
| | S0084XD,S0085XD,S0086XD,S0087XD,S0090XD,S0091XD,S0092XD | |
| | S0093XD,S0094XD,S0095XD,S0096XD,S0097XD,S0500XD,S0501XD | |
| | S0502XD,S0510XD,S0511XD,S0512XD,S100XXD,S1010XD,S1011XD | |
| | S1012XD,S1014XD,S1015XD,S1016XD,S1017XD,S1080XD,S1081XD | |
| | S1082XD,S1083XD,S1084XD,S1085XD,S1086XD,S1087XD,S1090XD | |
| | S1091XD,S1092XD,S1093XD,S1094XD,S1095XD,S1096XD,S1097XD | |
| | S2000XD,S2001XD,S2002XD,S20101D,S20102D,S20109D,S20111D | |
| | S20112D,S20119D,S20121D,S20122D,S20129D,S20141D,S20142D | |
| | S20149D,S20151D,S20152D,S20159D,S20161D,S20162D,S20169D | |
| | S20171D,S20172D,S20179D,S2020XD,S20211D,S20212D,S20213D | |

Continued on next page

**Supplementary Table S1:** (cont'd) A comprehensive mapping of diagnosis groups used in the experiments to their corresponding ICD-10 and ICD-9 codes.

| Diagnosis | ICD-10 | ICD-9 |
|---|---|---|
| | S20214D,S20219D,S20221D,S20222D,S20223D,S20224D,S20229D | |
| | S20301D,S20302D,S20303D,S20304D,S20309D,S20311D,S20312D | |
| | S20313D,S20314D,S20319D,S20321D,S20322D,S20323D,S20324D | |
| | S20329D,S20341D,S20342D,S20343D,S20344D,S20349D,S20351D | |
| | S20352D,S20353D,S20354D,S20359D,S20361D,S20362D,S20363D | |
| | S20364D,S20369D,S20371D,S20372D,S20373D,S20374D,S20379D | |
| | S20401D,S20402D,S20409D,S20411D,S20412D,S20419D,S20421D | |
| | S20422D,S20429D,S20441D,S20442D,S20449D,S20451D,S20452D | |
| | S20459D,S20461D,S20462D,S20469D,S20471D,S20472D,S20479D | |
| | S2090XD,S2091XD,S2092XD,S2094XD,S2095XD,S2096XD,S2097XD | |
| | S300XXD,S301XXD,S30201D,S30202D,S3021XD,S3022XD,S3023XD | |
| | S303XXD,S30810D,S30811D,S30812D,S30813D,S30814D,S30815D | |
| | S30816D,S30817D,S30820D,S30821D,S30822D,S30823D,S30824D | |
| | S30825D,S30826D,S30827D,S30840D,S30841D,S30842D,S30843D | |
| | S30844D,S30845D,S30846D,S30850D,S30851D,S30852D,S30853D | |
| | S30854D,S30855D,S30856D,S30857D,S30860D,S30861D,S30862D | |
| | S30863D,S30864D,S30865D,S30866D,S30867D,S30870D,S30871D | |
| | S30872D,S30873D,S30874D,S30875D,S30876D,S30877D,S3091XD | |
| | S3092XD,S3093XD,S3094XD,S3095XD,S3096XD,S3097XD,S3098XD | |
| | S40011D,S40012D,S40019D,S40021D,S40022D,S40029D,S40211D | |
| | S40212D,S40219D,S40221D,S40222D,S40229D,S40241D,S40242D | |
| | S40249D,S40251D,S40252D,S40259D,S40261D,S40262D,S40269D | |
| | S40271D,S40272D,S40279D,S40811D,S40812D,S40819D,S40821D | |
| | S40822D,S40829D,S40841D,S40842D,S40849D,S40851D,S40852D | |
| | S40859D,S40861D,S40862D,S40869D,S40871D,S40872D,S40879D | |
| | S40911D,S40912D,S40919D,S40921D,S40922D,S40929D,S5000XD | |
| | S5001XD,S5002XD,S5010XD,S5011XD,S5012XD,S50311D,S50312D | |
| | S50319D,S50321D,S50322D,S50329D,S50341D,S50342D,S50349D | |
| | S50351D,S50352D,S50359D,S50361D,S50362D,S50369D,S50371D | |
| | S50372D,S50379D,S50811D,S50812D,S50819D,S50821D,S50822D | |
| | S50829D,S50841D,S50842D,S50849D,S50851D,S50852D,S50859D | |
| | S50861D,S50862D,S50869D,S50871D,S50872D,S50879D,S50901D | |
| | S50902D,S50909D,S50911D,S50912D,S50919D,S6000XD,S60011D | |
| | S60012D,S60019D,S60021D,S60022D,S60029D,S60031D,S60032D | |
| | S60039D,S60041D,S60042D,S60049D,S60051D,S60052D,S60059D | |
| | S6010XD,S60111D,S60112D,S60119D,S60121D,S60122D,S60129D | |
| | S60131D,S60132D,S60139D,S60141D,S60142D,S60149D,S60151D | |
| | S60152D,S60159D,S60211D,S60212D,S60219D,S60221D,S60222D | |
| | S60229D,S60311D,S60312D,S60319D,S60321D,S60322D,S60329D | |
| | S60341D,S60342D,S60349D,S60351D,S60352D,S60359D,S60361D | |

Continued on next page

**Supplementary Table S1:** (cont'd) A comprehensive mapping of diagnosis groups used in the experiments to their corresponding ICD-10 and ICD-9 codes.

| Diagnosis | ICD-10 | ICD-9 |
|---|---|---|
| | S60362D,S60369D,S60371D,S60372D,S60379D,S60391D,S60392D | |
| | S60399D,S60410D,S60411D,S60412D,S60413D,S60414D,S60415D | |
| | S60416D,S60417D,S60418D,S60419D,S60420D,S60421D,S60422D | |
| | S60423D,S60424D,S60425D,S60426D,S60427D,S60428D,S60429D | |
| | S60440D,S60441D,S60442D,S60443D,S60444D,S60445D,S60446D | |
| | S60447D,S60448D,S60449D,S60450D,S60451D,S60452D,S60453D | |
| | S60454D,S60455D,S60456D,S60457D,S60458D,S60459D,S60460D | |
| | S60461D,S60462D,S60463D,S60464D,S60465D,S60466D,S60467D | |
| | S60468D,S60469D,S60470D,S60471D,S60472D,S60473D,S60474D | |
| | S60475D,S60476D,S60477D,S60478D,S60479D,S60511D,S60512D | |
| | S60519D,S60521D,S60522D,S60529D,S60541D,S60542D,S60549D | |
| | S60551D,S60552D,S60559D,S60561D,S60562D,S60569D,S60571D | |
| | S60572D,S60579D,S60811D,S60812D,S60819D,S60821D,S60822D | |
| | S60829D,S60841D,S60842D,S60849D,S60851D,S60852D,S60859D | |
| | S60861D,S60862D,S60869D,S60871D,S60872D,S60879D,S60911D | |
| | S60912D,S60919D,S60921D,S60922D,S60929D,S60931D,S60932D | |
| | S60939D,S60940D,S60941D,S60942D,S60943D,S60944D,S60945D | |
| | S60946D,S60947D,S60948D,S60949D,S7000XD,S7001XD,S7002XD | |
| | S7010XD,S7011XD,S7012XD,S70211D,S70212D,S70219D,S70221D | |
| | S70222D,S70229D,S70241D,S70242D,S70249D,S70251D,S70252D | |
| | S70259D,S70261D,S70262D,S70269D,S70271D,S70272D,S70279D | |
| | S70311D,S70312D,S70319D,S70321D,S70322D,S70329D,S70341D | |
| | S70342D,S70349D,S70351D,S70352D,S70359D,S70361D,S70362D | |
| | S70369D,S70371D,S70372D,S70379D,S70911D,S70912D,S70919D | |
| | S70921D,S70922D,S70929D,S8000XD,S8001XD,S8002XD,S8010XD | |
| | S8011XD,S8012XD,S80211D,S80212D,S80219D,S80221D,S80222D | |
| | S80229D,S80241D,S80242D,S80249D,S80251D,S80252D,S80259D | |
| | S80261D,S80262D,S80269D,S80271D,S80272D,S80279D,S80811D | |
| | S80812D,S80819D,S80821D,S80822D,S80829D,S80841D,S80842D | |
| | S80849D,S80851D,S80852D,S80859D,S80861D,S80862D,S80869D | |
| | S80871D,S80872D,S80879D,S80911D,S80912D,S80919D,S80921D | |
| | S80922D,S80929D,S9000XD,S9001XD,S9002XD,S90111D,S90112D | |
| | S90119D,S90121D,S90122D,S90129D,S90211D,S90212D,S90219D | |
| | S90221D,S90222D,S90229D,S9030XD,S9031XD,S9032XD,S90411D | |
| | S90412D,S90413D,S90414D,S90415D,S90416D,S90421D,S90422D | |
| | S90423D,S90424D,S90425D,S90426D,S90441D,S90442D,S90443D | |
| | S90444D,S90445D,S90446D,S90451D,S90452D,S90453D,S90454D | |
| | S90455D,S90456D,S90461D,S90462D,S90463D,S90464D,S90465D | |
| | S90466D,S90471D,S90472D,S90473D,S90474D,S90475D,S90476D | |
| | S90511D,S90512D,S90519D,S90521D,S90522D,S90529D,S90541D | |

Continued on next page

**Supplementary Table S1:** (cont'd) A comprehensive mapping of diagnosis groups used in the experiments to their corresponding ICD-10 and ICD-9 codes.

| Diagnosis | ICD-10 | ICD-9 |
|---|---|---|
| | S90542D,S90549D,S90551D,S90552D,S90559D,S90561D,S90562D<br>S90569D,S90571D,S90572D,S90579D,S90811D,S90812D,S90819D<br>S90821D,S90822D,S90829D,S90841D,S90842D,S90849D,S90851D<br>S90852D,S90859D,S90861D,S90862D,S90869D,S90871D,S90872D<br>S90879D,S90911D,S90912D,S90919D,S90921D,S90922D,S90929D<br>S90931D,S90932D,S90933D,S90934D,S90935D,S90936D | |
| Symptoms of mental and substance use conditions | R450,R451,R452,R453,R454,R455,R456,R457,R4581,R4582,R4583<br>R4584,R45850,R4586,R4587,R4589,R460,R461,R462,R463,R464,R465<br>R466,R467,R4681,R4689 | 3009,3079,3089,78095,78099,79921,79922,79923,79924,79925<br>79929,79989,V4039,V6285 |
| Syncope | G9001,R55 | 33701,7802 |
| Systemic lupus erythematosus and connective tissue disorders | H01121,H01122,H01123,H01124,H01125,H01126,H01129,L930,L931<br>L932,M300,M301,M302,M303,M308,M310,M311,M3110,M3111,M3119<br>M312,M3130,M3131,M314,M315,M316,M317,M318,M319,M320,M3210<br>M3211,M3212,M3213,M3214,M3215,M3219,M328,M329,M3300,M3301<br>M3302,M3303,M3309,M3310,M3311,M3312,M3313,M3319,M3320,M3321<br>M3322,M3329,M3390,M3391,M3392,M3393,M3399,M340,M341,M342<br>M3481,M3482,M3483,M3489,M349,M3500,M3501,M3502,M3503,M3504<br>M3505,M3506,M3507,M3508,M3509,M350A,M350B,M350C,M351,M352<br>M353,M354,M355,M356,M357,M358,M3581,M3589,M359,M360,M368 | 1361,3574,3596,37334,4200,42491,4460,4461,44620,44621,44629<br>4463,4464,4465,4466,4467,4475,5172,5178,58381,6954,7100,7101<br>7102,7103,7104,7105,7108,7109,71120,725,7285,72889,72930 |
| Tendon and synovial disorders | M1220,M12211,M12212,M12219,M12221,M12222,M12229,M12231,M12232<br>M12239,M12241,M12242,M12249,M12251,M12252,M12259,M12261<br>M12262,M12269,M12271,M12272,M12279,M1228,M1229,M6500,M65011<br>M65012,M65019,M65021,M65022,M65029,M65031,M65032,M65039<br>M65041,M65042,M65049,M65051,M65052,M65059,M65061,M65062<br>M65069,M65071,M65072,M65079,M6508,M6510,M65111,M65112,M65119<br>M65121,M65122,M65129,M65131,M65132,M65139,M65141,M65142<br>M65149,M65151,M65152,M65159,M65161,M65162,M65169,M65171<br>M65172,M65179,M6518,M6519,M6520,M65221,M65222,M65229,M65231<br>M65232,M65239,M65241,M65242,M65249,M65251,M65252,M65259<br>M65261,M65262,M65269,M65271,M65272,M65279,M6528,M6529,M6530<br>M65311,M65312,M65319,M65321,M65322,M65329,M65331,M65332<br>M65339,M65341,M65342,M65349,M65351,M65352,M65359,M654,M6580<br>M65811,M65812,M65819,M65821,M65822,M65829,M65831,M65832<br>M65839,M65841,M65842,M65849,M65851,M65852,M65859,M65861<br>M65862,M65869,M65871,M65872,M65879,M6588,M6589,M659,M660<br>M6610,M66111,M66112,M66119,M66121,M66122,M66129,M66131,M66132 | 71920,71921,71922,71923,71924,71925,71926,71927,71928,71929<br>72610,72611,72612,72613,7262,7265,72664,72669,72670,72671<br>72672,72679,7268,72690,72700,72701,72703,72704,72705,72706<br>72709,7272,72741,72742,72743,72750,72751,72759,72760,72761<br>72762,72763,72764,72765,72766,72767,72768,72769,72781,72782<br>72783,72789,7279 |

Continued on next page

**Supplementary Table S1:** (cont'd) A comprehensive mapping of diagnosis groups used in the experiments to their corresponding ICD-10 and ICD-9 codes.

| Diagnosis | ICD-10 | ICD-9 |
|---|---|---|
| | M66139,M66141,M66142,M66143,M66144,M66145,M66146,M66151<br>M66152,M66159,M66171,M66172,M66173,M66174,M66175,M66176<br>M66177,M66178,M66179,M6618,M6620,M66211,M66212,M66219,M66221<br>M66222,M66229,M66231,M66232,M66239,M66241,M66242,M66249<br>M66251,M66252,M66259,M66261,M66262,M66269,M66271,M66272<br>M66279,M6628,M6629,M6630,M66311,M66312,M66319,M66321,M66322<br>M66329,M66331,M66332,M66339,M66341,M66342,M66349,M66351<br>M66352,M66359,M66361,M66362,M66369,M66371,M66372,M66379,M6638<br>M6639,M6680,M66811,M66812,M66819,M66821,M66822,M66829,M66831<br>M66832,M66839,M66841,M66842,M66849,M66851,M66852,M66859<br>M66861,M66862,M66869,M66871,M66872,M66879,M6688,M6689,M669<br>M6700,M6701,M6702,M6720,M67211,M67212,M67219,M67221,M67222<br>M67229,M67231,M67232,M67239,M67241,M67242,M67249,M67251<br>M67252,M67259,M67261,M67262,M67269,M67271,M67272,M67279,M6728<br>M6729,M6730,M67311,M67312,M67319,M67321,M67322,M67329,M67331<br>M67332,M67339,M67341,M67342,M67349,M67351,M67352,M67359<br>M67361,M67362,M67369,M67371,M67372,M67379,M6738,M6739,M6740<br>M67411,M67412,M67419,M67421,M67422,M67429,M67431,M67432<br>M67439,M67441,M67442,M67449,M67451,M67452,M67459,M67461<br>M67462,M67469,M67471,M67472,M67479,M6748,M6749,M6750,M6751<br>M6752,M6780,M67811,M67812,M67813,M67814,M67819,M67821,M67822<br>M67823,M67824,M67829,M67831,M67832,M67833,M67834,M67839<br>M67841,M67842,M67843,M67844,M67849,M67851,M67852,M67853<br>M67854,M67859,M67861,M67862,M67863,M67864,M67869,M67871<br>M67872,M67873,M67874,M67879,M6788,M6789,M6790,M67911,M67912<br>M67919,M67921,M67922,M67929,M67931,M67932,M67939,M67941<br>M67942,M67949,M67951,M67952,M67959,M67961,M67962,M67969<br>M67971,M67972,M67979,M6798,M6799,M70031,M70032,M70039,M70041<br>M70042,M70049,M7120,M7121,M7122,M75100,M75101,M75102,M75110<br>M75111,M75112,M75120,M75121,M75122,M7520,M7521,M7522,M7530<br>M7531,M7532,M7600,M7601,M7602,M7610,M7611,M7612,M7650,M7651<br>M7652,M7660,M7661,M7662,M7670,M7671,M7672,M76821,M76822<br>M76829,M76891,M76892,M76899,M769,M7750,M7751,M7752,M778,M779 | |
| Thyroid disorders | A1881,B6731,E000,E001,E002,E009,E010,E011,E012,E018,E02,E030<br>E031,E032,E033,E034,E035,E038,E039,E040,E041,E042,E048,E049<br>E0500,E0501,E0510,E0511,E0520,E0521,E0530,E0531,E0540,E0541<br>E0580,E0581,E0590,E0591,E060,E061,E062,E063,E064,E065,E069<br>E070,E071,E0781,E0789,E079,E890,O905 | 01750,1222,2400,2409,2410,2411,2419,24200,24201,24210,24211<br>24220,24221,24230,24231,24240,24241,24280,24281,24290,24291<br>243,2440,2441,2442,2443,2448,2449,2450,2451,2452,2453,2454<br>2458,2459,2460,2461,2462,2463,2468,2469,64812,64814,78001<br>79094 |

Continued on next page

**Supplementary Table S1:** (cont'd) A comprehensive mapping of diagnosis groups used in the experiments to their corresponding ICD-10 and ICD-9 codes.

| Diagnosis | ICD-10 | ICD-9 |
|---|---|---|
| Tobacco-related disorders | F17200,F17203,F17208,F17209,F17210,F17213,F17218,F17219 F17220,F17223,F17228,F17229,F17290,F17293,F17298,F17299 O99330,O99331,O99332,O99333,O99334,O99335 | 2920,29289,2929,3051,64900,64901,64902,64903,64904 |
| Toxic effects, initial encounter | T510X1A,T510X2A,T510X3A,T510X4A,T511X1A,T511X2A,T511X3A T511X4A,T512X1A,T512X2A,T512X3A,T512X4A,T513X1A,T513X2A T513X3A,T513X4A,T518X1A,T518X2A,T518X3A,T518X4A,T5191XA T5192XA,T5193XA,T5194XA,T520X1A,T520X2A,T520X3A,T520X4A T521X1A,T521X2A,T521X3A,T521X4A,T522X1A,T522X2A,T522X3A T522X4A,T523X1A,T523X2A,T523X3A,T523X4A,T524X1A,T524X2A T524X3A,T524X4A,T528X1A,T528X2A,T528X3A,T528X4A,T5291XA T5292XA,T5293XA,T5294XA,T530X1A,T530X2A,T530X3A,T530X4A T531X1A,T531X2A,T531X3A,T531X4A,T532X1A,T532X2A,T532X3A T532X4A,T533X1A,T533X2A,T533X3A,T533X4A,T534X1A,T534X2A T534X3A,T534X4A,T535X1A,T535X2A,T535X3A,T535X4A,T536X1A T536X2A,T536X3A,T536X4A,T537X1A,T537X2A,T537X3A,T537X4A T5391XA,T5392XA,T5393XA,T5394XA,T540X1A,T540X2A,T540X3A T540X4A,T541X1A,T541X2A,T541X3A,T541X4A,T542X1A,T542X2A T542X3A,T542X4A,T543X1A,T543X2A,T543X3A,T543X4A,T5491XA T5492XA,T5493XA,T5494XA,T550X1A,T550X2A,T550X3A,T550X4A T551X1A,T551X2A,T551X3A,T551X4A,T560X1A,T560X2A,T560X3A T560X4A,T561X1A,T561X2A,T561X3A,T561X4A,T562X1A,T562X2A T562X3A,T562X4A,T563X1A,T563X2A,T563X3A,T563X4A,T564X1A T564X2A,T564X3A,T564X4A,T565X1A,T565X2A,T565X3A,T565X4A T566X1A,T566X2A,T566X3A,T566X4A,T567X1A,T567X2A,T567X3A T567X4A,T56811A,T56812A,T56813A,T56814A,T56891A,T56892A T56893A,T56894A,T5691XA,T5692XA,T5693XA,T5694XA,T570X1A T570X2A,T570X3A,T570X4A,T571X1A,T571X2A,T571X3A,T571X4A T572X1A,T572X2A,T572X3A,T572X4A,T573X1A,T573X2A,T573X3A T573X4A,T578X1A,T578X2A,T578X3A,T578X4A,T5791XA,T5792XA T5793XA,T5794XA,T5801XA,T5802XA,T5803XA,T5804XA,T5811XA T5812XA,T5813XA,T5814XA,T582X1A,T582X2A,T582X3A,T582X4A T588X1A,T588X2A,T588X3A,T588X4A,T5891XA,T5892XA,T5893XA T5894XA,T590X1A,T590X2A,T590X3A,T590X4A,T591X1A,T591X2A T591X3A,T591X4A,T592X1A,T592X2A,T592X3A,T592X4A,T593X1A T593X2A,T593X3A,T593X4A,T594X1A,T594X2A,T594X3A,T594X4A T595X1A,T595X2A,T595X3A,T595X4A,T596X1A,T596X2A,T596X3A T596X4A,T597X1A,T597X2A,T597X3A,T597X4A,T59811A,T59812A T59813A,T59814A,T59891A,T59892A,T59893A,T59894A,T5991XA T5992XA,T5993XA,T5994XA,T600X1A,T600X2A,T600X3A,T600X4A | 9800,9801,9802,9803,9808,9809,981,9820,9821,9822,9823,9824 9828,9830,9831,9832,9839,9849,9850,9851,9852,9853,9855,9856 9858,9859,986,9871,9872,9873,9874,9875,9876,9877,9878,9879 9880,9881,9882,9888,9889,9890,9891,9893,9894,9895,9896,9897 98982,98984,98989,9899,E8601,E8602,E8603,E8604,E8608,E8609 E8610,E8611,E8616,E8621,E8624,E8631,E8634,E8635,E8637,E8640 E8641,E8643,E8644,E8651,E8652,E8653,E8654,E8655,E8658,E8659 E8660,E8661,E8663,E8664,E8668,E8681,E8682,E8683,E8688,E8689 E8690,E8691,E8692,E8693,E8698,E9050,E9051,E9052,E9053,E9054 E9055,E9056,E9241,E9286,E9506,E9507,E9508,E9509,E9520,E9521 E9528,E9529,E961,E9621,E9622,E9629,E9806,E9807,E9808,E9809 E9820,E9821,E9828 |

Continued on next page

**Supplementary Table S1:** (cont'd) A comprehensive mapping of diagnosis groups used in the experiments to their corresponding ICD-10 and ICD-9 codes.

| Diagnosis | ICD-10 | ICD-9 |
|---|---|---|
| | T601X1A,T601X2A,T601X3A,T601X4A,T602X1A,T602X2A,T602X3A | |
| | T602X4A,T603X1A,T603X2A,T603X3A,T603X4A,T604X1A,T604X2A | |
| | T604X3A,T604X4A,T608X1A,T608X2A,T608X3A,T608X4A,T6091XA | |
| | T6092XA,T6093XA,T6094XA,T6101XA,T6102XA,T6103XA,T6104XA | |
| | T6111XA,T6112XA,T6113XA,T6114XA,T61771A,T61772A,T61773A | |
| | T61774A,T61781A,T61782A,T61783A,T61784A,T618X1A,T618X2A | |
| | T618X3A,T618X4A,T6191XA,T6192XA,T6193XA,T6194XA,T620X1A | |
| | T620X2A,T620X3A,T620X4A,T621X1A,T621X2A,T621X3A,T621X4A | |
| | T622X1A,T622X2A,T622X3A,T622X4A,T628X1A,T628X2A,T628X3A | |
| | T628X4A,T6291XA,T6292XA,T6293XA,T6294XA,T63001A,T63002A | |
| | T63003A,T63004A,T63011A,T63012A,T63013A,T63014A,T63021A | |
| | T63022A,T63023A,T63024A,T63031A,T63032A,T63033A,T63034A | |
| | T63041A,T63042A,T63043A,T63044A,T63061A,T63062A,T63063A | |
| | T63064A,T63071A,T63072A,T63073A,T63074A,T63081A,T63082A | |
| | T63083A,T63084A,T63091A,T63092A,T63093A,T63094A,T63111A | |
| | T63112A,T63113A,T63114A,T63121A,T63122A,T63123A,T63124A | |
| | T63191A,T63192A,T63193A,T63194A,T632X1A,T632X2A,T632X3A | |
| | T632X4A,T63301A,T63302A,T63303A,T63304A,T63311A,T63312A | |
| | T63313A,T63314A,T63321A,T63322A,T63323A,T63324A,T63331A | |
| | T63332A,T63333A,T63334A,T63391A,T63392A,T63393A,T63394A | |
| | T63411A,T63412A,T63413A,T63414A,T63421A,T63422A,T63423A | |
| | T63424A,T63431A,T63432A,T63433A,T63434A,T63441A,T63442A | |
| | T63443A,T63444A,T63451A,T63452A,T63453A,T63454A,T63461A | |
| | T63462A,T63463A,T63464A,T63481A,T63482A,T63483A,T63484A | |
| | T63511A,T63512A,T63513A,T63514A,T63591A,T63592A,T63593A | |
| | T63594A,T63611A,T63612A,T63613A,T63614A,T63621A,T63622A | |
| | T63623A,T63624A,T63631A,T63632A,T63633A,T63634A,T63691A | |
| | T63692A,T63693A,T63694A,T63711A,T63712A,T63713A,T63714A | |
| | T63791A,T63792A,T63793A,T63794A,T63811A,T63812A,T63813A | |
| | T63814A,T63821A,T63822A,T63823A,T63824A,T63831A,T63832A | |
| | T63833A,T63834A,T63891A,T63892A,T63893A,T63894A,T6391XA | |
| | T6392XA,T6393XA,T6394XA,T6401XA,T6402XA,T6403XA,T6404XA | |
| | T6481XA,T6482XA,T6483XA,T6484XA,T650X1A,T650X2A,T650X3A | |
| | T650X4A,T651X1A,T651X2A,T651X3A,T651X4A,T65211A,T65212A | |
| | T65213A,T65214A,T65221A,T65222A,T65223A,T65224A,T65291A | |
| | T65292A,T65293A,T65294A,T653X1A,T653X2A,T653X3A,T653X4A | |
| | T654X1A,T654X2A,T654X3A,T654X4A,T655X1A,T655X2A,T655X3A | |
| | T655X4A,T656X1A,T656X2A,T656X3A,T656X4A,T65811A,T65812A | |
| | T65813A,T65814A,T65821A,T65822A,T65823A,T65824A,T65831A | |
| | T65832A,T65833A,T65834A,T65891A,T65892A,T65893A,T65894A | |

Continued on next page

**Supplementary Table S1:** (cont'd) A comprehensive mapping of diagnosis groups used in the experiments to their corresponding ICD-10 and ICD-9 codes.

| Diagnosis | ICD-10 | ICD-9 |
|---|---|---|
| | T6591XA,T6592XA,T6593XA,T6594XA | |
| Toxic effects, subsequent encounter | T510X1D,T510X2D,T510X3D,T510X4D,T511X1D,T511X2D,T511X3D<br>T511X4D,T512X1D,T512X2D,T512X3D,T512X4D,T513X1D,T513X2D<br>T513X3D,T513X4D,T518X1D,T518X2D,T518X3D,T518X4D,T5191XD<br>T5192XD,T5193XD,T5194XD,T520X1D,T520X2D,T520X3D,T520X4D<br>T521X1D,T521X2D,T521X3D,T521X4D,T522X1D,T522X2D,T522X3D<br>T522X4D,T523X1D,T523X2D,T523X3D,T523X4D,T524X1D,T524X2D<br>T524X3D,T524X4D,T528X1D,T528X2D,T528X3D,T528X4D,T5291XD<br>T5292XD,T5293XD,T5294XD,T530X1D,T530X2D,T530X3D,T530X4D<br>T531X1D,T531X2D,T531X3D,T531X4D,T532X1D,T532X2D,T532X3D<br>T532X4D,T533X1D,T533X2D,T533X3D,T533X4D,T534X1D,T534X2D<br>T534X3D,T534X4D,T535X1D,T535X2D,T535X3D,T535X4D,T536X1D<br>T536X2D,T536X3D,T536X4D,T537X1D,T537X2D,T537X3D,T537X4D<br>T5391XD,T5392XD,T5393XD,T5394XD,T540X1D,T540X2D,T540X3D<br>T540X4D,T541X1D,T541X2D,T541X3D,T541X4D,T542X1D,T542X2D<br>T542X3D,T542X4D,T543X1D,T543X2D,T543X3D,T543X4D,T5491XD<br>T5492XD,T5493XD,T5494XD,T550X1D,T550X2D,T550X3D,T550X4D<br>T551X1D,T551X2D,T551X3D,T551X4D,T560X1D,T560X2D,T560X3D<br>T560X4D,T561X1D,T561X2D,T561X3D,T561X4D,T562X1D,T562X2D<br>T562X3D,T562X4D,T563X1D,T563X2D,T563X3D,T563X4D,T564X1D<br>T564X2D,T564X3D,T564X4D,T565X1D,T565X2D,T565X3D,T565X4D<br>T566X1D,T566X2D,T566X3D,T566X4D,T567X1D,T567X2D,T567X3D<br>T567X4D,T56811D,T56812D,T56813D,T56814D,T56891D,T56892D<br>T56893D,T56894D,T5691XD,T5692XD,T5693XD,T5694XD,T570X1D<br>T570X2D,T570X3D,T570X4D,T571X1D,T571X2D,T571X3D,T571X4D<br>T572X1D,T572X2D,T572X3D,T572X4D,T573X1D,T573X2D,T573X3D<br>T573X4D,T578X1D,T578X2D,T578X3D,T578X4D,T5791XD,T5792XD<br>T5793XD,T5794XD,T5801XD,T5802XD,T5803XD,T5804XD,T5811XD<br>T5812XD,T5813XD,T5814XD,T582X1D,T582X2D,T582X3D,T582X4D<br>T588X1D,T588X2D,T588X3D,T588X4D,T5891XD,T5892XD,T5893XD<br>T5894XD,T590X1D,T590X2D,T590X3D,T590X4D,T591X1D,T591X2D<br>T591X3D,T591X4D,T592X1D,T592X2D,T592X3D,T592X4D,T593X1D<br>T593X2D,T593X3D,T593X4D,T594X1D,T594X2D,T594X3D,T594X4D<br>T595X1D,T595X2D,T595X3D,T595X4D,T596X1D,T596X2D,T596X3D<br>T596X4D,T597X1D,T597X2D,T597X3D,T597X4D,T59811D,T59812D<br>T59813D,T59814D,T59891D,T59892D,T59893D,T59894D,T5991XD<br>T5992XD,T5993XD,T5994XD,T600X1D,T600X2D,T600X3D,T600X4D<br>T601X1D,T601X2D,T601X3D,T601X4D,T602X1D,T602X2D,T602X3D<br>T602X4D,T603X1D,T603X2D,T603X3D,T603X4D,T604X1D,T604X2D | V5889 |

Continued on next page

**Supplementary Table S1:** (cont'd) A comprehensive mapping of diagnosis groups used in the experiments to their corresponding ICD-10 and ICD-9 codes.

| Diagnosis | ICD-10 | ICD-9 |
|---|---|---|
| | T604X3D,T604X4D,T608X1D,T608X2D,T608X3D,T608X4D,T6091XD T6092XD,T6093XD,T6094XD,T6101XD,T6102XD,T6103XD,T6104XD T6111XD,T6112XD,T6113XD,T6114XD,T61771D,T61772D,T61773D T61774D,T61781D,T61782D,T61783D,T61784D,T618X1D,T618X2D T618X3D,T618X4D,T6191XD,T6192XD,T6193XD,T6194XD,T620X1D T620X2D,T620X3D,T620X4D,T621X1D,T621X2D,T621X3D,T621X4D T622X1D,T622X2D,T622X3D,T622X4D,T628X1D,T628X2D,T628X3D T628X4D,T6291XD,T6292XD,T6293XD,T6294XD,T63001D,T63002D T63003D,T63004D,T63011D,T63012D,T63013D,T63014D,T63021D T63022D,T63023D,T63024D,T63031D,T63032D,T63033D,T63034D T63041D,T63042D,T63043D,T63044D,T63061D,T63062D,T63063D T63064D,T63071D,T63072D,T63073D,T63074D,T63081D,T63082D T63083D,T63084D,T63091D,T63092D,T63093D,T63094D,T63111D T63112D,T63113D,T63114D,T63121D,T63122D,T63123D,T63124D T63191D,T63192D,T63193D,T63194D,T632X1D,T632X2D,T632X3D T632X4D,T63301D,T63302D,T63303D,T63304D,T63311D,T63312D T63313D,T63314D,T63321D,T63322D,T63323D,T63324D,T63331D T63332D,T63333D,T63334D,T63391D,T63392D,T63393D,T63394D T63411D,T63412D,T63413D,T63414D,T63421D,T63422D,T63423D T63424D,T63431D,T63432D,T63433D,T63434D,T63441D,T63442D T63443D,T63444D,T63451D,T63452D,T63453D,T63454D,T63461D T63462D,T63463D,T63464D,T63481D,T63482D,T63483D,T63484D T63511D,T63512D,T63513D,T63514D,T63591D,T63592D,T63593D T63594D,T63611D,T63612D,T63613D,T63614D,T63621D,T63622D T63623D,T63624D,T63631D,T63632D,T63633D,T63634D,T63691D T63692D,T63693D,T63694D,T63711D,T63712D,T63713D,T63714D T63791D,T63792D,T63793D,T63794D,T63811D,T63812D,T63813D T63814D,T63821D,T63822D,T63823D,T63824D,T63831D,T63832D T63833D,T63834D,T63891D,T63892D,T63893D,T63894D,T6391XD T6392XD,T6393XD,T6394XD,T6401XD,T6402XD,T6403XD,T6404XD T6481XD,T6482XD,T6483XD,T6484XD,T650X1D,T650X2D,T650X3D T650X4D,T651X1D,T651X2D,T651X3D,T651X4D,T65211D,T65212D T65213D,T65214D,T65221D,T65222D,T65223D,T65224D,T65291D T65292D,T65293D,T65294D,T653X1D,T653X2D,T653X3D,T653X4D T654X1D,T654X2D,T654X3D,T654X4D,T655X1D,T655X2D,T655X3D T655X4D,T656X1D,T656X2D,T656X3D,T656X4D,T65811D,T65812D T65813D,T65814D,T65821D,T65822D,T65823D,T65824D,T65831D T65832D,T65833D,T65834D,T65891D,T65892D,T65893D,T65894D T6591XD,T6592XD,T6593XD,T6594XD | |

Continued on next page

**Supplementary Table S1:** (cont'd) A comprehensive mapping of diagnosis groups used in the experiments to their corresponding ICD-10 and ICD-9 codes.

| Diagnosis | ICD-10 | ICD-9 |
|---:|---:|---:|
| Transient cerebral ischemia | G450,G451,G452,G453,G454,G458,G459 | 36234,4350,4351,4352,4353,4358,4359,4377 |
| Trauma- and stressor-related disorders | F430,F4310,F4311,F4312,F4320,F4321,F4322,F4323,F4324,F4325<br>F4329,F438,F4381,F4389,F439,F440,F441,F442,F444,F445,F446<br>F447,F4481,F4489,F449,F481,F941,F942 | 2982,30011,30012,30013,30014,30015,30016,30019,3006,3089,3090<br>3091,30924,30928,30929,3093,3094,30981,30989,3099,31389 |
| Traumatic arthropathy | M1250,M12511,M12512,M12519,M12521,M12522,M12529,M12531,M12532<br>M12539,M12541,M12542,M12549,M12551,M12552,M12559,M12561<br>M12562,M12569,M12571,M12572,M12579,M1258,M1259 | 71610,71611,71612,71613,71614,71615,71616,71617,71618,71619 |
| Traumatic brain injury (TBI); concussion, initial encounter | S060X0A,S060X1A,S060X2A,S060X3A,S060X4A,S060X5A,S060X6A<br>S060X7A,S060X8A,S060X9A,S060XAA,S061X0A,S061X1A,S061X2A<br>S061X3A,S061X4A,S061X5A,S061X6A,S061X7A,S061X8A,S061X9A<br>S061XAA,S062X0A,S062X1A,S062X2A,S062X3A,S062X4A,S062X5A<br>S062X6A,S062X7A,S062X8A,S062X9A,S062XAA,S06300A,S06301A<br>S06302A,S06303A,S06304A,S06305A,S06306A,S06307A,S06308A<br>S06309A,S0630AA,S06310A,S06311A,S06312A,S06313A,S06314A<br>S06315A,S06316A,S06317A,S06318A,S06319A,S0631AA,S06320A<br>S06321A,S06322A,S06323A,S06324A,S06325A,S06326A,S06327A<br>S06328A,S06329A,S0632AA,S06330A,S06331A,S06332A,S06333A<br>S06334A,S06335A,S06336A,S06337A,S06338A,S06339A,S0633AA<br>S06340A,S06341A,S06342A,S06343A,S06344A,S06345A,S06346A<br>S06347A,S06348A,S06349A,S0634AA,S06350A,S06351A,S06352A<br>S06353A,S06354A,S06355A,S06356A,S06357A,S06358A,S06359A<br>S0635AA,S06360A,S06361A,S06362A,S06363A,S06364A,S06365A<br>S06366A,S06367A,S06368A,S06369A,S0636AA,S06370A,S06371A<br>S06372A,S06373A,S06374A,S06375A,S06376A,S06377A,S06378A<br>S06379A,S0637AA,S06380A,S06381A,S06382A,S06383A,S06384A<br>S06385A,S06386A,S06387A,S06388A,S06389A,S0638AA,S064X0A<br>S064X1A,S064X2A,S064X3A,S064X4A,S064X5A,S064X6A,S064X7A<br>S064X8A,S064X9A,S064XAA,S065X0A,S065X1A,S065X2A,S065X3A<br>S065X4A,S065X5A,S065X6A,S065X7A,S065X8A,S065X9A,S065XAA<br>S066X0A,S066X1A,S066X2A,S066X3A,S066X4A,S066X5A,S066X6A<br>S066X7A,S066X8A,S066X9A,S066XAA,S06810A,S06811A,S06812A<br>S06813A,S06814A,S06815A,S06816A,S06817A,S06818A,S06819A<br>S0681AA,S06820A,S06821A,S06822A,S06823A,S06824A,S06825A<br>S06826A,S06827A,S06828A,S06829A,S0682AA,S06890A,S06891A<br>S06892A,S06893A,S06894A,S06895A,S06896A,S06897A,S06898A<br>S06899A,S0689AA,S068A0A,S068A1A,S068A2A,S068A3A,S068A4A | 3485,8500,85011,85012,8502,8503,8504,8505,85141,85142,85143<br>85144,85145,85146,85161,85162,85163,85164,85165,85166,85181<br>85182,85183,85184,85185,85186,85201,85202,85203,85204,85205<br>85206,85221,85222,85223,85224,85225,85226,85241,85242,85243<br>85244,85245,85246,85301,85302,85303,85304,85305,85306,85401<br>85402,85403,85404,85405,85406 |

Continued on next page

**Supplementary Table S1:** (cont'd) A comprehensive mapping of diagnosis groups used in the experiments to their corresponding ICD-10 and ICD-9 codes.

| Diagnosis | ICD-10 | ICD-9 |
|---|---|---|
| | S068A5A,S068A6A,S068A7A,S068A8A,S068A9A,S068AAA,S069X0A | |
| | S069X1A,S069X2A,S069X3A,S069X4A,S069X5A,S069X6A,S069X7A | |
| | S069X8A,S069X9A,S069XAA,S06A0XA,S06A1XA | |
| Traumatic brain | S060X0D,S060X1D,S060X2D,S060X3D,S060X4D,S060X5D,S060X6D | V5889 |
| injury (TBI); | S060X7D,S060X8D,S060X9D,S060XAD,S061X0D,S061X1D,S061X2D | |
| concussion, | S061X3D,S061X4D,S061X5D,S061X6D,S061X7D,S061X8D,S061X9D | |
| subsequent encounter | S061XAD,S062X0D,S062X1D,S062X2D,S062X3D,S062X4D,S062X5D | |
| | S062X6D,S062X7D,S062X8D,S062X9D,S062XAD,S06300D,S06301D | |
| | S06302D,S06303D,S06304D,S06305D,S06306D,S06307D,S06308D | |
| | S06309D,S0630AD,S06310D,S06311D,S06312D,S06313D,S06314D | |
| | S06315D,S06316D,S06317D,S06318D,S06319D,S0631AD,S06320D | |
| | S06321D,S06322D,S06323D,S06324D,S06325D,S06326D,S06327D | |
| | S06328D,S06329D,S0632AD,S06330D,S06331D,S06332D,S06333D | |
| | S06334D,S06335D,S06336D,S06337D,S06338D,S06339D,S0633AD | |
| | S06340D,S06341D,S06342D,S06343D,S06344D,S06345D,S06346D | |
| | S06347D,S06348D,S06349D,S0634AD,S06350D,S06351D,S06352D | |
| | S06353D,S06354D,S06355D,S06356D,S06357D,S06358D,S06359D | |
| | S0635AD,S06360D,S06361D,S06362D,S06363D,S06364D,S06365D | |
| | S06366D,S06367D,S06368D,S06369D,S0636AD,S06370D,S06371D | |
| | S06372D,S06373D,S06374D,S06375D,S06376D,S06377D,S06378D | |
| | S06379D,S0637AD,S06380D,S06381D,S06382D,S06383D,S06384D | |
| | S06385D,S06386D,S06387D,S06388D,S06389D,S0638AD,S064X0D | |
| | S064X1D,S064X2D,S064X3D,S064X4D,S064X5D,S064X6D,S064X7D | |
| | S064X8D,S064X9D,S064XAD,S065X0D,S065X1D,S065X2D,S065X3D | |
| | S065X4D,S065X5D,S065X6D,S065X7D,S065X8D,S065X9D,S065XAD | |
| | S066X0D,S066X1D,S066X2D,S066X3D,S066X4D,S066X5D,S066X6D | |
| | S066X7D,S066X8D,S066X9D,S066XAD,S06810D,S06811D,S06812D | |
| | S06813D,S06814D,S06815D,S06816D,S06817D,S06818D,S06819D | |
| | S0681AD,S06820D,S06821D,S06822D,S06823D,S06824D,S06825D | |
| | S06826D,S06827D,S06828D,S06829D,S0682AD,S06890D,S06891D | |
| | S06892D,S06893D,S06894D,S06895D,S06896D,S06897D,S06898D | |
| | S06899D,S0689AD,S068A0D,S068A1D,S068A2D,S068A3D,S068A4D | |
| | S068A5D,S068A6D,S068A9D,S068AAD,S069X0D,S069X1D,S069X2D | |
| | S069X3D,S069X4D,S069X5D,S069X6D,S069X7D,S069X8D,S069X9D | |
| | S069XAD,S06A0XD,S06A1XD | |
| Tuberculosis | A150,A154,A155,A156,A157,A158,A159,A170,A171,A1781,A1782 | 01090,01130,01190,01200,01210,01220,01230,01280,01300,01310 |
| | A1783,A1789,A179,A1801,A1802,A1803,A1809,A1810,A1811,A1812 | 01320,01330,01340,01350,01360,01362,01380,01400,01480,01500 |
| | A1813,A1814,A1815,A1816,A1817,A1818,A182,A1831,A1832,A1839 | 01510,01520,01560,01570,01580,01590,01600,01610,01620,01630 |

Continued on next page

**Supplementary Table S1:** (cont'd) A comprehensive mapping of diagnosis groups used in the experiments to their corresponding ICD-10 and ICD-9 codes.

| Diagnosis | ICD-10 | ICD-9 |
|---|---|---|
| | A184,A1850,A1851,A1852,A1853,A1854,A1859,A186,A187,A1881 A1882,A1883,A1884,A1885,A1889,A190,A191,A192,A198,A199,J65 O98011,O98012,O98013,O98019,O9802,O9803,P370 | 01650,01660,01670,01690,01700,01710,01720,01730,01740,01750 01760,01770,01780,01790,01800,01880,01890,36313,36411,37031 37059,37909,4200,4220,42491,4258,505,6014,64730,64731,64732 64733,64734,71145,71146,71148,72081,73088,73740,7712 |
| Uncomplicated pregnancy, delivery or puerperium | O4202,O4212,O4292,O6020X0,O6020X1,O6020X2,O6020X3,O6020X4 O6020X5,O6020X9,O6022X0,O6022X1,O6022X2,O6022X3,O6022X4 O6022X5,O6022X9,O6023X0,O6023X1,O6023X2,O6023X3,O6023X4 O6023X5,O6023X9,O80,Z3201,Z331,Z333,Z3400,Z3401,Z3402,Z3403 Z3480,Z3481,Z3482,Z3483,Z3490,Z3491,Z3492,Z3493,Z390,Z391 Z392 | 64420,64421,650,65811,65821,V220,V221,V222,V240,V241,V242 V7242 |
| Urinary incontinence | F980,F981,N393,N3941,N3942,N3943,N3944,N3945,N3946,N39490 N39491,N39492,N39498,R32,R3981 | 3076,3077,6256,78830,78831,78832,78833,78834,78835,78836 78837,78838,78839,78891 |
| Urinary system cancers - all other types | C688,C689,D0910,D0919 | 1898,1899,2339 |
| Urinary system cancers - bladder | C670,C671,C672,C673,C674,C675,C676,C677,C678,C679,D090 | 1880,1881,1882,1883,1884,1885,1886,1887,1888,1889,2337 |
| Urinary system cancers - kidney | C641,C642,C649 | 1890 |
| Urinary system cancers - ureter and renal pelvis | C651,C652,C659,C661,C662,C669 | 1891,1892 |
| Urinary system cancers - urethra | C680,C681 | 1893,1894 |
| Urinary tract infections | A0225,A1810,A1813,A3685,A5275,A5401,A5601,A5903,B3741,B3749 N10,N110,N111,N118,N119,N12,N136,N151,N3000,N3001,N3010,N3011 N3020,N3021,N3030,N3031,N3040,N3041,N3080,N3081,N3090,N3091 N340,N341,N342,N343,N390,O2300,O2301,O2302,O2303,O2310,O2311 O2312,O2313,O2320,O2321,O2322,O2323,O2330,O2331,O2332,O2333 O2340,O2341,O2342,O2343,O8620,O8621,O8622,O8629 | 00329,01630,01690,03284,0954,09811,09831,09940,09953,1122 13102,59000,59001,59010,59011,5902,59080,5933,5934,5950,5951 5952,5953,5954,59581,59582,59589,5959,5970,59780,59781,59789 5990,64660,64661,64663,64664 |
| Uveitis and ocular | H15001,H15002,H15003,H15009,H15011,H15012,H15013,H15019 | 36000,36001,36002,36003,36004,36011,36012,36013,36019,36300 |

Continued on next page

**Supplementary Table S1:** (cont'd) A comprehensive mapping of diagnosis groups used in the experiments to their corresponding ICD-10 and ICD-9 codes.

| Diagnosis | ICD-10 | ICD-9 |
|---|---|---|
| inflammation | H15021,H15022,H15023,H15029,H15031,H15032,H15033,H15039<br>H15041,H15042,H15043,H15049,H15051,H15052,H15053,H15059<br>H15091,H15092,H15093,H15099,H15101,H15102,H15103,H15109<br>H15111,H15112,H15113,H15119,H15121,H15122,H15123,H15129,H2000<br>H20011,H20012,H20013,H20019,H20021,H20022,H20023,H20029<br>H20031,H20032,H20033,H20039,H20041,H20042,H20043,H20049<br>H20051,H20052,H20053,H20059,H2010,H2011,H2012,H2013,H2020<br>H2021,H2022,H2023,H20811,H20812,H20813,H20819,H20821,H20822<br>H20823,H20829,H209,H30001,H30002,H30003,H30009,H30011,H30012<br>H30013,H30019,H30021,H30022,H30023,H30029,H30031,H30032<br>H30033,H30039,H30041,H30042,H30043,H30049,H30101,H30102<br>H30103,H30109,H30111,H30112,H30113,H30119,H30121,H30122<br>H30123,H30129,H30131,H30132,H30133,H30139,H30141,H30142<br>H30143,H30149,H3020,H3021,H3022,H3023,H30811,H30812,H30813<br>H30819,H30891,H30892,H30893,H30899,H3090,H3091,H3092,H3093<br>H44001,H44002,H44003,H44009,H44011,H44012,H44013,H44019<br>H44021,H44022,H44023,H44029,H44111,H44112,H44113,H44119<br>H44121,H44122,H44123,H44129,H44131,H44132,H44133,H44139,H4419 | 36301,36303,36304,36305,36306,36307,36308,36310,36311,36312<br>36313,36314,36315,36320,36321,36322,36400,36401,36402,36403<br>36404,36405,36410,36421,36423,36424,3643,37900,37901,37902<br>37903,37904,37905,37906,37907,37909 |
| Varicose veins of lower extremity | I83001,I83002,I83003,I83004,I83005,I83008,I83009,I83011<br>I83012,I83013,I83014,I83015,I83018,I83019,I83021,I83022<br>I83023,I83024,I83025,I83028,I83029,I8310,I8311,I8312,I83201<br>I83202,I83203,I83204,I83205,I83208,I83209,I83211,I83212<br>I83213,I83214,I83215,I83218,I83219,I83221,I83222,I83223<br>I83224,I83225,I83228,I83229,I83811,I83812,I83813,I83819<br>I83891,I83892,I83893,I83899,I8390,I8391,I8392,I8393,O2200<br>O2201,O2202,O2203,O874 | 4540,4541,4542,4548,4549,67100,67101,67102,67103,67104 |
| Vesicoureteral reflux | N1370,N1371,N13721,N13722,N13729,N13731,N13732,N13739 | 59370,59371,59372,59373 |
| Viral infection | A080,A0811,A0819,A082,A0831,A0832,A0839,A084,A088,A6000,A6001<br>A6002,A6003,A6004,A6009,A601,A609,A630,A800,A801,A802,A8030<br>A8039,A804,A809,A8100,A8101,A8109,A811,A812,A8181,A8182,A8183<br>A8189,A819,A820,A821,A829,A830,A831,A832,A833,A834,A835,A836<br>A838,A839,A840,A841,A848,A8481,A8489,A849,A850,A851,A852,A858<br>A86,A870,A871,A872,A878,A879,A880,A881,A888,A89,A90,A91,A920<br>A921,A922,A9230,A9231,A9232,A9239,A924,A925,A928,A929,A930<br>A931,A932,A938,A94,A950,A951,A959,A960,A961,A962,A968,A969<br>A980,A981,A982,A983,A984,A985,A988,A99,B000,B001,B002,B003<br>B004,B0050,B0051,B0052,B0053,B0059,B007,B0081,B0082,B0089 | 00861,00862,00863,00864,00865,00866,00867,00869,0088,04510<br>04520,04590,0460,04611,04619,0462,0463,04671,04672,04679,0468<br>0469,0470,0471,0478,0479,048,0490,0491,0498,0499,0500,0501<br>0509,05101,05102,0511,0512,0519,0520,0521,0522,0527,0528,0529<br>0530,05310,05311,05312,05313,05314,05319,05320,05321,05322<br>05329,05371,05379,0538,0539,0540,05410,05411,05412,05413<br>05419,0542,0543,05440,05441,05442,05443,05444,05449,0545,0546<br>05471,05472,05473,05474,05479,0548,0549,0550,0551,0552,05571<br>05579,0559,05600,05601,05609,05671,05679,0569,0570,0578,0579<br>05810,05811,05812,05821,05829,05881,05882,05889,05900,05901 |

Continued on next page

**Supplementary Table S1:** (cont'd) A comprehensive mapping of diagnosis groups used in the experiments to their corresponding ICD-10 and ICD-9 codes.

| Diagnosis | ICD-10 | ICD-9 |
|---|---|---|
| | B009,B010,B0111,B0112,B012,B0181,B0189,B019,B020,B021,B0221<br>B0222,B0223,B0224,B0229,B0230,B0231,B0232,B0233,B0234,B0239<br>B027,B028,B029,B03,B04,B050,B051,B052,B053,B054,B0581,B0589<br>B059,B0600,B0601,B0602,B0609,B0681,B0682,B0689,B069,B070,B078<br>B079,B08010,B08011,B0802,B0803,B0804,B0809,B081,B0820,B0821<br>B0822,B083,B084,B085,B0860,B0861,B0862,B0869,B0870,B0871<br>B0872,B0879,B088,B09,B1001,B1009,B1081,B1082,B1089,B150,B159<br>B160,B161,B162,B169,B170,B1710,B1711,B172,B178,B179,B180,B181<br>B182,B188,B189,B190,B1910,B1911,B1920,B1921,B199,B250,B251<br>B252,B258,B259,B260,B261,B262,B263,B2681,B2682,B2683,B2684<br>B2685,B2689,B269,B2700,B2701,B2702,B2709,B2710,B2711,B2712<br>B2719,B2780,B2781,B2782,B2789,B2790,B2791,B2792,B2799,B300<br>B301,B302,B303,B308,B309,B330,B331,B3320,B3321,B3322,B3323<br>B3324,B333,B334,B338,B340,B341,B342,B343,B344,B348,B349,B970<br>B9710,B9711,B9712,B9719,B9721,B9729,B9730,B9731,B9732,B9733<br>B9734,B9739,B974,B975,B976,B977,B9781,B9789,J120,J121,J122<br>J123,J1281,J1289,J129,J203,J204,J205,J206,J207,J210,J211<br>O2640,O2641,O2642,O2643,P230,P350,P351,P352,P354,P358,P359 | 05909,05910,05911,05912,05919,05920,05921,05922,0598,0600<br>0601,0609,061,0620,0621,0622,0623,0624,0625,0628,0629,0630<br>0631,0632,0638,0639,064,0650,0651,0652,0653,0654,0659,0660<br>0661,0662,0663,06640,06641,06642,06649,0668,0669,0700,0701<br>07020,07021,07030,07031,07032,07033,07041,07049,07051,07052<br>07053,07054,07059,0706,07070,07071,0709,071,0720,0721,0722<br>0723,07271,07272,07279,0729,0740,0741,07420,07421,07422,07423<br>0743,0748,075,0771,0772,0773,0774,0778,07799,0780,07810,07811<br>07812,07819,0782,0784,0785,0786,0787,07881,07889,0790,0791<br>0792,0793,0794,07950,07951,07952,07959,0796,07981,07982,07983<br>07989,07999,4660,46611,46619,4800,4801,4802,4803,4808,4809<br>4841,4870,5731,5770,64680,64681,64683,7700,7710,7711,7712<br>7908 |
| Counseling related to sexual behavior or orientation | Z700,Z701,Z702,Z703,Z708,Z709 | V6549 |
| Encounter for prophylactic measures (excludes immunization) | Z2911,Z2912,Z2913,Z2914,Z293,Z298,Z299 | |
| Gestational weeks | Z3A00,Z3A01,Z3A08,Z3A09,Z3A10,Z3A11,Z3A12,Z3A13,Z3A14,Z3A15<br>Z3A16,Z3A17,Z3A18,Z3A19,Z3A20,Z3A21,Z3A22,Z3A23,Z3A24,Z3A25<br>Z3A26,Z3A27,Z3A28,Z3A29,Z3A30,Z3A31,Z3A32,Z3A33,Z3A34,Z3A35<br>Z3A36,Z3A37,Z3A38,Z3A39,Z3A40,Z3A41,Z3A42,Z3A49 | NoDx |
| Maternal outcome of delivery | Z370,Z371,Z372,Z373,Z374,Z3750,Z3751,Z3752,Z3753,Z3754,Z3759<br>Z3760,Z3761,Z3762,Z3763,Z3764,Z3769,Z377,Z379 | V270,V271,V272,V273,V274,V275,V276,V277,V279 |
| No immunization or underimmunization | Z2801,Z2802,Z2803,Z2804,Z2809,Z281,Z2820,Z2821,Z2829,Z283<br>Z28310,Z28311,Z2839,Z2881,Z2882,Z2883,Z2889,Z289 | V1583,V6400,V6401,V6402,V6403,V6404,V6405,V6406,V6407,V6408<br>V6409 |
| Organ transplant | Z940,Z941,Z942,Z943,Z944,Z945,Z946,Z947,Z9481,Z9482,Z9483 | V420,V421,V423,V424,V425,V426,V427,V4281,V4282,V4283,V4284 |

Continued on next page

**Supplementary Table S1:** (cont'd) A comprehensive mapping of diagnosis groups used in the experiments to their corresponding ICD-10 and ICD-9 codes.

| Diagnosis | ICD-10 | ICD-9 |
|---|---|---|
| status | Z9484,Z9489,Z949,Z9885 | V4289,V429,V4587 |
| Resistance to | Z1610,Z1611,Z1612,Z1619,Z1620,Z1621,Z1622,Z1623,Z1624,Z1629 | V090,V091,V092,V093,V094,V0950,V0951,V096,V0970,V0971,V0980 |
| antimicrobial drugs | Z1630,Z1631,Z1632,Z1633,Z16341,Z16342,Z1635,Z1639 | V0981,V0990,V0991 |
| Supervision of | O0900,O0901,O0902,O0903,O0910,O0911,O0912,O0913,O09211,O09212 | 65941,65943,65950,65951,65953,65960,65961,65963,V230,V231 |
| high-risk pregnancy | O09213,O09219,O09291,O09292,O09293,O09299,O0930,O0931,O0932 | V233,V2341,V2342,V2349,V235,V237,V2381,V2382,V2383,V2384 |
| | O0933,O0940,O0941,O0942,O0943,O09511,O09512,O09513,O09519 | V2385,V2386,V2389,V239 |
| | O09521,O09522,O09523,O09529,O09611,O09612,O09613,O09619 | |
| | O09621,O09622,O09623,O09629,O0970,O0971,O0972,O0973,O09811 | |
| | O09812,O09813,O09819,O09821,O09822,O09823,O09829,O09891 | |
| | O09892,O09893,O09899,O0990,O0991,O0992,O0993,O09A0,O09A1 | |
| | O09A2,O09A3 | |
| Any dental condition | A690,A691,B002,B084,B370,B3783,K000,K001,K002,K003,K004,K005 | 0542,0743,101,1120,5200,5201,5202,5203,5204,5205,5206,5207 |
| including traumatic | K006,K007,K008,K009,K010,K011,K023,K0251,K0252,K0253,K0261 | 5208,5209,52100,52101,52102,52103,52104,52105,52106,52107 |
| injury | K0262,K0263,K027,K029,K030,K031,K032,K033,K034,K035,K036,K037 | 52108,52110,52120,52130,52140,52141,52142,5215,5216,5217 |
| | K0381,K0389,K039,K040,K0401,K0402,K041,K042,K043,K044,K045 | 52181,52189,5219,5220,5221,5222,5223,5224,5225,5226,5227,5228 |
| | K046,K047,K048,K0490,K0499,K0500,K0501,K0510,K0511,K0520 | 5229,52300,52301,52310,52311,52320,52330,52331,52332,52333 |
| | K0521,K05211,K05212,K05213,K05219,K0522,K05221,K05222,K05223 | 52340,52341,52342,5235,5236,5238,5239,52400,52401,52402,52403 |
| | K05229,K0530,K0531,K05311,K05312,K05313,K05319,K0532,K05321 | 52404,52405,52406,52407,52409,52410,52411,52412,52419,52420 |
| | K05322,K05323,K05329,K054,K055,K056,K060,K06010,K06011,K06012 | 52421,52422,52423,52424,52425,52426,52427,52428,52429,52430 |
| | K06013,K06020,K06021,K06022,K06023,K061,K062,K063,K068,K069 | 52431,52432,52433,52434,52435,52436,52437,52439,5244,52450 |
| | K080,K08101,K08102,K08103,K08104,K08109,K08111,K08112,K08113 | 52451,52452,52453,52454,52455,52456,52457,52459,52460,52461 |
| | K08114,K08119,K08121,K08122,K08123,K08124,K08129,K08131 | 52462,52463,52469,52470,52471,52472,52473,52474,52479,52481 |
| | K08132,K08133,K08134,K08139,K08191,K08192,K08193,K08194 | 52482,52489,5249,5250,52510,52511,52512,52513,52519,52520 |
| | K08199,K0820,K0821,K0822,K0823,K0824,K0825,K0826,K083,K08401 | 52521,52522,52523,52524,52525,52526,5253,52540,52541,52542 |
| | K08402,K08403,K08404,K08409,K08411,K08412,K08413,K08414 | 52543,52544,52550,52551,52552,52553,52554,52560,52561,52562 |
| | K08419,K08421,K08422,K08423,K08424,K08429,K08431,K08432 | 52563,52564,52565,52566,52567,52569,52571,52572,52573,52579 |
| | K08433,K08434,K08439,K08491,K08492,K08493,K08494,K08499,K0850 | 5258,5259,5260,5261,5262,5263,5264,5265,52661,52662,52663 |
| | K0851,K0852,K08530,K08531,K08539,K0854,K0855,K0856,K0859,K088 | 52669,52681,52689,5269,5270,5271,5272,5273,5274,5275,5276 |
| | K0881,K0882,K0889,K089,K090,K091,K098,K099,K110,K111,K1120 | 5277,5278,5279,52800,52801,52802,52809,5281,5282,5283,5284 |
| | K1121,K1122,K1123,K113,K114,K115,K116,K117,K118,K119,K120 | 5285,5286,52871,52872,52879,5288,5289,5290,5291,5292,5293 |
| | K121,K122,K1230,K1231,K1232,K1233,K1239,K130,K131,K1321,K1322 | 5294,5295,5296,5298,5299,6820,73382,78492,80220,80221,80222 |
| | K1323,K1324,K1329,K133,K134,K135,K136,K1370,K1379,K140,K141 | 80223,80224,80225,80226,80227,80228,80230,80231,80232,80233 |
| | K142,K143,K144,K145,K146,K148,K149,M2600,M2601,M2602,M2603 | 80234,80235,80236,80237,80238,8024,8025,8028,8029,8300,8481 |
| | M2604,M2605,M2606,M2607,M2609,M2610,M2611,M2612,M2619,M2620 | 87341,87342,87343,87344,87349,87351,87352,87353,87354,87359 |
| | M26211,M26212,M26213,M26219,M26220,M26221,M2623,M2624,M2625 | 87360,87361,87362,87363,87364,87365,87370,87371,87372,87374 |
| | M2629,M2630,M2631,M2632,M2633,M2634,M2635,M2636,M2637,M2639 | 87375,8738,8739,9050,9056,9057,9060,9089,95909,V523,V534 |
| | M264,M2650,M2651,M2652,M2653,M2654,M2655,M2656,M2657,M2659 | V5419,V585,V5889,V722 |

Continued on next page

**Supplementary Table S1:** (cont'd) A comprehensive mapping of diagnosis groups used in the experiments to their corresponding ICD-10 and ICD-9 codes.

| Diagnosis | ICD-10 | ICD-9 |
|---|---|---|
| | M2660,M26601,M26602,M26603,M26609,M2661,M26611,M26612,M26613 | |
| | M26619,M2662,M26621,M26622,M26623,M26629,M2663,M26631,M26632 | |
| | M26633,M26639,M26641,M26642,M26643,M26649,M26651,M26652 | |
| | M26653,M26659,M2669,M2670,M2671,M2672,M2673,M2674,M2679,M2681 | |
| | M2682,M2689,M269,M270,M271,M272,M273,M2740,M2749,M2751,M2752 | |
| | M2753,M2759,M2761,M2762,M2763,M2769,M278,M279,M350C,M7911 | |
| | R682,R6884,S01401A,S01401D,S01401S,S01402A,S01402D,S01402S | |
| | S01409A,S01409D,S01409S,S01411A,S01411D,S01411S,S01412A | |
| | S01412D,S01412S,S01419A,S01419D,S01419S,S01421A,S01421D | |
| | S01421S,S01422A,S01422D,S01422S,S01429A,S01429D,S01429S | |
| | S01431A,S01431D,S01431S,S01432A,S01432D,S01432S,S01439A | |
| | S01439D,S01439S,S01441A,S01441D,S01441S,S01442A,S01442D | |
| | S01442S,S01449A,S01449D,S01449S,S01451A,S01451D,S01451S | |
| | S01452A,S01452D,S01452S,S01459A,S01459D,S01459S,S01501A | |
| | S01501D,S01501S,S01502A,S01502D,S01502S,S01511A,S01511D | |
| | S01511S,S01512A,S01512D,S01512S,S01521A,S01521D,S01521S | |
| | S01522A,S01522D,S01522S,S01531A,S01531D,S01531S,S01532A | |
| | S01532D,S01532S,S01541A,S01541D,S01541S,S01542A,S01542D | |
| | S01542S,S01551A,S01551D,S01551S,S01552A,S01552D,S01552S | |
| | S0180XA,S0180XD,S0180XS,S0181XA,S0181XD,S0181XS,S0182XA | |
| | S0182XD,S0182XS,S0190XA,S0190XD,S0190XS,S0191XA,S0191XD | |
| | S0191XS,S0192XA,S0192XD,S0192XS,S02401A,S02401B,S02401D | |
| | S02401G,S02401K,S02401S,S0240CA,S0240CB,S0240CD,S0240CG | |
| | S0240CK,S0240CS,S0240DA,S0240DB,S0240DD,S0240DG,S0240DK | |
| | S0240DS,S0242XA,S0242XB,S0242XD,S0242XG,S0242XK,S0242XS | |
| | S025XXA,S025XXB,S025XXD,S025XXG,S025XXK,S025XXS,S02600A | |
| | S02600B,S02600D,S02600G,S02600K,S02600S,S02601A,S02601B | |
| | S02601D,S02601G,S02601K,S02601S,S02602A,S02602B,S02602D | |
| | S02602G,S02602K,S02602S,S02609A,S02609B,S02609D,S02609G | |
| | S02609K,S02609S,S02610A,S02610B,S02610D,S02610G,S02610K | |
| | S02610S,S02611A,S02611B,S02611D,S02611G,S02611K,S02611S | |
| | S02612A,S02612B,S02612D,S02612G,S02612K,S02612S,S0261XA | |
| | S0261XB,S0261XD,S0261XG,S0261XK,S0261XS,S02620A,S02620B | |
| | S02620D,S02620G,S02620K,S02620S,S02621A,S02621B,S02621D | |
| | S02621G,S02621K,S02621S,S02622A,S02622B,S02622D,S02622G | |
| | S02622K,S02622S,S0262XA,S0262XB,S0262XD,S0262XG,S0262XK | |
| | S0262XS,S02630A,S02630B,S02630D,S02630G,S02630K,S02630S | |
| | S02631A,S02631B,S02631D,S02631G,S02631K,S02631S,S02632A | |
| | S02632B,S02632D,S02632G,S02632K,S02632S,S0263XA,S0263XB | |
| | S0263XD,S0263XG,S0263XK,S0263XS,S02640A,S02640B,S02640D | |

Continued on next page

**Supplementary Table S1:** (cont'd) A comprehensive mapping of diagnosis groups used in the experiments to their corresponding ICD-10 and ICD-9 codes.

| Diagnosis | ICD-10 | ICD-9 |
|---|---|---|
| | S02640G,S02640K,S02640S,S02641A,S02641B,S02641D,S02641G<br>S02641K,S02641S,S02642A,S02642B,S02642D,S02642G,S02642K<br>S02642S,S0264XA,S0264XB,S0264XD,S0264XG,S0264XK,S0264XS<br>S02650A,S02650B,S02650D,S02650G,S02650K,S02650S,S02651A<br>S02651B,S02651D,S02651G,S02651K,S02651S,S02652A,S02652B<br>S02652D,S02652G,S02652K,S02652S,S0265XA,S0265XB,S0265XD<br>S0265XG,S0265XK,S0265XS,S0266XA,S0266XB,S0266XD,S0266XG<br>S0266XK,S0266XS,S02670A,S02670B,S02670D,S02670G,S02670K<br>S02670S,S02671A,S02671B,S02671D,S02671G,S02671K,S02671S<br>S02672A,S02672B,S02672D,S02672G,S02672K,S02672S,S0267XA<br>S0267XB,S0267XD,S0267XG,S0267XK,S0267XS,S0269XA,S0269XB<br>S0269XD,S0269XG,S0269XK,S0269XS,S0300XA,S0300XD,S0300XS<br>S0301XA,S0301XD,S0301XS,S0302XA,S0302XD,S0302XS,S0303XA<br>S0303XD,S0303XS,S030XXA,S030XXD,S030XXS,S032XXA,S032XXD<br>S032XXS,S0340XA,S0340XD,S0340XS,S0341XA,S0341XD,S0341XS<br>S0342XA,S0342XD,S0342XS,S0343XA,S0343XD,S0343XS,S034XXA<br>S034XXD,S034XXS,S0993XA,S0993XD,S0993XS,Z0120,Z0121,Z463,Z464 | |
| External cause codes:<br>activity codes | Y9301,Y9302,Y9311,Y9312,Y9313,Y9314,Y9315,Y9316,Y9317,Y9318<br>Y9319,Y9321,Y9322,Y9323,Y9324,Y9329,Y9331,Y9332,Y9333,Y9334<br>Y9335,Y9339,Y9341,Y9342,Y9343,Y9344,Y9345,Y9349,Y9351,Y9352<br>Y9353,Y9354,Y9355,Y9356,Y9357,Y9359,Y9361,Y9362,Y9363,Y9364<br>Y9365,Y9366,Y9367,Y9368,Y9369,Y936A,Y9371,Y9372,Y9373,Y9374<br>Y9375,Y9379,Y9381,Y9382,Y9383,Y9384,Y9385,Y9389,Y939,Y93A1<br>Y93A2,Y93A3,Y93A4,Y93A5,Y93A6,Y93A9,Y93B1,Y93B2,Y93B3,Y93B4<br>Y93B9,Y93C1,Y93C2,Y93C9,Y93D1,Y93D2,Y93D3,Y93D9,Y93E1,Y93E2<br>Y93E3,Y93E4,Y93E5,Y93E6,Y93E8,Y93E9,Y93F1,Y93F2,Y93F9,Y93G1<br>Y93G2,Y93G3,Y93G9,Y93H1,Y93H2,Y93H3,Y93H9,Y93I1,Y93I9,Y93J1<br>Y93J2,Y93J3,Y93J4,Y93K1,Y93K2,Y93K3,Y93K9,Y990,Y991,Y992,Y998<br>Y999 | E0000,E0001,E0002,E0008,E0009,E0010,E0011,E0020,E0021,E0022<br>E0023,E0024,E0025,E0026,E0027,E0028,E0029,E0030,E0031,E0032<br>E0033,E0039,E0040,E0041,E0042,E0043,E0044,E0049,E0050,E0051<br>E0052,E0053,E0054,E0059,E0060,E0061,E0062,E0063,E0064,E0065<br>E0066,E0069,E0070,E0071,E0072,E0073,E0074,E0075,E0076,E0077<br>E0078,E0079,E0080,E0081,E0082,E0083,E0084,E0089,E0090,E0091<br>E0092,E0093,E0094,E0095,E0099,E0100,E0101,E0102,E0103,E0109<br>E0110,E0111,E0119,E0120,E0121,E0122,E0129,E0130,E0131,E0132<br>E0133,E0134,E0135,E0138,E0139,E0140,E0141,E0149,E0150,E0151<br>E0152,E0159,E0160,E0161,E0162,E0169,E0170,E0179,E0180,E0181<br>E0182,E0183,E0190,E0191,E0192,E0199,E0290,E0291,E0292,E0299<br>E030 |
| External cause codes:<br>bites; initial<br>encounter | T63001A,T63002A,T63003A,T63004A,T63011A,T63012A,T63013A<br>T63014A,T63021A,T63022A,T63023A,T63024A,T63031A,T63032A<br>T63033A,T63034A,T63041A,T63042A,T63043A,T63044A,T63061A<br>T63062A,T63063A,T63064A,T63071A,T63072A,T63073A,T63074A<br>T63081A,T63082A,T63083A,T63084A,T63091A,T63092A,T63093A<br>T63094A,T63111A,T63112A,T63113A,T63114A,T63121A,T63122A<br>T63123A,T63124A,T63191A,T63192A,T63193A,T63194A,T632X1A<br>T632X2A,T632X3A,T632X4A,T63301A,T63302A,T63303A,T63304A<br>T63311A,T63312A,T63313A,T63314A,T63321A,T63322A,T63323A | 9895,E9050,E9051,E9052,E9053,E9054,E9055,E9056,E9060,E9061<br>E9062,E9063,E9064,E9509,E9621,E9809 |

Continued on next page

**Supplementary Table S1:** (cont'd) A comprehensive mapping of diagnosis groups used in the experiments to their corresponding ICD-10 and ICD-9 codes.

| Diagnosis | ICD-10 | ICD-9 |
|---|---|---|
| | T63324A,T63331A,T63332A,T63333A,T63334A,T63391A,T63392A<br>T63393A,T63394A,T63411A,T63412A,T63413A,T63414A,T63421A<br>T63422A,T63423A,T63424A,T63431A,T63432A,T63433A,T63434A<br>T63441A,T63442A,T63443A,T63444A,T63451A,T63452A,T63453A<br>T63454A,T63461A,T63462A,T63463A,T63464A,T63481A,T63482A<br>T63483A,T63484A,T63511A,T63512A,T63513A,T63514A,T63591A<br>T63592A,T63593A,T63594A,T63611A,T63612A,T63613A,T63614A<br>T63621A,T63622A,T63623A,T63624A,T63631A,T63632A,T63633A<br>T63634A,T63691A,T63692A,T63693A,T63694A,T63811A,T63812A<br>T63813A,T63814A,T63821A,T63822A,T63823A,T63824A,T63831A<br>T63832A,T63833A,T63834A,T63891A,T63892A,T63893A,T63894A<br>T6391XA,T6392XA,T6393XA,T6394XA,W5301XA,W5311XA,W5321XA<br>W5381XA,W540XXA,W5501XA,W5511XA,W5521XA,W5531XA,W5541XA<br>W5551XA,W5581XA,W5601XA,W5611XA,W5621XA,W5631XA,W5641XA<br>W5651XA,W5681XA,W57XXXA,W5801XA,W5811XA,W5901XA,W5911XA<br>W5921XA,W5981XA,W6101XA,W6111XA,W6121XA,W6151XA,W6161XA<br>W6191XA | |
| External cause codes: complications of medical and surgical care, initial encounter | Y620,Y621,Y622,Y623,Y624,Y625,Y626,Y628,Y629,Y630,Y631,Y632<br>Y633,Y634,Y635,Y636,Y638,Y639,Y640,Y641,Y648,Y649,Y650,Y651<br>Y652,Y653,Y654,Y6551,Y6552,Y6553,Y658,Y66,Y69,Y700,Y701,Y702<br>Y703,Y708,Y710,Y711,Y712,Y713,Y718,Y720,Y721,Y722,Y723,Y728<br>Y730,Y731,Y732,Y733,Y738,Y740,Y741,Y742,Y743,Y748,Y750,Y751<br>Y752,Y753,Y758,Y760,Y761,Y762,Y763,Y768,Y770,Y771,Y7711,Y7719<br>Y772,Y773,Y778,Y780,Y781,Y782,Y783,Y788,Y790,Y791,Y792,Y793<br>Y798,Y800,Y801,Y802,Y803,Y808,Y810,Y811,Y812,Y813,Y818,Y828<br>Y829,Y830,Y831,Y832,Y833,Y834,Y835,Y836,Y838,Y839,Y840,Y841<br>Y842,Y843,Y844,Y845,Y846,Y847,Y848,Y849,Y95 | E8720,E8721,E8722,E8723,E8724,E8725,E8726,E8728,E8729,E8730<br>E8731,E8732,E8733,E8734,E8735,E8736,E8738,E8739,E8750,E8751<br>E8752,E8758,E8759,E8760,E8761,E8762,E8763,E8764,E8765,E8766<br>E8767,E8768,E8769,E8780,E8781,E8782,E8783,E8784,E8785,E8786<br>E8788,E8789,E8790,E8791,E8792,E8793,E8794,E8795,E8796,E8797<br>E8798,NoDx |
| External cause codes: cut pierce; initial encounter | W25XXXA,W260XXA,W261XXA,W262XXA,W268XXA,W269XXA,W270XXA<br>W271XXA,W272XXA,W273XXA,W274XXA,W275XXA,W278XXA,W28XXXA<br>W290XXA,W291XXA,W292XXA,W293XXA,W294XXA,W298XXA,W450XXA<br>W451XXA,W452XXA,W458XXA,W460XXA,W461XXA,X780XXA,X781XXA<br>X782XXA,X788XXA,X789XXA,X990XXA,X991XXA,X992XXA,X998XXA<br>X999XXA,Y280XXA,Y281XXA,Y282XXA,Y288XXA,Y289XXA,Y35401A<br>Y35402A,Y35403A,Y35409A,Y35411A,Y35412A,Y35413A,Y35419A<br>Y35491A,Y35492A,Y35493A,Y35499A | E9200,E9201,E9202,E9203,E9204,E9205,E9208,E956,E966,E974,E986 |
| External cause codes: drowning submersion; | T751XXA,V9000XA,V9001XA,V9002XA,V9003XA,V9004XA,V9005XA<br>V9006XA,V9008XA,V9009XA,V9010XA,V9011XA,V9012XA,V9013XA | 9941,E8309,E8329,E8830,E9102,E9104,E9108,E9109,E9174,E954<br>E964,E984 |

Continued on next page

**Supplementary Table S1:** (cont'd) A comprehensive mapping of diagnosis groups used in the experiments to their corresponding ICD-10 and ICD-9 codes.

| Diagnosis | ICD-10 | ICD-9 |
|---|---|---|
| initial encounter | V9014XA,V9015XA,V9016XA,V9018XA,V9019XA,V9020XA,V9021XA | |
| | V9022XA,V9023XA,V9024XA,V9025XA,V9026XA,V9027XA,V9028XA | |
| | V9029XA,V9030XA,V9031XA,V9032XA,V9033XA,V9034XA,V9035XA | |
| | V9036XA,V9037XA,V9038XA,V9039XA,V9080XA,V9081XA,V9082XA | |
| | V9083XA,V9084XA,V9085XA,V9086XA,V9087XA,V9088XA,V9089XA | |
| | V9200XA,V9201XA,V9202XA,V9203XA,V9204XA,V9205XA,V9206XA | |
| | V9207XA,V9208XA,V9209XA,V9210XA,V9211XA,V9212XA,V9213XA | |
| | V9214XA,V9215XA,V9216XA,V9219XA,V9220XA,V9221XA,V9222XA | |
| | V9223XA,V9224XA,V9225XA,V9226XA,V9227XA,V9228XA,V9229XA | |
| | W16011A,W16021A,W16031A,W16111A,W16121A,W16131A,W16211A | |
| | W16221A,W16311A,W16321A,W16331A,W1641XA,W16511A,W16521A | |
| | W16531A,W16611A,W16621A,W16711A,W16721A,W16811A,W16821A | |
| | W16831A,W1691XA,W22041A,W65XXXA,W67XXXA,W69XXXA,W73XXXA | |
| | W74XXXA,X710XXA,X711XXA,X712XXA,X713XXA,X718XXA,X719XXA | |
| | X920XXA,X921XXA,X922XXA,X923XXA,X928XXA,X929XXA,Y210XXA | |
| | Y211XXA,Y212XXA,Y213XXA,Y214XXA,Y218XXA,Y219XXA | |
| External cause codes: evidence of alcohol involvement | Y900,Y901,Y902,Y903,Y904,Y905,Y906,Y907,Y908,Y909 | NoDx |
| External cause codes: fall; initial encounter | V00111A,V00121A,V00131A,V00141A,V00151A,V00181A,V00211A | E8800,E8801,E8809,E8810,E8811,E882,E8830,E8831,E8832,E8839 |
| | V00221A,V00281A,V00311A,V00321A,V00381A,V00811A,V00821A | E8840,E8841,E8842,E8843,E8844,E8845,E8846,E8849,E8850,E8851 |
| | V00831A,V00841A,V00891A,W000XXA,W001XXA,W002XXA,W009XXA | E8852,E8853,E8854,E8859,E8860,E8880,E8881,E8888,E8889,E9579 |
| | W010XXA,W0110XA,W01110A,W01111A,W01118A,W01119A,W01190A | E9681,E9879 |
| | W01198A,W03XXXA,W04XXXA,W050XXA,W051XXA,W052XXA,W06XXXA | |
| | W07XXXA,W08XXXA,W090XXA,W091XXA,W092XXA,W098XXA,W100XXA | |
| | W101XXA,W102XXA,W108XXA,W109XXA,W11XXXA,W12XXXA,W130XXA | |
| | W131XXA,W132XXA,W133XXA,W134XXA,W138XXA,W139XXA,W14XXXA | |
| | W15XXXA,W16012A,W16022A,W16032A,W16112A,W16122A,W16132A | |
| | W16212A,W16222A,W16312A,W16322A,W16332A,W1642XA,W16512A | |
| | W16522A,W16532A,W16612A,W16622A,W16712A,W16722A,W16812A | |
| | W16822A,W16832A,W1692XA,W170XXA,W171XXA,W172XXA,W173XXA | |
| | W174XXA,W1781XA,W1782XA,W1789XA,W1811XA,W1812XA,W182XXA | |
| | W1830XA,W1831XA,W1839XA,W19XXXA,X80XXXA,Y01XXXA,Y30XXXA | |
| External cause codes: fire burn; initial encounter | T541X1A,T541X2A,T541X3A,T541X4A,T542X1A,T542X2A,T542X3A | 9831,9832,9839,E8902,E8903,E8908,E892,E8930,E8938,E894,E896 |
| | T542X4A,T543X1A,T543X2A,T543X3A,T543X4A,T5491XA,T5492XA | E897,E8980,E8981,E9240,E9241,E9242,E9248,E9507,E9581,E9582 |
| | T5493XA,T5494XA,X000XXA,X001XXA,X002XXA,X003XXA,X004XXA | E961,E9680,E9683,E9688,E9793,E9806,E9881,E9882,E9901,E9903 |
| | X005XXA,X008XXA,X010XXA,X011XXA,X013XXA,X014XXA,X018XXA | E9909 |

Continued on next page

**Supplementary Table S1:** (cont'd) A comprehensive mapping of diagnosis groups used in the experiments to their corresponding ICD-10 and ICD-9 codes.

| Diagnosis | ICD-10 | ICD-9 |
|---|---|---|
| | X020XXA,X021XXA,X022XXA,X023XXA,X024XXA,X025XXA,X028XXA X030XXA,X031XXA,X033XXA,X034XXA,X038XXA,X04XXXA,X05XXXA X060XXA,X061XXA,X062XXA,X063XXA,X0800XA,X0801XA,X0809XA X0810XA,X0811XA,X0819XA,X0820XA,X0821XA,X0829XA,X088XXA X100XXA,X101XXA,X102XXA,X110XXA,X111XXA,X118XXA,X12XXXA X130XXA,X131XXA,X140XXA,X141XXA,X150XXA,X151XXA,X152XXA X153XXA,X158XXA,X16XXXA,X17XXXA,X18XXXA,X19XXXA,X76XXXA X770XXA,X771XXA,X772XXA,X773XXA,X778XXA,X779XXA,X97XXXA X980XXA,X981XXA,X982XXA,X983XXA,X988XXA,X989XXA,Y26XXXA Y270XXA,Y271XXA,Y272XXA,Y273XXA,Y278XXA,Y279XXA,Y36300A Y36301A,Y36330A,Y36331A,Y36390A,Y36391A,Y37300A,Y37301A Y37330A,Y37331A,Y37390A,Y37391A,Y383X1A,Y383X2A,Y383X3A | |
| External cause codes: firearm; initial encounter | W320XXA,W321XXA,W3300XA,W3301XA,W3302XA,W3303XA,W3309XA W3310XA,W3311XA,W3312XA,W3313XA,W3319XA,W3400XA,W3409XA W3410XA,W3419XA,X72XXXA,X730XXA,X731XXA,X732XXA,X738XXA X739XXA,X748XXA,X749XXA,X93XXXA,X940XXA,X941XXA,X942XXA X948XXA,X949XXA,X958XXA,X959XXA,Y22XXXA,Y230XXA,Y231XXA Y232XXA,Y233XXA,Y238XXA,Y239XXA,Y248XXA,Y249XXA,Y35001A Y35002A,Y35003A,Y35009A,Y35011A,Y35012A,Y35013A,Y35019A Y35021A,Y35022A,Y35023A,Y35029A,Y35031A,Y35032A,Y35033A Y35039A,Y35091A,Y35092A,Y35093A,Y35099A,Y36420A,Y36421A Y36430A,Y36431A,Y3692XA,Y37420A,Y37421A,Y37430A,Y37431A Y3792XA,Y384X1A,Y384X2A,Y384X3A | E9220,E9221,E9222,E9223,E9228,E9229,E9550,E9551,E9552,E9553 E9554,E9559,E9650,E9651,E9652,E9653,E9654,E970,E9794,E9850 E9851,E9852,E9853,E9854,E9857,E9909,E9911,E9912 |
| External cause codes: machinery; initial encounter | W240XXA,W241XXA,W300XXA,W301XXA,W302XXA,W303XXA,W3081XA W3089XA,W309XXA,W310XXA,W311XXA,W312XXA,W313XXA,W3181XA W3182XA,W3183XA,W3189XA,W319XXA | E9190,E9191,E9192,E9193,E9194,E9195,E9196,E9197,E9198,E9199 |
| External cause codes: motor vehicle traffic (MVT); initial encounter | V0210XA,V0211XA,V0212XA,V02131A,V02138A,V0219XA,V0290XA V0291XA,V0292XA,V02931A,V02938A,V0299XA,V0310XA,V0311XA V0312XA,V03131A,V03138A,V0319XA,V0390XA,V0391XA,V0392XA V03931A,V03938A,V0399XA,V0410XA,V0411XA,V0412XA,V04131A V04138A,V0419XA,V0490XA,V0491XA,V0492XA,V04931A,V04938A V0499XA,V0920XA,V0921XA,V0929XA,V093XXA,V123XXA,V124XXA V125XXA,V129XXA,V133XXA,V134XXA,V135XXA,V139XXA,V143XXA V144XXA,V145XXA,V149XXA,V1940XA,V1949XA,V1950XA,V1959XA V1960XA,V1969XA,V199XXA,V2039XA,V203XXA,V2049XA,V204XXA V2059XA,V205XXA,V2099XA,V209XXA,V2139XA,V213XXA,V2149XA V214XXA,V2159XA,V215XXA,V2199XA,V219XXA,V2231XA,V2239XA | E8100,E8101,E8102,E8103,E8108,E8109,E8120,E8121,E8122,E8123 E8128,E8129,E8130,E8131,E8132,E8133,E8134,E8135,E8136,E8138 E8139,E8140,E8141,E8142,E8143,E8147,E8148,E8149,E8150,E8151 E8152,E8153,E8158,E8159,E8160,E8161,E8162,E8163,E8168,E8169 E8178,E8188,E8189,E8190,E8192,E8199,E8222,E8223,E8239,E8242 E8246,E8248,E8261,E8290,E846,E9580,E9585,E9685,E9885 |

Continued on next page

**Supplementary Table S1:** (cont'd) A comprehensive mapping of diagnosis groups used in the experiments to their corresponding ICD-10 and ICD-9 codes.

| Diagnosis | ICD-10 | ICD-9 |
|---|---|---|
| | V223XXA,V2241XA,V2249XA,V224XXA,V2251XA,V2259XA,V225XXA | |
| | V2291XA,V2299XA,V229XXA,V2331XA,V2339XA,V233XXA,V2341XA | |
| | V2349XA,V234XXA,V2351XA,V2359XA,V235XXA,V2391XA,V2399XA | |
| | V239XXA,V2431XA,V2439XA,V243XXA,V2441XA,V2449XA,V244XXA | |
| | V2451XA,V2459XA,V245XXA,V2491XA,V2499XA,V249XXA,V2539XA | |
| | V253XXA,V2549XA,V254XXA,V2559XA,V255XXA,V2599XA,V259XXA | |
| | V2639XA,V263XXA,V2649XA,V264XXA,V2659XA,V265XXA,V2699XA | |
| | V269XXA,V2739XA,V273XXA,V2749XA,V274XXA,V2759XA,V275XXA | |
| | V2799XA,V279XXA,V2839XA,V283XXA,V2849XA,V284XXA,V2859XA | |
| | V285XXA,V2899XA,V289XXA,V29401A,V29408A,V2940XA,V29491A | |
| | V29498A,V2949XA,V29501A,V29508A,V2950XA,V29591A,V29598A | |
| | V2959XA,V29601A,V29608A,V2960XA,V29691A,V29698A,V2969XA | |
| | V29818A,V2981XA,V29888A,V2988XA,V2991XA,V2999XA,V299XXA | |
| | V304XXA,V305XXA,V306XXA,V307XXA,V309XXA,V314XXA,V315XXA | |
| | V316XXA,V317XXA,V319XXA,V324XXA,V325XXA,V326XXA,V327XXA | |
| | V329XXA,V334XXA,V335XXA,V336XXA,V337XXA,V339XXA,V344XXA | |
| | V345XXA,V346XXA,V347XXA,V349XXA,V354XXA,V355XXA,V356XXA | |
| | V357XXA,V359XXA,V364XXA,V365XXA,V366XXA,V367XXA,V369XXA | |
| | V374XXA,V375XXA,V376XXA,V377XXA,V379XXA,V384XXA,V385XXA | |
| | V386XXA,V387XXA,V389XXA,V3940XA,V3949XA,V3950XA,V3959XA | |
| | V3960XA,V3969XA,V3981XA,V3989XA,V399XXA,V404XXA,V405XXA | |
| | V406XXA,V407XXA,V409XXA,V414XXA,V415XXA,V416XXA,V417XXA | |
| | V419XXA,V424XXA,V425XXA,V426XXA,V427XXA,V429XXA,V4341XA | |
| | V4342XA,V4343XA,V4344XA,V4351XA,V4352XA,V4353XA,V4354XA | |
| | V4361XA,V4362XA,V4363XA,V4364XA,V4371XA,V4372XA,V4373XA | |
| | V4374XA,V4391XA,V4392XA,V4393XA,V4394XA,V444XXA,V445XXA | |
| | V446XXA,V447XXA,V449XXA,V454XXA,V455XXA,V456XXA,V457XXA | |
| | V459XXA,V464XXA,V465XXA,V466XXA,V467XXA,V469XXA,V474XXA | |
| | V4751XA,V4752XA,V475XXA,V4761XA,V4762XA,V476XXA,V477XXA | |
| | V4791XA,V4792XA,V479XXA,V484XXA,V485XXA,V486XXA,V487XXA | |
| | V489XXA,V4940XA,V4949XA,V4950XA,V4959XA,V4960XA,V4969XA | |
| | V4981XA,V4988XA,V499XXA,V504XXA,V505XXA,V506XXA,V507XXA | |
| | V509XXA,V514XXA,V515XXA,V516XXA,V517XXA,V519XXA,V524XXA | |
| | V525XXA,V526XXA,V527XXA,V529XXA,V534XXA,V535XXA,V536XXA | |
| | V537XXA,V539XXA,V544XXA,V545XXA,V546XXA,V547XXA,V549XXA | |
| | V554XXA,V555XXA,V556XXA,V557XXA,V559XXA,V564XXA,V565XXA | |
| | V566XXA,V567XXA,V569XXA,V574XXA,V575XXA,V576XXA,V577XXA | |
| | V579XXA,V584XXA,V585XXA,V586XXA,V587XXA,V589XXA,V5940XA | |
| | V5949XA,V5950XA,V5959XA,V5960XA,V5969XA,V5981XA,V5988XA | |
| | V599XXA,V604XXA,V605XXA,V606XXA,V607XXA,V609XXA,V614XXA | |

Continued on next page

**Supplementary Table S1:** (cont'd) A comprehensive mapping of diagnosis groups used in the experiments to their corresponding ICD-10 and ICD-9 codes.

| Diagnosis | ICD-10 | ICD-9 |
|---|---|---|
| | V615XXA,V616XXA,V617XXA,V619XXA,V624XXA,V625XXA,V626XXA<br>V627XXA,V629XXA,V634XXA,V635XXA,V636XXA,V637XXA,V639XXA<br>V644XXA,V645XXA,V646XXA,V647XXA,V649XXA,V654XXA,V655XXA<br>V656XXA,V657XXA,V659XXA,V664XXA,V665XXA,V666XXA,V667XXA<br>V669XXA,V674XXA,V675XXA,V676XXA,V677XXA,V679XXA,V684XXA<br>V685XXA,V686XXA,V687XXA,V689XXA,V6940XA,V6949XA,V6950XA<br>V6959XA,V6960XA,V6969XA,V6981XA,V6988XA,V699XXA,V704XXA<br>V705XXA,V706XXA,V707XXA,V709XXA,V714XXA,V715XXA,V716XXA<br>V717XXA,V719XXA,V724XXA,V725XXA,V726XXA,V727XXA,V729XXA<br>V734XXA,V735XXA,V736XXA,V737XXA,V739XXA,V744XXA,V745XXA<br>V746XXA,V747XXA,V749XXA,V754XXA,V755XXA,V756XXA,V757XXA<br>V759XXA,V764XXA,V765XXA,V766XXA,V767XXA,V769XXA,V774XXA<br>V775XXA,V776XXA,V777XXA,V779XXA,V784XXA,V785XXA,V786XXA<br>V787XXA,V789XXA,V7940XA,V7949XA,V7950XA,V7959XA,V7960XA<br>V7969XA,V7981XA,V7988XA,V799XXA,V8031XA,V8032XA,V8041XA<br>V8042XA,V8051XA,V8052XA,V811XXA,V821XXA,V830XXA,V831XXA<br>V832XXA,V833XXA,V840XXA,V841XXA,V842XXA,V843XXA,V850XXA<br>V851XXA,V852XXA,V853XXA,V8601XA,V8602XA,V8603XA,V8604XA<br>V8605XA,V8606XA,V8609XA,V8611XA,V8612XA,V8613XA,V8614XA<br>V8615XA,V8616XA,V8619XA,V8621XA,V8622XA,V8623XA,V8624XA<br>V8625XA,V8626XA,V8629XA,V8631XA,V8632XA,V8633XA,V8634XA<br>V8635XA,V8636XA,V8639XA,V870XXA,V871XXA,V872XXA,V873XXA<br>V874XXA,V875XXA,V876XXA,V877XXA,V878XXA,V892XXA,X810XXA<br>X820XXA,X821XXA,X822XXA,X828XXA,Y020XXA,Y030XXA,Y038XXA<br>Y32XXXA | |
| External cause codes:<br>natural environment;<br>initial encounter | T63711A,T63712A,T63713A,T63714A,T63791A,T63792A,T63793A<br>T63794A,T65821A,T65822A,T65823A,T65824A,T730XXA,T731XXA<br>T732XXA,T738XXA,T739XXA,T7500XA,T7501XA,T7509XA,T7520XA<br>T7521XA,T7522XA,T7523XA,T7529XA,T753XXA,W420XXA,W429XXA<br>W5309XA,W5319XA,W5329XA,W5389XA,W541XXA,W548XXA,W5503XA<br>W5509XA,W5512XA,W5519XA,W5522XA,W5529XA,W5532XA,W5539XA<br>W5542XA,W5549XA,W5552XA,W5559XA,W5582XA,W5589XA,W5602XA<br>W5609XA,W5612XA,W5619XA,W5622XA,W5629XA,W5632XA,W5639XA<br>W5642XA,W5649XA,W5652XA,W5659XA,W5682XA,W5689XA,W5802XA<br>W5803XA,W5809XA,W5812XA,W5813XA,W5819XA,W5902XA,W5909XA<br>W5912XA,W5913XA,W5919XA,W5922XA,W5929XA,W5982XA,W5983XA<br>W5989XA,W60XXXA,W6102XA,W6109XA,W6112XA,W6119XA,W6122XA<br>W6129XA,W6132XA,W6133XA,W6139XA,W6142XA,W6143XA,W6149XA<br>W6152XA,W6159XA,W6162XA,W6169XA,W6192XA,W6199XA,W620XXA | 9895,98989,9940,9942,9943,9944,9946,9949,E9000,E9001,E9010<br>E9011,E9018,E9020,E9021,E9022,E9028,E903,E9041,E9042,E9043<br>E9049,E9056,E9061,E9063,E9068,E9069,E9080,E9081,E9082,E9083<br>E9084,E9088,E9089,E9090,E9091,E9092,E9093,E9094,E9098,E9208<br>E9268,E9280,E9281,E9286,E9288,E9509,E9583,E9621,E9809 |

Continued on next page

**Supplementary Table S1:** (cont'd) A comprehensive mapping of diagnosis groups used in the experiments to their corresponding ICD-10 and ICD-9 codes.

| Diagnosis | ICD-10 | ICD-9 |
|---|---|---|
| | W621XXA,W629XXA,W64XXXA,W92XXXA,W9301XA,W9302XA,W9311XA<br>W9312XA,W932XXA,W938XXA,W940XXA,W9411XA,W9412XA,W9421XA<br>W9422XA,W9423XA,W9429XA,W9431XA,W9432XA,W9439XA,W99XXXA<br>X30XXXA,X31XXXA,X32XXXA,X34XXXA,X35XXXA,X360XXA,X361XXA<br>X370XXA,X371XXA,X372XXA,X373XXA,X3741XA,X3742XA,X3743XA<br>X378XXA,X379XXA,X38XXXA,X3901XA,X3908XA,X398XXA,X52XXXA<br>X832XXA | |
| External cause codes:<br>other specified,<br>classifiable and NEC;<br>initial encounter | T1500XA,T1501XA,T1502XA,T1510XA,T1511XA,T1512XA,T1580XA<br>T1581XA,T1582XA,T1590XA,T1591XA,T1592XA,T161XXA,T162XXA<br>T169XXA,T170XXA,T171XXA,T17208A,T17218A,T17228A,T17298A<br>T17308A,T17318A,T17328A,T17398A,T17408A,T17418A,T17428A<br>T17498A,T17508A,T17518A,T17528A,T17598A,T17808A,T17818A<br>T17828A,T17898A,T17908A,T17918A,T17928A,T17998A,T180XXA<br>T18100A,T18108A,T18110A,T18118A,T18120A,T18128A,T18190A<br>T18198A,T182XXA,T183XXA,T184XXA,T185XXA,T188XXA,T189XXA<br>T190XXA,T191XXA,T192XXA,T193XXA,T194XXA,T198XXA,T199XXA<br>T7401XA,T7402XA,T7411XA,T7412XA,T7421XA,T7422XA,T7431XA<br>T7432XA,T744XXA,T7451XA,T7452XA,T7461XA,T7462XA,T7491XA<br>T7492XA,T754XXA,T7601XA,T7602XA,T7611XA,T7612XA,T7621XA<br>T7622XA,T7631XA,T7632XA,T7651XA,T7652XA,T7661XA,T7662XA<br>T7691XA,T7692XA,V00118A,V00128A,V00138A,V00148A,V00158A<br>V00188A,V00218A,V00228A,V00288A,V00318A,V00328A,V00388A<br>V00818A,V00828A,V00838A,V00848A,V00898A,W230XXA,W231XXA<br>W232XXA,W34010A,W34011A,W34018A,W34110A,W34111A,W34118A<br>W35XXXA,W361XXA,W362XXA,W363XXA,W368XXA,W369XXA,W370XXA<br>W378XXA,W38XXXA,W39XXXA,W400XXA,W401XXA,W408XXA,W409XXA<br>W4901XA,W4902XA,W4903XA,W4904XA,W4909XA,W499XXA,W85XXXA<br>W860XXA,W861XXA,W868XXA,W880XXA,W881XXA,W888XXA,W890XXA<br>W891XXA,W898XXA,W899XXA,W900XXA,W901XXA,W902XXA,W908XXA<br>X7401XA,X7402XA,X7409XA,X75XXXA,X818XXA,X831XXA,X838XXA<br>X9501XA,X9502XA,X9509XA,X960XXA,X961XXA,X962XXA,X963XXA<br>X964XXA,X968XXA,X969XXA,Y028XXA,Y0701,Y0702,Y0703,Y0704,Y0711<br>Y0712,Y0713,Y0714,Y07410,Y07411,Y07420,Y07421,Y07430,Y07432<br>Y07433,Y07434,Y07435,Y07436,Y07490,Y07491,Y07499,Y0750,Y07510<br>Y07511,Y07512,Y07513,Y07519,Y07521,Y07528,Y07529,Y0753,Y0759<br>Y076,Y079,Y0889XA,Y240XXA,Y25XXXA,Y31XXXA,Y33XXXA,Y35041A<br>Y35042A,Y35043A,Y35049A,Y35101A,Y35102A,Y35103A,Y35109A<br>Y35111A,Y35112A,Y35113A,Y35119A,Y35121A,Y35122A,Y35123A<br>Y35129A,Y35191A,Y35192A,Y35193A,Y35199A,Y35831A,Y35832A | 9300,9301,9302,9308,9309,931,932,9330,9331,9340,9341,9348<br>9349,9350,9351,9352,936,937,938,9390,9391,9392,9393,9399,9948<br>99550,99551,99552,99553,99554,99555,99580,99581,99582,99583<br>99584,E8843,E8845,E8849,E8850,E8851,E8852,E8853,E8854,E8869<br>E9040,E911,E912,E914,E915,E918,E9210,E9211,E9218,E9224,E9225<br>E9228,E9230,E9231,E9232,E9238,E9239,E9250,E9251,E9252,E9258<br>E9260,E9261,E9262,E9263,E9264,E9265,E9268,E9284,E9285,E9288<br>E9555,E9556,E9557,E9580,E9584,E9587,E9588,E9654,E9655,E9656<br>E9657,E9658,E9659,E9670,E9672,E9673,E9675,E9677,E9678,E9679<br>E9684,E9685,E9686,E9688,E970,E971,E975,E9790,E9792,E9795<br>E9796,E9798,E9799,E9855,E9856,E9880,E9888,E9900,E9902,E9910<br>E9913,E9914,E9915,E9916,E9917,E9918,E9919,E9920,E9921,E9922<br>E9923,E9928,E9929,E9930,E9931,E9932,E9933,E9934,E9935,E9936<br>E9937,E9938,E9939,E9951,E9952,E9958,E9959,E9960,E9961,E9962<br>E9963,E9968,E9969,E9971,E9973,E9980,E9981,E9988 |

Continued on next page

**Supplementary Table S1:** (cont'd) A comprehensive mapping of diagnosis groups used in the experiments to their corresponding ICD-10 and ICD-9 codes.

| Diagnosis | ICD-10 | ICD-9 |
|---|---|---|
| | Y35833A,Y35839A,Y35891A,Y35892A,Y35893A,Y36000A,Y36001A Y36010A,Y36011A,Y36020A,Y36021A,Y36030A,Y36031A,Y36040A Y36041A,Y36050A,Y36051A,Y36090A,Y36091A,Y36200A,Y36201A Y36210A,Y36211A,Y36220A,Y36221A,Y36230A,Y36231A,Y36240A Y36241A,Y36250A,Y36251A,Y36260A,Y36261A,Y36270A,Y36271A Y36290A,Y36291A,Y36310A,Y36311A,Y36320A,Y36321A,Y36410A Y36411A,Y36450A,Y36451A,Y36490A,Y36491A,Y36500A,Y36501A Y36510A,Y36511A,Y36520A,Y36521A,Y36530A,Y36531A,Y36540A Y36541A,Y36590A,Y36591A,Y366X0A,Y366X1A,Y36810A,Y36811A Y36820A,Y36821A,Y36880A,Y36881A,Y3691XA,Y37000A,Y37001A Y37010A,Y37011A,Y37020A,Y37021A,Y37030A,Y37031A,Y37040A Y37041A,Y37050A,Y37051A,Y37090A,Y37091A,Y37200A,Y37201A Y37210A,Y37211A,Y37220A,Y37221A,Y37230A,Y37231A,Y37240A Y37241A,Y37250A,Y37251A,Y37260A,Y37261A,Y37270A,Y37271A Y37290A,Y37291A,Y37310A,Y37311A,Y37320A,Y37321A,Y37410A Y37411A,Y37450A,Y37451A,Y37490A,Y37491A,Y37500A,Y37501A Y37510A,Y37511A,Y37520A,Y37521A,Y37530A,Y37531A,Y37540A Y37541A,Y37590A,Y37591A,Y376X0A,Y376X1A,Y3791XA,Y380X1A Y380X2A,Y380X3A,Y382X1A,Y382X2A,Y382X3A,Y385X1A,Y385X2A Y385X3A,Y386X1A,Y386X2A,Y386X3A,Y38811A,Y38812A,Y38891A Y38892A,Y38893A,Y389X1A,Y389X2A | |
| External cause codes: overexertion; initial encounter | T733XXA,W1840XA,W1841XA,W1842XA,W1843XA,W1849XA,X500XXA X501XXA,X503XXA,X509XXA | 9945,E8859,E9272 |
| External cause codes: pedal cyclist; not MVT; initial encounter | V100XXA,V101XXA,V102XXA,V103XXA,V104XXA,V105XXA,V109XXA V110XXA,V111XXA,V112XXA,V113XXA,V114XXA,V115XXA,V119XXA V120XXA,V121XXA,V122XXA,V130XXA,V131XXA,V132XXA,V140XXA V141XXA,V142XXA,V150XXA,V151XXA,V152XXA,V153XXA,V154XXA V155XXA,V159XXA,V160XXA,V161XXA,V162XXA,V163XXA,V164XXA V165XXA,V169XXA,V170XXA,V171XXA,V172XXA,V173XXA,V174XXA V175XXA,V179XXA,V180XXA,V181XXA,V182XXA,V183XXA,V184XXA V185XXA,V189XXA,V1900XA,V1909XA,V1910XA,V1919XA,V1920XA V1929XA,V193XXA,V1981XA,V1988XA,V2001XA,V2011XA,V2021XA V2031XA,V2041XA,V2051XA,V2091XA,V2101XA,V2111XA,V2121XA V2131XA,V2141XA,V2151XA,V2191XA,V2201XA,V2211XA,V2221XA V2301XA,V2311XA,V2321XA,V2401XA,V2411XA,V2421XA,V2501XA V2511XA,V2521XA,V2531XA,V2541XA,V2551XA,V2591XA,V2601XA V2611XA,V2621XA,V2631XA,V2641XA,V2651XA,V2691XA,V2701XA | E8013,E8226,E8261,E8269 |

Continued on next page

**Supplementary Table S1:** (cont'd) A comprehensive mapping of diagnosis groups used in the experiments to their corresponding ICD-10 and ICD-9 codes.

| Diagnosis | ICD-10 | ICD-9 |
| --- | --- | --- |
| | V2711XA,V2721XA,V2731XA,V2741XA,V2751XA,V2791XA,V2801XA V2811XA,V2821XA,V2831XA,V2841XA,V2851XA,V2891XA,V29001A V29091A,V29101A,V29191A,V29201A,V29291A,V2931XA,V29811A V29881A | |
| External cause codes: pedestrian; not MVT; initial encounter | V0100XA,V0101XA,V0102XA,V01031A,V01038A,V0109XA,V0110XA V0111XA,V0112XA,V01131A,V01138A,V0119XA,V0190XA,V0191XA V0192XA,V01931A,V01938A,V0199XA,V0200XA,V0201XA,V0202XA V02031A,V02038A,V0209XA,V0300XA,V0301XA,V0302XA,V03031A V03038A,V0309XA,V0400XA,V0401XA,V0402XA,V04031A,V04038A V0409XA,V0500XA,V0501XA,V0502XA,V05031A,V05038A,V0509XA V0510XA,V0511XA,V0512XA,V05131A,V05138A,V0519XA,V0590XA V0591XA,V0592XA,V05931A,V05938A,V0599XA,V0600XA,V0601XA V0602XA,V06031A,V06038A,V0609XA,V0610XA,V0611XA,V0612XA V06131A,V06138A,V0619XA,V0690XA,V0691XA,V0692XA,V06931A V06938A,V0699XA,V0900XA,V0901XA,V0909XA,V091XXA,V099XXA | E8052,E8072,E8107,E8227,E8260,E8290 |
| External cause codes: place of occurrence of the external cause | Y92000,Y92001,Y92002,Y92003,Y92007,Y92008,Y92009,Y92010 Y92011,Y92012,Y92013,Y92014,Y92015,Y92016,Y92017,Y92018 Y92019,Y92020,Y92021,Y92022,Y92023,Y92024,Y92025,Y92026 Y92027,Y92028,Y92029,Y92030,Y92031,Y92032,Y92038,Y92039 Y92040,Y92041,Y92042,Y92043,Y92044,Y92045,Y92046,Y92048 Y92049,Y92090,Y92091,Y92092,Y92093,Y92094,Y92095,Y92096 Y92098,Y92099,Y9210,Y92110,Y92111,Y92112,Y92113,Y92114,Y92115 Y92116,Y92118,Y92119,Y92120,Y92121,Y92122,Y92123,Y92124 Y92125,Y92126,Y92128,Y92129,Y92130,Y92131,Y92133,Y92135 Y92136,Y92137,Y92138,Y92139,Y92140,Y92141,Y92142,Y92143 Y92146,Y92147,Y92148,Y92149,Y92150,Y92151,Y92152,Y92153 Y92154,Y92155,Y92156,Y92157,Y92158,Y92159,Y92160,Y92161 Y92162,Y92163,Y92168,Y92169,Y92190,Y92191,Y92192,Y92193 Y92194,Y92195,Y92196,Y92197,Y92198,Y92199,Y92210,Y92211 Y92212,Y92213,Y92214,Y92215,Y92218,Y92219,Y9222,Y92230,Y92231 Y92232,Y92233,Y92234,Y92238,Y92239,Y92240,Y92241,Y92242 Y92243,Y92248,Y92250,Y92251,Y92252,Y92253,Y92254,Y92258,Y9226 Y9229,Y92310,Y92311,Y92312,Y92318,Y92320,Y92321,Y92322,Y92328 Y92330,Y92331,Y9234,Y9239,Y92410,Y92411,Y92412,Y92413,Y92414 Y92415,Y92480,Y92481,Y92482,Y92488,Y92510,Y92511,Y92512 Y92513,Y92520,Y92521,Y92522,Y92523,Y92524,Y92530,Y92531 Y92532,Y92538,Y9259,Y9261,Y9262,Y9263,Y9264,Y9265,Y9269,Y9271 Y9272,Y9273,Y9274,Y9279,Y92810,Y92811,Y92812,Y92813,Y92814 | E8490,E8491,E8492,E8493,E8494,E8495,E8496,E8497,E8498,E8499 |

Continued on next page

**Supplementary Table S1:** (cont'd) A comprehensive mapping of diagnosis groups used in the experiments to their corresponding ICD-10 and ICD-9 codes.

| Diagnosis | ICD-10 | ICD-9 |
|---|---|---|
| | Y92815,Y92816,Y92818,Y92820,Y92821,Y92828,Y92830,Y92831 | |
| | Y92832,Y92833,Y92834,Y92838,Y9284,Y9285,Y9286,Y9289,Y929 | |
| External cause codes: | T360X1A,T360X2A,T360X3A,T360X4A,T361X1A,T361X2A,T361X3A | 9600,9601,9602,9603,9604,9605,9606,9608,9609,9610,9614,9615 |
| poisoning by drug | T361X4A,T362X1A,T362X2A,T362X3A,T362X4A,T363X1A,T363X2A | 9616,9617,9618,9619,9620,9621,9622,9623,9624,9627,9628,9629 |
| | T363X3A,T363X4A,T364X1A,T364X2A,T364X3A,T364X4A,T365X1A | 9630,9631,9634,9635,9638,9639,9640,9642,9643,9644,9645,9646 |
| | T365X2A,T365X3A,T365X4A,T366X1A,T366X2A,T366X3A,T366X4A | 9648,9649,96500,96501,96502,96509,9651,9654,9655,96561,96569 |
| | T367X1A,T367X2A,T367X3A,T367X4A,T368X1A,T368X2A,T368X3A | 9657,9659,9660,9661,9662,9663,9664,9670,9678,9679,9680,9682 |
| | T368X4A,T3691XA,T3692XA,T3693XA,T3694XA,T370X1A,T370X2A | 9683,9684,9685,9689,96900,96901,96902,96903,96904,96905,96909 |
| | T370X3A,T370X4A,T371X1A,T371X2A,T371X3A,T371X4A,T372X1A | 9691,9692,9693,9694,9696,96970,96971,96972,96973,96979,9698 |
| | T372X2A,T372X3A,T372X4A,T373X1A,T373X2A,T373X3A,T373X4A | 9699,9700,9701,97081,9710,9711,9712,9713,9719,9720,9721,9722 |
| | T374X1A,T374X2A,T374X3A,T374X4A,T375X1A,T375X2A,T375X3A | 9723,9724,9725,9726,9727,9729,9730,9731,9732,9733,9734,9735 |
| | T375X4A,T378X1A,T378X2A,T378X3A,T378X4A,T3791XA,T3792XA | 9736,9738,9739,9742,9744,9745,9747,9750,9751,9752,9753,9754 |
| | T3793XA,T3794XA,T380X1A,T380X2A,T380X3A,T380X4A,T381X1A | 9755,9756,9757,9758,9760,9761,9762,9763,9764,9765,9766,9767 |
| | T381X2A,T381X3A,T381X4A,T382X1A,T382X2A,T382X3A,T382X4A | 9768,9769,9770,9772,9778,9779,9786,9788,9789,9790,9796,9799 |
| | T383X1A,T383X2A,T383X3A,T383X4A,T384X1A,T384X2A,T384X3A | E8500,E8501,E8502,E8503,E8504,E8505,E8506,E8507,E851,E8528 |
| | T384X4A,T385X1A,T385X2A,T385X3A,T385X4A,T386X1A,T386X2A | E8530,E8531,E8532,E8540,E8541,E8542,E8543,E8548,E8550,E8551 |
| | T386X3A,T386X4A,T387X1A,T387X2A,T387X3A,T387X4A,T38801A | E8552,E8553,E8554,E8555,E8556,E8558,E856,E857,E8580,E8581 |
| | T38802A,T38803A,T38804A,T38811A,T38812A,T38813A,T38814A | E8582,E8583,E8584,E8585,E8586,E8587,E8588,E9500,E9501,E9502 |
| | T38891A,T38892A,T38893A,T38894A,T38901A,T38902A,T38903A | E9503,E9504,E9528,E9620,E9800,E9801,E9802,E9803,E9804,E9805 |
| | T38904A,T38991A,T38992A,T38993A,T38994A,T39011A,T39012A | |
| | T39013A,T39014A,T39091A,T39092A,T39093A,T39094A,T391X1A | |
| | T391X2A,T391X3A,T391X4A,T392X1A,T392X2A,T392X3A,T392X4A | |
| | T39311A,T39312A,T39313A,T39314A,T39391A,T39392A,T39393A | |
| | T39394A,T394X1A,T394X2A,T394X3A,T394X4A,T398X1A,T398X2A | |
| | T398X3A,T398X4A,T3991XA,T3992XA,T3993XA,T3994XA,T400X1A | |
| | T400X2A,T400X3A,T400X4A,T401X1A,T401X2A,T401X3A,T401X4A | |
| | T402X1A,T402X2A,T402X3A,T402X4A,T403X1A,T403X2A,T403X3A | |
| | T403X4A,T40411A,T40412A,T40413A,T40414A,T40421A,T40422A | |
| | T40423A,T40424A,T40491A,T40492A,T40493A,T40494A,T404X1A | |
| | T404X2A,T404X3A,T404X4A,T405X1A,T405X2A,T405X3A,T405X4A | |
| | T40601A,T40602A,T40603A,T40604A,T40691A,T40692A,T40693A | |
| | T40694A,T40711A,T40712A,T40713A,T40714A,T40721A,T40722A | |
| | T40723A,T40724A,T407X1A,T407X2A,T407X3A,T407X4A,T408X1A | |
| | T408X2A,T408X3A,T408X4A,T40901A,T40902A,T40903A,T40904A | |
| | T40991A,T40992A,T40993A,T40994A,T410X1A,T410X2A,T410X3A | |
| | T410X4A,T411X1A,T411X2A,T411X3A,T411X4A,T41201A,T41202A | |
| | T41203A,T41204A,T41291A,T41292A,T41293A,T41294A,T413X1A | |
| | T413X2A,T413X3A,T413X4A,T4141XA,T4142XA,T4143XA,T4144XA | |
| | T415X1A,T415X2A,T415X3A,T415X4A,T420X1A,T420X2A,T420X3A | |

Continued on next page

**Supplementary Table S1:** (cont'd) A comprehensive mapping of diagnosis groups used in the experiments to their corresponding ICD-10 and ICD-9 codes.

| Diagnosis | ICD-10 | ICD-9 |
|---|---|---|
| | T420X4A,T421X1A,T421X2A,T421X3A,T421X4A,T422X1A,T422X2A | |
| | T422X3A,T422X4A,T423X1A,T423X2A,T423X3A,T423X4A,T424X1A | |
| | T424X2A,T424X3A,T424X4A,T425X1A,T425X2A,T425X3A,T425X4A | |
| | T426X1A,T426X2A,T426X3A,T426X4A,T4271XA,T4272XA,T4273XA | |
| | T4274XA,T428X1A,T428X2A,T428X3A,T428X4A,T43011A,T43012A | |
| | T43013A,T43014A,T43021A,T43022A,T43023A,T43024A,T431X1A | |
| | T431X2A,T431X3A,T431X4A,T43201A,T43202A,T43203A,T43204A | |
| | T43211A,T43212A,T43213A,T43214A,T43221A,T43222A,T43223A | |
| | T43224A,T43291A,T43292A,T43293A,T43294A,T433X1A,T433X2A | |
| | T433X3A,T433X4A,T434X1A,T434X2A,T434X3A,T434X4A,T43501A | |
| | T43502A,T43503A,T43504A,T43591A,T43592A,T43593A,T43594A | |
| | T43601A,T43602A,T43603A,T43604A,T43611A,T43612A,T43613A | |
| | T43614A,T43621A,T43622A,T43623A,T43624A,T43631A,T43632A | |
| | T43633A,T43634A,T43641A,T43642A,T43643A,T43644A,T43651A | |
| | T43652A,T43653A,T43654A,T43691A,T43692A,T43693A,T43694A | |
| | T438X1A,T438X2A,T438X3A,T438X4A,T4391XA,T4392XA,T4393XA | |
| | T4394XA,T440X1A,T440X2A,T440X3A,T440X4A,T441X1A,T441X2A | |
| | T441X3A,T441X4A,T442X1A,T442X2A,T442X3A,T442X4A,T443X1A | |
| | T443X2A,T443X3A,T443X4A,T444X1A,T444X2A,T444X3A,T444X4A | |
| | T445X1A,T445X2A,T445X3A,T445X4A,T446X1A,T446X2A,T446X3A | |
| | T446X4A,T447X1A,T447X2A,T447X3A,T447X4A,T448X1A,T448X2A | |
| | T448X3A,T448X4A,T44901A,T44902A,T44903A,T44904A,T44991A | |
| | T44992A,T44993A,T44994A,T450X1A,T450X2A,T450X3A,T450X4A | |
| | T451X1A,T451X2A,T451X3A,T451X4A,T452X1A,T452X2A,T452X3A | |
| | T452X4A,T453X1A,T453X2A,T453X3A,T453X4A,T454X1A,T454X2A | |
| | T454X3A,T454X4A,T45511A,T45512A,T45513A,T45514A,T45521A | |
| | T45522A,T45523A,T45524A,T45601A,T45602A,T45603A,T45604A | |
| | T45611A,T45612A,T45613A,T45614A,T45621A,T45622A,T45623A | |
| | T45624A,T45691A,T45692A,T45693A,T45694A,T457X1A,T457X2A | |
| | T457X3A,T457X4A,T458X1A,T458X2A,T458X3A,T458X4A,T4591XA | |
| | T4592XA,T4593XA,T4594XA,T460X1A,T460X2A,T460X3A,T460X4A | |
| | T461X1A,T461X2A,T461X3A,T461X4A,T462X1A,T462X2A,T462X3A | |
| | T462X4A,T463X1A,T463X2A,T463X3A,T463X4A,T464X1A,T464X2A | |
| | T464X3A,T464X4A,T465X1A,T465X2A,T465X3A,T465X4A,T466X1A | |
| | T466X2A,T466X3A,T466X4A,T467X1A,T467X2A,T467X3A,T467X4A | |
| | T468X1A,T468X2A,T468X3A,T468X4A,T46901A,T46902A,T46903A | |
| | T46904A,T46991A,T46992A,T46993A,T46994A,T470X1A,T470X2A | |
| | T470X3A,T470X4A,T471X1A,T471X2A,T471X3A,T471X4A,T472X1A | |
| | T472X2A,T472X3A,T472X4A,T473X1A,T473X2A,T473X3A,T473X4A | |
| | T474X1A,T474X2A,T474X3A,T474X4A,T475X1A,T475X2A,T475X3A | |

Continued on next page

**Supplementary Table S1:** (cont'd) A comprehensive mapping of diagnosis groups used in the experiments to their corresponding ICD-10 and ICD-9 codes.

| Diagnosis | ICD-10 | ICD-9 |
|---|---|---|
| | T475X4A,T476X1A,T476X2A,T476X3A,T476X4A,T477X1A,T477X2A<br>T477X3A,T477X4A,T478X1A,T478X2A,T478X3A,T478X4A,T4791XA<br>T4792XA,T4793XA,T4794XA,T480X1A,T480X2A,T480X3A,T480X4A<br>T481X1A,T481X2A,T481X3A,T481X4A,T48201A,T48202A,T48203A<br>T48204A,T48291A,T48292A,T48293A,T48294A,T483X1A,T483X2A<br>T483X3A,T483X4A,T484X1A,T484X2A,T484X3A,T484X4A,T485X1A<br>T485X2A,T485X3A,T485X4A,T486X1A,T486X2A,T486X3A,T486X4A<br>T48901A,T48902A,T48903A,T48904A,T48991A,T48992A,T48993A<br>T48994A,T490X1A,T490X2A,T490X3A,T490X4A,T491X1A,T491X2A<br>T491X3A,T491X4A,T492X1A,T492X2A,T492X3A,T492X4A,T493X1A<br>T493X2A,T493X3A,T493X4A,T494X1A,T494X2A,T494X3A,T494X4A<br>T495X1A,T495X2A,T495X3A,T495X4A,T496X1A,T496X2A,T496X3A<br>T496X4A,T497X1A,T497X2A,T497X3A,T497X4A,T498X1A,T498X2A<br>T498X3A,T498X4A,T4991XA,T4992XA,T4993XA,T4994XA,T500X1A<br>T500X2A,T500X3A,T500X4A,T501X1A,T501X2A,T501X3A,T501X4A<br>T502X1A,T502X2A,T502X3A,T502X4A,T503X1A,T503X2A,T503X3A<br>T503X4A,T504X1A,T504X2A,T504X3A,T504X4A,T505X1A,T505X2A<br>T505X3A,T505X4A,T506X1A,T506X2A,T506X3A,T506X4A,T507X1A<br>T507X2A,T507X3A,T507X4A,T508X1A,T508X2A,T508X3A,T508X4A<br>T50901A,T50902A,T50903A,T50904A,T50911A,T50912A,T50913A<br>T50914A,T50991A,T50992A,T50993A,T50994A,T50A11A,T50A12A<br>T50A13A,T50A14A,T50A21A,T50A22A,T50A23A,T50A24A,T50A91A<br>T50A92A,T50A93A,T50A94A,T50B11A,T50B12A,T50B13A,T50B14A<br>T50B91A,T50B92A,T50B93A,T50B94A,T50Z11A,T50Z12A,T50Z13A<br>T50Z14A,T50Z91A,T50Z92A,T50Z93A,T50Z94A | |
| External cause codes:<br>poisoning by non-drug | T510X1A,T510X2A,T510X3A,T510X4A,T511X1A,T511X2A,T511X3A<br>T511X4A,T512X1A,T512X2A,T512X3A,T512X4A,T513X1A,T513X2A<br>T513X3A,T513X4A,T518X1A,T518X2A,T518X3A,T518X4A,T5191XA<br>T5192XA,T5193XA,T5194XA,T520X1A,T520X2A,T520X3A,T520X4A<br>T521X1A,T521X2A,T521X3A,T521X4A,T522X1A,T522X2A,T522X3A<br>T522X4A,T523X1A,T523X2A,T523X3A,T523X4A,T524X1A,T524X2A<br>T524X3A,T524X4A,T528X1A,T528X2A,T528X3A,T528X4A,T5291XA<br>T5292XA,T5293XA,T5294XA,T530X1A,T530X2A,T530X3A,T530X4A<br>T531X1A,T531X2A,T531X3A,T531X4A,T532X1A,T532X2A,T532X3A<br>T532X4A,T533X1A,T533X2A,T533X3A,T533X4A,T534X1A,T534X2A<br>T534X3A,T534X4A,T535X1A,T535X2A,T535X3A,T535X4A,T536X1A<br>T536X2A,T536X3A,T536X4A,T537X1A,T537X2A,T537X3A,T537X4A<br>T5391XA,T5392XA,T5393XA,T5394XA,T540X1A,T540X2A,T540X3A<br>T540X4A,T550X1A,T550X2A,T550X3A,T550X4A,T551X1A,T551X2A | 9800,9801,9802,9803,9808,9809,981,9820,9821,9822,9823,9824<br>9828,9830,9839,9849,9850,9851,9852,9853,9855,9856,9858,9859<br>986,9871,9872,9873,9874,9875,9876,9877,9878,9879,9880,9881<br>9882,9888,9889,9890,9891,9893,9894,9896,9897,98982,98984<br>98989,9899,E8601,E8602,E8603,E8604,E8608,E8609,E8610,E8611<br>E8616,E8621,E8624,E8631,E8634,E8635,E8637,E8640,E8641,E8643<br>E8644,E8651,E8652,E8653,E8654,E8655,E8658,E8659,E8660,E8661<br>E8663,E8664,E8668,E8681,E8682,E8683,E8688,E8689,E8690,E8691<br>E8692,E8693,E8698,E9506,E9507,E9508,E9509,E9520,E9521,E9528<br>E9529,E961,E9621,E9622,E9629,E972,E9797,E9806,E9807,E9808<br>E9809,E9820,E9821,E9828,E9970,E9972,E9978,E9979 |

Continued on next page

**Supplementary Table S1:** (cont'd) A comprehensive mapping of diagnosis groups used in the experiments to their corresponding ICD-10 and ICD-9 codes.

| Diagnosis | ICD-10 | ICD-9 |
|---|---|---|
| | T551X3A,T551X4A,T560X1A,T560X2A,T560X3A,T560X4A,T561X1A | |
| | T561X2A,T561X3A,T561X4A,T562X1A,T562X2A,T562X3A,T562X4A | |
| | T563X1A,T563X2A,T563X3A,T563X4A,T564X1A,T564X2A,T564X3A | |
| | T564X4A,T565X1A,T565X2A,T565X3A,T565X4A,T566X1A,T566X2A | |
| | T566X3A,T566X4A,T567X1A,T567X2A,T567X3A,T567X4A,T56811A | |
| | T56812A,T56813A,T56814A,T56891A,T56892A,T56893A,T56894A | |
| | T5691XA,T5692XA,T5693XA,T5694XA,T570X1A,T570X2A,T570X3A | |
| | T570X4A,T571X1A,T571X2A,T571X3A,T571X4A,T572X1A,T572X2A | |
| | T572X3A,T572X4A,T573X1A,T573X2A,T573X3A,T573X4A,T578X1A | |
| | T578X2A,T578X3A,T578X4A,T5791XA,T5792XA,T5793XA,T5794XA | |
| | T5801XA,T5802XA,T5803XA,T5804XA,T5811XA,T5812XA,T5813XA | |
| | T5814XA,T582X1A,T582X2A,T582X3A,T582X4A,T588X1A,T588X2A | |
| | T588X3A,T588X4A,T5891XA,T5892XA,T5893XA,T5894XA,T590X1A | |
| | T590X2A,T590X3A,T590X4A,T591X1A,T591X2A,T591X3A,T591X4A | |
| | T592X1A,T592X2A,T592X3A,T592X4A,T593X1A,T593X2A,T593X3A | |
| | T593X4A,T594X1A,T594X2A,T594X3A,T594X4A,T595X1A,T595X2A | |
| | T595X3A,T595X4A,T596X1A,T596X2A,T596X3A,T596X4A,T597X1A | |
| | T597X2A,T597X3A,T597X4A,T59811A,T59812A,T59813A,T59814A | |
| | T59891A,T59892A,T59893A,T59894A,T5991XA,T5992XA,T5993XA | |
| | T5994XA,T600X1A,T600X2A,T600X3A,T600X4A,T601X1A,T601X2A | |
| | T601X3A,T601X4A,T602X1A,T602X2A,T602X3A,T602X4A,T603X1A | |
| | T603X2A,T603X3A,T603X4A,T604X1A,T604X2A,T604X3A,T604X4A | |
| | T608X1A,T608X2A,T608X3A,T608X4A,T6091XA,T6092XA,T6093XA | |
| | T6094XA,T6101XA,T6102XA,T6103XA,T6104XA,T6111XA,T6112XA | |
| | T6113XA,T6114XA,T61771A,T61772A,T61773A,T61774A,T61781A | |
| | T61782A,T61783A,T61784A,T618X1A,T618X2A,T618X3A,T618X4A | |
| | T6191XA,T6192XA,T6193XA,T6194XA,T620X1A,T620X2A,T620X3A | |
| | T620X4A,T621X1A,T621X2A,T621X3A,T621X4A,T622X1A,T622X2A | |
| | T622X3A,T622X4A,T628X1A,T628X2A,T628X3A,T628X4A,T6291XA | |
| | T6292XA,T6293XA,T6294XA,T6401XA,T6402XA,T6403XA,T6404XA | |
| | T6481XA,T6482XA,T6483XA,T6484XA,T650X1A,T650X2A,T650X3A | |
| | T650X4A,T651X1A,T651X2A,T651X3A,T651X4A,T65211A,T65212A | |
| | T65213A,T65214A,T65221A,T65222A,T65223A,T65224A,T65291A | |
| | T65292A,T65293A,T65294A,T653X1A,T653X2A,T653X3A,T653X4A | |
| | T654X1A,T654X2A,T654X3A,T654X4A,T655X1A,T655X2A,T655X3A | |
| | T655X4A,T656X1A,T656X2A,T656X3A,T656X4A,T65811A,T65812A | |
| | T65813A,T65814A,T65831A,T65832A,T65833A,T65834A,T65891A | |
| | T65892A,T65893A,T65894A,T6591XA,T6592XA,T6593XA,T6594XA | |
| | Y35201A,Y35202A,Y35203A,Y35209A,Y35211A,Y35212A,Y35213A | |
| | Y35219A,Y35291A,Y35292A,Y35293A,Y35299A,Y367X0A,Y367X1A | |

Continued on next page

**Supplementary Table S1:** (cont'd) A comprehensive mapping of diagnosis groups used in the experiments to their corresponding ICD-10 and ICD-9 codes.

| Diagnosis | ICD-10 | ICD-9 |
|---|---|---|
| | Y377X0A,Y377X1A,Y387X1A,Y387X2A,Y387X3A | |
| External cause codes: | V0001XS,V0002XS,V00031S,V00038S,V0009XS,V00111S,V00112S | E9290,E9291,E9293,E9294,E9295,E9298,E959,E969,E977,E9990 |
| sequela | V00118S,V00121S,V00122S,V00128S,V00131S,V00132S,V00138S | E9991 |
| | V00141S,V00142S,V00148S,V00151S,V00152S,V00158S,V00181S | |
| | V00182S,V00188S,V00211S,V00212S,V00218S,V00221S,V00222S | |
| | V00228S,V00281S,V00282S,V00288S,V00311S,V00312S,V00318S | |
| | V00321S,V00322S,V00328S,V00381S,V00382S,V00388S,V00811S | |
| | V00812S,V00818S,V00821S,V00822S,V00828S,V00831S,V00832S | |
| | V00838S,V00841S,V00842S,V00848S,V00891S,V00892S,V00898S | |
| | V0100XS,V0101XS,V0102XS,V01031S,V01038S,V0109XS,V0110XS | |
| | V0111XS,V0112XS,V01131S,V01138S,V0119XS,V0190XS,V0191XS | |
| | V0192XS,V01931S,V01938S,V0199XS,V0200XS,V0201XS,V0202XS | |
| | V02031S,V02038S,V0209XS,V0210XS,V0211XS,V0212XS,V02131S | |
| | V02138S,V0219XS,V0290XS,V0291XS,V0292XS,V02931S,V02938S | |
| | V0299XS,V0300XS,V0301XS,V0302XS,V03031S,V03038S,V0309XS | |
| | V0310XS,V0311XS,V0312XS,V03131S,V03138S,V0319XS,V0390XS | |
| | V0391XS,V0392XS,V03931S,V03938S,V0399XS,V0400XS,V0401XS | |
| | V0402XS,V04031S,V04038S,V0409XS,V0410XS,V0411XS,V0412XS | |
| | V04131S,V04138S,V0419XS,V0490XS,V0491XS,V0492XS,V04931S | |
| | V04938S,V0499XS,V0500XS,V0501XS,V0502XS,V05031S,V05038S | |
| | V0509XS,V0510XS,V0511XS,V0512XS,V05131S,V05138S,V0519XS | |
| | V0590XS,V0591XS,V0592XS,V05931S,V05938S,V0599XS,V0600XS | |
| | V0601XS,V0602XS,V06031S,V06038S,V0609XS,V0610XS,V0611XS | |
| | V0612XS,V06131S,V06138S,V0619XS,V0690XS,V0691XS,V0692XS | |
| | V06931S,V06938S,V0699XS,V0900XS,V0901XS,V0909XS,V091XXS | |
| | V0920XS,V0921XS,V0929XS,V093XXS,V099XXS,V100XXS,V101XXS | |
| | V102XXS,V103XXS,V104XXS,V105XXS,V109XXS,V110XXS,V111XXS | |
| | V112XXS,V113XXS,V114XXS,V115XXS,V119XXS,V120XXS,V121XXS | |
| | V122XXS,V123XXS,V124XXS,V125XXS,V129XXS,V130XXS,V131XXS | |
| | V132XXS,V133XXS,V134XXS,V135XXS,V139XXS,V140XXS,V141XXS | |
| | V142XXS,V143XXS,V144XXS,V145XXS,V149XXS,V150XXS,V151XXS | |
| | V152XXS,V153XXS,V154XXS,V155XXS,V159XXS,V160XXS,V161XXS | |
| | V162XXS,V163XXS,V164XXS,V165XXS,V169XXS,V170XXS,V171XXS | |
| | V172XXS,V173XXS,V174XXS,V175XXS,V179XXS,V180XXS,V181XXS | |
| | V182XXS,V183XXS,V184XXS,V185XXS,V189XXS,V1900XS,V1909XS | |
| | V1910XS,V1919XS,V1920XS,V1929XS,V193XXS,V1940XS,V1949XS | |
| | V1950XS,V1959XS,V1960XS,V1969XS,V1981XS,V1988XS,V199XXS | |
| | V2001XS,V2009XS,V200XXS,V2011XS,V2019XS,V201XXS,V2021XS | |
| | V2029XS,V202XXS,V2031XS,V2039XS,V203XXS,V2041XS,V2049XS | |

Continued on next page

**Supplementary Table S1:** (cont'd) A comprehensive mapping of diagnosis groups used in the experiments to their corresponding ICD-10 and ICD-9 codes.

| Diagnosis | ICD-10 | ICD-9 |
|---|---|---|
| | V204XXS,V2051XS,V2059XS,V205XXS,V2091XS,V2099XS,V209XXS | |
| | V2101XS,V2109XS,V210XXS,V2111XS,V2119XS,V211XXS,V2121XS | |
| | V2129XS,V212XXS,V2131XS,V2139XS,V213XXS,V2141XS,V2149XS | |
| | V214XXS,V2151XS,V2159XS,V215XXS,V2191XS,V2199XS,V219XXS | |
| | V2201XS,V2209XS,V220XXS,V2211XS,V2219XS,V221XXS,V2221XS | |
| | V2229XS,V222XXS,V2231XS,V2239XS,V223XXS,V2241XS,V2249XS | |
| | V224XXS,V2251XS,V2259XS,V225XXS,V2291XS,V2299XS,V229XXS | |
| | V2301XS,V2309XS,V230XXS,V2311XS,V2319XS,V231XXS,V2321XS | |
| | V2329XS,V232XXS,V2331XS,V2339XS,V233XXS,V2341XS,V2349XS | |
| | V234XXS,V2351XS,V2359XS,V235XXS,V2391XS,V2399XS,V239XXS | |
| | V2401XS,V2409XS,V240XXS,V2411XS,V2419XS,V241XXS,V2421XS | |
| | V2429XS,V242XXS,V2431XS,V2439XS,V243XXS,V2441XS,V2449XS | |
| | V244XXS,V2451XS,V2459XS,V245XXS,V2491XS,V2499XS,V249XXS | |
| | V2501XS,V2509XS,V250XXS,V2511XS,V2519XS,V251XXS,V2521XS | |
| | V2529XS,V252XXS,V2531XS,V2539XS,V253XXS,V2541XS,V2549XS | |
| | V254XXS,V2551XS,V2559XS,V255XXS,V2591XS,V2599XS,V259XXS | |
| | V2601XS,V2609XS,V260XXS,V2611XS,V2619XS,V261XXS,V2621XS | |
| | V2629XS,V262XXS,V2631XS,V2639XS,V263XXS,V2641XS,V2649XS | |
| | V264XXS,V2651XS,V2659XS,V265XXS,V2691XS,V2699XS,V269XXS | |
| | V2701XS,V2709XS,V270XXS,V2711XS,V2719XS,V271XXS,V2721XS | |
| | V2729XS,V272XXS,V2731XS,V2739XS,V273XXS,V2741XS,V2749XS | |
| | V274XXS,V2751XS,V2759XS,V275XXS,V2791XS,V2799XS,V279XXS | |
| | V2801XS,V2809XS,V280XXS,V2811XS,V2819XS,V281XXS,V2821XS | |
| | V2829XS,V282XXS,V2831XS,V2839XS,V283XXS,V2841XS,V2849XS | |
| | V284XXS,V2851XS,V2859XS,V285XXS,V2891XS,V2899XS,V289XXS | |
| | V29001S,V29008S,V2900XS,V29091S,V29098S,V2909XS,V29101S | |
| | V29108S,V2910XS,V29191S,V29198S,V2919XS,V29201S,V29208S | |
| | V2920XS,V29291S,V29298S,V2929XS,V2931XS,V2939XS,V293XXS | |
| | V29401S,V29408S,V2940XS,V29491S,V29498S,V2949XS,V29501S | |
| | V29508S,V2950XS,V29591S,V29598S,V2959XS,V29601S,V29608S | |
| | V2960XS,V29691S,V29698S,V2969XS,V29811S,V29818S,V2981XS | |
| | V29881S,V29888S,V2988XS,V2991XS,V2999XS,V299XXS,V300XXS | |
| | V301XXS,V302XXS,V303XXS,V304XXS,V305XXS,V306XXS,V307XXS | |
| | V309XXS,V310XXS,V311XXS,V312XXS,V313XXS,V314XXS,V315XXS | |
| | V316XXS,V317XXS,V319XXS,V320XXS,V321XXS,V322XXS,V323XXS | |
| | V324XXS,V325XXS,V326XXS,V327XXS,V329XXS,V330XXS,V331XXS | |
| | V332XXS,V333XXS,V334XXS,V335XXS,V336XXS,V337XXS,V339XXS | |
| | V340XXS,V341XXS,V342XXS,V343XXS,V344XXS,V345XXS,V346XXS | |
| | V347XXS,V349XXS,V350XXS,V351XXS,V352XXS,V353XXS,V354XXS | |
| | V355XXS,V356XXS,V357XXS,V359XXS,V360XXS,V361XXS,V362XXS | |

Continued on next page

**Supplementary Table S1:** (cont'd) A comprehensive mapping of diagnosis groups used in the experiments to their corresponding ICD-10 and ICD-9 codes.

| Diagnosis | ICD-10 | ICD-9 |
|---|---|---|
| | V363XXS,V364XXS,V365XXS,V366XXS,V367XXS,V369XXS,V370XXS | |
| | V371XXS,V372XXS,V373XXS,V374XXS,V375XXS,V376XXS,V377XXS | |
| | V379XXS,V380XXS,V381XXS,V382XXS,V383XXS,V384XXS,V385XXS | |
| | V386XXS,V387XXS,V389XXS,V3900XS,V3909XS,V3910XS,V3919XS | |
| | V3920XS,V3929XS,V393XXS,V3940XS,V3949XS,V3950XS,V3959XS | |
| | V3960XS,V3969XS,V3981XS,V3989XS,V399XXS,V400XXS,V401XXS | |
| | V402XXS,V403XXS,V404XXS,V405XXS,V406XXS,V407XXS,V409XXS | |
| | V410XXS,V411XXS,V412XXS,V413XXS,V414XXS,V415XXS,V416XXS | |
| | V417XXS,V419XXS,V420XXS,V421XXS,V422XXS,V423XXS,V424XXS | |
| | V425XXS,V426XXS,V427XXS,V429XXS,V4301XS,V4302XS,V4303XS | |
| | V4304XS,V4311XS,V4312XS,V4313XS,V4314XS,V4321XS,V4322XS | |
| | V4323XS,V4324XS,V4331XS,V4332XS,V4333XS,V4334XS,V4341XS | |
| | V4342XS,V4343XS,V4344XS,V4351XS,V4352XS,V4353XS,V4354XS | |
| | V4361XS,V4362XS,V4363XS,V4364XS,V4371XS,V4372XS,V4373XS | |
| | V4374XS,V4391XS,V4392XS,V4393XS,V4394XS,V440XXS,V441XXS | |
| | V442XXS,V443XXS,V444XXS,V445XXS,V446XXS,V447XXS,V449XXS | |
| | V450XXS,V451XXS,V452XXS,V453XXS,V454XXS,V455XXS,V456XXS | |
| | V457XXS,V459XXS,V460XXS,V461XXS,V462XXS,V463XXS,V464XXS | |
| | V465XXS,V466XXS,V467XXS,V469XXS,V4701XS,V4702XS,V470XXS | |
| | V4711XS,V4712XS,V471XXS,V472XXS,V4731XS,V4732XS,V473XXS | |
| | V474XXS,V4751XS,V4752XS,V475XXS,V4761XS,V4762XS,V476XXS | |
| | V477XXS,V4791XS,V4792XS,V479XXS,V480XXS,V481XXS,V482XXS | |
| | V483XXS,V484XXS,V485XXS,V486XXS,V487XXS,V489XXS,V4900XS | |
| | V4909XS,V4910XS,V4919XS,V4920XS,V4929XS,V493XXS,V4940XS | |
| | V4949XS,V4950XS,V4959XS,V4960XS,V4969XS,V4981XS,V4988XS | |
| | V499XXS,V500XXS,V501XXS,V502XXS,V503XXS,V504XXS,V505XXS | |
| | V506XXS,V507XXS,V509XXS,V510XXS,V511XXS,V512XXS,V513XXS | |
| | V514XXS,V515XXS,V516XXS,V517XXS,V519XXS,V520XXS,V521XXS | |
| | V522XXS,V523XXS,V524XXS,V525XXS,V526XXS,V527XXS,V529XXS | |
| | V530XXS,V531XXS,V532XXS,V533XXS,V534XXS,V535XXS,V536XXS | |
| | V537XXS,V539XXS,V540XXS,V541XXS,V542XXS,V543XXS,V544XXS | |
| | V545XXS,V546XXS,V547XXS,V549XXS,V550XXS,V551XXS,V552XXS | |
| | V553XXS,V554XXS,V555XXS,V556XXS,V557XXS,V559XXS,V560XXS | |
| | V561XXS,V562XXS,V563XXS,V564XXS,V565XXS,V566XXS,V567XXS | |
| | V569XXS,V570XXS,V571XXS,V572XXS,V573XXS,V574XXS,V575XXS | |
| | V576XXS,V577XXS,V579XXS,V580XXS,V581XXS,V582XXS,V583XXS | |
| | V584XXS,V585XXS,V586XXS,V587XXS,V589XXS,V5900XS,V5909XS | |
| | V5910XS,V5919XS,V5920XS,V5929XS,V593XXS,V5940XS,V5949XS | |
| | V5950XS,V5959XS,V5960XS,V5969XS,V5981XS,V5988XS,V599XXS | |
| | V600XXS,V601XXS,V602XXS,V603XXS,V604XXS,V605XXS,V606XXS | |

Continued on next page

**Supplementary Table S1:** (cont'd) A comprehensive mapping of diagnosis groups used in the experiments to their corresponding ICD-10 and ICD-9 codes.

| Diagnosis | ICD-10 | ICD-9 |
|---|---|---|
| | V607XXS,V609XXS,V610XXS,V611XXS,V612XXS,V613XXS,V614XXS | |
| | V615XXS,V616XXS,V617XXS,V619XXS,V620XXS,V621XXS,V622XXS | |
| | V623XXS,V624XXS,V625XXS,V626XXS,V627XXS,V629XXS,V630XXS | |
| | V631XXS,V632XXS,V633XXS,V634XXS,V635XXS,V636XXS,V637XXS | |
| | V639XXS,V640XXS,V641XXS,V642XXS,V643XXS,V644XXS,V645XXS | |
| | V646XXS,V647XXS,V649XXS,V650XXS,V651XXS,V652XXS,V653XXS | |
| | V654XXS,V655XXS,V656XXS,V657XXS,V659XXS,V660XXS,V661XXS | |
| | V662XXS,V663XXS,V664XXS,V665XXS,V666XXS,V667XXS,V669XXS | |
| | V670XXS,V671XXS,V672XXS,V673XXS,V674XXS,V675XXS,V676XXS | |
| | V677XXS,V679XXS,V680XXS,V681XXS,V682XXS,V683XXS,V684XXS | |
| | V685XXS,V686XXS,V687XXS,V689XXS,V6900XS,V6909XS,V6910XS | |
| | V6919XS,V6920XS,V6929XS,V693XXS,V6940XS,V6949XS,V6950XS | |
| | V6959XS,V6960XS,V6969XS,V6981XS,V6988XS,V699XXS,V700XXS | |
| | V701XXS,V702XXS,V703XXS,V704XXS,V705XXS,V706XXS,V707XXS | |
| | V709XXS,V710XXS,V711XXS,V712XXS,V713XXS,V714XXS,V715XXS | |
| | V716XXS,V717XXS,V719XXS,V720XXS,V721XXS,V722XXS,V723XXS | |
| | V724XXS,V725XXS,V726XXS,V727XXS,V729XXS,V730XXS,V731XXS | |
| | V732XXS,V733XXS,V734XXS,V735XXS,V736XXS,V737XXS,V739XXS | |
| | V740XXS,V741XXS,V742XXS,V743XXS,V744XXS,V745XXS,V746XXS | |
| | V747XXS,V749XXS,V750XXS,V751XXS,V752XXS,V753XXS,V754XXS | |
| | V755XXS,V756XXS,V757XXS,V759XXS,V760XXS,V761XXS,V762XXS | |
| | V763XXS,V764XXS,V765XXS,V766XXS,V767XXS,V769XXS,V770XXS | |
| | V771XXS,V772XXS,V773XXS,V774XXS,V775XXS,V776XXS,V777XXS | |
| | V779XXS,V780XXS,V781XXS,V782XXS,V783XXS,V784XXS,V785XXS | |
| | V786XXS,V787XXS,V789XXS,V7900XS,V7909XS,V7910XS,V7919XS | |
| | V7920XS,V7929XS,V793XXS,V7940XS,V7949XS,V7950XS,V7959XS | |
| | V7960XS,V7969XS,V7981XS,V7988XS,V799XXS,V80010S,V80018S | |
| | V8002XS,V8011XS,V8012XS,V8021XS,V8022XS,V8031XS,V8032XS | |
| | V8041XS,V8042XS,V8051XS,V8052XS,V8061XS,V8062XS,V80710S | |
| | V80711S,V80720S,V80721S,V80730S,V80731S,V80790S,V80791S | |
| | V8081XS,V8082XS,V80910S,V80918S,V80919S,V80920S,V80928S | |
| | V80929S,V810XXS,V811XXS,V812XXS,V813XXS,V814XXS,V815XXS | |
| | V816XXS,V817XXS,V8181XS,V8182XS,V8183XS,V8189XS,V819XXS | |
| | V820XXS,V821XXS,V822XXS,V823XXS,V824XXS,V825XXS,V826XXS | |
| | V827XXS,V828XXS,V829XXS,V830XXS,V831XXS,V832XXS,V833XXS | |
| | V834XXS,V835XXS,V836XXS,V837XXS,V839XXS,V840XXS,V841XXS | |
| | V842XXS,V843XXS,V844XXS,V845XXS,V846XXS,V847XXS,V849XXS | |
| | V850XXS,V851XXS,V852XXS,V853XXS,V854XXS,V855XXS,V856XXS | |
| | V857XXS,V859XXS,V8601XS,V8602XS,V8603XS,V8604XS,V8605XS | |
| | V8606XS,V8609XS,V8611XS,V8612XS,V8613XS,V8614XS,V8615XS | |

Continued on next page

**Supplementary Table S1:** (cont'd) A comprehensive mapping of diagnosis groups used in the experiments to their corresponding ICD-10 and ICD-9 codes.

| Diagnosis | ICD-10 | ICD-9 |
|---|---|---|
| | V8616XS,V8619XS,V8621XS,V8622XS,V8623XS,V8624XS,V8625XS | |
| | V8626XS,V8629XS,V8631XS,V8632XS,V8633XS,V8634XS,V8635XS | |
| | V8636XS,V8639XS,V8641XS,V8642XS,V8643XS,V8644XS,V8645XS | |
| | V8646XS,V8649XS,V8651XS,V8652XS,V8653XS,V8654XS,V8655XS | |
| | V8656XS,V8659XS,V8661XS,V8662XS,V8663XS,V8664XS,V8665XS | |
| | V8666XS,V8669XS,V8671XS,V8672XS,V8673XS,V8674XS,V8675XS | |
| | V8676XS,V8679XS,V8691XS,V8692XS,V8693XS,V8694XS,V8695XS | |
| | V8696XS,V8699XS,V870XXS,V871XXS,V872XXS,V873XXS,V874XXS | |
| | V875XXS,V876XXS,V877XXS,V878XXS,V879XXS,V880XXS,V881XXS | |
| | V882XXS,V883XXS,V884XXS,V885XXS,V886XXS,V887XXS,V888XXS | |
| | V889XXS,V890XXS,V891XXS,V892XXS,V893XXS,V899XXS,V9000XS | |
| | V9001XS,V9002XS,V9003XS,V9004XS,V9005XS,V9006XS,V9008XS | |
| | V9009XS,V9010XS,V9011XS,V9012XS,V9013XS,V9014XS,V9015XS | |
| | V9016XS,V9018XS,V9019XS,V9020XS,V9021XS,V9022XS,V9023XS | |
| | V9024XS,V9025XS,V9026XS,V9027XS,V9028XS,V9029XS,V9030XS | |
| | V9031XS,V9032XS,V9033XS,V9034XS,V9035XS,V9036XS,V9037XS | |
| | V9038XS,V9039XS,V9080XS,V9081XS,V9082XS,V9083XS,V9084XS | |
| | V9085XS,V9086XS,V9087XS,V9088XS,V9089XS,V9100XS,V9101XS | |
| | V9102XS,V9103XS,V9104XS,V9105XS,V9106XS,V9107XS,V9108XS | |
| | V9109XS,V9110XS,V9111XS,V9112XS,V9113XS,V9114XS,V9115XS | |
| | V9116XS,V9118XS,V9119XS,V9120XS,V9121XS,V9122XS,V9123XS | |
| | V9124XS,V9125XS,V9126XS,V9129XS,V9130XS,V9131XS,V9132XS | |
| | V9133XS,V9134XS,V9135XS,V9136XS,V9137XS,V9138XS,V9139XS | |
| | V9180XS,V9181XS,V9182XS,V9183XS,V9184XS,V9185XS,V9186XS | |
| | V9187XS,V9188XS,V9189XS,V9200XS,V9201XS,V9202XS,V9203XS | |
| | V9204XS,V9205XS,V9206XS,V9207XS,V9208XS,V9209XS,V9210XS | |
| | V9211XS,V9212XS,V9213XS,V9214XS,V9215XS,V9216XS,V9219XS | |
| | V9220XS,V9221XS,V9222XS,V9223XS,V9224XS,V9225XS,V9226XS | |
| | V9227XS,V9228XS,V9229XS,V9300XS,V9301XS,V9302XS,V9303XS | |
| | V9304XS,V9309XS,V9310XS,V9311XS,V9312XS,V9313XS,V9314XS | |
| | V9319XS,V9320XS,V9321XS,V9322XS,V9323XS,V9324XS,V9329XS | |
| | V9330XS,V9331XS,V9332XS,V9333XS,V9334XS,V9335XS,V9336XS | |
| | V9338XS,V9339XS,V9340XS,V9341XS,V9342XS,V9343XS,V9344XS | |
| | V9348XS,V9349XS,V9350XS,V9351XS,V9352XS,V9353XS,V9354XS | |
| | V9359XS,V9360XS,V9361XS,V9362XS,V9363XS,V9364XS,V9369XS | |
| | V9380XS,V9381XS,V9382XS,V9383XS,V9384XS,V9385XS,V9386XS | |
| | V9387XS,V9388XS,V9389XS,V940XXS,V9411XS,V9412XS,V9421XS | |
| | V9422XS,V9431XS,V9432XS,V944XXS,V94810S,V94811S,V94818S | |
| | V9489XS,V949XXS,V9500XS,V9501XS,V9502XS,V9503XS,V9504XS | |
| | V9505XS,V9509XS,V9510XS,V9511XS,V9512XS,V9513XS,V9514XS | |

Continued on next page

**Supplementary Table S1:** (cont'd) A comprehensive mapping of diagnosis groups used in the experiments to their corresponding ICD-10 and ICD-9 codes.

| Diagnosis | ICD-10 | ICD-9 |
|---|---|---|
| | V9515XS,V9519XS,V9520XS,V9521XS,V9522XS,V9523XS,V9524XS | |
| | V9525XS,V9529XS,V9530XS,V9531XS,V9532XS,V9533XS,V9534XS | |
| | V9535XS,V9539XS,V9540XS,V9541XS,V9542XS,V9543XS,V9544XS | |
| | V9545XS,V9549XS,V958XXS,V959XXS,V9600XS,V9601XS,V9602XS | |
| | V9603XS,V9604XS,V9605XS,V9609XS,V9610XS,V9611XS,V9612XS | |
| | V9613XS,V9614XS,V9615XS,V9619XS,V9620XS,V9621XS,V9622XS | |
| | V9623XS,V9624XS,V9625XS,V9629XS,V968XXS,V969XXS,V970XXS | |
| | V971XXS,V9721XS,V9722XS,V9729XS,V9731XS,V9732XS,V9733XS | |
| | V9739XS,V97810S,V97811S,V97818S,V9789XS,V980XXS,V981XXS | |
| | V982XXS,V983XXS,V988XXS,V99XXXS,W000XXS,W001XXS,W002XXS | |
| | W009XXS,W010XXS,W0110XS,W01110S,W01111S,W01118S,W01119S | |
| | W01190S,W01198S,W03XXXS,W04XXXS,W050XXS,W051XXS,W052XXS | |
| | W06XXXS,W07XXXS,W08XXXS,W090XXS,W091XXS,W092XXS,W098XXS | |
| | W100XXS,W101XXS,W102XXS,W108XXS,W109XXS,W11XXXS,W12XXXS | |
| | W130XXS,W131XXS,W132XXS,W133XXS,W134XXS,W138XXS,W139XXS | |
| | W14XXXS,W15XXXS,W16011S,W16012S,W16021S,W16022S,W16031S | |
| | W16032S,W16111S,W16112S,W16121S,W16122S,W16131S,W16132S | |
| | W16211S,W16212S,W16221S,W16222S,W16311S,W16312S,W16321S | |
| | W16322S,W16331S,W16332S,W1641XS,W1642XS,W16511S,W16512S | |
| | W16521S,W16522S,W16531S,W16532S,W16611S,W16612S,W16621S | |
| | W16622S,W16711S,W16712S,W16721S,W16722S,W16811S,W16812S | |
| | W16821S,W16822S,W16831S,W16832S,W1691XS,W1692XS,W170XXS | |
| | W171XXS,W172XXS,W173XXS,W174XXS,W1781XS,W1782XS,W1789XS | |
| | W1800XS,W1801XS,W1802XS,W1809XS,W1811XS,W1812XS,W182XXS | |
| | W1830XS,W1831XS,W1839XS,W1840XS,W1841XS,W1842XS,W1843XS | |
| | W1849XS,W19XXXS,W200XXS,W201XXS,W208XXS,W2100XS,W2101XS | |
| | W2102XS,W2103XS,W2104XS,W2105XS,W2106XS,W2107XS,W2109XS | |
| | W2111XS,W2112XS,W2113XS,W2119XS,W21210S,W21211S,W21220S | |
| | W21221S,W2131XS,W2132XS,W2139XS,W214XXS,W2181XS,W2189XS | |
| | W219XXS,W2201XS,W2202XS,W2203XS,W22041S,W22042S,W2209XS | |
| | W2210XS,W2211XS,W2212XS,W2219XS,W228XXS,W230XXS,W231XXS | |
| | W232XXS,W240XXS,W241XXS,W25XXXS,W260XXS,W261XXS,W262XXS | |
| | W268XXS,W269XXS,W270XXS,W271XXS,W272XXS,W273XXS,W274XXS | |
| | W275XXS,W278XXS,W28XXXS,W290XXS,W291XXS,W292XXS,W293XXS | |
| | W294XXS,W298XXS,W300XXS,W301XXS,W302XXS,W303XXS,W3081XS | |
| | W3089XS,W309XXS,W310XXS,W311XXS,W312XXS,W313XXS,W3181XS | |
| | W3182XS,W3183XS,W3189XS,W319XXS,W320XXS,W321XXS,W3300XS | |
| | W3301XS,W3302XS,W3303XS,W3309XS,W3310XS,W3311XS,W3312XS | |
| | W3313XS,W3319XS,W3400XS,W34010S,W34011S,W34018S,W3409XS | |
| | W3410XS,W34110S,W34111S,W34118S,W3419XS,W35XXXS,W361XXS | |

Continued on next page

**Supplementary Table S1:** (cont'd) A comprehensive mapping of diagnosis groups used in the experiments to their corresponding ICD-10 and ICD-9 codes.

| Diagnosis | ICD-10 | ICD-9 |
|---|---|---|
| | W362XXS,W363XXS,W368XXS,W369XXS,W370XXS,W378XXS,W38XXXS | |
| | W39XXXS,W400XXS,W401XXS,W408XXS,W409XXS,W420XXS,W429XXS | |
| | W450XXS,W451XXS,W452XXS,W458XXS,W460XXS,W461XXS,W4901XS | |
| | W4902XS,W4903XS,W4904XS,W4909XS,W499XXS,W500XXS,W501XXS | |
| | W502XXS,W503XXS,W504XXS,W51XXXS,W52XXXS,W5301XS,W5309XS | |
| | W5311XS,W5319XS,W5321XS,W5329XS,W5381XS,W5389XS,W540XXS | |
| | W541XXS,W548XXS,W5501XS,W5503XS,W5509XS,W5511XS,W5512XS | |
| | W5519XS,W5521XS,W5522XS,W5529XS,W5531XS,W5532XS,W5539XS | |
| | W5541XS,W5542XS,W5549XS,W5551XS,W5552XS,W5559XS,W5581XS | |
| | W5582XS,W5589XS,W5601XS,W5602XS,W5609XS,W5611XS,W5612XS | |
| | W5619XS,W5621XS,W5622XS,W5629XS,W5631XS,W5632XS,W5639XS | |
| | W5641XS,W5642XS,W5649XS,W5651XS,W5652XS,W5659XS,W5681XS | |
| | W5682XS,W5689XS,W57XXXS,W5801XS,W5802XS,W5803XS,W5809XS | |
| | W5811XS,W5812XS,W5813XS,W5819XS,W5901XS,W5902XS,W5909XS | |
| | W5911XS,W5912XS,W5913XS,W5919XS,W5921XS,W5922XS,W5929XS | |
| | W5981XS,W5982XS,W5983XS,W5989XS,W60XXXS,W6101XS,W6102XS | |
| | W6109XS,W6111XS,W6112XS,W6119XS,W6121XS,W6122XS,W6129XS | |
| | W6132XS,W6133XS,W6139XS,W6142XS,W6143XS,W6149XS,W6151XS | |
| | W6152XS,W6159XS,W6161XS,W6162XS,W6169XS,W6191XS,W6192XS | |
| | W6199XS,W620XXS,W621XXS,W629XXS,W64XXXS,W65XXXS,W67XXXS | |
| | W69XXXS,W73XXXS,W74XXXS,W85XXXS,W860XXS,W861XXS,W868XXS | |
| | W880XXS,W881XXS,W888XXS,W890XXS,W891XXS,W898XXS,W899XXS | |
| | W900XXS,W901XXS,W902XXS,W908XXS,W92XXXS,W9301XS,W9302XS | |
| | W9311XS,W9312XS,W932XXS,W938XXS,W940XXS,W9411XS,W9412XS | |
| | W9421XS,W9422XS,W9423XS,W9429XS,W9431XS,W9432XS,W9439XS | |
| | W99XXXS,X000XXS,X001XXS,X002XXS,X003XXS,X004XXS,X005XXS | |
| | X008XXS,X010XXS,X011XXS,X013XXS,X014XXS,X018XXS,X020XXS | |
| | X021XXS,X022XXS,X023XXS,X024XXS,X025XXS,X028XXS,X030XXS | |
| | X031XXS,X033XXS,X034XXS,X038XXS,X04XXXS,X05XXXS,X060XXS | |
| | X061XXS,X062XXS,X063XXS,X0800XS,X0801XS,X0809XS,X0810XS | |
| | X0811XS,X0819XS,X0820XS,X0821XS,X0829XS,X088XXS,X100XXS | |
| | X101XXS,X102XXS,X110XXS,X111XXS,X118XXS,X12XXXS,X130XXS | |
| | X131XXS,X140XXS,X141XXS,X150XXS,X151XXS,X152XXS,X153XXS | |
| | X158XXS,X16XXXS,X17XXXS,X18XXXS,X19XXXS,X30XXXS,X31XXXS | |
| | X32XXXS,X34XXXS,X35XXXS,X360XXS,X361XXS,X370XXS,X371XXS | |
| | X372XXS,X373XXS,X3741XS,X3742XS,X3743XS,X378XXS,X379XXS | |
| | X38XXXS,X3901XS,X3908XS,X398XXS,X500XXS,X501XXS,X503XXS | |
| | X509XXS,X52XXXS,X58XXXS,X710XXS,X711XXS,X712XXS,X713XXS | |
| | X718XXS,X719XXS,X72XXXS,X730XXS,X731XXS,X732XXS,X738XXS | |
| | X739XXS,X7401XS,X7402XS,X7409XS,X748XXS,X749XXS,X75XXXS | |

Continued on next page

**Supplementary Table S1:** (cont'd) A comprehensive mapping of diagnosis groups used in the experiments to their corresponding ICD-10 and ICD-9 codes.

| Diagnosis | ICD-10 | ICD-9 |
|---|---|---|
| | X76XXXS,X770XXS,X771XXS,X772XXS,X773XXS,X778XXS,X779XXS | |
| | X780XXS,X781XXS,X782XXS,X788XXS,X789XXS,X79XXXS,X80XXXS | |
| | X810XXS,X811XXS,X818XXS,X820XXS,X821XXS,X822XXS,X828XXS | |
| | X830XXS,X831XXS,X832XXS,X838XXS,X920XXS,X921XXS,X922XXS | |
| | X923XXS,X928XXS,X929XXS,X93XXXS,X940XXS,X941XXS,X942XXS | |
| | X948XXS,X949XXS,X9501XS,X9502XS,X9509XS,X958XXS,X959XXS | |
| | X960XXS,X961XXS,X962XXS,X963XXS,X964XXS,X968XXS,X969XXS | |
| | X97XXXS,X980XXS,X981XXS,X982XXS,X983XXS,X988XXS,X989XXS | |
| | X990XXS,X991XXS,X992XXS,X998XXS,X999XXS,Y00XXXS,Y01XXXS | |
| | Y020XXS,Y021XXS,Y028XXS,Y030XXS,Y038XXS,Y040XXS,Y041XXS | |
| | Y042XXS,Y048XXS,Y0801XS,Y0802XS,Y0809XS,Y0881XS,Y0889XS | |
| | Y210XXS,Y211XXS,Y212XXS,Y213XXS,Y214XXS,Y218XXS,Y219XXS | |
| | Y22XXXS,Y230XXS,Y231XXS,Y232XXS,Y233XXS,Y238XXS,Y239XXS | |
| | Y240XXS,Y248XXS,Y249XXS,Y25XXXS,Y26XXXS,Y270XXS,Y271XXS | |
| | Y272XXS,Y273XXS,Y278XXS,Y279XXS,Y280XXS,Y281XXS,Y282XXS | |
| | Y288XXS,Y289XXS,Y29XXXS,Y30XXXS,Y31XXXS,Y32XXXS,Y33XXXS | |
| | Y35001S,Y35002S,Y35003S,Y35009S,Y35011S,Y35012S,Y35013S | |
| | Y35019S,Y35021S,Y35022S,Y35023S,Y35029S,Y35031S,Y35032S | |
| | Y35033S,Y35039S,Y35041S,Y35042S,Y35043S,Y35049S,Y35091S | |
| | Y35092S,Y35093S,Y35099S,Y35101S,Y35102S,Y35103S,Y35109S | |
| | Y35111S,Y35112S,Y35113S,Y35119S,Y35121S,Y35122S,Y35123S | |
| | Y35129S,Y35191S,Y35192S,Y35193S,Y35199S,Y35201S,Y35202S | |
| | Y35203S,Y35209S,Y35211S,Y35212S,Y35213S,Y35219S,Y35291S | |
| | Y35292S,Y35293S,Y35299S,Y35301S,Y35302S,Y35303S,Y35309S | |
| | Y35311S,Y35312S,Y35313S,Y35319S,Y35391S,Y35392S,Y35393S | |
| | Y35399S,Y35401S,Y35402S,Y35403S,Y35409S,Y35411S,Y35412S | |
| | Y35413S,Y35419S,Y35491S,Y35492S,Y35493S,Y35499S,Y35811S | |
| | Y35812S,Y35813S,Y35819S,Y35831S,Y35832S,Y35833S,Y35839S | |
| | Y35891S,Y35892S,Y35893S,Y35899S,Y3591XS,Y3592XS,Y3593XS | |
| | Y3599XS,Y36000S,Y36001S,Y36010S,Y36011S,Y36020S,Y36021S | |
| | Y36030S,Y36031S,Y36040S,Y36041S,Y36050S,Y36051S,Y36090S | |
| | Y36091S,Y36100S,Y36101S,Y36110S,Y36111S,Y36120S,Y36121S | |
| | Y36130S,Y36131S,Y36140S,Y36141S,Y36190S,Y36191S,Y36200S | |
| | Y36201S,Y36210S,Y36211S,Y36220S,Y36221S,Y36230S,Y36231S | |
| | Y36240S,Y36241S,Y36250S,Y36251S,Y36260S,Y36261S,Y36270S | |
| | Y36271S,Y36290S,Y36291S,Y36300S,Y36301S,Y36310S,Y36311S | |
| | Y36320S,Y36321S,Y36330S,Y36331S,Y36390S,Y36391S,Y36410S | |
| | Y36411S,Y36420S,Y36421S,Y36430S,Y36431S,Y36440S,Y36441S | |
| | Y36450S,Y36451S,Y36460S,Y36461S,Y36470S,Y36471S,Y36490S | |
| | Y36491S,Y36500S,Y36501S,Y36510S,Y36511S,Y36520S,Y36521S | |

Continued on next page

**Supplementary Table S1:** (cont'd) A comprehensive mapping of diagnosis groups used in the experiments to their corresponding ICD-10 and ICD-9 codes.

| Diagnosis | ICD-10 | ICD-9 |
|---|---|---|
| | Y36530S,Y36531S,Y36540S,Y36541S,Y36590S,Y36591S,Y366X0S | |
| | Y366X1S,Y367X0S,Y367X1S,Y36810S,Y36811S,Y36820S,Y36821S | |
| | Y36880S,Y36881S,Y36890S,Y36891S,Y3690XS,Y3691XS,Y3692XS | |
| | Y37000S,Y37001S,Y37010S,Y37011S,Y37020S,Y37021S,Y37030S | |
| | Y37031S,Y37040S,Y37041S,Y37050S,Y37051S,Y37090S,Y37091S | |
| | Y37100S,Y37101S,Y37110S,Y37111S,Y37120S,Y37121S,Y37130S | |
| | Y37131S,Y37140S,Y37141S,Y37190S,Y37191S,Y37200S,Y37201S | |
| | Y37210S,Y37211S,Y37220S,Y37221S,Y37230S,Y37231S,Y37240S | |
| | Y37241S,Y37250S,Y37251S,Y37260S,Y37261S,Y37270S,Y37271S | |
| | Y37290S,Y37291S,Y37300S,Y37301S,Y37310S,Y37311S,Y37320S | |
| | Y37321S,Y37330S,Y37331S,Y37390S,Y37391S,Y37410S,Y37411S | |
| | Y37420S,Y37421S,Y37430S,Y37431S,Y37440S,Y37441S,Y37450S | |
| | Y37451S,Y37460S,Y37461S,Y37470S,Y37471S,Y37490S,Y37491S | |
| | Y37500S,Y37501S,Y37510S,Y37511S,Y37520S,Y37521S,Y37530S | |
| | Y37531S,Y37540S,Y37541S,Y37590S,Y37591S,Y376X0S,Y376X1S | |
| | Y377X0S,Y377X1S,Y3790XS,Y3791XS,Y3792XS,Y380X1S,Y380X2S | |
| | Y380X3S,Y381X1S,Y381X2S,Y381X3S,Y382X1S,Y382X2S,Y382X3S | |
| | Y383X1S,Y383X2S,Y383X3S,Y384X1S,Y384X2S,Y384X3S,Y385X1S | |
| | Y385X2S,Y385X3S,Y386X1S,Y386X2S,Y386X3S,Y387X1S,Y387X2S | |
| | Y387X3S,Y3880XS,Y38811S,Y38812S,Y38891S,Y38892S,Y38893S | |
| | Y389X1S,Y389X2S | |
| External cause codes: | V0001XA,V0002XA,V00031A,V00038A,V0009XA,V00112A,V00122A | E8843,E8845,E8849,E8850,E8851,E8852,E8853,E8854,E8859,E8880 |
| struck by; against; | V00132A,V00142A,V00152A,V00182A,V00212A,V00222A,V00282A | E8881,E916,E9170,E9171,E9173,E9174,E9178,E9179,E9283,E9588 |
| initial encounter | V00312A,V00322A,V00382A,V00812A,V00822A,V00832A,V00842A | E9600,E9682,E9687,E973,E975,E9888,E9950 |
| | V00892A,W1800XA,W1801XA,W1802XA,W1809XA,W200XXA,W201XXA | |
| | W208XXA,W2100XA,W2101XA,W2102XA,W2103XA,W2104XA,W2105XA | |
| | W2106XA,W2107XA,W2109XA,W2111XA,W2112XA,W2113XA,W2119XA | |
| | W21210A,W21211A,W21220A,W21221A,W2131XA,W2132XA,W2139XA | |
| | W214XXA,W2181XA,W2189XA,W219XXA,W2201XA,W2202XA,W2203XA | |
| | W22042A,W2209XA,W2210XA,W2211XA,W2212XA,W2219XA,W228XXA | |
| | W500XXA,W501XXA,W502XXA,W503XXA,W504XXA,W51XXXA,W52XXXA | |
| | X79XXXA,Y00XXXA,Y040XXA,Y041XXA,Y042XXA,Y048XXA,Y0801XA | |
| | Y0802XA,Y0809XA,Y29XXXA,Y35301A,Y35302A,Y35303A,Y35309A | |
| | Y35311A,Y35312A,Y35313A,Y35319A,Y35391A,Y35392A,Y35393A | |
| | Y35399A,Y35811A,Y35812A,Y35813A,Y35819A,Y36440A,Y36441A | |
| | Y37440A,Y37441A | |
| External cause codes: | V0001XD,V0002XD,V00031D,V00038D,V0009XD,V00111D,V00112D | E8001,E8011,E8013,E8018,E8019,E8021,E8031,E8041,E8052,E8061 |
| subsequent encounter | V00118D,V00121D,V00122D,V00128D,V00131D,V00132D,V00138D | E8071,E8072,E8100,E8101,E8102,E8103,E8107,E8108,E8109,E8120 |

Continued on next page

**Supplementary Table S1:** (cont'd) A comprehensive mapping of diagnosis groups used in the experiments to their corresponding ICD-10 and ICD-9 codes.

| Diagnosis | ICD-10 | ICD-9 |
|---|---|---|
| | V00141D,V00142D,V00148D,V00151D,V00152D,V00158D,V00181D | E8121,E8122,E8123,E8128,E8129,E8130,E8131,E8132,E8133,E8134 |
| | V00182D,V00188D,V00211D,V00212D,V00218D,V00221D,V00222D | E8135,E8136,E8138,E8139,E8140,E8141,E8142,E8143,E8147,E8148 |
| | V00228D,V00281D,V00282D,V00288D,V00311D,V00312D,V00318D | E8149,E8150,E8151,E8152,E8153,E8158,E8159,E8160,E8161,E8162 |
| | V00321D,V00322D,V00328D,V00381D,V00382D,V00388D,V00811D | E8163,E8168,E8169,E8178,E8188,E8189,E8190,E8192,E8199,E8200 |
| | V00812D,V00818D,V00821D,V00822D,V00828D,V00831D,V00832D | E8201,E8208,E8209,E8210,E8211,E8218,E8219,E8220,E8221,E8222 |
| | V00838D,V00841D,V00842D,V00848D,V00891D,V00892D,V00898D | E8223,E8224,E8226,E8227,E8228,E8229,E8230,E8231,E8232,E8233 |
| | V0100XD,V0101XD,V0102XD,V01031D,V01038D,V0109XD,V0110XD | E8238,E8239,E8242,E8246,E8248,E8250,E8251,E8252,E8253,E8258 |
| | V0111XD,V0112XD,V01131D,V01138D,V0119XD,V0190XD,V0191XD | E8259,E8260,E8261,E8262,E8263,E8269,E8272,E8273,E8282,E8290 |
| | V0192XD,V01931D,V01938D,V0199XD,V0200XD,V0201XD,V0202XD | E8294,E8299,E8309,E8319,E8329,E8359,E8369,E8379,E8380,E8381 |
| | V02031D,V02038D,V0209XD,V0210XD,V0211XD,V0212XD,V02131D | E8384,E8385,E8389,E8403,E8406,E8411,E8413,E8426,E8433,E8437 |
| | V02138D,V0219XD,V0290XD,V0291XD,V0292XD,V02931D,V02938D | E8439,E8441,E8443,E8447,E8449,E8450,E846,E847,E848,E8800 |
| | V0299XD,V0300XD,V0301XD,V0302XD,V03031D,V03038D,V0309XD | E8801,E8809,E8810,E8811,E882,E8830,E8831,E8832,E8839,E8840 |
| | V0310XD,V0311XD,V0312XD,V03131D,V03138D,V0319XD,V0390XD | E8841,E8842,E8843,E8844,E8845,E8846,E8849,E8850,E8851,E8852 |
| | V0391XD,V0392XD,V03931D,V03938D,V0399XD,V0400XD,V0401XD | E8853,E8854,E8859,E8860,E8869,E8880,E8881,E8888,E8889,E8902 |
| | V0402XD,V04031D,V04038D,V0409XD,V0410XD,V0411XD,V0412XD | E8903,E8908,E892,E8930,E8938,E894,E897,E8980,E8981,E9000 |
| | V04131D,V04138D,V0419XD,V0490XD,V0491XD,V0492XD,V04931D | E9001,E9010,E9011,E9018,E9020,E9021,E9022,E9028,E9060,E9061 |
| | V04938D,V0499XD,V0500XD,V0501XD,V0502XD,V05031D,V05038D | E9062,E9063,E9064,E9068,E9069,E9080,E9081,E9082,E9083,E9084 |
| | V0509XD,V0510XD,V0511XD,V0512XD,V05131D,V05138D,V0519XD | E9088,E9089,E9090,E9091,E9092,E9093,E9094,E9098,E9102,E9104 |
| | V0590XD,V0591XD,V0592XD,V05931D,V05938D,V0599XD,V0600XD | E9108,E9109,E916,E9170,E9171,E9173,E9174,E9179,E918,E9190 |
| | V0601XD,V0602XD,V06031D,V06038D,V0609XD,V0610XD,V0611XD | E9191,E9192,E9193,E9194,E9195,E9196,E9197,E9198,E9199,E9200 |
| | V0612XD,V06131D,V06138D,V0619XD,V0690XD,V0691XD,V0692XD | E9201,E9202,E9203,E9204,E9205,E9208,E9210,E9211,E9218,E9220 |
| | V06931D,V06938D,V0699XD,V0900XD,V0901XD,V0909XD,V091XXD | E9221,E9222,E9223,E9224,E9225,E9228,E9229,E9230,E9231,E9232 |
| | V0920XD,V0921XD,V0929XD,V093XXD,V099XXD,V100XXD,V101XXD | E9238,E9239,E9240,E9242,E9248,E9250,E9251,E9252,E9258,E9260 |
| | V102XXD,V103XXD,V104XXD,V105XXD,V109XXD,V110XXD,V111XXD | E9261,E9262,E9263,E9264,E9265,E9268,E9280,E9281,E9283,E9284 |
| | V112XXD,V113XXD,V114XXD,V115XXD,V119XXD,V120XXD,V121XXD | E9285,E9288,E954,E9550,E9551,E9552,E9553,E9554,E9555,E9556 |
| | V122XXD,V123XXD,V124XXD,V125XXD,V129XXD,V130XXD,V131XXD | E9557,E9559,E956,E9579,E9580,E9581,E9582,E9583,E9584,E9585 |
| | V132XXD,V133XXD,V134XXD,V135XXD,V139XXD,V140XXD,V141XXD | E9586,E9587,E9588,E9600,E964,E9650,E9651,E9652,E9653,E9654 |
| | V142XXD,V143XXD,V144XXD,V145XXD,V149XXD,V150XXD,V151XXD | E9655,E9656,E9657,E9658,E9659,E966,E9680,E9681,E9682,E9683 |
| | V152XXD,V153XXD,V154XXD,V155XXD,V159XXD,V160XXD,V161XXD | E9685,E9686,E9687,E9688,E970,E971,E972,E973,E974,E975,E976 |
| | V162XXD,V163XXD,V164XXD,V165XXD,V169XXD,V170XXD,V171XXD | E9790,E9791,E9792,E9793,E9794,E9795,E9796,E9797,E9798,E9799 |
| | V172XXD,V173XXD,V174XXD,V175XXD,V179XXD,V180XXD,V181XXD | E984,E9850,E9851,E9852,E9853,E9854,E9855,E9856,E9857,E986 |
| | V182XXD,V183XXD,V184XXD,V185XXD,V189XXD,V1900XD,V1909XD | E9879,E9880,E9881,E9882,E9885,E9888,E9900,E9901,E9902,E9903 |
| | V1910XD,V1919XD,V1920XD,V1929XD,V193XXD,V1940XD,V1949XD | E9909,E9910,E9911,E9912,E9913,E9914,E9915,E9916,E9917,E9918 |
| | V1950XD,V1959XD,V1960XD,V1969XD,V1981XD,V1988XD,V199XXD | E9919,E9920,E9921,E9922,E9923,E9928,E9929,E9930,E9931,E9932 |
| | V2001XD,V2009XD,V200XXD,V2011XD,V2019XD,V201XXD,V2021XD | E9933,E9934,E9935,E9936,E9937,E9938,E9939,E9940,E9941,E9942 |
| | V2029XD,V202XXD,V2031XD,V2039XD,V203XXD,V2041XD,V2049XD | E9943,E9948,E9949,E9950,E9951,E9952,E9953,E9954,E9958,E9959 |
| | V204XXD,V2051XD,V2059XD,V205XXD,V2091XD,V2099XD,V209XXD | E9960,E9961,E9962,E9963,E9968,E9969,E9970,E9971,E9972,E9973 |
| | V2101XD,V2109XD,V210XXD,V2111XD,V2119XD,V211XXD,V2121XD | E9978,E9979,E9980,E9981,E9988,E9989 |
| | V2129XD,V212XXD,V2131XD,V2139XD,V213XXD,V2141XD,V2149XD | |
| | V214XXD,V2151XD,V2159XD,V215XXD,V2191XD,V2199XD,V219XXD | |

Continued on next page

**Supplementary Table S1:** (cont'd) A comprehensive mapping of diagnosis groups used in the experiments to their corresponding ICD-10 and ICD-9 codes.

| Diagnosis | ICD-10 | ICD-9 |
|---|---|---|
| | V2201XD,V2209XD,V220XXD,V2211XD,V2219XD,V221XXD,V2221XD | |
| | V2229XD,V222XXD,V2231XD,V2239XD,V223XXD,V2241XD,V2249XD | |
| | V224XXD,V2251XD,V2259XD,V225XXD,V2291XD,V2299XD,V229XXD | |
| | V2301XD,V2309XD,V230XXD,V2311XD,V2319XD,V231XXD,V2321XD | |
| | V2329XD,V232XXD,V2331XD,V2339XD,V233XXD,V2341XD,V2349XD | |
| | V234XXD,V2351XD,V2359XD,V235XXD,V2391XD,V2399XD,V239XXD | |
| | V2401XD,V2409XD,V240XXD,V2411XD,V2419XD,V241XXD,V2421XD | |
| | V2429XD,V242XXD,V2431XD,V2439XD,V243XXD,V2441XD,V2449XD | |
| | V244XXD,V2451XD,V2459XD,V245XXD,V2491XD,V2499XD,V249XXD | |
| | V2501XD,V2509XD,V250XXD,V2511XD,V2519XD,V251XXD,V2521XD | |
| | V2529XD,V252XXD,V2531XD,V2539XD,V253XXD,V2541XD,V2549XD | |
| | V254XXD,V2551XD,V2559XD,V255XXD,V2591XD,V2599XD,V259XXD | |
| | V2601XD,V2609XD,V260XXD,V2611XD,V2619XD,V261XXD,V2621XD | |
| | V2629XD,V262XXD,V2631XD,V2639XD,V263XXD,V2641XD,V2649XD | |
| | V264XXD,V2651XD,V2659XD,V265XXD,V2691XD,V2699XD,V269XXD | |
| | V2701XD,V2709XD,V270XXD,V2711XD,V2719XD,V271XXD,V2721XD | |
| | V2729XD,V272XXD,V2731XD,V2739XD,V273XXD,V2741XD,V2749XD | |
| | V274XXD,V2751XD,V2759XD,V275XXD,V2791XD,V2799XD,V279XXD | |
| | V2801XD,V2809XD,V280XXD,V2811XD,V2819XD,V281XXD,V2821XD | |
| | V2829XD,V282XXD,V2831XD,V2839XD,V283XXD,V2841XD,V2849XD | |
| | V284XXD,V2851XD,V2859XD,V285XXD,V2891XD,V2899XD,V289XXD | |
| | V29001D,V29008D,V2900XD,V29091D,V29098D,V2909XD,V29101D | |
| | V29108D,V2910XD,V29191D,V29198D,V2919XD,V29201D,V29208D | |
| | V2920XD,V29291D,V29298D,V2929XD,V2931XD,V2939XD,V293XXD | |
| | V29401D,V29408D,V2940XD,V29491D,V29498D,V2949XD,V29501D | |
| | V29508D,V2950XD,V29591D,V29598D,V2959XD,V29601D,V29608D | |
| | V2960XD,V29691D,V29698D,V2969XD,V29811D,V29818D,V2981XD | |
| | V29881D,V29888D,V2988XD,V2991XD,V2999XD,V299XXD,V300XXD | |
| | V301XXD,V302XXD,V303XXD,V304XXD,V305XXD,V306XXD,V307XXD | |
| | V309XXD,V310XXD,V311XXD,V312XXD,V313XXD,V314XXD,V315XXD | |
| | V316XXD,V317XXD,V319XXD,V320XXD,V321XXD,V322XXD,V323XXD | |
| | V324XXD,V325XXD,V326XXD,V327XXD,V329XXD,V330XXD,V331XXD | |
| | V332XXD,V333XXD,V334XXD,V335XXD,V336XXD,V337XXD,V339XXD | |
| | V340XXD,V341XXD,V342XXD,V343XXD,V344XXD,V345XXD,V346XXD | |
| | V347XXD,V349XXD,V350XXD,V351XXD,V352XXD,V353XXD,V354XXD | |
| | V355XXD,V356XXD,V357XXD,V359XXD,V360XXD,V361XXD,V362XXD | |
| | V363XXD,V364XXD,V365XXD,V366XXD,V367XXD,V369XXD,V370XXD | |
| | V371XXD,V372XXD,V373XXD,V374XXD,V375XXD,V376XXD,V377XXD | |
| | V379XXD,V380XXD,V381XXD,V382XXD,V383XXD,V384XXD,V385XXD | |
| | V386XXD,V387XXD,V389XXD,V3900XD,V3909XD,V3910XD,V3919XD | |

Continued on next page

**Supplementary Table S1:** (cont'd) A comprehensive mapping of diagnosis groups used in the experiments to their corresponding ICD-10 and ICD-9 codes.

| Diagnosis | ICD-10 | ICD-9 |
|---|---|---|
| | V3920XD,V3929XD,V393XXD,V3940XD,V3949XD,V3950XD,V3959XD | |
| | V3960XD,V3969XD,V3981XD,V3989XD,V399XXD,V400XXD,V401XXD | |
| | V402XXD,V403XXD,V404XXD,V405XXD,V406XXD,V407XXD,V409XXD | |
| | V410XXD,V411XXD,V412XXD,V413XXD,V414XXD,V415XXD,V416XXD | |
| | V417XXD,V419XXD,V420XXD,V421XXD,V422XXD,V423XXD,V424XXD | |
| | V425XXD,V426XXD,V427XXD,V429XXD,V4301XD,V4302XD,V4303XD | |
| | V4304XD,V4311XD,V4312XD,V4313XD,V4314XD,V4321XD,V4322XD | |
| | V4323XD,V4324XD,V4331XD,V4332XD,V4333XD,V4334XD,V4341XD | |
| | V4342XD,V4343XD,V4344XD,V4351XD,V4352XD,V4353XD,V4354XD | |
| | V4361XD,V4362XD,V4363XD,V4364XD,V4371XD,V4372XD,V4373XD | |
| | V4374XD,V4391XD,V4392XD,V4393XD,V4394XD,V440XXD,V441XXD | |
| | V442XXD,V443XXD,V444XXD,V445XXD,V446XXD,V447XXD,V449XXD | |
| | V450XXD,V451XXD,V452XXD,V453XXD,V454XXD,V455XXD,V456XXD | |
| | V457XXD,V459XXD,V460XXD,V461XXD,V462XXD,V463XXD,V464XXD | |
| | V465XXD,V466XXD,V467XXD,V469XXD,V4701XD,V4702XD,V470XXD | |
| | V4711XD,V4712XD,V471XXD,V472XXD,V4731XD,V4732XD,V473XXD | |
| | V474XXD,V4751XD,V4752XD,V475XXD,V4761XD,V4762XD,V476XXD | |
| | V477XXD,V4791XD,V4792XD,V479XXD,V480XXD,V481XXD,V482XXD | |
| | V483XXD,V484XXD,V485XXD,V486XXD,V487XXD,V489XXD,V4900XD | |
| | V4909XD,V4910XD,V4919XD,V4920XD,V4929XD,V493XXD,V4940XD | |
| | V4949XD,V4950XD,V4959XD,V4960XD,V4969XD,V4981XD,V4988XD | |
| | V499XXD,V500XXD,V501XXD,V502XXD,V503XXD,V504XXD,V505XXD | |
| | V506XXD,V507XXD,V509XXD,V510XXD,V511XXD,V512XXD,V513XXD | |
| | V514XXD,V515XXD,V516XXD,V517XXD,V519XXD,V520XXD,V521XXD | |
| | V522XXD,V523XXD,V524XXD,V525XXD,V526XXD,V527XXD,V529XXD | |
| | V530XXD,V531XXD,V532XXD,V533XXD,V534XXD,V535XXD,V536XXD | |
| | V537XXD,V539XXD,V540XXD,V541XXD,V542XXD,V543XXD,V544XXD | |
| | V545XXD,V546XXD,V547XXD,V549XXD,V550XXD,V551XXD,V552XXD | |
| | V553XXD,V554XXD,V555XXD,V556XXD,V557XXD,V559XXD,V560XXD | |
| | V561XXD,V562XXD,V563XXD,V564XXD,V565XXD,V566XXD,V567XXD | |
| | V569XXD,V570XXD,V571XXD,V572XXD,V573XXD,V574XXD,V575XXD | |
| | V576XXD,V577XXD,V579XXD,V580XXD,V581XXD,V582XXD,V583XXD | |
| | V584XXD,V585XXD,V586XXD,V587XXD,V589XXD,V5900XD,V5909XD | |
| | V5910XD,V5919XD,V5920XD,V5929XD,V593XXD,V5940XD,V5949XD | |
| | V5950XD,V5959XD,V5960XD,V5969XD,V5981XD,V5988XD,V599XXD | |
| | V600XXD,V601XXD,V602XXD,V603XXD,V604XXD,V605XXD,V606XXD | |
| | V607XXD,V609XXD,V610XXD,V611XXD,V612XXD,V613XXD,V614XXD | |
| | V615XXD,V616XXD,V617XXD,V619XXD,V620XXD,V621XXD,V622XXD | |
| | V623XXD,V624XXD,V625XXD,V626XXD,V627XXD,V629XXD,V630XXD | |
| | V631XXD,V632XXD,V633XXD,V634XXD,V635XXD,V636XXD,V637XXD | |

Continued on next page

Supplementary Table S1: (cont'd) A comprehensive mapping of diagnosis groups used in the experiments to their corresponding ICD-10 and ICD-9 codes.

| Diagnosis | ICD-10 | ICD-9 |
|---|---|---|
| | V639XXD,V640XXD,V641XXD,V642XXD,V643XXD,V644XXD,V645XXD | |
| | V646XXD,V647XXD,V649XXD,V650XXD,V651XXD,V652XXD,V653XXD | |
| | V654XXD,V655XXD,V656XXD,V657XXD,V659XXD,V660XXD,V661XXD | |
| | V662XXD,V663XXD,V664XXD,V665XXD,V666XXD,V667XXD,V669XXD | |
| | V670XXD,V671XXD,V672XXD,V673XXD,V674XXD,V675XXD,V676XXD | |
| | V677XXD,V679XXD,V680XXD,V681XXD,V682XXD,V683XXD,V684XXD | |
| | V685XXD,V686XXD,V687XXD,V689XXD,V6900XD,V6909XD,V6910XD | |
| | V6919XD,V6920XD,V6929XD,V693XXD,V6940XD,V6949XD,V6950XD | |
| | V6959XD,V6960XD,V6969XD,V6981XD,V6988XD,V699XXD,V700XXD | |
| | V701XXD,V702XXD,V703XXD,V704XXD,V705XXD,V706XXD,V707XXD | |
| | V709XXD,V710XXD,V711XXD,V712XXD,V713XXD,V714XXD,V715XXD | |
| | V716XXD,V717XXD,V719XXD,V720XXD,V721XXD,V722XXD,V723XXD | |
| | V724XXD,V725XXD,V726XXD,V727XXD,V729XXD,V730XXD,V731XXD | |
| | V732XXD,V733XXD,V734XXD,V735XXD,V736XXD,V737XXD,V739XXD | |
| | V740XXD,V741XXD,V742XXD,V743XXD,V744XXD,V745XXD,V746XXD | |
| | V747XXD,V749XXD,V750XXD,V751XXD,V752XXD,V753XXD,V754XXD | |
| | V755XXD,V756XXD,V757XXD,V759XXD,V760XXD,V761XXD,V762XXD | |
| | V763XXD,V764XXD,V765XXD,V766XXD,V767XXD,V769XXD,V770XXD | |
| | V771XXD,V772XXD,V773XXD,V774XXD,V775XXD,V776XXD,V777XXD | |
| | V779XXD,V780XXD,V781XXD,V782XXD,V783XXD,V784XXD,V785XXD | |
| | V786XXD,V787XXD,V789XXD,V7900XD,V7909XD,V7910XD,V7919XD | |
| | V7920XD,V7929XD,V793XXD,V7940XD,V7949XD,V7950XD,V7959XD | |
| | V7960XD,V7969XD,V7981XD,V7988XD,V799XXD,V80010D,V80018D | |
| | V8002XD,V8011XD,V8012XD,V8021XD,V8022XD,V8031XD,V8032XD | |
| | V8041XD,V8042XD,V8051XD,V8052XD,V8061XD,V8062XD,V80710D | |
| | V80711D,V80720D,V80721D,V80730D,V80731D,V80790D,V80791D | |
| | V8081XD,V8082XD,V80910D,V80918D,V80919D,V80920D,V80928D | |
| | V80929D,V810XXD,V811XXD,V812XXD,V813XXD,V814XXD,V815XXD | |
| | V816XXD,V817XXD,V8181XD,V8182XD,V8183XD,V8189XD,V819XXD | |
| | V820XXD,V821XXD,V822XXD,V823XXD,V824XXD,V825XXD,V826XXD | |
| | V827XXD,V828XXD,V829XXD,V830XXD,V831XXD,V832XXD,V833XXD | |
| | V834XXD,V835XXD,V836XXD,V837XXD,V839XXD,V840XXD,V841XXD | |
| | V842XXD,V843XXD,V844XXD,V845XXD,V846XXD,V847XXD,V849XXD | |
| | V850XXD,V851XXD,V852XXD,V853XXD,V854XXD,V855XXD,V856XXD | |
| | V857XXD,V859XXD,V8601XD,V8602XD,V8603XD,V8604XD,V8605XD | |
| | V8606XD,V8609XD,V8611XD,V8612XD,V8613XD,V8614XD,V8615XD | |
| | V8616XD,V8619XD,V8621XD,V8622XD,V8623XD,V8624XD,V8625XD | |
| | V8626XD,V8629XD,V8631XD,V8632XD,V8633XD,V8634XD,V8635XD | |
| | V8636XD,V8639XD,V8641XD,V8642XD,V8643XD,V8644XD,V8645XD | |
| | V8646XD,V8649XD,V8651XD,V8652XD,V8653XD,V8654XD,V8655XD | |

Continued on next page

**Supplementary Table S1:** (cont'd) A comprehensive mapping of diagnosis groups used in the experiments to their corresponding ICD-10 and ICD-9 codes.

| Diagnosis | ICD-10 | ICD-9 |
|---|---|---|
| | V8656XD,V8659XD,V8661XD,V8662XD,V8663XD,V8664XD,V8665XD | |
| | V8666XD,V8669XD,V8671XD,V8672XD,V8673XD,V8674XD,V8675XD | |
| | V8676XD,V8679XD,V8691XD,V8692XD,V8693XD,V8694XD,V8695XD | |
| | V8696XD,V8699XD,V870XXD,V871XXD,V872XXD,V873XXD,V874XXD | |
| | V875XXD,V876XXD,V877XXD,V878XXD,V879XXD,V880XXD,V881XXD | |
| | V882XXD,V883XXD,V884XXD,V885XXD,V886XXD,V887XXD,V888XXD | |
| | V889XXD,V890XXD,V891XXD,V892XXD,V893XXD,V899XXD,V9000XD | |
| | V9001XD,V9002XD,V9003XD,V9004XD,V9005XD,V9006XD,V9008XD | |
| | V9009XD,V9010XD,V9011XD,V9012XD,V9013XD,V9014XD,V9015XD | |
| | V9016XD,V9018XD,V9019XD,V9020XD,V9021XD,V9022XD,V9023XD | |
| | V9024XD,V9025XD,V9026XD,V9027XD,V9028XD,V9029XD,V9030XD | |
| | V9031XD,V9032XD,V9033XD,V9034XD,V9035XD,V9036XD,V9037XD | |
| | V9038XD,V9039XD,V9080XD,V9081XD,V9082XD,V9083XD,V9084XD | |
| | V9085XD,V9086XD,V9087XD,V9088XD,V9089XD,V9100XD,V9101XD | |
| | V9102XD,V9103XD,V9104XD,V9105XD,V9106XD,V9107XD,V9108XD | |
| | V9109XD,V9110XD,V9111XD,V9112XD,V9113XD,V9114XD,V9115XD | |
| | V9116XD,V9118XD,V9119XD,V9120XD,V9121XD,V9122XD,V9123XD | |
| | V9124XD,V9125XD,V9126XD,V9129XD,V9130XD,V9131XD,V9132XD | |
| | V9133XD,V9134XD,V9135XD,V9136XD,V9137XD,V9138XD,V9139XD | |
| | V9180XD,V9181XD,V9182XD,V9183XD,V9184XD,V9185XD,V9186XD | |
| | V9187XD,V9188XD,V9189XD,V9200XD,V9201XD,V9202XD,V9203XD | |
| | V9204XD,V9205XD,V9206XD,V9207XD,V9208XD,V9209XD,V9210XD | |
| | V9211XD,V9212XD,V9213XD,V9214XD,V9215XD,V9216XD,V9219XD | |
| | V9220XD,V9221XD,V9222XD,V9223XD,V9224XD,V9225XD,V9226XD | |
| | V9227XD,V9228XD,V9229XD,V9300XD,V9301XD,V9302XD,V9303XD | |
| | V9304XD,V9309XD,V9310XD,V9311XD,V9312XD,V9313XD,V9314XD | |
| | V9319XD,V9320XD,V9321XD,V9322XD,V9323XD,V9324XD,V9329XD | |
| | V9330XD,V9331XD,V9332XD,V9333XD,V9334XD,V9335XD,V9336XD | |
| | V9338XD,V9339XD,V9340XD,V9341XD,V9342XD,V9343XD,V9344XD | |
| | V9348XD,V9349XD,V9350XD,V9351XD,V9352XD,V9353XD,V9354XD | |
| | V9359XD,V9360XD,V9361XD,V9362XD,V9363XD,V9364XD,V9369XD | |
| | V9380XD,V9381XD,V9382XD,V9383XD,V9384XD,V9385XD,V9386XD | |
| | V9387XD,V9388XD,V9389XD,V940XXD,V9411XD,V9412XD,V9421XD | |
| | V9422XD,V9431XD,V9432XD,V944XXD,V94810D,V94811D,V94818D | |
| | V9489XD,V949XXD,V9500XD,V9501XD,V9502XD,V9503XD,V9504XD | |
| | V9505XD,V9509XD,V9510XD,V9511XD,V9512XD,V9513XD,V9514XD | |
| | V9515XD,V9519XD,V9520XD,V9521XD,V9522XD,V9523XD,V9524XD | |
| | V9525XD,V9529XD,V9530XD,V9531XD,V9532XD,V9533XD,V9534XD | |
| | V9535XD,V9539XD,V9540XD,V9541XD,V9542XD,V9543XD,V9544XD | |
| | V9545XD,V9549XD,V958XXD,V959XXD,V9600XD,V9601XD,V9602XD | |

Continued on next page

**Supplementary Table S1:** (cont'd) A comprehensive mapping of diagnosis groups used in the experiments to their corresponding ICD-10 and ICD-9 codes.

| Diagnosis | ICD-10 | ICD-9 |
|---|---|---|
| | V9603XD,V9604XD,V9605XD,V9609XD,V9610XD,V9611XD,V9612XD | |
| | V9613XD,V9614XD,V9615XD,V9619XD,V9620XD,V9621XD,V9622XD | |
| | V9623XD,V9624XD,V9625XD,V9629XD,V968XXD,V969XXD,V970XXD | |
| | V971XXD,V9721XD,V9722XD,V9729XD,V9731XD,V9732XD,V9733XD | |
| | V9739XD,V97810D,V97811D,V97818D,V9789XD,V980XXD,V981XXD | |
| | V982XXD,V983XXD,V988XXD,V99XXXD,W000XXD,W001XXD,W002XXD | |
| | W009XXD,W010XXD,W0110XD,W01110D,W01111D,W01118D,W01119D | |
| | W01190D,W01198D,W03XXXD,W04XXXD,W050XXD,W051XXD,W052XXD | |
| | W06XXXD,W07XXXD,W08XXXD,W090XXD,W091XXD,W092XXD,W098XXD | |
| | W100XXD,W101XXD,W102XXD,W108XXD,W109XXD,W11XXXD,W12XXXD | |
| | W130XXD,W131XXD,W132XXD,W133XXD,W134XXD,W138XXD,W139XXD | |
| | W14XXXD,W15XXXD,W16011D,W16012D,W16021D,W16022D,W16031D | |
| | W16032D,W16111D,W16112D,W16121D,W16122D,W16131D,W16132D | |
| | W16211D,W16212D,W16221D,W16222D,W16311D,W16312D,W16321D | |
| | W16322D,W16331D,W16332D,W1641XD,W1642XD,W16511D,W16512D | |
| | W16521D,W16522D,W16531D,W16532D,W16611D,W16612D,W16621D | |
| | W16622D,W16711D,W16712D,W16721D,W16722D,W16811D,W16812D | |
| | W16821D,W16822D,W16831D,W16832D,W1691XD,W1692XD,W170XXD | |
| | W171XXD,W172XXD,W173XXD,W174XXD,W1781XD,W1782XD,W1789XD | |
| | W1800XD,W1801XD,W1802XD,W1809XD,W1811XD,W1812XD,W182XXD | |
| | W1830XD,W1831XD,W1839XD,W1840XD,W1841XD,W1842XD,W1843XD | |
| | W1849XD,W19XXXD,W200XXD,W201XXD,W208XXD,W2100XD,W2101XD | |
| | W2102XD,W2103XD,W2104XD,W2105XD,W2106XD,W2107XD,W2109XD | |
| | W2111XD,W2112XD,W2113XD,W2119XD,W21210D,W21211D,W21220D | |
| | W21221D,W2131XD,W2132XD,W2139XD,W214XXD,W2181XD,W2189XD | |
| | W219XXD,W2201XD,W2202XD,W2203XD,W22041D,W22042D,W2209XD | |
| | W2210XD,W2211XD,W2212XD,W2219XD,W228XXD,W230XXD,W231XXD | |
| | W232XXD,W240XXD,W241XXD,W25XXXD,W260XXD,W261XXD,W262XXD | |
| | W268XXD,W269XXD,W270XXD,W271XXD,W272XXD,W273XXD,W274XXD | |
| | W275XXD,W278XXD,W28XXXD,W290XXD,W291XXD,W292XXD,W293XXD | |
| | W294XXD,W298XXD,W300XXD,W301XXD,W302XXD,W303XXD,W3081XD | |
| | W3089XD,W309XXD,W310XXD,W311XXD,W312XXD,W313XXD,W3181XD | |
| | W3182XD,W3183XD,W3189XD,W319XXD,W320XXD,W321XXD,W3300XD | |
| | W3301XD,W3302XD,W3303XD,W3309XD,W3310XD,W3311XD,W3312XD | |
| | W3313XD,W3319XD,W3400XD,W34010D,W34011D,W34018D,W3409XD | |
| | W3410XD,W34110D,W34111D,W34118D,W3419XD,W35XXXD,W361XXD | |
| | W362XXD,W363XXD,W368XXD,W369XXD,W370XXD,W378XXD,W38XXXD | |
| | W39XXXD,W400XXD,W401XXD,W408XXD,W409XXD,W420XXD,W429XXD | |
| | W450XXD,W451XXD,W452XXD,W458XXD,W460XXD,W461XXD,W4901XD | |
| | W4902XD,W4903XD,W4904XD,W4909XD,W499XXD,W500XXD,W501XXD | |

Continued on next page

**Supplementary Table S1:** (cont'd) A comprehensive mapping of diagnosis groups used in the experiments to their corresponding ICD-10 and ICD-9 codes.

| Diagnosis | ICD-10 | ICD-9 |
|---|---|---|
| | W502XXD,W503XXD,W504XXD,W51XXXD,W52XXXD,W5301XD,W5309XD | |
| | W5311XD,W5319XD,W5321XD,W5329XD,W5381XD,W5389XD,W540XXD | |
| | W541XXD,W548XXD,W5501XD,W5503XD,W5509XD,W5511XD,W5512XD | |
| | W5519XD,W5521XD,W5522XD,W5529XD,W5531XD,W5532XD,W5539XD | |
| | W5541XD,W5542XD,W5549XD,W5551XD,W5552XD,W5559XD,W5581XD | |
| | W5582XD,W5589XD,W5601XD,W5602XD,W5609XD,W5611XD,W5612XD | |
| | W5619XD,W5621XD,W5622XD,W5629XD,W5631XD,W5632XD,W5639XD | |
| | W5641XD,W5642XD,W5649XD,W5651XD,W5652XD,W5659XD,W5681XD | |
| | W5682XD,W5689XD,W57XXXD,W5801XD,W5802XD,W5803XD,W5809XD | |
| | W5811XD,W5812XD,W5813XD,W5819XD,W5901XD,W5902XD,W5909XD | |
| | W5911XD,W5912XD,W5913XD,W5919XD,W5921XD,W5922XD,W5929XD | |
| | W5981XD,W5982XD,W5983XD,W5989XD,W60XXXD,W6101XD,W6102XD | |
| | W6109XD,W6111XD,W6112XD,W6119XD,W6121XD,W6122XD,W6129XD | |
| | W6132XD,W6133XD,W6139XD,W6142XD,W6143XD,W6149XD,W6151XD | |
| | W6152XD,W6159XD,W6161XD,W6162XD,W6169XD,W6191XD,W6192XD | |
| | W6199XD,W620XXD,W621XXD,W629XXD,W64XXXD,W65XXXD,W67XXXD | |
| | W69XXXD,W73XXXD,W74XXXD,W85XXXD,W860XXD,W861XXD,W868XXD | |
| | W880XXD,W881XXD,W888XXD,W890XXD,W891XXD,W898XXD,W899XXD | |
| | W900XXD,W901XXD,W902XXD,W908XXD,W92XXXD,W9301XD,W9302XD | |
| | W9311XD,W9312XD,W932XXD,W938XXD,W940XXD,W9411XD,W9412XD | |
| | W9421XD,W9422XD,W9423XD,W9429XD,W9431XD,W9432XD,W9439XD | |
| | W99XXXD,X000XXD,X001XXD,X002XXD,X003XXD,X004XXD,X005XXD | |
| | X008XXD,X010XXD,X011XXD,X013XXD,X014XXD,X018XXD,X020XXD | |
| | X021XXD,X022XXD,X023XXD,X024XXD,X025XXD,X028XXD,X030XXD | |
| | X031XXD,X033XXD,X034XXD,X038XXD,X04XXXD,X05XXXD,X060XXD | |
| | X061XXD,X062XXD,X063XXD,X0800XD,X0801XD,X0809XD,X0810XD | |
| | X0811XD,X0819XD,X0820XD,X0821XD,X0829XD,X088XXD,X100XXD | |
| | X101XXD,X102XXD,X110XXD,X111XXD,X118XXD,X12XXXD,X130XXD | |
| | X131XXD,X140XXD,X141XXD,X150XXD,X151XXD,X152XXD,X153XXD | |
| | X158XXD,X16XXXD,X17XXXD,X18XXXD,X19XXXD,X30XXXD,X31XXXD | |
| | X32XXXD,X34XXXD,X35XXXD,X360XXD,X361XXD,X370XXD,X371XXD | |
| | X372XXD,X373XXD,X3741XD,X3742XD,X3743XD,X378XXD,X379XXD | |
| | X38XXXD,X3901XD,X3908XD,X398XXD,X500XXD,X501XXD,X503XXD | |
| | X509XXD,X52XXXD,X58XXXD,X710XXD,X711XXD,X712XXD,X713XXD | |
| | X718XXD,X719XXD,X72XXXD,X730XXD,X731XXD,X732XXD,X738XXD | |
| | X739XXD,X7401XD,X7402XD,X7409XD,X748XXD,X749XXD,X75XXXD | |
| | X76XXXD,X770XXD,X771XXD,X772XXD,X773XXD,X778XXD,X779XXD | |
| | X780XXD,X781XXD,X782XXD,X788XXD,X789XXD,X79XXXD,X80XXXD | |
| | X810XXD,X811XXD,X818XXD,X820XXD,X821XXD,X822XXD,X828XXD | |
| | X830XXD,X831XXD,X832XXD,X838XXD,X920XXD,X921XXD,X922XXD | |

Continued on next page

**Supplementary Table S1:** (cont'd) A comprehensive mapping of diagnosis groups used in the experiments to their corresponding ICD-10 and ICD-9 codes.

| Diagnosis | ICD-10 | ICD-9 |
|---|---|---|
| | X923XXD,X928XXD,X929XXD,X93XXXD,X940XXD,X941XXD,X942XXD | |
| | X948XXD,X949XXD,X9501XD,X9502XD,X9509XD,X958XXD,X959XXD | |
| | X960XXD,X961XXD,X962XXD,X963XXD,X964XXD,X968XXD,X969XXD | |
| | X97XXXD,X980XXD,X981XXD,X982XXD,X983XXD,X988XXD,X989XXD | |
| | X990XXD,X991XXD,X992XXD,X998XXD,X999XXD,Y00XXXD,Y01XXXD | |
| | Y020XXD,Y021XXD,Y028XXD,Y030XXD,Y038XXD,Y040XXD,Y041XXD | |
| | Y042XXD,Y048XXD,Y0801XD,Y0802XD,Y0809XD,Y0881XD,Y0889XD | |
| | Y210XXD,Y211XXD,Y212XXD,Y213XXD,Y214XXD,Y218XXD,Y219XXD | |
| | Y22XXXD,Y230XXD,Y231XXD,Y232XXD,Y233XXD,Y238XXD,Y239XXD | |
| | Y240XXD,Y248XXD,Y249XXD,Y25XXXD,Y26XXXD,Y270XXD,Y271XXD | |
| | Y272XXD,Y273XXD,Y278XXD,Y279XXD,Y280XXD,Y281XXD,Y282XXD | |
| | Y288XXD,Y289XXD,Y29XXXD,Y30XXXD,Y31XXXD,Y32XXXD,Y33XXXD | |
| | Y35001D,Y35002D,Y35003D,Y35009D,Y35011D,Y35012D,Y35013D | |
| | Y35019D,Y35021D,Y35022D,Y35023D,Y35029D,Y35031D,Y35032D | |
| | Y35033D,Y35039D,Y35041D,Y35042D,Y35043D,Y35049D,Y35091D | |
| | Y35092D,Y35093D,Y35099D,Y35101D,Y35102D,Y35103D,Y35109D | |
| | Y35111D,Y35112D,Y35113D,Y35119D,Y35121D,Y35122D,Y35123D | |
| | Y35129D,Y35191D,Y35192D,Y35193D,Y35199D,Y35201D,Y35202D | |
| | Y35203D,Y35209D,Y35211D,Y35212D,Y35213D,Y35219D,Y35291D | |
| | Y35292D,Y35293D,Y35299D,Y35301D,Y35302D,Y35303D,Y35309D | |
| | Y35311D,Y35312D,Y35313D,Y35319D,Y35391D,Y35392D,Y35393D | |
| | Y35399D,Y35401D,Y35402D,Y35403D,Y35409D,Y35411D,Y35412D | |
| | Y35413D,Y35419D,Y35491D,Y35492D,Y35493D,Y35499D,Y35811D | |
| | Y35812D,Y35813D,Y35819D,Y35831D,Y35832D,Y35833D,Y35839D | |
| | Y35891D,Y35892D,Y35893D,Y35899D,Y3591XD,Y3592XD,Y3593XD | |
| | Y3599XD,Y36000D,Y36001D,Y36010D,Y36011D,Y36020D,Y36021D | |
| | Y36030D,Y36031D,Y36040D,Y36041D,Y36050D,Y36051D,Y36090D | |
| | Y36091D,Y36100D,Y36101D,Y36110D,Y36111D,Y36120D,Y36121D | |
| | Y36130D,Y36131D,Y36140D,Y36141D,Y36190D,Y36191D,Y36200D | |
| | Y36201D,Y36210D,Y36211D,Y36220D,Y36221D,Y36230D,Y36231D | |
| | Y36240D,Y36241D,Y36250D,Y36251D,Y36260D,Y36261D,Y36270D | |
| | Y36271D,Y36290D,Y36291D,Y36300D,Y36301D,Y36310D,Y36311D | |
| | Y36320D,Y36321D,Y36330D,Y36331D,Y36390D,Y36391D,Y36410D | |
| | Y36411D,Y36420D,Y36421D,Y36430D,Y36431D,Y36440D,Y36441D | |
| | Y36450D,Y36451D,Y36460D,Y36461D,Y36470D,Y36471D,Y36490D | |
| | Y36491D,Y36500D,Y36501D,Y36510D,Y36511D,Y36520D,Y36521D | |
| | Y36530D,Y36531D,Y36540D,Y36541D,Y36590D,Y36591D,Y366X0D | |
| | Y366X1D,Y367X0D,Y367X1D,Y36810D,Y36811D,Y36820D,Y36821D | |
| | Y36880D,Y36881D,Y36890D,Y36891D,Y3690XD,Y3691XD,Y3692XD | |
| | Y37000D,Y37001D,Y37010D,Y37011D,Y37020D,Y37021D,Y37030D | |

Continued on next page

**Supplementary Table S1:** (cont'd) A comprehensive mapping of diagnosis groups used in the experiments to their corresponding ICD-10 and ICD-9 codes.

| Diagnosis | ICD-10 | ICD-9 |
|---|---|---|
| | Y37031D,Y37040D,Y37041D,Y37050D,Y37051D,Y37090D,Y37091D | |
| | Y37100D,Y37101D,Y37110D,Y37111D,Y37120D,Y37121D,Y37130D | |
| | Y37131D,Y37140D,Y37141D,Y37190D,Y37191D,Y37200D,Y37201D | |
| | Y37210D,Y37211D,Y37220D,Y37221D,Y37230D,Y37231D,Y37240D | |
| | Y37241D,Y37250D,Y37251D,Y37260D,Y37261D,Y37270D,Y37271D | |
| | Y37290D,Y37291D,Y37300D,Y37301D,Y37310D,Y37311D,Y37320D | |
| | Y37321D,Y37330D,Y37331D,Y37390D,Y37391D,Y37410D,Y37411D | |
| | Y37420D,Y37421D,Y37430D,Y37431D,Y37440D,Y37441D,Y37450D | |
| | Y37451D,Y37460D,Y37461D,Y37470D,Y37471D,Y37490D,Y37491D | |
| | Y37500D,Y37501D,Y37510D,Y37511D,Y37520D,Y37521D,Y37530D | |
| | Y37531D,Y37540D,Y37541D,Y37590D,Y37591D,Y376X0D,Y376X1D | |
| | Y377X0D,Y377X1D,Y3790XD,Y3791XD,Y3792XD,Y380X1D,Y380X2D | |
| | Y380X3D,Y381X1D,Y381X2D,Y381X3D,Y382X1D,Y382X2D,Y382X3D | |
| | Y383X1D,Y383X2D,Y383X3D,Y384X1D,Y384X2D,Y384X3D,Y385X1D | |
| | Y385X2D,Y385X3D,Y386X1D,Y386X2D,Y386X3D,Y387X1D,Y387X2D | |
| | Y387X3D,Y3880XD,Y38811D,Y38812D,Y38891D,Y38892D,Y38893D | |
| | Y389X1D,Y389X2D | |
| External cause codes: | T17200A,T17210A,T17220A,T17290A,T17300A,T17310A,T17320A | 9330,9331,9340,9341,9348,9349,9947,E911,E912,E9130,E9131 |
| suffocation | T17390A,T17400A,T17410A,T17420A,T17490A,T17500A,T17510A | E9132,E9133,E9138,E9139,E9530,E9531,E9538,E963,E9830,E9831 |
| inhalation; initial | T17520A,T17590A,T17800A,T17810A,T17820A,T17890A,T17900A | E9838,E9953,E9954 |
| encounter | T17910A,T17920A,T17990A,T71111A,T71112A,T71113A,T71114A | |
| | T71121A,T71122A,T71123A,T71124A,T71131A,T71132A,T71133A | |
| | T71134A,T71141A,T71143A,T71144A,T71151A,T71152A,T71153A | |
| | T71154A,T71161A,T71162A,T71163A,T71164A,T71191A,T71192A | |
| | T71193A,T71194A,T7120XA,T7121XA,T71221A,T71222A,T71223A | |
| | T71224A,T71231A,T71232A,T71233A,T71234A,T7129XA,T719XXA | |
| | Y36460A,Y36461A,Y36470A,Y36471A,Y37460A,Y37461A,Y37470A | |
| | Y37471A | |
| External cause codes: | V2009XA,V200XXA,V2019XA,V201XXA,V2029XA,V202XXA,V2109XA | E8001,E8011,E8018,E8019,E8021,E8031,E8041,E8061,E8071,E8129 |
| transport; not MVT; | V210XXA,V2119XA,V211XXA,V2129XA,V212XXA,V2209XA,V220XXA | E8139,E8199,E8200,E8201,E8208,E8209,E8210,E8211,E8218,E8219 |
| initial encounter | V2219XA,V221XXA,V2229XA,V222XXA,V2309XA,V230XXA,V2319XA | E8220,E8221,E8222,E8223,E8224,E8228,E8229,E8230,E8231,E8232 |
| | V231XXA,V2329XA,V232XXA,V2409XA,V240XXA,V2419XA,V241XXA | E8233,E8238,E8239,E8248,E8250,E8251,E8252,E8253,E8258,E8259 |
| | V2429XA,V242XXA,V2509XA,V250XXA,V2519XA,V251XXA,V2529XA | E8262,E8263,E8272,E8273,E8282,E8294,E8299,E8319,E8359,E8369 |
| | V252XXA,V2609XA,V260XXA,V2619XA,V261XXA,V2629XA,V262XXA | E8379,E8380,E8381,E8384,E8385,E8389,E8401,E8403,E8406,E8411 |
| | V2709XA,V270XXA,V2719XA,V271XXA,V2729XA,V272XXA,V2809XA | E8413,E8426,E8433,E8437,E8439,E8441,E8443,E8447,E8449,E8450 |
| | V280XXA,V2819XA,V281XXA,V2829XA,V282XXA,V29008A,V2900XA | E846,E847,E848,E9580,E9586,E9685,E9791,E9940,E9941,E9942 |
| | V29098A,V2909XA,V29108A,V2910XA,V29198A,V2919XA,V29208A | E9943,E9948,E9949 |
| | V2920XA,V29298A,V2929XA,V2939XA,V293XXA,V300XXA,V301XXA | |

Continued on next page

**Supplementary Table S1:** (cont'd) A comprehensive mapping of diagnosis groups used in the experiments to their corresponding ICD-10 and ICD-9 codes.

| Diagnosis | ICD-10 | ICD-9 |
|---|---|---|
| | V302XXA,V303XXA,V310XXA,V311XXA,V312XXA,V313XXA,V320XXA | |
| | V321XXA,V322XXA,V323XXA,V330XXA,V331XXA,V332XXA,V333XXA | |
| | V340XXA,V341XXA,V342XXA,V343XXA,V350XXA,V351XXA,V352XXA | |
| | V353XXA,V360XXA,V361XXA,V362XXA,V363XXA,V370XXA,V371XXA | |
| | V372XXA,V373XXA,V380XXA,V381XXA,V382XXA,V383XXA,V3900XA | |
| | V3909XA,V3910XA,V3919XA,V3920XA,V3929XA,V393XXA,V400XXA | |
| | V401XXA,V402XXA,V403XXA,V410XXA,V411XXA,V412XXA,V413XXA | |
| | V420XXA,V421XXA,V422XXA,V423XXA,V4301XA,V4302XA,V4303XA | |
| | V4304XA,V4311XA,V4312XA,V4313XA,V4314XA,V4321XA,V4322XA | |
| | V4323XA,V4324XA,V4331XA,V4332XA,V4333XA,V4334XA,V440XXA | |
| | V441XXA,V442XXA,V443XXA,V450XXA,V451XXA,V452XXA,V453XXA | |
| | V460XXA,V461XXA,V462XXA,V463XXA,V4701XA,V4702XA,V470XXA | |
| | V4711XA,V4712XA,V471XXA,V472XXA,V4731XA,V4732XA,V473XXA | |
| | V480XXA,V481XXA,V482XXA,V483XXA,V4900XA,V4909XA,V4910XA | |
| | V4919XA,V4920XA,V4929XA,V493XXA,V500XXA,V501XXA,V502XXA | |
| | V503XXA,V510XXA,V511XXA,V512XXA,V513XXA,V520XXA,V521XXA | |
| | V522XXA,V523XXA,V530XXA,V531XXA,V532XXA,V533XXA,V540XXA | |
| | V541XXA,V542XXA,V543XXA,V550XXA,V551XXA,V552XXA,V553XXA | |
| | V560XXA,V561XXA,V562XXA,V563XXA,V570XXA,V571XXA,V572XXA | |
| | V573XXA,V580XXA,V581XXA,V582XXA,V583XXA,V5900XA,V5909XA | |
| | V5910XA,V5919XA,V5920XA,V5929XA,V593XXA,V600XXA,V601XXA | |
| | V602XXA,V603XXA,V610XXA,V611XXA,V612XXA,V613XXA,V620XXA | |
| | V621XXA,V622XXA,V623XXA,V630XXA,V631XXA,V632XXA,V633XXA | |
| | V640XXA,V641XXA,V642XXA,V643XXA,V650XXA,V651XXA,V652XXA | |
| | V653XXA,V660XXA,V661XXA,V662XXA,V663XXA,V670XXA,V671XXA | |
| | V672XXA,V673XXA,V680XXA,V681XXA,V682XXA,V683XXA,V6900XA | |
| | V6909XA,V6910XA,V6919XA,V6920XA,V6929XA,V693XXA,V700XXA | |
| | V701XXA,V702XXA,V703XXA,V710XXA,V711XXA,V712XXA,V713XXA | |
| | V720XXA,V721XXA,V722XXA,V723XXA,V730XXA,V731XXA,V732XXA | |
| | V733XXA,V740XXA,V741XXA,V742XXA,V743XXA,V750XXA,V751XXA | |
| | V752XXA,V753XXA,V760XXA,V761XXA,V762XXA,V763XXA,V770XXA | |
| | V771XXA,V772XXA,V773XXA,V780XXA,V781XXA,V782XXA,V783XXA | |
| | V7900XA,V7909XA,V7910XA,V7919XA,V7920XA,V7929XA,V793XXA | |
| | V80010A,V80018A,V8002XA,V8011XA,V8012XA,V8021XA,V8022XA | |
| | V8061XA,V8062XA,V80710A,V80711A,V80720A,V80721A,V80730A | |
| | V80731A,V80790A,V80791A,V8081XA,V8082XA,V80910A,V80918A | |
| | V80919A,V80920A,V80928A,V80929A,V810XXA,V812XXA,V813XXA | |
| | V814XXA,V815XXA,V816XXA,V817XXA,V8181XA,V8182XA,V8183XA | |
| | V8189XA,V819XXA,V820XXA,V822XXA,V823XXA,V824XXA,V825XXA | |
| | V826XXA,V827XXA,V828XXA,V829XXA,V834XXA,V835XXA,V836XXA | |

Continued on next page

**Supplementary Table S1:** (cont'd) A comprehensive mapping of diagnosis groups used in the experiments to their corresponding ICD-10 and ICD-9 codes.

| Diagnosis | ICD-10 | ICD-9 |
|---|---|---|
| | V837XXA,V839XXA,V844XXA,V845XXA,V846XXA,V847XXA,V849XXA | |
| | V854XXA,V855XXA,V856XXA,V857XXA,V859XXA,V8641XA,V8642XA | |
| | V8643XA,V8644XA,V8645XA,V8646XA,V8649XA,V8651XA,V8652XA | |
| | V8653XA,V8654XA,V8655XA,V8656XA,V8659XA,V8661XA,V8662XA | |
| | V8663XA,V8664XA,V8665XA,V8666XA,V8669XA,V8671XA,V8672XA | |
| | V8673XA,V8674XA,V8675XA,V8676XA,V8679XA,V8691XA,V8692XA | |
| | V8693XA,V8694XA,V8695XA,V8696XA,V8699XA,V879XXA,V880XXA | |
| | V881XXA,V882XXA,V883XXA,V884XXA,V885XXA,V886XXA,V887XXA | |
| | V888XXA,V889XXA,V890XXA,V891XXA,V893XXA,V899XXA,V9100XA | |
| | V9101XA,V9102XA,V9103XA,V9104XA,V9105XA,V9106XA,V9107XA | |
| | V9108XA,V9109XA,V9110XA,V9111XA,V9112XA,V9113XA,V9114XA | |
| | V9115XA,V9116XA,V9118XA,V9119XA,V9120XA,V9121XA,V9122XA | |
| | V9123XA,V9124XA,V9125XA,V9126XA,V9129XA,V9130XA,V9131XA | |
| | V9132XA,V9133XA,V9134XA,V9135XA,V9136XA,V9137XA,V9138XA | |
| | V9139XA,V9180XA,V9181XA,V9182XA,V9183XA,V9184XA,V9185XA | |
| | V9186XA,V9187XA,V9188XA,V9189XA,V9300XA,V9301XA,V9302XA | |
| | V9303XA,V9304XA,V9309XA,V9310XA,V9311XA,V9312XA,V9313XA | |
| | V9314XA,V9319XA,V9320XA,V9321XA,V9322XA,V9323XA,V9324XA | |
| | V9329XA,V9330XA,V9331XA,V9332XA,V9333XA,V9334XA,V9335XA | |
| | V9336XA,V9338XA,V9339XA,V9340XA,V9341XA,V9342XA,V9343XA | |
| | V9344XA,V9348XA,V9349XA,V9350XA,V9351XA,V9352XA,V9353XA | |
| | V9354XA,V9359XA,V9360XA,V9361XA,V9362XA,V9363XA,V9364XA | |
| | V9369XA,V9380XA,V9381XA,V9382XA,V9383XA,V9384XA,V9385XA | |
| | V9386XA,V9387XA,V9388XA,V9389XA,V940XXA,V9411XA,V9412XA | |
| | V9421XA,V9422XA,V9431XA,V9432XA,V944XXA,V94810A,V94811A | |
| | V94818A,V9489XA,V949XXA,V9500XA,V9501XA,V9502XA,V9503XA | |
| | V9504XA,V9505XA,V9509XA,V9510XA,V9511XA,V9512XA,V9513XA | |
| | V9514XA,V9515XA,V9519XA,V9520XA,V9521XA,V9522XA,V9523XA | |
| | V9524XA,V9525XA,V9529XA,V9530XA,V9531XA,V9532XA,V9533XA | |
| | V9534XA,V9535XA,V9539XA,V9540XA,V9541XA,V9542XA,V9543XA | |
| | V9544XA,V9545XA,V9549XA,V958XXA,V959XXA,V9600XA,V9601XA | |
| | V9602XA,V9603XA,V9604XA,V9605XA,V9609XA,V9610XA,V9611XA | |
| | V9612XA,V9613XA,V9614XA,V9615XA,V9619XA,V9620XA,V9621XA | |
| | V9622XA,V9623XA,V9624XA,V9625XA,V9629XA,V968XXA,V969XXA | |
| | V970XXA,V971XXA,V9721XA,V9722XA,V9729XA,V9731XA,V9732XA | |
| | V9733XA,V9739XA,V97810A,V97811A,V97818A,V9789XA,V980XXA | |
| | V981XXA,V982XXA,V983XXA,V988XXA,V99XXXA,X811XXA,X830XXA | |
| | Y021XXA,Y0881XA,Y36100A,Y36101A,Y36110A,Y36111A,Y36120A | |
| | Y36121A,Y36130A,Y36131A,Y36140A,Y36141A,Y36190A,Y36191A | |
| | Y37100A,Y37101A,Y37110A,Y37111A,Y37120A,Y37121A,Y37130A | |

Continued on next page

**Supplementary Table S1:** (cont'd) A comprehensive mapping of diagnosis groups used in the experiments to their corresponding ICD-10 and ICD-9 codes.

| Diagnosis | ICD-10 | ICD-9 |
|---|---|---|
| | Y37131A,Y37140A,Y37141A,Y37190A,Y37191A,Y381X1A,Y381X2A | |
| | Y381X3A | |
| External cause codes: | R4588,T1491,T1491XA,X58XXXA,Y09,Y35899A,Y3591XA,Y3592XA | 9599,E9288,E9589,E9689,E976,E9798,E9909,E9989 |
| unspecified mechanism | Y3593XA,Y3599XA,Y36890A,Y36891A,Y3690XA,Y3790XA,Y3880XA | |
| Socioeconomic | Z550,Z551,Z552,Z553,Z554,Z555,Z558,Z559,Z560,Z561,Z562,Z563 | V1585,V600,V601,V602,V603,V606,V6081,V6089,V609,V6103,V6106 |
| psychosocial factors | Z564,Z565,Z566,Z5681,Z5682,Z5689,Z569,Z570,Z571,Z572,Z5731 | V6107,V6108,V6109,V6129,V613,V6141,V6149,V615,V616,V617,V618 |
| | Z5739,Z574,Z575,Z576,Z577,Z578,Z579,Z586,Z590,Z5900,Z5901 | V619,V620,V621,V6222,V6229,V623,V624,V625,V6281,V6282,V6289 |
| | Z5902,Z591,Z592,Z593,Z594,Z5941,Z5948,Z595,Z596,Z597,Z598 | V629 |
| | Z59811,Z59812,Z59819,Z5982,Z5986,Z5987,Z5989,Z599,Z600,Z602 | |
| | Z603,Z604,Z605,Z608,Z609,Z620,Z621,Z6221,Z6222,Z6229,Z623 | |
| | Z626,Z62810,Z62811,Z62812,Z62813,Z62819,Z62820,Z62821,Z62822 | |
| | Z62890,Z62891,Z62898,Z629,Z630,Z631,Z6331,Z6332,Z634,Z635 | |
| | Z636,Z6371,Z6372,Z6379,Z638,Z639,Z640,Z641,Z644,Z650,Z651 | |
| | Z652,Z653,Z654,Z655,Z658,Z659 | |
| Underdosing of drugs | T360X6A,T361X6A,T362X6A,T363X6A,T364X6A,T365X6A,T366X6A | NoDx |
| and medicaments, | T367X6A,T368X6A,T3696XA,T370X6A,T371X6A,T372X6A,T373X6A | |
| initial encounter | T374X6A,T375X6A,T378X6A,T3796XA,T380X6A,T381X6A,T382X6A | |
| | T383X6A,T384X6A,T385X6A,T386X6A,T387X6A,T38806A,T38816A | |
| | T38896A,T38906A,T38996A,T39016A,T39096A,T391X6A,T392X6A | |
| | T39316A,T39396A,T394X6A,T398X6A,T3996XA,T400X6A,T402X6A | |
| | T403X6A,T40416A,T40426A,T40496A,T404X6A,T405X6A,T40606A | |
| | T40696A,T40716A,T40726A,T407X6A,T40906A,T40996A,T410X6A | |
| | T411X6A,T41206A,T41296A,T413X6A,T4146XA,T415X6A,T420X6A | |
| | T421X6A,T422X6A,T423X6A,T424X6A,T425X6A,T426X6A,T4276XA | |
| | T428X6A,T43016A,T43026A,T431X6A,T43206A,T43216A,T43226A | |
| | T43296A,T433X6A,T434X6A,T43506A,T43596A,T43606A,T43616A | |
| | T43626A,T43636A,T43656A,T43696A,T438X6A,T4396XA,T440X6A | |
| | T441X6A,T442X6A,T443X6A,T444X6A,T445X6A,T446X6A,T447X6A | |
| | T448X6A,T44906A,T44996A,T450X6A,T451X6A,T452X6A,T453X6A | |
| | T454X6A,T45516A,T45526A,T45606A,T45616A,T45626A,T45696A | |
| | T457X6A,T458X6A,T4596XA,T460X6A,T461X6A,T462X6A,T463X6A | |
| | T464X6A,T465X6A,T466X6A,T467X6A,T468X6A,T46906A,T46996A | |
| | T470X6A,T471X6A,T472X6A,T473X6A,T474X6A,T475X6A,T476X6A | |
| | T477X6A,T478X6A,T4796XA,T480X6A,T481X6A,T48206A,T48296A | |
| | T483X6A,T484X6A,T485X6A,T486X6A,T48906A,T48996A,T490X6A | |
| | T491X6A,T492X6A,T493X6A,T494X6A,T495X6A,T496X6A,T497X6A | |
| | T498X6A,T4996XA,T500X6A,T501X6A,T502X6A,T503X6A,T504X6A | |

Continued on next page

**Supplementary Table S1:** (cont'd) A comprehensive mapping of diagnosis groups used in the experiments to their corresponding ICD-10 and ICD-9 codes.

| Diagnosis | ICD-10 | ICD-9 |
|---|---|---|
| | T505X6A,T506X6A,T507X6A,T508X6A,T50906A,T50916A,T50996A<br>T50A16A,T50A26A,T50A96A,T50B16A,T50B96A,T50Z16A,T50Z96A | |
| Underdosing of drugs and medicaments, subsequent encounter | T360X6D,T361X6D,T362X6D,T363X6D,T364X6D,T365X6D,T366X6D<br>T367X6D,T368X6D,T3696XD,T370X6D,T371X6D,T372X6D,T373X6D<br>T374X6D,T375X6D,T378X6D,T3796XD,T380X6D,T381X6D,T382X6D<br>T383X6D,T384X6D,T385X6D,T386X6D,T387X6D,T38806D,T38816D<br>T38896D,T38906D,T38996D,T39016D,T39096D,T391X6D,T392X6D<br>T39316D,T39396D,T394X6D,T398X6D,T3996XD,T400X6D,T402X6D<br>T403X6D,T40416D,T40426D,T40496D,T404X6D,T405X6D,T40606D<br>T40696D,T40716D,T40726D,T407X6D,T40906D,T40996D,T410X6D<br>T411X6D,T41206D,T41296D,T413X6D,T4146XD,T415X6D,T420X6D<br>T421X6D,T422X6D,T423X6D,T424X6D,T425X6D,T426X6D,T4276XD<br>T428X6D,T43016D,T43026D,T431X6D,T43206D,T43216D,T43226D<br>T43296D,T433X6D,T434X6D,T43506D,T43596D,T43606D,T43616D<br>T43626D,T43636D,T43656D,T43696D,T438X6D,T4396XD,T440X6D<br>T441X6D,T442X6D,T443X6D,T444X6D,T445X6D,T446X6D,T447X6D<br>T448X6D,T44906D,T44996D,T450X6D,T451X6D,T452X6D,T453X6D<br>T454X6D,T45516D,T45526D,T45606D,T45616D,T45626D,T45696D<br>T457X6D,T458X6D,T4596XD,T460X6D,T461X6D,T462X6D,T463X6D<br>T464X6D,T465X6D,T466X6D,T467X6D,T468X6D,T46906D,T46996D<br>T470X6D,T471X6D,T472X6D,T473X6D,T474X6D,T475X6D,T476X6D<br>T477X6D,T478X6D,T4796XD,T480X6D,T481X6D,T48206D,T48296D<br>T483X6D,T484X6D,T485X6D,T486X6D,T48906D,T48996D,T490X6D<br>T491X6D,T492X6D,T493X6D,T494X6D,T495X6D,T496X6D,T497X6D<br>T498X6D,T4996XD,T500X6D,T501X6D,T502X6D,T503X6D,T504X6D<br>T505X6D,T506X6D,T507X6D,T508X6D,T50906D,T50916D,T50996D<br>T50A16D,T50A26D,T50A96D,T50B16D,T50B96D,T50Z16D,T50Z96D | NoDx |
| Vasculitis | D690,I776,L950,L951,L958,L959,M0520,M05211,M05212,M05219<br>M05221,M05222,M05229,M05231,M05232,M05239,M05241,M05242<br>M05249,M05251,M05252,M05259,M05261,M05262,M05269,M05271<br>M05272,M05279,M0529,M300,M301,M302,M303,M308,M310,M311,M3110<br>M3111,M3119,M312,M3130,M3131,M314,M315,M316,M317,M318,M319<br>M350B | 2870,4460,4461,44620,44621,44629,4463,4464,4465,4466,4467<br>4475,4476,69589,7091,7142 |
| Cannabis-related disorders; subsequent encounter | T40711D,T40712D,T40713D,T40714D,T40715D,T40721D,T40722D<br>T40723D,T40724D,T40725D,T407X1D,T407X2D,T407X3D,T407X4D<br>T407X5D | V5889 |

Continued on next page

**Supplementary Table S1:** (cont'd) A comprehensive mapping of diagnosis groups used in the experiments to their corresponding ICD-10 and ICD-9 codes.

| Diagnosis | ICD-10 | ICD-9 |
|---|---|---|
| Diabetes mellitus,<br>Type 1 | E1010,E1011,E1021,E1022,E1029,E10311,E10319,E10321,E103211<br>E103212,E103213,E103219,E10329,E103291,E103292,E103293<br>E103299,E10331,E103311,E103312,E103313,E103319,E10339,E103391<br>E103392,E103393,E103399,E10341,E103411,E103412,E103413<br>E103419,E10349,E103491,E103492,E103493,E103499,E10351,E103511<br>E103512,E103513,E103519,E103521,E103522,E103523,E103529<br>E103531,E103532,E103533,E103539,E103541,E103542,E103543<br>E103549,E103551,E103552,E103553,E103559,E10359,E103591<br>E103592,E103593,E103599,E1036,E1037X1,E1037X2,E1037X3,E1037X9<br>E1039,E1040,E1041,E1042,E1043,E1044,E1049,E1051,E1052,E1059<br>E10610,E10618,E10620,E10621,E10622,E10628,E10630,E10638<br>E10641,E10649,E1065,E1069,E108,E109,O24011,O24012,O24013<br>O24019,O2402,O2403 | 25001,25011,25031,25041,25051,25061,25071,25081,25091,34989<br>3535,3559,3572,36201,36202,36204,36205,36206,36207,36641<br>44381,5238,5363,64800,64801,64803,64804,7135,71680,7854 |
| Diabetes mellitus,<br>Type 2 | E1100,E1101,E1110,E1111,E1121,E1122,E1129,E11311,E11319<br>E11321,E113211,E113212,E113213,E113219,E11329,E113291,E113292<br>E113293,E113299,E11331,E113311,E113312,E113313,E113319,E11339<br>E113391,E113392,E113393,E113399,E11341,E113411,E113412<br>E113413,E113419,E11349,E113491,E113492,E113493,E113499,E11351<br>E113511,E113512,E113513,E113519,E113521,E113522,E113523<br>E113529,E113531,E113532,E113533,E113539,E113541,E113542<br>E113543,E113549,E113551,E113552,E113553,E113559,E11359<br>E113591,E113592,E113593,E113599,E1136,E1137X1,E1137X2,E1137X3<br>E1137X9,E1139,E1140,E1141,E1142,E1143,E1144,E1149,E1151,E1152<br>E1159,E11610,E11618,E11620,E11621,E11622,E11628,E11630,E11638<br>E11641,E11649,E1165,E1169,E118,E119,O24111,O24112,O24113<br>O24119,O2412,O2413 | 25000,25020,25030,25040,25050,25060,25070,25080,25090,34989<br>3535,3559,3572,36201,36202,36204,36205,36206,36207,36641<br>44381,5238,5363,64800,64801,64803,64804,7135,71680,7854 |
| Diabetes mellitus,<br>due to underlying<br>condition, drug or<br>chemical induced, or<br>other specified type | E0800,E0801,E0810,E0811,E0821,E0822,E0829,E08311,E08319<br>E08321,E083211,E083212,E083213,E083219,E08329,E083291,E083292<br>E083293,E083299,E08331,E083311,E083312,E083313,E083319,E08339<br>E083391,E083392,E083393,E083399,E08341,E083411,E083412<br>E083413,E083419,E08349,E083491,E083492,E083493,E083499,E08351<br>E083511,E083512,E083513,E083519,E083521,E083522,E083523<br>E083529,E083531,E083532,E083533,E083539,E083541,E083542<br>E083543,E083549,E083551,E083552,E083553,E083559,E08359<br>E083591,E083592,E083593,E083599,E0836,E0837X1,E0837X2,E0837X3<br>E0837X9,E0839,E0840,E0841,E0842,E0843,E0844,E0849,E0851,E0852<br>E0859,E08610,E08618,E08620,E08621,E08622,E08628,E08630,E08638<br>E08641,E08649,E0865,E0869,E088,E089,E0900,E0901,E0910,E0911 | 24900,24910,24920,24930,24940,24950,24960,24970,24980,24990<br>25000,25010,25020,25030,25040,25050,25060,25070,25080,25090<br>2511,34989,3535,3559,3572,36201,36202,36204,36205,36206,36207<br>36641,44381,5238,5289,5363,58181,58381,7098,7135,71680,7854 |

Continued on next page

**Supplementary Table S1:** (cont'd) A comprehensive mapping of diagnosis groups used in the experiments to their corresponding ICD-10 and ICD-9 codes.

| Diagnosis | ICD-10 | ICD-9 |
|---|---|---|
| | E0921,E0922,E0929,E09311,E09319,E09321,E093211,E093212 | |
| | E093213,E093219,E09329,E093291,E093292,E093293,E093299,E09331 | |
| | E093311,E093312,E093313,E093319,E09339,E093391,E093392 | |
| | E093393,E093399,E09341,E093411,E093412,E093413,E093419,E09349 | |
| | E093491,E093492,E093493,E093499,E09351,E093511,E093512 | |
| | E093513,E093519,E093521,E093522,E093523,E093529,E093531 | |
| | E093532,E093533,E093539,E093541,E093542,E093543,E093549 | |
| | E093551,E093552,E093553,E093559,E09359,E093591,E093592 | |
| | E093593,E093599,E0936,E0937X1,E0937X2,E0937X3,E0937X9,E0939 | |
| | E0940,E0941,E0942,E0943,E0944,E0949,E0951,E0952,E0959,E09610 | |
| | E09618,E09620,E09621,E09622,E09628,E09630,E09638,E09641 | |
| | E09649,E0965,E0969,E098,E099,E1300,E1301,E1310,E1311,E1321 | |
| | E1322,E1329,E13311,E13319,E13321,E133211,E133212,E133213 | |
| | E133219,E13329,E133291,E133292,E133293,E133299,E13331,E133311 | |
| | E133312,E133313,E133319,E13339,E133391,E133392,E133393 | |
| | E133399,E13341,E133411,E133412,E133413,E133419,E13349,E133491 | |
| | E133492,E133493,E133499,E13351,E133511,E133512,E133513 | |
| | E133519,E133521,E133522,E133523,E133529,E133531,E133532 | |
| | E133533,E133539,E133541,E133542,E133543,E133549,E133551 | |
| | E133552,E133553,E133559,E13359,E133591,E133592,E133593 | |
| | E133599,E1336,E1339,E1340,E1341,E1342,E1343,E1344,E1349,E1351 | |
| | E1352,E1359,E13610,E13618,E13620,E13621,E13622,E13628,E13630 | |
| | E13638,E13641,E13649,E1365,E1369,E138,E139 | |
| External cause codes: | T1500XA,T1501XA,T1502XA,T1510XA,T1511XA,T1512XA,T1580XA | 9300,9301,9302,9308,9309,931,932,9330,9331,9340,9341,9348 |
| intent of injury, | T1581XA,T1582XA,T1590XA,T1591XA,T1592XA,T161XXA,T162XXA | 9349,9350,9351,9352,936,937,938,9390,9391,9392,9393,9399,9600 |
| accidental | T169XXA,T170XXA,T171XXA,T17200A,T17208A,T17210A,T17218A | 9601,9602,9603,9604,9605,9606,9608,9609,9610,9614,9615,9616 |
| unintentional | T17220A,T17228A,T17290A,T17298A,T17300A,T17308A,T17310A | 9617,9618,9619,9620,9621,9622,9623,9624,9627,9628,9629,9630 |
| | T17318A,T17320A,T17328A,T17390A,T17398A,T17400A,T17408A | 9631,9634,9635,9638,9639,9640,9642,9644,9645,9646,9648,9649 |
| | T17410A,T17418A,T17420A,T17428A,T17490A,T17498A,T17500A | 96500,96501,96502,96509,9651,9654,9655,96561,96569,9657,9659 |
| | T17508A,T17510A,T17518A,T17520A,T17528A,T17590A,T17598A | 9660,9661,9662,9663,9664,9670,9678,9679,9680,9682,9683,9684 |
| | T17800A,T17808A,T17810A,T17818A,T17820A,T17828A,T17890A | 9685,9689,96900,96901,96902,96903,96904,96905,96909,9691,9692 |
| | T17898A,T17900A,T17908A,T17910A,T17918A,T17920A,T17928A | 9693,9694,9696,96970,96971,96972,96973,96979,9698,9699,9700 |
| | T17990A,T17998A,T180XXA,T18100A,T18108A,T18110A,T18118A | 9701,97081,9710,9711,9712,9713,9719,9720,9721,9722,9723,9724 |
| | T18120A,T18128A,T18190A,T18198A,T182XXA,T183XXA,T184XXA | 9725,9726,9727,9729,9730,9731,9732,9733,9734,9735,9736,9738 |
| | T185XXA,T188XXA,T189XXA,T190XXA,T191XXA,T192XXA,T193XXA | 9739,9742,9744,9745,9747,9750,9751,9752,9753,9754,9755,9756 |
| | T194XXA,T198XXA,T199XXA,T360X1A,T361X1A,T362X1A,T363X1A | 9757,9758,9760,9761,9762,9763,9764,9765,9766,9767,9768,9769 |
| | T364X1A,T365X1A,T366X1A,T367X1A,T368X1A,T3691XA,T370X1A | 9770,9772,9778,9779,9786,9788,9789,9790,9796,9799,9800,9801 |
| | T371X1A,T372X1A,T373X1A,T374X1A,T375X1A,T378X1A,T3791XA | 9802,9803,9808,9809,981,9820,9821,9822,9823,9824,9828,9830 |
| | T380X1A,T381X1A,T382X1A,T383X1A,T384X1A,T385X1A,T386X1A | 9831,9832,9839,9849,9850,9851,9852,9853,9855,9856,9858,9859 |

Continued on next page

**Supplementary Table S1:** (cont'd) A comprehensive mapping of diagnosis groups used in the experiments to their corresponding ICD-10 and ICD-9 codes.

| Diagnosis | ICD-10 | ICD-9 |
|---|---|---|
| | T387X1A,T38801A,T38811A,T38891A,T38901A,T38991A,T39011A | 986,9871,9872,9873,9874,9875,9876,9877,9878,9879,9880,9881 |
| | T39091A,T391X1A,T392X1A,T39311A,T39391A,T394X1A,T398X1A | 9882,9888,9889,9890,9891,9893,9894,9895,9896,9897,98982,98984 |
| | T3991XA,T400X1A,T401X1A,T402X1A,T403X1A,T40411A,T40421A | 98989,9899,9940,9942,9943,9944,9945,9946,9947,9948,9949,E8001 |
| | T40491A,T404X1A,T405X1A,T40601A,T40691A,T40711A,T40721A | E8011,E8013,E8018,E8019,E8021,E8031,E8041,E8052,E8061,E8071 |
| | T407X1A,T408X1A,T40901A,T40991A,T410X1A,T411X1A,T41201A | E8072,E8100,E8101,E8102,E8103,E8107,E8108,E8109,E8120,E8121 |
| | T41291A,T413X1A,T4141XA,T415X1A,T420X1A,T421X1A,T422X1A | E8122,E8123,E8128,E8129,E8130,E8131,E8132,E8133,E8134,E8135 |
| | T423X1A,T424X1A,T425X1A,T426X1A,T4271XA,T428X1A,T43011A | E8136,E8138,E8139,E8140,E8141,E8142,E8143,E8147,E8148,E8149 |
| | T43021A,T431X1A,T43201A,T43211A,T43221A,T43291A,T433X1A | E8150,E8151,E8152,E8153,E8158,E8159,E8160,E8161,E8162,E8163 |
| | T434X1A,T43501A,T43591A,T43601A,T43611A,T43621A,T43631A | E8168,E8169,E8178,E8188,E8189,E8190,E8192,E8199,E8200,E8201 |
| | T43641A,T43651A,T43691A,T438X1A,T4391XA,T440X1A,T441X1A | E8208,E8209,E8210,E8211,E8218,E8219,E8220,E8221,E8222,E8223 |
| | T442X1A,T443X1A,T444X1A,T445X1A,T446X1A,T447X1A,T448X1A | E8224,E8226,E8227,E8228,E8229,E8230,E8231,E8232,E8233,E8238 |
| | T44901A,T44991A,T450X1A,T451X1A,T452X1A,T453X1A,T454X1A | E8239,E8242,E8246,E8248,E8250,E8251,E8252,E8253,E8258,E8259 |
| | T45511A,T45521A,T45601A,T45611A,T45621A,T45691A,T457X1A | E8260,E8261,E8262,E8263,E8269,E8272,E8273,E8282,E8290,E8294 |
| | T458X1A,T4591XA,T460X1A,T461X1A,T462X1A,T463X1A,T464X1A | E8299,E8309,E8319,E8329,E8359,E8369,E8379,E8380,E8381,E8384 |
| | T465X1A,T466X1A,T467X1A,T468X1A,T46901A,T46991A,T470X1A | E8385,E8389,E8401,E8403,E8406,E8411,E8413,E8426,E8433,E8437 |
| | T471X1A,T472X1A,T473X1A,T474X1A,T475X1A,T476X1A,T477X1A | E8439,E8441,E8443,E8447,E8449,E8450,E846,E847,E848,E8500 |
| | T478X1A,T4791XA,T480X1A,T481X1A,T48201A,T48291A,T483X1A | E8501,E8502,E8503,E8504,E8505,E8506,E8507,E851,E8528,E8530 |
| | T484X1A,T485X1A,T486X1A,T48901A,T48991A,T490X1A,T491X1A | E8531,E8532,E8540,E8541,E8542,E8543,E8548,E8550,E8551,E8552 |
| | T492X1A,T493X1A,T494X1A,T495X1A,T496X1A,T497X1A,T498X1A | E8553,E8554,E8555,E8556,E8558,E856,E857,E8580,E8581,E8582 |
| | T4991XA,T500X1A,T501X1A,T502X1A,T503X1A,T504X1A,T505X1A | E8583,E8584,E8585,E8586,E8587,E8588,E8601,E8602,E8603,E8604 |
| | T506X1A,T507X1A,T508X1A,T50901A,T50911A,T50991A,T50A11A | E8608,E8609,E8610,E8611,E8616,E8621,E8624,E8631,E8634,E8635 |
| | T50A21A,T50A91A,T50B11A,T50B91A,T50Z11A,T50Z91A,T510X1A | E8637,E8640,E8641,E8643,E8644,E8651,E8652,E8653,E8654,E8655 |
| | T511X1A,T512X1A,T513X1A,T518X1A,T5191XA,T520X1A,T521X1A | E8658,E8659,E8660,E8661,E8663,E8664,E8668,E8681,E8682,E8683 |
| | T522X1A,T523X1A,T524X1A,T528X1A,T5291XA,T530X1A,T531X1A | E8688,E8689,E8690,E8691,E8692,E8693,E8698,E8800,E8801,E8809 |
| | T532X1A,T533X1A,T534X1A,T535X1A,T536X1A,T537X1A,T5391XA | E8810,E8811,E882,E8830,E8831,E8832,E8839,E8840,E8841,E8842 |
| | T540X1A,T541X1A,T542X1A,T543X1A,T5491XA,T550X1A,T551X1A | E8843,E8844,E8845,E8846,E8849,E8850,E8851,E8852,E8853,E8854 |
| | T560X1A,T561X1A,T562X1A,T563X1A,T564X1A,T565X1A,T566X1A | E8859,E8860,E8869,E8880,E8881,E8888,E8889,E8902,E8903,E8908 |
| | T567X1A,T56811A,T56891A,T5691XA,T570X1A,T571X1A,T572X1A | E892,E8930,E8938,E894,E896,E897,E8980,E8981,E9000,E9001,E9010 |
| | T573X1A,T578X1A,T5791XA,T5801XA,T5811XA,T582X1A,T588X1A | E9011,E9018,E9020,E9021,E9022,E9028,E903,E9041,E9042,E9043 |
| | T5891XA,T590X1A,T591X1A,T592X1A,T593X1A,T594X1A,T595X1A | E9049,E9050,E9051,E9052,E9053,E9054,E9055,E9056,E9060,E9061 |
| | T596X1A,T597X1A,T59811A,T59891A,T5991XA,T600X1A,T601X1A | E9062,E9063,E9064,E9068,E9069,E9080,E9081,E9082,E9083,E9084 |
| | T602X1A,T603X1A,T604X1A,T608X1A,T6091XA,T6101XA,T6111XA | E9088,E9089,E9090,E9091,E9092,E9093,E9094,E9098,E9102,E9104 |
| | T61771A,T61781A,T618X1A,T6191XA,T620X1A,T621X1A,T622X1A | E9108,E9109,E911,E912,E9130,E9131,E9132,E9133,E9138,E9139 |
| | T628X1A,T6291XA,T63001A,T63011A,T63021A,T63031A,T63041A | E914,E915,E916,E9170,E9171,E9173,E9174,E9178,E9179,E918,E9190 |
| | T63061A,T63071A,T63081A,T63091A,T63111A,T63121A,T63191A | E9191,E9192,E9193,E9194,E9195,E9196,E9197,E9198,E9199,E9200 |
| | T632X1A,T63301A,T63311A,T63321A,T63331A,T63391A,T63411A | E9201,E9202,E9203,E9204,E9205,E9208,E9210,E9211,E9218,E9220 |
| | T63421A,T63431A,T63441A,T63451A,T63461A,T63481A,T63511A | E9221,E9222,E9223,E9224,E9225,E9228,E9229,E9230,E9231,E9232 |
| | T63591A,T63611A,T63621A,T63631A,T63691A,T63711A,T63791A | E9238,E9239,E9240,E9241,E9242,E9248,E9250,E9251,E9252,E9258 |
| | T63811A,T63821A,T63831A,T63891A,T6391XA,T6401XA,T6481XA | E9260,E9261,E9262,E9263,E9264,E9265,E9268,E9272,E9280,E9281 |
| | T650X1A,T651X1A,T65211A,T65221A,T65291A,T653X1A,T654X1A | E9283,E9284,E9285,E9286,E9288 |

Continued on next page

**Supplementary Table S1:** (cont'd) A comprehensive mapping of diagnosis groups used in the experiments to their corresponding ICD-10 and ICD-9 codes.

| Diagnosis | ICD-10 | ICD-9 |
|---|---|---|
| | T655X1A,T656X1A,T65811A,T65821A,T65831A,T65891A,T6591XA | |
| | T71111A,T71121A,T71131A,T71141A,T71151A,T71161A,T71191A | |
| | T7120XA,T7121XA,T71221A,T71231A,T7129XA,T719XXA,T730XXA | |
| | T731XXA,T732XXA,T733XXA,T738XXA,T739XXA,T7500XA,T7501XA | |
| | T7509XA,T7520XA,T7521XA,T7522XA,T7523XA,T7529XA,T753XXA | |
| | T754XXA,V0001XA,V0002XA,V00031A,V00038A,V0009XA,V00111A | |
| | V00112A,V00118A,V00121A,V00122A,V00128A,V00131A,V00132A | |
| | V00138A,V00141A,V00142A,V00148A,V00151A,V00152A,V00158A | |
| | V00181A,V00182A,V00188A,V00211A,V00212A,V00218A,V00221A | |
| | V00222A,V00228A,V00281A,V00282A,V00288A,V00311A,V00312A | |
| | V00318A,V00321A,V00322A,V00328A,V00381A,V00382A,V00388A | |
| | V00811A,V00812A,V00818A,V00821A,V00822A,V00828A,V00831A | |
| | V00832A,V00838A,V00841A,V00842A,V00848A,V00891A,V00892A | |
| | V00898A,V0100XA,V0101XA,V0102XA,V01031A,V01038A,V0109XA | |
| | V0110XA,V0111XA,V0112XA,V01131A,V01138A,V0119XA,V0190XA | |
| | V0191XA,V0192XA,V01931A,V01938A,V0199XA,V0200XA,V0201XA | |
| | V0202XA,V02031A,V02038A,V0209XA,V0210XA,V0211XA,V0212XA | |
| | V02131A,V02138A,V0219XA,V0290XA,V0291XA,V0292XA,V02931A | |
| | V02938A,V0299XA,V0300XA,V0301XA,V0302XA,V03031A,V03038A | |
| | V0309XA,V0310XA,V0311XA,V0312XA,V03131A,V03138A,V0319XA | |
| | V0390XA,V0391XA,V0392XA,V03931A,V03938A,V0399XA,V0400XA | |
| | V0401XA,V0402XA,V04031A,V04038A,V0409XA,V0410XA,V0411XA | |
| | V0412XA,V04131A,V04138A,V0419XA,V0490XA,V0491XA,V0492XA | |
| | V04931A,V04938A,V0499XA,V0500XA,V0501XA,V0502XA,V05031A | |
| | V05038A,V0509XA,V0510XA,V0511XA,V0512XA,V05131A,V05138A | |
| | V0519XA,V0590XA,V0591XA,V0592XA,V05931A,V05938A,V0599XA | |
| | V0600XA,V0601XA,V0602XA,V06031A,V06038A,V0609XA,V0610XA | |
| | V0611XA,V0612XA,V06131A,V06138A,V0619XA,V0690XA,V0691XA | |
| | V0692XA,V06931A,V06938A,V0699XA,V0900XA,V0901XA,V0909XA | |
| | V091XXA,V0920XA,V0921XA,V0929XA,V093XXA,V099XXA,V100XXA | |
| | V101XXA,V102XXA,V103XXA,V104XXA,V105XXA,V109XXA,V110XXA | |
| | V111XXA,V112XXA,V113XXA,V114XXA,V115XXA,V119XXA,V120XXA | |
| | V121XXA,V122XXA,V123XXA,V124XXA,V125XXA,V129XXA,V130XXA | |
| | V131XXA,V132XXA,V133XXA,V134XXA,V135XXA,V139XXA,V140XXA | |
| | V141XXA,V142XXA,V143XXA,V144XXA,V145XXA,V149XXA,V150XXA | |
| | V151XXA,V152XXA,V153XXA,V154XXA,V155XXA,V159XXA,V160XXA | |
| | V161XXA,V162XXA,V163XXA,V164XXA,V165XXA,V169XXA,V170XXA | |
| | V171XXA,V172XXA,V173XXA,V174XXA,V175XXA,V179XXA,V180XXA | |
| | V181XXA,V182XXA,V183XXA,V184XXA,V185XXA,V189XXA,V1900XA | |
| | V1909XA,V1910XA,V1919XA,V1920XA,V1929XA,V193XXA,V1940XA | |

Continued on next page

**Supplementary Table S1:** (cont'd) A comprehensive mapping of diagnosis groups used in the experiments to their corresponding ICD-10 and ICD-9 codes.

| Diagnosis | ICD-10 | ICD-9 |
|---|---|---|
| | V1949XA,V1950XA,V1959XA,V1960XA,V1969XA,V1981XA,V1988XA | |
| | V199XXA,V2001XA,V2009XA,V200XXA,V2011XA,V2019XA,V201XXA | |
| | V2021XA,V2029XA,V202XXA,V2031XA,V2039XA,V203XXA,V2041XA | |
| | V2049XA,V204XXA,V2051XA,V2059XA,V205XXA,V2091XA,V2099XA | |
| | V209XXA,V2101XA,V2109XA,V210XXA,V2111XA,V2119XA,V211XXA | |
| | V2121XA,V2129XA,V212XXA,V2131XA,V2139XA,V213XXA,V2141XA | |
| | V2149XA,V214XXA,V2151XA,V2159XA,V215XXA,V2191XA,V2199XA | |
| | V219XXA,V2201XA,V2209XA,V220XXA,V2211XA,V2219XA,V221XXA | |
| | V2221XA,V2229XA,V222XXA,V2231XA,V2239XA,V223XXA,V2241XA | |
| | V2249XA,V224XXA,V2251XA,V2259XA,V225XXA,V2291XA,V2299XA | |
| | V229XXA,V2301XA,V2309XA,V230XXA,V2311XA,V2319XA,V231XXA | |
| | V2321XA,V2329XA,V232XXA,V2331XA,V2339XA,V233XXA,V2341XA | |
| | V2349XA,V234XXA,V2351XA,V2359XA,V235XXA,V2391XA,V2399XA | |
| | V239XXA,V2401XA,V2409XA,V240XXA,V2411XA,V2419XA,V241XXA | |
| | V2421XA,V2429XA,V242XXA,V2431XA,V2439XA,V243XXA,V2441XA | |
| | V2449XA,V244XXA,V2451XA,V2459XA,V245XXA,V2491XA,V2499XA | |
| | V249XXA,V2501XA,V2509XA,V250XXA,V2511XA,V2519XA,V251XXA | |
| | V2521XA,V2529XA,V252XXA,V2531XA,V2539XA,V253XXA,V2541XA | |
| | V2549XA,V254XXA,V2551XA,V2559XA,V255XXA,V2591XA,V2599XA | |
| | V259XXA,V2601XA,V2609XA,V260XXA,V2611XA,V2619XA,V261XXA | |
| | V2621XA,V2629XA,V262XXA,V2631XA,V2639XA,V263XXA,V2641XA | |
| | V2649XA,V264XXA,V2651XA,V2659XA,V265XXA,V2691XA,V2699XA | |
| | V269XXA,V2701XA,V2709XA,V270XXA,V2711XA,V2719XA,V271XXA | |
| | V2721XA,V2729XA,V272XXA,V2731XA,V2739XA,V273XXA,V2741XA | |
| | V2749XA,V274XXA,V2751XA,V2759XA,V275XXA,V2791XA,V2799XA | |
| | V279XXA,V2801XA,V2809XA,V280XXA,V2811XA,V2819XA,V281XXA | |
| | V2821XA,V2829XA,V282XXA,V2831XA,V2839XA,V283XXA,V2841XA | |
| | V2849XA,V284XXA,V2851XA,V2859XA,V285XXA,V2891XA,V2899XA | |
| | V289XXA,V29001A,V29008A,V2900XA,V29091A,V29098A,V2909XA | |
| | V29101A,V29108A,V2910XA,V29191A,V29198A,V2919XA,V29201A | |
| | V29208A,V2920XA,V29291A,V29298A,V2929XA,V2931XA,V2939XA | |
| | V293XXA,V29401A,V29408A,V2940XA,V29491A,V29498A,V2949XA | |
| | V29501A,V29508A,V2950XA,V29591A,V29598A,V2959XA,V29601A | |
| | V29608A,V2960XA,V29691A,V29698A,V2969XA,V29811A,V29818A | |
| | V2981XA,V29881A,V29888A,V2988XA,V2991XA,V2999XA,V299XXA | |
| | V300XXA,V301XXA,V302XXA,V303XXA,V304XXA,V305XXA,V306XXA | |
| | V307XXA,V309XXA,V310XXA,V311XXA,V312XXA,V313XXA,V314XXA | |
| | V315XXA,V316XXA,V317XXA,V319XXA,V320XXA,V321XXA,V322XXA | |
| | V323XXA,V324XXA,V325XXA,V326XXA,V327XXA,V329XXA,V330XXA | |
| | V331XXA,V332XXA,V333XXA,V334XXA,V335XXA,V336XXA,V337XXA | |

Continued on next page

**Supplementary Table S1:** (cont'd) A comprehensive mapping of diagnosis groups used in the experiments to their corresponding ICD-10 and ICD-9 codes.

| Diagnosis | ICD-10 | ICD-9 |
|---|---|---|
| | V339XXA,V340XXA,V341XXA,V342XXA,V343XXA,V344XXA,V345XXA | |
| | V346XXA,V347XXA,V349XXA,V350XXA,V351XXA,V352XXA,V353XXA | |
| | V354XXA,V355XXA,V356XXA,V357XXA,V359XXA,V360XXA,V361XXA | |
| | V362XXA,V363XXA,V364XXA,V365XXA,V366XXA,V367XXA,V369XXA | |
| | V370XXA,V371XXA,V372XXA,V373XXA,V374XXA,V375XXA,V376XXA | |
| | V377XXA,V379XXA,V380XXA,V381XXA,V382XXA,V383XXA,V384XXA | |
| | V385XXA,V386XXA,V387XXA,V389XXA,V3900XA,V3909XA,V3910XA | |
| | V3919XA,V3920XA,V3929XA,V393XXA,V3940XA,V3949XA,V3950XA | |
| | V3959XA,V3960XA,V3969XA,V3981XA,V3989XA,V399XXA,V400XXA | |
| | V401XXA,V402XXA,V403XXA,V404XXA,V405XXA,V406XXA,V407XXA | |
| | V409XXA,V410XXA,V411XXA,V412XXA,V413XXA,V414XXA,V415XXA | |
| | V416XXA,V417XXA,V419XXA,V420XXA,V421XXA,V422XXA,V423XXA | |
| | V424XXA,V425XXA,V426XXA,V427XXA,V429XXA,V4301XA,V4302XA | |
| | V4303XA,V4304XA,V4311XA,V4312XA,V4313XA,V4314XA,V4321XA | |
| | V4322XA,V4323XA,V4324XA,V4331XA,V4332XA,V4333XA,V4334XA | |
| | V4341XA,V4342XA,V4343XA,V4344XA,V4351XA,V4352XA,V4353XA | |
| | V4354XA,V4361XA,V4362XA,V4363XA,V4364XA,V4371XA,V4372XA | |
| | V4373XA,V4374XA,V4391XA,V4392XA,V4393XA,V4394XA,V440XXA | |
| | V441XXA,V442XXA,V443XXA,V444XXA,V445XXA,V446XXA,V447XXA | |
| | V449XXA,V450XXA,V451XXA,V452XXA,V453XXA,V454XXA,V455XXA | |
| | V456XXA,V457XXA,V459XXA,V460XXA,V461XXA,V462XXA,V463XXA | |
| | V464XXA,V465XXA,V466XXA,V467XXA,V469XXA,V4701XA,V4702XA | |
| | V470XXA,V4711XA,V4712XA,V471XXA,V472XXA,V4731XA,V4732XA | |
| | V473XXA,V474XXA,V4751XA,V4752XA,V475XXA,V4761XA,V4762XA | |
| | V476XXA,V477XXA,V4791XA,V4792XA,V479XXA,V480XXA,V481XXA | |
| | V482XXA,V483XXA,V484XXA,V485XXA,V486XXA,V487XXA,V489XXA | |
| | V4900XA,V4909XA,V4910XA,V4919XA,V4920XA,V4929XA,V493XXA | |
| | V4940XA,V4949XA,V4950XA,V4959XA,V4960XA,V4969XA,V4981XA | |
| | V4988XA,V499XXA,V500XXA,V501XXA,V502XXA,V503XXA,V504XXA | |
| | V505XXA,V506XXA,V507XXA,V509XXA,V510XXA,V511XXA,V512XXA | |
| | V513XXA,V514XXA,V515XXA,V516XXA,V517XXA,V519XXA,V520XXA | |
| | V521XXA,V522XXA,V523XXA,V524XXA,V525XXA,V526XXA,V527XXA | |
| | V529XXA,V530XXA,V531XXA,V532XXA,V533XXA,V534XXA,V535XXA | |
| | V536XXA,V537XXA,V539XXA,V540XXA,V541XXA,V542XXA,V543XXA | |
| | V544XXA,V545XXA,V546XXA,V547XXA,V549XXA,V550XXA,V551XXA | |
| | V552XXA,V553XXA,V554XXA,V555XXA,V556XXA,V557XXA,V559XXA | |
| | V560XXA,V561XXA,V562XXA,V563XXA,V564XXA,V565XXA,V566XXA | |
| | V567XXA,V569XXA,V570XXA,V571XXA,V572XXA,V573XXA,V574XXA | |
| | V575XXA,V576XXA,V577XXA,V579XXA,V580XXA,V581XXA,V582XXA | |
| | V583XXA,V584XXA,V585XXA,V586XXA,V587XXA,V589XXA,V5900XA | |

Continued on next page

**Supplementary Table S1:** (cont'd) A comprehensive mapping of diagnosis groups used in the experiments to their corresponding ICD-10 and ICD-9 codes.

| Diagnosis | ICD-10 | ICD-9 |
|---|---|---|
| | V5909XA,V5910XA,V5919XA,V5920XA,V5929XA,V593XXA,V5940XA<br>V5949XA,V5950XA,V5959XA,V5960XA,V5969XA,V5981XA,V5988XA<br>V599XXA,V600XXA,V601XXA,V602XXA,V603XXA,V604XXA,V605XXA<br>V606XXA,V607XXA,V609XXA,V610XXA,V611XXA,V612XXA,V613XXA<br>V614XXA,V615XXA,V616XXA,V617XXA,V619XXA,V620XXA,V621XXA<br>V622XXA,V623XXA,V624XXA,V625XXA,V626XXA,V627XXA,V629XXA<br>V630XXA,V631XXA,V632XXA,V633XXA,V634XXA,V635XXA,V636XXA<br>V637XXA,V639XXA,V640XXA,V641XXA,V642XXA,V643XXA,V644XXA<br>V645XXA,V646XXA,V647XXA,V649XXA,V650XXA,V651XXA,V652XXA<br>V653XXA,V654XXA,V655XXA,V656XXA,V657XXA,V659XXA,V660XXA<br>V661XXA,V662XXA,V663XXA,V664XXA,V665XXA,V666XXA,V667XXA<br>V669XXA,V670XXA,V671XXA,V672XXA,V673XXA,V674XXA,V675XXA<br>V676XXA,V677XXA,V679XXA,V680XXA,V681XXA,V682XXA,V683XXA<br>V684XXA,V685XXA,V686XXA,V687XXA,V689XXA,V6900XA,V6909XA<br>V6910XA,V6919XA,V6920XA,V6929XA,V693XXA,V6940XA,V6949XA<br>V6950XA,V6959XA,V6960XA,V6969XA,V6981XA,V6988XA,V699XXA<br>V700XXA,V701XXA,V702XXA,V703XXA,V704XXA,V705XXA,V706XXA<br>V707XXA,V709XXA,V710XXA,V711XXA,V712XXA,V713XXA,V714XXA<br>V715XXA,V716XXA,V717XXA,V719XXA,V720XXA,V721XXA,V722XXA<br>V723XXA,V724XXA,V725XXA,V726XXA,V727XXA,V729XXA,V730XXA<br>V731XXA,V732XXA,V733XXA,V734XXA,V735XXA,V736XXA,V737XXA<br>V739XXA,V740XXA,V741XXA,V742XXA,V743XXA,V744XXA,V745XXA<br>V746XXA,V747XXA,V749XXA,V750XXA,V751XXA,V752XXA,V753XXA<br>V754XXA,V755XXA,V756XXA,V757XXA,V759XXA,V760XXA,V761XXA<br>V762XXA,V763XXA,V764XXA,V765XXA,V766XXA,V767XXA,V769XXA<br>V770XXA,V771XXA,V772XXA,V773XXA,V774XXA,V775XXA,V776XXA<br>V777XXA,V779XXA,V780XXA,V781XXA,V782XXA,V783XXA,V784XXA<br>V785XXA,V786XXA,V787XXA,V789XXA,V7900XA,V7909XA,V7910XA<br>V7919XA,V7920XA,V7929XA,V793XXA,V7940XA,V7949XA,V7950XA<br>V7959XA,V7960XA,V7969XA,V7981XA,V7988XA,V799XXA,V80010A<br>V80018A,V8002XA,V8011XA,V8012XA,V8021XA,V8022XA,V8031XA<br>V8032XA,V8041XA,V8042XA,V8051XA,V8052XA,V8061XA,V8062XA<br>V80710A,V80711A,V80720A,V80721A,V80730A,V80731A,V80790A<br>V80791A,V8081XA,V8082XA,V80910A,V80918A,V80919A,V80920A<br>V80928A,V80929A,V810XXA,V811XXA,V812XXA,V813XXA,V814XXA<br>V815XXA,V816XXA,V817XXA,V8181XA,V8182XA,V8183XA,V8189XA<br>V819XXA,V820XXA,V821XXA,V822XXA,V823XXA,V824XXA,V825XXA<br>V826XXA,V827XXA,V828XXA,V829XXA,V830XXA,V831XXA,V832XXA<br>V833XXA,V834XXA,V835XXA,V836XXA,V837XXA,V839XXA,V840XXA<br>V841XXA,V842XXA,V843XXA,V844XXA,V845XXA,V846XXA,V847XXA | |

Continued on next page

**Supplementary Table S1:** (cont'd) A comprehensive mapping of diagnosis groups used in the experiments to their corresponding ICD-10 and ICD-9 codes.

| Diagnosis | ICD-10 | ICD-9 |
|---|---|---|
| | V849XXA,V850XXA,V851XXA,V852XXA,V853XXA,V854XXA,V855XXA | |
| | V856XXA,V857XXA,V859XXA,V8601XA,V8602XA,V8603XA,V8604XA | |
| | V8605XA,V8606XA,V8609XA,V8611XA,V8612XA,V8613XA,V8614XA | |
| | V8615XA,V8616XA,V8619XA,V8621XA,V8622XA,V8623XA,V8624XA | |
| | V8625XA,V8626XA,V8629XA,V8631XA,V8632XA,V8633XA,V8634XA | |
| | V8635XA,V8636XA,V8639XA,V8641XA,V8642XA,V8643XA,V8644XA | |
| | V8645XA,V8646XA,V8649XA,V8651XA,V8652XA,V8653XA,V8654XA | |
| | V8655XA,V8656XA,V8659XA,V8661XA,V8662XA,V8663XA,V8664XA | |
| | V8665XA,V8666XA,V8669XA,V8671XA,V8672XA,V8673XA,V8674XA | |
| | V8675XA,V8676XA,V8679XA,V8691XA,V8692XA,V8693XA,V8694XA | |
| | V8695XA,V8696XA,V8699XA,V870XXA,V871XXA,V872XXA,V873XXA | |
| | V874XXA,V875XXA,V876XXA,V877XXA,V878XXA,V879XXA,V880XXA | |
| | V881XXA,V882XXA,V883XXA,V884XXA,V885XXA,V886XXA,V887XXA | |
| | V888XXA,V889XXA,V890XXA,V891XXA,V892XXA,V893XXA,V899XXA | |
| | V9000XA,V9001XA,V9002XA,V9003XA,V9004XA,V9005XA,V9006XA | |
| | V9008XA,V9009XA,V9010XA,V9011XA,V9012XA,V9013XA,V9014XA | |
| | V9015XA,V9016XA,V9018XA,V9019XA,V9020XA,V9021XA,V9022XA | |
| | V9023XA,V9024XA,V9025XA,V9026XA,V9027XA,V9028XA,V9029XA | |
| | V9030XA,V9031XA,V9032XA,V9033XA,V9034XA,V9035XA,V9036XA | |
| | V9037XA,V9038XA,V9039XA,V9080XA,V9081XA,V9082XA,V9083XA | |
| | V9084XA,V9085XA,V9086XA,V9087XA,V9088XA,V9089XA,V9100XA | |
| | V9101XA,V9102XA,V9103XA,V9104XA,V9105XA,V9106XA,V9107XA | |
| | V9108XA,V9109XA,V9110XA,V9111XA,V9112XA,V9113XA,V9114XA | |
| | V9115XA,V9116XA,V9118XA,V9119XA,V9120XA,V9121XA,V9122XA | |
| | V9123XA,V9124XA,V9125XA,V9126XA,V9129XA,V9130XA,V9131XA | |
| | V9132XA,V9133XA,V9134XA,V9135XA,V9136XA,V9137XA,V9138XA | |
| | V9139XA,V9180XA,V9181XA,V9182XA,V9183XA,V9184XA,V9185XA | |
| | V9186XA,V9187XA,V9188XA,V9189XA,V9200XA,V9201XA,V9202XA | |
| | V9203XA,V9204XA,V9205XA,V9206XA,V9207XA,V9208XA,V9209XA | |
| | V9210XA,V9211XA,V9212XA,V9213XA,V9214XA,V9215XA,V9216XA | |
| | V9219XA,V9220XA,V9221XA,V9222XA,V9223XA,V9224XA,V9225XA | |
| | V9226XA,V9227XA,V9228XA,V9229XA,V9300XA,V9301XA,V9302XA | |
| | V9303XA,V9304XA,V9309XA,V9310XA,V9311XA,V9312XA,V9313XA | |
| | V9314XA,V9319XA,V9320XA,V9321XA,V9322XA,V9323XA,V9324XA | |
| | V9329XA,V9330XA,V9331XA,V9332XA,V9333XA,V9334XA,V9335XA | |
| | V9336XA,V9338XA,V9339XA,V9340XA,V9341XA,V9342XA,V9343XA | |
| | V9344XA,V9348XA,V9349XA,V9350XA,V9351XA,V9352XA,V9353XA | |
| | V9354XA,V9359XA,V9360XA,V9361XA,V9362XA,V9363XA,V9364XA | |
| | V9369XA,V9380XA,V9381XA,V9382XA,V9383XA,V9384XA,V9385XA | |
| | V9386XA,V9387XA,V9388XA,V9389XA,V940XXA,V9411XA,V9412XA | |

Continued on next page

**Supplementary Table S1:** (cont'd) A comprehensive mapping of diagnosis groups used in the experiments to their corresponding ICD-10 and ICD-9 codes.

| Diagnosis | ICD-10 | ICD-9 |
|---|---|---|
| | V9421XA,V9422XA,V9431XA,V9432XA,V944XXA,V94810A,V94811A | |
| | V94818A,V9489XA,V949XXA,V9500XA,V9501XA,V9502XA,V9503XA | |
| | V9504XA,V9505XA,V9509XA,V9510XA,V9511XA,V9512XA,V9513XA | |
| | V9514XA,V9515XA,V9519XA,V9520XA,V9521XA,V9522XA,V9523XA | |
| | V9524XA,V9525XA,V9529XA,V9530XA,V9531XA,V9532XA,V9533XA | |
| | V9534XA,V9535XA,V9539XA,V9540XA,V9541XA,V9542XA,V9543XA | |
| | V9544XA,V9545XA,V9549XA,V958XXA,V959XXA,V9600XA,V9601XA | |
| | V9602XA,V9603XA,V9604XA,V9605XA,V9609XA,V9610XA,V9611XA | |
| | V9612XA,V9613XA,V9614XA,V9615XA,V9619XA,V9620XA,V9621XA | |
| | V9622XA,V9623XA,V9624XA,V9625XA,V9629XA,V968XXA,V969XXA | |
| | V970XXA,V971XXA,V9721XA,V9722XA,V9729XA,V9731XA,V9732XA | |
| | V9733XA,V9739XA,V97810A,V97811A,V97818A,V9789XA,V980XXA | |
| | V981XXA,V982XXA,V983XXA,V988XXA,V99XXXA,W000XXA,W001XXA | |
| | W002XXA,W009XXA,W010XXA,W0110XA,W01110A,W01111A,W01118A | |
| | W01119A,W01190A,W01198A,W03XXXA,W04XXXA,W050XXA,W051XXA | |
| | W052XXA,W06XXXA,W07XXXA,W08XXXA,W090XXA,W091XXA,W092XXA | |
| | W098XXA,W100XXA,W101XXA,W102XXA,W108XXA,W109XXA,W11XXXA | |
| | W12XXXA,W130XXA,W131XXA,W132XXA,W133XXA,W134XXA,W138XXA | |
| | W139XXA,W14XXXA,W15XXXA,W16011A,W16012A,W16021A,W16022A | |
| | W16031A,W16032A,W16111A,W16112A,W16121A,W16122A,W16131A | |
| | W16132A,W16211A,W16212A,W16221A,W16222A,W16311A,W16312A | |
| | W16321A,W16322A,W16331A,W16332A,W1641XA,W1642XA,W16511A | |
| | W16512A,W16521A,W16522A,W16531A,W16532A,W16611A,W16612A | |
| | W16621A,W16622A,W16711A,W16712A,W16721A,W16722A,W16811A | |
| | W16812A,W16821A,W16822A,W16831A,W16832A,W1691XA,W1692XA | |
| | W170XXA,W171XXA,W172XXA,W173XXA,W174XXA,W1781XA,W1782XA | |
| | W1789XA,W1800XA,W1801XA,W1802XA,W1809XA,W1811XA,W1812XA | |
| | W182XXA,W1830XA,W1831XA,W1839XA,W1840XA,W1841XA,W1842XA | |
| | W1843XA,W1849XA,W19XXXA,W200XXA,W201XXA,W208XXA,W2100XA | |
| | W2101XA,W2102XA,W2103XA,W2104XA,W2105XA,W2106XA,W2107XA | |
| | W2109XA,W2111XA,W2112XA,W2113XA,W2119XA,W21210A,W21211A | |
| | W21220A,W21221A,W2131XA,W2132XA,W2139XA,W214XXA,W2181XA | |
| | W2189XA,W219XXA,W2201XA,W2202XA,W2203XA,W22041A,W22042A | |
| | W2209XA,W2210XA,W2211XA,W2212XA,W2219XA,W228XXA,W230XXA | |
| | W231XXA,W232XXA,W240XXA,W241XXA,W25XXXA,W260XXA,W261XXA | |
| | W262XXA,W268XXA,W269XXA,W270XXA,W271XXA,W272XXA,W273XXA | |
| | W274XXA,W275XXA,W278XXA,W28XXXA,W290XXA,W291XXA,W292XXA | |
| | W293XXA,W294XXA,W298XXA,W300XXA,W301XXA,W302XXA,W303XXA | |
| | W3081XA,W3089XA,W309XXA,W310XXA,W311XXA,W312XXA,W313XXA | |
| | W3181XA,W3182XA,W3183XA,W3189XA,W319XXA,W320XXA,W321XXA | |

Continued on next page

**Supplementary Table S1:** (cont'd) A comprehensive mapping of diagnosis groups used in the experiments to their corresponding ICD-10 and ICD-9 codes.

| Diagnosis | ICD-10 | ICD-9 |
|---|---|---|
| | W3300XA,W3301XA,W3302XA,W3303XA,W3309XA,W3310XA,W3311XA | |
| | W3312XA,W3313XA,W3319XA,W3400XA,W34010A,W34011A,W34018A | |
| | W3409XA,W3410XA,W34110A,W34111A,W34118A,W3419XA,W35XXXA | |
| | W361XXA,W362XXA,W363XXA,W368XXA,W369XXA,W370XXA,W378XXA | |
| | W38XXXA,W39XXXA,W400XXA,W401XXA,W408XXA,W409XXA,W420XXA | |
| | W429XXA,W450XXA,W451XXA,W452XXA,W458XXA,W460XXA,W461XXA | |
| | W4901XA,W4902XA,W4903XA,W4904XA,W4909XA,W499XXA,W500XXA | |
| | W501XXA,W502XXA,W503XXA,W504XXA,W51XXXA,W52XXXA,W5301XA | |
| | W5309XA,W5311XA,W5319XA,W5321XA,W5329XA,W5381XA,W5389XA | |
| | W540XXA,W541XXA,W548XXA,W5501XA,W5503XA,W5509XA,W5511XA | |
| | W5512XA,W5519XA,W5521XA,W5522XA,W5529XA,W5531XA,W5532XA | |
| | W5539XA,W5541XA,W5542XA,W5549XA,W5551XA,W5552XA,W5559XA | |
| | W5581XA,W5582XA,W5589XA,W5601XA,W5602XA,W5609XA,W5611XA | |
| | W5612XA,W5619XA,W5621XA,W5622XA,W5629XA,W5631XA,W5632XA | |
| | W5639XA,W5641XA,W5642XA,W5649XA,W5651XA,W5652XA,W5659XA | |
| | W5681XA,W5682XA,W5689XA,W57XXXA,W5801XA,W5802XA,W5803XA | |
| | W5809XA,W5811XA,W5812XA,W5813XA,W5819XA,W5901XA,W5902XA | |
| | W5909XA,W5911XA,W5912XA,W5913XA,W5919XA,W5921XA,W5922XA | |
| | W5929XA,W5981XA,W5982XA,W5983XA,W5989XA,W60XXXA,W6101XA | |
| | W6102XA,W6109XA,W6111XA,W6112XA,W6119XA,W6121XA,W6122XA | |
| | W6129XA,W6132XA,W6133XA,W6139XA,W6142XA,W6143XA,W6149XA | |
| | W6151XA,W6152XA,W6159XA,W6161XA,W6162XA,W6169XA,W6191XA | |
| | W6192XA,W6199XA,W620XXA,W621XXA,W629XXA,W64XXXA,W65XXXA | |
| | W67XXXA,W69XXXA,W73XXXA,W74XXXA,W85XXXA,W860XXA,W861XXA | |
| | W868XXA,W880XXA,W881XXA,W888XXA,W890XXA,W891XXA,W898XXA | |
| | W899XXA,W900XXA,W901XXA,W902XXA,W908XXA,W92XXXA,W9301XA | |
| | W9302XA,W9311XA,W9312XA,W932XXA,W938XXA,W940XXA,W9411XA | |
| | W9412XA,W9421XA,W9422XA,W9423XA,W9429XA,W9431XA,W9432XA | |
| | W9439XA,W99XXXA,X000XXA,X001XXA,X002XXA,X003XXA,X004XXA | |
| | X005XXA,X008XXA,X010XXA,X011XXA,X013XXA,X014XXA,X018XXA | |
| | X020XXA,X021XXA,X022XXA,X023XXA,X024XXA,X025XXA,X028XXA | |
| | X030XXA,X031XXA,X033XXA,X034XXA,X038XXA,X04XXXA,X05XXXA | |
| | X060XXA,X061XXA,X062XXA,X063XXA,X0800XA,X0801XA,X0809XA | |
| | X0810XA,X0811XA,X0819XA,X0820XA,X0821XA,X0829XA,X088XXA | |
| | X100XXA,X101XXA,X102XXA,X110XXA,X111XXA,X118XXA,X12XXXA | |
| | X130XXA,X131XXA,X140XXA,X141XXA,X150XXA,X151XXA,X152XXA | |
| | X153XXA,X158XXA,X16XXXA,X17XXXA,X18XXXA,X19XXXA,X30XXXA | |
| | X31XXXA,X32XXXA,X34XXXA,X35XXXA,X360XXA,X361XXA,X370XXA | |
| | X371XXA,X372XXA,X373XXA,X3741XA,X3742XA,X3743XA,X378XXA | |
| | X379XXA,X38XXXA,X3901XA,X3908XA,X398XXA,X500XXA,X501XXA | |

Continued on next page

**Supplementary Table S1:** (cont'd) A comprehensive mapping of diagnosis groups used in the experiments to their corresponding ICD-10 and ICD-9 codes.

| Diagnosis | ICD-10 | ICD-9 |
|---|---|---|
| | X503XXA,X509XXA,X52XXXA,X58XXXA | |
| External cause codes:<br>intent of injury,<br>assault | T360X3A,T361X3A,T362X3A,T363X3A,T364X3A,T365X3A,T366X3A<br>T367X3A,T368X3A,T3693XA,T370X3A,T371X3A,T372X3A,T373X3A<br>T374X3A,T375X3A,T378X3A,T3793XA,T380X3A,T381X3A,T382X3A<br>T383X3A,T384X3A,T385X3A,T386X3A,T387X3A,T38803A,T38813A<br>T38893A,T38903A,T38993A,T39013A,T39093A,T391X3A,T392X3A<br>T39313A,T39393A,T394X3A,T398X3A,T3993XA,T400X3A,T401X3A<br>T402X3A,T403X3A,T40413A,T40423A,T40493A,T404X3A,T405X3A<br>T40603A,T40693A,T40713A,T40723A,T407X3A,T408X3A,T40903A<br>T40993A,T410X3A,T411X3A,T41203A,T41293A,T413X3A,T4143XA<br>T415X3A,T420X3A,T421X3A,T422X3A,T423X3A,T424X3A,T425X3A<br>T426X3A,T4273XA,T428X3A,T43013A,T43023A,T431X3A,T43203A<br>T43213A,T43223A,T43293A,T433X3A,T434X3A,T43503A,T43593A<br>T43603A,T43613A,T43623A,T43633A,T43643A,T43653A,T43693A<br>T438X3A,T4393XA,T440X3A,T441X3A,T442X3A,T443X3A,T444X3A<br>T445X3A,T446X3A,T447X3A,T448X3A,T44903A,T44993A,T450X3A<br>T451X3A,T452X3A,T453X3A,T454X3A,T45513A,T45523A,T45603A<br>T45613A,T45623A,T45693A,T457X3A,T458X3A,T4593XA,T460X3A<br>T461X3A,T462X3A,T463X3A,T464X3A,T465X3A,T466X3A,T467X3A<br>T468X3A,T46903A,T46993A,T470X3A,T471X3A,T472X3A,T473X3A<br>T474X3A,T475X3A,T476X3A,T477X3A,T478X3A,T4793XA,T480X3A<br>T481X3A,T48203A,T48293A,T483X3A,T484X3A,T485X3A,T486X3A<br>T48903A,T48993A,T490X3A,T491X3A,T492X3A,T493X3A,T494X3A<br>T495X3A,T496X3A,T497X3A,T498X3A,T4993XA,T500X3A,T501X3A<br>T502X3A,T503X3A,T504X3A,T505X3A,T506X3A,T507X3A,T508X3A<br>T50903A,T50913A,T50993A,T50A13A,T50A23A,T50A93A,T50B13A<br>T50B93A,T50Z13A,T50Z93A,T510X3A,T511X3A,T512X3A,T513X3A<br>T518X3A,T5193XA,T520X3A,T521X3A,T522X3A,T523X3A,T524X3A<br>T528X3A,T5293XA,T530X3A,T531X3A,T532X3A,T533X3A,T534X3A<br>T535X3A,T536X3A,T537X3A,T5393XA,T540X3A,T541X3A,T542X3A<br>T543X3A,T5493XA,T550X3A,T551X3A,T560X3A,T561X3A,T562X3A<br>T563X3A,T564X3A,T565X3A,T566X3A,T567X3A,T56813A,T56893A<br>T5693XA,T570X3A,T571X3A,T572X3A,T573X3A,T578X3A,T5793XA<br>T5803XA,T5813XA,T582X3A,T588X3A,T5893XA,T590X3A,T591X3A<br>T592X3A,T593X3A,T594X3A,T595X3A,T596X3A,T597X3A,T59813A<br>T59893A,T5993XA,T600X3A,T601X3A,T602X3A,T603X3A,T604X3A<br>T608X3A,T6093XA,T6103XA,T6113XA,T61773A,T61783A,T618X3A<br>T6193XA,T620X3A,T621X3A,T622X3A,T628X3A,T6293XA,T63003A<br>T63013A,T63023A,T63033A,T63043A,T63063A,T63073A,T63083A | 9600,9601,9602,9603,9604,9605,9606,9608,9609,9610,9614,9615<br>9616,9617,9618,9619,9620,9621,9622,9623,9624,9627,9628,9629<br>9630,9631,9634,9635,9638,9639,9640,9642,9643,9644,9645,9646<br>9648,9649,96500,96501,96502,96509,9651,9654,9655,96561,96569<br>9657,9659,9660,9661,9662,9663,9664,9670,9678,9679,9680,9682<br>9683,9684,9685,9689,96900,96901,96902,96903,96904,96905,96909<br>9691,9692,9693,9694,9696,96970,96971,96972,96973,96979,9698<br>9699,9700,9701,97081,9710,9711,9712,9713,9719,9720,9721,9722<br>9723,9724,9725,9726,9727,9729,9730,9731,9732,9733,9734,9735<br>9736,9738,9739,9742,9744,9745,9747,9750,9752,9753,9754,9755<br>9756,9757,9758,9760,9761,9762,9763,9764,9765,9766,9767,9768<br>9769,9770,9772,9778,9779,9786,9788,9789,9790,9796,9799,9800<br>9801,9802,9803,9808,9809,981,9820,9821,9822,9823,9824,9828<br>9830,9831,9832,9839,9849,9850,9851,9852,9853,9855,9856,9858<br>9859,986,9871,9872,9873,9874,9875,9876,9877,9878,9879,9880<br>9881,9882,9888,9889,9890,9891,9893,9894,9895,9896,9897,98982<br>98984,98989,9899,9947,99550,99551,99552,99553,99554,99555<br>99580,99581,99582,99583,99584,E9040,E9600,E961,E9620,E9621<br>E9622,E9629,E963,E964,E9650,E9651,E9652,E9653,E9654,E9655<br>E9656,E9657,E9658,E9659,E966,E9670,E9672,E9673,E9675,E9677<br>E9678,E9679,E9680,E9681,E9682,E9683,E9684,E9685,E9686,E9687<br>E9688,E9689,E9790,E9791,E9792,E9793,E9794,E9795,E9796,E9797<br>E9798,E9799 |

Continued on next page

**Supplementary Table S1:** (cont'd) A comprehensive mapping of diagnosis groups used in the experiments to their corresponding ICD-10 and ICD-9 codes.

| Diagnosis | ICD-10 | ICD-9 |
|---|---|---|
| | T63093A,T63113A,T63123A,T63193A,T632X3A,T63303A,T63313A | |
| | T63323A,T63333A,T63393A,T63413A,T63423A,T63433A,T63443A | |
| | T63453A,T63463A,T63483A,T63513A,T63593A,T63613A,T63623A | |
| | T63633A,T63693A,T63713A,T63793A,T63813A,T63823A,T63833A | |
| | T63893A,T6393XA,T6403XA,T6483XA,T650X3A,T651X3A,T65213A | |
| | T65223A,T65293A,T653X3A,T654X3A,T655X3A,T656X3A,T65813A | |
| | T65823A,T65833A,T65893A,T6593XA,T71113A,T71123A,T71133A | |
| | T71143A,T71153A,T71163A,T71193A,T71223A,T71233A,T7401XA | |
| | T7402XA,T7411XA,T7412XA,T7421XA,T7422XA,T7431XA,T7432XA | |
| | T744XXA,T7451XA,T7452XA,T7461XA,T7462XA,T7491XA,T7492XA | |
| | T7601XA,T7602XA,T7611XA,T7612XA,T7621XA,T7622XA,T7631XA | |
| | T7632XA,T7651XA,T7652XA,T7661XA,T7662XA,T7691XA,T7692XA | |
| | X920XXA,X921XXA,X922XXA,X923XXA,X928XXA,X929XXA,X93XXXA | |
| | X940XXA,X941XXA,X942XXA,X948XXA,X949XXA,X9501XA,X9502XA | |
| | X9509XA,X958XXA,X959XXA,X960XXA,X961XXA,X962XXA,X963XXA | |
| | X964XXA,X968XXA,X969XXA,X97XXXA,X980XXA,X981XXA,X982XXA | |
| | X983XXA,X988XXA,X989XXA,X990XXA,X991XXA,X992XXA,X998XXA | |
| | X999XXA,Y00XXXA,Y01XXXA,Y020XXA,Y021XXA,Y028XXA,Y030XXA | |
| | Y038XXA,Y040XXA,Y041XXA,Y042XXA,Y048XXA,Y0701,Y0702,Y0703 | |
| | Y0704,Y0711,Y0712,Y0713,Y0714,Y07410,Y07411,Y07420,Y07421 | |
| | Y07430,Y07432,Y07433,Y07434,Y07435,Y07436,Y07490,Y07491 | |
| | Y07499,Y0750,Y07510,Y07511,Y07512,Y07513,Y07519,Y07521,Y07528 | |
| | Y07529,Y0753,Y0759,Y076,Y079,Y0801XA,Y0802XA,Y0809XA,Y0881XA | |
| | Y0889XA,Y09,Y380X1A,Y380X2A,Y380X3A,Y381X1A,Y381X2A,Y381X3A | |
| | Y382X1A,Y382X2A,Y382X3A,Y383X1A,Y383X2A,Y383X3A,Y384X1A | |
| | Y384X2A,Y384X3A,Y385X1A,Y385X2A,Y385X3A,Y386X1A,Y386X2A | |
| | Y386X3A,Y387X1A,Y387X2A,Y387X3A,Y3880XA,Y38811A,Y38812A | |
| | Y38891A,Y38892A,Y38893A,Y389X1A,Y389X2A | |
| External cause codes: | Y35001A,Y35002A,Y35003A,Y35009A,Y35011A,Y35012A,Y35013A | E970,E971,E972,E973,E974,E975,E976,E9900,E9901,E9902,E9903 |
| intent of injury, | Y35019A,Y35021A,Y35022A,Y35023A,Y35029A,Y35031A,Y35032A | E9909,E9910,E9911,E9912,E9913,E9914,E9915,E9916,E9917,E9918 |
| legal intervention | Y35033A,Y35039A,Y35041A,Y35042A,Y35043A,Y35049A,Y35091A | E9919,E9920,E9921,E9922,E9923,E9928,E9929,E9930,E9931,E9932 |
| war | Y35092A,Y35093A,Y35099A,Y35101A,Y35102A,Y35103A,Y35109A | E9933,E9934,E9935,E9936,E9937,E9938,E9939,E9940,E9941,E9942 |
| | Y35111A,Y35112A,Y35113A,Y35119A,Y35121A,Y35122A,Y35123A | E9943,E9948,E9949,E9950,E9951,E9952,E9953,E9954,E9958,E9959 |
| | Y35129A,Y35191A,Y35192A,Y35193A,Y35199A,Y35201A,Y35202A | E9960,E9961,E9962,E9963,E9968,E9969,E9970,E9971,E9972,E9973 |
| | Y35203A,Y35209A,Y35211A,Y35212A,Y35213A,Y35219A,Y35291A | E9978,E9979,E9980,E9981,E9988,E9989 |
| | Y35292A,Y35293A,Y35299A,Y35301A,Y35302A,Y35303A,Y35309A | |
| | Y35311A,Y35312A,Y35313A,Y35319A,Y35391A,Y35392A,Y35393A | |
| | Y35399A,Y35401A,Y35402A,Y35403A,Y35409A,Y35411A,Y35412A | |
| | Y35413A,Y35419A,Y35491A,Y35492A,Y35493A,Y35499A,Y35811A | |

Continued on next page

**Supplementary Table S1:** (cont'd) A comprehensive mapping of diagnosis groups used in the experiments to their corresponding ICD-10 and ICD-9 codes.

| Diagnosis | ICD-10 | ICD-9 |
|---|---|---|
| | Y35812A,Y35813A,Y35819A,Y35831A,Y35832A,Y35833A,Y35839A | |
| | Y35891A,Y35892A,Y35893A,Y35899A,Y3591XA,Y3592XA,Y3593XA | |
| | Y3599XA,Y36000A,Y36001A,Y36010A,Y36011A,Y36020A,Y36021A | |
| | Y36030A,Y36031A,Y36040A,Y36041A,Y36050A,Y36051A,Y36090A | |
| | Y36091A,Y36100A,Y36101A,Y36110A,Y36111A,Y36120A,Y36121A | |
| | Y36130A,Y36131A,Y36140A,Y36141A,Y36190A,Y36191A,Y36200A | |
| | Y36201A,Y36210A,Y36211A,Y36220A,Y36221A,Y36230A,Y36231A | |
| | Y36240A,Y36241A,Y36250A,Y36251A,Y36260A,Y36261A,Y36270A | |
| | Y36271A,Y36290A,Y36291A,Y36300A,Y36301A,Y36310A,Y36311A | |
| | Y36320A,Y36321A,Y36330A,Y36331A,Y36390A,Y36391A,Y36410A | |
| | Y36411A,Y36420A,Y36421A,Y36430A,Y36431A,Y36440A,Y36441A | |
| | Y36450A,Y36451A,Y36460A,Y36461A,Y36470A,Y36471A,Y36490A | |
| | Y36491A,Y36500A,Y36501A,Y36510A,Y36511A,Y36520A,Y36521A | |
| | Y36530A,Y36531A,Y36540A,Y36541A,Y36590A,Y36591A,Y366X0A | |
| | Y366X1A,Y367X0A,Y367X1A,Y36810A,Y36811A,Y36820A,Y36821A | |
| | Y36880A,Y36881A,Y36890A,Y36891A,Y3690XA,Y3691XA,Y3692XA | |
| | Y37000A,Y37001A,Y37010A,Y37011A,Y37020A,Y37021A,Y37030A | |
| | Y37031A,Y37040A,Y37041A,Y37050A,Y37051A,Y37090A,Y37091A | |
| | Y37100A,Y37101A,Y37110A,Y37111A,Y37120A,Y37121A,Y37130A | |
| | Y37131A,Y37140A,Y37141A,Y37190A,Y37191A,Y37200A,Y37201A | |
| | Y37210A,Y37211A,Y37220A,Y37221A,Y37230A,Y37231A,Y37240A | |
| | Y37241A,Y37250A,Y37251A,Y37260A,Y37261A,Y37270A,Y37271A | |
| | Y37290A,Y37291A,Y37300A,Y37301A,Y37310A,Y37311A,Y37320A | |
| | Y37321A,Y37330A,Y37331A,Y37390A,Y37391A,Y37410A,Y37411A | |
| | Y37420A,Y37421A,Y37430A,Y37431A,Y37440A,Y37441A,Y37450A | |
| | Y37451A,Y37460A,Y37461A,Y37470A,Y37471A,Y37490A,Y37491A | |
| | Y37500A,Y37501A,Y37510A,Y37511A,Y37520A,Y37521A,Y37530A | |
| | Y37531A,Y37540A,Y37541A,Y37590A,Y37591A,Y376X0A,Y376X1A | |
| | Y377X0A,Y377X1A,Y3790XA,Y3791XA,Y3792XA | |
| External cause codes: | R4588,T1491,T1491XA,T360X2A,T361X2A,T362X2A,T363X2A,T364X2A | 9599,9600,9601,9602,9603,9604,9605,9606,9608,9609,9610,9614 |
| intent of injury, | T365X2A,T366X2A,T367X2A,T368X2A,T3692XA,T370X2A,T371X2A | 9615,9616,9617,9618,9619,9620,9621,9622,9623,9624,9627,9628 |
| self-harm | T372X2A,T373X2A,T374X2A,T375X2A,T378X2A,T3792XA,T380X2A | 9629,9630,9631,9634,9635,9638,9639,9640,9642,9643,9644,9645 |
| | T381X2A,T382X2A,T383X2A,T384X2A,T385X2A,T386X2A,T387X2A | 9646,9648,9649,96500,96501,96502,96509,9651,9654,9655,96561 |
| | T38802A,T38812A,T38892A,T38902A,T38992A,T39012A,T39092A | 96569,9657,9659,9660,9661,9662,9663,9664,9670,9678,9679,9680 |
| | T391X2A,T392X2A,T39312A,T39392A,T394X2A,T398X2A,T3992XA | 9682,9683,9684,9685,9689,96900,96901,96902,96903,96904,96905 |
| | T400X2A,T401X2A,T402X2A,T403X2A,T40412A,T40422A,T40492A | 96909,9691,9692,9693,9694,9696,96970,96971,96972,96973,96979 |
| | T404X2A,T405X2A,T40602A,T40692A,T40712A,T40722A,T407X2A | 9698,9699,9700,9701,97081,9710,9711,9712,9713,9719,9720,9721 |
| | T408X2A,T40902A,T40992A,T410X2A,T411X2A,T41202A,T41292A | 9722,9723,9724,9725,9726,9727,9729,9730,9731,9732,9733,9734 |
| | T413X2A,T4142XA,T415X2A,T420X2A,T421X2A,T422X2A,T423X2A | 9735,9736,9738,9739,9742,9744,9745,9747,9750,9752,9753,9754 |

Continued on next page

**Supplementary Table S1:** (cont'd) A comprehensive mapping of diagnosis groups used in the experiments to their corresponding ICD-10 and ICD-9 codes.

| Diagnosis | ICD-10 | ICD-9 |
|---|---|---|
| | T424X2A,T425X2A,T426X2A,T4272XA,T428X2A,T43012A,T43022A | 9755,9756,9757,9758,9760,9761,9762,9763,9764,9765,9766,9767 |
| | T431X2A,T43202A,T43212A,T43222A,T43292A,T433X2A,T434X2A | 9768,9769,9770,9772,9778,9779,9786,9788,9789,9790,9796,9799 |
| | T43502A,T43592A,T43602A,T43612A,T43622A,T43632A,T43642A | 9800,9801,9802,9803,9808,9809,981,9820,9821,9822,9823,9824 |
| | T43652A,T43692A,T438X2A,T4392XA,T440X2A,T441X2A,T442X2A | 9828,9830,9831,9832,9839,9849,9850,9851,9852,9853,9855,9856 |
| | T443X2A,T444X2A,T445X2A,T446X2A,T447X2A,T448X2A,T44902A | 9858,9859,986,9871,9872,9873,9874,9875,9876,9877,9878,9879 |
| | T44992A,T450X2A,T451X2A,T452X2A,T453X2A,T454X2A,T45512A | 9880,9881,9882,9888,9889,9890,9891,9893,9894,9895,9896,9897 |
| | T45522A,T45602A,T45612A,T45622A,T45692A,T457X2A,T458X2A | 98982,98984,98989,9899,9947,E9500,E9501,E9502,E9503,E9504 |
| | T4592XA,T460X2A,T461X2A,T462X2A,T463X2A,T464X2A,T465X2A | E9506,E9507,E9508,E9509,E9520,E9521,E9528,E9529,E9530,E9531 |
| | T466X2A,T467X2A,T468X2A,T46902A,T46992A,T470X2A,T471X2A | E9538,E954,E9550,E9551,E9552,E9553,E9554,E9555,E9556,E9557 |
| | T472X2A,T473X2A,T474X2A,T475X2A,T476X2A,T477X2A,T478X2A | E9559,E956,E9579,E9580,E9581,E9582,E9583,E9584,E9585,E9586 |
| | T4792XA,T480X2A,T481X2A,T48202A,T48292A,T483X2A,T484X2A | E9587,E9588,E9589 |
| | T485X2A,T486X2A,T48902A,T48992A,T490X2A,T491X2A,T492X2A | |
| | T493X2A,T494X2A,T495X2A,T496X2A,T497X2A,T498X2A,T4992XA | |
| | T500X2A,T501X2A,T502X2A,T503X2A,T504X2A,T505X2A,T506X2A | |
| | T507X2A,T508X2A,T50902A,T50912A,T50992A,T50A12A,T50A22A | |
| | T50A92A,T50B12A,T50B92A,T50Z12A,T50Z92A,T510X2A,T511X2A | |
| | T512X2A,T513X2A,T518X2A,T5192XA,T520X2A,T521X2A,T522X2A | |
| | T523X2A,T524X2A,T528X2A,T5292XA,T530X2A,T531X2A,T532X2A | |
| | T533X2A,T534X2A,T535X2A,T536X2A,T537X2A,T5392XA,T540X2A | |
| | T541X2A,T542X2A,T543X2A,T5492XA,T550X2A,T551X2A,T560X2A | |
| | T561X2A,T562X2A,T563X2A,T564X2A,T565X2A,T566X2A,T567X2A | |
| | T56812A,T56892A,T5692XA,T570X2A,T571X2A,T572X2A,T573X2A | |
| | T578X2A,T5792XA,T5802XA,T5812XA,T582X2A,T588X2A,T5892XA | |
| | T590X2A,T591X2A,T592X2A,T593X2A,T594X2A,T595X2A,T596X2A | |
| | T597X2A,T59812A,T59892A,T5992XA,T600X2A,T601X2A,T602X2A | |
| | T603X2A,T604X2A,T608X2A,T6092XA,T6102XA,T6112XA,T61772A | |
| | T61782A,T618X2A,T6192XA,T620X2A,T621X2A,T622X2A,T628X2A | |
| | T6292XA,T63002A,T63012A,T63022A,T63032A,T63042A,T63062A | |
| | T63072A,T63082A,T63092A,T63112A,T63122A,T63192A,T632X2A | |
| | T63302A,T63312A,T63322A,T63332A,T63392A,T63412A,T63422A | |
| | T63432A,T63442A,T63452A,T63462A,T63482A,T63512A,T63592A | |
| | T63612A,T63622A,T63632A,T63692A,T63712A,T63792A,T63812A | |
| | T63822A,T63832A,T63892A,T6392XA,T6402XA,T6482XA,T650X2A | |
| | T651X2A,T65212A,T65222A,T65292A,T653X2A,T654X2A,T655X2A | |
| | T656X2A,T65812A,T65822A,T65832A,T65892A,T6592XA,T71112A | |
| | T71122A,T71132A,T71152A,T71162A,T71192A,T71222A,T71232A | |
| | X710XXA,X711XXA,X712XXA,X713XXA,X718XXA,X719XXA,X72XXXA | |
| | X730XXA,X731XXA,X732XXA,X738XXA,X739XXA,X7401XA,X7402XA | |
| | X7409XA,X748XXA,X749XXA,X75XXXA,X76XXXA,X770XXA,X771XXA | |
| | X772XXA,X773XXA,X778XXA,X779XXA,X780XXA,X781XXA,X782XXA | |

Continued on next page

**Supplementary Table S1:** (cont'd) A comprehensive mapping of diagnosis groups used in the experiments to their corresponding ICD-10 and ICD-9 codes.

| Diagnosis | ICD-10 | ICD-9 |
|---|---|---|
| | X788XXA,X789XXA,X79XXXA,X80XXXA,X810XXA,X811XXA,X818XXA | |
| | X820XXA,X821XXA,X822XXA,X828XXA,X830XXA,X831XXA,X832XXA | |
| | X838XXA | |
| External cause codes: | T360X4A,T361X4A,T362X4A,T363X4A,T364X4A,T365X4A,T366X4A | 9600,9601,9602,9603,9604,9605,9606,9608,9609,9610,9614,9615 |
| intent of injury, | T367X4A,T368X4A,T3694XA,T370X4A,T371X4A,T372X4A,T373X4A | 9616,9617,9618,9619,9620,9621,9622,9623,9624,9627,9628,9629 |
| undetermined | T374X4A,T375X4A,T378X4A,T3794XA,T380X4A,T381X4A,T382X4A | 9630,9631,9634,9635,9638,9639,9640,9642,9644,9645,9646,9648 |
| | T383X4A,T384X4A,T385X4A,T386X4A,T387X4A,T38804A,T38814A | 9649,96500,96501,96502,96509,9651,9654,9655,96561,96569,9657 |
| | T38894A,T38904A,T38994A,T39014A,T39094A,T391X4A,T392X4A | 9659,9660,9661,9662,9663,9664,9670,9678,9679,9680,9682,9683 |
| | T39314A,T39394A,T394X4A,T398X4A,T3994XA,T400X4A,T401X4A | 9684,9685,9689,96900,96901,96902,96903,96904,96905,96909,9691 |
| | T402X4A,T403X4A,T40414A,T40424A,T40494A,T404X4A,T405X4A | 9692,9693,9694,9696,96970,96971,96972,96973,96979,9698,9699 |
| | T40604A,T40694A,T40714A,T40724A,T407X4A,T408X4A,T40904A | 9700,9701,97081,9710,9711,9712,9713,9719,9720,9721,9722,9723 |
| | T40994A,T410X4A,T411X4A,T41204A,T41294A,T413X4A,T4144XA | 9724,9725,9726,9727,9729,9730,9731,9732,9733,9734,9735,9736 |
| | T415X4A,T420X4A,T421X4A,T422X4A,T423X4A,T424X4A,T425X4A | 9738,9739,9742,9744,9745,9747,9750,9752,9753,9754,9755,9756 |
| | T426X4A,T4274XA,T428X4A,T43014A,T43024A,T431X4A,T43204A | 9757,9758,9760,9761,9762,9763,9764,9765,9766,9767,9768,9769 |
| | T43214A,T43224A,T43294A,T433X4A,T434X4A,T43504A,T43594A | 9770,9772,9778,9779,9786,9788,9789,9790,9796,9799,9800,9801 |
| | T43604A,T43614A,T43624A,T43634A,T43644A,T43654A,T43694A | 9802,9803,9808,9809,981,9820,9821,9822,9823,9824,9828,9830 |
| | T438X4A,T4394XA,T440X4A,T441X4A,T442X4A,T443X4A,T444X4A | 9831,9832,9839,9849,9850,9851,9852,9853,9855,9856,9858,9859 |
| | T445X4A,T446X4A,T447X4A,T448X4A,T44904A,T44994A,T450X4A | 986,9871,9872,9873,9874,9875,9876,9877,9878,9879,9880,9881 |
| | T451X4A,T452X4A,T453X4A,T454X4A,T45514A,T45524A,T45604A | 9882,9888,9889,9890,9891,9893,9894,9895,9896,9897,98982,98984 |
| | T45614A,T45624A,T45694A,T457X4A,T458X4A,T4594XA,T460X4A | 98989,9899,9941,9947,E8585,E8698,E9104,E9108,E9241,E9286 |
| | T461X4A,T462X4A,T463X4A,T464X4A,T465X4A,T466X4A,T467X4A | E9502,E9504,E9800,E9801,E9802,E9803,E9804,E9805,E9806,E9807 |
| | T468X4A,T46904A,T46994A,T470X4A,T471X4A,T472X4A,T473X4A | E9808,E9809,E9820,E9821,E9828,E9830,E9831,E9838,E984,E9850 |
| | T474X4A,T475X4A,T476X4A,T477X4A,T478X4A,T4794XA,T480X4A | E9851,E9852,E9853,E9854,E9855,E9856,E9857,E986,E9879,E9880 |
| | T481X4A,T48204A,T48294A,T483X4A,T484X4A,T485X4A,T486X4A | E9881,E9882,E9885,E9888 |
| | T48904A,T48994A,T490X4A,T491X4A,T492X4A,T493X4A,T494X4A | |
| | T495X4A,T496X4A,T497X4A,T498X4A,T4994XA,T500X4A,T501X4A | |
| | T502X4A,T503X4A,T504X4A,T505X4A,T506X4A,T507X4A,T508X4A | |
| | T50904A,T50914A,T50994A,T50A14A,T50A24A,T50A94A,T50B14A | |
| | T50B94A,T50Z14A,T50Z94A,T510X4A,T511X4A,T512X4A,T513X4A | |
| | T518X4A,T5194XA,T520X4A,T521X4A,T522X4A,T523X4A,T524X4A | |
| | T528X4A,T5294XA,T530X4A,T531X4A,T532X4A,T533X4A,T534X4A | |
| | T535X4A,T536X4A,T537X4A,T5394XA,T540X4A,T541X4A,T542X4A | |
| | T543X4A,T5494XA,T550X4A,T551X4A,T560X4A,T561X4A,T562X4A | |
| | T563X4A,T564X4A,T565X4A,T566X4A,T567X4A,T56814A,T56894A | |
| | T5694XA,T570X4A,T571X4A,T572X4A,T573X4A,T578X4A,T5794XA | |
| | T5804XA,T5814XA,T582X4A,T588X4A,T5894XA,T590X4A,T591X4A | |
| | T592X4A,T593X4A,T594X4A,T595X4A,T596X4A,T597X4A,T59814A | |
| | T59894A,T5994XA,T600X4A,T601X4A,T602X4A,T603X4A,T604X4A | |
| | T608X4A,T6094XA,T6104XA,T6114XA,T61774A,T61784A,T618X4A | |

Continued on next page

**Supplementary Table S1:** (cont'd) A comprehensive mapping of diagnosis groups used in the experiments to their corresponding ICD-10 and ICD-9 codes.

| Diagnosis | ICD-10 | ICD-9 |
|---|---|---|
| | T6194XA,T620X4A,T621X4A,T622X4A,T628X4A,T6294XA,T63004A | |
| | T63014A,T63024A,T63034A,T63044A,T63064A,T63074A,T63084A | |
| | T63094A,T63114A,T63124A,T63194A,T632X4A,T63304A,T63314A | |
| | T63324A,T63334A,T63394A,T63414A,T63424A,T63434A,T63444A | |
| | T63454A,T63464A,T63484A,T63514A,T63594A,T63614A,T63624A | |
| | T63634A,T63694A,T63714A,T63794A,T63814A,T63824A,T63834A | |
| | T63894A,T6394XA,T6404XA,T6484XA,T650X4A,T651X4A,T65214A | |
| | T65224A,T65294A,T653X4A,T654X4A,T655X4A,T656X4A,T65814A | |
| | T65824A,T65834A,T65894A,T6594XA,T71114A,T71124A,T71134A | |
| | T71144A,T71154A,T71164A,T71194A,T71224A,T71234A,T751XXA | |
| | Y210XXA,Y211XXA,Y212XXA,Y213XXA,Y214XXA,Y218XXA,Y219XXA | |
| | Y22XXXA,Y230XXA,Y231XXA,Y232XXA,Y233XXA,Y238XXA,Y239XXA | |
| | Y240XXA,Y248XXA,Y249XXA,Y25XXXA,Y26XXXA,Y270XXA,Y271XXA | |
| | Y272XXA,Y273XXA,Y278XXA,Y279XXA,Y280XXA,Y281XXA,Y282XXA | |
| | Y288XXA,Y289XXA,Y29XXXA,Y30XXXA,Y31XXXA,Y32XXXA,Y33XXXA | |
| Hallucinogen-related | T408X1D,T408X2D,T408X3D,T408X4D,T40901D,T40902D,T40903D | V5889 |
| disorders; subsequent | T40904D,T40905D,T40991D,T40992D,T40993D,T40994D,T40995D | |
| encounter | T43641D,T43642D,T43643D,T43644D | |
| Inhalant-related | T410X1D,T410X2D,T410X3D,T410X4D,T410X5D | V5889 |
| disorders; subsequent | | |
| encounter | | |
| Nontraumatic dental | A690,A691,B002,B084,B370,B3783,K000,K001,K002,K003,K004,K005 | 0542,0743,101,1120,5200,5201,5202,5203,5204,5205,5206,5207 |
| conditions | K006,K007,K008,K009,K010,K011,K023,K0251,K0252,K0253,K0261 | 5208,5209,52100,52101,52102,52103,52104,52105,52106,52107 |
| | K0262,K0263,K027,K029,K030,K031,K032,K033,K034,K035,K036,K037 | 52108,52110,52120,52130,52140,52141,52142,5215,5216,5217 |
| | K0381,K0389,K039,K040,K0401,K0402,K041,K042,K043,K044,K045 | 52181,52189,5219,5220,5221,5222,5223,5224,5225,5226,5227,5228 |
| | K046,K047,K048,K0490,K0499,K0500,K0501,K0510,K0511,K0520 | 5229,52300,52301,52310,52311,52320,52330,52331,52332,52333 |
| | K0521,K05211,K05212,K05213,K05219,K0522,K05221,K05222,K05223 | 52340,52341,52342,5235,5236,5238,5239,52400,52401,52402,52403 |
| | K05229,K0530,K0531,K05311,K05312,K05313,K05319,K0532,K05321 | 52404,52405,52406,52407,52409,52410,52411,52412,52419,52420 |
| | K05322,K05323,K05329,K054,K055,K056,K060,K06010,K06011,K06012 | 52421,52422,52423,52424,52425,52426,52427,52428,52429,52430 |
| | K06013,K06020,K06021,K06022,K06023,K061,K063,K068,K069,K080 | 52431,52432,52433,52434,52435,52436,52437,52439,5244,52450 |
| | K08101,K08102,K08103,K08104,K08109,K08121,K08122,K08123 | 52451,52452,52453,52454,52455,52456,52457,52459,52460,52461 |
| | K08124,K08129,K08131,K08132,K08133,K08134,K08139,K08191 | 52462,52463,52469,52470,52471,52472,52473,52474,52479,52481 |
| | K08192,K08193,K08194,K08199,K0820,K0821,K0822,K0823,K0824 | 52482,52489,5249,5250,52510,52512,52513,52519,52520,52521 |
| | K0825,K0826,K083,K08401,K08402,K08403,K08404,K08409,K08421 | 52522,52523,52524,52525,52526,5253,52540,52541,52542,52543 |
| | K08422,K08423,K08424,K08429,K08431,K08432,K08433,K08434 | 52544,52550,52551,52552,52553,52554,52560,52561,52562,52563 |
| | K08439,K08491,K08492,K08493,K08494,K08499,K0850,K0851,K0852 | 52564,52565,52566,52567,52569,52571,52572,52573,52579,5258 |
| | K08530,K08531,K08539,K0854,K0855,K0856,K0859,K088,K0889,K089 | 5259,5260,5261,5262,5263,5264,5265,52661,52662,52663,52669 |

Continued on next page

**Supplementary Table S1:** (cont'd) A comprehensive mapping of diagnosis groups used in the experiments to their corresponding ICD-10 and ICD-9 codes.

| Diagnosis | ICD-10 | ICD-9 |
|---|---|---|
| | K090,K091,K098,K099,K110,K111,K1120,K1121,K1122,K1123,K113 K114,K115,K116,K117,K118,K119,K120,K121,K122,K1230,K1231 K1232,K1233,K1239,K130,K131,K1321,K1322,K1323,K1324,K1329 K133,K134,K135,K136,K1370,K1379,K140,K141,K142,K143,K144,K145 K146,K148,K149,M2600,M2601,M2602,M2603,M2604,M2605,M2606 M2607,M2609,M2610,M2611,M2612,M2619,M2620,M26211,M26212 M26213,M26219,M26220,M26221,M2623,M2624,M2625,M2629,M2630 M2631,M2632,M2633,M2634,M2635,M2636,M2637,M2639,M264,M2650 M2651,M2652,M2653,M2654,M2655,M2656,M2657,M2659,M2660,M26601 M26602,M26603,M26609,M2661,M26611,M26612,M26613,M26619,M2662 M26621,M26622,M26623,M26629,M2663,M26631,M26632,M26633,M26639 M26641,M26642,M26643,M26649,M26651,M26652,M26653,M26659,M2669 M2670,M2671,M2672,M2673,M2674,M2679,M2681,M2682,M2689,M269 M270,M271,M272,M273,M2740,M2749,M2751,M2752,M2753,M2759,M2761 M2762,M2763,M2769,M278,M279,M350C,M7911,R682,R6884,Z0120 Z0121,Z463,Z464 | 52681,52689,5269,5270,5271,5272,5273,5274,5275,5276,5277,5278 5279,52800,52801,52802,52809,5281,5282,5283,5284,5285,5286 52871,52872,52879,5288,5289,5290,5291,5292,5293,5294,5295 5296,5298,5299,6820,78492,V523,V534,V585,V722 |
| Opioid-related disorders; subsequent encounter | T400X1D,T400X2D,T400X3D,T400X4D,T400X5D,T401X1D,T401X2D T401X3D,T401X4D,T402X1D,T402X2D,T402X3D,T402X4D,T402X5D T403X1D,T403X2D,T403X3D,T403X4D,T403X5D,T40411D,T40412D T40413D,T40414D,T40415D,T40421D,T40422D,T40423D,T40424D T40425D,T40491D,T40492D,T40493D,T40494D,T40495D,T404X1D T404X2D,T404X3D,T404X4D,T404X5D,T40601D,T40602D,T40603D T40604D,T40605D,T40691D,T40692D,T40693D,T40694D,T40695D | V5889 |
| Sedative-related disorders; subsequent encounter | T426X1D,T426X2D,T426X3D,T426X4D,T426X5D,T4271XD,T4272XD T4273XD,T4274XD,T4275XD | V5889 |
| Stimulant-related disorders; subsequent encounter | T405X1D,T405X2D,T405X3D,T405X4D,T405X5D,T43601D,T43602D T43603D,T43604D,T43605D,T43611D,T43612D,T43613D,T43614D T43615D,T43621D,T43622D,T43623D,T43624D,T43625D,T43631D T43632D,T43633D,T43634D,T43635D,T43641D,T43642D,T43643D T43644D,T43651D,T43652D,T43653D,T43654D,T43655D,T43691D T43692D,T43693D,T43694D,T43695D | V5889 |
| Caries, periodontitis, and other preventable dental conditions | B002,B084,B370,B3783,K003,K007,K023,K0251,K0252,K0253,K0261 K0262,K0263,K027,K029,K030,K031,K032,K036,K037,K0381,K0389 K039,K040,K0401,K042,K044,K045,K0500,K0501,K0510,K0511,K0520 K0521,K05211,K05212,K05213,K05219,K0522,K05221,K05222,K05223 | 0542,0743,1120,5203,5207,52100,52101,52102,52103,52104,52105 52106,52107,52108,52110,52120,52130,5217,52181,52189,5219 5220,5222,5224,5226,52300,52301,52310,52311,52320,52330,52331 52332,52333,52340,52341,52342,5235,5236,5238,5239,52510,52512 |

Continued on next page

**Supplementary Table S1:** (cont'd) A comprehensive mapping of diagnosis groups used in the experiments to their corresponding ICD-10 and ICD-9 codes.

| Diagnosis | ICD-10 | ICD-9 |
|---|---|---|
| | K05229,K0530,K0531,K05311,K05312,K05313,K05319,K0532,K05321 K05322,K05323,K05329,K054,K055,K056,K060,K06010,K06011,K06012 K06013,K06020,K06021,K06022,K06023,K061,K08101,K08102,K08103 K08104,K08109,K08121,K08122,K08123,K08124,K08129,K08131 K08132,K08133,K08134,K08139,K08191,K08192,K08193,K08194 K08199,K08401,K08402,K08403,K08404,K08409,K08421,K08422 K08423,K08424,K08429,K08431,K08432,K08433,K08434,K08439 K08491,K08492,K08493,K08494,K08499,K0850,K0851,K0852,K08530 K08531,K08539,K0854,K0855,K0856,K0859,K088,K0889,K089,M2751 M2752,M2753,M2759,M2761,M2762,M2763,M2769,M350C,R6884,Z0120 Z0121,Z463,Z464 | 52513,52519,52540,52541,52542,52543,52544,52550,52551,52552 52553,52554,52560,52561,52562,52563,52564,52565,52566,52567 52569,52571,52572,52573,52579,5258,5259,52661,52662,52663 52669,78492,V523,V534,V585,V722 |
| Chest pain | R070,R071,R0781,R0782,R0789,R079 | 7841,78650,78652,78659 |
| Neuropathic pain | M792 | 7292 |
| Pain | R52 | 33819,78096 |
| Pleuritic chest pain | R071 | 78652 |
| Unusual muscle pain | M791,M7910,M7911,M7912,M7918 | 7291 |
| Back pain | M5450,M5451,M549 | 7245 |
| Fatigue | G933,R5382,R5383 | 78071,78079 |
| Post exertional fatigue | T733XXA,T733XXD,T733XXS | 9094,9945,E9272,V5889 |
| Anosmia (Loss of Smell) | R430 | 7811 |
| Ageusia (Loss of Taste) | R432,R439 | 7811,V415 |
| Cough | R05,R051 ,R052,R053,R054,R058,R059 | 7862 |
| Dysguesia | R438 | 7811 |
| Ear pain | H9201,H9202,H9203,H9209 | 38870 |

Continued on next page

**Supplementary Table S1:** (cont'd) A comprehensive mapping of diagnosis groups used in the experiments to their corresponding ICD-10 and ICD-9 codes.

| Diagnosis | ICD-10 | ICD-9 |
|---|---|---|
| Hoarse voice | R490 | 78442 |
| Hyperacusis | H9323 | |
| Nasal congestion | R0981 | 47819 |
| Phlegm | R848 | 7929 |
| Sneezing | R067 | 78499 |
| Sore throat | J029 | 462 |
| Tinnitus | H931 | |
| Runny Nose | J3489 | 47819 |
| Lump in throat (Globus) | R0989 | 7859,7867 |
| Diseases of middle ear and mastoid (except otitis media) | H6500,H6501,H6502,H6503,H6504,H6505,H6506,H6507,H65111,H65112 H65113,H65114,H65115,H65116,H65117,H65119,H65191,H65192 H65193,H65194,H65195,H65196,H65197,H65199,H6520,H6521,H6522 H6523,H6530,H6531,H6532,H6533,H65411,H65412,H65413,H65419 H65491,H65492,H65493,H65499,H6590,H6591,H6592,H6593,H68001 H68002,H68003,H68009,H68011,H68012,H68013,H68019,H68021 H68022,H68023,H68029,H68101,H68102,H68103,H68109,H68111 H68112,H68113,H68119,H68121,H68122,H68123,H68129,H68131 H68132,H68133,H68139,H6900,H6901,H6902,H6903,H70001,H70002 H70003,H70009,H70011,H70012,H70013,H70019,H70091,H70092 H70093,H70099,H7010,H7011,H7012,H7013,H70201,H70202,H70203 H70209,H70211,H70212,H70213,H70219,H70221,H70222,H70223 H70229,H70811,H70812,H70813,H70819,H70891,H70892,H70893 H70899,H7090,H7091,H7092,H7093,H7100,H7101,H7102,H7103,H7110 H7111,H7112,H7113,H7120,H7121,H7122,H7123,H7130,H7131,H7132 H7133,H7190,H7191,H7192,H7193,H7200,H7201,H7202,H7203,H7210 H7211,H7212,H7213,H722X1,H722X2,H722X3,H722X9,H72811,H72812 H72813,H72819,H72821,H72822,H72823,H72829,H7290,H7291,H7292 H7293,H73001,H73002,H73003,H73009,H73011,H73012,H73013,H73019 H73091,H73092,H73093,H73099,H7401,H7402,H7403,H7409 | 38100,38101,38102,38103,38104,38105,38106,38110,38119,38120 38129,3813,3814,38150,38151,38152,38160,38161,38162,38163 3817,38300,38301,38302,3831,38320,38321,38322,38381,38389 3839,38400,38401,38409,38420,38421,38422,38423,38424,38425 38500,38501,38502,38503,38509,38530,38531,38532,38533,38535 |

Continued on next page

**Supplementary Table S1:** (cont'd) A comprehensive mapping of diagnosis groups used in the experiments to their corresponding ICD-10 and ICD-9 codes.

| Diagnosis | ICD-10 | ICD-9 |
|---|---|---|
| Stomache pain | K219,K259,K299,K30,K319,K5090,K5190,K589,R100,R1010,R1011 R1012,R1013,R102,R1030,R1031,R1032,R1033,R10811,R10812,R10813 R10814,R10815,R10816,R10817,R10819,R10821,R10822,R10823 R10824,R10825,R10826,R10827,R10829,R1083,R1084,R109 | 53081,53190,5368,5369,5379,5559,5569,5641,6089,6259,78900 78901,78902,78903,78904,78905,78906,78907,78909,78960,78961 78962,78963,78964,78965,78966,78967,78969,7897 |
| Bloating | R140 | 7873 |
| Bowel incontinence | R150,R151,R152,R159 | 78760,78761,78762,78763 |
| Constipation | K5900,K5901,K5902,K5903,K5904,K5909,K591,K592,K593,K5931 K5939,K594,K598,K5981 ,K5989,K599 | 56400,56401,56402,56409,5645,5646,5647,56481,56489,5649 |
| Diarrhoea | R197 | 78791 |
| Gastric reflux | K210,K2100,K2101,K219 | 53011,53081 |
| Vomiting | R1110,R1111,R1112,R1113,R1114,R1115 | 07882,5362,56987,78703,78704 |
| Weight gain | R635 | 7831 |
| Pancreatic disorders (excluding diabetes) | K860,K861,K862,K863,K868,K8681,K8689,K869 | 5771,5772,5778,5779 |
| Weight loss | R634 | 78321 |
| Anhedonia | R4584 | 78099 |
| Anorexia | R630 | 7830 |
| Depression | F320,F321,F322,F323,F324,F325,F328,F3281,F3289,F329,F32A,F330 F331,F332,F333,F3340,F3341,F3342,F338,F339 | 29620,29621,29622,29623,29624,29625,29626,29630,29631,29632 29633,29634,29635,29636,29682,29699,2980,311 |
| Increased appetite | R632 | 7836 |
| Loneliness | Z602,Z604 | V603,V624 |
| Mood swings bipolar | F3010,F3011,F3012,F3013,F302,F303,F304,F308,F309,F310,F3110 F3111,F3112,F3113,F312,F3130,F3131,F3132,F314,F315,F3160 F3161,F3162,F3163,F3164,F3170,F3171,F3172,F3173,F3174,F3175 | 29600,29601,29602,29603,29604,29605,29606,29640,29641,29642 29643,29644,29645,29646,29650,29651,29652,29653,29654,29655 29656,29660,29661,29662,29663,29664,29665,29666,2967,29680 |

Continued on next page

**Supplementary Table S1:** (cont'd) A comprehensive mapping of diagnosis groups used in the experiments to their corresponding ICD-10 and ICD-9 codes.

| Diagnosis | ICD-10 | ICD-9 |
|---|---|---|
| | F3176,F3177,F3178,F3181,F3189,F319,F340,F341,F348,F3481,F3489<br>F349,F39 | 29681,29689,29690,29699,3004,30110,30112,30113 |
| Post-traumatic stress disorder | F4310,F4311,F4312 | 30981 |
| Skipped Meals | Z594,Z724 | V602,V691 |
| Amnesia memory problems | R411,R412,R413 | 78093 |
| Brain fog cognitive deficit | F0100EZ,F0100FZ,F0100UZ,F0100YZ,F0100ZZ,F0101ZZ,F0103ZZ<br>F0104ZZ,F0105YZ,F0105ZZ,F0106YZ,F0106ZZ,F010GZZ,F0110EZ<br>F0110FZ,F0110UZ,F0110YZ,F0110ZZ,F0111ZZ,F0113ZZ,F0114ZZ<br>F0115YZ,F0115ZZ,F0116YZ,F0116ZZ,F011GZZ,F0120EZ,F0120FZ<br>F0120UZ,F0120YZ,F0120ZZ,F0121ZZ,F0123ZZ,F0124ZZ,F0125YZ<br>F0125ZZ,F0126YZ,F0126ZZ,F012GZZ,F0130EZ,F0130FZ,F0130UZ<br>F0130YZ,F0130ZZ,F0131ZZ,F0133ZZ,F0134ZZ,F0135YZ,F0135ZZ<br>F0136YZ,F0136ZZ,F013GZZ,F0150,F0151,F01511,F01518,F0152,F0153<br>F0154,F01A0,F01A11,F01A18,F01A2,F01A3,F01A4,F01B0,F01B11<br>F01B18,F01B2,F01B3,F01B4,F01C0,F01C11,F01C18,F01C2,F01C3<br>F01C4,F01D0EZ,F01D0FZ,F01D0UZ,F01D0YZ,F01D0ZZ,F01D1ZZ,F01D5YZ<br>F01D5ZZ,F01D6YZ,F01D6ZZ,F01F0EZ,F01F0FZ,F01F0UZ,F01F0YZ<br>F01F0ZZ,F01F1ZZ,F01F5YZ,F01F5ZZ,F01F6YZ,F01F6ZZ,F01G0EZ<br>F01G0FZ,F01G0UZ,F01G0YZ,F01G0ZZ,F01G1ZZ,F01G5YZ,F01G5ZZ<br>F01G6YZ,F01G6ZZ,F01H0EZ,F01H0FZ,F01H0UZ,F01H0YZ,F01H0ZZ<br>F01H1ZZ,F01H5YZ,F01H5ZZ,F01H6YZ,F01H6ZZ,F01J0EZ,F01J0FZ<br>F01J0UZ,F01J0YZ,F01J0ZZ,F01J1ZZ,F01J5YZ,F01J5ZZ,F01J6YZ<br>F01J6ZZ,F01K0EZ,F01K0FZ,F01K0UZ,F01K0YZ,F01K0ZZ,F01K1ZZ<br>F01K5YZ,F01K5ZZ,F01K6YZ,F01K6ZZ,F01L0EZ,F01L0FZ,F01L0UZ<br>F01L0YZ,F01L0ZZ,F01L1ZZ,F01L5YZ,F01L5ZZ,F01L6YZ,F01L6ZZ<br>F01M0EZ,F01M0FZ,F01M0UZ,F01M0YZ,F01M0ZZ,F01M1ZZ,F01M5YZ<br>F01M5ZZ,F01M6YZ,F01M6ZZ,F01N0EZ,F01N0FZ,F01N0UZ,F01N0YZ<br>F01N0ZZ,F01Z2KZ,F01Z2MZ,F01Z2NZ,F01Z2PZ,F01Z2QZ,F01Z2SZ<br>F01Z2YZ,F01Z2ZZ,F01Z77Z,F01Z9JZ,F01ZBEZ,F01ZBFZ,F01ZBUZ<br>F01ZBZZ,F01ZCEZ,F01ZCFZ,F01ZCUZ,F01ZCZZ,F01ZDEZ,F01ZDFZ<br>F01ZDUZ,F01ZDYZ,F01ZDZZ,F01ZFEZ,F01ZFFZ,F01ZFUZ,F01ZFZZ<br>F0209YZ,F0209ZZ,F020DYZ,F020DZZ,F021DYZ,F021DZZ,F022DYZ<br>F022DZZ,F023DYZ,F023DZZ,F024GCZ,F024GGZ,F024GYZ,F024GZZ<br>F025GCZ,F025GGZ,F025GYZ,F025GZZ,F026GCZ,F026GGZ,F026GYZ | 2900,29010,29011,29013,29021,29040,29041,2908,2909,2930,2931<br>29410,29411,29420,29421,2948,3079,31501,3310,33111,33119<br>33182,33183,34830,36816,78001,78002,78003,78009,78460,78461<br>78469,8915,9301,9302,9304,9305,9306,9307,9309,NoDx |

Continued on next page

**Supplementary Table S1:** (cont'd) A comprehensive mapping of diagnosis groups used in the experiments to their corresponding ICD-10 and ICD-9 codes.

| Diagnosis | ICD-10 | ICD-9 |
|---|---|---|
| | F026GZZ,F0277EZ,F0277GZ,F0277UZ,F0277YZ,F0277ZZ,F027GCZ F027GGZ,F027GYZ,F027GZZ,F0280,F0281,F02811,F02818,F0282,F0283 F0284,F028GCZ,F028GGZ,F028GYZ,F028GZZ,F029GCZ,F029GGZ,F029GYZ F029GZZ,F02A0,F02A11,F02A18,F02A2,F02A3,F02A4,F02B0,F02B11 F02B18,F02B2,F02B3,F02B4,F02BGCZ,F02BGGZ,F02BGYZ,F02BGZZ F02C0,F02C11,F02C18,F02C2,F02C3,F02C4,F02C7EZ,F02C7GZ,F02C7UZ F02C7YZ,F02C7ZZ,F02CGCZ,F02CGGZ,F02CGYZ,F02CGZZ,F02Z0EZ F02Z0FZ,F02Z0UZ,F02Z0ZZ,F02Z1EZ,F02Z1FZ,F02Z1UZ,F02Z1ZZ F02Z2YZ,F02Z2ZZ,F02Z3EZ,F02Z3FZ,F02Z3UZ,F02Z3ZZ,F02Z4EZ F02Z4FZ,F02Z4UZ,F02Z4ZZ,F02Z5KZ,F02Z5MZ,F02Z5NZ,F02Z5PZ F02Z5QZ,F02Z5SZ,F02Z5YZ,F02Z5ZZ,F02Z6ZZ,F02Z8YZ,F02Z8ZZ F02ZBEZ,F02ZBFZ,F02ZBUZ,F02ZBYZ,F02ZBZZ,F02ZCEZ,F02ZCFZ F02ZCUZ,F02ZCYZ,F02ZCZZ,F02ZFYZ,F02ZFZZ,F02ZHEZ,F02ZHFZ F02ZHGZ,F02ZHUZ,F02ZHYZ,F02ZHZZ,F0390,F0391,F03911,F03918 F0392,F0393,F0394,F03A0,F03A11,F03A18,F03A2,F03A3,F03A4,F03B0 F03B11,F03B18,F03B2,F03B3,F03B4,F03C0,F03C11,F03C18,F03C2 F03C3,F03C4,F05,F068,G300,G301,G308,G309,G3101,G3109,G3183 G3184,G9340,R400,R401,R4020,R402110,R402111,R402112,R402113 R402114,R402120,R402121,R402122,R402123,R402124,R402130 R402131,R402132,R402133,R402134,R402140,R402141,R402142 R402143,R402144,R402210,R402211,R402212,R402213,R402214 R402220,R402221,R402222,R402223,R402224,R402230,R402231 R402232,R402233,R402234,R402240,R402241,R402242,R402243 R402244,R402250,R402251,R402252,R402253,R402254,R402310 R402311,R402312,R402313,R402314,R402320,R402321,R402322 R402323,R402324,R402330,R402331,R402332,R402333,R402334 R402340,R402341,R402342,R402343,R402344,R402350,R402351 R402352,R402353,R402354,R402360,R402361,R402362,R402363 R402364,R40241,R402410,R402411,R402412,R402413,R402414,R40242 R402420,R402421,R402422,R402423,R402424,R40243,R402430 R402431,R402432,R402433,R402434,R40244,R402440,R402441 R402442,R402443,R402444,R402A,R403,R404,R451,R480,R481,R482 R483,R488,R489 | |
| Dementia | F003GKZ,F003GMZ,F003GPZ,F003GYZ,F003GZZ,F00Z01Z,F00Z02Z F00Z0KZ,F00Z0ZZ,F00Z11Z,F00Z12Z,F00Z19Z,F00Z1KZ,F00Z1ZZ F00Z21Z,F00Z22Z,F00Z29Z,F00Z2KZ,F00Z2ZZ,F00Z31Z,F00Z32Z F00Z3KZ,F00Z3ZZ,F00Z41Z,F00Z42Z,F00Z4ZZ,F00Z51Z,F00Z52Z F00Z59Z,F00Z5KZ,F00Z6KZ,F00Z6MZ,F00Z6PZ,F00Z6YZ,F00Z6ZZ F00Z7KZ,F00Z7MZ,F00Z7PZ,F00Z7YZ,F00Z7ZZ,F00Z8KZ,F00Z8MZ | 2900,29010,29011,29013,29021,29040,29041,2908,2909,29410 29411,29420,29421,8915,9301,9302,9303,9304,9305,9306,9307 9309,9375 |

Continued on next page

**Supplementary Table S1:** (cont'd) A comprehensive mapping of diagnosis groups used in the experiments to their corresponding ICD-10 and ICD-9 codes.

| Diagnosis | ICD-10 | ICD-9 |
|---|---|---|
| | F00Z8PZ,F00Z8YZ,F00Z8ZZ,F00Z9KZ,F00Z9PZ,F00Z9QZ,F00Z9YZ | |
| | F00Z9ZZ,F00ZBKZ,F00ZBNZ,F00ZBPZ,F00ZBQZ,F00ZBTZ,F00ZBYZ | |
| | F00ZBZZ,F00ZCKZ,F00ZCMZ,F00ZCPZ,F00ZCYZ,F00ZCZZ,F00ZDKZ | |
| | F00ZDNZ,F00ZDPZ,F00ZDQZ,F00ZDSZ,F00ZDTZ,F00ZDYZ,F00ZDZZ | |
| | F00ZFKZ,F00ZFNZ,F00ZFPZ,F00ZFSZ,F00ZFTZ,F00ZFYZ,F00ZFZZ | |
| | F00ZGKZ,F00ZGMZ,F00ZGPZ,F00ZGYZ,F00ZGZZ,F00ZHYZ,F00ZHZZ | |
| | F00ZJTZ,F00ZJWZ,F00ZJYZ,F00ZKKZ,F00ZKPZ,F00ZKYZ,F00ZKZZ | |
| | F00ZLKZ,F00ZLMZ,F00ZLPZ,F00ZLYZ,F00ZLZZ,F00ZMKZ,F00ZMPZ | |
| | F00ZMSZ,F00ZMVZ,F00ZMYZ,F00ZMZZ,F00ZNNZ,F00ZNPZ,F00ZNQZ | |
| | F00ZNSZ,F00ZNTZ,F00ZNYZ,F00ZPYZ,F00ZPZZ,F00ZQ1Z,F00ZQ2Z | |
| | F00ZQKZ,F00ZQZZ,F00ZR1Z,F00ZR2Z,F00ZRKZ,F00ZRZZ,F00ZS1Z | |
| | F00ZS2Z,F00ZSKZ,F00ZSZZ,F00ZT1Z,F00ZT2Z,F00ZTKZ,F00ZTZZ | |
| | F00ZV1Z,F00ZV2Z,F00ZVKZ,F00ZVZZ,F00ZW1Z,F00ZW2Z,F00ZWKZ | |
| | F00ZWZZ,F00ZXZZ,F0100EZ,F0100FZ,F0100UZ,F0100YZ,F0100ZZ | |
| | F0101ZZ,F0103ZZ,F0104ZZ,F0105YZ,F0105ZZ,F0106YZ,F0106ZZ | |
| | F010GZZ,F0110EZ,F0110FZ,F0110UZ,F0110YZ,F0110ZZ,F0111ZZ | |
| | F0113ZZ,F0114ZZ,F0115YZ,F0115ZZ,F0116YZ,F0116ZZ,F011GZZ | |
| | F0120EZ,F0120FZ,F0120UZ,F0120YZ,F0120ZZ,F0121ZZ,F0123ZZ | |
| | F0124ZZ,F0125YZ,F0125ZZ,F0126YZ,F0126ZZ,F012GZZ,F0130EZ | |
| | F0130FZ,F0130UZ,F0130YZ,F0130ZZ,F0131ZZ,F0133ZZ,F0134ZZ | |
| | F0135YZ,F0135ZZ,F0136YZ,F0136ZZ,F013GZZ,F0150,F0151,F01511 | |
| | F01518,F0152,F0153,F0154,F01A0,F01A11,F01A18,F01A2,F01A3 | |
| | F01A4,F01B0,F01B11,F01B18,F01B2,F01B3,F01B4,F01C0,F01C11 | |
| | F01C18,F01C2,F01C3,F01C4,F01D0EZ,F01D0FZ,F01D0UZ,F01D0YZ | |
| | F01D0ZZ,F01D1ZZ,F01D5YZ,F01D5ZZ,F01D6YZ,F01D6ZZ,F01F0EZ | |
| | F01F0FZ,F01F0UZ,F01F0YZ,F01F0ZZ,F01F1ZZ,F01F5YZ,F01F5ZZ | |
| | F01F6YZ,F01F6ZZ,F01G0EZ,F01G0FZ,F01G0UZ,F01G0YZ,F01G0ZZ | |
| | F01G1ZZ,F01G5YZ,F01G5ZZ,F01G6YZ,F01G6ZZ,F01H0EZ,F01H0FZ | |
| | F01H0UZ,F01H0YZ,F01H0ZZ,F01H1ZZ,F01H5YZ,F01H5ZZ,F01H6YZ | |
| | F01H6ZZ,F01J0EZ,F01J0FZ,F01J0UZ,F01J0YZ,F01J0ZZ,F01J1ZZ | |
| | F01J5YZ,F01J5ZZ,F01J6YZ,F01J6ZZ,F01K0EZ,F01K0FZ,F01K0UZ | |
| | F01K0YZ,F01K0ZZ,F01K1ZZ,F01K5YZ,F01K5ZZ,F01K6YZ,F01K6ZZ | |
| | F01L0EZ,F01L0FZ,F01L0UZ,F01L0YZ,F01L0ZZ,F01L1ZZ,F01L5YZ | |
| | F01L5ZZ,F01L6YZ,F01L6ZZ,F01M0EZ,F01M0FZ,F01M0UZ,F01M0YZ | |
| | F01M0ZZ,F01M1ZZ,F01M5YZ,F01M5ZZ,F01M6YZ,F01M6ZZ,F01N0EZ | |
| | F01N0FZ,F01N0UZ,F01N0YZ,F01N0ZZ,F01Z2KZ,F01Z2MZ,F01Z2NZ | |
| | F01Z2PZ,F01Z2QZ,F01Z2SZ,F01Z2YZ,F01Z2ZZ,F01Z77Z,F01Z9JZ | |
| | F01ZBEZ,F01ZBFZ,F01ZBUZ,F01ZBZZ,F01ZCEZ,F01ZCFZ,F01ZCUZ | |
| | F01ZCZZ,F01ZDEZ,F01ZDFZ,F01ZDUZ,F01ZDYZ,F01ZDZZ,F01ZFEZ | |
| | F01ZFFZ,F01ZFUZ,F01ZFZZ,F0209YZ,F0209ZZ,F020DYZ,F020DZZ | |

Continued on next page

**Supplementary Table S1:** (cont'd) A comprehensive mapping of diagnosis groups used in the experiments to their corresponding ICD-10 and ICD-9 codes.

| Diagnosis | ICD-10 | ICD-9 |
|---|---|---|
| | F021DYZ,F021DZZ,F022DYZ,F022DZZ,F023DYZ,F023DZZ,F024GCZ<br>F024GGZ,F024GYZ,F024GZZ,F025GCZ,F025GGZ,F025GYZ,F025GZZ<br>F026GCZ,F026GGZ,F026GYZ,F026GZZ,F0277EZ,F0277GZ,F0277UZ<br>F0277YZ,F0277ZZ,F027GCZ,F027GGZ,F027GYZ,F027GZZ,F0280,F0281<br>F02811,F02818,F0282,F0283,F0284,F028GCZ,F028GGZ,F028GYZ<br>F028GZZ,F029GCZ,F029GGZ,F029GYZ,F029GZZ,F02A0,F02A11,F02A18<br>F02A2,F02A3,F02A4,F02B0,F02B11,F02B18,F02B2,F02B3,F02B4<br>F02BGCZ,F02BGGZ,F02BGYZ,F02BGZZ,F02C0,F02C11,F02C18,F02C2<br>F02C3,F02C4,F02C7EZ,F02C7GZ,F02C7UZ,F02C7YZ,F02C7ZZ,F02CGCZ<br>F02CGGZ,F02CGYZ,F02CGZZ,F02Z0EZ,F02Z0FZ,F02Z0UZ,F02Z0ZZ<br>F02Z1EZ,F02Z1FZ,F02Z1UZ,F02Z1ZZ,F02Z2YZ,F02Z2ZZ,F02Z3EZ<br>F02Z3FZ,F02Z3UZ,F02Z3ZZ,F02Z4EZ,F02Z4FZ,F02Z4UZ,F02Z4ZZ<br>F02Z5KZ,F02Z5MZ,F02Z5NZ,F02Z5PZ,F02Z5QZ,F02Z5SZ,F02Z5YZ<br>F02Z5ZZ,F02Z6ZZ,F02Z8YZ,F02Z8ZZ,F02ZBEZ,F02ZBFZ,F02ZBUZ<br>F02ZBYZ,F02ZBZZ,F02ZCEZ,F02ZCFZ,F02ZCUZ,F02ZCYZ,F02ZCZZ<br>F02ZFYZ,F02ZFZZ,F02ZHEZ,F02ZHFZ,F02ZHGZ,F02ZHUZ,F02ZHYZ<br>F02ZHZZ,F0390,F0391,F03911,F03918,F0392,F0393,F0394,F03A0<br>F03A11,F03A18,F03A2,F03A3,F03A4,F03B0,F03B11,F03B18,F03B2<br>F03B3,F03B4,F03C0,F03C11,F03C18,F03C2,F03C3,F03C4 | |
| Difficulty understanding | R41841 | 79952 |
| Dysarthria | R471 | 78451 |
| Dysphasia | R4702 | 78459 |
| Reading difficulty | F810 | 31500,31502 |
| Intracranial haemorrhage | I6000,I6001,I6002,I6010,I6011,I6012,I602,I6020,I6021,I6022<br>I6030,I6031,I6032,I604,I6050,I6051,I6052,I606,I607,I608,I609<br>I610,I611,I612,I613,I614,I615,I616,I618,I619,I6200,I6201<br>I6202,I6203,I621,I629 | 430,431,4320,4321,4329 |
| Delirium | F05 | 2930,2931 |
| Alzheimer's | G300,G301,G308,G309 | 3310 |
| Apraxia | R482 | 78469 |

Continued on next page

**Supplementary Table S1:** (cont'd) A comprehensive mapping of diagnosis groups used in the experiments to their corresponding ICD-10 and ICD-9 codes.

| Diagnosis | ICD-10 | ICD-9 |
|---|---|---|
| Balance difficulty | R2681 | 7812 |
| Tremors | R250,R251,R252,R253,R258,R259 | 72982,7810 |
| Heavy arms or legs | R29898 | 72989,78199 |
| Dystonia | G2401,G2402,G2409,G241,G242,G243,G244,G245,G248,G249 | 3336,33372,33379,33381,33382,33383,33385,33389 |
| Myoclonus | G253 | 3332 |
| Parkinson's | G20,G20A1,G20A2,G20B1,G20B2,G20C | 3320 |
| Excessive sleep | G4710,G4711,G4712,G4713,G4714,G4719 | 32710,32711,32712,32713,32714,32719,78054 |
| Insomnia | F510,G470 | |
| Somnolence | R400 | 78009 |
| Diplopia | H532 | 3682 |
| Dry eye | H0412 | |
| Eye pain | H5710,H5711,H5712,H5713 | 37991 |
| Flashing lights and floaters | H53001,H53002,H53003,H53009,H53011,H53012,H53013,H53019 H53021,H53022,H53023,H53029,H53031,H53032,H53033,H53039 H53041,H53042,H53043,H53049,H5310,H5311,H53121,H53122,H53123 H53129,H53131,H53132,H53133,H53139,H53141,H53142,H53143 H53149,H5315,H5316,H5319,H532,H5330,H5331,H5332,H5333,H5334 H5340,H53411,H53412,H53413,H53419,H53421,H53422,H53423,H53429 H53431,H53432,H53433,H53439,H53451,H53452,H53453,H53459 H53461,H53462,H53469,H5347,H53481,H53482,H53483,H53489,H5350 H5351,H5352,H5353,H5354,H5355,H5359,H5360,H5361,H5362,H5363 H5369,H5371,H5372,H538,H539 | 36800,36801,36802,36803,36810,36811,36812,36813,36814,36815 36816,3682,36830,36831,36832,36833,36834,36840,36841,36842 36843,36844,36845,36846,36847,36851,36852,36853,36854,36855 36859,36860,36861,36862,36863,36869,3688,3689 |
| Itchy eyes | H01111,H01112,H01113,H01114,H01115,H01116,H01119,H01121 H01122,H01123,H01124,H01125,H01126,H01129,H01131,H01132 H01133,H01134,H01135,H01136,H01139,H01141,H01142,H01143 H01144,H01145,H01146,H01149 | 37331,37332,37333,37334 |

Continued on next page

**Supplementary Table S1:** (cont'd) A comprehensive mapping of diagnosis groups used in the experiments to their corresponding ICD-10 and ICD-9 codes.

| Diagnosis | ICD-10 | ICD-9 |
|---|---|---|
| Photophobia | H5319 | 36815 |
| Red eye | H10011,H10012,H10013,H10019,H10021,H10022,H10023,H10029,H1010 H1011,H1012,H1013,H10211,H10212,H10213,H10219,H10221,H10222 H10223,H10229,H10231,H10232,H10233,H10239,H1030,H1031,H1032 H1033,H10401,H10402,H10403,H10409,H10411,H10412,H10413,H10419 H10421,H10422,H10423,H10429,H10431,H10432,H10433,H10439,H1044 H1045,H10501,H10502,H10503,H10509,H10511,H10512,H10513,H10519 H10521,H10522,H10523,H10529,H10531,H10532,H10533,H10539 H10811,H10812,H10813,H10819,H10821,H10822,H10823,H10829,H1089 H109 | 37200,37201,37202,37203,37204,37205,37206,37210,37211,37212 37213,37214,37220,37221,37222,37230,37234,37239 |
| Watery eyes | H04201,H04202,H04203,H04209,H04211,H04212,H04213,H04219 H04221,H04222,H04223,H04229 | 37520,37521,37522 |
| Neuro-ophthalmologic disorders | H4600,H4601,H4602,H4603,H4610,H4611,H4612,H4613,H462,H463 H468,H469,H47011,H47012,H47013,H47019,H47021,H47022,H47023 H47029,H47031,H47032,H47033,H47039,H47091,H47092,H47093 H47099,H4710,H4711,H4712,H4713,H47141,H47142,H47143,H47149 H4720,H47211,H47212,H47213,H47219,H4722,H47231,H47232,H47233 H47239,H47291,H47292,H47293,H47299,H47311,H47312,H47313 H47319,H47321,H47322,H47323,H47329,H47331,H47332,H47333 H47339,H47391,H47392,H47393,H47399,H4741,H4742,H4743,H4749 H47511,H47512,H47519,H47521,H47522,H47529,H47531,H47532 H47539,H47611,H47612,H47619,H47621,H47622,H47629,H47631 H47632,H47639,H47641,H47642,H47649,H479 | 37700,37701,37702,37703,37704,37710,37711,37712,37713,37714 37715,37716,37721,37722,37723,37724,37730,37731,37732,37733 37734,37739,37741,37742,37743,37749,37751,37752,37753,37754 37761,37762,37763,37771,37772,37773,37775,3779 |
| Ejaculation difficulty | N5311,N5312,N5313,N5314,N5319 | 60887,60889 |
| Menorrhagia | N920,N922,N924 | 6262,6263,6270 |
| Menstrual changes | N920,N921,N922,N923,N924,N925,N926 | 6262,6263,6264,6265,6266,6268,6269,6270 |
| Premenstrual syndrome | N943 | 6254 |
| Vaginal discharge | N898,N930,N931,N938,N939 | 6234,6235,6236,6238,6267,6268,6269 |
| Vaginal dryness | N952 | 6273 |

Continued on next page

**Supplementary Table S1:** (cont'd) A comprehensive mapping of diagnosis groups used in the experiments to their corresponding ICD-10 and ICD-9 codes.

| Diagnosis | ICD-10 | ICD-9 |
|---:|---:|---:|
| Anorgasm | F5231,F5232 | 30273,30274 |
| Reduced libido | R6882 | 79981 |
| Cardiogenic shock | R570,R571,R578,R579 | 78550,78551,78559 |
| Atrial Fibrillation | I48 | |
| Deep vein thrombosis | I82401,I82402,I82403,I82409,I82411,I82412,I82413,I82419<br>I82421,I82422,I82423,I82429,I82431,I82432,I82433,I82439<br>I82441,I82442,I82443,I82449,I82451,I82452,I82453,I82459<br>I82461,I82462,I82463,I82469,I82491,I82492,I82493,I82499<br>I824Y1,I824Y2,I824Y3,I824Y9,I824Z1,I824Z2,I824Z3,I824Z9<br>I82629 | 45340,45341,45342,45382 |
| Superficial vein thrombosis | I82619,I82811,I82812,I82813,I82819 | 4536,45381 |
| Pulmonary embolism | I2601,I2602,I2609,I2690,I2692,I2693,I2694,I2699 | 4150,41512,41513,41519 |
| Phlebitis; thrombophlebitis and thromboembolism | I8000,I8001,I8002,I8003,I8010,I8011,I8012,I8013,I80201,I80202<br>I80203,I80209,I80211,I80212,I80213,I80219,I80221,I80222<br>I80223,I80229,I80231,I80232,I80233,I80239,I80241,I80242<br>I80243,I80249,I80251,I80252,I80253,I80259,I80291,I80292<br>I80293,I80299,I803,I808,I809 | 4510,45111,45119,4512,45181,45189,4519 |
| TIA | G450,G451,G452,G453,G454,G458,G459 | 36234,4350,4351,4352,4353,4358,4359,4377 |
| Ischemic stroke | I6300,I63011,I63012,I63013,I63019,I6302,I63031,I63032,I63033<br>I63039,I6309,I6310,I63111,I63112,I63113,I63119,I6312,I63131<br>I63132,I63133,I63139,I6319,I6320,I63211,I63212,I63213,I63219<br>I6322,I63231,I63232,I63233,I63239,I6329,I6330,I63311,I63312<br>I63313,I63319,I63321,I63322,I63323,I63329,I63331,I63332<br>I63333,I63339,I63341,I63342,I63343,I63349,I6339,I6340,I63411<br>I63412,I63413,I63419,I63421,I63422,I63423,I63429,I63431<br>I63432,I63433,I63439,I63441,I63442,I63443,I63449,I6349,I6350<br>I63511,I63512,I63513,I63519,I63521,I63522,I63523,I63529<br>I63531,I63532,I63533,I63539,I63541,I63542,I63543,I63549,I6359<br>I636,I638,I6381,I6389,I639 | 43301,43311,43321,43331,43381,43391,43401,43411,43491 |

Continued on next page

**Supplementary Table S1:** (cont'd) A comprehensive mapping of diagnosis groups used in the experiments to their corresponding ICD-10 and ICD-9 codes.

| Diagnosis | ICD-10 | ICD-9 |
|---|---|---|
| Ischemic cardiomyopathy | I255 | 4148 |
| Hemorrhagic stroke | I610,I611,I612,I613,I614,I615,I616,I618,I619 | 431 |
| Cold extremities | R209 | 7820 |
| Limb swelling | R220,R221,R222,R2230,R2231,R2232,R2233,R2240,R2241,R2242 R2243,R229 | 7822,7842,7866 |
| Orthostatic hypotension | I951 | 4580 |
| Palpitations | R002 | 7851 |
| Presyncope | O14 | |
| Tachycardia | ROOO | |
| Bradycardia | R001 | 42781,42789 |
| Paresthesia (tingling limbs) | R202 | 7820 |
| Flushing | R232 | 78262 |
| Ventricular arrhythmia | I4901,I4902,I491,I492,I493,I4940,I4949,I495,I498,I499 | 4270,42741,42742,42760,42761,42769,42781,42789,4279 |
| Atrial flutter | I483,I484,I4892 | 42732 |
| Encounter for observation and examination for conditions ruled out (excludes infectious disease, neoplasm, mental disorders) | Z036,Z0371,Z0372,Z0373,Z0374,Z0375,Z0379,Z03810,Z03818,Z0389 | V7182,V7183,V7189,V8901,V8902,V8903,V8904,V8905,V8909 |
| Sexually transmitted | A64 | 0999 |

Continued on next page

**Supplementary Table S1:** (cont'd) A comprehensive mapping of diagnosis groups used in the experiments to their corresponding ICD-10 and ICD-9 codes.

| Diagnosis | ICD-10 | ICD-9 |
|---|---|---|
| infections (excluding<br>HIV and hepatitis) | | |
| Asthenia | R531 | 78079 |
| Joint pain | M2550,M2551,M25511,M25512,M25519,M25521,M25522,M25529,M25531<br>M25532,M25539,M25541,M25542,M25549,M25551,M25552,M25559<br>M25561,M25562,M25569,M25571,M25572,M25579,M2559 | 71940,71941,71942,71943,71945,71946,71947 |
| Joint stiffness | M2560,M25611,M25612,M25619,M25621,M25622,M25629,M25631,M25632<br>M25639,M25641,M25642,M25649,M25651,M25652,M25659,M25661<br>M25662,M25669,M25671,M25672,M25673,M25674,M25675,M25676,M2569 | 71950,71951,71952,71953,71954,71955,71956,71957 |
| Myalgia | M791 | 7291 |
| Myopathy | G720,G721,G722,G723,G7241,G7249,G7281,G7289,G729 | 3593,3594,35971,35979,35981,35989,3599 |
| Muscle cramping | R252 | 72982,7810 |
| Muscle twitch | M6283 | |
| Paraesthesia | R202 | 7820 |
| Myoneural junction<br>muscle disease | G7000,G7001,G701,G702,G7080,G7081,G7089,G709,G710,G7100,G7101<br>G7102,G7109,G7111,G7112,G7113,G7114,G7119,G712,G7120,G7121<br>G71220,G71228,G7129,G713,G718,G719,G720,G721,G722,G723,G7241<br>G7249,G7281,G7289,G729,G731,G733,G737 | 35800,35801,3581,3582,35830,35831,35839,3588,3589,3590,3591<br>35921,35922,35923,35924,35929,3593,3594,3595,35971,35979<br>35981,35989,3599 |
| Other and unspecified<br>arthropathy | M1200,M12011,M12012,M12019,M12021,M12022,M12029,M12031,M12032<br>M12039,M12041,M12042,M12049,M12051,M12052,M12059,M12061<br>M12062,M12069,M12071,M12072,M12079,M1208,M1209,M1210,M12111<br>M12112,M12119,M12121,M12122,M12129,M12131,M12132,M12139<br>M12141,M12142,M12149,M12151,M12152,M12159,M12161,M12162<br>M12169,M12171,M12172,M12179,M1218,M1219,M1220,M12211,M12212<br>M12219,M12221,M12222,M12229,M12231,M12232,M12239,M12241<br>M12242,M12249,M12251,M12252,M12259,M12261,M12262,M12269<br>M12271,M12272,M12279,M1228,M1229,M1230,M12311,M12312,M12319<br>M12321,M12322,M12329,M12331,M12332,M12339,M12341,M12342<br>M12349,M12351,M12352,M12359,M12361,M12362,M12369,M12371<br>M12372,M12379,M1238,M1239,M1240,M12411,M12412,M12419,M12421 | 7144,71600,71601,71602,71603,71604,71605,71606,71607,71608<br>71609,71610,71611,71612,71613,71614,71615,71616,71617,71618<br>71619,71640,71641,71642,71643,71644,71645,71646,71647,71648<br>71649,71680,71681,71682,71683,71684,71685,71686,71687,71688<br>71689,71690,71920,71921,71922,71923,71924,71925,71926,71927<br>71928,71929,71930,71931,71932,71933,71934,71935,71936,71937<br>71938,71939 |

Continued on next page

**Supplementary Table S1:** (cont'd) A comprehensive mapping of diagnosis groups used in the experiments to their corresponding ICD-10 and ICD-9 codes.

| Diagnosis | ICD-10 | ICD-9 |
|---|---|---|
| | M12422,M12429,M12431,M12432,M12439,M12441,M12442,M12449<br>M12451,M12452,M12459,M12461,M12462,M12469,M12471,M12472<br>M12479,M1248,M1249,M1250,M12511,M12512,M12519,M12521,M12522<br>M12529,M12531,M12532,M12539,M12541,M12542,M12549,M12551<br>M12552,M12559,M12561,M12562,M12569,M12571,M12572,M12579,M1258<br>M1259,M1280,M12811,M12812,M12819,M12821,M12822,M12829,M12831<br>M12832,M12839,M12841,M12842,M12849,M12851,M12852,M12859<br>M12861,M12862,M12869,M12871,M12872,M12879,M1288,M1289,M129 | |
| Spondylopathies spondyloarthropathy (including infective) | M4600,M4601,M4602,M4603,M4604,M4605,M4606,M4607,M4608,M4609<br>M461,M4620,M4621,M4622,M4623,M4624,M4625,M4626,M4627,M4628<br>M4630,M4631,M4632,M4633,M4634,M4635,M4636,M4637,M4638,M4639<br>M4640,M4641,M4642,M4643,M4644,M4645,M4646,M4647,M4648,M4649<br>M4650,M4651,M4652,M4653,M4654,M4655,M4656,M4657,M4658,M4659<br>M4680,M4681,M4682,M4683,M4684,M4685,M4686,M4687,M4688,M4689<br>M4690,M4691,M4692,M4693,M4694,M4695,M4696,M4697,M4698,M4699 | 7201,7202,72089,7209,72290,72291,72292,72293,73028,73098 |
| Acquired deformities (excluding foot) | M2100,M21021,M21022,M21029,M21051,M21052,M21059,M21061,M21062<br>M21069,M21071,M21072,M21079,M2110,M21121,M21122,M21129,M21151<br>M21152,M21159,M21161,M21162,M21169,M21171,M21172,M21179,M2120<br>M21211,M21212,M21219,M21221,M21222,M21229,M21231,M21232<br>M21239,M21241,M21242,M21249,M21251,M21252,M21259,M21261<br>M21262,M21269,M21271,M21272,M21279,M21331,M21332,M21339<br>M21511,M21512,M21519,M21521,M21522,M21529,M2170,M21721,M21722<br>M21729,M21731,M21732,M21733,M21734,M21739,M21751,M21752<br>M21759,M21761,M21762,M21763,M21764,M21769,M2180,M21821,M21822<br>M21829,M21831,M21832,M21839,M21851,M21852,M21859,M21861<br>M21862,M21869,M2190,M21921,M21922,M21929,M21931,M21932,M21939<br>M21941,M21942,M21949,M21951,M21952,M21959,M21961,M21962<br>M21969 | 73600,73601,73602,73605,73606,73607,73609,73630,73631,73632<br>73639,73641,73642,7366,73670,73671,73679,73681,73689,7369 |
| Crystal arthropathies (excluding gout) | M119 | 71290 |
| Breast cancer - ductal carcinoma in situ (DCIS) | D0500,D0501,D0502,D0510,D0511,D0512,D0580,D0581,D0582,D0590<br>D0591,D0592 | 2330 |
| Leukemia - acute lymphoblastic | C9100,C9101,C9102 | 20400,20401,20402 |

Continued on next page

**Supplementary Table S1:** (cont'd) A comprehensive mapping of diagnosis groups used in the experiments to their corresponding ICD-10 and ICD-9 codes.

| Diagnosis | ICD-10 | ICD-9 |
|---|---|---|
| leukemia (ALL) | | |
| Leukemia - acute myeloid leukemia (AML) | C9200 | 20500 |
| Leukemia - chronic lymphocytic leukemia (CLL) | C9110,C9111,C9112 | 20410,20411,20412 |
| Leukemia - chronic myeloid leukemia (CML) | C9210,C9211,C9212 | 20510,20511,20512 |
| Myelodysplastic syndrome (MDS) | D469 | 23875 |
| Transverse myelitis | G373 | 34120,34121 |
| Other specified hereditary and degenerative nervous system conditions | G3189 | 33189 |
| Paralysis (other than cerebral palsy) | G8100,G8101,G8102,G8103,G8104,G8110,G8111,G8112,G8113,G8114 G8190,G8191,G8192,G8193,G8194,G8220,G8221,G8222,G8250,G8251 G8252,G8253,G8254,G830,G8310,G8311,G8312,G8313,G8314,G8320 G8321,G8322,G8323,G8324,G8330,G8331,G8332,G8333,G8334,G834 G835,G8381,G8382,G8383,G8384,G8389,G839 | 34200,34201,34202,34210,34211,34212,34280,34281,34282,34290 34291,34292,34400,34401,34402,34403,34404,34409,3441,3442 34430,34431,34432,34440,34441,34442,3445,34460,34461,34481 34489,3449 |
| Other specified nervous system disorders | G890,G8911,G8912,G8918,G8921,G8922,G8928,G8929,G893,G894 G9001,G9009,G901,G902,G903,G904,G9050,G90511,G90512,G90513 G90519,G90521,G90522,G90523,G90529,G9059,G908,G909,G90A,G90B G910,G911,G912,G913,G914,G918,G919,G92,G9200,G9201,G9202 G9203,G9204,G9205,G928,G929,G930,G931,G932,G933,G9331,G9332 G9339,G9340,G9341,G9342,G9343,G9344,G9349,G935,G936,G937 G9381,G9382,G9389,G939,G94,G950,G9511,G9519,G9520,G9529,G9581 G9589,G959,G960,G9600,G9601,G9602,G9608 ,G9609,G9611,G9612 G9619,G96191,G96198,G968,G96810,G96811,G96819,G9689,G969,G970 G971,G972,G9731,G9732,G9741,G9748,G9749,G9751,G9752,G9761 | 32371,32372,3313,3314,3315,3317,33181,3330,3360,3361,3363 3368,3369,33700,33701,33709,3371,33720,33721,33722,33729,3373 3379,3380,33811,33812,33818,33821,33822,33828,33829,3383,3384 3480,3481,3482,34830,34831,34839,3484,3485,34881,34882,34889 3489,3490,3491,3492,34931,34939,34981,34982,34989,3499,38861 7428,78079,99701,99702,99709,99811,99812,9982 |

Continued on next page

**Supplementary Table S1:** (cont'd) A comprehensive mapping of diagnosis groups used in the experiments to their corresponding ICD-10 and ICD-9 codes.

| Diagnosis | ICD-10 | ICD-9 |
|---|---|---|
| | G9762,G9763,G9764,G9781,G9782,G9783,G9784,G980,G988,G990,G992<br>G998 | |
| Bells palsy | G510 | 3510 |
| Dysautonomia | G9001,G9009,G901,G902,G903,G904,G9050,G90511,G90512,G90513<br>G90519,G90521,G90522,G90523,G90529,G9059,G908,G909,G90A,G90B<br>G990,G998 | 3330,33700,33701,33709,3371,33720,33721,33722,33729,3373,3379<br>34989,7428 |
| Peripheral neuropathy | G629 | 3579 |
| Shortness of Breath | R0602 | 78605 |
| Orthopnoea | R0601 | 78602 |
| Paroxysmal nocturnal | D595 | 2832 |
| Dyspnoea | R0600 | 78609 |
| Shortness of breath | R0602 | 78605 |
| Tachypnoea | R0682 | 78606 |
| Wheezing | R062 | 78607 |
| Pneumonia (except that caused by tuberculosis) | J180,J181,J182,J188,J189 | 481,485,486,514 |
| Hypoxemia | R0902 | 79902 |
| Dry and scaly skin | L853 | 7068 |
| Hair loss | L659 | 70400 |
| Hives | L509 | 7089 |
| Itchy skin | L299 | 6989 |
| Nail changes | L609 | 7039 |

Continued on next page

**Supplementary Table S1:** (cont'd) A comprehensive mapping of diagnosis groups used in the experiments to their corresponding ICD-10 and ICD-9 codes.

| Diagnosis | ICD-10 | ICD-9 |
|---|---|---|
| Purpura | D690,D691,D692,D693,D6941,D6942,D6949,D6951,D6959,D696,D698<br>D699 | 2870,2871,2872,28731,28732,28733,28739,28741,28749,2875,2878<br>2879 |
| Rash | R21 | 7821 |
| Allergies | T7840XA | 9953 |
| Angioedema | T783 | |
| Body ache | R52 | 33819,78096 |
| Chills and shivering | R6883 | 78064 |
| Dizziness | R42 | 7804 |
| Dry mouth | R682 | 5277 |
| Haemoptysis | R042 | 78630 |
| Hallucinations | R443 | 7801 |
| Hot flashes | R232 | 78262 |
| Mouth ulcer | K1230 | 52800 |
| Neurasthenia | F480 | |
| Polydipsia | R631 | 7835 |
| Polyuria | R358,R3581,R3589 | 78842 |
| Epilepsy Seizures | G40001,G40009,G40011,G40019,G40101,G40109,G40111,G40119<br>G40201,G40209,G40211,G40219,G40301,G40309,G40311,G40319<br>G40401,G40409,G40411,G40419,G4042 ,G40501,G40509,G40801<br>G40802,G40803,G40804,G40811,G40812,G40813,G40814,G40821<br>G40822,G40823,G40824,G40833,G40834,G4089,G40901,G40909,G40911<br>G40919,G40A01,G40A09,G40A11,G40A19,G40B01,G40B09,G40B11<br>G40B19,G40C01,G40C09,G40C11,G40C19 | 34500,34501,34510,34511,3452,3453,34540,34541,34550,34551<br>34560,34561,34570,34571,34580,34581,34590,34591 |

Continued on next page

**Supplementary Table S1:** (cont'd) A comprehensive mapping of diagnosis groups used in the experiments to their corresponding ICD-10 and ICD-9 codes.

| Diagnosis | ICD-10 | ICD-9 |
|---|---|---|
| Sweating | R61 | 7808 |
| Swelling of lymph nodes | R599 | 7856 |
| Urinary retention | R339 | 78820 |
| Vertigo | H814 | |
| Guillain-Barre syndrome | G610 | 3570 |
| Nerve nerve root plexus disorders | G50,G51,G52,G53,G54,G55,G56,G57,G58,G59 | 3529,3538,3559 |
| Parkinsonism | G20,G21 | 3320 |
| Psychotic disorder | F20,F21,F22,F23,F24,F25,F26,F27,F28,F29 | 2970,2971,2972,2973,2983,2984,2988,2989,30122 |
| ADULT AND CHILD ABUSE NEGLECT AND OTHER MALTREATMENT CONFIRMED | T7401XA,T7401XD,T7401XS,T7402XA,T7402XD,T7402XS,T7411XA T7411XD,T7411XS,T7412XA,T7412XD,T7412XS,T7421XA,T7421XD T7421XS,T7422XA,T7422XD,T7422XS,T7431XA,T7431XD,T7431XS T7432XA,T7432XD,T7432XS,T744XXA,T744XXD,T744XXS,T7451XA T7451XD,T7451XS,T7452XA,T7452XD,T7452XS,T7461XA,T7461XD T7461XS,T7462XA,T7462XD,T7462XS,T7491XA,T7491XD,T7491XS T7492XA,T7492XD,T7492XS | 9099,99550,99551,99552,99553,99554,99555,99580,99581,99582 99583,99584,E9040,V5889 |
| LIGHTNING DROWNING SUBMERSION ELECTROCUTION VIBRATION | T7500XA,T7500XD,T7500XS,T7501XA,T7501XD,T7501XS,T7509XA T7509XD,T7509XS,T751XXA,T751XXD,T751XXS,T7520XA,T7520XD T7520XS,T7521XA,T7521XD,T7521XS,T7522XA,T7522XD,T7522XS T7523XA,T7523XD,T7523XS,T7529XA,T7529XD,T7529XS,T754XXA T754XXD,T754XXS | 9094,9940,9941,9948,9949,V5889 |
| OTHER AND UNSPECIFIED EFFECTS OF OTHER EXTERNAL CAUSES | T7500XA,T7500XD,T7500XS,T7501XA,T7501XD,T7501XS,T7509XA T7509XD,T7509XS,T751XXA,T751XXD,T751XXS,T7520XA,T7520XD T7520XS,T7521XA,T7521XD,T7521XS,T7522XA,T7522XD,T7522XS T7523XA,T7523XD,T7523XS,T7529XA,T7529XD,T7529XS,T753XXA T753XXD,T753XXS,T754XXA,T754XXD,T754XXS,T7581XA,T7581XD T7581XS,T7582XA,T7582XD,T7582XS,T7589XA,T7589XD,T7589XS | 9094,9940,9941,9946,9948,9949,E903,V5889 |

Continued on next page

**Supplementary Table S1:** (cont'd) A comprehensive mapping of diagnosis groups used in the experiments to their corresponding ICD-10 and ICD-9 codes.

| Diagnosis | ICD-10 | ICD-9 |
|---|---|---|
| ADULT AND CHILD ABUSE<br>NEGLECT AND OTHER<br>MALTREATMENT<br>SUSPECTED | T7601XA,T7601XD,T7601XS,T7602XA,T7602XD,T7602XS,T7611XA<br>T7611XD,T7611XS,T7612XA,T7612XD,T7612XS,T7621XA,T7621XD<br>T7621XS,T7622XA,T7622XD,T7622XS,T7631XA,T7631XD,T7631XS<br>T7632XA,T7632XD,T7632XS,T7651XA,T7651XD,T7651XS,T7652XA<br>T7652XD,T7652XS,T7661XA,T7661XD,T7661XS,T7662XA,T7662XD<br>T7662XS,T7691XA,T7691XD,T7691XS,T7692XA,T7692XD,T7692XS | 9099,99550,99551,99552,99553,99554,99580,99581,99582,99583<br>99584,V5889 |
| TOXIC EFFECT OF<br>CONTACT WITH VENOMOUS<br>ANIMALS AND PLANTS | T63001A,T63001D,T63001S,T63002A,T63002D,T63002S,T63003A<br>T63003D,T63003S,T63004A,T63004D,T63004S,T63011A,T63011D<br>T63011S,T63012A,T63012D,T63012S,T63013A,T63013D,T63013S<br>T63014A,T63014D,T63014S,T63021A,T63021D,T63021S,T63022A<br>T63022D,T63022S,T63023A,T63023D,T63023S,T63024A,T63024D<br>T63024S,T63031A,T63031D,T63031S,T63032A,T63032D,T63032S<br>T63033A,T63033D,T63033S,T63034A,T63034D,T63034S,T63041A<br>T63041D,T63041S,T63042A,T63042D,T63042S,T63043A,T63043D<br>T63043S,T63044A,T63044D,T63044S,T63061A,T63061D,T63061S<br>T63062A,T63062D,T63062S,T63063A,T63063D,T63063S,T63064A<br>T63064D,T63064S,T63071A,T63071D,T63071S,T63072A,T63072D<br>T63072S,T63073A,T63073D,T63073S,T63074A,T63074D,T63074S<br>T63081A,T63081D,T63081S,T63082A,T63082D,T63082S,T63083A<br>T63083D,T63083S,T63084A,T63084D,T63084S,T63091A,T63091D<br>T63091S,T63092A,T63092D,T63092S,T63093A,T63093D,T63093S<br>T63094A,T63094D,T63094S,T63111A,T63111D,T63111S,T63112A<br>T63112D,T63112S,T63113A,T63113D,T63113S,T63114A,T63114D<br>T63114S,T63121A,T63121D,T63121S,T63122A,T63122D,T63122S<br>T63123A,T63123D,T63123S,T63124A,T63124D,T63124S,T63191A<br>T63191D,T63191S,T63192A,T63192D,T63192S,T63193A,T63193D<br>T63193S,T63194A,T63194D,T63194S,T632X1A,T632X1D,T632X1S<br>T632X2A,T632X2D,T632X2S,T632X3A,T632X3D,T632X3S,T632X4A<br>T632X4D,T632X4S,T63301A,T63301D,T63301S,T63302A,T63302D<br>T63302S,T63303A,T63303D,T63303S,T63304A,T63304D,T63304S<br>T63311A,T63311D,T63311S,T63312A,T63312D,T63312S,T63313A<br>T63313D,T63313S,T63314A,T63314D,T63314S,T63321A,T63321D<br>T63321S,T63322A,T63322D,T63322S,T63323A,T63323D,T63323S<br>T63324A,T63324D,T63324S,T63331A,T63331D,T63331S,T63332A<br>T63332D,T63332S,T63333A,T63333D,T63333S,T63334A,T63334D<br>T63334S,T63391A,T63391D,T63391S,T63392A,T63392D,T63392S<br>T63393A,T63393D,T63393S,T63394A,T63394D,T63394S,T63411A<br>T63411D,T63411S,T63412A,T63412D,T63412S,T63413A,T63413D<br>T63413S,T63414A,T63414D,T63414S,T63421A,T63421D,T63421S | 9091,9895,E9050,E9051,E9052,E9053,E9054,E9055,E9056,E9292<br>E9509,E959,E9621,E969,E9809,E989,V5889 |

Continued on next page

**Supplementary Table S1:** (cont'd) A comprehensive mapping of diagnosis groups used in the experiments to their corresponding ICD-10 and ICD-9 codes.

| Diagnosis | ICD-10 | ICD-9 |
|---|---|---|
| | T63422A,T63422D,T63422S,T63423A,T63423D,T63423S,T63424A T63424D,T63424S,T63431A,T63431D,T63431S,T63432A,T63432D T63432S,T63433A,T63433D,T63433S,T63434A,T63434D,T63434S T63441A,T63441D,T63441S,T63442A,T63442D,T63442S,T63443A T63443D,T63443S,T63444A,T63444D,T63444S,T63451A,T63451D T63451S,T63452A,T63452D,T63452S,T63453A,T63453D,T63453S T63454A,T63454D,T63454S,T63461A,T63461D,T63461S,T63462A T63462D,T63462S,T63463A,T63463D,T63463S,T63464A,T63464D T63464S,T63481A,T63481D,T63481S,T63482A,T63482D,T63482S T63483A,T63483D,T63483S,T63484A,T63484D,T63484S,T63511A T63511D,T63511S,T63512A,T63512D,T63512S,T63513A,T63513D T63513S,T63514A,T63514D,T63514S,T63591A,T63591D,T63591S T63592A,T63592D,T63592S,T63593A,T63593D,T63593S,T63594A T63594D,T63594S,T63611A,T63611D,T63611S,T63612A,T63612D T63612S,T63613A,T63613D,T63613S,T63614A,T63614D,T63614S T63621A,T63621D,T63621S,T63622A,T63622D,T63622S,T63623A T63623D,T63623S,T63624A,T63624D,T63624S,T63631A,T63631D T63631S,T63632A,T63632D,T63632S,T63633A,T63633D,T63633S T63634A,T63634D,T63634S,T63691A,T63691D,T63691S,T63692A T63692D,T63692S,T63693A,T63693D,T63693S,T63694A,T63694D T63694S,T63711A,T63711D,T63711S,T63712A,T63712D,T63712S T63713A,T63713D,T63713S,T63714A,T63714D,T63714S,T63791A T63791D,T63791S,T63792A,T63792D,T63792S,T63793A,T63793D T63793S,T63794A,T63794D,T63794S,T63811A,T63811D,T63811S T63812A,T63812D,T63812S,T63813A,T63813D,T63813S,T63814A T63814D,T63814S,T63821A,T63821D,T63821S,T63822A,T63822D T63822S,T63823A,T63823D,T63823S,T63824A,T63824D,T63824S T63831A,T63831D,T63831S,T63832A,T63832D,T63832S,T63833A T63833D,T63833S,T63834A,T63834D,T63834S,T63891A,T63891D T63891S,T63892A,T63892D,T63892S,T63893A,T63893D,T63893S T63894A,T63894D,T63894S,T6391XA,T6391XD,T6391XS,T6392XA T6392XD,T6392XS,T6393XA,T6393XD,T6393XS,T6394XA,T6394XD T6394XS | |
| CONTACT STRIKE OR BITTEN BY DOG INITIAL ENCOUNTER | W540XXA,W540XXD,W540XXS,W541XXA,W541XXD,W541XXS,W548XXA W548XXD,W548XXS | E9060,E9068,E9295 |
| CONTACT STRIKE OR BITTEN BY RODENT | W5301XA,W5301XD,W5301XS,W5309XA,W5309XD,W5309XS,W5311XA W5311XD,W5311XS,W5319XA,W5319XD,W5319XS,W5321XA,W5321XD | E9061,E9063,E9068,E9295 |

Continued on next page

**Supplementary Table S1:** (cont'd) A comprehensive mapping of diagnosis groups used in the experiments to their corresponding ICD-10 and ICD-9 codes.

| Diagnosis | ICD-10 | ICD-9 |
|---|---|---|
| | W5321XS,W5329XA,W5329XD,W5329XS,W5381XA,W5381XD,W5381XS W5389XA,W5389XD,W5389XS | |
| CONTACT STRIKE OR BITTEN BY MARINE MAMMALS | W5601XA,W5601XD,W5601XS,W5602XA,W5602XD,W5602XS,W5609XA W5609XD,W5609XS,W5611XA,W5611XD,W5611XS,W5612XA,W5612XD W5612XS,W5619XA,W5619XD,W5619XS,W5621XA,W5621XD,W5621XS W5622XA,W5622XD,W5622XS,W5629XA,W5629XD,W5629XS,W5631XA W5631XD,W5631XS,W5632XA,W5632XD,W5632XS,W5639XA,W5639XD W5639XS,W5641XA,W5641XD,W5641XS,W5642XA,W5642XD,W5642XS W5649XA,W5649XD,W5649XS,W5651XA,W5651XD,W5651XS,W5652XA W5652XD,W5652XS,W5659XA,W5659XD,W5659XS,W5681XA,W5681XD W5681XS,W5682XA,W5682XD,W5682XS,W5689XA,W5689XD,W5689XS | E9063,E9068,E9295 |
| CONTACT STRIKE OR BITTEN BY OTHER MAMMALS EG CATS | W5501XA,W5501XD,W5501XS,W5503XA,W5503XD,W5503XS,W5509XA W5509XD,W5509XS,W5511XA,W5511XD,W5511XS,W5512XA,W5512XD W5512XS,W5519XA,W5519XD,W5519XS,W5521XA,W5521XD,W5521XS W5522XA,W5522XD,W5522XS,W5529XA,W5529XD,W5529XS,W5531XA W5531XD,W5531XS,W5532XA,W5532XD,W5532XS,W5539XA,W5539XD W5539XS,W5541XA,W5541XD,W5541XS,W5542XA,W5542XD,W5542XS W5549XA,W5549XD,W5549XS,W5551XA,W5551XD,W5551XS,W5552XA W5552XD,W5552XS,W5559XA,W5559XD,W5559XS,W5581XA,W5581XD W5581XS,W5582XA,W5582XD,W5582XS,W5589XA,W5589XD,W5589XS | E9063,E9068,E9295 |
| CONTACT STRIKE OR BITTEN BY WILD BIRD | W6101XA,W6101XD,W6101XS,W6102XA,W6102XD,W6102XS,W6109XA W6109XD,W6109XS,W6111XA,W6111XD,W6111XS,W6112XA,W6112XD W6112XS,W6119XA,W6119XD,W6119XS,W6121XA,W6121XD,W6121XS W6122XA,W6122XD,W6122XS,W6129XA,W6129XD,W6129XS,W6132XA W6132XD,W6132XS,W6133XA,W6133XD,W6133XS,W6139XA,W6139XD W6139XS,W6142XA,W6142XD,W6142XS,W6143XA,W6143XD,W6143XS W6149XA,W6149XD,W6149XS,W6151XA,W6151XD,W6151XS,W6152XA W6152XD,W6152XS,W6159XA,W6159XD,W6159XS,W6161XA,W6161XD W6161XS,W6162XA,W6162XD,W6162XS,W6169XA,W6169XD,W6169XS W6191XA,W6191XD,W6191XS,W6192XA,W6192XD,W6192XS,W6199XA W6199XD,W6199XS | E9063,E9068,E9295 |
| BITTEN OR STUNG BY NONVENOMOUS INSECT | W57XXXA,W57XXXD,W57XXXS | E9064,E9295 |
| CATACLYSM SUCH AS EXCESSIVE HEAT OR | X30XXXA,X30XXXD,X30XXXS,X31XXXA,X31XXXD,X31XXXS,X32XXXA X32XXXD,X32XXXS,X34XXXA,X34XXXD,X34XXXS,X35XXXA,X35XXXD | E9000,E9010,E9080,E9081,E9082,E9083,E9084,E9088,E9089,E9090 E9091,E9092,E9093,E9094,E9098,E9268,E9288,E9295,E9298 |

Continued on next page

**Supplementary Table S1:** (cont'd) A comprehensive mapping of diagnosis groups used in the experiments to their corresponding ICD-10 and ICD-9 codes.

| Diagnosis | ICD-10 | ICD-9 |
|---|---|---|
| COLD SUNLIGHT | X35XXXS,X360XXA,X360XXD,X360XXS,X361XXA,X361XXD,X361XXS | |
| EARTHQUAKE VOLCANO | X370XXA,X370XXD,X370XXS,X371XXA,X371XXD,X371XXS,X372XXA | |
| FLOOD | X372XXD,X372XXS,X373XXA,X373XXD,X373XXS,X3741XA,X3741XD | |
| | X3741XS,X3742XA,X3742XD,X3742XS,X3743XA,X3743XD,X3743XS | |
| | X378XXA,X378XXD,X378XXS,X379XXA,X379XXD,X379XXS,X38XXXA | |
| | X38XXXD,X38XXXS,X3901XA,X3901XD,X3901XS,X3908XA,X3908XD | |
| | X3908XS,X398XXA,X398XXD,X398XXS | |
| STRUCK BY THROWN | W200XXA,W200XXD,W200XXS,W201XXA,W201XXD,W201XXS,W208XXA | E916,E9170,E9173,E9174,E918,E9192,E9196,E9298 |
| PROJECTED OR FALLING | W208XXD,W208XXS,W2100XA,W2100XD,W2100XS,W2101XA,W2101XD | |
| OBJECT OR OTHER | W2101XS,W2102XA,W2102XD,W2102XS,W2103XA,W2103XD,W2103XS | |
| OBJECT | W2104XA,W2104XD,W2104XS,W2105XA,W2105XD,W2105XS,W2106XA | |
| | W2106XD,W2106XS,W2107XA,W2107XD,W2107XS,W2109XA,W2109XD | |
| | W2109XS,W2111XA,W2111XD,W2111XS,W2112XA,W2112XD,W2112XS | |
| | W2113XA,W2113XD,W2113XS,W2119XA,W2119XD,W2119XS,W21210A | |
| | W21210D,W21210S,W21211A,W21211D,W21211S,W21220A,W21220D | |
| | W21220S,W21221A,W21221D,W21221S,W2131XA,W2131XD,W2131XS | |
| | W2132XA,W2132XD,W2132XS,W2139XA,W2139XD,W2139XS,W214XXA | |
| | W214XXD,W214XXS,W2181XA,W2181XD,W2181XS,W2189XA,W2189XD | |
| | W2189XS,W219XXA,W219XXD,W219XXS,W2201XA,W2201XD,W2201XS | |
| | W2202XA,W2202XD,W2202XS,W2203XA,W2203XD,W2203XS,W22041A | |
| | W22041D,W22041S,W22042A,W22042D,W22042S,W2209XA,W2209XD | |
| | W2209XS,W2210XA,W2210XD,W2210XS,W2211XA,W2211XD,W2211XS | |
| | W2212XA,W2212XD,W2212XS,W2219XA,W2219XD,W2219XS,W228XXA | |
| | W228XXD,W228XXS,W230XXA,W230XXD,W230XXS,W231XXA,W231XXD | |
| | W231XXS,W232XXA,W232XXD,W232XXS,W240XXA,W240XXD,W240XXS | |
| | W241XXA,W241XXD,W241XXS | |
| CONTACT WITH OTHER | W25XXXA,W25XXXD,W25XXXS,W260XXA,W260XXD,W260XXS,W261XXA | E9190,E9191,E9193,E9194,E9195,E9197,E9198,E9199,E9200,E9201 |
| AND MACHINERY HAND | W261XXD,W261XXS,W262XXA,W262XXD,W262XXS,W268XXA,W268XXD | E9202,E9203,E9204,E9208,E9298 |
| TOOL OR GLASS | W268XXS,W269XXA,W269XXD,W269XXS,W270XXA,W270XXD,W270XXS | |
| | W271XXA,W271XXD,W271XXS,W272XXA,W272XXD,W272XXS,W273XXA | |
| | W273XXD,W273XXS,W274XXA,W274XXD,W274XXS,W275XXA,W275XXD | |
| | W275XXS,W278XXA,W278XXD,W278XXS,W28XXXA,W28XXXD,W28XXXS | |
| | W290XXA,W290XXD,W290XXS,W291XXA,W291XXD,W291XXS,W292XXA | |
| | W292XXD,W292XXS,W293XXA,W293XXD,W293XXS,W294XXA,W294XXD | |
| | W294XXS,W298XXA,W298XXD,W298XXS,W300XXA,W300XXD,W300XXS | |
| | W301XXA,W301XXD,W301XXS,W302XXA,W302XXD,W302XXS,W303XXA | |
| | W303XXD,W303XXS,W3081XA,W3081XD,W3081XS,W3089XA,W3089XD | |
| | W3089XS,W309XXA,W309XXD,W309XXS,W310XXA,W310XXD,W310XXS | |

Continued on next page

**Supplementary Table S1:** (cont'd) A comprehensive mapping of diagnosis groups used in the experiments to their corresponding ICD-10 and ICD-9 codes.

| Diagnosis | ICD-10 | ICD-9 |
|---|---|---|
| | W311XXA,W311XXD,W311XXS,W312XXA,W312XXD,W312XXS,W313XXA<br>W313XXD,W313XXS,W3181XA,W3181XD,W3181XS,W3182XA,W3182XD<br>W3182XS,W3183XA,W3183XD,W3183XS,W3189XA,W3189XD,W3189XS<br>W319XXA,W319XXD,W319XXS | |
| EXPLOSION OF PRESSURIZED DEVICE GAS CYLINDER BOILER | W35XXXA,W35XXXD,W35XXXS,W361XXA,W361XXD,W361XXS,W362XXA<br>W362XXD,W362XXS,W363XXA,W363XXD,W363XXS,W368XXA,W368XXD<br>W368XXS,W369XXA,W369XXD,W369XXS,W370XXA,W370XXD,W370XXS<br>W378XXA,W378XXD,W378XXS,W38XXXA,W38XXXD,W38XXXS | E9210,E9211,E9218,E9298 |
| EXPLOSION OF OTHER MATERIALS | W400XXA,W400XXD,W400XXS,W401XXA,W401XXD,W401XXS,W408XXA<br>W408XXD,W408XXS,W409XXA,W409XXD,W409XXS | E9231,E9232,E9238,E9239,E9298 |
| DISCHARGE OF FIREWORK | W39XXXA,W39XXXD,W39XXXS | E9230,E9298 |
| HIT STRUCK KICKED TWISTED BITTEN OR SCRATCHED BY ANOTHER PERSON | W500XXA,W500XXD,W500XXS,W501XXA,W501XXD,W501XXS,W502XXA<br>W502XXD,W502XXS,W503XXA,W503XXD,W503XXS,W504XXA,W504XXD<br>W504XXS | E9179,E9283,E9298 |
| ACCIDENTAL BUMPING INTO ANOTHE PERSON | W51XXXA,W51XXXD,W51XXXS | E9179,E9298 |
| CRUSHED PUSHED OR STEPPED ON BY CROWD | W52XXXA,W52XXXD,W52XXXS | E9171,E9298 |
| DISCHARGE FROM OTHER AND UNSPECIFIED FIREARMS | W320XXA,W320XXD,W320XXS,W321XXA,W321XXD,W321XXS,W3300XA<br>W3300XD,W3300XS,W3301XA,W3301XD,W3301XS,W3302XA,W3302XD<br>W3302XS,W3303XA,W3303XD,W3303XS,W3309XA,W3309XD,W3309XS<br>W3310XA,W3310XD,W3310XS,W3311XA,W3311XD,W3311XS,W3312XA<br>W3312XD,W3312XS,W3313XA,W3313XD,W3313XS,W3319XA,W3319XD<br>W3319XS,W3400XA,W3400XD,W3400XS,W34010A,W34010D,W34010S<br>W34011A,W34011D,W34011S,W34018A,W34018D,W34018S,W3409XA<br>W3409XD,W3409XS,W3410XA,W3410XD,W3410XS,W34110A,W34110D<br>W34110S,W34111A,W34111D,W34111S,W34118A,W34118D,W34118S<br>W3419XA,W3419XD,W3419XS | E9220,E9221,E9222,E9223,E9224,E9225,E9228,E9229,E9298 |
| FOOD POSIONING | A050,A051,A052,A053,A054,A055,A058,A059 | 0050,0051,0052,0054,00581,00589,0059 |
| BENIGN NEOPLASMS | D100,D101,D102,D1030,D1039,D104,D105,D106,D107,D109,D110,D117 | 2100,2101,2102,2103,2104,2105,2106,2107,2108,2109,2110,2111 |

Continued on next page

**Supplementary Table S1:** (cont'd) A comprehensive mapping of diagnosis groups used in the experiments to their corresponding ICD-10 and ICD-9 codes.

| Diagnosis | ICD-10 | ICD-9 |
|---|---|---|
| | D119,D120,D121,D122,D123,D124,D125,D126,D127,D128,D129,D130 D131,D132,D1330,D1339,D134,D135,D136,D137,D139,D1391,D1399 D140,D141,D142,D1430,D1431,D1432,D144,D150,D151,D152,D157 D159,D1600,D1601,D1602,D1610,D1611,D1612,D1620,D1621,D1622 D1630,D1631,D1632,D164,D165,D166,D167,D168,D169,D170,D171 D1720,D1721,D1722,D1723,D1724,D1730,D1739,D174,D175,D176 D1771,D1772,D1779,D179,D1800,D1801,D1802,D1803,D1809,D181 D190,D191,D197,D199,D200,D201,D210,D2110,D2111,D2112,D2120 D2121,D2122,D213,D214,D215,D216,D219,D220,D2210,D2211,D22111 D22112,D2212,D22121,D22122,D2220,D2221,D2222,D2230,D2239,D224 D225,D2260,D2261,D2262,D2270,D2271,D2272,D229,D230,D2310 D2311,D23111,D23112,D2312,D23121,D23122,D2320,D2321,D2322 D2330,D2339,D234,D235,D2360,D2361,D2362,D2370,D2371,D2372 D239,D241,D242,D249,D250,D251,D252,D259,D260,D261,D267,D269 D270,D271,D279,D280,D281,D282,D287,D289,D290,D291,D2920,D2921 D2922,D2930,D2931,D2932,D294,D298,D299,D3000,D3001,D3002 D3010,D3011,D3012,D3020,D3021,D3022,D303,D304,D308,D309,D3100 D3101,D3102,D3110,D3111,D3112,D3120,D3121,D3122,D3130,D3131 D3132,D3140,D3141,D3142,D3150,D3151,D3152,D3160,D3161,D3162 D3190,D3191,D3192,D320,D321,D329,D330,D331,D332,D333,D334 D337,D339,D34,D3500,D3501,D3502,D351,D352,D353,D354,D355,D356 D357,D359,D360,D3610,D3611,D3612,D3613,D3614,D3615,D3616 D3617,D367,D369 | 2112,2113,2114,2115,2116,2117,2118,2119,2120,2121,2122,2123 2124,2125,2126,2127,2128,2129,2130,2131,2132,2133,2134,2135 2136,2137,2138,2139,2140,2141,2142,2143,2144,2148,2149,2150 2152,2153,2154,2155,2156,2157,2159,2160,2161,2162,2163,2164 2165,2166,2167,2168,2169,217,2180,2181,2182,2189,2190,2191 2198,2199,220,2210,2211,2212,2218,2219,2220,2221,2222,2223 2224,2228,2229,2230,2231,2232,2233,22381,22389,2239,2240,2241 2242,2243,2244,2245,2246,2247,2248,2249,2250,2251,2252,2253 2254,2258,2259,226,2270,2271,2273,2274,2275,2276,2278,2279 22800,22801,22802,22804,22809,2281,2290,2298,2299 |
| SCARRING | L905 | 7092 |
| FITTING OR ADJUSTMENT OF ORTHODONTIC OR DENTAL PROSTHETIC | Z463,Z464 | V523,V534,V585 |
| FITTING OR ADJUSTMENT OF HEARING AID | Z461 | V532 |
| FITTING OR ADJUSTMENT OF CASTS OR BANDAGES | Z4689,Z4801 | V537,V538,V5399,V5831 |
| Encounter for supervision of normal pregnancy | Z3400,Z3401,Z3402,Z3403,Z3480,Z3481,Z3482,Z3483,Z3490,Z3491 Z3492,Z3493 | V220,V221 |

Continued on next page

**Supplementary Table S1:** (cont'd) A comprehensive mapping of diagnosis groups used in the experiments to their corresponding ICD-10 and ICD-9 codes.

| Diagnosis | ICD-10 | ICD-9 |
|---|---|---|
| Encounter for supervision of normal first pregnancy TRIMESTER unspecified | Z3400 | V220 |
| Encounter for supervision of normal first pregnancy TRIMESTER 1 | Z3401 | V220 |
| Encounter for supervision of normal first pregnancy TRIMESTER 2 | Z3402 | V220 |
| Encounter for supervision of normal first pregnancy TRIMESTER 3 | Z3403 | V220 |
| Encounter for supervision of other normal pregnancy TRIMESTER unspecified | Z3480 | V221 |
| Encounter for supervision of other normal pregnancy TRIMESTER 1 | Z3481 | V221 |
| Encounter for supervision of other normal pregnancy TRIMESTER 2 | Z3482 | V221 |
| Encounter for supervision of other normal pregnancy TRIMESTER 3 | Z3483 | V221 |

Continued on next page

**Supplementary Table S1:** (cont'd) A comprehensive mapping of diagnosis groups used in the experiments to their corresponding ICD-10 and ICD-9 codes.

| Diagnosis | ICD-10 | ICD-9 |
|---|---|---|
| Encounter for supervision of normal pregnancy unspecified TRIMESTER unspecified | Z3490 | V221 |
| Encounter for supervision of normal pregnancy unspecified TRIMESTER 1 | Z3491 | V221 |
| Encounter for supervision of normal pregnancy unspecified TRIMESTER 2 | Z3492 | V221 |
| Encounter for supervision of normal pregnancy unspecified TRIMESTER 3 | Z3493 | V221 |
